# Comparative Safety of Second-Line Antihyperglycemic Agents in Older Adults with Type 2 Diabetes: A Multinational Real-World Evidence From LEGEND-T2DM

**DOI:** 10.1101/2025.05.08.25327248

**Authors:** Chungsoo Kim, Fan Bu, Clair Blacketer, Anna Ostropolets, Talita Duarte-Salles, Benjamin Viernes, Thomas Falconer, Andrea Pistillo, Jing Li, Can Yin, Mui Van Zandt, Paul Nagy, Akihiko Nishimura, Evan Minty, Seng Chan You, Mitsuaki Sawano, Shoko Sawano, Ja Young Jeon, Arya Aminorroaya, Lovedeep S. Dhingra, Aline F. Pedroso, Phyllis Thangraraj, David A. Dorr, Nicole Pratt, Kenneth K.C. Man, Wallis C.Y. Lau, Daniel R. Morales, Rohan Khera, Martijn Schuemie, Patrick B. Ryan, George Hripcsak, Harlan M. Krumholz, Marc A. Suchard, Yuan Lu

## Abstract

**Background:** As prescribing of newer antihyperglycemic agents expands, there remains limited comparative safety data for older adults—a population particularly vulnerable to adverse drug events and underrepresented in clinical trials. We aimed to evaluate the real-world safety of second-line antihyperglycemic agents among older adults with type 2 diabetes.

**Methods:** We conducted a multinational cohort study using nine harmonized electronic health record and claims databases from the U.S. and Europe, applying a consistent analytical framework based on the LEGEND-T2DM initiative. Among adults aged ≥65 years who initiated a second-line agent after metformin monotherapy, we compared safety outcomes across four drug classes: GLP-1 receptor agonists (GLP1RAs), SGLT2 inhibitors (SGLT2Is), DPP-4 inhibitors (DPP4Is), and sulfonylureas (SUs). We used propensity score adjustment, empirical calibration, and prespecified diagnostics to estimate hazard ratios (HRs) for 18 safety outcomes.

**Results:** In a cohort of 1.8 million older adults, both GLP1RAs and SGLT2Is were linked to significantly lower risks of hypoglycemia (HR 0.21 [95% CI, 0.16–0.27] for GLP1RA vs SU; HR 0.21 [0.13– 0.33] for SGLT2I vs SU) and hyperkalemia (HR 0.63 [0.50–0.81] for GLP1RA vs SU; HR 0.75 [0.63–0.90] for SGLT2I vs SU) and peripheral edema (HR 0.81 [0.71–0.92] for GLP1RAs vs. DPP4Is; HR 0.62 [0.46–0.84] for SGLT2Is vs. SU). However, SGLT2Is were associated with a higher risk of diabetic ketoacidosis compared to both GLP1RAs (HR 2.03 [1.38–2.99]) and SUs (HR 1.64 [1.27–2.11]). GLP1RAs had significantly higher risks of nausea (HR 0.63 [0.55–0.72]) and vomiting (HR 0.63 [0.57–0.69]) relative to SGLT2Is. Results were consistent across both on-treatment and intent-to-treat sensitivity analyses.

**Conclusion:** In older adults with type 2 diabetes, GLP1RAs and SGLT2Is demonstrated more favorable safety profiles than SUs and DPP4Is across multiple clinically relevant outcomes. These results support more informed, safety-conscious prescribing in a population underrepresented in clinical trials yet highly susceptible to adverse effects.

## INTRODUCTION

As the global population ages, the burden of type 2 diabetes mellitus among older adults continues to rise. Second-line antihyperglycemic agents play a central role in managing hyperglycemia and preventing complications once metformin monotherapy is insufficient. In recent years, glucagon-like peptide-1 receptor agonists (GLP1RAs) and sodium-glucose cotransporter-2 inhibitors (SGLT2Is) have gained substantial attention for their cardiovascular and renal benefits, prompting guideline endorsements for broader use in high-risk patients.^1,2^ However, despite increasing uptake, a critical knowledge gap persists: the comparative safety of these agents—particularly in older adults—remains insufficiently understood.

Older adults are uniquely vulnerable to adverse drug events due to multimorbidity, frailty, and polypharmacy.^1,3^ Yet, they remain underrepresented in randomized trials. The small sample sizes and selective populations of these trials limit their ability to detect rare, severe, or population-specific safety events—leaving clinicians with limited guidance on how to weigh safety considerations when initiating second-line therapy in real-world settings. Furthermore, much of the existing evidence is fragmented or focused on single outcomes, rather than offering a comprehensive assessment of safety across drug classes.^4–6^

The need to address this gap is urgent. As prescribing patterns shift toward newer agents such as GLP1RA and the older adult population increases,^7^ clinicians are making treatment decisions with incomplete safety data for a population at highest risk. Without robust, comparative evidence, efforts to personalize diabetes care and minimize avoidable harm fall short. To address this unmet need, we leveraged the LEGEND-T2DM initiative^8–10^—a multinational, large-scale, real-world study using harmonized data from nine electronic health record and claims databases—to evaluate the comparative safety of four major second-line drug classes: GLP1RAs, SGLT2Is, dipeptidyl peptidase-4 inhibitors (DPP4Is), and sulfonylureas (SUs). Using rigorous methods including large-scale propensity score adjustment, empirical calibration, and pre-specified diagnostics, we systematically assessed a broad range of clinically relevant safety outcomes. This study offers one of the most comprehensive safety evaluations to date of second-line antihyperglycemic agents in older adults. The findings aim to inform safer prescribing, reduce avoidable harms, and support evidence-based, patient-centered care in a population often left out of trials but central to clinical practice.

## METHODS

### Data source and study design

This study is part of the LEGEND-T2DM initiative, led by the Observational Health Data Sciences and Informatics (OHDSI) research collaborative. We conducted a retrospective observational cohort study using international databases standardized to the Common Data Model (CDM). A total of nine databases from the United States, Germany, France, and Spain were included, all mapped to version 5.3.1 of the Observational Medical Outcomes Partnership (OMOP) CDM.^11^ These comprised five U.S. insurance claims databases and several additional databases containing electronic health records (EHRs). All data were de-identified, and informed patient consent was waived. Ethical approvals for data use were obtained as required. Potential overlap of patients across some de-identified U.S. data sources was not explicitly addressed. Basic information, including data coverage periods and contributing entities for each source, is presented in Supplementary Table 1.

Detailed definitions of the study population, exposures, outcomes, follow-up strategies, and statistical methods were prespecified, registered, and published prior to study execution.^12,13^ The study utilized OHDSI’s Health Analytics Data-to-Evidence Suite, a suite of open-source tools for epidemiologic research. All study results were fully disclosed to support transparency and open science. The study followed the LEGEND principles and was reported in accordance with STROBE guidelines.^9,14^

### Study population and exposure

The study population included older adults (aged ≥65 years) with a diagnosis of T2DM who initiated a second-line antihyperglycemic agent following first-line metformin monotherapy (Figure 1). Second-line agents were defined as GLP1RAs, SGLT2Is, DPP4Is, and SUs.

**Figure 1.**
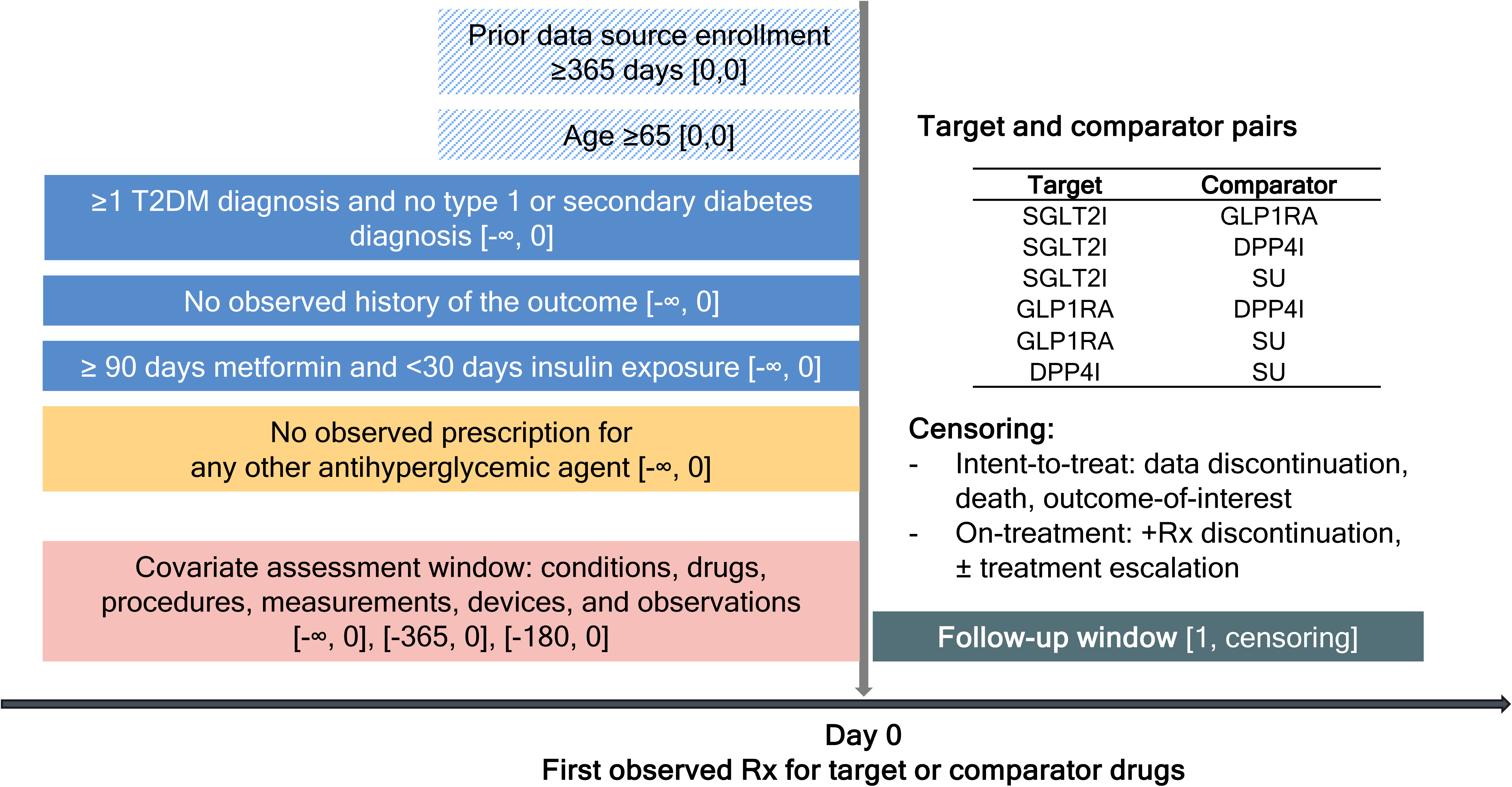
Study design scheme

We employed a new-user, active-comparator design, with the index date defined as the date of the first prescription for a second-line agent. Patients were excluded if they met any of the following criteria: (1) less than one year of continuous observation prior to the index date; (2) a diagnosis of type 1 diabetes or secondary diabetes mellitus; (3) prior use of antihyperglycemic agents other than the specified second-line agents; or (4) fewer than 90 days of metformin monotherapy or evidence of long-term insulin use (defined as ≥30 days of continuous use).

### Outcome and follow-up

All safety outcomes were selected based on the 2018 American Diabetes Association guidelines and prior randomized controlled trials.^15^ We evaluated eighteen outcomes, grouped into three categories: (1) *Metabolic and endocrine complications* (abnormal weight gain, abnormal weight loss, diabetic ketoacidosis, hyperkalemia, hypoglycemia, and hypotension); (2) *Organ system complications* (acute pancreatitis, nausea, vomiting, diarrhea, bone fracture, joint pain, lower extremity amputation, peripheral edema, genitourinary infection, and photosensitivity); and (3) *Systemic complications* (all-cause mortality and venous thromboembolism). Outcome definitions were based on prior work and have been previously implemented and validated.^16^ For each outcome, individuals with a prior history of the event were excluded. Follow-up was assessed using three approaches: (1) on-treatment regardless of treatment escalation (OT1), (2) on-treatment with censoring upon treatment escalation (OT2), and (3) intent-to-treat (ITT). In the OT1 approach, patients were followed from the day after the index date until the earliest occurrence of treatment discontinuation, outcome event, or end of observation. Treatment was considered continuous if a new prescription was recorded within 30 days of the previous one. Discontinuation was defined as the absence of a new prescription following the end of the prior prescription. The OT2 approach followed the same rules as OT1, but additionally censored follow-up upon initiation of any new antihyperglycemic agent. In contrast, the ITT approach followed patients from the index date until the end of observation or outcome occurrence, regardless of treatment discontinuation or escalation.

### Statistical analysis

Large-scale propensity scores (PS) were estimated using L1-penalized logistic regression for each treatment comparison within each database, incorporating all available covariates across clinical data domains (including demographics, conditions, drugs, procedures, measurements, devices, and observations) over multiple time windows prior to treatment initiation (index date, 6 months prior, 1 year prior, and any time prior).^17^

Confounding was addressed using PS stratification into 10 strata and variable-ratio matching for each pairwise comparison between second-line agents. To enhance generalizability, we prioritized reporting results based on PS stratification. Hazard ratios (HRs) for each outcome were estimated using Cox proportional hazards models within each database. Empirical calibration of HRs was conducted using negative control outcomes to correct for residual systematic error and to maintain nominal statistical properties despite the observational study design.^18^

In total, we conducted 38,232 analyses, based on 9 databases × 6 comparison pairs × 118 outcomes (18 safety outcomes plus negative controls) × 2 PS methods × 3 follow-up approaches. Statistical significance was defined as a two-sided p-value <0.05. In addition, we applied Bonferroni correction for multiple testing in the meta-analytic results (Threshold with p < 0.00277778 from 18 outcomes hypotheses for each comparison), and both unadjusted and adjusted values are reported.

Prior to evidence synthesis, we applied prespecified study diagnostics to ensure reliability. First, to confirm empirical equipoise, at least 25% of patients in each comparison within each database were required to have preference scores between 0.3 and 0.7.^19^ Second, all covariates were required to have a maximum standardized mean difference (SMD) <0.15 following PS adjustment, indicating adequate balance. Third, we required a minimum detectable relative risk (MDRR) <4.0 and a minimum sample size of ≥1,000 patients per treatment group to ensure sufficient statistical power.^20^ Only comparisons and databases that met all diagnostic criteria were included in the meta-analyses.

Pooled HR estimates were obtained using a random-effects meta-analysis of the empirically calibrated database-specific HRs. All analyses were performed in a controlled computing environment using OHDSI and other R packages (version 4.2.3).^21^ The full analytic code and results are publicly available at: https://github.com/ohdsi-studies/LegendT2dm, https://data.ohdsi.org/LegendT2dmClassEvidenceExplorer/.

## RESULTS

### Characteristics

A total of 1,808,003 adults aged 65 years or older met the eligibility criteria across the nine databases. There were more male patients at 984,590 (54.5%) than female patients, and the frequent age group was 65–74 years at 1,211,729 (67.0%). Among these, 73,603 (4.1%) patients initiated a GLP1RA, 173,465 (9.6%) initiated a SGLT2I, 485,016 (26.8%) initiated a DPP4I, and 1,075,919 (59.5%) received a SU as second-line therapy. The most commonly prescribed agents included dulaglutide (45.4%), semaglutide (36.1%), and exenatide (17.8%) among GLP1RAs; empagliflozin (58.6%), dapagliflozin (22.0%), and canagliflozin (18.9%) among SGLT2Is; sitagliptin (84.3%) among DPP4Is; and glipizide (53.9%) and glimepiride (35.3%) among SUs.

In the on-treatment follow-up, the median days of follow-up periods ranged from 55 to 202 days for GLP1RAs and 59 to 246 days for SGLT2Is, with longer follow-up durations observed for DPP4Is (86–719 days) and SUs (84–741 days) (Table 1). Table 2 presents baseline characteristics for the GLP1RA and SGLT2I groups in the US Open Claims database before and after propensity score stratification. Detailed baseline characteristics for all comparisons and databases, both before and after adjustment, are provided in Supplementary Tables 2–36.

**Table 1.** Population size and follow-up time for each drug class within each data source. We report the total patient counts and median and interquartile (IQR) times. When executing comparative studies, we included only within-database populations with > 1,000 new users per treatment-arm.

|  | Patients | On-treatment<br>time (in days) |  | Total follow-up<br>time (in days) |  |
| --- | --- | --- | --- | --- | --- |
|  |  | Median | IQR | Median | IQR |
| **SGLT2I** |  |  |  |  |  |
| DAG | 2,727 | 157 | (97 - 372) | 598 | (293 - 1,155) |
| MDCR | 3,511 | 121 | (38 - 344) | 419 | (156 - 880) |
| ODOD | 11,619 | 114 | (55 - 273) | 390 | (146 - 799) |
| OEHR | 7,828 | 59 | (29 - 99) | 596 | (295 - 1,081) |
| SIDIAP | 3,746 | 246 | (84 - 617) | 498 | (155 - 875) |
| USOC | 135,495 | 125 | (59 - 309) | 681 | (293 - 1,342) |
| VA | 8,539 | 144 | (71 - 306) | 283 | (124 - 592) |
| **Total:** | **173,465** |  |  |  |  |
| **GLP1RA** |  |  |  |  |  |
| MDCR | 2,135 | 108 | (36 - 300) | 513 | (159 - 1,129) |
| ODOD | 5,627 | 95 | (40 - 219) | 345 | (124 - 728) |
| OEHR | 3,573 | 55 | (29 - 102) | 527 | (273 - 983) |
| USOC | 62,268 | 105 | (50 - 258) | 585 | (241 - 1,130) |
| **Total:** | **73,603** |  |  |  |  |
| **DPP4I** |  |  |  |  |  |
| CCAE | 2,089 | 86 | (29 - 144) | 110 | (56 - 180) |
| DAG | 12,158 | 219 | (97 - 509) | 1,174 | (564 - 2,065) |
| MDCD | 1,210 | 130 | (56 - 330) | 834 | (357 - 1,539) |
| MDCR | 17,961 | 191 | (87 - 491) | 815 | (333 - 1,533) |
| ODOD | 27,244 | 157 | (63 - 375) | 832 | (342 - 1,581) |
| OEHR | 23,345 | 90 | (89 - 194) | 1,076 | (537 - 1,827) |
| SIDIAP | 26,465 | 719 | (200 - 1,464) | 1,207 | (567 - 2,137) |
| USOC | 358,700 | 158 | (59 - 416) | 1,575 | (806 - 2,430) |
| VA | 15,844 | 235 | (90 - 523) | 756 | (348 - 1,337) |
| **Total:** | **485,016** |  |  |  |  |
| **SU** |  |  |  |  |  |
| CCAE | 3,916 | 84 | (29 - 146) | 113 | (57 - 189) |
| DAG | 4,149 | 211 | (119 - 467) | 2,043 | (1,003 - 3,325) |
| MDCD | 2,488 | 126 | (48 - 311) | 1,020 | (464 - 1,874) |
| MDCR | 43,297 | 186 | (72 - 496) | 890 | (354 - 1,780) |
| ODOD | 66,309 | 193 | (89 - 504) | 882 | (352 - 1,709) |
| OEHR | 58,387 | 102 | (89 - 231) | 1,219 | (587 - 2,042) |
| SIDIAP | 8,667 | 741 | (188 - 1,615) | 2,879 | (1,674 - 3,808) |
| USOC | 741,462 | 203 | (89 - 547) | 1,498 | (717 - 2,419) |
| VA | 147,244 | 224 | (90 - 544) | 2,071 | (1,043 - 3,330) |
| **Total:** | **1,075,919** |  |  |  |  |

Abbreviations: IQR: interquartile range; DPP4I: dipeptidyl peptidase inhibitor 4; GLP1RA: glucagon-like peptide-1 receptor agonist; SGLT2I: sodium-glucose cotransporter 2 inhibitor; SU: sulfonylurea.

**Table 2.** Baseline patient characteristics for SGLT2I (T) and GLP1RA (C) new-users in the USOC data source. We report the proportion of initiators satisfying selected baseline characteristics and the standardized difference of population proportions (StdDiff) before and after propensity score adjustment. Less extreme StdDiffs through matching and stratification suggest improved balance between patient cohorts through adjustment.

| Characteristic | Before adjustment |  |  | After matching |  |  | After stratification |  |  |
| --- | --- | --- | --- | --- | --- | --- | --- | --- | --- |
|  | T (%) | C (%) | StdDiff | T (%) | C (%) | StdDiff | T (%) | C (%) | StdDiff |
| **Age group** |  |  |  |  |  |  |  |  |  |
| 65 - 69 | 43.1 | 49.1 | 0.12 | 44.9 | 44.9 | 0.00 | 44.9 | 44.9 | 0.00 |
| 70 - 74 | 29.2 | 29.8 | 0.01 | 30.1 | 29.9 | 0.00 | 29.5 | 29.4 | 0.00 |
| 75 - 79 | 17.5 | 14.2 | -0.09 | 14.7 | 14.8 | 0.00 | 16.4 | 16.6 | 0.00 |
| 80 - 84 | 9.5 | 6.4 | -0.12 | 6.7 | 6.8 | 0.00 | 8.5 | 8.6 | 0.00 |
| 85 - 89 | 0.8 | 0.5 | -0.03 | 0.5 | 0.5 | 0.00 | 0.7 | 0.6 | -0.01 |
| Gender: female | 44.7 | 57.3 | 0.25 | 55.3 | 54.7 | -0.01 | 48.8 | 48.3 | -0.01 |
| **Medical history: General** |  |  |  |  |  |  |  |  |  |
| Chronic liver disease | 0.9 | 0.9 | -0.01 | 0.9 | 0.9 | 0.00 | 0.9 | 0.9 | 0.00 |
| Chronic obstructive lung disease | 7.2 | 7.1 | 0.00 | 7.1 | 6.9 | -0.01 | 7.3 | 7.1 | 0.00 |
| Dementia | 1.0 | 0.8 | -0.02 | 0.9 | 0.9 | 0.00 | 1.0 | 1.0 | 0.00 |
| Depressive disorder | 6.0 | 8.8 | 0.10 | 8.0 | 7.7 | -0.01 | 6.9 | 7.0 | 0.00 |
| Hyperlipidemia | 48.5 | 46.1 | -0.05 | 45.8 | 45.6 | 0.00 | 47.8 | 47.8 | 0.00 |
| Hypertensive disorder | 54.8 | 53.5 | -0.03 | 53.1 | 52.7 | -0.01 | 54.5 | 54.6 | 0.00 |
| Obesity | 9.6 | 16.2 | 0.20 | 13.8 | 13.4 | -0.01 | 11.6 | 11.9 | 0.01 |
| Osteoarthritis | 18.0 | 22.1 | 0.10 | 20.9 | 20.5 | -0.01 | 19.3 | 19.4 | 0.00 |
| Renal impairment | 10.4 | 9.4 | -0.03 | 9.7 | 9.2 | -0.02 | 10.2 | 10.0 | -0.01 |
| Rheumatoid arthritis | 1.2 | 1.5 | 0.03 | 1.4 | 1.3 | -0.01 | 1.3 | 1.3 | 0.00 |
| **Medical history: Cardiovascular disease** |  |  |  |  |  |  |  |  |  |
| Cerebrovascular disease | 4.7 | 3.9 | -0.04 | 4.0 | 3.9 | 0.00 | 4.5 | 4.5 | 0.00 |
| Coronary arteriosclerosis | 16.4 | 11.4 | -0.14 | 12.0 | 11.7 | -0.01 | 15.0 | 14.7 | -0.01 |
| Heart failure | 8.6 | 5.0 | -0.14 | 5.2 | 5.0 | -0.01 | 7.6 | 6.8 | -0.03 |
| Ischemic heart disease | 7.2 | 4.6 | -0.11 | 4.8 | 4.7 | -0.01 | 6.4 | 6.1 | -0.01 |
| Peripheral vascular disease | 7.5 | 6.7 | -0.03 | 6.8 | 6.6 | -0.01 | 7.3 | 7.3 | 0.00 |
| **Medical history: Neoplasms** |  |  |  |  |  |  |  |  |  |
| Malignant neoplastic disease | 9.1 | 8.7 | -0.02 | 8.8 | 8.5 | -0.01 | 9.0 | 8.8 | -0.01 |
| Malignant tumor of breast | 1.5 | 1.8 | 0.02 | 1.8 | 1.7 | 0.00 | 1.6 | 1.5 | -0.01 |
| Malignant tumor of colon | 0.4 | 0.3 | -0.01 | 0.4 | 0.3 | 0.00 | 0.4 | 0.3 | 0.00 |
| Malignant tumor of lung | 0.3 | 0.3 | -0.01 | 0.3 | 0.3 | 0.00 | 0.3 | 0.3 | 0.00 |
| Malignant tumor of urinary bladder | 0.4 | 0.4 | 0.00 | 0.4 | 0.4 | 0.00 | 0.4 | 0.5 | 0.01 |
| Primary malignant neoplasm of prostate | 2.3 | 1.8 | -0.03 | 1.9 | 1.9 | 0.00 | 2.2 | 2.2 | 0.00 |
| **Medication use** |  |  |  |  |  |  |  |  |  |
| Agents acting on the renin-angiotensin system | 74.4 | 70.2 | -0.10 | 71.0 | 70.9 | 0.00 | 73.2 | 73.0 | 0.00 |
| Antibacterials for systemic use | 55.4 | 59.1 | 0.07 | 58.5 | 57.8 | -0.01 | 56.7 | 56.3 | -0.01 |
| Antiinflammatory and antirheumatic products | 33.6 | 36.0 | 0.05 | 35.5 | 34.9 | -0.01 | 34.5 | 34.0 | -0.01 |
| Antithrombotic agents | 28.7 | 22.2 | -0.15 | 22.6 | 22.7 | 0.00 | 26.6 | 26.4 | 0.00 |
| Beta blocking agents | 47.5 | 41.1 | -0.13 | 41.8 | 41.7 | 0.00 | 45.5 | 45.4 | 0.00 |
| Calcium channel blockers | 33.2 | 31.1 | -0.04 | 31.5 | 31.5 | 0.00 | 32.5 | 32.8 | 0.01 |
| Diuretics | 47.9 | 49.4 | 0.03 | 49.3 | 48.5 | -0.02 | 48.6 | 48.1 | -0.01 |
| Drugs for obstructive airway diseases | 39.5 | 44.3 | 0.10 | 43.4 | 42.7 | -0.01 | 41.2 | 41.0 | 0.00 |
| Lipid modifying agents | 81.1 | 77.0 | -0.10 | 77.7 | 77.8 | 0.00 | 79.8 | 79.8 | 0.00 |
| Opioids | 22.8 | 26.2 | 0.08 | 25.4 | 24.8 | -0.01 | 24.0 | 23.8 | -0.01 |

Abbreviations: SGLT2I: sodium-glucose transporter-2 inhibitor; GLP1RA: glucagon-like peptide-1 receptor agonist; StdDiff: standardized difference of mean.

### Study Diagnostics

Following study diagnostics, the number of databases included for each drug comparison varied. For GLP1RA comparisons, a median of 2.0 (interquartile range [IQR] 1.0–2.0) databases were included for GLP1RA vs. DPP4I, and 1.0 (1.0–1.0) for GLP1RA vs. SU. For SGLT2I comparisons, a median of 4.0 (IQR 2.0–4.0) databases were included for SGLT2I vs. GLP1RA, 6.0 (5.0–7.0) for SGLT2I vs. DPP4I, and 3.0 (3.0–3.0) for SGLT2I vs. SU. Finally, for the DPP4I vs. SU comparison, a median of 6.5 (5.0–8.0) databases were included. Detailed diagnostic criteria—including thresholds for empirical equipoise, SMD, and MDRR—for each database-comparison pair are available in Supplementary Figure 36–53.

### Safety Outcomes

Meta-analytic results from the primary analysis (on-treatment follow-up [OT1] with propensity score stratification) are presented in Figure 2. Compared with SUs and DPP4Is, both GLP1RAs and SGLT2Is were associated with more favorable safety profiles across multiple metabolic and organ-related complications.

**Figure 2.**
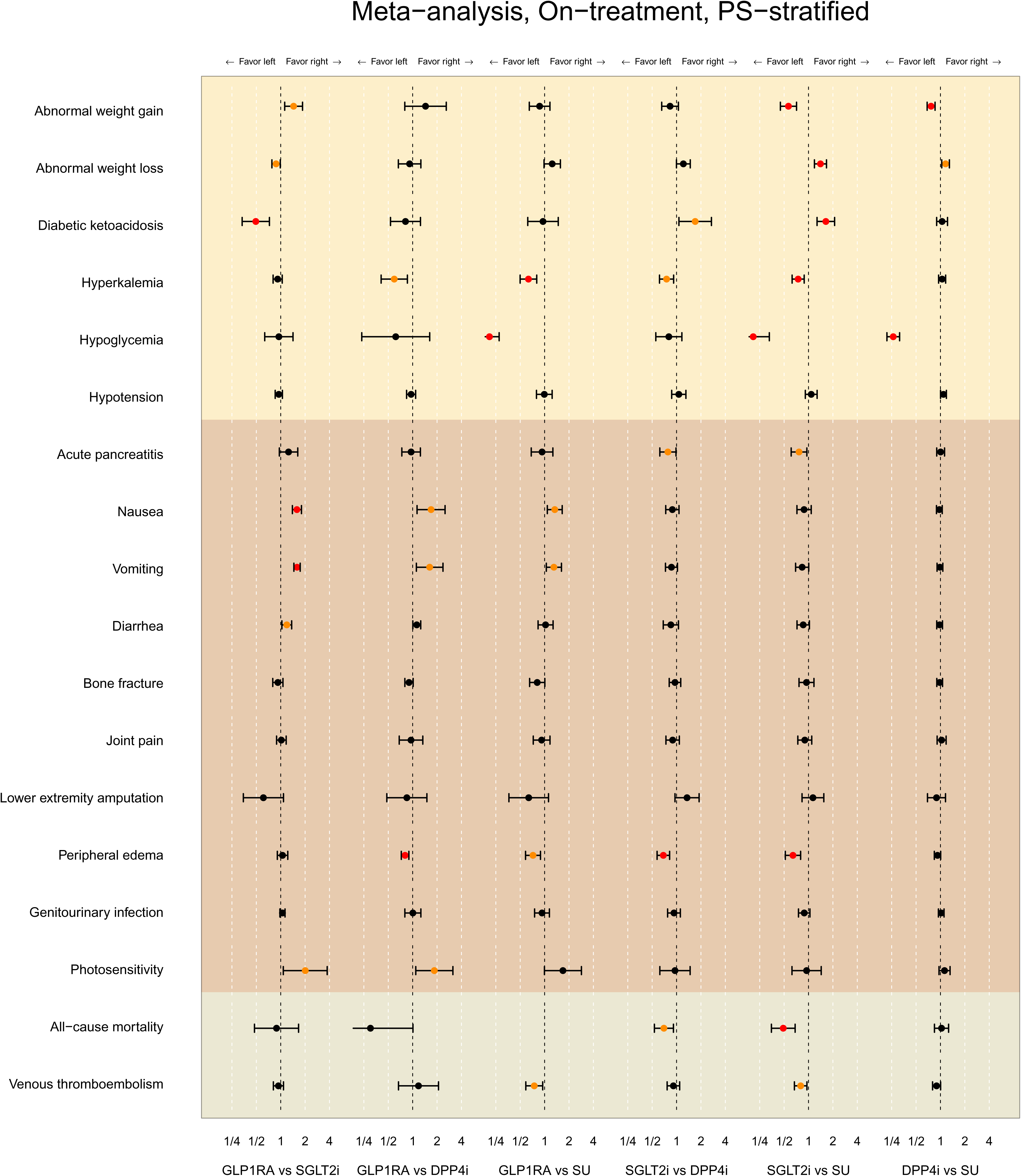
Meta-analytic safety profiles comparing new users of SGLT2I to GLP1RA, DPP4I, and SU across 18 outcomes. Points and lines identify HR estimates with their 95% CIs, respectively. Outcomes in orange signify that the p<0.05 and outcomes in red mean statistically significant after Bonferroni correction (p<0.00277778) for the multiple testing. The result for all-cause mortality in the GLP1RA vs SU comparison was not presented because there was no valid result after the study diagnostic process.

**Figure 3.**
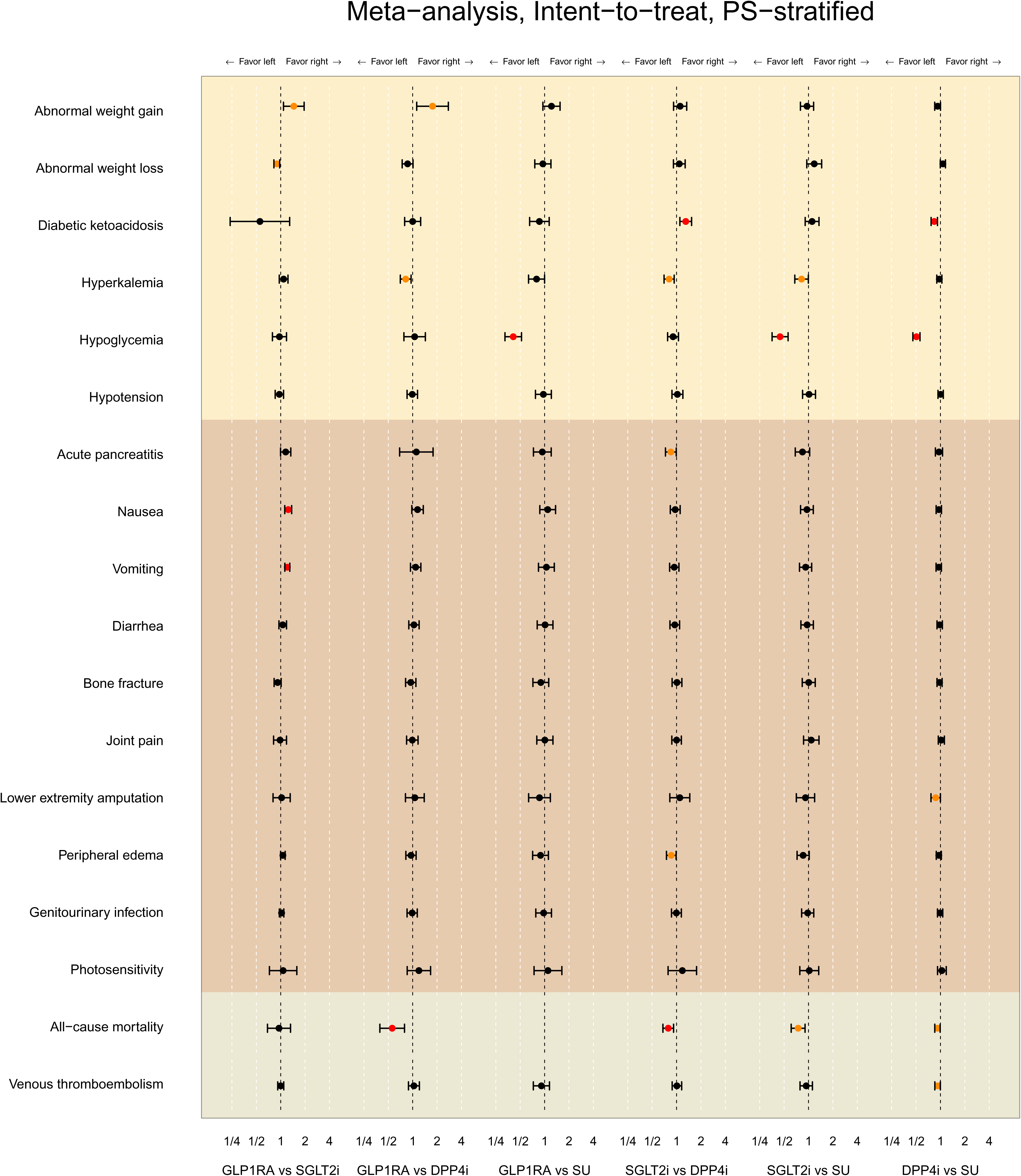
Safety profiles comparing new users of SGLT2I to GLP1RA, DPP4I, and SU across 18 outcomes from the intent-to-treat follow-up. Points and lines identify HR estimates with their 95% CIs, respectively. Outcomes in orange signify that the p<0.05 and outcomes in red mean statistically significant after Bonferroni correction (p<0.00277778) for the multiple testing. The result for all-cause mortality in the GLP1RA vs SU comparison was not presented because there was no valid result after the study diagnostic process.

GLP1RA use was associated with a substantially lower risk of hypoglycemia compared to SU (HR 0.21 [95% CI, 0.16–0.28]), similar to SGLT2I (HR 0.20 [0.12–0.34]) and DPP4I (HR 0.26 [0.21–0.32]). Additionally, both GLP1RAs and SGLT2Is had significantly lower risks of hyperkalemia compared to SU (HR 0.63 [0.50–0.81] and 0.75 [0.63–0.90], respectively), with no meaningful difference observed between the two newer classes. SGLT2Is, in particular, demonstrated lower risk of abnormal weight gain (HR 0.57 [0.45–0.71]) and higher incidence of abnormal weight loss (HR 1.41 [1.18–1.67]) than SUs, reflecting their known weight-reducing properties. For peripheral edema, both GLP1RAs and SGLT2Is demonstrated lower risks than DPP4Is (HR 0.81 [0.72–0.90] and 0.69 [0.57–0.82], respectively), and were also favorable compared to SUs (HR 0.72 [0.58–0.90] and 0.62 [0.46–0.84], respectively).

However, SGLT2Is showed a significantly increased risk compared to GLP1RAs (HR 2.03 [1.38–2.99]) and SUs (HR 1.64 [1.27–2.10]). GLP1RAs had shown high risks of nausea and vomiting (HR 1.58 [1.45–1.72] and 1.58 [1.44–1.74], respectively) compared to SGLT2Is.

No significant differences across drug classes were observed for bone fracture, joint pain, lower extremity amputation, or venous thromboembolism. Importantly, SGLT2Is were associated with a significantly lower all-cause mortality risk compared to SUs (HR 0.49 [0.35– 0.68]), reinforcing the growing evidence supporting their long-term safety benefits in older adults.

### Sensitivity Analysis

Findings from sensitivity analyses—using the OT2 follow-up design, which censors patients at treatment escalation—are presented in Supplementary Tables 216–323 and were largely consistent with the primary analysis. GLP1RAs continued to show a favorable safety profile, including a significantly lower risk of hyperkalemia compared to SUs (HR 0.62 [0.49–0.77]) and reduced risk of peripheral edema compared to SUs (HR 0.71 [0.58–0.86]). SGLT2Is also maintained a lower risk of peripheral edema compared to both DPP4Is and SUs (HR 0.70 [0.60– 0.83] and HR 0.65 [0.52–0.80], respectively).

However, SGLT2Is remained associated with a significantly increased risk of diabetic ketoacidosis relative to both GLP1RAs (HR 1.99 [1.31–3.01]) and SUs (HR 1.70 [1.28–2.27]). In contrast, GLP1RAs continued to demonstrate inferior gastrointestinal tolerability, with significantly higher risks of nausea (HR 1.63 [1.50–1.77]) and vomiting (HR 1.62 [1.47–1.79]) compared to SGLT2Is. Across all drug classes, the risk of hypoglycemia remained significantly lower compared to SU, further reinforcing the advantages of GLP1RAs (HR 0.21 [0.16–0.27]), SGLT2Is (HR 0.17 [0.09–0.30]), and DPP4Is (HR 0.17 [0.13–0.21]).

In the ITT analysis, which introduces greater bias toward the null due to persistent exposure misclassification, fewer relative risk estimates met the Bonferroni-corrected significance threshold. Nonetheless, key safety signals remained consistent. GLP1RAs continued to show higher risks of nausea (HR 1.24 [1.10–1.40]) and vomiting (HR 1.22 [1.10–1.34]) compared to SGLT2Is. Importantly, GLP1RAs and SGLT2Is showed a significantly lower risk of all-cause mortality compared to DPP4Is (HR 0.56 [0.39–0.79] and 0.79 [0.67–0.93], respectively), highlighting their potential long-term benefit in older adults.

## DISCUSSION

This study provides new and comprehensive insight into the comparative safety of second-line antihyperglycemic agents in older adults—a population at high risk for drug-related complications yet historically underrepresented in clinical trials. By analyzing over 1.8 million patients across multiple international databases, we found that GLP1RAs and SGLT2Is generally offer more favorable safety profiles compared to SUs and DPP4Is. Importantly, we identified clinically relevant differences across a broad set of outcomes—not just the well-known risks like hypoglycemia or gastrointestinal side effects,^22^ but also less frequently assessed events in clinical trials such as peripheral edema, diabetic ketoacidosis, and hyperkalemia.^23–27^ These findings help move the field beyond efficacy comparisons to a more nuanced, safety-centered understanding of treatment choices in older adults with type 2 diabetes.

The observed differences in safety profiles across drug classes likely reflect their distinct pharmacologic mechanisms and how those interact with age-related vulnerabilities. Sulfonylureas, which stimulate insulin secretion irrespective of glucose levels, predictably increase the risk of hypoglycemia, especially in older adults with fluctuating food intake or renal impairment. The weight gain and edema associated with sulfonylureas and DPP4Is may be tied to fluid retention and insulin-mediated anabolic effects. In contrast, SGLT2Is promote glucosuria and osmotic diuresis, which can reduce body weight and blood pressure but also predispose to volume depletion and ketoacidosis in frail or volume-sensitive patients. The lower incidence of peripheral edema observed with GLP1RAs and SGLT2Is may be due to their favorable renal and vascular effects. SGLT2I was associated with an elevated risk of diabetic ketoacidosis, which previous retrospective studies have similarly identified^28–32^ but rarely detected in RCTs.^23,24^ In a context where previous studies have limited evidence on hyperkalemia with SGLT2I and GLP1RA,^25–27^ our study showed a significantly lower risk of hyperkalemia with SGLT2I and GLP1RA. However, potential differences between SGLT2I and GLP1RA suggested in previous literature were not observed in this study.^26^ Bone fracture, which is specifically important to older adults and reported SGLT2Is can potentially cause in a previous trial, did not show significantly different risks in any comparison in line with other observational studies.^31,33^ GLP1RAs, acting through central and gastrointestinal pathways, are known to cause nausea and vomiting, especially during initiation. Incretin-based glucose lowering agents have been reported to cause acute pancreatitis potentially,^34–36^ however, no significant difference was found between SGLT2I and GLP1RA for acute pancreatitis. However, previous studies have shown that liraglutide strongly associated with pancreatitis among GLP1RAs,^37–40^ further studies on individual ingredient are needed because we had very low rate of liraglutide in the population used GLP1RA. Finally, the signals for reduced all-cause mortality among GLP1RA and SGLT2I users align with emerging evidence of their cardioprotective and anti-inflammatory properties, which may translate into broader safety benefits in older populations. These results emphasize that prioritizing SGLT2Is and GLP1RAs as second-line treatment in older adults over the current pattern of prescribing DPP4s and SUs with T2DM may be potentially beneficial.^7^

Our study extends the literature in three important ways. First, we focus explicitly on safety outcomes in older adults, a population frequently excluded or underrepresented in randomized controlled trials but disproportionately vulnerable to adverse drug events due to multimorbidity, frailty, and polypharmacy. Prior studies have either aggregated findings across age groups or relied on post hoc subgroup analyses, limiting the ability to draw robust conclusions specific to older adults. Second, we conduct a comprehensive, class-wide comparison of four major second-line antihyperglycemic drug classes—GLP1RA, SGLT2Is, DPP4Is, and SUs—rather than focusing on isolated pairwise comparisons or single adverse events. Previous real-world studies have typically concentrated on one or two outcomes (e.g., hypoglycemia or heart failure) or compared only a subset of these drug classes. Our study fills this gap by systematically examining 18 clinically relevant outcomes, allowing for a more balanced and practical evaluation of the relative safety of each class. Third, we applied a rigorous and transparent analytic framework, using harmonized multinational real-world data, large-scale propensity score adjustment, and empirical calibration. While prior observational studies often rely on a single database or apply variable methods across studies, our approach— anchored in the LEGEND and OHDSI frameworks—ensures consistency, reproducibility, and generalizability of the findings across diverse healthcare settings. This methodological strength not only confirms previously documented risks (such as hypoglycemia with sulfonylureas) but also uncovers less-recognized differences, such as the gastrointestinal tolerability advantages of SGLT2Is over GLP1RAs and the mortality benefits of SGLT2Is compared to sulfonylureas. These nuanced insights are particularly valuable for tailoring treatment decisions in older adults, where safety considerations often take precedence over modest differences in glycemic control.

These findings have important clinical and policy implications. First, they reinforce current guideline recommendations—such as those from the American Diabetes Association and European Association for the Study of Diabetes—that prioritize GLP1RAs and SGLT2Is over SUs and DPP4Is, particularly in older adults. These preferences have largely been based on cardiometabolic benefits, but our results add further justification by demonstrating superior safety profiles across a wide range of adverse outcomes. In clinical practice, this means that clinicians should prioritize newer agents—especially in patients with multiple comorbidities, frailty, or a history of adverse drug reactions. Second, the findings highlight the need to reevaluate entrenched prescribing patterns that continue to favor SUs, which—despite their low cost—pose disproportionate risks of hypoglycemia, weight gain, and mortality in older adults. These patterns may reflect legacy habits, formulary limitations, or clinician concerns about the cost or side effects of newer agents. Addressing these barriers may require targeted educational efforts, updated formularies, or value-based pricing strategies to ensure that safety and effectiveness guide treatment choices. Third, these results can inform shared decision-making by providing patients and caregivers with more nuanced information about potential risks and tolerability profiles. For example, the lower rates of nausea and vomiting associated with SGLT2Is compared to GLP1RAs may be meaningful to patients who previously discontinued medications due to gastrointestinal side effects. Likewise, awareness of the increased risk of diabetic ketoacidosis with SGLT2Is may prompt closer monitoring or alternative choices in certain subgroups, such as those with low insulin reserve or volume depletion. At a systems level, this study underscores the feasibility and value of multinational, large-scale real-world safety surveillance. The use of harmonized data and transparent, reproducible methods provides a model for generating actionable evidence that is both rigorous and patient-centered. These approaches can be extended to other drug classes, populations, or outcomes—supporting a more responsive and evidence-driven healthcare system, especially for populations such as older adults who are often excluded from clinical trials.

Several limitations should be considered when interpreting our findings. First, as with all observational studies, residual confounding cannot be fully excluded, despite our use of large-scale propensity score adjustment and empirical calibration. Second, we examined drug classes rather than individual agents, which may obscure important within-class heterogeneity— particularly relevant for GLP1RAs, where agents differ in gastrointestinal tolerability and cardiovascular effects. Third, medication exposure was based on prescription or dispensing records, which may not reflect actual patient adherence. Additionally, we did not incorporate laboratory data (e.g., glucose or electrolyte levels) or patient-reported outcomes, limiting the granularity of some safety assessments. Finally, certain outcomes of interest—such as cognitive decline or neuropsychiatric effects—were not included due to limitations in the underlying data or their absence at the time of protocol development. While these limitations may attenuate some findings or leave specific questions unanswered, they do not diminish the overall value of this large-scale, real-world evaluation of safety in a high-risk population.

In conclusion, GLP1RAs and SGLT2Is are generally safer than SUs and DPP4Is in older adults with type 2 diabetes, with lower risks of hypoglycemia, hyperkalemia, edema, and mortality. These findings provide robust, real-world evidence to support safer prescribing choices in a high-risk population. Incorporating these safety considerations into clinical decision-making can improve outcomes and reduce avoidable harm in older adults.

## Data Availability

All data produced are available online at https://data.ohdsi.org/LegendT2dmClassEvidenceExplorer/

https://ohdsi-studies.github.io/LegendT2dm/Protocol.html

https://data.ohdsi.org/LegendT2dmClassEvidenceExplorer/

## Funding

This study was partially funded through National Institutes of Health grants R01 HL167858, K23 HL153775, R01 LM006910, R01 HG006139, and R01 HL169954 and the U.S. Department of Veterans Affairs under the research priority to “Put VA Data to Work for Veterans” (VA ORD 22-D4V). The funders had no role in the design and conduct of the protocol; preparation, review, or approval of the manuscript; and decision to submit the manuscript for publication.

## Disclosure

Dr. Khera is an associate editor at JAMA; and has received support from the National Institutes of Health (awards R01 AG089981, R01 HL167858 and K23 HL153775) and the Doris Duke Charitable Foundation (award 2022060); has received research support, through Yale University, from Bristol Myers Squibb, Novo Nordisk, and BridgeBio; is a coinventor of U.S. provisional patent applications WO2023230345A1, US20220336048A1, 63/177,117, 63/346,610, 63/428,569, 63/484,426, 63/508,315, 63/580,137, 63/606,203, 63/619,241, and 63/562,335, unrelated to the present work; and is a cofounder of Evidence2Health and Ensight-AI, precision health platforms to improve evidence-based cardiovascular care and cardiovascular diagnostics. Dr. DuVall has received grants from Alnylam Pharmaceuticals, AstraZeneca Pharmaceuticals, Biodesix, Celgene, Cerner Enviza, GlaxoSmithKline, Janssen Pharmaceuticals, Novartis International, and Parexel International through the University of Utah or the Western Institute for Veteran Research (outside the submitted work). In the past 3 years, Dr. Krumholz has received expenses and/or personal fees from UnitedHealth, Element Science, Aetna, Reality Labs, Tesseract/4Catalyst, F-Prime, the Siegfried and Jensen Law Firm, the Arnold and Porter Law Firm, and the Martin/Baughman Law Firm; is a cofounder of Refactor Health, HugoHealth, and Ensight-AI; and is associated with contracts, through Yale New Haven Hospital, from the Centers for Medicare and Medicaid Services and, through Yale University, from Johnson & Johnson. Dr. Thangaraj is a coinventor on provisional patent 63/606,203, unrelated to the present work, and is funded by grant 5T32 HL155000-03. Ms. Blacketer, Dr. Ostropolets, and Dr. Ryan are employees of Johnson & Johnson. Dr. Schuemie is an employee and shareholder of Johnson & Johnson. Dr. Suchard receives additional contracts and grants from the US Food & Drug Administration, the US National Institutes of Health and Janssen Research & Development outside the scope of this work. Dr. Lau received research fundings from Diabetes UK outside the scope of this work. Dr. You reports being a chief executive officer of PHI Digital Healthcare; and grants from Daiichi Sankyo. He is a coinventor of granted Korea Patent DP-2023-1223 and DP-2023-0920, and pending Patent Applications DP-2024-0909, DP-2024-0908, DP-2022-1658, DP-2022-1478, and DP-2022-1365, unrelated to current work. Dr. Man has received support from the C.W. Maplethorpe Fellowship, the National Institute of Health Research, European Commission Framework Horizon 2020, the Hong Kong Research Grant Council, and the Innovation and Technology Commission of the Hong Kong Special Administration Region Government outside the submitted work). Dr. Morales is supported by a Wellcome Trust Clinical Research Fellowship (214588/Z/18/Z). Dr. Lu received support from the National Heart, Lung, and Blood Institute of the National Institutes of Health (under awards R01HL69954 and R01HL169171), the Patient-Centered Outcomes Research Institute (under award HM-2022C2-28354), Sentara Research Foundation, and Novartis through Yale University. All other authors have reported that they have no relationships relevant to the contents of this paper to disclose.

## Notes

### Clinical Protocols

https://ohdsi-studies.github.io/LegendT2dm/Protocol.html

## REFERENCES

1 American Diabetes Association Professional Practice Committee; 9. Pharmacologic Approaches to Glycemic Treatment: Standards of Care in Diabetes—2024. Diabetes Care 47, S158–S178 (2023). 10.2337/dc24-S009

2 Qaseem, A. et al. Newer Pharmacologic Treatments in Adults With Type 2 Diabetes: A Clinical Guideline From the American College of Physicians. Annals of Internal Medicine (2024). 10.7326/M23-2788

3 American Diabetes Association Professional Practice Committee; 13. Older Adults: Standards of Care in Diabetes—2024. Diabetes Care 47, S244–S257 (2023). 10.2337/dc24-S013

4 Engler, C. et al. Long-term trends in the prescription of antidiabetic drugs: real-world evidence from the Diabetes Registry Tyrol 2012–2018. BMJ Open Diabetes Research &amp; Care 8, e001279 (2020). 10.1136/bmjdrc-2020-001279

5 Schernthaner, G. et al. Worldwide inertia to the use of cardiorenal protective glucose-lowering drugs (SGLT2i and GLP-1 RA) in high-risk patients with type 2 diabetes. Cardiovascular Diabetology 19, 185 (2020). 10.1186/s12933-020-01154-w

6 Bellary, S. & Barnett, A. H. SGLT2 inhibitors in older adults: overcoming the age barrier. The Lancet Healthy Longevity 4, e127–e128 (2023). 10.1016/S2666-7568(23)00039-9

7 Khera, R. et al. Multinational patterns of second line antihyperglycaemic drug initiation across cardiovascular risk groups: federated pharmacoepidemiological evaluation in LEGEND-T2DM. BMJ Medicine 2, e000651 (2023). 10.1136/bmjmed-2023-000651

8 Suchard, M. A. et al. Comprehensive comparative effectiveness and safety of first-line antihypertensive drug classes: a systematic, multinational, large-scale analysis. The Lancet 394, 1816–1826 (2019). 10.1016/S0140-6736(19)32317-7

9 Schuemie, M. J. et al. Principles of Large-scale Evidence Generation and Evaluation across a Network of Databases (LEGEND). Journal of the American Medical Informatics Association 27, 1331–1337 (2020). 10.1093/jamia/ocaa103

10 Khera, R. et al. Comparative Effectiveness of Second-Line Antihyperglycemic Agents for Cardiovascular Outcomes: A Multinational, Federated Analysis of LEGEND-T2DM. Journal of the American College of Cardiology 84, 904–917 (2024). 10.1016/j.jacc.2024.05.069

11 Hripcsak, G. et al. in MEDINFO 2015: eHealth-enabled Health 574–578 (IOS Press, 2015).

12 Hripcsak, G., et al. RESEARCH PROTOCOL Large-scale evidence generation and evaluation across a network of databases for type 2 diabetes mellitus, <https://ohdsi-studies.github.io/LegendT2dm/Protocol.html> (2021).

13 Khera, R. et al. Large-scale evidence generation and evaluation across a network of databases for type 2 diabetes mellitus (LEGEND-T2DM): a protocol for a series of multinational, real-world comparative cardiovascular effectiveness and safety studies. BMJ Open 12, e057977 (2022). 10.1136/bmjopen-2021-057977

14 von Elm, E. et al. The Strengthening the Reporting of Observational Studies in Epidemiology (STROBE) statement: guidelines for reporting observational studies. The Lancet 370, 1453–1457 (2007). 10.1016/S0140-6736(07)61602-X

15 Diabetes, A. A. Updates to the Standards of Medical Care in Diabetes—2018. Diabetes Care 41, 2045–2047 (2018). 10.2337/dc18-su09

16 Rao, G. (2024).

17 Tian, Y., Schuemie, M. J. & Suchard, M. A. Evaluating large-scale propensity score performance through real-world and synthetic data experiments. International Journal of Epidemiology 47, 2005–2014 (2018). 10.1093/ije/dyy120

18 Schuemie, M. J., Ryan, P. B., DuMouchel, W., Suchard, M. A. & Madigan, D. Interpreting observational studies: why empirical calibration is needed to correct p-values. Statistics in Medicine 33, 209–218 (2014). 10.1002/sim.5925

19 Walker, A. M. et al. A tool for assessing the feasibility of comparative effectiveness research. Comparative Effectiveness Research 3, 11–20 (2013). 10.2147/CER.S40357

20 Armstrong, B. A SIMPLE ESTIMATOR OF MINIMUM DETECTABLE RELATIVE RISK, SAMPLE SIZE, OR POWER IN COHORT STUDIES. American Journal of Epidemiology 126, 356–358 (1987). 10.1093/aje/126.2.356

21 Schuemie, M. et al. Health-Analytics Data to Evidence Suite (HADES): Open-Source Software for Observational Research. Stud Health Technol Inform 310, 966–970 (2024). 10.3233/shti231108

22 The GRADE Study Research Group, Glycemia Reduction in Type 2 Diabetes — Glycemic Outcomes. New England Journal of Medicine 387, 1063–1074 (2022). doi:10.1056/NEJMoa2200433

23 Zinman, B. et al. Empagliflozin, Cardiovascular Outcomes, and Mortality in Type 2 Diabetes. New England Journal of Medicine 373, 2117–2128 (2015). doi:10.1056/NEJMoa1504720

24 Neal, B. et al. Canagliflozin and Cardiovascular and Renal Events in Type 2 Diabetes. New England Journal of Medicine 377, 644–657 (2017). doi:10.1056/NEJMoa1611925

25 Neuen, B. L. et al. Sodium-Glucose Cotransporter 2 Inhibitors and Risk of Hyperkalemia in People With Type 2 Diabetes: A Meta-Analysis of Individual Participant Data From Randomized, Controlled Trials. Circulation 145, 1460–1470 (2022). doi:10.1161/CIRCULATIONAHA.121.057736

26 Fu, E. L. et al. SGLT-2 inhibitors, GLP-1 receptor agonists, and DPP-4 inhibitors and risk of hyperkalemia among people with type 2 diabetes in clinical practice: population based cohort study. BMJ 385, e078483 (2024). 10.1136/bmj-2023-078483

27 Huang, T. et al. GLP-1RA vs DPP-4i Use and Rates of Hyperkalemia and RAS Blockade Discontinuation in Type 2 Diabetes. JAMA Internal Medicine 184, 1195–1203 (2024). 10.1001/jamainternmed.2024.3806

28 Fralick, M., Schneeweiss, S. & Patorno, E. Risk of Diabetic Ketoacidosis after Initiation of an SGLT2 Inhibitor. New England Journal of Medicine 376, 2300–2302 (2017). doi:10.1056/NEJMc1701990

29 Douros, A. et al. Sodium–Glucose Cotransporter-2 Inhibitors and the Risk for Diabetic Ketoacidosis. Annals of Internal Medicine 173, 417–425 (2020). 10.7326/M20-0289

30 Fralick, M., Colacci, M., Thiruchelvam, D., Gomes, T. & Redelmeier, D. A. Sodium-glucose co-transporter-2 inhibitors versus dipeptidyl peptidase-4 inhibitors and the risk of heart failure: A nationwide cohort study of older adults with diabetes mellitus. Diabetes, Obesity and Metabolism 23, 950–960 (2021). 10.1111/dom.14300

31 Patorno, E. et al. Comparative Effectiveness and Safety of Sodium–Glucose Cotransporter 2 Inhibitors Versus Glucagon-Like Peptide 1 Receptor Agonists in Older Adults. Diabetes Care 44, 826–835 (2021). 10.2337/dc20-1464

32 Goldman, A. et al. The real-world safety profile of sodium-glucose co-transporter-2 inhibitors among older adults (≥ 75 years): a retrospective, pharmacovigilance study. Cardiovascular Diabetology 22, 16 (2023). 10.1186/s12933-023-01743-5

33 Zhuo, M. et al. Association of Sodium-Glucose Cotransporter–2 Inhibitors With Fracture Risk in Older Adults With Type 2 Diabetes. JAMA Network Open 4, e2130762–e2130762 (2021). 10.1001/jamanetworkopen.2021.30762

34 Liu, L., Chen, J., Wang, L., Chen, C. & Chen, L. Association between different GLP-1 receptor agonists and gastrointestinal adverse reactions: A real-world disproportionality study based on FDA adverse event reporting system database. Frontiers in Endocrinology 13 (2022). 10.3389/fendo.2022.1043789

35 Alenzi, K. A., Alsuhaibani, D., Batarfi, B. & Alshammari, T. M. Pancreatitis with use of new diabetic medications: a real-world data study using the post-marketing FDA adverse event reporting system (FAERS) database. Frontiers in Pharmacology 15 (2024). 10.3389/fphar.2024.1364110

36 Singh, A. K., Gangopadhyay, K. K. & Singh, R. Risk of acute pancreatitis with incretin-based therapy: a systematic review and updated meta-analysis of cardiovascular outcomes trials. Expert Review of Clinical Pharmacology 13, 461–468 (2020). 10.1080/17512433.2020.1736041

37 Marso, S. P. et al. Semaglutide and Cardiovascular Outcomes in Patients with Type 2 Diabetes. New England Journal of Medicine 375, 1834–1844 (2016). doi:10.1056/NEJMoa1607141

38 Gerstein, H. C. et al. Dulaglutide and cardiovascular outcomes in type 2 diabetes (REWIND): a double-blind, randomised placebo-controlled trial. The Lancet 394, 121–130 (2019). 10.1016/S0140-6736(19)31149-3

39 Holman, R. R. et al. Effects of Once-Weekly Exenatide on Cardiovascular Outcomes in Type 2 Diabetes. New England Journal of Medicine 377, 1228–1239 (2017). doi:10.1056/NEJMoa1612917

40 Abd El Aziz, M., Cahyadi, O., Meier, J. J., Schmidt, W. E. & Nauck, M. A. Incretin-based glucose-lowering medications and the risk of acute pancreatitis and malignancies: a meta-analysis based on cardiovascular outcomes trials. Diabetes, Obesity and Metabolism 22, 699–704 (2020). 10.1111/dom.13924

