## Supplementary file for "Comparative Safety of Second-Line Antihyperglycemic Agents in Older Adults with Type 2 Diabetes: A Multinational Real-World Evidence From LEGEND-T2DM"

### Supplementary Material

**4 Negative control outcomes** **144**

**5 Study diagnostics for final inclusion in meta-analysis** **148**

**6 Exposures, outcomes and risk across data sources and outcomes** **166**

8 Bibliography 382

### 1 Data sources

**Supplementary Table 1:** Data sources and the populations they cover.

| Data source | Population | Patients | History | Data capture process and short description |
|---|---|---|---|---|
| | | **Administrative claims** | | |
| IBM MarketScan Commercial Claims and Encounters (CCAE) | Commercially insured, < 65 years | 142M | 2000 – | Adjudicated health insurance claims (e.g. inpatient, outpatient, and outpatient pharmacy) from large employers and health plans who provide private healthcare coverage to employees, their spouses and dependents. |
| IBM MarketScan Medicare Supplemental Database (MDCR) | Commercially insured, 65$+$ years | 10M | 2000 – | Adjudicated health insurance claims of retirees with primary or Medicare supplemental coverage through privately insured fee-for-service, point-of-service or capitated health plans. |
| IBM MarketScan Multi-State Medicaid Database (MDCD) | Medicaid enrollees, racially diverse | 26M | 2006 – | Adjudicated health insurance claims for Medicaid enrollees from multiple states and includes hospital discharge diagnoses, outpatient diagnoses and procedures, and outpatient pharmacy claims. |
| IQVIA Open Claims (Open Claims) | General | 160M | 2010 – | Pre-adjudicated claims at the anonymized patient-level collected from office-based physicians and specialists via office management software and clearinghouse switch sources for the purpose of reimbursement. |
| Optum Clinformatics Data Mart (OptumDOD) | Commercially or Medicare insured | 85M | 2000 – | Inpatient and outpatient healthcare insurance claims. |
| | | **Electronic health records (EHRs)** | | |
| IQVIA Disease Analyzer Germany (DAG) | Germany, general | 37M | 1992 – | Collection from patient management software used by general practitioners and selected specialists to document patients' medical records within their office-based practice during a visit. |
| Optum Electronic Health Records (OptumEHR) | US, general | 93M | 2006 – | Clinical information, prescriptions, lab results, vital signs, body measurements, diagnoses and procedures derived from clinical notes using natural language processing. |
| Information System for Research in Primary Care (SIDIAP) | 80% of all Catalonia (Spain) | 7.7M | 2006 – | Primary care partially linked to inpatient data with pharmacy dispensations and primary care laboratories. Healthcare is universal and taxpayer funded in the region, and PCPs are gatekeeps for all care and responsible for repeat prescriptions. |
| US Department of Veterans Affairs (VA) | Veterans, older, racially diverse | 12M | 2000 – | National VA health care system, the largest integrated provider of medical services in the US, provided at 170 VA medical centers and 1,063 outpatient sites. |

### 2 Baseline patient characteristics across data sources

#### 2.1 GLP1RA vs SGLT2I

##### 2.1.1 OptumDOD

**Supplementary Table 2:** Baseline patient characteristics for GLP1RA (T) and SGLT2I (C) new-users in the OptumDOD data source. We report proportion of initiators satisfying selected base-line characteristics and the standardized difference of population proportions (StdDiff) before and after propensity score adjustment. Less extreme StdDiffs through matching and stratification suggest improved balance between patient cohorts through adjustment.

| | Before adjustment | | | After matching | | | After stratification | | |
|---|---|---|---|---|---|---|---|---|---|
| Characteristic | T (%) | C (%) | StdDiff | T (%) | C (%) | StdDiff | T (%) | C (%) | StdDiff |
| **Age group** | | | | | | | | | |
| 65 - 69 | 46.5 | 40.3 | 0.12 | 45.7 | 43.4 | 0.05 | 43.4 | 41.5 | 0.04 |
| 70 - 74 | 33.3 | 33.8 | -0.01 | 33.3 | 34.1 | -0.02 | 33.6 | 34.0 | -0.01 |
| 75 - 79 | 14.7 | 16.9 | -0.06 | 15.0 | 15.6 | -0.02 | 16.3 | 16.4 | 0.00 |
| 80 - 84 | 4.0 | 6.2 | -0.10 | 4.3 | 4.6 | -0.01 | 5.2 | 5.7 | -0.02 |
| 85 - 89 | 1.4 | 2.4 | -0.07 | 1.5 | 2.1 | -0.04 | 1.5 | 2.2 | -0.05 |
| 90 - 94 | 0.1 | 0.3 | -0.05 | 0.1 | 0.3 | -0.04 | <0.1 | 0.3 | -0.05 |
| Gender: female | 57.8 | 45.2 | 0.26 | 55.1 | 57.4 | -0.05 | 47.8 | 49.7 | -0.04 |
| **Medical history: General** | | | | | | | | | |
| Acute respiratory disease | 22.8 | 22.6 | 0.00 | 22.4 | 23.2 | -0.02 | 22.8 | 23.0 | 0.00 |
| Attention deficit hyperactivity disorder | 0.6 | 0.3 | 0.05 | 0.4 | 0.4 | 0.00 | 0.4 | 0.4 | 0.01 |
| Chronic liver disease | 2.0 | 1.5 | 0.04 | 2.0 | 1.6 | 0.03 | 1.8 | 1.5 | 0.02 |
| Chronic obstructive lung disease | 12.7 | 12.3 | 0.01 | 12.4 | 12.7 | -0.01 | 12.9 | 12.6 | 0.01 |
| Crohn's disease | 0.1 | 0.3 | -0.04 | 0.1 | 0.3 | -0.03 | 0.1 | 0.3 | -0.04 |
| Dementia | 2.1 | 1.9 | 0.01 | 2.1 | 1.9 | 0.01 | 2.1 | 1.9 | 0.01 |
| Depressive disorder | 20.1 | 15.5 | 0.12 | 18.7 | 20.7 | -0.05 | 16.1 | 17.7 | -0.04 |
| Gastroesophageal reflux disease | 24.1 | 22.1 | 0.05 | 23.5 | 25.0 | -0.04 | 22.3 | 23.4 | -0.03 |
| Gastrointestinal hemorrhage | 2.3 | 2.5 | -0.01 | 2.1 | 2.3 | -0.02 | 2.3 | 2.5 | -0.01 |
| Human immunodeficiency virus infection | 0.3 | 0.2 | 0.01 | 0.3 | 0.2 | 0.02 | 0.3 | 0.2 | 0.01 |
| Hyperlipidemia | 84.6 | 86.3 | -0.05 | 84.6 | 84.1 | 0.01 | 86.0 | 85.5 | 0.01 |
| Hypertensive disorder | 86.0 | 87.1 | -0.03 | 86.0 | 86.3 | -0.01 | 86.7 | 86.9 | -0.01 |
| Lesion of liver | 1.6 | 1.4 | 0.02 | 1.7 | 1.5 | 0.02 | 1.5 | 1.4 | 0.01 |
| Obesity | 39.6 | 26.3 | 0.29 | 35.3 | 38.4 | -0.06 | 29.2 | 31.6 | -0.05 |
| Osteoarthritis | 40.0 | 34.0 | 0.12 | 38.2 | 39.9 | -0.03 | 35.9 | 36.5 | -0.01 |
| Pneumonia | 4.1 | 4.3 | -0.01 | 4.1 | 4.3 | -0.01 | 4.5 | 4.4 | 0.00 |
| Psoriasis | 2.3 | 2.0 | 0.02 | 2.3 | 2.2 | 0.00 | 2.1 | 2.0 | 0.01 |
| Renal impairment | 20.5 | 19.1 | 0.04 | 20.1 | 20.3 | 0.00 | 20.1 | 19.7 | 0.01 |
| Rheumatoid arthritis | 2.4 | 2.0 | 0.03 | 2.4 | 2.4 | 0.00 | 2.1 | 2.2 | -0.01 |
| Schizophrenia | 0.2 | 0.2 | 0.01 | 0.2 | 0.1 | 0.01 | 0.2 | 0.2 | 0.01 |
| Ulcerative colitis | 0.4 | 0.4 | -0.01 | 0.4 | 0.5 | -0.02 | 0.5 | 0.4 | 0.01 |
| Urinary tract infectious disease | 11.3 | 8.5 | 0.09 | 10.4 | 10.5 | 0.00 | 9.7 | 9.3 | 0.01 |
| Viral hepatitis C | 0.7 | 0.5 | 0.03 | 0.7 | 0.5 | 0.03 | 0.7 | 0.5 | 0.03 |
| Visual system disorder | 54.2 | 54.8 | -0.01 | 54.5 | 54.5 | 0.00 | 54.3 | 54.8 | -0.01 |
| **Medical history: Cardiovascular disease** | | | | | | | | | |
| Atrial fibrillation | 9.8 | 11.6 | -0.06 | 10.2 | 9.7 | 0.01 | 11.2 | 11.0 | 0.01 |
| Cerebrovascular disease | 8.4 | 9.3 | -0.03 | 8.3 | 8.4 | 0.00 | 9.5 | 9.0 | 0.02 |
| Coronary arteriosclerosis | 20.2 | 27.8 | -0.18 | 20.9 | 19.9 | 0.02 | 26.3 | 25.1 | 0.03 |
| Heart disease | 40.7 | 45.7 | -0.10 | 41.1 | 39.2 | 0.04 | 46.0 | 43.7 | 0.05 |
| Heart failure | 9.8 | 12.6 | -0.09 | 9.9 | 9.2 | 0.02 | 12.5 | 11.6 | 0.03 |
| Ischemic heart disease | 10.1 | 14.5 | -0.14 | 10.5 | 10.4 | 0.00 | 13.0 | 13.1 | 0.00 |
| Peripheral vascular disease | 16.9 | 18.2 | -0.03 | 17.1 | 16.6 | 0.01 | 17.9 | 17.7 | 0.00 |
| Pulmonary embolism | 1.2 | 1.1 | 0.01 | 1.2 | 1.1 | 0.01 | 1.1 | 1.1 | 0.00 |
| Venous thrombosis | 2.0 | 1.9 | 0.01 | 1.9 | 1.7 | 0.02 | 2.0 | 1.8 | 0.01 |
| **Medical history: Neoplasms** | | | | | | | | | |
| Hematologic neoplasm | 0.9 | 1.2 | -0.03 | 0.9 | 1.1 | -0.03 | 1.0 | 1.2 | -0.02 |
| Malignant lymphoma | 0.6 | 0.7 | -0.02 | 0.6 | 0.6 | 0.00 | 0.6 | 0.7 | 0.00 |
| Malignant neoplasm of anorectum | 0.2 | 0.2 | 0.00 | 0.2 | 0.2 | 0.01 | 0.2 | 0.2 | 0.01 |
| Malignant neoplastic disease | 14.2 | 14.5 | -0.01 | 14.7 | 14.2 | 0.02 | 14.6 | 14.3 | 0.01 |
| Malignant tumor of breast | 3.0 | 2.0 | 0.06 | 3.0 | 2.8 | 0.01 | 2.5 | 2.3 | 0.01 |
| Malignant tumor of colon | 0.7 | 0.7 | 0.00 | 0.7 | 0.5 | 0.03 | 0.7 | 0.6 | 0.01 |
| Malignant tumor of lung | 0.4 | 0.5 | -0.01 | 0.4 | 0.4 | 0.01 | 0.6 | 0.4 | 0.02 |
| Malignant tumor of urinary bladder | 0.6 | 0.6 | 0.00 | 0.6 | 0.5 | 0.01 | 0.6 | 0.5 | 0.01 |
| Primary malignant neoplasm of prostate | 3.1 | 3.5 | -0.02 | 3.4 | 3.0 | 0.02 | 3.7 | 3.3 | 0.02 |

| (Continued) | Before ajustment | | | After matching | | | After stratification | | |
|---|---|---|---|---|---|---|---|---|---|
| Characteristic | T (%) | C (%) | StdDiff | T (%) | C (%) | StdDiff | T (%) | C (%) | StdDiff |
| **Medication use** | | | | | | | | | |
| Agents acting on the renin-angiotensin system | 76.2 | 78.1 | -0.05 | 76.5 | 77.1 | -0.01 | 78.1 | 77.8 | 0.01 |
| Antibacterials for systemic use | 59.9 | 56.6 | 0.07 | 58.5 | 60.0 | -0.03 | 57.4 | 58.0 | -0.01 |
| Antidepressants | 38.5 | 28.2 | 0.22 | 35.6 | 37.7 | -0.04 | 30.5 | 32.1 | -0.03 |
| Antiepileptics | 24.8 | 19.1 | 0.14 | 23.2 | 24.0 | -0.02 | 20.9 | 21.2 | -0.01 |
| Antiinflammatory and antirheumatic products | 33.6 | 29.6 | 0.09 | 32.3 | 34.0 | -0.04 | 30.3 | 31.4 | -0.02 |
| Antineoplastic agents | 7.3 | 6.4 | 0.04 | 7.1 | 7.0 | 0.00 | 6.9 | 6.7 | 0.01 |
| Antipsoriatics | 0.7 | 0.5 | 0.03 | 0.7 | 0.6 | 0.01 | 0.8 | 0.6 | 0.03 |
| Antithrombotic agents | 20.0 | 24.6 | -0.11 | 20.5 | 20.2 | 0.01 | 23.7 | 23.2 | 0.01 |
| Beta blocking agents | 41.5 | 47.2 | -0.11 | 42.0 | 41.1 | 0.02 | 45.9 | 45.3 | 0.01 |
| Calcium channel blockers | 32.7 | 33.4 | -0.02 | 32.8 | 33.0 | 0.00 | 33.2 | 33.3 | 0.00 |
| Diuretics | 53.4 | 49.2 | 0.09 | 51.5 | 52.2 | -0.01 | 50.1 | 50.6 | -0.01 |
| Drugs for acid related disorders | 35.9 | 32.3 | 0.08 | 34.9 | 35.5 | -0.01 | 32.9 | 33.6 | -0.01 |
| Drugs for obstructive airway diseases | 47.8 | 42.0 | 0.12 | 46.1 | 47.6 | -0.03 | 43.9 | 44.3 | -0.01 |
| Immunosuppressants | 3.6 | 3.2 | 0.02 | 3.5 | 3.8 | -0.01 | 3.2 | 3.5 | -0.01 |
| Lipid modifying agents | 82.3 | 85.8 | -0.10 | 83.3 | 83.2 | 0.00 | 85.2 | 84.9 | 0.01 |
| Opioids | 28.2 | 26.3 | 0.04 | 26.7 | 28.3 | -0.04 | 26.1 | 27.4 | -0.03 |
| Psycholeptics | 26.8 | 23.7 | 0.07 | 25.3 | 26.3 | -0.02 | 23.7 | 24.9 | -0.03 |
| Psychostimulants | 1.8 | 1.1 | 0.06 | 1.5 | 1.4 | 0.00 | 1.4 | 1.3 | 0.01 |

##### 2.1.2 MDCR

**Supplementary Table 3:** Baseline patient characteristics for GLP1RA (T) and SGLT2I (C) new-users in the MDCR data source. We report proportion of initiators satisfying selected base-line characteristics and the standardized difference of population proportions (StdDiff) before and after propensity score adjustment. Less extreme StdDiffs through matching and stratification suggest improved balance between patient cohorts through adjustment.

| | Before adjustment | | | After matching | | | After stratification | | |
|---|---|---|---|---|---|---|---|---|---|
| Characteristic | T (%) | C (%) | StdDiff | T (%) | C (%) | StdDiff | T (%) | C (%) | StdDiff |
| **Age group** | | | | | | | | | |
| 65 - 69 | 44.0 | 39.3 | 0.10 | 41.9 | 41.5 | 0.01 | 40.9 | 40.0 | 0.02 |
| 70 - 74 | 36.6 | 35.6 | 0.02 | 36.5 | 35.9 | 0.01 | 35.9 | 35.7 | 0.00 |
| 75 - 79 | 13.3 | 17.3 | -0.11 | 13.7 | 15.2 | -0.04 | 15.0 | 16.6 | -0.04 |
| 80 - 84 | 4.6 | 5.6 | -0.04 | 6.0 | 5.3 | 0.03 | 5.7 | 5.5 | 0.01 |
| 85 - 89 | 1.1 | 1.8 | -0.06 | 1.5 | 1.7 | -0.02 | 1.9 | 1.7 | 0.01 |
| 90 - 94 | 0.3 | 0.4 | -0.01 | 0.5 | 0.4 | 0.02 | 0.5 | 0.4 | 0.02 |
| Gender: female | 55.2 | 41.5 | 0.28 | 50.9 | 53.7 | -0.06 | 43.3 | 45.7 | -0.05 |
| **Medical history: General** | | | | | | | | | |
| Acute respiratory disease | 22.8 | 22.8 | 0.00 | 22.8 | 24.7 | -0.04 | 21.7 | 23.4 | -0.04 |
| Attention deficit hyperactivity disorder | 0.3 | 0.2 | 0.03 | 0.4 | 0.2 | 0.04 | <0.3 | 0.2 | 0.01 |
| Chronic liver disease | 1.9 | 1.9 | 0.00 | 1.6 | 2.1 | -0.04 | 1.5 | 2.0 | -0.04 |
| Chronic obstructive lung disease | 10.9 | 9.9 | 0.03 | 10.4 | 10.9 | -0.01 | 10.1 | 10.2 | 0.00 |
| Crohn's disease | 0.3 | 0.3 | -0.01 | <0.4 | 0.2 | -0.01 | <0.3 | 0.3 | -0.01 |
| Dementia | 1.4 | 1.2 | 0.01 | 1.9 | 1.6 | 0.02 | 1.7 | 1.3 | 0.03 |
| Depressive disorder | 13.1 | 9.3 | 0.12 | 14.7 | 15.1 | -0.01 | 11.4 | 11.4 | 0.00 |
| Gastroesophageal reflux disease | 14.8 | 17.3 | -0.07 | 17.9 | 20.4 | -0.06 | 16.4 | 18.5 | -0.06 |
| Gastrointestinal hemorrhage | 2.3 | 2.6 | -0.02 | 1.7 | 2.8 | -0.07 | 1.7 | 2.7 | -0.06 |
| Human immunodeficiency virus infection | <0.2 | <0.1 | -0.02 | <0.4 | <0.1 | -0.01 | <0.3 | <0.1 | -0.01 |
| Hyperlipidemia | 67.7 | 81.3 | -0.32 | 81.6 | 81.2 | 0.01 | 81.7 | 81.4 | 0.01 |
| Hypertensive disorder | 76.4 | 83.8 | -0.18 | 82.6 | 84.1 | -0.04 | 83.7 | 83.9 | 0.00 |
| Lesion of liver | 0.7 | 1.2 | -0.05 | 0.8 | 1.2 | -0.04 | 0.8 | 1.2 | -0.04 |
| Obesity | 28.7 | 23.7 | 0.11 | 33.9 | 38.1 | -0.09 | 25.6 | 28.8 | -0.07 |
| Osteoarthritis | 37.3 | 31.8 | 0.12 | 38.5 | 39.4 | -0.02 | 33.8 | 34.4 | -0.01 |
| Pneumonia | 3.5 | 3.7 | -0.01 | 3.7 | 4.2 | -0.03 | 3.6 | 3.9 | -0.01 |
| Psoriasis | 2.8 | 2.3 | 0.03 | 3.2 | 2.4 | 0.05 | 3.2 | 2.3 | 0.05 |
| Renal impairment | 11.2 | 11.9 | -0.02 | 13.6 | 13.0 | 0.02 | 13.4 | 12.3 | 0.03 |
| Rheumatoid arthritis | 1.8 | 1.5 | 0.02 | 1.9 | 1.8 | 0.01 | 1.6 | 1.6 | 0.00 |
| Schizophrenia | <0.2 | <0.1 | 0.02 | <0.4 | <0.1 | 0.02 | <0.3 | <0.1 | 0.00 |
| Ulcerative colitis | 0.2 | 0.5 | -0.04 | 0.4 | 0.4 | -0.01 | 0.3 | 0.4 | -0.01 |
| Urinary tract infectious disease | 10.0 | 7.1 | 0.10 | 10.3 | 9.0 | 0.04 | 8.7 | 7.7 | 0.04 |
| Viral hepatitis C | <0.2 | 0.5 | -0.06 | <0.4 | 0.5 | -0.04 | <0.3 | 0.4 | -0.04 |
| Visual system disorder | 53.2 | 55.1 | -0.04 | 55.5 | 56.4 | -0.02 | 55.2 | 55.6 | -0.01 |
| **Medical history: Cardiovascular disease** | | | | | | | | | |
| Atrial fibrillation | 8.5 | 12.1 | -0.12 | 9.3 | 10.5 | -0.04 | 10.9 | 11.5 | -0.02 |
| Cerebrovascular disease | 9.6 | 11.0 | -0.04 | 10.0 | 10.6 | -0.02 | 10.2 | 10.8 | -0.02 |
| Coronary arteriosclerosis | 21.9 | 28.9 | -0.16 | 23.5 | 22.7 | 0.02 | 27.9 | 26.8 | 0.02 |
| Heart disease | 40.2 | 47.4 | -0.15 | 41.9 | 42.6 | -0.01 | 46.3 | 45.7 | 0.01 |
| Heart failure | 7.3 | 9.9 | -0.09 | 7.8 | 9.8 | -0.07 | 8.3 | 9.8 | -0.05 |
| Ischemic heart disease | 9.3 | 12.0 | -0.09 | 9.3 | 10.5 | -0.04 | 9.8 | 11.5 | -0.06 |
| Peripheral vascular disease | 11.5 | 13.8 | -0.07 | 12.1 | 13.8 | -0.05 | 12.0 | 13.8 | -0.05 |
| Pulmonary embolism | 0.8 | 0.9 | 0.00 | 0.8 | 1.2 | -0.04 | 0.7 | 0.9 | -0.03 |
| Venous thrombosis | 1.8 | 2.0 | -0.01 | 1.9 | 2.1 | -0.01 | 1.7 | 2.0 | -0.02 |
| **Medical history: Neoplasms** | | | | | | | | | |
| Hematologic neoplasm | 1.5 | 1.4 | 0.01 | 1.9 | 1.3 | 0.05 | 1.7 | 1.4 | 0.03 |
| Malignant lymphoma | 0.7 | 1.1 | -0.03 | 0.9 | 1.1 | -0.03 | 0.8 | 1.1 | -0.03 |
| Malignant neoplasm of anorectum | <0.2 | 0.3 | -0.04 | <0.4 | 0.2 | -0.02 | <0.3 | 0.2 | -0.03 |
| Malignant neoplastic disease | 17.2 | 16.9 | 0.01 | 18.3 | 16.4 | 0.05 | 19.0 | 16.7 | 0.06 |
| Malignant tumor of breast | 3.7 | 2.5 | 0.07 | 3.1 | 3.3 | -0.01 | 2.7 | 2.9 | -0.01 |
| Malignant tumor of colon | 0.3 | 0.6 | -0.04 | <0.4 | 0.6 | -0.05 | 0.4 | 0.6 | -0.04 |
| Malignant tumor of lung | 0.5 | 0.3 | 0.02 | 0.6 | 0.4 | 0.03 | 0.7 | 0.4 | 0.05 |
| Malignant tumor of urinary bladder | 1.4 | 1.0 | 0.04 | 1.7 | 0.8 | 0.08 | 1.8 | 0.9 | 0.07 |
| Primary malignant neoplasm of prostate | 3.9 | 4.3 | -0.02 | 4.5 | 3.4 | 0.05 | 5.3 | 4.0 | 0.06 |
| **Medication use** | | | | | | | | | |
| Agents acting on the renin-angiotensin system | 76.4 | 76.2 | 0.00 | 75.5 | 75.1 | 0.01 | 76.5 | 75.8 | 0.02 |
| Antibacterials for systemic use | 63.4 | 60.6 | 0.06 | 62.2 | 63.8 | -0.03 | 60.3 | 61.7 | -0.03 |
| Antidepressants | 35.7 | 24.2 | 0.25 | 33.1 | 36.3 | -0.07 | 26.3 | 28.5 | -0.05 |
| Antiepileptics | 19.1 | 15.8 | 0.09 | 19.0 | 20.0 | -0.03 | 16.8 | 17.3 | -0.01 |
| Antiinflammatory and antirheumatic products | 34.3 | 29.5 | 0.10 | 32.8 | 35.0 | -0.05 | 29.1 | 31.2 | -0.04 |

| (Continued) | Before ajustment | | | After matching | | | After stratification | | |
|---|---|---|---|---|---|---|---|---|---|
| Characteristic | T (%) | C (%) | StdDiff | T (%) | C (%) | StdDiff | T (%) | C (%) | StdDiff |
| Antineoplastic agents | 10.1 | 7.9 | 0.07 | 9.3 | 9.1 | 0.00 | 9.1 | 8.3 | 0.03 |
| Antipsoriatics | 0.9 | 0.8 | 0.02 | 0.7 | 0.7 | 0.00 | 0.7 | 0.8 | 0.00 |
| Antithrombotic agents | 22.4 | 25.1 | -0.06 | 24.8 | 24.3 | 0.01 | 24.8 | 24.3 | 0.01 |
| Beta blocking agents | 44.8 | 48.5 | -0.07 | 45.4 | 44.7 | 0.01 | 48.8 | 47.3 | 0.03 |
| Calcium channel blockers | 31.4 | 32.8 | -0.03 | 29.8 | 31.5 | -0.04 | 31.0 | 32.4 | -0.03 |
| Diuretics | 54.7 | 48.8 | 0.12 | 51.3 | 51.8 | -0.01 | 49.2 | 49.8 | -0.01 |
| Drugs for acid related disorders | 35.1 | 33.8 | 0.03 | 34.7 | 37.2 | -0.05 | 32.8 | 34.9 | -0.04 |
| Drugs for obstructive airway diseases | 49.6 | 44.1 | 0.11 | 48.9 | 48.1 | 0.02 | 45.6 | 45.6 | 0.00 |
| Immunosuppressants | 4.5 | 3.6 | 0.05 | 5.2 | 4.7 | 0.03 | 4.6 | 3.9 | 0.03 |
| Lipid modifying agents | 82.5 | 83.9 | -0.04 | 83.3 | 82.1 | 0.03 | 83.8 | 83.3 | 0.01 |
| Opioids | 28.3 | 24.9 | 0.08 | 26.8 | 28.4 | -0.04 | 24.7 | 26.3 | -0.04 |
| Psycholeptics | 26.5 | 23.0 | 0.08 | 24.3 | 26.5 | -0.05 | 21.9 | 24.2 | -0.06 |
| Psychostimulants | 1.8 | 1.5 | 0.03 | 1.9 | 2.3 | -0.03 | 1.5 | 1.8 | -0.02 |

##### 2.1.3 Open Claims

**Supplementary Table 4:** Baseline patient characteristics for GLP1RA (T) and SGLT2I (C) new-users in the Open Claims data source. We report proportion of initiators satisfying selected base-line characteristics and the standardized difference of population proportions (StdDiff) before and after propensity score adjustment. Less extreme StdDiffs through matching and stratification suggest improved balance between patient cohorts through adjustment.

| | Before adjustment | | | After matching | | | After stratification | | |
|---|---|---|---|---|---|---|---|---|---|
| Characteristic | T (%) | C (%) | StdDiff | T (%) | C (%) | StdDiff | T (%) | C (%) | StdDiff |
| **Age group** | | | | | | | | | |
| 65 - 69 | 49.1 | 43.1 | 0.12 | 44.9 | 44.9 | 0.00 | 44.9 | 44.9 | 0.00 |
| 70 - 74 | 29.8 | 29.2 | 0.01 | 29.9 | 30.1 | 0.00 | 29.4 | 29.5 | 0.00 |
| 75 - 79 | 14.2 | 17.5 | -0.09 | 14.8 | 14.7 | 0.00 | 16.6 | 16.4 | 0.00 |
| 80 - 84 | 6.4 | 9.5 | -0.12 | 6.8 | 6.7 | 0.00 | 8.6 | 8.5 | 0.00 |
| 85 - 89 | 0.5 | 0.8 | -0.03 | 0.5 | 0.5 | 0.00 | 0.6 | 0.7 | -0.01 |
| Gender: female | 57.3 | 44.7 | 0.25 | 54.7 | 55.3 | -0.01 | 48.3 | 48.8 | -0.01 |
| **Medical history: General** | | | | | | | | | |
| Acute respiratory disease | 11.8 | 11.5 | 0.01 | 11.3 | 11.5 | -0.01 | 11.7 | 11.7 | 0.00 |
| Attention deficit hyperactivity disorder | 0.4 | 0.2 | 0.03 | 0.3 | 0.3 | 0.00 | 0.3 | 0.3 | 0.00 |
| Chronic liver disease | 0.9 | 0.9 | -0.01 | 0.9 | 0.9 | 0.00 | 0.9 | 0.9 | 0.00 |
| Chronic obstructive lung disease | 7.1 | 7.2 | 0.00 | 6.9 | 7.1 | -0.01 | 7.1 | 7.3 | 0.00 |
| Crohn's disease | 0.1 | 0.2 | 0.00 | 0.1 | 0.2 | -0.01 | 0.1 | 0.2 | -0.01 |
| Dementia | 0.8 | 1.0 | -0.02 | 0.9 | 0.9 | 0.00 | 1.0 | 1.0 | 0.00 |
| Depressive disorder | 8.8 | 6.0 | 0.10 | 7.7 | 8.0 | -0.01 | 7.0 | 6.9 | 0.00 |
| Gastroesophageal reflux disease | 10.0 | 8.9 | 0.04 | 9.4 | 9.6 | -0.01 | 9.4 | 9.3 | 0.00 |
| Gastrointestinal hemorrhage | 1.3 | 1.4 | -0.01 | 1.3 | 1.2 | 0.00 | 1.4 | 1.4 | 0.00 |
| Human immunodeficiency virus infection | 0.2 | 0.2 | 0.00 | 0.2 | 0.2 | -0.01 | 0.2 | 0.2 | 0.00 |
| Hyperlipidemia | 46.1 | 48.5 | -0.05 | 45.6 | 45.8 | 0.00 | 47.8 | 47.8 | 0.00 |
| Hypertensive disorder | 53.5 | 54.8 | -0.03 | 52.7 | 53.1 | -0.01 | 54.6 | 54.5 | 0.00 |
| Lesion of liver | 0.8 | 0.8 | 0.00 | 0.8 | 0.8 | 0.00 | 0.8 | 0.8 | 0.00 |
| Obesity | 16.2 | 9.6 | 0.20 | 13.4 | 13.8 | -0.01 | 11.9 | 11.6 | 0.01 |
| Osteoarthritis | 22.1 | 18.0 | 0.10 | 20.5 | 20.9 | -0.01 | 19.4 | 19.3 | 0.00 |
| Pneumonia | 2.5 | 2.8 | -0.01 | 2.5 | 2.5 | 0.00 | 2.6 | 2.7 | 0.00 |
| Psoriasis | 1.2 | 1.0 | 0.03 | 1.2 | 1.1 | 0.00 | 1.1 | 1.1 | 0.01 |
| Renal impairment | 9.4 | 10.4 | -0.03 | 9.2 | 9.7 | -0.02 | 10.0 | 10.2 | -0.01 |
| Rheumatoid arthritis | 1.5 | 1.2 | 0.03 | 1.3 | 1.4 | -0.01 | 1.3 | 1.3 | 0.00 |
| Schizophrenia | 0.1 | 0.1 | 0.00 | 0.1 | 0.2 | 0.00 | 0.1 | 0.2 | 0.00 |
| Urinary tract infectious disease | 6.3 | 5.0 | 0.06 | 5.8 | 5.9 | -0.01 | 5.8 | 5.9 | -0.01 |
| Viral hepatitis C | 0.2 | 0.3 | -0.01 | 0.2 | 0.2 | 0.00 | 0.2 | 0.3 | 0.00 |
| Visual system disorder | 25.2 | 25.0 | 0.00 | 24.8 | 24.9 | 0.00 | 25.0 | 25.1 | 0.00 |
| **Medical history: Cardiovascular disease** | | | | | | | | | |
| Atrial fibrillation | 6.2 | 8.1 | -0.07 | 6.2 | 6.3 | 0.00 | 7.3 | 7.5 | -0.01 |
| Cerebrovascular disease | 3.9 | 4.7 | -0.04 | 3.9 | 4.0 | 0.00 | 4.5 | 4.5 | 0.00 |
| Coronary arteriosclerosis | 11.4 | 16.4 | -0.14 | 11.7 | 12.0 | -0.01 | 14.7 | 15.0 | -0.01 |
| Heart disease | 24.1 | 30.3 | -0.14 | 24.2 | 24.6 | -0.01 | 28.0 | 28.5 | -0.01 |
| Heart failure | 5.0 | 8.6 | -0.14 | 5.0 | 5.2 | -0.01 | 6.8 | 7.6 | -0.03 |
| Ischemic heart disease | 4.6 | 7.2 | -0.11 | 4.7 | 4.8 | -0.01 | 6.1 | 6.4 | -0.01 |
| Peripheral vascular disease | 6.7 | 7.5 | -0.03 | 6.6 | 6.8 | -0.01 | 7.3 | 7.3 | 0.00 |
| Pulmonary embolism | 0.7 | 0.6 | 0.02 | 0.7 | 0.6 | 0.00 | 0.7 | 0.6 | 0.00 |
| Venous thrombosis | 0.9 | 1.0 | -0.01 | 0.9 | 0.9 | 0.00 | 1.0 | 1.0 | 0.00 |
| **Medical history: Neoplasms** | | | | | | | | | |
| Hematologic neoplasm | 0.8 | 0.9 | 0.00 | 0.8 | 0.9 | 0.00 | 0.9 | 0.9 | 0.00 |
| Malignant lymphoma | 0.5 | 0.6 | -0.01 | 0.5 | 0.6 | -0.01 | 0.5 | 0.6 | -0.01 |
| Malignant neoplasm of anorectum | 0.1 | 0.1 | -0.01 | 0.1 | 0.1 | 0.00 | 0.1 | 0.1 | 0.00 |
| Malignant neoplastic disease | 8.7 | 9.1 | -0.02 | 8.5 | 8.8 | -0.01 | 8.8 | 9.0 | -0.01 |
| Malignant tumor of breast | 1.8 | 1.5 | 0.02 | 1.7 | 1.8 | 0.00 | 1.5 | 1.6 | -0.01 |
| Malignant tumor of colon | 0.3 | 0.4 | -0.01 | 0.3 | 0.4 | 0.00 | 0.3 | 0.4 | 0.00 |
| Malignant tumor of lung | 0.3 | 0.3 | -0.01 | 0.3 | 0.3 | 0.00 | 0.3 | 0.3 | 0.00 |
| Malignant tumor of urinary bladder | 0.4 | 0.4 | 0.00 | 0.4 | 0.4 | 0.00 | 0.5 | 0.4 | 0.01 |
| Primary malignant neoplasm of prostate | 1.8 | 2.3 | -0.03 | 1.9 | 1.9 | 0.00 | 2.2 | 2.2 | 0.00 |
| **Medication use** | | | | | | | | | |
| Agents acting on the renin-angiotensin system | 70.2 | 74.4 | -0.10 | 70.9 | 71.0 | 0.00 | 73.0 | 73.2 | 0.00 |
| Antibacterials for systemic use | 59.1 | 55.4 | 0.07 | 57.8 | 58.5 | -0.01 | 56.3 | 56.7 | -0.01 |
| Antidepressants | 37.3 | 27.2 | 0.22 | 34.5 | 35.6 | -0.02 | 30.4 | 30.6 | -0.01 |
| Antiepileptics | 24.1 | 18.9 | 0.13 | 22.4 | 22.9 | -0.01 | 20.6 | 20.6 | 0.00 |
| Antiinflammatory and antirheumatic products | 36.0 | 33.6 | 0.05 | 34.9 | 35.5 | -0.01 | 34.0 | 34.5 | -0.01 |
| Antineoplastic agents | 7.9 | 6.5 | 0.06 | 7.3 | 7.5 | -0.01 | 6.8 | 7.0 | -0.01 |
| Antipsoriatics | 0.8 | 0.7 | 0.01 | 0.7 | 0.8 | 0.00 | 0.7 | 0.7 | 0.00 |

| (Continued) | Before ajustment | | | After matching | | | After stratification | | |
|---|---|---|---|---|---|---|---|---|---|
| Characteristic | T (%) | C (%) | StdDiff | T (%) | C (%) | StdDiff | T (%) | C (%) | StdDiff |
| Antithrombotic agents | 22.2 | 28.7 | -0.15 | 22.7 | 22.6 | 0.00 | 26.4 | 26.6 | 0.00 |
| Beta blocking agents | 41.1 | 47.5 | -0.13 | 41.7 | 41.8 | 0.00 | 45.4 | 45.5 | 0.00 |
| Calcium channel blockers | 31.1 | 33.2 | -0.04 | 31.5 | 31.5 | 0.00 | 32.8 | 32.5 | 0.01 |
| Diuretics | 49.4 | 47.9 | 0.03 | 48.5 | 49.3 | -0.02 | 48.1 | 48.6 | -0.01 |
| Drugs for acid related disorders | 38.9 | 37.1 | 0.04 | 38.0 | 38.6 | -0.01 | 37.6 | 37.8 | 0.00 |
| Drugs for obstructive airway diseases | 44.3 | 39.5 | 0.10 | 42.7 | 43.4 | -0.01 | 41.0 | 41.2 | 0.00 |
| Immunosuppressants | 4.0 | 3.3 | 0.04 | 3.7 | 3.8 | 0.00 | 3.5 | 3.5 | 0.00 |
| Lipid modifying agents | 77.0 | 81.1 | -0.10 | 77.8 | 77.7 | 0.00 | 79.8 | 79.8 | 0.00 |
| Opioids | 26.2 | 22.8 | 0.08 | 24.8 | 25.4 | -0.01 | 23.8 | 24.0 | -0.01 |
| Psycholeptics | 25.2 | 20.9 | 0.10 | 23.7 | 24.3 | -0.01 | 22.2 | 22.4 | 0.00 |
| Psychostimulants | 2.6 | 1.5 | 0.07 | 2.2 | 2.2 | 0.00 | 1.9 | 1.8 | 0.00 |

##### 2.1.4 OptumEHR

**Supplementary Table 5:** Baseline patient characteristics for GLP1RA (T) and SGLT2I (C) new-users in the OptumEHR data source. We report proportion of initiators satisfying selected base-line characteristics and the standardized difference of population proportions (StdDiff) before and after propensity score adjustment. Less extreme StdDiffs through matching and stratification suggest improved balance between patient cohorts through adjustment.

| | Before adjustment | | | After matching | | | After stratification | | |
|---|---|---|---|---|---|---|---|---|---|
| Characteristic | T (%) | C (%) | StdDiff | T (%) | C (%) | StdDiff | T (%) | C (%) | StdDiff |
| **Age group** | | | | | | | | | |
| 65 - 69 | 54.8 | 48.5 | 0.13 | 54.3 | 51.5 | 0.06 | 52.9 | 49.6 | 0.07 |
| 70 - 74 | 29.1 | 29.1 | 0.00 | 29.1 | 30.3 | -0.03 | 28.1 | 29.5 | -0.03 |
| 75 - 79 | 11.7 | 14.5 | -0.08 | 12.0 | 12.5 | -0.01 | 13.5 | 13.8 | -0.01 |
| 80 - 84 | 3.4 | 6.1 | -0.12 | 3.7 | 3.8 | -0.01 | 4.6 | 5.4 | -0.04 |
| 85 - 89 | 0.9 | 1.7 | -0.07 | 1.0 | 1.9 | -0.08 | 1.0 | 1.8 | -0.07 |
| Gender: female | 55.6 | 43.5 | 0.24 | 53.3 | 55.7 | -0.05 | 45.1 | 47.7 | -0.05 |
| **Race** | | | | | | | | | |
| Asian | 0.9 | 2.0 | -0.09 | 1.0 | 1.3 | -0.03 | 1.4 | 1.8 | -0.03 |
| Black or African American | 7.2 | 7.8 | -0.02 | 7.4 | 7.2 | 0.01 | 7.4 | 7.6 | -0.01 |
| White | 86.9 | 83.7 | 0.09 | 86.3 | 86.4 | 0.00 | 85.0 | 84.7 | 0.01 |
| **Ethnicity** | | | | | | | | | |
| Hispanic or Latino | 3.1 | 3.9 | -0.04 | 3.1 | 3.8 | -0.04 | 3.5 | 3.9 | -0.02 |
| Not Hispanic or Latino | 87.8 | 85.7 | 0.06 | 87.3 | 86.7 | 0.02 | 86.7 | 86.1 | 0.02 |
| **Medical history: General** | | | | | | | | | |
| Attention deficit hyperactivity disorder | 0.4 | 0.2 | 0.04 | 0.4 | 0.2 | 0.03 | 0.4 | 0.2 | 0.03 |
| Chronic liver disease | 2.0 | 1.6 | 0.03 | 2.0 | 1.8 | 0.02 | 2.0 | 1.7 | 0.03 |
| Chronic obstructive lung disease | 8.4 | 8.1 | 0.01 | 7.8 | 9.1 | -0.05 | 7.4 | 8.4 | -0.04 |
| Crohn's disease | <0.1 | 0.3 | -0.04 | <0.2 | 0.4 | -0.06 | 0.1 | 0.4 | -0.05 |
| Dementia | 1.0 | 1.2 | -0.01 | 1.0 | 1.3 | -0.03 | 1.0 | 1.2 | -0.03 |
| Depressive disorder | 16.0 | 11.1 | 0.14 | 14.3 | 15.2 | -0.03 | 12.5 | 12.7 | -0.01 |
| Gastroesophageal reflux disease | 20.0 | 18.8 | 0.03 | 19.0 | 20.7 | -0.04 | 18.4 | 19.6 | -0.03 |
| Gastrointestinal hemorrhage | 1.5 | 1.1 | 0.03 | 1.4 | 1.4 | 0.00 | 1.3 | 1.2 | 0.00 |
| Human immunodeficiency virus infection | 0.1 | 0.1 | 0.00 | <0.2 | 0.1 | 0.00 | <0.1 | 0.1 | 0.00 |
| Hyperlipidemia | 71.0 | 73.7 | -0.06 | 70.8 | 69.5 | 0.03 | 73.2 | 72.3 | 0.02 |
| Hypertensive disorder | 72.3 | 74.8 | -0.06 | 72.0 | 72.1 | 0.00 | 74.8 | 74.0 | 0.02 |
| Lesion of liver | 0.9 | 1.1 | -0.02 | 0.9 | 1.2 | -0.03 | 1.0 | 1.1 | -0.01 |
| Obesity | 35.3 | 24.5 | 0.24 | 32.9 | 35.8 | -0.06 | 26.7 | 28.6 | -0.04 |
| Osteoarthritis | 25.4 | 21.6 | 0.09 | 24.4 | 24.7 | -0.01 | 23.1 | 22.8 | 0.01 |
| Pneumonia | 2.3 | 2.0 | 0.02 | 2.1 | 2.2 | 0.00 | 2.2 | 2.1 | 0.01 |
| Psoriasis | 1.9 | 1.2 | 0.05 | 1.8 | 1.4 | 0.03 | 1.9 | 1.3 | 0.05 |
| Renal impairment | 10.8 | 10.0 | 0.02 | 10.5 | 10.8 | -0.01 | 10.6 | 10.3 | 0.01 |
| Rheumatoid arthritis | 1.1 | 1.5 | -0.04 | 1.2 | 1.9 | -0.06 | 1.1 | 1.7 | -0.05 |
| Schizophrenia | <0.1 | <0.1 | -0.01 | <0.2 | <0.1 | 0.00 | <0.1 | <0.1 | 0.00 |
| Ulcerative colitis | 0.3 | 0.3 | 0.00 | 0.4 | 0.3 | 0.01 | 0.5 | 0.3 | 0.02 |
| Urinary tract infectious disease | 5.5 | 4.3 | 0.06 | 5.1 | 5.2 | 0.00 | 4.8 | 4.6 | 0.01 |
| Viral hepatitis C | 0.6 | 0.3 | 0.06 | 0.7 | 0.4 | 0.05 | 0.8 | 0.3 | 0.07 |
| Visual system disorder | 16.1 | 16.0 | 0.00 | 16.0 | 16.5 | -0.02 | 15.3 | 16.3 | -0.03 |
| **Medical history: Cardiovascular disease** | | | | | | | | | |
| Atrial fibrillation | 8.1 | 8.6 | -0.02 | 8.1 | 8.0 | 0.00 | 8.3 | 8.4 | 0.00 |
| Cerebrovascular disease | 5.0 | 6.0 | -0.04 | 5.1 | 5.1 | 0.00 | 5.4 | 5.7 | -0.01 |
| Coronary arteriosclerosis | 17.5 | 24.1 | -0.16 | 18.2 | 17.4 | 0.02 | 22.3 | 21.9 | 0.01 |
| Heart disease | 33.3 | 37.8 | -0.09 | 33.3 | 32.1 | 0.03 | 36.9 | 36.0 | 0.02 |
| Heart failure | 5.8 | 7.2 | -0.06 | 5.7 | 6.6 | -0.04 | 6.4 | 7.0 | -0.02 |
| Ischemic heart disease | 7.3 | 9.6 | -0.08 | 7.5 | 7.4 | 0.00 | 8.7 | 8.9 | -0.01 |
| Peripheral vascular disease | 8.1 | 9.9 | -0.06 | 8.4 | 9.1 | -0.02 | 9.1 | 9.7 | -0.02 |
| Pulmonary embolism | 0.9 | 0.5 | 0.04 | 0.8 | 0.6 | 0.01 | 0.9 | 0.6 | 0.03 |
| Venous thrombosis | 0.9 | 1.3 | -0.04 | 0.9 | 1.5 | -0.05 | 1.1 | 1.4 | -0.03 |
| **Medical history: Neoplasms** | | | | | | | | | |
| Hematologic neoplasm | 1.1 | 1.0 | 0.01 | 1.0 | 1.2 | -0.01 | 1.0 | 1.1 | -0.01 |
| Malignant lymphoma | 0.7 | 0.7 | 0.00 | 0.7 | 0.6 | 0.01 | 0.6 | 0.7 | -0.01 |
| Malignant neoplasm of anorectum | 0.2 | 0.1 | 0.01 | 0.2 | 0.1 | 0.01 | 0.2 | 0.1 | 0.03 |
| Malignant neoplastic disease | 10.3 | 10.2 | 0.00 | 10.5 | 10.3 | 0.01 | 10.5 | 10.3 | 0.01 |
| Malignant tumor of breast | 2.0 | 1.7 | 0.02 | 1.8 | 2.4 | -0.04 | 1.5 | 1.9 | -0.03 |
| Malignant tumor of colon | 0.5 | 0.4 | 0.01 | 0.5 | 0.5 | 0.00 | 0.4 | 0.5 | -0.01 |
| Malignant tumor of lung | <0.1 | 0.4 | -0.07 | <0.2 | 0.4 | -0.09 | <0.1 | 0.4 | -0.08 |
| Malignant tumor of urinary bladder | 0.7 | 0.5 | 0.02 | 0.8 | 0.5 | 0.04 | 0.8 | 0.5 | 0.04 |
| Primary malignant neoplasm of prostate | 2.1 | 2.7 | -0.04 | 2.9 | 2.5 | 0.02 | 2.9 | 2.5 | 0.02 |
| **Medication use** | | | | | | | | | |

| (Continued) | Before ajustment | | | After matching | | | After stratification | | |
|---|---|---|---|---|---|---|---|---|---|
| Characteristic | T (%) | C (%) | StdDiff | T (%) | C (%) | StdDiff | T (%) | C (%) | StdDiff |
| Agents acting on the renin-angiotensin system | 72.0 | 74.2 | -0.05 | 72.5 | 71.7 | 0.02 | 74.6 | 73.3 | 0.03 |
| Antibacterials for systemic use | 43.8 | 40.7 | 0.06 | 42.4 | 42.6 | 0.00 | 41.6 | 41.5 | 0.00 |
| Antidepressants | 35.7 | 26.2 | 0.21 | 33.1 | 35.6 | -0.05 | 28.1 | 29.7 | -0.04 |
| Antiepileptics | 22.0 | 17.0 | 0.12 | 21.1 | 21.6 | -0.01 | 18.5 | 18.7 | 0.00 |
| Antiinflammatory and antirheumatic products | 63.4 | 62.7 | 0.01 | 63.0 | 62.9 | 0.00 | 63.1 | 63.0 | 0.00 |
| Antineoplastic agents | 7.2 | 5.4 | 0.07 | 6.7 | 6.1 | 0.02 | 6.2 | 5.7 | 0.02 |
| Antipsoriatics | 0.4 | 0.4 | 0.01 | 0.4 | 0.5 | -0.02 | 0.4 | 0.4 | 0.00 |
| Antithrombotic agents | 53.2 | 56.0 | -0.06 | 55.3 | 55.3 | 0.00 | 55.3 | 55.3 | 0.00 |
| Beta blocking agents | 43.7 | 47.6 | -0.08 | 44.0 | 44.2 | 0.00 | 46.1 | 46.6 | -0.01 |
| Calcium channel blockers | 29.9 | 31.0 | -0.02 | 29.7 | 30.6 | -0.02 | 29.9 | 30.8 | -0.02 |
| Diuretics | 50.4 | 46.2 | 0.08 | 49.1 | 50.0 | -0.02 | 48.0 | 47.6 | 0.01 |
| Drugs for acid related disorders | 46.6 | 42.8 | 0.08 | 45.3 | 47.5 | -0.04 | 42.7 | 44.5 | -0.04 |
| Drugs for obstructive airway diseases | 47.7 | 42.4 | 0.11 | 46.4 | 47.6 | -0.03 | 43.2 | 44.3 | -0.02 |
| Immunosuppressants | 4.2 | 3.0 | 0.06 | 4.0 | 3.8 | 0.01 | 4.0 | 3.3 | 0.04 |
| Lipid modifying agents | 80.8 | 83.7 | -0.08 | 81.6 | 81.4 | 0.00 | 81.6 | 81.4 | 0.00 |
| Opioids | 28.0 | 24.6 | 0.08 | 26.3 | 27.0 | -0.02 | 25.5 | 25.6 | 0.00 |
| Psycholeptics | 30.4 | 26.1 | 0.10 | 28.6 | 30.1 | -0.03 | 26.6 | 27.6 | -0.02 |
| Psychostimulants | 2.9 | 1.5 | 0.10 | 2.8 | 2.0 | 0.05 | 2.2 | 1.7 | 0.04 |

#### 2.2 GLP1RA vs DPP4I

##### 2.2.1 OptumDOD

**Supplementary Table 6:** Baseline patient characteristics for GLP1RA (T) and DPP4I (C) new-users in the Optum-DOD data source. We report proportion of initiators satisfying selected base-line characteristics and the standardized difference of population proportions (StdDiff) before and after propensity score adjustment. Less extreme StdDiffs through matching and stratification suggest improved balance between patient cohorts through adjustment.

| | Before adjustment | | | After matching | | | After stratification | | |
|---|---|---|---|---|---|---|---|---|---|
| Characteristic | T (%) | C (%) | StdDiff | T (%) | C (%) | StdDiff | T (%) | C (%) | StdDiff |
| **Age group** | | | | | | | | | |
| 65 - 69 | 46.5 | 32.6 | -0.29 | 44.0 | 42.6 | -0.03 | 34.8 | 34.6 | 0.00 |
| 70 - 74 | 33.3 | 30.6 | -0.06 | 33.6 | 34.9 | 0.03 | 30.8 | 31.3 | 0.01 |
| 75 - 79 | 14.7 | 19.4 | 0.13 | 16.1 | 15.9 | 0.00 | 19.1 | 18.7 | -0.01 |
| 80 - 84 | 4.0 | 11.3 | 0.28 | 4.6 | 4.4 | -0.01 | 10.1 | 10.0 | 0.00 |
| 85 - 89 | 1.4 | 5.6 | 0.23 | 1.6 | 1.7 | 0.01 | 5.2 | 4.9 | -0.01 |
| 90 - 94 | 0.1 | 0.5 | 0.07 | 0.1 | 0.5 | 0.07 | <0.1 | 0.5 | 0.08 |
| Gender: female | 57.8 | 53.4 | -0.09 | 56.3 | 58.4 | 0.04 | 55.4 | 54.1 | -0.02 |
| **Medical history: General** | | | | | | | | | |
| Acute respiratory disease | 22.8 | 24.7 | 0.04 | 22.9 | 23.1 | 0.00 | 25.2 | 24.5 | -0.01 |
| Attention deficit hyperactivity disorder | 0.6 | 0.2 | -0.06 | 0.5 | 0.4 | -0.02 | 0.3 | 0.3 | 0.00 |
| Chronic liver disease | 2.0 | 2.8 | 0.05 | 2.1 | 1.8 | -0.02 | 2.0 | 2.6 | 0.04 |
| Chronic obstructive lung disease | 12.7 | 13.0 | 0.01 | 12.3 | 12.9 | 0.02 | 12.8 | 13.1 | 0.01 |
| Crohn’s disease | 0.1 | 0.3 | 0.04 | 0.1 | 0.3 | 0.02 | 0.1 | 0.3 | 0.04 |
| Dementia | 2.1 | 4.6 | 0.14 | 2.2 | 2.5 | 0.02 | 3.9 | 4.2 | 0.02 |
| Depressive disorder | 20.1 | 14.6 | -0.15 | 17.9 | 19.7 | 0.04 | 15.1 | 15.6 | 0.01 |
| Gastroesophageal reflux disease | 24.1 | 21.3 | -0.07 | 23.1 | 24.4 | 0.03 | 23.4 | 21.9 | -0.04 |
| Gastrointestinal hemorrhage | 2.3 | 3.3 | 0.06 | 2.3 | 2.3 | 0.00 | 2.7 | 3.2 | 0.03 |
| Human immunodeficiency virus infection | 0.3 | 0.2 | -0.03 | 0.3 | 0.2 | -0.01 | 0.1 | 0.2 | 0.01 |
| Hyperlipidemia | 84.6 | 85.4 | 0.02 | 84.6 | 85.6 | 0.03 | 86.1 | 85.3 | -0.02 |
| Hypertensive disorder | 86.0 | 87.2 | 0.03 | 85.7 | 87.4 | 0.05 | 88.0 | 87.1 | -0.03 |
| Lesion of liver | 1.6 | 1.4 | -0.02 | 1.6 | 1.4 | -0.02 | 1.0 | 1.4 | 0.03 |
| Obesity | 39.6 | 18.3 | -0.48 | 32.7 | 36.0 | 0.07 | 20.7 | 22.0 | 0.03 |
| Osteoarthritis | 40.0 | 35.2 | -0.10 | 38.3 | 39.8 | 0.03 | 37.8 | 36.1 | -0.04 |
| Pneumonia | 4.1 | 5.5 | 0.06 | 4.1 | 4.2 | 0.01 | 7.7 | 5.2 | -0.10 |
| Psoriasis | 2.3 | 1.5 | -0.06 | 2.3 | 1.9 | -0.03 | 1.8 | 1.6 | -0.01 |
| Renal impairment | 20.5 | 24.9 | 0.10 | 21.0 | 21.7 | 0.01 | 25.7 | 24.4 | -0.03 |
| Rheumatoid arthritis | 2.4 | 2.1 | -0.02 | 2.3 | 2.2 | 0.00 | 3.0 | 2.1 | -0.06 |
| Schizophrenia | 0.2 | 0.3 | 0.03 | 0.2 | 0.3 | 0.02 | 0.1 | 0.3 | 0.04 |
| Ulcerative colitis | 0.4 | 0.5 | 0.01 | 0.4 | 0.5 | 0.02 | 0.6 | 0.5 | -0.01 |
| Urinary tract infectious disease | 11.3 | 15.2 | 0.12 | 11.5 | 12.2 | 0.02 | 17.4 | 14.6 | -0.08 |
| Viral hepatitis C | 0.7 | 0.5 | -0.03 | 0.7 | 0.5 | -0.03 | 0.7 | 0.5 | -0.03 |
| Visual system disorder | 54.2 | 54.3 | 0.00 | 53.8 | 54.8 | 0.02 | 53.7 | 54.4 | 0.01 |
| **Medical history: Cardiovascular disease** | | | | | | | | | |
| Atrial fibrillation | 9.8 | 11.4 | 0.05 | 9.7 | 9.5 | -0.01 | 11.7 | 11.1 | -0.02 |
| Cerebrovascular disease | 8.4 | 10.8 | 0.08 | 8.4 | 7.9 | -0.02 | 10.2 | 10.4 | 0.01 |
| Coronary arteriosclerosis | 20.2 | 23.3 | 0.07 | 20.4 | 20.2 | 0.00 | 24.2 | 22.8 | -0.03 |
| Heart disease | 40.7 | 43.2 | 0.05 | 40.3 | 39.8 | -0.01 | 46.5 | 42.8 | -0.07 |
| Heart failure | 9.8 | 11.4 | 0.05 | 9.5 | 10.0 | 0.02 | 12.4 | 11.1 | -0.04 |
| Ischemic heart disease | 10.1 | 12.2 | 0.07 | 10.0 | 10.6 | 0.02 | 12.9 | 12.0 | -0.03 |
| Peripheral vascular disease | 16.9 | 17.5 | 0.02 | 17.2 | 17.7 | 0.01 | 17.3 | 17.5 | 0.00 |
| Pulmonary embolism | 1.2 | 1.0 | -0.02 | 1.1 | 1.3 | 0.02 | 1.0 | 1.1 | 0.01 |
| Venous thrombosis | 2.0 | 2.2 | 0.02 | 2.0 | 1.6 | -0.03 | 3.1 | 2.2 | -0.06 |
| **Medical history: Neoplasms** | | | | | | | | | |
| Hematologic neoplasm | 0.9 | 1.7 | 0.07 | 0.9 | 1.6 | 0.06 | 1.1 | 1.6 | 0.05 |
| Malignant lymphoma | 0.6 | 0.8 | 0.03 | 0.6 | 0.7 | 0.02 | 0.6 | 0.8 | 0.03 |
| Malignant neoplasm of anorectum | 0.2 | 0.3 | 0.02 | 0.2 | 0.1 | -0.02 | 0.1 | 0.2 | 0.03 |
| Malignant neoplastic disease | 14.2 | 16.1 | 0.05 | 14.6 | 14.9 | 0.01 | 14.9 | 15.8 | 0.03 |
| Malignant tumor of breast | 3.0 | 2.8 | -0.01 | 3.0 | 3.0 | 0.00 | 2.5 | 2.8 | 0.02 |
| Malignant tumor of colon | 0.7 | 0.9 | 0.01 | 0.8 | 0.5 | -0.03 | 0.7 | 0.8 | 0.01 |
| Malignant tumor of lung | 0.4 | 0.6 | 0.03 | 0.4 | 0.4 | 0.00 | 0.9 | 0.6 | -0.03 |
| Malignant tumor of urinary bladder | 0.6 | 0.8 | 0.03 | 0.6 | 0.6 | 0.00 | 0.6 | 0.6 | 0.00 |
| Primary malignant neoplasm of prostate | 3.1 | 3.9 | 0.04 | 3.4 | 3.0 | -0.02 | 4.2 | 3.7 | -0.02 |
| **Medication use** | | | | | | | | | |
| Agents acting on the renin-angiotensin system | 76.2 | 77.0 | 0.02 | 76.3 | 76.9 | 0.01 | 78.3 | 76.9 | -0.04 |
| Antibacterials for systemic use | 59.9 | 59.7 | 0.00 | 59.2 | 60.6 | 0.03 | 60.1 | 60.0 | 0.00 |

| (Continued) | Before ajustment | | | After matching | | | After stratification | | |
|---|---|---|---|---|---|---|---|---|---|
| Characteristic | T (%) | C (%) | StdDiff | T (%) | C (%) | StdDiff | T (%) | C (%) | StdDiff |
| Antidepressants | 38.5 | 27.5 | -0.24 | 35.5 | 38.0 | 0.05 | 29.9 | 29.5 | -0.01 |
| Antiepileptics | 24.8 | 18.8 | -0.15 | 23.0 | 23.4 | 0.01 | 20.1 | 19.8 | -0.01 |
| Antiinflammatory and antirheumatic products | 33.6 | 29.3 | -0.09 | 32.2 | 33.1 | 0.02 | 29.0 | 30.1 | 0.02 |
| Antineoplastic agents | 7.3 | 7.3 | 0.00 | 7.1 | 7.3 | 0.01 | 7.1 | 7.3 | 0.01 |
| Antipsoriatics | 0.7 | 0.6 | -0.01 | 0.7 | 0.7 | -0.01 | 0.7 | 0.6 | -0.01 |
| Antithrombotic agents | 20.0 | 22.5 | 0.06 | 19.8 | 19.8 | 0.00 | 22.2 | 22.1 | 0.00 |
| Beta blocking agents | 41.5 | 45.5 | 0.08 | 41.5 | 41.9 | 0.01 | 47.7 | 45.0 | -0.06 |
| Calcium channel blockers | 32.7 | 35.4 | 0.06 | 32.8 | 33.1 | 0.01 | 34.3 | 35.0 | 0.02 |
| Diuretics | 53.4 | 49.3 | -0.08 | 51.8 | 52.9 | 0.02 | 53.4 | 50.0 | -0.07 |
| Drugs for acid related disorders | 35.9 | 34.4 | -0.03 | 35.3 | 36.7 | 0.03 | 35.6 | 34.7 | -0.02 |
| Drugs for obstructive airway diseases | 47.8 | 41.3 | -0.13 | 46.2 | 47.1 | 0.02 | 43.6 | 42.6 | -0.02 |
| Immunosuppressants | 3.6 | 3.1 | -0.03 | 3.2 | 3.2 | 0.00 | 3.3 | 3.1 | -0.01 |
| Lipid modifying agents | 82.3 | 81.8 | -0.01 | 82.1 | 83.9 | 0.05 | 80.3 | 82.1 | 0.04 |
| Opioids | 28.2 | 27.1 | -0.02 | 27.2 | 27.9 | 0.02 | 26.8 | 27.3 | 0.01 |
| Psycholeptics | 26.8 | 24.2 | -0.06 | 25.6 | 26.4 | 0.02 | 22.2 | 24.6 | 0.06 |
| Psychostimulants | 1.8 | 1.0 | -0.07 | 1.6 | 1.5 | 0.00 | 1.3 | 1.1 | -0.02 |

##### 2.2.2 MDCR

**Supplementary Table 7:** Baseline patient characteristics for GLP1RA (T) and DPP4I (C) new-users in the MDCR data source. We report proportion of initiators satisfying selected base-line characteristics and the standardized difference of population proportions (StdDiff) before and after propensity score adjustment. Less extreme StdDiffs through matching and stratification suggest improved balance between patient cohorts through adjustment.

| | Before adjustment | | | After matching | | | After stratification | | |
|---|---|---|---|---|---|---|---|---|---|
| Characteristic | T (%) | C (%) | StdDiff | T (%) | C (%) | StdDiff | T (%) | C (%) | StdDiff |
| **Age group** | | | | | | | | | |
| 65 - 69 | 44.0 | 31.2 | -0.27 | 43.0 | 39.6 | -0.07 | 38.3 | 32.1 | -0.13 |
| 70 - 74 | 36.6 | 30.5 | -0.13 | 36.1 | 37.6 | 0.03 | 30.4 | 31.2 | 0.02 |
| 75 - 79 | 13.3 | 19.3 | 0.16 | 14.0 | 14.9 | 0.03 | 18.5 | 18.8 | 0.01 |
| 80 - 84 | 4.6 | 11.6 | 0.26 | 5.3 | 4.8 | -0.02 | 8.3 | 11.0 | 0.09 |
| 85 - 89 | 1.1 | 5.4 | 0.25 | 1.3 | 1.2 | -0.01 | 4.4 | 5.1 | 0.03 |
| 90 - 94 | 0.3 | 1.6 | 0.13 | 0.4 | 1.5 | 0.12 | <0.1 | 1.6 | 0.16 |
| Gender: female | 55.2 | 48.1 | -0.14 | 53.9 | 54.8 | 0.02 | 43.9 | 48.7 | 0.10 |
| **Medical history: General** | | | | | | | | | |
| Acute respiratory disease | 22.8 | 23.1 | 0.01 | 23.3 | 21.9 | -0.03 | 23.0 | 23.1 | 0.00 |
| Attention deficit hyperactivity disorder | 0.3 | 0.2 | -0.03 | <0.2 | 0.4 | 0.03 | <0.1 | 0.2 | 0.02 |
| Chronic liver disease | 1.9 | 2.3 | 0.03 | 2.1 | 2.0 | -0.01 | 2.4 | 2.3 | 0.00 |
| Chronic obstructive lung disease | 10.9 | 10.5 | -0.01 | 10.8 | 10.2 | -0.02 | 11.2 | 10.5 | -0.02 |
| Crohn's disease | 0.3 | 0.3 | 0.00 | 0.2 | 0.3 | 0.01 | 0.2 | 0.3 | 0.01 |
| Dementia | 1.4 | 2.9 | 0.10 | 1.5 | 2.1 | 0.04 | 1.6 | 2.8 | 0.08 |
| Depressive disorder | 13.1 | 7.7 | -0.18 | 12.1 | 12.6 | 0.02 | 8.3 | 8.1 | -0.01 |
| Gastroesophageal reflux disease | 14.8 | 13.0 | -0.05 | 14.6 | 17.5 | 0.08 | 9.6 | 13.3 | 0.12 |
| Gastrointestinal hemorrhage | 2.3 | 3.2 | 0.05 | 2.1 | 2.9 | 0.05 | 2.3 | 3.1 | 0.05 |
| Human immunodeficiency virus infection | <0.2 | 0.1 | 0.01 | <0.2 | <0.3 | 0.03 | <0.1 | 0.1 | 0.03 |
| Hyperlipidemia | 67.7 | 63.3 | -0.09 | 69.3 | 70.4 | 0.03 | 62.0 | 64.1 | 0.04 |
| Hypertensive disorder | 76.4 | 75.0 | -0.03 | 77.7 | 78.6 | 0.02 | 80.7 | 75.3 | -0.13 |
| Lesion of liver | 0.7 | 0.8 | 0.01 | 0.8 | 1.2 | 0.04 | 0.3 | 0.8 | 0.07 |
| Obesity | 28.7 | 10.0 | -0.48 | 24.7 | 26.6 | 0.04 | 11.7 | 11.8 | 0.00 |
| Osteoarthritis | 37.3 | 30.4 | -0.15 | 35.9 | 34.5 | -0.03 | 30.2 | 31.0 | 0.02 |
| Pneumonia | 3.5 | 4.9 | 0.07 | 3.6 | 2.8 | -0.04 | 3.2 | 4.8 | 0.08 |
| Psoriasis | 2.8 | 1.7 | -0.07 | 2.7 | 2.0 | -0.05 | 1.9 | 1.7 | -0.01 |
| Renal impairment | 11.2 | 14.0 | 0.09 | 11.8 | 13.4 | 0.05 | 12.2 | 13.9 | 0.05 |
| Rheumatoid arthritis | 1.8 | 1.8 | 0.00 | 1.7 | 1.5 | -0.01 | 1.5 | 1.8 | 0.03 |
| Schizophrenia | <0.2 | 0.1 | -0.01 | 0.2 | 0.1 | -0.03 | 0.2 | 0.1 | -0.03 |
| Ulcerative colitis | 0.2 | 0.5 | 0.05 | 0.3 | 0.5 | 0.04 | 0.2 | 0.5 | 0.05 |
| Urinary tract infectious disease | 10.0 | 10.7 | 0.02 | 10.1 | 9.1 | -0.03 | 9.1 | 10.5 | 0.05 |
| Viral hepatitis C | <0.2 | 0.3 | 0.03 | <0.2 | 0.5 | 0.06 | <0.1 | 0.3 | 0.04 |
| Visual system disorder | 53.2 | 52.4 | -0.02 | 53.7 | 54.7 | 0.02 | 56.2 | 52.6 | -0.07 |
| **Medical history: Cardiovascular disease** | | | | | | | | | |
| Atrial fibrillation | 8.5 | 12.1 | 0.12 | 8.4 | 9.3 | 0.03 | 11.0 | 11.8 | 0.02 |
| Cerebrovascular disease | 9.6 | 12.7 | 0.10 | 9.8 | 9.9 | 0.00 | 12.0 | 12.4 | 0.01 |
| Coronary arteriosclerosis | 21.9 | 25.2 | 0.08 | 21.8 | 22.4 | 0.01 | 26.7 | 24.8 | -0.04 |
| Heart disease | 40.2 | 45.5 | 0.11 | 40.2 | 41.7 | 0.03 | 46.5 | 45.0 | -0.03 |
| Heart failure | 7.3 | 9.4 | 0.08 | 6.9 | 8.0 | 0.04 | 9.0 | 9.2 | 0.01 |
| Ischemic heart disease | 9.3 | 11.2 | 0.06 | 9.0 | 9.8 | 0.03 | 10.6 | 11.0 | 0.01 |
| Peripheral vascular disease | 11.5 | 12.8 | 0.04 | 11.9 | 12.9 | 0.03 | 11.5 | 12.8 | 0.04 |
| Pulmonary embolism | 0.8 | 0.9 | 0.01 | 0.8 | 1.1 | 0.03 | 1.5 | 0.9 | -0.05 |
| Venous thrombosis | 1.8 | 2.3 | 0.03 | 1.9 | 2.1 | 0.02 | 2.0 | 2.2 | 0.01 |
| **Medical history: Neoplasms** | | | | | | | | | |
| Hematologic neoplasm | 1.5 | 1.8 | 0.03 | 1.5 | 2.1 | 0.05 | 1.3 | 1.8 | 0.04 |
| Malignant lymphoma | 0.7 | 1.0 | 0.03 | 0.6 | 0.8 | 0.03 | 0.5 | 1.0 | 0.05 |
| Malignant neoplasm of anorectum | <0.2 | 0.4 | 0.06 | <0.2 | 0.5 | 0.07 | <0.1 | 0.4 | 0.06 |
| Malignant neoplastic disease | 17.2 | 19.2 | 0.05 | 17.7 | 20.1 | 0.06 | 17.3 | 19.2 | 0.05 |
| Malignant tumor of breast | 3.7 | 2.9 | -0.04 | 3.7 | 3.6 | 0.00 | 2.3 | 3.0 | 0.04 |
| Malignant tumor of colon | 0.3 | 0.9 | 0.08 | 0.3 | 0.8 | 0.06 | 0.1 | 0.9 | 0.11 |
| Malignant tumor of lung | 0.5 | 0.6 | 0.02 | 0.6 | 0.6 | 0.01 | 0.7 | 0.6 | 0.00 |
| Malignant tumor of urinary bladder | 1.4 | 1.2 | -0.01 | 1.5 | 1.2 | -0.03 | 1.6 | 1.2 | -0.03 |
| Primary malignant neoplasm of prostate | 3.9 | 4.5 | 0.03 | 4.1 | 4.4 | 0.02 | 5.9 | 4.4 | -0.07 |
| **Medication use** | | | | | | | | | |
| Agents acting on the renin-angiotensin system | 76.4 | 75.8 | -0.01 | 77.0 | 76.0 | -0.02 | 78.0 | 75.8 | -0.05 |
| Antibacterials for systemic use | 63.4 | 62.5 | -0.02 | 64.0 | 64.2 | 0.00 | 58.4 | 62.4 | 0.08 |
| Antidepressants | 35.7 | 24.9 | -0.24 | 34.0 | 36.4 | 0.05 | 25.5 | 25.9 | 0.01 |
| Antiepileptics | 19.1 | 14.6 | -0.12 | 18.0 | 19.5 | 0.04 | 14.2 | 15.0 | 0.02 |
| Antiinflammatory and antirheumatic products | 34.3 | 28.1 | -0.14 | 33.4 | 30.9 | -0.05 | 26.4 | 28.5 | 0.05 |

| (Continued) | Before ajustment | | | After matching | | | After stratification | | |
|---|---|---|---|---|---|---|---|---|---|
| Characteristic | T (%) | C (%) | StdDiff | T (%) | C (%) | StdDiff | T (%) | C (%) | StdDiff |
| Antineoplastic agents | 10.1 | 8.7 | -0.05 | 9.7 | 9.8 | 0.00 | 8.0 | 8.8 | 0.03 |
| Antipsoriatics | 0.9 | 0.8 | -0.02 | 0.8 | 0.7 | -0.01 | 0.7 | 0.7 | 0.01 |
| Antithrombotic agents | 22.4 | 25.4 | 0.07 | 22.4 | 23.7 | 0.03 | 26.1 | 25.1 | -0.02 |
| Beta blocking agents | 44.8 | 49.7 | 0.10 | 45.4 | 44.1 | -0.03 | 53.1 | 49.3 | -0.08 |
| Calcium channel blockers | 31.4 | 35.0 | 0.08 | 31.6 | 33.2 | 0.04 | 36.1 | 34.7 | -0.03 |
| Diuretics | 54.7 | 51.4 | -0.06 | 53.5 | 53.9 | 0.01 | 52.3 | 51.5 | -0.02 |
| Drugs for acid related disorders | 35.1 | 33.3 | -0.04 | 34.7 | 36.1 | 0.03 | 29.9 | 33.5 | 0.08 |
| Drugs for obstructive airway diseases | 49.6 | 41.5 | -0.16 | 48.8 | 47.6 | -0.03 | 42.6 | 42.3 | -0.01 |
| Immunosuppressants | 4.5 | 3.4 | -0.06 | 4.2 | 3.9 | -0.01 | 3.8 | 3.4 | -0.02 |
| Lipid modifying agents | 82.5 | 81.2 | -0.04 | 82.2 | 82.1 | 0.00 | 82.6 | 81.3 | -0.03 |
| Opioids | 28.3 | 27.5 | -0.02 | 28.5 | 27.0 | -0.03 | 26.7 | 27.5 | 0.02 |
| Psycholeptics | 26.5 | 26.0 | -0.01 | 26.6 | 27.0 | 0.01 | 27.1 | 26.0 | -0.03 |
| Psychostimulants | 1.8 | 1.3 | -0.04 | 1.7 | 1.9 | 0.01 | 0.7 | 1.3 | 0.06 |

##### 2.2.3 Open Claims

**Supplementary Table 8:** Baseline patient characteristics for GLP1RA (T) and DPP4I (C) new-users in the Open Claims data source. We report proportion of initiators satisfying selected base-line characteristics and the standardized difference of population proportions (StdDiff) before and after propensity score adjustment. Less extreme StdDiffs through matching and stratification suggest improved balance between patient cohorts through adjustment.

| | Before adjustment | | | After matching | | | After stratification | | |
|---|---|---|---|---|---|---|---|---|---|
| Characteristic | T (%) | C (%) | StdDiff | T (%) | C (%) | StdDiff | T (%) | C (%) | StdDiff |
| **Age group** | | | | | | | | | |
| 65 - 69 | 49.1 | 36.1 | -0.27 | 45.9 | 46.4 | 0.01 | 39.0 | 38.6 | -0.01 |
| 70 - 74 | 29.8 | 27.5 | -0.05 | 30.1 | 30.0 | 0.00 | 28.2 | 28.1 | 0.00 |
| 75 - 79 | 14.2 | 25.5 | 0.29 | 15.8 | 15.3 | -0.01 | 22.3 | 23.3 | 0.02 |
| 80 - 84 | 6.4 | 10.6 | 0.15 | 7.6 | 7.7 | 0.00 | 10.2 | 9.7 | -0.02 |
| 85 - 89 | 0.5 | 0.3 | -0.04 | 0.5 | 0.6 | 0.01 | 0.3 | 0.3 | 0.00 |
| Gender: female | 57.3 | 53.7 | -0.07 | 55.6 | 55.6 | 0.00 | 54.8 | 54.1 | -0.01 |
| **Medical history: General** | | | | | | | | | |
| Acute respiratory disease | 11.8 | 13.3 | 0.05 | 11.5 | 11.9 | 0.01 | 13.5 | 13.0 | -0.01 |
| Attention deficit hyperactivity disorder | 0.4 | 0.1 | -0.04 | 0.3 | 0.2 | 0.00 | 0.2 | 0.2 | 0.00 |
| Chronic liver disease | 0.9 | 1.4 | 0.05 | 1.3 | 1.2 | -0.01 | 1.3 | 1.2 | -0.01 |
| Chronic obstructive lung disease | 7.1 | 7.8 | 0.03 | 6.9 | 7.2 | 0.01 | 7.7 | 7.8 | 0.00 |
| Crohn’s disease | 0.1 | 0.2 | 0.00 | 0.1 | 0.2 | 0.01 | 0.1 | 0.2 | 0.01 |
| Dementia | 0.8 | 2.3 | 0.12 | 1.0 | 1.1 | 0.01 | 2.0 | 2.0 | 0.00 |
| Depressive disorder | 8.8 | 6.3 | -0.09 | 7.5 | 7.8 | 0.01 | 7.1 | 6.8 | -0.01 |
| Gastroesophageal reflux disease | 10.0 | 9.3 | -0.02 | 9.4 | 9.6 | 0.01 | 9.7 | 9.4 | -0.01 |
| Gastrointestinal hemorrhage | 1.3 | 1.9 | 0.05 | 1.3 | 1.2 | 0.00 | 1.8 | 1.7 | -0.01 |
| Human immunodeficiency virus infection | 0.2 | 0.2 | 0.00 | 0.2 | 0.2 | 0.02 | 0.1 | 0.2 | 0.01 |
| Hyperlipidemia | 46.1 | 48.9 | 0.06 | 45.2 | 45.8 | 0.01 | 45.2 | 45.8 | 0.01 |
| Hypertensive disorder | 53.5 | 57.5 | 0.08 | 52.8 | 53.0 | 0.00 | 56.9 | 56.6 | -0.01 |
| Lesion of liver | 0.8 | 0.9 | 0.01 | 0.8 | 0.8 | 0.00 | 0.8 | 0.8 | 0.01 |
| Obesity | 16.2 | 7.4 | -0.28 | 12.2 | 12.4 | 0.01 | 9.8 | 8.8 | -0.04 |
| Osteoarthritis | 22.1 | 19.8 | -0.06 | 20.2 | 20.5 | 0.01 | 20.5 | 20.0 | -0.01 |
| Pneumonia | 2.5 | 3.4 | 0.05 | 2.5 | 2.6 | 0.00 | 3.3 | 3.2 | 0.00 |
| Psoriasis | 1.2 | 0.8 | -0.04 | 1.1 | 1.1 | -0.01 | 1.0 | 0.9 | -0.01 |
| Renal impairment | 9.4 | 12.9 | 0.11 | 9.7 | 9.8 | 0.00 | 12.1 | 12.3 | 0.01 |
| Rheumatoid arthritis | 1.5 | 1.4 | -0.01 | 1.4 | 1.4 | 0.00 | 1.4 | 1.4 | 0.00 |
| Schizophrenia | 0.1 | 0.2 | 0.01 | 0.2 | 0.2 | 0.01 | 0.2 | 0.2 | 0.01 |
| Ulcerative colitis | 0.2 | 0.2 | 0.00 | 0.2 | 0.3 | 0.01 | 0.2 | 0.3 | 0.01 |
| Urinary tract infectious disease | 6.3 | 8.0 | 0.06 | 6.2 | 6.5 | 0.01 | 7.8 | 7.7 | 0.00 |
| Viral hepatitis C | 0.2 | 0.3 | 0.02 | 0.2 | 0.2 | 0.00 | 0.3 | 0.3 | 0.01 |
| Visual system disorder | 25.2 | 27.0 | 0.04 | 24.8 | 24.9 | 0.00 | 26.7 | 26.6 | 0.00 |
| **Medical history: Cardiovascular disease** | | | | | | | | | |
| Atrial fibrillation | 6.2 | 7.8 | 0.06 | 7.3 | 7.5 | 0.01 | 7.3 | 7.5 | 0.01 |
| Cerebrovascular disease | 3.9 | 6.1 | 0.10 | 4.1 | 4.1 | 0.00 | 5.6 | 5.7 | 0.00 |
| Coronary arteriosclerosis | 11.4 | 14.0 | 0.08 | 11.4 | 11.4 | 0.00 | 13.6 | 13.5 | 0.00 |
| Heart disease | 24.1 | 28.5 | 0.10 | 23.8 | 23.9 | 0.00 | 27.5 | 27.6 | 0.00 |
| Heart failure | 5.0 | 7.0 | 0.08 | 4.9 | 5.1 | 0.01 | 6.2 | 6.6 | 0.01 |
| Ischemic heart disease | 4.6 | 6.2 | 0.07 | 5.5 | 5.8 | 0.01 | 5.5 | 5.8 | 0.01 |
| Peripheral vascular disease | 6.7 | 7.6 | 0.04 | 7.6 | 7.4 | -0.01 | 7.6 | 7.4 | -0.01 |
| Pulmonary embolism | 0.7 | 0.7 | -0.01 | 0.7 | 0.7 | 0.00 | 0.6 | 0.7 | 0.00 |
| Venous thrombosis | 0.9 | 1.3 | 0.04 | 0.9 | 0.9 | 0.00 | 1.2 | 1.2 | 0.00 |
| **Medical history: Neoplasms** | | | | | | | | | |
| Hematologic neoplasm | 0.8 | 1.1 | 0.03 | 0.9 | 0.9 | 0.00 | 1.1 | 1.1 | 0.00 |
| Malignant lymphoma | 0.5 | 0.6 | 0.01 | 0.5 | 0.5 | 0.00 | 0.7 | 0.6 | -0.01 |
| Malignant neoplasm of anorectum | 0.1 | 0.2 | 0.02 | 0.1 | 0.1 | 0.00 | 0.2 | 0.2 | 0.01 |
| Malignant neoplastic disease | 8.7 | 10.5 | 0.06 | 8.8 | 8.9 | 0.00 | 10.4 | 10.1 | -0.01 |
| Malignant tumor of breast | 1.8 | 1.9 | 0.01 | 1.8 | 1.7 | -0.01 | 2.0 | 1.9 | -0.01 |
| Malignant tumor of colon | 0.3 | 0.5 | 0.04 | 0.3 | 0.4 | 0.01 | 0.4 | 0.5 | 0.02 |
| Malignant tumor of lung | 0.3 | 0.5 | 0.03 | 0.3 | 0.3 | 0.01 | 0.4 | 0.4 | 0.00 |
| Malignant tumor of urinary bladder | 0.4 | 0.6 | 0.03 | 0.7 | 0.6 | -0.01 | 0.7 | 0.6 | -0.01 |
| Primary malignant neoplasm of prostate | 1.8 | 2.4 | 0.04 | 1.9 | 2.0 | 0.00 | 2.5 | 2.2 | -0.02 |
| **Medication use** | | | | | | | | | |
| Agents acting on the renin-angiotensin system | 70.2 | 73.7 | 0.08 | 70.7 | 70.8 | 0.00 | 72.6 | 73.0 | 0.01 |
| Antibacterials for systemic use | 59.1 | 58.6 | -0.01 | 57.9 | 57.7 | 0.00 | 58.7 | 58.6 | 0.00 |
| Antidepressants | 37.3 | 27.5 | -0.21 | 33.6 | 34.3 | 0.02 | 30.3 | 29.4 | -0.02 |
| Antiepileptics | 24.1 | 19.7 | -0.10 | 22.2 | 22.7 | 0.01 | 21.2 | 20.5 | -0.02 |
| Antiinflammatory and antirheumatic products | 36.0 | 34.3 | -0.04 | 34.8 | 35.1 | 0.01 | 35.1 | 34.6 | -0.01 |
| Antineoplastic agents | 7.9 | 6.7 | -0.05 | 7.2 | 7.3 | 0.00 | 7.2 | 6.8 | -0.01 |

| (Continued) | Before ajustment | | | After matching | | | After stratification | | |
|---|---|---|---|---|---|---|---|---|---|
| Characteristic | T (%) | C (%) | StdDiff | T (%) | C (%) | StdDiff | T (%) | C (%) | StdDiff |
| Antipsoriatics | 0.8 | 0.8 | 0.00 | 0.8 | 0.7 | 0.00 | 0.8 | 0.7 | 0.00 |
| Antithrombotic agents | 22.2 | 27.4 | 0.12 | 22.7 | 22.4 | -0.01 | 26.0 | 26.3 | 0.01 |
| Beta blocking agents | 41.1 | 45.9 | 0.10 | 41.6 | 41.5 | 0.00 | 44.9 | 44.9 | 0.00 |
| Calcium channel blockers | 31.1 | 34.6 | 0.07 | 31.7 | 32.0 | 0.01 | 33.1 | 33.9 | 0.02 |
| Diuretics | 49.4 | 47.6 | -0.04 | 48.0 | 47.9 | 0.00 | 47.6 | 47.8 | 0.00 |
| Drugs for acid related disorders | 38.9 | 39.4 | 0.01 | 38.2 | 38.5 | 0.01 | 39.4 | 39.4 | 0.00 |
| Drugs for obstructive airway diseases | 44.3 | 39.3 | -0.10 | 42.2 | 42.7 | 0.01 | 40.1 | 40.3 | 0.01 |
| Immunosuppressants | 4.0 | 3.6 | -0.02 | 3.8 | 3.9 | 0.01 | 3.6 | 3.7 | 0.00 |
| Lipid modifying agents | 77.0 | 78.0 | 0.03 | 77.4 | 77.1 | -0.01 | 78.0 | 77.7 | -0.01 |
| Opioids | 26.2 | 25.7 | -0.01 | 24.9 | 25.2 | 0.01 | 26.0 | 25.6 | -0.01 |
| Psycholeptics | 25.2 | 23.1 | -0.05 | 23.5 | 23.9 | 0.01 | 24.0 | 23.4 | -0.01 |
| Psychostimulants | 2.6 | 1.4 | -0.08 | 2.0 | 2.1 | 0.00 | 1.7 | 1.5 | -0.01 |

##### 2.2.4 OptumEHR

**Supplementary Table 9:** Baseline patient characteristics for GLP1RA (T) and DPP4I (C) new-users in the OptumEHR data source. We report proportion of initiators satisfying selected base-line characteristics and the standardized difference of population proportions (StdDiff) before and after propensity score adjustment. Less extreme StdDiffs through matching and stratification suggest improved balance between patient cohorts through adjustment.

| | Before adjustment | | | After matching | | | After stratification | | |
|---|---|---|---|---|---|---|---|---|---|
| Characteristic | T (%) | C (%) | StdDiff | T (%) | C (%) | StdDiff | T (%) | C (%) | StdDiff |
| **Age group** | | | | | | | | | |
| 65 - 69 | 54.8 | 40.1 | -0.30 | 53.5 | 52.2 | -0.03 | 43.2 | 41.9 | -0.03 |
| 70 - 74 | 29.1 | 27.7 | -0.03 | 29.3 | 31.6 | 0.05 | 27.4 | 28.0 | 0.01 |
| 75 - 79 | 11.7 | 18.2 | 0.18 | 12.4 | 11.9 | -0.02 | 16.6 | 17.3 | 0.02 |
| 80 - 84 | 3.4 | 11.0 | 0.30 | 3.8 | 3.3 | -0.03 | 9.9 | 10.0 | 0.00 |
| 85 - 89 | 0.9 | 3.0 | 0.15 | 1.0 | 1.0 | 0.00 | 2.9 | 2.8 | -0.01 |
| Gender: female | 55.6 | 52.0 | -0.07 | 55.0 | 56.1 | 0.02 | 52.3 | 52.4 | 0.00 |
| **Race** | | | | | | | | | |
| Asian | 0.9 | 2.5 | 0.12 | 1.0 | 1.6 | 0.05 | 1.4 | 2.4 | 0.07 |
| Black or African American | 7.2 | 8.9 | 0.06 | 7.5 | 7.7 | 0.01 | 8.9 | 8.8 | -0.01 |
| White | 86.9 | 82.0 | -0.13 | 86.2 | 86.6 | 0.01 | 82.8 | 82.6 | -0.01 |
| **Ethnicity** | | | | | | | | | |
| Hispanic or Latino | 3.1 | 4.4 | 0.06 | 3.2 | 3.1 | -0.01 | 4.1 | 4.3 | 0.01 |
| Not Hispanic or Latino | 87.8 | 87.0 | -0.03 | 87.5 | 87.9 | 0.01 | 86.3 | 87.0 | 0.02 |
| **Medical history: General** | | | | | | | | | |
| Acute respiratory disease | 14.8 | 15.7 | 0.03 | 14.5 | 15.8 | 0.03 | 14.3 | 15.6 | 0.04 |
| Attention deficit hyperactivity disorder | 0.4 | 0.2 | -0.03 | 0.3 | 0.3 | 0.00 | 0.3 | 0.2 | 0.00 |
| Chronic obstructive lung disease | 8.4 | 8.4 | 0.00 | 8.1 | 8.9 | 0.03 | 8.4 | 8.5 | 0.00 |
| Crohn’s disease | <0.1 | 0.2 | 0.02 | <0.1 | 0.2 | 0.04 | 0.1 | 0.2 | 0.02 |
| Dementia | 1.0 | 2.4 | 0.10 | 1.1 | 1.1 | 0.00 | 1.3 | 2.2 | 0.07 |
| Depressive disorder | 16.0 | 11.3 | -0.14 | 14.4 | 15.0 | 0.01 | 12.4 | 11.8 | -0.02 |
| Gastroesophageal reflux disease | 20.0 | 18.3 | -0.04 | 18.9 | 19.4 | 0.01 | 18.4 | 18.6 | 0.00 |
| Gastrointestinal hemorrhage | 1.5 | 1.7 | 0.01 | 1.5 | 1.4 | 0.00 | 2.7 | 1.6 | -0.07 |
| Human immunodeficiency virus infection | 0.1 | 0.1 | -0.01 | <0.1 | <0.2 | 0.00 | <0.1 | 0.1 | 0.02 |
| Hyperlipidemia | 71.0 | 72.3 | 0.03 | 70.2 | 69.9 | -0.01 | 73.7 | 72.0 | -0.04 |
| Hypertensive disorder | 72.3 | 74.8 | 0.06 | 72.0 | 72.5 | 0.01 | 76.0 | 74.5 | -0.04 |
| Lesion of liver | 0.9 | 0.8 | -0.01 | 0.8 | 0.8 | 0.00 | 1.0 | 0.8 | -0.02 |
| Obesity | 35.3 | 17.4 | -0.42 | 31.0 | 33.5 | 0.05 | 18.9 | 19.6 | 0.02 |
| Osteoarthritis | 25.4 | 22.8 | -0.06 | 24.4 | 25.6 | 0.03 | 23.9 | 23.2 | -0.02 |
| Pneumonia | 2.3 | 2.7 | 0.03 | 2.1 | 2.4 | 0.02 | 2.5 | 2.6 | 0.01 |
| Psoriasis | 1.9 | 1.2 | -0.05 | 1.7 | 1.9 | 0.01 | 1.1 | 1.3 | 0.02 |
| Renal impairment | 10.8 | 12.9 | 0.07 | 10.8 | 11.1 | 0.01 | 12.4 | 12.7 | 0.01 |
| Rheumatoid arthritis | 1.1 | 1.4 | 0.02 | 1.2 | 1.4 | 0.02 | 1.2 | 1.4 | 0.02 |
| Schizophrenia | <0.1 | 0.2 | 0.05 | <0.1 | <0.2 | 0.03 | <0.1 | 0.2 | 0.05 |
| Ulcerative colitis | 0.3 | 0.2 | -0.02 | 0.3 | 0.3 | 0.00 | 0.2 | 0.3 | 0.00 |
| Urinary tract infectious disease | 5.5 | 7.5 | 0.08 | 5.6 | 5.7 | 0.00 | 7.4 | 7.3 | -0.01 |
| Viral hepatitis C | 0.6 | 0.3 | -0.04 | 0.6 | 0.4 | -0.04 | 0.8 | 0.3 | -0.06 |
| Visual system disorder | 16.1 | 17.3 | 0.03 | 15.6 | 15.9 | 0.01 | 14.3 | 17.1 | 0.08 |
| **Medical history: Cardiovascular disease** | | | | | | | | | |
| Atrial fibrillation | 8.1 | 9.4 | 0.04 | 8.6 | 9.2 | 0.02 | 8.6 | 9.2 | 0.02 |
| Cerebrovascular disease | 5.0 | 5.9 | 0.04 | 4.9 | 4.8 | 0.00 | 6.0 | 5.8 | -0.01 |
| Coronary arteriosclerosis | 17.5 | 19.0 | 0.04 | 16.9 | 17.3 | 0.01 | 18.8 | 18.7 | 0.00 |
| Heart disease | 33.3 | 34.8 | 0.03 | 32.4 | 31.7 | -0.01 | 35.5 | 34.4 | -0.02 |
| Heart failure | 5.8 | 6.1 | 0.01 | 5.4 | 5.3 | 0.00 | 5.7 | 6.0 | 0.01 |
| Ischemic heart disease | 7.3 | 7.6 | 0.01 | 7.1 | 6.7 | -0.02 | 7.7 | 7.5 | -0.01 |
| Peripheral vascular disease | 8.1 | 7.5 | -0.03 | 7.7 | 7.6 | 0.00 | 7.4 | 7.5 | 0.01 |
| Pulmonary embolism | 0.9 | 0.7 | -0.02 | 0.8 | 0.6 | -0.02 | 1.0 | 0.7 | -0.03 |
| Venous thrombosis | 0.9 | 1.4 | 0.05 | 0.9 | 1.0 | 0.02 | 1.1 | 1.4 | 0.03 |
| **Medical history: Neoplasms** | | | | | | | | | |
| Hematologic neoplasm | 1.1 | 1.2 | 0.01 | 1.1 | 0.9 | -0.01 | 1.3 | 1.1 | -0.02 |
| Malignant lymphoma | 0.7 | 0.7 | 0.00 | 0.6 | 0.6 | -0.01 | 0.4 | 0.7 | 0.04 |
| Malignant neoplasm of anorectum | 0.2 | 0.2 | 0.00 | 0.2 | <0.2 | -0.01 | 0.8 | 0.2 | -0.09 |
| Malignant neoplastic disease | 10.3 | 12.0 | 0.05 | 10.4 | 10.6 | 0.01 | 12.4 | 11.8 | -0.02 |
| Malignant tumor of breast | 2.0 | 2.2 | 0.01 | 2.0 | 2.3 | 0.02 | 1.8 | 2.2 | 0.03 |
| Malignant tumor of colon | 0.5 | 0.6 | 0.01 | 0.5 | 0.6 | 0.02 | 0.4 | 0.6 | 0.03 |
| Malignant tumor of lung | <0.1 | 0.5 | 0.09 | <0.1 | 0.5 | 0.09 | <0.1 | 0.4 | 0.09 |
| Malignant tumor of urinary bladder | 0.7 | 0.7 | -0.01 | 0.7 | 0.4 | -0.04 | 0.8 | 0.6 | -0.01 |
| Primary malignant neoplasm of prostate | 2.1 | 2.8 | 0.05 | 2.1 | 2.2 | 0.00 | 3.8 | 2.8 | -0.06 |
| **Medication use** | | | | | | | | | |

| (Continued) | Before ajustment | | | After matching | | | After stratification | | |
|---|---|---|---|---|---|---|---|---|---|
| Characteristic | T (%) | C (%) | StdDiff | T (%) | C (%) | StdDiff | T (%) | C (%) | StdDiff |
| Agents acting on the renin-angiotensin system | 72.0 | 72.1 | 0.00 | 72.1 | 72.4 | 0.01 | 73.9 | 72.1 | -0.04 |
| Antibacterials for systemic use | 43.8 | 42.8 | -0.02 | 42.6 | 43.2 | 0.01 | 42.3 | 42.8 | 0.01 |
| Antidepressants | 35.7 | 26.7 | -0.20 | 33.6 | 35.5 | 0.04 | 27.8 | 27.9 | 0.00 |
| Antiepileptics | 22.0 | 17.1 | -0.12 | 20.9 | 21.2 | 0.01 | 18.4 | 17.7 | -0.02 |
| Antiinflammatory and antirheumatic products | 63.4 | 60.3 | -0.06 | 62.3 | 62.3 | 0.00 | 62.9 | 60.6 | -0.05 |
| Antineoplastic agents | 7.2 | 6.0 | -0.05 | 6.9 | 6.6 | -0.01 | 6.7 | 6.1 | -0.02 |
| Antipsoriatics | 0.4 | 0.6 | 0.03 | 0.4 | 0.6 | 0.04 | 0.3 | 0.7 | 0.05 |
| Antithrombotic agents | 53.2 | 54.6 | 0.03 | 52.6 | 52.3 | -0.01 | 56.4 | 54.4 | -0.04 |
| Beta blocking agents | 43.7 | 45.7 | 0.04 | 43.5 | 44.0 | 0.01 | 45.9 | 45.4 | -0.01 |
| Calcium channel blockers | 29.9 | 31.2 | 0.03 | 30.0 | 29.8 | 0.00 | 32.7 | 31.1 | -0.04 |
| Diuretics | 50.4 | 47.6 | -0.06 | 49.5 | 48.6 | -0.02 | 50.6 | 47.9 | -0.05 |
| Drugs for acid related disorders | 46.6 | 44.7 | -0.04 | 45.3 | 46.5 | 0.03 | 45.7 | 45.0 | -0.01 |
| Drugs for obstructive airway diseases | 47.7 | 42.3 | -0.11 | 46.3 | 47.2 | 0.02 | 40.0 | 43.0 | 0.06 |
| Immunosuppressants | 4.2 | 3.0 | -0.07 | 4.1 | 3.6 | -0.02 | 3.9 | 3.1 | -0.04 |
| Lipid modifying agents | 80.8 | 79.7 | -0.03 | 80.4 | 81.1 | 0.02 | 80.9 | 79.8 | -0.03 |
| Opioids | 28.0 | 27.0 | -0.02 | 27.3 | 26.6 | -0.02 | 27.6 | 26.9 | -0.01 |
| Psycholeptics | 30.4 | 28.2 | -0.05 | 29.0 | 29.5 | 0.01 | 27.8 | 28.5 | 0.02 |
| Psychostimulants | 2.9 | 1.7 | -0.08 | 2.6 | 2.5 | -0.01 | 2.2 | 1.8 | -0.03 |

#### 2.3 GLP1RA vs SU

##### 2.3.1 OptumDOD

**Supplementary Table 10:** Baseline patient characteristics for GLP1RA (T) and SU (C) new-users in the OptumDOD data source. We report proportion of initiators satisfying selected base-line characteristics and the standardized difference of population proportions (StdDiff) before and after propensity score adjustment. Less extreme StdDiffs through matching and stratification suggest improved balance between patient cohorts through adjustment.

| | Before adjustment | | | After matching | | | After stratification | | |
|---|---|---|---|---|---|---|---|---|---|
| Characteristic | T (%) | C (%) | StdDiff | T (%) | C (%) | StdDiff | T (%) | C (%) | StdDiff |
| **Age group** | | | | | | | | | |
| 65 - 69 | 46.5 | 32.8 | 0.28 | 45.5 | 45.3 | 0.00 | 33.5 | 33.8 | -0.01 |
| 70 - 74 | 33.3 | 31.2 | 0.04 | 33.2 | 33.8 | -0.01 | 31.2 | 31.4 | 0.00 |
| 75 - 79 | 14.7 | 20.0 | -0.14 | 15.4 | 15.1 | 0.01 | 20.4 | 19.6 | 0.02 |
| 80 - 84 | 4.0 | 10.9 | -0.26 | 4.3 | 4.0 | 0.01 | 9.7 | 10.3 | -0.02 |
| 85 - 89 | 1.4 | 4.8 | -0.20 | 1.5 | 1.5 | 0.00 | 5.2 | 4.6 | 0.03 |
| 90 - 94 | 0.1 | 0.3 | -0.05 | 0.1 | 0.4 | -0.05 | 0.1 | 0.4 | -0.05 |
| Gender: female | 57.8 | 49.6 | 0.16 | 56.1 | 56.9 | -0.02 | 49.5 | 50.2 | -0.01 |
| **Medical history: General** | | | | | | | | | |
| Acute respiratory disease | 22.8 | 23.2 | -0.01 | 22.4 | 22.9 | -0.01 | 25.1 | 23.2 | 0.04 |
| Attention deficit hyperactivity disorder | 0.6 | 0.2 | 0.07 | 0.4 | 0.4 | 0.01 | 0.2 | 0.2 | 0.01 |
| Chronic liver disease | 2.0 | 2.5 | -0.03 | 2.1 | 1.9 | 0.01 | 1.8 | 2.4 | -0.04 |
| Chronic obstructive lung disease | 12.7 | 13.7 | -0.03 | 12.7 | 13.1 | -0.01 | 13.7 | 13.7 | 0.00 |
| Crohn's disease | 0.1 | 0.2 | -0.03 | 0.1 | 0.2 | -0.02 | 0.1 | 0.2 | -0.04 |
| Dementia | 2.1 | 4.1 | -0.12 | 2.1 | 2.3 | -0.01 | 3.7 | 4.0 | -0.01 |
| Depressive disorder | 20.1 | 13.8 | 0.17 | 18.7 | 20.0 | -0.03 | 14.0 | 14.2 | 0.00 |
| Gastroesophageal reflux disease | 24.1 | 19.3 | 0.12 | 23.1 | 24.2 | -0.03 | 21.2 | 19.7 | 0.04 |
| Gastrointestinal hemorrhage | 2.3 | 3.3 | -0.06 | 2.2 | 2.5 | -0.02 | 2.5 | 3.2 | -0.04 |
| Human immunodeficiency virus infection | 0.3 | 0.1 | 0.04 | 0.3 | 0.2 | 0.02 | 0.1 | 0.1 | -0.01 |
| Hyperlipidemia | 84.6 | 82.4 | 0.06 | 84.4 | 84.7 | -0.01 | 84.7 | 82.6 | 0.06 |
| Hypertensive disorder | 86.0 | 85.2 | 0.02 | 85.9 | 85.9 | 0.00 | 87.3 | 85.3 | 0.06 |
| Lesion of liver | 1.6 | 1.5 | 0.00 | 1.6 | 1.5 | 0.01 | 1.2 | 1.5 | -0.03 |
| Obesity | 39.6 | 17.8 | 0.50 | 35.7 | 38.3 | -0.05 | 18.6 | 19.3 | -0.02 |
| Osteoarthritis | 40.0 | 32.1 | 0.16 | 38.6 | 39.8 | -0.03 | 34.7 | 32.7 | 0.04 |
| Pneumonia | 4.1 | 5.6 | -0.07 | 4.1 | 4.1 | 0.00 | 8.2 | 5.5 | 0.11 |
| Psoriasis | 2.3 | 1.5 | 0.06 | 2.3 | 2.0 | 0.02 | 1.2 | 1.5 | -0.03 |
| Renal impairment | 20.5 | 24.3 | -0.09 | 20.8 | 20.8 | 0.00 | 27.3 | 24.0 | 0.07 |
| Rheumatoid arthritis | 2.4 | 2.1 | 0.02 | 2.2 | 2.4 | -0.01 | 2.9 | 2.1 | 0.05 |
| Schizophrenia | 0.2 | 0.2 | -0.01 | 0.2 | 0.2 | 0.00 | 0.1 | 0.2 | -0.03 |
| Ulcerative colitis | 0.4 | 0.4 | 0.00 | 0.4 | 0.3 | 0.01 | 0.4 | 0.3 | 0.01 |
| Urinary tract infectious disease | 11.3 | 13.6 | -0.07 | 11.0 | 11.3 | -0.01 | 17.7 | 13.4 | 0.12 |
| Viral hepatitis C | 0.7 | 0.5 | 0.02 | 0.7 | 0.6 | 0.01 | 0.5 | 0.5 | 0.00 |
| Visual system disorder | 54.2 | 51.9 | 0.04 | 53.8 | 53.6 | 0.00 | 51.6 | 52.1 | -0.01 |
| **Medical history: Cardiovascular disease** | | | | | | | | | |
| Atrial fibrillation | 9.8 | 11.0 | -0.04 | 9.6 | 9.5 | 0.00 | 12.8 | 10.8 | 0.06 |
| Cerebrovascular disease | 8.4 | 10.3 | -0.07 | 8.3 | 8.2 | 0.01 | 10.8 | 10.1 | 0.02 |
| Coronary arteriosclerosis | 20.2 | 23.0 | -0.07 | 20.4 | 20.6 | -0.01 | 25.9 | 22.8 | 0.07 |
| Heart disease | 40.7 | 42.1 | -0.03 | 40.0 | 40.1 | 0.00 | 46.8 | 41.9 | 0.10 |
| Heart failure | 9.8 | 11.5 | -0.06 | 9.8 | 9.4 | 0.01 | 13.6 | 11.3 | 0.07 |
| Ischemic heart disease | 10.1 | 12.3 | -0.07 | 10.0 | 10.3 | -0.01 | 13.8 | 12.1 | 0.05 |
| Peripheral vascular disease | 16.9 | 16.1 | 0.02 | 17.2 | 17.1 | 0.00 | 17.0 | 16.2 | 0.02 |
| Pulmonary embolism | 1.2 | 1.1 | 0.01 | 1.2 | 1.1 | 0.00 | 1.0 | 1.1 | -0.01 |
| Venous thrombosis | 2.0 | 2.3 | -0.02 | 2.0 | 1.9 | 0.00 | 3.2 | 2.2 | 0.06 |
| **Medical history: Neoplasms** | | | | | | | | | |
| Hematologic neoplasm | 0.9 | 1.8 | -0.08 | 0.9 | 1.1 | -0.03 | 1.0 | 1.8 | -0.07 |
| Malignant lymphoma | 0.6 | 0.8 | -0.03 | 0.5 | 0.7 | -0.02 | 0.5 | 0.8 | -0.04 |
| Malignant neoplasm of anorectum | 0.2 | 0.3 | -0.03 | 0.2 | 0.3 | -0.02 | 0.1 | 0.3 | -0.05 |
| Malignant neoplastic disease | 14.2 | 15.8 | -0.04 | 14.3 | 14.0 | 0.01 | 15.8 | 15.6 | 0.00 |
| Malignant tumor of breast | 3.0 | 2.5 | 0.03 | 2.9 | 2.8 | 0.01 | 2.2 | 2.5 | -0.03 |
| Malignant tumor of colon | 0.7 | 0.9 | -0.02 | 0.8 | 0.6 | 0.02 | 0.9 | 0.9 | 0.00 |
| Malignant tumor of lung | 0.4 | 0.8 | -0.06 | 0.4 | 0.4 | 0.00 | 1.1 | 0.8 | 0.04 |
| Malignant tumor of urinary bladder | 0.6 | 0.9 | -0.04 | 0.6 | 0.7 | -0.02 | 1.3 | 0.9 | 0.04 |
| Primary malignant neoplasm of prostate | 3.1 | 3.7 | -0.03 | 3.3 | 2.9 | 0.02 | 4.9 | 3.6 | 0.06 |
| **Medication use** | | | | | | | | | |
| Agents acting on the renin-angiotensin system | 76.2 | 75.6 | 0.01 | 76.4 | 76.3 | 0.00 | 79.3 | 75.7 | 0.09 |
| Antibacterials for systemic use | 59.9 | 57.4 | 0.05 | 58.7 | 59.2 | -0.01 | 58.4 | 57.5 | 0.02 |

| (Continued) | Before ajustment | | | After matching | | | After stratification | | |
|---|---|---|---|---|---|---|---|---|---|
| Characteristic | T (%) | C (%) | StdDiff | T (%) | C (%) | StdDiff | T (%) | C (%) | StdDiff |
| Antidepressants | 38.5 | 25.8 | 0.28 | 36.4 | 37.8 | -0.03 | 36.4 | 37.8 | -0.03 |
| Antiepileptics | 24.8 | 17.5 | 0.18 | 23.4 | 24.3 | -0.02 | 17.9 | 18.0 | 0.00 |
| Antiinflammatory and antirheumatic products | 33.6 | 26.6 | 0.15 | 31.9 | 33.4 | -0.03 | 25.8 | 27.1 | -0.03 |
| Antineoplastic agents | 7.3 | 6.3 | 0.04 | 7.0 | 6.9 | 0.00 | 6.6 | 6.4 | 0.01 |
| Antipsoriatics | 0.7 | 0.5 | 0.02 | 0.7 | 0.7 | 0.00 | 0.6 | 0.5 | 0.01 |
| Antithrombotic agents | 20.0 | 21.5 | -0.04 | 19.9 | 20.0 | 0.00 | 23.7 | 21.4 | 0.06 |
| Beta blocking agents | 41.5 | 45.5 | -0.08 | 41.5 | 41.7 | 0.00 | 49.9 | 45.2 | 0.09 |
| Calcium channel blockers | 32.7 | 34.0 | -0.03 | 32.8 | 33.0 | 0.00 | 31.5 | 33.9 | -0.05 |
| Diuretics | 53.4 | 49.8 | 0.07 | 52.6 | 52.9 | -0.01 | 52.6 | 50.0 | 0.05 |
| Drugs for acid related disorders | 35.9 | 31.8 | 0.09 | 35.2 | 35.6 | -0.01 | 35.3 | 32.1 | 0.07 |
| Drugs for obstructive airway diseases | 47.8 | 37.7 | 0.20 | 46.1 | 47.2 | -0.02 | 39.5 | 38.4 | 0.02 |
| Immunosuppressants | 3.6 | 2.5 | 0.06 | 3.2 | 3.0 | 0.01 | 3.2 | 2.5 | 0.04 |
| Lipid modifying agents | 82.3 | 78.7 | 0.09 | 82.0 | 82.4 | -0.01 | 79.4 | 79.0 | 0.01 |
| Opioids | 28.2 | 26.3 | 0.04 | 27.3 | 28.2 | -0.02 | 24.5 | 26.4 | -0.04 |
| Psycholeptics | 26.8 | 22.8 | 0.09 | 25.4 | 26.2 | -0.02 | 19.3 | 23.0 | -0.09 |
| Psychostimulants | 1.8 | 0.7 | 0.10 | 1.4 | 1.2 | 0.02 | 1.0 | 0.7 | 0.03 |

##### 2.3.2 MDCR

**Supplementary Table 11:** Baseline patient characteristics for GLP1RA (T) and SU (C) new-users in the MDCR data source. We report proportion of initiators satisfying selected base-line characteristics and the standardized difference of population proportions (StdDiff) before and after propensity score adjustment. Less extreme StdDiffs through matching and stratification suggest improved balance between patient cohorts through adjustment.

| | Before adjustment | | | After matching | | | After stratification | | |
|---|---|---|---|---|---|---|---|---|---|
| Characteristic | T (%) | C (%) | StdDiff | T (%) | C (%) | StdDiff | T (%) | C (%) | StdDiff |
| **Age group** | | | | | | | | | |
| 65 - 69 | 44.0 | 28.1 | 0.34 | 43.1 | 44.4 | -0.03 | 29.5 | 29.9 | -0.01 |
| 70 - 74 | 36.6 | 30.2 | 0.14 | 36.7 | 35.1 | 0.04 | 33.9 | 30.2 | 0.08 |
| 75 - 79 | 13.3 | 21.3 | -0.21 | 13.7 | 13.4 | 0.01 | 19.7 | 20.2 | -0.01 |
| 80 - 84 | 4.6 | 12.9 | -0.30 | 5.0 | 4.6 | 0.02 | 7.4 | 12.3 | -0.17 |
| 85 - 89 | 1.1 | 5.6 | -0.26 | 1.1 | 1.4 | -0.03 | 9.4 | 5.5 | 0.15 |
| 90 - 94 | 0.3 | 1.6 | -0.13 | 0.3 | 0.8 | -0.06 | <0.1 | 1.6 | -0.17 |
| Gender: female | 55.2 | 46.8 | 0.17 | 54.1 | 54.6 | -0.01 | 38.0 | 46.8 | -0.18 |
| **Medical history: General** | | | | | | | | | |
| Acute respiratory disease | 22.8 | 21.0 | 0.04 | 22.7 | 22.4 | 0.01 | 22.6 | 21.7 | 0.02 |
| Attention deficit hyperactivity disorder | 0.3 | 0.1 | 0.05 | 0.4 | 0.5 | -0.01 | <0.1 | 0.1 | -0.02 |
| Chronic liver disease | 1.9 | 2.1 | -0.01 | 2.1 | 1.8 | 0.02 | 1.5 | 2.2 | -0.06 |
| Chronic obstructive lung disease | 10.9 | 10.9 | 0.00 | 10.8 | 10.7 | 0.00 | 15.1 | 11.3 | 0.11 |
| Crohn's disease | 0.3 | 0.3 | 0.00 | 0.3 | 0.3 | 0.00 | 0.2 | 0.3 | -0.01 |
| Dementia | 1.4 | 3.0 | -0.11 | 1.4 | 1.5 | -0.01 | 4.6 | 3.1 | 0.08 |
| Depressive disorder | 13.1 | 6.7 | 0.22 | 11.9 | 13.1 | -0.04 | 10.6 | 7.7 | 0.10 |
| Gastroesophageal reflux disease | 14.8 | 10.1 | 0.14 | 14.2 | 14.5 | -0.01 | 9.4 | 11.3 | -0.06 |
| Gastrointestinal hemorrhage | 2.3 | 3.5 | -0.07 | 2.1 | 2.4 | -0.02 | 2.2 | 3.4 | -0.07 |
| Hyperlipidemia | 67.7 | 49.4 | 0.38 | 66.0 | 67.8 | -0.04 | 53.5 | 54.7 | -0.02 |
| Hypertensive disorder | 76.4 | 66.4 | 0.22 | 75.7 | 77.3 | -0.04 | 75.9 | 69.8 | 0.14 |
| Lesion of liver | 0.7 | 1.1 | -0.04 | 0.8 | 1.2 | -0.04 | 0.3 | 1.2 | -0.11 |
| Obesity | 28.7 | 7.7 | 0.57 | 24.6 | 26.7 | -0.05 | 10.6 | 9.6 | 0.03 |
| Osteoarthritis | 37.3 | 26.3 | 0.24 | 35.6 | 36.1 | -0.01 | 28.0 | 28.0 | 0.00 |
| Pneumonia | 3.5 | 5.9 | -0.11 | 3.5 | 3.9 | -0.02 | 2.9 | 5.8 | -0.14 |
| Psoriasis | 2.8 | 1.3 | 0.11 | 2.7 | 2.2 | 0.03 | 1.5 | 1.4 | 0.01 |
| Renal impairment | 11.2 | 12.4 | -0.04 | 11.2 | 11.6 | -0.01 | 15.4 | 13.7 | 0.05 |
| Rheumatoid arthritis | 1.8 | 1.5 | 0.02 | 1.7 | 1.9 | -0.01 | 1.0 | 1.6 | -0.06 |
| Ulcerative colitis | 0.2 | 0.4 | -0.03 | <0.3 | 0.3 | -0.01 | 0.1 | 0.4 | -0.05 |
| Urinary tract infectious disease | 10.0 | 10.4 | -0.01 | 9.5 | 9.0 | 0.02 | 8.4 | 10.5 | -0.07 |
| Viral hepatitis C | <0.2 | 0.2 | -0.02 | <0.3 | 0.3 | -0.02 | 0.2 | 0.3 | -0.01 |
| Visual system disorder | 53.2 | 48.1 | 0.10 | 52.7 | 53.6 | -0.02 | 53.9 | 49.0 | 0.10 |
| **Medical history: Cardiovascular disease** | | | | | | | | | |
| Atrial fibrillation | 8.5 | 11.4 | -0.10 | 8.7 | 8.6 | 0.00 | 13.2 | 11.5 | 0.05 |
| Cerebrovascular disease | 9.6 | 12.0 | -0.08 | 9.5 | 9.7 | -0.01 | 15.6 | 11.8 | 0.11 |
| Coronary arteriosclerosis | 21.9 | 24.1 | -0.05 | 26.9 | 24.1 | 0.06 | 26.9 | 24.1 | 0.06 |
| Heart disease | 40.2 | 43.8 | -0.07 | 40.0 | 40.4 | -0.01 | 45.2 | 43.6 | 0.03 |
| Heart failure | 7.3 | 10.6 | -0.12 | 7.3 | 7.5 | -0.01 | 12.0 | 10.3 | 0.05 |
| Ischemic heart disease | 9.3 | 11.8 | -0.08 | 9.5 | 9.2 | 0.01 | 11.5 | 11.4 | 0.00 |
| Peripheral vascular disease | 11.5 | 10.8 | 0.02 | 11.5 | 11.6 | 0.00 | 13.5 | 11.2 | 0.07 |
| Pulmonary embolism | 0.8 | 0.9 | 0.00 | 0.9 | 1.0 | -0.01 | 4.0 | 0.9 | 0.20 |
| Venous thrombosis | 1.8 | 2.2 | -0.03 | 1.9 | 1.8 | 0.01 | 1.5 | 2.3 | -0.06 |
| **Medical history: Neoplasms** | | | | | | | | | |
| Hematologic neoplasm | 1.5 | 1.8 | -0.02 | 1.5 | 1.6 | -0.01 | 1.2 | 1.9 | -0.05 |
| Malignant lymphoma | 0.7 | 0.8 | -0.01 | 0.7 | 1.0 | -0.04 | 0.4 | 0.9 | -0.05 |
| Malignant neoplasm of anorectum | <0.2 | 0.4 | -0.06 | <0.3 | 0.2 | -0.03 | <0.1 | 0.4 | -0.06 |
| Malignant neoplastic disease | 17.2 | 18.1 | -0.02 | 17.3 | 17.7 | -0.01 | 16.7 | 18.4 | -0.04 |
| Malignant tumor of breast | 3.7 | 2.7 | 0.06 | 3.5 | 3.7 | -0.01 | 1.7 | 2.8 | -0.07 |
| Malignant tumor of colon | 0.3 | 0.9 | -0.07 | <0.3 | 0.5 | -0.04 | 0.1 | 0.9 | -0.10 |
| Malignant tumor of lung | 0.5 | 0.8 | -0.04 | 0.5 | 0.6 | -0.01 | 0.8 | 0.9 | -0.01 |
| Malignant tumor of urinary bladder | 1.4 | 1.1 | 0.02 | 1.4 | 1.0 | 0.04 | 1.4 | 1.0 | 0.04 |
| Primary malignant neoplasm of prostate | 3.9 | 4.5 | -0.03 | 4.1 | 4.0 | 0.00 | 7.6 | 4.5 | 0.13 |
| **Medication use** | | | | | | | | | |
| Agents acting on the renin-angiotensin system | 76.4 | 72.3 | 0.09 | 77.3 | 76.2 | 0.03 | 71.6 | 74.6 | -0.07 |
| Antibacterials for systemic use | 63.4 | 60.6 | 0.06 | 63.2 | 62.2 | 0.02 | 55.8 | 61.3 | -0.11 |
| Antidepressants | 35.7 | 24.2 | 0.25 | 34.0 | 35.2 | -0.03 | 28.0 | 25.3 | 0.06 |
| Antiepileptics | 19.1 | 13.8 | 0.14 | 17.9 | 18.9 | -0.03 | 15.6 | 14.9 | 0.02 |
| Antiinflammatory and antirheumatic products | 34.3 | 26.8 | 0.16 | 33.0 | 33.3 | -0.01 | 28.1 | 27.3 | 0.02 |
| Antineoplastic agents | 10.1 | 8.8 | 0.04 | 9.7 | 9.5 | 0.01 | 6.4 | 8.4 | -0.08 |
| Antipsoriatics | 0.9 | 0.7 | 0.03 | 0.8 | 0.9 | -0.01 | 0.5 | 0.7 | -0.02 |

| (Continued) | Before ajustment | | | After matching | | | After stratification | | |
|---|---|---|---|---|---|---|---|---|---|
| Characteristic | T (%) | C (%) | StdDiff | T (%) | C (%) | StdDiff | T (%) | C (%) | StdDiff |
| Antithrombotic agents | 22.4 | 25.2 | -0.07 | 22.6 | 22.2 | 0.01 | 27.7 | 25.6 | 0.05 |
| Beta blocking agents | 44.8 | 48.6 | -0.08 | 45.2 | 44.9 | 0.01 | 52.7 | 49.6 | 0.06 |
| Calcium channel blockers | 31.4 | 33.4 | -0.04 | 31.3 | 30.9 | 0.01 | 29.0 | 34.1 | -0.11 |
| Diuretics | 54.7 | 53.2 | 0.03 | 54.1 | 53.5 | 0.01 | 54.7 | 53.5 | 0.02 |
| Drugs for acid related disorders | 35.1 | 31.7 | 0.07 | 34.8 | 34.1 | 0.01 | 29.9 | 32.2 | -0.05 |
| Drugs for obstructive airway diseases | 49.6 | 37.5 | 0.24 | 47.8 | 48.4 | -0.01 | 43.0 | 39.5 | 0.07 |
| Immunosuppressants | 4.5 | 2.7 | 0.10 | 4.2 | 4.0 | 0.01 | 3.4 | 3.0 | 0.03 |
| Lipid modifying agents | 82.5 | 75.4 | 0.17 | 82.1 | 82.2 | 0.00 | 82.1 | 78.6 | 0.09 |
| Opioids | 28.3 | 27.9 | 0.01 | 28.2 | 29.2 | -0.02 | 30.5 | 28.6 | 0.04 |
| Psycholeptics | 26.5 | 24.9 | 0.04 | 26.0 | 26.8 | -0.02 | 29.7 | 25.4 | 0.10 |
| Psychostimulants | 1.8 | 1.1 | 0.06 | 1.7 | 1.9 | -0.01 | 0.5 | 1.2 | -0.07 |

##### 2.3.3 Open Claims

**Supplementary Table 12:** Baseline patient characteristics for GLP1RA (T) and SU (C) new-users in the Open Claims data source. We report proportion of initiators satisfying selected base-line characteristics and the standardized difference of population proportions (StdDiff) before and after propensity score adjustment. Less extreme StdDiffs through matching and stratification suggest improved balance between patient cohorts through adjustment.

| | Before adjustment | | | After matching | | | After stratification | | |
|---|---|---|---|---|---|---|---|---|---|
| Characteristic | T (%) | C (%) | StdDiff | T (%) | C (%) | StdDiff | T (%) | C (%) | StdDiff |
| **Age group** | | | | | | | | | |
| 65 - 69 | 49.1 | 36.6 | 0.26 | 48.0 | 48.1 | 0.00 | 38.3 | 37.8 | 0.01 |
| 70 - 74 | 29.8 | 28.1 | 0.04 | 30.0 | 30.2 | 0.00 | 29.0 | 28.5 | 0.01 |
| 75 - 79 | 14.2 | 25.6 | -0.29 | 14.7 | 14.3 | 0.01 | 23.2 | 24.4 | -0.03 |
| 80 - 84 | 6.4 | 9.5 | -0.11 | 6.8 | 6.9 | -0.01 | 9.4 | 9.1 | 0.01 |
| 85 - 89 | 0.5 | 0.2 | 0.04 | 0.5 | 0.5 | 0.00 | 0.3 | 0.3 | 0.00 |
| Gender: female | 57.3 | 49.6 | 0.16 | 56.0 | 55.6 | 0.01 | 49.4 | 50.2 | -0.01 |
| **Medical history: General** | | | | | | | | | |
| Acute respiratory disease | 11.8 | 12.6 | -0.02 | 11.4 | 11.6 | -0.01 | 13.2 | 12.5 | 0.02 |
| Attention deficit hyperactivity disorder | 0.4 | 0.1 | 0.05 | 0.3 | 0.3 | 0.00 | 0.1 | 0.1 | 0.00 |
| Chronic liver disease | 0.9 | 1.3 | -0.04 | 0.9 | 0.9 | 0.00 | 1.3 | 1.2 | 0.01 |
| Chronic obstructive lung disease | 7.1 | 8.1 | -0.04 | 7.0 | 7.3 | -0.01 | 8.2 | 8.0 | 0.01 |
| Crohn’s disease | 0.1 | 0.2 | 0.00 | 0.1 | 0.2 | -0.01 | 0.1 | 0.2 | -0.01 |
| Dementia | 0.8 | 1.9 | -0.09 | 0.9 | 1.0 | -0.01 | 1.8 | 1.8 | 0.00 |
| Depressive disorder | 8.8 | 6.0 | 0.11 | 8.0 | 8.3 | -0.01 | 6.5 | 6.3 | 0.01 |
| Gastroesophageal reflux disease | 10.0 | 8.4 | 0.06 | 9.4 | 9.8 | -0.01 | 8.7 | 8.6 | 0.00 |
| Gastrointestinal hemorrhage | 1.3 | 1.7 | -0.04 | 1.2 | 1.3 | 0.00 | 1.7 | 1.7 | 0.00 |
| Human immunodeficiency virus infection | 0.2 | 0.1 | 0.01 | 0.2 | 0.1 | 0.00 | 0.1 | 0.1 | 0.01 |
| Hyperlipidemia | 46.1 | 44.8 | 0.03 | 44.9 | 45.3 | -0.01 | 45.5 | 44.9 | 0.01 |
| Hypertensive disorder | 53.5 | 54.2 | -0.01 | 52.5 | 52.7 | 0.00 | 54.5 | 54.1 | 0.01 |
| Lesion of liver | 0.8 | 0.9 | -0.01 | 0.8 | 0.8 | 0.00 | 0.8 | 0.9 | -0.01 |
| Obesity | 16.2 | 7.3 | 0.28 | 13.9 | 14.2 | -0.01 | 8.6 | 8.1 | 0.02 |
| Osteoarthritis | 22.1 | 17.9 | 0.11 | 20.7 | 21.2 | -0.01 | 18.8 | 18.2 | 0.01 |
| Pneumonia | 2.5 | 3.5 | -0.06 | 2.5 | 2.6 | -0.01 | 2.5 | 2.6 | -0.01 |
| Psoriasis | 1.2 | 0.8 | 0.05 | 1.1 | 1.2 | -0.01 | 0.9 | 0.8 | 0.01 |
| Renal impairment | 9.4 | 12.5 | -0.10 | 9.4 | 9.7 | -0.01 | 12.0 | 12.2 | -0.01 |
| Rheumatoid arthritis | 1.5 | 1.2 | 0.02 | 1.4 | 1.5 | -0.01 | 1.3 | 1.2 | 0.00 |
| Schizophrenia | 0.1 | 0.2 | -0.01 | 0.1 | 0.2 | 0.00 | 0.1 | 0.2 | 0.00 |
| Ulcerative colitis | 0.2 | 0.2 | 0.01 | 0.2 | 0.2 | 0.00 | 0.2 | 0.2 | 0.00 |
| Urinary tract infectious disease | 6.3 | 6.9 | -0.02 | 6.0 | 6.4 | -0.01 | 7.1 | 6.9 | 0.01 |
| Viral hepatitis C | 0.2 | 0.3 | -0.01 | 0.2 | 0.2 | 0.00 | 0.3 | 0.3 | 0.00 |
| Visual system disorder | 25.2 | 25.2 | 0.00 | 24.9 | 25.1 | 0.00 | 24.9 | 25.1 | 0.00 |
| **Medical history: Cardiovascular disease** | | | | | | | | | |
| Atrial fibrillation | 6.2 | 7.4 | -0.05 | 6.1 | 6.3 | -0.01 | 7.5 | 7.3 | 0.01 |
| Cerebrovascular disease | 3.9 | 5.4 | -0.07 | 3.9 | 4.0 | -0.01 | 5.1 | 5.2 | -0.01 |
| Coronary arteriosclerosis | 11.4 | 13.4 | -0.06 | 11.2 | 11.4 | -0.01 | 13.9 | 13.2 | 0.02 |
| Heart disease | 24.1 | 27.0 | -0.07 | 23.5 | 23.7 | -0.01 | 27.4 | 26.7 | 0.01 |
| Heart failure | 5.0 | 6.9 | -0.08 | 4.9 | 5.2 | -0.01 | 4.9 | 5.2 | -0.01 |
| Ischemic heart disease | 4.6 | 5.9 | -0.06 | 4.5 | 4.7 | -0.01 | 5.9 | 5.8 | 0.00 |
| Peripheral vascular disease | 6.7 | 7.3 | -0.02 | 6.6 | 6.9 | -0.01 | 7.5 | 7.2 | 0.01 |
| Pulmonary embolism | 0.7 | 0.7 | 0.01 | 0.7 | 0.7 | 0.00 | 0.7 | 0.7 | 0.00 |
| Venous thrombosis | 0.9 | 1.3 | -0.03 | 0.9 | 0.9 | 0.00 | 1.3 | 1.2 | 0.01 |
| **Medical history: Neoplasms** | | | | | | | | | |
| Hematologic neoplasm | 0.8 | 1.2 | -0.03 | 0.8 | 0.9 | -0.01 | 1.1 | 1.2 | -0.01 |
| Malignant lymphoma | 0.5 | 0.7 | -0.02 | 0.5 | 0.6 | -0.01 | 0.7 | 0.7 | 0.00 |
| Malignant neoplasm of anorectum | 0.1 | 0.2 | -0.02 | 0.1 | 0.1 | 0.01 | 0.2 | 0.2 | 0.01 |
| Malignant neoplastic disease | 8.7 | 10.3 | -0.06 | 8.5 | 8.8 | -0.01 | 10.2 | 10.1 | 0.00 |
| Malignant tumor of breast | 1.8 | 1.7 | 0.01 | 1.7 | 1.7 | 0.00 | 1.7 | 1.7 | 0.00 |
| Malignant tumor of colon | 0.3 | 0.6 | -0.04 | 0.3 | 0.3 | 0.00 | 0.3 | 0.3 | 0.00 |
| Malignant tumor of lung | 0.3 | 0.5 | -0.04 | 0.3 | 0.3 | 0.00 | 0.3 | 0.3 | 0.00 |
| Malignant tumor of urinary bladder | 0.4 | 0.6 | -0.02 | 0.4 | 0.4 | 0.00 | 0.7 | 0.6 | 0.02 |
| Primary malignant neoplasm of prostate | 1.8 | 2.3 | -0.03 | 1.9 | 2.0 | -0.01 | 2.5 | 2.3 | 0.02 |
| **Medication use** | | | | | | | | | |
| Agents acting on the renin-angiotensin system | 70.2 | 72.5 | -0.05 | 70.2 | 70.7 | -0.01 | 72.2 | 72.5 | -0.01 |
| Antibacterials for systemic use | 59.1 | 56.0 | 0.06 | 58.0 | 58.5 | -0.01 | 56.8 | 56.3 | 0.01 |
| Antidepressants | 37.3 | 26.4 | 0.23 | 35.2 | 36.1 | -0.02 | 28.5 | 27.5 | 0.02 |
| Antiepileptics | 24.1 | 19.0 | 0.12 | 22.7 | 23.8 | -0.03 | 20.0 | 19.7 | 0.01 |
| Antiinflammatory and antirheumatic products | 36.0 | 31.1 | 0.10 | 34.8 | 35.4 | -0.01 | 31.7 | 31.5 | 0.00 |
| Antineoplastic agents | 7.9 | 5.6 | 0.09 | 7.2 | 7.5 | -0.01 | 6.2 | 5.8 | 0.02 |

| (Continued) | Before ajustment | | | After matching | | | After stratification | | |
|---|---|---|---|---|---|---|---|---|---|
| Characteristic | T (%) | C (%) | StdDiff | T (%) | C (%) | StdDiff | T (%) | C (%) | StdDiff |
| Antipsoriatics | 0.8 | 0.6 | 0.02 | 0.7 | 0.7 | 0.01 | 0.6 | 0.6 | 0.00 |
| Antithrombotic agents | 22.2 | 25.1 | -0.07 | 22.0 | 22.3 | -0.01 | 25.2 | 24.9 | 0.01 |
| Beta blocking agents | 41.1 | 45.1 | -0.08 | 41.0 | 41.9 | -0.02 | 45.4 | 44.8 | 0.01 |
| Calcium channel blockers | 31.1 | 32.9 | -0.04 | 31.2 | 31.1 | 0.00 | 32.5 | 32.9 | -0.01 |
| Diuretics | 49.4 | 47.7 | 0.03 | 48.7 | 48.7 | 0.00 | 48.0 | 47.7 | 0.01 |
| Drugs for acid related disorders | 38.9 | 36.1 | 0.06 | 37.8 | 38.9 | -0.02 | 36.3 | 36.5 | 0.00 |
| Drugs for obstructive airway diseases | 44.3 | 36.1 | 0.17 | 42.6 | 43.2 | -0.01 | 42.6 | 43.2 | -0.01 |
| Immunosuppressants | 4.0 | 2.7 | 0.07 | 3.7 | 3.9 | -0.01 | 3.0 | 2.9 | 0.00 |
| Lipid modifying agents | 77.0 | 75.1 | 0.04 | 76.6 | 77.7 | -0.03 | 75.5 | 75.3 | 0.00 |
| Opioids | 26.2 | 24.9 | 0.03 | 25.2 | 25.6 | -0.01 | 26.0 | 25.1 | 0.02 |
| Psycholeptics | 25.2 | 21.8 | 0.08 | 23.8 | 24.6 | -0.02 | 22.7 | 22.2 | 0.01 |
| Psychostimulants | 2.6 | 1.2 | 0.10 | 1.4 | 1.2 | 0.01 | 1.4 | 1.2 | 0.01 |

##### 2.3.4 OptumEHR

**Supplementary Table 13:** Baseline patient characteristics for GLP1RA (T) and SU (C) new-users in the OptumEHR data source. We report proportion of initiators satisfying selected base-line characteristics and the standardized difference of population proportions (StdDiff) before and after propensity score adjustment. Less extreme StdDiffs through matching and stratification suggest improved balance between patient cohorts through adjustment.

| | Before adjustment | | | After matching | | | After stratification | | |
|---|---|---|---|---|---|---|---|---|---|
| Characteristic | T (%) | C (%) | StdDiff | T (%) | C (%) | StdDiff | T (%) | C (%) | StdDiff |
| **Age group** | | | | | | | | | |
| 65 - 69 | 54.8 | 37.7 | 0.35 | 53.7 | 53.8 | 0.00 | 39.1 | 38.6 | 0.01 |
| 70 - 74 | 29.1 | 28.0 | 0.03 | 29.4 | 30.0 | -0.01 | 28.2 | 28.1 | 0.00 |
| 75 - 79 | 11.7 | 20.3 | -0.24 | 12.1 | 11.5 | 0.02 | 18.6 | 19.8 | -0.03 |
| 80 - 84 | 3.4 | 11.6 | -0.31 | 3.7 | 3.5 | 0.01 | 11.2 | 11.1 | 0.00 |
| 85 - 89 | 0.9 | 2.4 | -0.12 | 1.0 | 1.2 | -0.02 | 2.9 | 2.4 | 0.03 |
| Gender: female | 55.6 | 47.4 | 0.16 | 54.4 | 54.9 | -0.01 | 45.8 | 47.8 | -0.04 |
| **Race** | | | | | | | | | |
| Asian | 0.9 | 2.1 | -0.10 | 1.0 | 1.3 | -0.03 | 1.0 | 2.0 | -0.08 |
| Black or African American | 7.2 | 7.6 | -0.01 | 7.3 | 7.0 | 0.01 | 8.9 | 7.6 | 0.05 |
| White | 86.9 | 84.4 | 0.07 | 86.8 | 86.9 | 0.00 | 84.9 | 84.5 | 0.01 |
| **Ethnicity** | | | | | | | | | |
| Hispanic or Latino | 3.1 | 4.1 | -0.05 | 3.1 | 3.5 | -0.02 | 3.7 | 4.0 | -0.02 |
| Not Hispanic or Latino | 87.8 | 88.5 | -0.02 | 88.1 | 87.5 | 0.02 | 89.9 | 88.5 | 0.04 |
| **Medical history: General** | | | | | | | | | |
| Acute respiratory disease | 14.8 | 14.6 | 0.01 | 14.5 | 15.4 | -0.02 | 12.3 | 14.6 | -0.07 |
| Attention deficit hyperactivity disorder | 0.4 | 0.2 | 0.04 | 0.3 | 0.5 | -0.02 | 0.2 | 0.2 | 0.00 |
| Chronic liver disease | 2.0 | 2.2 | -0.01 | 2.1 | 1.8 | 0.02 | 2.2 | 2.1 | 0.00 |
| Chronic obstructive lung disease | 8.4 | 8.8 | -0.01 | 8.4 | 8.7 | -0.01 | 8.8 | 8.8 | 0.00 |
| Crohn's disease | <0.1 | 0.2 | -0.03 | <0.2 | 0.4 | -0.06 | 0.1 | 0.2 | -0.04 |
| Dementia | 1.0 | 2.4 | -0.10 | 1.1 | 1.2 | -0.01 | 1.3 | 2.3 | -0.08 |
| Depressive disorder | 16.0 | 11.0 | 0.15 | 14.8 | 15.6 | -0.02 | 12.2 | 11.2 | 0.03 |
| Gastroesophageal reflux disease | 20.0 | 16.4 | 0.09 | 19.0 | 19.9 | -0.02 | 16.6 | 16.6 | 0.00 |
| Gastrointestinal hemorrhage | 1.5 | 1.7 | -0.02 | 1.5 | 1.3 | 0.01 | 2.0 | 1.7 | 0.02 |
| Human immunodeficiency virus infection | 0.1 | 0.1 | 0.02 | <0.2 | 0.1 | 0.00 | <0.1 | 0.1 | -0.02 |
| Hyperlipidemia | 71.0 | 68.7 | 0.05 | 69.7 | 70.0 | -0.01 | 70.3 | 68.8 | 0.03 |
| Hypertensive disorder | 72.3 | 72.4 | 0.00 | 71.9 | 72.2 | -0.01 | 75.2 | 72.3 | 0.06 |
| Lesion of liver | 0.9 | 1.0 | -0.01 | 0.9 | 1.1 | -0.02 | 0.6 | 1.0 | -0.04 |
| Obesity | 35.3 | 16.0 | 0.45 | 31.8 | 34.4 | -0.06 | 17.5 | 16.9 | 0.01 |
| Osteoarthritis | 25.4 | 21.5 | 0.09 | 24.3 | 25.2 | -0.02 | 22.4 | 21.6 | 0.02 |
| Pneumonia | 2.3 | 3.1 | -0.05 | 2.1 | 2.1 | 0.00 | 1.9 | 3.0 | -0.08 |
| Psoriasis | 1.9 | 1.2 | 0.05 | 1.8 | 1.9 | -0.01 | 0.9 | 1.3 | -0.03 |
| Renal impairment | 10.8 | 13.8 | -0.09 | 10.8 | 11.2 | -0.01 | 14.8 | 13.6 | 0.03 |
| Rheumatoid arthritis | 1.1 | 1.4 | -0.02 | 1.2 | 1.7 | -0.04 | 2.1 | 1.4 | 0.05 |
| Schizophrenia | <0.1 | 0.1 | -0.04 | <0.2 | 0.2 | -0.04 | <0.1 | 0.1 | -0.05 |
| Ulcerative colitis | 0.3 | 0.3 | 0.01 | 0.3 | 0.4 | -0.01 | 0.2 | 0.3 | -0.02 |
| Urinary tract infectious disease | 5.5 | 7.0 | -0.06 | 5.7 | 5.6 | 0.00 | 8.0 | 6.9 | 0.04 |
| Viral hepatitis C | 0.6 | 0.3 | 0.05 | 0.7 | 0.4 | 0.04 | 0.7 | 0.3 | 0.05 |
| Visual system disorder | 16.1 | 19.3 | -0.09 | 15.4 | 15.9 | -0.01 | 15.5 | 19.1 | -0.10 |
| **Medical history: Cardiovascular disease** | | | | | | | | | |
| Atrial fibrillation | 8.1 | 9.3 | -0.04 | 8.1 | 7.8 | 0.01 | 9.9 | 9.2 | 0.02 |
| Cerebrovascular disease | 5.0 | 5.7 | -0.03 | 4.8 | 4.8 | 0.00 | 5.2 | 5.6 | -0.02 |
| Coronary arteriosclerosis | 17.5 | 18.7 | -0.03 | 16.8 | 16.8 | 0.00 | 19.6 | 18.6 | 0.03 |
| Heart disease | 33.3 | 33.9 | -0.01 | 32.5 | 32.1 | 0.01 | 35.7 | 33.8 | 0.04 |
| Heart failure | 5.8 | 6.8 | -0.04 | 5.6 | 6.0 | -0.01 | 5.8 | 6.7 | -0.04 |
| Ischemic heart disease | 7.3 | 7.8 | -0.02 | 7.1 | 6.7 | 0.01 | 7.3 | 7.7 | -0.02 |
| Peripheral vascular disease | 8.1 | 7.8 | 0.01 | 7.9 | 8.3 | -0.02 | 6.9 | 7.9 | -0.04 |
| Pulmonary embolism | 0.9 | 0.8 | 0.01 | 0.8 | 0.7 | 0.01 | 2.1 | 0.8 | 0.11 |
| Venous thrombosis | 0.9 | 1.6 | -0.06 | 0.9 | 1.0 | -0.02 | 0.7 | 1.5 | -0.08 |
| **Medical history: Neoplasms** | | | | | | | | | |
| Hematologic neoplasm | 1.1 | 1.2 | -0.01 | 1.1 | 1.1 | 0.00 | 1.1 | 1.2 | -0.01 |
| Malignant lymphoma | 0.7 | 0.7 | 0.00 | 0.6 | 0.6 | 0.00 | 0.4 | 0.7 | -0.04 |
| Malignant neoplasm of anorectum | 0.2 | 0.2 | -0.01 | <0.2 | 0.1 | 0.00 | 0.7 | 0.2 | 0.07 |
| Malignant neoplastic disease | 10.3 | 12.1 | -0.06 | 10.5 | 10.3 | 0.00 | 13.4 | 12.0 | 0.04 |
| Malignant tumor of breast | 2.0 | 1.9 | 0.00 | 2.0 | 2.1 | 0.00 | 1.1 | 2.0 | -0.07 |
| Malignant tumor of colon | 0.5 | 0.7 | -0.02 | 0.5 | 0.5 | 0.00 | 0.4 | 0.6 | -0.04 |
| Malignant tumor of lung | <0.1 | 0.5 | -0.09 | <0.2 | 0.4 | -0.08 | <0.1 | 0.5 | -0.10 |
| Primary malignant neoplasm of prostate | 2.1 | 3.0 | -0.06 | 2.2 | 2.1 | 0.01 | 6.1 | 3.0 | 0.15 |
| **Medication use** | | | | | | | | | |

| (Continued) | Before ajustment | | | After matching | | | After stratification | | |
|---|---|---|---|---|---|---|---|---|---|
| Characteristic | T (%) | C (%) | StdDiff | T (%) | C (%) | StdDiff | T (%) | C (%) | StdDiff |
| Agents acting on the renin-angiotensin system | 72.0 | 71.1 | 0.02 | 72.2 | 71.7 | 0.01 | 71.7 | 71.2 | 0.01 |
| Antibacterials for systemic use | 43.8 | 41.6 | 0.04 | 42.7 | 42.5 | 0.00 | 41.7 | 41.6 | 0.00 |
| Antidepressants | 35.7 | 25.6 | 0.22 | 33.6 | 35.5 | -0.04 | 27.6 | 26.1 | 0.03 |
| Antiepileptics | 22.0 | 16.6 | 0.14 | 21.1 | 21.7 | -0.02 | 20.2 | 16.9 | 0.09 |
| Antiinflammatory and antirheumatic products | 63.4 | 60.7 | 0.06 | 63.4 | 60.8 | 0.06 | 63.4 | 60.8 | 0.06 |
| Antineoplastic agents | 7.2 | 5.2 | 0.08 | 6.7 | 6.8 | 0.00 | 5.2 | 5.3 | 0.00 |
| Antipsoriatics | 0.4 | 0.5 | -0.02 | 0.4 | 0.6 | -0.02 | 0.3 | 0.5 | -0.03 |
| Antithrombotic agents | 53.2 | 56.1 | -0.06 | 52.7 | 52.6 | 0.00 | 58.5 | 55.9 | 0.05 |
| Beta blocking agents | 43.7 | 46.7 | -0.06 | 43.9 | 43.3 | 0.01 | 47.9 | 46.6 | 0.03 |
| Calcium channel blockers | 29.9 | 29.8 | 0.00 | 29.9 | 30.5 | -0.01 | 33.8 | 29.9 | 0.08 |
| Diuretics | 50.4 | 48.8 | 0.03 | 49.8 | 50.0 | 0.00 | 54.6 | 48.8 | 0.12 |
| Drugs for acid related disorders | 46.6 | 43.5 | 0.06 | 45.6 | 46.0 | -0.01 | 46.4 | 43.6 | 0.06 |
| Drugs for obstructive airway diseases | 47.7 | 40.6 | 0.14 | 46.7 | 47.3 | -0.01 | 37.8 | 40.9 | -0.06 |
| Immunosuppressants | 4.2 | 2.6 | 0.09 | 4.0 | 3.8 | 0.01 | 3.0 | 2.6 | 0.02 |
| Lipid modifying agents | 80.8 | 77.9 | 0.07 | 80.0 | 80.4 | -0.01 | 80.3 | 78.0 | 0.06 |
| Opioids | 28.0 | 27.8 | 0.00 | 27.5 | 27.1 | 0.01 | 31.5 | 27.7 | 0.08 |
| Psycholeptics | 30.4 | 27.0 | 0.07 | 29.1 | 30.0 | -0.02 | 25.8 | 27.1 | -0.03 |
| Psychostimulants | 2.9 | 1.3 | 0.12 | 2.5 | 2.2 | 0.02 | 1.6 | 1.3 | 0.02 |

#### 2.4 SGLT2I vs DPP4I

##### 2.4.1 OptumDOD

**Supplementary Table 14:** Baseline patient characteristics for SGLT2I (T) and DPP4I (C) new-users in the Optum-DOD data source. We report proportion of initiators satisfying selected base-line characteristics and the standardized difference of population proportions (StdDiff) before and after propensity score adjustment. Less extreme StdDiffs through matching and stratification suggest improved balance between patient cohorts through adjustment.

| | Before adjustment | | | After matching | | | After stratification | | |
|---|---|---|---|---|---|---|---|---|---|
| Characteristic | T (%) | C (%) | StdDiff | T (%) | C (%) | StdDiff | T (%) | C (%) | StdDiff |
| **Age group** | | | | | | | | | |
| 65 - 69 | 40.3 | 32.6 | -0.16 | 38.3 | 37.7 | -0.01 | 36.0 | 34.3 | -0.04 |
| 70 - 74 | 33.8 | 30.6 | -0.07 | 33.5 | 34.4 | 0.02 | 32.1 | 32.2 | 0.00 |
| 75 - 79 | 16.9 | 19.4 | 0.07 | 17.9 | 17.5 | -0.01 | 17.5 | 18.4 | 0.02 |
| 80 - 84 | 6.2 | 11.3 | 0.18 | 7.2 | 6.8 | -0.01 | 9.6 | 9.6 | 0.00 |
| 85 - 89 | 2.4 | 5.6 | 0.17 | 2.9 | 2.9 | 0.00 | 4.6 | 4.9 | 0.01 |
| 90 - 94 | 0.3 | 0.5 | 0.03 | 0.3 | 0.6 | 0.05 | 0.2 | 0.6 | 0.06 |
| Gender: female | 45.2 | 53.4 | 0.16 | 48.7 | 48.5 | 0.00 | 51.8 | 50.8 | -0.02 |
| **Medical history: General** | | | | | | | | | |
| Acute respiratory disease | 22.6 | 24.7 | 0.05 | 23.0 | 22.5 | -0.01 | 24.2 | 23.7 | -0.01 |
| Attention deficit hyperactivity disorder | 0.3 | 0.2 | -0.01 | 0.3 | 0.3 | 0.01 | 0.2 | 0.3 | 0.01 |
| Chronic liver disease | 1.5 | 2.8 | 0.09 | 1.5 | 1.5 | 0.00 | 2.3 | 2.2 | -0.01 |
| Chronic obstructive lung disease | 12.3 | 13.0 | 0.02 | 12.0 | 12.5 | 0.01 | 12.4 | 13.0 | 0.02 |
| Dementia | 1.9 | 4.6 | 0.16 | 2.2 | 2.3 | 0.01 | 4.2 | 4.0 | -0.01 |
| Depressive disorder | 15.5 | 14.6 | -0.03 | 15.2 | 14.9 | -0.01 | 15.8 | 15.6 | 0.00 |
| Gastroesophageal reflux disease | 22.1 | 21.3 | -0.02 | 21.9 | 21.9 | 0.00 | 23.1 | 22.6 | -0.01 |
| Gastrointestinal hemorrhage | 2.5 | 3.3 | 0.05 | 2.5 | 2.4 | 0.00 | 3.4 | 3.0 | -0.03 |
| Human immunodeficiency virus infection | 0.2 | 0.2 | -0.01 | 0.2 | 0.2 | 0.00 | 0.3 | 0.2 | -0.01 |
| Hyperlipidemia | 86.3 | 85.4 | -0.03 | 85.5 | 86.2 | 0.02 | 84.8 | 85.8 | 0.03 |
| Hypertensive disorder | 87.1 | 87.2 | 0.00 | 86.4 | 87.1 | 0.02 | 86.5 | 87.4 | 0.03 |
| Lesion of liver | 1.4 | 1.4 | 0.00 | 1.4 | 1.3 | -0.01 | 1.6 | 1.4 | -0.01 |
| Obesity | 26.3 | 18.3 | -0.19 | 23.9 | 24.5 | 0.01 | 21.9 | 22.6 | 0.02 |
| Osteoarthritis | 34.0 | 35.2 | 0.03 | 34.3 | 34.3 | 0.00 | 35.5 | 35.3 | 0.00 |
| Pneumonia | 4.3 | 5.5 | 0.05 | 4.3 | 4.4 | 0.01 | 5.2 | 5.3 | 0.00 |
| Psoriasis | 2.0 | 1.5 | -0.04 | 1.8 | 1.7 | -0.01 | 1.7 | 1.6 | 0.00 |
| Renal impairment | 19.1 | 24.9 | 0.14 | 20.1 | 18.8 | -0.03 | 22.6 | 23.7 | 0.03 |
| Rheumatoid arthritis | 2.0 | 2.1 | 0.01 | 2.0 | 1.9 | -0.01 | 2.0 | 2.0 | 0.00 |
| Schizophrenia | 0.2 | 0.3 | 0.03 | 0.2 | 0.3 | 0.03 | 0.1 | 0.3 | 0.04 |
| Ulcerative colitis | 0.4 | 0.5 | 0.01 | 0.4 | 0.4 | 0.00 | 0.5 | 0.4 | -0.01 |
| Urinary tract infectious disease | 8.5 | 15.2 | 0.21 | 9.7 | 9.6 | 0.00 | 13.1 | 12.9 | 0.00 |
| Viral hepatitis C | 0.5 | 0.5 | 0.00 | 0.4 | 0.5 | 0.01 | 0.4 | 0.5 | 0.01 |
| Visual system disorder | 54.8 | 54.3 | -0.01 | 55.1 | 55.3 | 0.00 | 54.9 | 55.1 | 0.00 |
| **Medical history: Cardiovascular disease** | | | | | | | | | |
| Atrial fibrillation | 11.6 | 11.4 | 0.00 | 10.4 | 10.6 | 0.01 | 11.0 | 11.6 | 0.02 |
| Cerebrovascular disease | 9.3 | 10.8 | 0.05 | 9.0 | 9.4 | 0.02 | 9.7 | 10.1 | 0.01 |
| Coronary arteriosclerosis | 27.8 | 23.3 | -0.10 | 23.7 | 24.3 | 0.01 | 23.4 | 24.4 | 0.02 |
| Heart disease | 45.7 | 43.2 | -0.05 | 41.7 | 42.0 | 0.01 | 42.9 | 43.7 | 0.02 |
| Heart failure | 12.6 | 11.4 | -0.04 | 10.2 | 10.3 | 0.00 | 11.1 | 11.6 | 0.02 |
| Ischemic heart disease | 14.5 | 12.2 | -0.07 | 12.4 | 12.2 | 0.00 | 12.7 | 12.8 | 0.00 |
| Peripheral vascular disease | 18.2 | 17.5 | -0.02 | 17.4 | 17.7 | 0.01 | 17.9 | 18.4 | 0.01 |
| Pulmonary embolism | 1.1 | 1.0 | -0.01 | 1.1 | 1.0 | -0.02 | 1.2 | 1.1 | -0.01 |
| Venous thrombosis | 1.9 | 2.2 | 0.03 | 1.8 | 1.8 | 0.00 | 2.2 | 2.0 | -0.01 |
| **Medical history: Neoplasms** | | | | | | | | | |
| Hematologic neoplasm | 1.2 | 1.7 | 0.04 | 1.3 | 1.5 | 0.02 | 1.3 | 1.7 | 0.03 |
| Malignant lymphoma | 0.7 | 0.8 | 0.01 | 0.7 | 0.8 | 0.01 | 0.6 | 0.9 | 0.03 |
| Malignant neoplasm of anorectum | 0.2 | 0.3 | 0.02 | 0.2 | 0.2 | 0.01 | 0.3 | 0.2 | -0.02 |
| Malignant neoplastic disease | 14.5 | 16.1 | 0.05 | 14.6 | 14.5 | 0.00 | 14.7 | 15.5 | 0.02 |
| Malignant tumor of breast | 2.0 | 2.8 | 0.05 | 2.2 | 2.2 | 0.00 | 2.2 | 2.6 | 0.03 |
| Malignant tumor of colon | 0.7 | 0.9 | 0.02 | 0.7 | 0.7 | -0.01 | 1.0 | 0.7 | -0.03 |
| Malignant tumor of lung | 0.5 | 0.6 | 0.02 | 0.5 | 0.5 | 0.01 | 0.4 | 0.6 | 0.03 |
| Malignant tumor of urinary bladder | 0.6 | 0.8 | 0.03 | 0.6 | 0.6 | 0.01 | 0.6 | 0.8 | 0.02 |
| Primary malignant neoplasm of prostate | 3.5 | 3.9 | 0.02 | 3.5 | 3.4 | 0.00 | 3.4 | 3.7 | 0.02 |
| **Medication use** | | | | | | | | | |
| Agents acting on the renin-angiotensin system | 78.1 | 77.0 | -0.03 | 76.0 | 77.3 | 0.03 | 76.0 | 77.3 | 0.03 |
| Antibacterials for systemic use | 56.6 | 59.7 | 0.06 | 56.9 | 57.2 | 0.01 | 58.9 | 58.5 | -0.01 |
| Antidepressants | 28.2 | 27.5 | -0.01 | 27.9 | 27.8 | 0.00 | 28.1 | 28.3 | 0.00 |

| (Continued) | Before ajustment | | | After matching | | | After stratification | | |
|---|---|---|---|---|---|---|---|---|---|
| Characteristic | T (%) | C (%) | StdDiff | T (%) | C (%) | StdDiff | T (%) | C (%) | StdDiff |
| Antiepileptics | 19.1 | 18.8 | -0.01 | 19.2 | 19.1 | 0.00 | 20.1 | 20.1 | 0.00 |
| Antiinflammatory and antirheumatic products | 29.6 | 29.3 | -0.01 | 29.7 | 29.9 | 0.00 | 30.4 | 30.0 | -0.01 |
| Antineoplastic agents | 6.4 | 7.3 | 0.03 | 6.5 | 6.7 | 0.01 | 7.0 | 7.1 | 0.00 |
| Antipsoriatics | 0.5 | 0.6 | 0.01 | 0.5 | 0.5 | 0.00 | 0.4 | 0.6 | 0.03 |
| Antithrombotic agents | 24.6 | 22.5 | -0.05 | 23.2 | 23.2 | 0.00 | 23.2 | 23.2 | 0.00 |
| Beta blocking agents | 47.2 | 45.5 | -0.03 | 44.4 | 44.7 | 0.01 | 44.9 | 45.9 | 0.02 |
| Calcium channel blockers | 33.4 | 35.4 | 0.04 | 34.0 | 33.9 | 0.00 | 35.0 | 35.3 | 0.01 |
| Diuretics | 49.2 | 49.3 | 0.00 | 47.9 | 48.0 | 0.00 | 48.1 | 48.8 | 0.01 |
| Drugs for acid related disorders | 32.3 | 34.4 | 0.04 | 32.8 | 33.0 | 0.00 | 35.1 | 34.4 | -0.01 |
| Drugs for obstructive airway diseases | 42.0 | 41.3 | -0.01 | 41.8 | 42.0 | 0.00 | 43.2 | 42.2 | -0.02 |
| Immunosuppressants | 3.2 | 3.1 | -0.01 | 3.3 | 2.9 | -0.02 | 3.4 | 3.0 | -0.02 |
| Lipid modifying agents | 85.8 | 81.8 | -0.11 | 84.4 | 85.2 | 0.03 | 82.4 | 83.7 | 0.03 |
| Opioids | 26.3 | 27.1 | 0.02 | 25.6 | 25.8 | 0.01 | 26.8 | 26.8 | 0.00 |
| Psycholeptics | 23.7 | 24.2 | 0.01 | 23.3 | 23.5 | 0.00 | 24.7 | 24.2 | -0.01 |
| Psychostimulants | 1.1 | 1.0 | -0.02 | 1.2 | 1.2 | 0.00 | 1.2 | 1.1 | -0.01 |

##### 2.4.2 MDCR

**Supplementary Table 15:** Baseline patient characteristics for SGLT2I (T) and DPP4I (C) new-users in the MDCR data source. We report proportion of initiators satisfying selected base-line characteristics and the standardized difference of population proportions (StdDiff) before and after propensity score adjustment. Less extreme StdDiffs through matching and stratification suggest improved balance between patient cohorts through adjustment.

| | Before adjustment | | | After matching | | | After stratification | | |
|---|---|---|---|---|---|---|---|---|---|
| Characteristic | T (%) | C (%) | StdDiff | T (%) | C (%) | StdDiff | T (%) | C (%) | StdDiff |
| **Age group** | | | | | | | | | |
| 65 - 69 | 39.3 | 31.2 | -0.17 | 39.2 | 36.0 | -0.07 | 35.4 | 33.1 | -0.05 |
| 70 - 74 | 35.6 | 30.5 | -0.11 | 34.4 | 35.4 | 0.02 | 32.3 | 31.9 | -0.01 |
| 75 - 79 | 17.3 | 19.3 | 0.05 | 17.4 | 17.9 | 0.01 | 17.9 | 18.6 | 0.02 |
| 80 - 84 | 5.6 | 11.6 | 0.22 | 6.4 | 6.6 | 0.01 | 8.9 | 9.7 | 0.03 |
| 85 - 89 | 1.8 | 5.4 | 0.20 | 2.1 | 2.4 | 0.02 | 3.8 | 4.7 | 0.04 |
| 90 - 94 | 0.4 | 1.6 | 0.13 | 0.4 | 1.3 | 0.09 | 1.6 | 1.6 | 0.00 |
| 95 - 99 | <0.1 | 0.3 | 0.06 | <0.1 | 0.4 | 0.07 | <0.1 | 0.3 | 0.07 |
| Gender: female | 41.5 | 48.1 | 0.13 | 43.0 | 44.2 | 0.03 | 48.0 | 46.7 | -0.03 |
| **Medical history: General** | | | | | | | | | |
| Acute respiratory disease | 22.8 | 23.1 | 0.01 | 23.5 | 22.5 | -0.02 | 25.4 | 24.0 | -0.03 |
| Attention deficit hyperactivity disorder | 0.2 | 0.2 | 0.00 | 0.2 | 0.3 | 0.01 | 0.2 | 0.2 | 0.02 |
| Chronic liver disease | 1.9 | 2.3 | 0.03 | 2.0 | 1.7 | -0.03 | 2.3 | 2.3 | 0.00 |
| Chronic obstructive lung disease | 9.9 | 10.5 | 0.02 | 9.9 | 9.7 | 0.00 | 10.3 | 10.7 | 0.01 |
| Crohn’s disease | 0.3 | 0.3 | -0.01 | 0.3 | 0.2 | -0.03 | 0.3 | 0.3 | 0.00 |
| Dementia | 1.2 | 2.9 | 0.12 | 1.3 | 1.9 | 0.05 | 2.1 | 3.0 | 0.06 |
| Depressive disorder | 9.3 | 7.7 | -0.06 | 9.0 | 9.5 | 0.02 | 9.1 | 9.6 | 0.02 |
| Gastroesophageal reflux disease | 17.3 | 13.0 | -0.12 | 17.1 | 16.9 | 0.00 | 16.7 | 16.8 | 0.00 |
| Gastrointestinal hemorrhage | 2.6 | 3.2 | 0.03 | 2.7 | 2.0 | -0.04 | 3.0 | 2.9 | -0.01 |
| Human immunodeficiency virus infection | <0.1 | 0.1 | -0.01 | <0.1 | <0.2 | 0.01 | <0.1 | 0.1 | 0.03 |
| Hyperlipidemia | 81.3 | 63.3 | -0.41 | 80.5 | 80.9 | 0.01 | 78.4 | 78.3 | 0.00 |
| Hypertensive disorder | 83.8 | 75.0 | -0.22 | 84.0 | 83.2 | -0.02 | 84.0 | 83.2 | -0.02 |
| Lesion of liver | 1.2 | 0.8 | -0.04 | 1.1 | 1.1 | 0.00 | 1.2 | 1.0 | -0.02 |
| Obesity | 23.7 | 10.0 | -0.37 | 20.9 | 21.1 | 0.00 | 16.8 | 17.1 | 0.01 |
| Osteoarthritis | 31.8 | 30.4 | -0.03 | 32.0 | 32.6 | 0.01 | 33.3 | 33.2 | 0.00 |
| Pneumonia | 3.7 | 4.9 | 0.06 | 3.5 | 3.0 | -0.03 | 4.5 | 4.6 | 0.00 |
| Psoriasis | 2.3 | 1.7 | -0.04 | 2.2 | 2.1 | -0.01 | 2.3 | 1.9 | -0.03 |
| Renal impairment | 11.9 | 14.0 | 0.06 | 12.2 | 12.1 | 0.00 | 12.4 | 15.7 | 0.09 |
| Rheumatoid arthritis | 1.5 | 1.8 | 0.02 | 1.4 | 1.8 | 0.03 | 1.4 | 1.9 | 0.04 |
| Schizophrenia | <0.1 | 0.1 | 0.00 | <0.1 | <0.2 | 0.01 | <0.1 | 0.1 | 0.01 |
| Ulcerative colitis | 0.5 | 0.5 | 0.01 | 0.5 | 0.4 | 0.00 | 0.4 | 0.6 | 0.02 |
| Urinary tract infectious disease | 7.1 | 10.7 | 0.13 | 7.5 | 7.6 | 0.00 | 10.4 | 10.2 | -0.01 |
| Viral hepatitis C | 0.5 | 0.3 | -0.03 | 0.4 | 0.3 | 0.00 | 0.3 | 0.3 | 0.01 |
| Visual system disorder | 55.1 | 52.4 | -0.05 | 54.7 | 56.0 | 0.03 | 54.7 | 54.9 | 0.00 |
| **Medical history: Cardiovascular disease** | | | | | | | | | |
| Atrial fibrillation | 12.1 | 12.1 | 0.00 | 11.8 | 11.3 | -0.01 | 12.5 | 12.8 | 0.01 |
| Cerebrovascular disease | 11.0 | 12.7 | 0.05 | 10.6 | 11.0 | 0.01 | 12.2 | 11.8 | -0.01 |
| Coronary arteriosclerosis | 28.9 | 25.2 | -0.08 | 26.3 | 26.7 | 0.01 | 24.3 | 25.4 | 0.03 |
| Heart disease | 47.4 | 45.5 | -0.04 | 45.0 | 44.3 | -0.01 | 46.4 | 45.1 | -0.03 |
| Heart failure | 9.9 | 9.4 | -0.01 | 9.0 | 8.3 | -0.03 | 9.0 | 8.3 | -0.03 |
| Ischemic heart disease | 12.0 | 11.2 | -0.03 | 11.6 | 11.7 | 0.00 | 11.6 | 11.1 | -0.02 |
| Peripheral vascular disease | 13.8 | 12.8 | -0.03 | 12.9 | 13.3 | 0.01 | 12.7 | 14.0 | 0.04 |
| Pulmonary embolism | 0.9 | 0.9 | 0.00 | 0.8 | 0.9 | 0.01 | 1.4 | 1.1 | -0.03 |
| Venous thrombosis | 2.0 | 2.3 | 0.02 | 2.0 | 1.8 | -0.02 | 2.4 | 2.3 | -0.01 |
| **Medical history: Neoplasms** | | | | | | | | | |
| Hematologic neoplasm | 1.4 | 1.8 | 0.03 | 1.5 | 1.5 | 0.01 | 1.3 | 2.0 | 0.05 |
| Malignant lymphoma | 1.1 | 1.0 | -0.01 | 1.0 | 0.8 | -0.02 | 0.8 | 1.0 | 0.02 |
| Malignant neoplasm of anorectum | 0.3 | 0.4 | 0.02 | 0.3 | 0.3 | 0.01 | 0.4 | 0.3 | -0.03 |
| Malignant neoplastic disease | 16.9 | 19.2 | 0.06 | 16.9 | 17.5 | 0.02 | 18.1 | 18.8 | 0.02 |
| Malignant tumor of breast | 2.5 | 2.9 | 0.02 | 2.7 | 2.7 | 0.00 | 2.9 | 2.8 | -0.01 |
| Malignant tumor of colon | 0.6 | 0.9 | 0.04 | 0.6 | 0.5 | -0.01 | 0.6 | 0.9 | 0.04 |
| Malignant tumor of lung | 0.3 | 0.6 | 0.04 | 0.4 | 0.8 | 0.06 | 0.2 | 0.7 | 0.06 |
| Malignant tumor of urinary bladder | 1.0 | 1.2 | 0.03 | 1.0 | 1.0 | 0.00 | 1.0 | 1.3 | 0.03 |
| Primary malignant neoplasm of prostate | 4.3 | 4.5 | 0.01 | 4.4 | 4.7 | 0.01 | 4.4 | 4.6 | 0.01 |
| **Medication use** | | | | | | | | | |
| Agents acting on the renin-angiotensin system | 76.2 | 75.8 | -0.01 | 76.2 | 75.1 | -0.03 | 75.5 | 75.6 | 0.00 |
| Antibacterials for systemic use | 60.6 | 62.5 | 0.04 | 61.1 | 59.5 | -0.03 | 63.0 | 61.5 | -0.03 |
| Antidepressants | 24.2 | 24.9 | 0.02 | 24.4 | 24.3 | 0.00 | 24.1 | 25.0 | 0.02 |
| Antiepileptics | 15.8 | 14.6 | -0.03 | 15.8 | 14.7 | -0.03 | 16.3 | 15.3 | -0.03 |

| (Continued) | Before ajustment | | | After matching | | | After stratification | | |
|---|---|---|---|---|---|---|---|---|---|
| Characteristic | T (%) | C (%) | StdDiff | T (%) | C (%) | StdDiff | T (%) | C (%) | StdDiff |
| Antiinflammatory and antirheumatic products | 29.5 | 28.1 | -0.03 | 28.4 | 28.8 | 0.01 | 28.4 | 28.8 | 0.01 |
| Antineoplastic agents | 7.9 | 8.7 | 0.03 | 8.2 | 7.8 | -0.02 | 8.5 | 7.9 | -0.02 |
| Antipsoriatics | 0.8 | 0.8 | 0.00 | 0.7 | 0.5 | -0.02 | 0.6 | 0.6 | 0.01 |
| Antithrombotic agents | 25.1 | 25.4 | 0.00 | 24.4 | 24.4 | 0.00 | 24.8 | 24.9 | 0.00 |
| Beta blocking agents | 48.5 | 49.7 | 0.02 | 47.2 | 48.1 | 0.02 | 50.0 | 48.8 | -0.03 |
| Calcium channel blockers | 32.8 | 35.0 | 0.04 | 33.0 | 33.3 | 0.01 | 34.1 | 34.7 | 0.01 |
| Diuretics | 48.8 | 51.4 | 0.05 | 48.6 | 48.1 | -0.01 | 48.6 | 49.0 | 0.01 |
| Drugs for acid related disorders | 33.8 | 33.3 | -0.01 | 33.6 | 33.3 | -0.01 | 34.4 | 33.5 | -0.02 |
| Drugs for obstructive airway diseases | 44.1 | 41.5 | -0.05 | 43.7 | 44.6 | 0.02 | 43.7 | 43.6 | 0.00 |
| Immunosuppressants | 3.6 | 3.4 | -0.01 | 3.6 | 3.7 | 0.00 | 3.4 | 3.6 | 0.01 |
| Lipid modifying agents | 83.9 | 81.2 | -0.07 | 83.1 | 83.8 | 0.02 | 81.9 | 82.0 | 0.00 |
| Opioids | 24.9 | 27.5 | 0.06 | 25.5 | 24.6 | -0.02 | 27.9 | 26.3 | -0.04 |
| Psycholeptics | 23.0 | 26.0 | 0.07 | 23.2 | 23.2 | 0.00 | 25.0 | 24.1 | -0.02 |
| Psychostimulants | 1.5 | 1.3 | -0.02 | 1.7 | 1.3 | -0.03 | 1.7 | 1.2 | -0.04 |

##### 2.4.3 Open Claims

**Supplementary Table 16:** Baseline patient characteristics for SGLT2I (T) and DPP4I (C) new-users in the Open Claims data source. We report proportion of initiators satisfying selected base-line characteristics and the standardized difference of population proportions (StdDiff) before and after propensity score adjustment. Less extreme StdDiffs through matching and stratification suggest improved balance between patient cohorts through adjustment.

| | Before adjustment | | | After matching | | | After stratification | | |
|---|---|---|---|---|---|---|---|---|---|
| Characteristic | T (%) | C (%) | StdDiff | T (%) | C (%) | StdDiff | T (%) | C (%) | StdDiff |
| **Age group** | | | | | | | | | |
| 65 - 69 | 43.1 | 36.0 | -0.14 | 41.7 | 42.1 | 0.01 | 39.8 | 38.9 | -0.02 |
| 70 - 74 | 29.2 | 27.6 | -0.04 | 28.8 | 29.0 | 0.00 | 28.8 | 29.0 | 0.00 |
| 75 - 79 | 17.5 | 25.6 | 0.20 | 18.4 | 17.7 | -0.02 | 21.6 | 22.4 | 0.02 |
| 80 - 84 | 9.5 | 10.5 | 0.03 | 10.6 | 10.5 | 0.00 | 10.0 | 10.1 | 0.00 |
| 85 - 89 | 0.8 | 0.3 | -0.07 | 0.5 | 0.6 | 0.01 | 0.4 | 0.5 | 0.01 |
| Gender: female | 44.7 | 53.6 | 0.18 | 48.4 | 48.5 | 0.00 | 51.3 | 51.0 | -0.01 |
| **Medical history: General** | | | | | | | | | |
| Acute respiratory disease | 11.5 | 13.4 | 0.06 | 11.9 | 12.2 | 0.01 | 12.8 | 12.8 | 0.00 |
| Attention deficit hyperactivity disorder | 0.2 | 0.1 | -0.01 | 0.2 | 0.2 | 0.00 | 0.2 | 0.2 | 0.00 |
| Chronic liver disease | 0.9 | 1.3 | 0.04 | 1.0 | 1.0 | 0.00 | 1.3 | 1.2 | -0.01 |
| Chronic obstructive lung disease | 7.2 | 7.9 | 0.03 | 6.9 | 7.0 | 0.00 | 7.5 | 7.6 | 0.00 |
| Crohn’s disease | 0.2 | 0.2 | 0.00 | 0.2 | 0.1 | 0.00 | 0.2 | 0.1 | 0.00 |
| Dementia | 1.0 | 2.3 | 0.10 | 1.2 | 1.2 | 0.00 | 1.9 | 1.9 | 0.00 |
| Depressive disorder | 6.0 | 6.3 | 0.01 | 6.0 | 6.2 | 0.00 | 6.4 | 6.2 | -0.01 |
| Gastroesophageal reflux disease | 8.9 | 9.3 | 0.01 | 9.0 | 9.0 | 0.00 | 9.4 | 9.2 | -0.01 |
| Gastrointestinal hemorrhage | 1.4 | 1.9 | 0.04 | 1.4 | 1.4 | 0.00 | 1.7 | 1.7 | 0.00 |
| Human immunodeficiency virus infection | 0.2 | 0.2 | 0.00 | 0.2 | 0.2 | 0.01 | 0.2 | 0.2 | 0.01 |
| Hyperlipidemia | 48.5 | 49.1 | 0.01 | 48.1 | 48.2 | 0.00 | 49.2 | 48.7 | -0.01 |
| Hypertensive disorder | 54.8 | 57.6 | 0.06 | 54.8 | 54.9 | 0.00 | 56.5 | 56.5 | 0.00 |
| Lesion of liver | 0.8 | 0.8 | 0.01 | 0.8 | 0.8 | 0.00 | 0.8 | 0.8 | 0.00 |
| Obesity | 9.6 | 7.4 | -0.08 | 8.7 | 9.1 | 0.01 | 8.4 | 8.3 | 0.00 |
| Osteoarthritis | 18.0 | 19.7 | 0.05 | 18.2 | 18.5 | 0.01 | 19.4 | 19.2 | -0.01 |
| Pneumonia | 2.8 | 3.4 | 0.04 | 2.6 | 2.7 | 0.00 | 3.1 | 3.2 | 0.00 |
| Psoriasis | 1.0 | 0.9 | -0.01 | 0.9 | 0.9 | 0.00 | 0.9 | 0.9 | 0.00 |
| Renal impairment | 10.4 | 12.9 | 0.08 | 9.9 | 9.6 | -0.01 | 11.2 | 11.7 | 0.01 |
| Rheumatoid arthritis | 1.2 | 1.4 | 0.02 | 1.4 | 1.3 | -0.01 | 1.4 | 1.3 | -0.01 |
| Schizophrenia | 0.1 | 0.2 | 0.01 | 0.2 | 0.2 | 0.00 | 0.2 | 0.2 | 0.00 |
| Ulcerative colitis | 0.2 | 0.2 | 0.01 | 0.2 | 0.2 | 0.00 | 0.3 | 0.2 | 0.00 |
| Urinary tract infectious disease | 5.0 | 8.0 | 0.12 | 5.6 | 5.7 | 0.01 | 7.2 | 7.0 | -0.01 |
| Viral hepatitis C | 0.3 | 0.3 | 0.01 | 0.3 | 0.3 | 0.00 | 0.3 | 0.3 | 0.00 |
| Visual system disorder | 25.0 | 27.0 | 0.05 | 25.4 | 25.7 | 0.01 | 26.3 | 26.4 | 0.00 |
| **Medical history: Cardiovascular disease** | | | | | | | | | |
| Atrial fibrillation | 8.1 | 7.8 | -0.01 | 7.0 | 7.0 | 0.00 | 7.5 | 7.6 | 0.00 |
| Cerebrovascular disease | 4.7 | 6.1 | 0.06 | 4.8 | 4.7 | 0.00 | 5.5 | 5.6 | 0.00 |
| Coronary arteriosclerosis | 16.4 | 14.1 | -0.06 | 14.0 | 14.0 | 0.00 | 14.3 | 14.4 | 0.00 |
| Heart disease | 30.3 | 28.6 | -0.04 | 27.1 | 26.9 | 0.00 | 28.4 | 28.3 | 0.00 |
| Heart failure | 8.6 | 6.9 | -0.06 | 7.0 | 6.9 | 0.00 | 7.0 | 6.9 | 0.00 |
| Ischemic heart disease | 7.2 | 6.1 | -0.04 | 6.0 | 5.8 | 0.00 | 6.2 | 6.1 | 0.00 |
| Peripheral vascular disease | 7.5 | 7.6 | 0.00 | 7.1 | 7.2 | 0.00 | 7.5 | 7.6 | 0.00 |
| Pulmonary embolism | 0.6 | 0.7 | 0.01 | 0.7 | 0.6 | 0.00 | 0.7 | 0.6 | 0.00 |
| Venous thrombosis | 1.0 | 1.3 | 0.03 | 1.0 | 1.0 | 0.00 | 1.2 | 1.2 | 0.00 |
| **Medical history: Neoplasms** | | | | | | | | | |
| Hematologic neoplasm | 0.9 | 1.1 | 0.02 | 0.9 | 0.9 | 0.00 | 1.0 | 1.0 | 0.00 |
| Malignant lymphoma | 0.6 | 0.6 | 0.00 | 0.6 | 0.6 | 0.00 | 0.6 | 0.6 | 0.00 |
| Malignant neoplasm of anorectum | 0.1 | 0.2 | 0.01 | 0.2 | 0.2 | 0.00 | 0.2 | 0.2 | 0.00 |
| Malignant neoplastic disease | 9.1 | 10.5 | 0.05 | 9.3 | 9.2 | 0.00 | 10.1 | 10.0 | -0.01 |
| Malignant tumor of breast | 1.5 | 1.9 | 0.03 | 1.7 | 1.5 | -0.01 | 1.9 | 1.7 | -0.01 |
| Malignant tumor of colon | 0.4 | 0.6 | 0.03 | 0.4 | 0.4 | 0.00 | 0.5 | 0.5 | 0.00 |
| Malignant tumor of lung | 0.3 | 0.4 | 0.02 | 0.4 | 0.3 | 0.00 | 0.4 | 0.4 | 0.00 |
| Malignant tumor of urinary bladder | 0.4 | 0.6 | 0.03 | 0.4 | 0.4 | 0.00 | 0.5 | 0.5 | 0.00 |
| Primary malignant neoplasm of prostate | 2.3 | 2.3 | 0.00 | 2.2 | 2.3 | 0.00 | 2.3 | 2.3 | 0.00 |
| **Medication use** | | | | | | | | | |
| Agents acting on the renin-angiotensin system | 74.4 | 73.6 | -0.02 | 73.1 | 73.1 | 0.00 | 73.0 | 73.5 | 0.01 |
| Antibacterials for systemic use | 55.4 | 58.8 | 0.07 | 56.3 | 56.5 | 0.00 | 58.3 | 57.7 | -0.01 |
| Antidepressants | 27.2 | 27.6 | 0.01 | 27.3 | 27.2 | 0.00 | 27.9 | 27.5 | -0.01 |
| Antiepileptics | 18.9 | 19.8 | 0.02 | 19.0 | 19.1 | 0.00 | 19.6 | 19.6 | 0.00 |
| Antiinflammatory and antirheumatic products | 33.6 | 34.4 | 0.02 | 34.0 | 34.4 | 0.01 | 34.6 | 34.4 | 0.00 |
| Antineoplastic agents | 6.5 | 6.7 | 0.01 | 6.5 | 6.5 | 0.00 | 6.9 | 6.7 | -0.01 |

| (Continued) | Before ajustment | | | After matching | | | After stratification | | |
|---|---|---|---|---|---|---|---|---|---|
| Characteristic | T (%) | C (%) | StdDiff | T (%) | C (%) | StdDiff | T (%) | C (%) | StdDiff |
| Antipsoriatics | 0.7 | 0.8 | 0.01 | 0.7 | 0.7 | 0.00 | 0.7 | 0.7 | 0.00 |
| Antithrombotic agents | 28.7 | 27.5 | -0.03 | 26.4 | 25.9 | -0.01 | 26.4 | 25.9 | -0.01 |
| Beta blocking agents | 47.5 | 45.8 | -0.03 | 44.8 | 44.1 | -0.01 | 45.4 | 45.4 | 0.00 |
| Calcium channel blockers | 33.2 | 34.6 | 0.03 | 33.3 | 32.8 | -0.01 | 33.6 | 33.9 | 0.01 |
| Diuretics | 47.9 | 47.6 | -0.01 | 46.3 | 45.9 | -0.01 | 47.0 | 47.1 | 0.00 |
| Drugs for acid related disorders | 37.1 | 39.4 | 0.05 | 37.3 | 37.2 | 0.00 | 38.8 | 38.6 | 0.00 |
| Drugs for obstructive airway diseases | 39.5 | 39.3 | 0.00 | 39.4 | 39.4 | 0.00 | 39.9 | 39.5 | -0.01 |
| Immunosuppressants | 3.3 | 3.6 | 0.01 | 3.3 | 3.5 | 0.01 | 3.6 | 3.6 | 0.00 |
| Lipid modifying agents | 81.1 | 78.0 | -0.08 | 79.6 | 79.2 | -0.01 | 78.4 | 78.7 | 0.01 |
| Opioids | 22.8 | 25.8 | 0.07 | 23.3 | 23.4 | 0.00 | 25.2 | 24.8 | -0.01 |
| Psycholeptics | 20.9 | 23.1 | 0.05 | 21.1 | 21.4 | 0.01 | 22.7 | 22.5 | -0.01 |
| Psychostimulants | 1.5 | 1.4 | -0.01 | 1.5 | 1.5 | 0.00 | 1.6 | 1.4 | -0.01 |

##### 2.4.4 OptumEHR

**Supplementary Table 17:** Baseline patient characteristics for SGLT2I (T) and DPP4I (C) new-users in the OptumEHR data source. We report proportion of initiators satisfying selected base-line characteristics and the standardized difference of population proportions (StdDiff) before and after propensity score adjustment. Less extreme StdDiffs through matching and stratification suggest improved balance between patient cohorts through adjustment.

| | Before adjustment | | | After matching | | | After stratification | | |
|---|---|---|---|---|---|---|---|---|---|
| Characteristic | T (%) | C (%) | StdDiff | T (%) | C (%) | StdDiff | T (%) | C (%) | StdDiff |
| **Age group** | | | | | | | | | |
| 65 - 69 | 48.5 | 40.1 | -0.17 | 47.5 | 46.5 | -0.02 | 43.7 | 42.1 | -0.03 |
| 70 - 74 | 29.1 | 27.7 | -0.03 | 28.9 | 30.1 | 0.03 | 27.8 | 28.3 | 0.01 |
| 75 - 79 | 14.5 | 18.2 | 0.10 | 15.1 | 15.0 | 0.00 | 16.1 | 16.5 | 0.01 |
| 80 - 84 | 6.1 | 11.0 | 0.18 | 6.6 | 6.3 | -0.01 | 9.9 | 10.1 | 0.00 |
| 85 - 89 | 1.7 | 3.0 | 0.08 | 1.9 | 2.1 | 0.01 | 2.5 | 3.0 | 0.03 |
| Gender: female | 43.5 | 52.0 | 0.17 | 45.8 | 45.1 | -0.01 | 51.8 | 49.8 | -0.04 |
| **Race** | | | | | | | | | |
| Asian | 2.0 | 2.5 | 0.04 | 2.1 | 2.4 | 0.02 | 1.9 | 2.5 | 0.04 |
| Black or African American | 7.8 | 8.9 | 0.04 | 8.1 | 8.4 | 0.01 | 8.8 | 8.9 | 0.00 |
| White | 83.7 | 82.0 | -0.04 | 83.1 | 83.3 | 0.00 | 82.7 | 82.1 | -0.02 |
| **Ethnicity** | | | | | | | | | |
| Hispanic or Latino | 3.9 | 4.4 | 0.02 | 4.1 | 3.8 | -0.02 | 4.2 | 4.3 | 0.01 |
| Not Hispanic or Latino | 85.7 | 87.0 | 0.04 | 85.5 | 85.4 | 0.00 | 86.7 | 86.0 | -0.02 |
| **Medical history: General** | | | | | | | | | |
| Acute respiratory disease | 14.8 | 15.7 | 0.03 | 15.0 | 15.6 | 0.02 | 15.0 | 15.6 | 0.02 |
| Attention deficit hyperactivity disorder | 0.2 | 0.2 | 0.01 | 0.2 | 0.2 | 0.00 | 0.2 | 0.2 | 0.00 |
| Chronic liver disease | 1.6 | 2.0 | 0.03 | 1.7 | 1.5 | -0.01 | 2.0 | 1.8 | -0.01 |
| Chronic obstructive lung disease | 8.1 | 8.4 | 0.01 | 7.7 | 8.0 | 0.01 | 8.1 | 8.5 | 0.01 |
| Crohn’s disease | 0.3 | 0.2 | -0.02 | 0.3 | 0.2 | -0.02 | 0.3 | 0.2 | -0.03 |
| Dementia | 1.2 | 2.4 | 0.09 | 1.3 | 1.5 | 0.02 | 2.0 | 2.2 | 0.02 |
| Depressive disorder | 11.1 | 11.3 | 0.00 | 11.0 | 10.9 | 0.00 | 11.8 | 11.6 | -0.01 |
| Gastroesophageal reflux disease | 18.8 | 18.3 | -0.01 | 18.6 | 18.6 | 0.00 | 19.6 | 18.9 | -0.02 |
| Gastrointestinal hemorrhage | 1.1 | 1.7 | 0.04 | 1.2 | 1.1 | 0.00 | 1.3 | 1.5 | 0.02 |
| Human immunodeficiency virus infection | 0.1 | 0.1 | -0.01 | 0.1 | 0.1 | 0.00 | 0.1 | 0.1 | 0.00 |
| Hyperlipidemia | 73.7 | 72.3 | -0.03 | 72.4 | 73.3 | 0.02 | 71.7 | 72.2 | 0.01 |
| Hypertensive disorder | 74.8 | 74.8 | 0.00 | 74.1 | 75.3 | 0.03 | 75.1 | 74.9 | 0.00 |
| Lesion of liver | 1.1 | 0.8 | -0.03 | 1.1 | 0.9 | -0.02 | 1.3 | 0.8 | -0.04 |
| Obesity | 24.5 | 17.4 | -0.18 | 22.7 | 23.6 | 0.02 | 20.5 | 20.1 | -0.01 |
| Osteoarthritis | 21.6 | 22.8 | 0.03 | 21.7 | 21.8 | 0.00 | 23.5 | 22.9 | -0.01 |
| Pneumonia | 2.0 | 2.7 | 0.05 | 2.0 | 2.2 | 0.01 | 2.4 | 2.5 | 0.00 |
| Renal impairment | 10.0 | 12.9 | 0.09 | 10.2 | 10.0 | -0.01 | 11.7 | 12.4 | 0.02 |
| Rheumatoid arthritis | 1.5 | 1.4 | -0.01 | 1.6 | 1.3 | -0.03 | 1.8 | 1.4 | -0.03 |
| Schizophrenia | <0.1 | 0.2 | 0.04 | <0.1 | 0.1 | 0.03 | <0.1 | 0.2 | 0.05 |
| Ulcerative colitis | 0.3 | 0.2 | -0.02 | 0.4 | 0.3 | -0.02 | 0.4 | 0.3 | -0.01 |
| Urinary tract infectious disease | 4.3 | 7.5 | 0.14 | 4.6 | 4.8 | 0.01 | 7.3 | 6.6 | -0.03 |
| Viral hepatitis C | 0.3 | 0.3 | 0.01 | 0.3 | 0.4 | 0.02 | 0.2 | 0.4 | 0.04 |
| Visual system disorder | 16.0 | 17.3 | 0.03 | 17.1 | 16.8 | -0.01 | 17.1 | 16.8 | -0.01 |
| **Medical history: Cardiovascular disease** | | | | | | | | | |
| Atrial fibrillation | 8.6 | 9.4 | 0.03 | 8.2 | 8.7 | 0.02 | 8.6 | 9.2 | 0.02 |
| Cerebrovascular disease | 6.0 | 5.9 | 0.00 | 5.6 | 5.1 | -0.02 | 6.1 | 5.7 | -0.02 |
| Coronary arteriosclerosis | 24.1 | 19.0 | -0.12 | 21.2 | 21.7 | 0.01 | 18.8 | 20.0 | 0.03 |
| Heart disease | 37.8 | 34.8 | -0.06 | 35.1 | 35.6 | 0.01 | 33.8 | 35.3 | 0.03 |
| Heart failure | 7.2 | 6.1 | -0.04 | 6.2 | 5.7 | -0.02 | 5.9 | 6.1 | 0.01 |
| Ischemic heart disease | 9.6 | 7.6 | -0.07 | 8.5 | 8.6 | 0.00 | 7.8 | 8.1 | 0.01 |
| Peripheral vascular disease | 9.9 | 7.5 | -0.09 | 8.9 | 8.7 | 0.00 | 8.9 | 8.1 | -0.03 |
| Pulmonary embolism | 0.5 | 0.7 | 0.02 | 0.5 | 0.7 | 0.02 | 0.8 | 0.7 | -0.01 |
| Venous thrombosis | 1.3 | 1.4 | 0.01 | 1.3 | 1.2 | 0.00 | 1.6 | 1.3 | -0.02 |
| **Medical history: Neoplasms** | | | | | | | | | |
| Hematologic neoplasm | 1.0 | 1.2 | 0.01 | 1.1 | 1.0 | 0.00 | 1.3 | 1.1 | -0.01 |
| Malignant lymphoma | 0.7 | 0.7 | 0.00 | 0.7 | 0.7 | 0.00 | 0.7 | 0.7 | 0.00 |
| Malignant neoplasm of anorectum | 0.1 | 0.2 | 0.01 | 0.1 | 0.1 | 0.00 | 0.2 | 0.2 | -0.01 |
| Malignant neoplastic disease | 10.2 | 12.0 | 0.06 | 10.4 | 10.9 | 0.02 | 11.3 | 11.6 | 0.01 |
| Malignant tumor of breast | 1.7 | 2.2 | 0.04 | 1.8 | 1.8 | 0.00 | 2.1 | 2.1 | 0.00 |
| Malignant tumor of colon | 0.4 | 0.6 | 0.02 | 0.4 | 0.5 | 0.01 | 0.5 | 0.6 | 0.00 |
| Malignant tumor of lung | 0.4 | 0.5 | 0.01 | 0.4 | 0.4 | 0.01 | 0.4 | 0.5 | 0.01 |
| Malignant tumor of urinary bladder | 0.5 | 0.7 | 0.01 | 0.6 | 0.5 | -0.01 | 0.6 | 0.6 | 0.00 |
| Primary malignant neoplasm of prostate | 2.7 | 2.8 | 0.01 | 2.7 | 2.9 | 0.01 | 2.6 | 2.8 | 0.01 |
| **Medication use** | | | | | | | | | |

| (Continued) | Before ajustment | | | After matching | | | After stratification | | |
|---|---|---|---|---|---|---|---|---|---|
| Characteristic | T (%) | C (%) | StdDiff | T (%) | C (%) | StdDiff | T (%) | C (%) | StdDiff |
| Agents acting on the renin-angiotensin system | 74.2 | 72.1 | -0.05 | 73.6 | 73.8 | 0.00 | 72.5 | 72.7 | 0.00 |
| Antibacterials for systemic use | 40.7 | 42.8 | 0.04 | 40.9 | 41.3 | 0.01 | 43.7 | 42.3 | -0.03 |
| Antidepressants | 26.2 | 26.7 | 0.01 | 26.4 | 25.6 | -0.02 | 27.7 | 26.8 | -0.02 |
| Antiepileptics | 17.0 | 17.1 | 0.00 | 17.2 | 17.2 | 0.00 | 18.6 | 17.5 | -0.03 |
| Antiinflammatory and antirheumatic products | 62.7 | 60.3 | -0.05 | 61.7 | 62.3 | 0.01 | 61.7 | 62.3 | 0.01 |
| Antineoplastic agents | 5.4 | 6.0 | 0.03 | 5.4 | 5.5 | 0.01 | 6.0 | 5.8 | -0.01 |
| Antipsoriatics | 0.4 | 0.6 | 0.04 | 0.4 | 0.7 | 0.04 | 0.6 | 0.7 | 0.01 |
| Antithrombotic agents | 56.0 | 54.6 | -0.03 | 54.5 | 55.2 | 0.01 | 53.8 | 55.0 | 0.02 |
| Beta blocking agents | 47.6 | 45.7 | -0.04 | 46.0 | 45.7 | -0.01 | 45.5 | 46.0 | 0.01 |
| Calcium channel blockers | 31.0 | 31.2 | 0.01 | 30.6 | 30.7 | 0.00 | 31.0 | 31.3 | 0.01 |
| Diuretics | 46.2 | 47.6 | 0.03 | 45.7 | 45.0 | -0.01 | 47.1 | 46.5 | -0.01 |
| Drugs for acid related disorders | 42.8 | 44.7 | 0.04 | 43.0 | 42.3 | -0.01 | 45.5 | 44.3 | -0.03 |
| Drugs for obstructive airway diseases | 42.4 | 42.3 | 0.00 | 42.3 | 43.3 | 0.02 | 44.0 | 42.8 | -0.02 |
| Immunosuppressants | 3.0 | 3.0 | 0.00 | 3.0 | 3.0 | 0.00 | 2.9 | 3.1 | 0.01 |
| Lipid modifying agents | 83.7 | 79.7 | -0.10 | 82.6 | 82.8 | 0.00 | 80.5 | 80.8 | 0.01 |
| Opioids | 24.6 | 27.0 | 0.05 | 24.5 | 24.8 | 0.01 | 26.9 | 26.2 | -0.02 |
| Psycholeptics | 26.1 | 28.2 | 0.05 | 26.2 | 26.7 | 0.01 | 28.1 | 27.7 | -0.01 |
| Psychostimulants | 1.5 | 1.7 | 0.01 | 1.6 | 1.6 | 0.00 | 1.6 | 1.7 | 0.00 |

### 2.4.5 VA

**Supplementary Table 18:** Baseline patient characteristics for SGLT2I (T) and DPP4I (C) new-users in the VA data source. We report proportion of initiators satisfying selected base-line characteristics and the standardized difference of population proportions (StdDiff) before and after propensity score adjustment. Less extreme StdDiffs through matching and stratification suggest improved balance between patient cohorts through adjustment.

| | Before adjustment | | | After matching | | | After stratification | | |
|---|---|---|---|---|---|---|---|---|---|
| Characteristic | T (%) | C (%) | StdDiff | T (%) | C (%) | StdDiff | T (%) | C (%) | StdDiff |
| **Age group** | | | | | | | | | |
| 65 - 69 | 27.3 | 33.2 | 0.13 | 28.3 | 27.6 | -0.01 | 33.0 | 30.8 | -0.05 |
| 70 - 74 | 45.1 | 38.4 | -0.14 | 44.8 | 43.7 | -0.02 | 42.7 | 40.9 | -0.04 |
| 75 - 79 | 19.5 | 15.0 | -0.12 | 17.8 | 18.2 | 0.01 | 14.7 | 16.6 | 0.05 |
| 80 - 84 | 5.3 | 7.5 | 0.09 | 5.7 | 6.6 | 0.04 | 5.9 | 6.7 | 0.03 |
| 85 - 89 | 2.2 | 4.1 | 0.11 | 2.6 | 2.6 | 0.00 | 3.0 | 3.4 | 0.02 |
| 90 - 94 | 0.5 | 1.5 | 0.10 | 0.6 | 0.9 | 0.04 | 0.6 | 1.3 | 0.06 |
| 95 - 99 | <0.1 | 0.3 | 0.06 | 0.1 | 0.4 | 0.06 | <0.1 | 0.4 | 0.07 |
| 100 - 104 | <0.1 | <0.1 | -0.02 | <0.1 | <0.1 | -0.01 | <0.1 | <0.1 | -0.01 |
| Gender: female | 2.5 | 2.7 | 0.02 | 2.8 | 2.8 | 0.00 | 2.8 | 2.8 | 0.00 |
| **Race** | | | | | | | | | |
| Asian | 0.9 | 1.3 | 0.04 | 0.8 | 0.9 | 0.01 | 0.6 | 1.1 | 0.06 |
| Black or African American | 10.5 | 10.7 | 0.01 | 10.0 | 10.1 | 0.00 | 10.0 | 10.1 | 0.00 |
| White | 81.3 | 80.6 | -0.02 | 81.7 | 81.7 | 0.00 | 82.8 | 81.6 | -0.03 |
| Unknown | 5.7 | 5.9 | 0.01 | 5.9 | 5.7 | 0.00 | 5.3 | 5.7 | 0.02 |
| Native Hawaiian or Other Pacific Islander | 0.9 | 0.9 | 0.00 | 0.9 | 0.9 | 0.00 | 0.8 | 0.9 | 0.01 |
| American Indian or Alaska Native | 0.8 | 0.7 | -0.01 | 0.7 | 0.7 | 0.00 | 0.6 | 0.7 | 0.01 |
| **Ethnicity** | | | | | | | | | |
| Hispanic or Latino | 4.6 | 4.4 | -0.01 | 4.4 | 4.6 | 0.01 | 3.7 | 4.5 | 0.04 |
| Not Hispanic or Latino | 92.8 | 93.2 | 0.01 | 93.0 | 93.2 | 0.01 | 93.9 | 93.2 | -0.03 |
| **Medical history: General** | | | | | | | | | |
| Acute respiratory disease | 8.0 | 7.4 | -0.02 | 7.5 | 7.2 | -0.01 | 7.4 | 7.3 | 0.00 |
| Attention deficit hyperactivity disorder | 0.3 | 0.3 | 0.00 | 0.2 | 0.2 | 0.01 | 0.2 | 0.3 | 0.01 |
| Chronic liver disease | 2.4 | 1.8 | -0.04 | 2.2 | 1.8 | -0.03 | 2.2 | 1.8 | -0.03 |
| Chronic obstructive lung disease | 17.0 | 14.5 | -0.07 | 15.0 | 15.5 | 0.01 | 14.5 | 15.4 | 0.03 |
| Crohn's disease | 0.2 | 0.2 | 0.00 | 0.2 | <0.2 | -0.01 | 0.1 | 0.2 | 0.00 |
| Dementia | 1.8 | 2.6 | 0.06 | 1.8 | 2.3 | 0.03 | 2.0 | 2.4 | 0.03 |
| Depressive disorder | 17.5 | 16.2 | -0.03 | 16.4 | 16.3 | 0.00 | 15.8 | 16.3 | 0.01 |
| Gastroesophageal reflux disease | 20.7 | 19.7 | -0.03 | 19.6 | 20.4 | 0.02 | 19.6 | 20.4 | 0.02 |
| Gastrointestinal hemorrhage | 1.4 | 1.1 | -0.03 | 1.3 | 1.1 | -0.02 | 1.2 | 1.1 | -0.01 |
| Human immunodeficiency virus infection | 0.2 | 0.3 | 0.01 | 0.2 | 0.3 | 0.00 | 0.2 | 0.3 | 0.02 |
| Hyperlipidemia | 74.5 | 74.2 | -0.01 | 72.8 | 74.0 | 0.03 | 74.7 | 74.6 | 0.00 |
| Hypertensive disorder | 80.7 | 78.3 | -0.06 | 78.9 | 79.3 | 0.01 | 78.6 | 79.0 | 0.01 |
| Lesion of liver | 2.0 | 1.3 | -0.05 | 2.0 | 1.6 | -0.03 | 2.1 | 1.6 | -0.04 |
| Obesity | 25.9 | 21.7 | -0.10 | 23.4 | 23.5 | 0.00 | 22.2 | 22.8 | 0.02 |
| Osteoarthritis | 20.8 | 20.0 | -0.02 | 19.9 | 19.2 | -0.02 | 19.1 | 19.9 | 0.02 |
| Pneumonia | 2.1 | 1.6 | -0.04 | 1.8 | 1.7 | -0.01 | 1.5 | 1.6 | 0.01 |
| Psoriasis | 1.8 | 1.7 | -0.01 | 1.6 | 1.6 | 0.00 | 1.7 | 1.7 | 0.00 |
| Renal impairment | 12.6 | 11.4 | -0.03 | 10.9 | 11.7 | 0.02 | 9.9 | 11.5 | 0.05 |
| Rheumatoid arthritis | 0.9 | 0.9 | -0.01 | 0.9 | 0.8 | -0.01 | 0.8 | 0.8 | 0.00 |
| Schizophrenia | 0.5 | 0.7 | 0.02 | 0.4 | 0.5 | 0.02 | 0.4 | 0.5 | 0.02 |
| Ulcerative colitis | 0.4 | 0.5 | 0.01 | 0.4 | 0.3 | -0.01 | 0.3 | 0.4 | 0.02 |
| Urinary tract infectious disease | 1.6 | 2.6 | 0.07 | 1.5 | 1.9 | 0.03 | 1.8 | 2.2 | 0.03 |
| Viral hepatitis C | 1.1 | 0.9 | -0.02 | 1.0 | 0.8 | -0.02 | 0.8 | 0.8 | 0.00 |
| Visual system disorder | 47.4 | 48.9 | 0.03 | 47.2 | 47.8 | 0.01 | 45.8 | 48.0 | 0.04 |
| **Medical history: Cardiovascular disease** | | | | | | | | | |
| Atrial fibrillation | 16.8 | 10.8 | -0.17 | 12.8 | 14.4 | 0.05 | 11.3 | 13.0 | 0.05 |
| Cerebrovascular disease | 5.9 | 4.2 | -0.08 | 5.1 | 4.7 | -0.02 | 4.2 | 4.7 | 0.02 |
| Coronary arteriosclerosis | 41.3 | 21.0 | -0.45 | 29.1 | 32.2 | 0.07 | 25.6 | 28.6 | 0.07 |
| Heart disease | 58.9 | 37.2 | -0.44 | 46.0 | 49.4 | 0.07 | 39.9 | 44.8 | 0.10 |
| Heart failure | 16.2 | 5.4 | -0.35 | 7.5 | 8.4 | 0.03 | 8.3 | 7.9 | -0.02 |
| Ischemic heart disease | 16.5 | 8.5 | -0.24 | 10.1 | 11.0 | 0.03 | 9.1 | 10.5 | 0.05 |
| Peripheral vascular disease | 8.4 | 6.3 | -0.08 | 7.2 | 7.8 | 0.02 | 6.6 | 7.3 | 0.03 |
| Pulmonary embolism | 0.8 | 0.6 | -0.03 | 0.8 | 0.8 | 0.00 | 0.8 | 0.7 | 0.00 |
| Venous thrombosis | 1.1 | 1.4 | 0.03 | 1.2 | 1.3 | 0.01 | 1.1 | 1.5 | 0.03 |
| **Medical history: Neoplasms** | | | | | | | | | |
| Hematologic neoplasm | 1.3 | 1.4 | 0.01 | 1.3 | 1.4 | 0.00 | 1.3 | 1.4 | 0.00 |
| Malignant lymphoma | 1.0 | 0.9 | -0.01 | 1.0 | 0.7 | -0.03 | 0.8 | 0.9 | 0.01 |
| Malignant neoplasm of anorectum | <0.1 | 0.1 | 0.00 | <0.1 | <0.2 | 0.03 | <0.1 | <0.2 | 0.03 |

| (Continued) | Before ajustment | | | After matching | | | After stratification | | |
|---|---|---|---|---|---|---|---|---|---|
| Characteristic | T (%) | C (%) | StdDiff | T (%) | C (%) | StdDiff | T (%) | C (%) | StdDiff |
| Malignant neoplastic disease | 11.8 | 12.5 | 0.02 | 11.8 | 11.6 | 0.00 | 11.2 | 12.2 | 0.03 |
| Malignant tumor of breast | <0.1 | 0.1 | 0.01 | <0.1 | <0.2 | 0.01 | <0.1 | <0.1 | 0.02 |
| Malignant tumor of colon | 0.5 | 0.6 | 0.02 | 0.5 | 0.5 | -0.01 | 0.4 | 0.5 | 0.01 |
| Malignant tumor of lung | 0.6 | 0.5 | -0.01 | 0.6 | 0.5 | -0.01 | 0.5 | 0.5 | 0.00 |
| Malignant tumor of urinary bladder | 0.8 | 0.9 | 0.01 | 0.7 | 1.0 | 0.03 | 1.1 | 1.0 | -0.01 |
| Primary malignant neoplasm of prostate | 4.4 | 5.2 | 0.03 | 4.5 | 4.6 | 0.01 | 4.5 | 4.9 | 0.02 |
| **Medication use** | | | | | | | | | |
| Agents acting on the renin-angiotensin system | 70.6 | 65.5 | -0.11 | 66.5 | 67.3 | 0.01 | 65.0 | 66.6 | 0.03 |
| Antibacterials for systemic use | 25.6 | 26.2 | 0.01 | 23.9 | 25.4 | 0.03 | 25.0 | 25.5 | 0.01 |
| Antidepressants | 30.4 | 29.4 | -0.02 | 29.4 | 29.8 | 0.01 | 30.4 | 29.9 | -0.01 |
| Antiepileptics | 24.4 | 24.3 | 0.00 | 23.7 | 23.3 | -0.01 | 24.0 | 24.2 | 0.00 |
| Antiinflammatory and antirheumatic products | 48.5 | 41.9 | -0.13 | 44.1 | 44.4 | 0.00 | 43.8 | 44.0 | 0.00 |
| Antineoplastic agents | 5.1 | 4.5 | -0.03 | 5.0 | 4.8 | -0.01 | 5.9 | 4.5 | -0.06 |
| Antipsoriatics | 0.9 | 1.0 | 0.00 | 0.9 | 0.9 | 0.01 | 0.7 | 0.9 | 0.03 |
| Antithrombotic agents | 50.2 | 36.8 | -0.27 | 42.0 | 43.9 | 0.04 | 40.0 | 41.3 | 0.03 |
| Beta blocking agents | 56.9 | 41.4 | -0.31 | 46.6 | 49.3 | 0.06 | 42.8 | 46.2 | 0.07 |
| Calcium channel blockers | 31.2 | 29.8 | -0.03 | 30.0 | 30.5 | 0.01 | 27.7 | 29.6 | 0.04 |
| Diuretics | 44.9 | 38.1 | -0.14 | 39.0 | 39.7 | 0.01 | 38.8 | 39.9 | 0.02 |
| Drugs for acid related disorders | 44.0 | 42.4 | -0.03 | 41.3 | 42.5 | 0.02 | 43.3 | 42.9 | -0.01 |
| Drugs for obstructive airway diseases | 41.8 | 38.2 | -0.07 | 39.4 | 39.7 | 0.01 | 39.1 | 39.6 | 0.01 |
| Immunosuppressants | 2.9 | 2.4 | -0.03 | 2.7 | 2.3 | -0.02 | 2.3 | 2.3 | 0.00 |
| Lipid modifying agents | 85.1 | 80.0 | -0.14 | 81.9 | 83.0 | 0.03 | 81.0 | 81.6 | 0.01 |
| Opioids | 13.6 | 14.0 | 0.01 | 12.1 | 12.2 | 0.00 | 14.0 | 13.6 | -0.01 |
| Psycholeptics | 20.3 | 18.8 | -0.04 | 18.6 | 18.9 | 0.01 | 19.7 | 18.8 | -0.02 |
| Psychostimulants | 1.0 | 0.9 | 0.00 | 0.9 | 1.0 | 0.01 | 0.9 | 1.0 | 0.01 |

##### 2.4.6 GermanyDA

**Supplementary Table 19:** Baseline patient characteristics for SGLT2I (T) and DPP4I (C) new-users in the GermanyDA data source. We report proportion of initiators satisfying selected base-line characteristics and the standardized difference of population proportions (StdDiff) before and after propensity score adjustment. Less extreme StdDiffs through matching and stratification suggest improved balance between patient cohorts through adjustment.

| | Before adjustment | | | After matching | | | After stratification | | |
|---|---|---|---|---|---|---|---|---|---|
| Characteristic | T (%) | C (%) | StdDiff | T (%) | C (%) | StdDiff | T (%) | C (%) | StdDiff |
| **Age group** | | | | | | | | | |
| 65 - 69 | 32.1 | 27.0 | -0.11 | 32.2 | 31.5 | -0.02 | 32.4 | 28.2 | -0.09 |
| 70 - 74 | 26.9 | 25.4 | -0.03 | 25.9 | 27.2 | 0.03 | 23.7 | 24.7 | 0.02 |
| 75 - 79 | 20.8 | 22.9 | 0.05 | 21.0 | 20.4 | -0.02 | 21.3 | 22.4 | 0.03 |
| 80 - 84 | 14.3 | 16.0 | 0.05 | 14.6 | 14.6 | 0.00 | 15.7 | 16.1 | 0.01 |
| 85 - 89 | 5.0 | 6.8 | 0.07 | 5.3 | 5.2 | 0.00 | 5.5 | 6.7 | 0.05 |
| 90 - 94 | 0.8 | 1.8 | 0.09 | 0.8 | 1.1 | 0.03 | 1.3 | 1.7 | 0.04 |
| Gender: female | 38.2 | 48.6 | 0.21 | 41.6 | 40.0 | -0.03 | 47.7 | 45.9 | -0.04 |
| **Medical history: General** | | | | | | | | | |
| Acute respiratory disease | 10.3 | 12.7 | 0.08 | 10.5 | 10.0 | -0.02 | 12.2 | 11.7 | -0.01 |
| Attention deficit hyperactivity disorder | <0.1 | 0.2 | 0.03 | <0.1 | 0.2 | 0.05 | <0.1 | 0.2 | 0.03 |
| Chronic liver disease | 0.5 | 0.5 | 0.00 | 0.3 | 0.3 | -0.01 | 0.3 | 0.5 | 0.03 |
| Chronic obstructive lung disease | 6.7 | 7.0 | 0.01 | 6.2 | 6.3 | 0.00 | 6.3 | 6.8 | 0.02 |
| Crohn's disease | <0.1 | 0.1 | 0.03 | <0.1 | <0.1 | 0.03 | <0.1 | 0.1 | 0.01 |
| Dementia | 2.0 | 3.6 | 0.09 | 2.3 | 2.2 | -0.01 | 3.0 | 3.5 | 0.03 |
| Depressive disorder | 7.0 | 8.1 | 0.04 | 7.1 | 6.5 | -0.02 | 7.6 | 7.7 | 0.01 |
| Gastroesophageal reflux disease | 2.0 | 2.7 | 0.04 | 1.9 | 2.2 | 0.02 | 2.0 | 2.5 | 0.03 |
| Gastrointestinal hemorrhage | 0.7 | 0.9 | 0.01 | 0.7 | 0.8 | 0.01 | 0.5 | 0.9 | 0.04 |
| Hyperlipidemia | 24.8 | 25.0 | 0.01 | 23.5 | 23.6 | 0.00 | 22.1 | 23.5 | 0.03 |
| Hypertensive disorder | 45.0 | 51.3 | 0.13 | 43.8 | 43.2 | -0.01 | 43.7 | 47.2 | 0.07 |
| Lesion of liver | 0.6 | 0.7 | 0.02 | 0.5 | 0.5 | 0.01 | 0.5 | 0.7 | 0.03 |
| Obesity | 7.6 | 6.4 | -0.05 | 6.9 | 6.4 | -0.02 | 6.7 | 6.6 | 0.00 |
| Osteoarthritis | 11.6 | 13.4 | 0.05 | 11.9 | 11.4 | -0.02 | 12.2 | 12.6 | 0.01 |
| Pneumonia | 1.8 | 2.6 | 0.05 | 1.7 | 2.1 | 0.03 | 1.9 | 2.4 | 0.03 |
| Psoriasis | 1.5 | 1.1 | -0.03 | 1.5 | 1.3 | -0.02 | 1.8 | 1.2 | -0.05 |
| Renal impairment | 7.8 | 9.7 | 0.07 | 7.6 | 6.8 | -0.03 | 7.3 | 9.1 | 0.07 |
| Rheumatoid arthritis | 1.5 | 1.4 | 0.00 | 1.5 | 1.0 | -0.05 | 1.5 | 1.3 | -0.01 |
| Schizophrenia | 0.1 | 0.1 | 0.00 | 0.1 | <0.1 | -0.02 | <0.1 | 0.1 | 0.00 |
| Ulcerative colitis | 0.1 | 0.3 | 0.03 | 0.1 | <0.1 | 0.01 | 0.2 | 0.3 | 0.02 |
| Urinary tract infectious disease | 4.8 | 6.5 | 0.07 | 5.1 | 5.3 | 0.01 | 5.2 | 6.1 | 0.04 |
| Visual system disorder | 8.5 | 10.5 | 0.07 | 8.4 | 8.9 | 0.02 | 8.9 | 9.6 | 0.02 |
| **Medical history: Cardiovascular disease** | | | | | | | | | |
| Atrial fibrillation | 5.1 | 4.6 | -0.02 | 4.5 | 4.3 | -0.01 | 4.0 | 4.8 | 0.04 |
| Cerebrovascular disease | 5.6 | 6.2 | 0.03 | 5.4 | 5.6 | 0.01 | 4.8 | 6.0 | 0.06 |
| Coronary arteriosclerosis | 9.3 | 5.6 | -0.14 | 7.2 | 7.4 | 0.01 | 6.0 | 6.5 | 0.02 |
| Heart disease | 37.3 | 33.9 | -0.07 | 32.9 | 31.9 | -0.02 | 31.1 | 33.5 | 0.05 |
| Heart failure | 11.9 | 10.2 | -0.06 | 9.6 | 8.9 | -0.02 | 9.6 | 10.1 | 0.01 |
| Ischemic heart disease | 17.0 | 14.4 | -0.07 | 14.2 | 13.9 | -0.01 | 12.8 | 13.8 | 0.03 |
| Peripheral vascular disease | 7.8 | 6.9 | -0.03 | 7.1 | 6.5 | -0.03 | 6.2 | 7.0 | 0.04 |
| Pulmonary embolism | 0.7 | 0.8 | 0.02 | 0.6 | 0.9 | 0.03 | 0.4 | 0.9 | 0.06 |
| Venous thrombosis | 1.6 | 1.9 | 0.02 | 1.6 | 1.9 | 0.02 | 1.9 | 1.9 | 0.00 |
| **Medical history: Neoplasms** | | | | | | | | | |
| Hematologic neoplasm | 0.3 | 0.5 | 0.03 | 0.3 | 0.3 | 0.00 | 0.5 | 0.5 | 0.00 |
| Malignant lymphoma | 0.2 | 0.4 | 0.03 | 0.2 | 0.3 | 0.01 | 0.5 | 0.4 | -0.02 |
| Malignant neoplasm of anorectum | 0.1 | 0.3 | 0.04 | <0.1 | 0.2 | 0.04 | <0.1 | 0.3 | 0.05 |
| Malignant neoplastic disease | 5.6 | 7.2 | 0.07 | 5.6 | 5.4 | -0.01 | 6.3 | 6.8 | 0.02 |
| Malignant tumor of breast | 0.7 | 0.8 | 0.00 | 0.7 | 0.5 | -0.03 | 0.8 | 0.6 | -0.03 |
| Malignant tumor of colon | 0.5 | 0.6 | 0.01 | 0.6 | 0.5 | -0.01 | 0.5 | 0.6 | 0.00 |
| Malignant tumor of lung | <0.1 | 0.2 | 0.03 | <0.1 | <0.1 | 0.00 | <0.1 | 0.2 | 0.03 |
| Malignant tumor of urinary bladder | 0.3 | 0.3 | 0.00 | 0.3 | <0.1 | -0.03 | 0.5 | 0.2 | -0.04 |
| Primary malignant neoplasm of prostate | 1.1 | 1.2 | 0.01 | 1.2 | 0.8 | -0.04 | 1.1 | 1.2 | 0.01 |
| **Medication use** | | | | | | | | | |
| Agents acting on the renin-angiotensin system | 77.8 | 74.8 | -0.07 | 75.5 | 75.7 | 0.00 | 72.8 | 75.2 | 0.06 |
| Antibacterials for systemic use | 20.2 | 25.0 | 0.12 | 20.9 | 19.4 | -0.04 | 24.0 | 23.4 | -0.01 |
| Antidepressants | 10.1 | 11.1 | 0.03 | 10.5 | 10.3 | -0.01 | 11.0 | 11.1 | 0.00 |
| Antiepileptics | 5.4 | 6.1 | 0.03 | 5.4 | 5.6 | 0.01 | 6.0 | 6.3 | 0.01 |
| Antiinflammatory and antirheumatic products | 43.4 | 44.8 | 0.03 | 41.8 | 42.7 | 0.02 | 41.5 | 43.9 | 0.05 |
| Antineoplastic agents | 1.3 | 1.4 | 0.01 | 1.4 | 1.4 | 0.00 | 1.8 | 1.4 | -0.03 |
| Antipsoriatics | 0.6 | 0.4 | -0.02 | 0.5 | 0.6 | 0.00 | 0.5 | 0.5 | -0.01 |

| (Continued) | Before ajustment | | | After matching | | | After stratification | | |
|---|---|---|---|---|---|---|---|---|---|
| Characteristic | T (%) | C (%) | StdDiff | T (%) | C (%) | StdDiff | T (%) | C (%) | StdDiff |
| Antithrombotic agents | 47.4 | 42.5 | -0.10 | 43.2 | 42.9 | -0.01 | 39.6 | 43.6 | 0.08 |
| Beta blocking agents | 59.2 | 56.5 | -0.05 | 56.4 | 55.5 | -0.02 | 53.1 | 56.9 | 0.08 |
| Calcium channel blockers | 41.2 | 38.6 | -0.05 | 40.6 | 40.1 | -0.01 | 39.5 | 39.8 | 0.01 |
| Diuretics | 59.2 | 60.0 | 0.01 | 57.0 | 57.2 | 0.00 | 56.3 | 58.9 | 0.05 |
| Drugs for acid related disorders | 38.1 | 39.7 | 0.03 | 37.5 | 36.7 | -0.02 | 38.3 | 39.8 | 0.03 |
| Drugs for obstructive airway diseases | 18.1 | 17.8 | -0.01 | 17.9 | 17.7 | -0.01 | 17.3 | 18.0 | 0.02 |
| Immunosuppressants | 0.7 | 0.8 | 0.00 | 0.8 | 0.7 | -0.01 | 0.5 | 0.7 | 0.02 |
| Lipid modifying agents | 61.9 | 51.6 | -0.21 | 57.1 | 57.7 | 0.01 | 51.6 | 54.5 | 0.06 |
| Opioids | 10.7 | 13.3 | 0.08 | 11.2 | 10.1 | -0.04 | 12.2 | 12.7 | 0.01 |
| Psycholeptics | 9.5 | 11.8 | 0.07 | 9.8 | 10.4 | 0.02 | 11.6 | 11.4 | -0.01 |
| Psychostimulants | 0.2 | 0.3 | 0.03 | 0.2 | 0.3 | 0.02 | 0.2 | 0.3 | 0.02 |

##### 2.4.7 SIDIAP

**Supplementary Table 20:** Baseline patient characteristics for SGLT2I (T) and DPP4I (C) new-users in the SIDIAP data source. We report proportion of initiators satisfying selected base-line characteristics and the standardized difference of population proportions (StdDiff) before and after propensity score adjustment. Less extreme StdDiffs through matching and stratification suggest improved balance between patient cohorts through adjustment.

| | Before adjustment | | | After matching | | | After stratification | | |
|---|---|---|---|---|---|---|---|---|---|
| Characteristic | T (%) | C (%) | StdDiff | T (%) | C (%) | StdDiff | T (%) | C (%) | StdDiff |
| **Age group** | | | | | | | | | |
| 65 - 69 | 37.2 | 26.3 | -0.24 | 36.9 | 37.0 | 0.00 | 30.0 | 26.9 | -0.07 |
| 70 - 74 | 27.9 | 25.6 | -0.05 | 28.2 | 28.9 | 0.02 | 28.3 | 25.8 | -0.06 |
| 75 - 79 | 20.3 | 20.8 | 0.01 | 19.9 | 19.1 | -0.02 | 19.5 | 20.5 | 0.03 |
| 80 - 84 | 9.5 | 15.1 | 0.17 | 9.5 | 9.1 | -0.01 | 12.7 | 14.5 | 0.05 |
| 85 - 89 | 4.1 | 8.9 | 0.20 | 4.3 | 4.3 | 0.00 | 6.8 | 8.9 | 0.08 |
| 90 - 94 | 0.9 | 2.8 | 0.14 | 1.0 | 1.2 | 0.01 | 2.3 | 2.9 | 0.03 |
| 95 - 99 | 0.1 | 0.5 | 0.06 | 0.2 | 0.3 | 0.03 | 0.3 | 0.6 | 0.04 |
| Gender: female | 37.9 | 47.1 | 0.19 | 40.8 | 40.5 | -0.01 | 48.6 | 45.7 | -0.06 |
| **Medical history: General** | | | | | | | | | |
| Acute respiratory disease | 13.6 | 13.4 | 0.00 | 13.5 | 13.3 | 0.00 | 14.1 | 14.5 | 0.01 |
| Chronic liver disease | 0.2 | 0.4 | 0.03 | 0.3 | <0.2 | -0.02 | 0.2 | 0.3 | 0.03 |
| Chronic obstructive lung disease | 1.9 | 1.8 | -0.01 | 1.7 | 1.5 | -0.01 | 1.1 | 1.7 | 0.04 |
| Dementia | 0.4 | 1.1 | 0.07 | 0.5 | 0.5 | 0.00 | 1.0 | 0.9 | -0.02 |
| Depressive disorder | 1.4 | 2.4 | 0.07 | 1.4 | 2.1 | 0.05 | 2.5 | 2.1 | -0.03 |
| Gastroesophageal reflux disease | 1.2 | 1.3 | 0.01 | 1.1 | 1.3 | 0.02 | 1.6 | 1.5 | -0.01 |
| Gastrointestinal hemorrhage | 1.0 | 1.8 | 0.07 | 1.1 | 1.3 | 0.03 | 1.7 | 1.7 | 0.00 |
| Hyperlipidemia | 2.0 | 2.7 | 0.04 | 2.3 | 2.2 | 0.00 | 2.3 | 2.2 | 0.00 |
| Hypertensive disorder | 3.3 | 3.6 | 0.01 | 3.4 | 3.0 | -0.02 | 3.3 | 3.0 | -0.02 |
| Lesion of liver | 0.3 | 0.3 | 0.01 | 0.3 | <0.2 | -0.04 | 0.2 | 0.3 | 0.02 |
| Obesity | 5.9 | 6.7 | 0.04 | 6.1 | 5.6 | -0.02 | 7.7 | 6.4 | -0.05 |
| Osteoarthritis | 4.0 | 5.9 | 0.09 | 4.1 | 4.5 | 0.02 | 4.2 | 5.3 | 0.05 |
| Pneumonia | 1.9 | 1.8 | 0.00 | 2.1 | 2.0 | -0.01 | 2.1 | 2.0 | -0.01 |
| Psoriasis | 0.3 | 0.3 | 0.00 | 0.3 | 0.3 | -0.01 | 0.3 | 0.2 | -0.02 |
| Renal impairment | 2.5 | 6.1 | 0.18 | 2.7 | 2.8 | 0.01 | 4.3 | 5.9 | 0.07 |
| Rheumatoid arthritis | 0.1 | 0.1 | -0.01 | 0.1 | <0.2 | -0.01 | 0.1 | 0.1 | -0.01 |
| Urinary tract infectious disease | 6.9 | 8.3 | 0.06 | 8.5 | 9.0 | 0.02 | 8.5 | 9.0 | 0.02 |
| Viral hepatitis C | <0.1 | 0.2 | 0.05 | <0.1 | <0.2 | 0.03 | <0.1 | 0.2 | 0.05 |
| Visual system disorder | 14.6 | 18.6 | 0.11 | 15.1 | 14.9 | 0.00 | 16.5 | 18.5 | 0.05 |
| **Medical history: Cardiovascular disease** | | | | | | | | | |
| Atrial fibrillation | 3.6 | 2.5 | -0.07 | 2.7 | 3.0 | 0.01 | 2.2 | 2.6 | 0.03 |
| Cerebrovascular disease | 1.2 | 1.7 | 0.04 | 1.3 | 1.3 | 0.00 | 1.3 | 1.5 | 0.02 |
| Heart disease | 17.0 | 11.5 | -0.16 | 12.3 | 12.8 | 0.01 | 10.2 | 11.7 | 0.05 |
| Heart failure | 4.8 | 2.8 | -0.11 | 3.1 | 2.7 | -0.03 | 2.8 | 2.8 | 0.00 |
| Ischemic heart disease | 6.6 | 2.8 | -0.18 | 3.8 | 4.2 | 0.02 | 3.8 | 4.2 | 0.02 |
| Peripheral vascular disease | 1.2 | 1.1 | -0.01 | 1.0 | 0.8 | -0.02 | 0.9 | 1.0 | 0.01 |
| Pulmonary embolism | 0.2 | 0.3 | 0.02 | 0.2 | 0.3 | 0.02 | 0.3 | 0.4 | 0.02 |
| Venous thrombosis | 1.0 | 1.1 | 0.01 | 1.1 | 0.5 | -0.06 | 1.9 | 1.1 | -0.07 |
| **Medical history: Neoplasms** | | | | | | | | | |
| Hematologic neoplasm | <0.1 | 0.1 | 0.02 | <0.1 | <0.2 | 0.02 | 0.3 | 0.2 | -0.02 |
| Malignant neoplasm of anorectum | <0.1 | 0.1 | 0.02 | <0.1 | <0.2 | 0.03 | <0.1 | 0.1 | 0.01 |
| Malignant neoplastic disease | 2.4 | 3.9 | 0.09 | 2.4 | 2.8 | 0.03 | 3.1 | 3.7 | 0.03 |
| Malignant tumor of breast | <0.1 | 0.3 | 0.04 | 0.1 | <0.2 | -0.02 | 0.3 | 0.3 | 0.00 |
| Malignant tumor of colon | 0.2 | 0.4 | 0.04 | 0.2 | <0.2 | -0.04 | 0.5 | 0.4 | -0.02 |
| Malignant tumor of lung | <0.1 | 0.1 | 0.00 | <0.1 | 0.1 | 0.01 | <0.1 | 0.1 | 0.01 |
| Malignant tumor of urinary bladder | <0.1 | 0.3 | 0.05 | <0.1 | <0.2 | 0.01 | 0.1 | 0.3 | 0.04 |
| Primary malignant neoplasm of prostate | 0.2 | 0.5 | 0.04 | 0.3 | 0.5 | 0.04 | 0.2 | 0.4 | 0.04 |
| **Medication use** | | | | | | | | | |
| Agents acting on the renin-angiotensin system | 76.1 | 69.7 | -0.14 | 73.0 | 74.1 | 0.03 | 68.8 | 71.1 | 0.05 |
| Antibacterials for systemic use | 39.7 | 41.6 | 0.04 | 39.7 | 38.9 | -0.02 | 41.8 | 42.9 | 0.02 |
| Antidepressants | 21.2 | 23.2 | 0.05 | 22.0 | 22.6 | 0.01 | 24.2 | 23.7 | -0.01 |
| Antiepileptics | 10.0 | 10.4 | 0.01 | 10.2 | 10.0 | 0.00 | 11.0 | 10.9 | -0.01 |
| Antiinflammatory and antirheumatic products | 55.4 | 53.2 | -0.04 | 52.7 | 53.5 | 0.02 | 52.8 | 51.2 | -0.03 |
| Antineoplastic agents | 1.8 | 2.3 | 0.04 | 1.8 | 2.2 | 0.03 | 1.7 | 2.1 | 0.03 |
| Antipsoriatics | 2.0 | 1.8 | -0.02 | 2.0 | 2.2 | 0.02 | 1.3 | 1.9 | 0.05 |
| Antithrombotic agents | 57.8 | 46.0 | -0.24 | 51.5 | 52.6 | 0.02 | 41.4 | 47.7 | 0.13 |
| Beta blocking agents | 49.4 | 32.6 | -0.35 | 42.0 | 43.3 | 0.03 | 32.3 | 35.8 | 0.07 |
| Calcium channel blockers | 32.0 | 30.8 | -0.03 | 31.6 | 31.2 | -0.01 | 28.6 | 31.4 | 0.06 |
| Diuretics | 61.2 | 56.6 | -0.10 | 57.7 | 58.2 | 0.01 | 56.0 | 57.5 | 0.03 |

| (Continued) | Before ajustment | | | After matching | | | After stratification | | |
|---|---|---|---|---|---|---|---|---|---|
| Characteristic | T (%) | C (%) | StdDiff | T (%) | C (%) | StdDiff | T (%) | C (%) | StdDiff |
| Drugs for acid related disorders | 63.7 | 62.8 | -0.02 | 60.9 | 60.3 | -0.01 | 61.3 | 62.5 | 0.02 |
| Drugs for obstructive airway diseases | 34.1 | 30.7 | -0.07 | 33.2 | 34.0 | 0.02 | 31.9 | 32.0 | 0.00 |
| Immunosuppressants | 1.4 | 1.4 | 0.00 | 1.4 | 1.8 | 0.03 | 1.2 | 1.5 | 0.03 |
| Lipid modifying agents | 74.3 | 66.7 | -0.17 | 71.4 | 71.9 | 0.01 | 68.0 | 67.9 | 0.00 |
| Opioids | 20.7 | 20.8 | 0.00 | 20.9 | 19.9 | -0.02 | 22.0 | 21.6 | -0.01 |
| Psycholeptics | 30.6 | 35.3 | 0.10 | 31.1 | 31.9 | 0.02 | 36.1 | 35.0 | -0.02 |
| Psychostimulants | 9.5 | 4.4 | -0.20 | 6.5 | 7.5 | 0.04 | 4.3 | 4.6 | 0.01 |

#### 2.5 SGLT2I vs SU

##### 2.5.1 OptumDOD

**Supplementary Table 21:** Baseline patient characteristics for SGLT2I (T) and SU (C) new-users in the OptumDOD data source. We report proportion of initiators satisfying selected base-line characteristics and the standardized difference of population proportions (StdDiff) before and after propensity score adjustment. Less extreme StdDiffs through matching and stratification suggest improved balance between patient cohorts through adjustment.

| | Before adjustment | | | After matching | | | After stratification | | |
|---|---|---|---|---|---|---|---|---|---|
| Characteristic | T (%) | C (%) | StdDiff | T (%) | C (%) | StdDiff | T (%) | C (%) | StdDiff |
| **Age group** | | | | | | | | | |
| 65 - 69 | 40.3 | 32.8 | 0.16 | 39.8 | 40.0 | 0.00 | 36.2 | 34.3 | 0.04 |
| 70 - 74 | 33.8 | 31.2 | 0.06 | 33.9 | 34.1 | -0.01 | 32.8 | 32.5 | 0.00 |
| 75 - 79 | 16.9 | 20.0 | -0.08 | 17.1 | 16.9 | 0.00 | 17.5 | 18.7 | -0.03 |
| 80 - 84 | 6.2 | 10.9 | -0.16 | 6.4 | 6.2 | 0.01 | 8.9 | 9.5 | -0.02 |
| 85 - 89 | 2.4 | 4.8 | -0.13 | 2.5 | 2.4 | 0.00 | 4.4 | 4.6 | -0.01 |
| 90 - 94 | 0.3 | 0.3 | 0.00 | 0.3 | 0.3 | -0.01 | 0.2 | 0.3 | -0.03 |
| Gender: female | 45.2 | 49.6 | -0.09 | 46.0 | 46.1 | 0.00 | 49.4 | 49.0 | 0.01 |
| **Medical history: General** | | | | | | | | | |
| Acute respiratory disease | 22.6 | 23.2 | -0.01 | 23.9 | 22.8 | 0.03 | 23.9 | 22.8 | 0.03 |
| Attention deficit hyperactivity disorder | 0.3 | 0.2 | 0.03 | 0.3 | 0.3 | 0.01 | 0.2 | 0.2 | 0.00 |
| Chronic liver disease | 1.5 | 2.5 | -0.07 | 1.5 | 1.6 | -0.01 | 2.8 | 2.1 | 0.04 |
| Chronic obstructive lung disease | 12.3 | 13.7 | -0.04 | 12.0 | 12.2 | -0.01 | 12.0 | 12.2 | -0.01 |
| Crohn’s disease | 0.3 | 0.2 | 0.01 | 0.3 | 0.2 | 0.02 | 0.3 | 0.2 | 0.01 |
| Dementia | 1.9 | 4.1 | -0.13 | 2.0 | 2.0 | 0.00 | 4.0 | 3.8 | 0.01 |
| Depressive disorder | 15.5 | 13.8 | 0.05 | 15.4 | 15.3 | 0.00 | 15.7 | 15.2 | 0.01 |
| Gastroesophageal reflux disease | 22.1 | 19.3 | 0.07 | 21.5 | 21.4 | 0.00 | 21.7 | 20.9 | 0.02 |
| Gastrointestinal hemorrhage | 2.5 | 3.3 | -0.05 | 2.4 | 2.4 | 0.00 | 3.5 | 2.9 | 0.03 |
| Human immunodeficiency virus infection | 0.2 | 0.1 | 0.03 | 0.2 | 0.2 | 0.00 | 0.2 | 0.1 | 0.00 |
| Hyperlipidemia | 86.3 | 82.4 | 0.11 | 85.7 | 86.2 | -0.01 | 82.4 | 83.7 | -0.03 |
| Hypertensive disorder | 87.1 | 85.2 | 0.05 | 86.6 | 86.4 | 0.01 | 85.0 | 85.6 | -0.02 |
| Lesion of liver | 1.4 | 1.5 | -0.01 | 1.3 | 1.4 | 0.00 | 2.0 | 1.6 | 0.03 |
| Obesity | 26.3 | 17.8 | 0.21 | 25.1 | 25.7 | -0.01 | 21.1 | 21.6 | -0.01 |
| Osteoarthritis | 34.0 | 32.1 | 0.04 | 33.5 | 33.5 | 0.00 | 34.2 | 33.1 | 0.02 |
| Pneumonia | 4.3 | 5.6 | -0.06 | 4.1 | 4.1 | 0.00 | 5.4 | 5.2 | 0.01 |
| Psoriasis | 2.0 | 1.5 | 0.04 | 1.8 | 1.8 | 0.00 | 1.7 | 1.6 | 0.00 |
| Renal impairment | 19.1 | 24.3 | -0.13 | 19.1 | 18.3 | 0.02 | 22.8 | 24.0 | -0.03 |
| Rheumatoid arthritis | 2.0 | 2.1 | -0.01 | 2.0 | 2.0 | 0.00 | 1.8 | 2.1 | -0.02 |
| Schizophrenia | 0.2 | 0.2 | -0.02 | 0.1 | 0.1 | 0.00 | 0.1 | 0.2 | -0.03 |
| Ulcerative colitis | 0.4 | 0.4 | 0.01 | 0.4 | 0.3 | 0.01 | 0.4 | 0.3 | 0.01 |
| Urinary tract infectious disease | 8.5 | 13.6 | -0.16 | 12.8 | 12.2 | 0.02 | 12.8 | 12.2 | 0.02 |
| Viral hepatitis C | 0.5 | 0.5 | -0.01 | 0.5 | 0.5 | -0.01 | 0.5 | 0.6 | -0.01 |
| Visual system disorder | 54.8 | 51.9 | 0.06 | 54.5 | 54.4 | 0.00 | 52.7 | 53.4 | -0.01 |
| **Medical history: Cardiovascular disease** | | | | | | | | | |
| Atrial fibrillation | 11.6 | 11.0 | 0.02 | 10.7 | 10.7 | 0.00 | 10.4 | 10.9 | -0.02 |
| Cerebrovascular disease | 9.3 | 10.3 | -0.03 | 8.9 | 8.9 | 0.00 | 8.9 | 9.5 | -0.02 |
| Coronary arteriosclerosis | 27.8 | 23.0 | 0.11 | 25.5 | 25.8 | -0.01 | 21.7 | 23.4 | -0.04 |
| Heart disease | 45.7 | 42.1 | 0.07 | 43.5 | 43.3 | 0.00 | 40.6 | 41.8 | -0.03 |
| Heart failure | 12.6 | 11.5 | 0.03 | 10.8 | 10.9 | 0.00 | 10.5 | 11.2 | -0.02 |
| Ischemic heart disease | 14.5 | 12.3 | 0.06 | 11.7 | 12.1 | -0.01 | 11.7 | 12.1 | -0.01 |
| Peripheral vascular disease | 18.2 | 16.1 | 0.06 | 17.5 | 17.4 | 0.00 | 16.6 | 17.0 | -0.01 |
| Pulmonary embolism | 1.1 | 1.1 | 0.00 | 1.1 | 1.0 | 0.01 | 1.4 | 1.1 | 0.03 |
| Venous thrombosis | 1.9 | 2.3 | -0.03 | 2.3 | 2.2 | 0.01 | 2.3 | 2.2 | 0.01 |
| **Medical history: Neoplasms** | | | | | | | | | |
| Hematologic neoplasm | 1.2 | 1.8 | -0.05 | 1.2 | 1.4 | -0.01 | 1.3 | 1.7 | -0.04 |
| Malignant lymphoma | 0.7 | 0.8 | -0.01 | 0.7 | 0.8 | -0.01 | 0.6 | 0.8 | -0.03 |
| Malignant neoplasm of anorectum | 0.2 | 0.3 | -0.04 | 0.2 | 0.3 | -0.02 | 0.4 | 0.3 | 0.02 |
| Malignant neoplastic disease | 14.5 | 15.8 | -0.04 | 14.3 | 14.3 | 0.00 | 14.6 | 15.3 | -0.02 |
| Malignant tumor of breast | 2.0 | 2.5 | -0.03 | 2.1 | 2.2 | -0.01 | 2.0 | 2.5 | -0.03 |
| Malignant tumor of colon | 0.7 | 0.9 | -0.02 | 0.7 | 0.6 | 0.01 | 1.3 | 0.8 | 0.04 |
| Malignant tumor of lung | 0.5 | 0.8 | -0.04 | 0.4 | 0.5 | -0.01 | 0.5 | 0.8 | -0.04 |
| Malignant tumor of urinary bladder | 0.6 | 0.9 | -0.04 | 0.6 | 0.7 | -0.02 | 0.5 | 0.9 | -0.04 |
| Primary malignant neoplasm of prostate | 3.5 | 3.7 | -0.01 | 3.5 | 3.4 | 0.01 | 3.2 | 3.5 | -0.02 |
| **Medication use** | | | | | | | | | |
| Agents acting on the renin-angiotensin system | 78.1 | 75.6 | 0.06 | 77.4 | 77.5 | 0.00 | 75.0 | 76.3 | -0.03 |
| Antibacterials for systemic use | 56.6 | 57.4 | -0.02 | 56.1 | 56.0 | 0.00 | 57.4 | 56.6 | 0.02 |

| (Continued) | Before ajustment | | | After matching | | | After stratification | | |
|---|---|---|---|---|---|---|---|---|---|
| Characteristic | T (%) | C (%) | StdDiff | T (%) | C (%) | StdDiff | T (%) | C (%) | StdDiff |
| Antidepressants | 28.2 | 25.8 | 0.05 | 27.7 | 27.8 | 0.00 | 27.3 | 27.0 | 0.01 |
| Antiepileptics | 19.1 | 17.5 | 0.04 | 18.9 | 19.3 | -0.01 | 19.6 | 19.3 | 0.01 |
| Antiinflammatory and antirheumatic products | 29.6 | 26.6 | 0.07 | 29.2 | 30.0 | -0.02 | 28.9 | 27.8 | 0.03 |
| Antineoplastic agents | 6.4 | 6.3 | 0.00 | 6.3 | 6.4 | 0.00 | 6.6 | 6.3 | 0.01 |
| Antipsoriatics | 0.5 | 0.5 | 0.00 | 0.5 | 0.5 | 0.00 | 0.4 | 0.5 | -0.03 |
| Antithrombotic agents | 24.6 | 21.5 | 0.07 | 23.1 | 23.0 | 0.00 | 21.6 | 21.9 | -0.01 |
| Beta blocking agents | 47.2 | 45.5 | 0.03 | 45.5 | 45.7 | -0.01 | 44.5 | 45.1 | -0.01 |
| Calcium channel blockers | 33.4 | 34.0 | -0.01 | 33.4 | 33.6 | 0.00 | 33.5 | 34.1 | -0.01 |
| Diuretics | 49.2 | 49.8 | -0.01 | 48.4 | 48.2 | 0.00 | 47.4 | 48.5 | -0.02 |
| Drugs for acid related disorders | 32.3 | 31.8 | 0.01 | 32.0 | 31.9 | 0.00 | 33.2 | 32.5 | 0.01 |
| Drugs for obstructive airway diseases | 42.0 | 37.7 | 0.09 | 41.2 | 41.8 | -0.01 | 40.9 | 39.0 | 0.04 |
| Immunosuppressants | 3.2 | 2.5 | 0.04 | 3.1 | 2.9 | 0.01 | 3.1 | 2.7 | 0.03 |
| Lipid modifying agents | 85.8 | 78.7 | 0.19 | 85.0 | 85.5 | -0.01 | 79.7 | 80.9 | -0.03 |
| Opioids | 26.3 | 26.3 | 0.00 | 26.6 | 26.3 | 0.01 | 26.6 | 26.3 | 0.01 |
| Psycholeptics | 23.7 | 22.8 | 0.02 | 23.2 | 23.0 | 0.00 | 24.5 | 23.4 | 0.02 |
| Psychostimulants | 1.1 | 0.7 | 0.05 | 1.1 | 0.9 | 0.02 | 1.1 | 0.8 | 0.03 |

##### 2.5.2 MDCR

**Supplementary Table 22:** Baseline patient characteristics for SGLT2I (T) and SU (C) new-users in the MDCR data source. We report proportion of initiators satisfying selected base-line characteristics and the standardized difference of population proportions (StdDiff) before and after propensity score adjustment. Less extreme StdDiffs through matching and stratification suggest improved balance between patient cohorts through adjustment.

| | Before adjustment | | | After matching | | | After stratification | | |
|---|---|---|---|---|---|---|---|---|---|
| Characteristic | T (%) | C (%) | StdDiff | T (%) | C (%) | StdDiff | T (%) | C (%) | StdDiff |
| **Age group** | | | | | | | | | |
| 65 - 69 | 39.3 | 28.1 | 0.24 | 38.3 | 36.7 | 0.03 | 34.2 | 32.2 | 0.04 |
| 70 - 74 | 35.6 | 30.2 | 0.12 | 35.6 | 36.6 | -0.02 | 31.6 | 31.6 | 0.00 |
| 75 - 79 | 17.3 | 21.3 | -0.10 | 17.6 | 17.9 | -0.01 | 17.9 | 19.1 | -0.03 |
| 80 - 84 | 5.6 | 12.9 | -0.25 | 6.1 | 5.8 | 0.01 | 11.0 | 10.5 | 0.02 |
| 85 - 89 | 1.8 | 5.6 | -0.20 | 2.0 | 2.0 | 0.00 | 4.2 | 4.9 | -0.04 |
| 90 - 94 | 0.4 | 1.6 | -0.13 | 0.4 | 0.8 | -0.06 | 1.3 | 1.5 | -0.02 |
| 95 - 99 | <0.1 | 0.2 | -0.04 | <0.2 | 0.1 | -0.02 | <0.1 | 0.2 | -0.04 |
| Gender: female | 41.5 | 46.8 | -0.11 | 42.4 | 42.8 | -0.01 | 46.1 | 45.3 | 0.02 |
| **Medical history: General** | | | | | | | | | |
| Acute respiratory disease | 22.8 | 21.0 | 0.04 | 22.7 | 22.7 | 0.00 | 25.7 | 23.2 | 0.06 |
| Attention deficit hyperactivity disorder | 0.2 | 0.1 | 0.02 | 0.2 | 0.4 | -0.03 | 0.2 | 0.2 | -0.02 |
| Chronic liver disease | 1.9 | 2.1 | -0.01 | 2.1 | 1.6 | 0.04 | 2.2 | 2.2 | 0.00 |
| Chronic obstructive lung disease | 9.9 | 10.9 | -0.04 | 9.9 | 10.2 | -0.01 | 10.9 | 11.6 | -0.02 |
| Crohn's disease | 0.3 | 0.3 | 0.02 | 0.4 | 0.3 | 0.01 | 0.3 | 0.3 | 0.00 |
| Dementia | 1.2 | 3.0 | -0.13 | 1.2 | 1.5 | -0.03 | 1.9 | 3.5 | -0.10 |
| Depressive disorder | 9.3 | 6.7 | 0.10 | 9.2 | 9.7 | -0.02 | 12.4 | 10.3 | 0.06 |
| Gastroesophageal reflux disease | 17.3 | 10.1 | 0.21 | 16.7 | 16.7 | 0.00 | 15.4 | 16.1 | -0.02 |
| Gastrointestinal hemorrhage | 2.6 | 3.5 | -0.05 | 2.6 | 2.2 | 0.03 | 2.2 | 3.0 | -0.05 |
| Hyperlipidemia | 81.3 | 49.4 | 0.71 | 80.1 | 81.7 | -0.04 | 75.2 | 75.6 | -0.01 |
| Hypertensive disorder | 83.8 | 66.4 | 0.41 | 83.1 | 82.7 | 0.01 | 82.7 | 81.8 | 0.02 |
| Lesion of liver | 1.2 | 1.1 | 0.01 | 1.1 | 1.2 | -0.01 | 1.1 | 1.3 | -0.02 |
| Obesity | 23.7 | 7.7 | 0.45 | 22.2 | 22.8 | -0.01 | 16.6 | 17.5 | -0.02 |
| Osteoarthritis | 31.8 | 26.3 | 0.12 | 31.1 | 31.9 | -0.02 | 32.4 | 32.2 | 0.00 |
| Pneumonia | 3.7 | 5.9 | -0.10 | 3.5 | 3.5 | 0.00 | 6.2 | 5.2 | 0.04 |
| Psoriasis | 2.3 | 1.3 | 0.07 | 2.0 | 2.1 | -0.01 | 2.0 | 2.1 | -0.01 |
| Renal impairment | 11.9 | 12.4 | -0.01 | 12.3 | 11.9 | 0.01 | 13.3 | 17.1 | -0.10 |
| Rheumatoid arthritis | 1.5 | 1.5 | 0.00 | 1.4 | 1.6 | -0.02 | 1.3 | 1.7 | -0.03 |
| Schizophrenia | <0.1 | 0.1 | -0.01 | <0.1 | 0.1 | -0.02 | <0.1 | 0.1 | -0.02 |
| Ulcerative colitis | 0.5 | 0.4 | 0.01 | 0.4 | 0.3 | 0.01 | 0.4 | 0.4 | 0.00 |
| Urinary tract infectious disease | 7.1 | 10.4 | -0.12 | 7.3 | 7.4 | 0.00 | 11.4 | 10.6 | 0.03 |
| Viral hepatitis C | 0.5 | 0.2 | 0.04 | 0.5 | 0.2 | 0.04 | 0.3 | 0.3 | 0.00 |
| Visual system disorder | 55.1 | 48.1 | 0.14 | 54.6 | 54.4 | 0.00 | 53.3 | 52.9 | 0.01 |
| **Medical history: Cardiovascular disease** | | | | | | | | | |
| Atrial fibrillation | 12.1 | 11.4 | 0.02 | 11.5 | 11.0 | 0.02 | 10.9 | 12.3 | -0.04 |
| Cerebrovascular disease | 11.0 | 12.0 | -0.03 | 10.0 | 9.8 | 0.01 | 10.8 | 11.1 | -0.01 |
| Coronary arteriosclerosis | 28.9 | 24.1 | 0.11 | 26.2 | 27.1 | -0.02 | 21.4 | 25.3 | -0.09 |
| Heart disease | 47.4 | 43.8 | 0.07 | 44.3 | 44.3 | 0.00 | 43.9 | 44.3 | -0.01 |
| Heart failure | 9.9 | 10.6 | -0.03 | 8.8 | 7.9 | 0.03 | 9.9 | 10.0 | 0.00 |
| Ischemic heart disease | 12.0 | 11.8 | 0.01 | 11.3 | 11.0 | 0.01 | 10.4 | 11.3 | -0.03 |
| Peripheral vascular disease | 13.8 | 10.8 | 0.09 | 12.7 | 12.6 | 0.00 | 11.4 | 12.8 | -0.04 |
| Pulmonary embolism | 0.9 | 0.9 | 0.00 | 0.9 | 0.9 | -0.01 | 1.7 | 1.1 | 0.05 |
| Venous thrombosis | 2.0 | 2.2 | -0.02 | 2.1 | 1.9 | 0.01 | 2.6 | 2.4 | 0.01 |
| **Medical history: Neoplasms** | | | | | | | | | |
| Hematologic neoplasm | 1.4 | 1.8 | -0.03 | 1.4 | 1.6 | -0.01 | 1.1 | 2.1 | -0.08 |
| Malignant lymphoma | 1.1 | 0.8 | 0.03 | 0.9 | 1.0 | -0.02 | 0.9 | 1.1 | -0.02 |
| Malignant neoplasm of anorectum | 0.3 | 0.4 | -0.02 | 0.3 | 0.3 | 0.00 | 0.5 | 0.3 | 0.02 |
| Malignant neoplastic disease | 16.9 | 18.1 | -0.03 | 17.1 | 18.6 | -0.04 | 17.1 | 18.6 | -0.04 |
| Malignant tumor of breast | 2.5 | 2.7 | -0.01 | 2.6 | 2.5 | 0.01 | 2.7 | 2.8 | -0.01 |
| Malignant tumor of colon | 0.6 | 0.9 | -0.03 | 0.6 | 0.5 | 0.01 | 0.7 | 0.8 | -0.01 |
| Malignant tumor of lung | 0.3 | 0.8 | -0.06 | 0.4 | 0.6 | -0.04 | 0.3 | 0.9 | -0.08 |
| Malignant tumor of urinary bladder | 1.0 | 1.1 | -0.02 | 1.0 | 0.8 | 0.01 | 1.2 | 1.1 | 0.01 |
| Primary malignant neoplasm of prostate | 4.3 | 4.5 | -0.01 | 4.3 | 4.2 | 0.01 | 4.0 | 4.5 | -0.02 |
| **Medication use** | | | | | | | | | |
| Agents acting on the renin-angiotensin system | 76.2 | 72.3 | 0.09 | 75.9 | 75.6 | 0.01 | 76.9 | 75.0 | 0.05 |
| Antibacterials for systemic use | 60.6 | 60.6 | 0.00 | 59.6 | 59.9 | -0.01 | 60.1 | 60.7 | -0.01 |
| Antidepressants | 24.2 | 24.2 | 0.00 | 24.2 | 24.3 | 0.00 | 25.9 | 24.9 | 0.02 |
| Antiepileptics | 15.8 | 13.8 | 0.06 | 15.4 | 15.4 | 0.00 | 17.1 | 16.0 | 0.03 |
| Antiinflammatory and antirheumatic products | 29.5 | 26.8 | 0.06 | 29.1 | 29.6 | -0.01 | 29.2 | 27.8 | 0.03 |

| (Continued) | Before ajustment | | | After matching | | | After stratification | | |
|---|---|---|---|---|---|---|---|---|---|
| Characteristic | T (%) | C (%) | StdDiff | T (%) | C (%) | StdDiff | T (%) | C (%) | StdDiff |
| Antineoplastic agents | 7.9 | 8.8 | -0.03 | 7.7 | 7.8 | 0.00 | 8.8 | 7.8 | 0.04 |
| Antipsoriatics | 0.8 | 0.7 | 0.01 | 0.7 | 0.8 | -0.01 | 0.4 | 0.7 | -0.04 |
| Antithrombotic agents | 25.1 | 25.2 | 0.00 | 23.7 | 23.8 | 0.00 | 22.9 | 25.3 | -0.06 |
| Beta blocking agents | 48.5 | 48.6 | 0.00 | 47.2 | 47.4 | 0.00 | 50.0 | 49.0 | 0.02 |
| Calcium channel blockers | 32.8 | 33.4 | -0.01 | 32.7 | 32.5 | 0.00 | 34.7 | 34.3 | 0.01 |
| Diuretics | 48.8 | 53.2 | -0.09 | 48.2 | 47.3 | 0.02 | 49.7 | 49.9 | 0.00 |
| Drugs for acid related disorders | 33.8 | 31.7 | 0.04 | 33.0 | 32.2 | 0.01 | 32.4 | 32.1 | 0.01 |
| Drugs for obstructive airway diseases | 44.1 | 37.5 | 0.13 | 43.1 | 43.4 | 0.00 | 39.4 | 41.5 | -0.04 |
| Immunosuppressants | 3.6 | 2.7 | 0.05 | 3.4 | 3.5 | -0.01 | 3.1 | 3.4 | -0.01 |
| Lipid modifying agents | 83.9 | 75.4 | 0.21 | 82.7 | 83.5 | -0.02 | 81.8 | 80.3 | 0.04 |
| Opioids | 24.9 | 27.9 | -0.07 | 24.7 | 25.3 | -0.01 | 31.3 | 28.0 | 0.07 |
| Psycholeptics | 23.0 | 24.9 | -0.04 | 23.0 | 22.7 | 0.01 | 25.6 | 24.3 | 0.03 |
| Psychostimulants | 1.5 | 1.1 | 0.04 | 1.5 | 1.3 | 0.01 | 1.3 | 1.2 | 0.01 |

##### 2.5.3 Open Claims

**Supplementary Table 23:** Baseline patient characteristics for SGLT2I (T) and SU (C) new-users in the Open Claims data source. We report proportion of initiators satisfying selected base-line characteristics and the standardized difference of population proportions (StdDiff) before and after propensity score adjustment. Less extreme StdDiffs through matching and stratification suggest improved balance between patient cohorts through adjustment.

| | Before adjustment | | | After matching | | | After stratification | | |
|---|---|---|---|---|---|---|---|---|---|
| Characteristic | T (%) | C (%) | StdDiff | T (%) | C (%) | StdDiff | T (%) | C (%) | StdDiff |
| **Age group** | | | | | | | | | |
| 65 - 69 | 43.1 | 36.4 | 0.14 | 42.8 | 43.4 | -0.01 | 40.0 | 38.1 | 0.04 |
| 70 - 74 | 29.2 | 28.3 | 0.02 | 29.2 | 29.5 | -0.01 | 28.5 | 28.5 | 0.00 |
| 75 - 79 | 17.5 | 25.6 | -0.20 | 17.7 | 16.9 | 0.02 | 22.0 | 23.6 | -0.04 |
| 80 - 84 | 9.5 | 9.4 | 0.00 | 9.6 | 9.5 | 0.00 | 9.1 | 9.5 | -0.01 |
| 85 - 89 | 0.8 | 0.2 | 0.07 | 0.6 | 0.6 | 0.00 | 0.3 | 0.4 | 0.00 |
| Gender: female | 44.7 | 49.5 | -0.10 | 45.6 | 46.2 | -0.01 | 48.5 | 48.6 | 0.00 |
| **Medical history: General** | | | | | | | | | |
| Acute respiratory disease | 11.5 | 12.6 | -0.03 | 11.3 | 11.6 | -0.01 | 12.6 | 12.5 | 0.00 |
| Attention deficit hyperactivity disorder | 0.2 | 0.1 | 0.02 | 0.2 | 0.2 | 0.00 | 0.2 | 0.1 | 0.01 |
| Chronic liver disease | 0.9 | 1.3 | -0.03 | 0.9 | 1.0 | 0.00 | 1.3 | 1.2 | 0.01 |
| Chronic obstructive lung disease | 7.2 | 8.1 | -0.03 | 6.9 | 7.0 | 0.00 | 7.9 | 7.9 | 0.00 |
| Crohn's disease | 0.2 | 0.2 | 0.00 | 0.2 | 0.2 | 0.00 | 0.2 | 0.2 | 0.01 |
| Dementia | 1.0 | 1.9 | -0.07 | 1.0 | 1.1 | 0.00 | 1.7 | 1.7 | 0.00 |
| Depressive disorder | 6.0 | 6.0 | 0.00 | 6.2 | 6.1 | 0.00 | 6.2 | 6.1 | 0.00 |
| Gastroesophageal reflux disease | 8.9 | 8.4 | 0.02 | 8.7 | 9.2 | -0.02 | 8.7 | 8.6 | 0.00 |
| Gastrointestinal hemorrhage | 1.4 | 1.7 | -0.03 | 1.3 | 1.4 | 0.00 | 1.6 | 1.6 | 0.00 |
| Human immunodeficiency virus infection | 0.2 | 0.1 | 0.01 | 0.2 | 0.1 | 0.00 | 0.1 | 0.1 | 0.00 |
| Hyperlipidemia | 48.5 | 44.8 | 0.07 | 47.4 | 47.5 | 0.00 | 46.3 | 45.6 | 0.01 |
| Hypertensive disorder | 54.8 | 54.2 | 0.01 | 54.0 | 53.8 | 0.00 | 54.8 | 54.3 | 0.01 |
| Lesion of liver | 0.8 | 0.9 | -0.02 | 0.7 | 0.8 | 0.00 | 0.7 | 0.8 | 0.00 |
| Obesity | 9.6 | 7.3 | 0.08 | 9.1 | 9.4 | -0.01 | 8.2 | 7.9 | 0.01 |
| Osteoarthritis | 18.0 | 18.0 | 0.00 | 17.6 | 17.6 | 0.00 | 18.5 | 18.1 | 0.01 |
| Pneumonia | 2.8 | 3.5 | -0.04 | 2.6 | 2.7 | 0.00 | 2.6 | 2.7 | 0.00 |
| Psoriasis | 1.0 | 0.8 | 0.02 | 0.9 | 0.9 | 0.00 | 0.8 | 0.8 | 0.00 |
| Renal impairment | 10.4 | 12.5 | -0.07 | 9.9 | 9.0 | 0.03 | 11.5 | 11.9 | -0.01 |
| Rheumatoid arthritis | 1.2 | 1.2 | 0.00 | 1.1 | 1.1 | 0.00 | 1.4 | 1.2 | 0.01 |
| Schizophrenia | 0.1 | 0.2 | -0.01 | 0.1 | 0.1 | 0.00 | 0.2 | 0.2 | 0.00 |
| Ulcerative colitis | 0.2 | 0.2 | 0.00 | 0.2 | 0.2 | 0.00 | 0.2 | 0.2 | 0.00 |
| Urinary tract infectious disease | 5.0 | 6.9 | -0.08 | 5.0 | 4.9 | 0.00 | 7.0 | 6.6 | 0.02 |
| Viral hepatitis C | 0.3 | 0.3 | 0.00 | 0.2 | 0.3 | 0.00 | 0.2 | 0.3 | -0.01 |
| Visual system disorder | 25.0 | 25.0 | 0.00 | 24.8 | 24.4 | 0.01 | 24.8 | 24.4 | 0.01 |
| **Medical history: Cardiovascular disease** | | | | | | | | | |
| Atrial fibrillation | 8.1 | 7.4 | 0.02 | 7.3 | 7.2 | 0.00 | 7.3 | 7.2 | 0.00 |
| Cerebrovascular disease | 4.7 | 5.4 | -0.03 | 5.3 | 5.2 | 0.00 | 5.3 | 5.2 | 0.00 |
| Coronary arteriosclerosis | 16.4 | 13.4 | 0.08 | 14.9 | 14.0 | 0.02 | 13.8 | 13.7 | 0.00 |
| Heart disease | 30.3 | 27.0 | 0.07 | 28.0 | 27.0 | 0.02 | 27.5 | 27.2 | 0.01 |
| Heart failure | 8.6 | 6.8 | 0.06 | 7.0 | 6.4 | 0.02 | 7.0 | 6.4 | 0.02 |
| Ischemic heart disease | 7.2 | 5.9 | 0.05 | 6.3 | 6.0 | 0.02 | 5.9 | 6.0 | 0.00 |
| Peripheral vascular disease | 7.5 | 7.2 | 0.01 | 7.2 | 7.1 | 0.00 | 7.4 | 7.2 | 0.00 |
| Pulmonary embolism | 0.6 | 0.7 | -0.01 | 0.6 | 0.6 | 0.00 | 0.7 | 0.6 | 0.01 |
| Venous thrombosis | 1.0 | 1.2 | -0.02 | 1.0 | 0.9 | 0.00 | 1.3 | 1.2 | 0.01 |
| **Medical history: Neoplasms** | | | | | | | | | |
| Hematologic neoplasm | 0.9 | 1.1 | -0.03 | 1.1 | 1.1 | 0.00 | 1.1 | 1.1 | 0.00 |
| Malignant lymphoma | 0.6 | 0.7 | -0.01 | 0.6 | 0.6 | -0.01 | 0.7 | 0.7 | 0.00 |
| Malignant neoplasm of anorectum | 0.1 | 0.2 | -0.01 | 0.2 | 0.1 | 0.01 | 0.2 | 0.2 | 0.01 |
| Malignant neoplastic disease | 9.1 | 10.3 | -0.04 | 9.0 | 9.0 | 0.00 | 10.1 | 10.1 | 0.00 |
| Malignant tumor of breast | 1.5 | 1.7 | -0.02 | 1.5 | 1.5 | 0.00 | 1.7 | 1.7 | 0.00 |
| Malignant tumor of colon | 0.4 | 0.5 | -0.02 | 0.4 | 0.4 | 0.00 | 0.5 | 0.5 | 0.01 |
| Malignant tumor of lung | 0.3 | 0.6 | -0.03 | 0.3 | 0.4 | -0.01 | 0.5 | 0.5 | 0.00 |
| Malignant tumor of urinary bladder | 0.4 | 0.6 | -0.02 | 0.4 | 0.4 | 0.00 | 0.5 | 0.5 | 0.00 |
| Primary malignant neoplasm of prostate | 2.3 | 2.3 | 0.00 | 2.3 | 2.3 | 0.00 | 2.3 | 2.3 | 0.00 |
| **Medication use** | | | | | | | | | |
| Agents acting on the renin-angiotensin system | 74.4 | 72.6 | 0.04 | 73.5 | 73.0 | 0.01 | 72.3 | 72.8 | -0.01 |
| Antibacterials for systemic use | 55.4 | 56.1 | -0.01 | 54.9 | 55.2 | -0.01 | 57.6 | 56.2 | 0.03 |
| Antidepressants | 27.2 | 26.4 | 0.02 | 26.8 | 27.4 | -0.01 | 27.1 | 26.8 | 0.01 |
| Antiepileptics | 18.9 | 19.3 | -0.01 | 18.6 | 18.8 | 0.00 | 19.9 | 19.1 | 0.02 |
| Antiinflammatory and antirheumatic products | 33.6 | 31.0 | 0.06 | 33.2 | 33.0 | 0.00 | 32.9 | 31.9 | 0.02 |
| Antineoplastic agents | 6.5 | 5.6 | 0.04 | 6.3 | 6.3 | 0.00 | 6.2 | 5.8 | 0.02 |

| (Continued) | Before ajustment | | | After matching | | | After stratification | | |
|---|---|---|---|---|---|---|---|---|---|
| Characteristic | T (%) | C (%) | StdDiff | T (%) | C (%) | StdDiff | T (%) | C (%) | StdDiff |
| Antipsoriatics | 0.7 | 0.6 | 0.01 | 0.6 | 0.6 | 0.00 | 0.7 | 0.6 | 0.00 |
| Antithrombotic agents | 28.7 | 25.2 | 0.08 | 26.8 | 25.9 | 0.02 | 25.4 | 25.5 | 0.00 |
| Beta blocking agents | 47.5 | 45.3 | 0.04 | 45.6 | 44.7 | 0.02 | 44.8 | 45.2 | -0.01 |
| Calcium channel blockers | 33.2 | 32.9 | 0.01 | 32.9 | 32.6 | 0.01 | 32.1 | 33.1 | -0.02 |
| Diuretics | 47.9 | 47.7 | 0.00 | 46.7 | 46.3 | 0.01 | 47.1 | 47.5 | -0.01 |
| Drugs for acid related disorders | 37.1 | 36.1 | 0.02 | 36.3 | 36.4 | 0.00 | 36.9 | 36.5 | 0.01 |
| Drugs for obstructive airway diseases | 39.5 | 36.0 | 0.07 | 37.8 | 37.1 | 0.01 | 37.8 | 37.1 | 0.01 |
| Immunosuppressants | 3.3 | 2.8 | 0.03 | 3.1 | 3.0 | 0.00 | 3.1 | 2.9 | 0.01 |
| Lipid modifying agents | 81.1 | 75.1 | 0.14 | 80.1 | 79.6 | 0.01 | 75.5 | 76.1 | -0.01 |
| Opioids | 22.8 | 24.9 | -0.05 | 22.5 | 22.6 | 0.00 | 25.5 | 24.7 | 0.02 |
| Psycholeptics | 20.9 | 21.9 | -0.02 | 20.5 | 20.7 | 0.00 | 22.5 | 21.7 | 0.02 |
| Psychostimulants | 1.5 | 1.2 | 0.03 | 1.4 | 1.4 | 0.00 | 1.3 | 1.2 | 0.01 |

##### 2.5.4 OptumEHR

**Supplementary Table 24:** Baseline patient characteristics for SGLT2I (T) and SU (C) new-users in the OptumEHR data source. We report proportion of initiators satisfying selected base-line characteristics and the standardized difference of population proportions (StdDiff) before and after propensity score adjustment. Less extreme StdDiffs through matching and stratification suggest improved balance between patient cohorts through adjustment.

| | Before adjustment | | | After matching | | | After stratification | | |
|---|---|---|---|---|---|---|---|---|---|
| Characteristic | T (%) | C (%) | StdDiff | T (%) | C (%) | StdDiff | T (%) | C (%) | StdDiff |
| **Age group** | | | | | | | | | |
| 65 - 69 | 48.5 | 37.7 | 0.22 | 47.9 | 48.4 | -0.01 | 40.4 | 39.6 | 0.02 |
| 70 - 74 | 29.1 | 28.0 | 0.03 | 29.2 | 29.4 | 0.00 | 27.0 | 28.4 | -0.03 |
| 75 - 79 | 14.5 | 20.3 | -0.16 | 14.8 | 14.5 | 0.01 | 17.7 | 17.5 | 0.01 |
| 80 - 84 | 6.1 | 11.6 | -0.19 | 6.4 | 5.9 | 0.02 | 12.5 | 11.6 | 0.03 |
| 85 - 89 | 1.7 | 2.4 | -0.05 | 1.8 | 1.8 | 0.00 | 2.3 | 2.9 | -0.04 |
| Gender: female | 43.5 | 47.4 | -0.08 | 44.1 | 44.0 | 0.00 | 47.1 | 46.6 | 0.01 |
| **Race** | | | | | | | | | |
| Asian | 2.0 | 2.1 | -0.01 | 2.0 | 2.2 | -0.02 | 1.5 | 2.1 | -0.05 |
| Black or African American | 7.8 | 7.6 | 0.01 | 7.8 | 7.8 | 0.00 | 7.1 | 7.8 | -0.03 |
| White | 83.7 | 84.4 | -0.02 | 83.9 | 83.7 | 0.00 | 86.3 | 84.0 | 0.06 |
| **Ethnicity** | | | | | | | | | |
| Hispanic or Latino | 3.9 | 4.1 | -0.01 | 4.0 | 4.2 | -0.01 | 3.5 | 4.2 | -0.03 |
| Not Hispanic or Latino | 85.7 | 88.5 | -0.09 | 86.5 | 86.3 | 0.01 | 89.7 | 88.0 | 0.05 |
| **Medical history: General** | | | | | | | | | |
| Acute respiratory disease | 14.8 | 14.6 | 0.01 | 14.9 | 15.1 | 0.00 | 13.8 | 15.0 | -0.03 |
| Attention deficit hyperactivity disorder | 0.2 | 0.2 | 0.01 | 0.2 | 0.3 | -0.02 | 0.2 | 0.2 | 0.01 |
| Chronic liver disease | 1.6 | 2.2 | -0.04 | 1.7 | 1.8 | -0.01 | 1.8 | 2.1 | -0.03 |
| Chronic obstructive lung disease | 8.1 | 8.8 | -0.03 | 8.0 | 8.1 | 0.00 | 9.8 | 8.9 | 0.03 |
| Crohn’s disease | 0.3 | 0.2 | 0.02 | 0.3 | 0.3 | 0.00 | 0.3 | 0.2 | 0.00 |
| Dementia | 1.2 | 2.4 | -0.09 | 1.2 | 1.2 | 0.00 | 1.9 | 2.2 | -0.03 |
| Depressive disorder | 11.1 | 11.0 | 0.00 | 11.2 | 11.5 | -0.01 | 11.7 | 11.4 | 0.01 |
| Gastroesophageal reflux disease | 18.8 | 16.4 | 0.06 | 18.6 | 18.4 | 0.01 | 18.4 | 17.6 | 0.02 |
| Gastrointestinal hemorrhage | 1.1 | 1.7 | -0.05 | 1.1 | 1.2 | -0.01 | 1.6 | 1.6 | 0.00 |
| Human immunodeficiency virus infection | 0.1 | 0.1 | 0.01 | 0.1 | 0.1 | -0.01 | 0.1 | 0.1 | -0.01 |
| Hyperlipidemia | 73.7 | 68.7 | 0.11 | 72.4 | 73.3 | -0.02 | 69.4 | 68.9 | 0.01 |
| Hypertensive disorder | 74.8 | 72.4 | 0.06 | 74.0 | 74.6 | -0.01 | 71.9 | 72.5 | -0.01 |
| Lesion of liver | 1.1 | 1.0 | 0.01 | 1.1 | 1.0 | 0.01 | 2.0 | 1.0 | 0.08 |
| Obesity | 24.5 | 16.0 | 0.21 | 23.7 | 24.4 | -0.02 | 19.2 | 18.6 | 0.02 |
| Osteoarthritis | 21.6 | 21.5 | 0.00 | 21.5 | 21.8 | -0.01 | 23.4 | 22.1 | 0.03 |
| Pneumonia | 2.0 | 3.1 | -0.07 | 2.0 | 2.1 | 0.00 | 3.2 | 2.9 | 0.02 |
| Psoriasis | 1.2 | 1.2 | 0.00 | 1.3 | 1.6 | -0.02 | 1.1 | 1.4 | -0.03 |
| Renal impairment | 10.0 | 13.8 | -0.12 | 10.2 | 9.8 | 0.01 | 14.3 | 14.1 | 0.01 |
| Rheumatoid arthritis | 1.5 | 1.4 | 0.01 | 1.5 | 1.4 | 0.00 | 1.6 | 1.4 | 0.01 |
| Schizophrenia | <0.1 | 0.1 | -0.04 | <0.1 | 0.1 | -0.04 | <0.1 | 0.1 | -0.04 |
| Ulcerative colitis | 0.3 | 0.3 | 0.01 | 0.4 | 0.3 | 0.00 | 0.5 | 0.3 | 0.02 |
| Urinary tract infectious disease | 4.3 | 7.0 | -0.12 | 4.4 | 4.4 | 0.00 | 8.0 | 6.4 | 0.06 |
| Viral hepatitis C | 0.3 | 0.3 | -0.01 | 0.3 | 0.3 | -0.01 | 0.2 | 0.4 | -0.03 |
| Visual system disorder | 16.0 | 19.3 | -0.09 | 16.1 | 16.2 | 0.00 | 20.1 | 18.8 | 0.03 |
| **Medical history: Cardiovascular disease** | | | | | | | | | |
| Atrial fibrillation | 8.6 | 9.3 | -0.03 | 8.1 | 8.2 | 0.00 | 9.3 | 9.2 | 0.00 |
| Cerebrovascular disease | 6.0 | 5.7 | 0.01 | 5.6 | 5.3 | 0.01 | 5.5 | 5.6 | 0.00 |
| Coronary arteriosclerosis | 24.1 | 18.7 | 0.13 | 21.9 | 22.3 | -0.01 | 18.5 | 19.1 | -0.01 |
| Heart disease | 37.8 | 33.9 | 0.08 | 35.5 | 35.6 | 0.00 | 34.2 | 34.2 | 0.00 |
| Heart failure | 7.2 | 6.8 | 0.01 | 6.4 | 6.1 | 0.01 | 6.5 | 6.7 | -0.01 |
| Ischemic heart disease | 9.6 | 7.8 | 0.07 | 8.7 | 8.8 | 0.00 | 7.5 | 8.1 | -0.02 |
| Peripheral vascular disease | 9.9 | 7.8 | 0.07 | 9.1 | 9.2 | 0.00 | 9.4 | 8.4 | 0.04 |
| Pulmonary embolism | 0.5 | 0.8 | -0.03 | 0.5 | 0.7 | -0.03 | 1.1 | 0.8 | 0.03 |
| Venous thrombosis | 1.3 | 1.6 | -0.02 | 1.3 | 1.2 | 0.01 | 1.9 | 1.5 | 0.03 |
| **Medical history: Neoplasms** | | | | | | | | | |
| Hematologic neoplasm | 1.0 | 1.2 | -0.02 | 1.0 | 1.0 | 0.00 | 1.6 | 1.2 | 0.04 |
| Malignant lymphoma | 0.7 | 0.7 | 0.00 | 0.7 | 0.6 | 0.01 | 0.8 | 0.7 | 0.01 |
| Malignant neoplasm of anorectum | 0.1 | 0.2 | -0.02 | 0.1 | 0.1 | 0.00 | 0.5 | 0.2 | 0.06 |
| Malignant neoplastic disease | 10.2 | 12.1 | -0.06 | 10.2 | 10.3 | 0.00 | 12.3 | 11.8 | 0.02 |
| Malignant tumor of breast | 1.7 | 1.9 | -0.02 | 1.7 | 1.8 | 0.00 | 1.8 | 1.9 | -0.01 |
| Malignant tumor of colon | 0.4 | 0.7 | -0.03 | 0.4 | 0.4 | 0.00 | 0.5 | 0.6 | -0.01 |
| Malignant tumor of lung | 0.4 | 0.5 | -0.02 | 0.3 | 0.5 | -0.02 | 0.7 | 0.5 | 0.02 |
| Malignant tumor of urinary bladder | 0.5 | 0.7 | -0.02 | 0.5 | 0.6 | -0.01 | 1.0 | 0.7 | 0.03 |
| Primary malignant neoplasm of prostate | 2.7 | 3.0 | -0.02 | 2.7 | 2.6 | 0.00 | 2.7 | 2.9 | -0.01 |

| (Continued) | Before ajustment | | | After matching | | | After stratification | | |
|---|---|---|---|---|---|---|---|---|---|
| Characteristic | T (%) | C (%) | StdDiff | T (%) | C (%) | StdDiff | T (%) | C (%) | StdDiff |
| **Medication use** | | | | | | | | | |
| Agents acting on the renin-angiotensin system | 74.2 | 71.1 | 0.07 | 73.6 | 73.9 | -0.01 | 68.7 | 71.5 | -0.06 |
| Antibacterials for systemic use | 40.7 | 41.6 | -0.02 | 40.8 | 40.9 | 0.00 | 44.7 | 41.6 | 0.06 |
| Antidepressants | 26.2 | 25.6 | 0.01 | 26.2 | 26.4 | -0.01 | 27.4 | 26.0 | 0.03 |
| Antiepileptics | 17.0 | 16.6 | 0.01 | 17.1 | 17.2 | 0.00 | 18.1 | 17.6 | 0.01 |
| Antiinflammatory and antirheumatic products | 62.7 | 60.7 | 0.04 | 60.5 | 61.3 | -0.02 | 60.5 | 61.3 | -0.02 |
| Antineoplastic agents | 5.4 | 5.2 | 0.01 | 5.3 | 5.6 | -0.02 | 5.2 | 5.3 | 0.00 |
| Antipsoriatics | 0.4 | 0.5 | -0.03 | 0.4 | 0.5 | -0.02 | 0.9 | 0.5 | 0.04 |
| Antithrombotic agents | 56.0 | 56.1 | 0.00 | 53.7 | 56.1 | -0.05 | 53.7 | 56.1 | -0.05 |
| Beta blocking agents | 47.6 | 46.7 | 0.02 | 46.4 | 46.5 | 0.00 | 45.1 | 46.5 | -0.03 |
| Calcium channel blockers | 31.0 | 29.8 | 0.02 | 30.6 | 30.8 | 0.00 | 29.3 | 30.3 | -0.02 |
| Diuretics | 46.2 | 48.8 | -0.05 | 45.8 | 45.5 | 0.01 | 47.1 | 47.5 | -0.01 |
| Drugs for acid related disorders | 42.8 | 43.5 | -0.01 | 42.6 | 42.2 | 0.01 | 44.2 | 43.9 | 0.01 |
| Drugs for obstructive airway diseases | 42.4 | 40.6 | 0.04 | 41.8 | 42.4 | -0.01 | 43.7 | 41.7 | 0.04 |
| Immunosuppressants | 3.0 | 2.6 | 0.03 | 2.9 | 3.1 | -0.01 | 2.6 | 2.8 | -0.02 |
| Lipid modifying agents | 83.7 | 77.9 | 0.15 | 82.8 | 83.2 | -0.01 | 78.2 | 78.9 | -0.02 |
| Opioids | 24.6 | 27.8 | -0.07 | 24.6 | 24.7 | 0.00 | 30.2 | 27.6 | 0.06 |
| Psycholeptics | 26.1 | 27.0 | -0.02 | 25.9 | 26.3 | -0.01 | 29.4 | 26.9 | 0.06 |
| Psychostimulants | 1.5 | 1.3 | 0.02 | 1.5 | 1.4 | 0.01 | 1.6 | 1.3 | 0.03 |

### 2.5.5 VA

**Supplementary Table 25:** Baseline patient characteristics for SGLT2I (T) and SU (C) new-users in the VA data source. We report proportion of initiators satisfying selected base-line characteristics and the standardized difference of population proportions (StdDiff) before and after propensity score adjustment. Less extreme StdDiffs through matching and stratification suggest improved balance between patient cohorts through adjustment.

| | Before adjustment | | | After matching | | | After stratification | | |
|---|---|---|---|---|---|---|---|---|---|
| Characteristic | T (%) | C (%) | StdDiff | T (%) | C (%) | StdDiff | T (%) | C (%) | StdDiff |
| **Age group** | | | | | | | | | |
| 65 - 69 | 27.3 | 40.7 | -0.28 | 28.6 | 27.9 | 0.01 | 38.5 | 39.1 | -0.01 |
| 70 - 74 | 45.1 | 28.6 | 0.35 | 45.0 | 44.6 | 0.01 | 43.3 | 37.3 | 0.12 |
| 75 - 79 | 19.5 | 16.5 | 0.08 | 18.2 | 18.3 | 0.00 | 11.4 | 13.4 | -0.06 |
| 80 - 84 | 5.3 | 9.6 | -0.16 | 5.3 | 5.6 | -0.01 | 4.4 | 6.0 | -0.07 |
| 85 - 89 | 2.2 | 3.7 | -0.09 | 2.3 | 2.5 | -0.02 | 2.1 | 3.0 | -0.06 |
| 90 - 94 | 0.5 | 0.8 | -0.03 | 0.5 | 0.7 | -0.03 | 0.2 | 1.0 | -0.09 |
| 95 - 99 | <0.1 | 0.1 | -0.01 | <0.2 | 0.2 | -0.04 | <0.1 | 0.2 | -0.05 |
| Gender: female | 2.5 | 1.6 | 0.06 | 2.6 | 2.5 | 0.01 | 2.0 | 2.1 | -0.01 |
| **Race** | | | | | | | | | |
| Asian | 0.9 | 0.5 | 0.04 | 0.7 | 0.7 | 0.01 | 0.4 | 0.7 | -0.04 |
| Black or African American | 10.5 | 8.4 | 0.07 | 9.6 | 9.6 | 0.00 | 8.1 | 10.0 | -0.07 |
| White | 81.3 | 79.0 | 0.06 | 82.2 | 82.1 | 0.00 | 83.8 | 81.8 | 0.05 |
| Unknown | 5.7 | 10.7 | -0.18 | 6.0 | 6.1 | 0.00 | 6.6 | 5.9 | 0.03 |
| Native Hawaiian or Other Pacific Islander | 0.9 | 0.8 | 0.01 | 0.9 | 0.7 | 0.02 | 0.5 | 0.8 | -0.04 |
| American Indian or Alaska Native | 0.8 | 0.6 | 0.02 | 0.7 | 0.8 | -0.02 | 0.6 | 0.8 | -0.02 |
| **Ethnicity** | | | | | | | | | |
| Hispanic or Latino | 4.6 | 4.5 | 0.00 | 4.8 | 4.8 | 0.00 | 3.6 | 5.3 | -0.08 |
| Not Hispanic or Latino | 92.8 | 88.7 | 0.14 | 92.6 | 92.6 | 0.00 | 94.5 | 92.3 | 0.09 |
| **Medical history: General** | | | | | | | | | |
| Acute respiratory disease | 8.0 | 7.5 | 0.02 | 7.9 | 7.4 | 0.02 | 8.7 | 8.5 | 0.01 |
| Attention deficit hyperactivity disorder | 0.3 | 0.2 | 0.04 | <0.2 | 0.4 | -0.03 | 0.2 | 0.3 | -0.03 |
| Chronic liver disease | 2.4 | 1.7 | 0.05 | 2.4 | 2.3 | 0.00 | 1.6 | 2.5 | -0.06 |
| Chronic obstructive lung disease | 17.0 | 13.8 | 0.09 | 15.7 | 15.9 | 0.00 | 14.8 | 15.1 | -0.01 |
| Crohn’s disease | 0.2 | 0.2 | 0.00 | 0.2 | 0.3 | -0.01 | 0.1 | 0.2 | -0.04 |
| Dementia | 1.8 | 1.7 | 0.00 | 1.7 | 1.9 | -0.01 | 2.0 | 2.2 | -0.01 |
| Depressive disorder | 17.5 | 14.5 | 0.08 | 16.8 | 17.0 | 0.00 | 18.1 | 17.0 | 0.03 |
| Gastroesophageal reflux disease | 20.7 | 19.2 | 0.04 | 20.3 | 20.8 | -0.01 | 19.1 | 20.7 | -0.04 |
| Gastrointestinal hemorrhage | 1.4 | 1.3 | 0.01 | 1.3 | 1.2 | 0.01 | 0.8 | 1.2 | -0.04 |
| Human immunodeficiency virus infection | 0.2 | 0.1 | 0.02 | 0.2 | 0.3 | -0.02 | 0.1 | 0.2 | -0.04 |
| Hyperlipidemia | 74.5 | 74.9 | -0.01 | 73.5 | 73.7 | 0.00 | 73.9 | 73.3 | 0.01 |
| Hypertensive disorder | 80.7 | 81.0 | -0.01 | 77.1 | 78.9 | -0.04 | 77.1 | 78.9 | -0.04 |
| Lesion of liver | 2.0 | 1.0 | 0.09 | 2.0 | 2.1 | -0.01 | 1.6 | 1.7 | -0.01 |
| Obesity | 25.9 | 22.3 | 0.08 | 23.9 | 24.3 | -0.01 | 20.0 | 23.0 | -0.07 |
| Osteoarthritis | 20.8 | 21.5 | -0.02 | 19.7 | 19.8 | 0.00 | 20.4 | 20.4 | 0.00 |
| Pneumonia | 2.1 | 1.6 | 0.04 | 1.9 | 1.9 | 0.00 | 0.9 | 1.9 | -0.08 |
| Psoriasis | 1.8 | 1.5 | 0.02 | 1.7 | 1.5 | 0.01 | 2.8 | 1.6 | 0.08 |
| Renal impairment | 12.6 | 7.9 | 0.15 | 11.2 | 11.6 | -0.01 | 6.8 | 10.9 | -0.14 |
| Rheumatoid arthritis | 0.9 | 0.9 | 0.00 | 1.0 | 0.8 | 0.01 | 0.6 | 0.9 | -0.04 |
| Schizophrenia | 0.5 | 0.6 | -0.02 | 0.5 | 0.5 | 0.00 | 0.7 | 0.6 | 0.01 |
| Ulcerative colitis | 0.4 | 0.4 | 0.01 | 0.4 | 0.4 | 0.00 | 0.2 | 0.4 | -0.05 |
| Urinary tract infectious disease | 1.6 | 2.7 | -0.08 | 1.6 | 1.8 | -0.01 | 4.6 | 2.6 | 0.11 |
| Viral hepatitis C | 1.1 | 0.9 | 0.02 | 1.1 | 1.3 | -0.02 | 0.6 | 1.4 | -0.08 |
| Visual system disorder | 47.4 | 46.4 | 0.02 | 46.8 | 46.2 | 0.01 | 38.7 | 49.7 | -0.22 |
| **Medical history: Cardiovascular disease** | | | | | | | | | |
| Atrial fibrillation | 16.8 | 8.9 | 0.24 | 14.3 | 14.8 | -0.01 | 8.7 | 10.5 | -0.06 |
| Cerebrovascular disease | 5.9 | 6.1 | -0.01 | 5.3 | 5.0 | 0.01 | 3.1 | 4.8 | -0.08 |
| Coronary arteriosclerosis | 41.3 | 21.8 | 0.43 | 34.0 | 35.8 | -0.04 | 22.3 | 23.7 | -0.03 |
| Heart disease | 58.9 | 40.9 | 0.37 | 51.2 | 52.8 | -0.03 | 32.3 | 40.1 | -0.16 |
| Heart failure | 16.2 | 6.6 | 0.31 | 10.4 | 11.1 | -0.02 | 6.6 | 7.1 | -0.02 |
| Ischemic heart disease | 16.5 | 14.9 | 0.04 | 12.3 | 12.5 | -0.01 | 6.8 | 10.6 | -0.13 |
| Peripheral vascular disease | 8.4 | 6.8 | 0.06 | 7.8 | 7.8 | 0.00 | 5.7 | 6.6 | -0.04 |
| Pulmonary embolism | 0.8 | 0.5 | 0.04 | 0.8 | 0.7 | 0.00 | 0.5 | 0.7 | -0.03 |
| Venous thrombosis | 1.1 | 1.1 | -0.01 | 1.1 | 1.4 | -0.03 | 0.7 | 1.4 | -0.07 |
| **Medical history: Neoplasms** | | | | | | | | | |
| Hematologic neoplasm | 1.3 | 1.0 | 0.02 | 1.3 | 1.4 | -0.01 | 0.8 | 1.3 | -0.05 |
| Malignant lymphoma | 1.0 | 0.6 | 0.04 | 1.0 | 1.0 | 0.01 | 0.6 | 0.9 | -0.04 |
| Malignant neoplasm of anorectum | <0.1 | 0.2 | -0.03 | <0.2 | 0.1 | -0.01 | <0.1 | 0.1 | -0.03 |
| Malignant neoplastic disease | 11.8 | 13.9 | -0.06 | 12.1 | 11.7 | 0.01 | 8.0 | 12.8 | -0.16 |

| (Continued) | Before ajustment | | | After matching | | | After stratification | | |
|---|---|---|---|---|---|---|---|---|---|
| Characteristic | T (%) | C (%) | StdDiff | T (%) | C (%) | StdDiff | T (%) | C (%) | StdDiff |
| Malignant tumor of breast | <0.1 | 0.1 | -0.02 | <0.2 | 0.1 | -0.02 | <0.1 | 0.1 | -0.04 |
| Malignant tumor of colon | 0.5 | 1.0 | -0.06 | 0.5 | 0.5 | 0.01 | 0.2 | 0.6 | -0.06 |
| Malignant tumor of lung | 0.6 | 0.3 | 0.04 | 0.6 | 0.5 | 0.01 | 0.4 | 0.5 | -0.02 |
| Malignant tumor of urinary bladder | 0.8 | 1.0 | -0.02 | 0.8 | 0.7 | 0.01 | 0.7 | 0.9 | -0.02 |
| Primary malignant neoplasm of prostate | 4.4 | 6.5 | -0.09 | 4.6 | 4.5 | 0.01 | 2.9 | 5.2 | -0.12 |
| **Medication use** | | | | | | | | | |
| Agents acting on the renin-angiotensin system | 70.6 | 73.7 | -0.07 | 69.0 | 68.5 | 0.01 | 64.4 | 69.0 | -0.10 |
| Antibacterials for systemic use | 25.6 | 27.9 | -0.05 | 24.8 | 24.5 | 0.01 | 26.9 | 28.7 | -0.04 |
| Antidepressants | 30.4 | 26.8 | 0.08 | 30.0 | 30.3 | -0.01 | 39.0 | 31.1 | 0.17 |
| Antiepileptics | 24.4 | 18.0 | 0.16 | 29.1 | 25.1 | 0.09 | 29.1 | 25.1 | 0.09 |
| Antiinflammatory and antirheumatic products | 48.5 | 39.7 | 0.18 | 45.7 | 45.5 | 0.00 | 47.6 | 43.7 | 0.08 |
| Antineoplastic agents | 5.1 | 3.5 | 0.08 | 5.2 | 4.9 | 0.01 | 4.8 | 4.3 | 0.02 |
| Antipsoriatics | 0.9 | 0.7 | 0.02 | 0.9 | 1.1 | -0.02 | 0.7 | 0.9 | -0.01 |
| Antithrombotic agents | 50.2 | 35.8 | 0.30 | 45.4 | 45.8 | -0.01 | 39.8 | 38.5 | 0.03 |
| Beta blocking agents | 56.9 | 47.2 | 0.19 | 51.8 | 52.3 | -0.01 | 43.2 | 45.9 | -0.06 |
| Calcium channel blockers | 31.2 | 30.4 | 0.02 | 27.3 | 29.6 | -0.05 | 27.3 | 29.6 | -0.05 |
| Diuretics | 44.9 | 47.1 | -0.04 | 41.9 | 42.2 | 0.00 | 43.1 | 42.6 | 0.01 |
| Drugs for acid related disorders | 44.0 | 42.4 | 0.03 | 42.8 | 42.9 | 0.00 | 43.5 | 43.2 | 0.01 |
| Drugs for obstructive airway diseases | 41.8 | 34.1 | 0.16 | 40.7 | 40.5 | 0.01 | 40.9 | 39.0 | 0.04 |
| Immunosuppressants | 2.9 | 1.6 | 0.09 | 2.9 | 2.5 | 0.02 | 2.0 | 2.3 | -0.03 |
| Lipid modifying agents | 85.1 | 81.1 | 0.11 | 84.0 | 83.3 | 0.02 | 82.9 | 81.4 | 0.04 |
| Opioids | 13.6 | 16.8 | -0.09 | 12.8 | 12.8 | 0.00 | 20.1 | 16.3 | 0.10 |
| Psycholeptics | 20.3 | 17.3 | 0.08 | 19.2 | 18.9 | 0.01 | 22.4 | 19.4 | 0.07 |
| Psychostimulants | 1.0 | 0.8 | 0.01 | 1.0 | 0.9 | 0.01 | 0.8 | 1.0 | -0.02 |

##### 2.5.6 GermanyDA

**Supplementary Table 26:** Baseline patient characteristics for SGLT2I (T) and SU (C) new-users in the GermanyDA data source. We report proportion of initiators satisfying selected base-line characteristics and the standardized difference of population proportions (StdDiff) before and after propensity score adjustment. Less extreme StdDiffs through matching and stratification suggest improved balance between patient cohorts through adjustment.

| | Before adjustment | | | After matching | | | After stratification | | |
|---|---|---|---|---|---|---|---|---|---|
| Characteristic | T (%) | C (%) | StdDiff | T (%) | C (%) | StdDiff | T (%) | C (%) | StdDiff |
| **Age group** | | | | | | | | | |
| 65 - 69 | 32.1 | 27.7 | 0.10 | 33.3 | 26.8 | 0.14 | 32.3 | 36.7 | -0.09 |
| 70 - 74 | 26.9 | 28.8 | -0.04 | 22.0 | 24.0 | -0.05 | 24.9 | 23.8 | 0.03 |
| 75 - 79 | 20.8 | 24.0 | -0.08 | 22.8 | 23.5 | -0.02 | 22.0 | 21.3 | 0.02 |
| 80 - 84 | 14.3 | 13.9 | 0.01 | 15.6 | 16.9 | -0.04 | 14.8 | 11.8 | 0.09 |
| 85 - 89 | 5.0 | 4.6 | 0.02 | 5.3 | 6.5 | -0.05 | 5.1 | 5.0 | 0.00 |
| 90 - 94 | 0.8 | 0.9 | -0.02 | 1.0 | 2.2 | -0.10 | 0.9 | 1.3 | -0.04 |
| Gender: female | 38.2 | 50.4 | -0.25 | 44.2 | 45.7 | -0.03 | 40.8 | 40.4 | 0.01 |
| **Medical history: General** | | | | | | | | | |
| Acute respiratory disease | 10.3 | 13.5 | -0.10 | 12.7 | 10.3 | 0.07 | 11.0 | 10.8 | 0.00 |
| Attention deficit hyperactivity disorder | <0.1 | 0.5 | -0.09 | <0.1 | 0.4 | -0.07 | <0.1 | 0.4 | -0.07 |
| Chronic liver disease | 0.5 | 0.3 | 0.03 | <0.6 | 0.3 | 0.00 | 0.4 | 0.2 | 0.03 |
| Chronic obstructive lung disease | 6.7 | 7.2 | -0.02 | 6.5 | 6.6 | 0.00 | 6.5 | 5.4 | 0.05 |
| Crohn’s disease | <0.1 | <0.1 | -0.01 | <0.1 | <0.2 | 0.00 | <0.1 | <0.2 | 0.00 |
| Dementia | 2.0 | 3.8 | -0.11 | 2.5 | 3.4 | -0.06 | 2.3 | 2.5 | -0.01 |
| Depressive disorder | 7.0 | 8.9 | -0.07 | 8.9 | 7.1 | 0.06 | 7.3 | 9.9 | -0.09 |
| Gastroesophageal reflux disease | 2.0 | 2.6 | -0.04 | 2.0 | 2.1 | 0.00 | 1.9 | 1.7 | 0.02 |
| Gastrointestinal hemorrhage | 0.7 | 1.1 | -0.04 | 0.6 | 0.8 | -0.03 | 0.6 | 0.5 | 0.02 |
| Hyperlipidemia | 24.8 | 28.4 | -0.08 | 21.0 | 22.0 | -0.02 | 23.9 | 24.0 | 0.00 |
| Hypertensive disorder | 45.0 | 61.2 | -0.33 | 46.0 | 45.7 | 0.01 | 45.3 | 44.9 | 0.01 |
| Lesion of liver | 0.6 | 0.7 | -0.01 | 0.6 | 0.7 | -0.02 | 0.5 | 2.1 | -0.14 |
| Obesity | 7.6 | 6.7 | 0.04 | 8.2 | 6.6 | 0.06 | 7.9 | 10.6 | -0.10 |
| Osteoarthritis | 11.6 | 16.0 | -0.13 | 14.0 | 12.7 | 0.04 | 11.9 | 14.7 | -0.08 |
| Pneumonia | 1.8 | 2.1 | -0.02 | 1.9 | 1.7 | 0.01 | 1.7 | 1.6 | 0.01 |
| Psoriasis | 1.5 | 1.3 | 0.02 | 1.9 | 1.5 | 0.03 | 1.6 | 0.8 | 0.08 |
| Renal impairment | 7.8 | 6.6 | 0.05 | 6.5 | 7.6 | -0.04 | 6.9 | 6.7 | 0.01 |
| Schizophrenia | 0.1 | 0.2 | -0.03 | <0.1 | <0.2 | 0.01 | <0.1 | <0.2 | 0.01 |
| Ulcerative colitis | 0.1 | 0.3 | -0.03 | <0.6 | 0.2 | -0.03 | <0.1 | 0.5 | -0.08 |
| Urinary tract infectious disease | 4.8 | 6.1 | -0.06 | 5.6 | 5.3 | 0.02 | 5.0 | 5.1 | 0.00 |
| Visual system disorder | 8.5 | 11.3 | -0.09 | 9.4 | 9.6 | -0.01 | 8.5 | 7.3 | 0.04 |
| **Medical history: Cardiovascular disease** | | | | | | | | | |
| Atrial fibrillation | 5.1 | 3.2 | 0.10 | 3.0 | 4.3 | -0.07 | 4.4 | 3.3 | 0.06 |
| Cerebrovascular disease | 5.6 | 6.4 | -0.03 | 5.2 | 5.5 | -0.01 | 5.2 | 7.2 | -0.09 |
| Coronary arteriosclerosis | 9.3 | 4.1 | 0.21 | 5.1 | 5.0 | 0.00 | 7.9 | 8.7 | -0.03 |
| Heart disease | 37.3 | 36.9 | 0.01 | 27.6 | 28.4 | -0.02 | 34.3 | 34.0 | 0.01 |
| Heart failure | 11.9 | 12.0 | 0.00 | 8.6 | 9.1 | -0.02 | 10.9 | 8.7 | 0.07 |
| Ischemic heart disease | 17.0 | 18.6 | -0.04 | 10.4 | 11.2 | -0.03 | 15.3 | 14.0 | 0.04 |
| Peripheral vascular disease | 7.8 | 7.0 | 0.03 | 6.7 | 8.2 | -0.06 | 7.3 | 8.4 | -0.04 |
| Pulmonary embolism | 0.7 | 0.5 | 0.03 | <0.6 | 0.4 | -0.06 | 0.6 | 0.3 | 0.04 |
| Venous thrombosis | 1.6 | 1.5 | 0.00 | 1.5 | 1.1 | 0.03 | 1.5 | 2.3 | -0.06 |
| **Medical history: Neoplasms** | | | | | | | | | |
| Hematologic neoplasm | 0.3 | 0.4 | -0.01 | <0.6 | 0.4 | 0.01 | 0.3 | 0.3 | 0.01 |
| Malignant lymphoma | 0.2 | 0.4 | -0.03 | <0.6 | <0.2 | 0.04 | 0.2 | <0.2 | 0.04 |
| Malignant neoplasm of anorectum | 0.1 | 0.3 | -0.03 | <0.1 | <0.2 | 0.01 | <0.1 | <0.2 | 0.01 |
| Malignant neoplastic disease | 5.6 | 7.9 | -0.09 | 5.6 | 6.3 | -0.03 | 5.3 | 5.5 | -0.01 |
| Malignant tumor of breast | 0.7 | 1.2 | -0.05 | <0.6 | 0.9 | -0.05 | 0.7 | 0.6 | 0.01 |
| Malignant tumor of colon | 0.5 | 0.7 | -0.02 | 0.6 | 0.6 | 0.00 | 0.5 | 0.7 | -0.03 |
| Malignant tumor of lung | <0.1 | 0.2 | -0.04 | <0.1 | <0.2 | -0.02 | <0.1 | <0.2 | -0.02 |
| Primary malignant neoplasm of prostate | 1.1 | 1.5 | -0.03 | 0.9 | 1.6 | -0.06 | 1.2 | 1.8 | -0.04 |
| **Medication use** | | | | | | | | | |
| Agents acting on the renin-angiotensin system | 77.8 | 72.4 | 0.12 | 73.8 | 73.0 | 0.02 | 75.9 | 77.8 | -0.04 |
| Antibacterials for systemic use | 20.2 | 26.5 | -0.15 | 25.3 | 22.0 | 0.08 | 21.1 | 20.4 | 0.02 |
| Antidepressants | 10.1 | 10.4 | -0.01 | 11.4 | 9.9 | 0.05 | 10.3 | 13.4 | -0.10 |
| Antiepileptics | 5.4 | 4.2 | 0.06 | 6.5 | 4.5 | 0.09 | 5.4 | 5.9 | -0.02 |
| Antiinflammatory and antirheumatic products | 43.4 | 48.5 | -0.10 | 41.1 | 41.7 | -0.01 | 42.9 | 45.3 | -0.05 |
| Antineoplastic agents | 1.3 | 1.0 | 0.03 | 1.0 | 1.3 | -0.03 | 1.4 | 1.0 | 0.04 |
| Antipsoriatics | 0.6 | 0.3 | 0.04 | 0.8 | 0.2 | 0.08 | 0.5 | <0.2 | 0.07 |
| Antithrombotic agents | 47.4 | 38.9 | 0.17 | 34.2 | 37.3 | -0.06 | 43.4 | 44.2 | -0.02 |
| Beta blocking agents | 59.2 | 53.3 | 0.12 | 52.5 | 53.0 | -0.01 | 56.1 | 57.3 | -0.02 |

| (Continued) | Before ajustment | | | After matching | | | After stratification | | |
|---|---|---|---|---|---|---|---|---|---|
| Characteristic | T (%) | C (%) | StdDiff | T (%) | C (%) | StdDiff | T (%) | C (%) | StdDiff |
| Calcium channel blockers | 41.2 | 36.4 | 0.10 | 39.6 | 39.7 | 0.00 | 40.7 | 42.8 | -0.04 |
| Diuretics | 59.2 | 62.0 | -0.06 | 56.8 | 55.3 | 0.03 | 57.7 | 60.2 | -0.05 |
| Drugs for acid related disorders | 38.1 | 34.4 | 0.08 | 35.8 | 36.2 | -0.01 | 37.7 | 41.7 | -0.08 |
| Drugs for obstructive airway diseases | 18.1 | 16.5 | 0.04 | 16.3 | 15.6 | 0.02 | 17.5 | 17.6 | 0.00 |
| Immunosuppressants | 0.7 | 0.5 | 0.03 | <0.6 | 0.3 | -0.02 | 0.6 | 0.3 | 0.06 |
| Lipid modifying agents | 61.9 | 43.9 | 0.37 | 43.7 | 47.9 | -0.09 | 56.6 | 60.7 | -0.08 |
| Opioids | 10.7 | 13.7 | -0.09 | 12.2 | 11.3 | 0.03 | 11.0 | 11.7 | -0.02 |
| Psycholeptics | 9.5 | 13.1 | -0.11 | 11.1 | 10.5 | 0.02 | 9.9 | 10.6 | -0.02 |
| Psychostimulants | 0.2 | 0.7 | -0.08 | <0.6 | <0.2 | 0.03 | 0.2 | <0.2 | 0.00 |

##### 2.5.7 SIDIAP

**Supplementary Table 27:** Baseline patient characteristics for SGLT2I (T) and SU (C) new-users in the SIDIAP data source. We report proportion of initiators satisfying selected base-line characteristics and the standardized difference of population proportions (StdDiff) before and after propensity score adjustment. Less extreme StdDiffs through matching and stratification suggest improved balance between patient cohorts through adjustment.

| | Before adjustment | | | After matching | | | After stratification | | |
|---|---|---|---|---|---|---|---|---|---|
| Characteristic | T (%) | C (%) | StdDiff | T (%) | C (%) | StdDiff | T (%) | C (%) | StdDiff |
| **Age group** | | | | | | | | | |
| 65 - 69 | 37.2 | 31.6 | 0.12 | 44.1 | 36.8 | 0.15 | 39.4 | 41.8 | -0.05 |
| 70 - 74 | 27.9 | 27.2 | 0.01 | 26.9 | 29.1 | -0.05 | 29.7 | 26.4 | 0.07 |
| 75 - 79 | 20.3 | 20.8 | -0.01 | 15.7 | 17.6 | -0.05 | 17.8 | 15.2 | 0.07 |
| 80 - 84 | 9.5 | 12.7 | -0.10 | 8.4 | 9.6 | -0.04 | 8.6 | 9.5 | -0.03 |
| 85 - 89 | 4.1 | 5.9 | -0.09 | 4.1 | 4.8 | -0.04 | 3.6 | 4.8 | -0.06 |
| 90 - 94 | 0.9 | 1.5 | -0.06 | 0.6 | 1.7 | -0.10 | 0.8 | 2.1 | -0.11 |
| 95 - 99 | 0.1 | 0.2 | -0.01 | <0.5 | 0.5 | -0.05 | 0.1 | 0.2 | -0.02 |
| Gender: female | 37.9 | 49.9 | -0.24 | 46.2 | 45.7 | 0.01 | 44.0 | 50.2 | -0.12 |
| **Medical history: General** | | | | | | | | | |
| Acute respiratory disease | 13.6 | 10.4 | 0.10 | 14.4 | 14.4 | 0.00 | 13.8 | 11.2 | 0.08 |
| Chronic liver disease | 0.2 | 0.4 | -0.03 | <0.5 | 0.3 | -0.04 | 0.2 | <0.2 | 0.01 |
| Chronic obstructive lung disease | 1.9 | 2.0 | 0.00 | 1.1 | 1.6 | -0.04 | 1.5 | 1.0 | 0.04 |
| Dementia | 0.4 | 1.3 | -0.10 | 0.6 | 1.1 | -0.05 | 0.4 | 0.7 | -0.04 |
| Depressive disorder | 1.4 | 3.4 | -0.13 | 2.7 | 2.6 | 0.00 | 2.7 | 2.4 | 0.02 |
| Gastroesophageal reflux disease | 1.2 | 0.7 | 0.04 | 1.1 | 1.4 | -0.03 | 1.9 | 1.9 | 0.00 |
| Gastrointestinal hemorrhage | 1.0 | 1.3 | -0.02 | 2.1 | 1.2 | 0.07 | 1.6 | 0.9 | 0.06 |
| Hyperlipidemia | 2.0 | 3.6 | -0.10 | 2.4 | 2.5 | -0.01 | 1.9 | 2.2 | -0.02 |
| Hypertensive disorder | 3.3 | 6.5 | -0.15 | 3.6 | 3.1 | 0.03 | 3.4 | 2.7 | 0.04 |
| Lesion of liver | 0.3 | 0.4 | -0.03 | <0.5 | 0.7 | -0.06 | 0.2 | 0.5 | -0.05 |
| Obesity | 5.9 | 8.1 | -0.09 | 9.6 | 7.7 | 0.07 | 7.9 | 5.9 | 0.08 |
| Osteoarthritis | 4.0 | 6.9 | -0.13 | 4.8 | 4.7 | 0.00 | 4.3 | 4.1 | 0.01 |
| Pneumonia | 1.9 | 1.7 | 0.02 | 1.5 | 1.9 | -0.03 | 1.5 | 1.5 | 0.00 |
| Psoriasis | 0.3 | 0.6 | -0.04 | <0.5 | 0.8 | -0.06 | 0.3 | 0.4 | 0.00 |
| Renal impairment | 2.5 | 2.9 | -0.03 | 2.8 | 2.5 | 0.02 | 2.3 | 2.2 | 0.01 |
| Rheumatoid arthritis | 0.1 | 0.1 | 0.00 | <0.5 | <0.1 | 0.06 | 0.1 | <0.2 | 0.02 |
| Urinary tract infectious disease | 6.9 | 4.4 | 0.11 | 7.5 | 6.9 | 0.02 | 7.3 | 5.9 | 0.06 |
| Viral hepatitis C | <0.1 | 0.1 | -0.04 | <0.1 | <0.2 | -0.01 | <0.1 | <0.2 | -0.01 |
| Visual system disorder | 14.6 | 18.4 | -0.10 | 19.1 | 18.7 | 0.01 | 17.1 | 17.7 | -0.02 |
| **Medical history: Cardiovascular disease** | | | | | | | | | |
| Atrial fibrillation | 3.6 | 2.1 | 0.09 | 1.8 | 1.7 | 0.00 | 2.7 | 1.4 | 0.09 |
| Cerebrovascular disease | 1.2 | 1.8 | -0.05 | 1.5 | 1.8 | -0.02 | 1.7 | 1.0 | 0.06 |
| Heart disease | 17.0 | 11.2 | 0.17 | 7.8 | 9.4 | -0.06 | 13.4 | 9.4 | 0.13 |
| Heart failure | 4.8 | 2.7 | 0.11 | 1.1 | 1.5 | -0.03 | 3.3 | 1.4 | 0.13 |
| Ischemic heart disease | 6.6 | 3.4 | 0.15 | 1.8 | 2.7 | -0.06 | 5.2 | 2.5 | 0.14 |
| Peripheral vascular disease | 1.2 | 1.2 | 0.00 | 1.0 | 1.0 | 0.00 | 1.0 | 0.7 | 0.03 |
| Pulmonary embolism | 0.2 | 0.2 | -0.01 | <0.5 | 0.5 | -0.01 | 0.2 | 0.4 | -0.04 |
| Venous thrombosis | 1.0 | 1.2 | -0.01 | 2.0 | 1.0 | 0.08 | 2.0 | 0.7 | 0.12 |
| **Medical history: Neoplasms** | | | | | | | | | |
| Hematologic neoplasm | <0.1 | 0.1 | -0.02 | <0.5 | <0.1 | 0.04 | 0.2 | <0.2 | 0.04 |
| Malignant neoplasm of anorectum | <0.1 | 0.1 | -0.03 | 0.1 | <0.2 | 0.03 | 0.1 | <0.2 | 0.03 |
| Malignant neoplastic disease | 2.4 | 3.8 | -0.08 | 2.3 | 3.3 | -0.07 | 2.8 | 3.1 | -0.02 |
| Malignant tumor of breast | <0.1 | 0.5 | -0.07 | <0.5 | 0.3 | -0.02 | 0.2 | 0.2 | -0.01 |
| Malignant tumor of colon | 0.2 | 0.5 | -0.04 | <0.5 | 0.2 | 0.00 | 0.2 | 0.2 | -0.01 |
| Malignant tumor of lung | <0.1 | <0.1 | 0.00 | <0.1 | <0.2 | -0.02 | <0.1 | <0.2 | -0.02 |
| Malignant tumor of urinary bladder | <0.1 | 0.3 | -0.04 | <0.1 | 0.6 | -0.09 | <0.1 | 0.6 | -0.09 |
| Primary malignant neoplasm of prostate | 0.2 | 0.4 | -0.04 | <0.5 | 0.4 | -0.06 | 0.1 | 0.3 | -0.04 |
| **Medication use** | | | | | | | | | |
| Agents acting on the renin-angiotensin system | 76.1 | 63.2 | 0.28 | 65.6 | 66.9 | -0.03 | 71.8 | 71.2 | 0.01 |
| Antibacterials for systemic use | 39.7 | 33.4 | 0.13 | 41.0 | 41.2 | 0.00 | 40.7 | 37.7 | 0.06 |
| Antidepressants | 21.2 | 20.8 | 0.01 | 25.0 | 23.7 | 0.03 | 23.2 | 21.0 | 0.05 |
| Antiepileptics | 10.0 | 7.3 | 0.10 | 10.1 | 11.1 | -0.03 | 10.8 | 10.6 | 0.01 |
| Antiinflammatory and antirheumatic products | 55.4 | 56.9 | -0.03 | 48.8 | 49.0 | 0.00 | 53.3 | 46.1 | 0.14 |
| Antineoplastic agents | 1.8 | 1.8 | 0.00 | 2.2 | 2.5 | -0.02 | 1.8 | 1.6 | 0.01 |
| Antipsoriatics | 2.0 | 1.1 | 0.07 | 1.3 | 1.1 | 0.02 | 2.1 | 0.7 | 0.12 |
| Antithrombotic agents | 57.8 | 41.5 | 0.33 | 38.1 | 41.4 | -0.07 | 48.6 | 49.6 | -0.02 |
| Beta blocking agents | 49.4 | 26.6 | 0.48 | 30.9 | 33.8 | -0.06 | 42.5 | 45.6 | -0.06 |
| Calcium channel blockers | 32.0 | 24.4 | 0.17 | 25.4 | 27.7 | -0.05 | 28.2 | 23.8 | 0.10 |
| Diuretics | 61.2 | 52.4 | 0.18 | 53.9 | 54.5 | -0.01 | 58.6 | 59.2 | -0.01 |

| (Continued) | Before ajustment | | | After matching | | | After stratification | | |
|---|---|---|---|---|---|---|---|---|---|
| Characteristic | T (%) | C (%) | StdDiff | T (%) | C (%) | StdDiff | T (%) | C (%) | StdDiff |
| Drugs for acid related disorders | 63.7 | 59.4 | 0.09 | 57.0 | 58.6 | -0.03 | 61.3 | 63.7 | -0.05 |
| Drugs for obstructive airway diseases | 34.1 | 25.9 | 0.18 | 30.0 | 30.0 | 0.00 | 32.8 | 28.0 | 0.10 |
| Immunosuppressants | 1.4 | 0.8 | 0.05 | 1.5 | 1.6 | 0.00 | 1.2 | 0.9 | 0.03 |
| Lipid modifying agents | 74.3 | 59.3 | 0.32 | 63.4 | 66.1 | -0.06 | 70.5 | 61.7 | 0.19 |
| Opioids | 20.7 | 17.0 | 0.09 | 21.8 | 21.4 | 0.01 | 20.7 | 19.7 | 0.03 |
| Psycholeptics | 30.6 | 33.3 | -0.06 | 34.3 | 33.7 | 0.01 | 33.2 | 30.8 | 0.05 |
| Psychostimulants | 9.5 | 5.0 | 0.17 | 3.2 | 3.5 | -0.02 | 6.8 | 4.9 | 0.08 |

#### 2.6 DPP4I vs SU

##### 2.6.1 CCAE

**Supplementary Table 28:** Baseline patient characteristics for DPP4I (T) and SU (C) new-users in the CCAE data source. We report proportion of initiators satisfying selected base-line characteristics and the standardized difference of population proportions (StdDiff) before and after propensity score adjustment. Less extreme StdDiffs through matching and stratification suggest improved balance between patient cohorts through adjustment.

| | Before adjustment | | | After matching | | | After stratification | | |
|---|---|---|---|---|---|---|---|---|---|
| Characteristic | T (%) | C (%) | StdDiff | T (%) | C (%) | StdDiff | T (%) | C (%) | StdDiff |
| Gender: female | 45.1 | 43.2 | 0.04 | 44.4 | 44.8 | -0.01 | 43.5 | 43.8 | -0.01 |
| **Medical history: General** | | | | | | | | | |
| Acute respiratory disease | 26.1 | 22.1 | 0.09 | 24.7 | 24.9 | 0.00 | 23.3 | 23.8 | -0.01 |
| Attention deficit hyperactivity disorder | 0.3 | 0.2 | 0.01 | 0.3 | 0.3 | 0.00 | 0.3 | 0.3 | 0.00 |
| Chronic liver disease | 3.0 | 2.9 | 0.00 | 3.0 | 2.8 | 0.01 | 2.7 | 2.9 | -0.01 |
| Chronic obstructive lung disease | 6.5 | 6.1 | 0.02 | 6.3 | 5.9 | 0.01 | 6.3 | 5.9 | 0.02 |
| Crohn's disease | 0.4 | 0.3 | 0.02 | 0.3 | 0.3 | 0.00 | 0.3 | 0.3 | 0.00 |
| Dementia | 0.3 | 0.3 | -0.01 | 0.3 | 0.3 | -0.01 | 0.2 | 0.4 | -0.02 |
| Depressive disorder | 8.5 | 7.5 | 0.04 | 8.5 | 8.4 | 0.00 | 8.5 | 8.4 | 0.00 |
| Gastroesophageal reflux disease | 12.8 | 10.1 | 0.09 | 12.1 | 12.5 | -0.01 | 11.7 | 12.0 | -0.01 |
| Gastrointestinal hemorrhage | 2.2 | 1.9 | 0.02 | 2.1 | 1.7 | 0.03 | 1.9 | 1.8 | 0.01 |
| Human immunodeficiency virus infection | <0.2 | 0.2 | -0.01 | <0.3 | 0.3 | -0.03 | <0.2 | 0.2 | -0.03 |
| Hyperlipidemia | 74.0 | 67.5 | 0.14 | 73.0 | 73.9 | -0.02 | 72.0 | 72.2 | 0.00 |
| Hypertensive disorder | 76.1 | 69.4 | 0.15 | 75.1 | 75.7 | -0.01 | 73.4 | 73.7 | -0.01 |
| Lesion of liver | 1.1 | 1.5 | -0.03 | 1.1 | 1.3 | -0.03 | 1.0 | 1.4 | -0.04 |
| Obesity | 14.0 | 11.0 | 0.09 | 13.4 | 13.2 | 0.00 | 12.8 | 13.0 | -0.01 |
| Osteoarthritis | 23.5 | 20.3 | 0.08 | 23.0 | 23.1 | 0.00 | 22.2 | 21.9 | 0.01 |
| Pneumonia | 2.5 | 2.9 | -0.02 | 2.6 | 3.0 | -0.02 | 2.4 | 3.0 | -0.04 |
| Psoriasis | 1.5 | 1.5 | 0.01 | 1.5 | 1.9 | -0.03 | 1.3 | 1.8 | -0.04 |
| Renal impairment | 6.9 | 6.7 | 0.01 | 6.9 | 7.3 | -0.01 | 6.7 | 7.2 | -0.02 |
| Rheumatoid arthritis | 1.6 | 1.0 | 0.05 | 1.6 | 1.3 | 0.03 | 1.5 | 1.1 | 0.03 |
| Schizophrenia | <0.2 | 0.2 | -0.01 | <0.3 | 0.2 | 0.00 | <0.2 | 0.2 | -0.01 |
| Ulcerative colitis | 0.4 | 0.5 | -0.02 | 0.4 | 0.5 | -0.02 | 0.4 | 0.5 | -0.02 |
| Urinary tract infectious disease | 8.1 | 8.0 | 0.00 | 8.1 | 8.3 | -0.01 | 7.8 | 8.1 | -0.01 |
| Viral hepatitis C | 0.4 | 0.4 | 0.01 | 0.4 | 0.3 | 0.03 | 0.4 | 0.3 | 0.01 |
| Visual system disorder | 36.1 | 33.6 | 0.05 | 35.5 | 35.8 | -0.01 | 33.9 | 35.2 | -0.03 |
| **Medical history: Cardiovascular disease** | | | | | | | | | |
| Atrial fibrillation | 4.3 | 4.7 | -0.02 | 4.2 | 4.4 | -0.01 | 4.2 | 4.5 | -0.01 |
| Cerebrovascular disease | 4.8 | 5.0 | -0.01 | 4.5 | 5.0 | -0.02 | 5.0 | 4.9 | 0.00 |
| Coronary arteriosclerosis | 15.1 | 14.6 | 0.01 | 15.2 | 14.9 | 0.01 | 14.6 | 14.8 | -0.01 |
| Heart disease | 28.4 | 27.0 | 0.03 | 27.6 | 27.3 | 0.01 | 26.7 | 27.0 | -0.01 |
| Heart failure | 3.7 | 3.4 | 0.02 | 3.9 | 3.0 | 0.05 | 3.7 | 3.2 | 0.03 |
| Ischemic heart disease | 6.3 | 6.6 | -0.01 | 6.3 | 6.3 | 0.00 | 6.0 | 6.3 | -0.01 |
| Peripheral vascular disease | 5.6 | 4.5 | 0.05 | 5.4 | 4.4 | 0.05 | 4.9 | 4.3 | 0.03 |
| Pulmonary embolism | 0.5 | 0.5 | 0.01 | 0.5 | 0.5 | 0.00 | 0.5 | 0.5 | -0.01 |
| Venous thrombosis | 1.6 | 1.1 | 0.04 | 1.6 | 1.2 | 0.04 | 1.5 | 1.2 | 0.03 |
| **Medical history: Neoplasms** | | | | | | | | | |
| Hematologic neoplasm | 2.0 | 1.2 | 0.06 | 1.8 | 1.2 | 0.05 | 2.0 | 1.2 | 0.06 |
| Malignant lymphoma | 1.0 | 0.5 | 0.05 | 1.1 | 0.5 | 0.06 | 1.0 | 0.5 | 0.06 |
| Malignant neoplasm of anorectum | <0.2 | 0.3 | -0.03 | <0.3 | 0.1 | 0.01 | <0.2 | 0.2 | -0.01 |
| Malignant neoplastic disease | 12.9 | 12.4 | 0.02 | 12.7 | 13.1 | -0.01 | 12.0 | 12.6 | -0.02 |
| Malignant tumor of breast | 2.5 | 2.2 | 0.02 | 2.5 | 2.6 | -0.01 | 2.1 | 2.4 | -0.03 |
| Malignant tumor of colon | 0.4 | 0.7 | -0.05 | 0.4 | 0.6 | -0.04 | 0.3 | 0.6 | -0.04 |
| Malignant tumor of lung | 0.5 | 0.4 | 0.02 | 0.5 | 0.3 | 0.04 | 0.4 | 0.3 | 0.02 |
| Malignant tumor of urinary bladder | 0.6 | 0.6 | 0.00 | 0.4 | 0.5 | -0.01 | 0.5 | 0.5 | 0.00 |
| Primary malignant neoplasm of prostate | 2.2 | 2.8 | -0.03 | 2.3 | 3.3 | -0.06 | 2.3 | 3.0 | -0.05 |
| **Medication use** | | | | | | | | | |
| Agents acting on the renin-angiotensin system | 75.4 | 72.5 | 0.07 | 75.3 | 73.6 | 0.04 | 74.2 | 73.6 | 0.01 |
| Antibacterials for systemic use | 60.5 | 55.8 | 0.10 | 59.0 | 59.7 | -0.01 | 57.0 | 57.7 | -0.01 |
| Antidepressants | 23.8 | 23.5 | 0.01 | 23.9 | 24.7 | -0.02 | 22.9 | 24.2 | -0.03 |
| Antiepileptics | 11.7 | 11.3 | 0.01 | 11.7 | 12.2 | -0.01 | 11.4 | 12.0 | -0.02 |
| Antiinflammatory and antirheumatic products | 32.2 | 27.8 | 0.10 | 31.4 | 31.0 | 0.01 | 29.2 | 29.7 | -0.01 |
| Antineoplastic agents | 7.7 | 5.6 | 0.08 | 6.9 | 6.4 | 0.02 | 6.1 | 5.8 | 0.01 |
| Antipsoriatics | 0.5 | 0.5 | -0.01 | 0.4 | 0.6 | -0.03 | 0.4 | 0.6 | -0.03 |
| Antithrombotic agents | 15.6 | 15.7 | 0.00 | 15.1 | 15.7 | -0.02 | 15.1 | 15.7 | -0.02 |
| Beta blocking agents | 36.4 | 37.3 | -0.02 | 35.9 | 36.6 | -0.01 | 36.6 | 37.4 | -0.02 |
| Calcium channel blockers | 28.6 | 27.0 | 0.03 | 27.0 | 27.9 | -0.02 | 27.0 | 27.9 | -0.02 |

| (Continued) | Before ajustment | | | After matching | | | After stratification | | |
|---|---|---|---|---|---|---|---|---|---|
| Characteristic | T (%) | C (%) | StdDiff | T (%) | C (%) | StdDiff | T (%) | C (%) | StdDiff |
| Diuretics | 46.9 | 46.4 | 0.01 | 47.1 | 46.6 | 0.01 | 46.2 | 46.8 | -0.01 |
| Drugs for acid related disorders | 29.7 | 26.1 | 0.08 | 28.8 | 28.0 | 0.02 | 27.1 | 27.4 | -0.01 |
| Drugs for obstructive airway diseases | 39.3 | 34.9 | 0.09 | 38.1 | 38.4 | -0.01 | 36.6 | 37.1 | -0.01 |
| Immunosuppressants | 3.7 | 2.3 | 0.08 | 3.4 | 3.1 | 0.02 | 3.0 | 2.9 | 0.00 |
| Lipid modifying agents | 78.4 | 74.7 | 0.09 | 77.7 | 77.6 | 0.00 | 76.5 | 77.1 | -0.01 |
| Opioids | 26.9 | 24.7 | 0.05 | 27.1 | 25.7 | 0.03 | 26.3 | 25.2 | 0.02 |
| Psycholeptics | 23.0 | 23.6 | -0.01 | 23.4 | 24.2 | -0.02 | 23.6 | 24.3 | -0.02 |
| Psychostimulants | 1.8 | 1.7 | 0.00 | 2.0 | 1.7 | 0.02 | 1.8 | 1.6 | 0.01 |

##### 2.6.2 OptumDOD

**Supplementary Table 29:** Baseline patient characteristics for DPP4I (T) and SU (C) new-users in the OptumDOD data source. We report proportion of initiators satisfying selected base-line characteristics and the standardized difference of population proportions (StdDiff) before and after propensity score adjustment. Less extreme StdDiffs through matching and stratification suggest improved balance between patient cohorts through adjustment.

| | Before adjustment | | | After matching | | | After stratification | | |
|---|---|---|---|---|---|---|---|---|---|
| Characteristic | T (%) | C (%) | StdDiff | T (%) | C (%) | StdDiff | T (%) | C (%) | StdDiff |
| **Age group** | | | | | | | | | |
| 65 - 69 | 32.6 | 32.8 | 0.00 | 32.6 | 32.9 | 0.00 | 32.9 | 32.8 | 0.00 |
| 70 - 74 | 30.6 | 31.2 | -0.01 | 30.7 | 30.6 | 0.00 | 30.9 | 31.0 | 0.00 |
| 75 - 79 | 19.4 | 20.0 | -0.01 | 19.4 | 19.5 | 0.00 | 19.4 | 19.5 | 0.00 |
| 80 - 84 | 11.3 | 10.9 | 0.01 | 11.2 | 11.1 | 0.00 | 11.2 | 11.1 | 0.00 |
| 85 - 89 | 5.6 | 4.8 | 0.04 | 5.6 | 5.6 | 0.00 | 5.0 | 5.0 | 0.00 |
| 90 - 94 | 0.5 | 0.3 | 0.03 | 0.5 | 0.3 | 0.02 | 0.4 | 0.3 | 0.02 |
| Gender: female | 53.4 | 49.6 | 0.07 | 52.9 | 53.6 | -0.01 | 50.4 | 50.9 | -0.01 |
| **Medical history: General** | | | | | | | | | |
| Acute respiratory disease | 24.7 | 23.2 | 0.04 | 24.4 | 24.6 | -0.01 | 23.5 | 23.7 | 0.00 |
| Attention deficit hyperactivity disorder | 0.2 | 0.2 | 0.02 | 0.2 | 0.2 | 0.01 | 0.2 | 0.2 | 0.01 |
| Chronic liver disease | 2.8 | 2.5 | 0.02 | 2.7 | 2.6 | 0.00 | 2.6 | 2.5 | 0.00 |
| Chronic obstructive lung disease | 13.0 | 13.7 | -0.02 | 13.0 | 13.1 | 0.00 | 13.4 | 13.6 | 0.00 |
| Crohn’s disease | 0.3 | 0.2 | 0.01 | 0.3 | 0.3 | 0.01 | 0.3 | 0.2 | 0.01 |
| Dementia | 4.6 | 4.1 | 0.02 | 4.3 | 4.3 | 0.00 | 4.3 | 4.3 | 0.00 |
| Depressive disorder | 14.6 | 13.8 | 0.02 | 14.5 | 14.7 | -0.01 | 13.7 | 14.1 | -0.01 |
| Gastroesophageal reflux disease | 21.3 | 19.3 | 0.05 | 21.0 | 21.5 | -0.01 | 19.8 | 20.0 | 0.00 |
| Gastrointestinal hemorrhage | 3.3 | 3.3 | 0.00 | 3.2 | 3.4 | -0.01 | 3.1 | 3.3 | -0.01 |
| Human immunodeficiency virus infection | 0.2 | 0.1 | 0.01 | 0.2 | 0.1 | 0.00 | 0.1 | 0.1 | 0.00 |
| Hyperlipidemia | 85.4 | 82.4 | 0.08 | 85.1 | 85.7 | -0.02 | 82.8 | 83.4 | -0.02 |
| Hypertensive disorder | 87.2 | 85.2 | 0.06 | 87.0 | 87.3 | -0.01 | 85.7 | 85.9 | -0.01 |
| Lesion of liver | 1.4 | 1.5 | -0.01 | 1.4 | 1.5 | -0.01 | 1.4 | 1.5 | -0.01 |
| Obesity | 18.3 | 17.8 | 0.01 | 18.3 | 18.6 | -0.01 | 17.9 | 18.0 | 0.00 |
| Osteoarthritis | 35.2 | 32.1 | 0.06 | 34.6 | 35.2 | -0.01 | 32.6 | 33.1 | -0.01 |
| Pneumonia | 5.5 | 5.6 | -0.01 | 5.4 | 5.4 | 0.00 | 5.5 | 5.6 | 0.00 |
| Psoriasis | 1.5 | 1.5 | 0.00 | 1.5 | 1.6 | -0.01 | 1.4 | 1.5 | -0.01 |
| Renal impairment | 24.9 | 24.3 | 0.01 | 24.8 | 25.0 | 0.00 | 24.5 | 24.5 | 0.00 |
| Rheumatoid arthritis | 2.1 | 2.1 | 0.00 | 2.1 | 2.2 | -0.01 | 2.1 | 2.1 | 0.00 |
| Schizophrenia | 0.3 | 0.2 | 0.01 | 0.3 | 0.2 | 0.02 | 0.3 | 0.2 | 0.02 |
| Ulcerative colitis | 0.5 | 0.4 | 0.01 | 0.4 | 0.4 | 0.01 | 0.4 | 0.4 | 0.01 |
| Urinary tract infectious disease | 15.2 | 13.6 | 0.04 | 14.9 | 15.2 | -0.01 | 14.0 | 14.1 | 0.00 |
| Viral hepatitis C | 0.5 | 0.5 | 0.00 | 0.5 | 0.6 | -0.01 | 0.5 | 0.5 | 0.00 |
| Visual system disorder | 54.3 | 51.9 | 0.05 | 53.9 | 54.6 | -0.01 | 52.0 | 52.8 | -0.02 |
| **Medical history: Cardiovascular disease** | | | | | | | | | |
| Atrial fibrillation | 11.4 | 11.0 | 0.01 | 11.3 | 11.4 | 0.00 | 11.1 | 11.1 | 0.00 |
| Cerebrovascular disease | 10.8 | 10.3 | 0.02 | 10.7 | 10.9 | -0.01 | 10.3 | 10.5 | -0.01 |
| Coronary arteriosclerosis | 23.3 | 23.0 | 0.01 | 23.0 | 23.3 | -0.01 | 23.0 | 23.1 | 0.00 |
| Heart disease | 43.2 | 42.1 | 0.02 | 42.8 | 43.3 | -0.01 | 42.2 | 42.5 | -0.01 |
| Heart failure | 11.4 | 11.5 | 0.00 | 11.5 | 11.5 | 0.00 | 11.5 | 11.5 | 0.00 |
| Ischemic heart disease | 12.2 | 12.3 | 0.00 | 12.1 | 12.4 | -0.01 | 12.2 | 12.4 | 0.00 |
| Peripheral vascular disease | 17.5 | 16.1 | 0.04 | 17.3 | 17.6 | -0.01 | 16.5 | 16.6 | 0.00 |
| Pulmonary embolism | 1.0 | 1.1 | 0.00 | 1.0 | 1.1 | 0.00 | 1.0 | 1.1 | 0.00 |
| Venous thrombosis | 2.2 | 2.3 | 0.00 | 2.2 | 2.3 | -0.01 | 2.2 | 2.3 | -0.01 |
| **Medical history: Neoplasms** | | | | | | | | | |
| Hematologic neoplasm | 1.7 | 1.8 | -0.01 | 1.7 | 1.7 | 0.00 | 1.7 | 1.8 | 0.00 |
| Malignant lymphoma | 0.8 | 0.8 | 0.00 | 0.8 | 0.8 | 0.00 | 0.8 | 0.8 | 0.00 |
| Malignant neoplasm of anorectum | 0.3 | 0.3 | -0.01 | 0.3 | 0.3 | -0.01 | 0.3 | 0.3 | -0.01 |
| Malignant neoplastic disease | 16.1 | 15.8 | 0.01 | 16.1 | 16.3 | -0.01 | 15.7 | 16.0 | -0.01 |
| Malignant tumor of breast | 2.8 | 2.5 | 0.02 | 2.7 | 2.8 | 0.00 | 2.7 | 2.8 | 0.00 |
| Malignant tumor of colon | 0.9 | 0.9 | 0.00 | 0.8 | 0.9 | 0.00 | 0.9 | 0.9 | 0.00 |
| Malignant tumor of lung | 0.6 | 0.8 | -0.02 | 0.6 | 0.7 | -0.01 | 0.7 | 0.8 | -0.01 |
| Malignant tumor of urinary bladder | 0.8 | 0.9 | -0.01 | 0.8 | 0.9 | 0.00 | 0.9 | 0.9 | 0.00 |
| Primary malignant neoplasm of prostate | 3.9 | 3.7 | 0.01 | 3.9 | 3.9 | 0.00 | 3.7 | 3.8 | 0.00 |
| **Medication use** | | | | | | | | | |
| Agents acting on the renin-angiotensin system | 77.0 | 75.6 | 0.03 | 76.8 | 77.1 | -0.01 | 76.0 | 76.1 | 0.00 |
| Antibacterials for systemic use | 59.7 | 57.4 | 0.05 | 59.3 | 59.7 | -0.01 | 57.6 | 58.2 | -0.01 |
| Antidepressants | 27.5 | 25.8 | 0.04 | 27.2 | 27.6 | -0.01 | 26.2 | 26.4 | 0.00 |
| Antiepileptics | 18.8 | 17.5 | 0.03 | 18.6 | 19.1 | -0.01 | 17.9 | 18.0 | 0.00 |
| Antiinflammatory and antirheumatic products | 29.3 | 26.6 | 0.06 | 28.8 | 29.3 | -0.01 | 26.9 | 27.5 | -0.01 |

| (Continued) | Before ajustment | | | After matching | | | After stratification | | |
|---|---|---|---|---|---|---|---|---|---|
| Characteristic | T (%) | C (%) | StdDiff | T (%) | C (%) | StdDiff | T (%) | C (%) | StdDiff |
| Antineoplastic agents | 7.3 | 6.3 | 0.04 | 7.1 | 7.1 | 0.00 | 7.1 | 7.1 | 0.00 |
| Antipsoriatics | 0.6 | 0.5 | 0.01 | 0.6 | 0.7 | -0.01 | 0.5 | 0.6 | 0.00 |
| Antithrombotic agents | 22.5 | 21.5 | 0.02 | 22.3 | 22.6 | -0.01 | 21.8 | 21.9 | 0.00 |
| Beta blocking agents | 45.5 | 45.5 | 0.00 | 45.4 | 45.5 | 0.00 | 45.5 | 45.6 | 0.00 |
| Calcium channel blockers | 35.4 | 34.0 | 0.03 | 35.2 | 35.8 | -0.01 | 34.1 | 34.5 | -0.01 |
| Diuretics | 49.3 | 49.8 | -0.01 | 49.2 | 49.2 | 0.00 | 49.8 | 49.7 | 0.00 |
| Drugs for acid related disorders | 34.4 | 31.8 | 0.06 | 33.8 | 34.4 | -0.01 | 32.4 | 32.7 | -0.01 |
| Drugs for obstructive airway diseases | 41.3 | 37.7 | 0.07 | 40.7 | 41.4 | -0.01 | 38.3 | 38.9 | -0.01 |
| Immunosuppressants | 3.1 | 2.5 | 0.04 | 2.7 | 2.6 | 0.00 | 2.7 | 2.6 | 0.00 |
| Lipid modifying agents | 81.8 | 78.7 | 0.08 | 81.4 | 82.0 | -0.01 | 79.2 | 79.7 | -0.01 |
| Opioids | 27.1 | 26.3 | 0.02 | 26.5 | 26.5 | 0.00 | 26.5 | 26.5 | 0.00 |
| Psycholeptics | 24.2 | 22.8 | 0.03 | 23.9 | 24.3 | -0.01 | 23.2 | 23.3 | 0.00 |
| Psychostimulants | 1.0 | 0.7 | 0.03 | 0.9 | 0.9 | 0.01 | 0.8 | 0.7 | 0.01 |

##### 2.6.3 MDCR

**Supplementary Table 30:** Baseline patient characteristics for DPP4I (T) and SU (C) new-users in the MDCR data source. We report proportion of initiators satisfying selected base-line characteristics and the standardized difference of population proportions (StdDiff) before and after propensity score adjustment. Less extreme StdDiffs through matching and stratification suggest improved balance between patient cohorts through adjustment.

| | Before adjustment | | | After matching | | | After stratification | | |
|---|---|---|---|---|---|---|---|---|---|
| Characteristic | T (%) | C (%) | StdDiff | T (%) | C (%) | StdDiff | T (%) | C (%) | StdDiff |
| **Age group** | | | | | | | | | |
| 65 - 69 | 31.2 | 28.1 | 0.07 | 31.0 | 31.0 | 0.00 | 29.8 | 30.2 | -0.01 |
| 70 - 74 | 30.5 | 30.2 | 0.01 | 30.3 | 30.5 | 0.00 | 29.9 | 30.2 | -0.01 |
| 75 - 79 | 19.3 | 21.3 | -0.05 | 20.0 | 19.9 | 0.00 | 20.0 | 19.9 | 0.00 |
| 80 - 84 | 11.6 | 12.9 | -0.04 | 11.7 | 11.9 | 0.00 | 12.5 | 12.3 | 0.01 |
| 85 - 89 | 5.4 | 5.6 | -0.01 | 5.5 | 5.6 | 0.00 | 5.7 | 5.6 | 0.00 |
| 90 - 94 | 1.6 | 1.6 | 0.00 | 1.7 | 1.4 | 0.02 | 1.8 | 1.5 | 0.02 |
| 95 - 99 | 0.3 | 0.2 | 0.01 | 0.3 | 0.2 | 0.02 | 0.3 | 0.2 | 0.02 |
| Gender: female | 48.1 | 46.8 | 0.03 | 47.5 | 47.9 | -0.01 | 46.6 | 46.8 | 0.00 |
| **Medical history: General** | | | | | | | | | |
| Acute respiratory disease | 23.1 | 21.0 | 0.05 | 22.5 | 22.6 | 0.00 | 22.8 | 22.2 | 0.01 |
| Attention deficit hyperactivity disorder | 0.2 | 0.1 | 0.02 | 0.2 | 0.2 | 0.00 | 0.2 | 0.2 | 0.00 |
| Chronic liver disease | 2.3 | 2.1 | 0.02 | 2.3 | 2.3 | 0.00 | 2.4 | 2.3 | 0.00 |
| Chronic obstructive lung disease | 10.5 | 10.9 | -0.02 | 10.5 | 10.5 | 0.00 | 11.7 | 11.1 | 0.02 |
| Crohn’s disease | 0.3 | 0.3 | 0.00 | 0.3 | 0.3 | -0.01 | 0.3 | 0.3 | 0.00 |
| Dementia | 2.9 | 3.0 | -0.01 | 2.9 | 3.1 | -0.01 | 3.1 | 3.2 | 0.00 |
| Depressive disorder | 7.7 | 6.7 | 0.04 | 7.6 | 7.9 | -0.01 | 7.6 | 7.9 | -0.01 |
| Gastroesophageal reflux disease | 13.0 | 10.1 | 0.09 | 12.4 | 12.9 | -0.01 | 11.8 | 12.3 | -0.01 |
| Gastrointestinal hemorrhage | 3.2 | 3.5 | -0.02 | 3.2 | 3.3 | -0.01 | 3.3 | 3.4 | 0.00 |
| Hyperlipidemia | 63.3 | 49.4 | 0.28 | 61.9 | 63.2 | -0.03 | 57.9 | 59.4 | -0.03 |
| Hypertensive disorder | 75.0 | 66.4 | 0.19 | 74.2 | 74.8 | -0.01 | 72.4 | 72.7 | -0.01 |
| Lesion of liver | 0.8 | 1.1 | -0.03 | 0.8 | 0.9 | -0.01 | 1.0 | 1.1 | -0.01 |
| Obesity | 10.0 | 7.7 | 0.08 | 10.0 | 10.2 | -0.01 | 9.4 | 9.8 | -0.01 |
| Osteoarthritis | 30.4 | 26.3 | 0.09 | 29.7 | 30.3 | -0.01 | 29.0 | 29.1 | 0.00 |
| Pneumonia | 4.9 | 5.9 | -0.04 | 5.0 | 4.8 | 0.01 | 5.9 | 5.5 | 0.02 |
| Psoriasis | 1.7 | 1.3 | 0.03 | 1.6 | 1.7 | 0.00 | 1.6 | 1.5 | 0.01 |
| Renal impairment | 14.0 | 12.4 | 0.05 | 14.1 | 14.0 | 0.00 | 14.9 | 14.3 | 0.02 |
| Rheumatoid arthritis | 1.8 | 1.5 | 0.02 | 1.8 | 1.7 | 0.01 | 1.8 | 1.7 | 0.01 |
| Schizophrenia | 0.1 | 0.1 | 0.00 | 0.1 | 0.1 | 0.00 | 0.1 | 0.1 | 0.00 |
| Ulcerative colitis | 0.5 | 0.4 | 0.02 | 0.5 | 0.4 | 0.03 | 0.5 | 0.4 | 0.02 |
| Urinary tract infectious disease | 10.7 | 10.4 | 0.01 | 10.5 | 10.6 | 0.00 | 10.8 | 10.7 | 0.00 |
| Viral hepatitis C | 0.3 | 0.2 | 0.00 | 0.3 | 0.2 | 0.01 | 0.3 | 0.3 | 0.00 |
| Visual system disorder | 52.4 | 48.1 | 0.09 | 51.4 | 52.2 | -0.02 | 49.7 | 50.4 | -0.01 |
| **Medical history: Cardiovascular disease** | | | | | | | | | |
| Atrial fibrillation | 12.1 | 11.4 | 0.02 | 11.9 | 11.9 | 0.00 | 12.5 | 11.9 | 0.02 |
| Cerebrovascular disease | 12.7 | 12.0 | 0.02 | 12.3 | 12.4 | 0.00 | 12.6 | 12.1 | 0.01 |
| Coronary arteriosclerosis | 25.2 | 24.1 | 0.03 | 24.7 | 25.3 | -0.01 | 25.3 | 24.8 | 0.01 |
| Heart disease | 45.5 | 43.8 | 0.03 | 44.5 | 45.3 | -0.01 | 45.5 | 44.5 | 0.02 |
| Heart failure | 9.4 | 10.6 | -0.04 | 9.5 | 9.3 | 0.00 | 10.9 | 10.0 | 0.03 |
| Ischemic heart disease | 11.2 | 11.8 | -0.02 | 10.9 | 11.3 | -0.01 | 11.6 | 11.4 | 0.01 |
| Peripheral vascular disease | 12.8 | 10.8 | 0.06 | 12.4 | 12.6 | -0.01 | 12.1 | 12.0 | 0.00 |
| Pulmonary embolism | 0.9 | 0.9 | 0.00 | 0.9 | 0.9 | 0.00 | 1.0 | 1.0 | 0.01 |
| Venous thrombosis | 2.3 | 2.2 | 0.00 | 2.3 | 2.3 | 0.00 | 2.4 | 2.4 | 0.00 |
| **Medical history: Neoplasms** | | | | | | | | | |
| Hematologic neoplasm | 1.8 | 1.8 | 0.00 | 1.8 | 1.8 | 0.00 | 1.9 | 1.9 | 0.00 |
| Malignant lymphoma | 1.0 | 0.8 | 0.02 | 1.0 | 0.9 | 0.01 | 1.0 | 0.9 | 0.01 |
| Malignant neoplasm of anorectum | 0.4 | 0.4 | -0.01 | 0.4 | 0.3 | 0.01 | 0.4 | 0.3 | 0.01 |
| Malignant neoplastic disease | 19.2 | 18.1 | 0.03 | 19.1 | 19.0 | 0.00 | 19.1 | 18.8 | 0.01 |
| Malignant tumor of breast | 2.9 | 2.7 | 0.01 | 2.9 | 2.9 | 0.00 | 2.8 | 2.8 | 0.00 |
| Malignant tumor of colon | 0.9 | 0.9 | 0.00 | 1.0 | 0.8 | 0.02 | 1.0 | 0.8 | 0.01 |
| Malignant tumor of lung | 0.6 | 0.8 | -0.02 | 0.6 | 0.7 | -0.01 | 0.7 | 0.8 | -0.01 |
| Malignant tumor of urinary bladder | 1.2 | 1.1 | 0.01 | 1.2 | 1.2 | 0.00 | 1.2 | 1.2 | 0.00 |
| Primary malignant neoplasm of prostate | 4.5 | 4.5 | 0.00 | 4.5 | 4.6 | 0.00 | 4.5 | 4.6 | 0.00 |
| **Medication use** | | | | | | | | | |
| Agents acting on the renin-angiotensin system | 75.8 | 72.3 | 0.08 | 75.6 | 75.4 | 0.00 | 75.2 | 75.2 | 0.00 |
| Antibacterials for systemic use | 62.5 | 60.6 | 0.04 | 61.7 | 62.2 | -0.01 | 61.9 | 61.7 | 0.00 |
| Antidepressants | 24.9 | 24.2 | 0.02 | 24.7 | 24.7 | 0.00 | 25.1 | 24.9 | 0.00 |
| Antiepileptics | 14.6 | 13.8 | 0.02 | 14.5 | 14.8 | -0.01 | 15.3 | 14.9 | 0.01 |
| Antiinflammatory and antirheumatic products | 28.1 | 26.8 | 0.03 | 27.6 | 27.7 | 0.00 | 27.0 | 27.3 | -0.01 |

| (Continued) | Before ajustment | | | After matching | | | After stratification | | |
|---|---|---|---|---|---|---|---|---|---|
| Characteristic | T (%) | C (%) | StdDiff | T (%) | C (%) | StdDiff | T (%) | C (%) | StdDiff |
| Antineoplastic agents | 8.7 | 8.8 | 0.00 | 8.6 | 8.5 | 0.00 | 8.6 | 8.4 | 0.01 |
| Antipsoriatics | 0.8 | 0.7 | 0.01 | 0.7 | 0.9 | -0.02 | 0.7 | 0.8 | -0.01 |
| Antithrombotic agents | 25.4 | 25.2 | 0.00 | 25.3 | 25.3 | 0.00 | 26.5 | 25.7 | 0.02 |
| Beta blocking agents | 49.7 | 48.6 | 0.02 | 49.5 | 49.5 | 0.00 | 50.6 | 50.1 | 0.01 |
| Calcium channel blockers | 35.0 | 33.4 | 0.03 | 34.7 | 34.7 | 0.00 | 34.7 | 34.7 | 0.00 |
| Diuretics | 51.4 | 53.2 | -0.03 | 51.6 | 51.1 | 0.01 | 53.2 | 52.5 | 0.01 |
| Drugs for acid related disorders | 33.3 | 31.7 | 0.03 | 32.7 | 32.5 | 0.00 | 32.7 | 32.5 | 0.00 |
| Drugs for obstructive airway diseases | 41.5 | 37.5 | 0.08 | 40.7 | 41.1 | -0.01 | 40.4 | 40.2 | 0.00 |
| Immunosuppressants | 3.4 | 2.7 | 0.04 | 3.3 | 3.1 | 0.01 | 3.3 | 3.1 | 0.01 |
| Lipid modifying agents | 81.2 | 75.4 | 0.14 | 80.5 | 80.7 | 0.00 | 79.2 | 79.8 | -0.02 |
| Opioids | 27.5 | 27.9 | -0.01 | 28.8 | 28.2 | 0.01 | 28.8 | 28.2 | 0.01 |
| Psycholeptics | 26.0 | 24.9 | 0.03 | 25.9 | 25.5 | 0.01 | 25.9 | 25.5 | 0.01 |
| Psychostimulants | 1.3 | 1.1 | 0.02 | 1.3 | 1.2 | 0.00 | 1.3 | 1.2 | 0.01 |

##### 2.6.4 MDCD

**Supplementary Table 31:** Baseline patient characteristics for DPP4I (T) and SU (C) new-users in the MDCD data source. We report proportion of initiators satisfying selected base-line characteristics and the standardized difference of population proportions (StdDiff) before and after propensity score adjustment. Less extreme StdDiffs through matching and stratification suggest improved balance between patient cohorts through adjustment.

| | Before adjustment | | | After matching | | | After stratification | | |
|---|---|---|---|---|---|---|---|---|---|
| Characteristic | T (%) | C (%) | StdDiff | T (%) | C (%) | StdDiff | T (%) | C (%) | StdDiff |
| **Age group** | | | | | | | | | |
| 65 - 69 | 43.1 | 47.4 | -0.09 | 43.4 | 46.9 | -0.07 | 42.4 | 47.3 | -0.10 |
| 70 - 74 | 23.8 | 23.6 | 0.00 | 23.4 | 23.7 | -0.01 | 22.9 | 23.7 | -0.02 |
| 75 - 79 | 17.0 | 14.4 | 0.07 | 17.0 | 15.2 | 0.05 | 17.1 | 14.6 | 0.07 |
| 80 - 84 | 10.7 | 9.2 | 0.05 | 11.0 | 9.3 | 0.06 | 12.4 | 9.2 | 0.10 |
| 85 - 89 | 5.2 | 5.0 | 0.01 | 4.9 | 4.7 | 0.01 | 5.0 | 5.0 | 0.00 |
| 90 - 94 | <0.4 | 0.3 | -0.01 | <0.4 | <0.2 | 0.02 | <0.4 | 0.2 | -0.01 |
| Gender: female | 69.3 | 65.0 | 0.09 | 68.4 | 68.5 | 0.00 | 66.5 | 66.3 | 0.00 |
| **Race** | | | | | | | | | |
| Black or African American | 17.9 | 17.9 | 0.00 | 18.1 | 18.3 | -0.01 | 18.7 | 18.0 | 0.02 |
| White | 63.9 | 61.1 | 0.06 | 63.3 | 61.4 | 0.04 | 62.8 | 61.4 | 0.03 |
| **Ethnicity** | | | | | | | | | |
| Hispanic or Latino | 5.2 | 5.2 | 0.00 | 5.4 | 4.8 | 0.02 | 5.5 | 5.0 | 0.02 |
| **Medical history: General** | | | | | | | | | |
| Acute respiratory disease | 26.3 | 30.1 | -0.08 | 26.4 | 26.7 | -0.01 | 28.3 | 29.1 | -0.02 |
| Attention deficit hyperactivity disorder | <0.4 | 0.4 | -0.04 | <0.4 | 0.4 | -0.07 | <0.4 | 0.4 | -0.06 |
| Chronic liver disease | 2.1 | 3.2 | -0.07 | 2.1 | 2.7 | -0.04 | 2.2 | 3.0 | -0.05 |
| Chronic obstructive lung disease | 27.4 | 25.5 | 0.04 | 27.1 | 26.3 | 0.02 | 27.4 | 25.9 | 0.03 |
| Crohn's disease | <0.4 | <0.2 | -0.02 | <0.4 | <0.2 | 0.00 | <0.4 | <0.2 | -0.02 |
| Dementia | 9.8 | 8.0 | 0.06 | 9.6 | 9.0 | 0.02 | 9.8 | 8.3 | 0.05 |
| Depressive disorder | 20.6 | 18.9 | 0.04 | 20.2 | 21.3 | -0.03 | 18.2 | 19.9 | -0.04 |
| Gastroesophageal reflux disease | 24.2 | 22.1 | 0.05 | 24.6 | 25.0 | -0.01 | 22.1 | 23.3 | -0.03 |
| Gastrointestinal hemorrhage | 3.6 | 3.5 | 0.00 | 3.7 | 3.7 | 0.00 | 3.9 | 3.7 | 0.01 |
| Human immunodeficiency virus infection | <0.4 | 0.3 | -0.05 | <0.4 | 0.4 | -0.06 | <0.4 | 0.3 | -0.03 |
| Hyperlipidemia | 69.8 | 63.5 | 0.13 | 68.4 | 70.3 | -0.04 | 63.1 | 66.0 | -0.06 |
| Hypertensive disorder | 83.3 | 79.7 | 0.09 | 82.6 | 82.7 | 0.00 | 81.7 | 80.9 | 0.02 |
| Lesion of liver | 1.1 | 1.5 | -0.04 | 1.1 | 1.3 | -0.01 | 1.3 | 1.5 | -0.01 |
| Obesity | 16.9 | 14.3 | 0.07 | 16.3 | 16.5 | -0.01 | 15.6 | 15.3 | 0.01 |
| Osteoarthritis | 42.2 | 38.4 | 0.08 | 41.2 | 41.6 | -0.01 | 38.9 | 39.8 | -0.02 |
| Pneumonia | 9.8 | 9.5 | 0.01 | 9.5 | 7.9 | 0.06 | 10.7 | 9.0 | 0.06 |
| Psoriasis | 1.1 | 0.9 | 0.01 | 1.0 | 0.8 | 0.02 | 0.9 | 0.9 | 0.00 |
| Renal impairment | 20.7 | 18.2 | 0.06 | 20.4 | 20.1 | 0.01 | 18.9 | 19.1 | 0.00 |
| Rheumatoid arthritis | 3.2 | 2.3 | 0.06 | 3.2 | 2.2 | 0.06 | 3.1 | 2.2 | 0.06 |
| Schizophrenia | 3.3 | 3.4 | 0.00 | 3.3 | 3.4 | 0.00 | 3.2 | 3.4 | -0.01 |
| Ulcerative colitis | <0.4 | 0.2 | -0.02 | <0.4 | 0.2 | 0.00 | <0.4 | 0.2 | 0.00 |
| Urinary tract infectious disease | 14.2 | 15.0 | -0.02 | 13.9 | 14.6 | -0.02 | 13.6 | 14.8 | -0.03 |
| Viral hepatitis C | 0.5 | 1.4 | -0.09 | 0.5 | 1.2 | -0.08 | 0.5 | 1.4 | -0.09 |
| Visual system disorder | 46.7 | 44.9 | 0.04 | 46.5 | 48.7 | -0.04 | 43.2 | 46.3 | -0.06 |
| **Medical history: Cardiovascular disease** | | | | | | | | | |
| Atrial fibrillation | 11.2 | 10.3 | 0.03 | 11.2 | 9.4 | 0.06 | 12.2 | 10.1 | 0.07 |
| Cerebrovascular disease | 11.9 | 10.8 | 0.03 | 11.6 | 11.4 | 0.00 | 12.0 | 11.1 | 0.03 |
| Coronary arteriosclerosis | 23.1 | 22.7 | 0.01 | 23.2 | 23.2 | 0.00 | 22.3 | 23.0 | -0.01 |
| Heart failure | 17.9 | 17.5 | 0.01 | 17.5 | 17.3 | 0.01 | 18.0 | 17.6 | 0.01 |
| Ischemic heart disease | 12.1 | 13.4 | -0.04 | 12.2 | 13.2 | -0.03 | 12.3 | 13.3 | -0.03 |
| Peripheral vascular disease | 17.9 | 15.8 | 0.06 | 17.5 | 17.7 | 0.00 | 16.3 | 16.8 | -0.01 |
| Pulmonary embolism | 0.6 | 1.0 | -0.05 | 0.5 | 1.3 | -0.08 | 0.4 | 1.1 | -0.08 |
| Venous thrombosis | 2.1 | 2.0 | 0.01 | 1.8 | 2.4 | -0.05 | 1.6 | 2.2 | -0.05 |
| **Medical history: Neoplasms** | | | | | | | | | |
| Hematologic neoplasm | 1.7 | 1.6 | 0.01 | 1.8 | 1.3 | 0.04 | 1.8 | 1.5 | 0.02 |
| Malignant lymphoma | 0.5 | 0.5 | 0.00 | 0.5 | 0.4 | 0.03 | 0.5 | 0.4 | 0.00 |
| Malignant neoplasm of anorectum | 0.5 | 0.3 | 0.03 | <0.4 | 0.3 | 0.03 | 0.7 | 0.3 | 0.06 |
| Malignant neoplastic disease | 10.3 | 11.1 | -0.02 | 10.4 | 11.5 | -0.04 | 10.2 | 11.2 | -0.03 |
| Malignant tumor of breast | 2.6 | 2.5 | 0.01 | 2.5 | 2.9 | -0.02 | 2.3 | 2.7 | -0.02 |
| Malignant tumor of colon | 1.2 | 0.9 | 0.03 | 1.0 | 0.9 | 0.01 | 1.1 | 0.9 | 0.02 |
| Malignant tumor of lung | 0.7 | 0.9 | -0.03 | 0.7 | 0.8 | -0.01 | 0.5 | 0.9 | -0.05 |
| Malignant tumor of urinary bladder | <0.4 | 0.8 | -0.09 | <0.4 | 0.8 | -0.09 | <0.4 | 0.8 | -0.08 |
| Primary malignant neoplasm of prostate | 1.4 | 1.9 | -0.04 | 1.5 | 1.8 | -0.03 | 1.7 | 1.8 | -0.01 |
| **Medication use** | | | | | | | | | |
| Agents acting on the renin-angiotensin system | 72.5 | 70.4 | 0.05 | 72.5 | 72.6 | 0.00 | 70.3 | 71.4 | -0.02 |

| (Continued) | Before ajustment | | | After matching | | | After stratification | | |
|---|---|---|---|---|---|---|---|---|---|
| Characteristic | T (%) | C (%) | StdDiff | T (%) | C (%) | StdDiff | T (%) | C (%) | StdDiff |
| Antibacterials for systemic use | 58.4 | 58.0 | 0.01 | 58.1 | 57.4 | 0.01 | 57.6 | 57.7 | 0.00 |
| Antidepressants | 41.9 | 38.5 | 0.07 | 40.9 | 43.0 | -0.04 | 38.7 | 40.3 | -0.03 |
| Antiepileptics | 30.5 | 28.6 | 0.04 | 29.1 | 31.9 | -0.06 | 27.1 | 29.8 | -0.06 |
| Antiinflammatory and antirheumatic products | 49.3 | 44.6 | 0.09 | 48.2 | 48.7 | -0.01 | 45.7 | 46.2 | -0.01 |
| Antineoplastic agents | 7.5 | 6.0 | 0.06 | 7.5 | 6.2 | 0.05 | 7.5 | 6.0 | 0.06 |
| Antipsoriatics | 0.6 | 0.7 | -0.01 | 0.6 | 0.7 | 0.00 | 0.8 | 0.7 | 0.01 |
| Antithrombotic agents | 40.9 | 37.2 | 0.08 | 40.1 | 41.1 | -0.02 | 38.6 | 38.8 | 0.00 |
| Beta blocking agents | 47.4 | 48.8 | -0.03 | 47.1 | 50.3 | -0.06 | 47.9 | 49.6 | -0.03 |
| Calcium channel blockers | 37.6 | 36.7 | 0.02 | 37.3 | 38.1 | -0.02 | 35.9 | 37.4 | -0.03 |
| Diuretics | 55.5 | 55.5 | 0.00 | 55.8 | 55.5 | 0.01 | 55.8 | 55.6 | 0.01 |
| Drugs for acid related disorders | 55.4 | 49.7 | 0.11 | 54.6 | 55.7 | -0.02 | 51.0 | 52.0 | -0.02 |
| Drugs for obstructive airway diseases | 50.7 | 49.0 | 0.03 | 49.7 | 52.1 | -0.05 | 48.4 | 50.0 | -0.03 |
| Immunosuppressants | 4.1 | 2.2 | 0.11 | 4.3 | 2.3 | 0.11 | 3.9 | 2.2 | 0.10 |
| Lipid modifying agents | 78.3 | 73.0 | 0.12 | 77.6 | 79.4 | -0.04 | 72.1 | 75.2 | -0.07 |
| Opioids | 39.0 | 38.2 | 0.02 | 38.7 | 38.1 | 0.01 | 36.3 | 38.3 | -0.04 |
| Psycholeptics | 36.5 | 35.4 | 0.02 | 36.8 | 36.8 | 0.00 | 36.5 | 35.9 | 0.01 |
| Psychostimulants | 1.5 | 1.4 | 0.01 | 1.4 | 1.2 | 0.02 | 1.4 | 1.4 | 0.00 |

##### 2.6.5 Open Claims

**Supplementary Table 32:** Baseline patient characteristics for DPP4I (T) and SU (C) new-users in the Open Claims data source. We report proportion of initiators satisfying selected base-line characteristics and the standardized difference of population proportions (StdDiff) before and after propensity score adjustment. Less extreme StdDiffs through matching and stratification suggest improved balance between patient cohorts through adjustment.

| | Before adjustment | | | After matching | | | After stratification | | |
|---|---|---|---|---|---|---|---|---|---|
| Characteristic | T (%) | C (%) | StdDiff | T (%) | C (%) | StdDiff | T (%) | C (%) | StdDiff |
| **Age group** | | | | | | | | | |
| 65 - 69 | 36.1 | 36.4 | -0.01 | 36.1 | 36.3 | 0.00 | 36.1 | 36.3 | 0.00 |
| 70 - 74 | 27.5 | 28.2 | -0.02 | 27.6 | 27.7 | 0.00 | 27.7 | 28.2 | -0.01 |
| 75 - 79 | 25.6 | 25.6 | 0.00 | 25.6 | 25.2 | 0.01 | 25.8 | 25.4 | 0.01 |
| 80 - 84 | 10.5 | 9.5 | 0.04 | 10.4 | 10.6 | 0.00 | 9.8 | 9.9 | 0.00 |
| 85 - 89 | 0.3 | 0.2 | 0.01 | 0.3 | 0.2 | 0.02 | 0.3 | 0.2 | 0.01 |
| Gender: female | 53.6 | 49.4 | 0.08 | 53.3 | 53.6 | 0.00 | 51.0 | 50.8 | 0.00 |
| **Medical history: General** | | | | | | | | | |
| Acute respiratory disease | 13.4 | 12.6 | 0.03 | 13.2 | 13.3 | 0.00 | 12.9 | 12.8 | 0.00 |
| Attention deficit hyperactivity disorder | 0.1 | 0.1 | 0.01 | 0.2 | 0.1 | 0.01 | 0.1 | 0.1 | 0.00 |
| Chronic liver disease | 1.3 | 1.3 | 0.00 | 1.3 | 1.4 | 0.00 | 1.3 | 1.4 | 0.00 |
| Chronic obstructive lung disease | 7.8 | 8.1 | -0.01 | 7.8 | 8.0 | -0.01 | 8.0 | 8.0 | 0.00 |
| Crohn's disease | 0.2 | 0.2 | 0.00 | 0.1 | 0.2 | -0.01 | 0.1 | 0.2 | 0.00 |
| Dementia | 2.3 | 1.9 | 0.03 | 2.2 | 2.2 | 0.00 | 2.1 | 2.0 | 0.00 |
| Depressive disorder | 6.3 | 6.0 | 0.01 | 6.2 | 6.3 | 0.00 | 6.1 | 6.0 | 0.00 |
| Gastroesophageal reflux disease | 9.4 | 8.4 | 0.03 | 9.2 | 9.2 | 0.00 | 8.8 | 8.7 | 0.00 |
| Gastrointestinal hemorrhage | 1.9 | 1.7 | 0.01 | 1.8 | 1.9 | -0.01 | 1.8 | 1.8 | 0.00 |
| Human immunodeficiency virus infection | 0.2 | 0.1 | 0.01 | 0.2 | 0.1 | 0.01 | 0.2 | 0.1 | 0.01 |
| Hyperlipidemia | 49.0 | 44.6 | 0.09 | 48.5 | 48.3 | 0.00 | 46.3 | 45.9 | 0.01 |
| Hypertensive disorder | 57.5 | 54.2 | 0.07 | 57.1 | 57.2 | 0.00 | 55.3 | 55.2 | 0.00 |
| Lesion of liver | 0.8 | 0.9 | -0.01 | 0.9 | 0.9 | 0.00 | 0.9 | 0.9 | 0.00 |
| Obesity | 7.4 | 7.2 | 0.01 | 7.4 | 7.5 | -0.01 | 7.2 | 7.4 | -0.01 |
| Osteoarthritis | 19.7 | 18.0 | 0.04 | 19.5 | 19.3 | 0.00 | 18.8 | 18.4 | 0.01 |
| Pneumonia | 3.4 | 3.5 | 0.00 | 3.4 | 3.5 | 0.00 | 3.5 | 3.5 | 0.00 |
| Psoriasis | 0.9 | 0.8 | 0.01 | 0.8 | 0.8 | 0.00 | 0.8 | 0.8 | 0.00 |
| Renal impairment | 12.9 | 12.5 | 0.01 | 12.8 | 13.0 | 0.00 | 12.6 | 12.5 | 0.00 |
| Rheumatoid arthritis | 1.4 | 1.2 | 0.02 | 1.3 | 1.3 | 0.00 | 1.3 | 1.3 | 0.00 |
| Schizophrenia | 0.2 | 0.2 | 0.01 | 0.2 | 0.2 | 0.00 | 0.2 | 0.2 | 0.01 |
| Ulcerative colitis | 0.2 | 0.2 | 0.01 | 0.3 | 0.2 | 0.00 | 0.2 | 0.2 | 0.01 |
| Urinary tract infectious disease | 8.0 | 7.0 | 0.04 | 7.8 | 7.9 | 0.00 | 7.3 | 7.3 | 0.00 |
| Viral hepatitis C | 0.3 | 0.3 | 0.00 | 0.3 | 0.3 | 0.00 | 0.3 | 0.3 | 0.00 |
| Visual system disorder | 27.0 | 25.1 | 0.04 | 25.8 | 25.8 | 0.00 | 25.8 | 25.8 | 0.00 |
| **Medical history: Cardiovascular disease** | | | | | | | | | |
| Atrial fibrillation | 7.8 | 7.5 | 0.01 | 7.8 | 8.0 | -0.01 | 7.5 | 7.7 | -0.01 |
| Cerebrovascular disease | 6.1 | 5.4 | 0.03 | 6.0 | 5.9 | 0.00 | 5.7 | 5.6 | 0.01 |
| Coronary arteriosclerosis | 14.1 | 13.4 | 0.02 | 13.9 | 14.1 | 0.00 | 13.5 | 13.6 | 0.00 |
| Heart disease | 28.6 | 27.1 | 0.04 | 28.3 | 28.5 | 0.00 | 27.4 | 27.6 | 0.00 |
| Heart failure | 7.0 | 6.9 | 0.01 | 6.9 | 7.1 | -0.01 | 6.9 | 7.0 | 0.00 |
| Ischemic heart disease | 6.1 | 5.9 | 0.01 | 6.1 | 6.2 | -0.01 | 5.9 | 6.0 | 0.00 |
| Peripheral vascular disease | 7.6 | 7.3 | 0.01 | 7.5 | 7.3 | 0.00 | 7.4 | 7.3 | 0.01 |
| Pulmonary embolism | 0.7 | 0.7 | 0.00 | 0.7 | 0.7 | 0.00 | 0.6 | 0.7 | 0.00 |
| Venous thrombosis | 1.3 | 1.3 | 0.00 | 1.3 | 1.3 | 0.00 | 1.3 | 1.3 | 0.00 |
| **Medical history: Neoplasms** | | | | | | | | | |
| Hematologic neoplasm | 1.1 | 1.2 | 0.00 | 1.1 | 1.1 | 0.00 | 1.2 | 1.2 | 0.00 |
| Malignant lymphoma | 0.6 | 0.7 | 0.00 | 0.7 | 0.6 | 0.00 | 0.7 | 0.6 | 0.00 |
| Malignant neoplasm of anorectum | 0.2 | 0.2 | 0.00 | 0.2 | 0.2 | 0.00 | 0.2 | 0.2 | 0.00 |
| Malignant neoplastic disease | 10.5 | 10.3 | 0.01 | 10.5 | 10.6 | 0.00 | 10.5 | 10.6 | 0.00 |
| Malignant tumor of breast | 1.9 | 1.7 | 0.01 | 1.9 | 1.9 | 0.00 | 1.8 | 1.8 | 0.00 |
| Malignant tumor of colon | 0.6 | 0.5 | 0.00 | 0.5 | 0.6 | 0.00 | 0.6 | 0.6 | 0.00 |
| Malignant tumor of lung | 0.5 | 0.5 | -0.01 | 0.4 | 0.5 | 0.00 | 0.5 | 0.5 | 0.00 |
| Malignant tumor of urinary bladder | 0.6 | 0.6 | 0.00 | 0.6 | 0.6 | 0.00 | 0.6 | 0.6 | 0.00 |
| Primary malignant neoplasm of prostate | 2.4 | 2.4 | 0.00 | 2.3 | 2.4 | 0.00 | 2.3 | 2.4 | 0.00 |
| **Medication use** | | | | | | | | | |
| Agents acting on the renin-angiotensin system | 73.7 | 72.6 | 0.02 | 73.4 | 73.5 | 0.00 | 72.9 | 72.7 | 0.00 |
| Antibacterials for systemic use | 58.8 | 56.1 | 0.05 | 58.5 | 58.8 | -0.01 | 57.0 | 56.9 | 0.00 |
| Antidepressants | 27.6 | 26.4 | 0.03 | 27.3 | 27.5 | 0.00 | 26.9 | 26.7 | 0.00 |
| Antiepileptics | 19.8 | 19.2 | 0.02 | 19.6 | 19.7 | 0.00 | 19.4 | 19.3 | 0.00 |
| Antiinflammatory and antirheumatic products | 34.3 | 31.1 | 0.07 | 33.8 | 34.1 | 0.00 | 32.3 | 32.2 | 0.00 |
| Antineoplastic agents | 6.7 | 5.6 | 0.05 | 6.4 | 6.4 | 0.00 | 6.0 | 6.0 | 0.00 |

| (Continued) | Before ajustment | | | After matching | | | After stratification | | |
|---|---|---|---|---|---|---|---|---|---|
| Characteristic | T (%) | C (%) | StdDiff | T (%) | C (%) | StdDiff | T (%) | C (%) | StdDiff |
| Antipsoriatics | 0.8 | 0.6 | 0.02 | 0.8 | 0.8 | -0.01 | 0.7 | 0.6 | 0.00 |
| Antithrombotic agents | 27.6 | 25.3 | 0.05 | 27.0 | 27.3 | -0.01 | 26.0 | 25.9 | 0.00 |
| Beta blocking agents | 45.9 | 45.4 | 0.01 | 45.7 | 45.5 | 0.00 | 45.7 | 45.3 | 0.01 |
| Calcium channel blockers | 34.6 | 33.0 | 0.03 | 34.4 | 34.3 | 0.00 | 33.7 | 33.6 | 0.00 |
| Diuretics | 47.7 | 47.6 | 0.00 | 47.6 | 47.7 | 0.00 | 47.8 | 47.6 | 0.00 |
| Drugs for acid related disorders | 39.5 | 36.2 | 0.07 | 38.8 | 39.3 | -0.01 | 37.3 | 37.1 | 0.00 |
| Drugs for obstructive airway diseases | 39.4 | 36.2 | 0.07 | 38.9 | 39.0 | 0.00 | 37.4 | 37.1 | 0.01 |
| Immunosuppressants | 3.6 | 2.7 | 0.05 | 3.4 | 3.3 | 0.00 | 3.2 | 2.9 | 0.01 |
| Lipid modifying agents | 78.0 | 74.9 | 0.07 | 77.7 | 77.8 | 0.00 | 76.0 | 76.0 | 0.00 |
| Opioids | 25.8 | 25.1 | 0.02 | 25.5 | 25.8 | -0.01 | 25.3 | 25.4 | 0.00 |
| Psycholeptics | 23.1 | 21.8 | 0.03 | 22.9 | 23.0 | 0.00 | 22.4 | 22.2 | 0.01 |
| Psychostimulants | 1.4 | 1.2 | 0.02 | 1.4 | 1.3 | 0.00 | 1.3 | 1.2 | 0.00 |

##### 2.6.6 OptumEHR

**Supplementary Table 33:** Baseline patient characteristics for DPP4I (T) and SU (C) new-users in the OptumEHR data source. We report proportion of initiators satisfying selected base-line characteristics and the standardized difference of population proportions (StdDiff) before and after propensity score adjustment. Less extreme StdDiffs through matching and stratification suggest improved balance between patient cohorts through adjustment.

| | Before adjustment | | | After matching | | | After stratification | | |
|---|---|---|---|---|---|---|---|---|---|
| Characteristic | T (%) | C (%) | StdDiff | T (%) | C (%) | StdDiff | T (%) | C (%) | StdDiff |
| **Age group** | | | | | | | | | |
| 65 - 69 | 40.1 | 37.7 | 0.05 | 40.0 | 40.2 | 0.00 | 40.0 | 40.2 | 0.00 |
| 70 - 74 | 27.7 | 28.0 | -0.01 | 27.7 | 27.7 | 0.00 | 27.7 | 27.9 | 0.00 |
| 75 - 79 | 18.2 | 20.3 | -0.06 | 18.3 | 18.1 | 0.01 | 19.8 | 19.7 | 0.00 |
| 80 - 84 | 11.0 | 11.6 | -0.02 | 11.0 | 11.0 | 0.00 | 11.0 | 11.0 | 0.00 |
| 85 - 89 | 3.0 | 2.4 | 0.04 | 3.0 | 3.0 | 0.00 | 2.7 | 2.6 | 0.01 |
| Gender: female | 52.0 | 47.4 | 0.09 | 51.2 | 51.9 | -0.01 | 48.6 | 48.8 | -0.01 |
| **Race** | | | | | | | | | |
| Asian | 2.5 | 2.1 | 0.03 | 2.5 | 2.3 | 0.01 | 2.1 | 2.2 | 0.00 |
| Black or African American | 8.9 | 7.6 | 0.05 | 8.8 | 9.1 | -0.01 | 7.8 | 8.1 | -0.01 |
| White | 82.0 | 84.4 | -0.06 | 82.3 | 82.2 | 0.00 | 84.1 | 83.7 | 0.01 |
| **Ethnicity** | | | | | | | | | |
| Hispanic or Latino | 4.4 | 4.1 | 0.01 | 4.4 | 4.5 | -0.01 | 3.9 | 4.2 | -0.01 |
| Not Hispanic or Latino | 87.0 | 88.5 | -0.05 | 87.3 | 87.4 | 0.00 | 88.4 | 88.1 | 0.01 |
| **Medical history: General** | | | | | | | | | |
| Acute respiratory disease | 15.7 | 14.6 | 0.03 | 15.4 | 15.7 | -0.01 | 15.2 | 15.0 | 0.00 |
| Attention deficit hyperactivity disorder | 0.2 | 0.2 | 0.01 | 0.2 | 0.2 | 0.00 | 0.2 | 0.2 | 0.00 |
| Chronic liver disease | 2.0 | 2.2 | -0.01 | 2.0 | 2.1 | -0.01 | 2.1 | 2.1 | 0.00 |
| Chronic obstructive lung disease | 8.4 | 8.8 | -0.01 | 8.3 | 8.4 | 0.00 | 8.8 | 8.7 | 0.00 |
| Crohn’s disease | 0.2 | 0.2 | -0.01 | 0.2 | 0.2 | -0.01 | 0.2 | 0.2 | -0.01 |
| Depressive disorder | 11.3 | 11.0 | 0.01 | 11.1 | 11.4 | -0.01 | 11.1 | 11.1 | 0.00 |
| Gastroesophageal reflux disease | 18.3 | 16.4 | 0.05 | 17.6 | 18.2 | -0.01 | 17.2 | 17.0 | 0.00 |
| Gastrointestinal hemorrhage | 1.7 | 1.7 | 0.00 | 1.6 | 1.7 | 0.00 | 1.6 | 1.7 | -0.01 |
| Human immunodeficiency virus infection | 0.1 | 0.1 | 0.01 | 0.1 | 0.1 | 0.00 | 0.1 | 0.1 | 0.00 |
| Hyperlipidemia | 72.3 | 68.7 | 0.08 | 71.3 | 72.2 | -0.02 | 69.6 | 69.9 | -0.01 |
| Hypertensive disorder | 74.8 | 72.4 | 0.06 | 74.2 | 74.7 | -0.01 | 72.8 | 73.2 | -0.01 |
| Lesion of liver | 0.8 | 1.0 | -0.02 | 0.8 | 0.9 | -0.01 | 0.9 | 1.0 | -0.01 |
| Obesity | 17.4 | 16.0 | 0.04 | 16.9 | 17.1 | -0.01 | 16.6 | 16.4 | 0.01 |
| Osteoarthritis | 22.8 | 21.5 | 0.03 | 22.2 | 22.7 | -0.01 | 22.0 | 21.9 | 0.00 |
| Pneumonia | 2.7 | 3.1 | -0.03 | 2.7 | 2.8 | 0.00 | 3.0 | 3.0 | 0.00 |
| Psoriasis | 1.2 | 1.2 | 0.00 | 1.2 | 1.3 | 0.00 | 1.2 | 1.3 | -0.01 |
| Renal impairment | 12.9 | 13.8 | -0.03 | 12.9 | 13.0 | 0.00 | 13.9 | 13.6 | 0.01 |
| Schizophrenia | 0.2 | 0.1 | 0.01 | 0.2 | 0.1 | 0.01 | 0.2 | 0.1 | 0.01 |
| Ulcerative colitis | 0.2 | 0.3 | -0.01 | 0.2 | 0.3 | -0.01 | 0.2 | 0.3 | -0.01 |
| Urinary tract infectious disease | 7.5 | 7.0 | 0.02 | 7.3 | 7.2 | 0.00 | 7.2 | 7.1 | 0.01 |
| Viral hepatitis C | 0.3 | 0.3 | 0.00 | 0.3 | 0.3 | 0.00 | 0.3 | 0.3 | 0.00 |
| Visual system disorder | 17.3 | 19.3 | -0.05 | 17.3 | 17.2 | 0.00 | 19.0 | 18.8 | 0.01 |
| **Medical history: Cardiovascular disease** | | | | | | | | | |
| Atrial fibrillation | 9.4 | 9.3 | 0.00 | 9.3 | 9.3 | 0.00 | 9.7 | 9.3 | 0.01 |
| Cerebrovascular disease | 5.9 | 5.7 | 0.01 | 5.7 | 5.9 | -0.01 | 5.7 | 5.8 | 0.00 |
| Coronary arteriosclerosis | 19.0 | 18.7 | 0.01 | 18.6 | 18.7 | 0.00 | 18.7 | 18.8 | 0.00 |
| Heart disease | 34.8 | 33.9 | 0.02 | 34.3 | 34.5 | 0.00 | 34.5 | 34.1 | 0.01 |
| Heart failure | 6.1 | 6.8 | -0.03 | 6.1 | 6.1 | 0.00 | 6.8 | 6.6 | 0.01 |
| Ischemic heart disease | 7.6 | 7.8 | -0.01 | 7.5 | 7.3 | 0.01 | 7.9 | 7.7 | 0.01 |
| Peripheral vascular disease | 7.5 | 7.8 | -0.01 | 7.3 | 7.4 | -0.01 | 7.8 | 7.7 | 0.00 |
| Pulmonary embolism | 0.7 | 0.8 | -0.01 | 0.7 | 0.7 | 0.00 | 0.8 | 0.8 | 0.00 |
| Venous thrombosis | 1.4 | 1.6 | -0.01 | 1.4 | 1.4 | 0.00 | 1.6 | 1.5 | 0.00 |
| **Medical history: Neoplasms** | | | | | | | | | |
| Hematologic neoplasm | 1.2 | 1.2 | -0.01 | 1.2 | 1.2 | 0.00 | 1.2 | 1.2 | -0.01 |
| Malignant lymphoma | 0.7 | 0.7 | 0.00 | 0.7 | 0.7 | 0.00 | 0.7 | 0.7 | 0.00 |
| Malignant neoplasm of anorectum | 0.2 | 0.2 | -0.01 | 0.2 | 0.2 | 0.00 | 0.2 | 0.2 | 0.01 |
| Malignant neoplastic disease | 12.0 | 12.1 | -0.01 | 11.9 | 12.0 | 0.00 | 12.1 | 12.1 | 0.00 |
| Malignant tumor of breast | 2.2 | 1.9 | 0.02 | 2.1 | 2.3 | -0.01 | 1.9 | 2.0 | -0.01 |
| Malignant tumor of colon | 0.6 | 0.7 | -0.01 | 0.6 | 0.6 | 0.00 | 0.6 | 0.6 | 0.00 |
| Malignant tumor of lung | 0.5 | 0.5 | 0.00 | 0.4 | 0.5 | -0.01 | 0.5 | 0.5 | 0.00 |
| Malignant tumor of urinary bladder | 0.7 | 0.7 | -0.01 | 0.6 | 0.7 | 0.00 | 0.7 | 0.7 | 0.00 |
| Primary malignant neoplasm of prostate | 2.8 | 3.0 | -0.01 | 2.9 | 3.0 | 0.00 | 2.9 | 3.0 | 0.00 |
| **Medication use** | | | | | | | | | |
| Agents acting on the renin-angiotensin system | 72.1 | 71.1 | 0.02 | 71.7 | 71.7 | 0.00 | 71.5 | 71.4 | 0.00 |

| (Continued) | Before ajustment | | | After matching | | | After stratification | | |
|---|---|---|---|---|---|---|---|---|---|
| Characteristic | T (%) | C (%) | StdDiff | T (%) | C (%) | StdDiff | T (%) | C (%) | StdDiff |
| Antibacterials for systemic use | 42.8 | 41.6 | 0.03 | 42.3 | 42.5 | 0.00 | 42.4 | 42.0 | 0.01 |
| Antidepressants | 26.7 | 25.6 | 0.03 | 26.4 | 26.6 | -0.01 | 26.0 | 25.9 | 0.00 |
| Antiepileptics | 17.1 | 16.6 | 0.01 | 17.0 | 16.7 | 0.01 | 17.0 | 16.7 | 0.01 |
| Antiinflammatory and antirheumatic products | 60.3 | 60.7 | -0.01 | 59.9 | 59.9 | 0.00 | 60.7 | 60.5 | 0.00 |
| Antineoplastic agents | 6.0 | 5.2 | 0.03 | 5.8 | 6.0 | -0.01 | 5.6 | 5.5 | 0.00 |
| Antipsoriatics | 0.6 | 0.5 | 0.01 | 0.6 | 0.5 | 0.01 | 0.6 | 0.5 | 0.01 |
| Antithrombotic agents | 54.6 | 56.1 | -0.03 | 54.5 | 54.1 | 0.01 | 56.3 | 55.6 | 0.01 |
| Beta blocking agents | 45.7 | 46.7 | -0.02 | 45.4 | 45.3 | 0.00 | 47.0 | 46.4 | 0.01 |
| Calcium channel blockers | 31.2 | 29.8 | 0.03 | 30.9 | 31.1 | 0.00 | 30.4 | 30.3 | 0.00 |
| Diuretics | 47.6 | 48.8 | -0.02 | 48.9 | 48.5 | 0.01 | 48.9 | 48.5 | 0.01 |
| Drugs for acid related disorders | 44.7 | 43.5 | 0.03 | 44.1 | 44.5 | -0.01 | 44.0 | 43.9 | 0.00 |
| Drugs for obstructive airway diseases | 42.3 | 40.6 | 0.03 | 41.7 | 42.2 | -0.01 | 41.3 | 41.2 | 0.00 |
| Immunosuppressants | 3.0 | 2.6 | 0.02 | 2.9 | 2.9 | 0.00 | 2.8 | 2.7 | 0.01 |
| Lipid modifying agents | 79.7 | 77.9 | 0.04 | 79.1 | 79.3 | 0.00 | 78.3 | 78.4 | 0.00 |
| Opioids | 27.0 | 27.8 | -0.02 | 26.8 | 27.1 | -0.01 | 28.1 | 27.6 | 0.01 |
| Psycholeptics | 28.2 | 27.0 | 0.03 | 27.9 | 28.1 | 0.00 | 27.6 | 27.4 | 0.00 |
| Psychostimulants | 1.7 | 1.3 | 0.03 | 1.6 | 1.5 | 0.01 | 1.4 | 1.3 | 0.00 |

### 2.6.7 VA

**Supplementary Table 34:** Baseline patient characteristics for DPP4I (T) and SU (C) new-users in the VA data source. We report proportion of initiators satisfying selected base-line characteristics and the standardized difference of population proportions (StdDiff) before and after propensity score adjustment. Less extreme StdDiffs through matching and stratification suggest improved balance between patient cohorts through adjustment.

| | Before adjustment | | | After matching | | | After stratification | | |
|---|---|---|---|---|---|---|---|---|---|
| Characteristic | T (%) | C (%) | StdDiff | T (%) | C (%) | StdDiff | T (%) | C (%) | StdDiff |
| **Age group** | | | | | | | | | |
| 65 - 69 | 33.2 | 40.7 | -0.16 | 33.7 | 33.1 | 0.01 | 40.4 | 43.1 | -0.05 |
| 70 - 74 | 38.4 | 28.6 | 0.21 | 38.3 | 38.8 | -0.01 | 27.5 | 29.8 | -0.05 |
| 75 - 79 | 15.0 | 16.5 | -0.04 | 14.8 | 15.0 | -0.01 | 16.2 | 14.2 | 0.06 |
| 80 - 84 | 7.5 | 9.6 | -0.07 | 7.5 | 7.3 | 0.01 | 8.3 | 8.0 | 0.01 |
| 85 - 89 | 4.1 | 3.7 | 0.02 | 4.0 | 4.1 | 0.00 | 4.9 | 3.8 | 0.05 |
| 90 - 94 | 1.5 | 0.8 | 0.07 | 1.4 | 1.5 | 0.00 | 2.3 | 1.0 | 0.10 |
| 95 - 99 | 0.3 | 0.1 | 0.04 | 0.3 | 0.2 | 0.02 | 0.4 | 0.1 | 0.05 |
| Gender: female | 2.7 | 1.6 | 0.08 | 2.7 | 2.6 | 0.01 | 2.1 | 1.8 | 0.02 |
| **Race** | | | | | | | | | |
| Asian | 1.3 | 0.5 | 0.08 | 1.2 | 1.1 | 0.00 | 1.3 | 0.7 | 0.07 |
| Black or African American | 10.7 | 8.4 | 0.08 | 10.7 | 10.9 | -0.01 | 8.8 | 9.5 | -0.02 |
| White | 80.6 | 79.0 | 0.04 | 80.6 | 80.3 | 0.01 | 81.1 | 81.2 | 0.00 |
| Unknown | 5.9 | 10.7 | -0.17 | 6.0 | 6.1 | 0.00 | 6.7 | 7.1 | -0.02 |
| Native Hawaiian or Other Pacific Islander | 0.9 | 0.8 | 0.01 | 0.9 | 0.8 | 0.01 | 1.6 | 0.8 | 0.07 |
| American Indian or Alaska Native | 0.7 | 0.6 | 0.01 | 0.7 | 0.8 | -0.01 | 0.5 | 0.7 | -0.02 |
| **Ethnicity** | | | | | | | | | |
| Hispanic or Latino | 4.4 | 4.5 | 0.00 | 4.4 | 4.4 | 0.00 | 4.7 | 4.9 | -0.01 |
| Not Hispanic or Latino | 93.2 | 88.7 | 0.16 | 93.1 | 93.2 | -0.01 | 91.6 | 91.8 | -0.01 |
| **Medical history: General** | | | | | | | | | |
| Acute respiratory disease | 7.4 | 7.5 | 0.00 | 7.3 | 7.1 | 0.01 | 6.6 | 7.9 | -0.05 |
| Attention deficit hyperactivity disorder | 0.3 | 0.2 | 0.03 | 0.3 | 0.3 | 0.01 | 0.2 | 0.2 | 0.00 |
| Chronic liver disease | 1.8 | 1.7 | 0.01 | 1.8 | 1.9 | -0.01 | 1.5 | 2.0 | -0.04 |
| Chronic obstructive lung disease | 14.5 | 13.8 | 0.02 | 14.4 | 14.5 | 0.00 | 14.8 | 14.4 | 0.01 |
| Crohn's disease | 0.2 | 0.2 | 0.00 | 0.2 | 0.2 | -0.01 | 0.2 | 0.2 | 0.00 |
| Dementia | 2.6 | 1.7 | 0.06 | 2.6 | 2.6 | 0.00 | 2.7 | 1.9 | 0.05 |
| Depressive disorder | 16.2 | 14.5 | 0.05 | 20.1 | 16.2 | 0.10 | 20.1 | 16.2 | 0.10 |
| Gastroesophageal reflux disease | 19.7 | 19.2 | 0.01 | 19.7 | 19.7 | 0.00 | 22.9 | 20.4 | 0.06 |
| Gastrointestinal hemorrhage | 1.1 | 1.3 | -0.01 | 1.1 | 1.0 | 0.01 | 1.1 | 1.2 | -0.01 |
| Human immunodeficiency virus infection | 0.3 | 0.1 | 0.03 | 0.3 | 0.3 | 0.00 | 0.2 | 0.2 | -0.01 |
| Hyperlipidemia | 74.2 | 74.9 | -0.01 | 73.9 | 74.0 | 0.00 | 76.8 | 76.5 | 0.01 |
| Hypertensive disorder | 78.3 | 81.0 | -0.07 | 78.2 | 78.0 | 0.00 | 80.5 | 80.9 | -0.01 |
| Lesion of liver | 1.3 | 1.0 | 0.03 | 1.3 | 1.4 | -0.01 | 0.8 | 1.2 | -0.05 |
| Obesity | 21.7 | 22.3 | -0.01 | 21.6 | 21.7 | 0.00 | 25.6 | 23.7 | 0.04 |
| Osteoarthritis | 20.0 | 21.5 | -0.04 | 19.7 | 19.7 | 0.00 | 24.4 | 21.6 | 0.07 |
| Psoriasis | 1.7 | 1.5 | 0.01 | 1.7 | 1.5 | 0.01 | 1.8 | 1.6 | 0.01 |
| Renal impairment | 11.4 | 7.9 | 0.12 | 11.2 | 11.7 | -0.02 | 12.4 | 9.6 | 0.09 |
| Rheumatoid arthritis | 0.9 | 0.9 | 0.00 | 0.8 | 0.9 | -0.02 | 0.8 | 0.9 | -0.02 |
| Schizophrenia | 0.7 | 0.6 | 0.01 | 0.6 | 0.6 | 0.00 | 0.5 | 0.7 | -0.02 |
| Ulcerative colitis | 0.5 | 0.4 | 0.01 | 0.4 | 0.5 | 0.00 | 0.6 | 0.4 | 0.03 |
| Urinary tract infectious disease | 2.6 | 2.7 | 0.00 | 2.6 | 2.5 | 0.00 | 2.6 | 2.8 | -0.01 |
| Viral hepatitis C | 0.9 | 0.9 | 0.00 | 1.0 | 1.1 | -0.01 | 0.8 | 1.1 | -0.04 |
| Visual system disorder | 48.9 | 46.4 | 0.05 | 48.9 | 48.4 | 0.01 | 47.6 | 48.6 | -0.02 |
| **Medical history: Cardiovascular disease** | | | | | | | | | |
| Atrial fibrillation | 10.8 | 8.9 | 0.06 | 10.6 | 10.9 | -0.01 | 14.0 | 9.2 | 0.15 |
| Cerebrovascular disease | 4.2 | 6.1 | -0.09 | 4.2 | 4.2 | 0.00 | 8.5 | 5.6 | 0.12 |
| Coronary arteriosclerosis | 21.0 | 21.8 | -0.02 | 21.0 | 21.4 | -0.01 | 22.5 | 20.3 | 0.06 |
| Heart disease | 37.2 | 40.9 | -0.07 | 37.2 | 37.6 | -0.01 | 48.0 | 39.0 | 0.18 |
| Heart failure | 5.4 | 6.6 | -0.05 | 5.4 | 5.5 | 0.00 | 8.8 | 6.2 | 0.10 |
| Ischemic heart disease | 8.5 | 14.9 | -0.20 | 8.6 | 8.4 | 0.01 | 17.1 | 13.7 | 0.09 |
| Peripheral vascular disease | 6.3 | 6.8 | -0.02 | 6.2 | 6.4 | 0.00 | 6.9 | 6.4 | 0.02 |
| Pulmonary embolism | 0.6 | 0.5 | 0.01 | 0.6 | 0.6 | -0.01 | 1.3 | 0.6 | 0.07 |
| Venous thrombosis | 1.4 | 1.1 | 0.02 | 1.4 | 1.3 | 0.01 | 2.4 | 1.3 | 0.08 |
| **Medical history: Neoplasms** | | | | | | | | | |
| Hematologic neoplasm | 1.4 | 1.0 | 0.04 | 1.4 | 1.4 | 0.00 | 1.4 | 1.2 | 0.02 |
| Malignant lymphoma | 0.9 | 0.6 | 0.04 | 0.9 | 1.0 | -0.01 | 0.6 | 0.7 | -0.02 |
| Malignant neoplasm of anorectum | 0.1 | 0.2 | -0.02 | <0.1 | 0.1 | 0.00 | <0.1 | 0.1 | 0.00 |
| Malignant neoplastic disease | 12.5 | 13.9 | -0.04 | 12.5 | 12.5 | 0.00 | 14.6 | 13.9 | 0.02 |
| Malignant tumor of breast | 0.1 | 0.1 | -0.01 | <0.1 | 0.1 | -0.01 | 0.1 | 0.1 | 0.01 |

| (Continued) | Before ajustment | | | After matching | | | After stratification | | |
|---|---|---|---|---|---|---|---|---|---|
| Characteristic | T (%) | C (%) | StdDiff | T (%) | C (%) | StdDiff | T (%) | C (%) | StdDiff |
| Malignant tumor of colon | 0.6 | 1.0 | -0.04 | 0.6 | 0.6 | 0.00 | 1.2 | 0.8 | 0.03 |
| Malignant tumor of lung | 0.5 | 0.3 | 0.03 | 0.5 | 0.4 | 0.01 | 0.3 | 0.4 | -0.01 |
| Malignant tumor of urinary bladder | 0.9 | 1.0 | -0.01 | 0.9 | 0.9 | 0.00 | 0.8 | 1.0 | -0.03 |
| Primary malignant neoplasm of prostate | 5.2 | 6.5 | -0.06 | 5.2 | 5.0 | 0.01 | 5.9 | 6.1 | -0.01 |
| **Medication use** | | | | | | | | | |
| Agents acting on the renin-angiotensin system | 65.5 | 73.7 | -0.18 | 65.7 | 65.7 | 0.00 | 69.7 | 71.6 | -0.04 |
| Antibacterials for systemic use | 26.2 | 27.9 | -0.04 | 26.0 | 25.8 | 0.00 | 28.7 | 28.5 | 0.00 |
| Antidepressants | 29.4 | 26.8 | 0.06 | 33.1 | 29.0 | 0.09 | 33.1 | 29.0 | 0.09 |
| Antiepileptics | 24.3 | 18.0 | 0.15 | 24.3 | 24.5 | 0.00 | 23.0 | 21.5 | 0.04 |
| Antiinflammatory and antirheumatic products | 41.9 | 39.7 | 0.04 | 41.7 | 42.1 | -0.01 | 39.3 | 40.6 | -0.03 |
| Antineoplastic agents | 4.5 | 3.5 | 0.05 | 4.4 | 4.4 | 0.00 | 4.8 | 3.8 | 0.05 |
| Antipsoriatics | 1.0 | 0.7 | 0.03 | 0.9 | 1.0 | 0.00 | 1.9 | 0.8 | 0.10 |
| Antithrombotic agents | 36.8 | 35.8 | 0.02 | 36.6 | 37.1 | -0.01 | 40.9 | 35.6 | 0.11 |
| Beta blocking agents | 41.4 | 47.2 | -0.12 | 41.7 | 42.0 | -0.01 | 46.8 | 46.2 | 0.01 |
| Calcium channel blockers | 29.8 | 30.4 | -0.01 | 29.6 | 29.8 | 0.00 | 31.9 | 30.2 | 0.04 |
| Diuretics | 38.1 | 47.1 | -0.18 | 41.9 | 45.4 | -0.07 | 41.9 | 45.4 | -0.07 |
| Drugs for obstructive airway diseases | 38.2 | 34.1 | 0.09 | 38.0 | 38.1 | 0.00 | 37.2 | 36.3 | 0.02 |
| Immunosuppressants | 2.4 | 1.6 | 0.06 | 2.4 | 2.3 | 0.00 | 2.6 | 1.9 | 0.04 |
| Lipid modifying agents | 80.0 | 81.1 | -0.03 | 80.0 | 80.2 | -0.01 | 80.8 | 82.3 | -0.04 |
| Opioids | 14.0 | 16.8 | -0.08 | 14.0 | 13.7 | 0.01 | 18.1 | 17.3 | 0.02 |
| Psycholeptics | 18.8 | 17.3 | 0.04 | 18.6 | 18.4 | 0.00 | 20.3 | 18.7 | 0.04 |
| Psychostimulants | 0.9 | 0.8 | 0.01 | 0.9 | 0.9 | 0.00 | 0.5 | 0.9 | -0.04 |

##### 2.6.8 GermanyDA

**Supplementary Table 35:** Baseline patient characteristics for DPP4I (T) and SU (C) new-users in the GermanyDA data source. We report proportion of initiators satisfying selected base-line characteristics and the standardized difference of population proportions (StdDiff) before and after propensity score adjustment. Less extreme StdDiffs through matching and stratification suggest improved balance between patient cohorts through adjustment.

| | Before adjustment | | | After matching | | | After stratification | | |
|---|---|---|---|---|---|---|---|---|---|
| Characteristic | T (%) | C (%) | StdDiff | T (%) | C (%) | StdDiff | T (%) | C (%) | StdDiff |
| **Age group** | | | | | | | | | |
| 65 - 69 | 27.0 | 27.7 | -0.02 | 26.8 | 23.5 | 0.08 | 26.9 | 25.6 | 0.03 |
| 70 - 74 | 25.4 | 28.8 | -0.08 | 28.2 | 28.0 | 0.00 | 26.0 | 26.1 | 0.00 |
| 75 - 79 | 22.9 | 24.0 | -0.03 | 23.2 | 24.9 | -0.04 | 23.2 | 22.5 | 0.02 |
| 80 - 84 | 16.0 | 13.9 | 0.06 | 15.4 | 15.9 | -0.01 | 15.7 | 16.7 | -0.03 |
| 85 - 89 | 6.8 | 4.6 | 0.09 | 5.5 | 6.3 | -0.03 | 6.4 | 6.7 | -0.01 |
| 90 - 94 | 1.8 | 0.9 | 0.08 | 0.8 | 1.4 | -0.05 | 1.6 | 2.4 | -0.06 |
| Gender: female | 48.6 | 50.4 | -0.04 | 50.3 | 50.0 | 0.00 | 49.0 | 47.9 | 0.02 |
| **Medical history: General** | | | | | | | | | |
| Acute respiratory disease | 12.7 | 13.5 | -0.02 | 13.8 | 13.1 | 0.02 | 13.0 | 12.4 | 0.02 |
| Attention deficit hyperactivity disorder | 0.2 | 0.5 | -0.06 | <0.2 | 0.7 | -0.08 | 0.2 | 0.5 | -0.05 |
| Chronic liver disease | 0.5 | 0.3 | 0.03 | 0.3 | 0.2 | 0.01 | 0.5 | 0.2 | 0.04 |
| Chronic obstructive lung disease | 7.0 | 7.2 | -0.01 | 7.4 | 7.5 | 0.00 | 7.0 | 6.6 | 0.01 |
| Crohn's disease | 0.1 | <0.1 | 0.02 | <0.2 | <0.1 | 0.00 | 0.1 | <0.1 | 0.02 |
| Dementia | 3.6 | 3.8 | -0.01 | 3.4 | 4.0 | -0.04 | 3.5 | 3.7 | -0.01 |
| Depressive disorder | 8.1 | 8.9 | -0.03 | 8.7 | 9.2 | -0.02 | 8.2 | 8.6 | -0.01 |
| Gastroesophageal reflux disease | 2.7 | 2.6 | 0.01 | 3.1 | 2.6 | 0.03 | 2.8 | 2.3 | 0.03 |
| Gastrointestinal hemorrhage | 0.9 | 1.1 | -0.02 | 0.6 | 0.9 | -0.03 | 0.8 | 0.7 | 0.01 |
| Hyperlipidemia | 25.0 | 28.4 | -0.08 | 27.8 | 27.4 | 0.01 | 25.6 | 26.3 | -0.02 |
| Hypertensive disorder | 51.3 | 61.2 | -0.20 | 60.7 | 57.2 | 0.07 | 53.0 | 52.7 | 0.01 |
| Lesion of liver | 0.7 | 0.7 | 0.00 | 0.4 | 0.6 | -0.03 | 0.7 | 0.7 | 0.00 |
| Obesity | 6.4 | 6.7 | -0.01 | 6.7 | 5.9 | 0.03 | 6.4 | 6.6 | -0.01 |
| Osteoarthritis | 13.4 | 16.0 | -0.07 | 14.1 | 14.7 | -0.02 | 13.6 | 14.3 | -0.02 |
| Pneumonia | 2.6 | 2.1 | 0.03 | 2.7 | 2.3 | 0.03 | 2.6 | 2.2 | 0.02 |
| Psoriasis | 1.1 | 1.3 | -0.01 | 1.1 | 1.2 | -0.01 | 1.1 | 1.2 | -0.01 |
| Renal impairment | 9.7 | 6.6 | 0.12 | 6.6 | 7.5 | -0.03 | 9.2 | 9.6 | -0.01 |
| Rheumatoid arthritis | 1.4 | 1.5 | 0.00 | 1.5 | 1.4 | 0.01 | 1.4 | 1.2 | 0.01 |
| Schizophrenia | 0.1 | 0.2 | -0.03 | 0.2 | 0.2 | 0.00 | 0.1 | 0.2 | -0.02 |
| Ulcerative colitis | 0.3 | 0.3 | 0.00 | <0.2 | 0.3 | -0.03 | 0.3 | 0.6 | -0.05 |
| Urinary tract infectious disease | 6.5 | 6.1 | 0.02 | 6.2 | 6.3 | 0.00 | 6.5 | 6.4 | 0.00 |
| Viral hepatitis C | 0.1 | <0.1 | 0.02 | <0.2 | <0.1 | 0.03 | 0.1 | <0.1 | 0.02 |
| Visual system disorder | 10.5 | 11.3 | -0.02 | 11.7 | 11.1 | 0.02 | 10.7 | 10.6 | 0.00 |
| **Medical history: Cardiovascular disease** | | | | | | | | | |
| Atrial fibrillation | 4.6 | 3.2 | 0.07 | 3.2 | 3.9 | -0.04 | 4.4 | 4.7 | -0.01 |
| Cerebrovascular disease | 6.2 | 6.4 | -0.01 | 6.6 | 5.5 | 0.04 | 6.3 | 6.7 | -0.02 |
| Coronary arteriosclerosis | 5.6 | 4.1 | 0.07 | 4.2 | 4.7 | -0.02 | 5.3 | 5.7 | -0.02 |
| Heart disease | 33.9 | 36.9 | -0.06 | 35.8 | 34.3 | 0.03 | 34.3 | 34.5 | 0.00 |
| Heart failure | 10.2 | 12.0 | -0.06 | 10.3 | 10.5 | -0.01 | 10.3 | 11.1 | -0.03 |
| Ischemic heart disease | 14.4 | 18.6 | -0.11 | 17.0 | 15.2 | 0.05 | 14.8 | 14.4 | 0.01 |
| Peripheral vascular disease | 6.9 | 7.0 | 0.00 | 7.1 | 7.3 | -0.01 | 6.9 | 7.9 | -0.04 |
| Pulmonary embolism | 0.8 | 0.5 | 0.04 | 0.9 | 0.5 | 0.05 | 0.8 | 0.8 | 0.01 |
| Venous thrombosis | 1.9 | 1.5 | 0.03 | 1.6 | 1.3 | 0.03 | 1.8 | 1.4 | 0.03 |
| **Medical history: Neoplasms** | | | | | | | | | |
| Hematologic neoplasm | 0.5 | 0.4 | 0.02 | 0.6 | 0.5 | 0.01 | 0.5 | 0.4 | 0.02 |
| Malignant lymphoma | 0.4 | 0.4 | 0.00 | 0.3 | 0.4 | -0.02 | 0.4 | 0.3 | 0.01 |
| Malignant neoplasm of anorectum | 0.3 | 0.3 | 0.00 | 0.3 | 0.1 | 0.04 | 0.3 | 0.1 | 0.04 |
| Malignant neoplastic disease | 7.2 | 7.9 | -0.03 | 7.1 | 7.8 | -0.03 | 7.2 | 7.2 | 0.00 |
| Malignant tumor of breast | 0.8 | 1.2 | -0.05 | 1.0 | 1.2 | -0.02 | 0.8 | 0.9 | -0.01 |
| Malignant tumor of colon | 0.6 | 0.7 | -0.01 | 0.6 | 0.5 | 0.01 | 0.6 | 0.6 | 0.00 |
| Malignant tumor of lung | 0.2 | 0.2 | -0.01 | 0.3 | 0.2 | 0.01 | 0.2 | 0.3 | -0.03 |
| Malignant tumor of urinary bladder | 0.3 | 0.3 | 0.01 | 0.5 | 0.2 | 0.04 | 0.3 | 0.2 | 0.03 |
| Primary malignant neoplasm of prostate | 1.2 | 1.5 | -0.03 | 0.9 | 1.6 | -0.07 | 1.2 | 1.8 | -0.04 |
| **Medication use** | | | | | | | | | |
| Agents acting on the renin-angiotensin system | 74.8 | 72.4 | 0.05 | 75.3 | 73.9 | 0.03 | 74.6 | 75.6 | -0.02 |
| Antibacterials for systemic use | 25.0 | 26.5 | -0.04 | 26.2 | 26.4 | 0.00 | 25.3 | 25.1 | 0.00 |
| Antidepressants | 11.1 | 10.4 | 0.02 | 11.0 | 10.7 | 0.01 | 11.0 | 10.8 | 0.01 |
| Antiepileptics | 6.1 | 4.2 | 0.09 | 5.8 | 5.9 | 0.00 | 5.8 | 5.9 | 0.00 |
| Antiinflammatory and antirheumatic products | 44.8 | 48.5 | -0.07 | 44.6 | 46.6 | -0.04 | 45.3 | 46.1 | -0.02 |
| Antineoplastic agents | 1.4 | 1.0 | 0.04 | 1.1 | 1.1 | 0.00 | 1.3 | 1.4 | 0.00 |

| (Continued) | Before ajustment | | | After matching | | | After stratification | | |
|---|---|---|---|---|---|---|---|---|---|
| Characteristic | T (%) | C (%) | StdDiff | T (%) | C (%) | StdDiff | T (%) | C (%) | StdDiff |
| Antipsoriatics | 0.4 | 0.3 | 0.02 | 0.2 | 0.2 | -0.01 | 0.4 | 0.2 | 0.03 |
| Antithrombotic agents | 42.5 | 38.9 | 0.07 | 38.0 | 38.3 | -0.01 | 41.7 | 42.2 | -0.01 |
| Beta blocking agents | 56.5 | 53.3 | 0.06 | 55.4 | 55.0 | 0.01 | 55.4 | 55.0 | 0.01 |
| Calcium channel blockers | 38.6 | 36.4 | 0.05 | 38.4 | 37.5 | 0.02 | 38.2 | 39.3 | -0.02 |
| Diuretics | 60.0 | 62.0 | -0.04 | 63.3 | 61.4 | 0.04 | 60.6 | 60.1 | 0.01 |
| Drugs for acid related disorders | 39.7 | 34.4 | 0.11 | 36.4 | 36.4 | 0.00 | 39.0 | 38.6 | 0.01 |
| Drugs for obstructive airway diseases | 17.8 | 16.5 | 0.04 | 17.1 | 16.7 | 0.01 | 17.5 | 16.9 | 0.02 |
| Immunosuppressants | 0.8 | 0.5 | 0.03 | 0.5 | 0.5 | -0.01 | 0.7 | 0.4 | 0.04 |
| Lipid modifying agents | 51.6 | 43.9 | 0.15 | 46.2 | 46.8 | -0.01 | 50.5 | 51.8 | -0.03 |
| Opioids | 13.3 | 13.7 | -0.01 | 12.9 | 13.2 | -0.01 | 13.1 | 13.3 | 0.00 |
| Psycholeptics | 11.8 | 13.1 | -0.04 | 12.1 | 12.4 | -0.01 | 12.0 | 11.9 | 0.00 |
| Psychostimulants | 0.3 | 0.7 | -0.05 | 0.4 | 0.3 | 0.01 | 0.3 | 0.2 | 0.03 |

##### 2.6.9 SIDIAP

**Supplementary Table 36:** Baseline patient characteristics for DPP4I (T) and SU (C) new-users in the SIDIAP data source. We report proportion of initiators satisfying selected base-line characteristics and the standardized difference of population proportions (StdDiff) before and after propensity score adjustment. Less extreme StdDiffs through matching and stratification suggest improved balance between patient cohorts through adjustment.

| | Before adjustment | | | After matching | | | After stratification | | |
|---|---|---|---|---|---|---|---|---|---|
| Characteristic | T (%) | C (%) | StdDiff | T (%) | C (%) | StdDiff | T (%) | C (%) | StdDiff |
| **Age group** | | | | | | | | | |
| 65 - 69 | 26.3 | 31.6 | -0.12 | 31.6 | 32.2 | -0.01 | 27.8 | 30.8 | -0.06 |
| 70 - 74 | 25.6 | 27.2 | -0.04 | 27.5 | 26.6 | 0.02 | 26.1 | 28.0 | -0.04 |
| 75 - 79 | 20.8 | 20.8 | 0.00 | 20.0 | 20.3 | -0.01 | 20.7 | 19.6 | 0.03 |
| 80 - 84 | 15.1 | 12.7 | 0.07 | 12.5 | 12.9 | -0.01 | 14.3 | 13.1 | 0.04 |
| 85 - 89 | 8.9 | 5.9 | 0.11 | 6.0 | 6.2 | -0.01 | 8.1 | 6.5 | 0.06 |
| 90 - 94 | 2.8 | 1.5 | 0.09 | 2.0 | 1.7 | 0.02 | 2.5 | 1.8 | 0.05 |
| 95 - 99 | 0.5 | 0.2 | 0.05 | 0.3 | 0.2 | 0.02 | 0.4 | 0.3 | 0.03 |
| Gender: female | 47.1 | 49.9 | -0.06 | 49.0 | 49.0 | 0.00 | 47.7 | 47.2 | 0.01 |
| **Medical history: General** | | | | | | | | | |
| Acute respiratory disease | 13.4 | 10.4 | 0.09 | 11.2 | 11.3 | 0.00 | 12.9 | 12.0 | 0.03 |
| Chronic liver disease | 0.4 | 0.4 | 0.00 | 0.5 | 0.3 | 0.03 | 0.4 | 0.3 | 0.03 |
| Chronic obstructive lung disease | 1.8 | 2.0 | -0.01 | 2.1 | 1.9 | 0.02 | 1.8 | 1.7 | 0.01 |
| Dementia | 1.1 | 1.3 | -0.03 | 1.1 | 1.3 | -0.02 | 1.1 | 1.2 | -0.01 |
| Depressive disorder | 2.4 | 3.4 | -0.06 | 3.4 | 3.5 | 0.00 | 2.6 | 2.9 | -0.01 |
| Gastroesophageal reflux disease | 1.3 | 0.7 | 0.06 | 0.8 | 0.9 | -0.01 | 1.2 | 1.2 | -0.01 |
| Gastrointestinal hemorrhage | 1.8 | 1.3 | 0.04 | 1.6 | 1.5 | 0.01 | 1.7 | 1.2 | 0.05 |
| Hyperlipidemia | 2.7 | 3.6 | -0.05 | 3.6 | 3.6 | 0.00 | 3.0 | 3.1 | 0.00 |
| Hypertensive disorder | 3.6 | 6.5 | -0.14 | 5.3 | 5.3 | 0.00 | 4.2 | 4.2 | 0.00 |
| Lesion of liver | 0.3 | 0.4 | -0.01 | 0.3 | 0.4 | -0.02 | 0.4 | 0.6 | -0.04 |
| Obesity | 6.7 | 8.1 | -0.05 | 7.7 | 7.9 | -0.01 | 7.0 | 7.5 | -0.02 |
| Osteoarthritis | 5.9 | 6.9 | -0.04 | 7.3 | 6.8 | 0.02 | 6.1 | 5.8 | 0.01 |
| Pneumonia | 1.8 | 1.7 | 0.01 | 1.4 | 1.7 | -0.02 | 1.7 | 1.6 | 0.01 |
| Psoriasis | 0.3 | 0.6 | -0.04 | 0.4 | 0.6 | -0.03 | 0.3 | 0.7 | -0.05 |
| Renal impairment | 6.1 | 2.9 | 0.16 | 3.3 | 3.5 | -0.01 | 5.3 | 4.4 | 0.04 |
| Rheumatoid arthritis | 0.1 | 0.1 | -0.01 | 0.2 | 0.1 | 0.01 | 0.1 | 0.1 | 0.02 |
| Urinary tract infectious disease | 8.3 | 4.4 | 0.16 | 5.3 | 5.6 | -0.01 | 7.5 | 7.2 | 0.01 |
| Viral hepatitis C | 0.2 | 0.1 | 0.02 | 0.3 | 0.1 | 0.04 | 0.2 | 0.1 | 0.04 |
| Visual system disorder | 18.6 | 18.4 | 0.00 | 19.0 | 19.4 | -0.01 | 18.7 | 18.0 | 0.02 |
| **Medical history: Cardiovascular disease** | | | | | | | | | |
| Atrial fibrillation | 2.5 | 2.1 | 0.03 | 2.1 | 2.0 | 0.00 | 2.4 | 1.9 | 0.03 |
| Cerebrovascular disease | 1.7 | 1.8 | -0.01 | 1.7 | 1.9 | -0.01 | 1.7 | 1.7 | 0.00 |
| Heart disease | 11.5 | 11.2 | 0.01 | 11.1 | 11.5 | -0.01 | 11.3 | 10.8 | 0.01 |
| Heart failure | 2.8 | 2.7 | 0.00 | 2.6 | 2.9 | -0.02 | 2.7 | 2.3 | 0.03 |
| Ischemic heart disease | 2.8 | 3.4 | -0.03 | 3.1 | 3.3 | -0.01 | 2.8 | 2.9 | 0.00 |
| Peripheral vascular disease | 1.1 | 1.2 | -0.01 | 0.9 | 1.1 | -0.01 | 1.1 | 1.1 | 0.00 |
| Pulmonary embolism | 0.3 | 0.2 | 0.02 | 0.2 | 0.3 | -0.01 | 0.3 | 0.4 | -0.02 |
| Venous thrombosis | 1.1 | 1.2 | 0.00 | 1.4 | 1.2 | 0.01 | 1.2 | 1.0 | 0.01 |
| **Medical history: Neoplasms** | | | | | | | | | |
| Hematologic neoplasm | 0.1 | 0.1 | 0.00 | <0.1 | 0.1 | -0.02 | 0.1 | 0.1 | 0.02 |
| Malignant lymphoma | 0.1 | 0.1 | -0.01 | <0.1 | 0.1 | -0.01 | <0.1 | 0.1 | -0.01 |
| Malignant neoplasm of anorectum | 0.1 | 0.1 | -0.01 | 0.1 | 0.1 | 0.00 | 0.1 | 0.1 | 0.00 |
| Malignant neoplastic disease | 3.9 | 3.8 | 0.00 | 3.9 | 3.8 | 0.00 | 3.9 | 4.1 | -0.01 |
| Malignant tumor of breast | 0.3 | 0.5 | -0.03 | 0.3 | 0.4 | -0.01 | 0.3 | 0.3 | 0.00 |
| Malignant tumor of colon | 0.4 | 0.5 | 0.00 | 0.6 | 0.5 | 0.02 | 0.5 | 0.4 | 0.02 |
| Malignant tumor of lung | 0.1 | <0.1 | 0.01 | <0.1 | 0.1 | -0.01 | 0.1 | 0.1 | 0.00 |
| Malignant tumor of urinary bladder | 0.3 | 0.3 | 0.01 | 0.3 | 0.3 | 0.01 | 0.3 | 0.4 | -0.01 |
| Primary malignant neoplasm of prostate | 0.5 | 0.4 | 0.00 | 0.5 | 0.4 | 0.01 | 0.5 | 0.6 | -0.02 |
| **Medication use** | | | | | | | | | |
| Agents acting on the renin-angiotensin system | 69.7 | 63.2 | 0.14 | 65.4 | 65.8 | -0.01 | 68.3 | 66.1 | 0.05 |
| Antibacterials for systemic use | 41.6 | 33.4 | 0.17 | 40.1 | 40.1 | 0.00 | 40.1 | 40.1 | 0.00 |
| Antidepressants | 23.2 | 20.8 | 0.06 | 21.6 | 22.1 | -0.01 | 22.9 | 22.7 | 0.01 |
| Antiepileptics | 10.4 | 7.3 | 0.11 | 8.6 | 8.4 | 0.01 | 9.9 | 10.4 | -0.02 |
| Antiinflammatory and antirheumatic products | 53.2 | 56.9 | -0.07 | 58.8 | 57.9 | 0.02 | 54.3 | 53.5 | 0.02 |
| Antineoplastic agents | 2.3 | 1.8 | 0.04 | 2.4 | 2.1 | 0.01 | 2.3 | 2.7 | -0.02 |
| Antipsoriatics | 1.8 | 1.1 | 0.05 | 1.4 | 1.2 | 0.02 | 1.7 | 1.0 | 0.06 |
| Antithrombotic agents | 46.0 | 41.5 | 0.09 | 41.8 | 43.1 | -0.03 | 44.8 | 43.3 | 0.03 |
| Beta blocking agents | 32.6 | 26.6 | 0.13 | 27.4 | 28.5 | -0.02 | 31.3 | 31.0 | 0.01 |
| Calcium channel blockers | 30.8 | 24.4 | 0.14 | 26.6 | 26.4 | 0.01 | 29.2 | 28.4 | 0.02 |

| (Continued) | Before ajustment | | | After matching | | | After stratification | | |
|---|---|---|---|---|---|---|---|---|---|
| Characteristic | T (%) | C (%) | StdDiff | T (%) | C (%) | StdDiff | T (%) | C (%) | StdDiff |
| Diuretics | 56.6 | 52.4 | 0.08 | 53.3 | 54.1 | -0.01 | 55.7 | 53.8 | 0.04 |
| Drugs for acid related disorders | 62.8 | 59.4 | 0.07 | 61.4 | 62.0 | -0.01 | 62.2 | 60.9 | 0.03 |
| Drugs for obstructive airway diseases | 30.7 | 25.9 | 0.11 | 26.8 | 27.4 | -0.01 | 29.7 | 28.0 | 0.04 |
| Immunosuppressants | 1.4 | 0.8 | 0.06 | 0.8 | 0.9 | -0.01 | 1.3 | 1.2 | 0.00 |
| Lipid modifying agents | 66.7 | 59.3 | 0.15 | 63.4 | 63.7 | -0.01 | 65.4 | 64.8 | 0.01 |
| Opioids | 20.8 | 17.0 | 0.10 | 18.2 | 18.3 | 0.00 | 20.0 | 20.6 | -0.01 |
| Psycholeptics | 35.3 | 33.3 | 0.04 | 33.7 | 34.6 | -0.02 | 35.0 | 35.6 | -0.01 |
| Psychostimulants | 4.4 | 5.0 | -0.03 | 4.6 | 4.0 | 0.03 | 4.6 | 4.0 | 0.03 |

### 3 Cohort balance and systematic error diagnostics across data sources

#### 3.1 GLP1RA vs SGLT2I

##### 3.1.1 OptumDOD

**Supplementary Figure 1:** Preference score distributions, propensity score stratified (middle column) and matched (right column) covariate balance and negative control error distributions (bottom row) for GLP1RA vs SGLT2I in the OptumDOD data source.

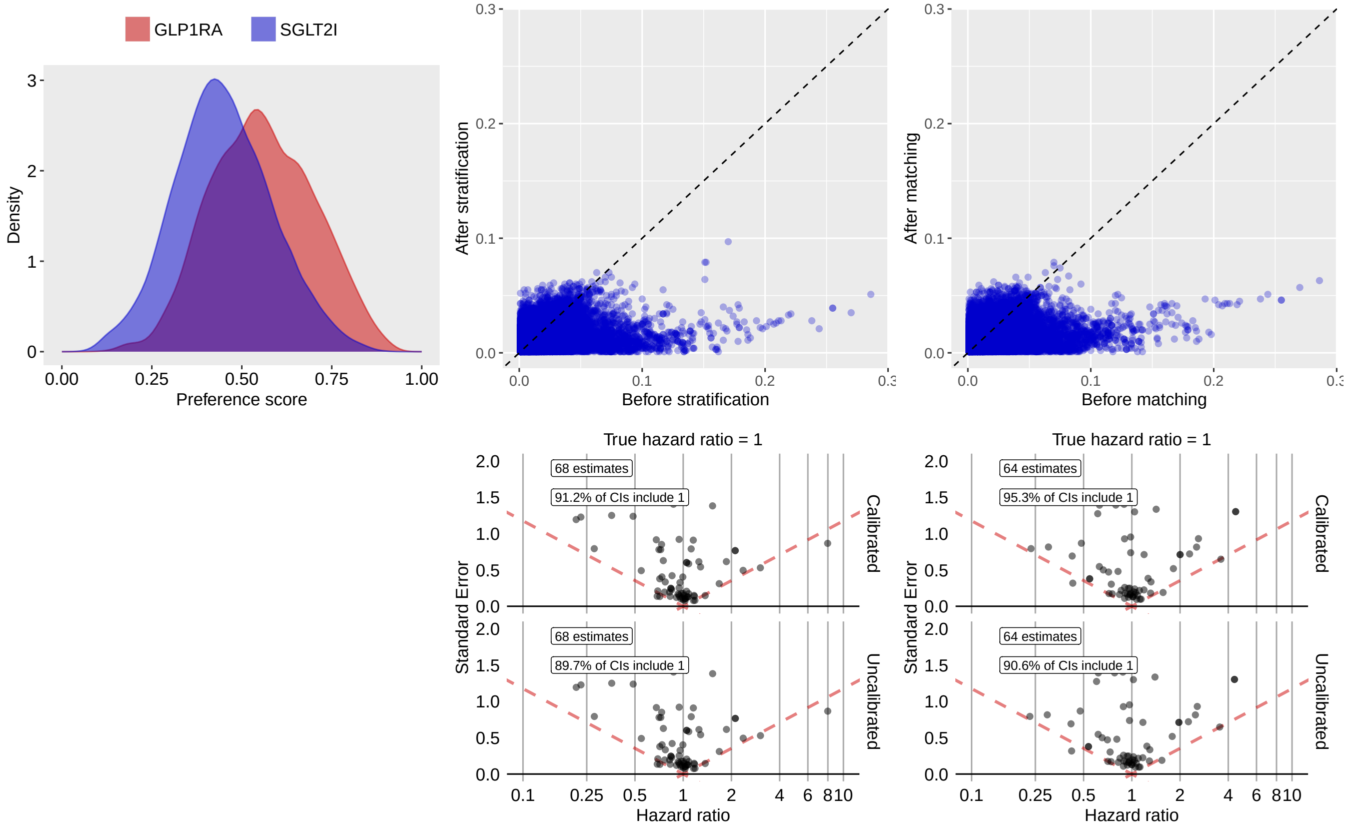

**Supplementary Table 37:** Most extreme patient characteristics balance diagnostics after stratification for GLP1RA vs SGLT2I in the OptumDOD data source.

| Characteristic (total count = 18142) | Before stratification T (%) | Before stratification C (%) | Before stratification SDf | After stratification T (%) | After stratification C (%) | After stratification SDf |
|---|---|---|---|---|---|---|
| drug_era group during day -365 through 0 days relative to index: sacubitril | 0.3 | 2.2 | -0.17 | 0.7 | 1.8 | -0.10 |
| drug_era group during day -30 through 0 days relative to index: sacubitril | 0.3 | 2.0 | -0.15 | 0.7 | 1.5 | -0.08 |
| drug_era group during day 0 through 0 days relative to index: sacubitril | 0.3 | 1.8 | -0.15 | 0.6 | 1.4 | -0.08 |
| condition_era group during day -365 through 0 days relative to index: Myocardial infarction due to demand ischemia | -0.1 | 0.3 | -0.07 | -0.1 | 0.3 | -0.07 |
| condition_era group during day -365 through 0 days relative to index: Polyp of nasal cavity and/or nasal sinus | -0.1 | 0.3 | -0.06 | -0.1 | 0.3 | -0.07 |

| (Continued) | T (%) | C (%) | SDf | T (%) | C (%) | SDf |
|---|---|---|---|---|---|---|
| observation during day -365 through 0 days relative to index: Set-up portable x-ray equipment | 0.8 | 0.3 | 0.07 | 0.7 | 0.3 | 0.07 |
| procedure_occurrence during day -365 through 0 days relative to index: Collection of venous blood by venipuncture or urine sample by catheterization from an individual in a skilled nursing facility (snf) or by a laboratory on behalf of a home health agency (hha) | 0.5 | 0.1 | 0.07 | 0.5 | 0.1 | 0.07 |
| condition_era group during day -30 through 0 days relative to index: Chronic congestive heart failure | 0.4 | 2.1 | -0.15 | 0.9 | 1.7 | -0.06 |
| observation during day -365 through 0 days relative to index: Transportation of portable x-ray equipment and personnel to home or nursing home, per trip to facility or location, more than one patient seen | 0.4 | 0.1 | 0.07 | 0.4 | 0.1 | 0.06 |
| condition_era group during day -30 through 0 days relative to index: Fluid volume disorder | 0.2 | 0.6 | -0.07 | 0.2 | 0.6 | -0.06 |

**Supplementary Table 38:** Most extreme patient characteristics balance diagnostics after matching for GLP1RA vs SGLT2I in the OptumDOD data source.

| | Before matching | | | After matching | | |
|---|---|---|---|---|---|---|
| Characteristic (total count = 17616) | T (%) | C (%) | SDf | T (%) | C (%) | SDf |
| observation during day -365 through 0 days relative to index: Transportation of portable x-ray equipment and personnel to home or nursing home, per trip to facility or location, more than one patient seen | 0.4 | 0.1 | 0.07 | 0.5 | 0.1 | 0.08 |
| procedure_occurrence during day -365 through 0 days relative to index: Collection of venous blood by venipuncture or urine sample by catheterization from an individual in a skilled nursing facility (snf) or by a laboratory on behalf of a home health agency (hha) | 0.5 | 0.1 | 0.07 | 0.6 | 0.1 | 0.08 |
| observation during day -365 through 0 days relative to index: Set-up portable x-ray equipment | 0.8 | 0.3 | 0.07 | 0.8 | 0.3 | 0.07 |
| condition_era group during day -365 through 0 days relative to index: Polyp of nasal cavity and/or nasal sinus | -0.1 | 0.3 | -0.06 | -0.1 | 0.3 | -0.07 |
| measurement during day -30 through 0 days relative to index: Insulin; total | 1.2 | 0.4 | 0.08 | 1.1 | 0.5 | 0.07 |
| drug_era group during day 0 through 0 days relative to index: Aminosalicylic acid and similar agents | 0.1 | 0.4 | -0.05 | -0.1 | 0.4 | -0.07 |
| measurement during day -365 through 0 days relative to index: Rheumatoid factor; qualitative | 0.2 | 0.4 | -0.04 | 0.1 | 0.5 | -0.07 |
| condition_era group during day -365 through 0 days relative to index: Myocardial infarction due to demand ischemia | -0.1 | 0.3 | -0.07 | -0.1 | 0.3 | -0.06 |
| condition_era group during day -365 through 0 days relative to index: Obesity | 39.6 | 26.3 | 0.29 | 35.3 | 38.4 | -0.06 |
| observation during day -365 through 0 days relative to index: History of malignant neoplasm of thyroid | 0.2 | 0.3 | -0.02 | 0.1 | 0.5 | -0.06 |

##### 3.1.2 MDCR

**Supplementary Figure 2:** Preference score distributions, propensity score stratified (middle column) and matched (right column) covariate balance and negative control error distributions (bottom row) for GLP1RA vs SGLT2I in the MDCR data source.

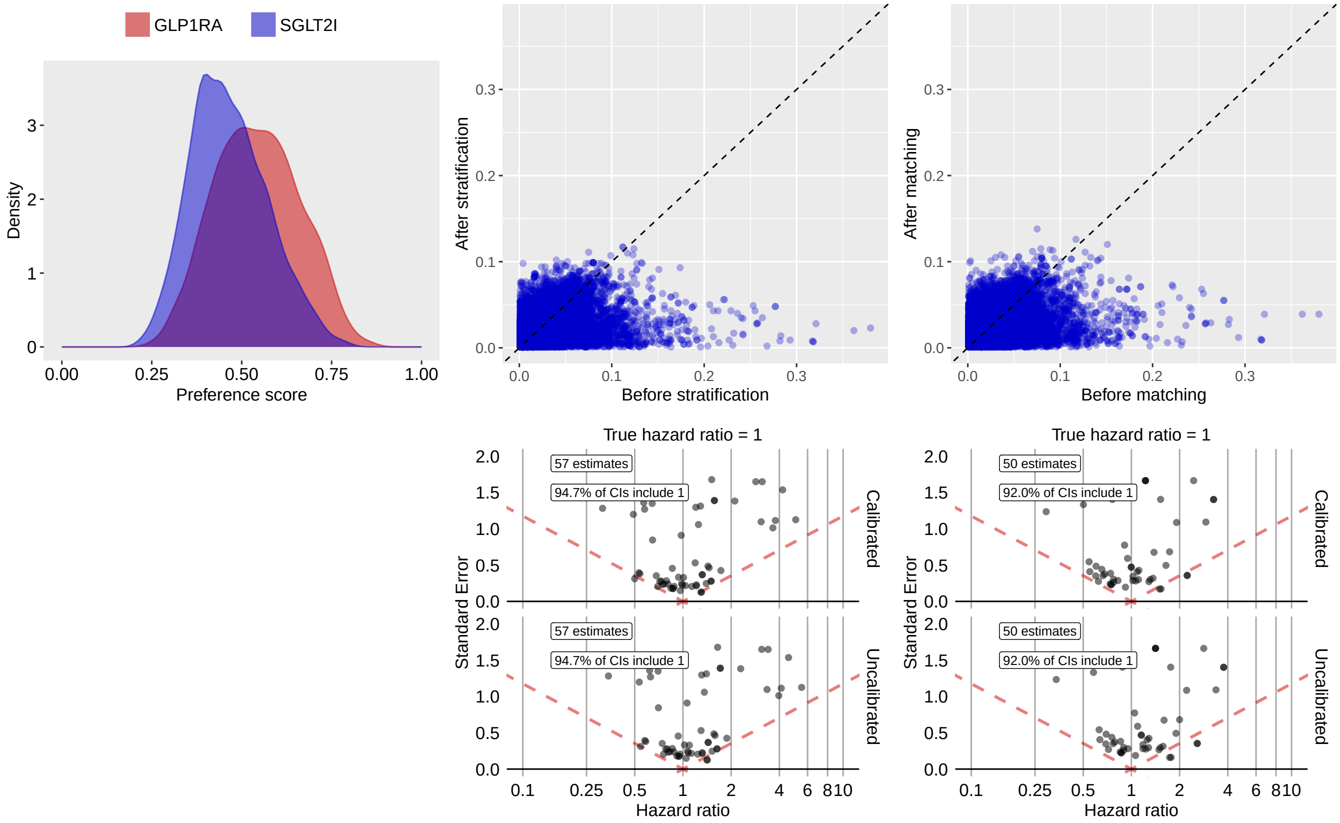

**Supplementary Table 39:** Most extreme patient characteristics balance diagnostics after stratification for GLP1RA vs SGLT2I in the MDCR data source.

| | Before stratification | | | After stratification | | |
|---|---|---|---|---|---|---|
| Characteristic (total count = 15383) | T (%) | C (%) | SDf | T (%) | C (%) | SDf |
| condition_era group during day -30 through 0 days relative to index: Acute myocardial infarction | -0.2 | 1.0 | -0.11 | -0.3 | 0.9 | -0.12 |
| condition_era group during day -30 through 0 days relative to index: Acute ischemic heart disease | -0.2 | 1.0 | -0.11 | -0.3 | 0.9 | -0.12 |
| condition_era group during day -365 through 0 days relative to index: Acute myocardial infarction | 0.8 | 2.3 | -0.12 | 0.8 | 2.1 | -0.12 |
| condition_era group during day -365 through 0 days relative to index: Congenital abnormality of lower limb and pelvic girdle | 0.4 | 1.0 | -0.07 | -0.3 | 1.1 | -0.11 |
| procedure_occurrence during day -365 through 0 days relative to index: Medical nutrition therapy; re-assessment and intervention, individual, face-to-face with the patient, each 15 minutes | 1.0 | 0.1 | 0.12 | 0.9 | -0.1 | 0.11 |
| condition_era group during day -365 through 0 days relative to index: Acute non-ST segment elevation myocardial infarction | 0.2 | 1.3 | -0.12 | 0.3 | 1.2 | -0.11 |
| condition_era group during day -365 through 0 days relative to index: Preinfarction syndrome | 1.4 | 2.8 | -0.10 | 1.2 | 2.7 | -0.11 |

| (Continued) | T (%) | C (%) | SDf | T (%) | C (%) | SDf |
|---|---|---|---|---|---|---|
| condition_era group during day -365 through 0 days relative to index: Anxiety state | 0.9 | 1.4 | -0.04 | 0.5 | 1.5 | -0.10 |
| observation during day -365 through 0 days relative to index: Most recent hemoglobin A1c level greater than 9.0% (DM) | 1.7 | 1.1 | 0.05 | 2.4 | 1.1 | 0.10 |
| observation during day -365 through 0 days relative to index: Bmi is documented within normal parameters and no follow-up plan is required | 0.6 | 2.1 | -0.13 | 0.9 | 2.1 | -0.10 |

**Supplementary Table 40:** Most extreme patient characteristics balance diagnostics after matching for GLP1RA vs SGLT2I in the MDCR data source.

| | Before matching | | | After matching | | |
|---|---|---|---|---|---|---|
| Characteristic (total count = 14779) | T (%) | C (%) | SDf | T (%) | C (%) | SDf |
| condition_era group during day -365 through 0 days relative to index: Congenital abnormality of lower limb and pelvic girdle | 0.4 | 1.0 | -0.07 | -0.4 | 1.3 | -0.14 |
| procedure_occurrence during day -365 through 0 days relative to index: Medical nutrition therapy; re-assessment and intervention, individual, face-to-face with the patient, each 15 minutes | 1.0 | 0.1 | 0.12 | 1.1 | -0.1 | 0.13 |
| condition_era group during day -365 through 0 days relative to index: Chronic systolic heart failure | 1.5 | 4.0 | -0.15 | 2.0 | 4.1 | -0.12 |
| condition_era group during day -30 through 0 days relative to index: Functional finding of gastrointestinal tract | 1.9 | 3.2 | -0.08 | 1.9 | 3.8 | -0.12 |
| condition_era group during day -365 through 0 days relative to index: Congenital anomaly of abdomen | -0.2 | 0.6 | -0.09 | -0.4 | 0.8 | -0.11 |
| condition_era group during day -365 through 0 days relative to index: Prediabetes | 3.0 | 1.6 | 0.09 | 3.9 | 2.0 | 0.11 |
| condition_era group during day -30 through 0 days relative to index: Altered bowel function | 1.7 | 2.7 | -0.07 | 1.6 | 3.3 | -0.11 |
| condition_era group during day -365 through 0 days relative to index: Hypertensive heart failure | 1.8 | 4.0 | -0.13 | 2.3 | 4.3 | -0.11 |
| drug_era group during day -30 through 0 days relative to index: nortriptyline | 0.3 | 0.6 | -0.04 | -0.4 | 0.9 | -0.11 |
| condition_era group during day -365 through 0 days relative to index: Congenital anomaly of digestive organ | -0.2 | 0.5 | -0.08 | -0.4 | 0.8 | -0.11 |

##### 3.1.3 Open Claims

**Supplementary Figure 3:** Preference score distributions, propensity score stratified (middle column) and matched (right column) covariate balance and negative control error distributions (bottom row) for GLP1RA vs SGLT2I in the Open Claims data source.

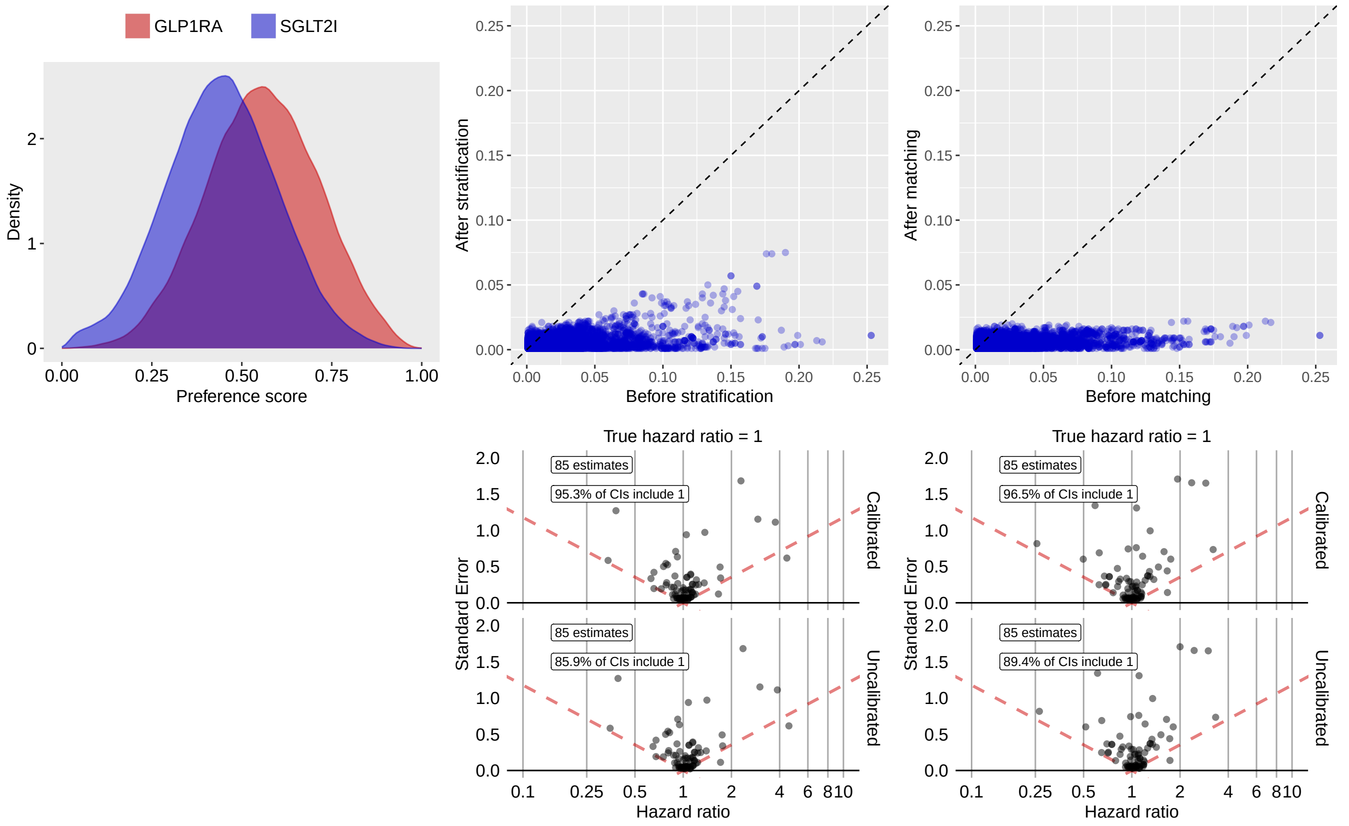

**Supplementary Table 41:** Most extreme patient characteristics balance diagnostics after stratification for GLP1RA vs SGLT2I in the Open Claims data source.

| | Before stratification | | | After stratification | | |
|---|---|---|---|---|---|---|
| Characteristic (total count = 10571) | T (%) | C (%) | SDf | T (%) | C (%) | SDf |
| drug_era group during day -365 through 0 days relative to index: sacubitril | 0.5 | 2.9 | -0.19 | 1.2 | 2.2 | -0.07 |
| drug_era group during day -30 through 0 days relative to index: sacubitril | 0.4 | 2.5 | -0.18 | 1.0 | 1.9 | -0.07 |
| drug_era group during day 0 through 0 days relative to index: sacubitril | 0.4 | 2.4 | -0.18 | 1.0 | 1.8 | -0.07 |
| condition_era group during day -30 through 0 days relative to index: Systolic dysfunction | 0.4 | 2.1 | -0.15 | 1.0 | 1.6 | -0.06 |
| condition_era group during day -30 through 0 days relative to index: Systolic heart failure | 0.4 | 2.1 | -0.15 | 1.0 | 1.6 | -0.06 |
| condition_era group during day -30 through 0 days relative to index: Chronic systolic heart failure | 0.3 | 1.6 | -0.13 | 0.8 | 1.3 | -0.05 |
| condition_era group during day -365 through 0 days relative to index: Systolic heart failure | 1.3 | 4.0 | -0.17 | 2.4 | 3.2 | -0.05 |
| condition_era group during day -365 through 0 days relative to index: Systolic dysfunction | 1.3 | 4.1 | -0.17 | 2.5 | 3.3 | -0.05 |

| (Continued) | T (%) | C (%) | SDf | T (%) | C (%) | SDf |
|---|---|---|---|---|---|---|
| condition_era group during day -30 through 0 days relative to index: Abnormal cardiovascular function | 1.0 | 3.0 | -0.14 | 1.7 | 2.4 | -0.05 |
| condition_era group during day -365 through 0 days relative to index: Chronic systolic heart failure | 1.0 | 3.3 | -0.15 | 2.0 | 2.6 | -0.04 |

**Supplementary Table 42:** Most extreme patient characteristics balance diagnostics after matching for GLP1RA vs SGLT2I in the Open Claims data source.

| | Before matching | | | After matching | | |
|---|---|---|---|---|---|---|
| Characteristic (total count = 10630) | T (%) | C (%) | SDf | T (%) | C (%) | SDf |
| drug_era group during day -365 through 0 days relative to index: PSYCHOANALEPTICS | 38.6 | 28.6 | 0.21 | 35.8 | 36.9 | -0.02 |
| drug_era group during day 0 through 0 days relative to index: NERVOUS SYSTEM | 47.3 | 39.6 | 0.16 | 45.1 | 46.2 | -0.02 |
| drug_era group during day -30 through 0 days relative to index: NERVOUS SYSTEM | 51.3 | 43.7 | 0.15 | 49.1 | 50.2 | -0.02 |
| drug_era group during day -365 through 0 days relative to index: ANTIDEPRESSANTS | 37.3 | 27.2 | 0.22 | 34.5 | 35.6 | -0.02 |
| drug_era group during day -365 through 0 days relative to index: NERVOUS SYSTEM | 69.1 | 62.2 | 0.14 | 67.1 | 68.1 | -0.02 |
| observation during day -365 through 0 days relative to index: Documentation that patient is a current non-tobacco user | 0.1 | 0.2 | -0.03 | 0.1 | 0.1 | -0.02 |
| drug_era group during day -30 through 0 days relative to index: ANTIDEPRESSANTS | 31.1 | 22.2 | 0.20 | 28.7 | 29.5 | -0.02 |
| drug_era group during day -365 through 0 days relative to index: Rauwolfia alkaloids and diuretics in combination | 62.6 | 59.3 | 0.07 | 61.3 | 62.2 | -0.02 |
| drug_era group during day -365 through 0 days relative to index: OTHER OTOLOGICALS | 41.8 | 39.2 | 0.05 | 40.3 | 41.2 | -0.02 |
| Charlson index - Romano adaptation | 422.6 | 428.9 | -0.02 | 419.6 | 424.6 | -0.02 |

##### 3.1.4 OptumEHR

**Supplementary Figure 4:** Preference score distributions, propensity score stratified (middle column) and matched (right column) covariate balance and negative control error distributions (bottom row) for GLP1RA vs SGLT2I in the OptumEHR data source.

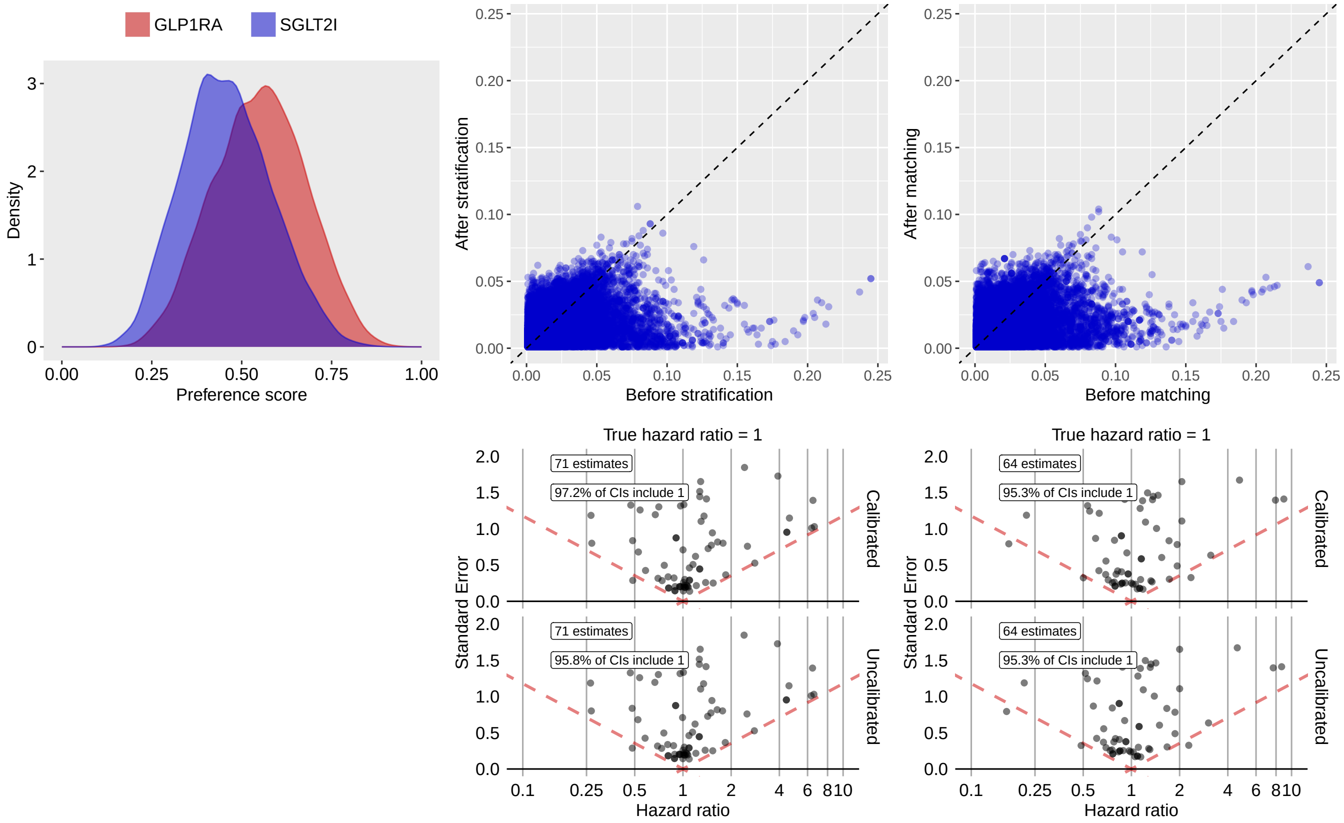

**Supplementary Table 43:** Most extreme patient characteristics balance diagnostics after stratification for GLP1RA vs SGLT2I in the OptumEHR data source.

| Characteristic (total count = 18674) | Before stratification | | | After stratification | | |
|---|---|---|---|---|---|---|
| | T (%) | C (%) | SDf | T (%) | C (%) | SDf |
| observation during day -365 through 0 days relative to index: Intensive Care - Psychiatric | 0.5 | 0.1 | 0.08 | 0.7 | -0.1 | 0.11 |
| condition_era group during day -365 through 0 days relative to index: Neoplasm of intrathoracic organs | -0.1 | 0.5 | -0.09 | -0.1 | 0.5 | -0.09 |
| condition_era group during day -365 through 0 days relative to index: Neoplasm of lower respiratory tract | -0.1 | 0.5 | -0.09 | -0.1 | 0.5 | -0.09 |
| condition_era group during day -365 through 0 days relative to index: Neoplasm of lung | -0.1 | 0.4 | -0.08 | -0.1 | 0.5 | -0.09 |
| drug_era group during day -365 through 0 days relative to index: sacubitril | 0.5 | 1.4 | -0.10 | 0.5 | 1.4 | -0.09 |
| condition_era group during day -365 through 0 days relative to index: Malignant neoplasm of lower respiratory tract | -0.1 | 0.4 | -0.08 | -0.1 | 0.4 | -0.08 |
| drug_era group during day 0 through 0 days relative to index: lovastatin | 1.7 | 2.4 | -0.05 | 1.2 | 2.3 | -0.08 |
| observation during day -365 through 0 days relative to index: Other Imaging Services - Positron Emission Tomography | -0.1 | 0.4 | -0.07 | -0.1 | 0.4 | -0.08 |

| (Continued) | T (%) | C (%) | SDf | T (%) | C (%) | SDf |
|---|---|---|---|---|---|---|
| condition_era group during day -365 through 0 days relative to index: Malignant tumor of lung | -0.1 | 0.4 | -0.07 | -0.1 | 0.4 | -0.08 |
| condition_era group during day -365 through 0 days relative to index: Imaging of thorax abnormal | 0.3 | 0.8 | -0.06 | 0.2 | 0.8 | -0.08 |

**Supplementary Table 44:** Most extreme patient characteristics balance diagnostics after matching for GLP1RA vs SGLT2I in the OptumEHR data source.

| | Before matching | | | After matching | | |
|---|---|---|---|---|---|---|
| Characteristic (total count = 17990) | T (%) | C (%) | SDf | T (%) | C (%) | SDf |
| condition_era group during day -365 through 0 days relative to index: Neoplasm of intrathoracic organs | -0.1 | 0.5 | -0.09 | -0.2 | 0.6 | -0.10 |
| condition_era group during day -365 through 0 days relative to index: Neoplasm of lower respiratory tract | -0.1 | 0.5 | -0.09 | -0.2 | 0.6 | -0.10 |
| condition_era group during day -365 through 0 days relative to index: Neoplasm of lung | -0.1 | 0.4 | -0.08 | -0.2 | 0.6 | -0.10 |
| condition_era group during day -365 through 0 days relative to index: Malignant neoplasm of lower respiratory tract | -0.1 | 0.4 | -0.08 | -0.2 | 0.5 | -0.09 |
| condition_era group during day -365 through 0 days relative to index: Malignant tumor of lung | -0.1 | 0.4 | -0.07 | -0.2 | 0.4 | -0.09 |
| drug_era group during day -365 through 0 days relative to index: sacubitril | 0.5 | 1.4 | -0.10 | 0.5 | 1.3 | -0.08 |
| condition_era group during day -365 through 0 days relative to index: Imaging of thorax abnormal | 0.3 | 0.8 | -0.06 | 0.2 | 0.8 | -0.08 |
| condition_era group during day -30 through 0 days relative to index: Metabolic syndrome X | 1.7 | 0.6 | 0.10 | 1.4 | 0.6 | 0.08 |
| observation during day -365 through 0 days relative to index: Intensive Care - Psychiatric | 0.5 | 0.1 | 0.08 | 0.5 | 0.1 | 0.08 |
| drug_era group during day -365 through 0 days relative to index: trospium | 0.4 | 0.1 | 0.08 | 0.5 | 0.1 | 0.08 |

#### 3.2 GLP1RA vs DPP4I

##### 3.2.1 OptumDOD

**Supplementary Figure 5:** Preference score distributions, propensity score stratified (middle column) and matched (right column) covariate balance and negative control error distributions (bottom row) for GLP1RA vs DPP4I in the OptumDOD data source.

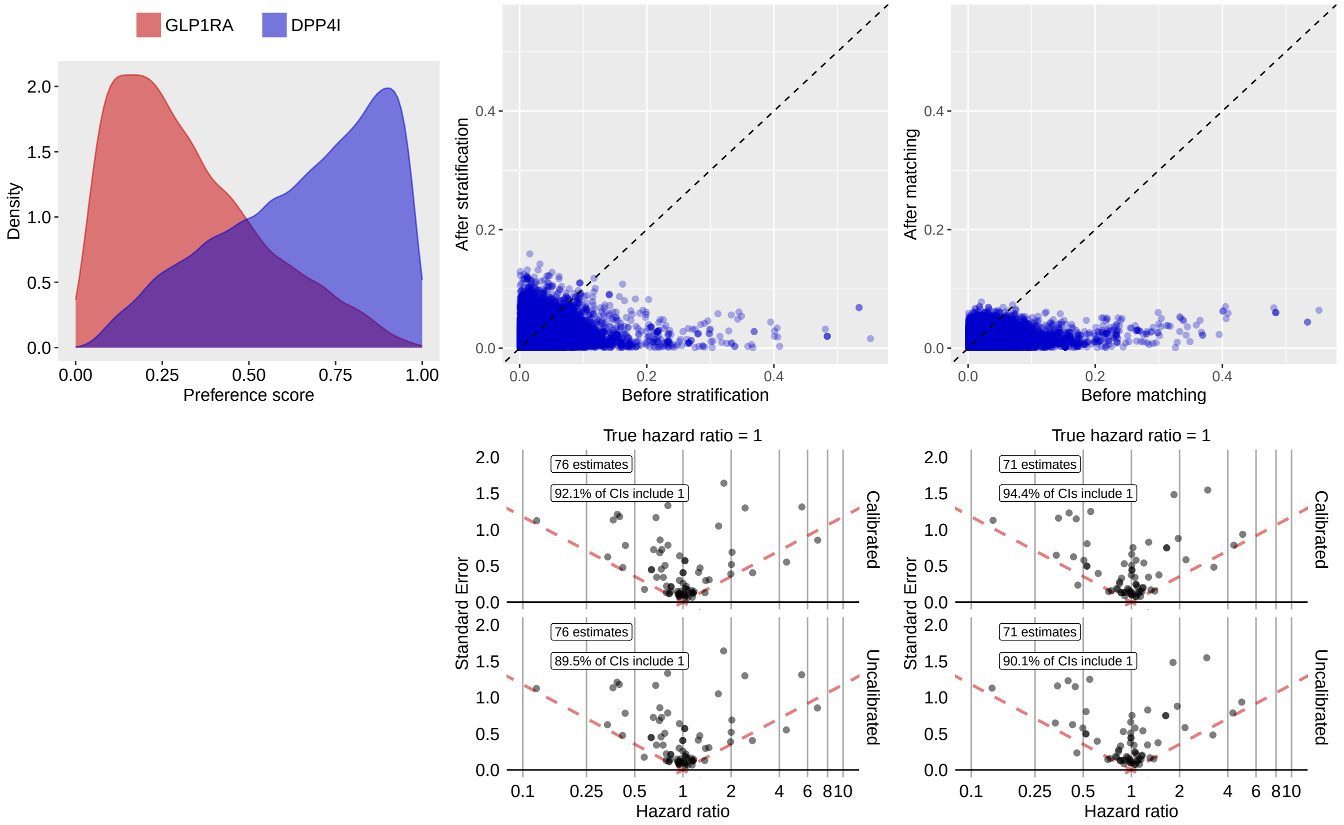

**Supplementary Table 45:** Most extreme patient characteristics balance diagnostics after stratification for GLP1RA vs DPP4I in the OptumDOD data source.

| | Before stratification | | | After stratification | | |
|---|---|---|---|---|---|---|
| Characteristic (total count = 18715) | T (%) | C (%) | SDf | T (%) | C (%) | SDf |
| measurement during day -365 through 0 days relative to index: Eye disorder screening | 0.1 | 0.2 | 0.02 | 1.7 | 0.2 | -0.16 |
| measurement during day -30 through 0 days relative to index: Lipase | 0.9 | 1.2 | 0.03 | 3.3 | 1.2 | -0.14 |
| procedure_occurrence during day -30 through 0 days relative to index: Transitional Care Management Services with the following required elements: Communication (direct contact, telephone, electronic) with the patient and/or caregiver within 2 business days of discharge Medical decision making of at least moderate complexity | 0.4 | 0.2 | -0.03 | 1.4 | 0.2 | -0.13 |
| condition_era group during day -30 through 0 days relative to index: Urinary tract obstruction | 2.1 | 2.3 | 0.01 | 4.6 | 2.2 | -0.13 |
| procedure_occurrence during day -365 through 0 days relative to index: Closed treatment of distal radial fracture (eg, Colles or Smith type) or epiphyseal separation, includes closed treatment of fracture of ulnar styloid, when performed; without manipulation | 0.1 | 0.1 | 0.00 | 1.1 | 0.1 | -0.13 |

| (Continued) | T (%) | C (%) | SDf | T (%) | C (%) | SDf |
|---|---|---|---|---|---|---|
| condition_era group during day -30 through 0 days relative to index: Hypertensive heart and renal disease with (congestive) heart failure | 0.5 | 0.9 | 0.05 | 2.5 | 0.9 | -0.13 |
| condition_era group during day -365 through 0 days relative to index: Tachypnea | 0.1 | 0.1 | 0.01 | 1.1 | 0.1 | -0.12 |
| condition_era group during day -365 through 0 days relative to index: Disorder due to and following fracture of upper limb | 0.1 | 0.1 | -0.00 | 1.0 | 0.1 | -0.12 |
| condition_era group during day -365 through 0 days relative to index: Acute pain | 0.4 | 0.7 | 0.05 | 2.0 | 0.6 | -0.12 |
| observation during day -30 through 0 days relative to index: HIPPS | 1.1 | 1.7 | 0.04 | 3.5 | 1.6 | -0.12 |

**Supplementary Table 46:** Most extreme patient characteristics balance diagnostics after matching for GLP1RA vs DPP4I in the OptumDOD data source.

| | Before matching | | | After matching | | |
|---|---|---|---|---|---|---|
| Characteristic (total count = 18042) | T (%) | C (%) | SDf | T (%) | C (%) | SDf |
| procedure_occurrence during day -365 through 0 days relative to index: Collection of venous blood by venipuncture or urine sample by catheterization from an individual in a skilled nursing facility (snf) or by a laboratory on behalf of a home health agency (hha) | 0.5 | 0.4 | -0.02 | 0.6 | 0.1 | -0.08 |
| age group: 90 - 94 | 0.1 | 0.5 | 0.07 | 0.1 | 0.5 | 0.07 |
| index year: 2020 | 28.8 | 12.7 | -0.41 | 29.3 | 26.2 | -0.07 |
| condition_era group during day -365 through 0 days relative to index: Shoulder synovitis | -0.1 | 0.1 | 0.02 | 0.1 | 0.4 | 0.07 |
| condition_era group during day -365 through 0 days relative to index: Synovitis/tenosynovitis - shoulder | -0.1 | 0.1 | 0.02 | 0.1 | 0.4 | 0.07 |
| condition_era group during day -365 through 0 days relative to index: Infectious disease of mucous membrane | 0.5 | 0.2 | -0.06 | 0.5 | 0.1 | -0.07 |
| measurement during day -365 through 0 days relative to index: Helicobacter pylori [Presence] in Stomach by urea breath test | 0.2 | 0.4 | 0.03 | 0.2 | 0.7 | 0.07 |
| measurement within normal range during day -365 through 0 days relative to index: Helicobacter pylori [Presence] in Stomach by urea breath test | 0.2 | 0.4 | 0.03 | 0.2 | 0.7 | 0.07 |
| condition_era group during day -365 through 0 days relative to index: Obesity | 39.6 | 18.3 | -0.48 | 32.7 | 36.0 | 0.07 |
| condition_era group during day -365 through 0 days relative to index: Postoperative hypothyroidism | 1.1 | 1.0 | -0.01 | 0.9 | 1.7 | 0.07 |

##### 3.2.2 MDCR

**Supplementary Figure 6:** Preference score distributions, propensity score stratified (middle column) and matched (right column) covariate balance and negative control error distributions (bottom row) for GLP1RA vs DPP4I in the MDCR data source.

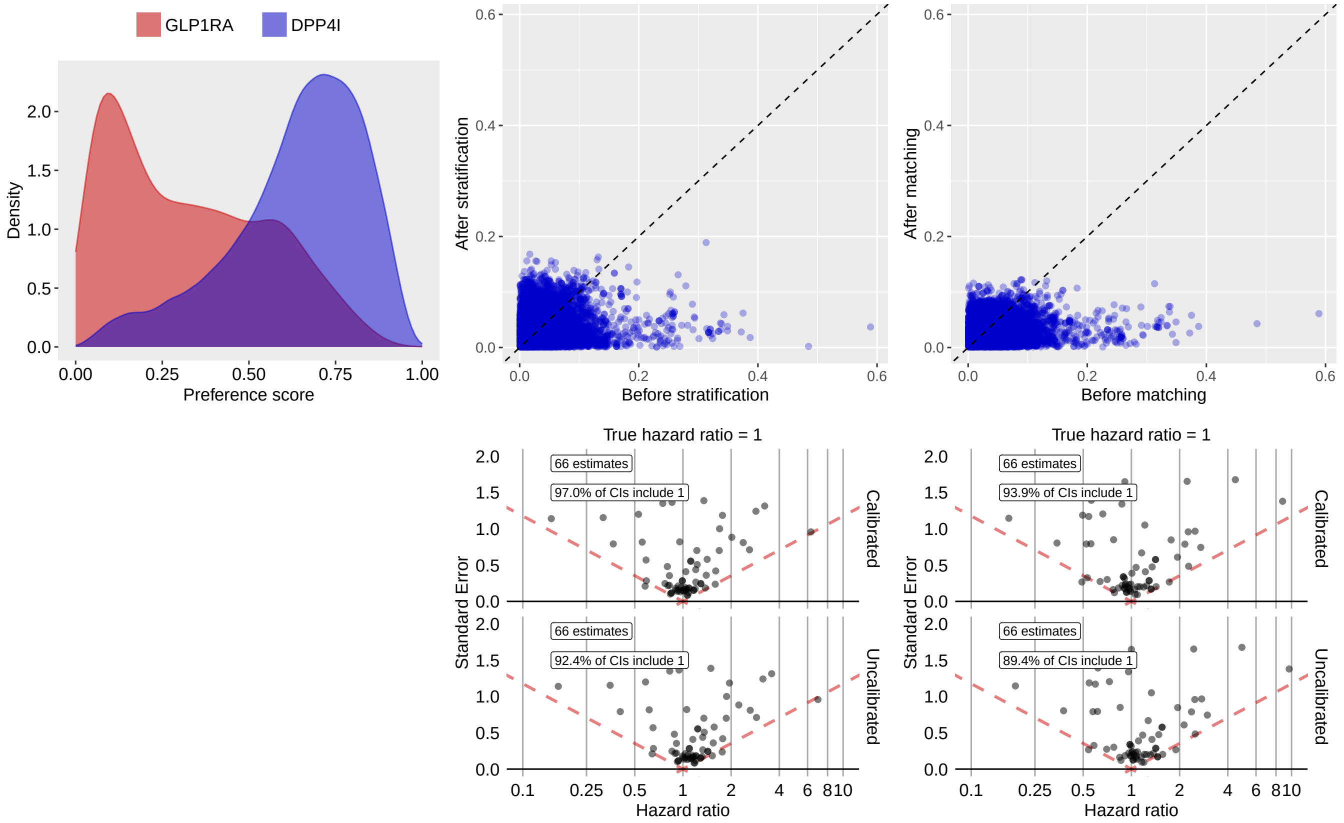

**Supplementary Table 47:** Most extreme patient characteristics balance diagnostics after stratification for GLP1RA vs DPP4I in the MDCR data source.

| Characteristic (total count = 13579) | Before stratification | | | After stratification | | |
|---|---|---|---|---|---|---|
| | T (%) | C (%) | SDf | T (%) | C (%) | SDf |
| index year: 2014 | 3.0 | 10.9 | 0.31 | 5.4 | 10.4 | 0.19 |
| procedure_occurrence during day -365 through 0 days relative to index: Radiologic examination, small intestine, including multiple serial images and scout abdominal radiograph(s), when performed; single-contrast (eg, barium) study | 0.4 | 0.3 | -0.02 | 2.1 | 0.3 | -0.17 |
| age group: 90 - 94 | 0.3 | 1.6 | 0.13 | -0.1 | 1.6 | 0.16 |
| observation during day -365 through 0 days relative to index: Abnormal weight loss | 0.5 | 1.9 | 0.13 | 0.3 | 1.9 | 0.16 |
| device_exposure during day -365 through 0 days relative to index: Nebulizer, with compressor | 1.9 | 1.3 | -0.05 | 3.7 | 1.3 | -0.16 |
| condition_era group during day -365 through 0 days relative to index: Superficial injury of head | 1.2 | 0.9 | -0.03 | 3.1 | 0.9 | -0.16 |
| drug_era group during day -365 through 0 days relative to index: sodium sulfate | 3.9 | 3.7 | -0.01 | 7.3 | 3.7 | -0.16 |
| condition_era group during day -365 through 0 days relative to index: Superficial injury of head and neck | 1.2 | 0.9 | -0.03 | 3.1 | 0.9 | -0.15 |

| (Continued) | T (%) | C (%) | SDf | T (%) | C (%) | SDf |
|---|---|---|---|---|---|---|
| drug_era group during day 0 through 0 days relative to index: ANTIBACTERIALS FOR SYSTEMIC USE | 6.9 | 8.5 | 0.06 | 4.6 | 8.4 | 0.15 |
| procedure_occurrence during day -365 through 0 days relative to index: Colonoscopy, flexible; with removal of tumor(s), polyp(s), or other lesion(s) by snare technique | 4.7 | 3.7 | -0.05 | 7.1 | 3.8 | -0.15 |

**Supplementary Table 48:** Most extreme patient characteristics balance diagnostics after matching for GLP1RA vs DPP4I in the MDCR data source.

| | Before matching | | | After matching | | |
|---|---|---|---|---|---|---|
| Characteristic (total count = 11906) | T (%) | C (%) | SDf | T (%) | C (%) | SDf |
| condition_era group during day -30 through 0 days relative to index: Hematologic neoplasm | -0.2 | 0.7 | 0.09 | -0.2 | 1.0 | 0.12 |
| condition_era group during day -30 through 0 days relative to index: Neoplasm affecting hematopoietic structure | -0.2 | 0.7 | 0.09 | -0.2 | 1.0 | 0.12 |
| condition_era group during day -30 through 0 days relative to index: Mass of lymphoreticular structure | -0.2 | 0.7 | 0.09 | -0.2 | 1.0 | 0.12 |
| age group: 90 - 94 | 0.3 | 1.6 | 0.13 | 0.4 | 1.5 | 0.12 |
| observation during day -365 through 0 days relative to index: Abnormal weight gain | 1.6 | 0.5 | -0.11 | 1.5 | 0.4 | -0.12 |
| condition_era group during day -365 through 0 days relative to index: Adjustment disorder with depressed mood | 0.8 | 0.9 | 0.01 | 0.7 | 2.0 | 0.12 |
| index year: 2014 | 3.0 | 10.9 | 0.31 | 3.7 | 6.1 | 0.12 |
| drug_era group during day -365 through 0 days relative to index: epinephrine | 2.2 | 1.2 | -0.08 | 2.4 | 1.0 | -0.12 |
| drug_era group during day -365 through 0 days relative to index: Alpha- and beta-adrenoreceptor agonists | 2.2 | 1.2 | -0.08 | 2.4 | 1.0 | -0.12 |
| condition_era group during day -30 through 0 days relative to index: Neoplasm of hematopoietic cell type | 0.3 | 0.9 | 0.08 | 0.2 | 1.2 | 0.12 |

##### 3.2.3 Open Claims

**Supplementary Figure 7:** Preference score distributions, propensity score stratified (middle column) and matched (right column) covariate balance and negative control error distributions (bottom row) for GLP1RA vs DPP4I in the Open Claims data source.

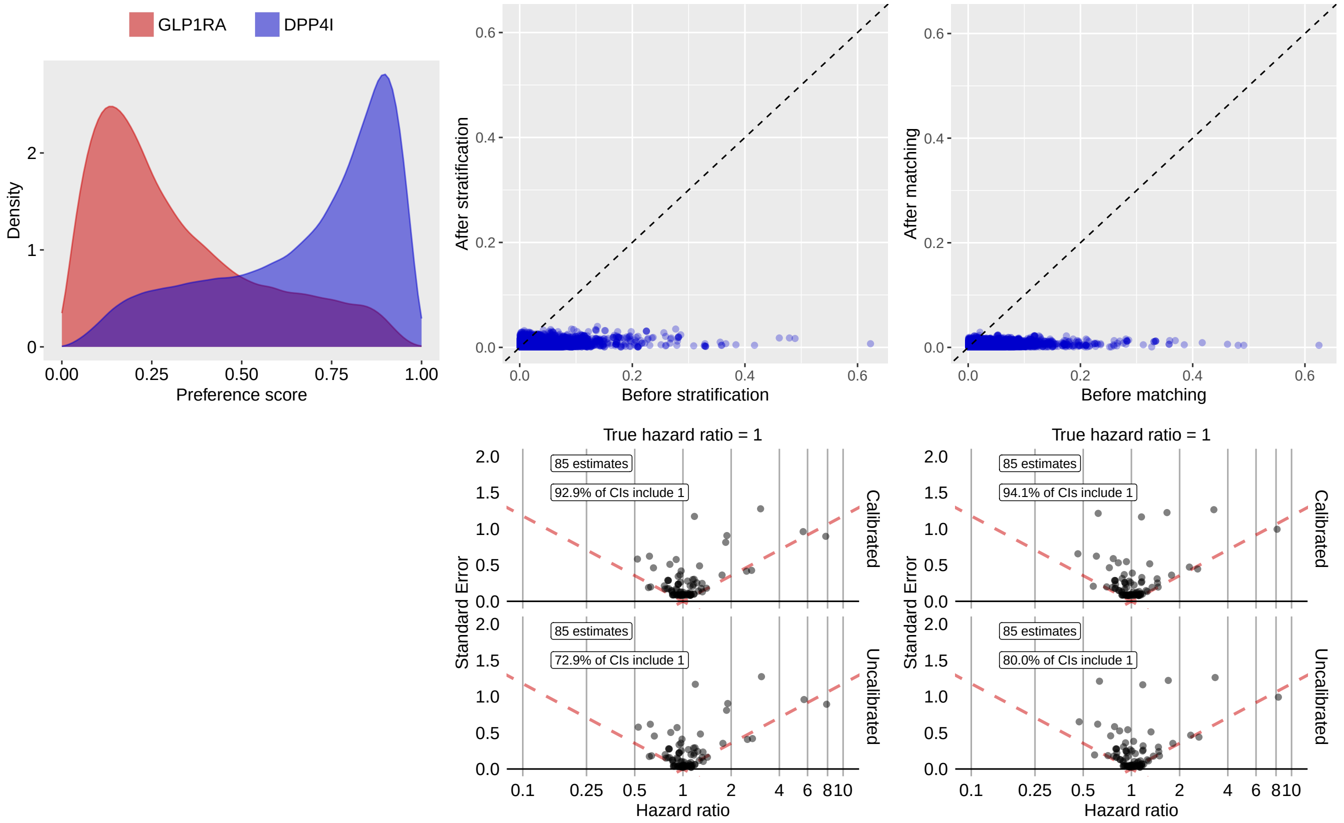

**Supplementary Table 49:** Most extreme patient characteristics balance diagnostics after stratification for GLP1RA vs DPP4I in the Open Claims data source.

| | Before stratification | | | After stratification | | |
|---|---|---|---|---|---|---|
| Characteristic (total count = 10927) | T (%) | C (%) | SDf | T (%) | C (%) | SDf |
| procedure_occurrence during day -30 through 0 days relative to index: Radiologic examination, chest; single view, frontal | 0.1 | 1.2 | 0.14 | 0.6 | 1.0 | 0.04 |
| condition_era group during day -365 through 0 days relative to index: Obesity | 16.2 | 7.4 | -0.28 | 9.8 | 8.8 | -0.04 |
| condition_era group during day -365 through 0 days relative to index: Thrombotic infarction | 0.1 | 0.7 | 0.09 | 0.4 | 0.6 | 0.04 |
| condition_era group during day -30 through 0 days relative to index: Acute renal impairment | 0.4 | 1.7 | 0.14 | 1.1 | 1.5 | 0.03 |
| condition_era group during day -365 through 0 days relative to index: Cerebral infarction due to thrombosis of cerebral arteries | 0.1 | 0.7 | 0.10 | 0.3 | 0.6 | 0.03 |
| condition_era group during day -30 through 0 days relative to index: Hyperkalemia | 0.2 | 0.5 | 0.06 | 0.2 | 0.4 | 0.03 |
| condition_era group during day -30 through 0 days relative to index: Acute renal failure syndrome | 0.4 | 1.7 | 0.14 | 1.1 | 1.5 | 0.03 |
| drug_era group during day -365 through 0 days relative to index: sodium polystyrene sulfonate | 0.1 | 0.2 | 0.04 | 0.1 | 0.2 | 0.03 |

| (Continued) | T (%) | C (%) | SDf | T (%) | C (%) | SDf |
|---|---|---|---|---|---|---|
| drug_era group during day -365 through 0 days relative to index: phentermine | 1.9 | 0.3 | -0.15 | 0.7 | 0.5 | -0.03 |
| condition_era group during day -30 through 0 days relative to index: Renal failure syndrome | 0.4 | 2.1 | 0.15 | 1.4 | 1.8 | 0.03 |

**Supplementary Table 50:** Most extreme patient characteristics balance diagnostics after matching for GLP1RA vs DPP4I in the Open Claims data source.

| | Before matching | | | After matching | | |
|---|---|---|---|---|---|---|
| Characteristic (total count = 10792) | T (%) | C (%) | SDf | T (%) | C (%) | SDf |
| Charlson index - Romano adaptation | 422.6 | 417.1 | -0.02 | 419.9 | 427.6 | 0.03 |
| drug_era group during day -365 through 0 days relative to index: alirocumab | 0.3 | 0.1 | -0.05 | 0.2 | 0.1 | -0.02 |
| drug_era group during day -365 through 0 days relative to index: ivermectin | 0.4 | 0.1 | -0.05 | 0.3 | 0.2 | -0.02 |
| drug_era group during day -365 through 0 days relative to index: Avermectines | 0.4 | 0.1 | -0.05 | 0.3 | 0.2 | -0.02 |
| drug_era group during day -365 through 0 days relative to index: ANTINEMATODAL AGENTS | 0.4 | 0.1 | -0.05 | 0.3 | 0.2 | -0.02 |
| drug_era group during day -365 through 0 days relative to index: Drugs for treatment of hyperkalemia and hyperphosphatemia | 0.1 | 0.1 | 0.02 | 0.1 | 0.1 | 0.02 |
| Diabetes Comorbidity Severity Index (DCSI) | 313.7 | 362.1 | 0.17 | 317.7 | 323.5 | 0.02 |
| condition_era group during day -365 through 0 days relative to index: Hyperglycemia | 18.4 | 13.7 | -0.13 | 17.9 | 18.7 | 0.02 |
| condition_era group during day -365 through 0 days relative to index: Hyperglycemia due to type 2 diabetes mellitus | 17.0 | 12.8 | -0.12 | 16.7 | 17.5 | 0.02 |
| condition_era group during day -365 through 0 days relative to index: Hyperglycemia due to diabetes mellitus | 17.0 | 12.8 | -0.12 | 16.7 | 17.5 | 0.02 |

##### 3.2.4 OptumEHR

**Supplementary Figure 8:** Preference score distributions, propensity score stratified (middle column) and matched (right column) covariate balance and negative control error distributions (bottom row) for GLP1RA vs DPP4I in the OptumEHR data source.

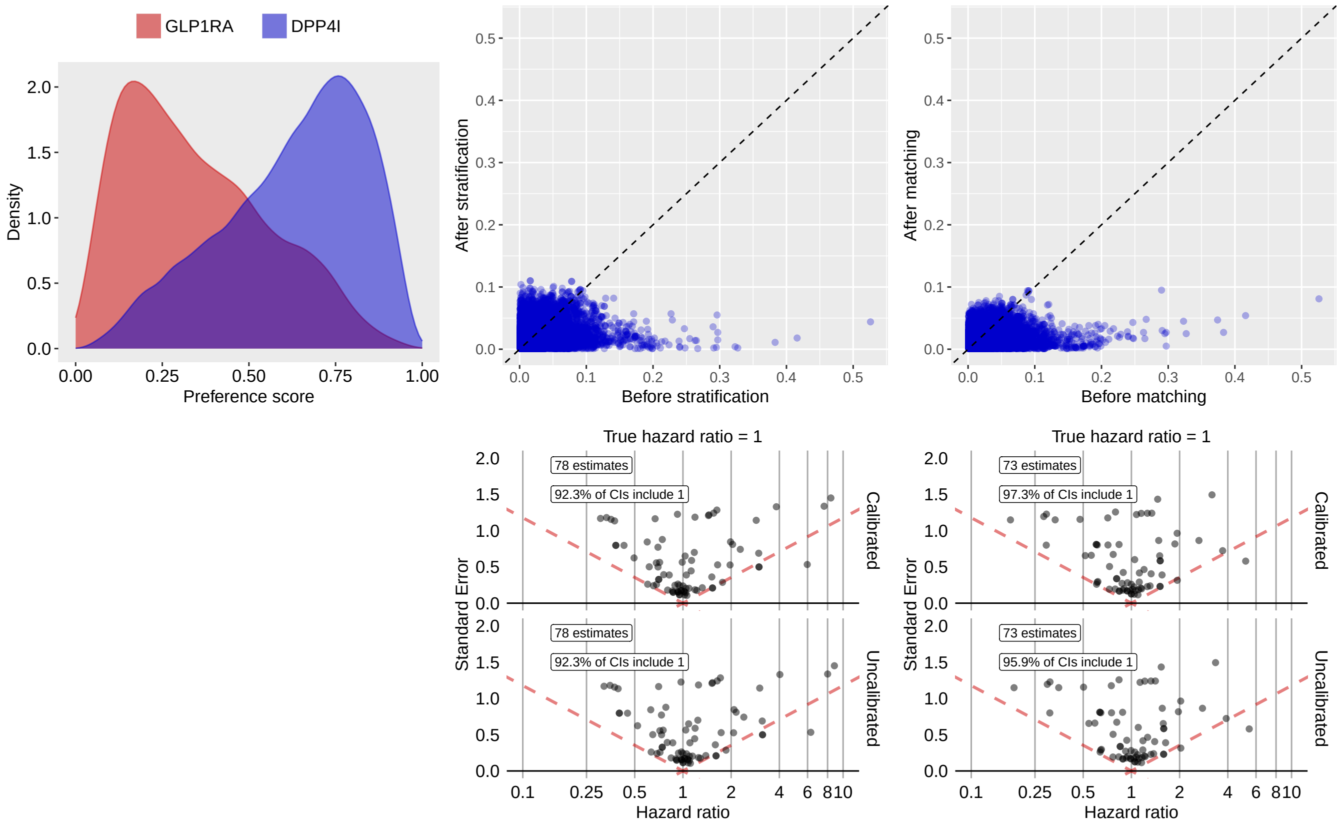

**Supplementary Table 51:** Most extreme patient characteristics balance diagnostics after stratification for GLP1RA vs DPP4I in the OptumEHR data source.

| Characteristic (total count = 14001) | Before stratification | | | After stratification | | |
|---|---|---|---|---|---|---|
| | T (%) | C (%) | SDf | T (%) | C (%) | SDf |
| drug_era group during day -30 through 0 days relative to index: nitrofurantoin | 1.3 | 1.1 | -0.02 | 2.6 | 1.1 | -0.11 |
| drug_era group during day -30 through 0 days relative to index: Nitrofuran derivatives | 1.3 | 1.1 | -0.02 | 2.6 | 1.1 | -0.11 |
| drug_era group during day -365 through 0 days relative to index: ascorbic acid | 6.4 | 4.6 | -0.08 | 7.4 | 4.8 | -0.11 |
| drug_era group during day -365 through 0 days relative to index: Ascorbic acid (vitamin C), plain | 6.4 | 4.6 | -0.08 | 7.4 | 4.8 | -0.11 |
| condition_era group during day -365 through 0 days relative to index: Displacement of intervertebral disc without myelopathy | -0.1 | 0.1 | -0.00 | 0.7 | 0.1 | -0.10 |
| procedure_occurrence during day -365 through 0 days relative to index: Complex cystometrogram (ie, calibrated electronic equipment); with voiding pressure studies (ie, bladder voiding pressure), any technique | 0.3 | 0.1 | -0.05 | 0.7 | 0.1 | -0.10 |
| condition_era group during day -30 through 0 days relative to index: Aneurysm of thoracic aorta | 0.4 | 0.2 | -0.05 | 0.9 | 0.2 | -0.10 |

| (Continued) | T (%) | C (%) | SDf | T (%) | C (%) | SDf |
|---|---|---|---|---|---|---|
| measurement during day -365 through 0 days relative to index: Vitamin B12 measurement | 0.5 | 0.5 | -0.01 | 1.4 | 0.5 | -0.10 |
| condition_era group during day -365 through 0 days relative to index: Pleural effusion | 0.3 | 0.8 | 0.06 | 0.1 | 0.7 | 0.10 |
| observation during day -30 through 0 days relative to index: Other Therapeutic Services - General Classification | 0.6 | 0.5 | -0.02 | 1.4 | 0.4 | -0.10 |

**Supplementary Table 52:** Most extreme patient characteristics balance diagnostics after matching for GLP1RA vs DPP4I in the OptumEHR data source.

| | Before matching | | | After matching | | |
|---|---|---|---|---|---|---|
| Characteristic (total count = 13122) | T (%) | C (%) | SDf | T (%) | C (%) | SDf |
| index year: 2019 | 24.1 | 12.9 | -0.29 | 25.1 | 21.1 | -0.10 |
| condition_era group during day -365 through 0 days relative to index: Neoplasm of intrathoracic organs | -0.1 | 0.5 | 0.09 | -0.1 | 0.5 | 0.09 |
| condition_era group during day -365 through 0 days relative to index: Neoplasm of lower respiratory tract | -0.1 | 0.5 | 0.09 | -0.1 | 0.5 | 0.09 |
| condition_era group during day -365 through 0 days relative to index: Malignant neoplasm of lower respiratory tract | -0.1 | 0.5 | 0.09 | -0.1 | 0.5 | 0.09 |
| condition_era group during day -365 through 0 days relative to index: Neoplasm of lung | -0.1 | 0.5 | 0.09 | -0.1 | 0.5 | 0.09 |
| condition_era group during day -365 through 0 days relative to index: Malignant tumor of lung | -0.1 | 0.5 | 0.09 | -0.1 | 0.5 | 0.09 |
| condition_era group during day -365 through 0 days relative to index: Primary cardiomyopathy | 0.6 | 1.1 | 0.05 | 0.7 | 0.2 | -0.09 |
| index year: 2020 | 29.4 | 9.3 | -0.53 | 26.6 | 30.3 | 0.08 |
| measurement during day -365 through 0 days relative to index: Sodium; urine | -0.1 | 0.5 | 0.10 | -0.1 | 0.4 | 0.08 |
| condition_era group during day -365 through 0 days relative to index: Primary malignant neoplasm of lung | -0.1 | 0.3 | 0.07 | -0.1 | 0.4 | 0.08 |

#### 3.3 GLP1RA vs SU

##### 3.3.1 OptumDOD

**Supplementary Figure 9:** Preference score distributions, propensity score stratified (middle column) and matched (right column) covariate balance and negative control error distributions (bottom row) for GLP1RA vs SU in the OptumDOD data source.

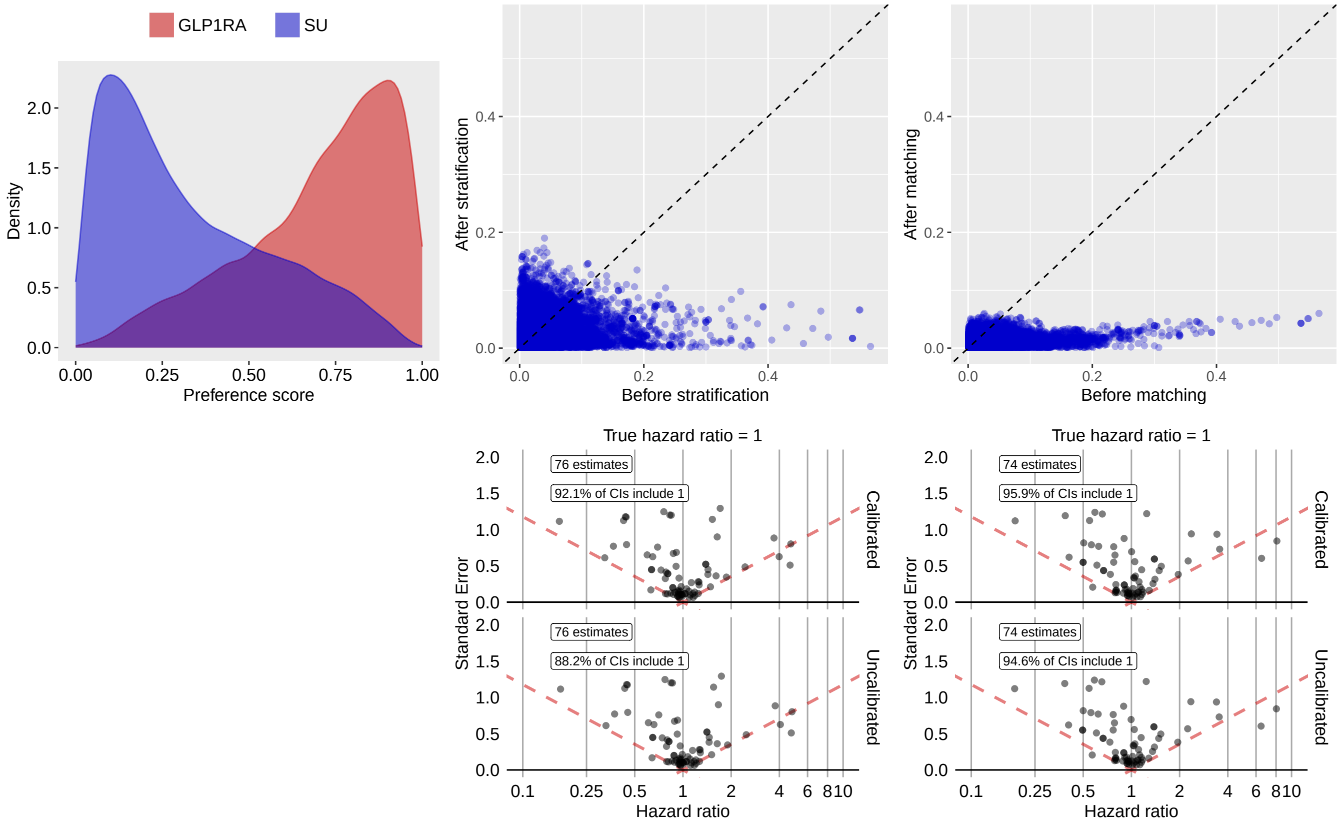

**Supplementary Table 53:** Most extreme patient characteristics balance diagnostics after stratification for GLP1RA vs SU in the OptumDOD data source.

| | Before stratification | | | After stratification | | |
|---|---|---|---|---|---|---|
| Characteristic (total count = 14965) | T (%) | C (%) | SDf | T (%) | C (%) | SDf |
| measurement during day -365 through 0 days relative to index: Eye disorder screening | 0.1 | 0.3 | -0.04 | 2.6 | 0.3 | 0.19 |
| measurement during day -30 through 0 days relative to index: Lipase | 0.9 | 1.3 | -0.04 | 4.1 | 1.3 | 0.17 |
| measurement during day -365 through 0 days relative to index: Amylase | 2.3 | 2.7 | -0.03 | 6.2 | 2.7 | 0.17 |
| condition_era group during day -30 through 0 days relative to index: Hypertensive heart and renal disease with (congestive) heart failure | 0.5 | 0.9 | -0.04 | 3.2 | 0.9 | 0.17 |
| condition_era group during day -365 through 0 days relative to index: Chronic combined systolic and diastolic heart failure | 0.7 | 1.0 | -0.03 | 3.4 | 1.0 | 0.17 |
| condition_era group during day -365 through 0 days relative to index: Hypertensive heart AND renal disease | 2.2 | 2.4 | -0.01 | 5.6 | 2.4 | 0.17 |
| condition_era group during day -30 through 0 days relative to index: Urinary tract obstruction | 2.1 | 2.2 | -0.01 | 5.3 | 2.2 | 0.16 |

| (Continued) | T (%) | C (%) | SDf | T (%) | C (%) | SDf |
|---|---|---|---|---|---|---|
| condition_era group during day -30 through 0 days relative to index: Hypertensive heart AND renal disease | 0.8 | 1.2 | -0.04 | 3.6 | 1.2 | 0.16 |
| condition_era group during day -30 through 0 days relative to index: Benign prostatic hypertrophy with outflow obstruction | 1.8 | 1.9 | -0.00 | 4.7 | 1.9 | 0.16 |
| condition_era group during day -30 through 0 days relative to index: Prostatic obstruction | 1.8 | 1.9 | -0.00 | 4.7 | 1.9 | 0.16 |

**Supplementary Table 54:** Most extreme patient characteristics balance diagnostics after matching for GLP1RA vs SU in the OptumDOD data source.

| | Before matching | | | After matching | | |
|---|---|---|---|---|---|---|
| Characteristic (total count = 16387) | T (%) | C (%) | SDf | T (%) | C (%) | SDf |
| index year: 2021 | 27.1 | 6.7 | 0.56 | 24.5 | 27.1 | -0.06 |
| measurement during day -365 through 0 days relative to index: Immunoassay for infectious agent antibody(ies), qualitative or semiquantitative, single step method (eg, reagent strip); severe acute respiratory syndrome coronavirus 2 (SARS-CoV-2) (Coronavirus disease [COVID-19]) | 0.1 | 0.1 | 0.03 | -0.1 | 0.2 | -0.06 |
| condition_era group during day -365 through 0 days relative to index: Functional diarrhea | 0.1 | 0.4 | -0.05 | 0.1 | 0.4 | -0.06 |
| measurement within normal range during day -365 through 0 days relative to index: traMADol [Mass/volume] in Urine | -0.1 | 0.1 | -0.03 | -0.1 | 0.2 | -0.06 |
| condition_era group during day -30 through 0 days relative to index: Dysthymia | 0.2 | 0.3 | -0.03 | 0.1 | 0.4 | -0.06 |
| age group: 90 - 94 | 0.1 | 0.3 | -0.05 | 0.1 | 0.4 | -0.05 |
| observation during day -30 through 0 days relative to index: Administrative reason for encounter | -0.1 | 0.2 | -0.04 | -0.1 | 0.3 | -0.05 |
| condition_era group during day -365 through 0 days relative to index: Obesity | 39.6 | 17.8 | 0.50 | 35.7 | 38.3 | -0.05 |
| drug_era group during day -365 through 0 days relative to index: megestrol | -0.1 | 0.4 | -0.08 | -0.1 | 0.2 | -0.05 |
| drug_era group during day -365 through 0 days relative to index: Pregnadien derivatives | -0.1 | 0.4 | -0.08 | -0.1 | 0.2 | -0.05 |

##### 3.3.2 MDCR

**Supplementary Figure 10:** Preference score distributions, propensity score stratified (middle column) and matched (right column) covariate balance and negative control error distributions (bottom row) for GLP1RA vs SU in the MDCR data source.

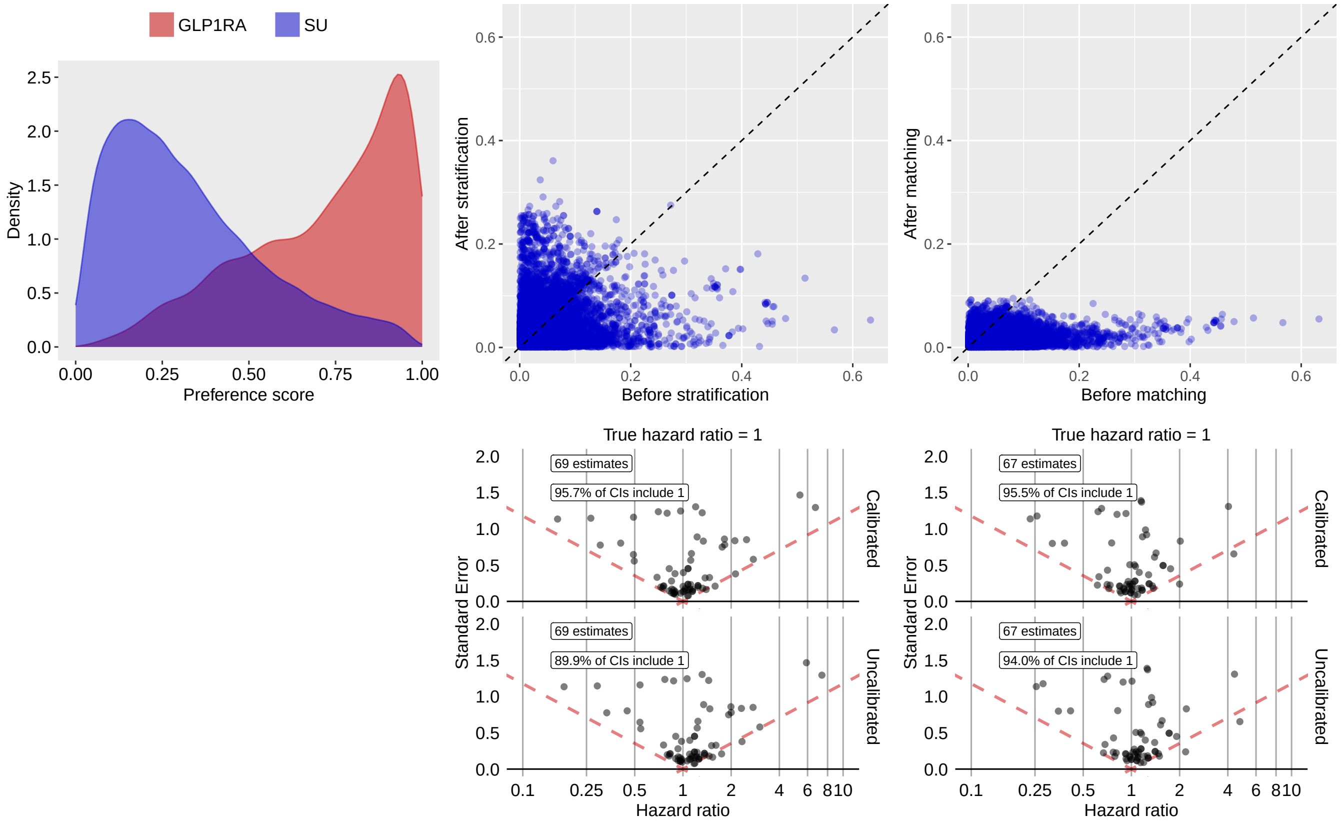

**Supplementary Table 55:** Most extreme patient characteristics balance diagnostics after stratification for GLP1RA vs SU in the MDCR data source.

| Characteristic (total count = 13120) | Before stratification | | | After stratification | | |
|---|---|---|---|---|---|---|
| | T (%) | C (%) | SDf | T (%) | C (%) | SDf |
| condition_era group during day -365 through 0 days relative to index: Joint effusion of ankle AND/OR foot | 0.7 | 0.3 | 0.06 | 6.9 | 0.3 | 0.36 |
| observation during day -365 through 0 days relative to index: 12-Lead ECG Performed (EM) | 0.5 | 0.8 | -0.04 | 7.1 | 0.9 | 0.32 |
| drug_era group during day -365 through 0 days relative to index: dipyridamole | 1.3 | 1.8 | -0.04 | 7.9 | 1.7 | 0.29 |
| drug_era group during day -30 through 0 days relative to index: ipratropium | 1.1 | 2.0 | -0.07 | 8.0 | 1.9 | 0.28 |
| index year: 2013 | 1.9 | 7.6 | -0.27 | 17.7 | 8.5 | 0.28 |
| condition_era group during day -365 through 0 days relative to index: Effusion of joint | 2.8 | 1.8 | 0.07 | 7.9 | 2.0 | 0.27 |
| procedure_occurrence during day -365 through 0 days relative to index: Radiologic examination, small intestine, including multiple serial images and scout abdominal radiograph(s), when performed; single-contrast (eg, barium) study | 0.4 | 0.3 | 0.02 | 4.3 | 0.2 | 0.27 |

| (Continued) | T (%) | C (%) | SDf | T (%) | C (%) | SDf |
|---|---|---|---|---|---|---|
| procedure_occurrence during day -365 through 0 days relative to index: Programming device evaluation (in person) with iterative adjustment of the implantable device to test the function of the device and select optimal permanent programmed values with analysis, review and report by a physician or other qualified health care | -0.2 | 0.3 | -0.01 | 4.2 | 0.3 | 0.27 |
| condition_era group during day -365 through 0 days relative to index: Syncope | 1.4 | 3.5 | -0.14 | 9.8 | 3.3 | 0.26 |
| condition_era group during day -365 through 0 days relative to index: Collapse | 1.4 | 3.5 | -0.14 | 9.8 | 3.3 | 0.26 |

**Supplementary Table 56:** Most extreme patient characteristics balance diagnostics after matching for GLP1RA vs SU in the MDCR data source.

| | Before matching | | | After matching | | |
|---|---|---|---|---|---|---|
| Characteristic (total count = 12217) | T (%) | C (%) | SDf | T (%) | C (%) | SDf |
| condition_era group during day -365 through 0 days relative to index: Limbic disorder | 1.0 | 0.4 | 0.08 | 1.0 | 0.3 | 0.10 |
| condition_era group during day -365 through 0 days relative to index: Pancreatitis | -0.2 | 0.7 | -0.11 | -0.3 | 0.6 | -0.09 |
| drug_era group during day 0 through 0 days relative to index: buspirone | 0.5 | 0.5 | 0.01 | 0.3 | 1.1 | -0.09 |
| drug_era group during day 0 through 0 days relative to index: Azaspirodecanedione derivatives | 0.5 | 0.5 | 0.01 | 0.3 | 1.1 | -0.09 |
| condition_era group during day -365 through 0 days relative to index: Congenital anomaly of abdomen | -0.2 | 0.4 | -0.06 | -0.3 | 0.5 | -0.09 |
| procedure_occurrence during day -365 through 0 days relative to index: Intensity modulated radiotherapy plan, including dose-volume histograms for target and critical structure partial tolerance specifications | -0.2 | 0.4 | -0.06 | -0.3 | 0.7 | -0.09 |
| condition_era group during day -365 through 0 days relative to index: Panic disorder | -0.2 | 0.3 | -0.04 | -0.3 | 0.8 | -0.09 |
| condition_era group during day -365 through 0 days relative to index: Panic | -0.2 | 0.3 | -0.04 | -0.3 | 0.8 | -0.09 |
| drug_era group during day -365 through 0 days relative to index: INSULINS AND ANALOGUES | 1.2 | 0.6 | 0.07 | 1.2 | 0.4 | 0.09 |
| drug_era group during day -365 through 0 days relative to index: Insulins and analogues for injection, long-acting | 1.0 | 0.2 | 0.10 | 1.0 | 0.3 | 0.09 |

##### 3.3.3 Open Claims

**Supplementary Figure 11:** Preference score distributions, propensity score stratified (middle column) and matched (right column) covariate balance and negative control error distributions (bottom row) for GLP1RA vs SU in the Open Claims data source.

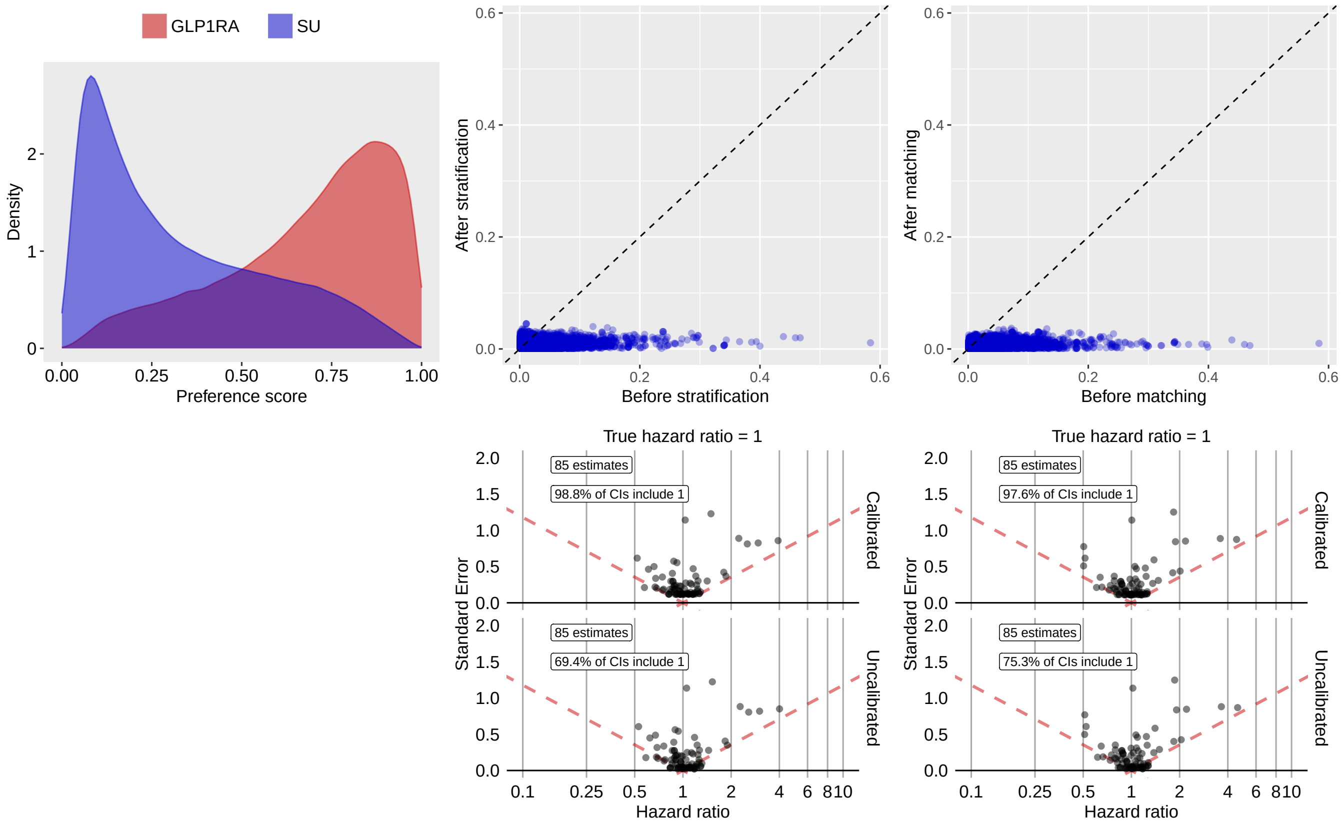

**Supplementary Table 57:** Most extreme patient characteristics balance diagnostics after stratification for GLP1RA vs SU in the Open Claims data source.

| | Before stratification | | | After stratification | | |
|---|---|---|---|---|---|---|
| Characteristic (total count = 10635) | T (%) | C (%) | SDf | T (%) | C (%) | SDf |
| condition_era group during day -365 through 0 days relative to index: Coronary artery spasm | 0.1 | 0.1 | 0.01 | 0.3 | 0.1 | 0.04 |
| condition_era group during day -365 through 0 days relative to index: Vasospasm | 0.1 | 0.1 | 0.01 | 0.3 | 0.1 | 0.04 |
| condition_era group during day -365 through 0 days relative to index: Arteriospasm | 0.1 | 0.1 | 0.01 | 0.3 | 0.1 | 0.04 |
| Diabetes Comorbidity Severity Index (DCSI) | 313.7 | 347.4 | -0.12 | 355.7 | 344.2 | 0.04 |
| procedure_occurrence during day -30 through 0 days relative to index: Radiologic examination, chest; single view, frontal | 0.1 | 1.3 | -0.15 | 0.8 | 1.2 | -0.04 |
| condition_era group during day -365 through 0 days relative to index: Contusion of upper limb | 0.5 | 0.4 | 0.00 | 0.7 | 0.4 | 0.04 |
| condition_era group during day -365 through 0 days relative to index: Contusion of lower limb | 0.9 | 0.8 | 0.01 | 1.2 | 0.8 | 0.04 |
| condition_era group during day -365 through 0 days relative to index: Open wound of forearm | 0.1 | 0.2 | -0.01 | 0.3 | 0.2 | 0.04 |

| (Continued) | T (%) | C (%) | SDf | T (%) | C (%) | SDf |
|---|---|---|---|---|---|---|
| drug_era group during day -365 through 0 days relative to index: phentermine | 1.9 | 0.3 | 0.15 | 0.6 | 0.4 | 0.03 |
| condition_era group during day -365 through 0 days relative to index: Thrombotic infarction | 0.1 | 0.7 | -0.09 | 0.4 | 0.6 | -0.03 |

**Supplementary Table 58:** Most extreme patient characteristics balance diagnostics after matching for GLP1RA vs SU in the Open Claims data source.

| | Before matching | | | After matching | | |
|---|---|---|---|---|---|---|
| Characteristic (total count = 10552) | T (%) | C (%) | SDf | T (%) | C (%) | SDf |
| condition_era group during day -30 through 0 days relative to index: Disorder due to type 2 diabetes mellitus | 17.7 | 15.0 | 0.07 | 17.6 | 19.0 | -0.04 |
| condition_era group during day -365 through 0 days relative to index: Disorder due to type 2 diabetes mellitus | 31.4 | 25.5 | 0.13 | 31.3 | 32.9 | -0.04 |
| Charlson index - Romano adaptation | 422.6 | 407.3 | 0.06 | 418.6 | 427.7 | -0.04 |
| condition_era group during day -30 through 0 days relative to index: Hyperglycemia | 10.4 | 8.4 | 0.07 | 10.4 | 11.4 | -0.03 |
| condition_era group during day -30 through 0 days relative to index: Complication due to diabetes mellitus | 21.0 | 19.0 | 0.05 | 20.9 | 22.3 | -0.03 |
| condition_era group during day -365 through 0 days relative to index: Complication due to diabetes mellitus | 36.7 | 33.0 | 0.08 | 36.5 | 38.1 | -0.03 |
| observation during day -30 through 0 days relative to index: Abnormal weight gain | 0.3 | 0.1 | 0.06 | 0.2 | 0.1 | 0.03 |
| condition_era group during day -30 through 0 days relative to index: Prediabetes | 1.0 | 0.1 | 0.12 | 0.7 | 0.4 | 0.03 |
| condition_era group during day -365 through 0 days relative to index: Hyperglycemia due to type 2 diabetes mellitus | 17.0 | 12.8 | 0.12 | 16.9 | 18.0 | -0.03 |
| condition_era group during day -365 through 0 days relative to index: Hyperglycemia due to diabetes mellitus | 17.0 | 12.8 | 0.12 | 16.9 | 18.0 | -0.03 |

##### 3.3.4 OptumEHR

**Supplementary Figure 12:** Preference score distributions, propensity score stratified (middle column) and matched (right column) covariate balance and negative control error distributions (bottom row) for GLP1RA vs SU in the OptumEHR data source.

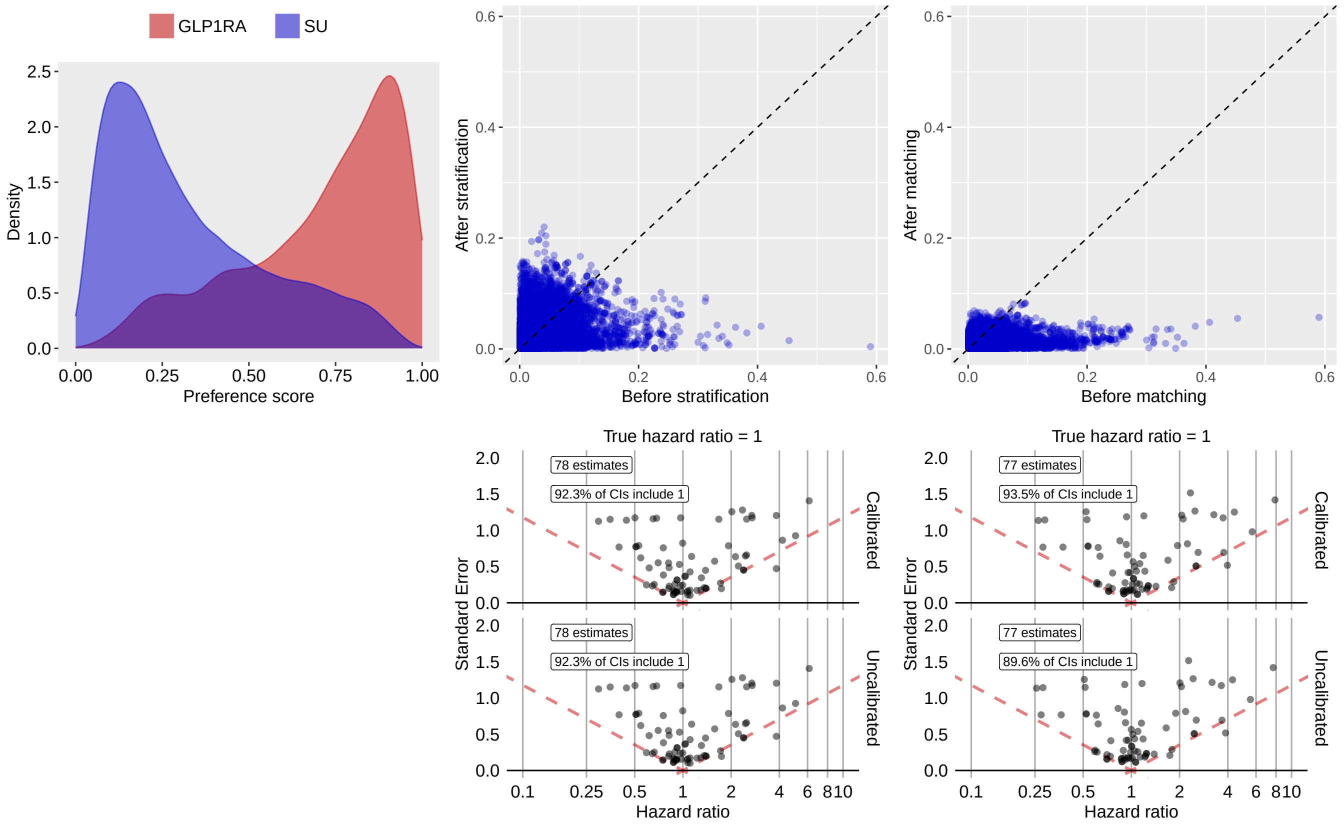

**Supplementary Table 59:** Most extreme patient characteristics balance diagnostics after stratification for GLP1RA vs SU in the OptumEHR data source.

| | Before stratification | | | After stratification | | |
|---|---|---|---|---|---|---|
| Characteristic (total count = 13921) | T (%) | C (%) | SDf | T (%) | C (%) | SDf |
| drug_era group during day 0 through 0 days relative to index: ceftriaxone | 0.4 | 0.7 | -0.04 | 4.0 | 0.7 | 0.22 |
| drug_era group during day -30 through 0 days relative to index: ceftriaxone | 0.6 | 1.0 | -0.04 | 4.3 | 1.0 | 0.21 |
| drug_era group during day 0 through 0 days relative to index: Third-generation cephalosporins | 0.6 | 1.0 | -0.04 | 4.2 | 1.0 | 0.20 |
| drug_era group during day -30 through 0 days relative to index: nitrofurantoin | 1.3 | 0.9 | 0.03 | 4.0 | 1.0 | 0.20 |
| drug_era group during day -30 through 0 days relative to index: Nitrofuran derivatives | 1.3 | 0.9 | 0.03 | 4.0 | 1.0 | 0.20 |
| procedure_occurrence during day -365 through 0 days relative to index: Measurement of post-voiding residual urine and/or bladder capacity by ultrasound, non-imaging | 1.9 | 1.7 | 0.02 | 5.2 | 1.7 | 0.19 |
| drug_era group during day -30 through 0 days relative to index: Third-generation cephalosporins | 0.9 | 1.4 | -0.04 | 4.5 | 1.3 | 0.19 |

| (Continued) | T (%) | C (%) | SDf | T (%) | C (%) | SDf |
|---|---|---|---|---|---|---|
| procedure_occurrence during day -365 through 0 days relative to index: Complex uroflowmetry (eg, calibrated electronic equipment) | 0.6 | 0.3 | 0.04 | 2.2 | 0.3 | 0.17 |
| observation during day -30 through 0 days relative to index: History of urinary tract infection | 0.3 | 0.2 | 0.03 | 1.9 | 0.2 | 0.17 |
| procedure_occurrence during day -365 through 0 days relative to index: Complex cystometrogram (ie, calibrated electronic equipment); with voiding pressure studies (ie, bladder voiding pressure), any technique | 0.3 | 0.1 | 0.06 | 1.6 | 0.1 | 0.17 |

**Supplementary Table 60:** Most extreme patient characteristics balance diagnostics after matching for GLP1RA vs SU in the OptumEHR data source.

| | Before matching | | | After matching | | |
|---|---|---|---|---|---|---|
| Characteristic (total count = 12843) | T (%) | C (%) | SDf | T (%) | C (%) | SDf |
| condition_era group during day -365 through 0 days relative to index: Neoplasm of intrathoracic organs | -0.1 | 0.5 | -0.10 | -0.2 | 0.4 | -0.08 |
| condition_era group during day -365 through 0 days relative to index: Neoplasm of lower respiratory tract | -0.1 | 0.5 | -0.10 | -0.2 | 0.4 | -0.08 |
| condition_era group during day -365 through 0 days relative to index: Neoplasm of lung | -0.1 | 0.5 | -0.09 | -0.2 | 0.4 | -0.08 |
| drug_era group during day -30 through 0 days relative to index: levetiracetam | -0.1 | 0.4 | -0.08 | -0.2 | 0.5 | -0.08 |
| condition_era group during day -365 through 0 days relative to index: Malignant neoplasm of lower respiratory tract | -0.1 | 0.5 | -0.09 | -0.2 | 0.4 | -0.08 |
| condition_era group during day -365 through 0 days relative to index: Malignant tumor of lung | -0.1 | 0.5 | -0.09 | -0.2 | 0.4 | -0.08 |
| drug_era group during day 0 through 0 days relative to index: levetiracetam | -0.1 | 0.4 | -0.07 | -0.2 | 0.4 | -0.07 |
| measurement during day -30 through 0 days relative to index: Hepatitis B virus surface Ab [Titer] in Serum | 0.1 | 0.4 | -0.05 | -0.2 | 0.6 | -0.07 |
| measurement above normal range during day -365 through 0 days relative to index: 3-epi-25-Hydroxyvitamin D3/25-hydroxyvitamin D.total in Serum or Plasma | 0.1 | 0.1 | 0.01 | -0.2 | 0.3 | -0.07 |
| condition_era group during day -365 through 0 days relative to index: Mass of pancreas | 0.2 | 0.5 | -0.05 | -0.2 | 0.5 | -0.07 |

#### 3.4 SGLT2I vs DPP4I

##### 3.4.1 OptumDOD

**Supplementary Figure 13:** Preference score distributions, propensity score stratified (middle column) and matched (right column) covariate balance and negative control error distributions (bottom row) for SGLT2I vs DPP4I in the OptumDOD data source.

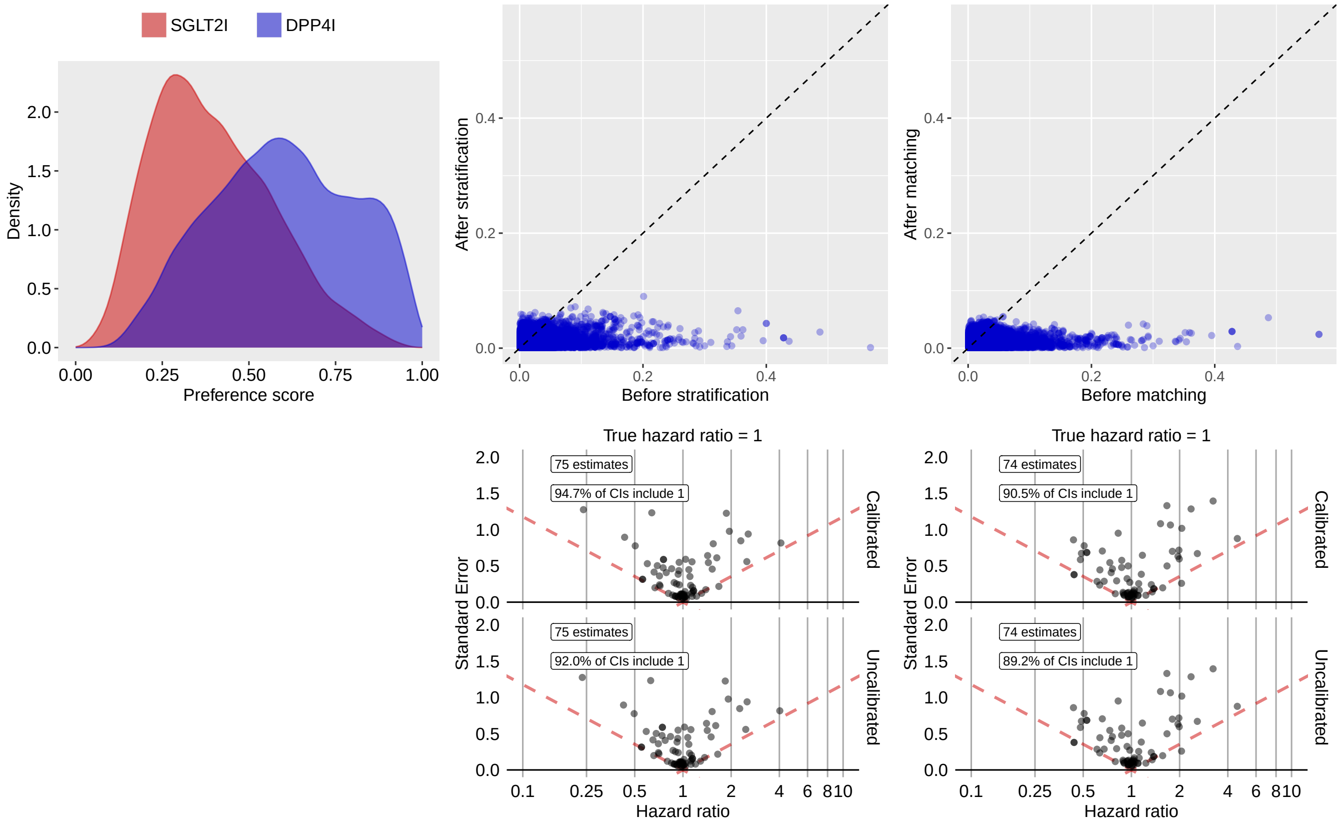

**Supplementary Table 61:** Most extreme patient characteristics balance diagnostics after stratification for SGLT2I vs DPP4I in the OptumDOD data source.

| | Before stratification | | | After stratification | | |
|---|---|---|---|---|---|---|
| Characteristic (total count = 16151) | T (%) | C (%) | SDf | T (%) | C (%) | SDf |
| procedure_occurrence during day -30 through 0 days relative to index: Radiologic examination, chest; single view, frontal | 0.2 | 2.6 | 0.20 | 0.7 | 1.6 | 0.09 |
| condition_era group during day -30 through 0 days relative to index: Acidosis | 0.4 | 1.2 | 0.09 | 0.5 | 1.1 | 0.07 |
| observation during day -30 through 0 days relative to index: Room & Board - Semi-private Two Bed (Medical or General) - Medical/Surgical/Gyn | 0.3 | 1.0 | 0.08 | 0.3 | 0.8 | 0.07 |
| procedure_occurrence during day -30 through 0 days relative to index: Ultrasound, retroperitoneal (eg, renal, aorta, nodes), real time with image documentation; complete | 0.4 | 1.5 | 0.11 | 0.6 | 1.2 | 0.07 |
| index year: 2013 | 0.3 | 6.7 | 0.35 | 2.2 | 3.3 | 0.07 |
| condition_era group during day -30 through 0 days relative to index: Hypertensive renal failure | 0.1 | 1.1 | 0.14 | 0.2 | 0.5 | 0.06 |
| age group: 90 - 94 | 0.3 | 0.5 | 0.03 | 0.2 | 0.6 | 0.06 |

| (Continued) | T (%) | C (%) | SDf | T (%) | C (%) | SDf |
|---|---|---|---|---|---|---|
| measurement during day -30 through 0 days relative to index: Immature granulocytes/100 leukocytes in Blood | 3.3 | 6.2 | 0.14 | 6.8 | 5.3 | -0.06 |
| measurement during day -30 through 0 days relative to index: Immature granulocytes [#/volume] in Blood | 2.9 | 5.7 | 0.14 | 6.3 | 4.9 | -0.06 |
| condition_era group during day -30 through 0 days relative to index: Chronic kidney disease stage 4 | 0.3 | 1.1 | 0.11 | 0.5 | 1.0 | 0.06 |

**Supplementary Table 62:** Most extreme patient characteristics balance diagnostics after matching for SGLT2I vs DPP4I in the OptumDOD data source.

| | Before matching | | | After matching | | |
|---|---|---|---|---|---|---|
| Characteristic (total count = 16374) | T (%) | C (%) | SDf | T (%) | C (%) | SDf |
| condition_era group during day -365 through 0 days relative to index: Central alveolar hypoventilation syndrome | 0.5 | 0.2 | -0.06 | 0.5 | 0.2 | -0.06 |
| condition_era group during day -365 through 0 days relative to index: Idiopathic sleep related non-obstructive alveolar hypoventilation | 0.5 | 0.1 | -0.06 | 0.4 | 0.1 | -0.05 |
| age group: 90 - 94 | 0.3 | 0.5 | 0.03 | 0.3 | 0.6 | 0.05 |
| index year: 2021 | 24.3 | 7.1 | -0.49 | 16.2 | 18.2 | 0.05 |
| condition_era group during day -365 through 0 days relative to index: Benign neoplasm of nervous system | 0.6 | 0.8 | 0.02 | 0.5 | 1.0 | 0.05 |
| measurement during day -365 through 0 days relative to index: Estradiol (E2) [Mass/volume] in Serum or Plasma | 0.5 | 0.2 | -0.04 | 0.5 | 0.2 | -0.05 |
| procedure_occurrence during day -365 through 0 days relative to index: 3-dimensional radiotherapy plan, including dose-volume histograms | 0.1 | 0.3 | 0.05 | 0.1 | 0.3 | 0.05 |
| condition_era group during day -365 through 0 days relative to index: Neoplasm of cerebrum | 0.1 | 0.3 | 0.04 | 0.1 | 0.3 | 0.05 |
| procedure_occurrence during day -365 through 0 days relative to index: Insertion or replacement of permanent implantable defibrillator system, with transvenous lead(s), single or dual chamber | 0.3 | 0.2 | -0.01 | 0.1 | 0.3 | 0.05 |
| condition_era group during day -30 through 0 days relative to index: Hyperosmolarity | 0.4 | 0.2 | -0.04 | 0.4 | 0.2 | -0.05 |

##### 3.4.2 MDCR

**Supplementary Figure 14:** Preference score distributions, propensity score stratified (middle column) and matched (right column) covariate balance and negative control error distributions (bottom row) for SGLT2I vs DPP4I in the MDCR data source.

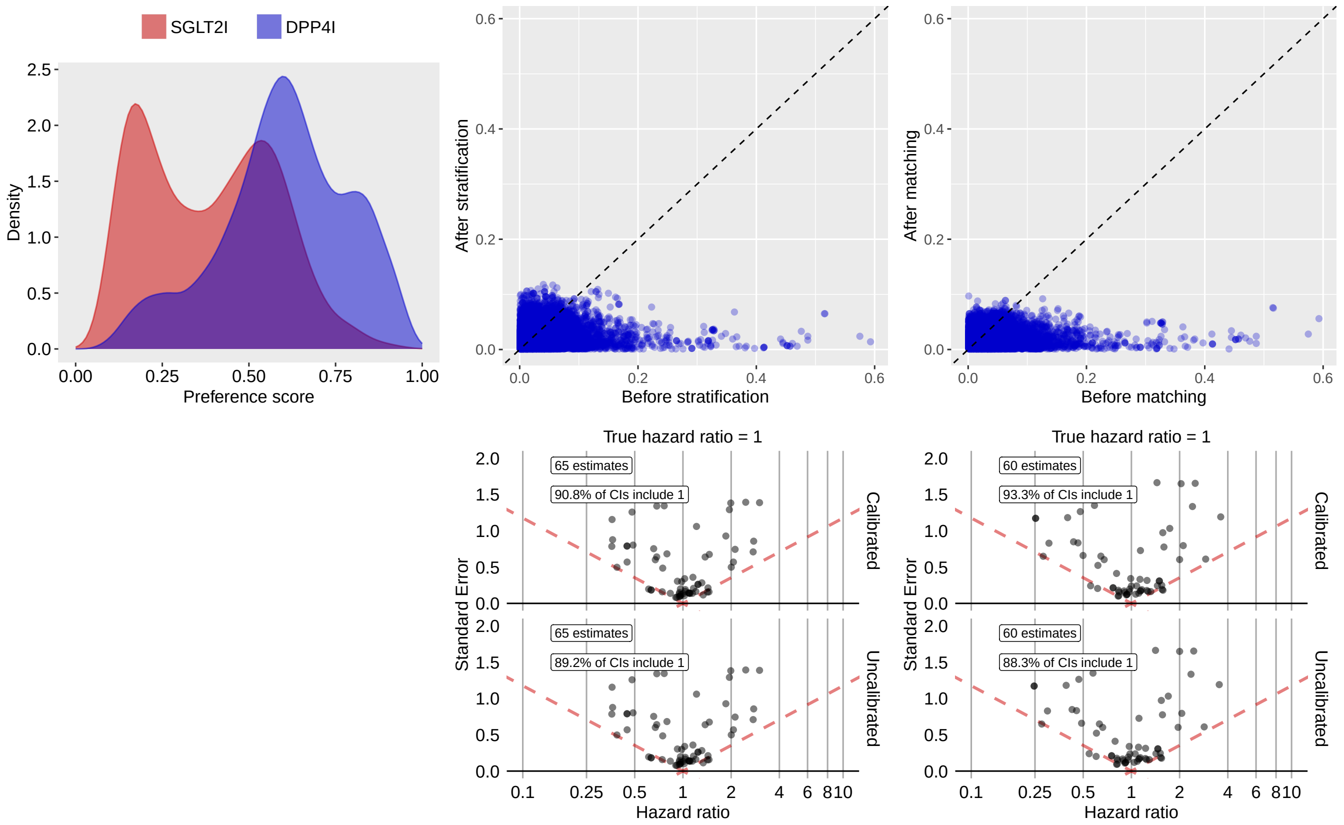

**Supplementary Table 63:** Most extreme patient characteristics balance diagnostics after stratification for SGLT2I vs DPP4I in the MDCR data source.

| Characteristic (total count = 13856) | Before stratification | | | After stratification | | |
|---|---|---|---|---|---|---|
| | T (%) | C (%) | SDf | T (%) | C (%) | SDf |
| condition_era group during day -30 through 0 days relative to index: Benign hypertensive heart disease without congestive heart failure | 0.2 | 0.5 | 0.04 | 1.3 | 0.2 | -0.12 |
| condition_era group during day -365 through 0 days relative to index: Atrophic gastritis | 0.7 | 1.2 | 0.06 | 2.3 | 0.9 | -0.12 |
| condition_era group during day -30 through 0 days relative to index: Renal impairment | 6.1 | 7.7 | 0.06 | 5.9 | 8.9 | 0.11 |
| condition_era group during day -30 through 0 days relative to index: Benign hypertensive heart disease | 0.3 | 0.5 | 0.04 | 1.3 | 0.3 | -0.11 |
| drug_era group during day 0 through 0 days relative to index: nitroglycerin | 1.2 | 0.9 | -0.03 | 1.7 | 0.6 | -0.11 |
| condition_era group during day -30 through 0 days relative to index: Acute nephropathy | 0.7 | 2.4 | 0.13 | 0.9 | 2.3 | 0.11 |
| condition_era group during day -30 through 0 days relative to index: Venous retinal branch occlusion | 0.2 | 0.1 | -0.02 | 0.9 | 0.1 | -0.11 |
| condition_era group during day -365 through 0 days relative to index: Paraplegia | -0.1 | 0.1 | -0.01 | 0.7 | 0.1 | -0.11 |

| (Continued) | T (%) | C (%) | SDf | T (%) | C (%) | SDf |
|---|---|---|---|---|---|---|
| condition_era group during day -365 through 0 days relative to index: Paralytic syndrome of both lower limbs | -0.1 | 0.1 | -0.01 | 0.7 | 0.1 | -0.11 |
| condition_era group during day -30 through 0 days relative to index: Acute renal failure syndrome | 0.7 | 2.3 | 0.13 | 0.9 | 2.2 | 0.10 |

**Supplementary Table 64:** Most extreme patient characteristics balance diagnostics after matching for SGLT2I vs DPP4I in the MDCR data source.

| | Before matching | | | After matching | | |
|---|---|---|---|---|---|---|
| Characteristic (total count = 13227) | T (%) | C (%) | SDf | T (%) | C (%) | SDf |
| observation during day -30 through 0 days relative to index: Review of all medications by a prescribing practitioner or clinical pharmacist (such as, prescriptions, OTCs, herbal therapies and supplements) documented in the medical record (COA) | 0.5 | 0.5 | 0.00 | 0.4 | 1.3 | 0.10 |
| age group: 90 - 94 | 0.4 | 1.6 | 0.13 | 0.4 | 1.3 | 0.09 |
| drug_era group during day -365 through 0 days relative to index: vancomycin | 1.0 | 0.5 | -0.06 | 1.0 | 0.3 | -0.09 |
| drug_era group during day -365 through 0 days relative to index: Glycopeptide antibacterials | 1.0 | 0.5 | -0.06 | 1.0 | 0.3 | -0.09 |
| observation during day -30 through 0 days relative to index: Medication list documented in medical record (COA) | 0.6 | 0.5 | -0.02 | 0.5 | 1.3 | 0.09 |
| measurement above normal range during day -365 through 0 days relative to index: Thyroxine (T4) free [Mass/volume] in Serum or Plasma | -0.1 | 0.2 | 0.06 | -0.1 | 0.4 | 0.08 |
| observation during day -365 through 0 days relative to index: Influenza immunization was not administered, reason not given | 1.1 | 1.6 | 0.05 | 1.2 | 2.3 | 0.08 |
| observation during day -365 through 0 days relative to index: Review of all medications by a prescribing practitioner or clinical pharmacist (such as, prescriptions, OTCs, herbal therapies and supplements) documented in the medical record (COA) | 1.4 | 0.8 | -0.06 | 1.2 | 2.2 | 0.08 |
| drug_era group during day -30 through 0 days relative to index: valacyclovir | 1.1 | 0.4 | -0.08 | 1.1 | 0.4 | -0.08 |
| drug_era group during day -365 through 0 days relative to index: Phenothiazines with piperazine structure | 0.3 | 0.9 | 0.08 | 0.3 | 0.9 | 0.08 |

##### 3.4.3 Open Claims

**Supplementary Figure 15:** Preference score distributions, propensity score stratified (middle column) and matched (right column) covariate balance and negative control error distributions (bottom row) for SGLT2I vs DPP4I in the Open Claims data source.

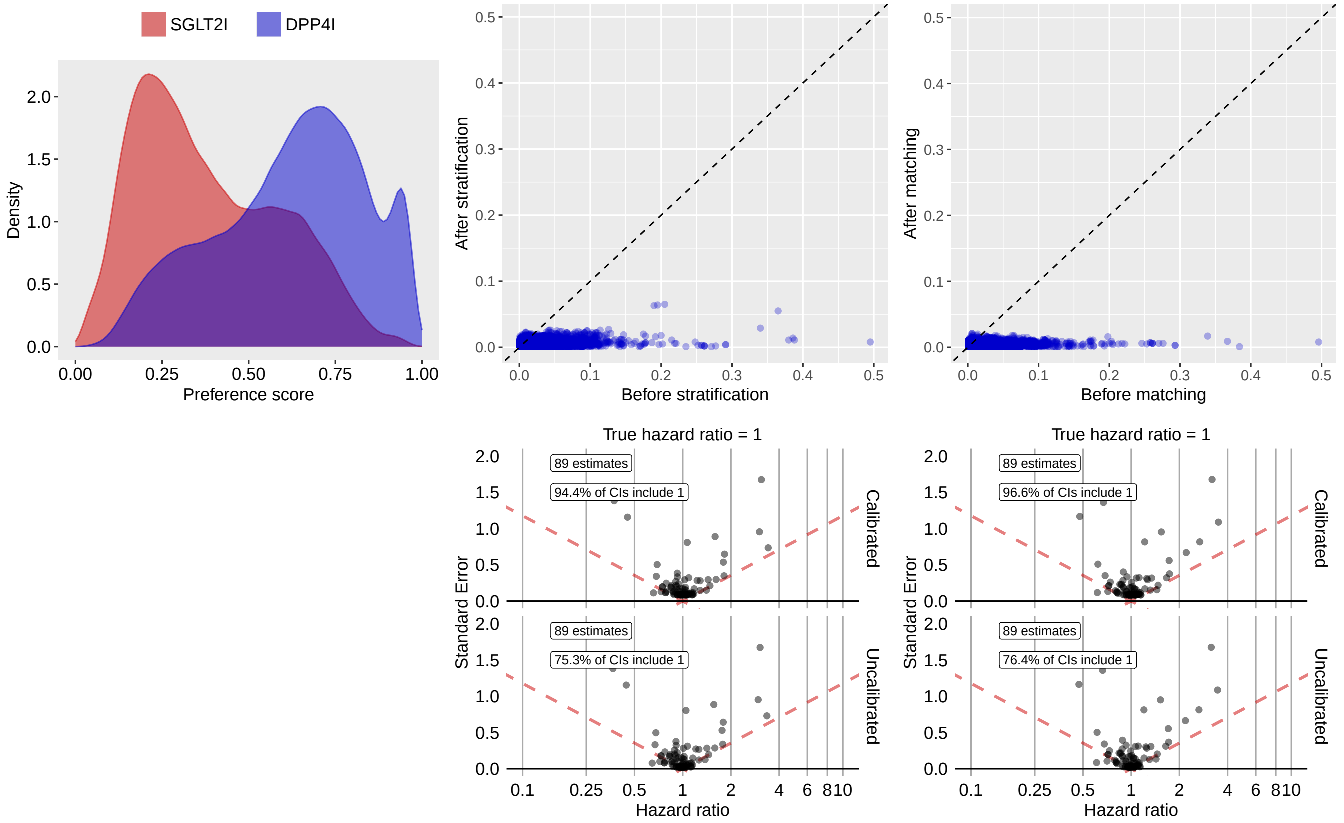

**Supplementary Table 65:** Most extreme patient characteristics balance diagnostics after stratification for SGLT2I vs DPP4I in the Open Claims data source.

| Characteristic (total count = 10771) | Before stratification | | | After stratification | | |
|---|---|---|---|---|---|---|
| | T (%) | C (%) | SDf | T (%) | C (%) | SDf |
| drug_era group during day -365 through 0 days relative to index: sacubitril | 2.9 | 0.3 | -0.20 | 1.4 | 0.7 | -0.07 |
| drug_era group during day -30 through 0 days relative to index: sacubitril | 2.5 | 0.3 | -0.20 | 1.2 | 0.6 | -0.06 |
| drug_era group during day 0 through 0 days relative to index: sacubitril | 2.4 | 0.2 | -0.19 | 1.1 | 0.6 | -0.06 |
| index year: 2013 | 0.4 | 7.3 | 0.36 | 3.5 | 4.6 | 0.06 |
| index year: 2014 | 2.8 | 11.3 | 0.34 | 9.1 | 8.3 | -0.03 |
| condition_era group during day -30 through 0 days relative to index: Renal failure syndrome | 0.6 | 2.1 | 0.13 | 1.3 | 1.6 | 0.03 |
| condition_era group during day -365 through 0 days relative to index: Chronic congestive heart failure | 2.2 | 0.8 | -0.11 | 1.3 | 1.0 | -0.03 |
| observation during day -30 through 0 days relative to index: Blood bank procedure | 0.1 | 0.3 | 0.04 | 0.1 | 0.2 | 0.03 |
| drug_era group during day -30 through 0 days relative to index: valsartan | 6.9 | 5.9 | -0.04 | 6.4 | 5.8 | -0.03 |

| (Continued) | T (%) | C (%) | SDf | T (%) | C (%) | SDf |
|---|---|---|---|---|---|---|
| condition_era group during day -30 through 0 days relative to index: Systolic heart failure | 2.1 | 0.9 | -0.10 | 1.3 | 1.0 | -0.03 |

**Supplementary Table 66:** Most extreme patient characteristics balance diagnostics after matching for SGLT2I vs DPP4I in the Open Claims data source.

| | Before matching | | | After matching | | |
|---|---|---|---|---|---|---|
| Characteristic (total count = 10563) | T (%) | C (%) | SDf | T (%) | C (%) | SDf |
| condition_era group during day -365 through 0 days relative to index: Primary malignant neoplasm of pancreas | 0.1 | 0.1 | -0.01 | 0.2 | 0.1 | -0.02 |
| condition_era group during day -365 through 0 days relative to index: Malignant tumor of pancreas | 0.1 | 0.1 | -0.01 | 0.2 | 0.1 | -0.02 |
| condition_era group during day -365 through 0 days relative to index: Mass of pancreas | 0.4 | 0.3 | -0.01 | 0.4 | 0.3 | -0.02 |
| condition_era group during day -365 through 0 days relative to index: Neoplasm of pancreas | 0.2 | 0.1 | -0.01 | 0.2 | 0.1 | -0.02 |
| drug_era group during day -365 through 0 days relative to index: evolocumab | 0.4 | 0.1 | -0.07 | 0.3 | 0.2 | -0.02 |
| condition_era group during day -365 through 0 days relative to index: Permanent atrial fibrillation | 0.4 | 0.1 | -0.06 | 0.3 | 0.2 | -0.02 |
| procedure_occurrence during day -365 through 0 days relative to index: Fluoroscopy of Multiple Coronary Arteries using Low Osmolar Contrast | 0.3 | 0.1 | -0.04 | 0.2 | 0.1 | -0.02 |
| drug_era group during day -365 through 0 days relative to index: liothyronine | 0.3 | 0.2 | -0.01 | 0.3 | 0.2 | -0.02 |
| drug_era group during day -30 through 0 days relative to index: mycophenolate mofetil | 0.1 | 0.1 | 0.01 | 0.1 | 0.1 | 0.02 |
| index year: 2014 | 2.8 | 11.3 | 0.34 | 3.6 | 3.3 | -0.02 |

##### 3.4.4 OptumEHR

**Supplementary Figure 16:** Preference score distributions, propensity score stratified (middle column) and matched (right column) covariate balance and negative control error distributions (bottom row) for SGLT2I vs DPP4I in the OptumEHR data source.

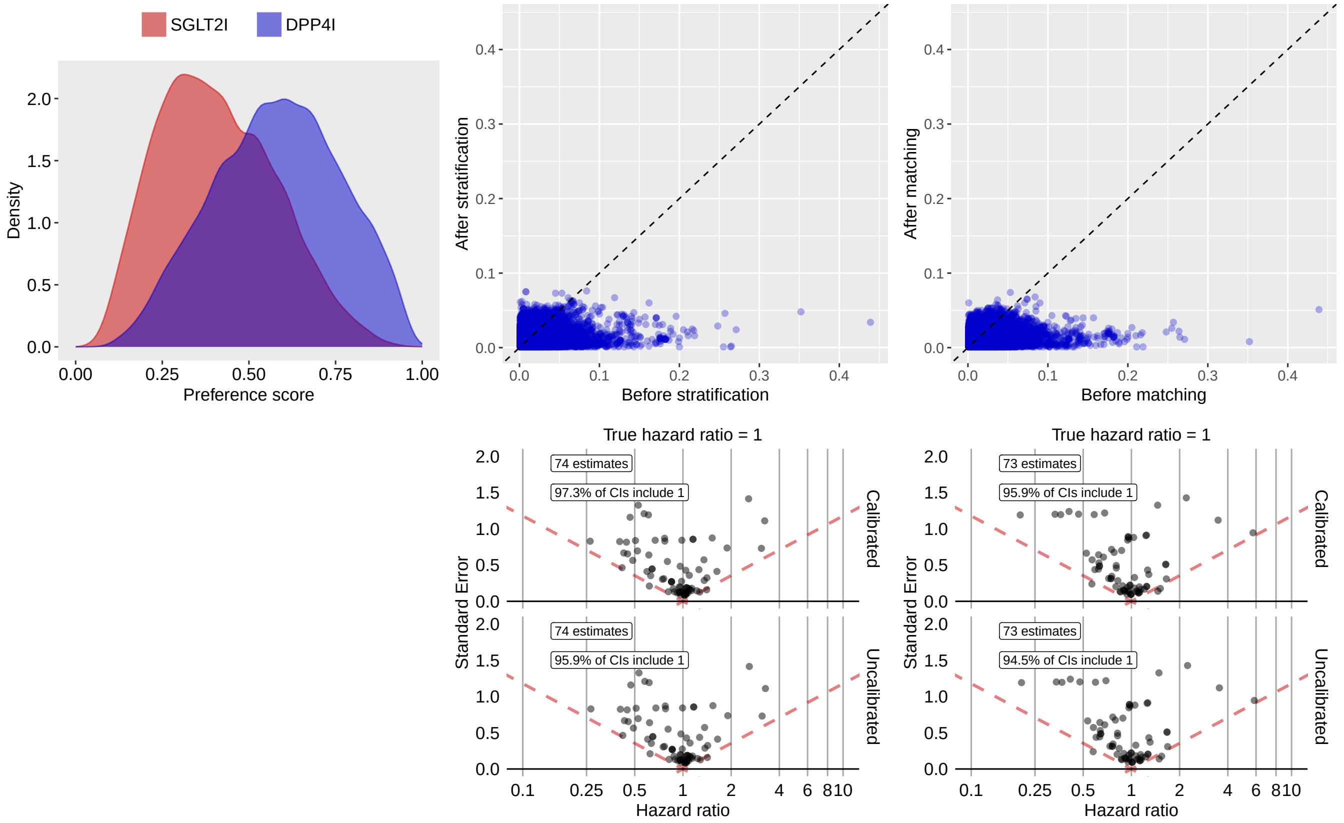

**Supplementary Table 67:** Most extreme patient characteristics balance diagnostics after stratification for SGLT2I vs DPP4I in the OptumEHR data source.

| | Before stratification | | | After stratification | | |
|---|---|---|---|---|---|---|
| Characteristic (total count = 14082) | T (%) | C (%) | SDf | T (%) | C (%) | SDf |
| condition_era group during day -365 through 0 days relative to index: Chronic kidney disease stage 4 | 0.2 | 0.7 | 0.08 | 0.2 | 0.7 | 0.08 |
| condition_era group during day -30 through 0 days relative to index: Allergic disorder of respiratory system | 0.1 | 0.1 | -0.01 | 0.4 | 0.1 | -0.07 |
| condition_era group during day -30 through 0 days relative to index: Allergic disorder of respiratory tract | 0.1 | 0.1 | -0.01 | 0.4 | 0.1 | -0.07 |
| observation during day -30 through 0 days relative to index: Abnormal weight gain | 0.5 | 0.2 | -0.05 | 0.6 | 0.2 | -0.07 |
| procedure_occurrence during day -365 through 0 days relative to index: Computer-aided detection (computer algorithm analysis of digital image data for lesion detection) with further review for interpretation, with or without digitization of film radiographic images; diagnostic mammography (List separately in addition to code | 0.5 | 0.9 | 0.04 | 1.5 | 0.7 | -0.07 |
| measurement below normal range during day -365 through 0 days relative to index: Alanine aminotransferase [Enzymatic activity/volume] in Serum or Plasma | 0.9 | 2.4 | 0.12 | 1.1 | 1.9 | 0.07 |

| (Continued) | T (%) | C (%) | SDf | T (%) | C (%) | SDf |
|---|---|---|---|---|---|---|
| condition_era group during day -30 through 0 days relative to index: Hypo-osmolality and or hyponatremia | 0.2 | 0.6 | 0.06 | 0.2 | 0.6 | 0.07 |
| condition_era group during day -365 through 0 days relative to index: Upper urinary tract infection | -0.1 | 0.3 | 0.07 | -0.1 | 0.2 | 0.06 |
| observation during day -365 through 0 days relative to index: Pharmacy - Other Pharmacy | -0.1 | 0.3 | 0.06 | -0.1 | 0.2 | 0.06 |
| measurement during day -30 through 0 days relative to index: Phosphorus inorganic (phosphate) | 0.2 | 0.7 | 0.07 | 0.2 | 0.6 | 0.06 |

**Supplementary Table 68:** Most extreme patient characteristics balance diagnostics after matching for SGLT2I vs DPP4I in the OptumEHR data source.

| | Before matching | | | After matching | | |
|---|---|---|---|---|---|---|
| Characteristic (total count = 14007) | T (%) | C (%) | SDf | T (%) | C (%) | SDf |
| observation during day -30 through 0 days relative to index: Abnormal weight gain | 0.5 | 0.2 | -0.05 | 0.5 | 0.1 | -0.07 |
| condition_era group during day -365 through 0 days relative to index: Chronic kidney disease stage 4 | 0.2 | 0.7 | 0.08 | 0.2 | 0.6 | 0.07 |
| condition_era group during day -365 through 0 days relative to index: Mood disorder in partial remission | 0.9 | 0.3 | -0.07 | 0.8 | 0.3 | -0.07 |
| condition_era group during day -365 through 0 days relative to index: Disorder in partial remission | 0.9 | 0.3 | -0.07 | 0.8 | 0.3 | -0.07 |
| observation during day -30 through 0 days relative to index: Most recent hemoglobin A1c (HbA1c) level greater than or equal to 7.0% and less than 8.0% (DM) | 0.7 | 0.4 | -0.04 | 0.5 | 1.1 | 0.06 |
| condition_era group during day -30 through 0 days relative to index: Chronic kidney disease stage 3A | 0.6 | 0.1 | -0.09 | 0.5 | 0.2 | -0.06 |
| condition_era group during day -365 through 0 days relative to index: Major depression in partial remission | 0.9 | 0.3 | -0.07 | 0.8 | 0.3 | -0.06 |
| measurement within normal range during day -365 through 0 days relative to index: Bicarbonate [Moles/volume] in Blood | 0.5 | 0.5 | -0.00 | 0.5 | 0.2 | -0.06 |
| measurement during day -30 through 0 days relative to index: Screening for cardiovascular system disease | 0.1 | 0.3 | 0.04 | 0.1 | 0.4 | 0.06 |
| procedure_occurrence during day -365 through 0 days relative to index: External electrocardiographic recording up to 48 hours by continuous rhythm recording and storage; review and interpretation by a physician or other qualified health care professional | 0.5 | 0.8 | 0.04 | 0.4 | 0.9 | 0.06 |

### 3.4.5 VA

**Supplementary Figure 17:** Preference score distributions, propensity score stratified (middle column) and matched (right column) covariate balance and negative control error distributions (bottom row) for SGLT2I vs DPP4I in the VA data source.

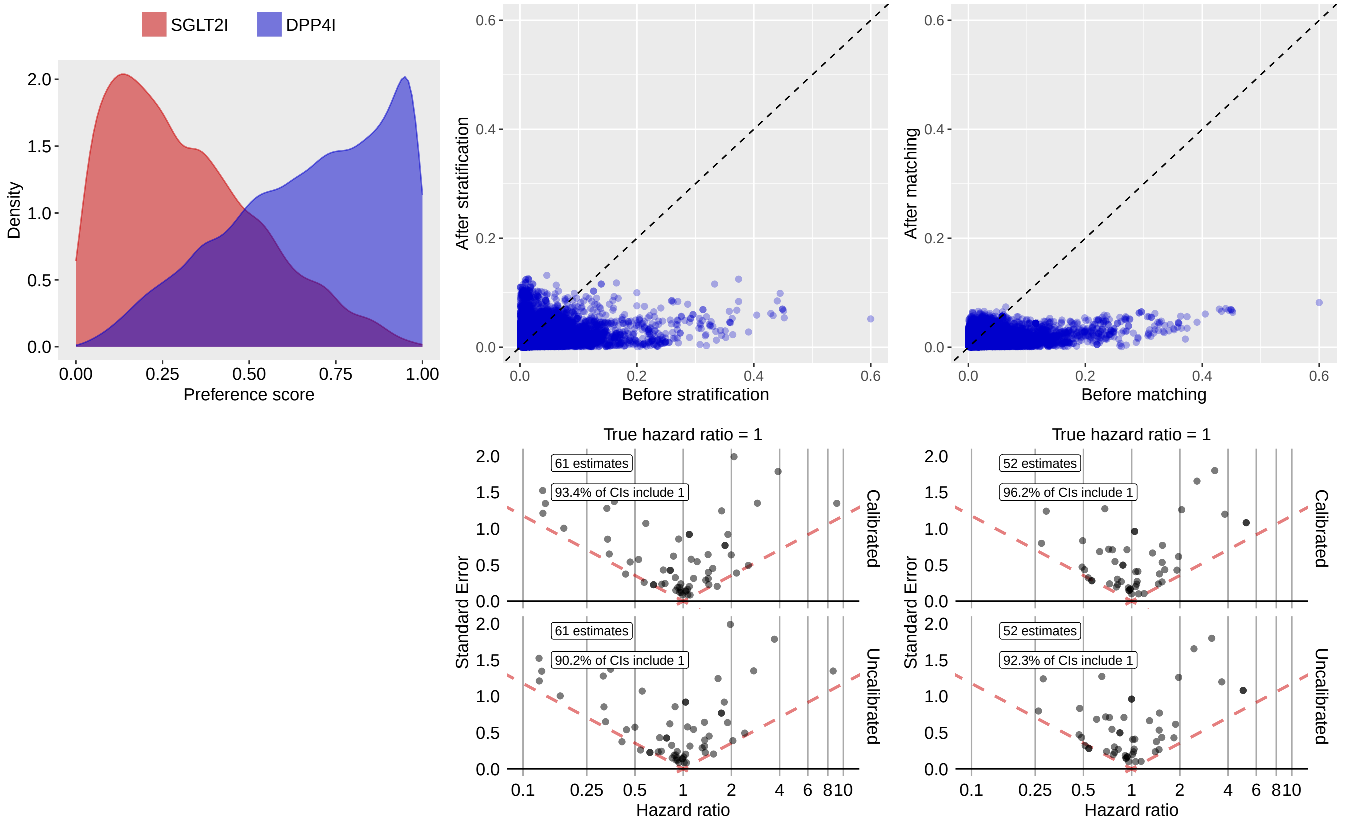

**Supplementary Table 69:** Most extreme patient characteristics balance diagnostics after stratification for SGLT2I vs DPP4I in the VA data source.

| Characteristic (total count = 23426) | Before stratification | | | After stratification | | |
|---|---|---|---|---|---|---|
| | T (%) | C (%) | SDf | T (%) | C (%) | SDf |
| measurement during day -30 through 0 days relative to index: Albumin [Mass/volume] in Urine | 1.6 | 2.2 | 0.05 | 4.3 | 2.0 | -0.13 |
| measurement during day -30 through 0 days relative to index: Thyroperoxidase Ab [Units/volume] in Serum or Plasma | 0.2 | 0.3 | 0.01 | 1.4 | 0.3 | -0.13 |
| procedure_occurrence during day -365 through 0 days relative to index: Counseling | -0.1 | 6.6 | 0.37 | 1.1 | 2.8 | 0.12 |
| measurement during day -30 through 0 days relative to index: Testosterone [Mass/volume] in Serum or Plasma | 1.3 | 1.5 | 0.01 | 3.2 | 1.4 | -0.12 |
| measurement within normal range during day -365 through 0 days relative to index: Thyroglobulin Ab [Units/volume] in Serum or Plasma | 0.2 | 0.2 | 0.01 | 1.2 | 0.2 | -0.12 |
| drug_era group during day 0 through 0 days relative to index: ANDROGENS | 1.4 | 1.2 | -0.01 | 3.0 | 1.2 | -0.12 |
| drug_era group during day 0 through 0 days relative to index: 3-oxoandrosten (4) derivatives | 1.4 | 1.2 | -0.01 | 3.0 | 1.2 | -0.12 |

| (Continued) | T (%) | C (%) | SDf | T (%) | C (%) | SDf |
|---|---|---|---|---|---|---|
| drug_era group during day 0 through 0 days relative to index: ANDROGENS AND FEMALE SEX HORMONES IN COMBINATION | 1.4 | 1.3 | -0.01 | 3.1 | 1.3 | -0.12 |
| drug_era group during day 0 through 0 days relative to index: Androgens and estrogens | 1.4 | 1.3 | -0.01 | 3.1 | 1.3 | -0.12 |
| procedure_occurrence during day -365 through 0 days relative to index: Physical therapy evaluation | -0.1 | 1.6 | 0.17 | 0.1 | 1.0 | 0.12 |

**Supplementary Table 70:** Most extreme patient characteristics balance diagnostics after matching for SGLT2I vs DPP4I in the VA data source.

| | Before matching | | | After matching | | |
|---|---|---|---|---|---|---|
| Characteristic (total count = 19952) | T (%) | C (%) | SDf | T (%) | C (%) | SDf |
| index year: 2021 | 40.0 | 14.4 | -0.60 | 28.8 | 32.6 | 0.08 |
| measurement during day -365 through 0 days relative to index: Thyroglobulin | 0.6 | 0.2 | -0.06 | 0.4 | -0.2 | -0.07 |
| condition_era group during day -365 through 0 days relative to index: Structural disorder of heart | 46.3 | 25.7 | -0.44 | 33.4 | 36.7 | 0.07 |
| condition_era group during day -365 through 0 days relative to index: Atherosclerosis of artery | 38.5 | 19.5 | -0.43 | 27.4 | 30.6 | 0.07 |
| condition_era group during day -365 through 0 days relative to index: Arteriosclerotic vascular disease | 42.4 | 22.0 | -0.45 | 30.2 | 33.4 | 0.07 |
| condition_era group during day -365 through 0 days relative to index: Vascular degeneration | 42.4 | 22.0 | -0.45 | 30.2 | 33.4 | 0.07 |
| condition_era group during day -365 through 0 days relative to index: Heart disease | 58.9 | 37.2 | -0.45 | 46.0 | 49.4 | 0.07 |
| condition_era group during day -365 through 0 days relative to index: Coronary arteriosclerosis | 41.3 | 21.0 | -0.45 | 29.1 | 32.2 | 0.07 |
| condition_era group during day -365 through 0 days relative to index: Coronary atherosclerosis | 37.5 | 18.5 | -0.43 | 26.4 | 29.4 | 0.07 |
| measurement above normal range during day -365 through 0 days relative to index: IgA [Mass/volume] in Serum or Plasma | -0.1 | 0.2 | 0.03 | -0.1 | 0.3 | 0.07 |

##### 3.4.6 GermanyDA

**Supplementary Figure 18:** Preference score distributions, propensity score stratified (middle column) and matched (right column) covariate balance and negative control error distributions (bottom row) for SGLT2I vs DPP4I in the GermanyDA data source.

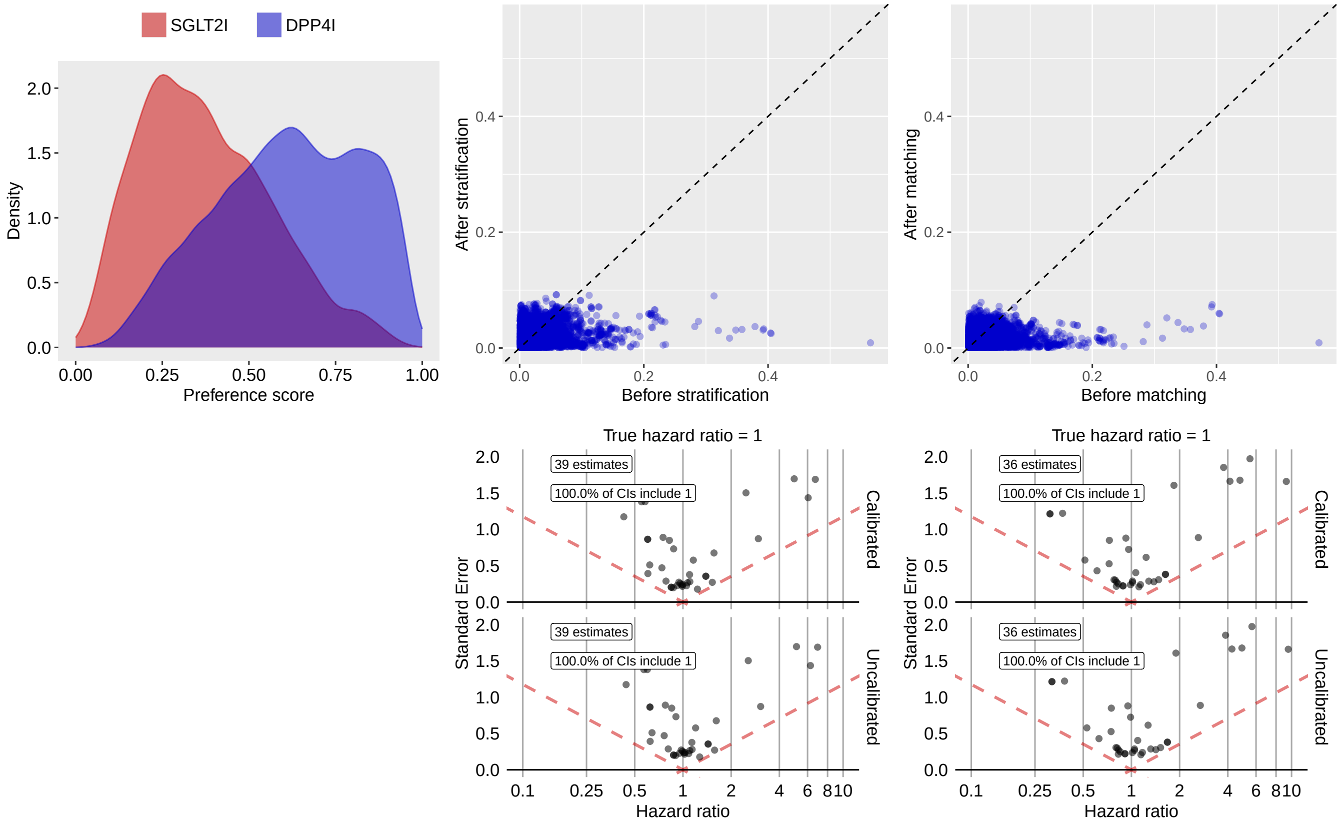

**Supplementary Table 71:** Most extreme patient characteristics balance diagnostics after stratification for SGLT2I vs DPP4I in the GermanyDA data source.

| | Before stratification | | | After stratification | | |
|---|---|---|---|---|---|---|
| Characteristic (total count = 6658) | T (%) | C (%) | SDf | T (%) | C (%) | SDf |
| drug_era group during day -365 through 0 days relative to index: flupirtine | -0.1 | 0.4 | 0.06 | 0.9 | 0.2 | -0.09 |
| drug_era group during day -365 through 0 days relative to index: Other analgesics and antipyretics | -0.1 | 0.4 | 0.06 | 0.9 | 0.2 | -0.09 |
| age group: 65 - 69 | 32.1 | 27.0 | -0.11 | 32.4 | 28.2 | -0.09 |
| index year: 2013 | 0.3 | 5.4 | 0.31 | 2.5 | 4.1 | 0.09 |
| condition_era group during day -365 through 0 days relative to index: Elevated levels of transaminase & lactic acid dehydrogenase | 0.7 | 0.4 | -0.04 | 1.2 | 0.4 | -0.09 |
| drug_era group during day -365 through 0 days relative to index: ANTITHROMBOTIC AGENTS | 47.4 | 42.5 | -0.10 | 39.6 | 43.6 | 0.08 |
| drug_era group during day -365 through 0 days relative to index: ANTITHROMBOTIC AGENTS | 47.4 | 42.5 | -0.10 | 39.6 | 43.6 | 0.08 |
| CHADS2 | 282.6 | 287.9 | 0.04 | 277.3 | 287.2 | 0.08 |
| drug_era group during day -365 through 0 days relative to index: ANALGESICS | 43.7 | 45.1 | 0.03 | 41.1 | 44.9 | 0.08 |

| (Continued) | T (%) | C (%) | SDf | T (%) | C (%) | SDf |
|---|---|---|---|---|---|---|
| drug_era group during day -365 through 0 days relative to index: BETA BLOCKING AGENTS | 59.2 | 56.5 | -0.05 | 53.1 | 56.9 | 0.08 |

**Supplementary Table 72:** Most extreme patient characteristics balance diagnostics after matching for SGLT2I vs DPP4I in the GermanyDA data source.

| | Before matching | | | After matching | | |
|---|---|---|---|---|---|---|
| Characteristic (total count = 6398) | T (%) | C (%) | SDf | T (%) | C (%) | SDf |
| condition_era group during day -365 through 0 days relative to index: Platelet disorder | 0.8 | 0.6 | -0.02 | 0.7 | 0.2 | -0.08 |
| drug_era group during day -365 through 0 days relative to index: Other viral vaccines | 14.1 | 3.2 | -0.39 | 8.7 | 10.9 | 0.07 |
| condition_era group during day -365 through 0 days relative to index: Thrombocytopenic disorder | 0.7 | 0.6 | -0.02 | 0.7 | 0.2 | -0.07 |
| condition_era group during day -365 through 0 days relative to index: Gangrenous disorder | 0.5 | 0.2 | -0.05 | 0.5 | -0.1 | -0.07 |
| drug_era group during day -365 through 0 days relative to index: SARS-CoV-2 (COVID-19) vaccine, mRNA spike protein | 14.0 | 3.2 | -0.39 | 8.7 | 10.8 | 0.07 |
| condition_era group during day -365 through 0 days relative to index: Monoarthritis | 0.2 | 0.6 | 0.07 | 0.2 | 0.6 | 0.07 |
| condition_era group during day -365 through 0 days relative to index: Primary malignant neoplasm of skin | 1.2 | 1.1 | -0.01 | 1.4 | 0.7 | -0.07 |
| condition_era group during day -30 through 0 days relative to index: Primary malignant neoplasm of soft tissues | 0.3 | 0.2 | -0.01 | 0.3 | -0.1 | -0.07 |
| measurement during day -365 through 0 days relative to index: Blood pressure taking | 0.4 | 0.3 | -0.01 | 0.4 | -0.1 | -0.07 |
| drug_era group during day 0 through 0 days relative to index: COMBINATIONS OF ANTIBACTERIALS | 1.5 | 1.7 | 0.02 | 1.6 | 0.9 | -0.07 |

##### 3.4.7 SIDIAP

**Supplementary Figure 19:** Preference score distributions, propensity score stratified (middle column) and matched (right column) covariate balance and negative control error distributions (bottom row) for SGLT2I vs DPP4I in the SIDIAP data source.

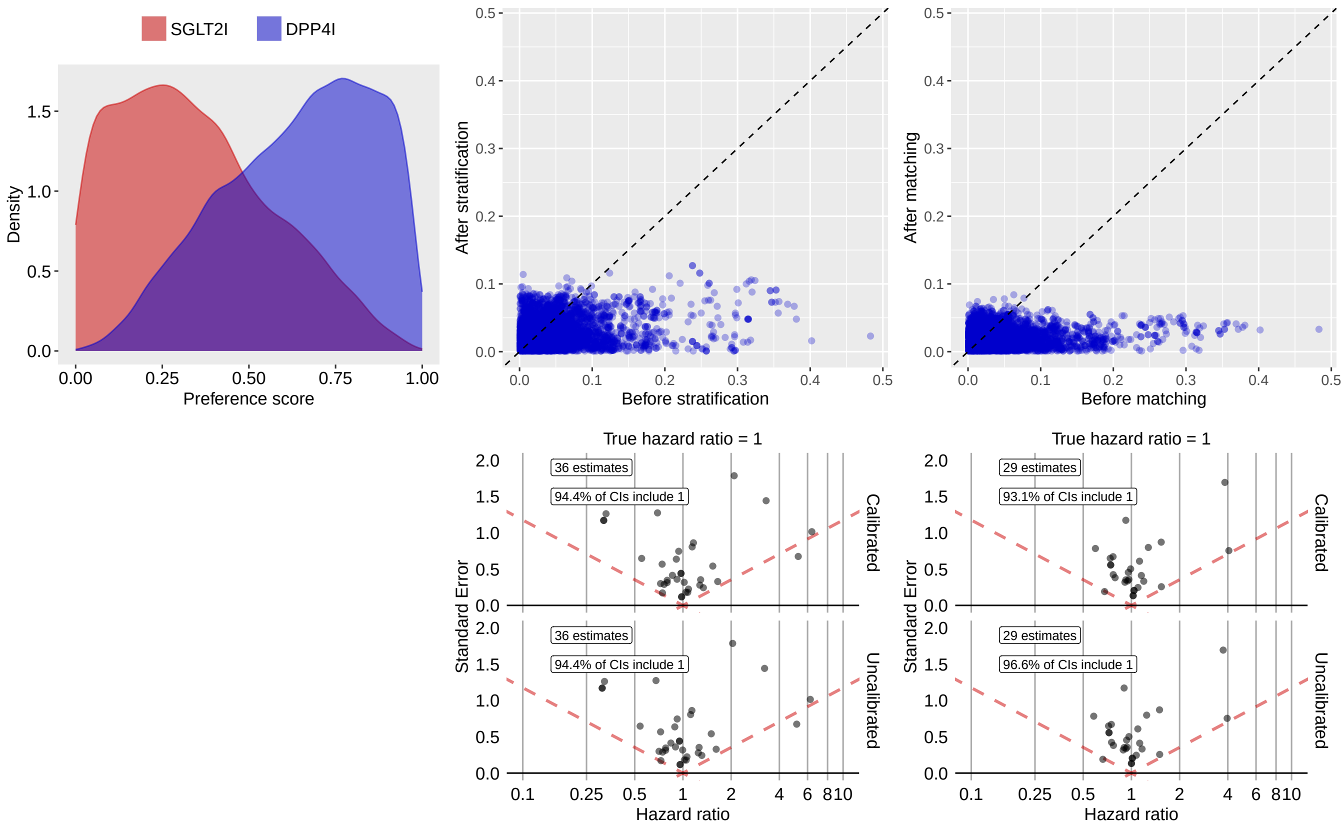

**Supplementary Table 73:** Most extreme patient characteristics balance diagnostics after stratification for SGLT2I vs DPP4I in the SIDIAP data source.

| | Before stratification | | | After stratification | | |
|---|---|---|---|---|---|---|
| Characteristic (total count = 6873) | T (%) | C (%) | SDf | T (%) | C (%) | SDf |
| drug_era group during day -365 through 0 days relative to index: ANTITHROMBOTIC AGENTS | 57.8 | 46.0 | -0.24 | 41.4 | 47.7 | 0.13 |
| drug_era group during day -365 through 0 days relative to index: ANTITHROMBOTIC AGENTS | 57.8 | 46.0 | -0.24 | 41.4 | 47.7 | 0.13 |
| drug_era group during day -30 through 0 days relative to index: ANTITHROMBOTIC AGENTS | 55.6 | 43.2 | -0.25 | 39.3 | 45.0 | 0.12 |
| drug_era group during day -30 through 0 days relative to index: ANTITHROMBOTIC AGENTS | 55.6 | 43.2 | -0.25 | 39.3 | 45.0 | 0.12 |
| Diabetes Comorbidity Severity Index (DCSI) | 147.7 | 128.4 | -0.12 | 123.6 | 141.4 | 0.12 |
| Charlson index - Romano adaptation | 257.3 | 256.4 | -0.01 | 247.7 | 267.8 | 0.11 |
| drug_era group during day -30 through 0 days relative to index: BLOOD AND BLOOD FORMING ORGANS | 59.2 | 49.0 | -0.21 | 45.1 | 50.7 | 0.11 |
| drug_era group during day 0 through 0 days relative to index: trimetazidine | 0.2 | 0.6 | 0.07 | 1.2 | 0.3 | -0.11 |
| drug_era group during day -30 through 0 days relative to index: sacubitril | 5.3 | 0.2 | -0.32 | 1.4 | 0.4 | -0.11 |

| (Continued) | T (%) | C (%) | SDf | T (%) | C (%) | SDf |
|---|---|---|---|---|---|---|
| drug_era group during day -365 through 0 days relative to index: sacubitril | 5.5 | 0.2 | -0.32 | 1.5 | 0.4 | -0.10 |

**Supplementary Table 74:** Most extreme patient characteristics balance diagnostics after matching for SGLT2I vs DPP4I in the SIDIAP data source.

| | Before matching | | | After matching | | |
|---|---|---|---|---|---|---|
| Characteristic (total count = 6471) | T (%) | C (%) | SDf | T (%) | C (%) | SDf |
| drug_era group during day -365 through 0 days relative to index: carteolol | -0.1 | 0.3 | 0.06 | -0.1 | 0.5 | 0.08 |
| procedure_occurrence during day -365 through 0 days relative to index: Injection of steroid | -0.1 | 0.4 | 0.08 | -0.1 | 0.4 | 0.08 |
| drug_era group during day -365 through 0 days relative to index: aceclofenac | 0.5 | 0.9 | 0.05 | 0.6 | -0.2 | -0.08 |
| condition_era group during day -365 through 0 days relative to index: Chronic gastritis | 0.4 | 0.6 | 0.04 | 0.4 | 1.1 | 0.08 |
| procedure_occurrence during day -30 through 0 days relative to index: Ultrasonography of Kidneys and Bladder | -0.1 | 0.1 | 0.01 | -0.1 | 0.3 | 0.07 |
| drug_era group during day -30 through 0 days relative to index: tobramycin | 0.9 | 0.8 | -0.00 | 0.9 | 0.4 | -0.07 |
| measurement during day -365 through 0 days relative to index: Thyrotropin receptor Ab [Units/volume] in Serum | 0.5 | 0.2 | -0.05 | 0.5 | -0.2 | -0.07 |
| condition_era group during day -30 through 0 days relative to index: Lesion of oral mucosa | -0.1 | 0.4 | 0.06 | -0.1 | 0.4 | 0.07 |
| condition_era group during day -365 through 0 days relative to index: Viral pneumonia | 0.1 | 0.2 | 0.01 | -0.1 | 0.4 | 0.07 |
| condition_era group during day -365 through 0 days relative to index: Viral lower respiratory infection | 0.1 | 0.2 | 0.01 | -0.1 | 0.4 | 0.07 |

#### 3.5 SGLT2I vs SU

##### 3.5.1 OptumDOD

**Supplementary Figure 20:** Preference score distributions, propensity score stratified (middle column) and matched (right column) covariate balance and negative control error distributions (bottom row) for SGLT2I vs SU in the OptumDOD data source.

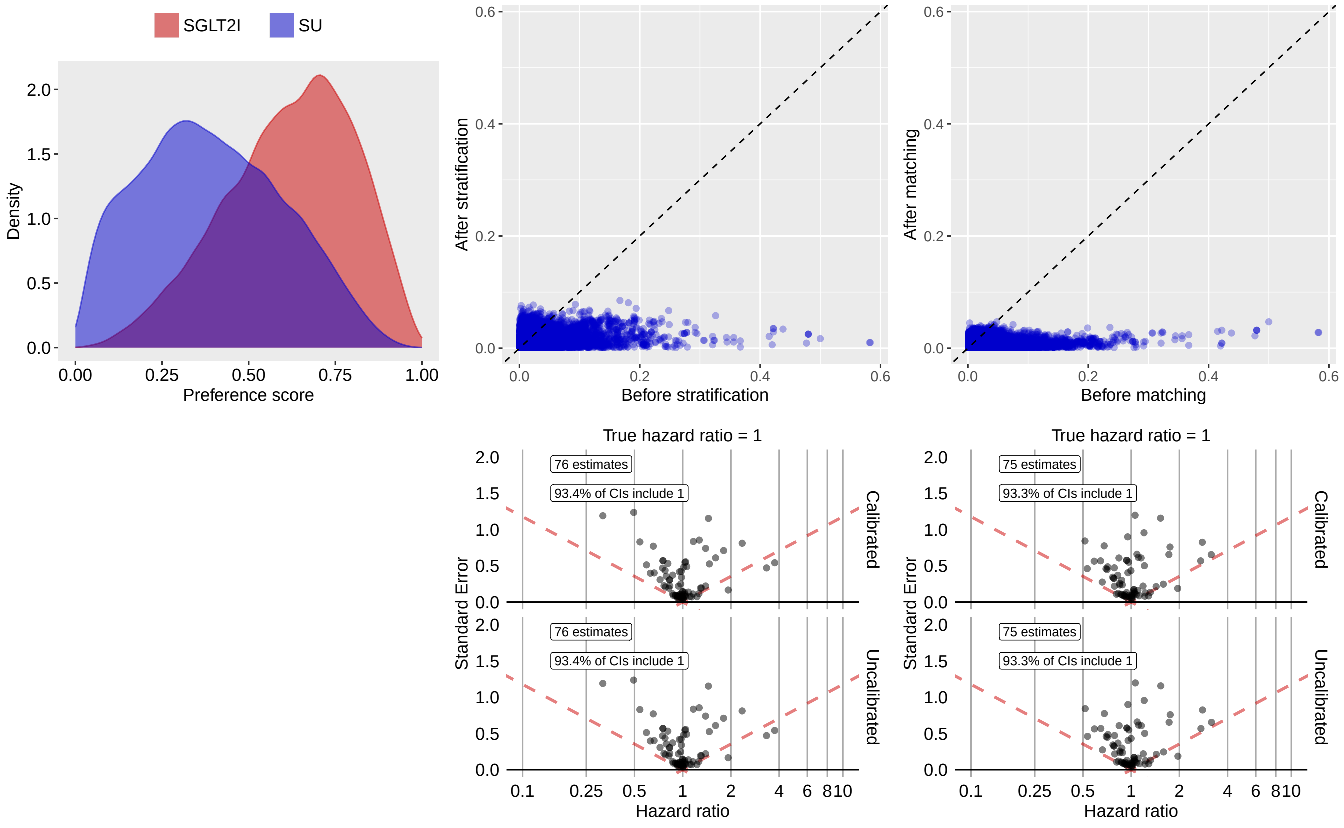

**Supplementary Table 75:** Most extreme patient characteristics balance diagnostics after stratification for SGLT2I vs SU in the OptumDOD data source.

| | Before stratification | | | After stratification | | |
|---|---|---|---|---|---|---|
| Characteristic (total count = 15708) | T (%) | C (%) | SDf | T (%) | C (%) | SDf |
| condition_era group during day -30 through 0 days relative to index: Hypertensive renal failure | 0.1 | 1.5 | -0.17 | 0.1 | 0.7 | -0.09 |
| measurement within normal range during day -365 through 0 days relative to index: Thyrotropin [Units/volume] in Serum or Plasma by Detection limit <= 0.05 mIU/L | 3.3 | 7.4 | -0.18 | 7.5 | 5.5 | 0.08 |
| observation during day -30 through 0 days relative to index: Room & Board - Semi-private Two Bed (Medical or General) - Medical/Surgical/Gyn | 0.3 | 1.1 | -0.09 | 0.3 | 0.8 | -0.08 |
| drug_era group during day -365 through 0 days relative to index: Anticholinergics | 1.6 | 1.9 | -0.02 | 2.8 | 1.7 | 0.08 |
| condition_era group during day -365 through 0 days relative to index: Closed fracture of intracapsular section of femur | 0.1 | 0.1 | -0.00 | 0.5 | 0.1 | 0.08 |
| condition_era group during day -365 through 0 days relative to index: Abdominal abscess | 0.5 | 0.4 | 0.02 | 1.0 | 0.4 | 0.07 |

| (Continued) | T (%) | C (%) | SDf | T (%) | C (%) | SDf |
|---|---|---|---|---|---|---|
| measurement during day -365 through 0 days relative to index: Thyrotropin [Units/volume] in Serum or Plasma by Detection limit $<=$ 0.05 mIU/L | 3.9 | 8.5 | -0.19 | 8.4 | 6.5 | 0.07 |
| measurement above normal range during day -365 through 0 days relative to index: Iron | 0.1 | 0.1 | -0.00 | 0.5 | 0.1 | 0.07 |
| drug_era group during day -365 through 0 days relative to index: BELLADONNA AND DERIVATIVES, PLAIN | 1.1 | 1.6 | -0.04 | 2.2 | 1.3 | 0.07 |
| observation during day -30 through 0 days relative to index: History of urinary tract infection | 0.2 | 0.3 | -0.01 | 0.8 | 0.3 | 0.07 |

**Supplementary Table 76:** Most extreme patient characteristics balance diagnostics after matching for SGLT2I vs SU in the OptumDOD data source.

| | Before matching | | | After matching | | |
|---|---|---|---|---|---|---|
| Characteristic (total count = 15497) | T (%) | C (%) | SDf | T (%) | C (%) | SDf |
| index year: 2021 | 24.3 | 6.7 | 0.50 | 21.7 | 23.7 | -0.05 |
| measurement above normal range during day -365 through 0 days relative to index: Microalbumin/Creatinine [Mass Ratio] in 24 hour Urine | 0.4 | 0.1 | 0.05 | 0.4 | 0.1 | 0.05 |
| index month: 3 | 10.2 | 10.1 | 0.00 | 10.1 | 11.5 | -0.04 |
| procedure_occurrence during day -30 through 0 days relative to index: External ocular photography with interpretation and report for documentation of medical progress (eg, close-up photography, slit lamp photography, goniophotography, stereo-photography) | 0.2 | 0.1 | 0.04 | 0.2 | 0.1 | 0.04 |
| measurement above normal range during day -365 through 0 days relative to index: Natriuretic peptide.B prohormone N-Terminal [Mass/volume] in Serum or Plasma | 0.6 | 0.1 | 0.07 | 0.5 | 0.2 | 0.04 |
| condition_era group during day -365 through 0 days relative to index: Arterial malformation | 0.1 | 0.3 | -0.04 | 0.1 | 0.2 | -0.04 |
| measurement during day -365 through 0 days relative to index: Microalbumin/Creatinine [Mass Ratio] in 24 hour Urine | 0.4 | 0.2 | 0.04 | 0.4 | 0.2 | 0.04 |
| condition_era group during day -365 through 0 days relative to index: Hypertrophic osteoarthropathy | 0.2 | 0.1 | 0.02 | 0.1 | 0.3 | -0.04 |
| drug_era group during day 0 through 0 days relative to index: pitavastatin | 0.4 | 0.1 | 0.06 | 0.4 | 0.2 | 0.04 |
| measurement during day -365 through 0 days relative to index: Hemoglobin [Mass/volume] in Urine by Automated test strip | 0.1 | 0.3 | -0.04 | 0.1 | 0.3 | -0.04 |

##### 3.5.2 MDCR

**Supplementary Figure 21:** Preference score distributions, propensity score stratified (middle column) and matched (right column) covariate balance and negative control error distributions (bottom row) for SGLT2I vs SU in the MDCR data source.

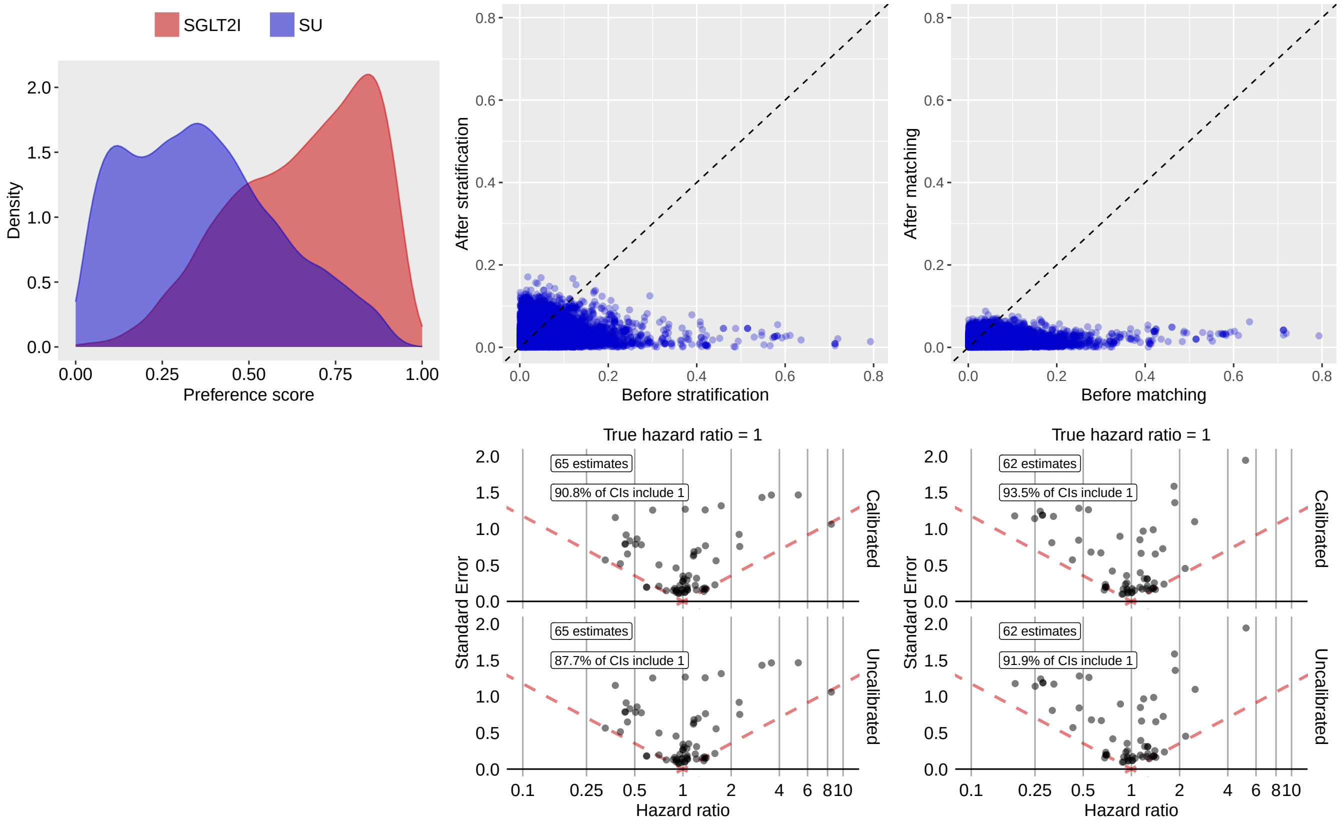

**Supplementary Table 77:** Most extreme patient characteristics balance diagnostics after stratification for SGLT2I vs SU in the MDCR data source.

| Characteristic (total count = 12974) | Before stratification | | | After stratification | | |
|---|---|---|---|---|---|---|
| | T (%) | C (%) | SDf | T (%) | C (%) | SDf |
| measurement during day -30 through 0 days relative to index: Myoglobin | -0.1 | 0.1 | -0.02 | 1.7 | 0.1 | 0.17 |
| procedure_occurrence during day -30 through 0 days relative to index: Office or other outpatient visit for the evaluation and management of a new patient, which requires these 3 key components: A comprehensive history; A comprehensive examination; Medical decision making of high complexity. Counseling and/or coordination of | 1.8 | 1.2 | 0.05 | 4.4 | 1.5 | 0.17 |
| condition_era group during day -30 through 0 days relative to index: Chronic pain | 3.0 | 1.3 | 0.12 | 6.0 | 2.6 | 0.17 |
| condition_era group during day -30 through 0 days relative to index: Benign hypertensive heart disease without congestive heart failure | 0.2 | 0.4 | -0.04 | 1.9 | 0.3 | 0.16 |
| condition_era group during day -30 through 0 days relative to index: Benign hypertensive heart disease | 0.3 | 0.5 | -0.04 | 2.0 | 0.3 | 0.15 |
| measurement during day -365 through 0 days relative to index: Triiodothyronine T3; free | 2.4 | 0.8 | 0.13 | 3.9 | 1.4 | 0.15 |

| (Continued) | T (%) | C (%) | SDf | T (%) | C (%) | SDf |
|---|---|---|---|---|---|---|
| condition_era group during day -365 through 0 days relative to index: Abnormal chest sounds | -0.1 | 0.4 | -0.07 | 1.7 | 0.2 | 0.15 |
| condition_era group during day -365 through 0 days relative to index: Lesion of eyelid | 2.5 | 1.4 | 0.08 | 4.4 | 1.9 | 0.14 |
| drug_era group during day -365 through 0 days relative to index: ANDROGENS AND FEMALE SEX HORMONES IN COMBINATION | 4.8 | 6.0 | -0.05 | 7.8 | 4.5 | 0.14 |
| drug_era group during day -365 through 0 days relative to index: Androgens and estrogens | 4.8 | 6.0 | -0.05 | 7.8 | 4.5 | 0.14 |

**Supplementary Table 78:** Most extreme patient characteristics balance diagnostics after matching for SGLT2I vs SU in the MDCR data source.

| | Before matching | | | After matching | | |
|---|---|---|---|---|---|---|
| Characteristic (total count = 12136) | T (%) | C (%) | SDf | T (%) | C (%) | SDf |
| procedure_occurrence during day -365 through 0 days relative to index: Multi-leaf collimator (MLC) device(s) for intensity modulated radiation therapy (IMRT), design and construction per IMRT plan | -0.1 | 0.3 | -0.04 | -0.2 | 0.6 | -0.09 |
| procedure_occurrence during day -365 through 0 days relative to index: Intensity modulated radiotherapy plan, including dose-volume histograms for target and critical structure partial tolerance specifications | 0.1 | 0.4 | -0.04 | -0.2 | 0.6 | -0.08 |
| measurement during day -365 through 0 days relative to index: C-peptide | 0.8 | 0.1 | 0.10 | 0.8 | 0.2 | 0.08 |
| procedure_occurrence during day -365 through 0 days relative to index: Ambulatory continuous glucose monitoring of interstitial tissue fluid via a subcutaneous sensor for a minimum of 72 hours; physician or other qualified health care professional (office) provided equipment, sensor placement, hook-up, calibration of monitor | 0.8 | 0.1 | 0.11 | 0.7 | 0.2 | 0.07 |
| measurement above normal range during day -365 through 0 days relative to index: INR in Platelet poor plasma by Coagulation assay | -0.1 | 0.3 | -0.07 | -0.2 | 0.3 | -0.07 |
| procedure_occurrence during day -365 through 0 days relative to index: Computed tomography guidance for placement of radiation therapy fields | 0.2 | 0.5 | -0.06 | 0.2 | 0.6 | -0.07 |
| measurement during day -30 through 0 days relative to index: Hemoglobin; glycosylated (A1C) by device cleared by FDA for home use | 0.5 | 0.3 | 0.03 | 0.6 | 0.2 | 0.07 |
| observation during day -30 through 0 days relative to index: Review of all medications by a prescribing practitioner or clinical pharmacist (such as, prescriptions, OTCs, herbal therapies and supplements) documented in the medical record (COA) | 0.5 | 0.2 | 0.04 | 0.4 | 1.0 | -0.07 |
| condition_era group during day -365 through 0 days relative to index: Acute otitis media | 0.4 | 0.5 | -0.01 | 0.3 | 0.8 | -0.07 |
| drug_era group during day -30 through 0 days relative to index: sacubitril | 1.5 | 0.1 | 0.16 | 1.0 | 0.4 | 0.07 |

##### 3.5.3 Open Claims

**Supplementary Figure 22:** Preference score distributions, propensity score stratified (middle column) and matched (right column) covariate balance and negative control error distributions (bottom row) for SGLT2I vs SU in the Open Claims data source.

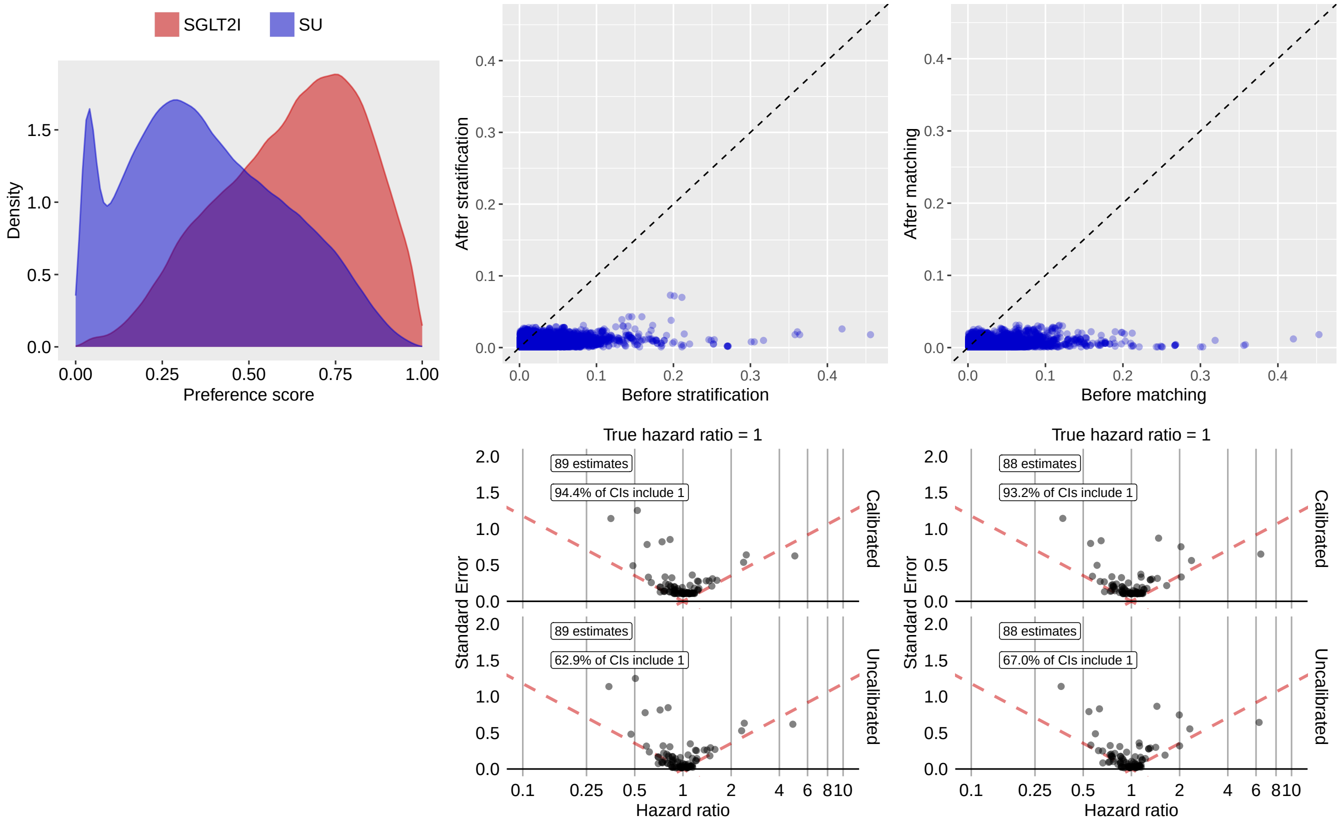

**Supplementary Table 79:** Most extreme patient characteristics balance diagnostics after stratification for SGLT2I vs SU in the Open Claims data source.

| | Before stratification | | | After stratification | | |
|---|---|---|---|---|---|---|
| Characteristic (total count = 10610) | T (%) | C (%) | SDf | T (%) | C (%) | SDf |
| drug_era group during day 0 through 0 days relative to index: sacubitril | 2.4 | 0.2 | 0.20 | 0.9 | 0.3 | 0.07 |
| drug_era group during day -30 through 0 days relative to index: sacubitril | 2.5 | 0.2 | 0.20 | 0.9 | 0.4 | 0.07 |
| drug_era group during day -365 through 0 days relative to index: sacubitril | 2.9 | 0.3 | 0.21 | 1.1 | 0.5 | 0.07 |
| drug_era group during day -365 through 0 days relative to index: icosapent ethyl | 2.0 | 0.3 | 0.16 | 0.8 | 0.5 | 0.04 |
| drug_era group during day -30 through 0 days relative to index: icosapent ethyl | 1.6 | 0.2 | 0.15 | 0.6 | 0.3 | 0.04 |
| drug_era group during day 0 through 0 days relative to index: icosapent ethyl | 1.5 | 0.2 | 0.14 | 0.6 | 0.3 | 0.04 |
| age group: 65 - 69 | 43.1 | 36.6 | 0.13 | 40.0 | 38.1 | 0.04 |
| age group: 75 - 79 | 17.5 | 25.5 | -0.20 | 22.0 | 23.6 | -0.04 |
| observation during day -365 through 0 days relative to index: Exposure to SARS-CoV-2 | 3.5 | 0.9 | 0.17 | 1.7 | 1.3 | 0.03 |

| (Continued) | T (%) | C (%) | SDf | T (%) | C (%) | SDf |
|---|---|---|---|---|---|---|
| drug_era group during day -365 through 0 days relative to index: Sulfonamides | 32.1 | 36.0 | -0.08 | 36.9 | 35.4 | 0.03 |

**Supplementary Table 80:** Most extreme patient characteristics balance diagnostics after matching for SGLT2I vs SU in the Open Claims data source.

| | Before matching | | | After matching | | |
|---|---|---|---|---|---|---|
| Characteristic (total count = 10418) | T (%) | C (%) | SDf | T (%) | C (%) | SDf |
| drug_era group during day -30 through 0 days relative to index: Aldosterone antagonists | 5.7 | 3.3 | 0.12 | 4.5 | 3.9 | 0.03 |
| condition_era group during day -365 through 0 days relative to index: Renal impairment | 10.4 | 12.5 | -0.07 | 9.9 | 9.0 | 0.03 |
| condition_era group during day -30 through 0 days relative to index: Heart failure | 4.2 | 3.0 | 0.06 | 3.2 | 2.7 | 0.03 |
| drug_era group during day -365 through 0 days relative to index: Aldosterone antagonists | 6.9 | 4.1 | 0.12 | 5.6 | 4.9 | 0.03 |
| drug_era group during day 0 through 0 days relative to index: Aldosterone antagonists | 5.4 | 3.1 | 0.12 | 4.3 | 3.7 | 0.03 |
| condition_era group during day -30 through 0 days relative to index: Abnormal cardiovascular function | 3.0 | 1.6 | 0.09 | 2.1 | 1.7 | 0.03 |
| condition_era group during day -30 through 0 days relative to index: Systolic dysfunction | 2.1 | 0.9 | 0.09 | 1.4 | 1.0 | 0.03 |
| drug_era group during day -30 through 0 days relative to index: spironolactone | 5.4 | 3.2 | 0.11 | 4.3 | 3.8 | 0.03 |
| drug_era group during day -30 through 0 days relative to index: Thiazides, combinations with psycholeptics and/or analgesics | 5.4 | 3.2 | 0.11 | 4.4 | 3.8 | 0.03 |
| condition_era group during day -30 through 0 days relative to index: Systolic heart failure | 2.1 | 0.9 | 0.10 | 1.3 | 1.0 | 0.03 |

##### 3.5.4 OptumEHR

**Supplementary Figure 23:** Preference score distributions, propensity score stratified (middle column) and matched (right column) covariate balance and negative control error distributions (bottom row) for SGLT2I vs SU in the OptumEHR data source.

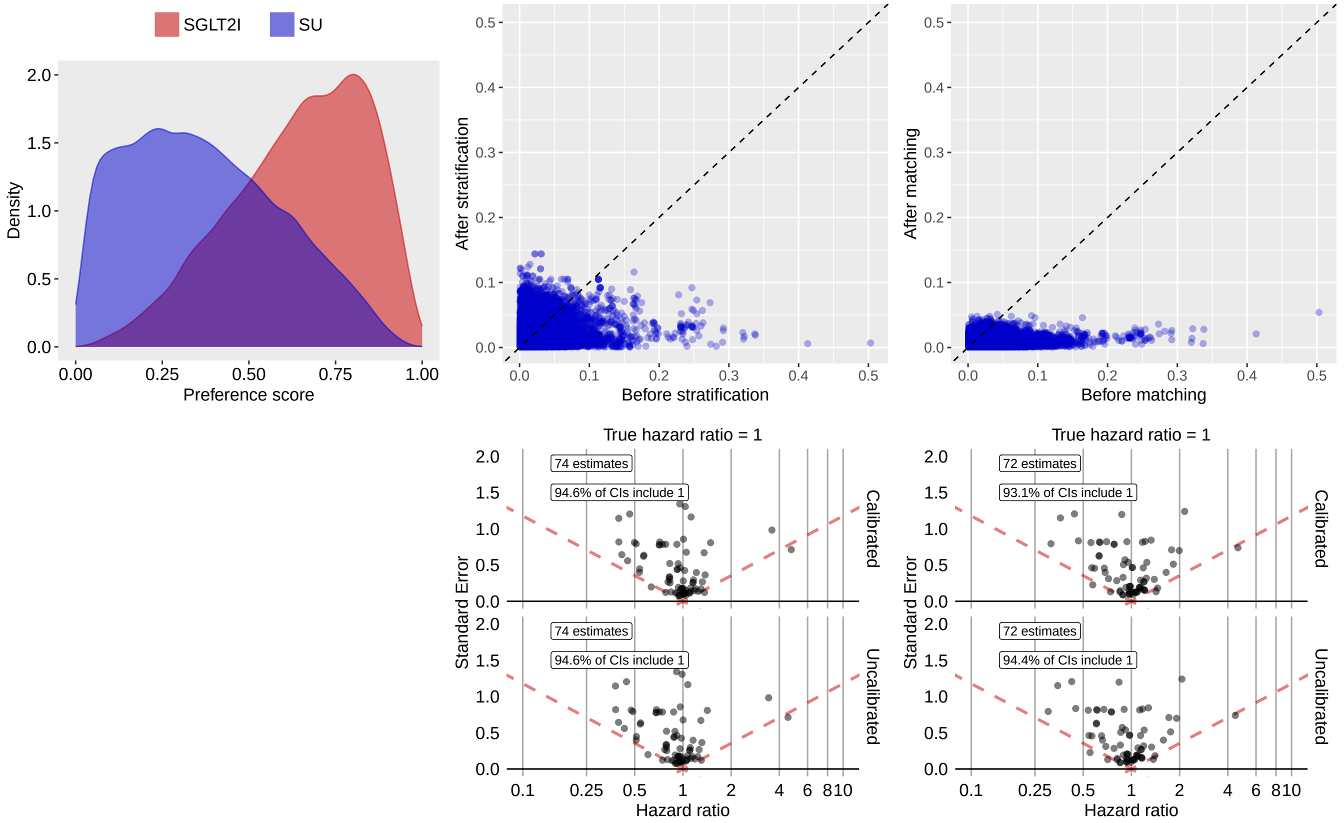

**Supplementary Table 81:** Most extreme patient characteristics balance diagnostics after stratification for SGLT2I vs SU in the OptumEHR data source.

| | Before stratification | | | After stratification | | |
|---|---|---|---|---|---|---|
| Characteristic (total count = 12519) | T (%) | C (%) | SDf | T (%) | C (%) | SDf |
| drug_era group during day -365 through 0 days relative to index: hyaluronate | 1.0 | 0.7 | 0.03 | 2.7 | 0.8 | 0.14 |
| drug_era group during day -365 through 0 days relative to index: Other drugs for disorders of the musculo-skeletal system | 1.0 | 0.7 | 0.03 | 2.7 | 0.8 | 0.14 |
| drug_era group during day -365 through 0 days relative to index: OTHER DRUGS FOR DISORDERS OF THE MUSCULO-SKELETAL SYSTEM | 1.1 | 0.9 | 0.02 | 2.9 | 0.9 | 0.14 |
| drug_era group during day -365 through 0 days relative to index: OTHER DRUGS FOR DISORDERS OF THE MUSCULO-SKELETAL SYSTEM | 1.1 | 0.9 | 0.02 | 2.9 | 0.9 | 0.14 |
| drug_era group during day 0 through 0 days relative to index: solifenacin | 0.6 | 0.5 | 0.01 | 1.8 | 0.4 | 0.13 |
| drug_era group during day -30 through 0 days relative to index: solifenacin | 0.7 | 0.6 | 0.01 | 1.8 | 0.5 | 0.12 |
| drug_era group during day -30 through 0 days relative to index: megestrol | 0.1 | 0.1 | 0.00 | 1.0 | 0.1 | 0.12 |

| (Continued) | T (%) | C (%) | SDf | T (%) | C (%) | SDf |
|---|---|---|---|---|---|---|
| drug_era group during day -30 through 0 days relative to index: Pregnadien derivatives | 0.1 | 0.1 | 0.00 | 1.0 | 0.1 | 0.12 |
| drug_era group during day -365 through 0 days relative to index: SURGICAL AIDS | 2.2 | 1.8 | 0.03 | 4.0 | 2.0 | 0.12 |
| drug_era group during day -365 through 0 days relative to index: Viscoelastic substances | 2.2 | 1.8 | 0.03 | 4.0 | 2.0 | 0.12 |

**Supplementary Table 82:** Most extreme patient characteristics balance diagnostics after matching for SGLT2I vs SU in the OptumEHR data source.

| | Before matching | | | After matching | | |
|---|---|---|---|---|---|---|
| Characteristic (total count = 12898) | T (%) | C (%) | SDf | T (%) | C (%) | SDf |
| index year: 2020 | 25.6 | 7.5 | 0.50 | 23.7 | 26.0 | -0.05 |
| measurement during day -365 through 0 days relative to index: Lipoprotein, blood; quantitation of lipoprotein particle number(s) (eg, by nuclear magnetic resonance spectroscopy), includes lipoprotein particle subclass(es), when performed | 0.3 | 0.1 | 0.04 | 0.3 | 0.1 | 0.05 |
| condition_era group during day -365 through 0 days relative to index: Psychotic disorder | 0.1 | 0.6 | -0.09 | -0.1 | 0.3 | -0.05 |
| condition_era group during day -30 through 0 days relative to index: Hyperuricemia without signs of inflammatory arthritis and tophaceous disease | 0.5 | 0.1 | 0.06 | 0.5 | 0.2 | 0.05 |
| measurement during day -365 through 0 days relative to index: Alcohol consumption screening | -0.1 | 0.1 | -0.03 | -0.1 | 0.2 | -0.05 |
| device_exposure during day -365 through 0 days relative to index: Tc-99m from non-highly enriched uranium source, full cost recovery add-on, per study dose | 0.2 | 0.1 | 0.01 | 0.1 | 0.4 | -0.05 |
| procedure_occurrence during day -365 through 0 days relative to index: Exercises education, guidance, and counseling | 0.2 | 0.2 | 0.01 | 0.2 | 0.5 | -0.05 |
| observation during day -365 through 0 days relative to index: Professional Fees - Consultation | -0.1 | 1.4 | -0.16 | -0.1 | 0.2 | -0.05 |
| drug_era group during day -365 through 0 days relative to index: evolocumab | 0.6 | 0.1 | 0.10 | 0.4 | 0.2 | 0.05 |
| condition_era group during day -365 through 0 days relative to index: Weakness of face muscles | 0.3 | 0.2 | 0.03 | 0.4 | 0.1 | 0.04 |

### 3.5.5 VA

**Supplementary Figure 24:** Preference score distributions, propensity score stratified (middle column) and matched (right column) covariate balance and negative control error distributions (bottom row) for SGLT2I vs SU in the VA data source.

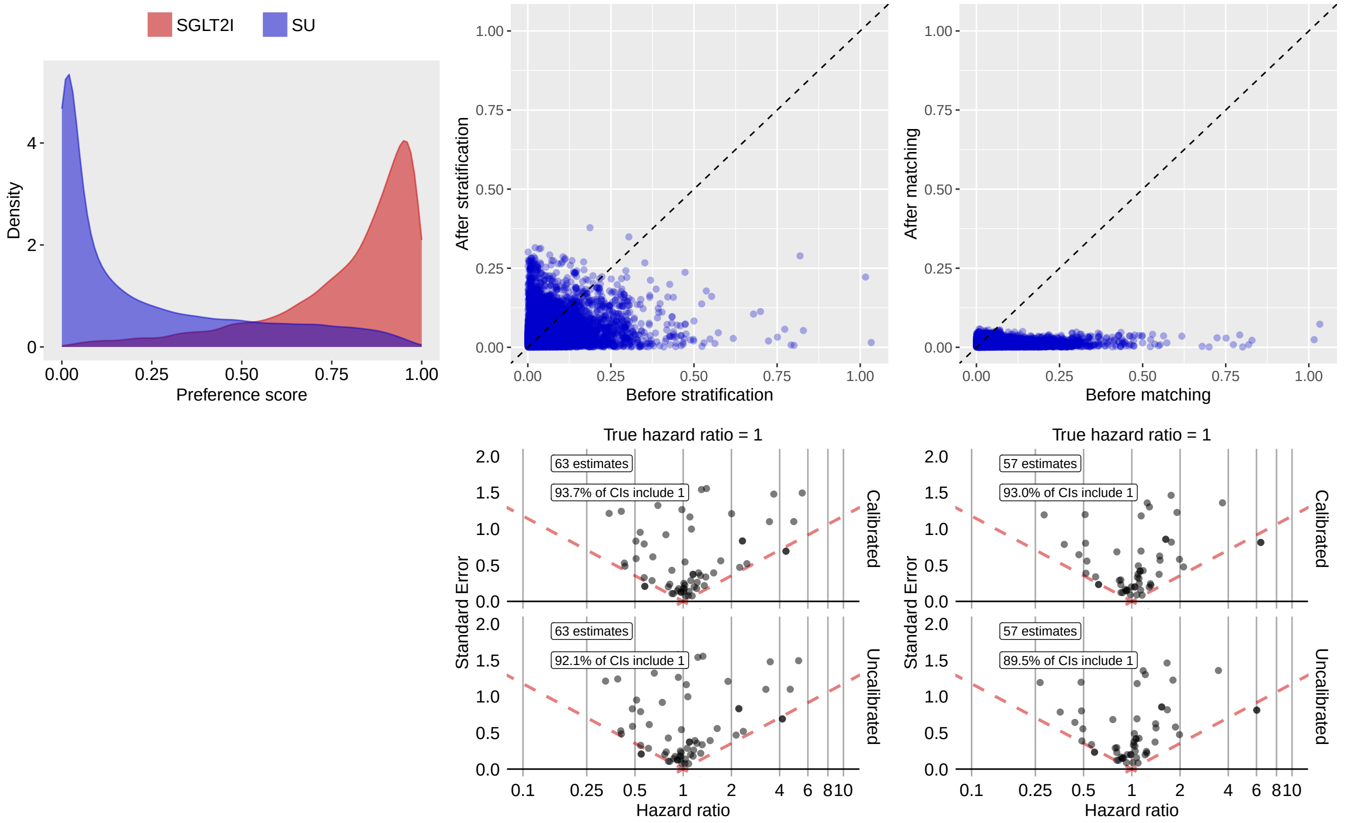

**Supplementary Table 83:** Most extreme patient characteristics balance diagnostics after stratification for SGLT2I vs SU in the VA data source.

| Characteristic (total count = 13185) | Before stratification | | | After stratification | | |
|---|---|---|---|---|---|---|
| | T (%) | C (%) | SDf | T (%) | C (%) | SDf |
| condition_era group during day -30 through 0 days relative to index: Chronic post-traumatic stress disorder | 5.3 | 1.9 | 0.19 | 16.6 | 5.1 | 0.38 |
| condition_era group during day -365 through 0 days relative to index: Chronic post-traumatic stress disorder | 11.7 | 3.7 | 0.30 | 23.0 | 10.2 | 0.35 |
| condition_era group during day -365 through 0 days relative to index: Talipes | 1.0 | 0.8 | 0.02 | 6.9 | 0.9 | 0.32 |
| condition_era group during day -30 through 0 days relative to index: Deformity of foot | 1.0 | 0.6 | 0.04 | 6.4 | 0.7 | 0.31 |
| condition_era group during day -30 through 0 days relative to index: Deformity of lower limb | 1.0 | 0.7 | 0.03 | 6.4 | 0.7 | 0.31 |
| measurement during day -365 through 0 days relative to index: Hemoglobin A1c/Hemoglobin.total in Blood | 94.4 | 94.4 | -0.00 | 88.2 | 96.2 | -0.30 |
| condition_era group during day -30 through 0 days relative to index: Posttraumatic stress disorder | 7.0 | 5.4 | 0.07 | 17.7 | 8.0 | 0.29 |
| observation during day -365 through 0 days relative to index: Requires influenza virus vaccination | -0.1 | 25.2 | -0.82 | 0.7 | 5.9 | -0.29 |

| (Continued) | T (%) | C (%) | SDf | T (%) | C (%) | SDf |
|---|---|---|---|---|---|---|
| measurement during day -365 through 0 days relative to index: Hemoglobin [Mass/volume] in Blood | 86.5 | 82.7 | 0.11 | 77.5 | 88.2 | -0.29 |
| measurement during day -30 through 0 days relative to index: Thyroperoxidase Ab [Units/volume] in Serum or Plasma | 0.2 | 0.1 | 0.02 | 4.5 | 0.2 | 0.29 |

**Supplementary Table 84:** Most extreme patient characteristics balance diagnostics after matching for SGLT2I vs SU in the VA data source.

| | Before matching | | | After matching | | |
|---|---|---|---|---|---|---|
| Characteristic (total count = 12365) | T (%) | C (%) | SDf | T (%) | C (%) | SDf |
| index year: 2021 | 40.0 | 2.5 | 1.03 | 30.3 | 33.7 | -0.07 |
| procedure_occurrence during day -365 through 0 days relative to index: Psychotherapy for crisis; first 60 minutes | -0.1 | 0.1 | 0.01 | -0.2 | 0.4 | -0.06 |
| measurement above normal range during day -365 through 0 days relative to index: Variant lymphocytes/100 leukocytes in Blood by Manual count | 0.2 | 0.2 | -0.01 | -0.2 | 0.3 | -0.06 |
| measurement during day -365 through 0 days relative to index: Character of Urine | -0.1 | 0.3 | -0.07 | -0.2 | 0.2 | -0.06 |
| measurement during day -365 through 0 days relative to index: Mycobacterium tuberculosis tuberculin stimulated gamma interferon/Mitogen stimulated gamma interferon [Units/volume] in control Blood | 0.4 | 0.2 | 0.04 | -0.2 | 0.5 | -0.06 |
| condition_era group during day -30 through 0 days relative to index: Cerebral degeneration presenting primarily with dementia | -0.1 | 0.3 | -0.07 | -0.2 | 0.2 | -0.05 |
| device_exposure during day -365 through 0 days relative to index: Dme delivery, set up, and/or dispensing service component of another hcpcs code | 0.5 | 0.1 | 0.07 | 0.4 | 0.2 | 0.05 |
| measurement during day -365 through 0 days relative to index: Testosterone Free/Testosterone.total in Serum or Plasma | 0.4 | 0.3 | 0.03 | 0.5 | 0.2 | 0.05 |
| measurement above normal range during day -365 through 0 days relative to index: IgA [Mass/volume] in Serum or Plasma | -0.1 | 0.1 | -0.02 | -0.2 | 0.2 | -0.05 |
| condition_era group during day -30 through 0 days relative to index: Cerebral degeneration | -0.1 | 0.3 | -0.07 | -0.2 | 0.2 | -0.05 |

##### 3.5.6 GermanyDA

**Supplementary Figure 25:** Preference score distributions, propensity score stratified (middle column) and matched (right column) covariate balance and negative control error distributions (bottom row) for SGLT2I vs SU in the GermanyDA data source.

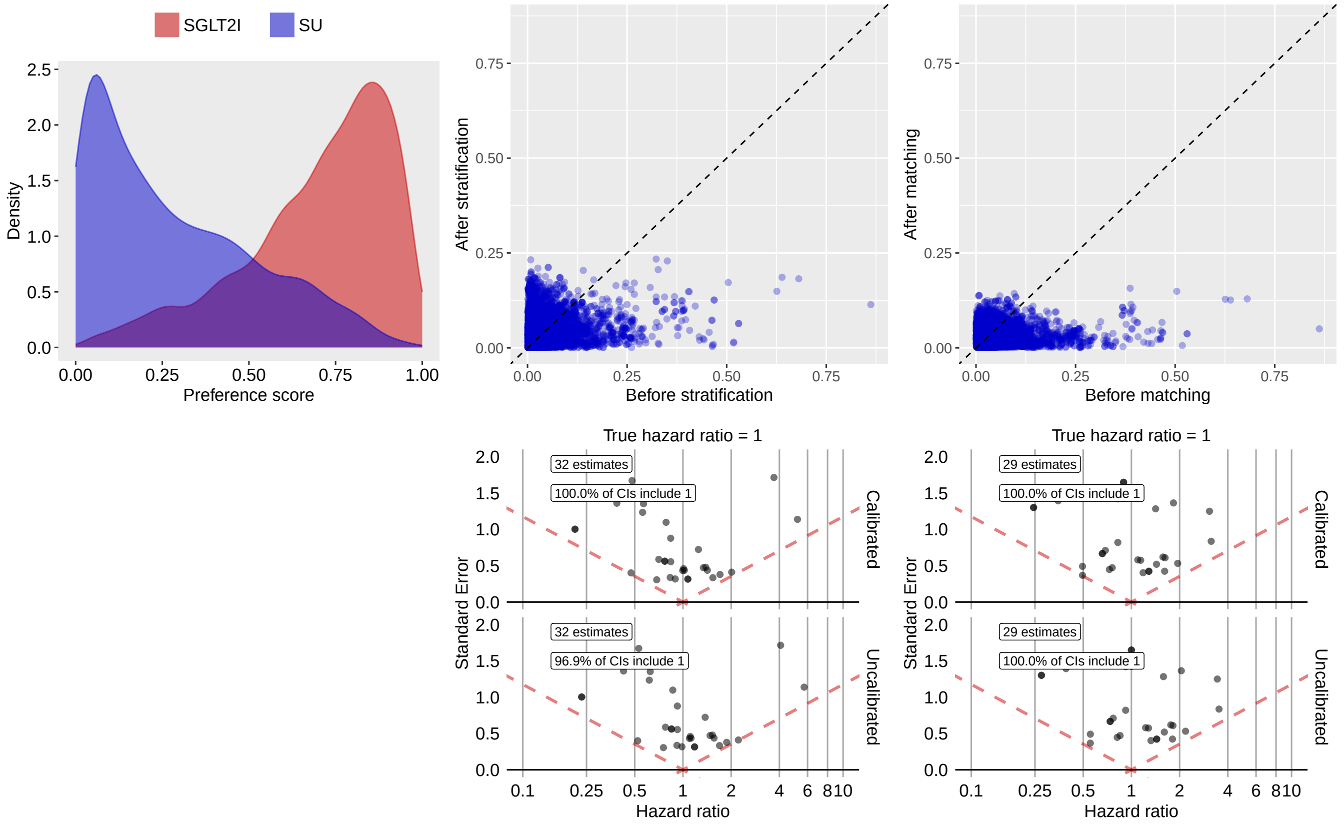

**Supplementary Table 85:** Most extreme patient characteristics balance diagnostics after stratification for SGLT2I vs SU in the GermanyDA data source.

| | Before stratification | | | After stratification | | |
|---|---|---|---|---|---|---|
| Characteristic (total count = 7041) | T (%) | C (%) | SDf | T (%) | C (%) | SDf |
| drug_era group during day 0 through 0 days relative to index: rosuvastatin | 5.1 | -0.1 | 0.32 | 3.8 | 0.4 | 0.23 |
| drug_era group during day -30 through 0 days relative to index: albuterol | 1.1 | 1.1 | -0.01 | 0.9 | 4.7 | -0.23 |
| drug_era group during day -365 through 0 days relative to index: rosuvastatin | 6.0 | -0.1 | 0.35 | 4.5 | 0.8 | 0.23 |
| condition_era group during day -30 through 0 days relative to index: Inflammation of skin and/or subcutaneous tissue | 1.2 | 1.8 | -0.05 | 1.2 | 4.7 | -0.21 |
| condition_era group during day -30 through 0 days relative to index: Inflammatory dermatosis | 1.2 | 1.8 | -0.05 | 1.2 | 4.7 | -0.21 |
| condition_era group during day -365 through 0 days relative to index: Kidney stone | 0.7 | 0.9 | -0.03 | 0.6 | 3.6 | -0.21 |
| drug_era group during day -30 through 0 days relative to index: rosuvastatin | 5.3 | -0.1 | 0.33 | 4.0 | 0.8 | 0.21 |
| measurement during day -30 through 0 days relative to index: Cholesterol [Mass/volume] in Body fluid | 31.5 | 25.2 | 0.14 | 30.6 | 40.3 | -0.20 |

| (Continued) | T (%) | C (%) | SDf | T (%) | C (%) | SDf |
|---|---|---|---|---|---|---|
| drug_era group during day -30 through 0 days relative to index: ADRENERGICS FOR SYSTEMIC USE | 1.6 | 1.7 | -0.01 | 1.4 | 4.9 | -0.20 |
| drug_era group during day -30 through 0 days relative to index: Selective beta-2-adrenoreceptor agonists | 1.6 | 1.7 | -0.01 | 1.4 | 4.9 | -0.20 |

**Supplementary Table 86:** Most extreme patient characteristics balance diagnostics after matching for SGLT2I vs SU in the GermanyDA data source.

| | Before matching | | | After matching | | |
|---|---|---|---|---|---|---|
| Characteristic (total count = 5519) | T (%) | C (%) | SDf | T (%) | C (%) | SDf |
| index year: 2019 | 13.1 | 2.8 | 0.39 | 7.1 | 11.6 | -0.16 |
| index year: 2020 | 19.2 | 3.7 | 0.50 | 10.4 | 15.4 | -0.15 |
| age group: 65 - 69 | 32.1 | 27.7 | 0.10 | 33.3 | 26.8 | 0.14 |
| condition_era group during day -365 through 0 days relative to index: Conjunctivitis | 1.3 | 1.4 | -0.01 | 2.1 | 0.5 | 0.14 |
| condition_era group during day -365 through 0 days relative to index: Inflammation of orbit | 1.3 | 1.4 | -0.01 | 2.1 | 0.5 | 0.14 |
| condition_era group during day -30 through 0 days relative to index: Open wound | 0.5 | 0.2 | 0.06 | 1.1 | -0.2 | 0.14 |
| condition_era group during day -365 through 0 days relative to index: Inflammatory disorder of the eye | 1.4 | 1.5 | -0.01 | 2.1 | 0.6 | 0.14 |
| drug_era group during day -365 through 0 days relative to index: molsidomine | 1.1 | 2.4 | -0.10 | -0.6 | 1.7 | -0.13 |
| drug_era group during day -365 through 0 days relative to index: Other vasodilators used in cardiac diseases | 1.1 | 2.4 | -0.10 | -0.6 | 1.7 | -0.13 |
| drug_era group during day -365 through 0 days relative to index: atorvastatin | 28.9 | 4.8 | 0.68 | 8.9 | 12.9 | -0.13 |

##### 3.5.7 SIDIAP

**Supplementary Figure 26:** Preference score distributions, propensity score stratified (middle column) and matched (right column) covariate balance and negative control error distributions (bottom row) for SGLT2I vs SU in the SIDIAP data source.

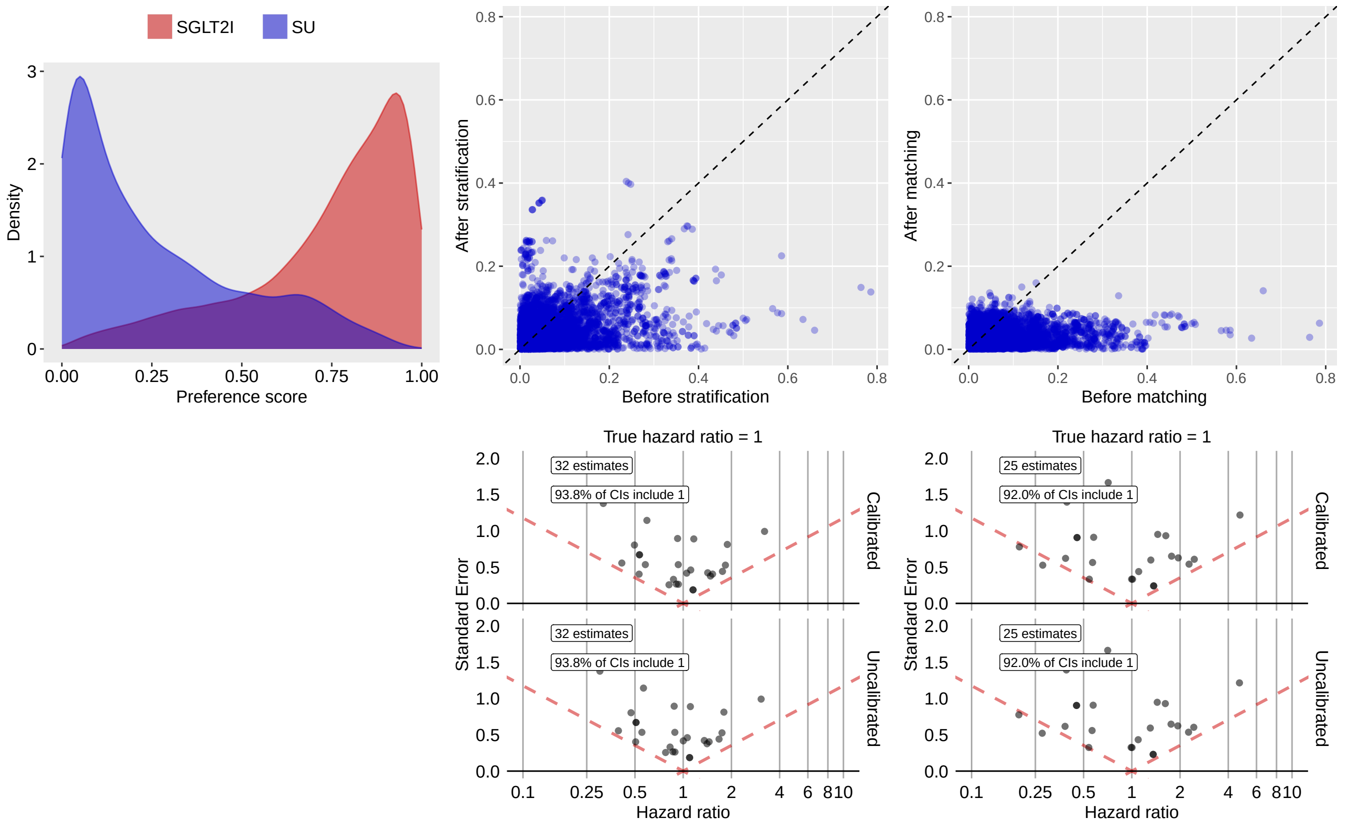

**Supplementary Table 87:** Most extreme patient characteristics balance diagnostics after stratification for SGLT2I vs SU in the SIDIAP data source.

| | Before stratification | | | After stratification | | |
|---|---|---|---|---|---|---|
| Characteristic (total count = 7396) | T (%) | C (%) | SDf | T (%) | C (%) | SDf |
| drug_era group during day 0 through 0 days relative to index: apixaban | 3.2 | 0.1 | 0.24 | 2.2 | 12.6 | -0.40 |
| drug_era group during day -30 through 0 days relative to index: apixaban | 3.3 | 0.1 | 0.24 | 2.2 | 12.6 | -0.40 |
| drug_era group during day -365 through 0 days relative to index: apixaban | 3.5 | 0.2 | 0.25 | 2.4 | 12.7 | -0.40 |
| drug_era group during day 0 through 0 days relative to index: digoxin | 3.3 | 4.2 | -0.05 | 3.0 | 12.5 | -0.36 |
| drug_era group during day 0 through 0 days relative to index: CARDIAC GLYCOSIDES | 3.3 | 4.2 | -0.05 | 3.0 | 12.5 | -0.36 |
| drug_era group during day 0 through 0 days relative to index: Digitalis glycosides | 3.3 | 4.2 | -0.05 | 3.0 | 12.5 | -0.36 |
| drug_era group during day -30 through 0 days relative to index: CARDIAC GLYCOSIDES | 3.6 | 4.4 | -0.04 | 3.2 | 12.6 | -0.35 |
| drug_era group during day -30 through 0 days relative to index: Digitalis glycosides | 3.6 | 4.4 | -0.04 | 3.2 | 12.6 | -0.35 |

| (Continued) | T (%) | C (%) | SDf | T (%) | C (%) | SDf |
|---|---|---|---|---|---|---|
| drug_era group during day -30 through 0 days relative to index: digoxin | 3.6 | 4.4 | -0.04 | 3.2 | 12.6 | -0.35 |
| drug_era group during day -365 through 0 days relative to index: CARDIAC GLYCOSIDES | 4.0 | 4.6 | -0.03 | 3.6 | 12.7 | -0.34 |

**Supplementary Table 88:** Most extreme patient characteristics balance diagnostics after matching for SGLT2I vs SU in the SIDIAP data source.

| | Before matching | | | After matching | | |
|---|---|---|---|---|---|---|
| Characteristic (total count = 6066) | T (%) | C (%) | SDf | T (%) | C (%) | SDf |
| procedure_occurrence during day -365 through 0 days relative to index: Replacement of Right Lens with Synthetic Substitute, Percutaneous Approach | 1.7 | 0.2 | 0.15 | -0.5 | 1.8 | -0.16 |
| age group: 65 - 69 | 37.2 | 31.6 | 0.12 | 44.1 | 36.8 | 0.15 |
| index year: 2019 | 22.1 | 1.8 | 0.66 | 10.9 | 15.7 | -0.14 |
| drug_era group during day -365 through 0 days relative to index: Antihistamines for topical use | 1.4 | 0.9 | 0.05 | 1.8 | 0.4 | 0.14 |
| Charlson index - Romano adaptation | 257.3 | 204.7 | 0.34 | 215.7 | 236.4 | -0.13 |
| drug_era group during day -365 through 0 days relative to index: pyrilamine | 1.3 | 0.8 | 0.05 | 1.6 | 0.4 | 0.13 |
| drug_era group during day -365 through 0 days relative to index: Substituted ethylene diamines | 1.3 | 0.8 | 0.05 | 1.6 | 0.4 | 0.13 |
| condition_era group during day -365 through 0 days relative to index: Movement disorder | 0.3 | 1.1 | -0.09 | -0.5 | 1.1 | -0.12 |
| condition_era group during day -365 through 0 days relative to index: System disorder of the nervous system | 0.3 | 1.1 | -0.09 | -0.5 | 1.1 | -0.12 |
| observation during day -365 through 0 days relative to index: Cares for dependent relative at home | 0.3 | 1.1 | -0.10 | -0.5 | 1.0 | -0.12 |

#### 3.6 DPP4I vs SU

##### 3.6.1 CCAE

**Supplementary Figure 27:** Preference score distributions, propensity score stratified (middle column) and matched (right column) covariate balance and negative control error distributions (bottom row) for DPP4I vs SU in the CCAE data source.

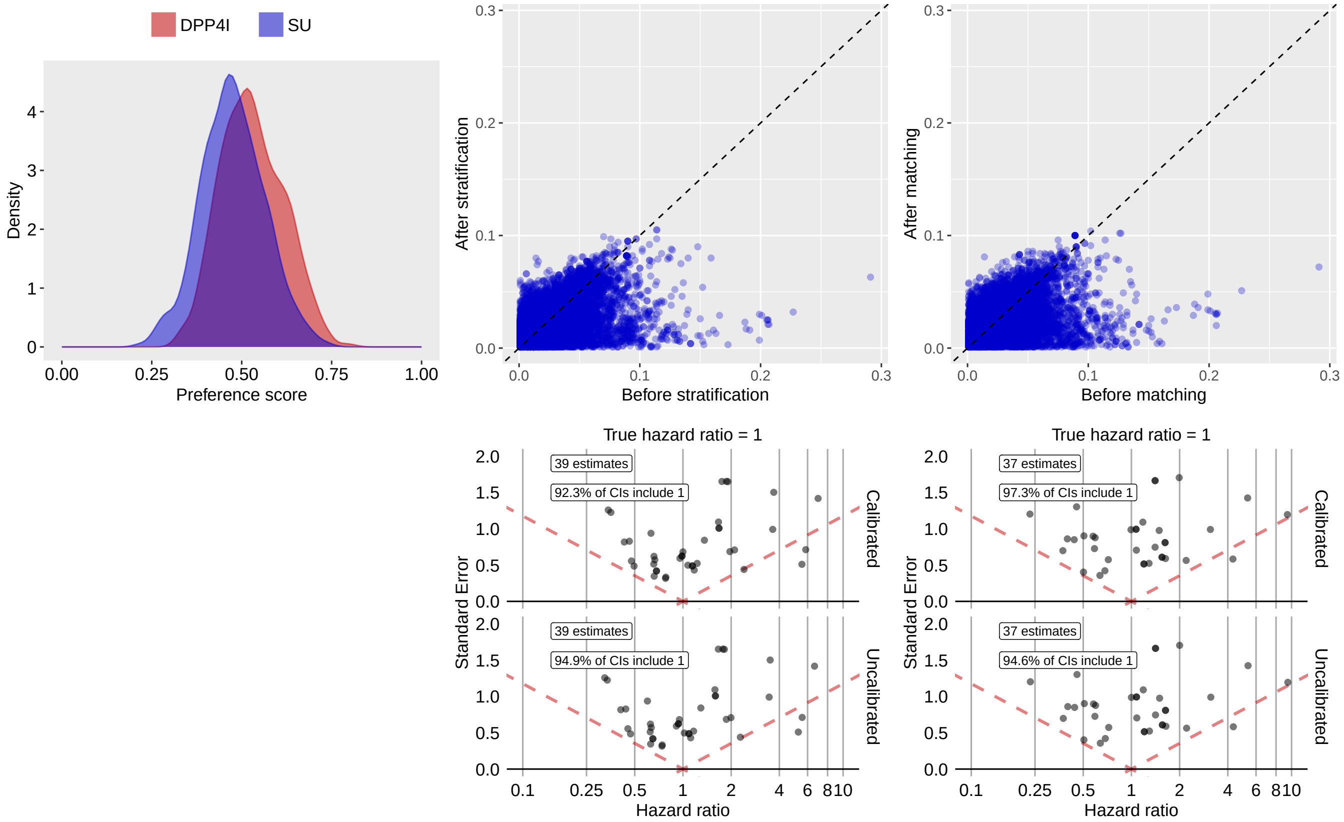

**Supplementary Table 89:** Most extreme patient characteristics balance diagnostics after stratification for DPP4I vs SU in the CCAE data source.

| | Before stratification | | | After stratification | | |
|---|---|---|---|---|---|---|
| Characteristic (total count = 15535) | T (%) | C (%) | SDf | T (%) | C (%) | SDf |
| measurement during day -365 through 0 days relative to index: Hemoglobin A1c/Hemoglobin.total in Blood by calculation | -0.2 | 0.9 | -0.11 | -0.2 | 0.8 | -0.10 |
| measurement above normal range during day -365 through 0 days relative to index: Hemoglobin A1c/Hemoglobin.total in Blood by calculation | -0.2 | 0.9 | -0.11 | -0.2 | 0.8 | -0.10 |
| drug_era group during day -30 through 0 days relative to index: metaxalone | 0.4 | -0.1 | 0.07 | 0.6 | -0.1 | 0.10 |
| drug_era group during day -365 through 0 days relative to index: dexlansoprazole | 0.9 | -0.1 | 0.11 | 0.8 | -0.1 | 0.10 |
| condition_era group during day -365 through 0 days relative to index: Atherosclerosis of artery of lower limb | 0.7 | -0.1 | 0.10 | 0.7 | -0.1 | 0.10 |
| condition_era group during day -365 through 0 days relative to index: Arteriosclerosis of artery of lower extremity | 0.7 | -0.1 | 0.10 | 0.7 | -0.1 | 0.10 |

| (Continued) | T (%) | C (%) | SDf | T (%) | C (%) | SDf |
|---|---|---|---|---|---|---|
| procedure_occurrence during day -30 through 0 days relative to index: Application of a modality to 1 or more areas; hot or cold packs | 0.7 | 0.2 | 0.08 | 0.9 | 0.2 | 0.10 |
| condition_era group during day -365 through 0 days relative to index: Pain in bilateral legs | 0.5 | -0.1 | 0.09 | 0.5 | -0.1 | 0.10 |
| condition_era group during day -365 through 0 days relative to index: Intermittent claudication of right lower limb co-occurrent and due to atherosclerosis | 0.5 | -0.1 | 0.09 | 0.5 | -0.1 | 0.10 |
| condition_era group during day -365 through 0 days relative to index: Intermittent claudication of bilateral lower limbs co-occurrent and due to atherosclerosis | 0.5 | -0.1 | 0.09 | 0.5 | -0.1 | 0.10 |

**Supplementary Table 90:** Most extreme patient characteristics balance diagnostics after matching for DPP4I vs SU in the CCAE data source.

| | Before matching | | | After matching | | |
|---|---|---|---|---|---|---|
| Characteristic (total count = 14577) | T (%) | C (%) | SDf | T (%) | C (%) | SDf |
| condition_era group during day -365 through 0 days relative to index: Anal polyp | 1.6 | 0.5 | 0.10 | 1.6 | 0.6 | 0.10 |
| drug_era group during day -365 through 0 days relative to index: nebivolol | 2.2 | 0.7 | 0.13 | 2.1 | 0.9 | 0.10 |
| drug_era group during day -30 through 0 days relative to index: nebivolol | 1.9 | 0.5 | 0.13 | 1.8 | 0.7 | 0.10 |
| condition_era group during day -365 through 0 days relative to index: Residual hemorrhoidal skin tags | 0.5 | -0.1 | 0.09 | 0.6 | -0.1 | 0.10 |
| condition_era group during day -365 through 0 days relative to index: Neoplasm of soft tissues of perineum | 0.5 | -0.1 | 0.09 | 0.6 | -0.1 | 0.10 |
| condition_era group during day -365 through 0 days relative to index: Skin tag | 0.5 | -0.1 | 0.09 | 0.6 | -0.1 | 0.10 |
| condition_era group during day -365 through 0 days relative to index: Anal skin tag | 0.5 | -0.1 | 0.09 | 0.6 | -0.1 | 0.10 |
| condition_era group during day -365 through 0 days relative to index: Neoplasm of skin of perineum | 0.5 | -0.1 | 0.09 | 0.6 | -0.1 | 0.10 |
| condition_era group during day -365 through 0 days relative to index: Benign neoplasm of soft tissues of perineum | 0.5 | -0.1 | 0.09 | 0.6 | -0.1 | 0.10 |
| condition_era group during day -365 through 0 days relative to index: Benign neoplasm of skin of perineum | 0.5 | -0.1 | 0.09 | 0.6 | -0.1 | 0.10 |

##### 3.6.2 OptumDOD

**Supplementary Figure 28:** Preference score distributions, propensity score stratified (middle column) and matched (right column) covariate balance and negative control error distributions (bottom row) for DPP4I vs SU in the OptumDOD data source.

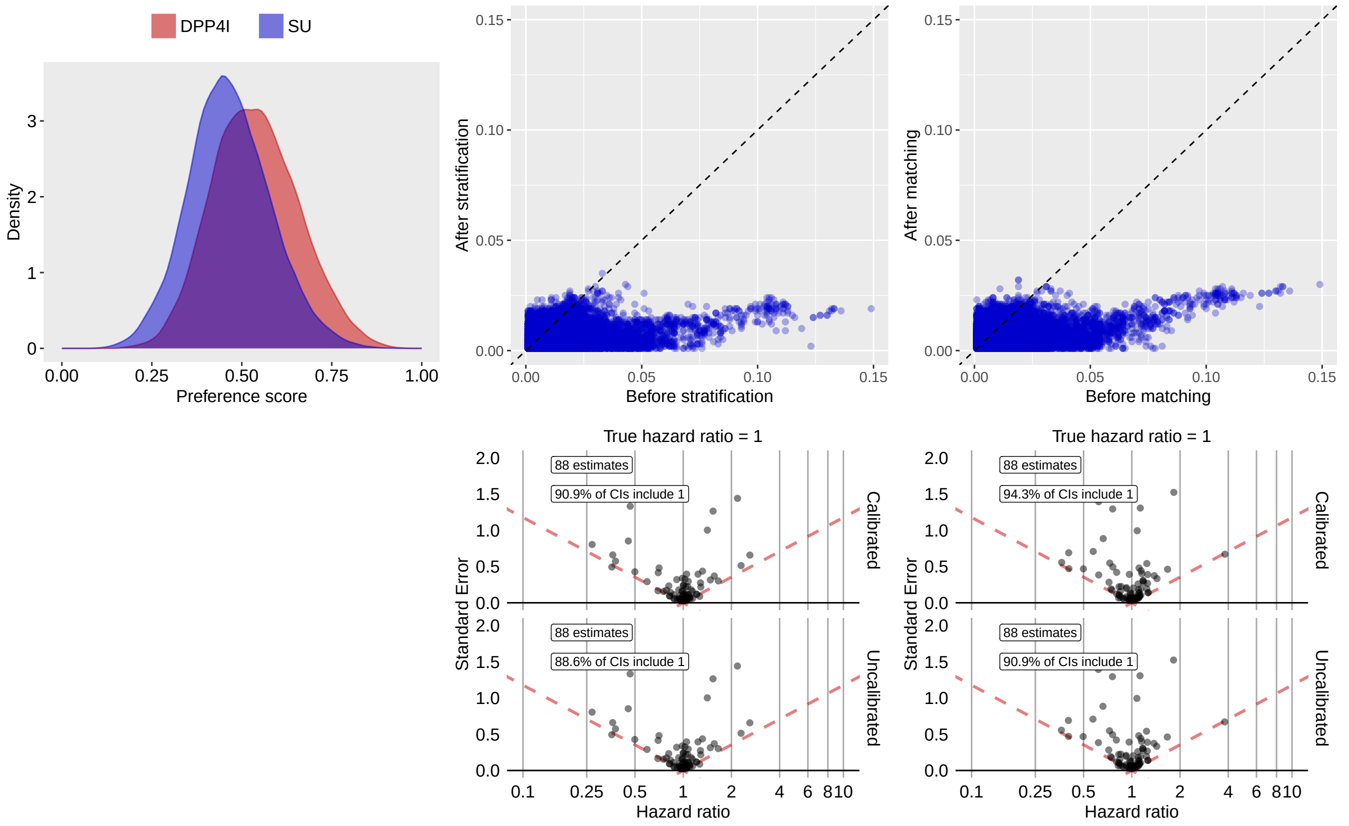

**Supplementary Table 91:** Most extreme patient characteristics balance diagnostics after stratification for DPP4I vs SU in the OptumDOD data source.

| Characteristic (total count = 16668) | Before stratification | | | After stratification | | |
|---|---|---|---|---|---|---|
| | T (%) | C (%) | SDf | T (%) | C (%) | SDf |
| measurement within normal range during day -365 through 0 days relative to index: Urinalysis; qualitative or semiquantitative, except immunoassays | 0.2 | 0.1 | 0.03 | 0.2 | 0.1 | 0.04 |
| procedure_occurrence during day -30 through 0 days relative to index: Computed tomography, pelvis; without contrast material | 0.1 | 0.2 | -0.04 | 0.1 | 0.2 | -0.03 |
| procedure_occurrence during day -30 through 0 days relative to index: Computed tomography, abdomen; without contrast material | 0.1 | 0.2 | -0.04 | 0.1 | 0.2 | -0.03 |
| measurement above normal range during day -365 through 0 days relative to index: Specific gravity of Urine by Refractometry automated | 0.1 | 0.2 | -0.03 | 0.1 | 0.2 | -0.03 |
| measurement above normal range during day -365 through 0 days relative to index: Microalbumin/Creatinine panel in random Urine | 0.1 | 0.2 | -0.03 | 0.1 | 0.2 | -0.03 |
| measurement during day -365 through 0 days relative to index: Hemoglobin [Mass/volume] in Urine by Automated test strip | 0.2 | 0.3 | -0.03 | 0.2 | 0.3 | -0.03 |

| (Continued) | T (%) | C (%) | SDf | T (%) | C (%) | SDf |
|---|---|---|---|---|---|---|
| measurement within normal range during day -365 through 0 days relative to index: Hemoglobin [Mass/volume] in Urine by Automated test strip | 0.2 | 0.3 | -0.03 | 0.2 | 0.3 | -0.03 |
| condition_era group during day -365 through 0 days relative to index: Temporal arteritis | 0.1 | 0.2 | -0.03 | 0.1 | 0.2 | -0.03 |
| procedure_occurrence during day -30 through 0 days relative to index: Hospital outpatient clinic visit specimen collection for severe acute respiratory syndrome coronavirus 2 (sars-cov-2) (coronavirus disease [covid-19]) | 0.2 | 0.1 | 0.03 | 0.2 | 0.1 | 0.03 |
| observation during day -365 through 0 days relative to index: Motor vehicle on road in collision with another motor vehicle | 0.2 | 0.3 | -0.03 | 0.2 | 0.3 | -0.03 |

**Supplementary Table 92:** Most extreme patient characteristics balance diagnostics after matching for DPP4I vs SU in the OptumDOD data source.

| | Before matching | | | After matching | | |
|---|---|---|---|---|---|---|
| Characteristic (total count = 16770) | T (%) | C (%) | SDf | T (%) | C (%) | SDf |
| condition_era group during day -30 through 0 days relative to index: Basal cell carcinoma of face | 0.1 | 0.2 | -0.02 | 0.1 | 0.2 | -0.03 |
| condition_era group during day -30 through 0 days relative to index: Carcinoma of face | 0.1 | 0.2 | -0.02 | 0.1 | 0.2 | -0.03 |
| measurement during day -365 through 0 days relative to index: Vitamin D; 25 hydroxy, includes fraction(s), if performed | 29.7 | 23.2 | 0.15 | 28.7 | 30.0 | -0.03 |
| condition_era group during day -365 through 0 days relative to index: Vitamin deficiency | 22.9 | 17.5 | 0.13 | 21.9 | 23.1 | -0.03 |
| condition_era group during day -365 through 0 days relative to index: Vitamin disease | 22.9 | 17.5 | 0.13 | 21.9 | 23.1 | -0.03 |
| condition_era group during day -365 through 0 days relative to index: Vitamin D deficiency | 19.8 | 14.8 | 0.13 | 18.9 | 20.1 | -0.03 |
| measurement during day -365 through 0 days relative to index: Urea nitrogen/Creatinine [Mass Ratio] in Serum or Plasma | 45.1 | 39.8 | 0.11 | 44.5 | 45.9 | -0.03 |
| drug_era group during day -365 through 0 days relative to index: aliskiren | 0.3 | 0.1 | 0.03 | 0.3 | 0.1 | 0.03 |
| drug_era group during day -365 through 0 days relative to index: OTHER AGENTS ACTING ON THE RENIN-ANGIOTENSIN SYSTEM | 0.3 | 0.1 | 0.03 | 0.3 | 0.1 | 0.03 |
| drug_era group during day -365 through 0 days relative to index: Renin-inhibitors | 0.3 | 0.1 | 0.03 | 0.3 | 0.1 | 0.03 |

##### 3.6.3 MDCR

**Supplementary Figure 29:** Preference score distributions, propensity score stratified (middle column) and matched (right column) covariate balance and negative control error distributions (bottom row) for DPP4I vs SU in the MDCR data source.

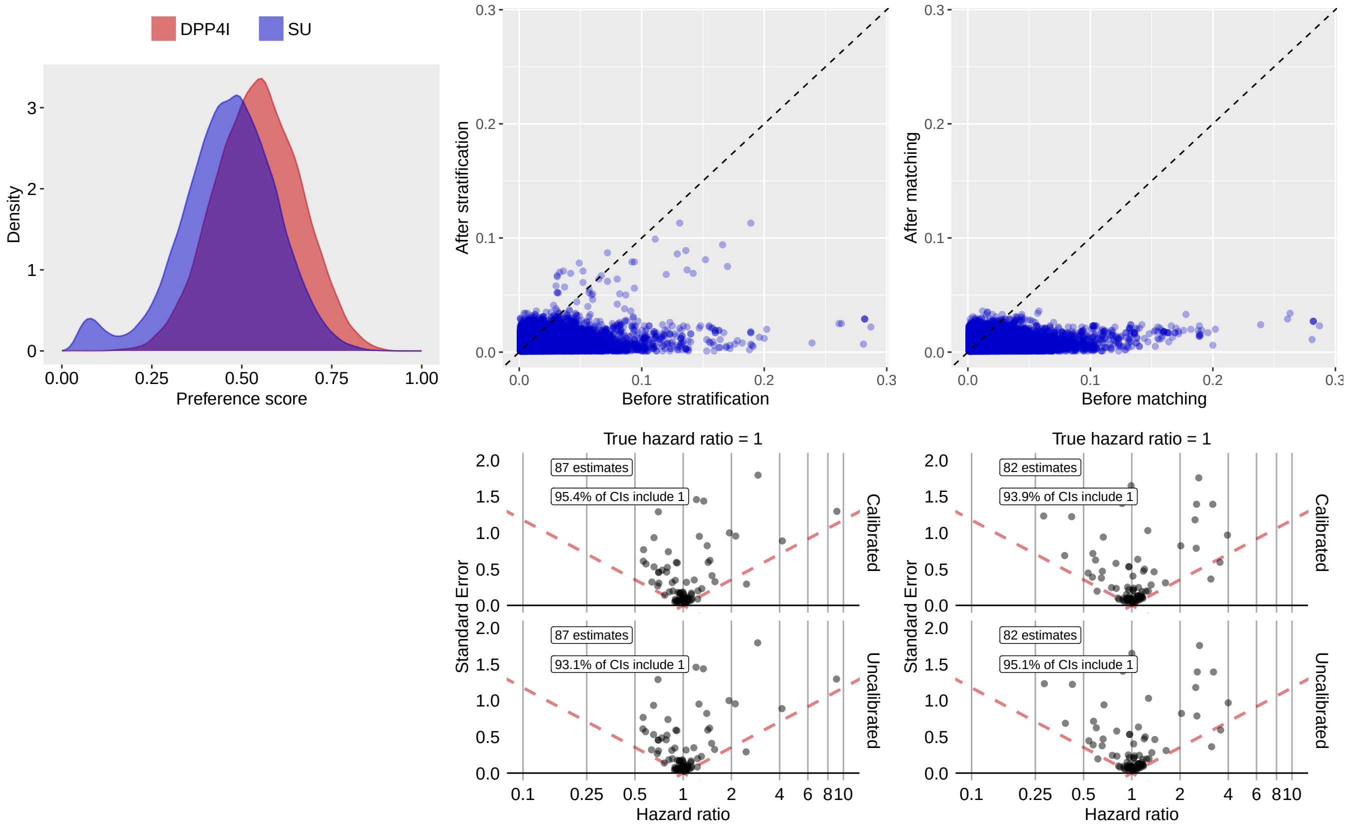

**Supplementary Table 93:** Most extreme patient characteristics balance diagnostics after stratification for DPP4I vs SU in the MDCR data source.

| Characteristic (total count = 13114) | Before stratification | | | After stratification | | |
|---|---|---|---|---|---|---|
| | T (%) | C (%) | SDf | T (%) | C (%) | SDf |
| measurement during day -365 through 0 days relative to index: Cholesterol, serum or whole blood, total | 0.5 | 2.9 | -0.19 | 0.8 | 2.2 | -0.11 |
| measurement during day -365 through 0 days relative to index: Immunoassay for infectious agent antibody, qualitative or semiquantitative, single step method (eg, reagent strip) | 0.1 | 1.0 | -0.13 | 0.1 | 0.9 | -0.11 |
| measurement during day -365 through 0 days relative to index: Hematocrit [Volume Fraction] of Blood | 0.1 | 0.8 | -0.11 | 0.1 | 0.7 | -0.10 |
| measurement during day -365 through 0 days relative to index: Potassium; serum, plasma or whole blood | 1.6 | 4.4 | -0.17 | 2.1 | 3.7 | -0.09 |
| measurement during day -30 through 0 days relative to index: Potassium; serum, plasma or whole blood | 0.4 | 1.7 | -0.14 | 0.5 | 1.4 | -0.09 |
| measurement above normal range during day -365 through 0 days relative to index: Cholesterol [Mass/volume] in Serum or Plasma | 1.4 | 2.4 | -0.07 | 1.4 | 2.7 | -0.09 |
| measurement during day -30 through 0 days relative to index: Cholesterol, serum or whole blood, total | 0.1 | 1.2 | -0.13 | 0.3 | 0.9 | -0.09 |

| (Continued) | T (%) | C (%) | SDf | T (%) | C (%) | SDf |
|---|---|---|---|---|---|---|
| measurement during day -30 through 0 days relative to index: Creatinine; blood | 1.1 | 3.3 | -0.15 | 1.6 | 2.8 | -0.08 |
| procedure_occurrence during day -30 through 0 days relative to index: General examination of patient | 2.1 | 3.7 | -0.09 | 2.2 | 3.5 | -0.08 |
| measurement during day -365 through 0 days relative to index: Blood count; blood smear, microscopic examination without manual differential WBC count | 0.1 | 0.7 | -0.09 | 0.1 | 0.6 | -0.08 |

**Supplementary Table 94:** Most extreme patient characteristics balance diagnostics after matching for DPP4I vs SU in the MDCR data source.

| | Before matching | | | After matching | | |
|---|---|---|---|---|---|---|
| Characteristic (total count = 13039) | T (%) | C (%) | SDf | T (%) | C (%) | SDf |
| drug_era group during day 0 through 0 days relative to index: febuxostat | 0.4 | 0.1 | 0.06 | 0.3 | 0.2 | 0.04 |
| procedure_occurrence during day -365 through 0 days relative to index: Injection procedure for sacroiliac joint; provision of anesthetic, steroid and/or other therapeutic agent, with or without arthrography | 0.1 | 0.2 | -0.01 | 0.1 | 0.3 | -0.04 |
| condition_era group during day -365 through 0 days relative to index: Panic disorder | 0.2 | 0.3 | -0.02 | 0.2 | 0.4 | -0.04 |
| condition_era group during day -365 through 0 days relative to index: Panic | 0.2 | 0.3 | -0.02 | 0.2 | 0.4 | -0.04 |
| procedure_occurrence during day -365 through 0 days relative to index: Office or other outpatient visit for the evaluation and management of an established patient, which requires at least 2 of these 3 key components: A detailed history; A detailed examination; Medical decision making of moderate complexity. Counseling and/o | 87.7 | 77.9 | 0.26 | 87.1 | 88.2 | -0.03 |
| drug_era group during day -30 through 0 days relative to index: febuxostat | 0.4 | 0.1 | 0.06 | 0.4 | 0.2 | 0.03 |
| procedure_occurrence during day -365 through 0 days relative to index: Electrocardiogram, routine ECG with at least 12 leads; with interpretation and report | 37.5 | 29.1 | 0.18 | 35.2 | 36.8 | -0.03 |
| condition_era group during day -30 through 0 days relative to index: Neoplasm of pancreas | 0.1 | 0.2 | -0.04 | 0.1 | 0.2 | -0.03 |
| measurement during day -365 through 0 days relative to index: Screening for cardiovascular system disease | 0.2 | 0.4 | -0.03 | 0.2 | 0.4 | -0.03 |
| procedure_occurrence during day -365 through 0 days relative to index: Electronic analysis of dual-chamber pacemaker system (includes evaluation of programmable parameters at rest and during activity where applicable, using electrocardiographic recording and interpretation of recordings at rest and during exercise, analysis | 0.2 | 0.3 | -0.02 | 0.2 | 0.1 | 0.03 |

##### 3.6.4 MDCD

**Supplementary Figure 30:** Preference score distributions, propensity score stratified (middle column) and matched (right column) covariate balance and negative control error distributions (bottom row) for DPP4I vs SU in the MDCD data source.

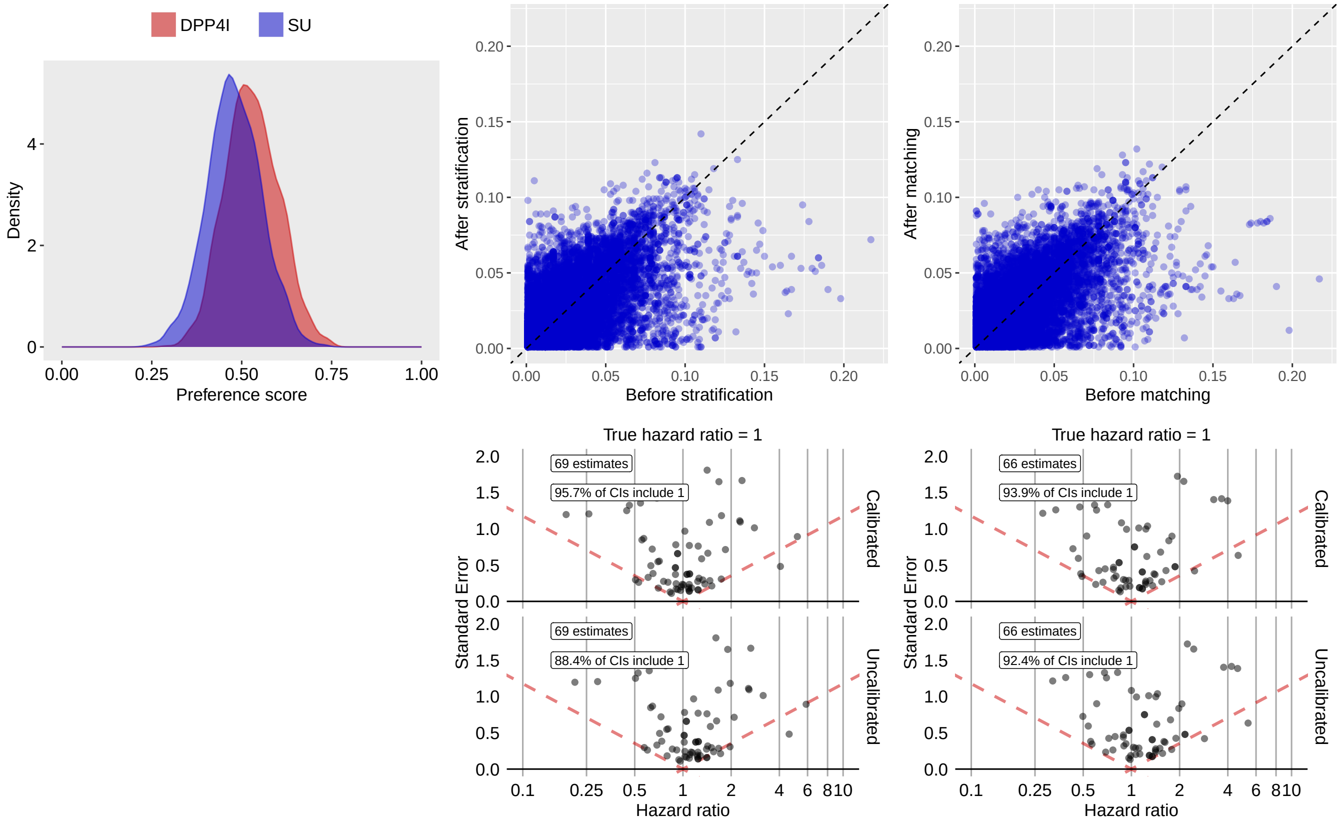

**Supplementary Table 95:** Most extreme patient characteristics balance diagnostics after stratification for DPP4I vs SU in the MDCD data source.

| Characteristic (total count = 14799) | Before stratification | | | After stratification | | |
|---|---|---|---|---|---|---|
| | T (%) | C (%) | SDf | T (%) | C (%) | SDf |
| procedure_occurrence during day -30 through 0 days relative to index: Ultrasound, soft tissues of head and neck (eg, thyroid, parathyroid, parotid), real time with image documentation | 0.9 | -0.2 | 0.11 | 1.3 | -0.2 | 0.14 |
| condition_era group during day -365 through 0 days relative to index: Inflammatory disease of liver | 0.7 | 2.3 | -0.13 | 0.7 | 2.1 | -0.12 |
| condition_era group during day -365 through 0 days relative to index: Cyst of kidney | 5.0 | 3.3 | 0.08 | 6.0 | 3.4 | 0.12 |
| condition_era group during day -365 through 0 days relative to index: Uterine prolapse | 0.9 | -0.2 | 0.12 | 0.9 | -0.2 | 0.12 |
| condition_era group during day -365 through 0 days relative to index: Renal mass | 5.4 | 3.8 | 0.08 | 6.3 | 3.8 | 0.12 |
| observation during day -365 through 0 days relative to index: Set-up portable x-ray equipment | 4.8 | 2.8 | 0.11 | 4.9 | 2.7 | 0.12 |
| drug_era group during day -30 through 0 days relative to index: terbinafine | 0.7 | -0.2 | 0.10 | 0.9 | -0.2 | 0.11 |

| (Continued) | T (%) | C (%) | SDf | T (%) | C (%) | SDf |
|---|---|---|---|---|---|---|
| drug_era group during day -30 through 0 days relative to index: ANTIFUNGALS FOR SYSTEMIC USE | 0.7 | -0.2 | 0.10 | 0.9 | -0.2 | 0.11 |
| drug_era group during day -30 through 0 days relative to index: Antifungals for systemic use | 0.7 | -0.2 | 0.10 | 0.9 | -0.2 | 0.11 |
| condition_era group during day -365 through 0 days relative to index: Infectious disease of abdomen | 2.3 | 3.8 | -0.09 | 1.9 | 3.7 | -0.11 |

**Supplementary Table 96:** Most extreme patient characteristics balance diagnostics after matching for DPP4I vs SU in the MDCD data source.

| | Before matching | | | After matching | | |
|---|---|---|---|---|---|---|
| Characteristic (total count = 14101) | T (%) | C (%) | SDf | T (%) | C (%) | SDf |
| drug_era group during day -30 through 0 days relative to index: Serotonin (5HT3) antagonists | 1.8 | 3.5 | -0.10 | 1.7 | 3.8 | -0.13 |
| drug_era group during day -30 through 0 days relative to index: ondansetron | 1.7 | 3.2 | -0.09 | 1.6 | 3.6 | -0.13 |
| drug_era group during day -30 through 0 days relative to index: ANTIEMETICS AND ANTINAUSEANTS | 2.0 | 3.5 | -0.10 | 1.8 | 3.9 | -0.12 |
| drug_era group during day -30 through 0 days relative to index: ANTIEMETICS AND ANTINAUSEANTS | 2.0 | 3.5 | -0.10 | 1.8 | 3.9 | -0.12 |
| procedure_occurrence during day -30 through 0 days relative to index: Ultrasound, soft tissues of head and neck (eg, thyroid, parathyroid, parotid), real time with image documentation | 0.9 | -0.2 | 0.11 | 1.0 | -0.2 | 0.12 |
| condition_era group during day -365 through 0 days relative to index: Uterine prolapse | 0.9 | -0.2 | 0.12 | 0.9 | -0.2 | 0.12 |
| procedure_occurrence during day -365 through 0 days relative to index: Limited bilateral noninvasive physiologic studies of upper or lower extremity arteries, (eg, for lower extremity: ankle/brachial indices at distal posterior tibial and anterior tibial/dorsalis pedis arteries plus bidirectional, Doppler waveform recording | 1.9 | 3.1 | -0.08 | 1.7 | 3.6 | -0.12 |
| observation during day -365 through 0 days relative to index: Set-up portable x-ray equipment | 4.8 | 2.8 | 0.11 | 4.7 | 2.6 | 0.12 |
| measurement during day -365 through 0 days relative to index: Deoxyribonucleic acid (DNA) antibody; native or double stranded | -0.4 | 0.6 | -0.09 | -0.4 | 0.9 | -0.12 |
| observation during day -365 through 0 days relative to index: H/O: risk factor | -0.4 | 0.7 | -0.08 | -0.4 | 0.9 | -0.12 |

##### 3.6.5 Open Claims

**Supplementary Figure 31:** Preference score distributions, propensity score stratified (middle column) and matched (right column) covariate balance and negative control error distributions (bottom row) for DPP4I vs SU in the Open Claims data source.

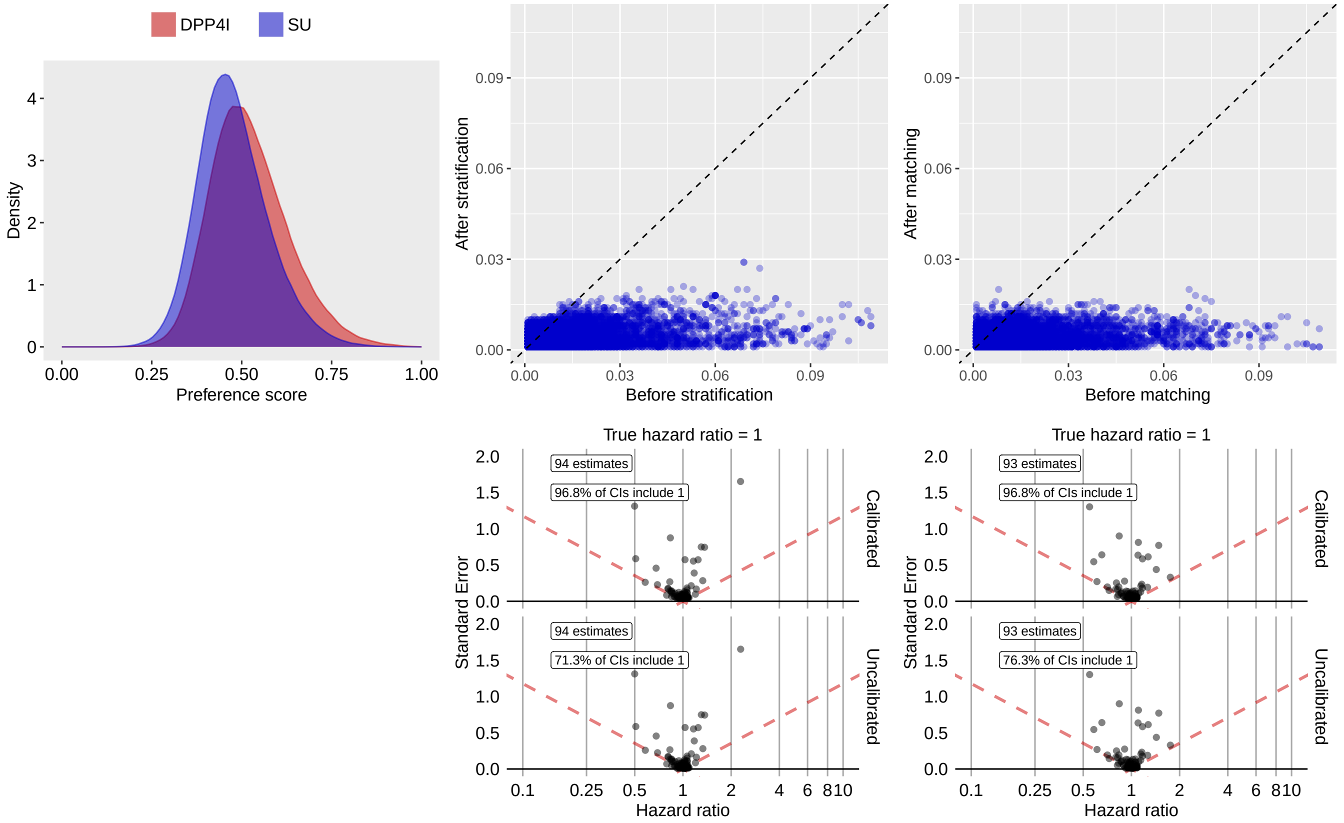

**Supplementary Table 97:** Most extreme patient characteristics balance diagnostics after stratification for DPP4I vs SU in the Open Claims data source.

| Characteristic (total count = 10908) | Before stratification | | | After stratification | | |
|---|---|---|---|---|---|---|
| | T (%) | C (%) | SDf | T (%) | C (%) | SDf |
| drug_era group during day -30 through 0 days relative to index: icosapent ethyl | 0.7 | 0.2 | 0.07 | 0.4 | 0.3 | 0.03 |
| drug_era group during day 0 through 0 days relative to index: icosapent ethyl | 0.6 | 0.2 | 0.07 | 0.4 | 0.2 | 0.03 |
| drug_era group during day -365 through 0 days relative to index: icosapent ethyl | 0.9 | 0.3 | 0.07 | 0.6 | 0.4 | 0.03 |
| observation during day -365 through 0 days relative to index: Personal care services, per 15 minutes, not for an inpatient or resident of a hospital, nursing facility, icf/mr or imd, part of the individualized plan of treatment (code may not be used to identify services provided by home health aide or certified n... | 0.5 | 0.2 | 0.05 | 0.3 | 0.2 | 0.02 |
| drug_era group during day -365 through 0 days relative to index: Combinations and complexes of aluminium, calcium and magnesium compounds | 2.2 | 1.3 | 0.07 | 1.7 | 1.4 | 0.02 |
| drug_era group during day -365 through 0 days relative to index: calcium carbonate | 2.0 | 1.1 | 0.07 | 1.5 | 1.3 | 0.02 |

| (Continued) | T (%) | C (%) | SDf | T (%) | C (%) | SDf |
|---|---|---|---|---|---|---|
| measurement during day -365 through 0 days relative to index: Lipoprotein, blood; quantitation of lipoprotein particle number(s) (eg, by nuclear magnetic resonance spectroscopy), includes lipoprotein particle subclass(es), when performed | 0.6 | 0.2 | 0.05 | 0.4 | 0.3 | 0.02 |
| observation during day -30 through 0 days relative to index: Personal care services, per 15 minutes, not for an inpatient or resident of a hospital, nursing facility, icf/mr or imd, part of the individualized plan of treatment (code may not be used to identify services provided by home health aide or certified n... | 0.4 | 0.1 | 0.05 | 0.3 | 0.2 | 0.02 |
| observation during day -365 through 0 days relative to index: Day care services, adult; per diem | 0.2 | 0.1 | 0.04 | 0.2 | 0.1 | 0.02 |
| drug_era group during day -30 through 0 days relative to index: Combinations and complexes of aluminium, calcium and magnesium compounds | 1.3 | 0.7 | 0.06 | 1.0 | 0.8 | 0.02 |

**Supplementary Table 98:** Most extreme patient characteristics balance diagnostics after matching for DPP4I vs SU in the Open Claims data source.

| | Before matching | | | After matching | | |
|---|---|---|---|---|---|---|
| Characteristic (total count = 10809) | T (%) | C (%) | SDf | T (%) | C (%) | SDf |
| drug_era group during day 0 through 0 days relative to index: icosapent ethyl | 0.6 | 0.2 | 0.07 | 0.5 | 0.3 | 0.02 |
| age group: 85 - 89 | 0.3 | 0.2 | 0.01 | 0.3 | 0.2 | 0.02 |
| drug_era group during day -30 through 0 days relative to index: icosapent ethyl | 0.7 | 0.2 | 0.07 | 0.5 | 0.4 | 0.02 |
| drug_era group during day -365 through 0 days relative to index: icosapent ethyl | 0.9 | 0.3 | 0.07 | 0.7 | 0.5 | 0.02 |
| measurement during day -365 through 0 days relative to index: Thyroxine; free | 7.4 | 5.6 | 0.07 | 7.2 | 6.8 | 0.02 |
| condition_era group during day -30 through 0 days relative to index: Goiter | 0.7 | 0.4 | 0.04 | 0.7 | 0.5 | 0.02 |
| condition_era group during day -30 through 0 days relative to index: Simple goiter | 0.6 | 0.4 | 0.04 | 0.6 | 0.5 | 0.02 |
| drug_era group during day -365 through 0 days relative to index: vilazodone | 0.1 | 0.1 | 0.02 | 0.1 | 0.1 | 0.02 |
| device_exposure during day -365 through 0 days relative to index: TRUETEST GLUCOSE TEST STRIP | 0.1 | 0.2 | -0.02 | 0.1 | 0.2 | -0.02 |
| drug_era group during day -30 through 0 days relative to index: oxycodone | 2.6 | 2.8 | -0.01 | 2.6 | 2.9 | -0.02 |

##### 3.6.6 OptumEHR

**Supplementary Figure 32:** Preference score distributions, propensity score stratified (middle column) and matched (right column) covariate balance and negative control error distributions (bottom row) for DPP4I vs SU in the OptumEHR data source.

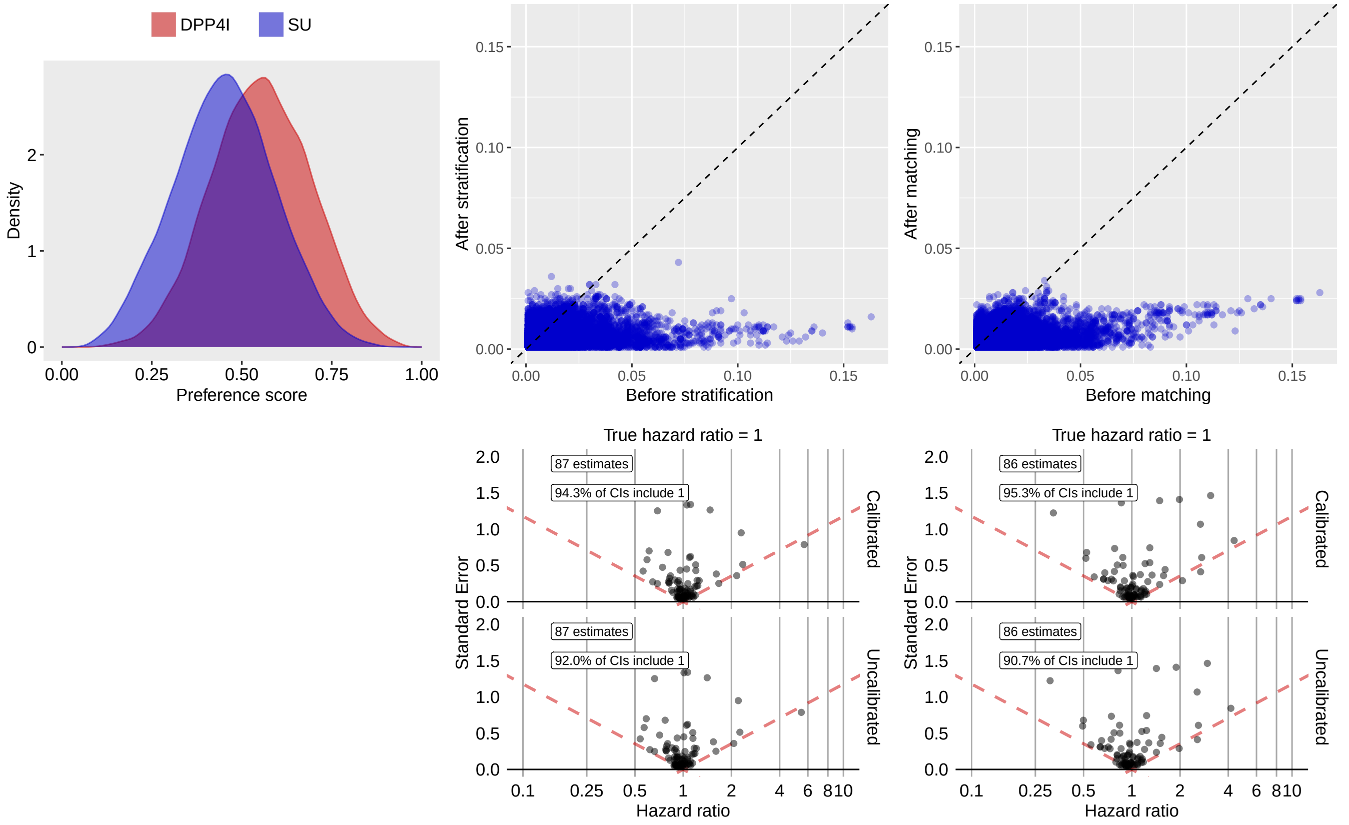

**Supplementary Table 99:** Most extreme patient characteristics balance diagnostics after stratification for DPP4I vs SU in the OptumEHR data source.

| Characteristic (total count = 12791) | Before stratification | | | After stratification | | |
|---|---|---|---|---|---|---|
| | T (%) | C (%) | SDf | T (%) | C (%) | SDf |
| measurement during day -365 through 0 days relative to index: Eye disorder screening | 0.1 | 0.6 | -0.07 | 0.2 | 0.5 | -0.04 |
| procedure_occurrence during day -365 through 0 days relative to index: Examination of research control | 0.2 | 0.1 | 0.01 | 0.3 | 0.1 | 0.04 |
| measurement during day -30 through 0 days relative to index: Lipoprotein.beta/Lipoprotein.alpha [Mass Ratio] in Serum or Plasma | 0.1 | 0.3 | -0.04 | 0.1 | 0.2 | -0.03 |
| procedure_occurrence during day -365 through 0 days relative to index: Rhythm ECG, 1-3 leads; with interpretation and report | 0.2 | 0.1 | 0.03 | 0.2 | 0.1 | 0.03 |
| condition_era group during day -30 through 0 days relative to index: Hypoglycemic disorder | 0.1 | 0.3 | -0.03 | 0.1 | 0.3 | -0.03 |
| condition_era group during day -365 through 0 days relative to index: Polyneuropathy due to drug | 0.1 | 0.2 | -0.03 | 0.1 | 0.2 | -0.03 |
| measurement during day -365 through 0 days relative to index: Blood count; red blood cell (RBC), automated | 0.1 | 0.1 | 0.02 | 0.2 | 0.1 | 0.03 |
| condition_era group during day -30 through 0 days relative to index: Open-angle glaucoma - borderline | 0.2 | 0.1 | 0.02 | 0.2 | 0.1 | 0.03 |

| (Continued) | T (%) | C (%) | SDf | T (%) | C (%) | SDf |
|---|---|---|---|---|---|---|
| measurement during day -30 through 0 days relative to index: Unlisted chemistry procedure | 0.1 | 0.1 | 0.01 | 0.2 | 0.1 | 0.03 |
| condition_era group during day -30 through 0 days relative to index: Hypoglycemia | 0.1 | 0.2 | -0.03 | 0.1 | 0.2 | -0.03 |

**Supplementary Table 100:** Most extreme patient characteristics balance diagnostics after matching for DPP4I vs SU in the OptumEHR data source.

| | Before matching | | | After matching | | |
|---|---|---|---|---|---|---|
| Characteristic (total count = 12638) | T (%) | C (%) | SDf | T (%) | C (%) | SDf |
| procedure_occurrence during day -365 through 0 days relative to index: Rhythm ECG, 1-3 leads; with interpretation and report | 0.2 | 0.1 | 0.03 | 0.2 | 0.1 | 0.03 |
| condition_era group during day -365 through 0 days relative to index: Tumor of oral cavity | 0.1 | 0.2 | -0.03 | 0.1 | 0.2 | -0.03 |
| condition_era group during day -365 through 0 days relative to index: Right inguinal hernia | 0.2 | 0.1 | 0.04 | 0.2 | 0.1 | 0.03 |
| condition_era group during day -365 through 0 days relative to index: Bilateral inguinal hernia | 0.2 | 0.1 | 0.03 | 0.2 | 0.1 | 0.03 |
| measurement during day -365 through 0 days relative to index: Measurement of liver enzyme | 0.1 | 0.1 | -0.02 | 0.1 | 0.1 | -0.03 |
| measurement during day -30 through 0 days relative to index: Body height | 69.3 | 61.6 | 0.16 | 68.4 | 69.8 | -0.03 |
| procedure_occurrence during day -365 through 0 days relative to index: Computed tomography, abdomen; without contrast material | 0.1 | 0.3 | -0.03 | 0.1 | 0.2 | -0.03 |
| condition_era group during day -365 through 0 days relative to index: Left inguinal hernia | 0.2 | 0.1 | 0.03 | 0.2 | 0.1 | 0.03 |
| condition_era group during day -365 through 0 days relative to index: Polyneuropathy due to drug | 0.1 | 0.2 | -0.03 | 0.1 | 0.1 | -0.03 |
| condition_era group during day -365 through 0 days relative to index: Inflammation of vulva | 0.1 | 0.1 | -0.02 | 0.1 | 0.1 | -0.03 |

### 3.6.7 VA

**Supplementary Figure 33:** Preference score distributions, propensity score stratified (middle column) and matched (right column) covariate balance and negative control error distributions (bottom row) for DPP4I vs SU in the VA data source.

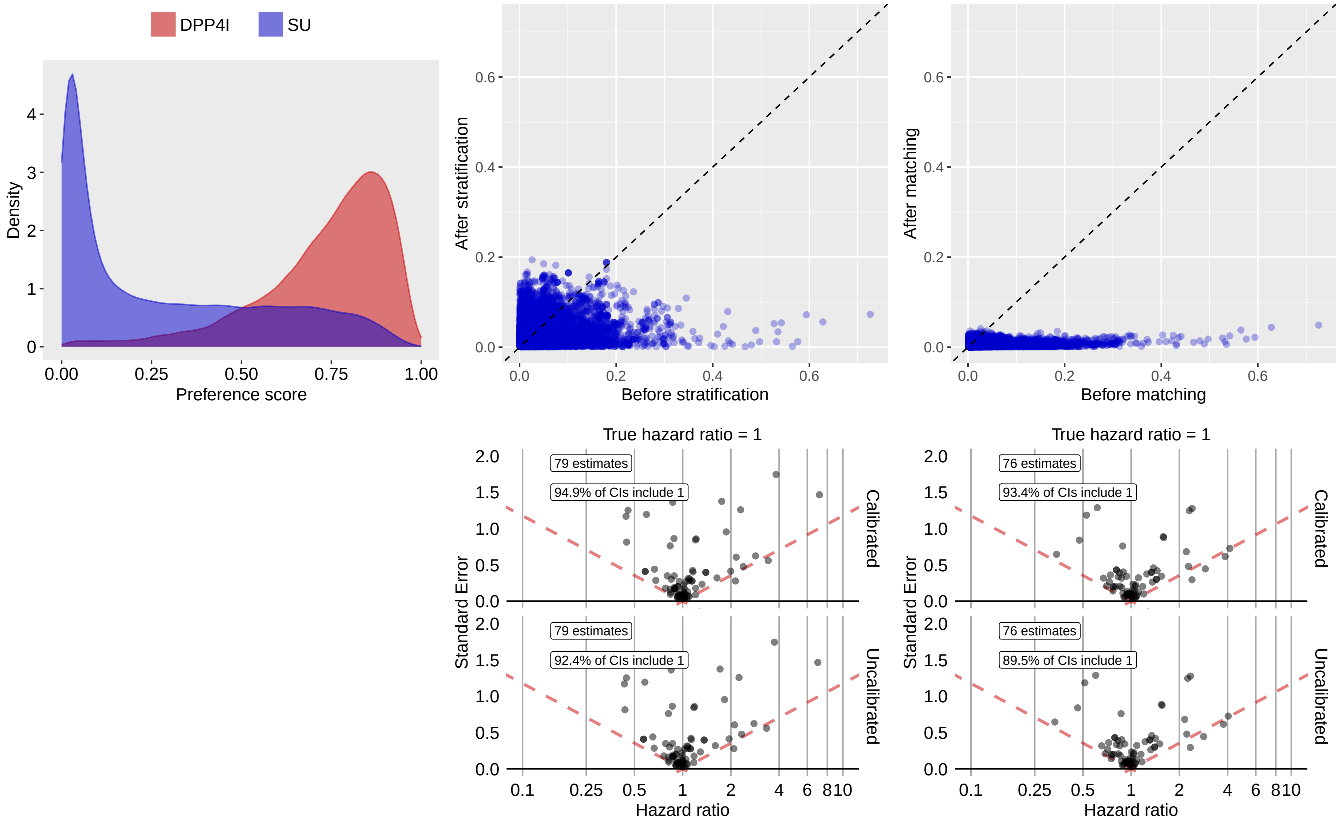

**Supplementary Table 101:** Most extreme patient characteristics balance diagnostics after stratification for DPP4I vs SU in the VA data source.

| Characteristic (total count = 12551) | Before stratification | | | After stratification | | |
|---|---|---|---|---|---|---|
| | T (%) | C (%) | SDf | T (%) | C (%) | SDf |
| observation during day -30 through 0 days relative to index: Discharge from medical service | 1.0 | 1.3 | -0.03 | 4.2 | 1.1 | 0.19 |
| drug_era group during day -365 through 0 days relative to index: Metformin | 86.3 | 91.9 | -0.18 | 83.8 | 90.1 | -0.19 |
| drug_era group during day -365 through 0 days relative to index: BLOOD GLUCOSE LOWERING DRUGS, EXCL. INSULINS | 86.3 | 91.9 | -0.18 | 83.8 | 90.1 | -0.19 |
| drug_era group during day -365 through 0 days relative to index: Biguanides | 86.3 | 91.9 | -0.18 | 83.8 | 90.1 | -0.19 |
| drug_era group during day -365 through 0 days relative to index: Combinations of oral blood glucose lowering drugs | 86.3 | 91.9 | -0.18 | 83.8 | 90.1 | -0.19 |
| observation during day -365 through 0 days relative to index: Discharge from medical service | 5.8 | 7.1 | -0.05 | 12.5 | 7.1 | 0.18 |
| condition_era group during day -365 through 0 days relative to index: Heart disease | 37.2 | 40.9 | -0.07 | 48.0 | 39.0 | 0.18 |
| drug_era group during day -365 through 0 days relative to index: DRUGS USED IN DIABETES | 86.6 | 92.1 | -0.18 | 84.7 | 90.4 | -0.17 |

| (Continued) | T (%) | C (%) | SDf | T (%) | C (%) | SDf |
|---|---|---|---|---|---|---|
| measurement during day -30 through 0 days relative to index: Methicillin resistant Staphylococcus aureus (MRSA) DNA [Presence] in Specimen by NAA with probe detection | 0.7 | 0.6 | 0.02 | 3.0 | 0.7 | 0.17 |
| drug_era group during day -30 through 0 days relative to index: DRUGS USED IN DIABETES | 74.5 | 78.8 | -0.10 | 70.0 | 77.3 | -0.17 |

**Supplementary Table 102:** Most extreme patient characteristics balance diagnostics after matching for DPP4I vs SU in the VA data source.

| | Before matching | | | After matching | | |
|---|---|---|---|---|---|---|
| Characteristic (total count = 12647) | T (%) | C (%) | SDf | T (%) | C (%) | SDf |
| procedure_occurrence during day -365 through 0 days relative to index: Counseling | 6.6 | 34.1 | -0.73 | 7.1 | 5.9 | 0.05 |
| observation during day -365 through 0 days relative to index: Requires influenza virus vaccination | 4.0 | 25.2 | -0.63 | 4.3 | 3.5 | 0.04 |
| measurement within normal range during day -365 through 0 days relative to index: Vancomycin [Mass/volume] in Serum or Plasma – trough | -0.1 | 0.1 | -0.03 | -0.1 | 0.1 | -0.04 |
| index year: 2020 | 22.8 | 4.2 | 0.56 | 21.3 | 22.8 | -0.04 |
| measurement below normal range during day -365 through 0 days relative to index: Prostate Specific Ag Free/Prostate specific Ag.total in Serum or Plasma | -0.1 | 0.1 | -0.04 | -0.1 | 0.1 | -0.04 |
| procedure_occurrence during day -30 through 0 days relative to index: Medical team conference with interdisciplinary team of health care professionals, patient and/or family not present, 30 minutes or more; participation by physician | -0.1 | 0.1 | -0.00 | -0.1 | 0.1 | -0.04 |
| index year: 2014 | 2.5 | 6.3 | -0.18 | 2.7 | 2.2 | 0.04 |
| measurement above normal range during day -365 through 0 days relative to index: Erythrocyte distribution width [Entitic volume] | 0.2 | 0.1 | 0.03 | 0.2 | 0.1 | 0.04 |
| condition_era group during day -365 through 0 days relative to index: Neutrophil count abnormal | -0.1 | 0.1 | -0.03 | -0.1 | 0.1 | -0.03 |
| drug_era group during day -30 through 0 days relative to index: Sodium polystyrene sulfonate | -0.1 | 0.1 | -0.05 | -0.1 | 0.1 | -0.03 |

##### 3.6.8 GermanyDA

**Supplementary Figure 34:** Preference score distributions, propensity score stratified (middle column) and matched (right column) covariate balance and negative control error distributions (bottom row) for DPP4I vs SU in the GermanyDA data source.

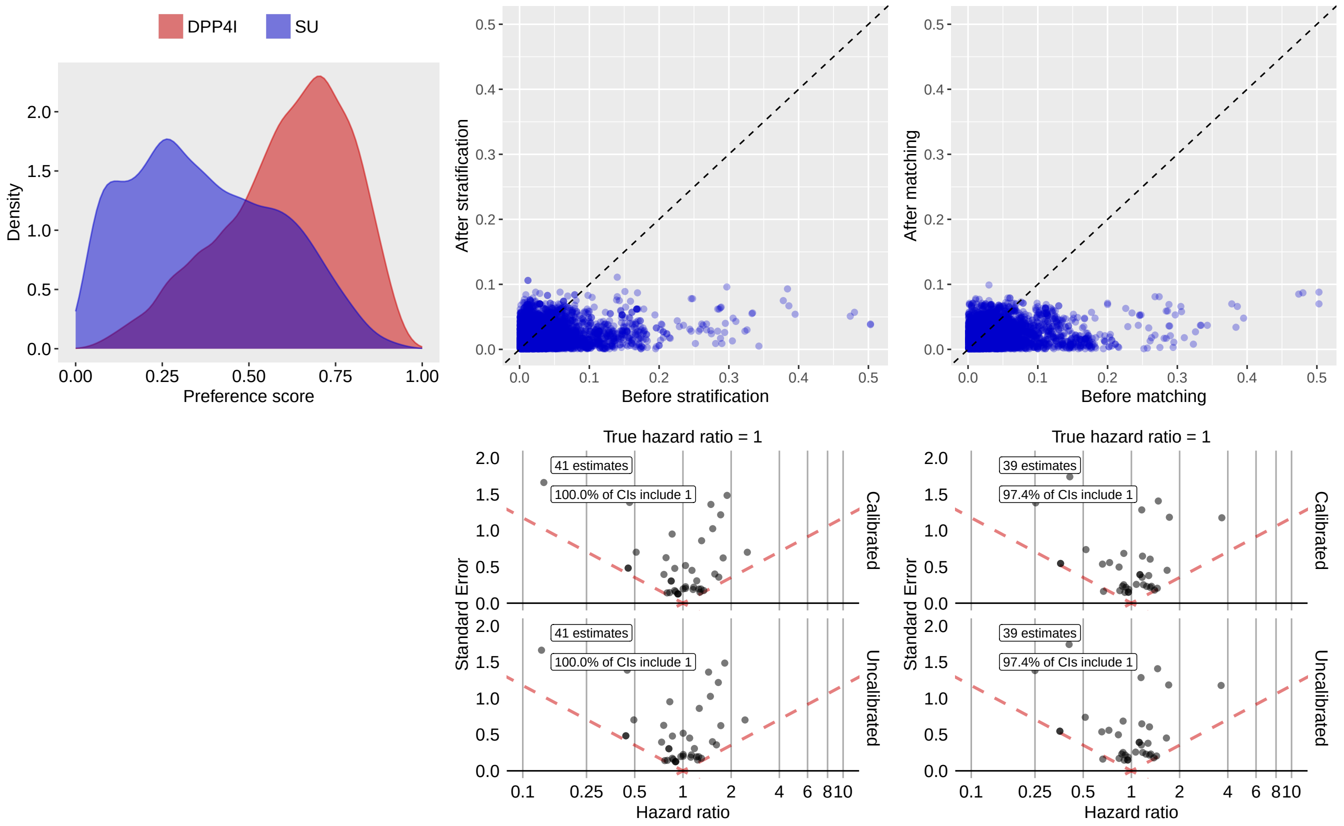

**Supplementary Table 103:** Most extreme patient characteristics balance diagnostics after stratification for DPP4I vs SU in the GermanyDA data source.

| | Before stratification | | | After stratification | | |
|---|---|---|---|---|---|---|
| Characteristic (total count = 6736) | T (%) | C (%) | SDf | T (%) | C (%) | SDf |
| drug_era group during day 0 through 0 days relative to index: rosuvastatin | 1.1 | -0.1 | 0.14 | 1.0 | 0.1 | 0.11 |
| condition_era group during day -365 through 0 days relative to index: Cerebral atherosclerosis | 0.9 | 0.8 | 0.01 | 0.9 | 2.2 | -0.11 |
| condition_era group during day -365 through 0 days relative to index: Cerebral arteriosclerosis | 0.9 | 0.8 | 0.01 | 0.9 | 2.2 | -0.11 |
| index year: 2020 | 11.4 | 3.7 | 0.30 | 10.4 | 7.7 | 0.10 |
| index year: 2007 | 0.3 | 7.6 | -0.38 | 0.6 | 1.5 | -0.09 |
| drug_era group during day -365 through 0 days relative to index: rosuvastatin | 1.3 | -0.1 | 0.15 | 1.2 | 0.4 | 0.09 |
| condition_era group during day -365 through 0 days relative to index: Endemic goiter | 0.9 | 0.4 | 0.06 | 0.9 | 0.2 | 0.09 |
| condition_era group during day -30 through 0 days relative to index: Disorder due to type 2 diabetes mellitus | 6.6 | 6.1 | 0.02 | 6.4 | 8.7 | -0.09 |
| measurement during day -365 through 0 days relative to index: Albumin [Mass/volume] in Body fluid | 2.4 | 0.4 | 0.17 | 2.1 | 1.0 | 0.09 |

| (Continued) | T (%) | C (%) | SDf | T (%) | C (%) | SDf |
|---|---|---|---|---|---|---|
| condition_era group during day -365 through 0 days relative to index: Atherosclerosis of artery | 3.2 | 2.7 | 0.03 | 3.1 | 4.8 | -0.09 |

**Supplementary Table 104:** Most extreme patient characteristics balance diagnostics after matching for DPP4I vs SU in the GermanyDA data source.

| | Before matching | | | After matching | | |
|---|---|---|---|---|---|---|
| Characteristic (total count = 6308) | T (%) | C (%) | SDf | T (%) | C (%) | SDf |
| condition_era group during day -365 through 0 days relative to index: Cervicocranial syndrome | 0.5 | 0.3 | 0.03 | 1.0 | 0.2 | 0.10 |
| measurement during day -365 through 0 days relative to index: Erythrocytes [#/volume] in Body fluid | 40.7 | 18.4 | 0.50 | 23.1 | 27.0 | -0.09 |
| measurement during day -365 through 0 days relative to index: Leukocytes [#/volume] in Body fluid | 41.1 | 19.6 | 0.48 | 24.9 | 28.7 | -0.09 |
| measurement during day -365 through 0 days relative to index: Platelets [#/volume] in Body fluid by Automated count | 41.4 | 20.1 | 0.47 | 25.6 | 29.4 | -0.09 |
| measurement during day -30 through 0 days relative to index: Leukocytes [#/volume] in Body fluid | 19.9 | 10.2 | 0.27 | 12.1 | 14.8 | -0.08 |
| measurement during day -30 through 0 days relative to index: Platelets [#/volume] in Body fluid by Automated count | 20.1 | 10.5 | 0.27 | 12.5 | 15.3 | -0.08 |
| condition_era group during day -365 through 0 days relative to index: Attention deficit hyperactivity disorder | 0.2 | 0.5 | -0.06 | -0.2 | 0.7 | -0.08 |
| condition_era group during day -365 through 0 days relative to index: Disorders of attention and motor control | 0.2 | 0.5 | -0.06 | -0.2 | 0.7 | -0.08 |
| condition_era group during day -365 through 0 days relative to index: Developmental disorder of motor function | 0.2 | 0.5 | -0.06 | -0.2 | 0.7 | -0.08 |
| condition_era group during day -30 through 0 days relative to index: Paroxysmal atrial fibrillation | 0.9 | 0.8 | 0.01 | 0.4 | 1.0 | -0.08 |

##### 3.6.9 SIDIAP

**Supplementary Figure 35:** Preference score distributions, propensity score stratified (middle column) and matched (right column) covariate balance and negative control error distributions (bottom row) for DPP4I vs SU in the SIDIAP data source.

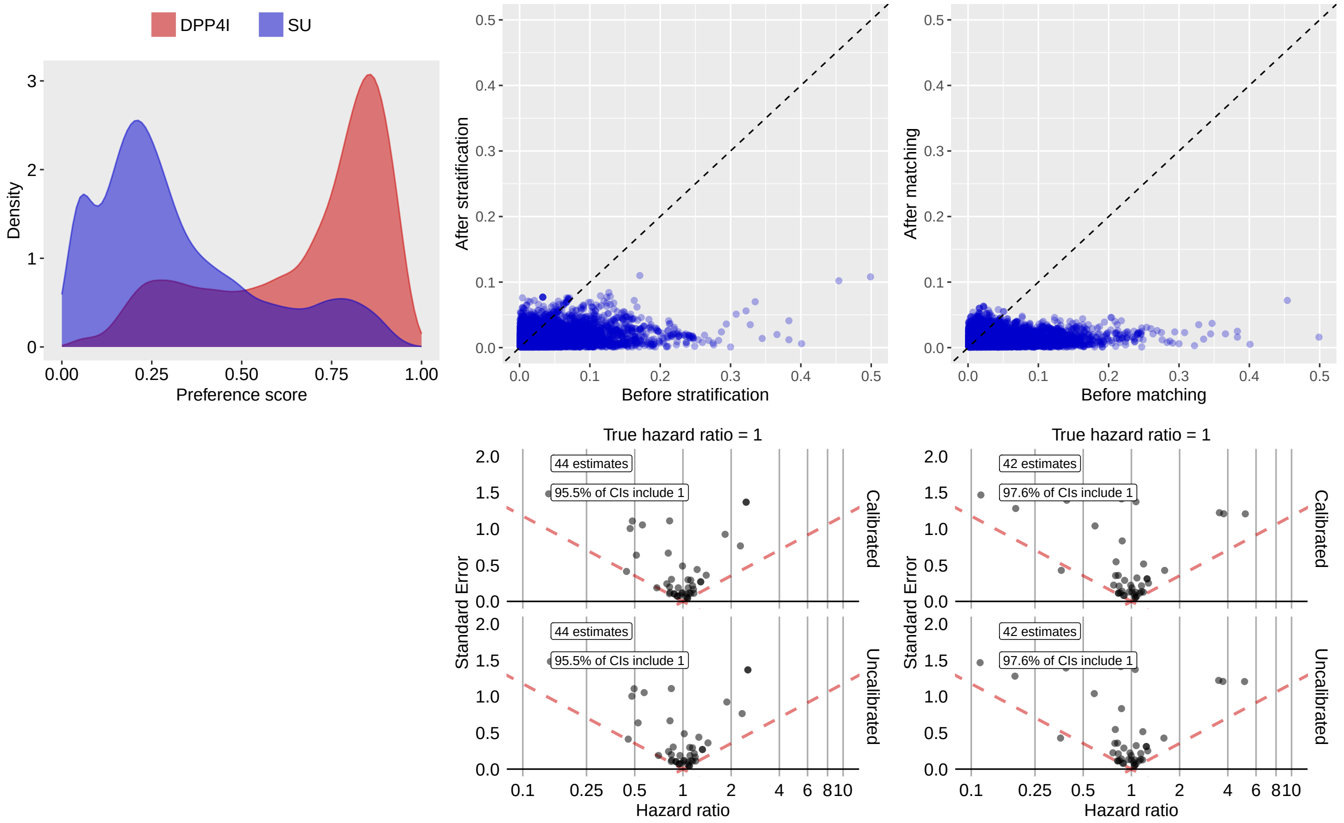

**Supplementary Table 105:** Most extreme patient characteristics balance diagnostics after stratification for DPP4I vs SU in the SIDIAP data source.

| | Before stratification | | | After stratification | | |
|---|---|---|---|---|---|---|
| Characteristic (total count = 6641) | T (%) | C (%) | SDf | T (%) | C (%) | SDf |
| procedure_occurrence during day -365 through 0 days relative to index: Plain Radiography of Chest | 1.5 | -0.1 | 0.17 | 1.2 | 0.3 | 0.11 |
| index year: 2008 | 0.7 | 13.0 | -0.50 | 2.4 | 4.3 | -0.11 |
| index year: 2019 | 13.6 | 1.8 | 0.45 | 11.1 | 8.1 | 0.10 |
| CHADS2VASc | 402.8 | 388.7 | 0.13 | 398.5 | 389.3 | 0.08 |
| drug_era group during day 0 through 0 days relative to index: High-ceiling diuretics and potassium-sparing agents | 15.0 | 11.1 | 0.12 | 13.9 | 11.3 | 0.08 |
| drug_era group during day 0 through 0 days relative to index: Sulfonamides and potassium in combination | 12.9 | 8.9 | 0.13 | 11.8 | 9.4 | 0.08 |
| drug_era group during day 0 through 0 days relative to index: furosemide | 12.7 | 8.8 | 0.12 | 11.6 | 9.2 | 0.08 |
| drug_era group during day 0 through 0 days relative to index: DRUGS USED IN DIABETES | 75.0 | 76.4 | -0.03 | 75.5 | 78.8 | -0.08 |
| drug_era group during day 0 through 0 days relative to index: BLOOD GLUCOSE LOWERING DRUGS, EXCL. INSULINS | 75.0 | 76.4 | -0.03 | 75.5 | 78.8 | -0.08 |

| (Continued) | T (%) | C (%) | SDf | T (%) | C (%) | SDf |
|---|---|---|---|---|---|---|
| drug_era group during day 0 through 0 days relative to index: metformin | 75.0 | 76.4 | -0.03 | 75.5 | 78.8 | -0.08 |

**Supplementary Table 106:** Most extreme patient characteristics balance diagnostics after matching for DPP4I vs SU in the SIDIAP data source.

| | Before matching | | | After matching | | |
|---|---|---|---|---|---|---|
| Characteristic (total count = 7075) | T (%) | C (%) | SDf | T (%) | C (%) | SDf |
| index year: 2019 | 13.6 | 1.8 | 0.45 | 3.8 | 2.5 | 0.07 |
| drug_era group during day 0 through 0 days relative to index: aliskiren | 0.2 | 0.1 | 0.02 | 0.5 | 0.1 | 0.06 |
| drug_era group during day 0 through 0 days relative to index: OTHER AGENTS ACTING ON THE RENIN-ANGIOTENSIN SYSTEM | 0.2 | 0.1 | 0.02 | 0.5 | 0.1 | 0.06 |
| drug_era group during day 0 through 0 days relative to index: Renin-inhibitors | 0.2 | 0.1 | 0.02 | 0.5 | 0.1 | 0.06 |
| condition_era group during day -30 through 0 days relative to index: Curvature of spine | 0.2 | 0.1 | 0.02 | 0.5 | 0.1 | 0.06 |
| drug_era group during day -30 through 0 days relative to index: aliskiren | 0.2 | 0.1 | 0.02 | 0.6 | 0.2 | 0.06 |
| drug_era group during day -30 through 0 days relative to index: OTHER AGENTS ACTING ON THE RENIN-ANGIOTENSIN SYSTEM | 0.2 | 0.1 | 0.02 | 0.6 | 0.2 | 0.06 |
| drug_era group during day -30 through 0 days relative to index: Renin-inhibitors | 0.2 | 0.1 | 0.02 | 0.6 | 0.2 | 0.06 |
| procedure_occurrence during day -30 through 0 days relative to index: Other oxygen enrichment | 0.3 | 0.1 | 0.04 | 0.4 | 0.1 | 0.06 |
| condition_era group during day -365 through 0 days relative to index: Eczema | 0.7 | 0.5 | 0.03 | 1.0 | 0.5 | 0.06 |

### 4 Negative control outcomes

We selected negative controls using a process similar to that outlined in Voss et al. [1]. We first constructed a list of all conditions that satisfy the following criteria with respect to all drug exposures in the LEGEND-T2DM study [2]:

- No Medline abstract where the MeSH terms suggests a drug-condition association [3],
- No mention of the drug-condition pair on a US product label in the “Adverse Drug Reactions” or “Postmarketing” section [4],
- No US spontaneous reports suggesting that the pair is in an adverse event relationship [5, 6],
- OMOP vocabulary does not suggest that the drug is indicated for the condition,
- Vocabulary conditional concepts are usable (i.e., not too broad, not suggestive of an adverse event relationship, no pregnancy related), and
- Exact condition concept itself is used in patient level data.

We then optimized the remaining condition concepts, such that parent concepts remove children as defined by the OMOP vocabulary and performed manual review to exclude any pairs that may still be in a causal relationship or too similar to the study outcome. This process led to a candidate list of 100 negative controls:

**Supplementary Table 107: Negative control outcomes.**

| Condition concept |
|---|
| Abnormal posture |
| Abrasion and/or friction burn of multiple sites |
| Absent kidney |
| Anomaly of jaw size |
| Benign paroxysmal positional vertigo |
| Bizarre personal appearance |
| Cachexia |
| Colostomy present |
| Complication of gastrostomy |
| Developmental delay |
| Deviated nasal septum |
| Epidermoid cyst |
| Exhaustion due to excessive exertion |
| Feces contents abnormal |
| Foreign body in ear |
| Foreskin deficient |
| Galactosemia |
| Ganglion cyst |
| Genetic disorder carrier |
| Impacted cerumen |
| Inadequate sleep hygiene |
| Jellyfish poisoning |
| Lagophthalmos |
| Lipid storage disease |
| Lymphangioma |
| Malingering |
| Marfan’s syndrome |
| Mechanical complication of internal orthopedic device, implant AND/OR graft |
| Minimal cognitive impairment |
| Nicotine dependence |
| Nonspecific tuberculin test reaction |
| Opioid abuse |

| |
|---|
| Opioid intoxication |
| Physiological development failure |
| Poisoning by tranquilizer |
| Social exclusion |
| Symbolic dysfunction |
| Tooth loss |
| Toxic effect of lead compound |
| Toxic effect of tobacco and nicotine |
| Tracheostomy present |
| Unsatisfactory tooth restoration |
| Abnormal pupil |
| Abrasion and/or friction burn of trunk without infection |
| Absence of breast |
| Acquired hallux valgus |
| Acquired keratoderma |
| Anal and rectal polyp |
| Burn of forearm |
| Calcaneal spur |
| Cannabis abuse |
| Changes in skin texture |
| Chondromalacia of patella |
| Cocaine abuse |
| Complication due to Crohn’s disease |
| Contact dermatitis |
| Contusion of knee |
| Crohn’s disease |
| Derangement of knee |
| Difficulty sleeping |
| Disproportion of reconstructed breast |
| Effects of hunger |
| Endometriosis |
| Feces contents abnormal |
| Foreign body in orifice |
| Ganglion cyst |
| Hammer toe |
| Hereditary thrombophilia |
| High risk sexual behavior |
| Homocystinuria |
| Impacted cerumen |
| Impingement syndrome of shoulder region |
| Ingrowing nail |
| Injury of knee |
| Kwashiorkor |
| Late effect of contusion |
| Late effect of motor vehicle accident |
| Macular drusen |
| Melena |
| Nicotine dependence |
| Noise effects on inner ear |
| Non-toxic multinodular goiter |
| Nonspecific tuberculin test reaction |
| Opioid abuse |
| Passing flatus |

| |
|---|
| Postviral fatigue syndrome |
| Presbyopia |
| Psychalgia |
| Ptotic breast |
| Regular astigmatism |
| Senile hyperkeratosis |
| Somatic dysfunction of lumbar region |
| Splinter of face without major open wound |
| Sprain of ankle |
| Strain of rotator cuff capsule |
| Tear film insufficiency |
| Tobacco dependence syndrome |
| Verruca vulgaris |
| Wrist joint pain |
| Wristdrop |

### 5 Study diagnostics for final inclusion in meta-analysis

**Supplementary Figure 36:** Study diagnostic figure across different comparison pairs and databases for Abnormal weight gain.

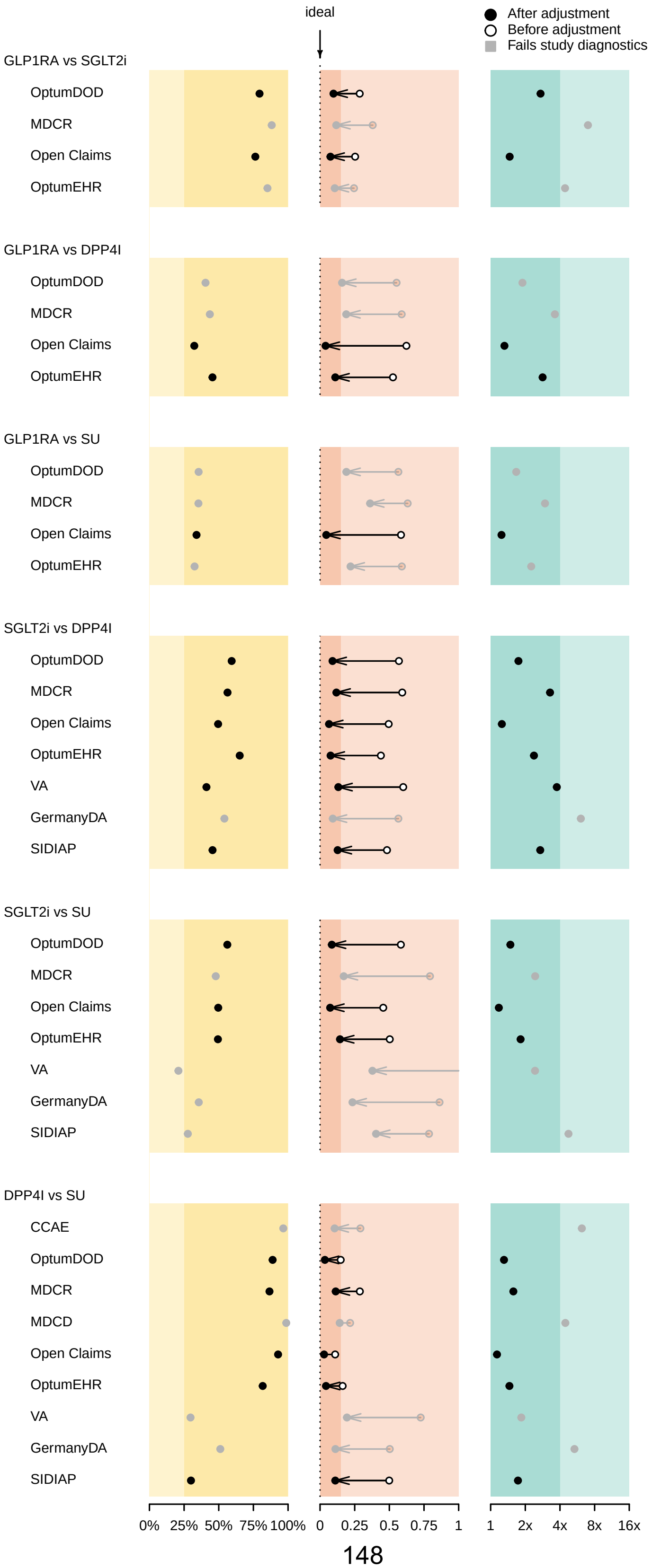

**Supplementary Figure 37:** Study diagnostic figure across different comparison pairs and databases for Abnormal weight loss.

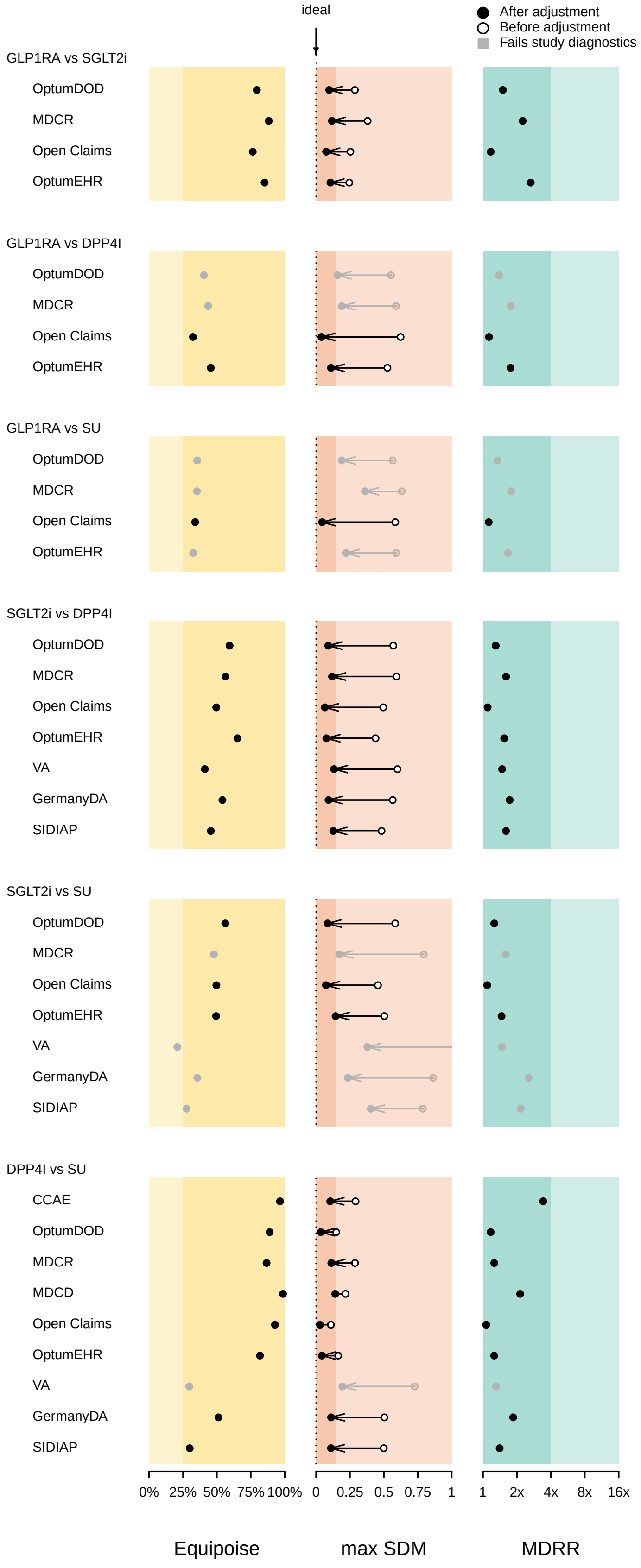

**Supplementary Figure 38:** Study diagnostic figure across different comparison pairs and databases for Acute pancreatitis.

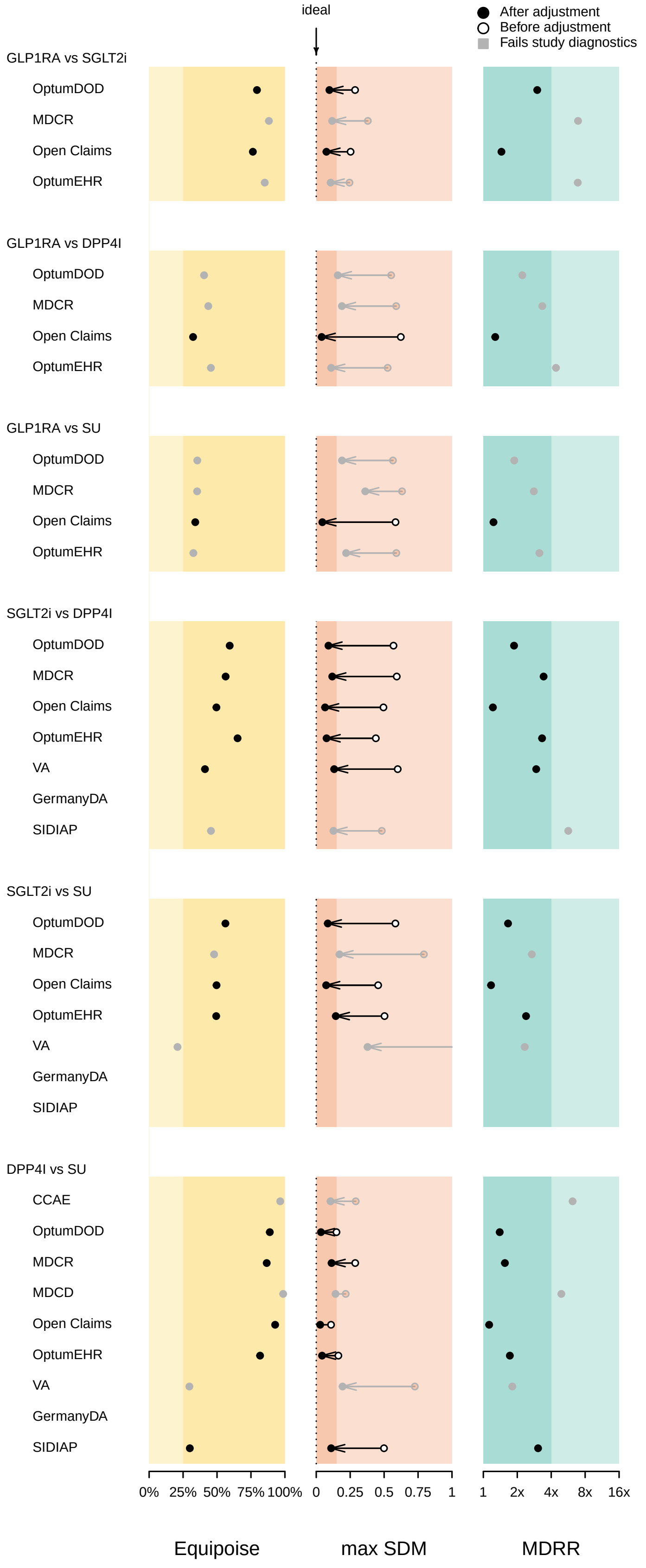

**Supplementary Figure 39:** Study diagnostic figure across different comparison pairs and databases for All-cause mortality.

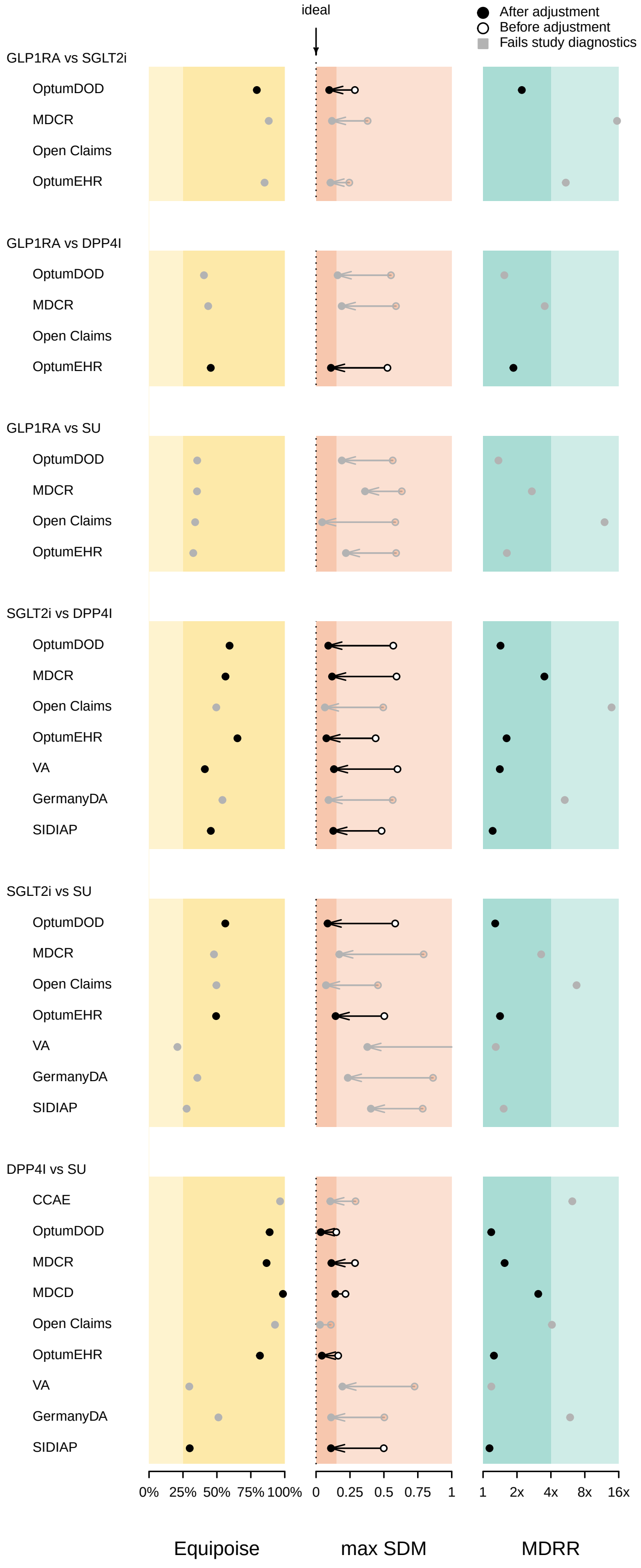

**Supplementary Figure 40:** Study diagnostic figure across different comparison pairs and databases for Bone fracture.

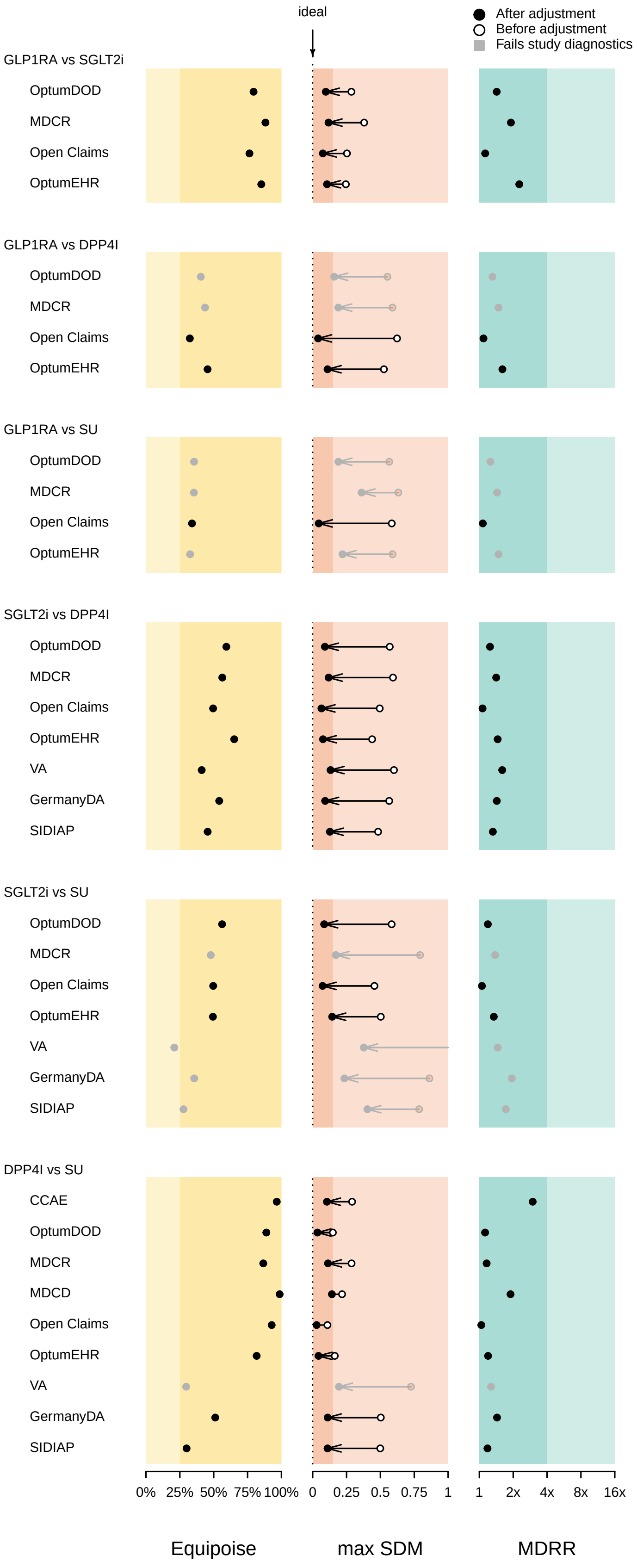

**Supplementary Figure 41:** Study diagnostic figure across different comparison pairs and databases for Diabetic ketoacidosis.

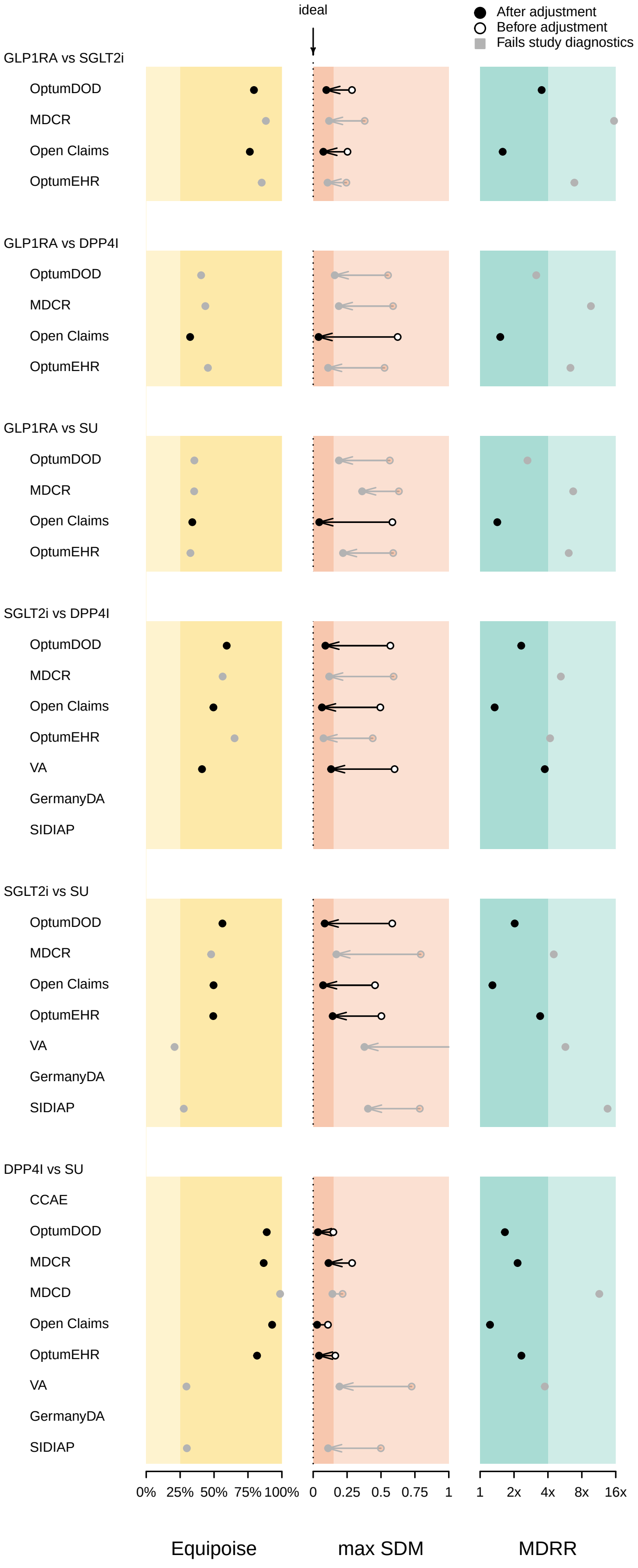

**Supplementary Figure 42:** Study diagnostic figure across different comparison pairs and databases for Diarrhea.

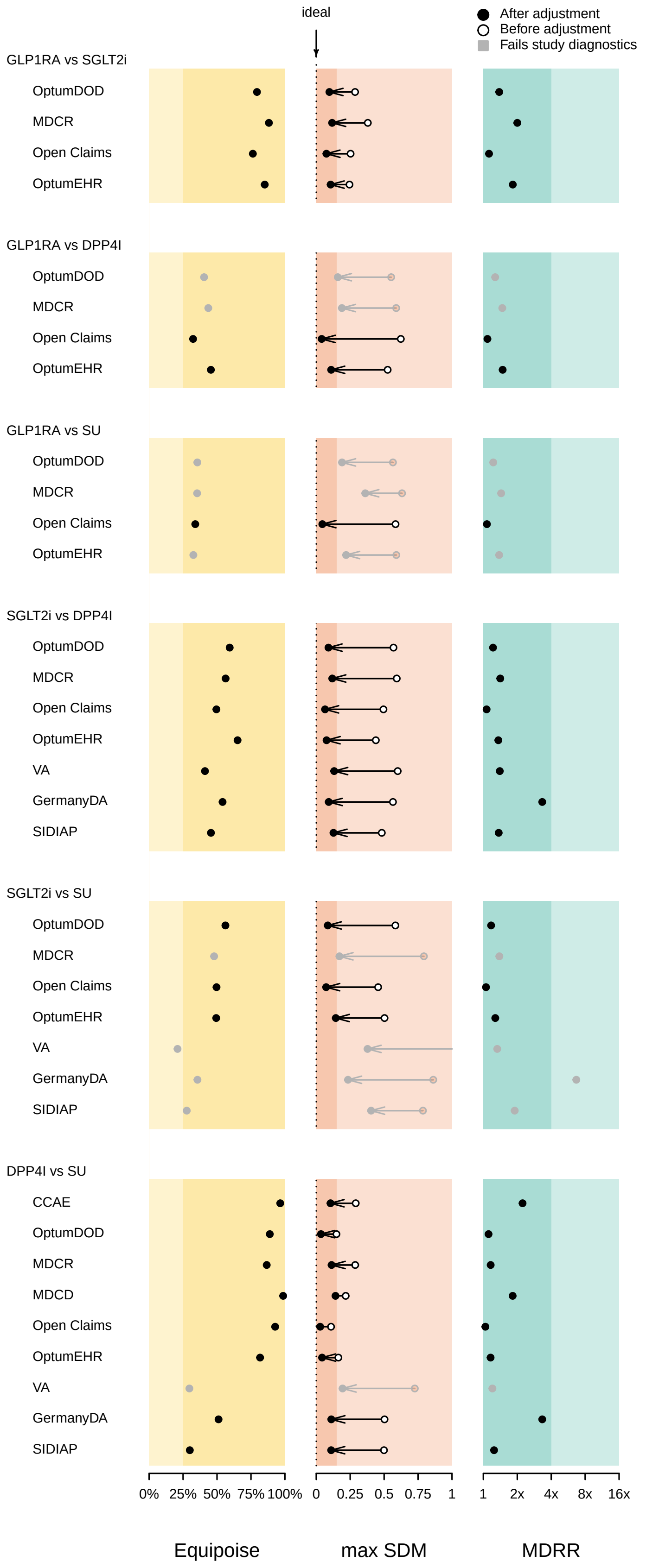

**Supplementary Figure 43:** Study diagnostic figure across different comparison pairs and databases for Genitourinary infection.

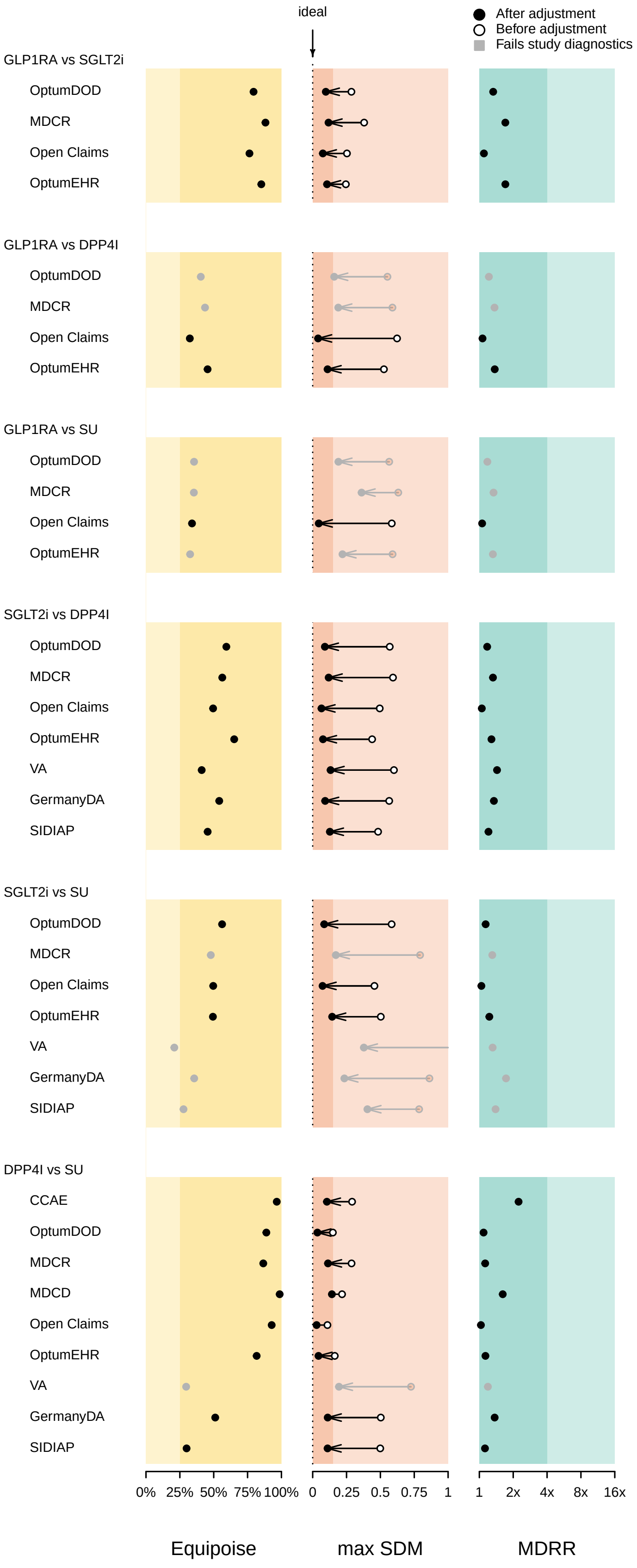

**Supplementary Figure 44:** Study diagnostic figure across different comparison pairs and databases for Hyperkalemia.

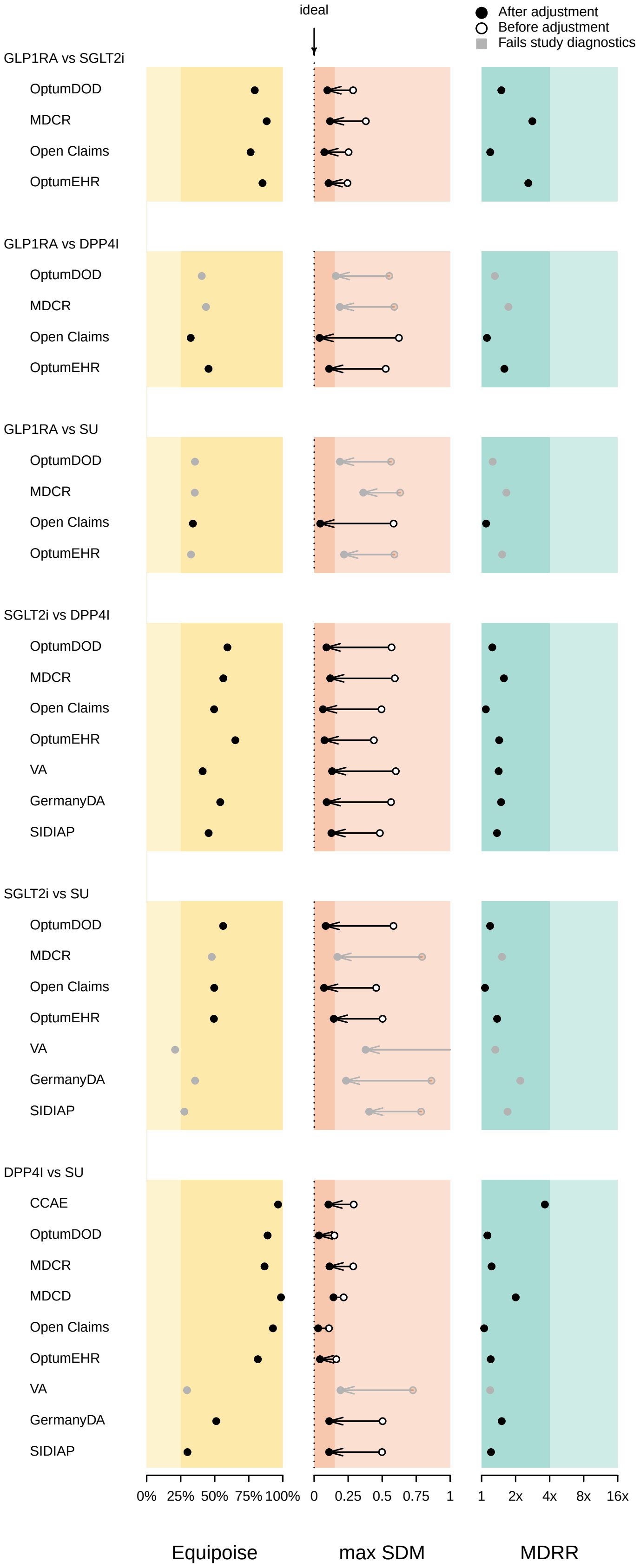

**Supplementary Figure 45:** Study diagnostic figure across different comparison pairs and databases for Hypoglycemia.

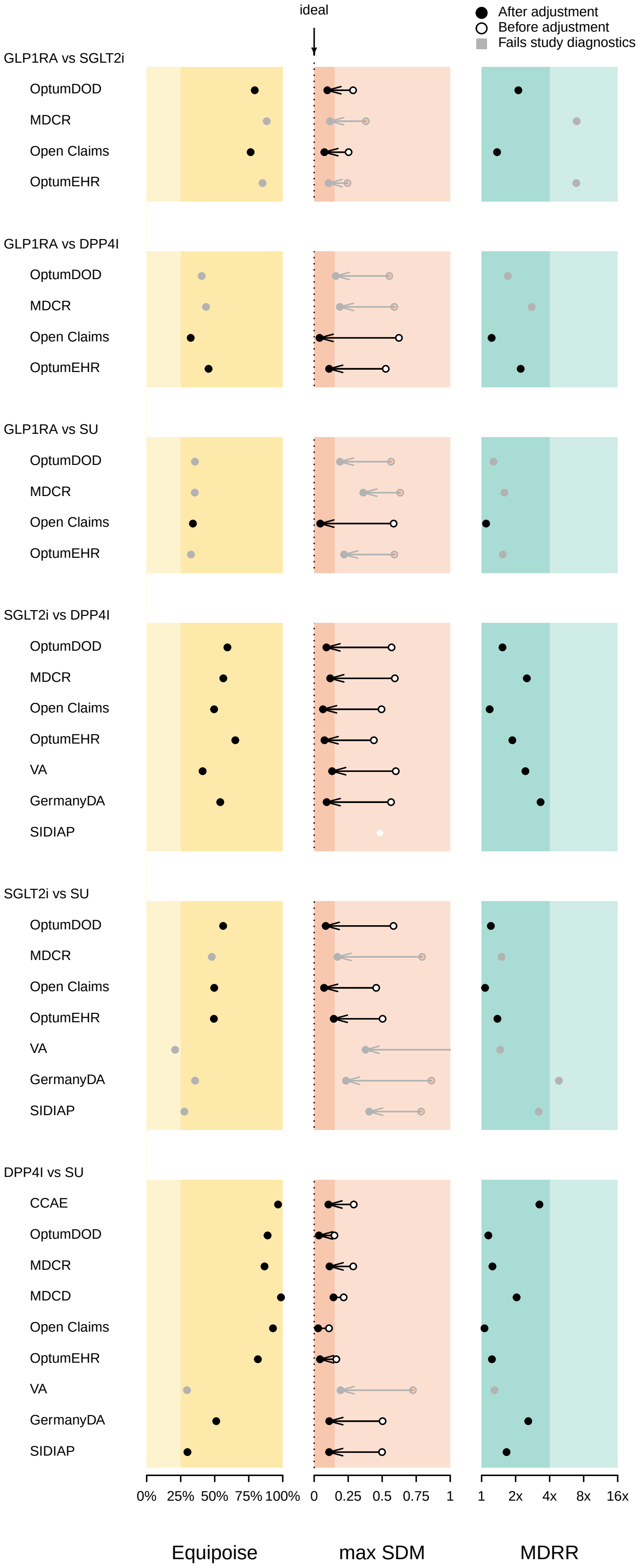

**Supplementary Figure 46:** Study diagnostic figure across different comparison pairs and databases for Hypotension.

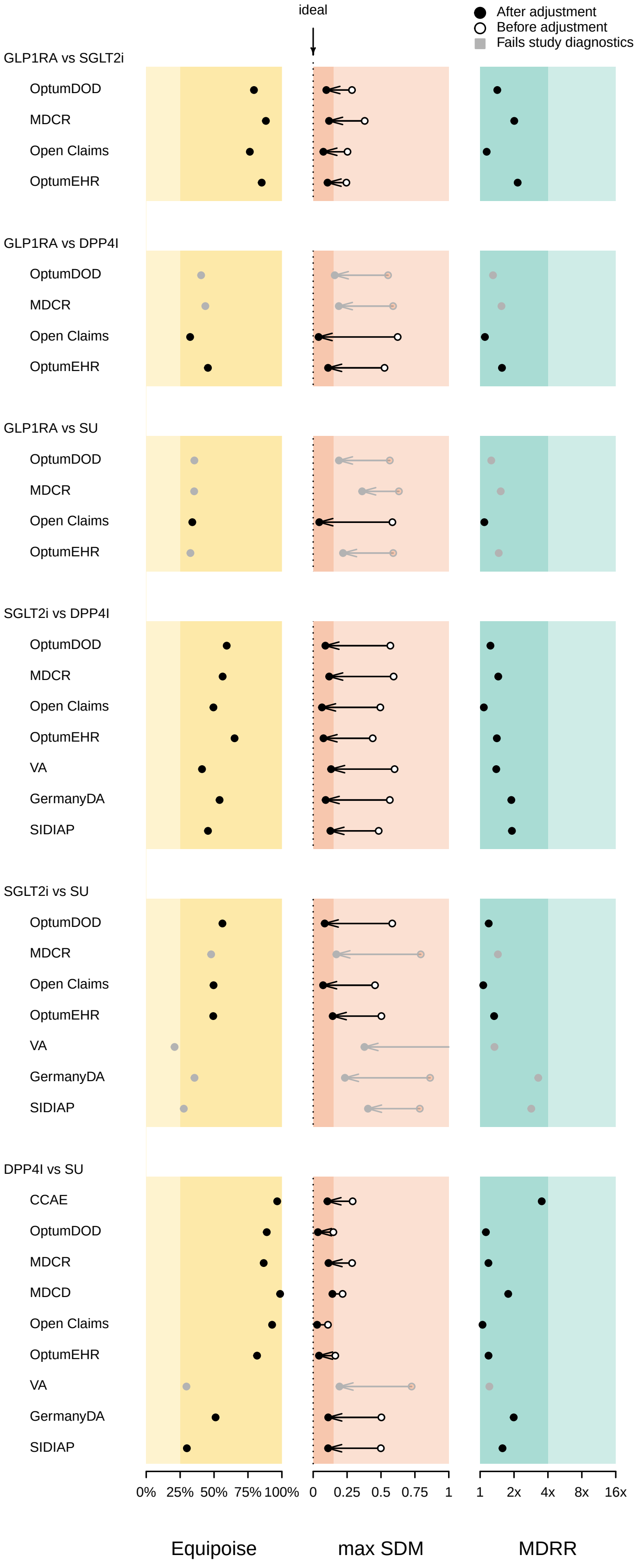

**Supplementary Figure 47:** Study diagnostic figure across different comparison pairs and databases for Joint pain.

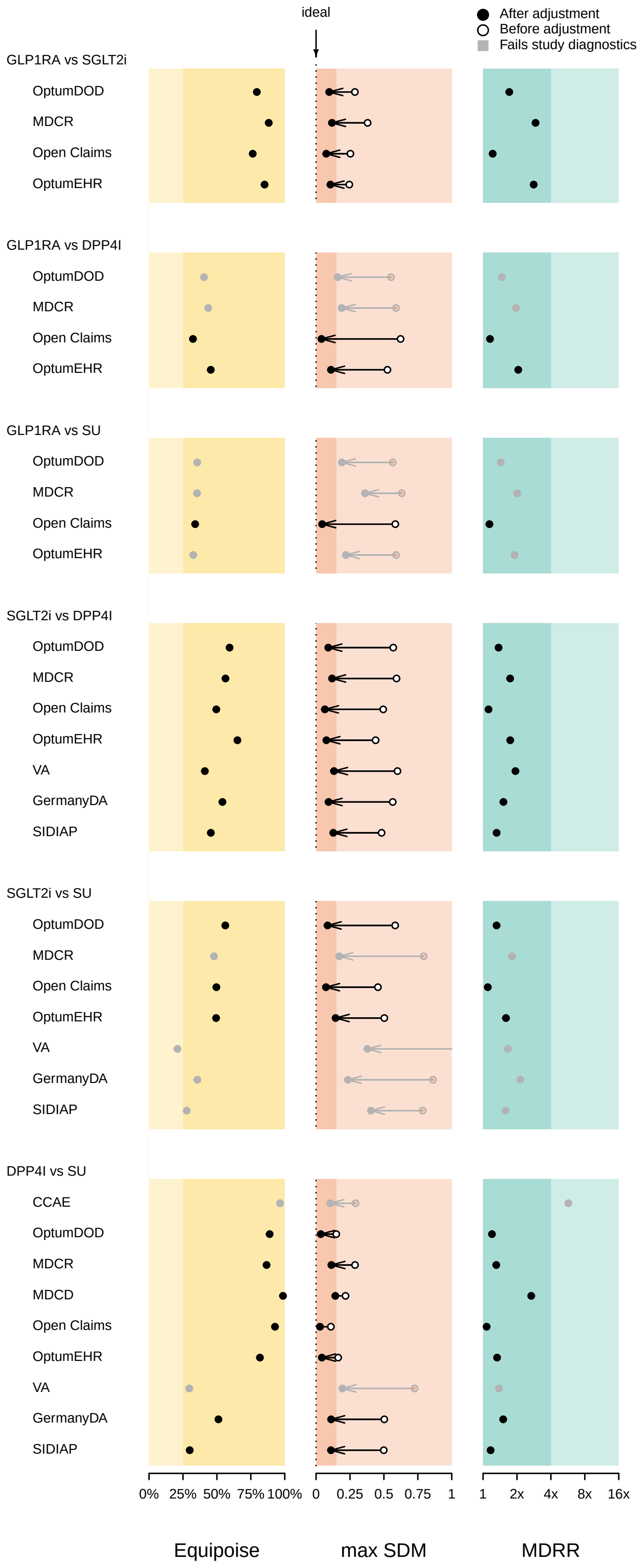

**Supplementary Figure 48:** Study diagnostic figure across different comparison pairs and databases for Lower extremity amputation.

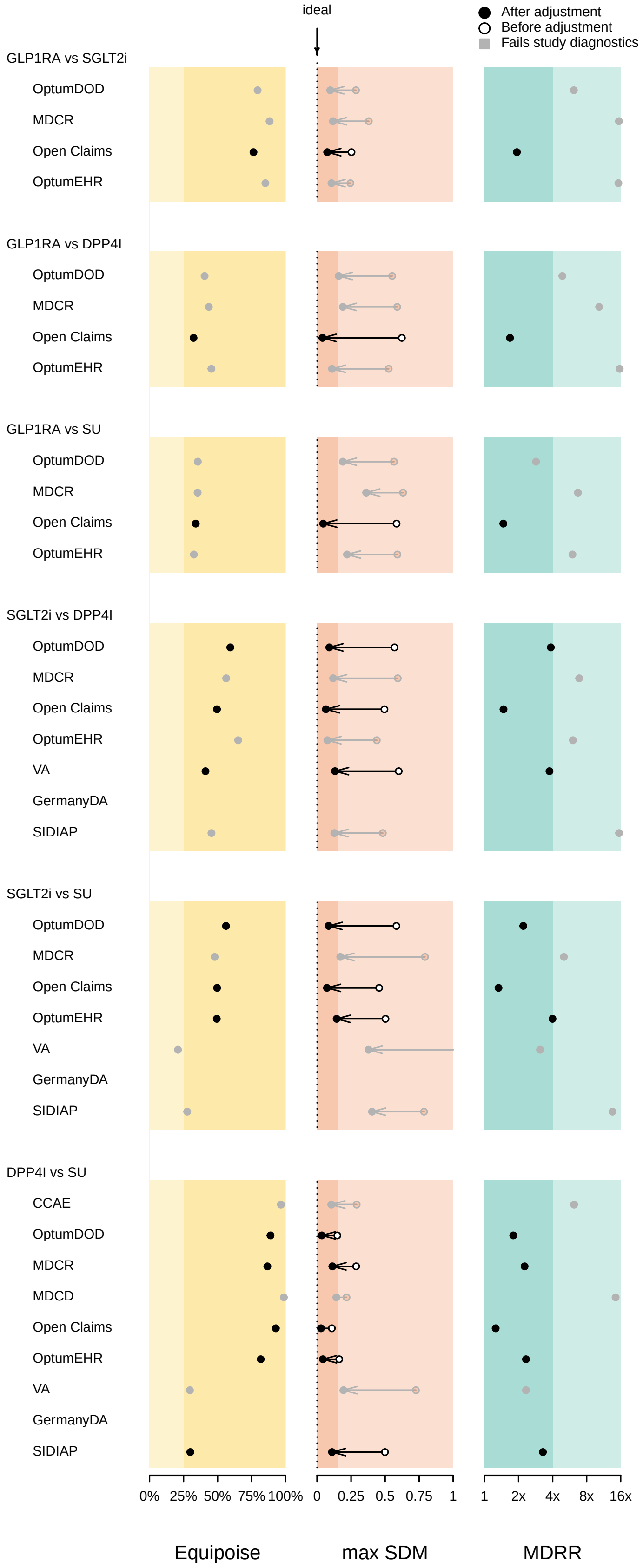

**Supplementary Figure 49:** Study diagnostic figure across different comparison pairs and databases for Nausea.

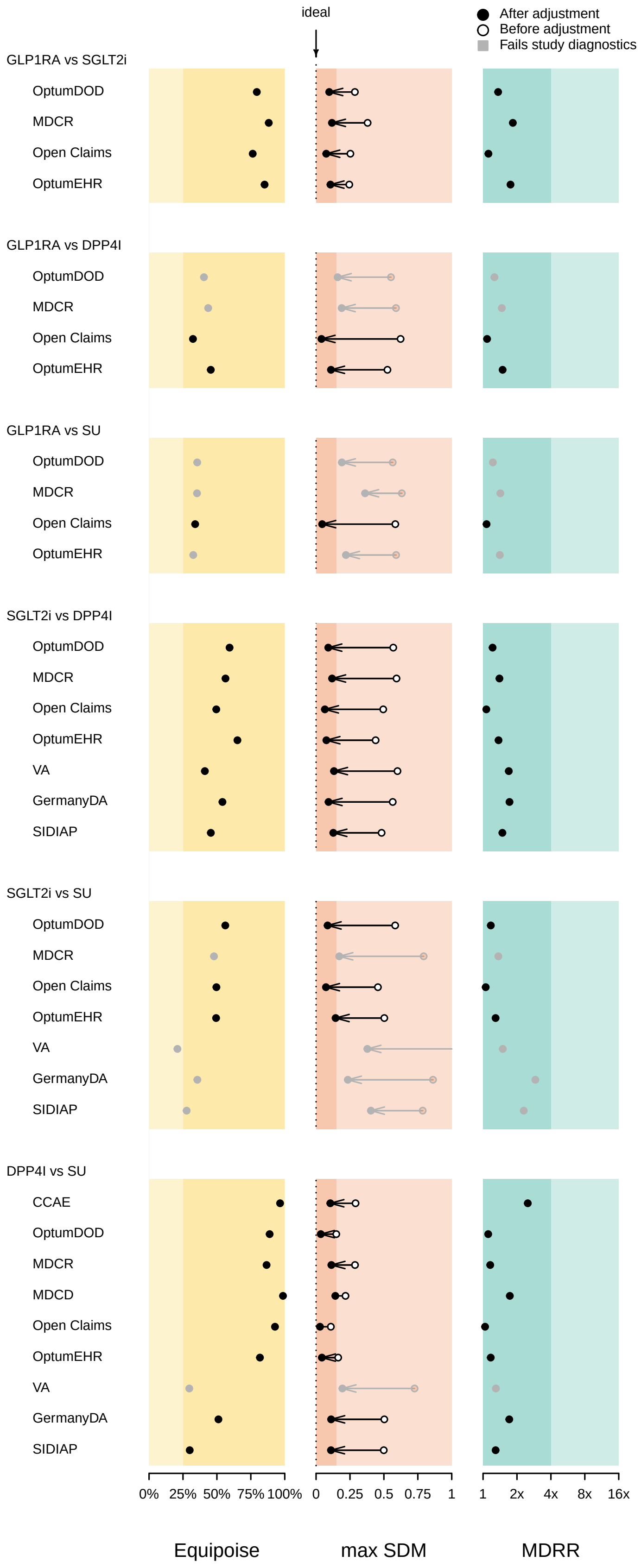

**Supplementary Figure 50:** Study diagnostic figure across different comparison pairs and databases for Peripheral edema.

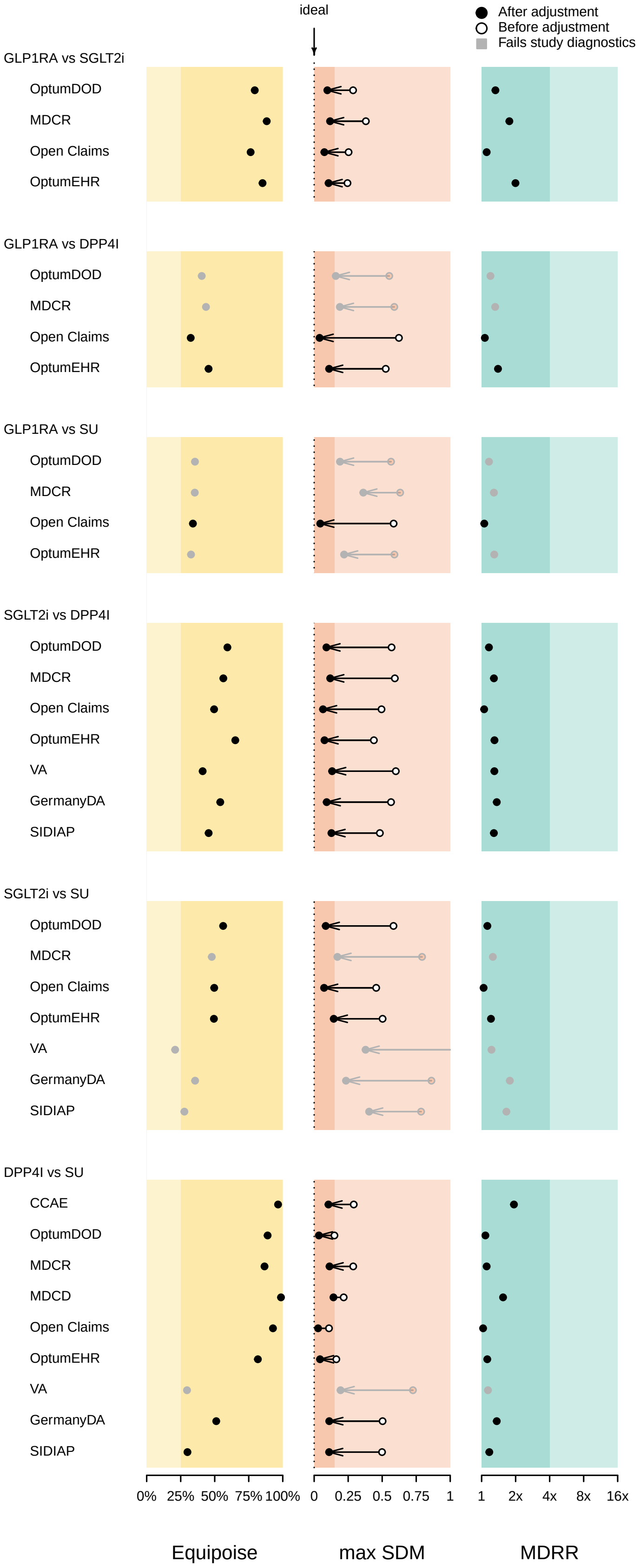

**Supplementary Figure 51:** Study diagnostic figure across different comparison pairs and databases for Photosensitivity.

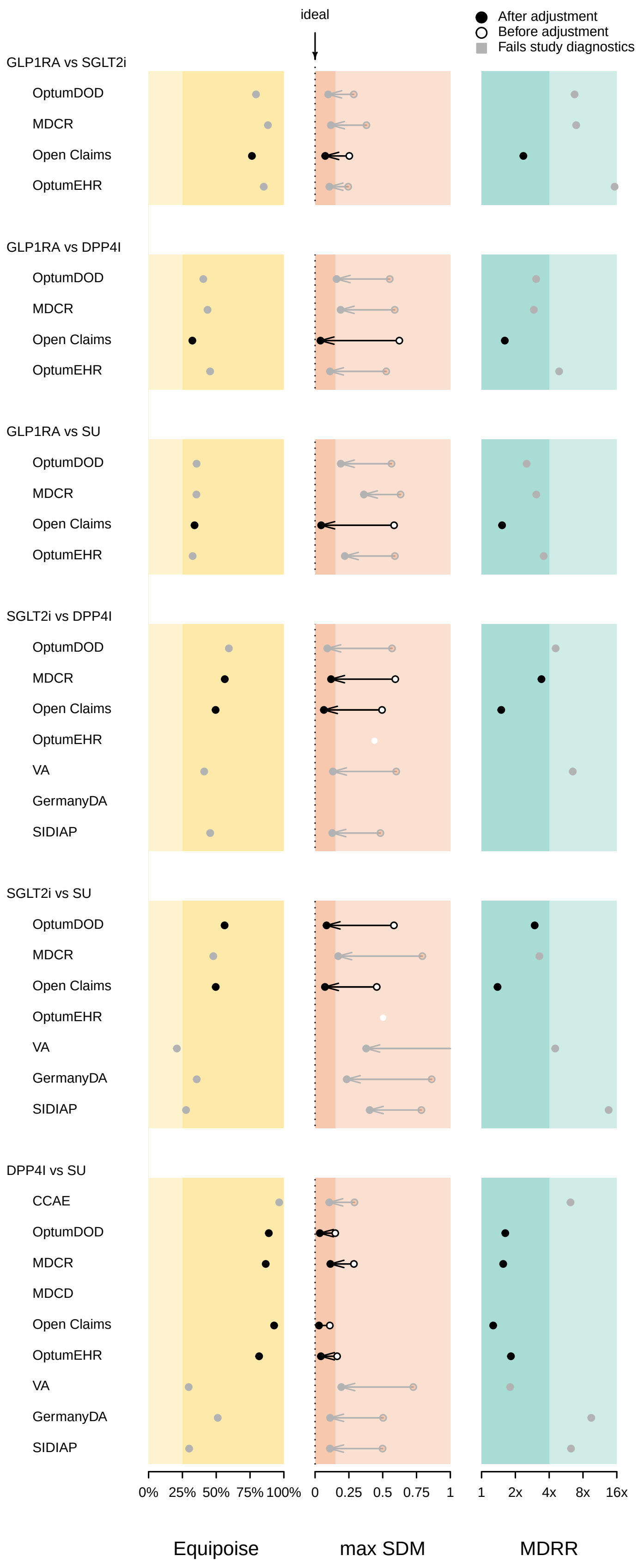

**Supplementary Figure 52:** Study diagnostic figure across different comparison pairs and databases for Venous thromboembolism.

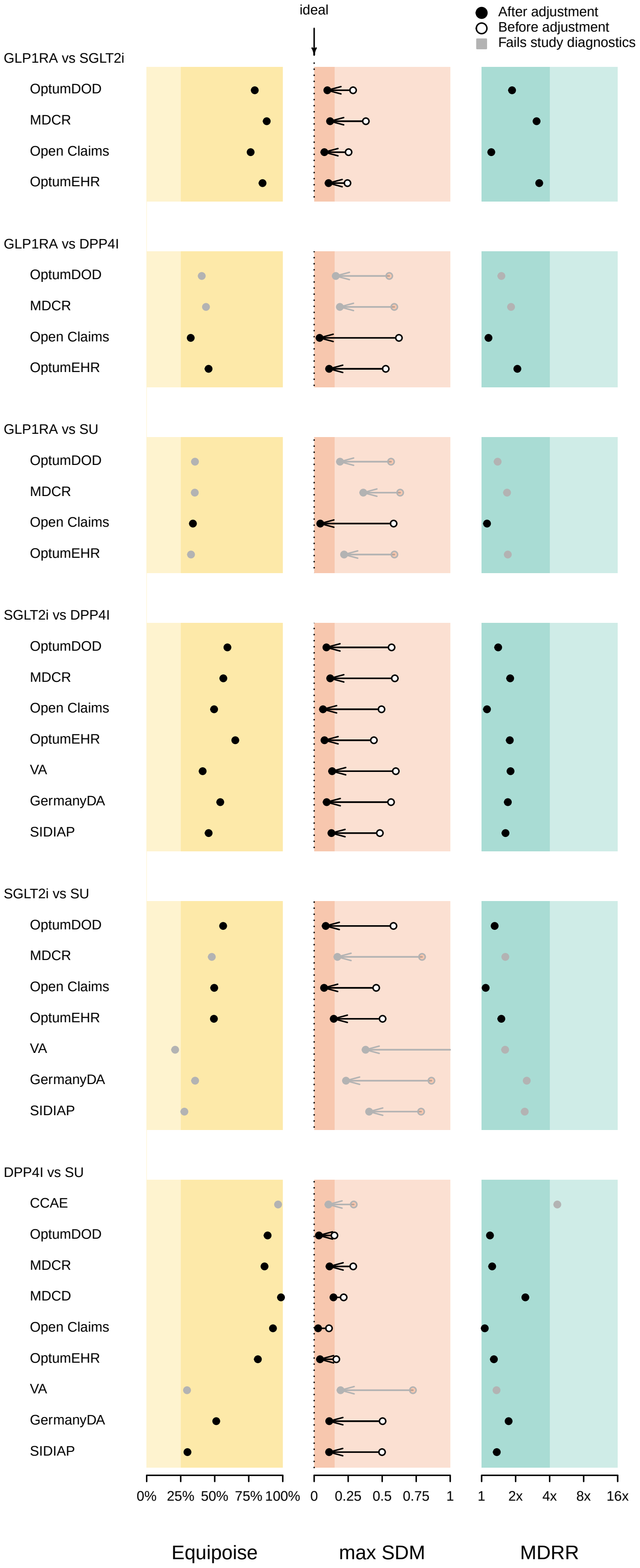

**Supplementary Figure 53:** Study diagnostic figure across different comparison pairs and databases for Vomiting.

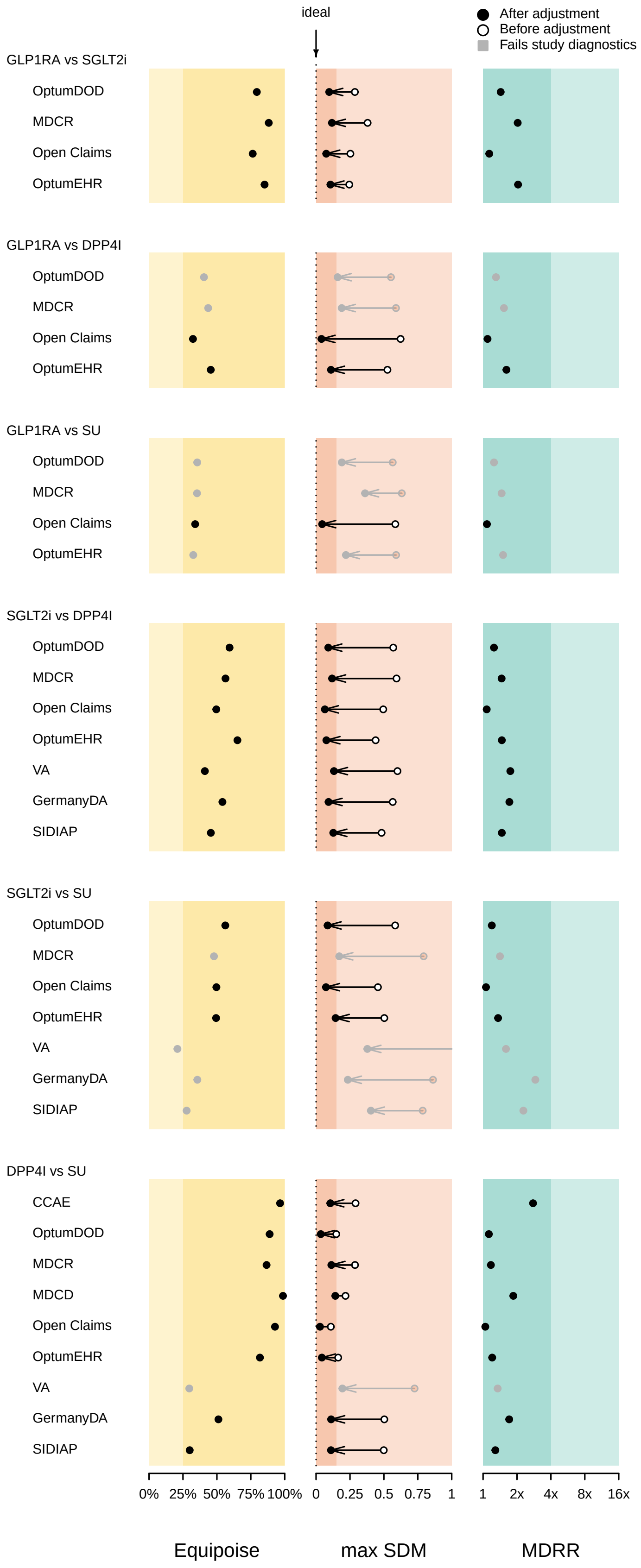

### 6 Exposures, outcomes and risk across data sources and outcomes

#### 6.1 GLP1RA vs SGLT2I

##### 6.1.1 Abnormal weight gain

**Supplementary Table 108:** Exposures, outcomes and risk estimates for Abnormal weight gain in GLP1RA (T) vs SGLT2I (C) new-user cohorts under four different propensity score (PS) adjustment / time-at-risk (TAR) designs across data sources. We report total patient exposure time in 1,000 patient-year (KYr) units, uncalibrated and calibrated hazard ratios (HRs) and their 95% confidence intervals (CIs). Greyed out entries did not pass study diagnostics and are included only for completeness.

| | Patients | | Exposure (KYr) | | Outcomes | | Uncalibrated | | Calibrated | |
|---|---|---|---|---|---|---|---|---|---|---|
| | T | C | T | C | T | C | HR (95% CI) | $p$ | HR (95% CI) | $p$ |
| **PS-stratification / on-treatment TAR** | | | | | | | | | | |
| OptumDOD | 5,009 | 11,275 | 2.5 | 6.7 | 17 | 20 | 1.36 (0.68 - 2.73) | 0.38 | 1.38 (0.69 - 2.78) | 0.36 |
| MDCR | 1,427 | 3,444 | 0.9 | 2.5 | 5 | <5 | 3.08 (0.77 - 12.9) | 0.12 | 2.68 (0.66 - 11.0) | 0.17 |
| Open Claims | 81,047 | 177,824 | 49.4 | 123.7 | 108 | 144 | 1.51 (1.15 - 1.97) | <0.01 | 1.46 (1.11 - 1.91) | <0.01 |
| OptumEHR | 3,184 | 7,560 | 0.9 | 2.1 | 9 | 8 | 2.80 (0.97 - 8.43) | 0.06 | 2.72 (0.92 - 8.04) | 0.07 |
| Meta-analysis | 86,056 | 189,099 | 52.0 | 130.5 | 125 | 164 | 1.49 (1.16 - 1.91) | <0.01 | 1.44 (1.11 - 1.85) | <0.01 |
| **PS-matching / on-treatment TAR** | | | | | | | | | | |
| OptumDOD | 4,556 | 11,259 | 2.3 | 6.7 | 13 | 20 | 0.64 (0.17 - 1.98) | 0.47 | 0.71 (0.21 - 2.43) | 0.59 |
| MDCR | 1,348 | 3,436 | 0.8 | 2.5 | 5 | <5 | | 0.21 | | 0.22 |
| Open Claims | 74,946 | 177,769 | 46.5 | 123.7 | 91 | 144 | 1.75 (1.17 - 2.60) | <0.01 | 1.66 (1.11 - 2.47) | 0.01 |
| OptumEHR | 3,008 | 7,548 | 0.8 | 2.1 | 8 | 7 | | | | |
| Meta-analysis | 79,502 | 189,028 | 48.8 | 130.4 | 104 | 164 | 1.25 (0.49 - 3.18) | 0.63 | 1.23 (0.48 - 3.12) | 0.66 |
| **PS-stratification / intent-to-treat TAR** | | | | | | | | | | |
| OptumDOD | 5,009 | 11,275 | 6.4 | 17.1 | 43 | 78 | 1.06 (0.71 - 1.56) | 0.78 | 1.06 (0.71 - 1.58) | 0.80 |
| MDCR | 1,427 | 3,444 | 1.9 | 5.8 | 18 | 18 | 2.91 (1.45 - 5.81) | <0.01 | 2.67 (1.34 - 5.35) | <0.01 |
| Open Claims | 81,047 | 177,824 | 167.5 | 438.9 | 469 | 750 | 1.34 (1.19 - 1.52) | <0.01 | 1.30 (1.12 - 1.51) | <0.01 |
| OptumEHR | 3,184 | 7,560 | 5.6 | 15.5 | 46 | 61 | 1.77 (1.17 - 2.66) | <0.01 | 1.78 (1.18 - 2.69) | <0.01 |
| Meta-analysis | 90,667 | 200,103 | 181.5 | 477.2 | 576 | 907 | 1.51 (1.09 - 2.08) | 0.01 | 1.47 (1.06 - 2.06) | 0.02 |
| **PS-matching / intent-to-treat TAR** | | | | | | | | | | |
| OptumDOD | 4,556 | 11,259 | 6.0 | 17.1 | 36 | 78 | 1.14 (0.65 - 1.97) | 0.63 | 1.16 (0.67 - 2.03) | 0.60 |
| MDCR | 1,348 | 3,436 | 1.8 | 5.8 | 18 | 18 | 3.88 (1.48 - 11.4) | <0.01 | 3.36 (1.20 - 9.41) | 0.03 |
| Open Claims | 74,946 | 177,769 | 158.2 | 438.8 | 419 | 750 | 1.39 (1.18 - 1.63) | <0.01 | 1.34 (1.13 - 1.59) | <0.01 |
| OptumEHR | 3,008 | 7,548 | 5.4 | 15.4 | 43 | 59 | 2.45 (1.44 - 4.21) | <0.01 | 2.53 (1.48 - 4.33) | <0.01 |
| Meta-analysis | 83,858 | 200,012 | 171.4 | 477.1 | 516 | 905 | 1.73 (1.12 - 2.68) | 0.01 | 1.71 (1.10 - 2.65) | 0.02 |

##### 6.1.2 Abnormal weight loss

**Supplementary Table 109:** Exposures, outcomes and risk estimates for Abnormal weight loss in GLP1RA (T) vs SGLT2I (C) new-user cohorts under four different propensity score (PS) adjustment / time-at-risk (TAR) designs across data sources. We report total patient exposure time in 1,000 patient-year (KYr) units, uncalibrated and calibrated hazard ratios (HRs) and their 95% confidence intervals (CIs). Greyed out entries did not pass study diagnostics and are included only for completeness.

| | Patients | | Exposure (KYr) | | Outcomes | | Uncalibrated | | Calibrated | |
|---|---|---|---|---|---|---|---|---|---|---|
| | T | C | T | C | T | C | HR (95% CI) | $p$ | HR (95% CI) | $p$ |
| **PS-stratification / on-treatment TAR** | | | | | | | | | | |
| OptumDOD | 5,123 | 11,094 | 2.6 | 6.6 | 59 | 163 | 0.97 (0.71 - 1.33) | 0.87 | 0.99 (0.72 - 1.36) | 0.88 |
| MDCR | 1,458 | 3,392 | 0.9 | 2.5 | 13 | 44 | 0.92 (0.46 - 1.71) | 0.79 | 0.80 (0.41 - 1.54) | 0.51 |
| Open Claims | 83,825 | 176,400 | 50.9 | 122.5 | 368 | 1,091 | 0.90 (0.80 - 1.02) | 0.11 | 0.87 (0.77 - 0.99) | 0.05 |
| OptumEHR | 3,296 | 7,530 | 0.9 | 2.1 | 9 | 30 | 0.61 (0.26 - 1.29) | 0.22 | 0.59 (0.27 - 1.31) | 0.20 |
| Meta-analysis | 93,702 | 198,416 | 55.3 | 133.7 | 449 | 1,328 | 0.90 (0.81 - 1.01) | 0.08 | 0.87 (0.77 - 0.99) | 0.03 |
| **PS-matching / on-treatment TAR** | | | | | | | | | | |
| OptumDOD | 4,578 | 11,078 | 2.4 | 6.6 | 55 | 163 | 1.06 (0.70 - 1.58) | 0.79 | 1.18 (0.78 - 1.78) | 0.44 |
| MDCR | 1,363 | 3,384 | 0.9 | 2.5 | 13 | 43 | 1.02 (0.45 - 2.17) | 0.96 | 0.96 (0.44 - 2.13) | 0.87 |
| Open Claims | 76,528 | 176,352 | 47.3 | 122.5 | 340 | 1,088 | 0.84 (0.71 - 0.99) | 0.04 | 0.80 (0.68 - 0.95) | 0.01 |
| OptumEHR | 3,081 | 7,518 | 0.8 | 2.1 | 7 | 30 | 0.75 (0.26 - 1.95) | 0.58 | 0.69 (0.25 - 1.90) | 0.48 |
| Meta-analysis | 85,550 | 198,332 | 51.4 | 133.6 | 415 | 1,324 | 0.87 (0.75 - 1.01) | 0.07 | 0.86 (0.73 - 1.00) | 0.05 |
| **PS-stratification / intent-to-treat TAR** | | | | | | | | | | |
| OptumDOD | 5,123 | 11,094 | 6.5 | 16.5 | 142 | 362 | 1.04 (0.85 - 1.28) | 0.69 | 1.04 (0.84 - 1.28) | 0.73 |
| MDCR | 1,458 | 3,392 | 1.9 | 5.6 | 29 | 111 | 0.82 (0.52 - 1.23) | 0.35 | 0.75 (0.49 - 1.15) | 0.20 |
| Open Claims | 83,825 | 176,400 | 171.5 | 433.7 | 1,212 | 3,566 | 0.91 (0.85 - 0.98) | 0.01 | 0.89 (0.79 - 0.99) | 0.05 |
| OptumEHR | 3,296 | 7,530 | 5.8 | 15.3 | 62 | 201 | 0.79 (0.58 - 1.05) | 0.12 | 0.79 (0.59 - 1.07) | 0.14 |
| Meta-analysis | 93,702 | 198,416 | 185.7 | 471.1 | 1,445 | 4,240 | 0.92 (0.86 - 0.98) | <0.01 | 0.90 (0.81 - 1.00) | 0.04 |
| **PS-matching / intent-to-treat TAR** | | | | | | | | | | |
| OptumDOD | 4,578 | 11,078 | 6.0 | 16.5 | 130 | 361 | 1.09 (0.84 - 1.41) | 0.51 | 1.11 (0.85 - 1.44) | 0.46 |
| MDCR | 1,363 | 3,384 | 1.8 | 5.6 | 27 | 110 | 1.08 (0.63 - 1.80) | 0.77 | 0.94 (0.55 - 1.60) | 0.82 |
| Open Claims | 76,528 | 176,352 | 160.4 | 433.6 | 1,115 | 3,563 | 0.89 (0.82 - 0.96) | <0.01 | 0.86 (0.77 - 0.95) | <0.01 |
| OptumEHR | 3,081 | 7,518 | 5.5 | 15.2 | 54 | 201 | 0.85 (0.58 - 1.22) | 0.40 | 0.88 (0.61 - 1.28) | 0.52 |
| Meta-analysis | 85,550 | 198,332 | 173.7 | 470.9 | 1,326 | 4,235 | 0.92 (0.82 - 1.04) | 0.18 | 0.91 (0.80 - 1.03) | 0.14 |

##### 6.1.3 Acute pancreatitis

**Supplementary Table 110:** Exposures, outcomes and risk estimates for Acute pancreatitis in GLP1RA (T) vs SGLT2I (C) new-user cohorts under four different propensity score (PS) adjustment / time-at-risk (TAR) designs across data sources. We report total patient exposure time in 1,000 patient-year (KYr) units, uncalibrated and calibrated hazard ratios (HRs) and their 95% confidence intervals (CIs). Greyed out entries did not pass study diagnostics and are included only for completeness.

| | Patients | | Exposure (KYr) | | Outcomes | | Uncalibrated | | Calibrated | |
|---|---|---|---|---|---|---|---|---|---|---|
| | T | C | T | C | T | C | HR (95% CI) | $p$ | HR (95% CI) | $p$ |
| **PS-stratification / on-treatment TAR** | | | | | | | | | | |
| OptumDOD | 5,302 | 11,512 | 2.7 | 6.9 | 9 | 21 | 1.06 (0.44 - 2.34) | 0.89 | 1.07 (0.47 - 2.47) | 0.87 |
| MDCR | 1,487 | 3,480 | 0.9 | 2.5 | <5 | <5 | 2.18 (0.36 - 11.8) | 0.38 | 1.90 (0.33 - 10.9) | 0.47 |
| Open Claims | 86,346 | 182,041 | 52.5 | 126.7 | 87 | 176 | 1.32 (1.00 - 1.72) | 0.05 | 1.27 (0.97 - 1.67) | 0.09 |
| OptumEHR | 3,385 | 7,801 | 0.9 | 2.2 | <5 | <5 | 2.25 (0.38 - 13.4) | 0.37 | 2.18 (0.37 - 12.9) | 0.39 |
| Meta-analysis | 91,648 | 193,553 | 55.2 | 133.6 | 96 | 197 | 1.29 (0.99 - 1.67) | 0.06 | 1.24 (0.96 - 1.62) | 0.11 |
| **PS-matching / on-treatment TAR** | | | | | | | | | | |
| OptumDOD | 4,737 | 11,496 | 2.5 | 6.9 | 7 | 21 | 0.62 (0.20 - 1.64) | 0.37 | 0.69 (0.24 - 1.99) | 0.49 |
| MDCR | 1,385 | 3,472 | 0.9 | 2.5 | <5 | <5 | 1.50 (0.18 - 10.5) | 0.70 | 1.42 (0.18 - 10.9) | 0.74 |
| Open Claims | 78,761 | 181,987 | 48.8 | 126.7 | 82 | 176 | 1.39 (0.97 - 2.00) | 0.07 | 1.33 (0.92 - 1.91) | 0.14 |
| OptumEHR | 3,162 | 7,789 | 0.8 | 2.2 | <5 | <5 | 0.00 (0.00 - 11.7) | 0.23 | 0.00 (0.00 - 91799) | 0.23 |
| Meta-analysis | 83,498 | 193,483 | 51.3 | 133.5 | 89 | 197 | 1.09 (0.52 - 2.27) | 0.83 | 1.06 (0.51 - 2.23) | 0.87 |
| **PS-stratification / intent-to-treat TAR** | | | | | | | | | | |
| OptumDOD | 5,302 | 11,512 | 6.8 | 17.6 | 26 | 54 | 1.22 (0.73 - 1.99) | 0.43 | 1.22 (0.74 - 2.02) | 0.44 |
| MDCR | 1,487 | 3,480 | 2.0 | 5.9 | 7 | 10 | 2.22 (0.74 - 6.27) | 0.14 | 2.04 (0.70 - 5.93) | 0.20 |
| Open Claims | 86,346 | 182,041 | 178.1 | 451.3 | 305 | 675 | 1.18 (1.02 - 1.36) | 0.02 | 1.14 (0.97 - 1.35) | 0.12 |
| OptumEHR | 3,385 | 7,801 | 6.0 | 16.1 | 10 | 20 | 1.15 (0.50 - 2.54) | 0.73 | 1.16 (0.51 - 2.63) | 0.72 |
| Meta-analysis | 95,033 | 201,354 | 191.0 | 485.0 | 341 | 749 | 1.18 (1.03 - 1.35) | 0.02 | 1.16 (0.99 - 1.35) | 0.07 |
| **PS-matching / intent-to-treat TAR** | | | | | | | | | | |
| OptumDOD | 4,737 | 11,496 | 6.3 | 17.6 | 21 | 54 | 0.88 (0.45 - 1.67) | 0.71 | 0.90 (0.46 - 1.74) | 0.75 |
| MDCR | 1,385 | 3,472 | 1.9 | 5.8 | 7 | 9 | 1.91 (0.44 - 7.87) | 0.38 | 1.66 (0.39 - 7.04) | 0.51 |
| Open Claims | 78,761 | 181,987 | 166.5 | 451.2 | 283 | 675 | 1.13 (0.95 - 1.35) | 0.16 | 1.09 (0.91 - 1.31) | 0.36 |
| OptumEHR | 3,162 | 7,789 | 5.7 | 16.0 | 9 | 20 | 1.90 (0.64 - 5.49) | 0.24 | 1.96 (0.67 - 5.76) | 0.22 |
| Meta-analysis | 86,660 | 201,272 | 178.4 | 484.8 | 313 | 749 | 1.13 (0.96 - 1.33) | 0.15 | 1.11 (0.94 - 1.32) | 0.23 |

##### 6.1.4 All-cause mortality

**Supplementary Table 111:** Exposures, outcomes and risk estimates for All-cause mortality in GLP1RA (T) vs SGLT2I (C) new-user cohorts under four different propensity score (PS) adjustment / time-at-risk (TAR) designs across data sources. We report total patient exposure time in 1,000 patient-year (KYr) units, uncalibrated and calibrated hazard ratios (HRs) and their 95% confidence intervals (CIs). Greyed out entries did not pass study diagnostics and are included only for completeness.

| | Patients | | Exposure (KYr) | | Outcomes | | Uncalibrated | | Calibrated | |
|---|---|---|---|---|---|---|---|---|---|---|
| | T | C | T | C | T | C | HR (95% CI) | $p$ | HR (95% CI) | $p$ |
| **PS-stratification / on-treatment TAR** | | | | | | | | | | |
| OptumDOD | 5,336 | 11,614 | 2.7 | 6.9 | 15 | 43 | 0.92 (0.48 - 1.67) | 0.78 | 0.93 (0.50 - 1.74) | 0.82 |
| MDCR | 1,493 | 3,510 | 0.9 | 2.6 | 0 | <5 | | | | |
| Open Claims | 87,367 | 184,807 | 53.1 | 128.6 | 0 | 0 | | | | |
| OptumEHR | 3,403 | 7,828 | 0.9 | 2.2 | <5 | 8 | 0.63 (0.13 - 2.40) | 0.54 | 0.62 (0.14 - 2.64) | 0.51 |
| Meta-analysis | 5,336 | 11,614 | 2.7 | 6.9 | 15 | 43 | 0.92 (0.48 - 1.67) | 0.78 | 0.88 (0.47 - 1.65) | 0.70 |
| **PS-matching / on-treatment TAR** | | | | | | | | | | |
| OptumDOD | 4,772 | 11,598 | 2.5 | 6.9 | 14 | 43 | 1.93 (0.79 - 4.59) | 0.14 | 2.16 (0.90 - 5.20) | 0.09 |
| MDCR | 1,393 | 3,502 | 0.9 | 2.6 | 0 | <5 | | | | |
| Open Claims | 79,722 | 184,753 | 49.4 | 128.6 | 0 | 0 | | | | |
| OptumEHR | 3,177 | 7,816 | 0.8 | 2.2 | <5 | 8 | 0.41 (0.02 - 2.97) | 0.49 | 0.38 (0.03 - 4.60) | 0.45 |
| Meta-analysis | 4,772 | 11,598 | 2.5 | 6.9 | 14 | 43 | 1.93 (0.79 - 4.59) | 0.14 | 1.89 (0.79 - 4.56) | 0.15 |
| **PS-stratification / intent-to-treat TAR** | | | | | | | | | | |
| OptumDOD | 5,336 | 11,614 | 6.9 | 17.7 | 134 | 359 | 1.11 (0.90 - 1.37) | 0.32 | 1.11 (0.90 - 1.38) | 0.39 |
| MDCR | 1,493 | 3,510 | 2.0 | 5.9 | <5 | <5 | 1.62 (0.20 - 10.2) | 0.63 | 1.49 (0.21 - 10.5) | 0.69 |
| Open Claims | 87,367 | 184,807 | 180.7 | 459.1 | 0 | <5 | | | | |
| OptumEHR | 3,403 | 7,828 | 6.1 | 16.1 | 37 | 119 | 0.79 (0.53 - 1.15) | 0.24 | 0.80 (0.54 - 1.18) | 0.26 |
| Meta-analysis | 8,739 | 19,442 | 12.9 | 33.9 | 171 | 478 | 0.98 (0.71 - 1.35) | 0.89 | 0.96 (0.68 - 1.33) | 0.79 |
| **PS-matching / intent-to-treat TAR** | | | | | | | | | | |
| OptumDOD | 4,772 | 11,598 | 6.3 | 17.7 | 128 | 354 | 0.98 (0.74 - 1.29) | 0.88 | 1.00 (0.75 - 1.32) | 0.89 |
| MDCR | 1,393 | 3,502 | 1.9 | 5.9 | <5 | <5 | 1.41 (0.05 - 36.7) | 0.83 | 1.22 (0.05 - 31.8) | 0.90 |
| Open Claims | 79,722 | 184,753 | 168.9 | 459.0 | 0 | <5 | | | | |
| OptumEHR | 3,177 | 7,816 | 5.7 | 16.1 | 34 | 118 | 0.87 (0.53 - 1.39) | 0.58 | 0.90 (0.56 - 1.46) | 0.67 |
| Meta-analysis | 7,949 | 19,414 | 12.1 | 33.8 | 162 | 472 | 0.95 (0.75 - 1.21) | 0.68 | 0.94 (0.73 - 1.20) | 0.60 |

##### 6.1.5 Bone fracture

**Supplementary Table 112:** Exposures, outcomes and risk estimates for Bone fracture in GLP1RA (T) vs SGLT2I (C) new-user cohorts under four different propensity score (PS) adjustment / time-at-risk (TAR) designs across data sources. We report total patient exposure time in 1,000 patient-year (KYr) units, uncalibrated and calibrated hazard ratios (HRs) and their 95% confidence intervals (CIs). Greyed out entries did not pass study diagnostics and are included only for completeness.

| | Patients | | Exposure (KYr) | | Outcomes | | Uncalibrated | | Calibrated | |
|---|---|---|---|---|---|---|---|---|---|---|
| | T | C | T | C | T | C | HR (95% CI) | $p$ | HR (95% CI) | $p$ |
| **PS-stratification / on-treatment TAR** | | | | | | | | | | |
| OptumDOD | 4,701 | 10,423 | 2.4 | 6.1 | 90 | 200 | 1.04 (0.79 - 1.35) | 0.77 | 1.06 (0.81 - 1.38) | 0.70 |
| MDCR | 1,295 | 3,143 | 0.8 | 2.3 | 18 | 73 | 0.59 (0.33 - 1.00) | 0.06 | 0.51 (0.30 - 0.89) | 0.02 |
| Open Claims | 70,647 | 155,987 | 42.7 | 108.3 | 754 | 1,810 | 0.98 (0.89 - 1.07) | 0.60 | 0.94 (0.86 - 1.04) | 0.27 |
| OptumEHR | 3,067 | 7,175 | 0.8 | 2.0 | 17 | 39 | 0.81 (0.43 - 1.45) | 0.49 | 0.79 (0.43 - 1.44) | 0.45 |
| Meta-analysis | 79,710 | 176,728 | 46.7 | 118.7 | 879 | 2,122 | 0.97 (0.89 - 1.05) | 0.44 | 0.93 (0.85 - 1.03) | 0.16 |
| **PS-matching / on-treatment TAR** | | | | | | | | | | |
| OptumDOD | 4,216 | 10,409 | 2.1 | 6.1 | 76 | 200 | 0.95 (0.65 - 1.37) | 0.77 | 1.06 (0.73 - 1.54) | 0.77 |
| MDCR | 1,212 | 3,136 | 0.7 | 2.2 | 17 | 73 | 0.67 (0.30 - 1.38) | 0.30 | 0.63 (0.30 - 1.36) | 0.26 |
| Open Claims | 65,162 | 155,863 | 40.1 | 108.3 | 685 | 1,807 | 0.97 (0.85 - 1.10) | 0.58 | 0.92 (0.81 - 1.04) | 0.24 |
| OptumEHR | 2,874 | 7,163 | 0.8 | 2.0 | 14 | 39 | 0.79 (0.35 - 1.65) | 0.54 | 0.73 (0.33 - 1.58) | 0.43 |
| Meta-analysis | 73,464 | 176,571 | 43.8 | 118.6 | 792 | 2,119 | 0.95 (0.84 - 1.07) | 0.40 | 0.93 (0.82 - 1.06) | 0.28 |
| **PS-stratification / intent-to-treat TAR** | | | | | | | | | | |
| OptumDOD | 4,701 | 10,423 | 5.9 | 15.3 | 198 | 493 | 0.94 (0.79 - 1.12) | 0.53 | 0.94 (0.79 - 1.13) | 0.58 |
| MDCR | 1,295 | 3,143 | 1.7 | 5.1 | 54 | 158 | 0.89 (0.64 - 1.22) | 0.48 | 0.82 (0.59 - 1.14) | 0.25 |
| Open Claims | 70,647 | 155,987 | 142.5 | 377.8 | 2,930 | 7,504 | 0.97 (0.93 - 1.02) | 0.25 | 0.95 (0.86 - 1.04) | 0.29 |
| OptumEHR | 3,067 | 7,175 | 5.4 | 14.4 | 91 | 281 | 0.76 (0.59 - 0.97) | 0.03 | 0.77 (0.60 - 0.98) | 0.05 |
| Meta-analysis | 79,710 | 176,728 | 155.4 | 412.6 | 3,273 | 8,436 | 0.94 (0.86 - 1.02) | 0.15 | 0.92 (0.81 - 1.03) | 0.16 |
| **PS-matching / intent-to-treat TAR** | | | | | | | | | | |
| OptumDOD | 4,216 | 10,409 | 5.4 | 15.3 | 177 | 493 | 0.91 (0.72 - 1.14) | 0.40 | 0.92 (0.73 - 1.16) | 0.51 |
| MDCR | 1,212 | 3,136 | 1.6 | 5.1 | 50 | 158 | 0.78 (0.51 - 1.17) | 0.24 | 0.67 (0.44 - 1.04) | 0.14 |
| Open Claims | 65,162 | 155,863 | 134.3 | 377.7 | 2,700 | 7,499 | 0.99 (0.94 - 1.05) | 0.73 | 0.96 (0.88 - 1.04) | 0.36 |
| OptumEHR | 2,874 | 7,163 | 5.1 | 14.4 | 85 | 281 | 0.81 (0.59 - 1.09) | 0.18 | 0.84 (0.62 - 1.14) | 0.28 |
| Meta-analysis | 73,464 | 176,571 | 146.4 | 412.4 | 3,012 | 8,431 | 0.94 (0.84 - 1.05) | 0.26 | 0.93 (0.82 - 1.04) | 0.21 |

##### 6.1.6 Diabetic ketoacidosis

**Supplementary Table 113:** Exposures, outcomes and risk estimates for Diabetic ketoacidosis in GLP1RA (T) vs SGLT2I (C) new-user cohorts under four different propensity score (PS) adjustment / time-at-risk (TAR) designs across data sources. We report total patient exposure time in 1,000 patient-year (KYr) units, uncalibrated and calibrated hazard ratios (HRs) and their 95% confidence intervals (CIs). Greyed out entries did not pass study diagnostics and are included only for completeness.

| | Patients | | Exposure (KYr) | | Outcomes | | Uncalibrated | | Calibrated | |
|---|---|---|---|---|---|---|---|---|---|---|
| | T | C | T | C | T | C | HR (95% CI) | $p$ | HR (95% CI) | $p$ |
| **PS-stratification / on-treatment TAR** | | | | | | | | | | |
| OptumDOD | 5,330 | 11,606 | 2.7 | 6.9 | <5 | 18 | 0.33 (0.08 - 1.03) | 0.10 | 0.34 (0.09 - 1.24) | 0.10 |
| MDCR | 1,492 | 3,506 | 0.9 | 2.6 | 0 | <5 | | | | |
| Open Claims | 87,275 | 184,585 | 53.0 | 128.5 | 32 | 137 | 0.53 (0.35 - 0.78) | <0.01 | 0.51 (0.34 - 0.77) | <0.01 |
| OptumEHR | 3,402 | 7,821 | 0.9 | 2.2 | <5 | <5 | 0.52 (0.03 - 3.69) | 0.60 | 0.50 (0.04 - 6.01) | 0.59 |
| Meta-analysis | 92,605 | 196,191 | 55.7 | 135.4 | <37 | 155 | 0.51 (0.35 - 0.75) | <0.01 | 0.49 (0.33 - 0.72) | <0.01 |
| **PS-matching / on-treatment TAR** | | | | | | | | | | |
| OptumDOD | 4,769 | 11,590 | 2.5 | 6.9 | <5 | 18 | 0.64 (0.14 - 2.23) | 0.53 | 0.71 (0.18 - 2.87) | 0.63 |
| MDCR | 1,391 | 3,498 | 0.9 | 2.5 | 0 | <5 | | | | |
| Open Claims | 79,633 | 184,531 | 49.3 | 128.4 | 28 | 136 | 0.50 (0.30 - 0.81) | <0.01 | 0.48 (0.29 - 0.78) | <0.01 |
| OptumEHR | 3,176 | 7,809 | 0.8 | 2.2 | <5 | <5 | 2.00 (0.07 - 56.4) | 0.68 | 1.85 (0.07 - 52.1) | 0.72 |
| Meta-analysis | 84,402 | 196,121 | 51.8 | 135.4 | <33 | 154 | 0.51 (0.32 - 0.82) | <0.01 | 0.50 (0.32 - 0.81) | <0.01 |
| **PS-stratification / intent-to-treat TAR** | | | | | | | | | | |
| OptumDOD | 5,330 | 11,606 | 6.9 | 17.7 | 6 | 44 | 0.33 (0.12 - 0.73) | 0.01 | 0.33 (0.13 - 0.80) | 0.01 |
| MDCR | 1,492 | 3,506 | 2.0 | 5.9 | 0 | 11 | | | | |
| Open Claims | 87,275 | 184,585 | 180.2 | 457.8 | 155 | 494 | 0.79 (0.65 - 0.96) | 0.02 | 0.77 (0.63 - 0.95) | 0.02 |
| OptumEHR | 3,402 | 7,821 | 6.1 | 16.1 | <5 | 11 | 0.38 (0.06 - 1.49) | 0.24 | 0.38 (0.07 - 1.95) | 0.25 |
| Meta-analysis | 92,605 | 196,191 | 187.1 | 475.5 | 161 | 538 | 0.57 (0.24 - 1.32) | 0.19 | 0.56 (0.24 - 1.30) | 0.17 |
| **PS-matching / intent-to-treat TAR** | | | | | | | | | | |
| OptumDOD | 4,769 | 11,590 | 6.3 | 17.7 | 5 | 44 | 0.27 (0.08 - 0.71) | 0.02 | 0.28 (0.09 - 0.83) | 0.02 |
| MDCR | 1,391 | 3,498 | 1.9 | 5.9 | 0 | 11 | | | | |
| Open Claims | 79,633 | 184,531 | 168.5 | 457.7 | 143 | 493 | 0.62 (0.48 - 0.79) | <0.01 | 0.60 (0.46 - 0.77) | <0.01 |
| OptumEHR | 3,176 | 7,809 | 5.7 | 16.1 | <5 | 11 | 0.48 (0.07 - 2.18) | 0.41 | 0.50 (0.09 - 2.80) | 0.43 |
| Meta-analysis | 84,402 | 196,121 | 174.8 | 475.4 | 148 | 537 | 0.49 (0.24 - 1.01) | 0.05 | 0.48 (0.24 - 1.00) | 0.05 |

##### 6.1.7 Diarrhea

**Supplementary Table 114:** Exposures, outcomes and risk estimates for Diarrhea in GLP1RA (T) vs SGLT2I (C) new-user cohorts under four different propensity score (PS) adjustment / time-at-risk (TAR) designs across data sources. We report total patient exposure time in 1,000 patient-year (KYr) units, uncalibrated and calibrated hazard ratios (HRs) and their 95% confidence intervals (CIs). Greyed out entries did not pass study diagnostics and are included only for completeness.

| | Patients | | Exposure (KYr) | | Outcomes | | Uncalibrated | | Calibrated | |
|---|---|---|---|---|---|---|---|---|---|---|
| | T | C | T | C | T | C | HR (95% CI) | $p$ | HR (95% CI) | $p$ |
| **PS-stratification / on-treatment TAR** | | | | | | | | | | |
| OptumDOD | 4,449 | 9,875 | 2.2 | 5.8 | 120 | 228 | 1.19 (0.94 - 1.51) | 0.14 | 1.21 (0.95 - 1.54) | 0.13 |
| MDCR | 1,281 | 3,032 | 0.8 | 2.2 | 21 | 57 | 0.96 (0.55 - 1.61) | 0.87 | 0.83 (0.49 - 1.43) | 0.52 |
| Open Claims | 72,445 | 158,033 | 43.8 | 109.5 | 964 | 1,724 | 1.31 (1.21 - 1.43) | <0.01 | 1.27 (1.16 - 1.39) | <0.01 |
| OptumEHR | 2,980 | 6,910 | 0.8 | 1.9 | 33 | 70 | 0.92 (0.59 - 1.42) | 0.72 | 0.90 (0.58 - 1.39) | 0.63 |
| Meta-analysis | 81,155 | 177,850 | 47.6 | 119.4 | 1,138 | 2,079 | 1.22 (1.05 - 1.40) | <0.01 | 1.17 (1.01 - 1.36) | 0.04 |
| **PS-matching / on-treatment TAR** | | | | | | | | | | |
| OptumDOD | 3,981 | 9,860 | 2.0 | 5.8 | 106 | 226 | 1.24 (0.90 - 1.70) | 0.18 | 1.39 (1.01 - 1.91) | 0.05 |
| MDCR | 1,195 | 3,025 | 0.7 | 2.2 | 20 | 57 | 0.91 (0.43 - 1.89) | 0.81 | 0.87 (0.41 - 1.83) | 0.72 |
| Open Claims | 66,618 | 157,992 | 40.9 | 109.4 | 861 | 1,722 | 1.37 (1.22 - 1.54) | <0.01 | 1.30 (1.16 - 1.46) | <0.01 |
| OptumEHR | 2,775 | 6,899 | 0.7 | 1.9 | 32 | 70 | 1.08 (0.64 - 1.81) | 0.76 | 1.00 (0.59 - 1.69) | 0.77 |
| Meta-analysis | 74,569 | 177,776 | 44.5 | 119.3 | 1,019 | 2,075 | 1.33 (1.20 - 1.48) | <0.01 | 1.30 (1.16 - 1.46) | <0.01 |
| **PS-stratification / intent-to-treat TAR** | | | | | | | | | | |
| OptumDOD | 4,449 | 9,875 | 5.4 | 14.4 | 276 | 576 | 1.13 (0.97 - 1.32) | 0.11 | 1.13 (0.97 - 1.33) | 0.18 |
| MDCR | 1,281 | 3,032 | 1.7 | 4.8 | 55 | 171 | 0.89 (0.64 - 1.22) | 0.48 | 0.82 (0.59 - 1.13) | 0.25 |
| Open Claims | 72,445 | 158,033 | 144.3 | 379.4 | 3,152 | 6,741 | 1.14 (1.09 - 1.19) | <0.01 | 1.11 (1.00 - 1.22) | 0.06 |
| OptumEHR | 2,980 | 6,910 | 5.1 | 13.6 | 141 | 341 | 0.95 (0.77 - 1.16) | 0.61 | 0.95 (0.77 - 1.18) | 0.68 |
| Meta-analysis | 81,155 | 177,850 | 156.4 | 412.2 | 3,624 | 7,829 | 1.09 (0.99 - 1.19) | 0.08 | 1.06 (0.94 - 1.20) | 0.33 |
| **PS-matching / intent-to-treat TAR** | | | | | | | | | | |
| OptumDOD | 3,981 | 9,860 | 5.0 | 14.3 | 248 | 572 | 1.33 (1.09 - 1.62) | <0.01 | 1.35 (1.10 - 1.65) | <0.01 |
| MDCR | 1,195 | 3,025 | 1.6 | 4.8 | 52 | 171 | 0.91 (0.60 - 1.35) | 0.63 | 0.78 (0.51 - 1.19) | 0.35 |
| Open Claims | 66,618 | 157,992 | 135.8 | 379.3 | 2,874 | 6,738 | 1.19 (1.13 - 1.26) | <0.01 | 1.15 (1.06 - 1.25) | <0.01 |
| OptumEHR | 2,775 | 6,899 | 4.8 | 13.6 | 134 | 341 | 0.92 (0.71 - 1.19) | 0.55 | 0.95 (0.74 - 1.23) | 0.73 |
| Meta-analysis | 74,569 | 177,776 | 147.1 | 412.1 | 3,308 | 7,822 | 1.13 (0.96 - 1.32) | 0.15 | 1.11 (0.94 - 1.31) | 0.22 |

##### 6.1.8 Genitourinary infection

**Supplementary Table 115:** Exposures, outcomes and risk estimates for Genitourinary infection in GLP1RA (T) vs SGLT2I (C) new-user cohorts under four different propensity score (PS) adjustment / time-at-risk (TAR) designs across data sources. We report total patient exposure time in 1,000 patient-year (KYr) units, uncalibrated and calibrated hazard ratios (HRs) and their 95% confidence intervals (CIs). Greyed out entries did not pass study diagnostics and are included only for completeness.

| | Patients | | Exposure (KYr) | | Outcomes | | Uncalibrated | | Calibrated | |
|---|---|---|---|---|---|---|---|---|---|---|
| | T | C | T | C | T | C | HR (95% CI) | $p$ | HR (95% CI) | $p$ |
| **PS-stratification / on-treatment TAR** | | | | | | | | | | |
| OptumDOD | 3,876 | 9,018 | 1.9 | 5.3 | 137 | 329 | 1.08 (0.87 - 1.34) | 0.46 | 1.10 (0.89 - 1.36) | 0.42 |
| MDCR | 1,118 | 2,894 | 0.6 | 2.1 | 42 | 96 | 1.21 (0.81 - 1.76) | 0.34 | 1.05 (0.71 - 1.55) | 0.79 |
| Open Claims | 60,348 | 137,023 | 36.9 | 96.5 | 1,313 | 2,911 | 1.07 (1.00 - 1.15) | 0.04 | 1.04 (0.96 - 1.12) | 0.40 |
| OptumEHR | 2,754 | 6,565 | 0.7 | 1.8 | 47 | 86 | 1.24 (0.84 - 1.81) | 0.27 | 1.21 (0.82 - 1.77) | 0.36 |
| Meta-analysis | 68,096 | 155,500 | 40.2 | 105.7 | 1,539 | 3,422 | 1.08 (1.02 - 1.15) | 0.01 | 1.05 (0.97 - 1.13) | 0.28 |
| **PS-matching / on-treatment TAR** | | | | | | | | | | |
| OptumDOD | 3,491 | 9,007 | 1.7 | 5.3 | 128 | 329 | 0.83 (0.62 - 1.11) | 0.21 | 0.93 (0.70 - 1.24) | 0.64 |
| MDCR | 1,054 | 2,886 | 0.6 | 2.1 | 40 | 95 | 1.15 (0.65 - 1.98) | 0.63 | 1.09 (0.62 - 1.90) | 0.78 |
| Open Claims | 56,132 | 136,980 | 34.9 | 96.5 | 1,172 | 2,909 | 1.01 (0.92 - 1.11) | 0.87 | 0.96 (0.87 - 1.06) | 0.48 |
| OptumEHR | 2,609 | 6,555 | 0.7 | 1.8 | 44 | 86 | 1.17 (0.68 - 1.96) | 0.57 | 1.08 (0.63 - 1.83) | 0.74 |
| Meta-analysis | 63,286 | 155,428 | 37.9 | 105.6 | 1,384 | 3,419 | 1.00 (0.91 - 1.09) | 0.95 | 0.98 (0.89 - 1.08) | 0.66 |
| **PS-stratification / intent-to-treat TAR** | | | | | | | | | | |
| OptumDOD | 3,876 | 9,018 | 4.4 | 12.6 | 329 | 825 | 1.06 (0.92 - 1.21) | 0.43 | 1.05 (0.91 - 1.22) | 0.55 |
| MDCR | 1,118 | 2,894 | 1.3 | 4.5 | 91 | 236 | 1.13 (0.87 - 1.45) | 0.35 | 1.04 (0.80 - 1.34) | 0.78 |
| Open Claims | 60,348 | 137,023 | 117.6 | 320.7 | 4,580 | 11,149 | 1.05 (1.01 - 1.09) | <0.01 | 1.02 (0.93 - 1.12) | 0.68 |
| OptumEHR | 2,754 | 6,565 | 4.6 | 12.6 | 178 | 421 | 1.00 (0.83 - 1.20) | 0.98 | 1.01 (0.84 - 1.22) | 0.80 |
| Meta-analysis | 68,096 | 155,500 | 127.9 | 350.4 | 5,178 | 12,631 | 1.05 (1.02 - 1.09) | <0.01 | 1.03 (0.94 - 1.12) | 0.53 |
| **PS-matching / intent-to-treat TAR** | | | | | | | | | | |
| OptumDOD | 3,491 | 9,007 | 4.1 | 12.6 | 303 | 823 | 0.95 (0.80 - 1.12) | 0.55 | 0.96 (0.81 - 1.15) | 0.71 |
| MDCR | 1,054 | 2,886 | 1.3 | 4.5 | 85 | 235 | 1.10 (0.79 - 1.53) | 0.55 | 0.96 (0.68 - 1.35) | 0.81 |
| Open Claims | 56,132 | 136,980 | 111.6 | 320.7 | 4,233 | 11,146 | 1.05 (1.00 - 1.09) | 0.04 | 1.01 (0.94 - 1.09) | 0.81 |
| OptumEHR | 2,609 | 6,555 | 4.4 | 12.5 | 163 | 421 | 0.93 (0.73 - 1.16) | 0.51 | 0.96 (0.76 - 1.21) | 0.71 |
| Meta-analysis | 63,286 | 155,428 | 121.3 | 350.3 | 4,784 | 12,625 | 1.03 (0.97 - 1.09) | 0.30 | 1.02 (0.94 - 1.09) | 0.69 |

##### 6.1.9 Hyperkalemia

**Supplementary Table 116:** Exposures, outcomes and risk estimates for Hyperkalemia in GLP1RA (T) vs SGLT2I (C) new-user cohorts under four different propensity score (PS) adjustment / time-at-risk (TAR) designs across data sources. We report total patient exposure time in 1,000 patient-year (KYr) units, uncalibrated and calibrated hazard ratios (HRs) and their 95% confidence intervals (CIs). Greyed out entries did not pass study diagnostics and are included only for completeness.

| | Patients | | Exposure (KYr) | | Outcomes | | Uncalibrated | | Calibrated | |
|---|---|---|---|---|---|---|---|---|---|---|
| | T | C | T | C | T | C | HR (95% CI) | $p$ | HR (95% CI) | $p$ |
| **PS-stratification / on-treatment TAR** | | | | | | | | | | |
| OptumDOD | 5,051 | 10,938 | 2.5 | 6.5 | 62 | 162 | 1.06 (0.78 - 1.44) | 0.69 | 1.08 (0.79 - 1.48) | 0.64 |
| MDCR | 1,457 | 3,408 | 0.9 | 2.5 | 10 | 25 | 1.01 (0.43 - 2.17) | 0.98 | 0.88 (0.39 - 1.97) | 0.76 |
| Open Claims | 84,511 | 178,385 | 51.2 | 124.1 | 293 | 871 | 0.93 (0.81 - 1.07) | 0.32 | 0.90 (0.78 - 1.04) | 0.17 |
| OptumEHR | 3,303 | 7,603 | 0.9 | 2.1 | 10 | 31 | 0.67 (0.30 - 1.36) | 0.29 | 0.65 (0.31 - 1.38) | 0.27 |
| Meta-analysis | 94,322 | 200,334 | 55.5 | 135.2 | 375 | 1,089 | 0.95 (0.83 - 1.07) | 0.38 | 0.91 (0.80 - 1.04) | 0.18 |
| **PS-matching / on-treatment TAR** | | | | | | | | | | |
| OptumDOD | 4,513 | 10,924 | 2.3 | 6.5 | 58 | 161 | 0.91 (0.59 - 1.38) | 0.66 | 1.02 (0.66 - 1.56) | 0.88 |
| MDCR | 1,357 | 3,400 | 0.8 | 2.5 | 9 | 24 | 1.20 (0.44 - 2.96) | 0.71 | 1.13 (0.44 - 2.93) | 0.81 |
| Open Claims | 77,107 | 178,338 | 47.6 | 124.1 | 274 | 870 | 0.96 (0.79 - 1.16) | 0.67 | 0.91 (0.75 - 1.10) | 0.37 |
| OptumEHR | 3,089 | 7,592 | 0.8 | 2.1 | 8 | 31 | 0.33 (0.08 - 1.02) | 0.10 | 0.31 (0.08 - 1.12) | 0.08 |
| Meta-analysis | 86,066 | 200,254 | 51.6 | 135.2 | 349 | 1,086 | 0.94 (0.79 - 1.11) | 0.48 | 0.92 (0.77 - 1.10) | 0.37 |
| **PS-stratification / intent-to-treat TAR** | | | | | | | | | | |
| OptumDOD | 5,051 | 10,938 | 6.3 | 16.5 | 179 | 400 | 1.25 (1.04 - 1.51) | 0.02 | 1.25 (1.03 - 1.52) | 0.04 |
| MDCR | 1,457 | 3,408 | 1.9 | 5.7 | 26 | 71 | 1.06 (0.65 - 1.68) | 0.81 | 0.97 (0.60 - 1.57) | 0.89 |
| Open Claims | 84,511 | 178,385 | 172.7 | 440.7 | 1,310 | 3,397 | 1.12 (1.04 - 1.19) | <0.01 | 1.08 (0.97 - 1.21) | 0.17 |
| OptumEHR | 3,303 | 7,603 | 5.8 | 15.4 | 54 | 176 | 0.85 (0.62 - 1.17) | 0.34 | 0.86 (0.62 - 1.19) | 0.37 |
| Meta-analysis | 94,322 | 200,334 | 186.8 | 478.3 | 1,569 | 4,044 | 1.12 (1.05 - 1.19) | <0.01 | 1.09 (0.99 - 1.21) | 0.08 |
| **PS-matching / intent-to-treat TAR** | | | | | | | | | | |
| OptumDOD | 4,513 | 10,924 | 5.8 | 16.4 | 168 | 398 | 1.20 (0.95 - 1.51) | 0.13 | 1.22 (0.96 - 1.54) | 0.12 |
| MDCR | 1,357 | 3,400 | 1.8 | 5.7 | 25 | 70 | 1.32 (0.70 - 2.41) | 0.38 | 1.14 (0.61 - 2.14) | 0.71 |
| Open Claims | 77,107 | 178,338 | 161.5 | 440.6 | 1,218 | 3,396 | 1.12 (1.03 - 1.22) | <0.01 | 1.08 (0.98 - 1.20) | 0.17 |
| OptumEHR | 3,089 | 7,592 | 5.5 | 15.4 | 50 | 176 | 0.76 (0.51 - 1.11) | 0.17 | 0.78 (0.53 - 1.16) | 0.24 |
| Meta-analysis | 86,066 | 200,254 | 174.6 | 478.2 | 1,461 | 4,040 | 1.12 (1.04 - 1.20) | <0.01 | 1.10 (1.01 - 1.20) | 0.04 |

##### 6.1.10 Hypoglycemia

**Supplementary Table 117:** Exposures, outcomes and risk estimates for Hypoglycemia in GLP1RA (T) vs SGLT2I (C) new-user cohorts under four different propensity score (PS) adjustment / time-at-risk (TAR) designs across data sources. We report total patient exposure time in 1,000 patient-year (KYr) units, uncalibrated and calibrated hazard ratios (HRs) and their 95% confidence intervals (CIs). Greyed out entries did not pass study diagnostics and are included only for completeness.

| | Patients | | Exposure (KYr) | | Outcomes | | Uncalibrated | | Calibrated | |
|---|---|---|---|---|---|---|---|---|---|---|
| | T | C | T | C | T | C | HR (95% CI) | $p$ | HR (95% CI) | $p$ |
| **PS-stratification / on-treatment TAR** | | | | | | | | | | |
| OptumDOD | 5,239 | 11,439 | 2.7 | 6.8 | 17 | 48 | 0.72 (0.39 - 1.27) | 0.27 | 0.73 (0.41 - 1.32) | 0.30 |
| MDCR | 1,473 | 3,474 | 0.9 | 2.5 | <5 | <5 | 1.60 (0.22 - 8.54) | 0.61 | 1.40 (0.22 - 8.79) | 0.72 |
| Open Claims | 85,575 | 181,772 | 52.0 | 126.5 | 116 | 247 | 1.12 (0.88 - 1.41) | 0.34 | 1.08 (0.85 - 1.37) | 0.52 |
| OptumEHR | 3,355 | 7,748 | 0.9 | 2.2 | <5 | <5 | 1.01 (0.14 - 5.29) | 1.00 | 0.98 (0.16 - 6.08) | 0.95 |
| Meta-analysis | 90,814 | 193,211 | 54.7 | 133.4 | 133 | 295 | 0.98 (0.66 - 1.46) | 0.92 | 0.94 (0.63 - 1.41) | 0.78 |
| **PS-matching / on-treatment TAR** | | | | | | | | | | |
| OptumDOD | 4,701 | 11,424 | 2.4 | 6.8 | 15 | 48 | 0.51 (0.20 - 1.16) | 0.14 | 0.57 (0.23 - 1.38) | 0.22 |
| MDCR | 1,373 | 3,466 | 0.9 | 2.5 | <5 | <5 | 2.07 (0.24 - 17.9) | 0.51 | 1.96 (0.23 - 17.0) | 0.55 |
| Open Claims | 78,221 | 181,718 | 48.4 | 126.5 | 103 | 247 | 1.51 (1.09 - 2.09) | 0.01 | 1.44 (1.04 - 1.99) | 0.03 |
| OptumEHR | 3,140 | 7,736 | 0.8 | 2.2 | <5 | <5 | 1.81 (0.08 - 19.0) | 0.67 | 1.67 (0.11 - 25.2) | 0.71 |
| Meta-analysis | 82,922 | 193,142 | 50.9 | 133.3 | 118 | 295 | 0.95 (0.33 - 2.73) | 0.93 | 0.93 (0.33 - 2.68) | 0.90 |
| **PS-stratification / intent-to-treat TAR** | | | | | | | | | | |
| OptumDOD | 5,239 | 11,439 | 6.7 | 17.3 | 56 | 171 | 0.81 (0.58 - 1.11) | 0.19 | 0.81 (0.58 - 1.12) | 0.21 |
| MDCR | 1,473 | 3,474 | 2.0 | 5.8 | 8 | 26 | 0.83 (0.34 - 1.82) | 0.67 | 0.77 (0.33 - 1.76) | 0.54 |
| Open Claims | 85,575 | 181,772 | 175.7 | 448.6 | 660 | 1,553 | 1.12 (1.02 - 1.23) | 0.02 | 1.09 (0.95 - 1.24) | 0.23 |
| OptumEHR | 3,355 | 7,748 | 5.9 | 15.9 | 29 | 82 | 0.90 (0.57 - 1.38) | 0.64 | 0.91 (0.58 - 1.41) | 0.67 |
| Meta-analysis | 95,642 | 204,433 | 190.4 | 487.6 | 753 | 1,832 | 0.98 (0.79 - 1.21) | 0.83 | 0.96 (0.76 - 1.20) | 0.70 |
| **PS-matching / intent-to-treat TAR** | | | | | | | | | | |
| OptumDOD | 4,701 | 11,424 | 6.2 | 17.3 | 52 | 170 | 0.69 (0.45 - 1.04) | 0.08 | 0.70 (0.46 - 1.07) | 0.11 |
| MDCR | 1,373 | 3,466 | 1.9 | 5.8 | 8 | 26 | 0.97 (0.36 - 2.44) | 0.95 | 0.84 (0.32 - 2.21) | 0.73 |
| Open Claims | 78,221 | 181,718 | 164.5 | 448.6 | 609 | 1,553 | 1.06 (0.94 - 1.21) | 0.35 | 1.03 (0.89 - 1.18) | 0.74 |
| OptumEHR | 3,140 | 7,736 | 5.6 | 15.8 | 26 | 82 | 0.58 (0.30 - 1.06) | 0.09 | 0.60 (0.32 - 1.13) | 0.12 |
| Meta-analysis | 87,435 | 204,344 | 178.2 | 487.4 | 695 | 1,831 | 0.84 (0.61 - 1.17) | 0.31 | 0.83 (0.60 - 1.16) | 0.27 |

##### 6.1.11 Hypotension

**Supplementary Table 118:** Exposures, outcomes and risk estimates for Hypotension in GLP1RA (T) vs SGLT2I (C) new-user cohorts under four different propensity score (PS) adjustment / time-at-risk (TAR) designs across data sources. We report total patient exposure time in 1,000 patient-year (KYr) units, uncalibrated and calibrated hazard ratios (HRs) and their 95% confidence intervals (CIs). Greyed out entries did not pass study diagnostics and are included only for completeness.

| | Patients | | Exposure (KYr) | | Outcomes | | Uncalibrated | | Calibrated | |
|---|---|---|---|---|---|---|---|---|---|---|
| | T | C | T | C | T | C | HR (95% CI) | $p$ | HR (95% CI) | $p$ |
| **PS-stratification / on-treatment TAR** | | | | | | | | | | |
| OptumDOD | 4,991 | 10,826 | 2.5 | 6.4 | 71 | 223 | 0.81 (0.61 - 1.08) | 0.16 | 0.82 (0.62 - 1.10) | 0.20 |
| MDCR | 1,417 | 3,342 | 0.9 | 2.4 | 17 | 60 | 0.80 (0.45 - 1.36) | 0.44 | 0.70 (0.40 - 1.22) | 0.22 |
| Open Claims | 82,501 | 174,134 | 50.0 | 121.2 | 538 | 1,455 | 1.01 (0.91 - 1.12) | 0.86 | 0.98 (0.87 - 1.09) | 0.68 |
| OptumEHR | 3,277 | 7,517 | 0.9 | 2.1 | 17 | 46 | 0.94 (0.51 - 1.65) | 0.83 | 0.91 (0.51 - 1.64) | 0.76 |
| Meta-analysis | 92,186 | 195,819 | 54.3 | 132.1 | 643 | 1,784 | 0.94 (0.81 - 1.10) | 0.46 | 0.91 (0.78 - 1.07) | 0.25 |
| **PS-matching / on-treatment TAR** | | | | | | | | | | |
| OptumDOD | 4,483 | 10,804 | 2.3 | 6.4 | 65 | 223 | 0.70 (0.47 - 1.02) | 0.07 | 0.79 (0.53 - 1.16) | 0.23 |
| MDCR | 1,319 | 3,334 | 0.8 | 2.4 | 17 | 60 | 0.95 (0.43 - 1.96) | 0.90 | 0.90 (0.42 - 1.92) | 0.79 |
| Open Claims | 75,427 | 174,062 | 46.6 | 121.1 | 486 | 1,447 | 0.95 (0.82 - 1.09) | 0.47 | 0.90 (0.78 - 1.04) | 0.20 |
| OptumEHR | 3,053 | 7,506 | 0.8 | 2.1 | 16 | 46 | 1.54 (0.76 - 3.02) | 0.22 | 1.42 (0.71 - 2.84) | 0.34 |
| Meta-analysis | 84,282 | 195,706 | 50.5 | 132.1 | 584 | 1,776 | 0.93 (0.77 - 1.10) | 0.39 | 0.91 (0.76 - 1.09) | 0.30 |
| **PS-stratification / intent-to-treat TAR** | | | | | | | | | | |
| OptumDOD | 4,991 | 10,826 | 6.3 | 16.2 | 184 | 518 | 0.89 (0.75 - 1.07) | 0.22 | 0.89 (0.74 - 1.07) | 0.28 |
| MDCR | 1,417 | 3,342 | 1.9 | 5.4 | 44 | 141 | 0.88 (0.62 - 1.25) | 0.50 | 0.81 (0.57 - 1.16) | 0.27 |
| Open Claims | 82,501 | 174,134 | 167.6 | 428.5 | 2,102 | 5,437 | 1.06 (1.00 - 1.12) | 0.04 | 1.03 (0.93 - 1.14) | 0.63 |
| OptumEHR | 3,277 | 7,517 | 5.7 | 15.1 | 82 | 230 | 0.93 (0.71 - 1.21) | 0.59 | 0.94 (0.72 - 1.22) | 0.64 |
| Meta-analysis | 92,186 | 195,819 | 181.4 | 465.3 | 2,412 | 6,326 | 0.98 (0.87 - 1.10) | 0.74 | 0.96 (0.83 - 1.11) | 0.57 |
| **PS-matching / intent-to-treat TAR** | | | | | | | | | | |
| OptumDOD | 4,483 | 10,804 | 5.8 | 16.1 | 171 | 516 | 0.74 (0.58 - 0.92) | <0.01 | 0.75 (0.59 - 0.94) | 0.02 |
| MDCR | 1,319 | 3,334 | 1.8 | 5.4 | 43 | 141 | 1.03 (0.66 - 1.56) | 0.91 | 0.89 (0.57 - 1.38) | 0.66 |
| Open Claims | 75,427 | 174,062 | 156.9 | 428.4 | 1,924 | 5,428 | 1.06 (1.00 - 1.14) | 0.06 | 1.03 (0.94 - 1.12) | 0.61 |
| OptumEHR | 3,053 | 7,506 | 5.4 | 15.1 | 77 | 229 | 0.93 (0.67 - 1.26) | 0.63 | 0.96 (0.70 - 1.31) | 0.78 |
| Meta-analysis | 84,282 | 195,706 | 169.9 | 465.1 | 2,215 | 6,314 | 0.93 (0.77 - 1.13) | 0.49 | 0.92 (0.76 - 1.12) | 0.42 |

##### 6.1.12 Joint pain

**Supplementary Table 119:** Exposures, outcomes and risk estimates for Joint pain in GLP1RA (T) vs SGLT2I (C) new-user cohorts under four different propensity score (PS) adjustment / time-at-risk (TAR) designs across data sources. We report total patient exposure time in 1,000 patient-year (KYr) units, uncalibrated and calibrated hazard ratios (HRs) and their 95% confidence intervals (CIs). Greyed out entries did not pass study diagnostics and are included only for completeness.

| | Patients | | Exposure (KYr) | | Outcomes | | Uncalibrated | | Calibrated | |
|---|---|---|---|---|---|---|---|---|---|---|
| | T | C | T | C | T | C | HR (95% CI) | $p$ | HR (95% CI) | $p$ |
| **PS-stratification / on-treatment TAR** | | | | | | | | | | |
| OptumDOD | 4,874 | 10,899 | 2.5 | 6.5 | 46 | 82 | 1.19 (0.80 - 1.74) | 0.39 | 1.20 (0.82 - 1.77) | 0.36 |
| MDCR | 1,390 | 3,368 | 0.9 | 2.5 | 8 | 25 | 0.61 (0.25 - 1.35) | 0.25 | 0.53 (0.23 - 1.24) | 0.15 |
| Open Claims | 79,450 | 172,024 | 48.5 | 120.0 | 314 | 623 | 1.06 (0.92 - 1.22) | 0.44 | 1.02 (0.88 - 1.18) | 0.78 |
| OptumEHR | 3,155 | 7,405 | 0.8 | 2.0 | 10 | 25 | 0.81 (0.35 - 1.71) | 0.59 | 0.78 (0.36 - 1.72) | 0.55 |
| Meta-analysis | 88,869 | 193,696 | 52.7 | 131.0 | 378 | 755 | 1.05 (0.92 - 1.20) | 0.47 | 1.01 (0.88 - 1.16) | 0.85 |
| **PS-matching / on-treatment TAR** | | | | | | | | | | |
| OptumDOD | 4,400 | 10,883 | 2.3 | 6.5 | 43 | 82 | 0.92 (0.52 - 1.58) | 0.76 | 1.03 (0.59 - 1.78) | 0.90 |
| MDCR | 1,312 | 3,360 | 0.8 | 2.4 | 7 | 25 | 0.33 (0.05 - 1.41) | 0.20 | 0.32 (0.06 - 1.69) | 0.18 |
| Open Claims | 73,133 | 171,974 | 45.4 | 120.0 | 277 | 623 | 0.92 (0.75 - 1.13) | 0.43 | 0.87 (0.71 - 1.08) | 0.23 |
| OptumEHR | 2,958 | 7,395 | 0.8 | 2.0 | 9 | 25 | 0.51 (0.11 - 1.82) | 0.35 | 0.47 (0.12 - 1.92) | 0.30 |
| Meta-analysis | 81,803 | 193,612 | 49.3 | 131.0 | 336 | 755 | 0.90 (0.74 - 1.09) | 0.27 | 0.88 (0.72 - 1.07) | 0.21 |
| **PS-stratification / intent-to-treat TAR** | | | | | | | | | | |
| OptumDOD | 4,874 | 10,899 | 6.2 | 16.4 | 121 | 224 | 1.20 (0.95 - 1.52) | 0.13 | 1.20 (0.94 - 1.52) | 0.17 |
| MDCR | 1,390 | 3,368 | 1.9 | 5.6 | 21 | 57 | 0.86 (0.50 - 1.45) | 0.59 | 0.79 (0.47 - 1.35) | 0.40 |
| Open Claims | 79,450 | 172,024 | 163.4 | 423.4 | 1,168 | 2,527 | 1.06 (0.98 - 1.14) | 0.13 | 1.03 (0.92 - 1.15) | 0.66 |
| OptumEHR | 3,155 | 7,405 | 5.6 | 15.0 | 46 | 147 | 0.72 (0.51 - 1.01) | 0.07 | 0.73 (0.51 - 1.03) | 0.08 |
| Meta-analysis | 88,869 | 193,696 | 177.0 | 460.4 | 1,356 | 2,955 | 1.00 (0.82 - 1.22) | 0.97 | 0.97 (0.78 - 1.21) | 0.82 |
| **PS-matching / intent-to-treat TAR** | | | | | | | | | | |
| OptumDOD | 4,400 | 10,883 | 5.8 | 16.4 | 108 | 224 | 0.94 (0.68 - 1.29) | 0.71 | 0.96 (0.69 - 1.32) | 0.79 |
| MDCR | 1,312 | 3,360 | 1.8 | 5.6 | 20 | 57 | 1.03 (0.49 - 2.12) | 0.94 | 0.89 (0.42 - 1.88) | 0.78 |
| Open Claims | 73,133 | 171,974 | 153.8 | 423.3 | 1,047 | 2,527 | 1.04 (0.95 - 1.14) | 0.42 | 1.00 (0.90 - 1.12) | 0.90 |
| OptumEHR | 2,958 | 7,395 | 5.3 | 14.9 | 42 | 147 | 0.68 (0.44 - 1.03) | 0.08 | 0.70 (0.46 - 1.08) | 0.12 |
| Meta-analysis | 81,803 | 193,612 | 166.6 | 460.3 | 1,217 | 2,955 | 0.96 (0.81 - 1.14) | 0.66 | 0.95 (0.79 - 1.13) | 0.56 |

##### 6.1.13 Lower extremity amputation

**Supplementary Table 120:** Exposures, outcomes and risk estimates for Lower extremity amputation in GLP1RA (T) vs SGLT2I (C) new-user cohorts under four different propensity score (PS) adjustment / time-at-risk (TAR) designs across data sources. We report total patient exposure time in 1,000 patient-year (KYr) units, uncalibrated and calibrated hazard ratios (HRs) and their 95% confidence intervals (CIs). Greyed out entries did not pass study diagnostics and are included only for completeness.

| | Patients | | Exposure (KYr) | | Outcomes | | Uncalibrated | | Calibrated | |
|---|---|---|---|---|---|---|---|---|---|---|
| | T | C | T | C | T | C | HR (95% CI) | $p$ | HR (95% CI) | $p$ |
| **PS-stratification / on-treatment TAR** | | | | | | | | | | |
| OptumDOD | 5,323 | 11,589 | 2.7 | 6.9 | <5 | 6 | 0.65 (0.09 - 3.23) | 0.64 | 0.66 (0.11 - 4.00) | 0.65 |
| MDCR | 1,492 | 3,500 | 0.9 | 2.6 | 0 | <5 | | | | |
| Open Claims | 87,174 | 184,471 | 53.0 | 128.3 | 16 | 67 | 0.63 (0.35 - 1.08) | 0.11 | 0.61 (0.34 - 1.08) | 0.09 |
| OptumEHR | 3,400 | 7,821 | 0.9 | 2.2 | 0 | <5 | | | | |
| Meta-analysis | 87,174 | 184,471 | 53.0 | 128.3 | 16 | 67 | 0.63 (0.35 - 1.08) | 0.11 | 0.61 (0.34 - 1.08) | 0.09 |
| **PS-matching / on-treatment TAR** | | | | | | | | | | |
| OptumDOD | 4,758 | 11,574 | 2.5 | 6.9 | <5 | 6 | | | | |
| MDCR | 1,392 | 3,492 | 0.9 | 2.5 | 0 | <5 | | | | |
| Open Claims | 79,536 | 184,417 | 49.3 | 128.3 | 16 | 67 | 0.58 (0.26 - 1.15) | 0.14 | 0.55 (0.26 - 1.14) | 0.11 |
| OptumEHR | 3,174 | 7,809 | 0.8 | 2.2 | 0 | <5 | | | | |
| Meta-analysis | 79,536 | 184,417 | 49.3 | 128.3 | 16 | 67 | 0.58 (0.26 - 1.15) | 0.14 | 0.56 (0.27 - 1.18) | 0.13 |
| **PS-stratification / intent-to-treat TAR** | | | | | | | | | | |
| OptumDOD | 5,323 | 11,589 | 6.9 | 17.8 | 10 | 21 | 1.40 (0.61 - 3.03) | 0.41 | 1.40 (0.63 - 3.12) | 0.42 |
| MDCR | 1,492 | 3,500 | 2.0 | 5.9 | <5 | 7 | 0.99 (0.15 - 4.25) | 0.99 | 0.91 (0.17 - 4.94) | 0.91 |
| Open Claims | 87,174 | 184,471 | 180.1 | 457.9 | 96 | 255 | 1.02 (0.80 - 1.31) | 0.85 | 0.99 (0.76 - 1.29) | 0.95 |
| OptumEHR | 3,400 | 7,821 | 6.1 | 16.2 | <5 | <5 | 2.17 (0.47 - 10.3) | 0.33 | 2.18 (0.46 - 10.2) | 0.32 |
| Meta-analysis | 92,497 | 196,060 | 187.0 | 475.7 | 106 | 276 | 1.05 (0.83 - 1.33) | 0.67 | 1.03 (0.80 - 1.32) | 0.82 |
| **PS-matching / intent-to-treat TAR** | | | | | | | | | | |
| OptumDOD | 4,758 | 11,574 | 6.3 | 17.7 | 8 | 20 | 0.90 (0.28 - 2.50) | 0.85 | 0.91 (0.30 - 2.74) | 0.87 |
| MDCR | 1,392 | 3,492 | 1.9 | 5.9 | <5 | 7 | 0.00 (0.00 - 1.78) | 0.21 | 0.00 (0.00 - 36470) | 0.20 |
| Open Claims | 79,536 | 184,417 | 168.4 | 457.9 | 90 | 255 | 1.12 (0.82 - 1.50) | 0.47 | 1.08 (0.79 - 1.47) | 0.63 |
| OptumEHR | 3,174 | 7,809 | 5.7 | 16.1 | <5 | <5 | | | | |
| Meta-analysis | 84,294 | 195,991 | 174.7 | 475.6 | 98 | 275 | 1.10 (0.82 - 1.47) | 0.52 | 1.08 (0.81 - 1.46) | 0.59 |

##### 6.1.14 Nausea

**Supplementary Table 121:** Exposures, outcomes and risk estimates for Nausea in GLP1RA (T) vs SGLT2I (C) new-user cohorts under four different propensity score (PS) adjustment / time-at-risk (TAR) designs across data sources. We report total patient exposure time in 1,000 patient-year (KYr) units, uncalibrated and calibrated hazard ratios (HRs) and their 95% confidence intervals (CIs). Greyed out entries did not pass study diagnostics and are included only for completeness.

| | Patients | | Exposure (KYr) | | Outcomes | | Uncalibrated | | Calibrated | |
|---|---|---|---|---|---|---|---|---|---|---|
| | T | C | T | C | T | C | HR (95% CI) | $p$ | HR (95% CI) | $p$ |
| **PS-stratification / on-treatment TAR** | | | | | | | | | | |
| OptumDOD | 4,438 | 9,971 | 2.2 | 5.9 | 173 | 215 | 1.77 (1.43 - 2.20) | <0.01 | 1.80 (1.44 - 2.24) | <0.01 |
| MDCR | 1,332 | 3,120 | 0.8 | 2.3 | 33 | 68 | 1.08 (0.68 - 1.68) | 0.73 | 0.94 (0.60 - 1.48) | 0.80 |
| Open Claims | 72,522 | 158,462 | 44.0 | 111.0 | 1,271 | 1,750 | 1.63 (1.51 - 1.76) | <0.01 | 1.58 (1.45 - 1.71) | <0.01 |
| OptumEHR | 3,028 | 7,090 | 0.8 | 2.0 | 57 | 62 | 1.84 (1.25 - 2.70) | <0.01 | 1.79 (1.22 - 2.63) | <0.01 |
| Meta-analysis | 81,320 | 178,643 | 47.8 | 121.1 | 1,534 | 2,095 | 1.64 (1.52 - 1.76) | <0.01 | 1.58 (1.45 - 1.72) | <0.01 |
| **PS-matching / on-treatment TAR** | | | | | | | | | | |
| OptumDOD | 3,990 | 9,957 | 2.0 | 5.9 | 150 | 215 | 2.02 (1.51 - 2.70) | <0.01 | 2.26 (1.69 - 3.03) | <0.01 |
| MDCR | 1,244 | 3,113 | 0.8 | 2.3 | 31 | 68 | 0.90 (0.47 - 1.66) | 0.74 | 0.85 (0.45 - 1.60) | 0.64 |
| Open Claims | 66,731 | 158,415 | 41.2 | 110.9 | 1,138 | 1,749 | 1.83 (1.65 - 2.03) | <0.01 | 1.74 (1.56 - 1.93) | <0.01 |
| OptumEHR | 2,835 | 7,079 | 0.7 | 2.0 | 53 | 62 | 1.98 (1.21 - 3.23) | <0.01 | 1.83 (1.12 - 2.99) | 0.02 |
| Meta-analysis | 74,800 | 178,564 | 44.7 | 121.1 | 1,372 | 2,094 | 1.82 (1.66 - 2.01) | <0.01 | 1.79 (1.61 - 1.98) | <0.01 |
| **PS-stratification / intent-to-treat TAR** | | | | | | | | | | |
| OptumDOD | 4,438 | 9,971 | 5.3 | 14.7 | 351 | 588 | 1.44 (1.24 - 1.65) | <0.01 | 1.43 (1.24 - 1.66) | <0.01 |
| MDCR | 1,332 | 3,120 | 1.7 | 5.1 | 82 | 171 | 1.26 (0.95 - 1.66) | 0.11 | 1.16 (0.87 - 1.53) | 0.35 |
| Open Claims | 72,522 | 158,462 | 144.9 | 384.3 | 3,678 | 7,063 | 1.24 (1.19 - 1.30) | <0.01 | 1.21 (1.10 - 1.33) | <0.01 |
| OptumEHR | 3,028 | 7,090 | 5.2 | 14.1 | 157 | 301 | 1.13 (0.92 - 1.39) | 0.23 | 1.14 (0.93 - 1.40) | 0.24 |
| Meta-analysis | 81,320 | 178,643 | 157.1 | 418.1 | 4,268 | 8,123 | 1.27 (1.17 - 1.38) | <0.01 | 1.24 (1.10 - 1.40) | <0.01 |
| **PS-matching / intent-to-treat TAR** | | | | | | | | | | |
| OptumDOD | 3,990 | 9,957 | 4.9 | 14.7 | 313 | 585 | 1.55 (1.30 - 1.86) | <0.01 | 1.58 (1.31 - 1.90) | <0.01 |
| MDCR | 1,244 | 3,113 | 1.6 | 5.0 | 77 | 171 | 1.03 (0.72 - 1.47) | 0.86 | 0.89 (0.61 - 1.30) | 0.64 |
| Open Claims | 66,731 | 158,415 | 136.4 | 384.3 | 3,352 | 7,062 | 1.34 (1.27 - 1.42) | <0.01 | 1.30 (1.20 - 1.40) | <0.01 |
| OptumEHR | 2,835 | 7,079 | 4.9 | 14.1 | 138 | 301 | 1.18 (0.92 - 1.51) | 0.18 | 1.22 (0.95 - 1.57) | 0.15 |
| Meta-analysis | 74,800 | 178,564 | 147.9 | 418.0 | 3,880 | 8,119 | 1.33 (1.19 - 1.49) | <0.01 | 1.31 (1.16 - 1.49) | <0.01 |

##### 6.1.15 Peripheral edema

**Supplementary Table 122:** Exposures, outcomes and risk estimates for Peripheral edema in GLP1RA (T) vs SGLT2I (C) new-user cohorts under four different propensity score (PS) adjustment / time-at-risk (TAR) designs across data sources. We report total patient exposure time in 1,000 patient-year (KYr) units, uncalibrated and calibrated hazard ratios (HRs) and their 95% confidence intervals (CIs). Greyed out entries did not pass study diagnostics and are included only for completeness.

| | Patients | | Exposure (KYr) | | Outcomes | | Uncalibrated | | Calibrated | |
|---|---|---|---|---|---|---|---|---|---|---|
| | T | C | T | C | T | C | HR (95% CI) | $p$ | HR (95% CI) | $p$ |
| **PS-stratification / on-treatment TAR** | | | | | | | | | | |
| OptumDOD | 3,713 | 8,491 | 1.8 | 5.0 | 125 | 343 | 0.91 (0.73 - 1.13) | 0.38 | 0.92 (0.74 - 1.15) | 0.48 |
| MDCR | 1,085 | 2,647 | 0.6 | 1.9 | 37 | 82 | 1.24 (0.81 - 1.88) | 0.31 | 1.09 (0.71 - 1.66) | 0.72 |
| Open Claims | 58,688 | 132,741 | 35.7 | 93.2 | 1,042 | 2,437 | 1.11 (1.02 - 1.19) | 0.01 | 1.07 (0.98 - 1.16) | 0.16 |
| OptumEHR | 2,573 | 6,272 | 0.7 | 1.7 | 33 | 47 | 1.48 (0.90 - 2.41) | 0.12 | 1.44 (0.88 - 2.36) | 0.16 |
| Meta-analysis | 66,059 | 150,151 | 38.8 | 101.8 | 1,237 | 2,909 | 1.08 (0.95 - 1.24) | 0.24 | 1.05 (0.91 - 1.21) | 0.53 |
| **PS-matching / on-treatment TAR** | | | | | | | | | | |
| OptumDOD | 3,349 | 8,482 | 1.7 | 5.0 | 114 | 343 | 0.93 (0.70 - 1.23) | 0.60 | 1.04 (0.78 - 1.38) | 0.78 |
| MDCR | 1,021 | 2,639 | 0.6 | 1.9 | 31 | 82 | 0.75 (0.38 - 1.39) | 0.37 | 0.71 (0.37 - 1.35) | 0.32 |
| Open Claims | 54,529 | 132,608 | 33.7 | 93.1 | 958 | 2,429 | 1.05 (0.94 - 1.16) | 0.40 | 0.99 (0.89 - 1.11) | 0.84 |
| OptumEHR | 2,426 | 6,262 | 0.6 | 1.7 | 30 | 47 | 1.87 (0.95 - 3.63) | 0.07 | 1.72 (0.88 - 3.37) | 0.13 |
| Meta-analysis | 61,325 | 149,991 | 36.6 | 101.7 | 1,133 | 2,901 | 1.04 (0.94 - 1.14) | 0.47 | 1.02 (0.92 - 1.13) | 0.74 |
| **PS-stratification / intent-to-treat TAR** | | | | | | | | | | |
| OptumDOD | 3,713 | 8,491 | 4.4 | 12.2 | 358 | 927 | 0.98 (0.86 - 1.12) | 0.81 | 0.98 (0.86 - 1.13) | 0.80 |
| MDCR | 1,085 | 2,647 | 1.3 | 4.1 | 98 | 261 | 1.10 (0.86 - 1.40) | 0.45 | 1.01 (0.79 - 1.30) | 0.84 |
| Open Claims | 58,688 | 132,741 | 115.1 | 316.7 | 4,132 | 10,335 | 1.10 (1.06 - 1.14) | <0.01 | 1.07 (0.97 - 1.18) | 0.19 |
| OptumEHR | 2,573 | 6,272 | 4.3 | 12.3 | 165 | 421 | 1.06 (0.87 - 1.28) | 0.56 | 1.07 (0.88 - 1.29) | 0.55 |
| Meta-analysis | 66,059 | 150,151 | 125.2 | 345.3 | 4,753 | 11,944 | 1.08 (1.01 - 1.15) | 0.02 | 1.05 (0.95 - 1.17) | 0.34 |
| **PS-matching / intent-to-treat TAR** | | | | | | | | | | |
| OptumDOD | 3,349 | 8,482 | 4.1 | 12.2 | 334 | 922 | 0.94 (0.79 - 1.10) | 0.44 | 0.95 (0.81 - 1.13) | 0.61 |
| MDCR | 1,021 | 2,639 | 1.3 | 4.1 | 88 | 261 | 0.96 (0.68 - 1.36) | 0.84 | 0.83 (0.58 - 1.20) | 0.44 |
| Open Claims | 54,529 | 132,608 | 109.1 | 316.6 | 3,846 | 10,321 | 1.11 (1.06 - 1.16) | <0.01 | 1.07 (0.99 - 1.15) | 0.15 |
| OptumEHR | 2,426 | 6,262 | 4.2 | 12.3 | 157 | 420 | 1.01 (0.79 - 1.28) | 0.94 | 1.04 (0.82 - 1.33) | 0.74 |
| Meta-analysis | 61,325 | 149,991 | 118.7 | 345.1 | 4,425 | 11,924 | 1.04 (0.93 - 1.16) | 0.51 | 1.02 (0.91 - 1.15) | 0.72 |

##### 6.1.16 Photosensitivity

**Supplementary Table 123:** Exposures, outcomes and risk estimates for Photosensitivity in GLP1RA (T) vs SGLT2I (C) new-user cohorts under four different propensity score (PS) adjustment / time-at-risk (TAR) designs across data sources. We report total patient exposure time in 1,000 patient-year (KYr) units, uncalibrated and calibrated hazard ratios (HRs) and their 95% confidence intervals (CIs). Greyed out entries did not pass study diagnostics and are included only for completeness.

| | Patients | | Exposure (KYr) | | Outcomes | | Uncalibrated | | Calibrated | |
|---|---|---|---|---|---|---|---|---|---|---|
| | T | C | T | C | T | C | HR (95% CI) | $p$ | HR (95% CI) | $p$ |
| **PS-stratification / on-treatment TAR** | | | | | | | | | | |
| OptumDOD | 5,293 | 11,516 | 2.7 | 6.9 | <5 | <5 | 1.60 (0.08 - 13.9) | 0.72 | 1.63 (0.12 - 22.1) | 0.72 |
| MDCR | 1,461 | 3,464 | 0.9 | 2.5 | <5 | <5 | 2.28 (0.27 - 15.2) | 0.42 | 1.99 (0.27 - 14.8) | 0.50 |
| Open Claims | 85,859 | 181,939 | 52.1 | 126.5 | 20 | 29 | 2.07 (1.10 - 3.82) | 0.02 | 2.00 (1.07 - 3.73) | 0.03 |
| OptumEHR | 3,366 | 7,758 | 0.9 | 2.2 | <5 | 0 | | | | |
| Meta-analysis | 85,859 | 181,939 | 52.1 | 126.5 | 20 | 29 | 2.07 (1.10 - 3.82) | 0.02 | 2.00 (1.07 - 3.73) | 0.03 |
| **PS-matching / on-treatment TAR** | | | | | | | | | | |
| OptumDOD | 4,739 | 11,501 | 2.5 | 6.9 | <5 | <5 | 2.36 (0.10 - 27.4) | 0.55 | 2.63 (0.16 - 43.3) | 0.50 |
| MDCR | 1,365 | 3,456 | 0.9 | 2.5 | <5 | <5 | 2.81 (0.30 - 25.7) | 0.36 | 2.66 (0.29 - 24.5) | 0.39 |
| Open Claims | 78,393 | 181,885 | 48.4 | 126.4 | 17 | 29 | 1.75 (0.82 - 3.70) | 0.15 | 1.66 (0.78 - 3.54) | 0.19 |
| OptumEHR | 3,146 | 7,746 | 0.8 | 2.2 | <5 | 0 | | | | |
| Meta-analysis | 78,393 | 181,885 | 48.4 | 126.4 | 17 | 29 | 1.75 (0.82 - 3.70) | 0.15 | 1.71 (0.80 - 3.65) | 0.16 |
| **PS-stratification / intent-to-treat TAR** | | | | | | | | | | |
| OptumDOD | 5,293 | 11,516 | 6.8 | 17.6 | <5 | 9 | 1.55 (0.39 - 5.18) | 0.51 | 1.54 (0.42 - 5.62) | 0.51 |
| MDCR | 1,461 | 3,464 | 1.9 | 5.8 | <5 | 9 | 0.72 (0.11 - 3.01) | 0.70 | 0.66 (0.12 - 3.54) | 0.63 |
| Open Claims | 85,859 | 181,939 | 177.2 | 451.1 | 42 | 97 | 1.10 (0.74 - 1.60) | 0.63 | 1.07 (0.72 - 1.58) | 0.75 |
| OptumEHR | 3,366 | 7,758 | 6.0 | 16.0 | 7 | 7 | 2.37 (0.77 - 7.35) | 0.13 | 2.39 (0.77 - 7.40) | 0.13 |
| Meta-analysis | 85,859 | 181,939 | 177.2 | 451.1 | 42 | 97 | 1.10 (0.74 - 1.60) | 0.63 | 1.07 (0.72 - 1.60) | 0.72 |
| **PS-matching / intent-to-treat TAR** | | | | | | | | | | |
| OptumDOD | 4,739 | 11,501 | 6.3 | 17.6 | <5 | 8 | 1.39 (0.18 - 8.02) | 0.73 | 1.42 (0.21 - 9.47) | 0.72 |
| MDCR | 1,365 | 3,456 | 1.9 | 5.8 | <5 | 9 | 0.92 (0.13 - 4.20) | 0.92 | 0.79 (0.14 - 4.53) | 0.79 |
| Open Claims | 78,393 | 181,885 | 165.8 | 451.1 | 36 | 97 | 1.05 (0.66 - 1.63) | 0.85 | 1.01 (0.64 - 1.59) | 0.97 |
| OptumEHR | 3,146 | 7,746 | 5.7 | 16.0 | 6 | 7 | 1.67 (0.39 - 6.67) | 0.48 | 1.72 (0.42 - 7.10) | 0.45 |
| Meta-analysis | 78,393 | 181,885 | 165.8 | 451.1 | 36 | 97 | 1.05 (0.66 - 1.63) | 0.85 | 1.03 (0.65 - 1.62) | 0.90 |

##### 6.1.17 Venous thromboembolism

**Supplementary Table 124:** Exposures, outcomes and risk estimates for Venous thromboembolism in GLP1RA (T) vs SGLT2I (C) new-user cohorts under four different propensity score (PS) adjustment / time-at-risk (TAR) designs across data sources. We report total patient exposure time in 1,000 patient-year (KYr) units, uncalibrated and calibrated hazard ratios (HRs) and their 95% confidence intervals (CIs). Greyed out entries did not pass study diagnostics and are included only for completeness.

| | Patients | | Exposure (KYr) | | Outcomes | | Uncalibrated | | Calibrated | |
|---|---|---|---|---|---|---|---|---|---|---|
| | T | C | T | C | T | C | HR (95% CI) | $p$ | HR (95% CI) | $p$ |
| **PS-stratification / on-treatment TAR** | | | | | | | | | | |
| OptumDOD | 5,063 | 11,177 | 2.6 | 6.7 | 28 | 67 | 0.89 (0.54 - 1.41) | 0.62 | 0.90 (0.56 - 1.45) | 0.67 |
| MDCR | 1,418 | 3,367 | 0.8 | 2.5 | 11 | 19 | 1.61 (0.72 - 3.41) | 0.23 | 1.40 (0.64 - 3.05) | 0.40 |
| Open Claims | 82,510 | 176,274 | 50.2 | 122.7 | 268 | 661 | 0.95 (0.82 - 1.11) | 0.54 | 0.92 (0.79 - 1.07) | 0.32 |
| OptumEHR | 3,263 | 7,536 | 0.9 | 2.1 | 10 | 17 | 1.01 (0.43 - 2.29) | 0.97 | 0.99 (0.43 - 2.28) | 0.89 |
| Meta-analysis | 92,254 | 198,354 | 54.5 | 133.9 | 317 | 764 | 0.97 (0.84 - 1.11) | 0.63 | 0.93 (0.81 - 1.08) | 0.35 |
| **PS-matching / on-treatment TAR** | | | | | | | | | | |
| OptumDOD | 4,546 | 11,162 | 2.4 | 6.6 | 25 | 67 | 1.20 (0.66 - 2.13) | 0.54 | 1.34 (0.75 - 2.42) | 0.33 |
| MDCR | 1,333 | 3,360 | 0.8 | 2.4 | 11 | 19 | 0.77 (0.26 - 2.03) | 0.62 | 0.73 (0.26 - 2.03) | 0.56 |
| Open Claims | 75,451 | 176,226 | 46.8 | 122.7 | 243 | 661 | 0.98 (0.80 - 1.20) | 0.87 | 0.93 (0.76 - 1.14) | 0.53 |
| OptumEHR | 3,046 | 7,524 | 0.8 | 2.1 | 8 | 17 | 0.99 (0.30 - 2.97) | 0.99 | 0.91 (0.29 - 2.90) | 0.86 |
| Meta-analysis | 84,376 | 198,272 | 50.8 | 133.9 | 287 | 764 | 0.99 (0.83 - 1.20) | 0.95 | 0.98 (0.81 - 1.18) | 0.80 |
| **PS-stratification / intent-to-treat TAR** | | | | | | | | | | |
| OptumDOD | 5,063 | 11,177 | 6.5 | 16.9 | 74 | 209 | 0.85 (0.64 - 1.12) | 0.25 | 0.85 (0.64 - 1.13) | 0.28 |
| MDCR | 1,418 | 3,367 | 1.9 | 5.6 | 26 | 61 | 1.30 (0.79 - 2.09) | 0.29 | 1.20 (0.73 - 1.95) | 0.49 |
| Open Claims | 82,510 | 176,274 | 169.3 | 435.3 | 1,185 | 2,914 | 1.04 (0.96 - 1.11) | 0.34 | 1.01 (0.90 - 1.12) | 0.90 |
| OptumEHR | 3,263 | 7,536 | 5.8 | 15.4 | 44 | 104 | 1.01 (0.69 - 1.46) | 0.95 | 1.02 (0.70 - 1.48) | 0.89 |
| Meta-analysis | 92,254 | 198,354 | 183.4 | 473.2 | 1,329 | 3,288 | 1.03 (0.96 - 1.10) | 0.43 | 1.00 (0.90 - 1.12) | 0.88 |
| **PS-matching / intent-to-treat TAR** | | | | | | | | | | |
| OptumDOD | 4,546 | 11,162 | 6.0 | 16.9 | 68 | 208 | 0.81 (0.56 - 1.13) | 0.23 | 0.82 (0.58 - 1.16) | 0.28 |
| MDCR | 1,333 | 3,360 | 1.8 | 5.6 | 26 | 61 | 1.07 (0.56 - 1.99) | 0.83 | 0.93 (0.49 - 1.77) | 0.83 |
| Open Claims | 75,451 | 176,226 | 158.5 | 435.2 | 1,082 | 2,913 | 1.05 (0.96 - 1.15) | 0.27 | 1.01 (0.91 - 1.13) | 0.81 |
| OptumEHR | 3,046 | 7,524 | 5.5 | 15.4 | 41 | 104 | 1.02 (0.64 - 1.60) | 0.93 | 1.05 (0.67 - 1.67) | 0.82 |
| Meta-analysis | 84,376 | 198,272 | 171.7 | 473.1 | 1,217 | 3,286 | 1.03 (0.92 - 1.14) | 0.63 | 1.01 (0.90 - 1.14) | 0.85 |

##### 6.1.18 Vomiting

**Supplementary Table 125:** Exposures, outcomes and risk estimates for Vomiting in GLP1RA (T) vs SGLT2I (C) new-user cohorts under four different propensity score (PS) adjustment / time-at-risk (TAR) designs across data sources. We report total patient exposure time in 1,000 patient-year (KYr) units, uncalibrated and calibrated hazard ratios (HRs) and their 95% confidence intervals (CIs). Greyed out entries did not pass study diagnostics and are included only for completeness.

| | Patients | | Exposure (KYr) | | Outcomes | | Uncalibrated | | Calibrated | |
|---|---|---|---|---|---|---|---|---|---|---|
| | T | C | T | C | T | C | HR (95% CI) | $p$ | HR (95% CI) | $p$ |
| **PS-stratification / on-treatment TAR** | | | | | | | | | | |
| OptumDOD | 4,765 | 10,476 | 2.4 | 6.2 | 126 | 155 | 1.83 (1.42 - 2.37) | <0.01 | 1.86 (1.44 - 2.41) | <0.01 |
| MDCR | 1,385 | 3,271 | 0.8 | 2.4 | 27 | 48 | 1.42 (0.85 - 2.33) | 0.18 | 1.24 (0.75 - 2.06) | 0.43 |
| Open Claims | 76,917 | 165,476 | 46.6 | 115.6 | 932 | 1,324 | 1.62 (1.48 - 1.77) | <0.01 | 1.56 (1.42 - 1.72) | <0.01 |
| OptumEHR | 3,174 | 7,398 | 0.8 | 2.1 | 33 | 40 | 1.87 (1.14 - 3.04) | 0.01 | 1.82 (1.11 - 2.97) | 0.02 |
| Meta-analysis | 86,241 | 186,621 | 50.7 | 126.3 | 1,118 | 1,567 | 1.64 (1.51 - 1.78) | <0.01 | 1.58 (1.44 - 1.74) | <0.01 |
| **PS-matching / on-treatment TAR** | | | | | | | | | | |
| OptumDOD | 4,261 | 10,462 | 2.2 | 6.2 | 107 | 155 | 1.80 (1.27 - 2.55) | <0.01 | 2.02 (1.42 - 2.85) | <0.01 |
| MDCR | 1,296 | 3,263 | 0.8 | 2.4 | 26 | 48 | 1.30 (0.65 - 2.54) | 0.45 | 1.23 (0.62 - 2.43) | 0.58 |
| Open Claims | 70,503 | 165,428 | 43.5 | 115.6 | 842 | 1,323 | 1.81 (1.60 - 2.04) | <0.01 | 1.72 (1.52 - 1.94) | <0.01 |
| OptumEHR | 2,968 | 7,387 | 0.8 | 2.1 | 29 | 40 | 1.80 (0.96 - 3.34) | 0.06 | 1.67 (0.89 - 3.11) | 0.13 |
| Meta-analysis | 79,028 | 186,540 | 47.3 | 126.3 | 1,004 | 1,566 | 1.79 (1.61 - 2.00) | <0.01 | 1.76 (1.56 - 1.98) | <0.01 |
| **PS-stratification / intent-to-treat TAR** | | | | | | | | | | |
| OptumDOD | 4,765 | 10,476 | 5.8 | 15.6 | 266 | 462 | 1.36 (1.16 - 1.60) | <0.01 | 1.36 (1.15 - 1.61) | <0.01 |
| MDCR | 1,385 | 3,271 | 1.8 | 5.4 | 61 | 124 | 1.31 (0.94 - 1.81) | 0.10 | 1.21 (0.87 - 1.67) | 0.29 |
| Open Claims | 76,917 | 165,476 | 154.9 | 403.7 | 2,771 | 5,532 | 1.23 (1.17 - 1.29) | <0.01 | 1.19 (1.08 - 1.31) | <0.01 |
| OptumEHR | 3,174 | 7,398 | 5.5 | 14.9 | 94 | 186 | 1.25 (0.96 - 1.62) | 0.10 | 1.25 (0.96 - 1.64) | 0.10 |
| Meta-analysis | 86,241 | 186,621 | 168.0 | 439.7 | 3,192 | 6,304 | 1.24 (1.18 - 1.31) | <0.01 | 1.22 (1.10 - 1.34) | <0.01 |
| **PS-matching / intent-to-treat TAR** | | | | | | | | | | |
| OptumDOD | 4,261 | 10,462 | 5.4 | 15.6 | 231 | 460 | 1.46 (1.17 - 1.81) | <0.01 | 1.48 (1.19 - 1.84) | <0.01 |
| MDCR | 1,296 | 3,263 | 1.7 | 5.4 | 57 | 124 | 1.13 (0.75 - 1.69) | 0.54 | 0.98 (0.64 - 1.49) | 0.87 |
| Open Claims | 70,503 | 165,428 | 145.4 | 403.7 | 2,545 | 5,530 | 1.27 (1.20 - 1.35) | <0.01 | 1.23 (1.13 - 1.34) | <0.01 |
| OptumEHR | 2,968 | 7,387 | 5.2 | 14.9 | 81 | 186 | 1.12 (0.81 - 1.54) | 0.48 | 1.16 (0.84 - 1.60) | 0.39 |
| Meta-analysis | 79,028 | 186,540 | 157.7 | 439.5 | 2,914 | 6,300 | 1.28 (1.21 - 1.35) | <0.01 | 1.26 (1.17 - 1.35) | <0.01 |

#### 6.2 GLP1RA vs DPP4I

##### 6.2.1 Abnormal weight gain

**Supplementary Table 126:** Exposures, outcomes and risk estimates for Abnormal weight gain in GLP1RA (T) vs DPP4I (C) new-user cohorts under four different propensity score (PS) adjustment / time-at-risk (TAR) designs across data sources. We report total patient exposure time in 1,000 patient-year (KYr) units, uncalibrated and calibrated hazard ratios (HRs) and their 95% confidence intervals (CIs). Greyed out entries did not pass study diagnostics and are included only for completeness.

| | Patients | | Exposure (KYr) | | Outcomes | | Uncalibrated | | Calibrated | |
|---|---|---|---|---|---|---|---|---|---|---|
| | T | C | T | C | T | C | HR (95% CI) | $p$ | HR (95% CI) | $p$ |
| **PS-stratification / on-treatment TAR** | | | | | | | | | | |
| OptumDOD | 5,281 | 26,500 | 2.7 | 22.0 | 18 | 122 | 1.18 (0.65 - 2.03) | 0.57 | 1.15 (0.65 - 2.03) | 0.63 |
| MDCR | 1,915 | 17,725 | 1.2 | 17.7 | 7 | 47 | 3.14 (1.19 - 7.21) | 0.01 | 2.79 (1.13 - 6.87) | 0.03 |
| Open Claims | 81,161 | 334,884 | 49.5 | 310.4 | 108 | 549 | 1.16 (0.91 - 1.47) | 0.23 | 1.15 (0.86 - 1.53) | 0.36 |
| OptumEHR | 3,350 | 22,797 | 0.9 | 11.8 | 10 | 55 | 2.18 (0.98 - 4.43) | 0.04 | 1.67 (0.78 - 3.57) | 0.19 |
| Meta-analysis | 84,511 | 357,681 | 50.4 | 322.2 | 118 | 604 | 1.43 (0.80 - 2.57) | 0.23 | 1.44 (0.80 - 2.61) | 0.23 |
| **PS-matching / on-treatment TAR** | | | | | | | | | | |
| OptumDOD | 4,536 | 25,289 | 2.5 | 21.0 | 14 | 113 | 0.98 (0.44 - 1.99) | 0.96 | 0.98 (0.46 - 2.07) | 0.93 |
| MDCR | 1,725 | 16,893 | 1.1 | 17.0 | 7 | 46 | 3.58 (1.17 - 9.62) | 0.02 | 3.19 (1.11 - 9.15) | 0.03 |
| Open Claims | 66,636 | 331,260 | 44.0 | 307.1 | 81 | 543 | 1.07 (0.75 - 1.50) | 0.70 | 1.04 (0.72 - 1.51) | 0.83 |
| OptumEHR | 3,089 | 22,640 | 0.9 | 11.7 | 9 | 54 | 2.20 (0.81 - 5.64) | 0.11 | 1.78 (0.67 - 4.71) | 0.26 |
| Meta-analysis | 75,986 | 396,082 | 48.5 | 356.8 | 111 | 756 | 1.47 (0.86 - 2.52) | 0.16 | 1.42 (0.81 - 2.48) | 0.22 |
| **PS-stratification / intent-to-treat TAR** | | | | | | | | | | |
| OptumDOD | 5,281 | 26,500 | 7.8 | 79.6 | 57 | 396 | 1.32 (0.95 - 1.79) | 0.09 | 1.32 (0.96 - 1.81) | 0.09 |
| MDCR | 1,915 | 17,725 | 3.9 | 50.5 | 28 | 178 | 1.67 (1.05 - 2.57) | 0.02 | 1.51 (0.97 - 2.37) | 0.07 |
| Open Claims | 81,161 | 334,884 | 168.5 | 1424.0 | 469 | 2,783 | 1.46 (1.31 - 1.63) | <0.01 | 1.44 (1.19 - 1.75) | <0.01 |
| OptumEHR | 3,350 | 22,797 | 6.9 | 77.9 | 56 | 281 | 2.27 (1.63 - 3.10) | <0.01 | 2.15 (1.56 - 2.97) | <0.01 |
| Meta-analysis | 84,511 | 357,681 | 175.3 | 1501.8 | 525 | 3,064 | 1.77 (1.16 - 2.71) | <0.01 | 1.76 (1.12 - 2.76) | 0.01 |
| **PS-matching / intent-to-treat TAR** | | | | | | | | | | |
| OptumDOD | 4,536 | 25,289 | 7.4 | 74.7 | 52 | 367 | 1.16 (0.79 - 1.68) | 0.44 | 1.18 (0.81 - 1.73) | 0.41 |
| MDCR | 1,725 | 16,893 | 3.8 | 48.5 | 28 | 172 | 2.13 (1.21 - 3.61) | <0.01 | 1.94 (1.12 - 3.36) | 0.02 |
| Open Claims | 66,636 | 331,260 | 156.7 | 1407.7 | 410 | 2,737 | 1.38 (1.22 - 1.57) | <0.01 | 1.36 (1.12 - 1.65) | <0.01 |
| OptumEHR | 3,089 | 22,640 | 6.6 | 77.1 | 52 | 278 | 2.15 (1.49 - 3.06) | <0.01 | 2.05 (1.43 - 2.93) | <0.01 |
| Meta-analysis | 75,986 | 396,082 | 174.5 | 1608.1 | 542 | 3,554 | 1.59 (1.19 - 2.11) | <0.01 | 1.54 (1.12 - 2.11) | <0.01 |

##### 6.2.2 Abnormal weight loss

**Supplementary Table 127:** Exposures, outcomes and risk estimates for Abnormal weight loss in GLP1RA (T) vs DPP4I (C) new-user cohorts under four different propensity score (PS) adjustment / time-at-risk (TAR) designs across data sources. We report total patient exposure time in 1,000 patient-year (KYr) units, uncalibrated and calibrated hazard ratios (HRs) and their 95% confidence intervals (CIs). Greyed out entries did not pass study diagnostics and are included only for completeness.

| | Patients | | Exposure (KYr) | | Outcomes | | Uncalibrated | | Calibrated | |
|---|---|---|---|---|---|---|---|---|---|---|
| | T | C | T | C | T | C | HR (95% CI) | $p$ | HR (95% CI) | $p$ |
| **PS-stratification / on-treatment TAR** | | | | | | | | | | |
| OptumDOD | 5,409 | 25,475 | 2.8 | 21.1 | 60 | 451 | 1.37 (1.00 - 1.85) | 0.05 | 1.34 (0.98 - 1.82) | 0.08 |
| MDCR | 1,954 | 17,148 | 1.2 | 17.1 | 16 | 251 | 1.01 (0.56 - 1.70) | 0.98 | 0.89 (0.51 - 1.56) | 0.70 |
| Open Claims | 83,942 | 326,105 | 51.0 | 301.2 | 368 | 2,882 | 0.97 (0.86 - 1.10) | 0.68 | 0.96 (0.79 - 1.18) | 0.73 |
| OptumEHR | 3,464 | 22,394 | 1.0 | 11.5 | 9 | 206 | 0.63 (0.29 - 1.21) | 0.21 | 0.49 (0.24 - 0.99) | 0.05 |
| Meta-analysis | 87,406 | 348,499 | 51.9 | 312.8 | 377 | 3,088 | 0.91 (0.67 - 1.24) | 0.55 | 0.92 (0.66 - 1.27) | 0.61 |
| **PS-matching / on-treatment TAR** | | | | | | | | | | |
| OptumDOD | 4,571 | 24,676 | 2.5 | 20.4 | 59 | 430 | 1.40 (0.95 - 2.02) | 0.08 | 1.39 (0.95 - 2.04) | 0.10 |
| MDCR | 1,735 | 16,470 | 1.2 | 16.5 | 15 | 233 | 1.32 (0.68 - 2.39) | 0.38 | 1.18 (0.63 - 2.21) | 0.62 |
| Open Claims | 67,430 | 323,455 | 44.6 | 298.8 | 327 | 2,826 | 0.94 (0.81 - 1.10) | 0.47 | 0.92 (0.75 - 1.13) | 0.44 |
| OptumEHR | 3,142 | 22,259 | 0.9 | 11.5 | 8 | 203 | 1.05 (0.44 - 2.18) | 0.91 | 0.84 (0.38 - 1.87) | 0.69 |
| Meta-analysis | 76,878 | 386,860 | 49.1 | 347.2 | 409 | 3,692 | 1.11 (0.86 - 1.43) | 0.44 | 1.07 (0.80 - 1.43) | 0.66 |
| **PS-stratification / intent-to-treat TAR** | | | | | | | | | | |
| OptumDOD | 5,409 | 25,475 | 7.9 | 74.4 | 163 | 1,658 | 1.05 (0.88 - 1.25) | 0.57 | 1.06 (0.88 - 1.27) | 0.58 |
| MDCR | 1,954 | 17,148 | 3.9 | 47.9 | 52 | 875 | 0.87 (0.64 - 1.16) | 0.38 | 0.79 (0.59 - 1.07) | 0.14 |
| Open Claims | 83,942 | 326,105 | 172.5 | 1365.6 | 1,215 | 14,182 | 0.87 (0.81 - 0.92) | <0.01 | 0.86 (0.72 - 1.01) | 0.07 |
| OptumEHR | 3,464 | 22,394 | 7.0 | 74.9 | 85 | 1,169 | 0.97 (0.76 - 1.22) | 0.80 | 0.92 (0.73 - 1.17) | 0.53 |
| Meta-analysis | 87,406 | 348,499 | 179.5 | 1440.4 | 1,300 | 15,351 | 0.87 (0.82 - 0.93) | <0.01 | 0.87 (0.74 - 1.01) | 0.08 |
| **PS-matching / intent-to-treat TAR** | | | | | | | | | | |
| OptumDOD | 4,571 | 24,676 | 7.4 | 71.4 | 157 | 1,559 | 1.07 (0.87 - 1.31) | 0.50 | 1.09 (0.89 - 1.34) | 0.45 |
| MDCR | 1,735 | 16,470 | 3.8 | 46.3 | 51 | 820 | 0.88 (0.62 - 1.22) | 0.47 | 0.80 (0.57 - 1.13) | 0.22 |
| Open Claims | 67,430 | 323,455 | 158.6 | 1354.3 | 1,125 | 13,954 | 0.87 (0.81 - 0.93) | <0.01 | 0.86 (0.73 - 1.01) | 0.06 |
| OptumEHR | 3,142 | 22,259 | 6.7 | 74.3 | 82 | 1,147 | 0.91 (0.69 - 1.18) | 0.50 | 0.87 (0.66 - 1.13) | 0.32 |
| Meta-analysis | 76,878 | 386,860 | 176.5 | 1546.3 | 1,415 | 17,480 | 0.92 (0.82 - 1.02) | 0.12 | 0.89 (0.75 - 1.06) | 0.18 |

##### 6.2.3 Acute pancreatitis

**Supplementary Table 128:** Exposures, outcomes and risk estimates for Acute pancreatitis in GLP1RA (T) vs DPP4I (C) new-user cohorts under four different propensity score (PS) adjustment / time-at-risk (TAR) designs across data sources. We report total patient exposure time in 1,000 patient-year (KYr) units, uncalibrated and calibrated hazard ratios (HRs) and their 95% confidence intervals (CIs). Greyed out entries did not pass study diagnostics and are included only for completeness.

| | Patients | | Exposure (KYr) | | Outcomes | | Uncalibrated | | Calibrated | |
|---|---|---|---|---|---|---|---|---|---|---|
| | T | C | T | C | T | C | HR (95% CI) | $p$ | HR (95% CI) | $p$ |
| **PS-stratification / on-treatment TAR** | | | | | | | | | | |
| OptumDOD | 5,591 | 26,987 | 2.9 | 22.5 | 9 | 78 | 1.37 (0.61 - 2.77) | 0.41 | 1.34 (0.63 - 2.86) | 0.45 |
| MDCR | 1,985 | 17,814 | 1.3 | 17.8 | <5 | 55 | 1.60 (0.44 - 4.42) | 0.42 | 1.42 (0.45 - 4.49) | 0.55 |
| Open Claims | 86,463 | 340,319 | 52.6 | 315.6 | 87 | 736 | 0.95 (0.73 - 1.21) | 0.69 | 0.94 (0.70 - 1.26) | 0.69 |
| OptumEHR | 3,555 | 23,251 | 1.0 | 12.1 | <5 | 26 | 1.50 (0.34 - 4.59) | 0.54 | 1.15 (0.31 - 4.22) | 0.83 |
| Meta-analysis | 86,463 | 340,319 | 52.6 | 315.6 | 87 | 736 | 0.95 (0.73 - 1.21) | 0.69 | 0.96 (0.73 - 1.26) | 0.76 |
| **PS-matching / on-treatment TAR** | | | | | | | | | | |
| OptumDOD | 4,723 | 25,870 | 2.6 | 21.6 | 9 | 74 | 1.06 (0.41 - 2.43) | 0.89 | 1.06 (0.43 - 2.59) | 0.90 |
| MDCR | 1,770 | 16,989 | 1.2 | 17.1 | <5 | 50 | 1.32 (0.29 - 4.30) | 0.68 | 1.18 (0.31 - 4.55) | 0.81 |
| Open Claims | 69,581 | 336,618 | 46.0 | 312.1 | 77 | 718 | 0.93 (0.68 - 1.24) | 0.62 | 0.90 (0.65 - 1.25) | 0.55 |
| OptumEHR | 3,229 | 23,093 | 0.9 | 12.0 | <5 | 26 | 0.72 (0.11 - 2.83) | 0.70 | 0.58 (0.11 - 2.99) | 0.52 |
| Meta-analysis | 76,074 | 379,477 | 49.8 | 350.8 | <91 | 842 | 0.95 (0.72 - 1.26) | 0.74 | 0.92 (0.67 - 1.26) | 0.61 |
| **PS-stratification / intent-to-treat TAR** | | | | | | | | | | |
| OptumDOD | 5,591 | 26,987 | 8.3 | 81.3 | 31 | 293 | 1.25 (0.82 - 1.85) | 0.27 | 1.26 (0.84 - 1.89) | 0.28 |
| MDCR | 1,985 | 17,814 | 4.1 | 50.6 | 15 | 174 | 1.29 (0.71 - 2.21) | 0.38 | 1.17 (0.66 - 2.07) | 0.59 |
| Open Claims | 86,463 | 340,319 | 179.1 | 1447.5 | 306 | 3,201 | 0.96 (0.84 - 1.09) | 0.52 | 0.95 (0.77 - 1.16) | 0.60 |
| OptumEHR | 3,555 | 23,251 | 7.3 | 79.9 | 13 | 120 | 1.58 (0.81 - 2.84) | 0.15 | 1.51 (0.81 - 2.81) | 0.20 |
| Meta-analysis | 90,018 | 363,570 | 186.4 | 1527.3 | 319 | 3,321 | 1.12 (0.71 - 1.76) | 0.63 | 1.11 (0.69 - 1.79) | 0.67 |
| **PS-matching / intent-to-treat TAR** | | | | | | | | | | |
| OptumDOD | 4,723 | 25,870 | 7.8 | 76.9 | 29 | 275 | 1.23 (0.76 - 1.91) | 0.38 | 1.25 (0.79 - 1.98) | 0.36 |
| MDCR | 1,770 | 16,989 | 4.0 | 48.6 | 15 | 164 | 1.21 (0.60 - 2.25) | 0.57 | 1.10 (0.57 - 2.13) | 0.78 |
| Open Claims | 69,581 | 336,618 | 164.9 | 1430.9 | 281 | 3,138 | 0.96 (0.83 - 1.10) | 0.55 | 0.94 (0.77 - 1.15) | 0.56 |
| OptumEHR | 3,229 | 23,093 | 7.0 | 79.2 | 12 | 120 | 1.44 (0.68 - 2.77) | 0.31 | 1.37 (0.68 - 2.76) | 0.38 |
| Meta-analysis | 79,303 | 402,570 | 183.7 | 1635.5 | 337 | 3,697 | 1.05 (0.86 - 1.28) | 0.65 | 1.02 (0.80 - 1.29) | 0.89 |

##### 6.2.4 All-cause mortality

**Supplementary Table 129:** Exposures, outcomes and risk estimates for All-cause mortality in GLP1RA (T) vs DPP4I (C) new-user cohorts under four different propensity score (PS) adjustment / time-at-risk (TAR) designs across data sources. We report total patient exposure time in 1,000 patient-year (KYr) units, uncalibrated and calibrated hazard ratios (HRs) and their 95% confidence intervals (CIs). Greyed out entries did not pass study diagnostics and are included only for completeness.

| | Patients | | Exposure (KYr) | | Outcomes | | Uncalibrated | | Calibrated | |
|---|---|---|---|---|---|---|---|---|---|---|
| | T | C | T | C | T | C | HR (95% CI) | $p$ | HR (95% CI) | $p$ |
| **PS-stratification / on-treatment TAR** | | | | | | | | | | |
| OptumDOD | 5,626 | 27,232 | 2.9 | 22.7 | 16 | 276 | 0.67 (0.37 - 1.13) | 0.16 | 0.65 (0.37 - 1.14) | 0.14 |
| MDCR | 1,993 | 17,961 | 1.3 | 18.0 | <5 | 50 | 1.74 (0.27 - 5.98) | 0.48 | 1.54 (0.33 - 7.19) | 0.58 |
| Open Claims | 87,486 | 344,544 | 53.1 | 319.7 | 0 | <5 | | | | |
| OptumEHR | 3,573 | 23,343 | 1.0 | 12.1 | <5 | 172 | 0.30 (0.07 - 0.81) | 0.05 | 0.23 (0.07 - 0.77) | 0.02 |
| Meta-analysis | 3,573 | 23,343 | 1.0 | 12.1 | <5 | 172 | 0.30 (0.07 - 0.81) | 0.05 | 0.30 (0.09 - 1.01) | 0.05 |
| **PS-matching / on-treatment TAR** | | | | | | | | | | |
| OptumDOD | 4,755 | 26,062 | 2.6 | 21.8 | 15 | 241 | 0.73 (0.35 - 1.38) | 0.36 | 0.72 (0.36 - 1.44) | 0.36 |
| MDCR | 1,778 | 17,109 | 1.2 | 17.2 | <5 | 41 | 1.28 (0.20 - 4.75) | 0.76 | 1.14 (0.23 - 5.62) | 0.87 |
| Open Claims | 70,428 | 340,897 | 46.6 | 316.3 | 0 | <5 | | | | |
| OptumEHR | 3,242 | 23,184 | 0.9 | 12.1 | <5 | 163 | 0.49 (0.11 - 1.41) | 0.26 | 0.39 (0.11 - 1.38) | 0.15 |
| Meta-analysis | 7,997 | 49,246 | 3.5 | 33.8 | <20 | 404 | 0.66 (0.36 - 1.21) | 0.18 | 0.64 (0.34 - 1.19) | 0.16 |
| **PS-stratification / intent-to-treat TAR** | | | | | | | | | | |
| OptumDOD | 5,626 | 27,232 | 8.4 | 81.2 | 169 | 3,230 | 0.85 (0.72 - 1.00) | 0.05 | 0.85 (0.72 - 1.01) | 0.08 |
| MDCR | 1,993 | 17,961 | 4.1 | 51.4 | 12 | 246 | 1.04 (0.54 - 1.82) | 0.90 | 0.94 (0.51 - 1.73) | 0.85 |
| Open Claims | 87,486 | 344,544 | 181.6 | 1472.9 | 0 | 23 | | | | |
| OptumEHR | 3,573 | 23,343 | 7.4 | 80.2 | 44 | 1,042 | 0.56 (0.40 - 0.77) | <0.01 | 0.54 (0.39 - 0.74) | <0.01 |
| Meta-analysis | 3,573 | 23,343 | 7.4 | 80.2 | 44 | 1,042 | 0.56 (0.40 - 0.77) | <0.01 | 0.56 (0.39 - 0.79) | <0.01 |
| **PS-matching / intent-to-treat TAR** | | | | | | | | | | |
| OptumDOD | 4,755 | 26,062 | 7.8 | 76.8 | 164 | 2,830 | 0.89 (0.73 - 1.07) | 0.22 | 0.90 (0.74 - 1.09) | 0.31 |
| MDCR | 1,778 | 17,109 | 4.0 | 49.3 | 12 | 214 | 1.30 (0.65 - 2.36) | 0.43 | 1.18 (0.62 - 2.25) | 0.62 |
| Open Claims | 70,428 | 340,897 | 167.3 | 1456.5 | 0 | 21 | | | | |
| OptumEHR | 3,242 | 23,184 | 7.0 | 79.5 | 43 | 1,015 | 0.57 (0.40 - 0.80) | <0.01 | 0.54 (0.38 - 0.77) | <0.01 |
| Meta-analysis | 9,775 | 66,355 | 18.9 | 205.6 | 219 | 4,059 | 0.82 (0.55 - 1.23) | 0.35 | 0.80 (0.53 - 1.22) | 0.30 |

##### 6.2.5 Bone fracture

**Supplementary Table 130:** Exposures, outcomes and risk estimates for Bone fracture in GLP1RA (T) vs DPP4I (C) new-user cohorts under four different propensity score (PS) adjustment / time-at-risk (TAR) designs across data sources. We report total patient exposure time in 1,000 patient-year (KYr) units, uncalibrated and calibrated hazard ratios (HRs) and their 95% confidence intervals (CIs). Greyed out entries did not pass study diagnostics and are included only for completeness.

| | Patients | | Exposure (KYr) | | Outcomes | | Uncalibrated | | Calibrated | |
|---|---|---|---|---|---|---|---|---|---|---|
| | T | C | T | C | T | C | HR (95% CI) | $p$ | HR (95% CI) | $p$ |
| **PS-stratification / on-treatment TAR** | | | | | | | | | | |
| OptumDOD | 4,964 | 24,065 | 2.5 | 19.6 | 96 | 681 | 1.17 (0.91 - 1.48) | 0.22 | 1.14 (0.89 - 1.46) | 0.33 |
| MDCR | 1,744 | 15,701 | 1.1 | 15.3 | 28 | 539 | 0.80 (0.52 - 1.18) | 0.29 | 0.71 (0.47 - 1.07) | 0.11 |
| Open Claims | 70,749 | 291,457 | 42.8 | 267.0 | 756 | 6,228 | 0.89 (0.82 - 0.97) | <0.01 | 0.88 (0.74 - 1.05) | 0.20 |
| OptumEHR | 3,230 | 21,423 | 0.9 | 10.9 | 20 | 289 | 1.01 (0.60 - 1.59) | 0.98 | 0.77 (0.47 - 1.25) | 0.31 |
| Meta-analysis | 73,979 | 312,880 | 43.7 | 277.9 | 776 | 6,517 | 0.90 (0.82 - 0.97) | <0.01 | 0.90 (0.79 - 1.04) | 0.14 |
| **PS-matching / on-treatment TAR** | | | | | | | | | | |
| OptumDOD | 4,206 | 22,818 | 2.3 | 18.5 | 82 | 624 | 1.07 (0.78 - 1.45) | 0.68 | 1.06 (0.78 - 1.46) | 0.72 |
| MDCR | 1,549 | 15,031 | 1.0 | 14.8 | 28 | 508 | 0.98 (0.60 - 1.51) | 0.92 | 0.87 (0.55 - 1.38) | 0.56 |
| Open Claims | 57,737 | 287,634 | 37.9 | 263.5 | 660 | 6,066 | 0.88 (0.79 - 0.98) | 0.02 | 0.85 (0.72 - 1.01) | 0.10 |
| OptumEHR | 2,949 | 21,287 | 0.8 | 10.9 | 19 | 283 | 1.03 (0.56 - 1.78) | 0.92 | 0.83 (0.46 - 1.48) | 0.55 |
| Meta-analysis | 66,441 | 346,770 | 42.0 | 307.7 | 789 | 7,481 | 0.91 (0.81 - 1.02) | 0.11 | 0.88 (0.73 - 1.05) | 0.16 |
| **PS-stratification / intent-to-treat TAR** | | | | | | | | | | |
| OptumDOD | 4,964 | 24,065 | 7.1 | 67.7 | 237 | 2,472 | 0.96 (0.83 - 1.11) | 0.59 | 0.96 (0.83 - 1.12) | 0.67 |
| MDCR | 1,744 | 15,701 | 3.4 | 42.0 | 125 | 1,608 | 1.04 (0.85 - 1.26) | 0.69 | 0.94 (0.77 - 1.15) | 0.57 |
| Open Claims | 70,749 | 291,457 | 143.2 | 1183.5 | 2,962 | 30,700 | 0.95 (0.92 - 0.99) | 0.03 | 0.94 (0.80 - 1.11) | 0.48 |
| OptumEHR | 3,230 | 21,423 | 6.5 | 70.1 | 116 | 1,629 | 0.93 (0.76 - 1.13) | 0.48 | 0.88 (0.72 - 1.08) | 0.27 |
| Meta-analysis | 73,979 | 312,880 | 149.7 | 1253.6 | 3,078 | 32,329 | 0.95 (0.92 - 0.99) | 0.02 | 0.95 (0.81 - 1.10) | 0.48 |
| **PS-matching / intent-to-treat TAR** | | | | | | | | | | |
| OptumDOD | 4,206 | 22,818 | 6.7 | 63.0 | 215 | 2,276 | 0.94 (0.79 - 1.12) | 0.50 | 0.95 (0.80 - 1.14) | 0.64 |
| MDCR | 1,549 | 15,031 | 3.3 | 40.6 | 123 | 1,510 | 1.08 (0.85 - 1.34) | 0.53 | 0.98 (0.78 - 1.23) | 0.84 |
| Open Claims | 57,737 | 287,634 | 132.6 | 1167.2 | 2,758 | 30,029 | 0.96 (0.91 - 1.00) | 0.05 | 0.94 (0.81 - 1.09) | 0.43 |
| OptumEHR | 2,949 | 21,287 | 6.2 | 69.6 | 112 | 1,602 | 0.91 (0.72 - 1.13) | 0.41 | 0.86 (0.69 - 1.08) | 0.23 |
| Meta-analysis | 66,441 | 346,770 | 148.8 | 1340.5 | 3,208 | 35,417 | 0.96 (0.92 - 1.00) | 0.04 | 0.93 (0.81 - 1.07) | 0.31 |

##### 6.2.6 Diabetic ketoacidosis

**Supplementary Table 131:** Exposures, outcomes and risk estimates for Diabetic ketoacidosis in GLP1RA (T) vs DPP4I (C) new-user cohorts under four different propensity score (PS) adjustment / time-at-risk (TAR) designs across data sources. We report total patient exposure time in 1,000 patient-year (KYr) units, uncalibrated and calibrated hazard ratios (HRs) and their 95% confidence intervals (CIs). Greyed out entries did not pass study diagnostics and are included only for completeness.

| | Patients | | Exposure (KYr) | | Outcomes | | Uncalibrated | | Calibrated | |
|---|---|---|---|---|---|---|---|---|---|---|
| | T | C | T | C | T | C | HR (95% CI) | $p$ | HR (95% CI) | $p$ |
| **PS-stratification / on-treatment TAR** | | | | | | | | | | |
| OptumDOD | 5,619 | 27,194 | 2.9 | 22.7 | <5 | 37 | 0.42 (0.09 - 1.35) | 0.20 | 0.41 (0.11 - 1.57) | 0.19 |
| MDCR | 1,992 | 17,928 | 1.3 | 17.9 | 0 | 17 | | | | |
| Open Claims | 87,394 | 344,081 | 53.1 | 319.1 | 32 | 254 | 0.81 (0.53 - 1.21) | 0.32 | 0.80 (0.51 - 1.25) | 0.34 |
| OptumEHR | 3,572 | 23,333 | 1.0 | 12.1 | <5 | 15 | 0.33 (0.02 - 2.14) | 0.37 | 0.25 (0.02 - 2.86) | 0.27 |
| Meta-analysis | 87,394 | 344,081 | 53.1 | 319.1 | 32 | 254 | 0.81 (0.53 - 1.21) | 0.32 | 0.82 (0.53 - 1.26) | 0.36 |
| **PS-matching / on-treatment TAR** | | | | | | | | | | |
| OptumDOD | 4,750 | 26,038 | 2.6 | 21.8 | <5 | 35 | 0.58 (0.13 - 1.95) | 0.43 | 0.57 (0.15 - 2.26) | 0.43 |
| MDCR | 1,777 | 17,083 | 1.2 | 17.2 | 0 | 15 | | | | |
| Open Claims | 70,336 | 340,456 | 46.5 | 315.8 | 27 | 251 | 0.87 (0.53 - 1.40) | 0.59 | 0.85 (0.51 - 1.41) | 0.53 |
| OptumEHR | 3,239 | 23,174 | 0.9 | 12.1 | <5 | 14 | 0.35 (0.02 - 2.38) | 0.41 | 0.29 (0.02 - 3.29) | 0.32 |
| Meta-analysis | 75,086 | 366,494 | 49.1 | 337.6 | <32 | 286 | 0.83 (0.53 - 1.32) | 0.44 | 0.81 (0.50 - 1.30) | 0.38 |
| **PS-stratification / intent-to-treat TAR** | | | | | | | | | | |
| OptumDOD | 5,619 | 27,194 | 8.4 | 82.3 | 7 | 156 | 0.40 (0.16 - 0.83) | 0.03 | 0.40 (0.18 - 0.90) | 0.03 |
| MDCR | 1,992 | 17,928 | 4.1 | 51.3 | <5 | 56 | 0.67 (0.11 - 2.30) | 0.61 | 0.60 (0.13 - 2.82) | 0.52 |
| Open Claims | 87,394 | 344,081 | 181.2 | 1468.1 | 156 | 1,543 | 1.01 (0.84 - 1.20) | 0.93 | 1.00 (0.79 - 1.26) | 0.94 |
| OptumEHR | 3,572 | 23,333 | 7.4 | 80.4 | 5 | 59 | 0.99 (0.32 - 2.45) | 0.98 | 0.94 (0.34 - 2.60) | 0.91 |
| Meta-analysis | 90,966 | 367,414 | 188.6 | 1548.5 | 161 | 1,602 | 1.01 (0.84 - 1.20) | 0.93 | 1.00 (0.80 - 1.26) | 0.94 |
| **PS-matching / intent-to-treat TAR** | | | | | | | | | | |
| OptumDOD | 4,750 | 26,038 | 7.9 | 77.6 | 7 | 147 | 0.46 (0.19 - 0.98) | 0.07 | 0.47 (0.20 - 1.08) | 0.08 |
| MDCR | 1,777 | 17,083 | 4.0 | 49.3 | <5 | 52 | 1.44 (0.22 - 5.52) | 0.66 | 1.31 (0.26 - 6.63) | 0.74 |
| Open Claims | 70,336 | 340,456 | 166.9 | 1451.8 | 145 | 1,518 | 0.96 (0.79 - 1.17) | 0.70 | 0.94 (0.74 - 1.21) | 0.65 |
| OptumEHR | 3,239 | 23,174 | 7.0 | 79.7 | 5 | 57 | 0.58 (0.16 - 1.62) | 0.35 | 0.55 (0.17 - 1.74) | 0.31 |
| Meta-analysis | 80,102 | 406,751 | 185.8 | 1658.4 | <162 | 1,774 | 0.81 (0.52 - 1.24) | 0.33 | 0.78 (0.50 - 1.23) | 0.29 |

##### 6.2.7 Diarrhea

**Supplementary Table 132:** Exposures, outcomes and risk estimates for Diarrhea in GLP1RA (T) vs DPP4I (C) new-user cohorts under four different propensity score (PS) adjustment / time-at-risk (TAR) designs across data sources. We report total patient exposure time in 1,000 patient-year (KYr) units, uncalibrated and calibrated hazard ratios (HRs) and their 95% confidence intervals (CIs). Greyed out entries did not pass study diagnostics and are included only for completeness.

| | Patients | | Exposure (KYr) | | Outcomes | | Uncalibrated | | Calibrated | |
|---|---|---|---|---|---|---|---|---|---|---|
| | T | C | T | C | T | C | HR (95% CI) | $p$ | HR (95% CI) | $p$ |
| **PS-stratification / on-treatment TAR** | | | | | | | | | | |
| OptumDOD | 4,704 | 22,558 | 2.4 | 18.1 | 125 | 813 | 1.06 (0.85 - 1.31) | 0.58 | 1.04 (0.83 - 1.29) | 0.76 |
| MDCR | 1,736 | 15,423 | 1.1 | 15.0 | 41 | 534 | 1.02 (0.71 - 1.43) | 0.91 | 0.91 (0.64 - 1.28) | 0.59 |
| Open Claims | 72,549 | 292,867 | 43.8 | 267.0 | 965 | 5,999 | 1.11 (1.03 - 1.20) | <0.01 | 1.10 (0.93 - 1.31) | 0.31 |
| OptumEHR | 3,136 | 20,427 | 0.8 | 10.2 | 36 | 401 | 1.14 (0.78 - 1.62) | 0.49 | 0.87 (0.60 - 1.26) | 0.50 |
| Meta-analysis | 75,685 | 313,294 | 44.7 | 277.2 | 1,001 | 6,400 | 1.12 (1.03 - 1.20) | <0.01 | 1.12 (0.98 - 1.29) | 0.08 |
| **PS-matching / on-treatment TAR** | | | | | | | | | | |
| OptumDOD | 3,968 | 21,606 | 2.2 | 17.4 | 108 | 766 | 1.07 (0.81 - 1.40) | 0.62 | 1.07 (0.81 - 1.40) | 0.67 |
| MDCR | 1,556 | 14,634 | 1.0 | 14.4 | 40 | 497 | 1.36 (0.90 - 2.00) | 0.13 | 1.21 (0.81 - 1.81) | 0.37 |
| Open Claims | 58,708 | 289,666 | 38.5 | 264.1 | 824 | 5,855 | 1.12 (1.01 - 1.24) | 0.03 | 1.09 (0.92 - 1.28) | 0.36 |
| OptumEHR | 2,846 | 20,254 | 0.8 | 10.1 | 35 | 395 | 1.26 (0.83 - 1.86) | 0.26 | 1.01 (0.68 - 1.52) | 0.80 |
| Meta-analysis | 67,078 | 346,160 | 42.5 | 306.0 | 1,007 | 7,513 | 1.13 (1.04 - 1.24) | <0.01 | 1.09 (0.93 - 1.29) | 0.29 |
| **PS-stratification / intent-to-treat TAR** | | | | | | | | | | |
| OptumDOD | 4,704 | 22,558 | 6.5 | 62.5 | 331 | 2,709 | 1.08 (0.94 - 1.22) | 0.27 | 1.08 (0.95 - 1.23) | 0.31 |
| MDCR | 1,736 | 15,423 | 3.4 | 40.9 | 124 | 1,699 | 0.88 (0.72 - 1.06) | 0.18 | 0.79 (0.65 - 0.96) | 0.03 |
| Open Claims | 72,549 | 292,867 | 145.0 | 1184.2 | 3,172 | 26,856 | 1.04 (1.00 - 1.08) | 0.06 | 1.03 (0.88 - 1.20) | 0.74 |
| OptumEHR | 3,136 | 20,427 | 6.0 | 65.1 | 193 | 1,999 | 1.11 (0.95 - 1.30) | 0.18 | 1.06 (0.90 - 1.25) | 0.53 |
| Meta-analysis | 75,685 | 313,294 | 151.0 | 1249.3 | 3,365 | 28,855 | 1.04 (1.00 - 1.09) | 0.03 | 1.04 (0.89 - 1.21) | 0.63 |
| **PS-matching / intent-to-treat TAR** | | | | | | | | | | |
| OptumDOD | 3,968 | 21,606 | 6.1 | 59.0 | 300 | 2,546 | 1.06 (0.92 - 1.23) | 0.41 | 1.08 (0.93 - 1.26) | 0.38 |
| MDCR | 1,556 | 14,634 | 3.3 | 39.1 | 122 | 1,606 | 0.93 (0.75 - 1.16) | 0.55 | 0.85 (0.68 - 1.06) | 0.18 |
| Open Claims | 58,708 | 289,666 | 133.7 | 1171.0 | 2,889 | 26,354 | 1.06 (1.01 - 1.10) | 0.02 | 1.04 (0.89 - 1.21) | 0.65 |
| OptumEHR | 2,846 | 20,254 | 5.7 | 64.4 | 186 | 1,975 | 1.05 (0.88 - 1.25) | 0.57 | 1.00 (0.84 - 1.20) | 0.83 |
| Meta-analysis | 67,078 | 346,160 | 148.8 | 1333.6 | 3,497 | 32,481 | 1.05 (1.01 - 1.10) | 0.01 | 1.02 (0.89 - 1.17) | 0.76 |

##### 6.2.8 Genitourinary infection

**Supplementary Table 133:** Exposures, outcomes and risk estimates for Genitourinary infection in GLP1RA (T) vs DPP4I (C) new-user cohorts under four different propensity score (PS) adjustment / time-at-risk (TAR) designs across data sources. We report total patient exposure time in 1,000 patient-year (KYr) units, uncalibrated and calibrated hazard ratios (HRs) and their 95% confidence intervals (CIs). Greyed out entries did not pass study diagnostics and are included only for completeness.

| | Patients | | Exposure (KYr) | | Outcomes | | Uncalibrated | | Calibrated | |
|---|---|---|---|---|---|---|---|---|---|---|
| | T | C | T | C | T | C | HR (95% CI) | $p$ | HR (95% CI) | $p$ |
| **PS-stratification / on-treatment TAR** | | | | | | | | | | |
| OptumDOD | 4,094 | 19,137 | 2.0 | 15.2 | 147 | 1,272 | 0.99 (0.82 - 1.20) | 0.95 | 0.97 (0.80 - 1.18) | 0.76 |
| MDCR | 1,515 | 13,747 | 0.9 | 13.2 | 60 | 852 | 1.07 (0.79 - 1.41) | 0.66 | 0.95 (0.71 - 1.26) | 0.71 |
| Open Claims | 60,438 | 247,138 | 37.0 | 222.7 | 1,315 | 10,579 | 0.93 (0.87 - 0.99) | 0.03 | 0.92 (0.78 - 1.09) | 0.37 |
| OptumEHR | 2,901 | 18,900 | 0.8 | 9.3 | 53 | 633 | 1.18 (0.86 - 1.58) | 0.28 | 0.90 (0.67 - 1.22) | 0.55 |
| Meta-analysis | 63,339 | 266,038 | 37.7 | 232.0 | 1,368 | 11,212 | 1.00 (0.81 - 1.23) | 0.99 | 1.01 (0.79 - 1.28) | 0.91 |
| **PS-matching / on-treatment TAR** | | | | | | | | | | |
| OptumDOD | 3,448 | 18,130 | 1.8 | 14.4 | 136 | 1,174 | 1.03 (0.81 - 1.29) | 0.83 | 1.02 (0.81 - 1.29) | 0.81 |
| MDCR | 1,355 | 13,361 | 0.8 | 12.9 | 58 | 820 | 1.02 (0.72 - 1.41) | 0.93 | 0.91 (0.64 - 1.28) | 0.59 |
| Open Claims | 49,225 | 243,775 | 32.6 | 219.7 | 1,150 | 10,239 | 0.93 (0.86 - 1.01) | 0.10 | 0.91 (0.78 - 1.06) | 0.27 |
| OptumEHR | 2,645 | 18,728 | 0.7 | 9.2 | 51 | 616 | 0.98 (0.67 - 1.40) | 0.92 | 0.79 (0.54 - 1.15) | 0.27 |
| Meta-analysis | 56,673 | 293,994 | 36.0 | 256.2 | 1,395 | 12,849 | 0.95 (0.88 - 1.02) | 0.15 | 0.91 (0.78 - 1.07) | 0.26 |
| **PS-stratification / intent-to-treat TAR** | | | | | | | | | | |
| OptumDOD | 4,094 | 19,137 | 5.3 | 49.1 | 402 | 3,947 | 0.99 (0.88 - 1.11) | 0.88 | 1.00 (0.89 - 1.12) | 0.85 |
| MDCR | 1,515 | 13,747 | 2.7 | 34.8 | 190 | 2,353 | 1.10 (0.94 - 1.28) | 0.24 | 0.99 (0.85 - 1.17) | 0.83 |
| Open Claims | 60,438 | 247,138 | 118.2 | 948.7 | 4,614 | 46,257 | 0.99 (0.96 - 1.02) | 0.62 | 0.98 (0.84 - 1.15) | 0.80 |
| OptumEHR | 2,901 | 18,900 | 5.4 | 57.9 | 240 | 2,753 | 1.02 (0.88 - 1.17) | 0.81 | 0.97 (0.84 - 1.12) | 0.69 |
| Meta-analysis | 63,339 | 266,038 | 123.6 | 1006.6 | 4,854 | 49,010 | 0.99 (0.96 - 1.03) | 0.67 | 0.99 (0.85 - 1.14) | 0.85 |
| **PS-matching / intent-to-treat TAR** | | | | | | | | | | |
| OptumDOD | 3,448 | 18,130 | 5.0 | 45.8 | 376 | 3,595 | 1.05 (0.92 - 1.19) | 0.47 | 1.06 (0.93 - 1.21) | 0.43 |
| MDCR | 1,355 | 13,361 | 2.6 | 34.0 | 185 | 2,266 | 1.07 (0.90 - 1.28) | 0.43 | 0.98 (0.82 - 1.17) | 0.79 |
| Open Claims | 49,225 | 243,775 | 109.2 | 936.2 | 4,297 | 45,056 | 0.99 (0.95 - 1.02) | 0.52 | 0.97 (0.83 - 1.13) | 0.70 |
| OptumEHR | 2,645 | 18,728 | 5.1 | 57.3 | 234 | 2,700 | 1.05 (0.89 - 1.22) | 0.57 | 0.99 (0.85 - 1.17) | 0.80 |
| Meta-analysis | 56,673 | 293,994 | 121.9 | 1073.2 | 5,092 | 53,617 | 1.00 (0.96 - 1.04) | 0.94 | 0.97 (0.85 - 1.12) | 0.70 |

##### 6.2.9 Hyperkalemia

**Supplementary Table 134:** Exposures, outcomes and risk estimates for Hyperkalemia in GLP1RA (T) vs DPP4I (C) new-user cohorts under four different propensity score (PS) adjustment / time-at-risk (TAR) designs across data sources. We report total patient exposure time in 1,000 patient-year (KYr) units, uncalibrated and calibrated hazard ratios (HRs) and their 95% confidence intervals (CIs). Greyed out entries did not pass study diagnostics and are included only for completeness.

| | Patients | | Exposure (KYr) | | Outcomes | | Uncalibrated | | Calibrated | |
|---|---|---|---|---|---|---|---|---|---|---|
| | T | C | T | C | T | C | HR (95% CI) | $p$ | HR (95% CI) | $p$ |
| **PS-stratification / on-treatment TAR** | | | | | | | | | | |
| OptumDOD | 5,330 | 25,331 | 2.7 | 20.6 | 65 | 688 | 0.80 (0.60 - 1.06) | 0.13 | 0.78 (0.59 - 1.04) | 0.10 |
| MDCR | 1,951 | 17,425 | 1.2 | 17.3 | 13 | 277 | 0.69 (0.36 - 1.22) | 0.23 | 0.61 (0.33 - 1.13) | 0.12 |
| Open Claims | 84,627 | 331,478 | 51.2 | 304.5 | 294 | 3,842 | 0.65 (0.57 - 0.74) | <0.01 | 0.64 (0.52 - 0.79) | <0.01 |
| OptumEHR | 3,469 | 22,492 | 1.0 | 11.4 | 10 | 305 | 0.42 (0.20 - 0.76) | <0.01 | 0.32 (0.17 - 0.62) | <0.01 |
| Meta-analysis | 88,096 | 353,970 | 52.2 | 316.0 | 304 | 4,147 | 0.59 (0.41 - 0.85) | <0.01 | 0.59 (0.41 - 0.87) | <0.01 |
| **PS-matching / on-treatment TAR** | | | | | | | | | | |
| OptumDOD | 4,496 | 24,476 | 2.4 | 19.9 | 61 | 654 | 0.94 (0.67 - 1.31) | 0.73 | 0.94 (0.67 - 1.32) | 0.73 |
| MDCR | 1,735 | 16,684 | 1.2 | 16.7 | 12 | 248 | 0.92 (0.42 - 1.79) | 0.81 | 0.82 (0.40 - 1.68) | 0.59 |
| Open Claims | 67,983 | 327,967 | 44.9 | 301.3 | 259 | 3,685 | 0.63 (0.54 - 0.74) | <0.01 | 0.62 (0.50 - 0.76) | <0.01 |
| OptumEHR | 3,137 | 22,354 | 0.9 | 11.4 | 8 | 303 | 0.31 (0.12 - 0.68) | <0.01 | 0.25 (0.11 - 0.61) | <0.01 |
| Meta-analysis | 77,351 | 391,481 | 49.3 | 349.3 | 340 | 4,890 | 0.70 (0.50 - 0.98) | 0.04 | 0.68 (0.47 - 0.98) | 0.04 |
| **PS-stratification / intent-to-treat TAR** | | | | | | | | | | |
| OptumDOD | 5,330 | 25,331 | 7.7 | 72.7 | 201 | 2,308 | 0.94 (0.80 - 1.09) | 0.42 | 0.94 (0.80 - 1.10) | 0.49 |
| MDCR | 1,951 | 17,425 | 4.0 | 48.7 | 51 | 903 | 0.86 (0.63 - 1.15) | 0.33 | 0.78 (0.58 - 1.05) | 0.12 |
| Open Claims | 84,627 | 331,478 | 173.6 | 1382.8 | 1,319 | 17,448 | 0.83 (0.78 - 0.88) | <0.01 | 0.82 (0.70 - 0.97) | 0.02 |
| OptumEHR | 3,469 | 22,492 | 7.0 | 74.6 | 79 | 1,289 | 0.74 (0.58 - 0.94) | 0.01 | 0.70 (0.55 - 0.90) | <0.01 |
| Meta-analysis | 88,096 | 353,970 | 180.6 | 1457.4 | 1,398 | 18,737 | 0.83 (0.78 - 0.88) | <0.01 | 0.82 (0.70 - 0.96) | 0.01 |
| **PS-matching / intent-to-treat TAR** | | | | | | | | | | |
| OptumDOD | 4,496 | 24,476 | 7.2 | 69.6 | 191 | 2,170 | 0.93 (0.78 - 1.12) | 0.46 | 0.95 (0.79 - 1.14) | 0.59 |
| MDCR | 1,735 | 16,684 | 3.9 | 47.0 | 50 | 823 | 0.85 (0.60 - 1.17) | 0.33 | 0.77 (0.55 - 1.08) | 0.14 |
| Open Claims | 67,983 | 327,967 | 159.7 | 1367.7 | 1,243 | 16,945 | 0.86 (0.80 - 0.92) | <0.01 | 0.84 (0.72 - 0.99) | 0.04 |
| OptumEHR | 3,137 | 22,354 | 6.7 | 74.0 | 75 | 1,273 | 0.72 (0.55 - 0.94) | 0.02 | 0.68 (0.52 - 0.90) | <0.01 |
| Meta-analysis | 77,351 | 391,481 | 177.5 | 1558.2 | 1,559 | 21,211 | 0.86 (0.81 - 0.91) | <0.01 | 0.83 (0.72 - 0.96) | 0.01 |

##### 6.2.10 Hypoglycemia

**Supplementary Table 135:** Exposures, outcomes and risk estimates for Hypoglycemia in GLP1RA (T) vs DPP4I (C) new-user cohorts under four different propensity score (PS) adjustment / time-at-risk (TAR) designs across data sources. We report total patient exposure time in 1,000 patient-year (KYr) units, uncalibrated and calibrated hazard ratios (HRs) and their 95% confidence intervals (CIs). Greyed out entries did not pass study diagnostics and are included only for completeness.

| | Patients | | Exposure (KYr) | | Outcomes | | Uncalibrated | | Calibrated | |
|---|---|---|---|---|---|---|---|---|---|---|
| | T | C | T | C | T | C | HR (95% CI) | $p$ | HR (95% CI) | $p$ |
| **PS-stratification / on-treatment TAR** | | | | | | | | | | |
| OptumDOD | 5,527 | 26,790 | 2.9 | 22.3 | 18 | 176 | 0.74 (0.42 - 1.24) | 0.28 | 0.72 (0.42 - 1.24) | 0.25 |
| MDCR | 1,968 | 17,761 | 1.2 | 17.7 | <5 | 79 | 0.55 (0.13 - 1.61) | 0.36 | 0.49 (0.14 - 1.73) | 0.27 |
| Open Claims | 85,691 | 338,560 | 52.1 | 313.8 | 117 | 1,045 | 0.82 (0.66 - 1.02) | 0.08 | 0.81 (0.62 - 1.06) | 0.15 |
| OptumEHR | 3,524 | 23,134 | 1.0 | 12.0 | <5 | 103 | 0.27 (0.04 - 0.89) | 0.09 | 0.21 (0.05 - 0.94) | 0.04 |
| Meta-analysis | 89,215 | 361,694 | 53.1 | 325.7 | <122 | 1,148 | 0.61 (0.23 - 1.60) | 0.32 | 0.62 (0.23 - 1.63) | 0.33 |
| **PS-matching / on-treatment TAR** | | | | | | | | | | |
| OptumDOD | 4,676 | 25,682 | 2.6 | 21.4 | 16 | 170 | 0.86 (0.45 - 1.57) | 0.64 | 0.86 (0.46 - 1.61) | 0.64 |
| MDCR | 1,756 | 16,935 | 1.2 | 17.0 | <5 | 72 | 0.21 (0.01 - 1.12) | 0.19 | 0.19 (0.02 - 1.88) | 0.16 |
| Open Claims | 69,093 | 335,043 | 45.7 | 310.6 | 100 | 1,026 | 0.86 (0.66 - 1.11) | 0.26 | 0.84 (0.62 - 1.12) | 0.25 |
| OptumEHR | 3,200 | 22,978 | 0.9 | 11.9 | <5 | 101 | 0.35 (0.05 - 1.22) | 0.18 | 0.28 (0.06 - 1.33) | 0.11 |
| Meta-analysis | 78,725 | 400,638 | 50.3 | 360.9 | <126 | 1,369 | 0.83 (0.65 - 1.05) | 0.12 | 0.80 (0.61 - 1.05) | 0.11 |
| **PS-stratification / intent-to-treat TAR** | | | | | | | | | | |
| OptumDOD | 5,527 | 26,790 | 8.2 | 79.6 | 66 | 966 | 0.76 (0.58 - 0.99) | 0.05 | 0.76 (0.58 - 1.00) | 0.06 |
| MDCR | 1,968 | 17,761 | 4.0 | 50.3 | 21 | 402 | 0.69 (0.42 - 1.06) | 0.11 | 0.62 (0.39 - 0.99) | 0.05 |
| Open Claims | 85,691 | 338,560 | 176.6 | 1429.3 | 668 | 8,066 | 0.96 (0.88 - 1.05) | 0.39 | 0.95 (0.80 - 1.13) | 0.58 |
| OptumEHR | 3,524 | 23,134 | 7.2 | 78.5 | 48 | 530 | 1.28 (0.92 - 1.74) | 0.12 | 1.22 (0.89 - 1.68) | 0.23 |
| Meta-analysis | 89,215 | 361,694 | 183.8 | 1507.9 | 716 | 8,596 | 1.07 (0.81 - 1.40) | 0.64 | 1.06 (0.78 - 1.44) | 0.72 |
| **PS-matching / intent-to-treat TAR** | | | | | | | | | | |
| OptumDOD | 4,676 | 25,682 | 7.7 | 75.3 | 62 | 899 | 0.85 (0.62 - 1.14) | 0.29 | 0.86 (0.63 - 1.17) | 0.35 |
| MDCR | 1,756 | 16,935 | 3.9 | 48.3 | 21 | 377 | 0.71 (0.41 - 1.18) | 0.21 | 0.65 (0.38 - 1.10) | 0.12 |
| Open Claims | 69,093 | 335,043 | 162.8 | 1413.9 | 630 | 7,889 | 0.97 (0.89 - 1.07) | 0.56 | 0.95 (0.80 - 1.14) | 0.61 |
| OptumEHR | 3,200 | 22,978 | 6.9 | 77.9 | 47 | 523 | 1.14 (0.77 - 1.63) | 0.50 | 1.08 (0.74 - 1.57) | 0.69 |
| Meta-analysis | 78,725 | 400,638 | 181.3 | 1615.4 | 760 | 9,688 | 0.96 (0.88 - 1.05) | 0.38 | 0.93 (0.80 - 1.09) | 0.40 |

##### 6.2.11 Hypotension

**Supplementary Table 136:** Exposures, outcomes and risk estimates for Hypotension in GLP1RA (T) vs DPP4I (C) new-user cohorts under four different propensity score (PS) adjustment / time-at-risk (TAR) designs across data sources. We report total patient exposure time in 1,000 patient-year (KYr) units, uncalibrated and calibrated hazard ratios (HRs) and their 95% confidence intervals (CIs). Greyed out entries did not pass study diagnostics and are included only for completeness.

| | Patients | | Exposure (KYr) | | Outcomes | | Uncalibrated | | Calibrated | |
|---|---|---|---|---|---|---|---|---|---|---|
| | T | C | T | C | T | C | HR (95% CI) | $p$ | HR (95% CI) | $p$ |
| **PS-stratification / on-treatment TAR** | | | | | | | | | | |
| OptumDOD | 5,272 | 25,208 | 2.7 | 20.6 | 76 | 715 | 0.96 (0.73 - 1.25) | 0.77 | 0.94 (0.71 - 1.23) | 0.65 |
| MDCR | 1,902 | 16,916 | 1.2 | 16.7 | 24 | 429 | 0.97 (0.61 - 1.49) | 0.90 | 0.86 (0.55 - 1.35) | 0.53 |
| Open Claims | 82,616 | 325,192 | 50.1 | 299.6 | 539 | 4,466 | 0.95 (0.86 - 1.06) | 0.36 | 0.94 (0.78 - 1.14) | 0.56 |
| OptumEHR | 3,445 | 22,476 | 1.0 | 11.5 | 19 | 322 | 0.86 (0.51 - 1.37) | 0.56 | 0.66 (0.40 - 1.09) | 0.12 |
| Meta-analysis | 86,061 | 347,668 | 51.0 | 311.1 | 558 | 4,788 | 0.95 (0.86 - 1.05) | 0.31 | 0.96 (0.83 - 1.11) | 0.57 |
| **PS-matching / on-treatment TAR** | | | | | | | | | | |
| OptumDOD | 4,460 | 24,064 | 2.4 | 19.7 | 65 | 645 | 0.90 (0.63 - 1.25) | 0.53 | 0.89 (0.64 - 1.25) | 0.54 |
| MDCR | 1,690 | 16,146 | 1.1 | 16.1 | 23 | 384 | 1.13 (0.65 - 1.86) | 0.65 | 1.01 (0.59 - 1.71) | 0.88 |
| Open Claims | 66,534 | 321,705 | 43.9 | 296.4 | 483 | 4,297 | 1.02 (0.90 - 1.16) | 0.71 | 1.00 (0.83 - 1.20) | 0.88 |
| OptumEHR | 3,127 | 22,351 | 0.9 | 11.4 | 18 | 320 | 1.12 (0.63 - 1.86) | 0.69 | 0.90 (0.52 - 1.55) | 0.72 |
| Meta-analysis | 75,811 | 384,266 | 48.3 | 343.6 | 589 | 5,646 | 1.02 (0.91 - 1.14) | 0.76 | 0.98 (0.82 - 1.17) | 0.83 |
| **PS-stratification / intent-to-treat TAR** | | | | | | | | | | |
| OptumDOD | 5,272 | 25,208 | 7.6 | 72.3 | 233 | 2,499 | 1.03 (0.88 - 1.19) | 0.72 | 1.03 (0.89 - 1.20) | 0.72 |
| MDCR | 1,902 | 16,916 | 3.8 | 46.4 | 88 | 1,415 | 0.97 (0.77 - 1.22) | 0.81 | 0.88 (0.70 - 1.11) | 0.30 |
| Open Claims | 82,616 | 325,192 | 168.5 | 1350.7 | 2,115 | 22,163 | 1.00 (0.95 - 1.05) | 0.94 | 0.99 (0.84 - 1.16) | 0.86 |
| OptumEHR | 3,445 | 22,476 | 6.9 | 74.3 | 111 | 1,429 | 0.98 (0.79 - 1.20) | 0.84 | 0.93 (0.75 - 1.15) | 0.53 |
| Meta-analysis | 86,061 | 347,668 | 175.3 | 1425.0 | 2,226 | 23,592 | 1.00 (0.95 - 1.05) | 0.90 | 0.99 (0.85 - 1.15) | 0.88 |
| **PS-matching / intent-to-treat TAR** | | | | | | | | | | |
| OptumDOD | 4,460 | 24,064 | 7.1 | 68.1 | 213 | 2,278 | 0.97 (0.81 - 1.15) | 0.75 | 0.99 (0.83 - 1.18) | 0.84 |
| MDCR | 1,690 | 16,146 | 3.7 | 44.7 | 86 | 1,301 | 1.01 (0.77 - 1.31) | 0.93 | 0.92 (0.71 - 1.20) | 0.57 |
| Open Claims | 66,534 | 321,705 | 155.1 | 1335.9 | 1,984 | 21,538 | 1.01 (0.96 - 1.07) | 0.70 | 0.99 (0.85 - 1.16) | 0.89 |
| OptumEHR | 3,127 | 22,351 | 6.6 | 73.8 | 108 | 1,413 | 1.04 (0.82 - 1.29) | 0.76 | 0.98 (0.78 - 1.24) | 0.84 |
| Meta-analysis | 75,811 | 384,266 | 172.5 | 1522.4 | 2,391 | 26,530 | 1.01 (0.96 - 1.06) | 0.73 | 0.98 (0.85 - 1.13) | 0.77 |

##### 6.2.12 Joint pain

**Supplementary Table 137:** Exposures, outcomes and risk estimates for Joint pain in GLP1RA (T) vs DPP4I (C) new-user cohorts under four different propensity score (PS) adjustment / time-at-risk (TAR) designs across data sources. We report total patient exposure time in 1,000 patient-year (KYr) units, uncalibrated and calibrated hazard ratios (HRs) and their 95% confidence intervals (CIs). Greyed out entries did not pass study diagnostics and are included only for completeness.

| | Patients | | Exposure (KYr) | | Outcomes | | Uncalibrated | | Calibrated | |
|---|---|---|---|---|---|---|---|---|---|---|
| | T | C | T | C | T | C | HR (95% CI) | $p$ | HR (95% CI) | $p$ |
| **PS-stratification / on-treatment TAR** | | | | | | | | | | |
| OptumDOD | 5,149 | 25,530 | 2.6 | 21.0 | 48 | 336 | 1.04 (0.73 - 1.45) | 0.82 | 1.02 (0.72 - 1.43) | 0.89 |
| MDCR | 1,869 | 17,206 | 1.2 | 17.1 | 8 | 187 | 0.45 (0.20 - 0.89) | 0.04 | 0.40 (0.19 - 0.85) | 0.02 |
| Open Claims | 79,565 | 322,730 | 48.6 | 298.3 | 314 | 2,141 | 0.87 (0.76 - 1.00) | 0.05 | 0.86 (0.70 - 1.06) | 0.19 |
| OptumEHR | 3,316 | 22,151 | 0.9 | 11.4 | 11 | 123 | 1.33 (0.66 - 2.43) | 0.40 | 1.02 (0.53 - 1.96) | 0.89 |
| Meta-analysis | 82,881 | 344,881 | 49.5 | 309.7 | 325 | 2,264 | 0.95 (0.68 - 1.31) | 0.74 | 0.95 (0.68 - 1.35) | 0.79 |
| **PS-matching / on-treatment TAR** | | | | | | | | | | |
| OptumDOD | 4,383 | 24,474 | 2.4 | 20.2 | 40 | 318 | 0.92 (0.58 - 1.42) | 0.72 | 0.92 (0.59 - 1.44) | 0.72 |
| MDCR | 1,678 | 16,305 | 1.1 | 16.3 | 6 | 183 | 0.45 (0.15 - 1.07) | 0.11 | 0.40 (0.15 - 1.07) | 0.07 |
| Open Claims | 64,635 | 319,079 | 42.8 | 295.0 | 271 | 2,111 | 0.82 (0.69 - 0.98) | 0.03 | 0.80 (0.65 - 1.00) | 0.06 |
| OptumEHR | 3,036 | 22,005 | 0.9 | 11.3 | 11 | 121 | 1.27 (0.50 - 2.90) | 0.59 | 1.02 (0.42 - 2.47) | 0.90 |
| Meta-analysis | 73,732 | 381,863 | 47.2 | 342.9 | 328 | 2,733 | 0.83 (0.71 - 0.98) | 0.02 | 0.81 (0.65 - 0.99) | 0.04 |
| **PS-stratification / intent-to-treat TAR** | | | | | | | | | | |
| OptumDOD | 5,149 | 25,530 | 7.5 | 75.2 | 146 | 1,093 | 1.11 (0.91 - 1.35) | 0.28 | 1.12 (0.92 - 1.36) | 0.29 |
| MDCR | 1,869 | 17,206 | 3.8 | 48.3 | 47 | 526 | 0.99 (0.71 - 1.35) | 0.96 | 0.90 (0.65 - 1.24) | 0.52 |
| Open Claims | 79,565 | 322,730 | 164.3 | 1358.0 | 1,178 | 9,467 | 1.01 (0.94 - 1.08) | 0.79 | 1.00 (0.84 - 1.18) | 0.91 |
| OptumEHR | 3,316 | 22,151 | 6.7 | 74.5 | 56 | 696 | 0.86 (0.64 - 1.14) | 0.32 | 0.82 (0.61 - 1.10) | 0.20 |
| Meta-analysis | 82,881 | 344,881 | 171.0 | 1432.5 | 1,234 | 10,163 | 1.00 (0.92 - 1.08) | 0.95 | 0.99 (0.84 - 1.17) | 0.89 |
| **PS-matching / intent-to-treat TAR** | | | | | | | | | | |
| OptumDOD | 4,383 | 24,474 | 7.1 | 71.1 | 130 | 1,041 | 1.04 (0.83 - 1.31) | 0.71 | 1.06 (0.84 - 1.33) | 0.65 |
| MDCR | 1,678 | 16,305 | 3.7 | 46.1 | 45 | 504 | 0.86 (0.58 - 1.24) | 0.43 | 0.78 (0.53 - 1.14) | 0.22 |
| Open Claims | 64,635 | 319,079 | 152.0 | 1342.0 | 1,080 | 9,319 | 1.03 (0.95 - 1.10) | 0.50 | 1.01 (0.85 - 1.19) | 0.90 |
| OptumEHR | 3,036 | 22,005 | 6.5 | 73.9 | 53 | 689 | 0.82 (0.59 - 1.11) | 0.21 | 0.77 (0.56 - 1.07) | 0.13 |
| Meta-analysis | 73,732 | 381,863 | 169.2 | 1533.1 | 1,308 | 11,553 | 1.01 (0.94 - 1.08) | 0.75 | 0.98 (0.85 - 1.14) | 0.81 |

##### 6.2.13 Lower extremity amputation

**Supplementary Table 138:** Exposures, outcomes and risk estimates for Lower extremity amputation in GLP1RA (T) vs DPP4I (C) new-user cohorts under four different propensity score (PS) adjustment / time-at-risk (TAR) designs across data sources. We report total patient exposure time in 1,000 patient-year (KYr) units, uncalibrated and calibrated hazard ratios (HRs) and their 95% confidence intervals (CIs). Greyed out entries did not pass study diagnostics and are included only for completeness.

| | Patients | | Exposure (KYr) | | Outcomes | | Uncalibrated | | Calibrated | |
|---|---|---|---|---|---|---|---|---|---|---|
| | T | C | T | C | T | C | HR (95% CI) | $p$ | HR (95% CI) | $p$ |
| **PS-stratification / on-treatment TAR** | | | | | | | | | | |
| OptumDOD | 5,613 | 27,172 | 2.9 | 22.7 | <5 | 17 | 0.93 (0.13 - 3.92) | 0.93 | 0.90 (0.16 - 4.95) | 0.91 |
| MDCR | 1,991 | 17,911 | 1.3 | 17.9 | 0 | 16 | | | | |
| Open Claims | 87,292 | 343,891 | 53.0 | 319.1 | 16 | 165 | 0.84 (0.46 - 1.43) | 0.55 | 0.83 (0.46 - 1.49) | 0.54 |
| OptumEHR | 3,570 | 23,317 | 1.0 | 12.1 | 0 | 9 | | | | |
| Meta-analysis | 87,292 | 343,891 | 53.0 | 319.1 | 16 | 165 | 0.84 (0.46 - 1.43) | 0.55 | 0.85 (0.48 - 1.51) | 0.58 |
| **PS-matching / on-treatment TAR** | | | | | | | | | | |
| OptumDOD | 4,744 | 26,015 | 2.6 | 21.8 | <5 | 17 | | | | |
| MDCR | 1,776 | 17,071 | 1.2 | 17.2 | 0 | 12 | | | | |
| Open Claims | 70,275 | 340,250 | 46.5 | 315.7 | 16 | 157 | 0.91 (0.46 - 1.69) | 0.77 | 0.88 (0.45 - 1.72) | 0.71 |
| OptumEHR | 3,238 | 23,158 | 0.9 | 12.1 | 0 | 9 | | | | |
| Meta-analysis | 70,275 | 340,250 | 46.5 | 315.7 | 16 | 157 | 0.91 (0.46 - 1.69) | 0.77 | 0.88 (0.45 - 1.71) | 0.70 |
| **PS-stratification / intent-to-treat TAR** | | | | | | | | | | |
| OptumDOD | 5,613 | 27,172 | 8.4 | 82.3 | 14 | 113 | 1.49 (0.77 - 2.71) | 0.21 | 1.50 (0.80 - 2.82) | 0.21 |
| MDCR | 1,991 | 17,911 | 4.1 | 51.2 | <5 | 64 | 1.32 (0.38 - 3.51) | 0.62 | 1.20 (0.39 - 3.66) | 0.75 |
| Open Claims | 87,292 | 343,891 | 181.1 | 1468.3 | 97 | 922 | 1.09 (0.86 - 1.36) | 0.48 | 1.07 (0.81 - 1.42) | 0.62 |
| OptumEHR | 3,570 | 23,317 | 7.4 | 80.4 | <5 | 51 | 0.78 (0.22 - 2.17) | 0.68 | 0.74 (0.23 - 2.36) | 0.62 |
| Meta-analysis | 90,862 | 367,208 | 188.4 | 1548.7 | <102 | 973 | 1.07 (0.86 - 1.34) | 0.55 | 1.06 (0.81 - 1.39) | 0.65 |
| **PS-matching / intent-to-treat TAR** | | | | | | | | | | |
| OptumDOD | 4,744 | 26,015 | 7.9 | 77.7 | 14 | 108 | 1.39 (0.60 - 2.93) | 0.41 | 1.41 (0.64 - 3.14) | 0.40 |
| MDCR | 1,776 | 17,071 | 4.0 | 49.2 | <5 | 58 | 1.58 (0.43 - 4.56) | 0.45 | 1.44 (0.44 - 4.70) | 0.55 |
| Open Claims | 70,275 | 340,250 | 166.8 | 1452.0 | 93 | 891 | 1.08 (0.83 - 1.38) | 0.56 | 1.06 (0.79 - 1.41) | 0.71 |
| OptumEHR | 3,238 | 23,158 | 7.0 | 79.7 | <5 | 51 | 0.85 (0.19 - 2.66) | 0.81 | 0.81 (0.22 - 3.03) | 0.76 |
| Meta-analysis | 80,033 | 406,494 | 185.7 | 1658.4 | <117 | 1,108 | 1.11 (0.88 - 1.40) | 0.38 | 1.08 (0.82 - 1.41) | 0.59 |

##### 6.2.14 Nausea

**Supplementary Table 139:** Exposures, outcomes and risk estimates for Nausea in GLP1RA (T) vs DPP4I (C) new-user cohorts under four different propensity score (PS) adjustment / time-at-risk (TAR) designs across data sources. We report total patient exposure time in 1,000 patient-year (KYr) units, uncalibrated and calibrated hazard ratios (HRs) and their 95% confidence intervals (CIs). Greyed out entries did not pass study diagnostics and are included only for completeness.

| | Patients | | Exposure (KYr) | | Outcomes | | Uncalibrated | | Calibrated | |
|---|---|---|---|---|---|---|---|---|---|---|
| | T | C | T | C | T | C | HR (95% CI) | $p$ | HR (95% CI) | $p$ |
| **PS-stratification / on-treatment TAR** | | | | | | | | | | |
| OptumDOD | 4,695 | 23,103 | 2.4 | 18.8 | 178 | 854 | 1.47 (1.21 - 1.77) | <0.01 | 1.43 (1.18 - 1.74) | <0.01 |
| MDCR | 1,799 | 15,971 | 1.1 | 15.8 | 44 | 541 | 1.01 (0.71 - 1.41) | 0.94 | 0.90 (0.64 - 1.27) | 0.56 |
| Open Claims | 72,632 | 297,396 | 44.1 | 273.9 | 1,274 | 6,036 | 1.41 (1.31 - 1.51) | <0.01 | 1.39 (1.17 - 1.66) | <0.01 |
| OptumEHR | 3,189 | 21,212 | 0.9 | 10.8 | 61 | 372 | 2.11 (1.54 - 2.83) | <0.01 | 1.61 (1.19 - 2.20) | <0.01 |
| Meta-analysis | 75,821 | 318,608 | 44.9 | 284.7 | 1,335 | 6,408 | 1.67 (1.13 - 2.47) | <0.01 | 1.69 (1.13 - 2.53) | 0.01 |
| **PS-matching / on-treatment TAR** | | | | | | | | | | |
| OptumDOD | 3,990 | 21,854 | 2.2 | 17.8 | 156 | 805 | 1.76 (1.38 - 2.22) | <0.01 | 1.75 (1.38 - 2.23) | <0.01 |
| MDCR | 1,612 | 15,197 | 1.0 | 15.1 | 40 | 494 | 1.05 (0.70 - 1.53) | 0.81 | 0.94 (0.63 - 1.39) | 0.74 |
| Open Claims | 58,946 | 293,827 | 38.8 | 270.6 | 1,093 | 5,920 | 1.51 (1.38 - 1.64) | <0.01 | 1.46 (1.25 - 1.72) | <0.01 |
| OptumEHR | 2,914 | 21,085 | 0.8 | 10.7 | 53 | 365 | 2.19 (1.49 - 3.17) | <0.01 | 1.77 (1.20 - 2.59) | <0.01 |
| Meta-analysis | 67,462 | 351,963 | 42.8 | 314.2 | 1,342 | 7,584 | 1.58 (1.24 - 2.00) | <0.01 | 1.52 (1.16 - 2.01) | <0.01 |
| **PS-stratification / intent-to-treat TAR** | | | | | | | | | | |
| OptumDOD | 4,695 | 23,103 | 6.4 | 64.7 | 406 | 2,822 | 1.28 (1.14 - 1.44) | <0.01 | 1.29 (1.14 - 1.45) | <0.01 |
| MDCR | 1,799 | 15,971 | 3.5 | 42.8 | 147 | 1,701 | 1.08 (0.90 - 1.29) | 0.41 | 0.98 (0.81 - 1.17) | 0.80 |
| Open Claims | 72,632 | 297,396 | 145.7 | 1210.0 | 3,699 | 28,481 | 1.13 (1.09 - 1.18) | <0.01 | 1.12 (0.96 - 1.31) | 0.17 |
| OptumEHR | 3,189 | 21,212 | 6.2 | 68.9 | 200 | 1,780 | 1.26 (1.07 - 1.47) | <0.01 | 1.19 (1.01 - 1.41) | 0.05 |
| Meta-analysis | 75,821 | 318,608 | 151.9 | 1278.9 | 3,899 | 30,261 | 1.16 (1.07 - 1.25) | <0.01 | 1.15 (0.97 - 1.36) | 0.10 |
| **PS-matching / intent-to-treat TAR** | | | | | | | | | | |
| OptumDOD | 3,990 | 21,854 | 6.0 | 60.1 | 370 | 2,623 | 1.35 (1.17 - 1.54) | <0.01 | 1.37 (1.19 - 1.57) | <0.01 |
| MDCR | 1,612 | 15,197 | 3.4 | 41.1 | 139 | 1,586 | 1.01 (0.81 - 1.25) | 0.93 | 0.92 (0.74 - 1.14) | 0.48 |
| Open Claims | 58,946 | 293,827 | 134.6 | 1194.8 | 3,356 | 27,929 | 1.16 (1.12 - 1.21) | <0.01 | 1.14 (0.98 - 1.33) | 0.10 |
| OptumEHR | 2,914 | 21,085 | 5.9 | 68.4 | 187 | 1,759 | 1.22 (1.01 - 1.45) | 0.03 | 1.16 (0.96 - 1.39) | 0.14 |
| Meta-analysis | 67,462 | 351,963 | 150.0 | 1364.4 | 4,052 | 33,897 | 1.19 (1.09 - 1.30) | <0.01 | 1.16 (0.99 - 1.36) | 0.07 |

##### 6.2.15 Peripheral edema

**Supplementary Table 140:** Exposures, outcomes and risk estimates for Peripheral edema in GLP1RA (T) vs DPP4I (C) new-user cohorts under four different propensity score (PS) adjustment / time-at-risk (TAR) designs across data sources. We report total patient exposure time in 1,000 patient-year (KYr) units, uncalibrated and calibrated hazard ratios (HRs) and their 95% confidence intervals (CIs). Greyed out entries did not pass study diagnostics and are included only for completeness.

| | Patients | | Exposure (KYr) | | Outcomes | | Uncalibrated | | Calibrated | |
|---|---|---|---|---|---|---|---|---|---|---|
| | T | C | T | C | T | C | HR (95% CI) | $p$ | HR (95% CI) | $p$ |
| **PS-stratification / on-treatment TAR** | | | | | | | | | | |
| OptumDOD | 3,920 | 19,394 | 2.0 | 15.0 | 136 | 1,589 | 0.69 (0.56 - 0.83) | <0.01 | 0.67 (0.55 - 0.82) | <0.01 |
| MDCR | 1,471 | 13,304 | 0.9 | 12.6 | 57 | 1,091 | 0.93 (0.69 - 1.22) | 0.61 | 0.82 (0.62 - 1.10) | 0.20 |
| Open Claims | 58,785 | 250,135 | 35.7 | 224.6 | 1,044 | 10,723 | 0.80 (0.74 - 0.85) | <0.01 | 0.79 (0.66 - 0.93) | 0.01 |
| OptumEHR | 2,721 | 18,902 | 0.7 | 9.2 | 37 | 603 | 0.94 (0.65 - 1.31) | 0.72 | 0.72 (0.51 - 1.02) | 0.08 |
| Meta-analysis | 61,506 | 269,037 | 36.4 | 233.7 | 1,081 | 11,326 | 0.80 (0.75 - 0.86) | <0.01 | 0.81 (0.71 - 0.92) | <0.01 |
| **PS-matching / on-treatment TAR** | | | | | | | | | | |
| OptumDOD | 3,331 | 18,604 | 1.8 | 14.4 | 117 | 1,487 | 0.60 (0.47 - 0.77) | <0.01 | 0.60 (0.47 - 0.77) | <0.01 |
| MDCR | 1,315 | 12,867 | 0.8 | 12.3 | 53 | 1,020 | 0.71 (0.49 - 1.00) | 0.06 | 0.63 (0.44 - 0.91) | 0.02 |
| Open Claims | 48,713 | 246,417 | 31.9 | 221.2 | 932 | 10,329 | 0.80 (0.73 - 0.87) | <0.01 | 0.78 (0.66 - 0.91) | <0.01 |
| OptumEHR | 2,522 | 18,727 | 0.7 | 9.1 | 34 | 590 | 0.91 (0.58 - 1.38) | 0.66 | 0.73 (0.47 - 1.14) | 0.20 |
| Meta-analysis | 55,881 | 296,615 | 35.2 | 257.0 | 1,136 | 13,426 | 0.74 (0.63 - 0.87) | <0.01 | 0.72 (0.58 - 0.89) | <0.01 |
| **PS-stratification / intent-to-treat TAR** | | | | | | | | | | |
| OptumDOD | 3,920 | 19,394 | 5.2 | 48.4 | 428 | 4,663 | 0.87 (0.78 - 0.97) | 0.01 | 0.87 (0.78 - 0.98) | 0.03 |
| MDCR | 1,471 | 13,304 | 2.6 | 32.4 | 229 | 3,001 | 1.08 (0.93 - 1.24) | 0.30 | 0.98 (0.84 - 1.13) | 0.75 |
| Open Claims | 58,785 | 250,135 | 115.7 | 965.9 | 4,173 | 45,288 | 0.96 (0.93 - 0.99) | 0.02 | 0.95 (0.81 - 1.11) | 0.51 |
| OptumEHR | 2,721 | 18,902 | 5.2 | 57.6 | 227 | 2,997 | 0.99 (0.85 - 1.14) | 0.85 | 0.94 (0.81 - 1.09) | 0.44 |
| Meta-analysis | 61,506 | 269,037 | 120.9 | 1023.5 | 4,400 | 48,285 | 0.96 (0.93 - 0.99) | 0.02 | 0.95 (0.82 - 1.11) | 0.54 |
| **PS-matching / intent-to-treat TAR** | | | | | | | | | | |
| OptumDOD | 3,331 | 18,604 | 4.9 | 46.0 | 393 | 4,346 | 0.81 (0.71 - 0.91) | <0.01 | 0.82 (0.72 - 0.93) | <0.01 |
| MDCR | 1,315 | 12,867 | 2.6 | 31.7 | 223 | 2,855 | 1.04 (0.88 - 1.23) | 0.64 | 0.95 (0.80 - 1.12) | 0.57 |
| Open Claims | 48,713 | 246,417 | 107.9 | 951.3 | 3,921 | 43,966 | 0.97 (0.93 - 1.00) | 0.07 | 0.95 (0.82 - 1.10) | 0.50 |
| OptumEHR | 2,522 | 18,727 | 5.0 | 57.0 | 220 | 2,940 | 0.97 (0.82 - 1.13) | 0.69 | 0.92 (0.78 - 1.08) | 0.35 |
| Meta-analysis | 55,881 | 296,615 | 120.3 | 1085.9 | 4,757 | 54,107 | 0.94 (0.85 - 1.04) | 0.21 | 0.91 (0.77 - 1.08) | 0.27 |

##### 6.2.16 Photosensitivity

**Supplementary Table 141:** Exposures, outcomes and risk estimates for Photosensitivity in GLP1RA (T) vs DPP4I (C) new-user cohorts under four different propensity score (PS) adjustment / time-at-risk (TAR) designs across data sources. We report total patient exposure time in 1,000 patient-year (KYr) units, uncalibrated and calibrated hazard ratios (HRs) and their 95% confidence intervals (CIs). Greyed out entries did not pass study diagnostics and are included only for completeness.

| | Patients | | Exposure (KYr) | | Outcomes | | Uncalibrated | | Calibrated | |
|---|---|---|---|---|---|---|---|---|---|---|
| | T | C | T | C | T | C | HR (95% CI) | $p$ | HR (95% CI) | $p$ |
| **PS-stratification / on-treatment TAR** | | | | | | | | | | |
| OptumDOD | 5,575 | 26,866 | 2.9 | 22.3 | <5 | 39 | 1.44 (0.31 - 4.68) | 0.60 | 1.41 (0.36 - 5.50) | 0.62 |
| MDCR | 1,951 | 17,593 | 1.2 | 17.5 | <5 | 71 | 0.75 (0.12 - 2.47) | 0.71 | 0.67 (0.15 - 3.01) | 0.60 |
| Open Claims | 85,977 | 339,304 | 52.1 | 314.4 | 21 | 193 | 1.84 (1.06 - 3.01) | 0.02 | 1.82 (1.05 - 3.13) | 0.03 |
| OptumEHR | 3,534 | 23,120 | 1.0 | 12.0 | <5 | 22 | 2.83 (0.75 - 8.20) | 0.09 | 2.17 (0.66 - 7.16) | 0.21 |
| Meta-analysis | 85,977 | 339,304 | 52.1 | 314.4 | 21 | 193 | 1.84 (1.06 - 3.01) | 0.02 | 1.85 (1.09 - 3.16) | 0.02 |
| **PS-matching / on-treatment TAR** | | | | | | | | | | |
| OptumDOD | 4,715 | 25,730 | 2.6 | 21.4 | <5 | 38 | 1.52 (0.22 - 6.54) | 0.63 | 1.51 (0.28 - 8.31) | 0.64 |
| MDCR | 1,735 | 16,778 | 1.2 | 16.8 | <5 | 66 | 1.03 (0.16 - 3.57) | 0.97 | 0.92 (0.20 - 4.30) | 0.91 |
| Open Claims | 69,216 | 335,699 | 45.7 | 311.1 | 18 | 190 | 2.35 (1.27 - 4.10) | <0.01 | 2.29 (1.26 - 4.16) | <0.01 |
| OptumEHR | 3,214 | 22,963 | 0.9 | 11.9 | <5 | 21 | 3.96 (0.93 - 14.3) | 0.05 | 3.20 (0.82 - 12.5) | 0.10 |
| Meta-analysis | 75,666 | 378,207 | 49.4 | 349.4 | <28 | 294 | 2.05 (1.22 - 3.46) | <0.01 | 1.98 (1.16 - 3.40) | 0.01 |
| **PS-stratification / intent-to-treat TAR** | | | | | | | | | | |
| OptumDOD | 5,575 | 26,866 | 8.3 | 80.8 | 10 | 99 | 1.64 (0.75 - 3.23) | 0.18 | 1.65 (0.79 - 3.42) | 0.18 |
| MDCR | 1,951 | 17,593 | 4.0 | 50.0 | 6 | 155 | 0.63 (0.24 - 1.34) | 0.29 | 0.57 (0.24 - 1.34) | 0.20 |
| Open Claims | 85,977 | 339,304 | 178.2 | 1446.9 | 43 | 504 | 1.17 (0.82 - 1.63) | 0.37 | 1.16 (0.79 - 1.68) | 0.45 |
| OptumEHR | 3,534 | 23,120 | 7.3 | 79.4 | 12 | 117 | 1.32 (0.67 - 2.37) | 0.39 | 1.25 (0.67 - 2.36) | 0.48 |
| Meta-analysis | 89,511 | 362,424 | 185.4 | 1526.4 | 55 | 621 | 1.20 (0.89 - 1.63) | 0.23 | 1.19 (0.85 - 1.67) | 0.30 |
| **PS-matching / intent-to-treat TAR** | | | | | | | | | | |
| OptumDOD | 4,715 | 25,730 | 7.8 | 76.3 | 9 | 96 | 1.63 (0.69 - 3.38) | 0.23 | 1.65 (0.74 - 3.66) | 0.22 |
| MDCR | 1,735 | 16,778 | 3.9 | 48.0 | 6 | 146 | 0.61 (0.23 - 1.37) | 0.28 | 0.56 (0.23 - 1.36) | 0.20 |
| Open Claims | 69,216 | 335,699 | 164.1 | 1430.7 | 39 | 497 | 1.14 (0.79 - 1.59) | 0.48 | 1.11 (0.76 - 1.63) | 0.58 |
| OptumEHR | 3,214 | 22,963 | 7.0 | 78.7 | 12 | 116 | 1.66 (0.81 - 3.11) | 0.14 | 1.57 (0.80 - 3.10) | 0.19 |
| Meta-analysis | 78,880 | 401,170 | 182.8 | 1633.8 | 66 | 855 | 1.19 (0.90 - 1.57) | 0.22 | 1.16 (0.85 - 1.57) | 0.35 |

##### 6.2.17 Venous thromboembolism

**Supplementary Table 142:** Exposures, outcomes and risk estimates for Venous thromboembolism in GLP1RA (T) vs DPP4I (C) new-user cohorts under four different propensity score (PS) adjustment / time-at-risk (TAR) designs across data sources. We report total patient exposure time in 1,000 patient-year (KYr) units, uncalibrated and calibrated hazard ratios (HRs) and their 95% confidence intervals (CIs). Greyed out entries did not pass study diagnostics and are included only for completeness.

| | Patients | | Exposure (KYr) | | Outcomes | | Uncalibrated | | Calibrated | |
|---|---|---|---|---|---|---|---|---|---|---|
| | T | C | T | C | T | C | HR (95% CI) | $p$ | HR (95% CI) | $p$ |
| **PS-stratification / on-treatment TAR** | | | | | | | | | | |
| OptumDOD | 5,345 | 26,088 | 2.8 | 21.6 | 30 | 313 | 0.76 (0.49 - 1.15) | 0.22 | 0.74 (0.48 - 1.15) | 0.19 |
| MDCR | 1,904 | 17,192 | 1.2 | 17.1 | 14 | 229 | 1.02 (0.54 - 1.79) | 0.94 | 0.91 (0.50 - 1.65) | 0.76 |
| Open Claims | 82,626 | 328,935 | 50.3 | 304.4 | 269 | 2,221 | 0.94 (0.82 - 1.09) | 0.42 | 0.93 (0.75 - 1.15) | 0.54 |
| OptumEHR | 3,430 | 22,530 | 1.0 | 11.6 | 13 | 116 | 1.71 (0.88 - 3.08) | 0.09 | 1.31 (0.70 - 2.46) | 0.41 |
| Meta-analysis | 86,056 | 351,465 | 51.3 | 316.0 | 282 | 2,337 | 1.17 (0.67 - 2.06) | 0.58 | 1.18 (0.67 - 2.10) | 0.57 |
| **PS-matching / on-treatment TAR** | | | | | | | | | | |
| OptumDOD | 4,544 | 24,889 | 2.5 | 20.6 | 27 | 290 | 0.87 (0.51 - 1.41) | 0.59 | 0.87 (0.52 - 1.44) | 0.59 |
| MDCR | 1,697 | 16,337 | 1.1 | 16.4 | 13 | 210 | 0.92 (0.45 - 1.74) | 0.80 | 0.82 (0.41 - 1.62) | 0.57 |
| Open Claims | 66,724 | 325,663 | 44.2 | 301.5 | 242 | 2,152 | 0.99 (0.83 - 1.18) | 0.94 | 0.97 (0.78 - 1.20) | 0.77 |
| OptumEHR | 3,126 | 22,391 | 0.9 | 11.5 | 12 | 112 | 1.92 (0.89 - 3.84) | 0.08 | 1.55 (0.74 - 3.23) | 0.26 |
| Meta-analysis | 76,091 | 389,280 | 48.7 | 350.0 | 294 | 2,764 | 1.01 (0.86 - 1.18) | 0.93 | 0.97 (0.79 - 1.20) | 0.79 |
| **PS-stratification / intent-to-treat TAR** | | | | | | | | | | |
| OptumDOD | 5,345 | 26,088 | 7.9 | 77.4 | 97 | 1,102 | 0.94 (0.75 - 1.18) | 0.62 | 0.95 (0.75 - 1.19) | 0.66 |
| MDCR | 1,904 | 17,192 | 3.8 | 48.1 | 58 | 758 | 1.06 (0.78 - 1.39) | 0.71 | 0.95 (0.71 - 1.27) | 0.76 |
| Open Claims | 82,626 | 328,935 | 170.2 | 1387.0 | 1,192 | 11,434 | 1.04 (0.98 - 1.11) | 0.22 | 1.03 (0.87 - 1.22) | 0.73 |
| OptumEHR | 3,430 | 22,530 | 7.0 | 76.3 | 58 | 642 | 1.08 (0.80 - 1.43) | 0.60 | 1.03 (0.77 - 1.37) | 0.85 |
| Meta-analysis | 86,056 | 351,465 | 177.2 | 1463.3 | 1,250 | 12,076 | 1.04 (0.98 - 1.11) | 0.19 | 1.04 (0.88 - 1.22) | 0.65 |
| **PS-matching / intent-to-treat TAR** | | | | | | | | | | |
| OptumDOD | 4,544 | 24,889 | 7.4 | 72.6 | 91 | 1,022 | 0.99 (0.76 - 1.27) | 0.95 | 1.01 (0.78 - 1.30) | 0.91 |
| MDCR | 1,697 | 16,337 | 3.7 | 46.2 | 57 | 702 | 0.97 (0.68 - 1.36) | 0.88 | 0.89 (0.63 - 1.25) | 0.51 |
| Open Claims | 66,724 | 325,663 | 156.9 | 1372.8 | 1,113 | 11,172 | 1.04 (0.97 - 1.12) | 0.25 | 1.02 (0.87 - 1.20) | 0.78 |
| OptumEHR | 3,126 | 22,391 | 6.7 | 75.7 | 57 | 630 | 1.16 (0.83 - 1.59) | 0.36 | 1.11 (0.80 - 1.53) | 0.56 |
| Meta-analysis | 76,091 | 389,280 | 174.7 | 1567.3 | 1,318 | 13,526 | 1.04 (0.97 - 1.11) | 0.23 | 1.01 (0.87 - 1.17) | 0.87 |

##### 6.2.18 Vomiting

**Supplementary Table 143:** Exposures, outcomes and risk estimates for Vomiting in GLP1RA (T) vs DPP4I (C) new-user cohorts under four different propensity score (PS) adjustment / time-at-risk (TAR) designs across data sources. We report total patient exposure time in 1,000 patient-year (KYr) units, uncalibrated and calibrated hazard ratios (HRs) and their 95% confidence intervals (CIs). Greyed out entries did not pass study diagnostics and are included only for completeness.

| | Patients | | Exposure (KYr) | | Outcomes | | Uncalibrated | | Calibrated | |
|---|---|---|---|---|---|---|---|---|---|---|
| | T | C | T | C | T | C | HR (95% CI) | $p$ | HR (95% CI) | $p$ |
| **PS-stratification / on-treatment TAR** | | | | | | | | | | |
| OptumDOD | 5,030 | 24,206 | 2.6 | 19.8 | 130 | 662 | 1.63 (1.30 - 2.03) | <0.01 | 1.59 (1.27 - 1.99) | <0.01 |
| MDCR | 1,861 | 16,422 | 1.2 | 16.2 | 35 | 436 | 1.06 (0.72 - 1.54) | 0.75 | 0.94 (0.64 - 1.39) | 0.77 |
| Open Claims | 77,029 | 308,143 | 46.7 | 284.4 | 933 | 4,902 | 1.39 (1.28 - 1.51) | <0.01 | 1.38 (1.15 - 1.64) | <0.01 |
| OptumEHR | 3,338 | 22,042 | 0.9 | 11.3 | 35 | 266 | 2.05 (1.37 - 2.99) | <0.01 | 1.57 (1.06 - 2.33) | 0.03 |
| Meta-analysis | 80,367 | 330,185 | 47.6 | 295.7 | 968 | 5,168 | 1.61 (1.11 - 2.33) | 0.01 | 1.62 (1.10 - 2.39) | 0.01 |
| **PS-matching / on-treatment TAR** | | | | | | | | | | |
| OptumDOD | 4,252 | 23,228 | 2.3 | 19.1 | 117 | 631 | 1.63 (1.23 - 2.13) | <0.01 | 1.62 (1.23 - 2.14) | <0.01 |
| MDCR | 1,660 | 15,632 | 1.1 | 15.5 | 32 | 392 | 1.12 (0.72 - 1.70) | 0.61 | 1.00 (0.65 - 1.54) | 0.86 |
| Open Claims | 62,159 | 304,538 | 41.0 | 281.1 | 801 | 4,775 | 1.44 (1.30 - 1.59) | <0.01 | 1.40 (1.18 - 1.65) | <0.01 |
| OptumEHR | 3,040 | 21,828 | 0.9 | 11.2 | 30 | 261 | 2.31 (1.39 - 3.71) | <0.01 | 1.86 (1.13 - 3.06) | 0.02 |
| Meta-analysis | 71,111 | 365,226 | 45.2 | 326.9 | 980 | 6,059 | 1.48 (1.31 - 1.67) | <0.01 | 1.43 (1.19 - 1.72) | <0.01 |
| **PS-stratification / intent-to-treat TAR** | | | | | | | | | | |
| OptumDOD | 5,030 | 24,206 | 7.0 | 68.8 | 302 | 2,324 | 1.26 (1.10 - 1.44) | <0.01 | 1.26 (1.10 - 1.45) | <0.01 |
| MDCR | 1,861 | 16,422 | 3.6 | 44.6 | 116 | 1,411 | 1.03 (0.83 - 1.25) | 0.80 | 0.93 (0.75 - 1.14) | 0.51 |
| Open Claims | 77,029 | 308,143 | 155.8 | 1267.6 | 2,788 | 23,815 | 1.09 (1.05 - 1.14) | <0.01 | 1.08 (0.92 - 1.27) | 0.36 |
| OptumEHR | 3,338 | 22,042 | 6.6 | 72.9 | 126 | 1,329 | 1.18 (0.96 - 1.43) | 0.11 | 1.12 (0.92 - 1.37) | 0.30 |
| Meta-analysis | 80,367 | 330,185 | 162.4 | 1340.5 | 2,914 | 25,144 | 1.10 (1.05 - 1.14) | <0.01 | 1.09 (0.94 - 1.27) | 0.27 |
| **PS-matching / intent-to-treat TAR** | | | | | | | | | | |
| OptumDOD | 4,252 | 23,228 | 6.6 | 65.3 | 277 | 2,182 | 1.26 (1.08 - 1.48) | <0.01 | 1.28 (1.09 - 1.50) | <0.01 |
| MDCR | 1,660 | 15,632 | 3.5 | 42.9 | 111 | 1,311 | 0.96 (0.76 - 1.22) | 0.77 | 0.88 (0.69 - 1.12) | 0.31 |
| Open Claims | 62,159 | 304,538 | 143.5 | 1252.4 | 2,549 | 23,261 | 1.11 (1.06 - 1.16) | <0.01 | 1.09 (0.94 - 1.27) | 0.28 |
| OptumEHR | 3,040 | 21,828 | 6.3 | 72.0 | 113 | 1,302 | 1.14 (0.90 - 1.41) | 0.26 | 1.08 (0.86 - 1.35) | 0.53 |
| Meta-analysis | 71,111 | 365,226 | 160.0 | 1432.5 | 3,050 | 28,056 | 1.12 (1.06 - 1.19) | <0.01 | 1.09 (0.94 - 1.26) | 0.24 |

#### 6.3 GLP1RA vs SU

##### 6.3.1 Abnormal weight gain

**Supplementary Table 144:** Exposures, outcomes and risk estimates for Abnormal weight gain in GLP1RA (T) vs SU (C) new-user cohorts under four different propensity score (PS) adjustment / time-at-risk (TAR) designs across data sources. We report total patient exposure time in 1,000 patient-year (KYr) units, uncalibrated and calibrated hazard ratios (HRs) and their 95% confidence intervals (CIs). Greyed out entries did not pass study diagnostics and are included only for completeness.

| | Patients | | Exposure (KYr) | | Outcomes | | Uncalibrated | | Calibrated | |
|---|---|---|---|---|---|---|---|---|---|---|
| | T | C | T | C | T | C | HR (95% CI) | $p$ | HR (95% CI) | $p$ |
| **PS-stratification / on-treatment TAR** | | | | | | | | | | |
| OptumDOD | 5,281 | 64,799 | 2.7 | 67.3 | 18 | 412 | 0.95 (0.55 - 1.54) | 0.85 | 0.93 (0.56 - 1.55) | 0.78 |
| MDCR | 2,056 | 36,650 | 1.3 | 36.4 | 7 | 125 | 1.20 (0.48 - 2.61) | 0.67 | 1.06 (0.46 - 2.49) | 0.89 |
| Open Claims | 81,161 | 689,985 | 49.5 | 796.3 | 108 | 1,657 | 0.91 (0.73 - 1.12) | 0.37 | 0.87 (0.65 - 1.17) | 0.36 |
| OptumEHR | 3,350 | 57,248 | 0.9 | 34.9 | 10 | 218 | 1.80 (0.85 - 3.42) | 0.10 | 1.50 (0.75 - 3.00) | 0.27 |
| Meta-analysis | 81,161 | 689,985 | 49.5 | 796.3 | 108 | 1,657 | 0.91 (0.73 - 1.12) | 0.37 | 0.87 (0.64 - 1.17) | 0.35 |
| **PS-matching / on-treatment TAR** | | | | | | | | | | |
| OptumDOD | 4,871 | 53,498 | 2.6 | 54.1 | 15 | 327 | 0.60 (0.26 - 1.22) | 0.20 | 0.60 (0.28 - 1.31) | 0.20 |
| MDCR | 1,896 | 28,137 | 1.3 | 28.3 | 7 | 85 | 1.25 (0.44 - 3.01) | 0.65 | 1.12 (0.43 - 2.92) | 0.82 |
| Open Claims | 75,457 | 642,107 | 47.2 | 740.2 | 88 | 1,477 | 0.76 (0.58 - 0.99) | 0.05 | 0.74 (0.54 - 1.01) | 0.07 |
| OptumEHR | 3,118 | 46,569 | 0.9 | 27.8 | 7 | 168 | 1.38 (0.49 - 3.28) | 0.51 | 1.10 (0.36 - 3.40) | 0.87 |
| Meta-analysis | 85,342 | 770,311 | 51.9 | 850.4 | 117 | 2,057 | 0.80 (0.63 - 1.01) | 0.06 | 0.76 (0.57 - 1.02) | 0.07 |
| **PS-stratification / intent-to-treat TAR** | | | | | | | | | | |
| OptumDOD | 5,281 | 64,799 | 7.8 | 209.8 | 57 | 1,067 | 1.25 (0.93 - 1.65) | 0.13 | 1.23 (0.92 - 1.64) | 0.19 |
| MDCR | 2,056 | 36,650 | 4.7 | 113.4 | 35 | 418 | 1.72 (1.15 - 2.50) | <0.01 | 1.57 (1.06 - 2.31) | 0.03 |
| Open Claims | 81,161 | 689,985 | 168.5 | 2828.5 | 469 | 5,713 | 1.24 (1.12 - 1.37) | <0.01 | 1.22 (0.95 - 1.55) | 0.12 |
| OptumEHR | 3,350 | 57,248 | 6.9 | 216.8 | 56 | 891 | 1.86 (1.36 - 2.48) | <0.01 | 1.86 (1.37 - 2.51) | <0.01 |
| Meta-analysis | 81,161 | 689,985 | 168.5 | 2828.5 | 469 | 5,713 | 1.24 (1.12 - 1.37) | <0.01 | 1.22 (0.95 - 1.56) | 0.12 |
| **PS-matching / intent-to-treat TAR** | | | | | | | | | | |
| OptumDOD | 4,871 | 53,498 | 7.5 | 161.3 | 52 | 837 | 1.19 (0.83 - 1.66) | 0.33 | 1.20 (0.85 - 1.70) | 0.32 |
| MDCR | 1,896 | 28,137 | 4.6 | 89.5 | 33 | 302 | 1.40 (0.86 - 2.19) | 0.16 | 1.28 (0.80 - 2.05) | 0.31 |
| Open Claims | 75,457 | 642,107 | 163.3 | 2573.7 | 424 | 5,046 | 1.13 (1.00 - 1.28) | 0.04 | 1.12 (0.88 - 1.42) | 0.38 |
| OptumEHR | 3,118 | 46,569 | 6.6 | 165.7 | 48 | 670 | 1.58 (1.11 - 2.21) | <0.01 | 1.63 (1.15 - 2.30) | <0.01 |
| Meta-analysis | 85,342 | 770,311 | 182.1 | 2990.3 | 557 | 6,855 | 1.24 (1.05 - 1.46) | 0.01 | 1.20 (0.95 - 1.52) | 0.12 |

##### 6.3.2 Abnormal weight loss

**Supplementary Table 145:** Exposures, outcomes and risk estimates for Abnormal weight loss in GLP1RA (T) vs SU (C) new-user cohorts under four different propensity score (PS) adjustment / time-at-risk (TAR) designs across data sources. We report total patient exposure time in 1,000 patient-year (KYr) units, uncalibrated and calibrated hazard ratios (HRs) and their 95% confidence intervals (CIs). Greyed out entries did not pass study diagnostics and are included only for completeness.

| | Patients | | Exposure (KYr) | | Outcomes | | Uncalibrated | | Calibrated | |
|---|---|---|---|---|---|---|---|---|---|---|
| | T | C | T | C | T | C | HR (95% CI) | $p$ | HR (95% CI) | $p$ |
| **PS-stratification / on-treatment TAR** | | | | | | | | | | |
| OptumDOD | 5,409 | 62,245 | 2.8 | 64.7 | 60 | 1,144 | 1.27 (0.94 - 1.67) | 0.11 | 1.24 (0.93 - 1.65) | 0.16 |
| MDCR | 2,094 | 35,329 | 1.4 | 35.1 | 16 | 435 | 1.01 (0.56 - 1.69) | 0.98 | 0.89 (0.51 - 1.54) | 0.68 |
| Open Claims | 83,942 | 672,909 | 51.0 | 777.3 | 368 | 5,606 | 1.30 (1.15 - 1.45) | <0.01 | 1.24 (0.98 - 1.57) | 0.08 |
| OptumEHR | 3,464 | 56,005 | 1.0 | 34.0 | 9 | 536 | 0.73 (0.34 - 1.37) | 0.38 | 0.61 (0.30 - 1.22) | 0.18 |
| Meta-analysis | 83,942 | 672,909 | 51.0 | 777.3 | 368 | 5,606 | 1.30 (1.15 - 1.45) | <0.01 | 1.24 (0.98 - 1.58) | 0.08 |
| **PS-matching / on-treatment TAR** | | | | | | | | | | |
| OptumDOD | 4,912 | 52,504 | 2.6 | 53.3 | 58 | 890 | 1.25 (0.88 - 1.73) | 0.20 | 1.25 (0.89 - 1.75) | 0.22 |
| MDCR | 1,909 | 27,582 | 1.3 | 27.7 | 15 | 315 | 0.91 (0.45 - 1.69) | 0.78 | 0.81 (0.42 - 1.59) | 0.55 |
| Open Claims | 77,147 | 630,400 | 48.1 | 727.2 | 338 | 4,924 | 1.23 (1.07 - 1.41) | <0.01 | 1.20 (0.97 - 1.48) | 0.13 |
| OptumEHR | 3,182 | 45,909 | 0.9 | 27.2 | 9 | 398 | 0.91 (0.40 - 1.81) | 0.81 | 0.73 (0.28 - 1.91) | 0.56 |
| Meta-analysis | 87,150 | 756,395 | 52.9 | 835.5 | 420 | 6,527 | 1.21 (1.07 - 1.37) | <0.01 | 1.16 (0.94 - 1.43) | 0.17 |
| **PS-stratification / intent-to-treat TAR** | | | | | | | | | | |
| OptumDOD | 5,409 | 62,245 | 7.9 | 195.9 | 163 | 4,099 | 1.12 (0.94 - 1.32) | 0.20 | 1.10 (0.92 - 1.30) | 0.34 |
| MDCR | 2,094 | 35,329 | 4.7 | 107.1 | 60 | 1,859 | 0.88 (0.66 - 1.14) | 0.35 | 0.80 (0.61 - 1.05) | 0.13 |
| Open Claims | 83,942 | 672,909 | 172.5 | 2720.9 | 1,215 | 25,887 | 0.97 (0.91 - 1.03) | 0.35 | 0.95 (0.76 - 1.20) | 0.68 |
| OptumEHR | 3,464 | 56,005 | 7.0 | 207.3 | 85 | 3,016 | 1.11 (0.88 - 1.38) | 0.38 | 1.11 (0.88 - 1.40) | 0.43 |
| Meta-analysis | 83,942 | 672,909 | 172.5 | 2720.9 | 1,215 | 25,887 | 0.97 (0.91 - 1.03) | 0.35 | 0.95 (0.75 - 1.20) | 0.68 |
| **PS-matching / intent-to-treat TAR** | | | | | | | | | | |
| OptumDOD | 4,912 | 52,504 | 7.6 | 155.5 | 156 | 3,073 | 1.08 (0.89 - 1.30) | 0.45 | 1.08 (0.89 - 1.31) | 0.44 |
| MDCR | 1,909 | 27,582 | 4.6 | 85.9 | 58 | 1,400 | 0.84 (0.60 - 1.15) | 0.30 | 0.77 (0.56 - 1.07) | 0.13 |
| Open Claims | 77,147 | 630,400 | 166.2 | 2498.9 | 1,156 | 22,438 | 0.98 (0.91 - 1.05) | 0.52 | 0.96 (0.77 - 1.20) | 0.74 |
| OptumEHR | 3,182 | 45,909 | 6.7 | 160.3 | 83 | 2,151 | 1.11 (0.86 - 1.41) | 0.43 | 1.14 (0.88 - 1.46) | 0.36 |
| Meta-analysis | 87,150 | 756,395 | 185.0 | 2900.6 | 1,453 | 29,062 | 0.99 (0.93 - 1.05) | 0.74 | 0.96 (0.80 - 1.15) | 0.66 |

##### 6.3.3 Acute pancreatitis

**Supplementary Table 146:** Exposures, outcomes and risk estimates for Acute pancreatitis in GLP1RA (T) vs SU (C) new-user cohorts under four different propensity score (PS) adjustment / time-at-risk (TAR) designs across data sources. We report total patient exposure time in 1,000 patient-year (KYr) units, uncalibrated and calibrated hazard ratios (HRs) and their 95% confidence intervals (CIs). Greyed out entries did not pass study diagnostics and are included only for completeness.

| | Patients | | Exposure (KYr) | | Outcomes | | Uncalibrated | | Calibrated | |
|---|---|---|---|---|---|---|---|---|---|---|
| | T | C | T | C | T | C | HR (95% CI) | $p$ | HR (95% CI) | $p$ |
| **PS-stratification / on-treatment TAR** | | | | | | | | | | |
| OptumDOD | 5,591 | 65,491 | 2.9 | 68.3 | 9 | 262 | 0.97 (0.44 - 1.88) | 0.93 | 0.95 (0.46 - 1.95) | 0.88 |
| MDCR | 2,126 | 36,544 | 1.4 | 36.3 | <5 | 137 | 0.90 (0.26 - 2.36) | 0.85 | 0.80 (0.27 - 2.40) | 0.69 |
| Open Claims | 86,463 | 696,817 | 52.6 | 804.1 | 87 | 1,771 | 0.97 (0.77 - 1.22) | 0.81 | 0.93 (0.68 - 1.27) | 0.65 |
| OptumEHR | 3,555 | 58,038 | 1.0 | 35.5 | <5 | 105 | 1.18 (0.28 - 3.36) | 0.80 | 0.98 (0.28 - 3.41) | 0.94 |
| Meta-analysis | 86,463 | 696,817 | 52.6 | 804.1 | 87 | 1,771 | 0.97 (0.77 - 1.22) | 0.81 | 0.93 (0.68 - 1.27) | 0.65 |
| **PS-matching / on-treatment TAR** | | | | | | | | | | |
| OptumDOD | 5,083 | 54,435 | 2.7 | 55.3 | 9 | 195 | 0.95 (0.35 - 2.18) | 0.92 | 0.95 (0.38 - 2.38) | 0.92 |
| MDCR | 1,939 | 28,098 | 1.3 | 28.3 | <5 | 101 | 1.04 (0.24 - 3.14) | 0.96 | 0.93 (0.26 - 3.37) | 0.91 |
| Open Claims | 79,335 | 650,891 | 49.6 | 750.3 | 82 | 1,575 | 1.04 (0.79 - 1.34) | 0.80 | 1.00 (0.74 - 1.37) | 0.93 |
| OptumEHR | 3,266 | 47,414 | 0.9 | 28.4 | <5 | 80 | 0.75 (0.11 - 2.98) | 0.73 | 0.60 (0.10 - 3.45) | 0.58 |
| Meta-analysis | 89,623 | 780,838 | 54.5 | 862.3 | <101 | 1,951 | 1.02 (0.80 - 1.31) | 0.86 | 0.98 (0.72 - 1.32) | 0.88 |
| **PS-stratification / intent-to-treat TAR** | | | | | | | | | | |
| OptumDOD | 5,591 | 65,491 | 8.3 | 212.8 | 31 | 796 | 1.20 (0.80 - 1.74) | 0.35 | 1.18 (0.80 - 1.74) | 0.41 |
| MDCR | 2,126 | 36,544 | 4.9 | 112.8 | 17 | 473 | 0.87 (0.50 - 1.41) | 0.60 | 0.79 (0.47 - 1.33) | 0.39 |
| Open Claims | 86,463 | 696,817 | 179.1 | 2855.2 | 306 | 6,504 | 0.96 (0.84 - 1.08) | 0.47 | 0.94 (0.73 - 1.21) | 0.62 |
| OptumEHR | 3,555 | 58,038 | 7.3 | 221.0 | 13 | 421 | 0.98 (0.52 - 1.69) | 0.94 | 0.98 (0.54 - 1.77) | 0.93 |
| Meta-analysis | 86,463 | 696,817 | 179.1 | 2855.2 | 306 | 6,504 | 0.96 (0.84 - 1.08) | 0.47 | 0.94 (0.72 - 1.21) | 0.62 |
| **PS-matching / intent-to-treat TAR** | | | | | | | | | | |
| OptumDOD | 5,083 | 54,435 | 8.0 | 165.2 | 31 | 592 | 1.36 (0.87 - 2.04) | 0.16 | 1.37 (0.89 - 2.10) | 0.15 |
| MDCR | 1,939 | 28,098 | 4.7 | 89.1 | 17 | 363 | 0.92 (0.48 - 1.60) | 0.78 | 0.84 (0.46 - 1.53) | 0.58 |
| Open Claims | 79,335 | 650,891 | 172.4 | 2611.4 | 293 | 5,694 | 0.99 (0.87 - 1.13) | 0.91 | 0.98 (0.76 - 1.25) | 0.85 |
| OptumEHR | 3,266 | 47,414 | 7.0 | 169.9 | 12 | 304 | 0.85 (0.39 - 1.62) | 0.65 | 0.87 (0.43 - 1.77) | 0.71 |
| Meta-analysis | 89,623 | 780,838 | 192.1 | 3035.6 | 353 | 6,953 | 1.01 (0.89 - 1.14) | 0.87 | 0.98 (0.80 - 1.21) | 0.85 |

##### 6.3.4 All-cause mortality

**Supplementary Table 147:** Exposures, outcomes and risk estimates for All-cause mortality in GLP1RA (T) vs SU (C) new-user cohorts under four different propensity score (PS) adjustment / time-at-risk (TAR) designs across data sources. We report total patient exposure time in 1,000 patient-year (KYr) units, uncalibrated and calibrated hazard ratios (HRs) and their 95% confidence intervals (CIs). Greyed out entries did not pass study diagnostics and are included only for completeness.

| | Patients | | Exposure (KYr) | | Outcomes | | Uncalibrated | | Calibrated | |
|---|---|---|---|---|---|---|---|---|---|---|
| | T | C | T | C | T | C | HR (95% CI) | $p$ | HR (95% CI) | $p$ |
| **PS-stratification / on-treatment TAR** | | | | | | | | | | |
| OptumDOD | 5,626 | 66,287 | 2.9 | 69.2 | 16 | 1,085 | 0.59 (0.34 - 0.95) | 0.04 | 0.58 (0.34 - 0.97) | 0.04 |
| MDCR | 2,135 | 36,988 | 1.4 | 36.9 | <5 | 148 | 1.20 (0.19 - 4.07) | 0.82 | 1.06 (0.23 - 4.92) | 0.94 |
| Open Claims | 87,486 | 707,423 | 53.1 | 817.0 | 0 | 13 | | | | |
| OptumEHR | 3,573 | 58,370 | 1.0 | 35.8 | <5 | 604 | 0.35 (0.09 - 0.91) | 0.08 | 0.29 (0.09 - 0.94) | 0.04 |
| **PS-matching / on-treatment TAR** | | | | | | | | | | |
| OptumDOD | 5,114 | 54,931 | 2.7 | 55.9 | 16 | 728 | 0.80 (0.44 - 1.35) | 0.44 | 0.80 (0.46 - 1.40) | 0.45 |
| MDCR | 1,949 | 28,390 | 1.3 | 28.6 | <5 | 81 | 1.62 (0.25 - 5.71) | 0.54 | 1.45 (0.31 - 6.88) | 0.64 |
| Open Claims | 80,327 | 659,963 | 50.2 | 761.2 | 0 | 10 | | | | |
| OptumEHR | 3,282 | 47,614 | 0.9 | 28.5 | <5 | 394 | 0.38 (0.09 - 1.05) | 0.12 | 0.30 (0.08 - 1.19) | 0.10 |
| Meta-analysis | 10,345 | 130,935 | 5.0 | 113.1 | <26 | 1,203 | 0.77 (0.47 - 1.24) | 0.27 | 0.73 (0.44 - 1.22) | 0.23 |
| **PS-stratification / intent-to-treat TAR** | | | | | | | | | | |
| OptumDOD | 5,626 | 66,287 | 8.4 | 212.1 | 169 | 10,047 | 0.76 (0.65 - 0.89) | <0.01 | 0.75 (0.64 - 0.88) | <0.01 |
| MDCR | 2,135 | 36,988 | 4.9 | 115.0 | 15 | 846 | 0.85 (0.48 - 1.41) | 0.57 | 0.78 (0.45 - 1.33) | 0.37 |
| Open Claims | 87,486 | 707,423 | 181.6 | 2912.9 | 0 | 66 | | | | |
| OptumEHR | 3,573 | 58,370 | 7.4 | 222.1 | 44 | 3,689 | 0.56 (0.40 - 0.75) | <0.01 | 0.56 (0.41 - 0.76) | <0.01 |
| Meta-analysis | 0 | 0 | 0.0 | 0.0 | 0 | 0 | | | | |
| **PS-matching / intent-to-treat TAR** | | | | | | | | | | |
| OptumDOD | 5,114 | 54,931 | 8.0 | 164.7 | 167 | 6,420 | 0.77 (0.65 - 0.92) | <0.01 | 0.78 (0.65 - 0.93) | <0.01 |
| MDCR | 1,949 | 28,390 | 4.8 | 90.7 | 15 | 518 | 1.18 (0.65 - 1.97) | 0.56 | 1.08 (0.62 - 1.88) | 0.79 |
| Open Claims | 80,327 | 659,963 | 174.9 | 2659.0 | 0 | 50 | | | | |
| OptumEHR | 3,282 | 47,614 | 7.0 | 170.5 | 42 | 2,313 | 0.62 (0.44 - 0.85) | <0.01 | 0.64 (0.46 - 0.88) | 0.01 |
| Meta-analysis | 10,345 | 130,935 | 19.8 | 426.0 | 224 | 9,251 | 0.77 (0.61 - 0.97) | 0.02 | 0.74 (0.56 - 0.99) | 0.04 |

##### 6.3.5 Bone fracture

**Supplementary Table 148:** Exposures, outcomes and risk estimates for Bone fracture in GLP1RA (T) vs SU (C) new-user cohorts under four different propensity score (PS) adjustment / time-at-risk (TAR) designs across data sources. We report total patient exposure time in 1,000 patient-year (KYr) units, uncalibrated and calibrated hazard ratios (HRs) and their 95% confidence intervals (CIs). Greyed out entries did not pass study diagnostics and are included only for completeness.

| | Patients | | Exposure (KYr) | | Outcomes | | Uncalibrated | | Calibrated | |
|---|---|---|---|---|---|---|---|---|---|---|
| | T | C | T | C | T | C | HR (95% CI) | $p$ | HR (95% CI) | $p$ |
| **PS-stratification / on-treatment TAR** | | | | | | | | | | |
| OptumDOD | 4,964 | 59,394 | 2.5 | 60.7 | 96 | 2,091 | 1.06 (0.85 - 1.32) | 0.58 | 1.04 (0.83 - 1.30) | 0.75 |
| MDCR | 1,876 | 32,430 | 1.2 | 31.6 | 33 | 1,115 | 0.77 (0.52 - 1.09) | 0.16 | 0.68 (0.47 - 0.98) | 0.05 |
| Open Claims | 70,749 | 605,180 | 42.8 | 692.1 | 756 | 15,715 | 0.85 (0.78 - 0.91) | <0.01 | 0.81 (0.65 - 1.01) | 0.07 |
| OptumEHR | 3,230 | 53,600 | 0.9 | 31.8 | 20 | 922 | 0.93 (0.56 - 1.44) | 0.75 | 0.77 (0.48 - 1.23) | 0.31 |
| Meta-analysis | 70,749 | 605,180 | 42.8 | 692.1 | 756 | 15,715 | 0.85 (0.78 - 0.91) | <0.01 | 0.81 (0.65 - 1.01) | 0.06 |
| **PS-matching / on-treatment TAR** | | | | | | | | | | |
| OptumDOD | 4,534 | 49,318 | 2.4 | 49.1 | 88 | 1,604 | 1.04 (0.79 - 1.36) | 0.76 | 1.05 (0.80 - 1.37) | 0.76 |
| MDCR | 1,721 | 25,443 | 1.1 | 25.0 | 33 | 847 | 0.81 (0.53 - 1.20) | 0.32 | 0.73 (0.48 - 1.10) | 0.14 |
| Open Claims | 65,480 | 562,671 | 40.6 | 642.6 | 700 | 14,163 | 0.82 (0.75 - 0.91) | <0.01 | 0.80 (0.66 - 0.96) | 0.03 |
| OptumEHR | 2,985 | 43,566 | 0.8 | 25.4 | 18 | 670 | 1.16 (0.67 - 1.88) | 0.58 | 0.92 (0.42 - 2.04) | 0.84 |
| Meta-analysis | 74,720 | 680,998 | 45.0 | 742.1 | 839 | 17,284 | 0.89 (0.76 - 1.05) | 0.16 | 0.85 (0.68 - 1.08) | 0.18 |
| **PS-stratification / intent-to-treat TAR** | | | | | | | | | | |
| OptumDOD | 4,964 | 59,394 | 7.1 | 179.0 | 237 | 6,544 | 0.89 (0.77 - 1.02) | 0.10 | 0.87 (0.76 - 1.01) | 0.10 |
| MDCR | 1,876 | 32,430 | 4.0 | 92.8 | 156 | 3,705 | 1.02 (0.86 - 1.20) | 0.84 | 0.93 (0.78 - 1.10) | 0.44 |
| Open Claims | 70,749 | 605,180 | 143.2 | 2364.7 | 2,962 | 61,857 | 0.92 (0.88 - 0.95) | <0.01 | 0.90 (0.72 - 1.13) | 0.36 |
| OptumEHR | 3,230 | 53,600 | 6.5 | 193.1 | 116 | 4,914 | 0.83 (0.68 - 1.00) | 0.06 | 0.83 (0.68 - 1.01) | 0.09 |
| Meta-analysis | 70,749 | 605,180 | 143.2 | 2364.7 | 2,962 | 61,857 | 0.92 (0.88 - 0.95) | <0.01 | 0.90 (0.71 - 1.13) | 0.36 |
| **PS-matching / intent-to-treat TAR** | | | | | | | | | | |
| OptumDOD | 4,534 | 49,318 | 6.9 | 139.1 | 222 | 4,907 | 0.91 (0.77 - 1.06) | 0.22 | 0.91 (0.77 - 1.07) | 0.30 |
| MDCR | 1,721 | 25,443 | 3.9 | 74.6 | 156 | 2,845 | 1.03 (0.84 - 1.24) | 0.79 | 0.94 (0.77 - 1.15) | 0.58 |
| Open Claims | 65,480 | 562,671 | 138.4 | 2153.8 | 2,848 | 54,377 | 0.92 (0.88 - 0.96) | <0.01 | 0.91 (0.73 - 1.12) | 0.38 |
| OptumEHR | 2,985 | 43,566 | 6.2 | 148.2 | 111 | 3,552 | 0.85 (0.68 - 1.04) | 0.13 | 0.87 (0.70 - 1.08) | 0.27 |
| Meta-analysis | 74,720 | 680,998 | 155.4 | 2515.7 | 3,337 | 65,681 | 0.92 (0.89 - 0.96) | <0.01 | 0.90 (0.75 - 1.06) | 0.21 |

##### 6.3.6 Diabetic ketoacidosis

**Supplementary Table 149:** Exposures, outcomes and risk estimates for Diabetic ketoacidosis in GLP1RA (T) vs SU (C) new-user cohorts under four different propensity score (PS) adjustment / time-at-risk (TAR) designs across data sources. We report total patient exposure time in 1,000 patient-year (KYr) units, uncalibrated and calibrated hazard ratios (HRs) and their 95% confidence intervals (CIs). Greyed out entries did not pass study diagnostics and are included only for completeness.

| | Patients | | Exposure (KYr) | | Outcomes | | Uncalibrated | | Calibrated | |
|---|---|---|---|---|---|---|---|---|---|---|
| | T | C | T | C | T | C | HR (95% CI) | $p$ | HR (95% CI) | $p$ |
| **PS-stratification / on-treatment TAR** | | | | | | | | | | |
| OptumDOD | 5,619 | 66,185 | 2.9 | 69.1 | <5 | 111 | 0.61 (0.14 - 1.79) | 0.44 | 0.59 (0.17 - 2.11) | 0.42 |
| MDCR | 2,134 | 36,952 | 1.4 | 36.8 | 0 | 42 | | | | |
| Open Claims | 87,394 | 706,213 | 53.1 | 815.6 | 32 | 616 | 1.00 (0.67 - 1.44) | 0.99 | 0.96 (0.62 - 1.47) | 0.84 |
| OptumEHR | 3,572 | 58,335 | 1.0 | 35.8 | <5 | 39 | 1.15 (0.06 - 5.97) | 0.91 | 0.95 (0.10 - 9.33) | 0.96 |
| Meta-analysis | 87,394 | 706,213 | 53.1 | 815.6 | 32 | 616 | 1.00 (0.67 - 1.44) | 0.99 | 0.96 (0.62 - 1.48) | 0.84 |
| **PS-matching / on-treatment TAR** | | | | | | | | | | |
| OptumDOD | 5,109 | 54,873 | 2.7 | 55.8 | <5 | 84 | 1.50 (0.31 - 5.04) | 0.57 | 1.50 (0.37 - 6.02) | 0.57 |
| MDCR | 1,949 | 28,376 | 1.3 | 28.6 | 0 | 26 | | | | |
| Open Claims | 80,238 | 658,824 | 50.1 | 760.0 | 31 | 537 | 0.98 (0.64 - 1.45) | 0.91 | 0.95 (0.61 - 1.47) | 0.82 |
| OptumEHR | 3,280 | 47,587 | 0.9 | 28.6 | <5 | 27 | 1.71 (0.09 - 9.26) | 0.65 | 1.36 (0.13 - 14.8) | 0.80 |
| Meta-analysis | 85,347 | 713,697 | 52.8 | 815.8 | <36 | 621 | 1.01 (0.68 - 1.50) | 0.95 | 0.97 (0.63 - 1.49) | 0.88 |
| **PS-stratification / intent-to-treat TAR** | | | | | | | | | | |
| OptumDOD | 5,619 | 66,185 | 8.4 | 216.0 | 7 | 490 | 0.44 (0.18 - 0.88) | 0.04 | 0.43 (0.20 - 0.94) | 0.04 |
| MDCR | 2,134 | 36,952 | 4.9 | 114.6 | <5 | 200 | 0.36 (0.06 - 1.15) | 0.18 | 0.32 (0.07 - 1.45) | 0.14 |
| Open Claims | 87,394 | 706,213 | 181.2 | 2901.5 | 156 | 3,708 | 0.88 (0.74 - 1.04) | 0.14 | 0.86 (0.65 - 1.14) | 0.30 |
| OptumEHR | 3,572 | 58,335 | 7.4 | 222.8 | 5 | 219 | 1.02 (0.35 - 2.31) | 0.97 | 1.02 (0.40 - 2.61) | 0.96 |
| Meta-analysis | 87,394 | 706,213 | 181.2 | 2901.5 | 156 | 3,708 | 0.88 (0.74 - 1.04) | 0.14 | 0.86 (0.65 - 1.14) | 0.30 |
| **PS-matching / intent-to-treat TAR** | | | | | | | | | | |
| OptumDOD | 5,109 | 54,873 | 8.1 | 167.2 | 7 | 351 | 0.55 (0.23 - 1.11) | 0.14 | 0.55 (0.25 - 1.22) | 0.14 |
| MDCR | 1,949 | 28,376 | 4.8 | 90.5 | <5 | 144 | 0.35 (0.06 - 1.21) | 0.18 | 0.32 (0.07 - 1.51) | 0.15 |
| Open Claims | 80,238 | 658,824 | 174.5 | 2648.8 | 152 | 3,236 | 0.87 (0.72 - 1.05) | 0.15 | 0.86 (0.65 - 1.14) | 0.29 |
| OptumEHR | 3,280 | 47,587 | 7.0 | 170.9 | 5 | 158 | 1.14 (0.39 - 2.62) | 0.79 | 1.17 (0.45 - 3.03) | 0.75 |
| Meta-analysis | 90,576 | 789,660 | 194.4 | 3077.4 | <169 | 3,889 | 0.85 (0.71 - 1.01) | 0.07 | 0.83 (0.65 - 1.05) | 0.12 |

##### 6.3.7 Diarrhea

**Supplementary Table 150:** Exposures, outcomes and risk estimates for Diarrhea in GLP1RA (T) vs SU (C) new-user cohorts under four different propensity score (PS) adjustment / time-at-risk (TAR) designs across data sources. We report total patient exposure time in 1,000 patient-year (KYr) units, uncalibrated and calibrated hazard ratios (HRs) and their 95% confidence intervals (CIs). Greyed out entries did not pass study diagnostics and are included only for completeness.

| | Patients | | Exposure (KYr) | | Outcomes | | Uncalibrated | | Calibrated | |
|---|---|---|---|---|---|---|---|---|---|---|
| | T | C | T | C | T | C | HR (95% CI) | $p$ | HR (95% CI) | $p$ |
| **PS-stratification / on-treatment TAR** | | | | | | | | | | |
| OptumDOD | 4,704 | 55,726 | 2.4 | 56.0 | 125 | 2,541 | 1.00 (0.81 - 1.21) | 0.96 | 0.97 (0.80 - 1.18) | 0.78 |
| MDCR | 1,864 | 32,189 | 1.2 | 31.3 | 46 | 1,101 | 1.01 (0.73 - 1.38) | 0.94 | 0.90 (0.65 - 1.24) | 0.52 |
| Open Claims | 72,549 | 609,869 | 43.8 | 694.2 | 965 | 14,458 | 1.07 (1.00 - 1.15) | 0.05 | 1.03 (0.83 - 1.28) | 0.81 |
| OptumEHR | 3,136 | 51,403 | 0.8 | 29.7 | 36 | 1,351 | 1.04 (0.72 - 1.45) | 0.85 | 0.86 (0.60 - 1.23) | 0.46 |
| Meta-analysis | 72,549 | 609,869 | 43.8 | 694.2 | 965 | 14,458 | 1.07 (1.00 - 1.15) | 0.05 | 1.03 (0.82 - 1.28) | 0.80 |
| **PS-matching / on-treatment TAR** | | | | | | | | | | |
| OptumDOD | 4,274 | 46,675 | 2.3 | 45.9 | 115 | 2,044 | 1.00 (0.78 - 1.26) | 1.00 | 1.00 (0.79 - 1.28) | 0.85 |
| MDCR | 1,722 | 24,974 | 1.1 | 24.6 | 45 | 825 | 1.22 (0.83 - 1.75) | 0.30 | 1.09 (0.75 - 1.59) | 0.66 |
| Open Claims | 67,070 | 567,394 | 41.6 | 645.2 | 889 | 13,011 | 1.07 (0.98 - 1.16) | 0.14 | 1.03 (0.86 - 1.24) | 0.73 |
| OptumEHR | 2,890 | 42,214 | 0.8 | 24.0 | 33 | 1,009 | 1.24 (0.83 - 1.79) | 0.28 | 0.99 (0.48 - 2.02) | 0.87 |
| Meta-analysis | 75,956 | 681,257 | 45.8 | 739.7 | 1,082 | 16,889 | 1.07 (0.99 - 1.16) | 0.08 | 1.03 (0.85 - 1.23) | 0.79 |
| **PS-stratification / intent-to-treat TAR** | | | | | | | | | | |
| OptumDOD | 4,704 | 55,726 | 6.5 | 165.0 | 331 | 7,254 | 1.02 (0.90 - 1.15) | 0.75 | 1.00 (0.89 - 1.13) | 0.86 |
| MDCR | 1,864 | 32,189 | 4.0 | 92.7 | 155 | 3,693 | 0.93 (0.78 - 1.10) | 0.39 | 0.85 (0.71 - 1.01) | 0.08 |
| Open Claims | 72,549 | 609,869 | 145.0 | 2380.8 | 3,172 | 52,286 | 1.03 (0.99 - 1.07) | 0.10 | 1.01 (0.81 - 1.27) | 0.90 |
| OptumEHR | 3,136 | 51,403 | 6.0 | 179.7 | 193 | 5,863 | 1.10 (0.94 - 1.27) | 0.24 | 1.10 (0.94 - 1.28) | 0.32 |
| Meta-analysis | 72,549 | 609,869 | 145.0 | 2380.8 | 3,172 | 52,286 | 1.03 (0.99 - 1.07) | 0.10 | 1.01 (0.81 - 1.27) | 0.90 |
| **PS-matching / intent-to-treat TAR** | | | | | | | | | | |
| OptumDOD | 4,274 | 46,675 | 6.2 | 129.7 | 312 | 5,703 | 1.03 (0.90 - 1.18) | 0.66 | 1.04 (0.90 - 1.19) | 0.64 |
| MDCR | 1,722 | 24,974 | 3.9 | 73.9 | 154 | 2,851 | 0.93 (0.75 - 1.13) | 0.46 | 0.85 (0.69 - 1.04) | 0.14 |
| Open Claims | 67,070 | 567,394 | 140.1 | 2169.7 | 3,022 | 46,378 | 1.05 (1.00 - 1.09) | 0.04 | 1.03 (0.83 - 1.27) | 0.79 |
| OptumEHR | 2,890 | 42,214 | 5.7 | 139.9 | 185 | 4,376 | 1.07 (0.90 - 1.26) | 0.42 | 1.10 (0.93 - 1.31) | 0.35 |
| Meta-analysis | 75,956 | 681,257 | 155.9 | 2513.2 | 3,673 | 59,308 | 1.04 (1.00 - 1.08) | 0.04 | 1.01 (0.85 - 1.20) | 0.88 |

##### 6.3.8 Genitourinary infection

**Supplementary Table 151:** Exposures, outcomes and risk estimates for Genitourinary infection in GLP1RA (T) vs SU (C) new-user cohorts under four different propensity score (PS) adjustment / time-at-risk (TAR) designs across data sources. We report total patient exposure time in 1,000 patient-year (KYr) units, uncalibrated and calibrated hazard ratios (HRs) and their 95% confidence intervals (CIs). Greyed out entries did not pass study diagnostics and are included only for completeness.

| | Patients | | Exposure (KYr) | | Outcomes | | Uncalibrated | | Calibrated | |
|---|---|---|---|---|---|---|---|---|---|---|
| | T | C | T | C | T | C | HR (95% CI) | $p$ | HR (95% CI) | $p$ |
| **PS-stratification / on-treatment TAR** | | | | | | | | | | |
| OptumDOD | 4,094 | 48,387 | 2.0 | 47.6 | 147 | 3,915 | 0.92 (0.77 - 1.09) | 0.35 | 0.90 (0.75 - 1.07) | 0.27 |
| MDCR | 1,633 | 28,860 | 1.0 | 27.9 | 66 | 1,765 | 1.06 (0.81 - 1.37) | 0.66 | 0.94 (0.72 - 1.23) | 0.66 |
| Open Claims | 60,438 | 528,093 | 37.0 | 600.1 | 1,315 | 24,920 | 0.97 (0.91 - 1.03) | 0.32 | 0.93 (0.75 - 1.15) | 0.51 |
| OptumEHR | 2,901 | 48,228 | 0.8 | 27.6 | 53 | 1,855 | 1.29 (0.96 - 1.70) | 0.09 | 1.07 (0.80 - 1.44) | 0.68 |
| Meta-analysis | 60,438 | 528,093 | 37.0 | 600.1 | 1,315 | 24,920 | 0.97 (0.91 - 1.03) | 0.32 | 0.93 (0.75 - 1.16) | 0.51 |
| **PS-matching / on-treatment TAR** | | | | | | | | | | |
| OptumDOD | 3,764 | 39,980 | 1.9 | 38.4 | 141 | 2,957 | 0.91 (0.74 - 1.12) | 0.39 | 0.91 (0.74 - 1.13) | 0.43 |
| MDCR | 1,519 | 22,564 | 1.0 | 22.0 | 63 | 1,261 | 1.12 (0.82 - 1.51) | 0.46 | 1.00 (0.74 - 1.37) | 0.82 |
| Open Claims | 56,324 | 490,658 | 35.3 | 556.9 | 1,219 | 22,175 | 0.92 (0.86 - 0.99) | 0.03 | 0.90 (0.75 - 1.07) | 0.27 |
| OptumEHR | 2,680 | 39,588 | 0.7 | 22.3 | 50 | 1,383 | 1.15 (0.83 - 1.58) | 0.39 | 0.92 (0.46 - 1.82) | 0.82 |
| Meta-analysis | 64,287 | 592,790 | 38.8 | 639.6 | 1,473 | 27,776 | 0.94 (0.88 - 1.00) | 0.06 | 0.90 (0.75 - 1.08) | 0.24 |
| **PS-stratification / intent-to-treat TAR** | | | | | | | | | | |
| OptumDOD | 4,094 | 48,387 | 5.3 | 130.8 | 402 | 10,427 | 0.97 (0.87 - 1.08) | 0.63 | 0.96 (0.86 - 1.07) | 0.54 |
| MDCR | 1,633 | 28,860 | 3.2 | 78.7 | 226 | 5,407 | 1.05 (0.91 - 1.21) | 0.50 | 0.96 (0.83 - 1.11) | 0.60 |
| Open Claims | 60,438 | 528,093 | 118.2 | 1963.4 | 4,614 | 90,895 | 0.99 (0.96 - 1.02) | 0.67 | 0.97 (0.78 - 1.22) | 0.82 |
| OptumEHR | 2,901 | 48,228 | 5.4 | 162.4 | 240 | 7,847 | 1.06 (0.92 - 1.21) | 0.42 | 1.06 (0.92 - 1.22) | 0.52 |
| Meta-analysis | 60,438 | 528,093 | 118.2 | 1963.4 | 4,614 | 90,895 | 0.99 (0.96 - 1.02) | 0.67 | 0.97 (0.77 - 1.22) | 0.81 |
| **PS-matching / intent-to-treat TAR** | | | | | | | | | | |
| OptumDOD | 3,764 | 39,980 | 5.1 | 101.1 | 390 | 7,752 | 1.01 (0.89 - 1.14) | 0.89 | 1.02 (0.90 - 1.15) | 0.80 |
| MDCR | 1,519 | 22,564 | 3.2 | 63.0 | 223 | 4,062 | 1.05 (0.89 - 1.24) | 0.53 | 0.97 (0.82 - 1.14) | 0.70 |
| Open Claims | 56,324 | 490,658 | 114.6 | 1791.1 | 4,426 | 79,503 | 0.98 (0.95 - 1.02) | 0.33 | 0.97 (0.78 - 1.20) | 0.77 |
| OptumEHR | 2,680 | 39,588 | 5.1 | 126.0 | 234 | 5,812 | 1.03 (0.89 - 1.20) | 0.67 | 1.06 (0.91 - 1.24) | 0.53 |
| Meta-analysis | 64,287 | 592,790 | 128.0 | 2081.2 | 5,273 | 97,129 | 0.99 (0.96 - 1.02) | 0.52 | 0.96 (0.81 - 1.14) | 0.64 |

##### 6.3.9 Hyperkalemia

**Supplementary Table 152:** Exposures, outcomes and risk estimates for Hyperkalemia in GLP1RA (T) vs SU (C) new-user cohorts under four different propensity score (PS) adjustment / time-at-risk (TAR) designs across data sources. We report total patient exposure time in 1,000 patient-year (KYr) units, uncalibrated and calibrated hazard ratios (HRs) and their 95% confidence intervals (CIs). Greyed out entries did not pass study diagnostics and are included only for completeness.

| | Patients | | Exposure (KYr) | | Outcomes | | Uncalibrated | | Calibrated | |
|---|---|---|---|---|---|---|---|---|---|---|
| | T | C | T | C | T | C | HR (95% CI) | $p$ | HR (95% CI) | $p$ |
| **PS-stratification / on-treatment TAR** | | | | | | | | | | |
| OptumDOD | 5,330 | 61,816 | 2.7 | 62.8 | 65 | 2,047 | 0.81 (0.61 - 1.04) | 0.11 | 0.79 (0.60 - 1.03) | 0.09 |
| MDCR | 2,092 | 35,848 | 1.3 | 35.4 | 13 | 579 | 0.67 (0.36 - 1.16) | 0.19 | 0.60 (0.33 - 1.07) | 0.09 |
| Open Claims | 84,627 | 682,223 | 51.2 | 780.0 | 294 | 9,278 | 0.66 (0.58 - 0.75) | <0.01 | 0.63 (0.50 - 0.80) | <0.01 |
| OptumEHR | 3,469 | 56,303 | 1.0 | 33.6 | 10 | 809 | 0.57 (0.28 - 1.03) | 0.09 | 0.48 (0.25 - 0.91) | 0.03 |
| Meta-analysis | 84,627 | 682,223 | 51.2 | 780.0 | 294 | 9,278 | 0.66 (0.58 - 0.75) | <0.01 | 0.63 (0.50 - 0.81) | <0.01 |
| **PS-matching / on-treatment TAR** | | | | | | | | | | |
| OptumDOD | 4,839 | 51,345 | 2.5 | 51.0 | 64 | 1,548 | 0.84 (0.61 - 1.12) | 0.25 | 0.84 (0.62 - 1.14) | 0.28 |
| MDCR | 1,912 | 27,765 | 1.3 | 27.7 | 12 | 406 | 0.77 (0.37 - 1.45) | 0.45 | 0.69 (0.35 - 1.36) | 0.29 |
| Open Claims | 77,661 | 637,350 | 48.4 | 728.1 | 278 | 8,069 | 0.64 (0.55 - 0.74) | <0.01 | 0.62 (0.50 - 0.77) | <0.01 |
| OptumEHR | 3,179 | 46,045 | 0.9 | 27.0 | 9 | 573 | 0.60 (0.26 - 1.20) | 0.19 | 0.48 (0.18 - 1.27) | 0.18 |
| Meta-analysis | 87,591 | 762,505 | 53.1 | 833.8 | 363 | 10,596 | 0.70 (0.58 - 0.84) | <0.01 | 0.67 (0.52 - 0.86) | <0.01 |
| **PS-stratification / intent-to-treat TAR** | | | | | | | | | | |
| OptumDOD | 5,330 | 61,816 | 7.7 | 190.7 | 201 | 6,120 | 0.91 (0.78 - 1.05) | 0.22 | 0.89 (0.77 - 1.04) | 0.21 |
| MDCR | 2,092 | 35,848 | 4.8 | 108.6 | 61 | 2,098 | 0.88 (0.67 - 1.15) | 0.37 | 0.80 (0.61 - 1.06) | 0.13 |
| Open Claims | 84,627 | 682,223 | 173.6 | 2735.1 | 1,319 | 36,551 | 0.81 (0.77 - 0.86) | <0.01 | 0.80 (0.63 - 1.00) | 0.06 |
| OptumEHR | 3,469 | 56,303 | 7.0 | 205.1 | 79 | 3,918 | 0.76 (0.60 - 0.95) | 0.02 | 0.76 (0.60 - 0.96) | 0.04 |
| Meta-analysis | 84,627 | 682,223 | 173.6 | 2735.1 | 1,319 | 36,551 | 0.81 (0.77 - 0.86) | <0.01 | 0.80 (0.63 - 1.00) | 0.05 |
| **PS-matching / intent-to-treat TAR** | | | | | | | | | | |
| OptumDOD | 4,839 | 51,345 | 7.4 | 148.5 | 195 | 4,444 | 1.00 (0.85 - 1.17) | 0.98 | 1.00 (0.85 - 1.19) | 0.87 |
| MDCR | 1,912 | 27,765 | 4.6 | 86.4 | 60 | 1,496 | 0.88 (0.64 - 1.20) | 0.45 | 0.81 (0.59 - 1.12) | 0.21 |
| Open Claims | 77,661 | 637,350 | 167.2 | 2505.5 | 1,283 | 31,278 | 0.82 (0.77 - 0.87) | <0.01 | 0.80 (0.65 - 1.00) | 0.05 |
| OptumEHR | 3,179 | 46,045 | 6.7 | 158.6 | 78 | 2,746 | 0.69 (0.53 - 0.89) | <0.01 | 0.71 (0.55 - 0.92) | 0.02 |
| Meta-analysis | 87,591 | 762,505 | 185.9 | 2899.1 | 1,616 | 39,964 | 0.85 (0.74 - 0.97) | 0.02 | 0.82 (0.66 - 1.02) | 0.08 |

##### 6.3.10 Hypoglycemia

**Supplementary Table 153:** Exposures, outcomes and risk estimates for Hypoglycemia in GLP1RA (T) vs SU (C) new-user cohorts under four different propensity score (PS) adjustment / time-at-risk (TAR) designs across data sources. We report total patient exposure time in 1,000 patient-year (KYr) units, uncalibrated and calibrated hazard ratios (HRs) and their 95% confidence intervals (CIs). Greyed out entries did not pass study diagnostics and are included only for completeness.

| | Patients | | Exposure (KYr) | | Outcomes | | Uncalibrated | | Calibrated | |
|---|---|---|---|---|---|---|---|---|---|---|
| | T | C | T | C | T | C | HR (95% CI) | $p$ | HR (95% CI) | $p$ |
| **PS-stratification / on-treatment TAR** | | | | | | | | | | |
| OptumDOD | 5,527 | 65,344 | 2.9 | 67.4 | 18 | 1,838 | 0.25 (0.15 - 0.39) | <0.01 | 0.24 (0.15 - 0.39) | <0.01 |
| MDCR | 2,108 | 36,653 | 1.4 | 36.3 | <5 | 699 | 0.13 (0.03 - 0.34) | <0.01 | 0.11 (0.03 - 0.37) | <0.01 |
| Open Claims | 85,691 | 695,883 | 52.1 | 800.2 | 117 | 9,472 | 0.22 (0.18 - 0.26) | <0.01 | 0.21 (0.16 - 0.28) | <0.01 |
| OptumEHR | 3,524 | 57,838 | 1.0 | 35.0 | <5 | 771 | 0.10 (0.02 - 0.33) | <0.01 | 0.09 (0.02 - 0.38) | <0.01 |
| Meta-analysis | 85,691 | 695,883 | 52.1 | 800.2 | 117 | 9,472 | 0.22 (0.18 - 0.26) | <0.01 | 0.21 (0.16 - 0.28) | <0.01 |
| **PS-matching / on-treatment TAR** | | | | | | | | | | |
| OptumDOD | 5,038 | 54,266 | 2.7 | 54.6 | 16 | 1,393 | 0.23 (0.13 - 0.38) | <0.01 | 0.23 (0.13 - 0.40) | <0.01 |
| MDCR | 1,928 | 28,184 | 1.3 | 28.2 | <5 | 475 | 0.12 (0.02 - 0.37) | <0.01 | 0.10 (0.02 - 0.46) | <0.01 |
| Open Claims | 78,843 | 649,860 | 49.2 | 746.6 | 108 | 8,262 | 0.21 (0.17 - 0.26) | <0.01 | 0.20 (0.16 - 0.27) | <0.01 |
| OptumEHR | 3,246 | 47,264 | 0.9 | 28.0 | <5 | 576 | 0.14 (0.02 - 0.43) | <0.01 | 0.11 (0.02 - 0.54) | <0.01 |
| Meta-analysis | 89,055 | 779,574 | 54.1 | 857.4 | <134 | 10,706 | 0.21 (0.17 - 0.25) | <0.01 | 0.20 (0.15 - 0.26) | <0.01 |
| **PS-stratification / intent-to-treat TAR** | | | | | | | | | | |
| OptumDOD | 5,527 | 65,344 | 8.2 | 203.9 | 66 | 4,714 | 0.36 (0.28 - 0.46) | <0.01 | 0.35 (0.27 - 0.45) | <0.01 |
| MDCR | 2,108 | 36,653 | 4.8 | 110.4 | 24 | 1,938 | 0.34 (0.22 - 0.51) | <0.01 | 0.31 (0.21 - 0.48) | <0.01 |
| Open Claims | 85,691 | 695,883 | 176.6 | 2789.2 | 668 | 31,797 | 0.42 (0.39 - 0.45) | <0.01 | 0.41 (0.32 - 0.52) | <0.01 |
| OptumEHR | 3,524 | 57,838 | 7.2 | 214.0 | 48 | 2,851 | 0.60 (0.44 - 0.80) | <0.01 | 0.60 (0.44 - 0.81) | <0.01 |
| Meta-analysis | 85,691 | 695,883 | 176.6 | 2789.2 | 668 | 31,797 | 0.42 (0.39 - 0.45) | <0.01 | 0.41 (0.32 - 0.52) | <0.01 |
| **PS-matching / intent-to-treat TAR** | | | | | | | | | | |
| OptumDOD | 5,038 | 54,266 | 7.9 | 158.7 | 63 | 3,457 | 0.36 (0.27 - 0.48) | <0.01 | 0.36 (0.27 - 0.49) | <0.01 |
| MDCR | 1,928 | 28,184 | 4.7 | 87.3 | 24 | 1,373 | 0.33 (0.20 - 0.52) | <0.01 | 0.30 (0.19 - 0.49) | <0.01 |
| Open Claims | 78,843 | 649,860 | 170.3 | 2553.9 | 651 | 27,420 | 0.42 (0.38 - 0.46) | <0.01 | 0.41 (0.33 - 0.52) | <0.01 |
| OptumEHR | 3,246 | 47,264 | 6.9 | 164.9 | 46 | 2,067 | 0.57 (0.40 - 0.78) | <0.01 | 0.58 (0.42 - 0.81) | <0.01 |
| Meta-analysis | 89,055 | 779,574 | 189.8 | 2964.8 | 784 | 34,317 | 0.42 (0.36 - 0.49) | <0.01 | 0.41 (0.32 - 0.51) | <0.01 |

##### 6.3.11 Hypotension

**Supplementary Table 154:** Exposures, outcomes and risk estimates for Hypotension in GLP1RA (T) vs SU (C) new-user cohorts under four different propensity score (PS) adjustment / time-at-risk (TAR) designs across data sources. We report total patient exposure time in 1,000 patient-year (KYr) units, uncalibrated and calibrated hazard ratios (HRs) and their 95% confidence intervals (CIs). Greyed out entries did not pass study diagnostics and are included only for completeness.

| | Patients | | Exposure (KYr) | | Outcomes | | Uncalibrated | | Calibrated | |
|---|---|---|---|---|---|---|---|---|---|---|
| | T | C | T | C | T | C | HR (95% CI) | $p$ | HR (95% CI) | $p$ |
| **PS-stratification / on-treatment TAR** | | | | | | | | | | |
| OptumDOD | 5,272 | 61,513 | 2.7 | 63.3 | 76 | 2,001 | 0.96 (0.74 - 1.22) | 0.73 | 0.93 (0.73 - 1.20) | 0.61 |
| MDCR | 2,042 | 34,961 | 1.3 | 34.5 | 26 | 821 | 1.13 (0.72 - 1.70) | 0.59 | 1.00 (0.65 - 1.53) | 0.89 |
| Open Claims | 82,616 | 670,684 | 50.1 | 771.6 | 539 | 10,235 | 1.04 (0.94 - 1.14) | 0.47 | 0.99 (0.79 - 1.24) | 0.90 |
| OptumEHR | 3,445 | 56,150 | 1.0 | 33.6 | 19 | 976 | 0.90 (0.54 - 1.40) | 0.65 | 0.74 (0.46 - 1.20) | 0.26 |
| Meta-analysis | 82,616 | 670,684 | 50.1 | 771.6 | 539 | 10,235 | 1.04 (0.94 - 1.14) | 0.47 | 0.99 (0.79 - 1.25) | 0.90 |
| **PS-matching / on-treatment TAR** | | | | | | | | | | |
| OptumDOD | 4,794 | 51,335 | 2.5 | 51.7 | 71 | 1,509 | 0.97 (0.71 - 1.30) | 0.82 | 0.97 (0.71 - 1.31) | 0.82 |
| MDCR | 1,865 | 26,924 | 1.3 | 26.9 | 26 | 532 | 1.11 (0.67 - 1.76) | 0.67 | 0.99 (0.61 - 1.61) | 0.88 |
| Open Claims | 75,965 | 625,422 | 47.3 | 718.6 | 495 | 8,771 | 0.98 (0.87 - 1.10) | 0.73 | 0.95 (0.78 - 1.15) | 0.64 |
| OptumEHR | 3,164 | 45,943 | 0.9 | 26.9 | 18 | 671 | 0.98 (0.57 - 1.60) | 0.94 | 0.78 (0.35 - 1.73) | 0.59 |
| Meta-analysis | 85,788 | 749,624 | 52.0 | 824.1 | 610 | 11,483 | 0.98 (0.89 - 1.09) | 0.76 | 0.94 (0.77 - 1.15) | 0.54 |
| **PS-stratification / intent-to-treat TAR** | | | | | | | | | | |
| OptumDOD | 5,272 | 61,513 | 7.6 | 189.4 | 233 | 6,676 | 1.00 (0.87 - 1.15) | 1.00 | 0.98 (0.85 - 1.14) | 0.82 |
| MDCR | 2,042 | 34,961 | 4.6 | 103.8 | 98 | 3,279 | 0.88 (0.71 - 1.09) | 0.25 | 0.80 (0.65 - 1.00) | 0.06 |
| Open Claims | 82,616 | 670,684 | 168.5 | 2683.5 | 2,115 | 44,836 | 0.99 (0.94 - 1.03) | 0.54 | 0.97 (0.77 - 1.21) | 0.77 |
| OptumEHR | 3,445 | 56,150 | 6.9 | 204.6 | 111 | 4,255 | 1.03 (0.84 - 1.25) | 0.76 | 1.03 (0.84 - 1.27) | 0.78 |
| Meta-analysis | 82,616 | 670,684 | 168.5 | 2683.5 | 2,115 | 44,836 | 0.99 (0.94 - 1.03) | 0.54 | 0.97 (0.77 - 1.22) | 0.77 |
| **PS-matching / intent-to-treat TAR** | | | | | | | | | | |
| OptumDOD | 4,794 | 51,335 | 7.3 | 148.6 | 224 | 4,847 | 1.04 (0.89 - 1.21) | 0.62 | 1.05 (0.89 - 1.23) | 0.61 |
| MDCR | 1,865 | 26,924 | 4.5 | 82.5 | 98 | 2,272 | 0.90 (0.70 - 1.14) | 0.38 | 0.82 (0.64 - 1.05) | 0.14 |
| Open Claims | 75,965 | 625,422 | 162.3 | 2452.3 | 2,024 | 38,035 | 0.99 (0.94 - 1.05) | 0.83 | 0.98 (0.79 - 1.21) | 0.85 |
| OptumEHR | 3,164 | 45,943 | 6.6 | 158.4 | 107 | 2,941 | 1.02 (0.81 - 1.26) | 0.88 | 1.04 (0.83 - 1.31) | 0.74 |
| Meta-analysis | 85,788 | 749,624 | 180.6 | 2841.7 | 2,453 | 48,095 | 1.00 (0.95 - 1.04) | 0.85 | 0.97 (0.81 - 1.15) | 0.70 |

##### 6.3.12 Joint pain

**Supplementary Table 155:** Exposures, outcomes and risk estimates for Joint pain in GLP1RA (T) vs SU (C) new-user cohorts under four different propensity score (PS) adjustment / time-at-risk (TAR) designs across data sources. We report total patient exposure time in 1,000 patient-year (KYr) units, uncalibrated and calibrated hazard ratios (HRs) and their 95% confidence intervals (CIs). Greyed out entries did not pass study diagnostics and are included only for completeness.

| | Patients | | Exposure (KYr) | | Outcomes | | Uncalibrated | | Calibrated | |
|---|---|---|---|---|---|---|---|---|---|---|
| | T | C | T | C | T | C | HR (95% CI) | $p$ | HR (95% CI) | $p$ |
| **PS-stratification / on-treatment TAR** | | | | | | | | | | |
| OptumDOD | 5,149 | 62,677 | 2.6 | 65.0 | 48 | 815 | 1.12 (0.81 - 1.53) | 0.47 | 1.10 (0.80 - 1.52) | 0.58 |
| MDCR | 2,007 | 35,645 | 1.3 | 35.4 | 8 | 311 | 0.54 (0.24 - 1.06) | 0.11 | 0.48 (0.23 - 1.01) | 0.06 |
| Open Claims | 79,565 | 668,637 | 48.6 | 771.7 | 314 | 4,519 | 0.96 (0.85 - 1.09) | 0.52 | 0.92 (0.72 - 1.17) | 0.50 |
| OptumEHR | 3,316 | 55,792 | 0.9 | 33.7 | 11 | 345 | 1.25 (0.63 - 2.24) | 0.49 | 1.04 (0.55 - 1.97) | 0.87 |
| Meta-analysis | 79,565 | 668,637 | 48.6 | 771.7 | 314 | 4,519 | 0.96 (0.85 - 1.09) | 0.52 | 0.92 (0.72 - 1.17) | 0.50 |
| **PS-matching / on-treatment TAR** | | | | | | | | | | |
| OptumDOD | 4,732 | 51,869 | 2.5 | 52.5 | 45 | 665 | 1.16 (0.76 - 1.71) | 0.48 | 1.16 (0.77 - 1.73) | 0.49 |
| MDCR | 1,835 | 27,180 | 1.3 | 27.3 | 6 | 242 | 0.28 (0.07 - 0.78) | 0.04 | 0.25 (0.07 - 0.85) | 0.03 |
| Open Claims | 73,542 | 622,035 | 46.1 | 717.2 | 287 | 4,126 | 0.92 (0.78 - 1.07) | 0.27 | 0.89 (0.71 - 1.11) | 0.34 |
| OptumEHR | 3,052 | 45,476 | 0.9 | 26.9 | 10 | 273 | 1.12 (0.52 - 2.21) | 0.76 | 0.90 (0.35 - 2.30) | 0.83 |
| Meta-analysis | 83,161 | 746,560 | 50.7 | 823.9 | 348 | 5,306 | 0.94 (0.81 - 1.08) | 0.36 | 0.89 (0.72 - 1.12) | 0.32 |
| **PS-stratification / intent-to-treat TAR** | | | | | | | | | | |
| OptumDOD | 5,149 | 62,677 | 7.5 | 199.8 | 146 | 2,453 | 1.21 (1.00 - 1.44) | 0.04 | 1.19 (0.98 - 1.43) | 0.10 |
| MDCR | 2,007 | 35,645 | 4.6 | 109.0 | 54 | 1,048 | 1.05 (0.77 - 1.40) | 0.75 | 0.96 (0.71 - 1.29) | 0.78 |
| Open Claims | 79,565 | 668,637 | 164.3 | 2714.4 | 1,178 | 17,056 | 1.03 (0.96 - 1.10) | 0.41 | 1.01 (0.80 - 1.27) | 0.91 |
| OptumEHR | 3,316 | 55,792 | 6.7 | 209.0 | 56 | 1,661 | 0.96 (0.72 - 1.26) | 0.77 | 0.96 (0.72 - 1.28) | 0.79 |
| Meta-analysis | 79,565 | 668,637 | 164.3 | 2714.4 | 1,178 | 17,056 | 1.03 (0.96 - 1.10) | 0.41 | 1.01 (0.80 - 1.27) | 0.91 |
| **PS-matching / intent-to-treat TAR** | | | | | | | | | | |
| OptumDOD | 4,732 | 51,869 | 7.3 | 154.3 | 138 | 1,955 | 1.11 (0.89 - 1.37) | 0.34 | 1.12 (0.90 - 1.39) | 0.34 |
| MDCR | 1,835 | 27,180 | 4.5 | 85.5 | 52 | 832 | 0.95 (0.66 - 1.34) | 0.78 | 0.87 (0.61 - 1.24) | 0.46 |
| Open Claims | 73,542 | 622,035 | 158.7 | 2471.3 | 1,115 | 15,340 | 1.05 (0.97 - 1.12) | 0.22 | 1.03 (0.83 - 1.28) | 0.80 |
| OptumEHR | 3,052 | 45,476 | 6.4 | 160.1 | 53 | 1,326 | 0.92 (0.66 - 1.24) | 0.59 | 0.94 (0.68 - 1.29) | 0.73 |
| Meta-analysis | 83,161 | 746,560 | 176.9 | 2871.2 | 1,358 | 19,453 | 1.04 (0.98 - 1.11) | 0.22 | 1.01 (0.85 - 1.21) | 0.88 |

##### 6.3.13 Lower extremity amputation

**Supplementary Table 156:** Exposures, outcomes and risk estimates for Lower extremity amputation in GLP1RA (T) vs SU (C) new-user cohorts under four different propensity score (PS) adjustment / time-at-risk (TAR) designs across data sources. We report total patient exposure time in 1,000 patient-year (KYr) units, uncalibrated and calibrated hazard ratios (HRs) and their 95% confidence intervals (CIs). Greyed out entries did not pass study diagnostics and are included only for completeness.

| | Patients | | Exposure (KYr) | | Outcomes | | Uncalibrated | | Calibrated | |
|---|---|---|---|---|---|---|---|---|---|---|
| | T | C | T | C | T | C | HR (95% CI) | $p$ | HR (95% CI) | $p$ |
| **PS-stratification / on-treatment TAR** | | | | | | | | | | |
| OptumDOD | 5,613 | 66,153 | 2.9 | 69.0 | <5 | 94 | 0.56 (0.09 - 1.94) | 0.46 | 0.55 (0.12 - 2.56) | 0.44 |
| MDCR | 2,133 | 36,894 | 1.4 | 36.7 | 0 | 42 | | | | |
| Open Claims | 87,292 | 705,868 | 53.0 | 815.0 | 16 | 532 | 0.67 (0.38 - 1.08) | 0.13 | 0.64 (0.36 - 1.12) | 0.12 |
| OptumEHR | 3,570 | 58,305 | 1.0 | 35.8 | 0 | 45 | | | | |
| Meta-analysis | 87,292 | 705,868 | 53.0 | 815.0 | 16 | 532 | 0.67 (0.38 - 1.08) | 0.13 | 0.64 (0.36 - 1.12) | 0.12 |
| **PS-matching / on-treatment TAR** | | | | | | | | | | |
| OptumDOD | 5,103 | 54,852 | 2.7 | 55.8 | <5 | 74 | 0.00 (0.00 - 0.71) | 0.20 | 0.00 (0.00 - 25895) | 0.20 |
| MDCR | 1,947 | 28,351 | 1.3 | 28.6 | 0 | 33 | | | | |
| Open Claims | 80,154 | 658,385 | 50.1 | 759.2 | 16 | 451 | 0.72 (0.38 - 1.27) | 0.29 | 0.70 (0.38 - 1.31) | 0.27 |
| OptumEHR | 3,280 | 47,533 | 0.9 | 28.5 | 0 | 28 | | | | |
| Meta-analysis | 85,257 | 713,237 | 52.8 | 815.0 | <21 | 525 | | 0.60 | | 0.60 |
| **PS-stratification / intent-to-treat TAR** | | | | | | | | | | |
| OptumDOD | 5,613 | 66,153 | 8.4 | 216.0 | 14 | 372 | 1.42 (0.77 - 2.42) | 0.24 | 1.39 (0.78 - 2.48) | 0.27 |
| MDCR | 2,133 | 36,894 | 4.9 | 114.5 | <5 | 177 | 0.69 (0.20 - 1.74) | 0.49 | 0.63 (0.22 - 1.83) | 0.40 |
| Open Claims | 87,292 | 705,868 | 181.1 | 2901.4 | 97 | 2,583 | 0.88 (0.71 - 1.09) | 0.25 | 0.86 (0.63 - 1.18) | 0.36 |
| OptumEHR | 3,570 | 58,305 | 7.4 | 222.5 | <5 | 236 | 0.70 (0.21 - 1.76) | 0.51 | 0.70 (0.24 - 2.04) | 0.51 |
| Meta-analysis | 87,292 | 705,868 | 181.1 | 2901.4 | 97 | 2,583 | 0.88 (0.71 - 1.09) | 0.25 | 0.86 (0.63 - 1.18) | 0.36 |
| **PS-matching / intent-to-treat TAR** | | | | | | | | | | |
| OptumDOD | 5,103 | 54,852 | 8.0 | 167.2 | 14 | 266 | 1.52 (0.77 - 2.75) | 0.20 | 1.53 (0.81 - 2.90) | 0.19 |
| MDCR | 1,947 | 28,351 | 4.8 | 90.4 | <5 | 121 | 1.02 (0.29 - 2.72) | 0.98 | 0.93 (0.30 - 2.85) | 0.90 |
| Open Claims | 80,154 | 658,385 | 174.4 | 2648.0 | 93 | 2,189 | 0.91 (0.71 - 1.14) | 0.42 | 0.89 (0.65 - 1.22) | 0.49 |
| OptumEHR | 3,280 | 47,533 | 7.0 | 170.6 | <5 | 145 | 0.81 (0.24 - 2.08) | 0.70 | 0.83 (0.28 - 2.47) | 0.74 |
| Meta-analysis | 90,484 | 789,121 | 194.3 | 3076.3 | <117 | 2,721 | 0.99 (0.75 - 1.30) | 0.93 | 0.96 (0.70 - 1.32) | 0.80 |

###### 6.3.14 Nausea

**Supplementary Table 157:** Exposures, outcomes and risk estimates for Nausea in GLP1RA (T) vs SU (C) new-user cohorts under four different propensity score (PS) adjustment / time-at-risk (TAR) designs across data sources. We report total patient exposure time in 1,000 patient-year (KYr) units, uncalibrated and calibrated hazard ratios (HRs) and their 95% confidence intervals (CIs). Greyed out entries did not pass study diagnostics and are included only for completeness.

| | Patients | | Exposure (KYr) | | Outcomes | | Uncalibrated | | Calibrated | |
|---|---|---|---|---|---|---|---|---|---|---|
| | T | C | T | C | T | C | HR (95% CI) | $p$ | HR (95% CI) | $p$ |
| **PS-stratification / on-treatment TAR** | | | | | | | | | | |
| OptumDOD | 4,695 | 56,566 | 2.4 | 57.8 | 178 | 2,589 | 1.52 (1.28 - 1.80) | <0.01 | 1.49 (1.25 - 1.77) | <0.01 |
| MDCR | 1,932 | 32,962 | 1.2 | 32.4 | 50 | 1,150 | 1.05 (0.76 - 1.42) | 0.74 | 0.93 (0.68 - 1.27) | 0.67 |
| Open Claims | 72,632 | 614,797 | 44.1 | 708.4 | 1,274 | 14,961 | 1.39 (1.31 - 1.49) | <0.01 | 1.34 (1.08 - 1.65) | 0.01 |
| OptumEHR | 3,189 | 53,253 | 0.9 | 31.4 | 61 | 1,194 | 2.08 (1.56 - 2.72) | <0.01 | 1.73 (1.30 - 2.30) | <0.01 |
| Meta-analysis | 72,632 | 614,797 | 44.1 | 708.4 | 1,274 | 14,961 | 1.39 (1.31 - 1.49) | <0.01 | 1.34 (1.07 - 1.66) | <0.01 |
| **PS-matching / on-treatment TAR** | | | | | | | | | | |
| OptumDOD | 4,285 | 46,818 | 2.2 | 46.6 | 163 | 2,003 | 1.59 (1.30 - 1.94) | <0.01 | 1.59 (1.30 - 1.96) | <0.01 |
| MDCR | 1,775 | 25,420 | 1.2 | 25.3 | 48 | 851 | 1.02 (0.69 - 1.46) | 0.93 | 0.91 (0.62 - 1.33) | 0.64 |
| Open Claims | 67,082 | 572,010 | 41.7 | 657.9 | 1,173 | 13,392 | 1.46 (1.35 - 1.58) | <0.01 | 1.42 (1.19 - 1.69) | <0.01 |
| OptumEHR | 2,932 | 43,666 | 0.8 | 25.3 | 56 | 889 | 2.08 (1.47 - 2.88) | <0.01 | 1.66 (0.83 - 3.31) | 0.22 |
| Meta-analysis | 76,074 | 687,914 | 46.0 | 755.1 | 1,440 | 17,135 | 1.51 (1.21 - 1.88) | <0.01 | 1.44 (1.09 - 1.90) | 0.01 |
| **PS-stratification / intent-to-treat TAR** | | | | | | | | | | |
| OptumDOD | 4,695 | 56,566 | 6.4 | 167.7 | 406 | 7,645 | 1.29 (1.15 - 1.43) | <0.01 | 1.26 (1.13 - 1.42) | <0.01 |
| MDCR | 1,932 | 32,962 | 4.2 | 95.4 | 170 | 3,934 | 0.93 (0.78 - 1.09) | 0.38 | 0.85 (0.71 - 1.00) | 0.07 |
| Open Claims | 72,632 | 614,797 | 145.7 | 2409.1 | 3,699 | 57,532 | 1.11 (1.07 - 1.15) | <0.01 | 1.09 (0.87 - 1.37) | 0.46 |
| OptumEHR | 3,189 | 53,253 | 6.2 | 190.7 | 200 | 5,087 | 1.28 (1.09 - 1.49) | <0.01 | 1.28 (1.09 - 1.50) | <0.01 |
| Meta-analysis | 72,632 | 614,797 | 145.7 | 2409.1 | 3,699 | 57,532 | 1.11 (1.07 - 1.15) | <0.01 | 1.09 (0.87 - 1.37) | 0.45 |
| **PS-matching / intent-to-treat TAR** | | | | | | | | | | |
| OptumDOD | 4,285 | 46,818 | 6.2 | 130.2 | 384 | 5,800 | 1.38 (1.22 - 1.55) | <0.01 | 1.39 (1.22 - 1.57) | <0.01 |
| MDCR | 1,775 | 25,420 | 4.1 | 75.6 | 163 | 3,007 | 0.90 (0.74 - 1.09) | 0.30 | 0.83 (0.68 - 1.01) | 0.08 |
| Open Claims | 67,082 | 572,010 | 140.7 | 2195.5 | 3,513 | 50,827 | 1.12 (1.08 - 1.17) | <0.01 | 1.11 (0.90 - 1.37) | 0.35 |
| OptumEHR | 2,932 | 43,666 | 5.9 | 148.2 | 189 | 3,779 | 1.28 (1.07 - 1.51) | <0.01 | 1.31 (1.10 - 1.56) | <0.01 |
| Meta-analysis | 76,074 | 687,914 | 156.8 | 2549.5 | 4,249 | 63,413 | 1.16 (0.98 - 1.38) | 0.08 | 1.13 (0.89 - 1.43) | 0.31 |

##### 6.3.15 Peripheral edema

**Supplementary Table 158:** Exposures, outcomes and risk estimates for Peripheral edema in GLP1RA (T) vs SU (C) new-user cohorts under four different propensity score (PS) adjustment / time-at-risk (TAR) designs across data sources. We report total patient exposure time in 1,000 patient-year (KYr) units, uncalibrated and calibrated hazard ratios (HRs) and their 95% confidence intervals (CIs). Greyed out entries did not pass study diagnostics and are included only for completeness.

| | Patients | | Exposure (KYr) | | Outcomes | | Uncalibrated | | Calibrated | |
|---|---|---|---|---|---|---|---|---|---|---|
| | T | C | T | C | T | C | HR (95% CI) | $p$ | HR (95% CI) | $p$ |
| **PS-stratification / on-treatment TAR** | | | | | | | | | | |
| OptumDOD | 3,920 | 48,322 | 2.0 | 46.6 | 136 | 4,879 | 0.66 (0.55 - 0.79) | <0.01 | 0.64 (0.54 - 0.77) | <0.01 |
| MDCR | 1,587 | 27,977 | 1.0 | 26.1 | 62 | 2,399 | 0.75 (0.57 - 0.97) | 0.04 | 0.66 (0.51 - 0.87) | <0.01 |
| Open Claims | 58,785 | 526,028 | 35.7 | 589.6 | 1,044 | 28,102 | 0.75 (0.70 - 0.80) | <0.01 | 0.72 (0.58 - 0.89) | <0.01 |
| OptumEHR | 2,721 | 47,402 | 0.7 | 26.3 | 37 | 2,271 | 0.72 (0.50 - 0.99) | 0.05 | 0.59 (0.42 - 0.84) | <0.01 |
| Meta-analysis | 58,785 | 526,028 | 35.7 | 589.6 | 1,044 | 28,102 | 0.75 (0.70 - 0.80) | <0.01 | 0.72 (0.58 - 0.90) | <0.01 |
| **PS-matching / on-treatment TAR** | | | | | | | | | | |
| OptumDOD | 3,601 | 40,002 | 1.9 | 37.7 | 126 | 3,746 | 0.65 (0.52 - 0.80) | <0.01 | 0.65 (0.52 - 0.81) | <0.01 |
| MDCR | 1,475 | 21,406 | 1.0 | 20.2 | 56 | 1,703 | 0.69 (0.48 - 0.96) | 0.03 | 0.62 (0.43 - 0.87) | <0.01 |
| Open Claims | 54,897 | 488,033 | 34.1 | 546.5 | 989 | 24,554 | 0.73 (0.68 - 0.79) | <0.01 | 0.71 (0.60 - 0.85) | <0.01 |
| OptumEHR | 2,528 | 38,649 | 0.7 | 21.2 | 35 | 1,629 | 0.71 (0.47 - 1.03) | 0.08 | 0.57 (0.28 - 1.16) | 0.18 |
| Meta-analysis | 62,501 | 588,090 | 37.6 | 625.7 | 1,206 | 31,632 | 0.72 (0.67 - 0.77) | <0.01 | 0.69 (0.58 - 0.83) | <0.01 |
| **PS-stratification / intent-to-treat TAR** | | | | | | | | | | |
| OptumDOD | 3,920 | 48,322 | 5.2 | 128.5 | 428 | 12,466 | 0.83 (0.75 - 0.92) | <0.01 | 0.82 (0.73 - 0.91) | <0.01 |
| MDCR | 1,587 | 27,977 | 3.1 | 72.7 | 275 | 7,000 | 0.99 (0.87 - 1.12) | 0.88 | 0.90 (0.79 - 1.03) | 0.18 |
| Open Claims | 58,785 | 526,028 | 115.7 | 1947.1 | 4,173 | 94,563 | 0.91 (0.88 - 0.94) | <0.01 | 0.89 (0.71 - 1.11) | 0.32 |
| OptumEHR | 2,721 | 47,402 | 5.2 | 155.1 | 227 | 9,366 | 0.91 (0.79 - 1.04) | 0.16 | 0.91 (0.78 - 1.05) | 0.27 |
| Meta-analysis | 58,785 | 526,028 | 115.7 | 1947.1 | 4,173 | 94,563 | 0.91 (0.88 - 0.94) | <0.01 | 0.89 (0.71 - 1.12) | 0.32 |
| **PS-matching / intent-to-treat TAR** | | | | | | | | | | |
| OptumDOD | 3,601 | 40,002 | 5.0 | 100.1 | 411 | 9,357 | 0.84 (0.75 - 0.94) | <0.01 | 0.84 (0.75 - 0.95) | 0.01 |
| MDCR | 1,475 | 21,406 | 3.1 | 57.2 | 266 | 5,168 | 0.94 (0.81 - 1.10) | 0.46 | 0.87 (0.74 - 1.01) | 0.10 |
| Open Claims | 54,897 | 488,033 | 112.3 | 1773.3 | 4,036 | 81,846 | 0.90 (0.87 - 0.94) | <0.01 | 0.89 (0.72 - 1.10) | 0.28 |
| OptumEHR | 2,528 | 38,649 | 5.0 | 120.6 | 221 | 6,716 | 0.90 (0.77 - 1.04) | 0.17 | 0.92 (0.79 - 1.08) | 0.43 |
| Meta-analysis | 62,501 | 588,090 | 125.3 | 2051.1 | 4,934 | 103,087 | 0.90 (0.87 - 0.93) | <0.01 | 0.87 (0.74 - 1.03) | 0.12 |

##### 6.3.16 Photosensitivity

**Supplementary Table 159:** Exposures, outcomes and risk estimates for Photosensitivity in GLP1RA (T) vs SU (C) new-user cohorts under four different propensity score (PS) adjustment / time-at-risk (TAR) designs across data sources. We report total patient exposure time in 1,000 patient-year (KYr) units, uncalibrated and calibrated hazard ratios (HRs) and their 95% confidence intervals (CIs). Greyed out entries did not pass study diagnostics and are included only for completeness.

| | Patients | | Exposure (KYr) | | Outcomes | | Uncalibrated | | Calibrated | |
|---|---|---|---|---|---|---|---|---|---|---|
| | T | C | T | C | T | C | HR (95% CI) | $p$ | HR (95% CI) | $p$ |
| **PS-stratification / on-treatment TAR** | | | | | | | | | | |
| OptumDOD | 5,575 | 65,490 | 2.9 | 68.2 | <5 | 122 | 1.51 (0.36 - 4.25) | 0.52 | 1.47 (0.43 - 5.08) | 0.54 |
| MDCR | 2,092 | 36,363 | 1.4 | 36.1 | <5 | 116 | 0.84 (0.20 - 2.43) | 0.79 | 0.75 (0.21 - 2.62) | 0.65 |
| Open Claims | 85,977 | 697,868 | 52.1 | 804.4 | 21 | 433 | 1.76 (1.05 - 2.78) | 0.02 | 1.68 (1.00 - 2.85) | 0.05 |
| OptumEHR | 3,534 | 57,741 | 1.0 | 35.3 | <5 | 84 | 4.59 (1.34 - 11.8) | <0.01 | 3.81 (1.28 - 11.3) | 0.02 |
| Meta-analysis | 85,977 | 697,868 | 52.1 | 804.4 | 21 | 433 | 1.76 (1.05 - 2.78) | 0.02 | 1.68 (0.99 - 2.86) | 0.05 |
| **PS-matching / on-treatment TAR** | | | | | | | | | | |
| OptumDOD | 5,067 | 54,291 | 2.7 | 55.1 | <5 | 81 | 1.88 (0.41 - 6.00) | 0.36 | 1.88 (0.49 - 7.23) | 0.36 |
| MDCR | 1,909 | 27,815 | 1.3 | 27.9 | <5 | 79 | 1.30 (0.30 - 3.97) | 0.69 | 1.17 (0.32 - 4.25) | 0.82 |
| Open Claims | 79,003 | 650,817 | 49.2 | 749.3 | 19 | 365 | 1.66 (0.92 - 2.83) | 0.08 | 1.61 (0.90 - 2.89) | 0.12 |
| OptumEHR | 3,251 | 47,209 | 0.9 | 28.3 | <5 | 52 | 3.44 (0.89 - 10.3) | 0.05 | 2.75 (0.70 - 10.8) | 0.17 |
| Meta-analysis | 85,979 | 732,923 | 53.2 | 832.3 | <29 | 525 | 1.63 (1.01 - 2.64) | 0.05 | 1.56 (0.93 - 2.60) | 0.09 |
| **PS-stratification / intent-to-treat TAR** | | | | | | | | | | |
| OptumDOD | 5,575 | 65,490 | 8.3 | 212.8 | 10 | 305 | 1.14 (0.54 - 2.12) | 0.71 | 1.12 (0.57 - 2.21) | 0.75 |
| MDCR | 2,092 | 36,363 | 4.8 | 112.1 | 8 | 369 | 0.52 (0.23 - 1.01) | 0.08 | 0.47 (0.22 - 0.99) | 0.05 |
| Open Claims | 85,977 | 697,868 | 178.2 | 2867.8 | 43 | 994 | 1.12 (0.80 - 1.54) | 0.49 | 1.10 (0.74 - 1.64) | 0.64 |
| OptumEHR | 3,534 | 57,741 | 7.3 | 219.7 | 12 | 327 | 2.15 (1.11 - 3.78) | 0.01 | 2.15 (1.17 - 3.97) | 0.02 |
| Meta-analysis | 85,977 | 697,868 | 178.2 | 2867.8 | 43 | 994 | 1.12 (0.80 - 1.54) | 0.49 | 1.10 (0.74 - 1.64) | 0.64 |
| **PS-matching / intent-to-treat TAR** | | | | | | | | | | |
| OptumDOD | 5,067 | 54,291 | 8.0 | 164.7 | 10 | 213 | 1.34 (0.63 - 2.54) | 0.41 | 1.35 (0.67 - 2.71) | 0.40 |
| MDCR | 1,909 | 27,815 | 4.7 | 88.1 | 8 | 268 | 0.53 (0.22 - 1.10) | 0.13 | 0.49 (0.22 - 1.10) | 0.08 |
| Open Claims | 79,003 | 650,817 | 171.7 | 2616.6 | 41 | 850 | 1.02 (0.70 - 1.44) | 0.91 | 1.00 (0.66 - 1.52) | 0.95 |
| OptumEHR | 3,251 | 47,209 | 7.0 | 169.1 | 12 | 230 | 2.46 (1.24 - 4.42) | <0.01 | 2.52 (1.34 - 4.76) | <0.01 |
| Meta-analysis | 89,230 | 780,132 | 191.3 | 3038.5 | 71 | 1,561 | 1.18 (0.67 - 2.09) | 0.57 | 1.15 (0.63 - 2.08) | 0.66 |

###### 6.3.17 Venous thromboembolism

**Supplementary Table 160:** Exposures, outcomes and risk estimates for Venous thromboembolism in GLP1RA (T) vs SU (C) new-user cohorts under four different propensity score (PS) adjustment / time-at-risk (TAR) designs across data sources. We report total patient exposure time in 1,000 patient-year (KYr) units, uncalibrated and calibrated hazard ratios (HRs) and their 95% confidence intervals (CIs). Greyed out entries did not pass study diagnostics and are included only for completeness.

| | Patients | | Exposure (KYr) | | Outcomes | | Uncalibrated | | Calibrated | |
|---|---|---|---|---|---|---|---|---|---|---|
| | T | C | T | C | T | C | HR (95% CI) | $p$ | HR (95% CI) | $p$ |
| **PS-stratification / on-treatment TAR** | | | | | | | | | | |
| OptumDOD | 5,345 | 63,537 | 2.8 | 65.8 | 30 | 999 | 0.88 (0.58 - 1.28) | 0.52 | 0.86 (0.58 - 1.27) | 0.45 |
| MDCR | 2,043 | 35,380 | 1.3 | 35.0 | 15 | 551 | 0.84 (0.47 - 1.42) | 0.55 | 0.75 (0.43 - 1.30) | 0.31 |
| Open Claims | 82,626 | 676,546 | 50.3 | 778.8 | 269 | 6,395 | 0.78 (0.68 - 0.89) | <0.01 | 0.75 (0.59 - 0.95) | 0.02 |
| OptumEHR | 3,430 | 56,341 | 1.0 | 34.0 | 13 | 497 | 1.34 (0.71 - 2.29) | 0.33 | 1.11 (0.62 - 1.99) | 0.73 |
| Meta-analysis | 82,626 | 676,546 | 50.3 | 778.8 | 269 | 6,395 | 0.78 (0.68 - 0.89) | <0.01 | 0.75 (0.58 - 0.95) | 0.02 |
| **PS-matching / on-treatment TAR** | | | | | | | | | | |
| OptumDOD | 4,865 | 52,372 | 2.6 | 52.7 | 28 | 720 | 0.89 (0.55 - 1.39) | 0.64 | 0.90 (0.56 - 1.43) | 0.65 |
| MDCR | 1,868 | 27,207 | 1.2 | 27.2 | 15 | 387 | 0.82 (0.41 - 1.51) | 0.55 | 0.73 (0.38 - 1.41) | 0.36 |
| Open Claims | 76,024 | 630,961 | 47.6 | 725.3 | 255 | 5,551 | 0.78 (0.67 - 0.91) | <0.01 | 0.76 (0.61 - 0.95) | 0.02 |
| OptumEHR | 3,157 | 46,144 | 0.9 | 27.3 | 11 | 355 | 1.36 (0.66 - 2.52) | 0.37 | 1.08 (0.44 - 2.66) | 0.86 |
| Meta-analysis | 85,914 | 756,684 | 52.3 | 832.6 | 309 | 7,013 | 0.81 (0.71 - 0.93) | <0.01 | 0.78 (0.62 - 0.97) | 0.02 |
| **PS-stratification / intent-to-treat TAR** | | | | | | | | | | |
| OptumDOD | 5,345 | 63,537 | 7.9 | 202.1 | 97 | 3,186 | 0.88 (0.71 - 1.09) | 0.26 | 0.87 (0.70 - 1.08) | 0.24 |
| MDCR | 2,043 | 35,380 | 4.6 | 106.9 | 73 | 1,954 | 0.97 (0.75 - 1.24) | 0.82 | 0.89 (0.69 - 1.14) | 0.37 |
| Open Claims | 82,626 | 676,546 | 170.2 | 2743.2 | 1,192 | 24,873 | 0.93 (0.88 - 0.99) | 0.03 | 0.91 (0.72 - 1.15) | 0.45 |
| OptumEHR | 3,430 | 56,341 | 7.0 | 210.7 | 58 | 2,056 | 0.98 (0.73 - 1.28) | 0.87 | 0.98 (0.74 - 1.29) | 0.86 |
| Meta-analysis | 82,626 | 676,546 | 170.2 | 2743.2 | 1,192 | 24,873 | 0.93 (0.88 - 0.99) | 0.03 | 0.91 (0.72 - 1.15) | 0.45 |
| **PS-matching / intent-to-treat TAR** | | | | | | | | | | |
| OptumDOD | 4,865 | 52,372 | 7.5 | 155.2 | 92 | 2,268 | 0.95 (0.74 - 1.20) | 0.69 | 0.96 (0.75 - 1.22) | 0.74 |
| MDCR | 1,868 | 27,207 | 4.4 | 84.4 | 73 | 1,417 | 0.92 (0.68 - 1.22) | 0.57 | 0.84 (0.62 - 1.13) | 0.27 |
| Open Claims | 76,024 | 630,961 | 164.0 | 2504.9 | 1,152 | 21,478 | 0.93 (0.86 - 0.99) | 0.03 | 0.91 (0.73 - 1.14) | 0.42 |
| OptumEHR | 3,157 | 46,144 | 6.7 | 162.9 | 56 | 1,486 | 0.94 (0.69 - 1.26) | 0.69 | 0.97 (0.71 - 1.31) | 0.83 |
| Meta-analysis | 85,914 | 756,684 | 182.7 | 2907.4 | 1,373 | 26,649 | 0.93 (0.87 - 0.99) | 0.02 | 0.90 (0.75 - 1.08) | 0.25 |

##### 6.3.18 Vomiting

**Supplementary Table 161:** Exposures, outcomes and risk estimates for Vomiting in GLP1RA (T) vs SU (C) new-user cohorts under four different propensity score (PS) adjustment / time-at-risk (TAR) designs across data sources. We report total patient exposure time in 1,000 patient-year (KYr) units, uncalibrated and calibrated hazard ratios (HRs) and their 95% confidence intervals (CIs). Greyed out entries did not pass study diagnostics and are included only for completeness.

| | Patients | | Exposure (KYr) | | Outcomes | | Uncalibrated | | Calibrated | |
|---|---|---|---|---|---|---|---|---|---|---|
| | T | C | T | C | T | C | HR (95% CI) | $p$ | HR (95% CI) | $p$ |
| **PS-stratification / on-treatment TAR** | | | | | | | | | | |
| OptumDOD | 5,030 | 59,035 | 2.6 | 60.8 | 130 | 2,033 | 1.40 (1.14 - 1.70) | <0.01 | 1.37 (1.12 - 1.67) | <0.01 |
| MDCR | 2,000 | 33,833 | 1.3 | 33.3 | 40 | 994 | 1.05 (0.73 - 1.46) | 0.78 | 0.93 (0.66 - 1.32) | 0.69 |
| Open Claims | 77,029 | 635,649 | 46.7 | 733.6 | 933 | 11,905 | 1.36 (1.26 - 1.46) | <0.01 | 1.30 (1.05 - 1.62) | 0.02 |
| OptumEHR | 3,338 | 55,066 | 0.9 | 33.0 | 35 | 836 | 1.83 (1.25 - 2.59) | <0.01 | 1.52 (1.05 - 2.20) | 0.04 |
| Meta-analysis | 77,029 | 635,649 | 46.7 | 733.6 | 933 | 11,905 | 1.36 (1.26 - 1.46) | <0.01 | 1.30 (1.04 - 1.63) | 0.02 |
| **PS-matching / on-treatment TAR** | | | | | | | | | | |
| OptumDOD | 4,567 | 49,167 | 2.4 | 49.3 | 120 | 1,557 | 1.49 (1.16 - 1.89) | <0.01 | 1.49 (1.17 - 1.90) | <0.01 |
| MDCR | 1,838 | 26,183 | 1.2 | 26.1 | 39 | 745 | 1.05 (0.69 - 1.55) | 0.81 | 0.94 (0.63 - 1.41) | 0.77 |
| Open Claims | 70,903 | 593,021 | 44.2 | 683.1 | 869 | 10,582 | 1.39 (1.27 - 1.52) | <0.01 | 1.35 (1.13 - 1.62) | <0.01 |
| OptumEHR | 3,065 | 45,087 | 0.9 | 26.5 | 34 | 608 | 1.86 (1.21 - 2.79) | <0.01 | 1.49 (0.71 - 3.10) | 0.36 |
| Meta-analysis | 80,373 | 713,458 | 48.6 | 785.0 | 1,062 | 13,492 | 1.40 (1.29 - 1.52) | <0.01 | 1.34 (1.11 - 1.62) | <0.01 |
| **PS-stratification / intent-to-treat TAR** | | | | | | | | | | |
| OptumDOD | 5,030 | 59,035 | 7.0 | 178.7 | 302 | 6,395 | 1.18 (1.04 - 1.34) | <0.01 | 1.16 (1.02 - 1.32) | 0.05 |
| MDCR | 2,000 | 33,833 | 4.4 | 98.9 | 133 | 3,463 | 0.89 (0.74 - 1.07) | 0.22 | 0.81 (0.67 - 0.98) | 0.04 |
| Open Claims | 77,029 | 635,649 | 155.8 | 2517.0 | 2,788 | 48,308 | 1.07 (1.03 - 1.12) | <0.01 | 1.05 (0.84 - 1.32) | 0.67 |
| OptumEHR | 3,338 | 55,066 | 6.6 | 200.7 | 126 | 3,852 | 1.16 (0.96 - 1.40) | 0.12 | 1.16 (0.96 - 1.41) | 0.17 |
| Meta-analysis | 77,029 | 635,649 | 155.8 | 2517.0 | 2,788 | 48,308 | 1.07 (1.03 - 1.12) | <0.01 | 1.05 (0.84 - 1.32) | 0.66 |
| **PS-matching / intent-to-treat TAR** | | | | | | | | | | |
| OptumDOD | 4,567 | 49,167 | 6.7 | 139.6 | 287 | 4,795 | 1.27 (1.10 - 1.46) | <0.01 | 1.28 (1.11 - 1.48) | <0.01 |
| MDCR | 1,838 | 26,183 | 4.3 | 78.6 | 130 | 2,617 | 0.85 (0.68 - 1.06) | 0.15 | 0.78 (0.63 - 0.98) | 0.04 |
| Open Claims | 70,903 | 593,021 | 150.1 | 2301.6 | 2,660 | 42,372 | 1.09 (1.04 - 1.14) | <0.01 | 1.07 (0.87 - 1.33) | 0.52 |
| OptumEHR | 3,065 | 45,087 | 6.3 | 155.2 | 121 | 2,832 | 1.14 (0.92 - 1.40) | 0.21 | 1.17 (0.95 - 1.45) | 0.18 |
| Meta-analysis | 80,373 | 713,458 | 167.4 | 2675.0 | 3,198 | 52,616 | 1.09 (0.95 - 1.26) | 0.23 | 1.06 (0.85 - 1.32) | 0.60 |

#### 6.4 SGLT2I vs DPP4I

##### 6.4.1 Abnormal weight gain

**Supplementary Table 162:** Exposures, outcomes and risk estimates for Abnormal weight gain in SGLT2I (T) vs DPP4I (C) new-user cohorts under four different propensity score (PS) adjustment / time-at-risk (TAR) designs across data sources. We report total patient exposure time in 1,000 patient-year (KYr) units, uncalibrated and calibrated hazard ratios (HRs) and their 95% confidence intervals (CIs). Greyed out entries did not pass study diagnostics and are included only for completeness.

| | Patients | | Exposure (KYr) | | Outcomes | | Uncalibrated | | Calibrated | |
|---|---|---|---|---|---|---|---|---|---|---|
| | T | C | T | C | T | C | HR (95% CI) | $p$ | HR (95% CI) | $p$ |
| **PS-stratification / on-treatment TAR** | | | | | | | | | | |
| OptumDOD | 11,275 | 21,260 | 6.7 | 16.9 | 20 | 92 | 0.62 (0.36 - 1.03) | 0.07 | 0.61 (0.36 - 1.03) | 0.07 |
| MDCR | 3,444 | 9,872 | 2.5 | 8.9 | <5 | 24 | 0.81 (0.22 - 2.34) | 0.73 | 0.84 (0.26 - 2.74) | 0.77 |
| Open Claims | 177,824 | 332,881 | 123.7 | 307.8 | 144 | 543 | 0.81 (0.66 - 1.00) | 0.05 | 0.83 (0.62 - 1.11) | 0.22 |
| OptumEHR | 7,560 | 19,569 | 2.1 | 10.3 | 8 | 44 | 1.26 (0.52 - 2.74) | 0.59 | 1.14 (0.49 - 2.63) | 0.76 |
| VA | 7,350 | 14,321 | 4.6 | 14.5 | <10 | <10 | 1.53 (0.43 - 5.55) | 0.51 | 1.59 (0.44 - 5.71) | 0.48 |
| GermanyDA | 4,137 | 10,716 | 3.2 | 11.6 | 0 | 12 | | | | |
| SIDIAP | 3,707 | 20,269 | 4.4 | 44.1 | <5 | 56 | 1.22 (0.36 - 3.15) | 0.72 | 1.26 (0.42 - 3.74) | 0.68 |
| Meta-analysis | 211,160 | 418,172 | 144.1 | 402.4 | <192 | <769 | 0.82 (0.69 - 0.98) | 0.03 | 0.83 (0.65 - 1.06) | 0.14 |
| **PS-matching / on-treatment TAR** | | | | | | | | | | |
| OptumDOD | 9,027 | 20,878 | 5.9 | 16.5 | 19 | 89 | 0.58 (0.29 - 1.08) | 0.10 | 0.55 (0.29 - 1.06) | 0.08 |
| MDCR | 2,867 | 9,846 | 2.3 | 8.9 | <5 | 24 | 1.19 (0.25 - 4.40) | 0.81 | 1.21 (0.29 - 5.09) | 0.80 |
| Open Claims | 135,354 | 331,887 | 103.0 | 307.0 | 122 | 540 | 0.87 (0.65 - 1.15) | 0.34 | 0.87 (0.62 - 1.23) | 0.45 |
| OptumEHR | 6,731 | 19,496 | 1.9 | 10.3 | 7 | 43 | 1.59 (0.45 - 5.16) | 0.46 | 1.49 (0.44 - 5.06) | 0.52 |
| VA | 4,515 | 13,918 | 3.3 | 13.8 | <10 | <10 | 1.85 (0.30 - 12.1) | 0.51 | 1.98 (0.31 - 12.6) | 0.47 |
| GermanyDA | 3,406 | 10,647 | 2.8 | 11.6 | 0 | 12 | | | | |
| SIDIAP | 3,081 | 19,652 | 3.9 | 42.1 | <5 | 51 | 1.85 (0.50 - 5.25) | 0.30 | 1.93 (0.60 - 6.25) | 0.28 |
| Meta-analysis | 157,060 | 401,759 | 117.0 | 384.8 | <158 | 747 | 0.88 (0.69 - 1.12) | 0.30 | 0.87 (0.63 - 1.21) | 0.41 |
| **PS-stratification / intent-to-treat TAR** | | | | | | | | | | |
| OptumDOD | 11,275 | 21,260 | 17.1 | 53.6 | 78 | 263 | 0.97 (0.74 - 1.27) | 0.84 | 0.99 (0.75 - 1.30) | 0.93 |
| MDCR | 3,444 | 9,872 | 5.8 | 21.4 | 18 | 83 | 1.09 (0.62 - 1.84) | 0.75 | 1.09 (0.63 - 1.89) | 0.75 |
| Open Claims | 177,824 | 332,881 | 438.9 | 1408.5 | 750 | 2,742 | 1.08 (0.99 - 1.18) | 0.09 | 1.10 (0.91 - 1.32) | 0.32 |
| OptumEHR | 7,560 | 19,569 | 15.5 | 56.1 | 61 | 200 | 1.37 (1.00 - 1.86) | 0.04 | 1.36 (1.00 - 1.85) | 0.05 |
| VA | 7,350 | 14,321 | 8.0 | 32.3 | 12 | 32 | 1.39 (0.60 - 3.05) | 0.43 | 1.46 (0.65 - 3.30) | 0.36 |
| GermanyDA | 4,137 | 10,716 | 8.7 | 37.0 | <5 | 35 | 0.12 (0.01 - 0.57) | 0.06 | 0.11 (0.01 - 1.07) | 0.06 |
| SIDIAP | 3,707 | 20,269 | 6.4 | 58.0 | <5 | 75 | 0.81 (0.24 - 2.03) | 0.70 | 0.83 (0.29 - 2.40) | 0.73 |
| Meta-analysis | 215,297 | 428,888 | 500.3 | 1666.8 | <929 | 3,430 | 1.08 (1.00 - 1.17) | 0.05 | 1.10 (0.93 - 1.30) | 0.24 |
| **PS-matching / intent-to-treat TAR** | | | | | | | | | | |
| OptumDOD | 9,027 | 20,878 | 15.8 | 52.2 | 74 | 256 | 0.98 (0.71 - 1.34) | 0.92 | 0.98 (0.71 - 1.35) | 0.91 |
| MDCR | 2,867 | 9,846 | 5.4 | 21.3 | 18 | 82 | 1.49 (0.80 - 2.66) | 0.19 | 1.52 (0.83 - 2.79) | 0.19 |
| Open Claims | 135,354 | 331,887 | 400.1 | 1403.8 | 702 | 2,720 | 1.08 (0.98 - 1.18) | 0.12 | 1.09 (0.91 - 1.31) | 0.37 |
| OptumEHR | 6,731 | 19,496 | 14.7 | 55.8 | 60 | 198 | 1.34 (0.94 - 1.88) | 0.10 | 1.31 (0.93 - 1.86) | 0.14 |
| VA | 4,515 | 13,918 | 6.0 | 30.3 | <10 | 30 | 1.65 (0.65 - 4.03) | 0.28 | 1.73 (0.69 - 4.31) | 0.25 |
| GermanyDA | 3,406 | 10,647 | 8.0 | 36.6 | <5 | 35 | | | | |
| SIDIAP | 3,081 | 19,652 | 5.8 | 55.2 | <5 | 69 | 0.93 (0.27 - 2.40) | 0.89 | 0.95 (0.32 - 2.84) | 0.92 |
| Meta-analysis | 161,575 | 415,677 | 447.9 | 1618.5 | <869 | 3,355 | 1.09 (1.00 - 1.19) | 0.04 | 1.11 (0.94 - 1.30) | 0.23 |

##### 6.4.2 Abnormal weight loss

**Supplementary Table 163:** Exposures, outcomes and risk estimates for Abnormal weight loss in SGLT2I (T) vs DPP4I (C) new-user cohorts under four different propensity score (PS) adjustment / time-at-risk (TAR) designs across data sources. We report total patient exposure time in 1,000 patient-year (KYr) units, uncalibrated and calibrated hazard ratios (HRs) and their 95% confidence intervals (CIs). Greyed out entries did not pass study diagnostics and are included only for completeness.

| | Patients | | Exposure (KYr) | | Outcomes | | Uncalibrated | | Calibrated | |
|---|---|---|---|---|---|---|---|---|---|---|
| | T | C | T | C | T | C | HR (95% CI) | $p$ | HR (95% CI) | $p$ |
| **PS-stratification / on-treatment TAR** | | | | | | | | | | |
| OptumDOD | 11,094 | 20,351 | 6.6 | 16.1 | 163 | 350 | 1.29 (1.05 - 1.58) | 0.02 | 1.27 (1.03 - 1.57) | 0.03 |
| MDCR | 3,392 | 9,519 | 2.5 | 8.5 | 44 | 139 | 1.32 (0.90 - 1.91) | 0.14 | 1.37 (0.94 - 2.00) | 0.11 |
| Open Claims | 176,400 | 324,139 | 122.5 | 298.7 | 1,091 | 2,851 | 1.11 (1.02 - 1.20) | 0.01 | 1.13 (0.91 - 1.40) | 0.29 |
| OptumEHR | 7,530 | 19,221 | 2.1 | 10.1 | 30 | 177 | 0.93 (0.60 - 1.39) | 0.73 | 0.84 (0.55 - 1.28) | 0.44 |
| VA | 7,184 | 13,921 | 4.5 | 14.1 | 71 | 160 | 1.74 (1.20 - 2.51) | <0.01 | 1.81 (1.25 - 2.62) | <0.01 |
| GermanyDA | 4,053 | 10,447 | 3.1 | 11.3 | 37 | 95 | 1.19 (0.76 - 1.83) | 0.43 | 1.10 (0.71 - 1.71) | 0.66 |
| SIDIAP | 3,673 | 20,124 | 4.4 | 43.6 | 35 | 239 | 1.16 (0.76 - 1.73) | 0.47 | 1.20 (0.79 - 1.81) | 0.40 |
| Meta-analysis | 213,326 | 417,722 | 145.7 | 402.4 | 1,471 | 4,011 | 1.20 (1.07 - 1.36) | <0.01 | 1.21 (0.98 - 1.49) | 0.07 |
| **PS-matching / on-treatment TAR** | | | | | | | | | | |
| OptumDOD | 8,849 | 20,010 | 5.7 | 15.8 | 149 | 338 | 1.15 (0.87 - 1.50) | 0.33 | 1.10 (0.83 - 1.45) | 0.52 |
| MDCR | 2,818 | 9,480 | 2.3 | 8.5 | 42 | 139 | 1.31 (0.80 - 2.10) | 0.27 | 1.33 (0.82 - 2.16) | 0.28 |
| Open Claims | 133,917 | 323,111 | 101.8 | 297.8 | 903 | 2,828 | 1.11 (0.99 - 1.23) | 0.06 | 1.11 (0.88 - 1.38) | 0.40 |
| OptumEHR | 6,698 | 19,157 | 1.9 | 10.0 | 27 | 176 | 1.03 (0.59 - 1.73) | 0.92 | 0.96 (0.56 - 1.65) | 0.85 |
| VA | 4,387 | 13,541 | 3.2 | 13.4 | 55 | 152 | 1.82 (1.15 - 2.83) | <0.01 | 1.95 (1.24 - 3.06) | <0.01 |
| GermanyDA | 3,336 | 10,392 | 2.7 | 11.2 | 28 | 95 | 0.94 (0.52 - 1.62) | 0.82 | 0.89 (0.50 - 1.57) | 0.68 |
| SIDIAP | 3,060 | 19,520 | 3.9 | 41.7 | 29 | 233 | 1.10 (0.65 - 1.78) | 0.72 | 1.15 (0.69 - 1.91) | 0.61 |
| Meta-analysis | 163,065 | 415,211 | 121.5 | 398.4 | 1,233 | 3,961 | 1.13 (1.03 - 1.24) | <0.01 | 1.12 (0.89 - 1.42) | 0.33 |
| **PS-stratification / intent-to-treat TAR** | | | | | | | | | | |
| OptumDOD | 11,094 | 20,351 | 16.5 | 50.1 | 362 | 1,152 | 1.02 (0.89 - 1.16) | 0.79 | 1.03 (0.91 - 1.18) | 0.64 |
| MDCR | 3,392 | 9,519 | 5.6 | 20.3 | 111 | 375 | 1.30 (1.03 - 1.63) | 0.02 | 1.30 (1.03 - 1.65) | 0.05 |
| Open Claims | 176,400 | 324,139 | 433.7 | 1350.8 | 3,566 | 14,002 | 0.94 (0.90 - 0.98) | <0.01 | 0.96 (0.81 - 1.13) | 0.63 |
| OptumEHR | 7,530 | 19,221 | 15.3 | 54.2 | 201 | 838 | 1.00 (0.85 - 1.18) | 0.99 | 0.99 (0.84 - 1.17) | 0.86 |
| VA | 7,184 | 13,921 | 7.7 | 31.2 | 114 | 374 | 1.29 (0.98 - 1.68) | 0.07 | 1.36 (1.03 - 1.78) | 0.03 |
| GermanyDA | 4,053 | 10,447 | 8.4 | 35.7 | 66 | 279 | 1.01 (0.74 - 1.36) | 0.93 | 0.98 (0.72 - 1.33) | 0.81 |
| SIDIAP | 3,673 | 20,124 | 6.3 | 57.3 | 55 | 322 | 1.29 (0.92 - 1.77) | 0.12 | 1.32 (0.95 - 1.83) | 0.11 |
| Meta-analysis | 213,326 | 417,722 | 493.5 | 1599.5 | 4,475 | 17,342 | 1.07 (0.96 - 1.19) | 0.23 | 1.09 (0.91 - 1.30) | 0.37 |
| **PS-matching / intent-to-treat TAR** | | | | | | | | | | |
| OptumDOD | 8,849 | 20,010 | 15.2 | 48.9 | 340 | 1,111 | 1.01 (0.87 - 1.18) | 0.86 | 1.01 (0.87 - 1.18) | 0.84 |
| MDCR | 2,818 | 9,480 | 5.2 | 20.2 | 109 | 373 | 1.44 (1.10 - 1.87) | <0.01 | 1.47 (1.11 - 1.94) | 0.03 |
| Open Claims | 133,917 | 323,111 | 394.8 | 1346.2 | 3,249 | 13,900 | 0.95 (0.91 - 0.99) | 0.02 | 0.96 (0.82 - 1.13) | 0.65 |
| OptumEHR | 6,698 | 19,157 | 14.5 | 53.9 | 193 | 827 | 1.02 (0.85 - 1.22) | 0.85 | 1.00 (0.83 - 1.20) | 0.86 |
| VA | 4,387 | 13,541 | 5.8 | 29.3 | 82 | 352 | 1.26 (0.91 - 1.72) | 0.16 | 1.31 (0.95 - 1.81) | 0.11 |
| GermanyDA | 3,336 | 10,392 | 7.8 | 35.4 | 57 | 276 | 0.97 (0.69 - 1.34) | 0.87 | 0.95 (0.68 - 1.32) | 0.73 |
| SIDIAP | 3,060 | 19,520 | 5.8 | 54.5 | 49 | 312 | 1.20 (0.80 - 1.76) | 0.37 | 1.23 (0.83 - 1.83) | 0.33 |
| Meta-analysis | 163,065 | 415,211 | 449.1 | 1588.4 | 4,079 | 17,151 | 1.06 (0.95 - 1.19) | 0.29 | 1.07 (0.90 - 1.28) | 0.44 |

##### 6.4.3 Acute pancreatitis

**Supplementary Table 164:** Exposures, outcomes and risk estimates for Acute pancreatitis in SGLT2I (T) vs DPP4I (C) new-user cohorts under four different propensity score (PS) adjustment / time-at-risk (TAR) designs across data sources. We report total patient exposure time in 1,000 patient-year (KYr) units, uncalibrated and calibrated hazard ratios (HRs) and their 95% confidence intervals (CIs). Greyed out entries did not pass study diagnostics and are included only for completeness.

| | Patients | | Exposure (KYr) | | Outcomes | | Uncalibrated | | Calibrated | |
|---|---|---|---|---|---|---|---|---|---|---|
| | T | C | T | C | T | C | HR (95% CI) | $p$ | HR (95% CI) | $p$ |
| **PS-stratification / on-treatment TAR** | | | | | | | | | | |
| OptumDOD | 11,512 | 21,644 | 6.9 | 17.2 | 21 | 67 | 0.96 (0.55 - 1.62) | 0.88 | 0.95 (0.55 - 1.63) | 0.85 |
| MDCR | 3,480 | 9,962 | 2.5 | 9.0 | <5 | 22 | 1.01 (0.28 - 2.82) | 0.98 | 1.05 (0.33 - 3.30) | 0.94 |
| Open Claims | 182,041 | 338,295 | 126.7 | 312.9 | 176 | 728 | 0.76 (0.63 - 0.91) | <0.01 | 0.77 (0.59 - 1.01) | 0.07 |
| OptumEHR | 7,801 | 19,970 | 2.2 | 10.6 | <5 | 22 | 0.73 (0.16 - 2.29) | 0.64 | 0.66 (0.18 - 2.47) | 0.54 |
| VA | 7,438 | 14,470 | 4.7 | 14.6 | <10 | 20 | 0.18 (0.01 - 1.11) | 0.16 | 0.19 (0.02 - 2.05) | 0.17 |
| GermanyDA | 4,175 | 10,792 | 3.2 | 11.7 | 0 | 0 | | | | |
| SIDIAP | 3,736 | 20,481 | 4.5 | 44.5 | 0 | 20 | | | | |
| Meta-analysis | 212,272 | 404,341 | 143.0 | 364.4 | <217 | 859 | 0.77 (0.65 - 0.92) | <0.01 | 0.78 (0.61 - 0.99) | 0.04 |
| **PS-matching / on-treatment TAR** | | | | | | | | | | |
| OptumDOD | 9,221 | 21,253 | 6.0 | 16.9 | 18 | 63 | 0.84 (0.42 - 1.59) | 0.61 | 0.80 (0.41 - 1.56) | 0.52 |
| MDCR | 2,902 | 9,936 | 2.3 | 8.9 | <5 | 22 | 1.41 (0.30 - 5.08) | 0.64 | 1.43 (0.34 - 5.94) | 0.62 |
| Open Claims | 138,478 | 337,302 | 105.4 | 312.1 | 149 | 724 | 0.75 (0.59 - 0.95) | 0.02 | 0.75 (0.55 - 1.02) | 0.08 |
| OptumEHR | 6,949 | 19,896 | 2.0 | 10.5 | <5 | 22 | 0.71 (0.16 - 2.26) | 0.61 | 0.66 (0.18 - 2.50) | 0.55 |
| VA | 4,551 | 14,060 | 3.4 | 14.0 | <10 | 20 | | | | |
| GermanyDA | 3,434 | 10,733 | 2.9 | 11.6 | 0 | 0 | | | | |
| SIDIAP | 3,110 | 19,850 | 3.9 | 42.5 | 0 | 19 | | | | |
| Meta-analysis | 157,550 | 388,387 | 115.7 | 348.5 | <177 | 831 | 0.77 (0.62 - 0.96) | 0.02 | 0.76 (0.56 - 1.04) | 0.08 |
| **PS-stratification / intent-to-treat TAR** | | | | | | | | | | |
| OptumDOD | 11,512 | 21,644 | 17.6 | 54.7 | 54 | 210 | 0.89 (0.64 - 1.22) | 0.50 | 0.91 (0.66 - 1.26) | 0.57 |
| MDCR | 3,480 | 9,962 | 5.9 | 21.5 | 10 | 56 | 1.07 (0.50 - 2.06) | 0.86 | 1.07 (0.52 - 2.17) | 0.86 |
| Open Claims | 182,041 | 338,295 | 451.3 | 1431.8 | 675 | 3,162 | 0.82 (0.75 - 0.90) | <0.01 | 0.84 (0.70 - 1.01) | 0.07 |
| OptumEHR | 7,801 | 19,970 | 16.1 | 57.6 | 20 | 83 | 1.26 (0.73 - 2.07) | 0.39 | 1.24 (0.74 - 2.09) | 0.41 |
| VA | 7,438 | 14,470 | 8.0 | 32.6 | <10 | 38 | 0.49 (0.13 - 1.44) | 0.24 | 0.51 (0.15 - 1.71) | 0.28 |
| GermanyDA | 4,175 | 10,792 | 8.8 | 37.4 | 0 | 0 | | | | |
| SIDIAP | 3,736 | 20,481 | 6.5 | 58.5 | 0 | 28 | | | | |
| Meta-analysis | 212,272 | 404,341 | 498.9 | 1598.2 | <769 | 3,549 | 0.84 (0.77 - 0.93) | <0.01 | 0.86 (0.72 - 1.02) | 0.08 |
| **PS-matching / intent-to-treat TAR** | | | | | | | | | | |
| OptumDOD | 9,221 | 21,253 | 16.2 | 53.2 | 51 | 202 | 0.84 (0.58 - 1.19) | 0.33 | 0.84 (0.58 - 1.20) | 0.34 |
| MDCR | 2,902 | 9,936 | 5.5 | 21.5 | 10 | 55 | 1.00 (0.44 - 2.08) | 1.00 | 1.02 (0.47 - 2.24) | 0.94 |
| Open Claims | 138,478 | 337,302 | 411.4 | 1427.1 | 624 | 3,143 | 0.82 (0.74 - 0.90) | <0.01 | 0.83 (0.69 - 0.99) | 0.05 |
| OptumEHR | 6,949 | 19,896 | 15.3 | 57.3 | 20 | 83 | 1.26 (0.71 - 2.13) | 0.40 | 1.24 (0.72 - 2.14) | 0.45 |
| VA | 4,551 | 14,060 | 6.1 | 30.6 | <10 | 37 | 0.35 (0.05 - 1.32) | 0.20 | 0.37 (0.07 - 1.83) | 0.22 |
| GermanyDA | 3,434 | 10,733 | 8.1 | 37.1 | 0 | 0 | | | | |
| SIDIAP | 3,110 | 19,850 | 5.9 | 55.6 | 0 | 27 | | | | |
| Meta-analysis | 162,101 | 402,447 | 454.4 | 1589.6 | <715 | 3,520 | 0.83 (0.76 - 0.91) | <0.01 | 0.84 (0.71 - 0.99) | 0.04 |

##### 6.4.4 All-cause mortality

**Supplementary Table 165:** Exposures, outcomes and risk estimates for All-cause mortality in SGLT2I (T) vs DPP4I (C) new-user cohorts under four different propensity score (PS) adjustment / time-at-risk (TAR) designs across data sources. We report total patient exposure time in 1,000 patient-year (KYr) units, uncalibrated and calibrated hazard ratios (HRs) and their 95% confidence intervals (CIs). Greyed out entries did not pass study diagnostics and are included only for completeness.

| | Patients | | Exposure (KYr) | | Outcomes | | Uncalibrated | | Calibrated | |
|---|---|---|---|---|---|---|---|---|---|---|
| | T | C | T | C | T | C | HR (95% CI) | $p$ | HR (95% CI) | $p$ |
| **PS-stratification / on-treatment TAR** | | | | | | | | | | |
| OptumDOD | 11,614 | 21,849 | 6.9 | 17.4 | 43 | 229 | 0.66 (0.46 - 0.94) | 0.03 | 0.66 (0.46 - 0.94) | 0.02 |
| MDCR | 3,510 | 10,031 | 2.6 | 9.0 | <5 | 21 | 1.36 (0.21 - 4.88) | 0.70 | 1.41 (0.29 - 6.74) | 0.67 |
| Open Claims | 184,807 | 342,501 | 128.6 | 317.0 | 0 | <5 | | | | |
| OptumEHR | 7,828 | 20,055 | 2.2 | 10.6 | 8 | 160 | 0.37 (0.16 - 0.72) | <0.01 | 0.33 (0.16 - 0.70) | <0.01 |
| VA | 7,529 | 14,605 | 4.7 | 14.7 | 65 | 233 | 0.63 (0.45 - 0.88) | <0.01 | 0.66 (0.47 - 0.92) | 0.02 |
| GermanyDA | 4,175 | 10,792 | 3.2 | 11.7 | <5 | 9 | 1.19 (0.23 - 4.88) | 0.82 | 1.10 (0.24 - 5.12) | 0.90 |
| SIDIAP | 3,746 | 20,524 | 4.5 | 44.7 | 95 | 1,466 | 0.82 (0.65 - 1.03) | 0.10 | 0.85 (0.67 - 1.07) | 0.20 |
| Meta-analysis | 34,227 | 87,064 | 20.9 | 96.5 | <216 | 2,109 | 0.70 (0.57 - 0.85) | <0.01 | 0.70 (0.54 - 0.92) | <0.01 |
| **PS-matching / on-treatment TAR** | | | | | | | | | | |
| OptumDOD | 9,301 | 21,451 | 6.1 | 17.1 | 35 | 216 | 0.55 (0.33 - 0.88) | 0.02 | 0.53 (0.32 - 0.86) | 0.01 |
| MDCR | 2,927 | 10,005 | 2.3 | 9.0 | <5 | 20 | 1.11 (0.17 - 4.40) | 0.90 | 1.13 (0.22 - 5.81) | 0.89 |
| Open Claims | 140,534 | 341,473 | 107.1 | 316.2 | 0 | <5 | | | | |
| OptumEHR | 6,967 | 19,980 | 2.0 | 10.6 | 7 | 156 | 0.46 (0.18 - 0.99) | 0.07 | 0.43 (0.18 - 1.00) | 0.05 |
| VA | 4,600 | 14,186 | 3.4 | 14.1 | 43 | 224 | 0.69 (0.43 - 1.05) | 0.09 | 0.73 (0.47 - 1.14) | 0.19 |
| GermanyDA | 3,434 | 10,733 | 2.9 | 11.6 | <5 | 9 | 0.85 (0.11 - 4.74) | 0.87 | 0.81 (0.12 - 5.25) | 0.83 |
| SIDIAP | 3,116 | 19,888 | 4.0 | 42.6 | 76 | 1,346 | 0.74 (0.55 - 0.98) | 0.04 | 0.78 (0.58 - 1.04) | 0.12 |
| Meta-analysis | 26,911 | 85,510 | 17.7 | 93.3 | <166 | 1,962 | 0.68 (0.55 - 0.83) | <0.01 | 0.67 (0.50 - 0.90) | <0.01 |
| **PS-stratification / intent-to-treat TAR** | | | | | | | | | | |
| OptumDOD | 11,614 | 21,849 | 17.7 | 54.9 | 359 | 2,180 | 0.78 (0.69 - 0.88) | <0.01 | 0.79 (0.70 - 0.90) | <0.01 |
| MDCR | 3,510 | 10,031 | 5.9 | 21.8 | <5 | 44 | 0.85 (0.20 - 2.43) | 0.80 | 0.85 (0.25 - 2.96) | 0.81 |
| Open Claims | 184,807 | 342,501 | 459.1 | 1456.9 | <5 | 22 | 0.92 (0.14 - 3.29) | 0.92 | 0.94 (0.19 - 4.52) | 0.94 |
| OptumEHR | 7,828 | 20,055 | 16.1 | 57.8 | 119 | 804 | 0.71 (0.58 - 0.87) | <0.01 | 0.70 (0.57 - 0.87) | <0.01 |
| VA | 7,529 | 14,605 | 8.1 | 32.9 | 184 | 895 | 0.72 (0.59 - 0.88) | <0.01 | 0.76 (0.63 - 0.93) | 0.01 |
| GermanyDA | 4,175 | 10,792 | 8.7 | 37.4 | 11 | 62 | 1.14 (0.54 - 2.23) | 0.72 | 1.10 (0.54 - 2.25) | 0.79 |
| SIDIAP | 3,746 | 20,524 | 6.5 | 58.7 | 202 | 2,923 | 0.84 (0.72 - 0.98) | 0.03 | 0.86 (0.74 - 1.01) | 0.11 |
| Meta-analysis | 223,209 | 440,357 | 522.2 | 1720.4 | <885 | 6,930 | 0.78 (0.72 - 0.84) | <0.01 | 0.79 (0.67 - 0.93) | <0.01 |
| **PS-matching / intent-to-treat TAR** | | | | | | | | | | |
| OptumDOD | 9,301 | 21,451 | 16.4 | 53.5 | 328 | 2,014 | 0.70 (0.61 - 0.81) | <0.01 | 0.70 (0.61 - 0.81) | <0.01 |
| MDCR | 2,927 | 10,005 | 5.6 | 21.7 | <5 | 43 | 0.87 (0.20 - 2.60) | 0.84 | 0.89 (0.25 - 3.21) | 0.87 |
| Open Claims | 140,534 | 341,473 | 418.5 | 1452.0 | <5 | 21 | 0.89 (0.14 - 3.21) | 0.89 | 0.90 (0.19 - 4.38) | 0.90 |
| OptumEHR | 6,967 | 19,980 | 15.3 | 57.5 | 115 | 790 | 0.74 (0.58 - 0.93) | <0.01 | 0.73 (0.58 - 0.91) | <0.01 |
| VA | 4,600 | 14,186 | 6.1 | 30.8 | 136 | 832 | 0.76 (0.59 - 0.96) | 0.02 | 0.79 (0.62 - 1.01) | 0.07 |
| GermanyDA | 3,434 | 10,733 | 8.1 | 37.0 | 9 | 62 | 1.08 (0.47 - 2.22) | 0.85 | 1.05 (0.48 - 2.28) | 0.89 |
| SIDIAP | 3,116 | 19,888 | 5.9 | 55.8 | 170 | 2,645 | 0.81 (0.67 - 0.97) | 0.02 | 0.83 (0.69 - 1.00) | 0.08 |
| Meta-analysis | 170,879 | 437,716 | 475.9 | 1708.3 | <768 | 6,407 | 0.75 (0.68 - 0.82) | <0.01 | 0.76 (0.64 - 0.89) | <0.01 |

##### 6.4.5 Bone fracture

**Supplementary Table 166:** Exposures, outcomes and risk estimates for Bone fracture in SGLT2I (T) vs DPP4I (C) new-user cohorts under four different propensity score (PS) adjustment / time-at-risk (TAR) designs across data sources. We report total patient exposure time in 1,000 patient-year (KYr) units, uncalibrated and calibrated hazard ratios (HRs) and their 95% confidence intervals (CIs). Greyed out entries did not pass study diagnostics and are included only for completeness.

| | Patients | | Exposure (KYr) | | Outcomes | | Uncalibrated | | Calibrated | |
|---|---|---|---|---|---|---|---|---|---|---|
| | T | C | T | C | T | C | HR (95% CI) | $p$ | HR (95% CI) | $p$ |
| **PS-stratification / on-treatment TAR** | | | | | | | | | | |
| OptumDOD | 10,423 | 19,212 | 6.1 | 15.0 | 200 | 527 | 1.09 (0.91 - 1.30) | 0.34 | 1.07 (0.90 - 1.29) | 0.46 |
| MDCR | 3,143 | 8,713 | 2.3 | 7.7 | 73 | 267 | 1.15 (0.86 - 1.53) | 0.34 | 1.19 (0.89 - 1.59) | 0.26 |
| Open Claims | 155,987 | 289,690 | 108.3 | 264.8 | 1,810 | 6,173 | 0.92 (0.87 - 0.98) | <0.01 | 0.94 (0.77 - 1.16) | 0.60 |
| OptumEHR | 7,175 | 18,312 | 2.0 | 9.5 | 39 | 237 | 1.11 (0.76 - 1.58) | 0.58 | 1.00 (0.69 - 1.45) | 0.85 |
| VA | 6,699 | 13,160 | 4.2 | 13.3 | 39 | 121 | 0.94 (0.58 - 1.50) | 0.81 | 0.98 (0.61 - 1.57) | 0.90 |
| GermanyDA | 3,627 | 9,166 | 2.8 | 9.8 | 50 | 253 | 0.92 (0.65 - 1.28) | 0.62 | 0.85 (0.60 - 1.19) | 0.38 |
| SIDIAP | 3,268 | 17,523 | 3.9 | 37.6 | 52 | 725 | 0.89 (0.64 - 1.19) | 0.44 | 0.91 (0.67 - 1.25) | 0.59 |
| Meta-analysis | 190,322 | 375,776 | 129.6 | 357.7 | 2,263 | 8,303 | 0.98 (0.89 - 1.07) | 0.63 | 0.99 (0.81 - 1.19) | 0.87 |
| **PS-matching / on-treatment TAR** | | | | | | | | | | |
| OptumDOD | 8,335 | 18,905 | 5.4 | 14.7 | 180 | 512 | 1.30 (1.01 - 1.66) | 0.04 | 1.24 (0.97 - 1.59) | 0.09 |
| MDCR | 2,618 | 8,623 | 2.1 | 7.6 | 70 | 261 | 1.41 (0.95 - 2.08) | 0.08 | 1.44 (0.97 - 2.13) | 0.09 |
| Open Claims | 118,920 | 288,961 | 90.2 | 264.2 | 1,571 | 6,140 | 0.94 (0.87 - 1.02) | 0.14 | 0.94 (0.76 - 1.17) | 0.61 |
| OptumEHR | 6,382 | 18,251 | 1.8 | 9.5 | 34 | 236 | 1.11 (0.69 - 1.75) | 0.66 | 1.04 (0.65 - 1.66) | 0.82 |
| VA | 4,146 | 12,701 | 3.0 | 12.6 | 24 | 110 | 0.80 (0.42 - 1.47) | 0.50 | 0.86 (0.46 - 1.62) | 0.65 |
| GermanyDA | 2,976 | 9,138 | 2.4 | 9.7 | 46 | 248 | 0.95 (0.62 - 1.42) | 0.79 | 0.90 (0.59 - 1.37) | 0.62 |
| SIDIAP | 2,700 | 17,009 | 3.4 | 36.1 | 42 | 671 | 0.95 (0.65 - 1.36) | 0.81 | 1.00 (0.69 - 1.45) | 0.80 |
| Meta-analysis | 146,077 | 373,588 | 108.4 | 354.5 | 1,967 | 8,178 | 1.05 (0.91 - 1.23) | 0.49 | 1.05 (0.80 - 1.36) | 0.74 |
| **PS-stratification / intent-to-treat TAR** | | | | | | | | | | |
| OptumDOD | 10,423 | 19,212 | 15.3 | 46.0 | 493 | 1,700 | 1.03 (0.92 - 1.15) | 0.62 | 1.04 (0.93 - 1.17) | 0.49 |
| MDCR | 3,143 | 8,713 | 5.1 | 18.1 | 158 | 646 | 1.03 (0.85 - 1.24) | 0.75 | 1.03 (0.85 - 1.25) | 0.79 |
| Open Claims | 155,987 | 289,690 | 377.8 | 1171.1 | 7,504 | 30,320 | 0.98 (0.95 - 1.00) | 0.08 | 0.99 (0.84 - 1.17) | 0.91 |
| OptumEHR | 7,175 | 18,312 | 14.4 | 50.9 | 281 | 1,130 | 1.15 (1.00 - 1.32) | 0.05 | 1.14 (0.99 - 1.32) | 0.09 |
| VA | 6,699 | 13,160 | 7.3 | 29.5 | 69 | 300 | 0.81 (0.58 - 1.12) | 0.22 | 0.85 (0.61 - 1.19) | 0.37 |
| GermanyDA | 3,627 | 9,166 | 7.4 | 30.4 | 137 | 768 | 0.93 (0.76 - 1.13) | 0.50 | 0.90 (0.74 - 1.10) | 0.37 |
| SIDIAP | 3,268 | 17,523 | 5.6 | 49.0 | 84 | 984 | 0.93 (0.73 - 1.18) | 0.56 | 0.95 (0.75 - 1.21) | 0.70 |
| Meta-analysis | 190,322 | 375,776 | 432.9 | 1395.1 | 8,726 | 35,848 | 1.00 (0.95 - 1.05) | 0.94 | 1.02 (0.87 - 1.18) | 0.85 |
| **PS-matching / intent-to-treat TAR** | | | | | | | | | | |
| OptumDOD | 8,335 | 18,905 | 14.1 | 45.0 | 458 | 1,649 | 1.06 (0.93 - 1.20) | 0.39 | 1.06 (0.93 - 1.20) | 0.43 |
| MDCR | 2,618 | 8,623 | 4.8 | 17.9 | 151 | 631 | 1.01 (0.81 - 1.26) | 0.91 | 1.03 (0.82 - 1.31) | 0.81 |
| Open Claims | 118,920 | 288,961 | 344.0 | 1168.1 | 7,073 | 30,143 | 0.98 (0.95 - 1.01) | 0.16 | 0.99 (0.84 - 1.16) | 0.89 |
| OptumEHR | 6,382 | 18,251 | 13.7 | 50.7 | 274 | 1,117 | 1.16 (0.99 - 1.36) | 0.06 | 1.14 (0.98 - 1.34) | 0.13 |
| VA | 4,146 | 12,701 | 5.5 | 27.3 | 45 | 274 | 0.75 (0.49 - 1.12) | 0.17 | 0.78 (0.52 - 1.19) | 0.26 |
| GermanyDA | 2,976 | 9,138 | 6.8 | 30.3 | 130 | 757 | 0.92 (0.74 - 1.14) | 0.47 | 0.90 (0.72 - 1.12) | 0.39 |
| SIDIAP | 2,700 | 17,009 | 5.1 | 46.8 | 73 | 912 | 0.96 (0.72 - 1.25) | 0.74 | 0.98 (0.74 - 1.29) | 0.79 |
| Meta-analysis | 146,077 | 373,588 | 394.0 | 1386.2 | 8,204 | 35,483 | 1.00 (0.95 - 1.06) | 0.92 | 1.01 (0.87 - 1.18) | 0.86 |

##### 6.4.6 Diabetic ketoacidosis

**Supplementary Table 167:** Exposures, outcomes and risk estimates for Diabetic ketoacidosis in SGLT2I (T) vs DPP4I (C) new-user cohorts under four different propensity score (PS) adjustment / time-at-risk (TAR) designs across data sources. We report total patient exposure time in 1,000 patient-year (KYr) units, uncalibrated and calibrated hazard ratios (HRs) and their 95% confidence intervals (CIs). Greyed out entries did not pass study diagnostics and are included only for completeness.

| | Patients | | Exposure (KYr) | | Outcomes | | Uncalibrated | | Calibrated | |
|---|---|---|---|---|---|---|---|---|---|---|
| | T | C | T | C | T | C | HR (95% CI) | $p$ | HR (95% CI) | $p$ |
| **PS-stratification / on-treatment TAR** | | | | | | | | | | |
| OptumDOD | 11,606 | 21,817 | 6.9 | 17.4 | 18 | 31 | 1.67 (0.86 - 3.19) | 0.12 | 1.65 (0.85 - 3.19) | 0.14 |
| MDCR | 3,506 | 10,008 | 2.6 | 9.0 | <5 | 10 | 1.52 (0.20 - 7.28) | 0.65 | 1.57 (0.26 - 9.39) | 0.62 |
| Open Claims | 184,585 | 342,039 | 128.5 | 316.5 | 137 | 252 | 1.54 (1.21 - 1.96) | <0.01 | 1.57 (1.15 - 2.16) | <0.01 |
| OptumEHR | 7,821 | 20,047 | 2.2 | 10.6 | <5 | 14 | 1.25 (0.32 - 4.15) | 0.73 | 1.13 (0.31 - 4.09) | 0.85 |
| VA | 7,524 | 14,600 | 4.7 | 14.7 | <10 | <10 | | 0.04 | | 0.04 |
| GermanyDA | 4,175 | 10,792 | 3.2 | 11.7 | 0 | 0 | | | | |
| SIDIAP | 3,746 | 20,524 | 4.5 | 44.7 | 0 | 0 | | | | |
| Meta-analysis | 203,715 | 378,456 | 140.1 | 348.6 | <165 | <293 | 1.58 (1.26 - 1.99) | <0.01 | 1.60 (1.21 - 2.12) | <0.01 |
| **PS-matching / on-treatment TAR** | | | | | | | | | | |
| OptumDOD | 9,292 | 21,424 | 6.1 | 17.1 | 15 | 30 | 1.67 (0.74 - 3.67) | 0.21 | 1.60 (0.72 - 3.56) | 0.25 |
| MDCR | 2,922 | 9,982 | 2.3 | 9.0 | <5 | 10 | 0.85 (0.04 - 6.64) | 0.90 | 0.87 (0.07 - 11.1) | 0.91 |
| Open Claims | 140,351 | 341,015 | 106.9 | 315.6 | 117 | 252 | 1.32 (0.98 - 1.78) | 0.07 | 1.32 (0.93 - 1.89) | 0.13 |
| OptumEHR | 6,961 | 19,973 | 2.0 | 10.6 | <5 | 14 | 0.83 (0.11 - 4.54) | 0.85 | 0.78 (0.12 - 5.00) | 0.79 |
| VA | 4,598 | 14,181 | 3.4 | 14.1 | <10 | <10 | | 0.10 | | 0.09 |
| GermanyDA | 3,434 | 10,733 | 2.9 | 11.6 | 0 | 0 | | | | |
| SIDIAP | 3,116 | 19,888 | 4.0 | 42.6 | 0 | 0 | | | | |
| Meta-analysis | 149,643 | 362,439 | 113.0 | 332.7 | 132 | 282 | 1.36 (1.03 - 1.80) | 0.03 | 1.35 (0.95 - 1.92) | 0.10 |
| **PS-stratification / intent-to-treat TAR** | | | | | | | | | | |
| OptumDOD | 11,606 | 21,817 | 17.7 | 55.3 | 44 | 112 | 1.42 (0.96 - 2.08) | 0.08 | 1.44 (0.98 - 2.13) | 0.07 |
| MDCR | 3,506 | 10,008 | 5.9 | 21.7 | 11 | 25 | 2.36 (1.05 - 5.01) | 0.03 | 2.36 (1.08 - 5.15) | 0.03 |
| Open Claims | 184,585 | 342,039 | 457.8 | 1452.2 | 494 | 1,520 | 1.26 (1.12 - 1.40) | <0.01 | 1.28 (1.05 - 1.56) | 0.02 |
| OptumEHR | 7,821 | 20,047 | 16.1 | 58.0 | 11 | 42 | 1.14 (0.53 - 2.29) | 0.72 | 1.13 (0.54 - 2.36) | 0.74 |
| VA | 7,524 | 14,600 | 8.1 | 32.9 | <10 | <10 | 1.77 (0.39 - 8.24) | 0.47 | 1.86 (0.40 - 8.58) | 0.43 |
| GermanyDA | 4,175 | 10,792 | 8.8 | 37.4 | 0 | 0 | | | | |
| SIDIAP | 3,746 | 20,524 | 6.5 | 58.7 | 0 | 0 | | | | |
| Meta-analysis | 215,042 | 408,511 | 505.7 | 1620.1 | <570 | <1,709 | 1.28 (1.15 - 1.42) | <0.01 | 1.30 (1.09 - 1.56) | <0.01 |
| **PS-matching / intent-to-treat TAR** | | | | | | | | | | |
| OptumDOD | 9,292 | 21,424 | 16.4 | 53.8 | 39 | 108 | 1.60 (1.03 - 2.47) | 0.03 | 1.60 (1.03 - 2.49) | 0.04 |
| MDCR | 2,922 | 9,982 | 5.6 | 21.7 | 11 | 25 | 1.69 (0.64 - 4.16) | 0.27 | 1.73 (0.68 - 4.40) | 0.26 |
| Open Claims | 140,351 | 341,015 | 417.2 | 1447.3 | 458 | 1,512 | 1.22 (1.08 - 1.38) | <0.01 | 1.24 (1.02 - 1.51) | 0.04 |
| OptumEHR | 6,961 | 19,973 | 15.3 | 57.6 | 11 | 42 | 1.23 (0.53 - 2.62) | 0.62 | 1.21 (0.54 - 2.69) | 0.65 |
| VA | 4,598 | 14,181 | 6.1 | 30.8 | <10 | <10 | | 0.06 | | 0.05 |
| GermanyDA | 3,434 | 10,733 | 8.1 | 37.1 | 0 | 0 | | | | |
| SIDIAP | 3,116 | 19,888 | 5.9 | 55.8 | 0 | 0 | | | | |
| Meta-analysis | 159,526 | 392,394 | 454.5 | 1580.4 | 519 | 1,687 | 1.27 (1.10 - 1.47) | <0.01 | 1.28 (1.05 - 1.57) | 0.02 |

##### 6.4.7 Diarrhea

**Supplementary Table 168:** Exposures, outcomes and risk estimates for Diarrhea in SGLT2I (T) vs DPP4I (C) new-user cohorts under four different propensity score (PS) adjustment / time-at-risk (TAR) designs across data sources. We report total patient exposure time in 1,000 patient-year (KYr) units, uncalibrated and calibrated hazard ratios (HRs) and their 95% confidence intervals (CIs). Greyed out entries did not pass study diagnostics and are included only for completeness.

| | Patients | | Exposure (KYr) | | Outcomes | | Uncalibrated | | Calibrated | |
|---|---|---|---|---|---|---|---|---|---|---|
| | T | C | T | C | T | C | HR (95% CI) | $p$ | HR (95% CI) | $p$ |
| **PS-stratification / on-treatment TAR** | | | | | | | | | | |
| OptumDOD | 9,875 | 17,929 | 5.8 | 13.8 | 228 | 648 | 0.93 (0.79 - 1.10) | 0.41 | 0.92 (0.78 - 1.09) | 0.35 |
| MDCR | 3,032 | 8,448 | 2.2 | 7.4 | 57 | 281 | 0.74 (0.54 - 1.00) | 0.05 | 0.76 (0.56 - 1.04) | 0.10 |
| Open Claims | 158,033 | 291,037 | 109.5 | 264.6 | 1,724 | 5,952 | 0.88 (0.83 - 0.93) | <0.01 | 0.90 (0.73 - 1.10) | 0.32 |
| OptumEHR | 6,910 | 17,477 | 1.9 | 8.9 | 70 | 341 | 1.10 (0.82 - 1.44) | 0.52 | 0.99 (0.75 - 1.32) | 0.81 |
| VA | 6,417 | 12,666 | 4.0 | 12.7 | 65 | 246 | 0.77 (0.55 - 1.06) | 0.12 | 0.80 (0.58 - 1.11) | 0.20 |
| GermanyDA | 4,111 | 10,663 | 3.2 | 11.5 | 6 | 21 | 2.05 (0.60 - 5.97) | 0.22 | 1.90 (0.60 - 5.98) | 0.27 |
| SIDIAP | 3,550 | 19,181 | 4.2 | 41.2 | 34 | 570 | 0.48 (0.33 - 0.69) | <0.01 | 0.50 (0.34 - 0.73) | <0.01 |
| Meta-analysis | 191,928 | 377,401 | 130.8 | 360.1 | 2,184 | 8,059 | 0.84 (0.69 - 1.01) | 0.06 | 0.84 (0.66 - 1.08) | 0.18 |
| **PS-matching / on-treatment TAR** | | | | | | | | | | |
| OptumDOD | 7,859 | 17,646 | 5.0 | 13.5 | 206 | 628 | 0.94 (0.75 - 1.16) | 0.56 | 0.90 (0.72 - 1.12) | 0.36 |
| MDCR | 2,517 | 8,362 | 2.0 | 7.3 | 52 | 275 | 0.63 (0.42 - 0.94) | 0.03 | 0.64 (0.43 - 0.97) | 0.05 |
| Open Claims | 119,369 | 290,175 | 90.5 | 263.9 | 1,471 | 5,921 | 0.90 (0.83 - 0.97) | <0.01 | 0.90 (0.73 - 1.11) | 0.35 |
| OptumEHR | 6,117 | 17,357 | 1.7 | 8.8 | 62 | 339 | 1.17 (0.80 - 1.67) | 0.41 | 1.09 (0.76 - 1.58) | 0.65 |
| VA | 3,980 | 12,179 | 2.9 | 12.0 | 45 | 234 | 0.78 (0.50 - 1.19) | 0.26 | 0.83 (0.54 - 1.29) | 0.43 |
| GermanyDA | 3,389 | 10,605 | 2.8 | 11.5 | 6 | 21 | 3.72 (1.02 - 12.8) | 0.04 | 3.53 (1.00 - 12.5) | 0.05 |
| SIDIAP | 2,946 | 18,612 | 3.7 | 39.4 | 25 | 546 | 0.38 (0.22 - 0.61) | <0.01 | 0.39 (0.24 - 0.66) | <0.01 |
| Meta-analysis | 146,177 | 374,936 | 108.8 | 356.4 | 1,867 | 7,964 | 0.84 (0.62 - 1.13) | 0.24 | 0.83 (0.57 - 1.20) | 0.32 |
| **PS-stratification / intent-to-treat TAR** | | | | | | | | | | |
| OptumDOD | 9,875 | 17,929 | 14.4 | 42.3 | 576 | 1,915 | 0.95 (0.86 - 1.05) | 0.35 | 0.97 (0.87 - 1.08) | 0.59 |
| MDCR | 3,032 | 8,448 | 4.8 | 17.3 | 171 | 716 | 0.97 (0.81 - 1.16) | 0.73 | 0.97 (0.80 - 1.17) | 0.79 |
| Open Claims | 158,033 | 291,037 | 379.4 | 1171.3 | 6,741 | 26,529 | 0.94 (0.92 - 0.97) | <0.01 | 0.96 (0.82 - 1.13) | 0.63 |
| OptumEHR | 6,910 | 17,477 | 13.6 | 47.4 | 341 | 1,385 | 0.98 (0.86 - 1.11) | 0.73 | 0.97 (0.85 - 1.10) | 0.64 |
| VA | 6,417 | 12,666 | 6.9 | 28.2 | 125 | 560 | 0.85 (0.67 - 1.08) | 0.18 | 0.90 (0.70 - 1.14) | 0.40 |
| GermanyDA | 4,111 | 10,663 | 8.6 | 36.8 | 11 | 52 | 1.17 (0.52 - 2.39) | 0.69 | 1.13 (0.53 - 2.41) | 0.75 |
| SIDIAP | 3,550 | 19,181 | 6.1 | 54.1 | 66 | 766 | 0.67 (0.51 - 0.88) | <0.01 | 0.69 (0.52 - 0.91) | 0.01 |
| Meta-analysis | 191,928 | 377,401 | 433.8 | 1397.4 | 8,031 | 31,923 | 0.94 (0.92 - 0.97) | <0.01 | 0.96 (0.83 - 1.11) | 0.56 |
| **PS-matching / intent-to-treat TAR** | | | | | | | | | | |
| OptumDOD | 7,859 | 17,646 | 13.2 | 41.3 | 536 | 1,855 | 0.94 (0.84 - 1.05) | 0.29 | 0.94 (0.83 - 1.06) | 0.35 |
| MDCR | 2,517 | 8,362 | 4.5 | 17.1 | 161 | 705 | 0.94 (0.76 - 1.16) | 0.58 | 0.96 (0.77 - 1.20) | 0.80 |
| Open Claims | 119,369 | 290,175 | 344.1 | 1167.7 | 6,267 | 26,376 | 0.95 (0.92 - 0.98) | <0.01 | 0.96 (0.82 - 1.12) | 0.60 |
| OptumEHR | 6,117 | 17,357 | 12.9 | 47.0 | 322 | 1,367 | 0.98 (0.85 - 1.13) | 0.82 | 0.97 (0.84 - 1.12) | 0.67 |
| VA | 3,980 | 12,179 | 5.2 | 25.9 | 91 | 517 | 0.92 (0.68 - 1.22) | 0.56 | 0.96 (0.72 - 1.28) | 0.78 |
| GermanyDA | 3,389 | 10,605 | 7.9 | 36.5 | 11 | 52 | 1.50 (0.66 - 3.16) | 0.31 | 1.46 (0.66 - 3.20) | 0.35 |
| SIDIAP | 2,946 | 18,612 | 5.6 | 51.6 | 56 | 734 | 0.57 (0.40 - 0.79) | <0.01 | 0.58 (0.41 - 0.82) | <0.01 |
| Meta-analysis | 146,177 | 374,936 | 393.5 | 1387.0 | 7,444 | 31,606 | 0.94 (0.92 - 0.97) | <0.01 | 0.95 (0.83 - 1.10) | 0.51 |

##### 6.4.8 Genitourinary infection

**Supplementary Table 169:** Exposures, outcomes and risk estimates for Genitourinary infection in SGLT2I (T) vs DPP4I (C) new-user cohorts under four different propensity score (PS) adjustment / time-at-risk (TAR) designs across data sources. We report total patient exposure time in 1,000 patient-year (KYr) units, uncalibrated and calibrated hazard ratios (HRs) and their 95% confidence intervals (CIs). Greyed out entries did not pass study diagnostics and are included only for completeness.

| | Patients | | Exposure (KYr) | | Outcomes | | Uncalibrated | | Calibrated | |
|---|---|---|---|---|---|---|---|---|---|---|
| | T | C | T | C | T | C | HR (95% CI) | $p$ | HR (95% CI) | $p$ |
| **PS-stratification / on-treatment TAR** | | | | | | | | | | |
| OptumDOD | 9,018 | 15,153 | 5.3 | 11.6 | 329 | 982 | 0.91 (0.79 - 1.04) | 0.19 | 0.90 (0.78 - 1.03) | 0.17 |
| MDCR | 2,894 | 7,564 | 2.1 | 6.6 | 96 | 409 | 0.96 (0.74 - 1.22) | 0.73 | 0.99 (0.77 - 1.27) | 0.84 |
| Open Claims | 137,023 | 245,572 | 96.5 | 220.8 | 2,911 | 10,481 | 0.87 (0.83 - 0.91) | <0.01 | 0.89 (0.72 - 1.09) | 0.29 |
| OptumEHR | 6,565 | 16,105 | 1.8 | 8.1 | 86 | 535 | 0.90 (0.70 - 1.14) | 0.39 | 0.81 (0.63 - 1.04) | 0.13 |
| VA | 6,609 | 12,626 | 4.2 | 12.8 | 48 | 218 | 0.60 (0.40 - 0.87) | <0.01 | 0.62 (0.42 - 0.92) | 0.02 |
| GermanyDA | 3,202 | 8,059 | 2.4 | 8.3 | 90 | 345 | 1.18 (0.91 - 1.53) | 0.20 | 1.10 (0.84 - 1.42) | 0.51 |
| SIDIAP | 2,815 | 15,113 | 3.4 | 31.7 | 147 | 1,550 | 1.02 (0.84 - 1.23) | 0.81 | 1.06 (0.87 - 1.28) | 0.58 |
| Meta-analysis | 168,126 | 320,192 | 115.7 | 299.8 | 3,707 | 14,520 | 0.91 (0.85 - 0.99) | 0.03 | 0.92 (0.77 - 1.11) | 0.39 |
| **PS-matching / on-treatment TAR** | | | | | | | | | | |
| OptumDOD | 7,055 | 14,990 | 4.5 | 11.5 | 287 | 956 | 0.93 (0.78 - 1.11) | 0.43 | 0.89 (0.75 - 1.07) | 0.24 |
| MDCR | 2,386 | 7,538 | 1.9 | 6.6 | 86 | 407 | 0.76 (0.55 - 1.04) | 0.10 | 0.77 (0.56 - 1.07) | 0.17 |
| Open Claims | 102,627 | 244,869 | 79.3 | 220.3 | 2,512 | 10,406 | 0.92 (0.87 - 0.98) | <0.01 | 0.92 (0.75 - 1.13) | 0.45 |
| OptumEHR | 5,785 | 16,004 | 1.6 | 8.1 | 81 | 522 | 1.03 (0.75 - 1.39) | 0.86 | 0.96 (0.71 - 1.31) | 0.76 |
| VA | 4,043 | 12,255 | 3.0 | 12.2 | 28 | 205 | 0.55 (0.32 - 0.90) | 0.02 | 0.59 (0.36 - 0.99) | 0.05 |
| GermanyDA | 2,640 | 8,032 | 2.2 | 8.3 | 81 | 343 | 1.16 (0.85 - 1.57) | 0.35 | 1.10 (0.80 - 1.50) | 0.54 |
| SIDIAP | 2,325 | 14,631 | 3.0 | 30.2 | 126 | 1,447 | 1.02 (0.81 - 1.27) | 0.84 | 1.07 (0.85 - 1.34) | 0.59 |
| Meta-analysis | 126,861 | 318,319 | 95.5 | 297.0 | 3,201 | 14,286 | 0.92 (0.88 - 0.97) | <0.01 | 0.92 (0.73 - 1.14) | 0.44 |
| **PS-stratification / intent-to-treat TAR** | | | | | | | | | | |
| OptumDOD | 9,018 | 15,153 | 12.6 | 33.8 | 825 | 2,713 | 0.97 (0.89 - 1.06) | 0.50 | 0.99 (0.90 - 1.08) | 0.78 |
| MDCR | 2,894 | 7,564 | 4.5 | 15.0 | 236 | 974 | 1.00 (0.86 - 1.17) | 0.98 | 1.00 (0.85 - 1.18) | 0.87 |
| Open Claims | 137,023 | 245,572 | 320.7 | 938.7 | 11,149 | 45,730 | 0.95 (0.93 - 0.97) | <0.01 | 0.97 (0.82 - 1.14) | 0.68 |
| OptumEHR | 6,565 | 16,105 | 12.6 | 42.6 | 421 | 1,884 | 0.94 (0.84 - 1.06) | 0.31 | 0.93 (0.83 - 1.05) | 0.29 |
| VA | 6,609 | 12,626 | 7.2 | 28.1 | 111 | 505 | 0.82 (0.63 - 1.05) | 0.12 | 0.86 (0.66 - 1.11) | 0.27 |
| GermanyDA | 3,202 | 8,059 | 6.3 | 25.4 | 221 | 939 | 1.17 (0.99 - 1.37) | 0.06 | 1.13 (0.96 - 1.33) | 0.19 |
| SIDIAP | 2,815 | 15,113 | 4.8 | 41.0 | 224 | 2,095 | 1.09 (0.94 - 1.27) | 0.25 | 1.12 (0.96 - 1.31) | 0.21 |
| Meta-analysis | 168,126 | 320,192 | 368.7 | 1124.6 | 13,187 | 54,840 | 0.98 (0.93 - 1.03) | 0.44 | 1.00 (0.85 - 1.16) | 0.91 |
| **PS-matching / intent-to-treat TAR** | | | | | | | | | | |
| OptumDOD | 7,055 | 14,990 | 11.5 | 33.3 | 756 | 2,647 | 0.94 (0.85 - 1.04) | 0.23 | 0.94 (0.85 - 1.04) | 0.30 |
| MDCR | 2,386 | 7,538 | 4.2 | 14.9 | 220 | 967 | 0.93 (0.77 - 1.11) | 0.44 | 0.95 (0.78 - 1.16) | 0.72 |
| Open Claims | 102,627 | 244,869 | 288.9 | 936.1 | 10,428 | 45,411 | 0.95 (0.93 - 0.98) | <0.01 | 0.97 (0.82 - 1.13) | 0.67 |
| OptumEHR | 5,785 | 16,004 | 11.9 | 42.3 | 408 | 1,844 | 0.95 (0.84 - 1.08) | 0.47 | 0.94 (0.82 - 1.07) | 0.38 |
| VA | 4,043 | 12,255 | 5.4 | 26.4 | 74 | 473 | 0.76 (0.55 - 1.03) | 0.08 | 0.79 (0.58 - 1.08) | 0.16 |
| GermanyDA | 2,640 | 8,032 | 5.8 | 25.3 | 210 | 934 | 1.19 (1.00 - 1.41) | 0.05 | 1.16 (0.97 - 1.38) | 0.17 |
| SIDIAP | 2,325 | 14,631 | 4.3 | 39.1 | 202 | 1,943 | 1.09 (0.91 - 1.30) | 0.33 | 1.12 (0.93 - 1.34) | 0.29 |
| Meta-analysis | 126,861 | 318,319 | 332.0 | 1117.3 | 12,298 | 54,219 | 0.96 (0.94 - 0.98) | <0.01 | 0.97 (0.84 - 1.12) | 0.65 |

##### 6.4.9 Hyperkalemia

**Supplementary Table 170:** Exposures, outcomes and risk estimates for Hyperkalemia in SGLT2I (T) vs DPP4I (C) new-user cohorts under four different propensity score (PS) adjustment / time-at-risk (TAR) designs across data sources. We report total patient exposure time in 1,000 patient-year (KYr) units, uncalibrated and calibrated hazard ratios (HRs) and their 95% confidence intervals (CIs). Greyed out entries did not pass study diagnostics and are included only for completeness.

| | Patients | | Exposure (KYr) | | Outcomes | | Uncalibrated | | Calibrated | |
|---|---|---|---|---|---|---|---|---|---|---|
| | T | C | T | C | T | C | HR (95% CI) | $p$ | HR (95% CI) | $p$ |
| **PS-stratification / on-treatment TAR** | | | | | | | | | | |
| OptumDOD | 10,938 | 20,150 | 6.5 | 15.7 | 162 | 565 | 0.83 (0.68 - 1.00) | 0.06 | 0.82 (0.67 - 0.99) | 0.05 |
| MDCR | 3,408 | 9,650 | 2.5 | 8.6 | 25 | 172 | 0.62 (0.38 - 0.97) | 0.05 | 0.64 (0.40 - 1.02) | 0.07 |
| Open Claims | 178,385 | 329,485 | 124.1 | 302.0 | 871 | 3,813 | 0.68 (0.63 - 0.74) | <0.01 | 0.70 (0.56 - 0.87) | <0.01 |
| OptumEHR | 7,603 | 19,295 | 2.1 | 10.0 | 31 | 271 | 0.58 (0.38 - 0.84) | <0.01 | 0.52 (0.35 - 0.78) | <0.01 |
| VA | 6,726 | 13,484 | 4.2 | 13.4 | 76 | 217 | 0.94 (0.67 - 1.30) | 0.69 | 0.97 (0.70 - 1.36) | 0.84 |
| GermanyDA | 3,819 | 9,818 | 3.0 | 10.5 | 45 | 202 | 0.67 (0.46 - 0.96) | 0.03 | 0.62 (0.43 - 0.90) | 0.01 |
| SIDIAP | 3,509 | 18,795 | 4.2 | 40.3 | 62 | 540 | 0.92 (0.67 - 1.24) | 0.59 | 0.95 (0.70 - 1.29) | 0.72 |
| Meta-analysis | 214,388 | 420,677 | 146.6 | 400.4 | 1,272 | 5,780 | 0.74 (0.66 - 0.84) | <0.01 | 0.75 (0.61 - 0.93) | <0.01 |
| **PS-matching / on-treatment TAR** | | | | | | | | | | |
| OptumDOD | 8,761 | 19,826 | 5.7 | 15.4 | 141 | 547 | 0.78 (0.60 - 1.01) | 0.06 | 0.75 (0.58 - 0.97) | 0.04 |
| MDCR | 2,844 | 9,629 | 2.3 | 8.6 | 23 | 172 | 0.56 (0.30 - 0.97) | 0.05 | 0.56 (0.31 - 1.01) | 0.07 |
| Open Claims | 136,020 | 328,574 | 103.5 | 301.2 | 688 | 3,769 | 0.66 (0.59 - 0.74) | <0.01 | 0.66 (0.53 - 0.83) | <0.01 |
| OptumEHR | 6,746 | 19,232 | 1.9 | 10.0 | 25 | 271 | 0.57 (0.33 - 0.92) | 0.03 | 0.53 (0.32 - 0.89) | 0.02 |
| VA | 4,211 | 13,135 | 3.1 | 12.8 | 47 | 208 | 1.01 (0.65 - 1.51) | 0.98 | 1.08 (0.71 - 1.64) | 0.74 |
| GermanyDA | 3,155 | 9,764 | 2.6 | 10.4 | 33 | 199 | 0.62 (0.38 - 0.97) | 0.04 | 0.58 (0.36 - 0.94) | 0.03 |
| SIDIAP | 2,919 | 18,303 | 3.7 | 38.7 | 43 | 513 | 0.88 (0.58 - 1.29) | 0.52 | 0.92 (0.61 - 1.38) | 0.69 |
| Meta-analysis | 164,656 | 418,463 | 122.7 | 397.1 | 1,000 | 5,679 | 0.71 (0.62 - 0.80) | <0.01 | 0.70 (0.55 - 0.90) | <0.01 |
| **PS-stratification / intent-to-treat TAR** | | | | | | | | | | |
| OptumDOD | 10,938 | 20,150 | 16.5 | 48.7 | 400 | 1,646 | 0.81 (0.72 - 0.91) | <0.01 | 0.83 (0.73 - 0.93) | <0.01 |
| MDCR | 3,408 | 9,650 | 5.7 | 20.4 | 71 | 427 | 0.77 (0.58 - 1.00) | 0.06 | 0.77 (0.59 - 1.01) | 0.09 |
| Open Claims | 178,385 | 329,485 | 440.7 | 1367.9 | 3,397 | 17,223 | 0.76 (0.73 - 0.79) | <0.01 | 0.77 (0.65 - 0.91) | <0.01 |
| OptumEHR | 7,603 | 19,295 | 15.4 | 54.0 | 176 | 927 | 0.78 (0.66 - 0.93) | <0.01 | 0.77 (0.65 - 0.92) | <0.01 |
| VA | 6,726 | 13,484 | 7.3 | 29.8 | 134 | 487 | 0.91 (0.71 - 1.16) | 0.45 | 0.96 (0.75 - 1.23) | 0.76 |
| GermanyDA | 3,819 | 9,818 | 7.8 | 32.8 | 120 | 634 | 0.83 (0.66 - 1.02) | 0.09 | 0.80 (0.64 - 1.00) | 0.06 |
| SIDIAP | 3,509 | 18,795 | 6.0 | 53.0 | 94 | 708 | 1.01 (0.78 - 1.28) | 0.95 | 1.03 (0.81 - 1.32) | 0.76 |
| Meta-analysis | 214,388 | 420,677 | 499.5 | 1606.6 | 4,392 | 22,052 | 0.80 (0.75 - 0.86) | <0.01 | 0.81 (0.69 - 0.96) | 0.01 |
| **PS-matching / intent-to-treat TAR** | | | | | | | | | | |
| OptumDOD | 8,761 | 19,826 | 15.2 | 47.6 | 361 | 1,583 | 0.78 (0.68 - 0.89) | <0.01 | 0.78 (0.68 - 0.90) | <0.01 |
| MDCR | 2,844 | 9,629 | 5.4 | 20.4 | 69 | 424 | 0.69 (0.50 - 0.94) | 0.02 | 0.71 (0.51 - 0.98) | 0.07 |
| Open Claims | 136,020 | 328,574 | 401.9 | 1363.9 | 3,076 | 17,055 | 0.75 (0.72 - 0.79) | <0.01 | 0.76 (0.65 - 0.90) | <0.01 |
| OptumEHR | 6,746 | 19,232 | 14.7 | 53.7 | 164 | 922 | 0.84 (0.69 - 1.01) | 0.06 | 0.82 (0.68 - 0.99) | 0.06 |
| VA | 4,211 | 13,135 | 5.6 | 28.1 | 88 | 461 | 0.97 (0.72 - 1.29) | 0.82 | 1.01 (0.75 - 1.36) | 0.87 |
| GermanyDA | 3,155 | 9,764 | 7.2 | 32.5 | 106 | 627 | 0.85 (0.67 - 1.08) | 0.20 | 0.83 (0.65 - 1.06) | 0.17 |
| SIDIAP | 2,919 | 18,303 | 5.5 | 50.8 | 71 | 665 | 0.97 (0.71 - 1.30) | 0.84 | 0.99 (0.73 - 1.35) | 0.81 |
| Meta-analysis | 164,656 | 418,463 | 455.5 | 1597.1 | 3,935 | 21,737 | 0.79 (0.74 - 0.85) | <0.01 | 0.80 (0.68 - 0.93) | <0.01 |

##### 6.4.10 Hypoglycemia

**Supplementary Table 171:** Exposures, outcomes and risk estimates for Hypoglycemia in SGLT2I (T) vs DPP4I (C) new-user cohorts under four different propensity score (PS) adjustment / time-at-risk (TAR) designs across data sources. We report total patient exposure time in 1,000 patient-year (KYr) units, uncalibrated and calibrated hazard ratios (HRs) and their 95% confidence intervals (CIs). Greyed out entries did not pass study diagnostics and are included only for completeness.

| | Patients | | Exposure (KYr) | | Outcomes | | Uncalibrated | | Calibrated | |
|---|---|---|---|---|---|---|---|---|---|---|
| | T | C | T | C | T | C | HR (95% CI) | $p$ | HR (95% CI) | $p$ |
| **PS-stratification / on-treatment TAR** | | | | | | | | | | |
| OptumDOD | 11,439 | 21,482 | 6.8 | 17.1 | 48 | 143 | 1.01 (0.70 - 1.44) | 0.95 | 1.00 (0.69 - 1.43) | 0.93 |
| MDCR | 3,474 | 9,898 | 2.5 | 8.9 | <5 | 43 | 0.39 (0.11 - 1.02) | 0.09 | 0.41 (0.14 - 1.21) | 0.11 |
| Open Claims | 181,772 | 336,560 | 126.5 | 311.2 | 247 | 1,035 | 0.76 (0.65 - 0.88) | <0.01 | 0.78 (0.60 - 1.00) | 0.06 |
| OptumEHR | 7,748 | 19,865 | 2.2 | 10.4 | <5 | 94 | 0.27 (0.08 - 0.67) | 0.02 | 0.24 (0.08 - 0.71) | <0.01 |
| VA | 7,425 | 14,475 | 4.7 | 14.6 | <10 | 34 | 1.56 (0.61 - 3.62) | 0.33 | 1.62 (0.67 - 3.93) | 0.29 |
| GermanyDA | 4,153 | 10,715 | 3.2 | 11.6 | <5 | 22 | 1.10 (0.25 - 3.37) | 0.88 | 1.02 (0.28 - 3.73) | 0.92 |
| SIDIAP | 3,739 | 20,488 | 4.5 | 44.5 | 0 | 33 | | | | |
| Meta-analysis | 216,011 | 412,995 | 145.9 | 373.9 | <320 | 1,371 | 0.80 (0.58 - 1.10) | 0.16 | 0.80 (0.56 - 1.15) | 0.23 |
| **PS-matching / on-treatment TAR** | | | | | | | | | | |
| OptumDOD | 9,150 | 21,098 | 6.0 | 16.8 | 43 | 139 | 1.11 (0.69 - 1.75) | 0.67 | 1.06 (0.67 - 1.69) | 0.80 |
| MDCR | 2,896 | 9,872 | 2.3 | 8.9 | <5 | 43 | 0.44 (0.07 - 1.77) | 0.33 | 0.45 (0.09 - 2.32) | 0.34 |
| Open Claims | 138,168 | 335,542 | 105.3 | 310.4 | 216 | 1,029 | 0.78 (0.63 - 0.96) | 0.02 | 0.78 (0.59 - 1.04) | 0.10 |
| OptumEHR | 6,900 | 19,792 | 1.9 | 10.4 | <5 | 94 | 0.39 (0.09 - 1.15) | 0.14 | 0.36 (0.10 - 1.30) | 0.12 |
| VA | 4,550 | 13,997 | 3.3 | 13.8 | <10 | 32 | 1.92 (0.56 - 5.76) | 0.27 | 2.06 (0.64 - 6.61) | 0.23 |
| GermanyDA | 3,415 | 10,656 | 2.8 | 11.5 | <5 | 22 | 0.38 (0.02 - 2.00) | 0.40 | 0.36 (0.04 - 3.54) | 0.38 |
| SIDIAP | 3,112 | 19,855 | 4.0 | 42.5 | 0 | 32 | | | | |
| Meta-analysis | 165,079 | 410,957 | 121.7 | 371.8 | <284 | 1,359 | 0.84 (0.65 - 1.10) | 0.20 | 0.84 (0.59 - 1.17) | 0.30 |
| **PS-stratification / intent-to-treat TAR** | | | | | | | | | | |
| OptumDOD | 11,439 | 21,482 | 17.3 | 53.6 | 171 | 670 | 0.96 (0.80 - 1.15) | 0.65 | 0.97 (0.81 - 1.17) | 0.79 |
| MDCR | 3,474 | 9,898 | 5.8 | 21.3 | 26 | 188 | 0.60 (0.38 - 0.91) | 0.02 | 0.60 (0.38 - 0.93) | 0.03 |
| Open Claims | 181,772 | 336,560 | 448.6 | 1414.1 | 1,553 | 7,931 | 0.88 (0.83 - 0.93) | <0.01 | 0.89 (0.75 - 1.06) | 0.21 |
| OptumEHR | 7,748 | 19,865 | 15.9 | 56.8 | 82 | 361 | 1.04 (0.80 - 1.34) | 0.77 | 1.03 (0.79 - 1.33) | 0.83 |
| VA | 7,425 | 14,475 | 8.0 | 32.5 | 21 | 115 | 0.93 (0.52 - 1.60) | 0.80 | 0.98 (0.56 - 1.72) | 0.93 |
| GermanyDA | 4,153 | 10,715 | 8.7 | 36.9 | 8 | 80 | 0.71 (0.31 - 1.42) | 0.37 | 0.68 (0.32 - 1.47) | 0.34 |
| SIDIAP | 3,739 | 20,488 | 6.5 | 58.5 | 0 | 74 | | | | |
| Meta-analysis | 216,011 | 412,995 | 504.3 | 1615.2 | 1,861 | 9,345 | 0.89 (0.84 - 0.93) | <0.01 | 0.90 (0.77 - 1.05) | 0.18 |
| **PS-matching / intent-to-treat TAR** | | | | | | | | | | |
| OptumDOD | 9,150 | 21,098 | 15.9 | 52.2 | 159 | 645 | 1.01 (0.82 - 1.25) | 0.89 | 1.02 (0.82 - 1.26) | 0.88 |
| MDCR | 2,896 | 9,872 | 5.5 | 21.2 | 25 | 188 | 0.61 (0.36 - 1.00) | 0.06 | 0.63 (0.37 - 1.05) | 0.10 |
| Open Claims | 138,168 | 335,542 | 408.7 | 1409.4 | 1,480 | 7,857 | 0.88 (0.83 - 0.94) | <0.01 | 0.89 (0.75 - 1.06) | 0.20 |
| OptumEHR | 6,900 | 19,792 | 15.1 | 56.4 | 80 | 360 | 1.09 (0.81 - 1.44) | 0.56 | 1.07 (0.80 - 1.42) | 0.66 |
| VA | 4,550 | 13,997 | 6.0 | 30.1 | 15 | 107 | 0.99 (0.45 - 2.00) | 0.98 | 1.04 (0.49 - 2.18) | 0.92 |
| GermanyDA | 3,415 | 10,656 | 8.0 | 36.6 | 8 | 80 | 0.83 (0.34 - 1.75) | 0.66 | 0.81 (0.36 - 1.84) | 0.62 |
| SIDIAP | 3,112 | 19,855 | 5.9 | 55.6 | 0 | 71 | | | | |
| Meta-analysis | 165,079 | 410,957 | 459.2 | 1606.0 | 1,767 | 9,237 | 0.91 (0.83 - 1.01) | 0.08 | 0.92 (0.78 - 1.10) | 0.37 |

###### 6.4.11 Hypotension

**Supplementary Table 172:** Exposures, outcomes and risk estimates for Hypotension in SGLT2I (T) vs DPP4I (C) new-user cohorts under four different propensity score (PS) adjustment / time-at-risk (TAR) designs across data sources. We report total patient exposure time in 1,000 patient-year (KYr) units, uncalibrated and calibrated hazard ratios (HRs) and their 95% confidence intervals (CIs). Greyed out entries did not pass study diagnostics and are included only for completeness.

| | Patients | | Exposure (KYr) | | Outcomes | | Uncalibrated | | Calibrated | |
|---|---|---|---|---|---|---|---|---|---|---|
| | T | C | T | C | T | C | HR (95% CI) | $p$ | HR (95% CI) | $p$ |
| **PS-stratification / on-treatment TAR** | | | | | | | | | | |
| OptumDOD | 10,826 | 20,117 | 6.4 | 15.7 | 223 | 551 | 1.12 (0.94 - 1.34) | 0.19 | 1.11 (0.93 - 1.32) | 0.28 |
| MDCR | 3,342 | 9,375 | 2.4 | 8.4 | 60 | 234 | 1.18 (0.85 - 1.62) | 0.31 | 1.22 (0.88 - 1.69) | 0.24 |
| Open Claims | 174,134 | 323,216 | 121.2 | 297.1 | 1,455 | 4,436 | 0.96 (0.89 - 1.02) | 0.20 | 0.98 (0.79 - 1.21) | 0.83 |
| OptumEHR | 7,517 | 19,273 | 2.1 | 10.0 | 46 | 287 | 0.87 (0.61 - 1.21) | 0.42 | 0.79 (0.56 - 1.11) | 0.19 |
| VA | 6,697 | 13,463 | 4.2 | 13.5 | 119 | 205 | 1.26 (0.94 - 1.69) | 0.12 | 1.31 (0.97 - 1.76) | 0.09 |
| GermanyDA | 4,027 | 10,466 | 3.1 | 11.3 | 20 | 76 | 0.73 (0.40 - 1.28) | 0.29 | 0.68 (0.38 - 1.20) | 0.19 |
| SIDIAP | 3,705 | 20,297 | 4.4 | 44.1 | 22 | 120 | 1.62 (0.91 - 2.76) | 0.09 | 1.67 (0.96 - 2.91) | 0.08 |
| Meta-analysis | 210,248 | 416,207 | 143.9 | 400.1 | 1,945 | 5,909 | 1.05 (0.93 - 1.19) | 0.42 | 1.06 (0.86 - 1.30) | 0.59 |
| **PS-matching / on-treatment TAR** | | | | | | | | | | |
| OptumDOD | 8,713 | 19,803 | 5.6 | 15.5 | 183 | 528 | 1.08 (0.85 - 1.35) | 0.53 | 1.03 (0.82 - 1.30) | 0.79 |
| MDCR | 2,781 | 9,357 | 2.2 | 8.3 | 52 | 231 | 1.20 (0.78 - 1.80) | 0.40 | 1.22 (0.80 - 1.85) | 0.40 |
| Open Claims | 133,224 | 322,265 | 101.4 | 296.3 | 1,166 | 4,394 | 0.97 (0.89 - 1.06) | 0.53 | 0.97 (0.78 - 1.21) | 0.81 |
| OptumEHR | 6,705 | 19,208 | 1.9 | 10.0 | 40 | 284 | 0.76 (0.50 - 1.15) | 0.21 | 0.72 (0.47 - 1.09) | 0.14 |
| VA | 4,192 | 13,096 | 3.1 | 12.9 | 66 | 198 | 1.35 (0.91 - 1.98) | 0.13 | 1.45 (0.98 - 2.14) | 0.08 |
| GermanyDA | 3,327 | 10,411 | 2.7 | 11.2 | 16 | 74 | 0.76 (0.38 - 1.46) | 0.43 | 0.73 (0.37 - 1.42) | 0.37 |
| SIDIAP | 3,086 | 19,670 | 3.9 | 42.0 | 19 | 113 | 1.71 (0.86 - 3.24) | 0.11 | 1.80 (0.92 - 3.49) | 0.10 |
| Meta-analysis | 162,028 | 413,810 | 120.9 | 396.2 | 1,542 | 5,822 | 1.02 (0.91 - 1.14) | 0.71 | 1.01 (0.80 - 1.29) | 0.89 |
| **PS-stratification / intent-to-treat TAR** | | | | | | | | | | |
| OptumDOD | 10,826 | 20,117 | 16.2 | 48.8 | 518 | 1,722 | 0.99 (0.89 - 1.10) | 0.86 | 1.01 (0.90 - 1.13) | 0.86 |
| MDCR | 3,342 | 9,375 | 5.4 | 19.7 | 141 | 598 | 1.12 (0.91 - 1.37) | 0.26 | 1.12 (0.91 - 1.38) | 0.36 |
| Open Claims | 174,134 | 323,216 | 428.5 | 1336.1 | 5,437 | 21,882 | 0.95 (0.92 - 0.98) | <0.01 | 0.97 (0.82 - 1.14) | 0.68 |
| OptumEHR | 7,517 | 19,273 | 15.1 | 54.0 | 230 | 974 | 0.96 (0.82 - 1.12) | 0.61 | 0.95 (0.81 - 1.11) | 0.54 |
| VA | 6,697 | 13,463 | 7.2 | 30.0 | 194 | 477 | 1.31 (1.05 - 1.62) | 0.02 | 1.38 (1.11 - 1.71) | <0.01 |
| GermanyDA | 4,027 | 10,466 | 8.4 | 35.9 | 38 | 203 | 0.75 (0.50 - 1.10) | 0.16 | 0.73 (0.49 - 1.08) | 0.13 |
| SIDIAP | 3,705 | 20,297 | 6.4 | 57.9 | 27 | 176 | 1.22 (0.75 - 1.94) | 0.41 | 1.25 (0.78 - 2.02) | 0.37 |
| Meta-analysis | 210,248 | 416,207 | 487.2 | 1582.2 | 6,585 | 26,032 | 1.01 (0.93 - 1.11) | 0.79 | 1.03 (0.87 - 1.22) | 0.74 |
| **PS-matching / intent-to-treat TAR** | | | | | | | | | | |
| OptumDOD | 8,713 | 19,803 | 15.0 | 47.8 | 463 | 1,653 | 0.93 (0.82 - 1.05) | 0.26 | 0.93 (0.82 - 1.06) | 0.31 |
| MDCR | 2,781 | 9,357 | 5.1 | 19.6 | 129 | 592 | 1.07 (0.84 - 1.36) | 0.56 | 1.10 (0.85 - 1.41) | 0.56 |
| Open Claims | 133,224 | 322,265 | 391.3 | 1332.0 | 4,920 | 21,679 | 0.94 (0.90 - 0.97) | <0.01 | 0.95 (0.81 - 1.11) | 0.53 |
| OptumEHR | 6,705 | 19,208 | 14.4 | 53.7 | 216 | 962 | 0.97 (0.81 - 1.15) | 0.72 | 0.95 (0.80 - 1.13) | 0.61 |
| VA | 4,192 | 13,096 | 5.5 | 28.2 | 122 | 450 | 1.29 (0.99 - 1.67) | 0.05 | 1.35 (1.04 - 1.75) | 0.03 |
| GermanyDA | 3,327 | 10,411 | 7.8 | 35.6 | 33 | 198 | 0.70 (0.45 - 1.06) | 0.11 | 0.68 (0.44 - 1.05) | 0.09 |
| SIDIAP | 3,086 | 19,670 | 5.8 | 55.0 | 23 | 163 | 1.02 (0.54 - 1.78) | 0.96 | 1.04 (0.57 - 1.89) | 0.87 |
| Meta-analysis | 162,028 | 413,810 | 444.8 | 1571.9 | 5,906 | 25,697 | 0.94 (0.91 - 0.98) | <0.01 | 0.95 (0.83 - 1.10) | 0.52 |

##### 6.4.12 Joint pain

**Supplementary Table 173:** Exposures, outcomes and risk estimates for Joint pain in SGLT2I (T) vs DPP4I (C) new-user cohorts under four different propensity score (PS) adjustment / time-at-risk (TAR) designs across data sources. We report total patient exposure time in 1,000 patient-year (KYr) units, uncalibrated and calibrated hazard ratios (HRs) and their 95% confidence intervals (CIs). Greyed out entries did not pass study diagnostics and are included only for completeness.

| | Patients | | Exposure (KYr) | | Outcomes | | Uncalibrated | | Calibrated | |
|---|---|---|---|---|---|---|---|---|---|---|
| | T | C | T | C | T | C | HR (95% CI) | $p$ | HR (95% CI) | $p$ |
| **PS-stratification / on-treatment TAR** | | | | | | | | | | |
| OptumDOD | 10,899 | 20,405 | 6.5 | 16.1 | 82 | 261 | 0.87 (0.66 - 1.14) | 0.33 | 0.86 (0.66 - 1.13) | 0.30 |
| MDCR | 3,368 | 9,564 | 2.5 | 8.6 | 25 | 108 | 0.76 (0.46 - 1.20) | 0.26 | 0.78 (0.49 - 1.26) | 0.33 |
| Open Claims | 172,024 | 320,780 | 120.0 | 295.8 | 623 | 2,126 | 0.84 (0.76 - 0.92) | <0.01 | 0.85 (0.68 - 1.07) | 0.19 |
| OptumEHR | 7,405 | 18,976 | 2.0 | 9.9 | 25 | 101 | 1.52 (0.93 - 2.41) | 0.09 | 1.37 (0.85 - 2.22) | 0.21 |
| VA | 6,617 | 12,920 | 4.2 | 13.2 | 25 | 55 | 1.10 (0.60 - 1.97) | 0.76 | 1.14 (0.63 - 2.07) | 0.67 |
| GermanyDA | 3,534 | 9,208 | 2.7 | 9.7 | 36 | 190 | 0.85 (0.56 - 1.25) | 0.42 | 0.78 (0.53 - 1.17) | 0.26 |
| SIDIAP | 2,847 | 16,060 | 3.4 | 34.2 | 63 | 737 | 0.94 (0.70 - 1.24) | 0.67 | 0.97 (0.73 - 1.29) | 0.76 |
| Meta-analysis | 206,694 | 407,913 | 141.3 | 387.5 | 879 | 3,578 | 0.87 (0.80 - 0.95) | <0.01 | 0.88 (0.73 - 1.06) | 0.18 |
| **PS-matching / on-treatment TAR** | | | | | | | | | | |
| OptumDOD | 8,712 | 20,040 | 5.7 | 15.8 | 74 | 253 | 0.82 (0.57 - 1.16) | 0.27 | 0.79 (0.55 - 1.12) | 0.19 |
| MDCR | 2,822 | 9,539 | 2.2 | 8.5 | 21 | 108 | 0.79 (0.40 - 1.47) | 0.47 | 0.80 (0.42 - 1.53) | 0.52 |
| Open Claims | 130,810 | 319,866 | 99.9 | 295.1 | 526 | 2,117 | 0.81 (0.70 - 0.92) | <0.01 | 0.81 (0.64 - 1.03) | 0.09 |
| OptumEHR | 6,588 | 18,909 | 1.8 | 9.9 | 22 | 100 | 1.42 (0.73 - 2.66) | 0.29 | 1.33 (0.69 - 2.54) | 0.40 |
| VA | 4,078 | 12,531 | 3.0 | 12.5 | 12 | 54 | 1.55 (0.62 - 3.67) | 0.33 | 1.66 (0.68 - 4.05) | 0.27 |
| GermanyDA | 2,887 | 9,166 | 2.3 | 9.7 | 34 | 189 | 0.82 (0.50 - 1.29) | 0.40 | 0.77 (0.48 - 1.24) | 0.32 |
| SIDIAP | 2,361 | 15,541 | 3.0 | 32.7 | 60 | 689 | 0.93 (0.66 - 1.29) | 0.69 | 0.98 (0.70 - 1.37) | 0.77 |
| Meta-analysis | 158,258 | 405,592 | 118.1 | 384.2 | 749 | 3,510 | 0.84 (0.75 - 0.94) | <0.01 | 0.84 (0.66 - 1.06) | 0.14 |
| **PS-stratification / intent-to-treat TAR** | | | | | | | | | | |
| OptumDOD | 10,899 | 20,405 | 16.4 | 50.5 | 224 | 779 | 0.92 (0.78 - 1.08) | 0.32 | 0.94 (0.79 - 1.10) | 0.46 |
| MDCR | 3,368 | 9,564 | 5.6 | 20.4 | 57 | 247 | 0.91 (0.66 - 1.23) | 0.53 | 0.91 (0.66 - 1.24) | 0.57 |
| Open Claims | 172,024 | 320,780 | 423.4 | 1343.2 | 2,527 | 9,345 | 0.98 (0.94 - 1.03) | 0.46 | 1.00 (0.85 - 1.18) | 0.92 |
| OptumEHR | 7,405 | 18,976 | 15.0 | 53.9 | 147 | 481 | 1.18 (0.96 - 1.43) | 0.11 | 1.17 (0.95 - 1.43) | 0.15 |
| VA | 6,617 | 12,920 | 7.2 | 29.2 | 38 | 139 | 0.91 (0.58 - 1.41) | 0.68 | 0.96 (0.61 - 1.50) | 0.85 |
| GermanyDA | 3,534 | 9,208 | 7.2 | 30.6 | 102 | 501 | 1.03 (0.81 - 1.29) | 0.81 | 1.00 (0.79 - 1.26) | 0.76 |
| SIDIAP | 2,847 | 16,060 | 5.0 | 45.1 | 99 | 936 | 1.06 (0.84 - 1.33) | 0.62 | 1.08 (0.86 - 1.37) | 0.53 |
| Meta-analysis | 206,694 | 407,913 | 479.7 | 1572.9 | 3,194 | 12,428 | 0.99 (0.95 - 1.03) | 0.57 | 1.00 (0.86 - 1.17) | 0.91 |
| **PS-matching / intent-to-treat TAR** | | | | | | | | | | |
| OptumDOD | 8,712 | 20,040 | 15.1 | 49.2 | 210 | 756 | 0.96 (0.80 - 1.15) | 0.66 | 0.96 (0.80 - 1.16) | 0.67 |
| MDCR | 2,822 | 9,539 | 5.3 | 20.3 | 53 | 247 | 0.89 (0.62 - 1.26) | 0.53 | 0.91 (0.63 - 1.32) | 0.66 |
| Open Claims | 130,810 | 319,866 | 385.7 | 1339.0 | 2,350 | 9,308 | 0.98 (0.93 - 1.03) | 0.35 | 0.99 (0.84 - 1.16) | 0.87 |
| OptumEHR | 6,588 | 18,909 | 14.2 | 53.6 | 142 | 479 | 1.17 (0.93 - 1.45) | 0.17 | 1.15 (0.92 - 1.43) | 0.25 |
| VA | 4,078 | 12,531 | 5.4 | 27.3 | 23 | 128 | 1.00 (0.54 - 1.76) | 1.00 | 1.04 (0.57 - 1.89) | 0.89 |
| GermanyDA | 2,887 | 9,166 | 6.6 | 30.4 | 99 | 498 | 1.11 (0.87 - 1.42) | 0.39 | 1.08 (0.84 - 1.39) | 0.56 |
| SIDIAP | 2,361 | 15,541 | 4.5 | 42.9 | 95 | 876 | 1.08 (0.83 - 1.38) | 0.58 | 1.10 (0.85 - 1.43) | 0.50 |
| Meta-analysis | 158,258 | 405,592 | 436.8 | 1562.7 | 2,972 | 12,292 | 0.99 (0.94 - 1.04) | 0.67 | 1.00 (0.86 - 1.16) | 0.91 |

##### 6.4.13 Lower extremity amputation

**Supplementary Table 174:** Exposures, outcomes and risk estimates for Lower extremity amputation in SGLT2I (T) vs DPP4I (C) new-user cohorts under four different propensity score (PS) adjustment / time-at-risk (TAR) designs across data sources. We report total patient exposure time in 1,000 patient-year (KYr) units, uncalibrated and calibrated hazard ratios (HRs) and their 95% confidence intervals (CIs). Greyed out entries did not pass study diagnostics and are included only for completeness.

| | Patients | | Exposure (KYr) | | Outcomes | | Uncalibrated | | Calibrated | |
|---|---|---|---|---|---|---|---|---|---|---|
| | T | C | T | C | T | C | HR (95% CI) | $p$ | HR (95% CI) | $p$ |
| **PS-stratification / on-treatment TAR** | | | | | | | | | | |
| OptumDOD | 11,589 | 21,794 | 6.9 | 17.4 | 6 | 13 | 1.61 (0.52 - 4.48) | 0.38 | 1.59 (0.54 - 4.64) | 0.40 |
| MDCR | 3,500 | 10,004 | 2.6 | 9.0 | <5 | 6 | 4.17 (0.74 - 19.0) | 0.08 | 4.31 (0.85 - 21.8) | 0.08 |
| Open Claims | 184,471 | 341,853 | 128.3 | 316.4 | 67 | 164 | 1.28 (0.92 - 1.77) | 0.14 | 1.31 (0.89 - 1.93) | 0.18 |
| OptumEHR | 7,821 | 20,033 | 2.2 | 10.6 | <5 | 7 | 0.50 (0.03 - 2.88) | 0.56 | 0.45 (0.04 - 4.70) | 0.51 |
| VA | 7,511 | 14,572 | 4.7 | 14.7 | <10 | <10 | 1.87 (0.50 - 7.07) | 0.36 | 1.94 (0.52 - 7.29) | 0.33 |
| GermanyDA | 4,175 | 10,792 | 3.2 | 11.7 | 0 | 0 | | | | |
| SIDIAP | 3,745 | 20,500 | 4.5 | 44.6 | 0 | 8 | | | | |
| Meta-analysis | 203,571 | 378,219 | 140.0 | 348.5 | <83 | <187 | 1.33 (0.98 - 1.81) | 0.07 | 1.34 (0.95 - 1.91) | 0.10 |
| **PS-matching / on-treatment TAR** | | | | | | | | | | |
| OptumDOD | 9,281 | 21,401 | 6.1 | 17.1 | 6 | 13 | 1.73 (0.48 - 5.83) | 0.39 | 1.66 (0.47 - 5.79) | 0.43 |
| MDCR | 2,921 | 9,978 | 2.3 | 9.0 | <5 | 6 | 2.90 (0.35 - 20.7) | 0.30 | 2.95 (0.39 - 22.6) | 0.30 |
| Open Claims | 140,306 | 340,833 | 106.8 | 315.5 | 55 | 160 | 1.88 (1.19 - 2.96) | <0.01 | 1.88 (1.14 - 3.09) | 0.01 |
| OptumEHR | 6,961 | 19,958 | 2.0 | 10.6 | <5 | 7 | | | | |
| VA | 4,587 | 14,155 | 3.4 | 14.0 | <10 | <10 | | 0.22 | | 0.20 |
| GermanyDA | 3,434 | 10,733 | 2.9 | 11.6 | 0 | 0 | | | | |
| SIDIAP | 3,114 | 19,867 | 3.9 | 42.5 | 0 | 7 | | | | |
| Meta-analysis | 140,306 | 340,833 | 106.8 | 315.5 | 55 | 160 | 1.88 (1.19 - 2.96) | <0.01 | 1.86 (1.12 - 3.08) | 0.02 |
| **PS-stratification / intent-to-treat TAR** | | | | | | | | | | |
| OptumDOD | 11,589 | 21,794 | 17.8 | 55.3 | 21 | 68 | 1.09 (0.63 - 1.82) | 0.74 | 1.11 (0.65 - 1.89) | 0.70 |
| MDCR | 3,500 | 10,004 | 5.9 | 21.7 | 7 | 22 | 1.50 (0.52 - 3.86) | 0.43 | 1.50 (0.55 - 4.12) | 0.43 |
| Open Claims | 184,471 | 341,853 | 457.9 | 1452.4 | 255 | 905 | 1.09 (0.93 - 1.27) | 0.27 | 1.11 (0.89 - 1.38) | 0.36 |
| OptumEHR | 7,821 | 20,033 | 16.2 | 58.0 | <5 | 33 | 0.36 (0.10 - 0.95) | 0.07 | 0.36 (0.12 - 1.07) | 0.07 |
| VA | 7,511 | 14,572 | 8.1 | 32.8 | 13 | 24 | 1.55 (0.66 - 3.55) | 0.30 | 1.64 (0.71 - 3.80) | 0.25 |
| GermanyDA | 4,175 | 10,792 | 8.8 | 37.4 | 0 | 0 | | | | |
| SIDIAP | 3,745 | 20,500 | 6.5 | 58.6 | 0 | 11 | | | | |
| Meta-analysis | 214,892 | 408,256 | 505.9 | 1620.1 | <301 | 1,052 | 1.09 (0.94 - 1.25) | 0.25 | 1.11 (0.90 - 1.35) | 0.33 |
| **PS-matching / intent-to-treat TAR** | | | | | | | | | | |
| OptumDOD | 9,281 | 21,401 | 16.4 | 53.8 | 20 | 68 | 1.16 (0.63 - 2.05) | 0.63 | 1.16 (0.64 - 2.09) | 0.63 |
| MDCR | 2,921 | 9,978 | 5.5 | 21.6 | 7 | 22 | 1.74 (0.37 - 6.00) | 0.44 | 1.77 (0.44 - 7.18) | 0.43 |
| Open Claims | 140,306 | 340,833 | 417.5 | 1447.5 | 234 | 890 | 1.18 (1.00 - 1.39) | 0.05 | 1.19 (0.95 - 1.50) | 0.13 |
| OptumEHR | 6,961 | 19,958 | 15.4 | 57.6 | <5 | 33 | 0.44 (0.10 - 1.33) | 0.21 | 0.43 (0.12 - 1.57) | 0.20 |
| VA | 4,587 | 14,155 | 6.1 | 30.8 | 11 | 23 | 1.16 (0.38 - 3.28) | 0.79 | 1.21 (0.41 - 3.57) | 0.73 |
| GermanyDA | 3,434 | 10,733 | 8.1 | 37.1 | 0 | 0 | | | | |
| SIDIAP | 3,114 | 19,867 | 5.9 | 55.7 | 0 | 10 | | | | |
| Meta-analysis | 164,056 | 406,325 | 460.8 | 1611.3 | <277 | 1,036 | 1.17 (1.00 - 1.36) | 0.05 | 1.18 (0.96 - 1.45) | 0.12 |

##### 6.4.14 Nausea

**Supplementary Table 175:** Exposures, outcomes and risk estimates for Nausea in SGLT2I (T) vs DPP4I (C) new-user cohorts under four different propensity score (PS) adjustment / time-at-risk (TAR) designs across data sources. We report total patient exposure time in 1,000 patient-year (KYr) units, uncalibrated and calibrated hazard ratios (HRs) and their 95% confidence intervals (CIs). Greyed out entries did not pass study diagnostics and are included only for completeness.

| | Patients | | Exposure (KYr) | | Outcomes | | Uncalibrated | | Calibrated | |
|---|---|---|---|---|---|---|---|---|---|---|
| | T | C | T | C | T | C | HR (95% CI) | $p$ | HR (95% CI) | $p$ |
| **PS-stratification / on-treatment TAR** | | | | | | | | | | |
| OptumDOD | 9,971 | 18,322 | 5.9 | 14.2 | 215 | 690 | 0.85 (0.71 - 1.00) | 0.05 | 0.83 (0.70 - 0.99) | 0.05 |
| MDCR | 3,120 | 8,773 | 2.3 | 7.8 | 68 | 294 | 0.94 (0.70 - 1.25) | 0.68 | 0.97 (0.73 - 1.30) | 0.83 |
| Open Claims | 158,462 | 295,534 | 111.0 | 271.6 | 1,750 | 5,973 | 0.88 (0.83 - 0.93) | <0.01 | 0.90 (0.73 - 1.11) | 0.33 |
| OptumEHR | 7,090 | 18,119 | 2.0 | 9.4 | 62 | 326 | 1.18 (0.87 - 1.57) | 0.28 | 1.06 (0.79 - 1.44) | 0.69 |
| VA | 6,967 | 13,696 | 4.4 | 13.9 | 30 | 97 | 0.82 (0.47 - 1.39) | 0.48 | 0.86 (0.50 - 1.47) | 0.58 |
| GermanyDA | 3,908 | 9,999 | 3.0 | 10.8 | 20 | 113 | 0.77 (0.45 - 1.26) | 0.33 | 0.71 (0.42 - 1.20) | 0.22 |
| SIDIAP | 3,589 | 19,649 | 4.3 | 42.6 | 17 | 368 | 0.51 (0.29 - 0.83) | 0.01 | 0.52 (0.31 - 0.88) | 0.02 |
| Meta-analysis | 193,107 | 384,092 | 132.9 | 370.1 | 2,162 | 7,861 | 0.88 (0.83 - 0.93) | <0.01 | 0.89 (0.74 - 1.06) | 0.18 |
| **PS-matching / on-treatment TAR** | | | | | | | | | | |
| OptumDOD | 7,961 | 18,031 | 5.2 | 14.0 | 192 | 671 | 0.82 (0.65 - 1.01) | 0.07 | 0.78 (0.63 - 0.97) | 0.04 |
| MDCR | 2,606 | 8,753 | 2.1 | 7.8 | 61 | 293 | 0.87 (0.60 - 1.24) | 0.46 | 0.88 (0.61 - 1.28) | 0.56 |
| Open Claims | 120,491 | 294,522 | 92.3 | 270.7 | 1,495 | 5,937 | 0.86 (0.80 - 0.93) | <0.01 | 0.86 (0.70 - 1.07) | 0.20 |
| OptumEHR | 6,300 | 17,988 | 1.8 | 9.3 | 57 | 322 | 1.29 (0.88 - 1.86) | 0.18 | 1.21 (0.83 - 1.76) | 0.35 |
| VA | 4,301 | 13,319 | 3.2 | 13.2 | 14 | 92 | 0.60 (0.24 - 1.29) | 0.23 | 0.64 (0.28 - 1.49) | 0.31 |
| GermanyDA | 3,205 | 9,959 | 2.6 | 10.7 | 17 | 113 | 0.66 (0.35 - 1.19) | 0.19 | 0.63 (0.34 - 1.17) | 0.15 |
| SIDIAP | 2,988 | 19,075 | 3.8 | 40.7 | 12 | 350 | 0.42 (0.20 - 0.78) | 0.01 | 0.44 (0.23 - 0.86) | 0.02 |
| Meta-analysis | 147,852 | 381,647 | 111.0 | 366.4 | 1,848 | 7,778 | 0.86 (0.80 - 0.92) | <0.01 | 0.85 (0.68 - 1.07) | 0.16 |
| **PS-stratification / intent-to-treat TAR** | | | | | | | | | | |
| OptumDOD | 9,971 | 18,322 | 14.7 | 43.4 | 588 | 2,003 | 0.94 (0.85 - 1.04) | 0.21 | 0.95 (0.86 - 1.06) | 0.42 |
| MDCR | 3,120 | 8,773 | 5.1 | 18.1 | 171 | 733 | 1.01 (0.84 - 1.21) | 0.88 | 1.01 (0.84 - 1.22) | 0.86 |
| Open Claims | 158,462 | 295,534 | 384.3 | 1196.9 | 7,063 | 28,122 | 0.95 (0.92 - 0.97) | <0.01 | 0.96 (0.82 - 1.14) | 0.67 |
| OptumEHR | 7,090 | 18,119 | 14.1 | 49.9 | 301 | 1,229 | 1.02 (0.89 - 1.16) | 0.81 | 1.01 (0.88 - 1.16) | 0.83 |
| VA | 6,967 | 13,696 | 7.6 | 30.6 | 54 | 254 | 0.70 (0.48 - 1.00) | 0.06 | 0.74 (0.51 - 1.07) | 0.11 |
| GermanyDA | 3,908 | 9,999 | 8.1 | 33.9 | 64 | 343 | 1.06 (0.79 - 1.41) | 0.70 | 1.02 (0.76 - 1.37) | 0.79 |
| SIDIAP | 3,589 | 19,649 | 6.2 | 55.8 | 34 | 526 | 0.68 (0.46 - 0.97) | 0.04 | 0.69 (0.47 - 1.01) | 0.06 |
| Meta-analysis | 193,107 | 384,092 | 440.0 | 1428.6 | 8,275 | 33,210 | 0.95 (0.92 - 0.97) | <0.01 | 0.96 (0.83 - 1.12) | 0.63 |
| **PS-matching / intent-to-treat TAR** | | | | | | | | | | |
| OptumDOD | 7,961 | 18,031 | 13.5 | 42.4 | 543 | 1,959 | 0.93 (0.83 - 1.04) | 0.22 | 0.93 (0.83 - 1.05) | 0.28 |
| MDCR | 2,606 | 8,753 | 4.8 | 18.0 | 160 | 731 | 1.03 (0.84 - 1.26) | 0.79 | 1.05 (0.84 - 1.31) | 0.74 |
| Open Claims | 120,491 | 294,522 | 349.6 | 1192.5 | 6,601 | 27,934 | 0.95 (0.92 - 0.98) | <0.01 | 0.96 (0.82 - 1.13) | 0.64 |
| OptumEHR | 6,300 | 17,988 | 13.4 | 49.4 | 287 | 1,207 | 1.06 (0.91 - 1.23) | 0.45 | 1.04 (0.89 - 1.21) | 0.64 |
| VA | 4,301 | 13,319 | 5.8 | 28.8 | 33 | 238 | 0.67 (0.42 - 1.04) | 0.09 | 0.70 (0.44 - 1.11) | 0.13 |
| GermanyDA | 3,205 | 9,959 | 7.5 | 33.7 | 60 | 338 | 1.11 (0.81 - 1.50) | 0.52 | 1.08 (0.79 - 1.47) | 0.65 |
| SIDIAP | 2,988 | 19,075 | 5.7 | 53.2 | 29 | 493 | 0.61 (0.38 - 0.92) | 0.03 | 0.62 (0.40 - 0.97) | 0.04 |
| Meta-analysis | 147,852 | 381,647 | 400.2 | 1418.2 | 7,713 | 32,900 | 0.95 (0.93 - 0.98) | <0.01 | 0.96 (0.83 - 1.11) | 0.60 |

##### 6.4.15 Peripheral edema

**Supplementary Table 176:** Exposures, outcomes and risk estimates for Peripheral edema in SGLT2I (T) vs DPP4I (C) new-user cohorts under four different propensity score (PS) adjustment / time-at-risk (TAR) designs across data sources. We report total patient exposure time in 1,000 patient-year (KYr) units, uncalibrated and calibrated hazard ratios (HRs) and their 95% confidence intervals (CIs). Greyed out entries did not pass study diagnostics and are included only for completeness.

| | Patients | | Exposure (KYr) | | Outcomes | | Uncalibrated | | Calibrated | |
|---|---|---|---|---|---|---|---|---|---|---|
| | T | C | T | C | T | C | HR (95% CI) | $p$ | HR (95% CI) | $p$ |
| **PS-stratification / on-treatment TAR** | | | | | | | | | | |
| OptumDOD | 8,491 | 15,272 | 5.0 | 11.4 | 343 | 1,195 | 0.76 (0.67 - 0.87) | <0.01 | 0.75 (0.66 - 0.86) | <0.01 |
| MDCR | 2,647 | 7,141 | 1.9 | 6.1 | 82 | 551 | 0.61 (0.47 - 0.78) | <0.01 | 0.63 (0.49 - 0.81) | <0.01 |
| Open Claims | 132,741 | 248,533 | 93.2 | 222.6 | 2,437 | 10,636 | 0.71 (0.68 - 0.75) | <0.01 | 0.73 (0.59 - 0.89) | <0.01 |
| OptumEHR | 6,272 | 16,106 | 1.7 | 8.0 | 47 | 518 | 0.59 (0.43 - 0.80) | <0.01 | 0.54 (0.39 - 0.74) | <0.01 |
| VA | 5,373 | 11,017 | 3.4 | 11.0 | 104 | 423 | 0.71 (0.54 - 0.92) | 0.01 | 0.74 (0.56 - 0.97) | 0.03 |
| GermanyDA | 3,389 | 8,757 | 2.6 | 9.3 | 56 | 353 | 0.60 (0.43 - 0.81) | <0.01 | 0.55 (0.40 - 0.76) | <0.01 |
| SIDIAP | 3,190 | 17,958 | 3.9 | 38.2 | 42 | 939 | 0.50 (0.35 - 0.68) | <0.01 | 0.51 (0.37 - 0.72) | <0.01 |
| Meta-analysis | 162,103 | 324,784 | 111.7 | 306.6 | 3,111 | 14,615 | 0.67 (0.61 - 0.74) | <0.01 | 0.68 (0.56 - 0.83) | <0.01 |
| **PS-matching / on-treatment TAR** | | | | | | | | | | |
| OptumDOD | 6,834 | 15,067 | 4.4 | 11.3 | 302 | 1,163 | 0.73 (0.61 - 0.87) | <0.01 | 0.70 (0.59 - 0.84) | <0.01 |
| MDCR | 2,188 | 7,101 | 1.7 | 6.1 | 72 | 544 | 0.66 (0.47 - 0.91) | 0.01 | 0.67 (0.48 - 0.93) | 0.03 |
| Open Claims | 102,215 | 247,958 | 78.2 | 222.2 | 2,088 | 10,565 | 0.71 (0.66 - 0.75) | <0.01 | 0.71 (0.58 - 0.87) | <0.01 |
| OptumEHR | 5,642 | 15,998 | 1.6 | 7.9 | 46 | 510 | 0.67 (0.45 - 0.96) | 0.04 | 0.63 (0.43 - 0.92) | 0.02 |
| VA | 3,400 | 10,752 | 2.5 | 10.6 | 70 | 403 | 0.65 (0.46 - 0.91) | 0.01 | 0.70 (0.50 - 0.98) | 0.05 |
| GermanyDA | 2,793 | 8,715 | 2.3 | 9.2 | 48 | 350 | 0.65 (0.43 - 0.96) | 0.04 | 0.62 (0.42 - 0.93) | 0.03 |
| SIDIAP | 2,656 | 17,306 | 3.4 | 36.3 | 37 | 874 | 0.57 (0.38 - 0.83) | <0.01 | 0.60 (0.41 - 0.88) | 0.01 |
| Meta-analysis | 125,728 | 322,897 | 94.1 | 303.5 | 2,663 | 14,409 | 0.70 (0.66 - 0.74) | <0.01 | 0.70 (0.56 - 0.87) | <0.01 |
| **PS-stratification / intent-to-treat TAR** | | | | | | | | | | |
| OptumDOD | 8,491 | 15,272 | 12.2 | 33.2 | 927 | 3,202 | 0.87 (0.80 - 0.94) | <0.01 | 0.88 (0.81 - 0.96) | 0.01 |
| MDCR | 2,647 | 7,141 | 4.1 | 13.7 | 261 | 1,249 | 0.86 (0.74 - 0.99) | 0.04 | 0.86 (0.74 - 1.00) | 0.13 |
| Open Claims | 132,741 | 248,533 | 316.7 | 955.7 | 10,335 | 44,728 | 0.88 (0.86 - 0.90) | <0.01 | 0.89 (0.76 - 1.05) | 0.18 |
| OptumEHR | 6,272 | 16,106 | 12.3 | 42.1 | 421 | 2,108 | 0.84 (0.75 - 0.94) | <0.01 | 0.83 (0.74 - 0.93) | <0.01 |
| VA | 5,373 | 11,017 | 5.9 | 24.0 | 198 | 865 | 0.83 (0.68 - 1.01) | 0.06 | 0.88 (0.72 - 1.06) | 0.22 |
| GermanyDA | 3,389 | 8,757 | 6.9 | 28.6 | 154 | 993 | 0.71 (0.59 - 0.85) | <0.01 | 0.69 (0.57 - 0.83) | <0.01 |
| SIDIAP | 3,190 | 17,958 | 5.6 | 49.9 | 83 | 1,247 | 0.69 (0.54 - 0.88) | <0.01 | 0.71 (0.56 - 0.90) | <0.01 |
| Meta-analysis | 162,103 | 324,784 | 363.7 | 1147.2 | 12,379 | 54,392 | 0.85 (0.82 - 0.89) | <0.01 | 0.87 (0.75 - 1.01) | 0.07 |
| **PS-matching / intent-to-treat TAR** | | | | | | | | | | |
| OptumDOD | 6,834 | 15,067 | 11.2 | 32.7 | 858 | 3,113 | 0.83 (0.76 - 0.91) | <0.01 | 0.83 (0.75 - 0.92) | <0.01 |
| MDCR | 2,188 | 7,101 | 3.8 | 13.7 | 246 | 1,238 | 0.84 (0.70 - 0.99) | 0.04 | 0.85 (0.71 - 1.03) | 0.24 |
| Open Claims | 102,215 | 247,958 | 289.1 | 953.7 | 9,678 | 44,460 | 0.89 (0.86 - 0.91) | <0.01 | 0.90 (0.76 - 1.05) | 0.19 |
| OptumEHR | 5,642 | 15,998 | 11.7 | 41.8 | 414 | 2,070 | 0.85 (0.75 - 0.96) | 0.01 | 0.83 (0.73 - 0.95) | 0.01 |
| VA | 3,400 | 10,752 | 4.5 | 22.8 | 143 | 813 | 0.81 (0.64 - 1.01) | 0.07 | 0.84 (0.67 - 1.06) | 0.17 |
| GermanyDA | 2,793 | 8,715 | 6.4 | 28.4 | 142 | 982 | 0.71 (0.58 - 0.87) | <0.01 | 0.69 (0.56 - 0.85) | <0.01 |
| SIDIAP | 2,656 | 17,306 | 5.1 | 47.1 | 77 | 1,153 | 0.76 (0.58 - 0.99) | 0.05 | 0.78 (0.60 - 1.02) | 0.10 |
| Meta-analysis | 125,728 | 322,897 | 331.9 | 1140.2 | 11,558 | 53,829 | 0.85 (0.80 - 0.89) | <0.01 | 0.85 (0.74 - 0.99) | 0.04 |

##### 6.4.16 Photosensitivity

**Supplementary Table 177:** Exposures, outcomes and risk estimates for Photosensitivity in SGLT2I (T) vs DPP4I (C) new-user cohorts under four different propensity score (PS) adjustment / time-at-risk (TAR) designs across data sources. We report total patient exposure time in 1,000 patient-year (KYr) units, uncalibrated and calibrated hazard ratios (HRs) and their 95% confidence intervals (CIs). Greyed out entries did not pass study diagnostics and are included only for completeness.

| | Patients | | Exposure (KYr) | | Outcomes | | Uncalibrated | | Calibrated | |
|---|---|---|---|---|---|---|---|---|---|---|
| | T | C | T | C | T | C | HR (95% CI) | $p$ | HR (95% CI) | $p$ |
| **PS-stratification / on-treatment TAR** | | | | | | | | | | |
| OptumDOD | 11,516 | 21,588 | 6.9 | 17.2 | <5 | 10 | 1.46 (0.27 - 5.80) | 0.63 | 1.44 (0.31 - 6.64) | 0.64 |
| MDCR | 3,464 | 9,818 | 2.5 | 8.8 | <5 | 22 | 0.90 (0.20 - 2.80) | 0.88 | 0.93 (0.25 - 3.45) | 0.92 |
| Open Claims | 181,939 | 337,293 | 126.5 | 311.8 | 29 | 183 | 0.95 (0.61 - 1.43) | 0.82 | 0.97 (0.61 - 1.55) | 0.90 |
| OptumEHR | 7,758 | 19,847 | 2.2 | 10.5 | 0 | 22 | | | | |
| VA | 7,386 | 14,331 | 4.6 | 14.5 | 0 | <10 | | | | |
| GermanyDA | 4,167 | 10,750 | 3.2 | 11.7 | 0 | 0 | | | | |
| SIDIAP | 3,737 | 20,476 | 4.4 | 44.6 | 0 | 6 | | | | |
| Meta-analysis | 185,403 | 347,111 | 129.0 | 320.6 | <34 | 205 | 0.95 (0.63 - 1.41) | 0.79 | 0.95 (0.62 - 1.47) | 0.83 |
| **PS-matching / on-treatment TAR** | | | | | | | | | | |
| OptumDOD | 9,215 | 21,198 | 6.0 | 16.9 | <5 | 10 | 0.96 (0.12 - 5.27) | 0.97 | 0.92 (0.14 - 5.98) | 0.93 |
| MDCR | 2,887 | 9,793 | 2.3 | 8.8 | <5 | 22 | 1.26 (0.25 - 4.84) | 0.76 | 1.28 (0.29 - 5.58) | 0.75 |
| Open Claims | 138,358 | 336,236 | 105.3 | 310.9 | 28 | 182 | 0.86 (0.51 - 1.41) | 0.57 | 0.86 (0.50 - 1.50) | 0.61 |
| OptumEHR | 6,908 | 19,770 | 1.9 | 10.4 | 0 | 22 | | | | |
| VA | 4,518 | 13,831 | 3.3 | 13.6 | 0 | <10 | | | | |
| GermanyDA | 3,424 | 10,691 | 2.8 | 11.6 | 0 | 0 | | | | |
| SIDIAP | 3,110 | 19,842 | 3.9 | 42.5 | 0 | 5 | | | | |
| Meta-analysis | 141,245 | 346,029 | 107.6 | 319.7 | <33 | 204 | 0.90 (0.55 - 1.46) | 0.67 | 0.89 (0.53 - 1.51) | 0.67 |
| **PS-stratification / intent-to-treat TAR** | | | | | | | | | | |
| OptumDOD | 11,516 | 21,588 | 17.6 | 54.6 | 9 | 24 | 2.09 (0.85 - 4.72) | 0.09 | 2.13 (0.90 - 5.00) | 0.08 |
| MDCR | 3,464 | 9,818 | 5.8 | 21.2 | 9 | 38 | 1.34 (0.58 - 2.82) | 0.47 | 1.34 (0.61 - 2.96) | 0.47 |
| Open Claims | 181,939 | 337,293 | 451.1 | 1431.3 | 97 | 487 | 1.02 (0.80 - 1.28) | 0.90 | 1.03 (0.78 - 1.37) | 0.81 |
| OptumEHR | 7,758 | 19,847 | 16.0 | 57.3 | 7 | 62 | 0.64 (0.26 - 1.37) | 0.30 | 0.64 (0.28 - 1.46) | 0.29 |
| VA | 7,386 | 14,331 | 8.0 | 32.2 | <10 | <10 | 3.69 (0.48 - 19.1) | 0.17 | 3.89 (0.61 - 24.6) | 0.15 |
| GermanyDA | 4,167 | 10,750 | 8.7 | 37.2 | 0 | 11 | | | | |
| SIDIAP | 3,737 | 20,476 | 6.5 | 58.6 | 0 | 10 | | | | |
| Meta-analysis | 212,063 | 402,877 | 498.5 | 1596.6 | <132 | <621 | 1.13 (0.82 - 1.55) | 0.46 | 1.15 (0.81 - 1.63) | 0.45 |
| **PS-matching / intent-to-treat TAR** | | | | | | | | | | |
| OptumDOD | 9,215 | 21,198 | 16.2 | 53.1 | 8 | 23 | 1.44 (0.49 - 3.73) | 0.48 | 1.44 (0.52 - 3.98) | 0.48 |
| MDCR | 2,887 | 9,793 | 5.5 | 21.2 | 9 | 38 | 1.57 (0.63 - 3.56) | 0.31 | 1.60 (0.67 - 3.83) | 0.30 |
| Open Claims | 138,358 | 336,236 | 411.2 | 1426.3 | 94 | 484 | 1.01 (0.78 - 1.29) | 0.96 | 1.02 (0.76 - 1.37) | 0.91 |
| OptumEHR | 6,908 | 19,770 | 15.2 | 56.9 | 7 | 62 | 0.66 (0.25 - 1.48) | 0.36 | 0.65 (0.26 - 1.59) | 0.34 |
| VA | 4,518 | 13,831 | 6.0 | 29.8 | <10 | <10 | 4.91 (0.51 - 37.2) | 0.15 | 5.13 (0.60 - 43.7) | 0.13 |
| GermanyDA | 3,424 | 10,691 | 8.1 | 36.9 | 0 | 11 | | | | |
| SIDIAP | 3,110 | 19,842 | 5.9 | 55.7 | 0 | 9 | | | | |
| Meta-analysis | 157,368 | 386,997 | 448.1 | 1557.5 | 118 | 607 | 1.03 (0.82 - 1.29) | 0.81 | 1.04 (0.80 - 1.36) | 0.78 |

##### 6.4.17 Venous thromboembolism

**Supplementary Table 178:** Exposures, outcomes and risk estimates for Venous thromboembolism in SGLT2I (T) vs DPP4I (C) new-user cohorts under four different propensity score (PS) adjustment / time-at-risk (TAR) designs across data sources. We report total patient exposure time in 1,000 patient-year (KYr) units, uncalibrated and calibrated hazard ratios (HRs) and their 95% confidence intervals (CIs). Greyed out entries did not pass study diagnostics and are included only for completeness.

| | Patients | | Exposure (KYr) | | Outcomes | | Uncalibrated | | Calibrated | |
|---|---|---|---|---|---|---|---|---|---|---|
| | T | C | T | C | T | C | HR (95% CI) | $p$ | HR (95% CI) | $p$ |
| **PS-stratification / on-treatment TAR** | | | | | | | | | | |
| OptumDOD | 11,177 | 20,859 | 6.7 | 16.6 | 67 | 236 | 0.78 (0.57 - 1.05) | 0.10 | 0.77 (0.57 - 1.04) | 0.09 |
| MDCR | 3,367 | 9,538 | 2.5 | 8.6 | 19 | 101 | 0.84 (0.48 - 1.41) | 0.53 | 0.87 (0.50 - 1.50) | 0.62 |
| Open Claims | 176,274 | 326,954 | 122.7 | 301.8 | 661 | 2,211 | 0.92 (0.84 - 1.02) | 0.11 | 0.94 (0.75 - 1.18) | 0.61 |
| OptumEHR | 7,536 | 19,333 | 2.1 | 10.1 | 17 | 101 | 1.03 (0.58 - 1.72) | 0.93 | 0.93 (0.54 - 1.60) | 0.79 |
| VA | 7,154 | 14,015 | 4.5 | 14.1 | 21 | 80 | 0.55 (0.30 - 0.97) | 0.05 | 0.57 (0.31 - 1.03) | 0.06 |
| GermanyDA | 3,859 | 9,869 | 3.0 | 10.6 | 27 | 109 | 1.16 (0.71 - 1.83) | 0.55 | 1.07 (0.67 - 1.73) | 0.75 |
| SIDIAP | 3,597 | 19,712 | 4.3 | 42.8 | 19 | 235 | 0.87 (0.50 - 1.43) | 0.61 | 0.90 (0.53 - 1.52) | 0.71 |
| Meta-analysis | 212,964 | 420,280 | 145.6 | 404.6 | 831 | 3,073 | 0.91 (0.83 - 0.99) | 0.02 | 0.91 (0.75 - 1.10) | 0.34 |
| **PS-matching / on-treatment TAR** | | | | | | | | | | |
| OptumDOD | 8,952 | 20,503 | 5.8 | 16.3 | 58 | 228 | 0.79 (0.53 - 1.15) | 0.23 | 0.76 (0.52 - 1.11) | 0.16 |
| MDCR | 2,809 | 9,513 | 2.2 | 8.5 | 18 | 101 | 1.37 (0.68 - 2.59) | 0.36 | 1.39 (0.71 - 2.70) | 0.35 |
| Open Claims | 134,282 | 325,993 | 102.3 | 301.0 | 559 | 2,187 | 0.87 (0.76 - 0.99) | 0.04 | 0.87 (0.69 - 1.10) | 0.28 |
| OptumEHR | 6,729 | 19,270 | 1.9 | 10.1 | 17 | 100 | 1.21 (0.61 - 2.29) | 0.56 | 1.14 (0.59 - 2.20) | 0.71 |
| VA | 4,363 | 13,526 | 3.2 | 13.3 | 10 | 76 | 0.22 (0.07 - 0.57) | <0.01 | 0.24 (0.08 - 0.71) | 0.01 |
| GermanyDA | 3,149 | 9,826 | 2.6 | 10.6 | 23 | 109 | 1.57 (0.86 - 2.79) | 0.13 | 1.49 (0.82 - 2.69) | 0.20 |
| SIDIAP | 2,991 | 19,131 | 3.8 | 40.9 | 18 | 217 | 0.98 (0.53 - 1.68) | 0.94 | 1.02 (0.57 - 1.82) | 0.86 |
| Meta-analysis | 163,275 | 417,762 | 121.8 | 400.7 | 703 | 3,018 | 0.93 (0.77 - 1.11) | 0.41 | 0.92 (0.69 - 1.22) | 0.55 |
| **PS-stratification / intent-to-treat TAR** | | | | | | | | | | |
| OptumDOD | 11,177 | 20,859 | 16.9 | 51.9 | 209 | 748 | 0.94 (0.80 - 1.12) | 0.51 | 0.96 (0.81 - 1.14) | 0.65 |
| MDCR | 3,367 | 9,538 | 5.6 | 20.4 | 61 | 294 | 0.92 (0.68 - 1.23) | 0.58 | 0.92 (0.68 - 1.24) | 0.62 |
| Open Claims | 176,274 | 326,954 | 435.3 | 1372.0 | 2,914 | 11,288 | 1.01 (0.96 - 1.05) | 0.78 | 1.03 (0.87 - 1.21) | 0.77 |
| OptumEHR | 7,536 | 19,333 | 15.4 | 55.2 | 104 | 432 | 1.05 (0.83 - 1.31) | 0.69 | 1.04 (0.82 - 1.31) | 0.77 |
| VA | 7,154 | 14,015 | 7.7 | 31.5 | 50 | 205 | 0.78 (0.52 - 1.15) | 0.21 | 0.82 (0.55 - 1.22) | 0.34 |
| GermanyDA | 3,859 | 9,869 | 8.0 | 33.5 | 63 | 348 | 0.90 (0.67 - 1.21) | 0.50 | 0.88 (0.65 - 1.18) | 0.40 |
| SIDIAP | 3,597 | 19,712 | 6.2 | 56.1 | 30 | 312 | 0.93 (0.60 - 1.38) | 0.72 | 0.95 (0.62 - 1.44) | 0.80 |
| Meta-analysis | 212,964 | 420,280 | 495.1 | 1620.5 | 3,431 | 13,627 | 1.00 (0.96 - 1.04) | 0.87 | 1.01 (0.87 - 1.18) | 0.86 |
| **PS-matching / intent-to-treat TAR** | | | | | | | | | | |
| OptumDOD | 8,952 | 20,503 | 15.6 | 50.7 | 189 | 721 | 0.91 (0.75 - 1.11) | 0.36 | 0.91 (0.75 - 1.11) | 0.38 |
| MDCR | 2,809 | 9,513 | 5.3 | 20.3 | 59 | 293 | 1.05 (0.73 - 1.48) | 0.78 | 1.07 (0.75 - 1.54) | 0.73 |
| Open Claims | 134,282 | 325,993 | 396.9 | 1367.6 | 2,708 | 11,189 | 1.00 (0.95 - 1.05) | 0.96 | 1.01 (0.86 - 1.19) | 0.87 |
| OptumEHR | 6,729 | 19,270 | 14.7 | 54.9 | 103 | 428 | 1.05 (0.81 - 1.34) | 0.71 | 1.03 (0.80 - 1.32) | 0.82 |
| VA | 4,363 | 13,526 | 5.8 | 29.1 | 33 | 188 | 0.78 (0.47 - 1.23) | 0.30 | 0.81 (0.50 - 1.31) | 0.40 |
| GermanyDA | 3,149 | 9,826 | 7.3 | 33.2 | 59 | 345 | 0.99 (0.71 - 1.35) | 0.94 | 0.96 (0.69 - 1.33) | 0.77 |
| SIDIAP | 2,991 | 19,131 | 5.6 | 53.5 | 29 | 287 | 0.97 (0.60 - 1.51) | 0.90 | 1.00 (0.62 - 1.59) | 0.87 |
| Meta-analysis | 163,275 | 417,762 | 451.2 | 1609.3 | 3,180 | 13,451 | 1.00 (0.95 - 1.04) | 0.86 | 1.01 (0.87 - 1.17) | 0.90 |

##### 6.4.18 Vomiting

**Supplementary Table 179:** Exposures, outcomes and risk estimates for Vomiting in SGLT2I (T) vs DPP4I (C) new-user cohorts under four different propensity score (PS) adjustment / time-at-risk (TAR) designs across data sources. We report total patient exposure time in 1,000 patient-year (KYr) units, uncalibrated and calibrated hazard ratios (HRs) and their 95% confidence intervals (CIs). Greyed out entries did not pass study diagnostics and are included only for completeness.

| | Patients | | Exposure (KYr) | | Outcomes | | Uncalibrated | | Calibrated | |
|---|---|---|---|---|---|---|---|---|---|---|
| | T | C | T | C | T | C | HR (95% CI) | $p$ | HR (95% CI) | $p$ |
| **PS-stratification / on-treatment TAR** | | | | | | | | | | |
| OptumDOD | 10,476 | 19,270 | 6.2 | 15.1 | 155 | 535 | 0.86 (0.71 - 1.05) | 0.14 | 0.85 (0.70 - 1.04) | 0.12 |
| MDCR | 3,271 | 9,092 | 2.4 | 8.1 | 48 | 232 | 0.82 (0.57 - 1.14) | 0.25 | 0.84 (0.60 - 1.19) | 0.35 |
| Open Claims | 165,476 | 306,254 | 115.6 | 282.0 | 1,324 | 4,849 | 0.86 (0.80 - 0.92) | <0.01 | 0.88 (0.71 - 1.09) | 0.25 |
| OptumEHR | 7,398 | 18,868 | 2.1 | 9.8 | 40 | 223 | 1.15 (0.79 - 1.63) | 0.47 | 1.04 (0.72 - 1.49) | 0.82 |
| VA | 7,106 | 13,967 | 4.5 | 14.1 | 25 | 88 | 0.78 (0.43 - 1.36) | 0.40 | 0.81 (0.46 - 1.44) | 0.48 |
| GermanyDA | 3,907 | 9,993 | 3.0 | 10.8 | 20 | 114 | 0.77 (0.45 - 1.26) | 0.32 | 0.71 (0.42 - 1.20) | 0.21 |
| SIDIAP | 3,583 | 19,596 | 4.3 | 42.5 | 19 | 389 | 0.54 (0.32 - 0.86) | 0.01 | 0.56 (0.34 - 0.91) | 0.02 |
| Meta-analysis | 201,217 | 397,040 | 138.1 | 382.3 | 1,631 | 6,430 | 0.86 (0.81 - 0.91) | <0.01 | 0.86 (0.72 - 1.03) | 0.11 |
| **PS-matching / on-treatment TAR** | | | | | | | | | | |
| OptumDOD | 8,391 | 18,971 | 5.5 | 14.9 | 142 | 521 | 0.91 (0.70 - 1.17) | 0.45 | 0.87 (0.67 - 1.12) | 0.29 |
| MDCR | 2,712 | 9,071 | 2.2 | 8.0 | 43 | 232 | 0.91 (0.60 - 1.37) | 0.67 | 0.93 (0.61 - 1.41) | 0.75 |
| Open Claims | 125,626 | 305,455 | 96.1 | 281.3 | 1,130 | 4,824 | 0.83 (0.76 - 0.91) | <0.01 | 0.84 (0.67 - 1.04) | 0.12 |
| OptumEHR | 6,581 | 18,727 | 1.9 | 9.8 | 38 | 221 | 1.37 (0.87 - 2.11) | 0.17 | 1.28 (0.82 - 2.00) | 0.30 |
| VA | 4,384 | 13,577 | 3.3 | 13.5 | 15 | 83 | 0.57 (0.22 - 1.25) | 0.19 | 0.61 (0.26 - 1.43) | 0.26 |
| GermanyDA | 3,205 | 9,953 | 2.6 | 10.7 | 17 | 114 | 0.66 (0.35 - 1.19) | 0.19 | 0.63 (0.34 - 1.17) | 0.16 |
| SIDIAP | 2,983 | 19,027 | 3.8 | 40.6 | 14 | 368 | 0.46 (0.24 - 0.82) | 0.01 | 0.48 (0.26 - 0.90) | 0.02 |
| Meta-analysis | 153,882 | 394,781 | 115.3 | 378.8 | 1,399 | 6,363 | 0.84 (0.78 - 0.91) | <0.01 | 0.84 (0.66 - 1.05) | 0.12 |
| **PS-stratification / intent-to-treat TAR** | | | | | | | | | | |
| OptumDOD | 10,476 | 19,270 | 15.6 | 46.5 | 462 | 1,610 | 0.99 (0.89 - 1.11) | 0.91 | 1.01 (0.90 - 1.13) | 0.85 |
| MDCR | 3,271 | 9,092 | 5.4 | 19.0 | 124 | 596 | 0.88 (0.71 - 1.08) | 0.22 | 0.88 (0.71 - 1.09) | 0.31 |
| Open Claims | 165,476 | 306,254 | 403.7 | 1254.0 | 5,532 | 23,515 | 0.92 (0.89 - 0.95) | <0.01 | 0.94 (0.80 - 1.11) | 0.46 |
| OptumEHR | 7,398 | 18,868 | 14.9 | 52.8 | 186 | 914 | 0.89 (0.75 - 1.05) | 0.18 | 0.88 (0.74 - 1.05) | 0.16 |
| VA | 7,106 | 13,967 | 7.7 | 31.3 | 54 | 216 | 0.93 (0.64 - 1.34) | 0.71 | 0.98 (0.68 - 1.43) | 0.90 |
| GermanyDA | 3,907 | 9,993 | 8.1 | 33.9 | 66 | 346 | 1.07 (0.80 - 1.42) | 0.63 | 1.04 (0.78 - 1.39) | 0.76 |
| SIDIAP | 3,583 | 19,596 | 6.2 | 55.7 | 37 | 547 | 0.71 (0.49 - 1.00) | 0.06 | 0.73 (0.51 - 1.04) | 0.10 |
| Meta-analysis | 201,217 | 397,040 | 461.7 | 1493.1 | 6,461 | 27,744 | 0.92 (0.90 - 0.95) | <0.01 | 0.94 (0.81 - 1.09) | 0.41 |
| **PS-matching / intent-to-treat TAR** | | | | | | | | | | |
| OptumDOD | 8,391 | 18,971 | 14.4 | 45.5 | 435 | 1,563 | 0.99 (0.87 - 1.13) | 0.89 | 0.99 (0.87 - 1.13) | 0.86 |
| MDCR | 2,712 | 9,071 | 5.0 | 18.9 | 116 | 594 | 0.88 (0.69 - 1.12) | 0.30 | 0.90 (0.70 - 1.16) | 0.52 |
| Open Claims | 125,626 | 305,455 | 367.2 | 1250.5 | 5,160 | 23,377 | 0.92 (0.89 - 0.96) | <0.01 | 0.94 (0.80 - 1.10) | 0.43 |
| OptumEHR | 6,581 | 18,727 | 14.2 | 52.2 | 173 | 900 | 0.88 (0.73 - 1.06) | 0.19 | 0.87 (0.72 - 1.05) | 0.16 |
| VA | 4,384 | 13,577 | 5.9 | 29.4 | 38 | 200 | 0.69 (0.42 - 1.10) | 0.13 | 0.72 (0.45 - 1.17) | 0.20 |
| GermanyDA | 3,205 | 9,953 | 7.5 | 33.7 | 61 | 341 | 1.10 (0.80 - 1.48) | 0.56 | 1.07 (0.78 - 1.46) | 0.69 |
| SIDIAP | 2,983 | 19,027 | 5.7 | 53.1 | 32 | 511 | 0.68 (0.44 - 1.00) | 0.06 | 0.70 (0.46 - 1.05) | 0.10 |
| Meta-analysis | 153,882 | 394,781 | 419.8 | 1483.4 | 6,015 | 27,486 | 0.93 (0.90 - 0.95) | <0.01 | 0.93 (0.81 - 1.08) | 0.35 |

#### 6.5 SGLT2I vs SU

##### 6.5.1 Abnormal weight gain

**Supplementary Table 180:** Exposures, outcomes and risk estimates for Abnormal weight gain in SGLT2I (T) vs SU (C) new-user cohorts under four different propensity score (PS) adjustment / time-at-risk (TAR) designs across data sources. We report total patient exposure time in 1,000 patient-year (KYr) units, uncalibrated and calibrated hazard ratios (HRs) and their 95% confidence intervals (CIs). Greyed out entries did not pass study diagnostics and are included only for completeness.

| | Patients | | Exposure (KYr) | | Outcomes | | Uncalibrated | | Calibrated | |
|---|---|---|---|---|---|---|---|---|---|---|
| | T | C | T | C | T | C | HR (95% CI) | $p$ | HR (95% CI) | $p$ |
| **PS-stratification / on-treatment TAR** | | | | | | | | | | |
| OptumDOD | 11,275 | 48,470 | 6.7 | 48.1 | 20 | 309 | 0.45 (0.27 - 0.72) | <0.01 | 0.45 (0.28 - 0.72) | <0.01 |
| MDCR | 3,444 | 15,316 | 2.5 | 13.5 | <5 | 61 | 0.35 (0.10 - 0.92) | 0.06 | 0.34 (0.11 - 1.03) | 0.06 |
| Open Claims | 177,824 | 682,286 | 123.7 | 783.1 | 144 | 1,635 | 0.58 (0.48 - 0.69) | <0.01 | 0.59 (0.45 - 0.77) | <0.01 |
| OptumEHR | 7,560 | 45,158 | 2.1 | 27.6 | 8 | 170 | 0.73 (0.31 - 1.46) | 0.41 | 0.71 (0.33 - 1.53) | 0.38 |
| VA | 7,350 | 52,957 | 4.6 | 52.4 | <10 | 83 | 0.78 (0.29 - 1.84) | 0.59 | 0.84 (0.33 - 2.12) | 0.71 |
| GermanyDA | 4,116 | 1,537 | 3.2 | 1.6 | 0 | <5 | | | | |
| SIDIAP | 3,707 | 2,113 | 4.4 | 4.7 | <5 | 9 | 0.46 (0.08 - 2.27) | 0.37 | 0.51 (0.10 - 2.74) | 0.43 |
| Meta-analysis | 196,659 | 775,914 | 132.6 | 858.8 | 172 | 2,114 | 0.57 (0.48 - 0.67) | <0.01 | 0.57 (0.45 - 0.71) | <0.01 |
| **PS-matching / on-treatment TAR** | | | | | | | | | | |
| OptumDOD | 10,483 | 47,455 | 6.4 | 47.1 | 20 | 294 | 0.42 (0.22 - 0.76) | <0.01 | 0.41 (0.22 - 0.78) | <0.01 |
| MDCR | 2,983 | 14,556 | 2.3 | 12.8 | <5 | 56 | 0.22 (0.03 - 0.81) | 0.06 | 0.21 (0.04 - 1.02) | 0.05 |
| Open Claims | 163,428 | 665,817 | 116.5 | 765.0 | 128 | 1,567 | 0.59 (0.47 - 0.73) | <0.01 | 0.60 (0.44 - 0.81) | <0.01 |
| OptumEHR | 7,048 | 42,481 | 2.0 | 25.8 | 5 | 151 | 0.56 (0.16 - 1.50) | 0.31 | 0.55 (0.18 - 1.69) | 0.31 |
| VA | 5,185 | 33,189 | 3.7 | 29.0 | <10 | 45 | 0.69 (0.16 - 2.17) | 0.58 | 0.74 (0.20 - 2.78) | 0.66 |
| GermanyDA | 4,057 | 883 | 3.2 | 0.8 | 0 | <5 | | | | |
| SIDIAP | 3,280 | 958 | 4.1 | 1.7 | <5 | <5 | 0.53 (0.10 - 3.07) | 0.47 | 0.67 (0.12 - 3.80) | 0.65 |
| Meta-analysis | 183,942 | 770,309 | 127.2 | 850.8 | <158 | 2,068 | 0.56 (0.45 - 0.68) | <0.01 | 0.56 (0.42 - 0.75) | <0.01 |
| **PS-stratification / intent-to-treat TAR** | | | | | | | | | | |
| OptumDOD | 11,275 | 48,470 | 17.1 | 123.0 | 78 | 663 | 0.85 (0.66 - 1.09) | 0.21 | 0.85 (0.66 - 1.10) | 0.23 |
| MDCR | 3,444 | 15,316 | 5.8 | 31.0 | 18 | 149 | 0.75 (0.43 - 1.26) | 0.30 | 0.75 (0.43 - 1.32) | 0.34 |
| Open Claims | 177,824 | 682,286 | 438.9 | 2771.6 | 750 | 5,583 | 0.93 (0.85 - 1.01) | 0.08 | 0.96 (0.77 - 1.19) | 0.69 |
| OptumEHR | 7,560 | 45,158 | 15.5 | 134.5 | 61 | 534 | 1.07 (0.79 - 1.42) | 0.67 | 1.12 (0.83 - 1.50) | 0.48 |
| VA | 7,350 | 52,957 | 8.0 | 170.2 | 12 | 233 | 0.89 (0.43 - 1.70) | 0.75 | 0.94 (0.48 - 1.87) | 0.87 |
| GermanyDA | 4,116 | 1,537 | 8.7 | 6.4 | <5 | 5 | 0.24 (0.01 - 2.19) | 0.29 | 0.22 (0.02 - 3.08) | 0.26 |
| SIDIAP | 3,707 | 2,113 | 6.4 | 8.9 | <5 | 15 | 0.34 (0.07 - 1.38) | 0.16 | 0.35 (0.08 - 1.63) | 0.18 |
| Meta-analysis | 196,659 | 775,914 | 471.4 | 3029.2 | 889 | 6,780 | 0.93 (0.86 - 1.00) | 0.06 | 0.96 (0.79 - 1.17) | 0.67 |
| **PS-matching / intent-to-treat TAR** | | | | | | | | | | |
| OptumDOD | 10,483 | 47,455 | 16.6 | 119.4 | 77 | 630 | 0.84 (0.62 - 1.11) | 0.23 | 0.83 (0.62 - 1.12) | 0.25 |
| MDCR | 2,983 | 14,556 | 5.4 | 29.5 | 18 | 136 | 0.61 (0.30 - 1.13) | 0.14 | 0.60 (0.31 - 1.17) | 0.15 |
| Open Claims | 163,428 | 665,817 | 425.2 | 2677.2 | 723 | 5,290 | 0.91 (0.83 - 1.00) | 0.05 | 0.94 (0.75 - 1.17) | 0.56 |
| OptumEHR | 7,048 | 42,481 | 14.9 | 122.1 | 57 | 466 | 0.98 (0.69 - 1.35) | 0.89 | 1.02 (0.73 - 1.42) | 0.91 |
| VA | 5,185 | 33,189 | 6.7 | 78.0 | <10 | 104 | 1.03 (0.44 - 2.14) | 0.93 | 1.10 (0.50 - 2.42) | 0.82 |
| GermanyDA | 4,057 | 883 | 8.6 | 3.0 | <5 | <5 | 0.71 (0.03 - 18.4) | 0.83 | 0.63 (0.02 - 16.4) | 0.78 |
| SIDIAP | 3,280 | 958 | 6.0 | 2.8 | <5 | 5 | 0.35 (0.07 - 1.53) | 0.18 | 0.35 (0.08 - 1.63) | 0.19 |
| Meta-analysis | 183,942 | 770,309 | 462.1 | 2948.2 | 875 | 6,522 | 0.91 (0.83 - 0.98) | 0.02 | 0.92 (0.75 - 1.14) | 0.46 |

##### 6.5.2 Abnormal weight loss

**Supplementary Table 181:** Exposures, outcomes and risk estimates for Abnormal weight loss in SGLT2I (T) vs SU (C) new-user cohorts under four different propensity score (PS) adjustment / time-at-risk (TAR) designs across data sources. We report total patient exposure time in 1,000 patient-year (KYr) units, uncalibrated and calibrated hazard ratios (HRs) and their 95% confidence intervals (CIs). Greyed out entries did not pass study diagnostics and are included only for completeness.

| | Patients | | Exposure (KYr) | | Outcomes | | Uncalibrated | | Calibrated | |
|---|---|---|---|---|---|---|---|---|---|---|
| | T | C | T | C | T | C | HR (95% CI) | $p$ | HR (95% CI) | $p$ |
| **PS-stratification / on-treatment TAR** | | | | | | | | | | |
| OptumDOD | 11,094 | 46,524 | 6.6 | 46.3 | 163 | 803 | 1.51 (1.25 - 1.82) | <0.01 | 1.48 (1.22 - 1.80) | <0.01 |
| MDCR | 3,392 | 14,708 | 2.5 | 13.0 | 44 | 196 | 1.62 (1.09 - 2.37) | 0.01 | 1.58 (1.07 - 2.35) | 0.04 |
| Open Claims | 176,400 | 665,366 | 122.5 | 764.5 | 1,091 | 5,468 | 1.40 (1.30 - 1.51) | <0.01 | 1.43 (1.15 - 1.76) | <0.01 |
| OptumEHR | 7,530 | 44,122 | 2.1 | 26.8 | 30 | 410 | 1.17 (0.76 - 1.72) | 0.46 | 1.14 (0.76 - 1.71) | 0.54 |
| VA | 7,184 | 51,321 | 4.5 | 50.9 | 71 | 419 | 1.78 (1.26 - 2.49) | <0.01 | 1.92 (1.37 - 2.70) | <0.01 |
| GermanyDA | 4,032 | 1,500 | 3.1 | 1.6 | 37 | 9 | 1.89 (0.64 - 6.13) | 0.27 | 1.79 (0.58 - 5.54) | 0.33 |
| SIDIAP | 3,673 | 2,113 | 4.4 | 4.6 | 35 | 22 | 1.33 (0.61 - 2.91) | 0.48 | 1.46 (0.67 - 3.19) | 0.36 |
| Meta-analysis | 195,024 | 756,012 | 131.2 | 837.6 | 1,284 | 6,681 | 1.41 (1.32 - 1.51) | <0.01 | 1.41 (1.18 - 1.67) | <0.01 |
| **PS-matching / on-treatment TAR** | | | | | | | | | | |
| OptumDOD | 10,284 | 45,740 | 6.3 | 45.6 | 152 | 777 | 1.46 (1.16 - 1.84) | <0.01 | 1.43 (1.13 - 1.81) | <0.01 |
| MDCR | 2,932 | 13,950 | 2.2 | 12.3 | 42 | 187 | 1.74 (1.08 - 2.72) | 0.02 | 1.65 (1.04 - 2.62) | 0.04 |
| Open Claims | 162,116 | 649,510 | 115.4 | 747.1 | 1,023 | 5,238 | 1.40 (1.27 - 1.53) | <0.01 | 1.43 (1.15 - 1.78) | <0.01 |
| OptumEHR | 7,005 | 41,529 | 1.9 | 25.1 | 29 | 363 | 0.96 (0.55 - 1.61) | 0.89 | 0.95 (0.55 - 1.64) | 0.85 |
| VA | 5,071 | 32,451 | 3.6 | 28.5 | 54 | 226 | 1.48 (0.94 - 2.29) | 0.08 | 1.60 (1.03 - 2.50) | 0.04 |
| GermanyDA | 3,974 | 869 | 3.1 | 0.8 | 37 | <5 | 3.85 (0.68 - 72.8) | 0.26 | 4.58 (0.44 - 47.3) | 0.20 |
| SIDIAP | 3,257 | 957 | 4.1 | 1.7 | 29 | 7 | 1.12 (0.38 - 3.76) | 0.84 | 1.42 (0.45 - 4.47) | 0.57 |
| Meta-analysis | 182,337 | 750,729 | 125.8 | 830.1 | 1,246 | 6,565 | 1.40 (1.29 - 1.52) | <0.01 | 1.41 (1.13 - 1.76) | <0.01 |
| **PS-stratification / intent-to-treat TAR** | | | | | | | | | | |
| OptumDOD | 11,094 | 46,524 | 16.5 | 116.0 | 362 | 2,354 | 1.20 (1.06 - 1.35) | <0.01 | 1.20 (1.06 - 1.36) | <0.01 |
| MDCR | 3,392 | 14,708 | 5.6 | 29.4 | 111 | 533 | 1.53 (1.20 - 1.92) | <0.01 | 1.52 (1.16 - 2.01) | <0.01 |
| Open Claims | 176,400 | 665,366 | 433.7 | 2667.0 | 3,566 | 25,180 | 1.05 (1.01 - 1.09) | 0.02 | 1.08 (0.87 - 1.33) | 0.50 |
| OptumEHR | 7,530 | 44,122 | 15.3 | 129.5 | 201 | 1,675 | 1.25 (1.06 - 1.47) | <0.01 | 1.31 (1.11 - 1.54) | <0.01 |
| VA | 7,184 | 51,321 | 7.7 | 163.2 | 114 | 1,880 | 1.23 (0.96 - 1.55) | 0.10 | 1.29 (1.01 - 1.64) | 0.05 |
| GermanyDA | 4,032 | 1,500 | 8.4 | 6.2 | 66 | 47 | 1.05 (0.63 - 1.75) | 0.85 | 0.95 (0.57 - 1.59) | 0.80 |
| SIDIAP | 3,673 | 2,113 | 6.3 | 8.8 | 55 | 45 | 1.52 (0.86 - 2.69) | 0.15 | 1.60 (0.90 - 2.82) | 0.12 |
| Meta-analysis | 195,024 | 756,012 | 465.5 | 2912.5 | 4,129 | 29,209 | 1.14 (1.02 - 1.28) | 0.03 | 1.17 (0.95 - 1.45) | 0.14 |
| **PS-matching / intent-to-treat TAR** | | | | | | | | | | |
| OptumDOD | 10,284 | 45,740 | 16.1 | 113.3 | 351 | 2,271 | 1.19 (1.04 - 1.36) | 0.01 | 1.19 (1.03 - 1.37) | 0.04 |
| MDCR | 2,932 | 13,950 | 5.2 | 27.9 | 107 | 496 | 1.28 (0.96 - 1.68) | 0.08 | 1.27 (0.96 - 1.68) | 0.13 |
| Open Claims | 162,116 | 649,510 | 420.0 | 2577.9 | 3,460 | 23,782 | 1.07 (1.03 - 1.12) | <0.01 | 1.10 (0.90 - 1.35) | 0.38 |
| OptumEHR | 7,005 | 41,529 | 14.7 | 117.8 | 197 | 1,449 | 1.17 (0.97 - 1.40) | 0.09 | 1.22 (1.01 - 1.47) | 0.05 |
| VA | 5,071 | 32,451 | 6.5 | 76.1 | 93 | 822 | 1.12 (0.83 - 1.47) | 0.45 | 1.18 (0.89 - 1.57) | 0.27 |
| GermanyDA | 3,974 | 869 | 8.4 | 2.9 | 66 | 22 | 1.08 (0.61 - 2.00) | 0.80 | 0.97 (0.53 - 1.76) | 0.80 |
| SIDIAP | 3,257 | 957 | 6.0 | 2.9 | 48 | 14 | 1.60 (0.77 - 3.58) | 0.23 | 1.61 (0.74 - 3.53) | 0.29 |
| Meta-analysis | 182,337 | 750,729 | 456.0 | 2836.9 | 4,115 | 27,998 | 1.13 (1.04 - 1.22) | <0.01 | 1.15 (0.93 - 1.42) | 0.19 |

##### 6.5.3 Acute pancreatitis

**Supplementary Table 182:** Exposures, outcomes and risk estimates for Acute pancreatitis in SGLT2I (T) vs SU (C) new-user cohorts under four different propensity score (PS) adjustment / time-at-risk (TAR) designs across data sources. We report total patient exposure time in 1,000 patient-year (KYr) units, uncalibrated and calibrated hazard ratios (HRs) and their 95% confidence intervals (CIs). Greyed out entries did not pass study diagnostics and are included only for completeness.

| | Patients | | Exposure (KYr) | | Outcomes | | Uncalibrated | | Calibrated | |
|---|---|---|---|---|---|---|---|---|---|---|
| | T | C | T | C | T | C | HR (95% CI) | $p$ | HR (95% CI) | $p$ |
| **PS-stratification / on-treatment TAR** | | | | | | | | | | |
| OptumDOD | 11,512 | 49,009 | 6.9 | 48.9 | 21 | 179 | 0.98 (0.59 - 1.55) | 0.93 | 0.96 (0.59 - 1.56) | 0.87 |
| MDCR | 3,480 | 15,305 | 2.5 | 13.5 | <5 | 48 | 0.56 (0.16 - 1.48) | 0.30 | 0.54 (0.18 - 1.66) | 0.29 |
| Open Claims | 182,041 | 689,073 | 126.7 | 790.8 | 176 | 1,739 | 0.75 (0.63 - 0.88) | <0.01 | 0.76 (0.58 - 0.98) | 0.04 |
| OptumEHR | 7,801 | 45,830 | 2.2 | 28.1 | <5 | 78 | 0.67 (0.16 - 1.93) | 0.53 | 0.66 (0.19 - 2.29) | 0.51 |
| VA | 7,438 | 53,201 | 4.7 | 52.7 | <10 | 92 | 0.25 (0.01 - 1.27) | 0.23 | 0.27 (0.03 - 2.60) | 0.26 |
| GermanyDA | 4,154 | 1,547 | 3.2 | 1.6 | 0 | 0 | | | | |
| SIDIAP | 3,736 | 2,128 | 4.5 | 4.7 | 0 | 0 | | | | |
| Meta-analysis | 201,354 | 783,912 | 135.8 | 867.8 | <202 | 1,996 | 0.77 (0.66 - 0.90) | <0.01 | 0.76 (0.61 - 0.96) | 0.02 |
| **PS-matching / on-treatment TAR** | | | | | | | | | | |
| OptumDOD | 10,688 | 47,974 | 6.6 | 47.9 | 20 | 176 | 0.99 (0.55 - 1.69) | 0.97 | 0.97 (0.55 - 1.70) | 0.90 |
| MDCR | 3,011 | 14,560 | 2.3 | 12.9 | <5 | 47 | 0.68 (0.19 - 1.96) | 0.52 | 0.65 (0.20 - 2.09) | 0.47 |
| Open Claims | 167,109 | 672,969 | 119.2 | 773.1 | 171 | 1,673 | 0.78 (0.64 - 0.95) | 0.02 | 0.80 (0.60 - 1.06) | 0.13 |
| OptumEHR | 7,259 | 43,132 | 2.0 | 26.3 | <5 | 74 | 0.93 (0.13 - 3.93) | 0.93 | 0.92 (0.17 - 4.98) | 0.92 |
| VA | 5,219 | 33,500 | 3.8 | 29.3 | <10 | 44 | 0.31 (0.02 - 1.63) | 0.31 | 0.33 (0.03 - 3.29) | 0.35 |
| GermanyDA | 4,095 | 892 | 3.2 | 0.8 | 0 | 0 | | | | |
| SIDIAP | 3,307 | 967 | 4.1 | 1.7 | 0 | 0 | | | | |
| Meta-analysis | 188,067 | 778,635 | 130.1 | 860.1 | <201 | 1,970 | 0.80 (0.66 - 0.96) | 0.02 | 0.81 (0.61 - 1.06) | 0.13 |
| **PS-stratification / intent-to-treat TAR** | | | | | | | | | | |
| OptumDOD | 11,512 | 49,009 | 17.6 | 124.8 | 54 | 468 | 1.00 (0.74 - 1.34) | 0.99 | 1.00 (0.74 - 1.36) | 0.95 |
| MDCR | 3,480 | 15,305 | 5.9 | 31.1 | 10 | 119 | 0.55 (0.26 - 1.03) | 0.09 | 0.55 (0.27 - 1.11) | 0.10 |
| Open Claims | 182,041 | 689,073 | 451.3 | 2798.0 | 675 | 6,370 | 0.79 (0.73 - 0.86) | <0.01 | 0.81 (0.65 - 1.02) | 0.07 |
| OptumEHR | 7,801 | 45,830 | 16.1 | 137.1 | 20 | 257 | 0.82 (0.49 - 1.30) | 0.43 | 0.86 (0.53 - 1.40) | 0.56 |
| VA | 7,438 | 53,201 | 8.0 | 170.9 | <10 | 289 | 0.62 (0.18 - 1.60) | 0.39 | 0.65 (0.22 - 1.95) | 0.44 |
| GermanyDA | 4,154 | 1,547 | 8.7 | 6.5 | 0 | 0 | | | | |
| SIDIAP | 3,736 | 2,128 | 6.5 | 9.0 | 0 | 0 | | | | |
| Meta-analysis | 201,354 | 783,912 | 485.0 | 3059.9 | 749 | 7,095 | 0.83 (0.71 - 0.97) | 0.02 | 0.86 (0.68 - 1.09) | 0.21 |
| **PS-matching / intent-to-treat TAR** | | | | | | | | | | |
| OptumDOD | 10,688 | 47,974 | 17.1 | 121.1 | 53 | 449 | 1.14 (0.82 - 1.57) | 0.43 | 1.14 (0.82 - 1.58) | 0.46 |
| MDCR | 3,011 | 14,560 | 5.5 | 29.5 | 10 | 113 | 0.62 (0.28 - 1.24) | 0.21 | 0.62 (0.29 - 1.31) | 0.22 |
| Open Claims | 167,109 | 672,969 | 437.0 | 2705.7 | 661 | 6,043 | 0.82 (0.75 - 0.90) | <0.01 | 0.84 (0.67 - 1.05) | 0.13 |
| OptumEHR | 7,259 | 43,132 | 15.5 | 124.5 | 19 | 234 | 0.78 (0.45 - 1.30) | 0.37 | 0.81 (0.48 - 1.39) | 0.46 |
| VA | 5,219 | 33,500 | 6.7 | 78.9 | <10 | 123 | 0.74 (0.21 - 1.90) | 0.58 | 0.78 (0.26 - 2.32) | 0.66 |
| GermanyDA | 4,095 | 892 | 8.7 | 3.0 | 0 | 0 | | | | |
| SIDIAP | 3,307 | 967 | 6.1 | 2.9 | 0 | 0 | | | | |
| Meta-analysis | 188,067 | 778,635 | 475.1 | 2980.8 | 743 | 6,839 | 0.87 (0.71 - 1.06) | 0.16 | 0.88 (0.67 - 1.17) | 0.39 |

##### 6.5.4 All-cause mortality

**Supplementary Table 183:** Exposures, outcomes and risk estimates for All-cause mortality in SGLT2I (T) vs SU (C) new-user cohorts under four different propensity score (PS) adjustment / time-at-risk (TAR) designs across data sources. We report total patient exposure time in 1,000 patient-year (KYr) units, uncalibrated and calibrated hazard ratios (HRs) and their 95% confidence intervals (CIs). Greyed out entries did not pass study diagnostics and are included only for completeness.

| | Patients | | Exposure (KYr) | | Outcomes | | Uncalibrated | | Calibrated | |
|---|---|---|---|---|---|---|---|---|---|---|
| | T | C | T | C | T | C | HR (95% CI) | $p$ | HR (95% CI) | $p$ |
| **PS-stratification / on-treatment TAR** | | | | | | | | | | |
| OptumDOD | 11,614 | 49,630 | 6.9 | 49.5 | 43 | 785 | 0.51 (0.36 - 0.70) | <0.01 | 0.50 (0.36 - 0.70) | <0.01 |
| MDCR | 3,510 | 15,524 | 2.6 | 13.7 | <5 | 32 | 1.54 (0.24 - 5.55) | 0.59 | 1.50 (0.31 - 7.24) | 0.61 |
| Open Claims | 184,807 | 699,596 | 128.6 | 803.5 | 0 | 13 | | | | |
| OptumEHR | 7,828 | 46,129 | 2.2 | 28.3 | 8 | 517 | 0.40 (0.18 - 0.77) | 0.01 | 0.39 (0.19 - 0.81) | 0.01 |
| VA | 7,529 | 53,862 | 4.7 | 53.4 | 65 | 1,015 | 0.60 (0.44 - 0.80) | <0.01 | 0.65 (0.48 - 0.88) | <0.01 |
| GermanyDA | 4,154 | 1,547 | 3.2 | 1.6 | <5 | 0 | | | | |
| SIDIAP | 3,746 | 2,136 | 4.5 | 4.7 | 95 | 96 | 0.79 (0.51 - 1.21) | 0.28 | 0.87 (0.56 - 1.33) | 0.54 |
| Meta-analysis | 19,442 | 95,759 | 9.1 | 77.9 | 51 | 1,302 | 0.49 (0.36 - 0.66) | <0.01 | 0.49 (0.35 - 0.68) | <0.01 |
| **PS-matching / on-treatment TAR** | | | | | | | | | | |
| OptumDOD | 10,783 | 48,541 | 6.6 | 48.5 | 43 | 737 | 0.60 (0.40 - 0.87) | <0.01 | 0.59 (0.40 - 0.86) | <0.01 |
| MDCR | 3,040 | 14,747 | 2.3 | 13.0 | <5 | 25 | 1.00 (0.14 - 4.21) | 1.00 | 0.95 (0.18 - 5.16) | 0.95 |
| Open Claims | 169,729 | 683,066 | 121.1 | 785.4 | 0 | 13 | | | | |
| OptumEHR | 7,282 | 43,385 | 2.0 | 26.4 | 7 | 451 | 0.49 (0.20 - 1.02) | 0.08 | 0.48 (0.22 - 1.09) | 0.09 |
| VA | 5,286 | 33,867 | 3.8 | 29.6 | 54 | 576 | 0.55 (0.37 - 0.80) | <0.01 | 0.60 (0.40 - 0.88) | 0.01 |
| GermanyDA | 4,095 | 892 | 3.2 | 0.8 | <5 | 0 | | | | |
| SIDIAP | 3,314 | 971 | 4.1 | 1.7 | 75 | 36 | 0.65 (0.36 - 1.17) | 0.15 | 0.82 (0.45 - 1.50) | 0.54 |
| Meta-analysis | 21,105 | 106,673 | 11.0 | 88.0 | <55 | 1,213 | 0.59 (0.42 - 0.83) | <0.01 | 0.60 (0.40 - 0.89) | 0.01 |
| **PS-stratification / intent-to-treat TAR** | | | | | | | | | | |
| OptumDOD | 11,614 | 49,630 | 17.7 | 125.8 | 359 | 5,334 | 0.73 (0.65 - 0.82) | <0.01 | 0.73 (0.65 - 0.82) | <0.01 |
| MDCR | 3,510 | 15,524 | 5.9 | 31.6 | <5 | 93 | 0.59 (0.14 - 1.71) | 0.41 | 0.59 (0.17 - 2.09) | 0.42 |
| Open Claims | 184,807 | 699,596 | 459.1 | 2854.6 | <5 | 65 | 0.57 (0.09 - 1.91) | 0.47 | 0.59 (0.13 - 2.71) | 0.49 |
| OptumEHR | 7,828 | 46,129 | 16.1 | 137.8 | 119 | 2,411 | 0.70 (0.57 - 0.84) | <0.01 | 0.73 (0.60 - 0.89) | <0.01 |
| VA | 7,529 | 53,862 | 8.1 | 173.4 | 184 | 5,499 | 0.71 (0.60 - 0.84) | <0.01 | 0.75 (0.63 - 0.89) | <0.01 |
| GermanyDA | 4,154 | 1,547 | 8.7 | 6.5 | 11 | 17 | 0.90 (0.34 - 2.29) | 0.83 | 0.82 (0.31 - 2.14) | 0.69 |
| SIDIAP | 3,746 | 2,136 | 6.5 | 9.0 | 202 | 362 | 0.73 (0.57 - 0.93) | 0.01 | 0.77 (0.60 - 0.98) | 0.05 |
| Meta-analysis | 204,249 | 795,355 | 493.0 | 3118.2 | <483 | 7,810 | 0.72 (0.65 - 0.80) | <0.01 | 0.74 (0.61 - 0.91) | <0.01 |
| **PS-matching / intent-to-treat TAR** | | | | | | | | | | |
| OptumDOD | 10,783 | 48,541 | 17.2 | 122.1 | 348 | 4,880 | 0.72 (0.63 - 0.82) | <0.01 | 0.72 (0.63 - 0.82) | <0.01 |
| MDCR | 3,040 | 14,747 | 5.5 | 30.0 | <5 | 77 | 0.65 (0.15 - 1.86) | 0.50 | 0.65 (0.18 - 2.26) | 0.50 |
| Open Claims | 169,729 | 683,066 | 444.7 | 2758.8 | <5 | 57 | 0.58 (0.09 - 1.95) | 0.48 | 0.59 (0.13 - 2.76) | 0.51 |
| OptumEHR | 7,282 | 43,385 | 15.5 | 125.0 | 115 | 2,015 | 0.67 (0.54 - 0.83) | <0.01 | 0.70 (0.56 - 0.87) | <0.01 |
| VA | 5,286 | 33,867 | 6.8 | 79.8 | 154 | 2,376 | 0.65 (0.53 - 0.80) | <0.01 | 0.69 (0.56 - 0.85) | <0.01 |
| GermanyDA | 4,095 | 892 | 8.7 | 3.0 | 11 | 5 | 1.21 (0.33 - 4.86) | 0.78 | 1.08 (0.28 - 4.15) | 0.89 |
| SIDIAP | 3,314 | 971 | 6.1 | 2.9 | 175 | 115 | 0.59 (0.44 - 0.81) | <0.01 | 0.60 (0.43 - 0.84) | 0.02 |
| Meta-analysis | 190,834 | 789,739 | 482.9 | 3035.8 | <473 | 7,029 | 0.71 (0.63 - 0.79) | <0.01 | 0.72 (0.58 - 0.90) | <0.01 |

##### 6.5.5 Bone fracture

**Supplementary Table 184:** Exposures, outcomes and risk estimates for Bone fracture in SGLT2I (T) vs SU (C) new-user cohorts under four different propensity score (PS) adjustment / time-at-risk (TAR) designs across data sources. We report total patient exposure time in 1,000 patient-year (KYr) units, uncalibrated and calibrated hazard ratios (HRs) and their 95% confidence intervals (CIs). Greyed out entries did not pass study diagnostics and are included only for completeness.

| | Patients | | Exposure (KYr) | | Outcomes | | Uncalibrated | | Calibrated | |
|---|---|---|---|---|---|---|---|---|---|---|
| | T | C | T | C | T | C | HR (95% CI) | $p$ | HR (95% CI) | $p$ |
| **PS-stratification / on-treatment TAR** | | | | | | | | | | |
| OptumDOD | 10,423 | 44,290 | 6.1 | 43.3 | 200 | 1,484 | 1.04 (0.88 - 1.21) | 0.68 | 1.02 (0.86 - 1.20) | 0.84 |
| MDCR | 3,143 | 13,509 | 2.3 | 11.6 | 73 | 419 | 1.00 (0.75 - 1.32) | 0.98 | 0.98 (0.73 - 1.31) | 0.85 |
| Open Claims | 155,987 | 598,249 | 108.3 | 680.6 | 1,810 | 15,403 | 0.87 (0.83 - 0.92) | <0.01 | 0.89 (0.72 - 1.09) | 0.26 |
| OptumEHR | 7,175 | 42,034 | 2.0 | 25.1 | 39 | 681 | 1.06 (0.74 - 1.48) | 0.73 | 1.04 (0.73 - 1.47) | 0.81 |
| VA | 6,699 | 48,394 | 4.2 | 47.8 | 39 | 479 | 1.17 (0.76 - 1.75) | 0.46 | 1.26 (0.83 - 1.91) | 0.28 |
| GermanyDA | 3,608 | 1,340 | 2.8 | 1.4 | 50 | 40 | 1.36 (0.76 - 2.43) | 0.30 | 1.29 (0.72 - 2.32) | 0.43 |
| SIDIAP | 3,268 | 1,874 | 3.9 | 4.0 | 52 | 63 | 0.78 (0.46 - 1.34) | 0.37 | 0.86 (0.50 - 1.48) | 0.60 |
| Meta-analysis | 173,585 | 684,573 | 116.5 | 749.0 | 2,049 | 17,568 | 0.95 (0.82 - 1.09) | 0.46 | 0.94 (0.76 - 1.17) | 0.60 |
| **PS-matching / on-treatment TAR** | | | | | | | | | | |
| OptumDOD | 9,696 | 43,445 | 5.9 | 42.5 | 194 | 1,436 | 1.06 (0.87 - 1.29) | 0.54 | 1.04 (0.85 - 1.27) | 0.73 |
| MDCR | 2,745 | 12,892 | 2.1 | 11.1 | 71 | 394 | 1.12 (0.78 - 1.59) | 0.53 | 1.07 (0.74 - 1.53) | 0.74 |
| Open Claims | 143,566 | 583,748 | 102.1 | 664.6 | 1,710 | 14,822 | 0.88 (0.82 - 0.94) | <0.01 | 0.90 (0.73 - 1.11) | 0.33 |
| OptumEHR | 6,659 | 39,434 | 1.8 | 23.4 | 38 | 601 | 1.04 (0.69 - 1.53) | 0.86 | 1.03 (0.69 - 1.54) | 0.83 |
| VA | 4,736 | 30,474 | 3.4 | 26.6 | 27 | 247 | 1.17 (0.68 - 1.93) | 0.55 | 1.27 (0.75 - 2.14) | 0.38 |
| GermanyDA | 3,561 | 791 | 2.8 | 0.8 | 50 | 21 | 0.94 (0.43 - 2.11) | 0.87 | 1.12 (0.50 - 2.47) | 0.68 |
| SIDIAP | 2,892 | 861 | 3.6 | 1.5 | 46 | 14 | 1.09 (0.49 - 2.60) | 0.84 | 1.38 (0.60 - 3.19) | 0.50 |
| Meta-analysis | 162,666 | 679,519 | 111.9 | 741.7 | 2,013 | 17,253 | 0.97 (0.84 - 1.12) | 0.68 | 0.98 (0.76 - 1.25) | 0.86 |
| **PS-stratification / intent-to-treat TAR** | | | | | | | | | | |
| OptumDOD | 10,423 | 44,290 | 15.3 | 106.9 | 493 | 3,846 | 1.00 (0.90 - 1.11) | 1.00 | 1.00 (0.90 - 1.11) | 0.88 |
| MDCR | 3,143 | 13,509 | 5.1 | 26.2 | 158 | 965 | 1.04 (0.86 - 1.25) | 0.67 | 1.04 (0.82 - 1.33) | 0.78 |
| Open Claims | 155,987 | 598,249 | 377.8 | 2318.2 | 7,504 | 60,364 | 0.94 (0.91 - 0.96) | <0.01 | 0.97 (0.79 - 1.19) | 0.74 |
| OptumEHR | 7,175 | 42,034 | 14.4 | 120.5 | 281 | 2,887 | 1.06 (0.93 - 1.21) | 0.40 | 1.11 (0.97 - 1.27) | 0.17 |
| VA | 6,699 | 48,394 | 7.3 | 153.4 | 69 | 1,668 | 0.93 (0.69 - 1.24) | 0.62 | 0.98 (0.73 - 1.31) | 0.86 |
| GermanyDA | 3,608 | 1,340 | 7.4 | 5.2 | 137 | 129 | 0.99 (0.73 - 1.36) | 0.97 | 0.90 (0.66 - 1.24) | 0.57 |
| SIDIAP | 3,268 | 1,874 | 5.6 | 7.7 | 84 | 121 | 0.96 (0.65 - 1.39) | 0.82 | 1.01 (0.69 - 1.47) | 0.80 |
| Meta-analysis | 173,585 | 684,573 | 407.5 | 2545.7 | 8,278 | 67,097 | 0.98 (0.91 - 1.05) | 0.49 | 1.01 (0.83 - 1.22) | 0.91 |
| **PS-matching / intent-to-treat TAR** | | | | | | | | | | |
| OptumDOD | 9,696 | 43,445 | 14.9 | 104.1 | 481 | 3,689 | 0.98 (0.87 - 1.09) | 0.67 | 0.97 (0.86 - 1.10) | 0.71 |
| MDCR | 2,745 | 12,892 | 4.8 | 25.1 | 154 | 893 | 1.08 (0.87 - 1.34) | 0.49 | 1.07 (0.86 - 1.34) | 0.61 |
| Open Claims | 143,566 | 583,748 | 366.1 | 2242.3 | 7,312 | 57,198 | 0.94 (0.92 - 0.97) | <0.01 | 0.97 (0.79 - 1.18) | 0.74 |
| OptumEHR | 6,659 | 39,434 | 13.8 | 109.2 | 272 | 2,522 | 1.06 (0.91 - 1.23) | 0.43 | 1.10 (0.95 - 1.29) | 0.25 |
| VA | 4,736 | 30,474 | 6.1 | 71.2 | 49 | 730 | 0.95 (0.66 - 1.33) | 0.77 | 1.01 (0.71 - 1.42) | 0.90 |
| GermanyDA | 3,561 | 791 | 7.4 | 2.6 | 136 | 58 | 0.97 (0.66 - 1.43) | 0.87 | 0.86 (0.59 - 1.27) | 0.51 |
| SIDIAP | 2,892 | 861 | 5.3 | 2.5 | 76 | 29 | 1.24 (0.75 - 2.11) | 0.41 | 1.25 (0.73 - 2.15) | 0.51 |
| Meta-analysis | 162,666 | 679,519 | 399.6 | 2480.7 | 8,219 | 64,302 | 0.97 (0.92 - 1.03) | 0.37 | 0.99 (0.81 - 1.22) | 0.91 |

##### 6.5.6 Diabetic ketoacidosis

**Supplementary Table 185:** Exposures, outcomes and risk estimates for Diabetic ketoacidosis in SGLT2I (T) vs SU (C) new-user cohorts under four different propensity score (PS) adjustment / time-at-risk (TAR) designs across data sources. We report total patient exposure time in 1,000 patient-year (KYr) units, uncalibrated and calibrated hazard ratios (HRs) and their 95% confidence intervals (CIs). Greyed out entries did not pass study diagnostics and are included only for completeness.

| | Patients | | Exposure (KYr) | | Outcomes | | Uncalibrated | | Calibrated | |
|---|---|---|---|---|---|---|---|---|---|---|
| | T | C | T | C | T | C | HR (95% CI) | $p$ | HR (95% CI) | $p$ |
| **PS-stratification / on-treatment TAR** | | | | | | | | | | |
| OptumDOD | 11,606 | 49,554 | 6.9 | 49.5 | 18 | 84 | 1.75 (0.95 - 3.07) | 0.06 | 1.72 (0.96 - 3.09) | 0.07 |
| MDCR | 3,506 | 15,501 | 2.6 | 13.7 | <5 | 18 | 0.90 (0.12 - 4.09) | 0.90 | 0.87 (0.15 - 5.13) | 0.88 |
| Open Claims | 184,585 | 698,404 | 128.5 | 802.2 | 137 | 608 | 1.60 (1.30 - 1.97) | <0.01 | 1.63 (1.22 - 2.17) | <0.01 |
| OptumEHR | 7,821 | 46,097 | 2.2 | 28.3 | <5 | 37 | 2.80 (0.77 - 7.86) | 0.08 | 2.73 (0.86 - 8.71) | 0.09 |
| VA | 7,524 | 53,832 | 4.7 | 53.3 | <10 | 14 | 2.15 (0.40 - 10.5) | 0.36 | 2.32 (0.45 - 11.9) | 0.31 |
| GermanyDA | 4,154 | 1,547 | 3.2 | 1.6 | 0 | 0 | | | | |
| SIDIAP | 3,746 | 2,136 | 4.5 | 4.7 | 0 | <5 | | | | |
| Meta-analysis | 204,012 | 794,055 | 137.6 | 880.1 | <160 | 729 | 1.64 (1.35 - 1.99) | <0.01 | 1.64 (1.27 - 2.11) | <0.01 |
| **PS-matching / on-treatment TAR** | | | | | | | | | | |
| OptumDOD | 10,774 | 48,470 | 6.6 | 48.4 | 16 | 81 | 2.47 (1.13 - 5.17) | 0.02 | 2.42 (1.13 - 5.17) | 0.02 |
| MDCR | 3,036 | 14,734 | 2.3 | 13.0 | <5 | 17 | 1.09 (0.06 - 6.98) | 0.94 | 1.04 (0.09 - 11.7) | 0.97 |
| Open Claims | 169,532 | 681,857 | 120.9 | 784.0 | 129 | 584 | 1.77 (1.37 - 2.27) | <0.01 | 1.81 (1.31 - 2.50) | <0.01 |
| OptumEHR | 7,275 | 43,358 | 2.0 | 26.5 | <5 | 34 | 2.83 (0.53 - 11.9) | 0.19 | 2.80 (0.59 - 13.3) | 0.20 |
| VA | 5,284 | 33,852 | 3.8 | 29.6 | <10 | <10 | 2.56 (0.42 - 13.2) | 0.29 | 2.77 (0.49 - 15.6) | 0.25 |
| GermanyDA | 4,095 | 892 | 3.2 | 0.8 | 0 | 0 | | | | |
| SIDIAP | 3,314 | 971 | 4.1 | 1.7 | 0 | <5 | | | | |
| Meta-analysis | 187,581 | 773,685 | 129.6 | 858.9 | <150 | 699 | 1.85 (1.46 - 2.34) | <0.01 | 1.86 (1.36 - 2.55) | <0.01 |
| **PS-stratification / intent-to-treat TAR** | | | | | | | | | | |
| OptumDOD | 11,606 | 49,554 | 17.7 | 126.6 | 44 | 310 | 1.25 (0.88 - 1.74) | 0.20 | 1.25 (0.89 - 1.76) | 0.21 |
| MDCR | 3,506 | 15,501 | 5.9 | 31.5 | 11 | 63 | 1.49 (0.70 - 2.92) | 0.28 | 1.49 (0.71 - 3.09) | 0.30 |
| Open Claims | 184,585 | 698,404 | 457.8 | 2843.6 | 494 | 3,609 | 1.06 (0.96 - 1.17) | 0.26 | 1.09 (0.87 - 1.37) | 0.46 |
| OptumEHR | 7,821 | 46,097 | 16.1 | 138.3 | 11 | 144 | 1.06 (0.52 - 1.94) | 0.86 | 1.11 (0.58 - 2.14) | 0.76 |
| VA | 7,524 | 53,832 | 8.1 | 173.2 | <10 | 113 | 0.71 (0.21 - 1.93) | 0.54 | 0.75 (0.25 - 2.24) | 0.60 |
| GermanyDA | 4,154 | 1,547 | 8.7 | 6.5 | 0 | 0 | | | | |
| SIDIAP | 3,746 | 2,136 | 6.5 | 9.0 | 0 | <5 | | | | |
| Meta-analysis | 204,012 | 794,055 | 491.6 | 3108.5 | 549 | 4,063 | 1.07 (0.98 - 1.18) | 0.14 | 1.11 (0.90 - 1.36) | 0.32 |
| **PS-matching / intent-to-treat TAR** | | | | | | | | | | |
| OptumDOD | 10,774 | 48,470 | 17.2 | 122.7 | 42 | 297 | 1.45 (0.98 - 2.08) | 0.05 | 1.44 (0.99 - 2.10) | 0.06 |
| MDCR | 3,036 | 14,734 | 5.5 | 29.9 | 11 | 62 | 1.41 (0.61 - 3.03) | 0.40 | 1.40 (0.63 - 3.12) | 0.42 |
| Open Claims | 169,532 | 681,857 | 443.4 | 2747.9 | 485 | 3,431 | 1.08 (0.96 - 1.20) | 0.19 | 1.10 (0.88 - 1.39) | 0.42 |
| OptumEHR | 7,275 | 43,358 | 15.5 | 125.4 | 11 | 129 | 1.18 (0.55 - 2.27) | 0.65 | 1.23 (0.60 - 2.48) | 0.57 |
| VA | 5,284 | 33,852 | 6.8 | 79.7 | <10 | 52 | 0.75 (0.17 - 2.30) | 0.67 | 0.79 (0.22 - 2.93) | 0.73 |
| GermanyDA | 4,095 | 892 | 8.7 | 3.0 | 0 | 0 | | | | |
| SIDIAP | 3,314 | 971 | 6.1 | 2.9 | 0 | <5 | | | | |
| Meta-analysis | 190,617 | 788,419 | 481.7 | 3025.9 | 549 | 3,919 | 1.17 (0.96 - 1.42) | 0.12 | 1.19 (0.90 - 1.57) | 0.21 |

##### 6.5.7 Diarrhea

**Supplementary Table 186:** Exposures, outcomes and risk estimates for Diarrhea in SGLT2I (T) vs SU (C) new-user cohorts under four different propensity score (PS) adjustment / time-at-risk (TAR) designs across data sources. We report total patient exposure time in 1,000 patient-year (KYr) units, uncalibrated and calibrated hazard ratios (HRs) and their 95% confidence intervals (CIs). Greyed out entries did not pass study diagnostics and are included only for completeness.

| | Patients | | Exposure (KYr) | | Outcomes | | Uncalibrated | | Calibrated | |
|---|---|---|---|---|---|---|---|---|---|---|
| | T | C | T | C | T | C | HR (95% CI) | $p$ | HR (95% CI) | $p$ |
| **PS-stratification / on-treatment TAR** | | | | | | | | | | |
| OptumDOD | 9,875 | 41,297 | 5.8 | 40.0 | 228 | 1,763 | 0.90 (0.77 - 1.04) | 0.16 | 0.88 (0.76 - 1.03) | 0.15 |
| MDCR | 3,032 | 13,000 | 2.2 | 11.2 | 57 | 421 | 0.69 (0.50 - 0.94) | 0.02 | 0.68 (0.49 - 0.93) | 0.03 |
| Open Claims | 158,033 | 602,791 | 109.5 | 682.4 | 1,724 | 14,147 | 0.84 (0.79 - 0.88) | <0.01 | 0.85 (0.69 - 1.05) | 0.13 |
| OptumEHR | 6,910 | 40,186 | 1.9 | 23.3 | 70 | 980 | 1.02 (0.77 - 1.32) | 0.89 | 1.00 (0.76 - 1.30) | 0.81 |
| VA | 6,417 | 46,833 | 4.0 | 45.6 | 65 | 854 | 0.81 (0.59 - 1.10) | 0.18 | 0.87 (0.64 - 1.19) | 0.41 |
| GermanyDA | 4,090 | 1,526 | 3.2 | 1.6 | 6 | <5 | 9.12 (1.98 - 41.7) | <0.01 | 8.65 (1.88 - 39.7) | <0.01 |
| SIDIAP | 3,550 | 1,997 | 4.2 | 4.3 | 34 | 49 | 0.41 (0.20 - 0.82) | 0.01 | 0.45 (0.22 - 0.91) | 0.03 |
| Meta-analysis | 174,818 | 684,274 | 117.2 | 745.7 | 2,022 | 16,890 | 0.86 (0.80 - 0.93) | <0.01 | 0.86 (0.72 - 1.03) | 0.10 |
| **PS-matching / on-treatment TAR** | | | | | | | | | | |
| OptumDOD | 9,161 | 40,356 | 5.5 | 39.1 | 219 | 1,711 | 0.85 (0.70 - 1.01) | 0.07 | 0.83 (0.69 - 0.99) | 0.06 |
| MDCR | 2,638 | 12,081 | 2.0 | 10.4 | 50 | 385 | 0.73 (0.48 - 1.08) | 0.13 | 0.70 (0.46 - 1.05) | 0.10 |
| Open Claims | 145,007 | 588,132 | 103.0 | 666.7 | 1,609 | 13,673 | 0.81 (0.76 - 0.86) | <0.01 | 0.83 (0.67 - 1.02) | 0.09 |
| OptumEHR | 6,404 | 37,784 | 1.8 | 21.8 | 63 | 874 | 1.06 (0.76 - 1.45) | 0.74 | 1.05 (0.75 - 1.45) | 0.78 |
| VA | 4,546 | 29,232 | 3.3 | 25.1 | 47 | 476 | 0.81 (0.54 - 1.17) | 0.28 | 0.87 (0.59 - 1.29) | 0.51 |
| GermanyDA | 4,031 | 878 | 3.2 | 0.8 | 6 | 0 | | | | |
| SIDIAP | 3,144 | 906 | 3.9 | 1.6 | 30 | 16 | 0.42 (0.14 - 1.26) | 0.12 | 0.54 (0.18 - 1.60) | 0.28 |
| Meta-analysis | 163,210 | 678,353 | 112.2 | 738.0 | 1,941 | 16,643 | 0.82 (0.77 - 0.87) | <0.01 | 0.82 (0.67 - 1.02) | 0.08 |
| **PS-stratification / intent-to-treat TAR** | | | | | | | | | | |
| OptumDOD | 9,875 | 41,297 | 14.4 | 98.1 | 576 | 4,359 | 0.92 (0.83 - 1.00) | 0.06 | 0.92 (0.83 - 1.01) | 0.12 |
| MDCR | 3,032 | 13,000 | 4.8 | 25.0 | 171 | 1,055 | 0.90 (0.75 - 1.08) | 0.27 | 0.90 (0.71 - 1.14) | 0.48 |
| Open Claims | 158,033 | 602,791 | 379.4 | 2333.1 | 6,741 | 51,016 | 0.92 (0.90 - 0.95) | <0.01 | 0.95 (0.77 - 1.17) | 0.64 |
| OptumEHR | 6,910 | 40,186 | 13.6 | 112.8 | 341 | 3,340 | 1.02 (0.90 - 1.16) | 0.70 | 1.07 (0.95 - 1.22) | 0.31 |
| VA | 6,417 | 46,833 | 6.9 | 146.1 | 125 | 2,816 | 0.84 (0.68 - 1.04) | 0.12 | 0.89 (0.71 - 1.10) | 0.31 |
| GermanyDA | 4,090 | 1,526 | 8.6 | 6.4 | 11 | 13 | 1.98 (0.70 - 5.34) | 0.19 | 1.80 (0.65 - 4.96) | 0.27 |
| SIDIAP | 3,550 | 1,997 | 6.1 | 8.2 | 66 | 117 | 0.51 (0.33 - 0.76) | <0.01 | 0.53 (0.35 - 0.81) | <0.01 |
| Meta-analysis | 174,818 | 684,274 | 407.4 | 2544.0 | 7,658 | 58,715 | 0.93 (0.90 - 0.95) | <0.01 | 0.96 (0.80 - 1.15) | 0.63 |
| **PS-matching / intent-to-treat TAR** | | | | | | | | | | |
| OptumDOD | 9,161 | 40,356 | 13.9 | 94.8 | 557 | 4,204 | 0.92 (0.83 - 1.02) | 0.12 | 0.92 (0.82 - 1.03) | 0.23 |
| MDCR | 2,638 | 12,081 | 4.6 | 23.2 | 159 | 962 | 0.85 (0.68 - 1.05) | 0.14 | 0.84 (0.67 - 1.06) | 0.20 |
| Open Claims | 145,007 | 588,132 | 367.2 | 2255.9 | 6,542 | 48,609 | 0.93 (0.91 - 0.96) | <0.01 | 0.96 (0.78 - 1.17) | 0.68 |
| OptumEHR | 6,404 | 37,784 | 13.1 | 102.6 | 324 | 2,923 | 0.97 (0.84 - 1.11) | 0.65 | 1.01 (0.88 - 1.16) | 0.83 |
| VA | 4,546 | 29,232 | 5.8 | 67.4 | 100 | 1,294 | 0.87 (0.67 - 1.11) | 0.28 | 0.92 (0.72 - 1.19) | 0.55 |
| GermanyDA | 4,031 | 878 | 8.5 | 3.0 | 11 | <5 | | 0.10 | | 0.13 |
| SIDIAP | 3,144 | 906 | 5.8 | 2.7 | 60 | 46 | 0.45 (0.26 - 0.76) | <0.01 | 0.46 (0.26 - 0.79) | 0.01 |
| Meta-analysis | 163,210 | 678,353 | 398.8 | 2476.5 | 7,582 | 56,698 | 0.93 (0.91 - 0.96) | <0.01 | 0.95 (0.78 - 1.16) | 0.63 |

##### 6.5.8 Genitourinary infection

**Supplementary Table 187:** Exposures, outcomes and risk estimates for Genitourinary infection in SGLT2I (T) vs SU (C) new-user cohorts under four different propensity score (PS) adjustment / time-at-risk (TAR) designs across data sources. We report total patient exposure time in 1,000 patient-year (KYr) units, uncalibrated and calibrated hazard ratios (HRs) and their 95% confidence intervals (CIs). Greyed out entries did not pass study diagnostics and are included only for completeness.

| | Patients | | Exposure (KYr) | | Outcomes | | Uncalibrated | | Calibrated | |
|---|---|---|---|---|---|---|---|---|---|---|
| | T | C | T | C | T | C | HR (95% CI) | $p$ | HR (95% CI) | $p$ |
| **PS-stratification / on-treatment TAR** | | | | | | | | | | |
| OptumDOD | 9,018 | 35,987 | 5.3 | 34.3 | 329 | 2,620 | 0.90 (0.79 - 1.02) | 0.09 | 0.88 (0.78 - 1.00) | 0.10 |
| MDCR | 2,894 | 11,879 | 2.1 | 10.1 | 96 | 618 | 0.96 (0.75 - 1.22) | 0.73 | 0.93 (0.72 - 1.20) | 0.67 |
| Open Claims | 137,023 | 521,831 | 96.5 | 590.0 | 2,911 | 24,445 | 0.89 (0.85 - 0.92) | <0.01 | 0.90 (0.73 - 1.10) | 0.32 |
| OptumEHR | 6,565 | 37,673 | 1.8 | 21.7 | 86 | 1,404 | 0.98 (0.77 - 1.24) | 0.88 | 0.96 (0.76 - 1.22) | 0.72 |
| VA | 6,609 | 46,264 | 4.2 | 45.4 | 48 | 923 | 0.61 (0.43 - 0.87) | <0.01 | 0.66 (0.46 - 0.94) | 0.03 |
| GermanyDA | 3,188 | 1,172 | 2.4 | 1.2 | 90 | 44 | 1.12 (0.70 - 1.80) | 0.65 | 1.06 (0.66 - 1.71) | 0.66 |
| SIDIAP | 2,815 | 1,678 | 3.4 | 3.4 | 147 | 158 | 0.96 (0.70 - 1.32) | 0.80 | 1.05 (0.77 - 1.45) | 0.67 |
| Meta-analysis | 152,606 | 595,491 | 103.6 | 646.0 | 3,326 | 28,469 | 0.89 (0.85 - 0.92) | <0.01 | 0.89 (0.75 - 1.05) | 0.15 |
| **PS-matching / on-treatment TAR** | | | | | | | | | | |
| OptumDOD | 8,360 | 35,704 | 5.0 | 34.1 | 314 | 2,575 | 0.98 (0.84 - 1.14) | 0.82 | 0.96 (0.82 - 1.12) | 0.65 |
| MDCR | 2,515 | 11,338 | 1.9 | 9.7 | 90 | 573 | 1.02 (0.74 - 1.37) | 0.91 | 0.97 (0.71 - 1.32) | 0.81 |
| Open Claims | 126,202 | 509,338 | 91.1 | 576.7 | 2,735 | 23,474 | 0.91 (0.87 - 0.96) | <0.01 | 0.93 (0.76 - 1.15) | 0.53 |
| OptumEHR | 6,100 | 35,316 | 1.7 | 20.2 | 82 | 1,250 | 1.04 (0.78 - 1.35) | 0.80 | 1.03 (0.78 - 1.36) | 0.78 |
| VA | 4,658 | 29,538 | 3.4 | 25.8 | 32 | 489 | 0.68 (0.43 - 1.04) | 0.09 | 0.73 (0.47 - 1.14) | 0.18 |
| GermanyDA | 3,143 | 685 | 2.4 | 0.6 | 90 | 22 | 1.15 (0.62 - 2.27) | 0.67 | 1.37 (0.71 - 2.64) | 0.38 |
| SIDIAP | 2,499 | 758 | 3.2 | 1.3 | 136 | 62 | 0.96 (0.63 - 1.45) | 0.83 | 1.21 (0.79 - 1.85) | 0.47 |
| Meta-analysis | 143,177 | 591,696 | 99.6 | 640.7 | 3,221 | 27,872 | 0.93 (0.88 - 0.99) | 0.02 | 0.94 (0.76 - 1.16) | 0.56 |
| **PS-stratification / intent-to-treat TAR** | | | | | | | | | | |
| OptumDOD | 9,018 | 35,987 | 12.6 | 80.8 | 825 | 6,095 | 0.96 (0.89 - 1.04) | 0.31 | 0.96 (0.88 - 1.04) | 0.43 |
| MDCR | 2,894 | 11,879 | 4.5 | 22.0 | 236 | 1,436 | 1.04 (0.89 - 1.21) | 0.65 | 1.04 (0.84 - 1.29) | 0.79 |
| Open Claims | 137,023 | 521,831 | 320.7 | 1925.1 | 11,149 | 88,715 | 0.95 (0.93 - 0.97) | <0.01 | 0.97 (0.79 - 1.20) | 0.79 |
| OptumEHR | 6,565 | 37,673 | 12.6 | 102.9 | 421 | 4,573 | 0.97 (0.87 - 1.08) | 0.57 | 1.02 (0.91 - 1.14) | 0.77 |
| VA | 6,609 | 46,264 | 7.2 | 144.4 | 111 | 2,989 | 0.81 (0.64 - 1.02) | 0.08 | 0.85 (0.68 - 1.08) | 0.21 |
| GermanyDA | 3,188 | 1,172 | 6.3 | 4.4 | 221 | 159 | 1.30 (1.00 - 1.69) | 0.05 | 1.18 (0.90 - 1.54) | 0.31 |
| SIDIAP | 2,815 | 1,678 | 4.8 | 6.4 | 224 | 334 | 0.93 (0.74 - 1.18) | 0.57 | 0.98 (0.78 - 1.24) | 0.69 |
| Meta-analysis | 152,606 | 595,491 | 345.9 | 2108.8 | 12,395 | 99,383 | 0.95 (0.93 - 0.97) | <0.01 | 0.98 (0.82 - 1.17) | 0.80 |
| **PS-matching / intent-to-treat TAR** | | | | | | | | | | |
| OptumDOD | 8,360 | 35,704 | 12.2 | 80.0 | 804 | 5,993 | 0.96 (0.88 - 1.04) | 0.33 | 0.95 (0.87 - 1.05) | 0.47 |
| MDCR | 2,515 | 11,338 | 4.2 | 21.0 | 221 | 1,323 | 1.02 (0.85 - 1.22) | 0.82 | 1.01 (0.84 - 1.22) | 0.84 |
| Open Claims | 126,202 | 509,338 | 310.6 | 1865.3 | 10,846 | 84,101 | 0.95 (0.93 - 0.97) | <0.01 | 0.98 (0.80 - 1.20) | 0.81 |
| OptumEHR | 6,100 | 35,316 | 12.0 | 93.3 | 414 | 3,976 | 0.98 (0.86 - 1.10) | 0.69 | 1.01 (0.90 - 1.15) | 0.79 |
| VA | 4,658 | 29,538 | 6.0 | 68.8 | 87 | 1,325 | 0.88 (0.67 - 1.13) | 0.33 | 0.93 (0.72 - 1.21) | 0.62 |
| GermanyDA | 3,143 | 685 | 6.3 | 2.2 | 221 | 63 | 1.48 (1.06 - 2.10) | 0.03 | 1.32 (0.93 - 1.87) | 0.17 |
| SIDIAP | 2,499 | 758 | 4.5 | 2.1 | 211 | 116 | 0.93 (0.69 - 1.24) | 0.61 | 0.93 (0.67 - 1.29) | 0.68 |
| Meta-analysis | 143,177 | 591,696 | 339.1 | 2059.5 | 12,285 | 95,393 | 0.95 (0.93 - 0.98) | <0.01 | 0.97 (0.80 - 1.18) | 0.79 |

##### 6.5.9 Hyperkalemia

**Supplementary Table 188:** Exposures, outcomes and risk estimates for Hyperkalemia in SGLT2I (T) vs SU (C) new-user cohorts under four different propensity score (PS) adjustment / time-at-risk (TAR) designs across data sources. We report total patient exposure time in 1,000 patient-year (KYr) units, uncalibrated and calibrated hazard ratios (HRs) and their 95% confidence intervals (CIs). Greyed out entries did not pass study diagnostics and are included only for completeness.

| | Patients | | Exposure (KYr) | | Outcomes | | Uncalibrated | | Calibrated | |
|---|---|---|---|---|---|---|---|---|---|---|
| | T | C | T | C | T | C | HR (95% CI) | $p$ | HR (95% CI) | $p$ |
| **PS-stratification / on-treatment TAR** | | | | | | | | | | |
| OptumDOD | 10,938 | 45,992 | 6.5 | 44.7 | 162 | 1,526 | 0.81 (0.68 - 0.97) | 0.02 | 0.80 (0.67 - 0.95) | 0.02 |
| MDCR | 3,408 | 14,878 | 2.5 | 13.0 | 25 | 274 | 0.62 (0.38 - 0.96) | 0.04 | 0.60 (0.38 - 0.97) | 0.05 |
| Open Claims | 178,385 | 674,573 | 124.1 | 767.2 | 871 | 9,089 | 0.73 (0.68 - 0.79) | <0.01 | 0.74 (0.60 - 0.92) | <0.01 |
| OptumEHR | 7,603 | 44,391 | 2.1 | 26.6 | 31 | 599 | 0.86 (0.57 - 1.24) | 0.43 | 0.84 (0.57 - 1.23) | 0.38 |
| VA | 6,726 | 49,326 | 4.2 | 48.1 | 76 | 875 | 0.98 (0.72 - 1.31) | 0.89 | 1.06 (0.78 - 1.43) | 0.73 |
| GermanyDA | 3,801 | 1,459 | 3.0 | 1.5 | 45 | 18 | 0.72 (0.36 - 1.51) | 0.38 | 0.69 (0.33 - 1.41) | 0.34 |
| SIDIAP | 3,509 | 2,023 | 4.2 | 4.4 | 62 | 59 | 0.66 (0.39 - 1.11) | 0.12 | 0.72 (0.43 - 1.23) | 0.25 |
| Meta-analysis | 196,926 | 764,956 | 132.7 | 838.4 | 1,064 | 11,214 | 0.75 (0.69 - 0.82) | <0.01 | 0.75 (0.63 - 0.90) | <0.01 |
| **PS-matching / on-treatment TAR** | | | | | | | | | | |
| OptumDOD | 10,178 | 45,090 | 6.2 | 43.8 | 154 | 1,480 | 0.82 (0.67 - 1.01) | 0.07 | 0.80 (0.65 - 0.99) | 0.06 |
| MDCR | 2,961 | 14,148 | 2.3 | 12.4 | 23 | 251 | 0.62 (0.34 - 1.06) | 0.10 | 0.59 (0.34 - 1.05) | 0.08 |
| Open Claims | 164,246 | 659,135 | 117.1 | 750.4 | 773 | 8,676 | 0.71 (0.65 - 0.78) | <0.01 | 0.73 (0.58 - 0.91) | <0.01 |
| OptumEHR | 7,051 | 41,907 | 2.0 | 25.0 | 27 | 538 | 0.89 (0.55 - 1.37) | 0.60 | 0.88 (0.56 - 1.39) | 0.61 |
| VA | 4,822 | 31,325 | 3.4 | 27.2 | 59 | 429 | 1.01 (0.70 - 1.44) | 0.94 | 1.10 (0.76 - 1.58) | 0.64 |
| GermanyDA | 3,746 | 828 | 2.9 | 0.8 | 44 | 11 | 0.91 (0.35 - 2.67) | 0.85 | 1.08 (0.39 - 2.99) | 0.74 |
| SIDIAP | 3,133 | 916 | 3.9 | 1.6 | 49 | 21 | 0.57 (0.27 - 1.23) | 0.15 | 0.72 (0.34 - 1.56) | 0.43 |
| Meta-analysis | 184,436 | 760,280 | 127.5 | 831.6 | 977 | 10,945 | 0.74 (0.67 - 0.83) | <0.01 | 0.75 (0.59 - 0.94) | 0.01 |
| **PS-stratification / intent-to-treat TAR** | | | | | | | | | | |
| OptumDOD | 10,938 | 45,992 | 16.5 | 112.3 | 400 | 3,784 | 0.81 (0.72 - 0.90) | <0.01 | 0.81 (0.72 - 0.90) | <0.01 |
| MDCR | 3,408 | 14,878 | 5.7 | 29.6 | 71 | 654 | 0.80 (0.61 - 1.05) | 0.11 | 0.80 (0.59 - 1.10) | 0.22 |
| Open Claims | 178,385 | 674,573 | 440.7 | 2681.1 | 3,397 | 35,603 | 0.76 (0.73 - 0.78) | <0.01 | 0.78 (0.63 - 0.96) | 0.02 |
| OptumEHR | 7,603 | 44,391 | 15.4 | 128.7 | 176 | 2,264 | 0.90 (0.76 - 1.06) | 0.22 | 0.94 (0.80 - 1.12) | 0.53 |
| VA | 6,726 | 49,326 | 7.3 | 154.6 | 134 | 2,756 | 1.02 (0.82 - 1.26) | 0.85 | 1.08 (0.87 - 1.34) | 0.54 |
| GermanyDA | 3,801 | 1,459 | 7.8 | 6.0 | 120 | 88 | 1.05 (0.74 - 1.51) | 0.78 | 0.96 (0.67 - 1.37) | 0.73 |
| SIDIAP | 3,509 | 2,023 | 6.0 | 8.3 | 94 | 114 | 0.79 (0.53 - 1.17) | 0.24 | 0.83 (0.56 - 1.23) | 0.37 |
| Meta-analysis | 196,926 | 764,956 | 472.6 | 2922.1 | 3,973 | 41,651 | 0.80 (0.73 - 0.87) | <0.01 | 0.82 (0.67 - 1.00) | 0.06 |
| **PS-matching / intent-to-treat TAR** | | | | | | | | | | |
| OptumDOD | 10,178 | 45,090 | 16.0 | 109.3 | 388 | 3,634 | 0.80 (0.71 - 0.91) | <0.01 | 0.80 (0.70 - 0.91) | <0.01 |
| MDCR | 2,961 | 14,148 | 5.4 | 28.2 | 68 | 601 | 0.79 (0.57 - 1.07) | 0.14 | 0.78 (0.57 - 1.08) | 0.17 |
| Open Claims | 164,246 | 659,135 | 427.2 | 2596.2 | 3,224 | 33,450 | 0.75 (0.72 - 0.79) | <0.01 | 0.77 (0.63 - 0.95) | 0.02 |
| OptumEHR | 7,051 | 41,907 | 14.8 | 117.8 | 170 | 1,942 | 0.90 (0.75 - 1.08) | 0.28 | 0.94 (0.78 - 1.13) | 0.54 |
| VA | 4,822 | 31,325 | 6.2 | 72.9 | 104 | 1,203 | 1.03 (0.80 - 1.31) | 0.83 | 1.09 (0.85 - 1.40) | 0.53 |
| GermanyDA | 3,746 | 828 | 7.8 | 2.7 | 119 | 45 | 1.08 (0.72 - 1.65) | 0.72 | 0.97 (0.63 - 1.47) | 0.73 |
| SIDIAP | 3,133 | 916 | 5.7 | 2.7 | 79 | 35 | 0.82 (0.51 - 1.35) | 0.44 | 0.83 (0.50 - 1.38) | 0.54 |
| Meta-analysis | 184,436 | 760,280 | 463.4 | 2851.4 | 3,850 | 39,627 | 0.79 (0.73 - 0.85) | <0.01 | 0.80 (0.65 - 0.99) | 0.04 |

##### 6.5.10 Hypoglycemia

**Supplementary Table 189:** Exposures, outcomes and risk estimates for Hypoglycemia in SGLT2I (T) vs SU (C) new-user cohorts under four different propensity score (PS) adjustment / time-at-risk (TAR) designs across data sources. We report total patient exposure time in 1,000 patient-year (KYr) units, uncalibrated and calibrated hazard ratios (HRs) and their 95% confidence intervals (CIs). Greyed out entries did not pass study diagnostics and are included only for completeness.

| | Patients | | Exposure (KYr) | | Outcomes | | Uncalibrated | | Calibrated | |
|---|---|---|---|---|---|---|---|---|---|---|
| | T | C | T | C | T | C | HR (95% CI) | $p$ | HR (95% CI) | $p$ |
| **PS-stratification / on-treatment TAR** | | | | | | | | | | |
| OptumDOD | 11,439 | 48,868 | 6.8 | 48.3 | 48 | 1,387 | 0.30 (0.22 - 0.40) | <0.01 | 0.30 (0.22 - 0.40) | <0.01 |
| MDCR | 3,474 | 15,356 | 2.5 | 13.4 | <5 | 310 | 0.09 (0.03 - 0.22) | <0.01 | 0.09 (0.03 - 0.26) | <0.01 |
| Open Claims | 181,772 | 688,223 | 126.5 | 787.2 | 247 | 9,337 | 0.19 (0.17 - 0.22) | <0.01 | 0.19 (0.15 - 0.25) | <0.01 |
| OptumEHR | 7,748 | 45,662 | 2.2 | 27.7 | <5 | 605 | 0.09 (0.03 - 0.22) | <0.01 | 0.09 (0.03 - 0.25) | <0.01 |
| VA | 7,425 | 53,312 | 4.7 | 52.6 | <10 | 496 | 0.25 (0.12 - 0.50) | <0.01 | 0.27 (0.13 - 0.57) | <0.01 |
| GermanyDA | 4,132 | 1,538 | 3.2 | 1.6 | <5 | 11 | 0.34 (0.06 - 1.49) | 0.18 | 0.33 (0.07 - 1.57) | 0.17 |
| SIDIAP | 3,739 | 2,133 | 4.5 | 4.7 | 0 | 25 | | | | |
| Meta-analysis | 200,959 | 782,753 | 135.5 | 863.2 | <300 | 11,329 | 0.20 (0.13 - 0.33) | <0.01 | 0.20 (0.12 - 0.34) | <0.01 |
| **PS-matching / on-treatment TAR** | | | | | | | | | | |
| OptumDOD | 10,623 | 47,820 | 6.5 | 47.3 | 45 | 1,333 | 0.28 (0.20 - 0.39) | <0.01 | 0.27 (0.19 - 0.39) | <0.01 |
| MDCR | 3,002 | 14,595 | 2.3 | 12.8 | <5 | 271 | 0.10 (0.02 - 0.28) | <0.01 | 0.10 (0.03 - 0.32) | <0.01 |
| Open Claims | 167,032 | 671,919 | 119.2 | 769.3 | 231 | 8,909 | 0.18 (0.15 - 0.21) | <0.01 | 0.18 (0.14 - 0.23) | <0.01 |
| OptumEHR | 7,209 | 42,966 | 2.0 | 25.9 | <5 | 542 | 0.11 (0.03 - 0.27) | <0.01 | 0.11 (0.04 - 0.32) | <0.01 |
| VA | 5,220 | 33,278 | 3.7 | 28.9 | <10 | 264 | 0.24 (0.08 - 0.55) | <0.01 | 0.26 (0.10 - 0.67) | <0.01 |
| GermanyDA | 4,074 | 889 | 3.2 | 0.8 | <5 | <5 | 0.71 (0.03 - 18.4) | 0.83 | 0.84 (0.03 - 21.9) | 0.90 |
| SIDIAP | 3,311 | 970 | 4.1 | 1.7 | 0 | 12 | | | | |
| Meta-analysis | 187,866 | 777,300 | 130.0 | 855.3 | <286 | 11,055 | 0.19 (0.13 - 0.28) | <0.01 | 0.19 (0.12 - 0.30) | <0.01 |
| **PS-stratification / intent-to-treat TAR** | | | | | | | | | | |
| OptumDOD | 11,439 | 48,868 | 17.3 | 120.4 | 171 | 3,028 | 0.47 (0.40 - 0.55) | <0.01 | 0.47 (0.40 - 0.56) | <0.01 |
| MDCR | 3,474 | 15,356 | 5.8 | 30.4 | 26 | 637 | 0.29 (0.19 - 0.43) | <0.01 | 0.29 (0.19 - 0.45) | <0.01 |
| Open Claims | 181,772 | 688,223 | 448.6 | 2734.5 | 1,553 | 30,972 | 0.39 (0.37 - 0.41) | <0.01 | 0.40 (0.33 - 0.50) | <0.01 |
| OptumEHR | 7,748 | 45,662 | 15.9 | 133.5 | 82 | 1,801 | 0.47 (0.37 - 0.59) | <0.01 | 0.49 (0.39 - 0.62) | <0.01 |
| VA | 7,425 | 53,312 | 8.0 | 169.0 | 21 | 1,421 | 0.34 (0.21 - 0.54) | <0.01 | 0.36 (0.22 - 0.58) | <0.01 |
| GermanyDA | 4,132 | 1,538 | 8.7 | 6.3 | 8 | 27 | 0.34 (0.13 - 0.85) | 0.03 | 0.31 (0.12 - 0.81) | 0.02 |
| SIDIAP | 3,739 | 2,133 | 6.5 | 8.8 | 0 | 44 | | | | |
| Meta-analysis | 200,959 | 782,753 | 481.8 | 2988.3 | 1,806 | 35,801 | 0.43 (0.37 - 0.50) | <0.01 | 0.44 (0.35 - 0.56) | <0.01 |
| **PS-matching / intent-to-treat TAR** | | | | | | | | | | |
| OptumDOD | 10,623 | 47,820 | 16.8 | 116.8 | 167 | 2,881 | 0.46 (0.38 - 0.55) | <0.01 | 0.46 (0.38 - 0.55) | <0.01 |
| MDCR | 3,002 | 14,595 | 5.4 | 28.9 | 23 | 578 | 0.23 (0.14 - 0.38) | <0.01 | 0.23 (0.14 - 0.39) | <0.01 |
| Open Claims | 167,032 | 671,919 | 434.5 | 2644.7 | 1,518 | 29,148 | 0.39 (0.37 - 0.41) | <0.01 | 0.40 (0.32 - 0.49) | <0.01 |
| OptumEHR | 7,209 | 42,966 | 15.2 | 121.3 | 81 | 1,580 | 0.43 (0.33 - 0.56) | <0.01 | 0.45 (0.35 - 0.58) | <0.01 |
| VA | 5,220 | 33,278 | 6.7 | 77.2 | 16 | 610 | 0.28 (0.14 - 0.50) | <0.01 | 0.30 (0.16 - 0.56) | <0.01 |
| GermanyDA | 4,074 | 889 | 8.7 | 3.0 | 8 | 10 | 0.34 (0.08 - 1.16) | 0.11 | 0.30 (0.08 - 1.16) | 0.09 |
| SIDIAP | 3,311 | 970 | 6.1 | 2.9 | 0 | 14 | | | | |
| Meta-analysis | 187,866 | 777,300 | 472.0 | 2911.7 | 1,789 | 34,187 | 0.40 (0.36 - 0.46) | <0.01 | 0.41 (0.33 - 0.52) | <0.01 |

##### 6.5.11 Hypotension

**Supplementary Table 190:** Exposures, outcomes and risk estimates for Hypotension in SGLT2I (T) vs SU (C) new-user cohorts under four different propensity score (PS) adjustment / time-at-risk (TAR) designs across data sources. We report total patient exposure time in 1,000 patient-year (KYr) units, uncalibrated and calibrated hazard ratios (HRs) and their 95% confidence intervals (CIs). Greyed out entries did not pass study diagnostics and are included only for completeness.

| | Patients | | Exposure (KYr) | | Outcomes | | Uncalibrated | | Calibrated | |
|---|---|---|---|---|---|---|---|---|---|---|
| | T | C | T | C | T | C | HR (95% CI) | $p$ | HR (95% CI) | $p$ |
| **PS-stratification / on-treatment TAR** | | | | | | | | | | |
| OptumDOD | 10,826 | 45,890 | 6.4 | 45.2 | 223 | 1,420 | 1.17 (1.00 - 1.37) | 0.05 | 1.15 (0.98 - 1.35) | 0.12 |
| MDCR | 3,342 | 14,481 | 2.4 | 12.7 | 60 | 330 | 1.48 (1.06 - 2.04) | 0.02 | 1.44 (1.04 - 2.02) | 0.05 |
| Open Claims | 174,134 | 663,104 | 121.2 | 758.8 | 1,455 | 10,044 | 1.07 (1.01 - 1.14) | 0.02 | 1.09 (0.89 - 1.34) | 0.42 |
| OptumEHR | 7,517 | 44,224 | 2.1 | 26.6 | 46 | 724 | 1.02 (0.73 - 1.39) | 0.93 | 0.99 (0.72 - 1.37) | 0.84 |
| VA | 6,697 | 49,540 | 4.2 | 48.6 | 119 | 750 | 1.37 (1.05 - 1.77) | 0.02 | 1.48 (1.14 - 1.92) | <0.01 |
| GermanyDA | 4,007 | 1,503 | 3.1 | 1.6 | 20 | 8 | 1.23 (0.33 - 5.09) | 0.77 | 1.17 (0.30 - 4.58) | 0.81 |
| SIDIAP | 3,705 | 2,123 | 4.4 | 4.7 | 22 | 9 | 2.11 (0.67 - 7.03) | 0.21 | 2.32 (0.72 - 7.51) | 0.17 |
| Meta-analysis | 192,477 | 753,218 | 129.7 | 830.7 | 1,724 | 12,188 | 1.08 (1.03 - 1.15) | <0.01 | 1.08 (0.91 - 1.28) | 0.37 |
| **PS-matching / on-treatment TAR** | | | | | | | | | | |
| OptumDOD | 10,106 | 45,167 | 6.2 | 44.6 | 209 | 1,384 | 1.24 (1.02 - 1.50) | 0.03 | 1.22 (1.00 - 1.47) | 0.07 |
| MDCR | 2,894 | 13,836 | 2.2 | 12.2 | 53 | 300 | 1.39 (0.91 - 2.08) | 0.12 | 1.32 (0.87 - 2.01) | 0.21 |
| Open Claims | 160,749 | 647,650 | 114.5 | 741.9 | 1,304 | 9,568 | 1.07 (0.99 - 1.15) | 0.08 | 1.09 (0.88 - 1.35) | 0.44 |
| OptumEHR | 6,986 | 41,566 | 1.9 | 24.8 | 41 | 634 | 1.05 (0.70 - 1.54) | 0.80 | 1.04 (0.70 - 1.55) | 0.82 |
| VA | 4,826 | 31,081 | 3.4 | 27.0 | 87 | 414 | 1.41 (1.02 - 1.93) | 0.04 | 1.52 (1.10 - 2.10) | 0.01 |
| GermanyDA | 3,951 | 866 | 3.1 | 0.8 | 19 | <5 | 1.03 (0.22 - 7.54) | 0.97 | 1.23 (0.21 - 7.15) | 0.80 |
| SIDIAP | 3,283 | 963 | 4.1 | 1.7 | 21 | <5 | 2.72 (0.43 - 53.2) | 0.42 | 3.44 (0.31 - 38.5) | 0.32 |
| Meta-analysis | 180,735 | 748,219 | 124.8 | 823.5 | 1,607 | 11,886 | 1.13 (1.01 - 1.27) | 0.04 | 1.14 (0.90 - 1.44) | 0.28 |
| **PS-stratification / intent-to-treat TAR** | | | | | | | | | | |
| OptumDOD | 10,826 | 45,890 | 16.2 | 112.3 | 518 | 3,966 | 0.99 (0.90 - 1.09) | 0.86 | 0.99 (0.89 - 1.10) | 0.85 |
| MDCR | 3,342 | 14,481 | 5.4 | 28.5 | 141 | 911 | 1.17 (0.96 - 1.43) | 0.12 | 1.17 (0.91 - 1.51) | 0.29 |
| Open Claims | 174,134 | 663,104 | 428.5 | 2630.2 | 5,437 | 43,719 | 0.95 (0.93 - 0.98) | <0.01 | 0.98 (0.80 - 1.21) | 0.86 |
| OptumEHR | 7,517 | 44,224 | 15.1 | 128.4 | 230 | 2,398 | 1.10 (0.95 - 1.27) | 0.21 | 1.15 (0.99 - 1.34) | 0.09 |
| VA | 6,697 | 49,540 | 7.2 | 155.6 | 194 | 2,693 | 1.43 (1.18 - 1.72) | <0.01 | 1.50 (1.24 - 1.82) | <0.01 |
| GermanyDA | 4,007 | 1,503 | 8.4 | 6.2 | 38 | 33 | 0.96 (0.48 - 1.90) | 0.90 | 0.87 (0.43 - 1.74) | 0.70 |
| SIDIAP | 3,705 | 2,123 | 6.4 | 8.9 | 27 | 16 | 1.53 (0.59 - 4.04) | 0.39 | 1.60 (0.61 - 4.18) | 0.34 |
| Meta-analysis | 192,477 | 753,218 | 459.8 | 2870.9 | 6,185 | 50,083 | 0.98 (0.92 - 1.05) | 0.62 | 1.01 (0.84 - 1.23) | 0.88 |
| **PS-matching / intent-to-treat TAR** | | | | | | | | | | |
| OptumDOD | 10,106 | 45,167 | 15.8 | 110.0 | 494 | 3,832 | 1.02 (0.91 - 1.14) | 0.76 | 1.01 (0.90 - 1.14) | 0.83 |
| MDCR | 2,894 | 13,836 | 5.1 | 27.3 | 129 | 829 | 1.27 (0.99 - 1.60) | 0.05 | 1.26 (0.99 - 1.61) | 0.10 |
| Open Claims | 160,749 | 647,650 | 415.8 | 2545.6 | 5,174 | 41,073 | 0.95 (0.92 - 0.98) | <0.01 | 0.97 (0.79 - 1.20) | 0.81 |
| OptumEHR | 6,986 | 41,566 | 14.5 | 116.7 | 219 | 2,051 | 1.07 (0.90 - 1.25) | 0.44 | 1.11 (0.94 - 1.31) | 0.26 |
| VA | 4,826 | 31,081 | 6.1 | 72.1 | 150 | 1,155 | 1.46 (1.17 - 1.80) | <0.01 | 1.54 (1.24 - 1.92) | <0.01 |
| GermanyDA | 3,951 | 866 | 8.3 | 2.9 | 37 | 8 | 1.34 (0.56 - 3.59) | 0.53 | 1.20 (0.47 - 3.05) | 0.71 |
| SIDIAP | 3,283 | 963 | 6.0 | 2.9 | 26 | <5 | 5.15 (0.95 - 96.0) | 0.16 | 5.19 (0.51 - 52.5) | 0.17 |
| Meta-analysis | 180,735 | 748,219 | 451.2 | 2799.6 | 6,016 | 47,785 | 1.02 (0.93 - 1.13) | 0.62 | 1.04 (0.84 - 1.30) | 0.69 |

##### 6.5.12 Joint pain

**Supplementary Table 191:** Exposures, outcomes and risk estimates for Joint pain in SGLT2I (T) vs SU (C) new-user cohorts under four different propensity score (PS) adjustment / time-at-risk (TAR) designs across data sources. We report total patient exposure time in 1,000 patient-year (KYr) units, uncalibrated and calibrated hazard ratios (HRs) and their 95% confidence intervals (CIs). Greyed out entries did not pass study diagnostics and are included only for completeness.

| | Patients | | Exposure (KYr) | | Outcomes | | Uncalibrated | | Calibrated | |
|---|---|---|---|---|---|---|---|---|---|---|
| | T | C | T | C | T | C | HR (95% CI) | $p$ | HR (95% CI) | $p$ |
| **PS-stratification / on-treatment TAR** | | | | | | | | | | |
| OptumDOD | 10,899 | 46,674 | 6.5 | 46.3 | 82 | 590 | 0.90 (0.70 - 1.15) | 0.42 | 0.89 (0.69 - 1.14) | 0.37 |
| MDCR | 3,368 | 14,797 | 2.5 | 13.0 | 25 | 123 | 0.95 (0.58 - 1.51) | 0.83 | 0.93 (0.57 - 1.51) | 0.77 |
| Open Claims | 172,024 | 661,104 | 120.0 | 758.8 | 623 | 4,453 | 0.88 (0.80 - 0.96) | <0.01 | 0.89 (0.72 - 1.11) | 0.32 |
| OptumEHR | 7,405 | 43,892 | 2.0 | 26.6 | 25 | 265 | 1.26 (0.79 - 1.94) | 0.31 | 1.23 (0.78 - 1.94) | 0.38 |
| VA | 6,617 | 47,471 | 4.2 | 46.9 | 25 | 260 | 1.14 (0.65 - 1.94) | 0.64 | 1.23 (0.71 - 2.12) | 0.46 |
| GermanyDA | 3,517 | 1,353 | 2.7 | 1.4 | 36 | 32 | 0.90 (0.48 - 1.72) | 0.75 | 0.86 (0.45 - 1.63) | 0.64 |
| SIDIAP | 2,847 | 1,738 | 3.4 | 3.7 | 63 | 95 | 0.67 (0.43 - 1.03) | 0.07 | 0.73 (0.47 - 1.13) | 0.19 |
| Meta-analysis | 190,328 | 751,670 | 128.6 | 831.8 | 730 | 5,308 | 0.89 (0.82 - 0.97) | <0.01 | 0.89 (0.74 - 1.07) | 0.20 |
| **PS-matching / on-treatment TAR** | | | | | | | | | | |
| OptumDOD | 10,129 | 45,692 | 6.2 | 45.3 | 82 | 574 | 0.86 (0.62 - 1.17) | 0.35 | 0.84 (0.61 - 1.15) | 0.31 |
| MDCR | 2,921 | 14,040 | 2.2 | 12.4 | 24 | 116 | 1.05 (0.54 - 1.94) | 0.88 | 1.00 (0.53 - 1.89) | 0.92 |
| Open Claims | 158,135 | 644,855 | 113.1 | 741.1 | 576 | 4,308 | 0.86 (0.77 - 0.96) | <0.01 | 0.88 (0.70 - 1.11) | 0.28 |
| OptumEHR | 6,872 | 41,316 | 1.9 | 24.9 | 23 | 248 | 1.35 (0.73 - 2.41) | 0.32 | 1.34 (0.74 - 2.44) | 0.36 |
| VA | 4,695 | 29,907 | 3.4 | 26.3 | 17 | 122 | 0.86 (0.39 - 1.72) | 0.69 | 0.93 (0.44 - 1.95) | 0.85 |
| GermanyDA | 3,453 | 771 | 2.7 | 0.7 | 36 | 12 | 1.54 (0.61 - 4.42) | 0.39 | 1.84 (0.68 - 4.96) | 0.25 |
| SIDIAP | 2,523 | 773 | 3.2 | 1.3 | 59 | 33 | 0.91 (0.49 - 1.70) | 0.76 | 1.15 (0.61 - 2.14) | 0.68 |
| Meta-analysis | 178,057 | 745,903 | 123.4 | 823.7 | 705 | 5,246 | 0.88 (0.79 - 0.97) | 0.01 | 0.88 (0.70 - 1.11) | 0.29 |
| **PS-stratification / intent-to-treat TAR** | | | | | | | | | | |
| OptumDOD | 10,899 | 46,674 | 16.4 | 117.1 | 224 | 1,517 | 0.98 (0.84 - 1.14) | 0.78 | 0.98 (0.84 - 1.14) | 0.80 |
| MDCR | 3,368 | 14,797 | 5.6 | 29.7 | 57 | 329 | 0.96 (0.69 - 1.30) | 0.78 | 0.96 (0.68 - 1.35) | 0.81 |
| Open Claims | 172,024 | 661,104 | 423.4 | 2659.8 | 2,527 | 16,716 | 0.98 (0.94 - 1.03) | 0.51 | 1.01 (0.82 - 1.25) | 0.90 |
| OptumEHR | 7,405 | 43,892 | 15.0 | 129.6 | 147 | 989 | 1.28 (1.05 - 1.55) | 0.01 | 1.34 (1.11 - 1.63) | <0.01 |
| VA | 6,617 | 47,471 | 7.2 | 151.8 | 38 | 754 | 1.14 (0.75 - 1.70) | 0.52 | 1.20 (0.80 - 1.81) | 0.39 |
| GermanyDA | 3,517 | 1,353 | 7.2 | 5.4 | 102 | 92 | 1.07 (0.74 - 1.54) | 0.72 | 0.97 (0.67 - 1.40) | 0.75 |
| SIDIAP | 2,847 | 1,738 | 5.0 | 6.9 | 99 | 169 | 0.93 (0.66 - 1.29) | 0.67 | 0.98 (0.70 - 1.36) | 0.76 |
| Meta-analysis | 190,328 | 751,670 | 454.8 | 2906.5 | 2,898 | 19,222 | 1.05 (0.90 - 1.23) | 0.51 | 1.09 (0.86 - 1.38) | 0.49 |
| **PS-matching / intent-to-treat TAR** | | | | | | | | | | |
| OptumDOD | 10,129 | 45,692 | 16.0 | 113.6 | 217 | 1,470 | 1.00 (0.84 - 1.18) | 0.97 | 0.99 (0.83 - 1.19) | 0.90 |
| MDCR | 2,921 | 14,040 | 5.3 | 28.2 | 55 | 308 | 0.94 (0.64 - 1.35) | 0.73 | 0.93 (0.64 - 1.36) | 0.73 |
| Open Claims | 158,135 | 644,855 | 410.3 | 2568.7 | 2,440 | 16,015 | 0.99 (0.94 - 1.04) | 0.74 | 1.02 (0.82 - 1.25) | 0.88 |
| OptumEHR | 6,872 | 41,316 | 14.4 | 117.7 | 141 | 892 | 1.30 (1.05 - 1.60) | 0.02 | 1.35 (1.09 - 1.67) | <0.01 |
| VA | 4,695 | 29,907 | 6.0 | 70.7 | 29 | 300 | 1.15 (0.71 - 1.79) | 0.56 | 1.22 (0.76 - 1.94) | 0.42 |
| GermanyDA | 3,453 | 771 | 7.1 | 2.5 | 102 | 37 | 1.17 (0.75 - 1.85) | 0.50 | 1.04 (0.66 - 1.64) | 0.75 |
| SIDIAP | 2,523 | 773 | 4.7 | 2.2 | 95 | 47 | 0.94 (0.62 - 1.43) | 0.77 | 0.95 (0.61 - 1.47) | 0.75 |
| Meta-analysis | 178,057 | 745,903 | 445.9 | 2828.2 | 2,853 | 18,685 | 1.04 (0.92 - 1.18) | 0.51 | 1.06 (0.84 - 1.34) | 0.60 |

##### 6.5.13 Lower extremity amputation

**Supplementary Table 192:** Exposures, outcomes and risk estimates for Lower extremity amputation in SGLT2I (T) vs SU (C) new-user cohorts under four different propensity score (PS) adjustment / time-at-risk (TAR) designs across data sources. We report total patient exposure time in 1,000 patient-year (KYr) units, uncalibrated and calibrated hazard ratios (HRs) and their 95% confidence intervals (CIs). Greyed out entries did not pass study diagnostics and are included only for completeness.

| | Patients | | Exposure (KYr) | | Outcomes | | Uncalibrated | | Calibrated | |
|---|---|---|---|---|---|---|---|---|---|---|
| | T | C | T | C | T | C | HR (95% CI) | $p$ | HR (95% CI) | $p$ |
| **PS-stratification / on-treatment TAR** | | | | | | | | | | |
| OptumDOD | 11,589 | 49,525 | 6.9 | 49.4 | 6 | 76 | 0.79 (0.30 - 1.74) | 0.60 | 0.78 (0.32 - 1.87) | 0.57 |
| MDCR | 3,500 | 15,493 | 2.6 | 13.7 | <5 | 15 | 1.17 (0.23 - 4.26) | 0.83 | 1.14 (0.27 - 4.89) | 0.86 |
| Open Claims | 184,471 | 698,060 | 128.3 | 801.6 | 67 | 521 | 1.19 (0.89 - 1.56) | 0.23 | 1.21 (0.86 - 1.70) | 0.29 |
| OptumEHR | 7,821 | 46,072 | 2.2 | 28.3 | <5 | 28 | 0.75 (0.04 - 4.04) | 0.81 | 0.74 (0.07 - 7.36) | 0.79 |
| VA | 7,511 | 53,702 | 4.7 | 53.2 | <10 | 47 | 1.56 (0.42 - 5.05) | 0.48 | 1.68 (0.49 - 5.80) | 0.41 |
| GermanyDA | 4,154 | 1,547 | 3.2 | 1.6 | 0 | 0 | | | | |
| SIDIAP | 3,745 | 2,134 | 4.5 | 4.7 | 0 | <5 | | | | |
| Meta-analysis | 203,881 | 793,657 | 137.5 | 879.3 | <78 | 625 | 1.14 (0.87 - 1.49) | 0.34 | 1.13 (0.83 - 1.55) | 0.43 |
| **PS-matching / on-treatment TAR** | | | | | | | | | | |
| OptumDOD | 10,760 | 48,449 | 6.6 | 48.4 | 6 | 74 | 0.89 (0.30 - 2.18) | 0.82 | 0.87 (0.32 - 2.36) | 0.78 |
| MDCR | 3,034 | 14,725 | 2.3 | 13.0 | <5 | 14 | 1.29 (0.17 - 6.59) | 0.78 | 1.23 (0.20 - 7.60) | 0.82 |
| Open Claims | 169,437 | 681,660 | 120.8 | 783.6 | 65 | 485 | 1.22 (0.87 - 1.69) | 0.23 | 1.25 (0.85 - 1.84) | 0.27 |
| OptumEHR | 7,278 | 43,336 | 2.0 | 26.4 | <5 | 25 | | | | |
| VA | 5,273 | 33,797 | 3.8 | 29.5 | <10 | 23 | 1.96 (0.38 - 7.34) | 0.37 | 2.12 (0.48 - 9.27) | 0.32 |
| GermanyDA | 4,095 | 892 | 3.2 | 0.8 | 0 | 0 | | | | |
| SIDIAP | 3,313 | 970 | 4.1 | 1.7 | 0 | <5 | | | | |
| Meta-analysis | 180,197 | 730,109 | 127.4 | 831.9 | 71 | 559 | 1.19 (0.86 - 1.62) | 0.29 | 1.19 (0.82 - 1.74) | 0.35 |
| **PS-stratification / intent-to-treat TAR** | | | | | | | | | | |
| OptumDOD | 11,589 | 49,525 | 17.8 | 126.6 | 21 | 214 | 0.89 (0.54 - 1.39) | 0.63 | 0.89 (0.55 - 1.43) | 0.63 |
| MDCR | 3,500 | 15,493 | 5.9 | 31.5 | 7 | 34 | 1.40 (0.51 - 3.34) | 0.48 | 1.40 (0.54 - 3.61) | 0.50 |
| Open Claims | 184,471 | 698,060 | 457.9 | 2843.4 | 255 | 2,509 | 0.92 (0.80 - 1.05) | 0.23 | 0.95 (0.74 - 1.21) | 0.66 |
| OptumEHR | 7,821 | 46,072 | 16.2 | 138.2 | <5 | 127 | 0.40 (0.12 - 1.01) | 0.09 | 0.42 (0.14 - 1.23) | 0.11 |
| VA | 7,511 | 53,702 | 8.1 | 172.5 | 13 | 200 | 2.17 (1.02 - 4.26) | 0.03 | 2.28 (1.11 - 4.67) | 0.03 |
| GermanyDA | 4,154 | 1,547 | 8.7 | 6.5 | 0 | 0 | | | | |
| SIDIAP | 3,745 | 2,134 | 6.5 | 9.0 | 0 | <5 | | | | |
| Meta-analysis | 203,881 | 793,657 | 491.8 | 3108.2 | <281 | 2,850 | 0.91 (0.79 - 1.03) | 0.14 | 0.93 (0.75 - 1.17) | 0.55 |
| **PS-matching / intent-to-treat TAR** | | | | | | | | | | |
| OptumDOD | 10,760 | 48,449 | 17.3 | 122.7 | 21 | 209 | 0.96 (0.56 - 1.57) | 0.88 | 0.96 (0.57 - 1.61) | 0.87 |
| MDCR | 3,034 | 14,725 | 5.5 | 29.9 | 6 | 31 | 0.81 (0.18 - 2.67) | 0.76 | 0.81 (0.21 - 3.13) | 0.76 |
| Open Claims | 169,437 | 681,660 | 443.6 | 2748.8 | 246 | 2,337 | 0.96 (0.83 - 1.11) | 0.59 | 0.98 (0.77 - 1.26) | 0.89 |
| OptumEHR | 7,278 | 43,336 | 15.5 | 125.3 | <5 | 106 | 0.48 (0.14 - 1.22) | 0.18 | 0.50 (0.17 - 1.46) | 0.20 |
| VA | 5,273 | 33,797 | 6.8 | 79.5 | 12 | 87 | 1.83 (0.76 - 3.98) | 0.15 | 1.94 (0.85 - 4.44) | 0.12 |
| GermanyDA | 4,095 | 892 | 8.7 | 3.0 | 0 | 0 | | | | |
| SIDIAP | 3,313 | 970 | 6.1 | 2.9 | 0 | <5 | | | | |
| Meta-analysis | 190,509 | 788,170 | 481.9 | 3026.7 | <278 | 2,683 | 0.95 (0.82 - 1.09) | 0.45 | 0.97 (0.76 - 1.23) | 0.78 |

##### 6.5.14 Nausea

**Supplementary Table 193:** Exposures, outcomes and risk estimates for Nausea in SGLT2I (T) vs SU (C) new-user cohorts under four different propensity score (PS) adjustment / time-at-risk (TAR) designs across data sources. We report total patient exposure time in 1,000 patient-year (KYr) units, uncalibrated and calibrated hazard ratios (HRs) and their 95% confidence intervals (CIs). Greyed out entries did not pass study diagnostics and are included only for completeness.

| | Patients | | Exposure (KYr) | | Outcomes | | Uncalibrated | | Calibrated | |
|---|---|---|---|---|---|---|---|---|---|---|
| | T | C | T | C | T | C | HR (95% CI) | $p$ | HR (95% CI) | $p$ |
| **PS-stratification / on-treatment TAR** | | | | | | | | | | |
| OptumDOD | 9,971 | 41,942 | 5.9 | 41.1 | 215 | 1,798 | 0.85 (0.73 - 0.99) | 0.04 | 0.84 (0.71 - 0.98) | 0.04 |
| MDCR | 3,120 | 13,382 | 2.3 | 11.6 | 68 | 456 | 0.89 (0.66 - 1.18) | 0.43 | 0.87 (0.65 - 1.17) | 0.43 |
| Open Claims | 158,462 | 607,675 | 111.0 | 696.3 | 1,750 | 14,690 | 0.84 (0.80 - 0.89) | <0.01 | 0.85 (0.69 - 1.05) | 0.14 |
| OptumEHR | 7,090 | 41,646 | 2.0 | 24.7 | 62 | 903 | 1.15 (0.86 - 1.51) | 0.32 | 1.13 (0.85 - 1.49) | 0.44 |
| VA | 6,967 | 50,402 | 4.4 | 49.9 | 30 | 423 | 0.75 (0.46 - 1.18) | 0.22 | 0.81 (0.51 - 1.29) | 0.38 |
| GermanyDA | 3,888 | 1,428 | 3.0 | 1.5 | 20 | 15 | 1.58 (0.61 - 4.19) | 0.35 | 1.50 (0.57 - 3.95) | 0.43 |
| SIDIAP | 3,589 | 2,054 | 4.3 | 4.5 | 17 | 32 | 0.48 (0.20 - 1.10) | 0.10 | 0.53 (0.22 - 1.26) | 0.16 |
| Meta-analysis | 175,523 | 691,263 | 118.9 | 762.1 | 2,027 | 17,391 | 0.89 (0.78 - 1.01) | 0.07 | 0.88 (0.72 - 1.09) | 0.24 |
| **PS-matching / on-treatment TAR** | | | | | | | | | | |
| OptumDOD | 9,287 | 41,144 | 5.7 | 40.4 | 206 | 1,748 | 0.90 (0.75 - 1.08) | 0.27 | 0.88 (0.73 - 1.06) | 0.22 |
| MDCR | 2,721 | 12,944 | 2.1 | 11.3 | 63 | 440 | 1.00 (0.69 - 1.42) | 0.99 | 0.95 (0.66 - 1.37) | 0.80 |
| Open Claims | 145,552 | 593,196 | 104.5 | 680.3 | 1,643 | 14,149 | 0.86 (0.80 - 0.91) | <0.01 | 0.88 (0.71 - 1.08) | 0.23 |
| OptumEHR | 6,581 | 39,191 | 1.8 | 23.1 | 58 | 813 | 1.10 (0.78 - 1.52) | 0.57 | 1.09 (0.78 - 1.52) | 0.66 |
| VA | 4,955 | 31,759 | 3.6 | 27.9 | 18 | 220 | 0.69 (0.35 - 1.25) | 0.25 | 0.75 (0.40 - 1.40) | 0.37 |
| GermanyDA | 3,830 | 831 | 3.0 | 0.8 | 20 | <5 | 3.39 (0.59 - 64.3) | 0.31 | 4.04 (0.39 - 42.2) | 0.25 |
| SIDIAP | 3,188 | 933 | 4.0 | 1.6 | 13 | 10 | 0.98 (0.26 - 4.01) | 0.98 | 1.24 (0.32 - 4.87) | 0.77 |
| Meta-analysis | 164,141 | 686,475 | 114.1 | 755.1 | 1,970 | 17,150 | 0.88 (0.82 - 0.95) | <0.01 | 0.89 (0.71 - 1.10) | 0.27 |
| **PS-stratification / intent-to-treat TAR** | | | | | | | | | | |
| OptumDOD | 9,971 | 41,942 | 14.7 | 99.7 | 588 | 4,581 | 0.92 (0.84 - 1.01) | 0.07 | 0.92 (0.83 - 1.01) | 0.14 |
| MDCR | 3,120 | 13,382 | 5.1 | 25.8 | 171 | 1,131 | 0.98 (0.82 - 1.17) | 0.82 | 0.98 (0.78 - 1.24) | 0.86 |
| Open Claims | 158,462 | 607,675 | 384.3 | 2360.9 | 7,063 | 56,251 | 0.91 (0.88 - 0.93) | <0.01 | 0.93 (0.76 - 1.15) | 0.52 |
| OptumEHR | 7,090 | 41,646 | 14.1 | 118.6 | 301 | 3,055 | 1.03 (0.90 - 1.17) | 0.65 | 1.08 (0.95 - 1.23) | 0.30 |
| VA | 6,967 | 50,402 | 7.6 | 160.2 | 54 | 1,508 | 0.73 (0.52 - 1.01) | 0.06 | 0.77 (0.56 - 1.07) | 0.13 |
| GermanyDA | 3,888 | 1,428 | 8.1 | 5.8 | 64 | 60 | 1.54 (0.97 - 2.43) | 0.07 | 1.40 (0.88 - 2.22) | 0.19 |
| SIDIAP | 3,589 | 2,054 | 6.2 | 8.5 | 34 | 74 | 0.59 (0.33 - 1.01) | 0.06 | 0.62 (0.35 - 1.08) | 0.10 |
| Meta-analysis | 175,523 | 691,263 | 413.1 | 2579.2 | 7,952 | 63,887 | 0.93 (0.88 - 0.98) | <0.01 | 0.95 (0.79 - 1.15) | 0.63 |
| **PS-matching / intent-to-treat TAR** | | | | | | | | | | |
| OptumDOD | 9,287 | 41,144 | 14.3 | 97.2 | 572 | 4,420 | 0.92 (0.83 - 1.02) | 0.13 | 0.92 (0.82 - 1.03) | 0.23 |
| MDCR | 2,721 | 12,944 | 4.7 | 25.0 | 162 | 1,082 | 1.06 (0.86 - 1.30) | 0.57 | 1.05 (0.85 - 1.30) | 0.69 |
| Open Claims | 145,552 | 593,196 | 372.1 | 2284.2 | 6,865 | 53,533 | 0.92 (0.89 - 0.95) | <0.01 | 0.94 (0.77 - 1.16) | 0.57 |
| OptumEHR | 6,581 | 39,191 | 13.5 | 108.1 | 289 | 2,686 | 1.08 (0.93 - 1.24) | 0.32 | 1.12 (0.97 - 1.29) | 0.17 |
| VA | 4,955 | 31,759 | 6.4 | 74.4 | 38 | 644 | 0.70 (0.46 - 1.02) | 0.08 | 0.74 (0.49 - 1.10) | 0.15 |
| GermanyDA | 3,830 | 831 | 8.1 | 2.8 | 63 | 18 | 1.60 (0.89 - 3.00) | 0.13 | 1.43 (0.78 - 2.63) | 0.28 |
| SIDIAP | 3,188 | 933 | 5.9 | 2.8 | 30 | 20 | 0.55 (0.26 - 1.15) | 0.12 | 0.55 (0.26 - 1.18) | 0.16 |
| Meta-analysis | 164,141 | 686,475 | 404.6 | 2514.5 | 7,888 | 61,721 | 0.96 (0.89 - 1.04) | 0.32 | 0.98 (0.80 - 1.21) | 0.86 |

##### 6.5.15 Peripheral edema

**Supplementary Table 194:** Exposures, outcomes and risk estimates for Peripheral edema in SGLT2I (T) vs SU (C) new-user cohorts under four different propensity score (PS) adjustment / time-at-risk (TAR) designs across data sources. We report total patient exposure time in 1,000 patient-year (KYr) units, uncalibrated and calibrated hazard ratios (HRs) and their 95% confidence intervals (CIs). Greyed out entries did not pass study diagnostics and are included only for completeness.

| | Patients | | Exposure (KYr) | | Outcomes | | Uncalibrated | | Calibrated | |
|---|---|---|---|---|---|---|---|---|---|---|
| | T | C | T | C | T | C | HR (95% CI) | $p$ | HR (95% CI) | $p$ |
| **PS-stratification / on-treatment TAR** | | | | | | | | | | |
| OptumDOD | 8,491 | 35,457 | 5.0 | 33.0 | 343 | 3,329 | 0.72 (0.64 - 0.81) | <0.01 | 0.71 (0.63 - 0.80) | <0.01 |
| MDCR | 2,647 | 11,067 | 1.9 | 9.1 | 82 | 877 | 0.54 (0.42 - 0.69) | <0.01 | 0.53 (0.41 - 0.69) | <0.01 |
| Open Claims | 132,741 | 519,881 | 93.2 | 579.7 | 2,437 | 27,521 | 0.67 (0.64 - 0.70) | <0.01 | 0.68 (0.56 - 0.84) | <0.01 |
| OptumEHR | 6,272 | 36,844 | 1.7 | 20.7 | 47 | 1,692 | 0.44 (0.32 - 0.59) | <0.01 | 0.43 (0.32 - 0.59) | <0.01 |
| VA | 5,373 | 40,662 | 3.4 | 38.8 | 104 | 1,776 | 0.65 (0.51 - 0.83) | <0.01 | 0.70 (0.55 - 0.90) | <0.01 |
| GermanyDA | 3,374 | 1,282 | 2.6 | 1.3 | 56 | 63 | 0.43 (0.25 - 0.72) | <0.01 | 0.40 (0.24 - 0.68) | <0.01 |
| SIDIAP | 3,190 | 1,916 | 3.9 | 4.1 | 42 | 89 | 0.59 (0.35 - 0.97) | 0.04 | 0.65 (0.39 - 1.07) | 0.11 |
| Meta-analysis | 147,504 | 592,182 | 99.9 | 633.4 | 2,827 | 32,542 | 0.62 (0.48 - 0.80) | <0.01 | 0.62 (0.46 - 0.84) | <0.01 |
| **PS-matching / on-treatment TAR** | | | | | | | | | | |
| OptumDOD | 7,985 | 34,773 | 4.8 | 32.4 | 324 | 3,220 | 0.68 (0.59 - 0.78) | <0.01 | 0.66 (0.57 - 0.77) | <0.01 |
| MDCR | 2,338 | 10,699 | 1.7 | 8.8 | 75 | 825 | 0.59 (0.43 - 0.80) | <0.01 | 0.56 (0.41 - 0.77) | <0.01 |
| Open Claims | 123,465 | 508,290 | 88.5 | 567.7 | 2,299 | 26,425 | 0.68 (0.65 - 0.72) | <0.01 | 0.70 (0.57 - 0.86) | <0.01 |
| OptumEHR | 5,847 | 34,817 | 1.6 | 19.5 | 47 | 1,520 | 0.47 (0.32 - 0.65) | <0.01 | 0.46 (0.32 - 0.66) | <0.01 |
| VA | 3,926 | 25,913 | 2.8 | 22.1 | 74 | 949 | 0.67 (0.49 - 0.90) | <0.01 | 0.73 (0.54 - 0.98) | 0.04 |
| GermanyDA | 3,323 | 734 | 2.6 | 0.7 | 55 | 31 | 0.42 (0.23 - 0.78) | <0.01 | 0.50 (0.27 - 0.93) | 0.05 |
| SIDIAP | 2,854 | 856 | 3.6 | 1.5 | 39 | 31 | 0.55 (0.26 - 1.13) | 0.11 | 0.69 (0.33 - 1.45) | 0.36 |
| Meta-analysis | 139,635 | 588,579 | 96.6 | 628.5 | 2,745 | 31,990 | 0.68 (0.64 - 0.71) | <0.01 | 0.68 (0.55 - 0.84) | <0.01 |
| **PS-stratification / intent-to-treat TAR** | | | | | | | | | | |
| OptumDOD | 8,491 | 35,457 | 12.2 | 77.5 | 927 | 7,449 | 0.84 (0.78 - 0.90) | <0.01 | 0.84 (0.77 - 0.91) | <0.01 |
| MDCR | 2,647 | 11,067 | 4.1 | 19.6 | 261 | 1,969 | 0.78 (0.67 - 0.89) | <0.01 | 0.78 (0.63 - 0.95) | 0.06 |
| Open Claims | 132,741 | 519,881 | 316.7 | 1910.0 | 10,335 | 92,244 | 0.83 (0.82 - 0.85) | <0.01 | 0.86 (0.70 - 1.05) | 0.15 |
| OptumEHR | 6,272 | 36,844 | 12.3 | 97.9 | 421 | 5,526 | 0.78 (0.70 - 0.87) | <0.01 | 0.82 (0.73 - 0.91) | <0.01 |
| VA | 5,373 | 40,662 | 5.9 | 122.7 | 198 | 5,052 | 0.78 (0.66 - 0.93) | <0.01 | 0.83 (0.69 - 0.98) | 0.04 |
| GermanyDA | 3,374 | 1,282 | 6.9 | 4.9 | 154 | 197 | 0.64 (0.48 - 0.84) | <0.01 | 0.58 (0.43 - 0.77) | <0.01 |
| SIDIAP | 3,190 | 1,916 | 5.6 | 7.7 | 83 | 173 | 0.90 (0.64 - 1.26) | 0.54 | 0.94 (0.67 - 1.33) | 0.72 |
| Meta-analysis | 147,504 | 592,182 | 341.2 | 2085.4 | 11,683 | 105,219 | 0.83 (0.82 - 0.85) | <0.01 | 0.86 (0.72 - 1.03) | 0.10 |
| **PS-matching / intent-to-treat TAR** | | | | | | | | | | |
| OptumDOD | 7,985 | 34,773 | 11.9 | 75.6 | 899 | 7,154 | 0.82 (0.75 - 0.89) | <0.01 | 0.81 (0.74 - 0.89) | <0.01 |
| MDCR | 2,338 | 10,699 | 3.9 | 19.0 | 245 | 1,865 | 0.76 (0.64 - 0.89) | <0.01 | 0.75 (0.63 - 0.90) | <0.01 |
| Open Claims | 123,465 | 508,290 | 308.0 | 1855.5 | 10,086 | 87,725 | 0.83 (0.81 - 0.85) | <0.01 | 0.85 (0.69 - 1.04) | 0.12 |
| OptumEHR | 5,847 | 34,817 | 11.8 | 90.3 | 411 | 4,856 | 0.75 (0.67 - 0.84) | <0.01 | 0.78 (0.69 - 0.88) | <0.01 |
| VA | 3,926 | 25,913 | 5.0 | 59.2 | 150 | 2,367 | 0.80 (0.65 - 0.97) | 0.03 | 0.85 (0.69 - 1.03) | 0.13 |
| GermanyDA | 3,323 | 734 | 6.9 | 2.3 | 153 | 85 | 0.54 (0.38 - 0.77) | <0.01 | 0.48 (0.34 - 0.69) | <0.01 |
| SIDIAP | 2,854 | 856 | 5.3 | 2.5 | 79 | 51 | 0.85 (0.55 - 1.29) | 0.44 | 0.85 (0.54 - 1.33) | 0.56 |
| Meta-analysis | 139,635 | 588,579 | 335.6 | 2040.3 | 11,641 | 101,600 | 0.82 (0.79 - 0.85) | <0.01 | 0.83 (0.68 - 1.02) | 0.07 |

##### 6.5.16 Photosensitivity

**Supplementary Table 195:** Exposures, outcomes and risk estimates for Photosensitivity in SGLT2I (T) vs SU (C) new-user cohorts under four different propensity score (PS) adjustment / time-at-risk (TAR) designs across data sources. We report total patient exposure time in 1,000 patient-year (KYr) units, uncalibrated and calibrated hazard ratios (HRs) and their 95% confidence intervals (CIs). Greyed out entries did not pass study diagnostics and are included only for completeness.

| | Patients | | Exposure (KYr) | | Outcomes | | Uncalibrated | | Calibrated | |
|---|---|---|---|---|---|---|---|---|---|---|
| | T | C | T | C | T | C | HR (95% CI) | $p$ | HR (95% CI) | $p$ |
| **PS-stratification / on-treatment TAR** | | | | | | | | | | |
| OptumDOD | 11,516 | 49,068 | 6.9 | 48.9 | <5 | 38 | 0.83 (0.19 - 2.60) | 0.79 | 0.82 (0.22 - 3.06) | 0.77 |
| MDCR | 3,464 | 15,230 | 2.5 | 13.4 | <5 | 32 | 0.75 (0.16 - 2.44) | 0.68 | 0.73 (0.19 - 2.83) | 0.65 |
| Open Claims | 181,939 | 690,142 | 126.5 | 791.2 | 29 | 413 | 0.96 (0.63 - 1.41) | 0.85 | 0.98 (0.62 - 1.53) | 0.92 |
| OptumEHR | 7,758 | 45,589 | 2.2 | 28.0 | 0 | 65 | | | | |
| VA | 7,386 | 52,854 | 4.6 | 52.3 | 0 | 32 | | | | |
| GermanyDA | 4,146 | 1,544 | 3.2 | 1.6 | 0 | <5 | | | | |
| SIDIAP | 3,737 | 2,132 | 4.4 | 4.7 | 0 | <5 | | | | |
| Meta-analysis | 193,455 | 739,210 | 133.4 | 840.1 | <34 | 451 | 0.95 (0.65 - 1.40) | 0.80 | 0.95 (0.62 - 1.44) | 0.80 |
| **PS-matching / on-treatment TAR** | | | | | | | | | | |
| OptumDOD | 10,694 | 48,019 | 6.6 | 47.9 | <5 | 34 | 2.63 (0.37 - 11.1) | 0.26 | 2.57 (0.47 - 14.0) | 0.28 |
| MDCR | 3,000 | 14,491 | 2.3 | 12.8 | <5 | 32 | 0.89 (0.19 - 3.01) | 0.86 | 0.84 (0.21 - 3.36) | 0.81 |
| Open Claims | 167,162 | 673,821 | 119.1 | 773.3 | 28 | 376 | 0.93 (0.58 - 1.44) | 0.76 | 0.95 (0.58 - 1.57) | 0.84 |
| OptumEHR | 7,222 | 42,897 | 2.0 | 26.1 | 0 | 56 | | | | |
| VA | 5,202 | 33,266 | 3.7 | 29.1 | 0 | 10 | | | | |
| GermanyDA | 4,087 | 891 | 3.2 | 0.8 | 0 | 0 | | | | |
| SIDIAP | 3,308 | 970 | 4.1 | 1.7 | 0 | 0 | | | | |
| Meta-analysis | 180,856 | 736,331 | 127.9 | 833.9 | <38 | 442 | 0.99 (0.65 - 1.51) | 0.95 | 1.00 (0.62 - 1.59) | 0.95 |
| **PS-stratification / intent-to-treat TAR** | | | | | | | | | | |
| OptumDOD | 11,516 | 49,068 | 17.6 | 125.2 | 9 | 79 | 0.85 (0.38 - 1.72) | 0.68 | 0.85 (0.40 - 1.82) | 0.68 |
| MDCR | 3,464 | 15,230 | 5.8 | 30.9 | 9 | 53 | 1.35 (0.57 - 2.89) | 0.47 | 1.35 (0.59 - 3.08) | 0.49 |
| Open Claims | 181,939 | 690,142 | 451.1 | 2810.5 | 97 | 960 | 1.00 (0.79 - 1.24) | 0.98 | 1.03 (0.76 - 1.39) | 0.87 |
| OptumEHR | 7,758 | 45,589 | 16.0 | 136.3 | 7 | 161 | 1.09 (0.45 - 2.25) | 0.84 | 1.14 (0.51 - 2.56) | 0.75 |
| VA | 7,386 | 52,854 | 8.0 | 170.0 | <10 | 60 | 1.47 (0.22 - 5.63) | 0.64 | 1.55 (0.31 - 7.81) | 0.60 |
| GermanyDA | 4,146 | 1,544 | 8.7 | 6.5 | 0 | <5 | | | | |
| SIDIAP | 3,737 | 2,132 | 6.5 | 9.0 | 0 | <5 | | | | |
| Meta-analysis | 201,213 | 784,799 | 484.7 | 3072.0 | 113 | 1,200 | 0.99 (0.81 - 1.22) | 0.93 | 1.02 (0.78 - 1.34) | 0.88 |
| **PS-matching / intent-to-treat TAR** | | | | | | | | | | |
| OptumDOD | 10,694 | 48,019 | 17.1 | 121.4 | 8 | 72 | 0.83 (0.30 - 1.95) | 0.70 | 0.83 (0.33 - 2.12) | 0.70 |
| MDCR | 3,000 | 14,491 | 5.5 | 29.4 | 9 | 50 | 0.98 (0.32 - 2.44) | 0.97 | 0.97 (0.35 - 2.68) | 0.95 |
| Open Claims | 167,162 | 673,821 | 437.0 | 2716.2 | 95 | 887 | 0.99 (0.77 - 1.26) | 0.93 | 1.01 (0.74 - 1.39) | 0.93 |
| OptumEHR | 7,222 | 42,897 | 15.4 | 123.8 | 7 | 132 | 1.09 (0.44 - 2.29) | 0.84 | 1.13 (0.49 - 2.58) | 0.77 |
| VA | 5,202 | 33,266 | 6.7 | 78.3 | <10 | 24 | 2.87 (0.43 - 11.0) | 0.20 | 3.04 (0.60 - 15.3) | 0.18 |
| GermanyDA | 4,087 | 891 | 8.7 | 3.0 | 0 | 0 | | | | |
| SIDIAP | 3,308 | 970 | 6.1 | 2.9 | 0 | 0 | | | | |
| Meta-analysis | 188,078 | 779,228 | 474.9 | 2990.8 | 119 | 1,141 | 0.99 (0.79 - 1.23) | 0.90 | 1.01 (0.75 - 1.35) | 0.94 |

##### 6.5.17 Venous thromboembolism

**Supplementary Table 196:** Exposures, outcomes and risk estimates for Venous thromboembolism in SGLT2I (T) vs SU (C) new-user cohorts under four different propensity score (PS) adjustment / time-at-risk (TAR) designs across data sources. We report total patient exposure time in 1,000 patient-year (KYr) units, uncalibrated and calibrated hazard ratios (HRs) and their 95% confidence intervals (CIs). Greyed out entries did not pass study diagnostics and are included only for completeness.

| | Patients | | Exposure (KYr) | | Outcomes | | Uncalibrated | | Calibrated | |
|---|---|---|---|---|---|---|---|---|---|---|
| | T | C | T | C | T | C | HR (95% CI) | $p$ | HR (95% CI) | $p$ |
| **PS-stratification / on-treatment TAR** | | | | | | | | | | |
| OptumDOD | 11,177 | 47,431 | 6.7 | 47.0 | 67 | 655 | 0.86 (0.65 - 1.12) | 0.26 | 0.84 (0.64 - 1.11) | 0.24 |
| MDCR | 3,367 | 14,720 | 2.5 | 12.9 | 19 | 203 | 0.66 (0.38 - 1.10) | 0.13 | 0.65 (0.38 - 1.10) | 0.13 |
| Open Claims | 176,274 | 668,987 | 122.7 | 766.0 | 661 | 6,288 | 0.80 (0.73 - 0.87) | <0.01 | 0.81 (0.65 - 1.00) | 0.06 |
| OptumEHR | 7,536 | 44,398 | 2.1 | 26.9 | 17 | 375 | 0.90 (0.52 - 1.45) | 0.68 | 0.88 (0.52 - 1.47) | 0.62 |
| VA | 7,154 | 51,672 | 4.5 | 51.0 | 21 | 299 | 0.63 (0.36 - 1.07) | 0.10 | 0.68 (0.39 - 1.18) | 0.17 |
| GermanyDA | 3,838 | 1,426 | 3.0 | 1.5 | 27 | 20 | 0.80 (0.36 - 1.77) | 0.58 | 0.76 (0.34 - 1.68) | 0.52 |
| SIDIAP | 3,597 | 2,051 | 4.3 | 4.5 | 19 | 25 | 0.56 (0.24 - 1.26) | 0.16 | 0.61 (0.27 - 1.40) | 0.25 |
| Meta-analysis | 194,987 | 760,816 | 131.5 | 839.9 | 745 | 7,318 | 0.80 (0.74 - 0.87) | <0.01 | 0.80 (0.67 - 0.96) | 0.02 |
| **PS-matching / on-treatment TAR** | | | | | | | | | | |
| OptumDOD | 10,388 | 46,555 | 6.4 | 46.2 | 62 | 624 | 0.85 (0.61 - 1.17) | 0.33 | 0.83 (0.60 - 1.16) | 0.29 |
| MDCR | 2,917 | 13,883 | 2.2 | 12.2 | 17 | 184 | 1.07 (0.56 - 1.91) | 0.84 | 1.02 (0.55 - 1.88) | 0.91 |
| Open Claims | 162,134 | 652,654 | 115.6 | 748.1 | 623 | 5,963 | 0.76 (0.69 - 0.85) | <0.01 | 0.78 (0.62 - 0.98) | 0.04 |
| OptumEHR | 7,012 | 41,767 | 1.9 | 25.2 | 16 | 321 | 1.05 (0.56 - 1.85) | 0.87 | 1.04 (0.57 - 1.90) | 0.87 |
| VA | 5,042 | 32,270 | 3.6 | 28.1 | 16 | 148 | 0.52 (0.24 - 1.01) | 0.07 | 0.56 (0.27 - 1.15) | 0.12 |
| GermanyDA | 3,782 | 828 | 2.9 | 0.8 | 27 | 9 | 0.62 (0.20 - 2.17) | 0.43 | 0.74 (0.22 - 2.44) | 0.63 |
| SIDIAP | 3,161 | 924 | 3.9 | 1.6 | 16 | 12 | 1.22 (0.41 - 4.05) | 0.74 | 1.54 (0.49 - 4.86) | 0.49 |
| Meta-analysis | 182,451 | 754,859 | 126.2 | 831.7 | 718 | 7,092 | 0.79 (0.71 - 0.89) | <0.01 | 0.80 (0.63 - 1.01) | 0.06 |
| **PS-stratification / intent-to-treat TAR** | | | | | | | | | | |
| OptumDOD | 11,177 | 47,431 | 16.9 | 118.9 | 209 | 1,824 | 0.92 (0.79 - 1.07) | 0.31 | 0.92 (0.79 - 1.08) | 0.35 |
| MDCR | 3,367 | 14,720 | 5.6 | 29.3 | 61 | 535 | 0.78 (0.58 - 1.04) | 0.10 | 0.78 (0.56 - 1.08) | 0.18 |
| Open Claims | 176,274 | 668,987 | 435.3 | 2688.6 | 2,914 | 24,317 | 0.91 (0.87 - 0.95) | <0.01 | 0.93 (0.76 - 1.15) | 0.53 |
| OptumEHR | 7,536 | 44,398 | 15.4 | 131.0 | 104 | 1,221 | 0.90 (0.72 - 1.11) | 0.32 | 0.94 (0.75 - 1.17) | 0.59 |
| VA | 7,154 | 51,672 | 7.7 | 165.0 | 50 | 1,152 | 0.85 (0.60 - 1.21) | 0.38 | 0.90 (0.63 - 1.28) | 0.57 |
| GermanyDA | 3,838 | 1,426 | 8.0 | 5.8 | 63 | 50 | 1.15 (0.71 - 1.86) | 0.56 | 1.05 (0.65 - 1.70) | 0.80 |
| SIDIAP | 3,597 | 2,051 | 6.2 | 8.6 | 30 | 43 | 0.86 (0.46 - 1.58) | 0.62 | 0.90 (0.49 - 1.67) | 0.74 |
| Meta-analysis | 194,987 | 760,816 | 467.6 | 2938.5 | 3,227 | 27,362 | 0.91 (0.87 - 0.94) | <0.01 | 0.94 (0.78 - 1.13) | 0.49 |
| **PS-matching / intent-to-treat TAR** | | | | | | | | | | |
| OptumDOD | 10,388 | 46,555 | 16.5 | 115.8 | 201 | 1,728 | 0.92 (0.77 - 1.08) | 0.31 | 0.91 (0.77 - 1.09) | 0.36 |
| MDCR | 2,917 | 13,883 | 5.3 | 27.6 | 57 | 482 | 0.75 (0.53 - 1.05) | 0.11 | 0.75 (0.53 - 1.06) | 0.13 |
| Open Claims | 162,134 | 652,654 | 421.7 | 2596.6 | 2,832 | 22,849 | 0.91 (0.87 - 0.95) | <0.01 | 0.93 (0.75 - 1.14) | 0.48 |
| OptumEHR | 7,012 | 41,767 | 14.8 | 119.0 | 98 | 1,058 | 0.82 (0.64 - 1.05) | 0.13 | 0.86 (0.67 - 1.10) | 0.25 |
| VA | 5,042 | 32,270 | 6.5 | 75.2 | 38 | 483 | 0.80 (0.52 - 1.20) | 0.31 | 0.85 (0.56 - 1.30) | 0.46 |
| GermanyDA | 3,782 | 828 | 7.9 | 2.8 | 63 | 20 | 1.69 (0.88 - 3.48) | 0.14 | 1.51 (0.76 - 3.02) | 0.26 |
| SIDIAP | 3,161 | 924 | 5.8 | 2.7 | 27 | 15 | 0.71 (0.32 - 1.57) | 0.39 | 0.71 (0.32 - 1.59) | 0.44 |
| Meta-analysis | 182,451 | 754,859 | 458.3 | 2859.0 | 3,188 | 26,117 | 0.90 (0.86 - 0.94) | <0.01 | 0.92 (0.75 - 1.12) | 0.41 |

##### 6.5.18 Vomiting

**Supplementary Table 197:** Exposures, outcomes and risk estimates for Vomiting in SGLT2I (T) vs SU (C) new-user cohorts under four different propensity score (PS) adjustment / time-at-risk (TAR) designs across data sources. We report total patient exposure time in 1,000 patient-year (KYr) units, uncalibrated and calibrated hazard ratios (HRs) and their 95% confidence intervals (CIs). Greyed out entries did not pass study diagnostics and are included only for completeness.

| | Patients | | Exposure (KYr) | | Outcomes | | Uncalibrated | | Calibrated | |
|---|---|---|---|---|---|---|---|---|---|---|
| | T | C | T | C | T | C | HR (95% CI) | $p$ | HR (95% CI) | $p$ |
| **PS-stratification / on-treatment TAR** | | | | | | | | | | |
| OptumDOD | 10,476 | 43,978 | 6.2 | 43.4 | 155 | 1,418 | 0.78 (0.65 - 0.93) | <0.01 | 0.77 (0.64 - 0.92) | <0.01 |
| MDCR | 3,271 | 13,927 | 2.4 | 12.2 | 48 | 379 | 0.84 (0.59 - 1.16) | 0.30 | 0.82 (0.58 - 1.15) | 0.31 |
| Open Claims | 165,476 | 628,373 | 115.6 | 721.2 | 1,324 | 11,672 | 0.83 (0.78 - 0.89) | <0.01 | 0.85 (0.69 - 1.04) | 0.13 |
| OptumEHR | 7,398 | 43,245 | 2.1 | 26.0 | 40 | 627 | 1.08 (0.76 - 1.51) | 0.65 | 1.06 (0.75 - 1.49) | 0.76 |
| VA | 7,106 | 51,346 | 4.5 | 50.9 | 25 | 310 | 0.70 (0.41 - 1.15) | 0.17 | 0.75 (0.45 - 1.27) | 0.29 |
| GermanyDA | 3,887 | 1,428 | 3.0 | 1.5 | 20 | 15 | 1.58 (0.61 - 4.20) | 0.35 | 1.50 (0.57 - 3.96) | 0.43 |
| SIDIAP | 3,583 | 2,045 | 4.3 | 4.5 | 19 | 31 | 0.48 (0.20 - 1.08) | 0.08 | 0.53 (0.23 - 1.21) | 0.14 |
| Meta-analysis | 183,350 | 715,596 | 123.9 | 790.6 | 1,519 | 13,717 | 0.83 (0.79 - 0.88) | <0.01 | 0.83 (0.70 - 0.99) | 0.03 |
| **PS-matching / on-treatment TAR** | | | | | | | | | | |
| OptumDOD | 9,734 | 43,246 | 6.0 | 42.7 | 151 | 1,373 | 0.87 (0.71 - 1.07) | 0.20 | 0.85 (0.69 - 1.05) | 0.17 |
| MDCR | 2,837 | 13,465 | 2.2 | 11.8 | 46 | 364 | 0.80 (0.51 - 1.20) | 0.30 | 0.76 (0.49 - 1.17) | 0.23 |
| Open Claims | 151,965 | 613,909 | 108.9 | 705.2 | 1,230 | 11,212 | 0.84 (0.78 - 0.91) | <0.01 | 0.86 (0.69 - 1.06) | 0.18 |
| OptumEHR | 6,867 | 40,661 | 1.9 | 24.3 | 39 | 558 | 1.44 (0.96 - 2.10) | 0.07 | 1.42 (0.96 - 2.11) | 0.10 |
| VA | 5,037 | 32,331 | 3.6 | 28.3 | 17 | 161 | 0.80 (0.38 - 1.56) | 0.53 | 0.86 (0.43 - 1.75) | 0.69 |
| GermanyDA | 3,829 | 831 | 3.0 | 0.8 | 20 | <5 | 3.39 (0.59 - 64.3) | 0.31 | 4.04 (0.39 - 42.2) | 0.25 |
| SIDIAP | 3,184 | 931 | 4.0 | 1.7 | 15 | 11 | 1.08 (0.30 - 4.30) | 0.91 | 1.36 (0.36 - 5.14) | 0.67 |
| Meta-analysis | 171,403 | 711,281 | 118.9 | 784.1 | 1,466 | 13,507 | 0.92 (0.75 - 1.12) | 0.39 | 0.93 (0.70 - 1.23) | 0.59 |
| **PS-stratification / intent-to-treat TAR** | | | | | | | | | | |
| OptumDOD | 10,476 | 43,978 | 15.6 | 106.3 | 462 | 3,769 | 0.92 (0.83 - 1.02) | 0.13 | 0.92 (0.83 - 1.03) | 0.20 |
| MDCR | 3,271 | 13,927 | 5.4 | 27.1 | 124 | 980 | 0.87 (0.71 - 1.07) | 0.19 | 0.87 (0.68 - 1.13) | 0.37 |
| Open Claims | 165,476 | 628,373 | 403.7 | 2466.9 | 5,532 | 47,154 | 0.89 (0.86 - 0.92) | <0.01 | 0.92 (0.74 - 1.13) | 0.41 |
| OptumEHR | 7,398 | 43,245 | 14.9 | 125.1 | 186 | 2,266 | 0.91 (0.77 - 1.06) | 0.24 | 0.95 (0.81 - 1.12) | 0.58 |
| VA | 7,106 | 51,346 | 7.7 | 163.8 | 54 | 1,154 | 0.95 (0.68 - 1.33) | 0.79 | 1.01 (0.72 - 1.41) | 0.91 |
| GermanyDA | 3,887 | 1,428 | 8.1 | 5.8 | 66 | 60 | 1.55 (0.98 - 2.45) | 0.06 | 1.41 (0.89 - 2.23) | 0.18 |
| SIDIAP | 3,583 | 2,045 | 6.2 | 8.5 | 37 | 77 | 0.57 (0.33 - 0.97) | 0.04 | 0.60 (0.35 - 1.03) | 0.07 |
| Meta-analysis | 183,350 | 715,596 | 434.3 | 2698.4 | 6,180 | 53,189 | 0.89 (0.87 - 0.92) | <0.01 | 0.92 (0.77 - 1.10) | 0.38 |
| **PS-matching / intent-to-treat TAR** | | | | | | | | | | |
| OptumDOD | 9,734 | 43,246 | 15.2 | 103.9 | 454 | 3,631 | 0.93 (0.83 - 1.04) | 0.21 | 0.93 (0.82 - 1.05) | 0.31 |
| MDCR | 2,837 | 13,465 | 5.0 | 26.2 | 118 | 931 | 0.89 (0.70 - 1.12) | 0.34 | 0.88 (0.69 - 1.13) | 0.40 |
| Open Claims | 151,965 | 613,909 | 390.9 | 2389.3 | 5,353 | 44,734 | 0.90 (0.88 - 0.93) | <0.01 | 0.93 (0.75 - 1.14) | 0.47 |
| OptumEHR | 6,867 | 40,661 | 14.3 | 113.7 | 178 | 1,977 | 0.91 (0.76 - 1.09) | 0.31 | 0.95 (0.79 - 1.14) | 0.60 |
| VA | 5,037 | 32,331 | 6.5 | 75.9 | 43 | 494 | 1.12 (0.74 - 1.64) | 0.58 | 1.18 (0.80 - 1.76) | 0.41 |
| GermanyDA | 3,829 | 831 | 8.0 | 2.8 | 65 | 18 | 1.59 (0.88 - 2.98) | 0.14 | 1.42 (0.77 - 2.61) | 0.29 |
| SIDIAP | 3,184 | 931 | 5.9 | 2.8 | 33 | 23 | 0.58 (0.28 - 1.18) | 0.14 | 0.59 (0.28 - 1.22) | 0.19 |
| Meta-analysis | 171,403 | 711,281 | 425.4 | 2633.1 | 6,103 | 51,273 | 0.91 (0.88 - 0.93) | <0.01 | 0.92 (0.76 - 1.13) | 0.44 |

#### 6.6 DPP4I vs SU

##### 6.6.1 Abnormal weight gain

**Supplementary Table 198:** Exposures, outcomes and risk estimates for Abnormal weight gain in DPP4I (T) vs SU (C) new-user cohorts under four different propensity score (PS) adjustment / time-at-risk (TAR) designs across data sources. We report total patient exposure time in 1,000 patient-year (KYr) units, uncalibrated and calibrated hazard ratios (HRs) and their 95% confidence intervals (CIs). Greyed out entries did not pass study diagnostics and are included only for completeness.

| | Patients | | Exposure (KYr) | | Outcomes | | Uncalibrated | | Calibrated | |
|---|---|---|---|---|---|---|---|---|---|---|
| | T | C | T | C | T | C | HR (95% CI) | $p$ | HR (95% CI) | $p$ |
| **PS-stratification / on-treatment TAR** | | | | | | | | | | |
| CCAE | 2,023 | 3,289 | 0.6 | 0.9 | <5 | <5 | 2.63 (0.43 - 20.3) | 0.33 | 2.94 (0.43 - 20.2) | 0.28 |
| OptumDOD | 26,503 | 64,803 | 22.0 | 67.3 | 122 | 412 | 0.80 (0.65 - 0.99) | 0.04 | 0.81 (0.65 - 1.00) | 0.06 |
| MDCR | 17,725 | 33,412 | 17.7 | 32.8 | 47 | 121 | 0.64 (0.44 - 0.90) | 0.01 | 0.64 (0.45 - 0.91) | 0.01 |
| MDCD | 1,168 | 2,416 | 0.8 | 1.7 | <5 | 11 | 0.53 (0.14 - 1.66) | 0.32 | 0.47 (0.14 - 1.60) | 0.23 |
| Open Claims | 334,949 | 690,159 | 310.5 | 796.5 | 550 | 1,657 | 0.75 (0.68 - 0.83) | <0.01 | 0.75 (0.64 - 0.87) | <0.01 |
| OptumEHR | 22,799 | 57,257 | 11.8 | 34.9 | 55 | 218 | 0.79 (0.58 - 1.08) | 0.15 | 0.84 (0.61 - 1.16) | 0.31 |
| VA | 15,017 | 108,943 | 15.5 | 122.7 | <10 | 186 | 0.28 (0.12 - 0.56) | <0.01 | 0.28 (0.13 - 0.61) | <0.01 |
| GermanyDA | 12,548 | 2,982 | 14.0 | 3.3 | 13 | 5 | 0.67 (0.20 - 2.49) | 0.53 | 0.78 (0.22 - 2.74) | 0.70 |
| SIDIAP | 26,193 | 8,059 | 70.1 | 22.4 | 107 | 39 | 0.99 (0.65 - 1.51) | 0.95 | 0.98 (0.63 - 1.53) | 0.90 |
| Meta-analysis | 428,169 | 853,690 | 432.1 | 953.9 | 881 | 2,447 | 0.76 (0.70 - 0.83) | <0.01 | 0.76 (0.68 - 0.86) | <0.01 |
| **PS-matching / on-treatment TAR** | | | | | | | | | | |
| CCAE | 1,842 | 3,216 | 0.5 | 0.9 | <5 | <5 | 2.56 (0.24 - 56.1) | 0.50 | 2.60 (0.17 - 39.9) | 0.49 |
| OptumDOD | 25,675 | 64,733 | 21.2 | 67.2 | 118 | 411 | 0.91 (0.69 - 1.19) | 0.49 | 0.93 (0.71 - 1.21) | 0.60 |
| MDCR | 16,554 | 32,528 | 16.5 | 31.9 | 40 | 120 | 0.48 (0.28 - 0.78) | <0.01 | 0.46 (0.27 - 0.77) | <0.01 |
| MDCD | 1,105 | 2,399 | 0.8 | 1.6 | <5 | 11 | | | | |
| Open Claims | 328,595 | 690,156 | 302.6 | 796.5 | 520 | 1,657 | 0.62 (0.54 - 0.71) | <0.01 | 0.63 (0.53 - 0.73) | <0.01 |
| OptumEHR | 21,633 | 57,215 | 11.0 | 34.8 | 51 | 218 | 0.74 (0.48 - 1.11) | 0.16 | 0.76 (0.50 - 1.16) | 0.21 |
| VA | 13,986 | 78,621 | 14.8 | 84.1 | <10 | 134 | 0.18 (0.04 - 0.52) | <0.01 | 0.18 (0.05 - 0.64) | <0.01 |
| GermanyDA | 12,512 | 2,552 | 14.0 | 2.8 | 13 | <5 | 0.81 (0.16 - 6.26) | 0.83 | 0.84 (0.13 - 5.27) | 0.86 |
| SIDIAP | 26,068 | 6,113 | 70.0 | 16.1 | 107 | 31 | 0.87 (0.52 - 1.51) | 0.62 | 0.93 (0.50 - 1.72) | 0.82 |
| Meta-analysis | 432,511 | 929,366 | 436.1 | 1030.7 | <846 | 2,571 | 0.68 (0.54 - 0.85) | <0.01 | 0.68 (0.53 - 0.87) | <0.01 |
| **PS-stratification / intent-to-treat TAR** | | | | | | | | | | |
| CCAE | 2,023 | 3,289 | 0.7 | 1.1 | 5 | 5 | 2.00 (0.53 - 7.54) | 0.31 | 2.11 (0.56 - 7.96) | 0.28 |
| OptumDOD | 26,503 | 64,803 | 79.6 | 209.9 | 396 | 1,067 | 0.92 (0.81 - 1.03) | 0.15 | 0.92 (0.81 - 1.04) | 0.19 |
| MDCR | 17,725 | 33,412 | 50.5 | 98.7 | 178 | 387 | 0.84 (0.70 - 1.01) | 0.07 | 0.85 (0.70 - 1.03) | 0.10 |
| MDCD | 1,168 | 2,416 | 3.4 | 8.4 | 21 | 55 | 0.81 (0.46 - 1.35) | 0.43 | 0.71 (0.42 - 1.22) | 0.22 |
| Open Claims | 334,949 | 690,159 | 1424.5 | 2829.8 | 2,786 | 5,717 | 0.93 (0.89 - 0.97) | <0.01 | 0.93 (0.84 - 1.03) | 0.16 |
| OptumEHR | 22,799 | 57,257 | 77.9 | 216.9 | 281 | 892 | 0.88 (0.76 - 1.01) | 0.08 | 0.92 (0.79 - 1.06) | 0.25 |
| VA | 15,017 | 108,943 | 36.7 | 588.1 | 36 | 745 | 0.76 (0.52 - 1.10) | 0.16 | 0.78 (0.53 - 1.14) | 0.21 |
| GermanyDA | 12,548 | 2,982 | 51.6 | 17.5 | 47 | 12 | 1.31 (0.66 - 2.78) | 0.46 | 1.36 (0.66 - 2.79) | 0.41 |
| SIDIAP | 26,193 | 8,059 | 105.6 | 58.3 | 159 | 106 | 0.93 (0.70 - 1.23) | 0.60 | 0.90 (0.68 - 1.20) | 0.50 |
| Meta-analysis | 441,885 | 859,088 | 1793.1 | 3439.5 | 3,868 | 8,236 | 0.92 (0.88 - 0.96) | <0.01 | 0.92 (0.85 - 1.00) | 0.05 |
| **PS-matching / intent-to-treat TAR** | | | | | | | | | | |
| CCAE | 1,842 | 3,216 | 0.6 | 1.1 | 5 | 5 | 1.58 (0.28 - 8.93) | 0.61 | 1.57 (0.28 - 8.86) | 0.61 |
| OptumDOD | 25,675 | 64,733 | 77.3 | 209.6 | 387 | 1,065 | 0.92 (0.79 - 1.06) | 0.26 | 0.92 (0.79 - 1.07) | 0.31 |
| MDCR | 16,554 | 32,528 | 47.4 | 96.2 | 161 | 382 | 0.79 (0.63 - 1.00) | 0.05 | 0.79 (0.62 - 1.00) | 0.05 |
| MDCD | 1,105 | 2,399 | 3.3 | 8.4 | 18 | 55 | 0.65 (0.32 - 1.27) | 0.23 | 0.57 (0.28 - 1.14) | 0.12 |
| Open Claims | 328,595 | 690,156 | 1395.5 | 2829.8 | 2,714 | 5,717 | 0.92 (0.87 - 0.97) | <0.01 | 0.92 (0.83 - 1.01) | 0.09 |
| OptumEHR | 21,633 | 57,215 | 74.3 | 216.6 | 263 | 891 | 0.85 (0.72 - 1.00) | 0.05 | 0.89 (0.75 - 1.05) | 0.17 |
| VA | 13,986 | 78,621 | 35.5 | 351.3 | 35 | 439 | 0.73 (0.47 - 1.10) | 0.15 | 0.75 (0.49 - 1.15) | 0.19 |
| GermanyDA | 12,512 | 2,552 | 51.6 | 14.0 | 47 | 8 | 1.55 (0.71 - 3.75) | 0.31 | 1.56 (0.68 - 3.59) | 0.30 |
| SIDIAP | 26,068 | 6,113 | 105.5 | 40.8 | 159 | 71 | 0.98 (0.71 - 1.36) | 0.90 | 0.99 (0.72 - 1.37) | 0.91 |
| Meta-analysis | 446,128 | 934,317 | 1790.3 | 3766.6 | 3,784 | 8,628 | 0.90 (0.86 - 0.95) | <0.01 | 0.91 (0.83 - 0.99) | 0.03 |

##### 6.6.2 Abnormal weight loss

**Supplementary Table 199:** Exposures, outcomes and risk estimates for Abnormal weight loss in DPP4I (T) vs SU (C) new-user cohorts under four different propensity score (PS) adjustment / time-at-risk (TAR) designs across data sources. We report total patient exposure time in 1,000 patient-year (KYr) units, uncalibrated and calibrated hazard ratios (HRs) and their 95% confidence intervals (CIs). Greyed out entries did not pass study diagnostics and are included only for completeness.

| | Patients | | Exposure (KYr) | | Outcomes | | Uncalibrated | | Calibrated | |
|---|---|---|---|---|---|---|---|---|---|---|
| | T | C | T | C | T | C | HR (95% CI) | $p$ | HR (95% CI) | $p$ |
| **PS-stratification / on-treatment TAR** | | | | | | | | | | |
| CCAE | 2,031 | 3,286 | 0.6 | 0.9 | 9 | 13 | 0.95 (0.37 - 2.35) | 0.92 | 1.06 (0.42 - 2.68) | 0.86 |
| OptumDOD | 25,478 | 62,249 | 21.1 | 64.7 | 451 | 1,144 | 1.13 (1.01 - 1.27) | 0.03 | 1.14 (1.01 - 1.29) | 0.05 |
| MDCR | 17,148 | 32,177 | 17.1 | 31.6 | 251 | 407 | 1.09 (0.93 - 1.29) | 0.29 | 1.10 (0.93 - 1.30) | 0.32 |
| MDCD | 1,115 | 2,298 | 0.8 | 1.6 | 19 | 43 | 0.90 (0.49 - 1.57) | 0.71 | 0.79 (0.44 - 1.40) | 0.44 |
| Open Claims | 326,168 | 673,081 | 301.3 | 777.5 | 2,885 | 5,608 | 1.25 (1.19 - 1.31) | <0.01 | 1.24 (1.09 - 1.41) | <0.01 |
| OptumEHR | 22,396 | 56,014 | 11.5 | 34.0 | 206 | 536 | 1.18 (0.99 - 1.40) | 0.06 | 1.25 (1.05 - 1.49) | 0.02 |
| VA | 14,599 | 105,915 | 15.1 | 119.5 | 168 | 871 | 1.34 (1.09 - 1.63) | <0.01 | 1.36 (1.11 - 1.67) | <0.01 |
| GermanyDA | 12,258 | 2,910 | 13.6 | 3.2 | 110 | 24 | 1.01 (0.63 - 1.68) | 0.98 | 1.18 (0.71 - 1.94) | 0.57 |
| SIDIAP | 26,053 | 8,065 | 69.6 | 22.4 | 313 | 66 | 0.87 (0.65 - 1.19) | 0.38 | 0.87 (0.62 - 1.22) | 0.45 |
| Meta-analysis | 432,647 | 840,080 | 435.5 | 935.8 | 4,244 | 7,841 | 1.14 (1.05 - 1.24) | <0.01 | 1.15 (1.02 - 1.30) | 0.03 |
| **PS-matching / on-treatment TAR** | | | | | | | | | | |
| CCAE | 1,848 | 3,215 | 0.5 | 0.9 | 7 | 13 | 0.78 (0.26 - 2.11) | 0.64 | 0.79 (0.28 - 2.25) | 0.68 |
| OptumDOD | 24,685 | 62,182 | 20.3 | 64.6 | 430 | 1,144 | 1.07 (0.92 - 1.25) | 0.39 | 1.09 (0.93 - 1.27) | 0.32 |
| MDCR | 15,980 | 31,323 | 15.8 | 30.7 | 239 | 404 | 1.10 (0.88 - 1.38) | 0.39 | 1.06 (0.85 - 1.33) | 0.64 |
| MDCD | 1,066 | 2,280 | 0.7 | 1.5 | 19 | 43 | 0.53 (0.20 - 1.23) | 0.16 | 0.50 (0.20 - 1.22) | 0.14 |
| Open Claims | 319,989 | 673,077 | 293.8 | 777.5 | 2,782 | 5,608 | 1.19 (1.12 - 1.27) | <0.01 | 1.20 (1.08 - 1.34) | <0.01 |
| OptumEHR | 21,223 | 55,973 | 10.8 | 33.9 | 191 | 535 | 1.23 (0.98 - 1.53) | 0.07 | 1.27 (1.01 - 1.59) | 0.05 |
| VA | 13,594 | 76,432 | 14.4 | 81.9 | 161 | 609 | 1.22 (0.94 - 1.58) | 0.13 | 1.23 (0.95 - 1.60) | 0.14 |
| GermanyDA | 12,223 | 2,498 | 13.5 | 2.7 | 110 | 21 | 1.83 (0.88 - 4.28) | 0.13 | 1.88 (0.85 - 4.16) | 0.13 |
| SIDIAP | 25,925 | 6,116 | 69.5 | 16.1 | 313 | 53 | 0.84 (0.57 - 1.26) | 0.39 | 0.89 (0.54 - 1.48) | 0.68 |
| Meta-analysis | 436,533 | 913,096 | 439.4 | 1009.8 | 4,252 | 8,430 | 1.16 (1.09 - 1.23) | <0.01 | 1.16 (1.05 - 1.29) | <0.01 |
| **PS-stratification / intent-to-treat TAR** | | | | | | | | | | |
| CCAE | 2,031 | 3,286 | 0.7 | 1.1 | 11 | 14 | 1.16 (0.49 - 2.71) | 0.73 | 1.22 (0.52 - 2.89) | 0.65 |
| OptumDOD | 25,478 | 62,249 | 74.4 | 195.9 | 1,658 | 4,099 | 1.06 (0.99 - 1.12) | 0.08 | 1.06 (0.99 - 1.13) | 0.18 |
| MDCR | 17,148 | 32,177 | 47.9 | 93.2 | 875 | 1,639 | 1.02 (0.94 - 1.12) | 0.59 | 1.03 (0.93 - 1.14) | 0.60 |
| MDCD | 1,115 | 2,298 | 3.1 | 7.8 | 99 | 197 | 1.14 (0.88 - 1.46) | 0.33 | 1.00 (0.77 - 1.30) | 0.87 |
| Open Claims | 326,168 | 673,081 | 1366.1 | 2722.1 | 14,188 | 25,901 | 1.07 (1.05 - 1.09) | <0.01 | 1.07 (0.97 - 1.17) | 0.17 |
| OptumEHR | 22,396 | 56,014 | 74.9 | 207.4 | 1,169 | 3,016 | 1.10 (1.02 - 1.18) | 0.01 | 1.14 (1.06 - 1.24) | <0.01 |
| VA | 14,599 | 105,915 | 35.4 | 562.8 | 419 | 6,434 | 0.97 (0.87 - 1.09) | 0.64 | 1.00 (0.89 - 1.12) | 0.88 |
| GermanyDA | 12,258 | 2,910 | 49.8 | 16.7 | 402 | 134 | 1.13 (0.91 - 1.41) | 0.28 | 1.17 (0.94 - 1.45) | 0.20 |
| SIDIAP | 26,053 | 8,065 | 104.7 | 58.1 | 480 | 205 | 0.94 (0.78 - 1.15) | 0.56 | 0.92 (0.76 - 1.12) | 0.43 |
| Meta-analysis | 432,647 | 840,080 | 1721.6 | 3302.4 | 18,882 | 35,205 | 1.07 (1.05 - 1.09) | <0.01 | 1.07 (0.99 - 1.15) | 0.09 |
| **PS-matching / intent-to-treat TAR** | | | | | | | | | | |
| CCAE | 1,848 | 3,215 | 0.6 | 1.1 | 9 | 14 | 1.12 (0.42 - 2.83) | 0.82 | 1.11 (0.43 - 2.88) | 0.81 |
| OptumDOD | 24,685 | 62,182 | 72.3 | 195.7 | 1,586 | 4,092 | 1.03 (0.96 - 1.11) | 0.39 | 1.04 (0.95 - 1.13) | 0.44 |
| MDCR | 15,980 | 31,323 | 44.8 | 90.8 | 817 | 1,618 | 1.04 (0.93 - 1.17) | 0.46 | 1.03 (0.92 - 1.16) | 0.63 |
| MDCD | 1,066 | 2,280 | 3.0 | 7.7 | 97 | 196 | 0.98 (0.70 - 1.34) | 0.88 | 0.85 (0.61 - 1.17) | 0.34 |
| Open Claims | 319,989 | 673,077 | 1338.5 | 2722.1 | 13,781 | 25,901 | 1.08 (1.05 - 1.11) | <0.01 | 1.08 (0.99 - 1.18) | 0.10 |
| OptumEHR | 21,223 | 55,973 | 71.3 | 207.1 | 1,108 | 3,012 | 1.08 (0.99 - 1.18) | 0.08 | 1.13 (1.03 - 1.24) | 0.02 |
| VA | 13,594 | 76,432 | 34.2 | 337.3 | 400 | 3,844 | 1.01 (0.88 - 1.15) | 0.92 | 1.03 (0.90 - 1.18) | 0.67 |
| GermanyDA | 12,223 | 2,498 | 49.7 | 13.4 | 402 | 108 | 1.12 (0.86 - 1.45) | 0.41 | 1.13 (0.87 - 1.46) | 0.43 |
| SIDIAP | 25,925 | 6,116 | 104.6 | 40.6 | 480 | 145 | 0.97 (0.78 - 1.23) | 0.83 | 0.99 (0.78 - 1.24) | 0.87 |
| Meta-analysis | 436,533 | 913,096 | 1719.2 | 3615.8 | 18,680 | 38,930 | 1.07 (1.04 - 1.10) | <0.01 | 1.07 (0.99 - 1.16) | 0.08 |

##### 6.6.3 Acute pancreatitis

**Supplementary Table 200:** Exposures, outcomes and risk estimates for Acute pancreatitis in DPP4I (T) vs SU (C) new-user cohorts under four different propensity score (PS) adjustment / time-at-risk (TAR) designs across data sources. We report total patient exposure time in 1,000 patient-year (KYr) units, uncalibrated and calibrated hazard ratios (HRs) and their 95% confidence intervals (CIs). Greyed out entries did not pass study diagnostics and are included only for completeness.

| | Patients | | Exposure (KYr) | | Outcomes | | Uncalibrated | | Calibrated | |
|---|---|---|---|---|---|---|---|---|---|---|
| | T | C | T | C | T | C | HR (95% CI) | $p$ | HR (95% CI) | $p$ |
| **PS-stratification / on-treatment TAR** | | | | | | | | | | |
| CCAE | 2,074 | 3,359 | 0.6 | 0.9 | <5 | <5 | 0.78 (0.11 - 4.11) | 0.79 | 0.87 (0.14 - 5.43) | 0.88 |
| OptumDOD | 26,990 | 65,495 | 22.5 | 68.3 | 78 | 262 | 0.87 (0.66 - 1.13) | 0.31 | 0.88 (0.67 - 1.15) | 0.35 |
| MDCR | 17,814 | 33,321 | 17.8 | 32.7 | 55 | 123 | 0.93 (0.66 - 1.29) | 0.65 | 0.93 (0.66 - 1.30) | 0.67 |
| MDCD | 1,190 | 2,425 | 0.8 | 1.7 | 6 | 8 | 1.47 (0.46 - 4.50) | 0.51 | 1.28 (0.41 - 4.04) | 0.67 |
| Open Claims | 340,386 | 696,991 | 315.7 | 804.4 | 738 | 1,772 | 1.02 (0.94 - 1.12) | 0.59 | 1.02 (0.88 - 1.18) | 0.80 |
| OptumEHR | 23,253 | 58,047 | 12.1 | 35.5 | 26 | 105 | 0.83 (0.52 - 1.30) | 0.44 | 0.88 (0.56 - 1.40) | 0.61 |
| VA | 15,177 | 109,510 | 15.7 | 123.5 | 21 | 189 | 1.10 (0.64 - 1.83) | 0.72 | 1.12 (0.66 - 1.91) | 0.67 |
| GermanyDA | 12,634 | 2,999 | 14.1 | 3.3 | 0 | 0 | | | | |
| SIDIAP | 26,418 | 8,085 | 70.7 | 22.6 | 29 | 6 | 1.19 (0.45 - 3.57) | 0.75 | 1.18 (0.42 - 3.35) | 0.76 |
| Meta-analysis | 434,861 | 861,939 | 438.8 | 963.6 | 926 | 2,268 | 0.99 (0.89 - 1.09) | 0.80 | 0.99 (0.87 - 1.13) | 0.88 |
| **PS-matching / on-treatment TAR** | | | | | | | | | | |
| CCAE | 1,880 | 3,286 | 0.5 | 0.9 | <5 | <5 | 0.46 (0.02 - 3.67) | 0.55 | 0.46 (0.04 - 5.97) | 0.56 |
| OptumDOD | 26,101 | 65,429 | 21.7 | 68.2 | 74 | 261 | 0.96 (0.68 - 1.35) | 0.84 | 0.98 (0.70 - 1.39) | 0.92 |
| MDCR | 16,586 | 32,348 | 16.5 | 31.8 | 51 | 118 | 0.67 (0.40 - 1.07) | 0.10 | 0.64 (0.39 - 1.04) | 0.08 |
| MDCD | 1,122 | 2,406 | 0.8 | 1.7 | 5 | 7 | 1.23 (0.24 - 5.35) | 0.79 | 1.16 (0.25 - 5.47) | 0.85 |
| Open Claims | 333,698 | 696,989 | 307.4 | 804.4 | 708 | 1,772 | 1.04 (0.92 - 1.17) | 0.54 | 1.05 (0.91 - 1.22) | 0.53 |
| OptumEHR | 22,038 | 58,004 | 11.3 | 35.5 | 25 | 105 | 1.28 (0.75 - 2.14) | 0.36 | 1.32 (0.78 - 2.23) | 0.31 |
| VA | 14,102 | 79,293 | 14.9 | 85.0 | 20 | 131 | 1.17 (0.60 - 2.15) | 0.64 | 1.18 (0.62 - 2.24) | 0.62 |
| GermanyDA | 12,597 | 2,566 | 14.0 | 2.8 | 0 | 0 | | | | |
| SIDIAP | 26,291 | 6,137 | 70.6 | 16.2 | 29 | 6 | 0.95 (0.34 - 2.90) | 0.93 | 1.01 (0.33 - 3.08) | 0.95 |
| Meta-analysis | 438,816 | 938,200 | 442.5 | 1041.1 | 907 | 2,393 | 1.02 (0.92 - 1.14) | 0.71 | 1.02 (0.89 - 1.17) | 0.75 |
| **PS-stratification / intent-to-treat TAR** | | | | | | | | | | |
| CCAE | 2,074 | 3,359 | 0.7 | 1.2 | <5 | <5 | 0.82 (0.11 - 4.28) | 0.83 | 0.86 (0.14 - 5.33) | 0.87 |
| OptumDOD | 26,990 | 65,495 | 81.3 | 212.8 | 293 | 796 | 0.97 (0.84 - 1.11) | 0.67 | 0.97 (0.84 - 1.12) | 0.70 |
| MDCR | 17,814 | 33,321 | 50.6 | 98.3 | 174 | 418 | 0.87 (0.72 - 1.04) | 0.14 | 0.87 (0.72 - 1.06) | 0.18 |
| MDCD | 1,190 | 2,425 | 3.5 | 8.5 | 18 | 36 | 1.59 (0.85 - 2.86) | 0.14 | 1.40 (0.76 - 2.57) | 0.29 |
| Open Claims | 340,386 | 696,991 | 1448.0 | 2856.5 | 3,203 | 6,508 | 0.97 (0.93 - 1.02) | 0.25 | 0.97 (0.88 - 1.08) | 0.61 |
| OptumEHR | 23,253 | 58,047 | 79.9 | 221.1 | 120 | 421 | 0.85 (0.68 - 1.05) | 0.13 | 0.88 (0.71 - 1.10) | 0.26 |
| VA | 15,177 | 109,510 | 37.1 | 591.4 | 42 | 847 | 0.96 (0.67 - 1.34) | 0.81 | 0.98 (0.69 - 1.39) | 0.92 |
| GermanyDA | 12,634 | 2,999 | 52.2 | 17.7 | 0 | 0 | | | | |
| SIDIAP | 26,418 | 8,085 | 106.5 | 58.8 | 45 | 16 | 1.38 (0.70 - 2.79) | 0.36 | 1.34 (0.67 - 2.68) | 0.41 |
| Meta-analysis | 436,051 | 864,364 | 1769.8 | 3455.9 | 3,853 | 8,195 | 0.97 (0.93 - 1.01) | 0.10 | 0.97 (0.89 - 1.05) | 0.44 |
| **PS-matching / intent-to-treat TAR** | | | | | | | | | | |
| CCAE | 1,880 | 3,286 | 0.6 | 1.1 | <5 | <5 | 0.46 (0.02 - 3.67) | 0.55 | 0.45 (0.04 - 5.84) | 0.55 |
| OptumDOD | 26,101 | 65,429 | 78.9 | 212.6 | 283 | 794 | 1.01 (0.85 - 1.19) | 0.92 | 1.01 (0.85 - 1.20) | 0.89 |
| MDCR | 16,586 | 32,348 | 47.3 | 95.5 | 165 | 405 | 1.01 (0.79 - 1.27) | 0.96 | 1.00 (0.78 - 1.27) | 0.94 |
| MDCD | 1,122 | 2,406 | 3.4 | 8.4 | 17 | 34 | 1.14 (0.53 - 2.31) | 0.73 | 0.99 (0.47 - 2.05) | 0.93 |
| Open Claims | 333,698 | 696,989 | 1417.6 | 2856.5 | 3,124 | 6,508 | 0.98 (0.93 - 1.03) | 0.34 | 0.98 (0.88 - 1.08) | 0.63 |
| OptumEHR | 22,038 | 58,004 | 76.2 | 220.8 | 114 | 421 | 0.91 (0.71 - 1.15) | 0.44 | 0.95 (0.74 - 1.22) | 0.70 |
| VA | 14,102 | 79,293 | 35.8 | 355.3 | 41 | 508 | 1.05 (0.72 - 1.50) | 0.78 | 1.08 (0.75 - 1.55) | 0.68 |
| GermanyDA | 12,597 | 2,566 | 52.1 | 14.1 | 0 | 0 | | | | |
| SIDIAP | 26,291 | 6,137 | 106.4 | 41.1 | 45 | 14 | 1.03 (0.52 - 2.13) | 0.94 | 1.04 (0.51 - 2.11) | 0.92 |
| Meta-analysis | 439,938 | 940,606 | 1765.4 | 3790.1 | 3,789 | 8,684 | 0.98 (0.94 - 1.02) | 0.36 | 0.98 (0.90 - 1.07) | 0.65 |

##### 6.6.4 All-cause mortality

**Supplementary Table 201:** Exposures, outcomes and risk estimates for All-cause mortality in DPP4I (T) vs SU (C) new-user cohorts under four different propensity score (PS) adjustment / time-at-risk (TAR) designs across data sources. We report total patient exposure time in 1,000 patient-year (KYr) units, uncalibrated and calibrated hazard ratios (HRs) and their 95% confidence intervals (CIs). Greyed out entries did not pass study diagnostics and are included only for completeness.

| | Patients | | Exposure (KYr) | | Outcomes | | Uncalibrated | | Calibrated | |
|---|---|---|---|---|---|---|---|---|---|---|
| | T | C | T | C | T | C | HR (95% CI) | $p$ | HR (95% CI) | $p$ |
| **PS-stratification / on-treatment TAR** | | | | | | | | | | |
| CCAE | 2,089 | 3,385 | 0.6 | 0.9 | <5 | <5 | 1.58 (0.06 - 19.8) | 0.76 | 1.76 (0.10 - 31.1) | 0.70 |
| OptumDOD | 27,235 | 66,291 | 22.7 | 69.2 | 276 | 1,085 | 0.84 (0.73 - 0.96) | 0.01 | 0.84 (0.73 - 0.97) | 0.03 |
| MDCR | 17,961 | 33,739 | 18.0 | 33.2 | 50 | 128 | 0.97 (0.68 - 1.36) | 0.87 | 0.97 (0.69 - 1.37) | 0.89 |
| MDCD | 1,210 | 2,481 | 0.8 | 1.7 | 15 | 13 | 2.56 (1.16 - 5.73) | 0.02 | 2.24 (1.01 - 4.99) | 0.05 |
| Open Claims | 344,611 | 707,599 | 319.8 | 817.2 | <5 | 13 | 0.34 (0.05 - 1.26) | 0.18 | 0.34 (0.07 - 1.67) | 0.18 |
| OptumEHR | 23,345 | 58,379 | 12.1 | 35.8 | 172 | 604 | 1.02 (0.85 - 1.21) | 0.85 | 1.08 (0.90 - 1.29) | 0.46 |
| VA | 15,312 | 110,592 | 15.8 | 124.8 | 245 | 2,318 | 0.81 (0.70 - 0.95) | <0.01 | 0.83 (0.71 - 0.97) | 0.02 |
| GermanyDA | 12,634 | 2,999 | 14.1 | 3.3 | 11 | <5 | 2.82 (0.43 - 56.6) | 0.40 | 3.29 (0.29 - 37.7) | 0.34 |
| SIDIAP | 26,465 | 8,098 | 70.9 | 22.6 | 2,104 | 429 | 1.15 (1.02 - 1.30) | 0.02 | 1.15 (0.95 - 1.39) | 0.25 |
| Meta-analysis | 98,292 | 172,101 | 124.6 | 162.6 | 2,622 | 2,285 | 1.02 (0.86 - 1.20) | 0.82 | 1.02 (0.85 - 1.24) | 0.82 |
| **PS-matching / on-treatment TAR** | | | | | | | | | | |
| CCAE | 1,896 | 3,312 | 0.5 | 0.9 | <5 | <5 | 1.35 (0.06 - 16.2) | 0.84 | 1.37 (0.08 - 23.3) | 0.83 |
| OptumDOD | 26,347 | 66,220 | 21.9 | 69.2 | 262 | 1,082 | 0.82 (0.68 - 0.99) | 0.04 | 0.84 (0.69 - 1.01) | 0.08 |
| MDCR | 16,748 | 32,853 | 16.7 | 32.3 | 49 | 124 | 1.26 (0.80 - 1.96) | 0.32 | 1.21 (0.77 - 1.89) | 0.42 |
| MDCD | 1,140 | 2,462 | 0.8 | 1.7 | 15 | 13 | 2.64 (0.74 - 9.51) | 0.14 | 2.49 (0.70 - 8.93) | 0.16 |
| Open Claims | 337,948 | 707,595 | 311.5 | 817.2 | <5 | 13 | 0.62 (0.08 - 3.23) | 0.60 | 0.63 (0.10 - 3.87) | 0.61 |
| OptumEHR | 22,131 | 58,336 | 11.4 | 35.7 | 169 | 604 | 1.00 (0.79 - 1.26) | 0.99 | 1.03 (0.82 - 1.30) | 0.79 |
| VA | 14,229 | 79,981 | 15.0 | 85.7 | 233 | 1,565 | 0.86 (0.71 - 1.05) | 0.14 | 0.87 (0.71 - 1.06) | 0.21 |
| GermanyDA | 12,597 | 2,566 | 14.0 | 2.8 | 11 | 0 | | | | |
| SIDIAP | 26,339 | 6,148 | 70.8 | 16.3 | 2,097 | 306 | 1.22 (1.04 - 1.44) | 0.02 | 1.30 (0.91 - 1.84) | 0.18 |
| Meta-analysis | 108,303 | 249,089 | 136.5 | 241.0 | 2,830 | 3,720 | 1.01 (0.85 - 1.20) | 0.89 | 1.01 (0.84 - 1.23) | 0.89 |
| **PS-stratification / intent-to-treat TAR** | | | | | | | | | | |
| CCAE | 2,089 | 3,385 | 0.7 | 1.2 | <5 | 7 | 1.15 (0.29 - 4.04) | 0.83 | 1.22 (0.33 - 4.53) | 0.77 |
| OptumDOD | 27,235 | 66,291 | 81.3 | 212.1 | 3,230 | 10,047 | 0.93 (0.89 - 0.96) | <0.01 | 0.93 (0.88 - 0.98) | 0.02 |
| MDCR | 17,961 | 33,739 | 51.4 | 100.2 | 246 | 698 | 0.94 (0.81 - 1.09) | 0.42 | 0.94 (0.80 - 1.11) | 0.50 |
| MDCD | 1,210 | 2,481 | 3.6 | 8.8 | 61 | 145 | 1.10 (0.80 - 1.50) | 0.55 | 0.97 (0.71 - 1.33) | 0.85 |
| Open Claims | 344,611 | 707,599 | 1473.4 | 2914.2 | 23 | 66 | 0.77 (0.46 - 1.23) | 0.29 | 0.77 (0.47 - 1.26) | 0.29 |
| OptumEHR | 23,345 | 58,379 | 80.3 | 222.2 | 1,042 | 3,691 | 0.90 (0.84 - 0.97) | <0.01 | 0.94 (0.87 - 1.01) | 0.15 |
| VA | 15,312 | 110,592 | 37.4 | 599.0 | 1,075 | 22,478 | 0.92 (0.86 - 0.99) | 0.02 | 0.94 (0.87 - 1.02) | 0.18 |
| GermanyDA | 12,634 | 2,999 | 52.2 | 17.7 | 84 | 36 | 0.86 (0.55 - 1.35) | 0.50 | 0.89 (0.57 - 1.39) | 0.60 |
| SIDIAP | 26,465 | 8,098 | 106.7 | 58.9 | 5,122 | 2,863 | 0.92 (0.87 - 0.97) | <0.01 | 0.90 (0.85 - 0.96) | <0.01 |
| Meta-analysis | 455,537 | 882,699 | 1856.6 | 3547.0 | 10,120 | 18,290 | 0.92 (0.89 - 0.94) | <0.01 | 0.92 (0.85 - 0.99) | 0.03 |
| **PS-matching / intent-to-treat TAR** | | | | | | | | | | |
| CCAE | 1,896 | 3,312 | 0.6 | 1.1 | <5 | 7 | 1.01 (0.20 - 4.46) | 0.99 | 1.01 (0.21 - 4.79) | 0.92 |
| OptumDOD | 26,347 | 66,220 | 78.8 | 211.9 | 3,144 | 10,005 | 0.93 (0.88 - 0.98) | <0.01 | 0.93 (0.87 - 0.99) | 0.06 |
| MDCR | 16,748 | 32,853 | 48.2 | 97.6 | 244 | 671 | 0.96 (0.79 - 1.15) | 0.65 | 0.95 (0.78 - 1.15) | 0.59 |
| MDCD | 1,140 | 2,462 | 3.4 | 8.7 | 58 | 141 | 0.89 (0.60 - 1.31) | 0.57 | 0.77 (0.52 - 1.15) | 0.22 |
| Open Claims | 337,948 | 707,595 | 1443.0 | 2914.2 | 23 | 66 | 0.81 (0.48 - 1.34) | 0.43 | 0.81 (0.49 - 1.36) | 0.44 |
| OptumEHR | 22,131 | 58,336 | 76.5 | 221.9 | 1,005 | 3,688 | 0.91 (0.83 - 0.99) | 0.03 | 0.95 (0.87 - 1.05) | 0.36 |
| VA | 14,229 | 79,981 | 36.1 | 358.7 | 1,042 | 12,250 | 0.92 (0.85 - 1.00) | 0.05 | 0.95 (0.87 - 1.03) | 0.28 |
| GermanyDA | 12,597 | 2,566 | 52.1 | 14.1 | 84 | 30 | 0.74 (0.46 - 1.21) | 0.22 | 0.75 (0.46 - 1.22) | 0.26 |
| SIDIAP | 26,339 | 6,148 | 106.6 | 41.2 | 5,110 | 1,972 | 0.92 (0.86 - 0.98) | 0.01 | 0.93 (0.87 - 1.00) | 0.11 |
| Meta-analysis | 458,848 | 959,250 | 1849.9 | 3881.1 | 10,936 | 29,558 | 0.92 (0.89 - 0.95) | <0.01 | 0.92 (0.85 - 1.00) | 0.04 |

##### 6.6.5 Bone fracture

**Supplementary Table 202:** Exposures, outcomes and risk estimates for Bone fracture in DPP4I (T) vs SU (C) new-user cohorts under four different propensity score (PS) adjustment / time-at-risk (TAR) designs across data sources. We report total patient exposure time in 1,000 patient-year (KYr) units, uncalibrated and calibrated hazard ratios (HRs) and their 95% confidence intervals (CIs). Greyed out entries did not pass study diagnostics and are included only for completeness.

| | Patients | | Exposure (KYr) | | Outcomes | | Uncalibrated | | Calibrated | |
|---|---|---|---|---|---|---|---|---|---|---|
| | T | C | T | C | T | C | HR (95% CI) | $p$ | HR (95% CI) | $p$ |
| **PS-stratification / on-treatment TAR** | | | | | | | | | | |
| CCAE | 1,879 | 3,098 | 0.5 | 0.8 | 7 | 21 | 0.53 (0.21 - 1.22) | 0.17 | 0.60 (0.24 - 1.45) | 0.27 |
| OptumDOD | 24,067 | 59,398 | 19.6 | 60.7 | 681 | 2,092 | 0.97 (0.89 - 1.07) | 0.56 | 0.98 (0.89 - 1.08) | 0.74 |
| MDCR | 15,701 | 29,545 | 15.3 | 28.4 | 539 | 1,008 | 0.98 (0.87 - 1.09) | 0.66 | 0.98 (0.87 - 1.10) | 0.74 |
| MDCD | 988 | 2,135 | 0.7 | 1.4 | 24 | 65 | 0.78 (0.47 - 1.26) | 0.33 | 0.69 (0.42 - 1.12) | 0.16 |
| Open Claims | 291,512 | 605,340 | 267.1 | 692.3 | 6,230 | 15,720 | 0.97 (0.94 - 1.00) | 0.05 | 0.97 (0.85 - 1.09) | 0.59 |
| OptumEHR | 21,425 | 53,609 | 10.9 | 31.8 | 289 | 922 | 0.97 (0.84 - 1.12) | 0.67 | 1.03 (0.89 - 1.19) | 0.75 |
| VA | 13,821 | 101,522 | 14.3 | 114.0 | 131 | 1,181 | 1.03 (0.83 - 1.28) | 0.78 | 1.05 (0.85 - 1.31) | 0.66 |
| GermanyDA | 10,829 | 2,636 | 11.8 | 2.9 | 306 | 75 | 0.95 (0.72 - 1.27) | 0.70 | 1.10 (0.82 - 1.49) | 0.59 |
| SIDIAP | 23,069 | 7,470 | 60.9 | 20.3 | 1,146 | 376 | 1.04 (0.91 - 1.19) | 0.58 | 1.04 (0.85 - 1.27) | 0.77 |
| Meta-analysis | 389,470 | 763,231 | 386.8 | 838.6 | 9,222 | 20,279 | 0.97 (0.95 - 1.00) | 0.04 | 0.98 (0.89 - 1.07) | 0.60 |
| **PS-matching / on-treatment TAR** | | | | | | | | | | |
| CCAE | 1,706 | 3,011 | 0.5 | 0.8 | 7 | 21 | 0.65 (0.22 - 1.65) | 0.39 | 0.66 (0.24 - 1.79) | 0.43 |
| OptumDOD | 23,325 | 59,331 | 18.9 | 60.6 | 656 | 2,087 | 0.91 (0.80 - 1.03) | 0.15 | 0.93 (0.82 - 1.06) | 0.31 |
| MDCR | 14,674 | 28,642 | 14.3 | 27.5 | 494 | 991 | 0.94 (0.80 - 1.11) | 0.48 | 0.91 (0.77 - 1.07) | 0.28 |
| MDCD | 945 | 2,119 | 0.6 | 1.4 | 23 | 65 | 0.93 (0.47 - 1.77) | 0.83 | 0.88 (0.45 - 1.70) | 0.71 |
| Open Claims | 285,984 | 605,337 | 260.4 | 692.3 | 6,025 | 15,720 | 0.98 (0.94 - 1.02) | 0.35 | 0.99 (0.90 - 1.09) | 0.84 |
| OptumEHR | 20,324 | 53,563 | 10.2 | 31.8 | 273 | 919 | 1.02 (0.85 - 1.23) | 0.81 | 1.06 (0.88 - 1.27) | 0.61 |
| VA | 12,914 | 73,134 | 13.7 | 78.2 | 127 | 776 | 1.05 (0.80 - 1.37) | 0.70 | 1.06 (0.81 - 1.39) | 0.68 |
| GermanyDA | 10,788 | 2,256 | 11.8 | 2.4 | 305 | 63 | 1.03 (0.72 - 1.51) | 0.87 | 1.06 (0.73 - 1.55) | 0.70 |
| SIDIAP | 22,974 | 5,624 | 60.8 | 14.5 | 1,146 | 266 | 1.05 (0.88 - 1.26) | 0.58 | 1.12 (0.78 - 1.60) | 0.58 |
| Meta-analysis | 393,634 | 833,017 | 391.2 | 909.5 | 9,056 | 20,908 | 0.98 (0.94 - 1.01) | 0.23 | 0.98 (0.89 - 1.07) | 0.65 |
| **PS-stratification / intent-to-treat TAR** | | | | | | | | | | |
| CCAE | 1,879 | 3,098 | 0.6 | 1.1 | 8 | 23 | 0.56 (0.23 - 1.24) | 0.18 | 0.59 (0.25 - 1.37) | 0.23 |
| OptumDOD | 24,067 | 59,398 | 67.7 | 179.0 | 2,472 | 6,546 | 0.98 (0.94 - 1.03) | 0.48 | 0.98 (0.93 - 1.04) | 0.65 |
| MDCR | 15,701 | 29,545 | 42.0 | 81.1 | 1,608 | 3,200 | 0.98 (0.92 - 1.04) | 0.44 | 0.98 (0.91 - 1.06) | 0.68 |
| MDCD | 988 | 2,135 | 2.7 | 6.9 | 111 | 278 | 0.95 (0.75 - 1.20) | 0.69 | 0.84 (0.66 - 1.06) | 0.18 |
| Open Claims | 291,512 | 605,340 | 1183.9 | 2365.9 | 30,711 | 61,885 | 0.97 (0.96 - 0.99) | <0.01 | 0.97 (0.88 - 1.06) | 0.53 |
| OptumEHR | 21,425 | 53,609 | 70.2 | 193.2 | 1,629 | 4,915 | 0.95 (0.90 - 1.01) | 0.12 | 0.99 (0.93 - 1.06) | 0.83 |
| VA | 13,821 | 101,522 | 33.6 | 535.9 | 351 | 6,113 | 1.06 (0.93 - 1.19) | 0.39 | 1.08 (0.95 - 1.23) | 0.26 |
| GermanyDA | 10,829 | 2,636 | 42.6 | 14.2 | 1,049 | 364 | 0.99 (0.87 - 1.13) | 0.91 | 1.03 (0.90 - 1.17) | 0.69 |
| SIDIAP | 23,069 | 7,470 | 90.7 | 50.6 | 1,864 | 1,108 | 0.99 (0.91 - 1.08) | 0.89 | 0.97 (0.89 - 1.06) | 0.57 |
| Meta-analysis | 391,468 | 766,235 | 1507.8 | 2904.2 | 39,495 | 78,405 | 0.97 (0.96 - 0.98) | <0.01 | 0.97 (0.90 - 1.05) | 0.46 |
| **PS-matching / intent-to-treat TAR** | | | | | | | | | | |
| CCAE | 1,706 | 3,011 | 0.6 | 1.0 | 8 | 23 | 0.71 (0.29 - 1.63) | 0.44 | 0.71 (0.29 - 1.69) | 0.45 |
| OptumDOD | 23,325 | 59,331 | 65.8 | 178.8 | 2,396 | 6,528 | 0.98 (0.93 - 1.05) | 0.62 | 0.99 (0.92 - 1.06) | 0.78 |
| MDCR | 14,674 | 28,642 | 39.5 | 78.6 | 1,507 | 3,142 | 0.95 (0.88 - 1.03) | 0.25 | 0.94 (0.86 - 1.03) | 0.25 |
| MDCD | 945 | 2,119 | 2.6 | 6.8 | 106 | 275 | 0.94 (0.70 - 1.26) | 0.68 | 0.82 (0.61 - 1.10) | 0.20 |
| Open Claims | 285,984 | 605,337 | 1160.1 | 2365.8 | 30,014 | 61,885 | 0.97 (0.95 - 0.98) | <0.01 | 0.97 (0.89 - 1.06) | 0.46 |
| OptumEHR | 20,324 | 53,563 | 66.9 | 193.0 | 1,564 | 4,902 | 0.96 (0.89 - 1.03) | 0.21 | 1.00 (0.93 - 1.08) | 0.87 |
| VA | 12,914 | 73,134 | 32.5 | 322.6 | 340 | 3,475 | 1.02 (0.88 - 1.17) | 0.82 | 1.04 (0.90 - 1.20) | 0.60 |
| GermanyDA | 10,788 | 2,256 | 42.5 | 11.5 | 1,046 | 288 | 0.92 (0.79 - 1.09) | 0.34 | 0.93 (0.79 - 1.10) | 0.50 |
| SIDIAP | 22,974 | 5,624 | 90.6 | 35.3 | 1,862 | 752 | 1.02 (0.92 - 1.13) | 0.75 | 1.03 (0.92 - 1.14) | 0.67 |
| Meta-analysis | 394,961 | 835,997 | 1506.1 | 3205.4 | 38,872 | 81,356 | 0.97 (0.95 - 0.98) | <0.01 | 0.97 (0.90 - 1.04) | 0.42 |

##### 6.6.6 Diabetic ketoacidosis

**Supplementary Table 203:** Exposures, outcomes and risk estimates for Diabetic ketoacidosis in DPP4I (T) vs SU (C) new-user cohorts under four different propensity score (PS) adjustment / time-at-risk (TAR) designs across data sources. We report total patient exposure time in 1,000 patient-year (KYr) units, uncalibrated and calibrated hazard ratios (HRs) and their 95% confidence intervals (CIs). Greyed out entries did not pass study diagnostics and are included only for completeness.

| | Patients | | Exposure (KYr) | | Outcomes | | Uncalibrated | | Calibrated | |
|---|---|---|---|---|---|---|---|---|---|---|
| | T | C | T | C | T | C | HR (95% CI) | $p$ | HR (95% CI) | $p$ |
| **PS-stratification / on-treatment TAR** | | | | | | | | | | |
| CCAE | 2,088 | 3,383 | 0.6 | 0.9 | 0 | 0 | | | | |
| OptumDOD | 27,197 | 66,189 | 22.7 | 69.1 | 37 | 111 | 1.02 (0.68 - 1.49) | 0.93 | 1.03 (0.69 - 1.52) | 0.90 |
| MDCR | 17,928 | 33,703 | 17.9 | 33.2 | 17 | 42 | 0.88 (0.48 - 1.55) | 0.67 | 0.88 (0.49 - 1.59) | 0.68 |
| MDCD | 1,207 | 2,474 | 0.8 | 1.7 | 0 | 6 | | | | |
| Open Claims | 344,148 | 706,389 | 319.2 | 815.9 | 254 | 616 | 1.04 (0.90 - 1.21) | 0.57 | 1.04 (0.86 - 1.26) | 0.70 |
| OptumEHR | 23,335 | 58,344 | 12.1 | 35.8 | 15 | 39 | 1.25 (0.65 - 2.30) | 0.49 | 1.32 (0.70 - 2.50) | 0.39 |
| VA | 15,307 | 110,551 | 15.8 | 124.8 | <10 | 32 | 0.45 (0.06 - 1.88) | 0.35 | 0.46 (0.08 - 2.46) | 0.36 |
| GermanyDA | 12,634 | 2,999 | 14.1 | 3.3 | 0 | 0 | | | | |
| SIDIAP | 26,465 | 8,098 | 70.9 | 22.6 | 0 | <5 | | | | |
| Meta-analysis | 412,608 | 864,625 | 372.0 | 954.0 | 323 | 808 | 1.04 (0.91 - 1.19) | 0.56 | 1.04 (0.89 - 1.23) | 0.60 |
| **PS-matching / on-treatment TAR** | | | | | | | | | | |
| CCAE | 1,895 | 3,310 | 0.5 | 0.9 | 0 | 0 | | | | |
| OptumDOD | 26,305 | 66,120 | 21.8 | 69.1 | 35 | 111 | 1.12 (0.66 - 1.85) | 0.67 | 1.14 (0.68 - 1.91) | 0.62 |
| MDCR | 16,716 | 32,819 | 16.6 | 32.3 | 16 | 42 | 0.68 (0.28 - 1.49) | 0.37 | 0.65 (0.28 - 1.51) | 0.32 |
| MDCD | 1,137 | 2,455 | 0.8 | 1.7 | 0 | 6 | | | | |
| Open Claims | 337,485 | 706,385 | 310.9 | 815.9 | 249 | 616 | 1.03 (0.83 - 1.26) | 0.79 | 1.04 (0.83 - 1.30) | 0.74 |
| OptumEHR | 22,121 | 58,301 | 11.4 | 35.8 | 14 | 39 | 0.53 (0.17 - 1.34) | 0.22 | 0.54 (0.19 - 1.52) | 0.25 |
| VA | 14,226 | 79,950 | 15.0 | 85.7 | <10 | 24 | | | | |
| GermanyDA | 12,597 | 2,566 | 14.0 | 2.8 | 0 | 0 | | | | |
| SIDIAP | 26,339 | 6,148 | 70.8 | 16.3 | 0 | <5 | | | | |
| Meta-analysis | 402,627 | 863,625 | 360.8 | 953.0 | 314 | 808 | 1.00 (0.83 - 1.20) | 0.98 | 1.00 (0.82 - 1.22) | 0.95 |
| **PS-stratification / intent-to-treat TAR** | | | | | | | | | | |
| CCAE | 2,088 | 3,383 | 0.7 | 1.2 | 0 | 0 | | | | |
| OptumDOD | 27,197 | 66,189 | 82.3 | 216.1 | 156 | 490 | 0.85 (0.70 - 1.02) | 0.08 | 0.85 (0.70 - 1.02) | 0.09 |
| MDCR | 17,928 | 33,703 | 51.3 | 99.9 | 56 | 183 | 0.64 (0.46 - 0.86) | <0.01 | 0.64 (0.47 - 0.88) | <0.01 |
| MDCD | 1,207 | 2,474 | 3.6 | 8.7 | 6 | 23 | 0.77 (0.27 - 1.84) | 0.58 | 0.68 (0.26 - 1.75) | 0.42 |
| Open Claims | 344,148 | 706,389 | 1468.7 | 2902.8 | 1,543 | 3,710 | 0.84 (0.79 - 0.90) | <0.01 | 0.84 (0.76 - 0.94) | <0.01 |
| OptumEHR | 23,335 | 58,344 | 80.4 | 222.9 | 59 | 219 | 0.83 (0.61 - 1.12) | 0.24 | 0.87 (0.64 - 1.18) | 0.37 |
| VA | 15,307 | 110,551 | 37.4 | 598.0 | <10 | 346 | 0.33 (0.14 - 0.65) | <0.01 | 0.33 (0.16 - 0.71) | <0.01 |
| GermanyDA | 12,634 | 2,999 | 52.2 | 17.7 | 0 | 0 | | | | |
| SIDIAP | 26,465 | 8,098 | 106.7 | 58.9 | 0 | <5 | | | | |
| Meta-analysis | 413,815 | 867,099 | 1686.2 | 3450.4 | 1,820 | 4,625 | 0.84 (0.79 - 0.88) | <0.01 | 0.84 (0.76 - 0.92) | <0.01 |
| **PS-matching / intent-to-treat TAR** | | | | | | | | | | |
| CCAE | 1,895 | 3,310 | 0.6 | 1.1 | 0 | 0 | | | | |
| OptumDOD | 26,305 | 66,120 | 79.8 | 215.8 | 154 | 490 | 0.85 (0.67 - 1.06) | 0.16 | 0.85 (0.68 - 1.07) | 0.18 |
| MDCR | 16,716 | 32,819 | 48.1 | 97.3 | 55 | 183 | 0.63 (0.41 - 0.94) | 0.03 | 0.62 (0.41 - 0.94) | 0.03 |
| MDCD | 1,137 | 2,455 | 3.4 | 8.6 | 6 | 23 | 0.85 (0.26 - 2.38) | 0.77 | 0.73 (0.24 - 2.22) | 0.59 |
| Open Claims | 337,485 | 706,385 | 1438.3 | 2902.8 | 1,519 | 3,710 | 0.88 (0.82 - 0.95) | <0.01 | 0.88 (0.79 - 0.98) | 0.03 |
| OptumEHR | 22,121 | 58,301 | 76.7 | 222.6 | 58 | 219 | 0.80 (0.56 - 1.13) | 0.22 | 0.84 (0.59 - 1.20) | 0.35 |
| VA | 14,226 | 79,950 | 36.1 | 358.2 | <10 | 220 | 0.42 (0.18 - 0.84) | 0.03 | 0.43 (0.20 - 0.92) | 0.03 |
| GermanyDA | 12,597 | 2,566 | 52.1 | 14.1 | 0 | 0 | | | | |
| SIDIAP | 26,339 | 6,148 | 106.6 | 41.2 | 0 | <5 | | | | |
| Meta-analysis | 417,990 | 946,030 | 1682.3 | 3805.3 | <1,802 | 4,845 | 0.84 (0.76 - 0.93) | <0.01 | 0.84 (0.74 - 0.95) | <0.01 |

##### 6.6.7 Diarrhea

**Supplementary Table 204:** Exposures, outcomes and risk estimates for Diarrhea in DPP4I (T) vs SU (C) new-user cohorts under four different propensity score (PS) adjustment / time-at-risk (TAR) designs across data sources. We report total patient exposure time in 1,000 patient-year (KYr) units, uncalibrated and calibrated hazard ratios (HRs) and their 95% confidence intervals (CIs). Greyed out entries did not pass study diagnostics and are included only for completeness.

| | Patients | | Exposure (KYr) | | Outcomes | | Uncalibrated | | Calibrated | |
|---|---|---|---|---|---|---|---|---|---|---|
| | T | C | T | C | T | C | HR (95% CI) | $p$ | HR (95% CI) | $p$ |
| **PS-stratification / on-treatment TAR** | | | | | | | | | | |
| CCAE | 1,822 | 2,998 | 0.5 | 0.8 | 17 | 35 | 0.73 (0.39 - 1.31) | 0.30 | 0.81 (0.44 - 1.50) | 0.53 |
| OptumDOD | 22,561 | 55,730 | 18.1 | 56.0 | 813 | 2,541 | 0.91 (0.84 - 0.99) | 0.03 | 0.92 (0.84 - 1.01) | 0.12 |
| MDCR | 15,423 | 29,180 | 15.0 | 27.9 | 534 | 1,010 | 0.93 (0.83 - 1.03) | 0.17 | 0.93 (0.83 - 1.04) | 0.26 |
| MDCD | 976 | 2,046 | 0.7 | 1.3 | 34 | 66 | 0.92 (0.59 - 1.41) | 0.71 | 0.81 (0.52 - 1.25) | 0.37 |
| Open Claims | 292,928 | 610,026 | 267.1 | 694.4 | 6,001 | 14,462 | 0.99 (0.95 - 1.02) | 0.35 | 0.98 (0.87 - 1.11) | 0.76 |
| OptumEHR | 20,428 | 51,411 | 10.2 | 29.7 | 401 | 1,351 | 0.92 (0.82 - 1.04) | 0.18 | 0.98 (0.86 - 1.11) | 0.76 |
| VA | 13,322 | 98,467 | 13.7 | 109.1 | 261 | 1,917 | 1.10 (0.94 - 1.28) | 0.24 | 1.12 (0.95 - 1.30) | 0.19 |
| GermanyDA | 12,474 | 2,955 | 13.8 | 3.3 | 28 | 7 | 1.11 (0.47 - 2.97) | 0.82 | 1.30 (0.52 - 3.27) | 0.59 |
| SIDIAP | 25,045 | 7,879 | 66.5 | 21.7 | 752 | 145 | 1.06 (0.87 - 1.31) | 0.58 | 1.06 (0.82 - 1.36) | 0.72 |
| Meta-analysis | 391,657 | 762,225 | 391.9 | 835.2 | 8,580 | 19,617 | 0.96 (0.91 - 1.00) | 0.05 | 0.96 (0.87 - 1.06) | 0.41 |
| **PS-matching / on-treatment TAR** | | | | | | | | | | |
| CCAE | 1,665 | 2,933 | 0.5 | 0.8 | 15 | 35 | 0.61 (0.27 - 1.29) | 0.22 | 0.62 (0.28 - 1.37) | 0.26 |
| OptumDOD | 21,835 | 55,648 | 17.5 | 55.9 | 777 | 2,540 | 0.91 (0.82 - 1.02) | 0.10 | 0.93 (0.83 - 1.04) | 0.25 |
| MDCR | 14,380 | 28,339 | 14.0 | 27.1 | 490 | 986 | 0.90 (0.77 - 1.05) | 0.17 | 0.86 (0.74 - 1.01) | 0.09 |
| MDCD | 938 | 2,030 | 0.6 | 1.3 | 33 | 63 | 0.74 (0.40 - 1.34) | 0.34 | 0.70 (0.38 - 1.29) | 0.27 |
| Open Claims | 287,496 | 610,023 | 260.5 | 694.4 | 5,794 | 14,462 | 0.95 (0.91 - 0.99) | 0.03 | 0.96 (0.88 - 1.06) | 0.47 |
| OptumEHR | 19,414 | 51,374 | 9.6 | 29.6 | 375 | 1,350 | 0.90 (0.77 - 1.05) | 0.20 | 0.93 (0.80 - 1.09) | 0.43 |
| VA | 12,415 | 70,431 | 13.1 | 74.1 | 247 | 1,324 | 1.14 (0.94 - 1.38) | 0.19 | 1.15 (0.94 - 1.40) | 0.21 |
| GermanyDA | 12,439 | 2,528 | 13.8 | 2.8 | 28 | 7 | 1.16 (0.40 - 4.27) | 0.80 | 1.20 (0.36 - 3.95) | 0.77 |
| SIDIAP | 24,934 | 5,941 | 66.4 | 15.5 | 751 | 115 | 0.98 (0.75 - 1.28) | 0.86 | 1.04 (0.69 - 1.56) | 0.84 |
| Meta-analysis | 395,516 | 829,247 | 395.9 | 901.6 | 8,510 | 20,882 | 0.95 (0.91 - 0.98) | <0.01 | 0.95 (0.87 - 1.04) | 0.26 |
| **PS-stratification / intent-to-treat TAR** | | | | | | | | | | |
| CCAE | 1,822 | 2,998 | 0.6 | 1.0 | 21 | 41 | 0.77 (0.44 - 1.32) | 0.36 | 0.81 (0.46 - 1.42) | 0.48 |
| OptumDOD | 22,561 | 55,730 | 62.6 | 165.0 | 2,709 | 7,255 | 0.94 (0.90 - 0.99) | <0.01 | 0.94 (0.89 - 1.00) | 0.09 |
| MDCR | 15,423 | 29,180 | 40.9 | 80.4 | 1,699 | 3,276 | 0.99 (0.93 - 1.06) | 0.82 | 1.00 (0.92 - 1.08) | 0.87 |
| MDCD | 976 | 2,046 | 2.6 | 6.5 | 120 | 283 | 0.95 (0.76 - 1.19) | 0.67 | 0.84 (0.67 - 1.05) | 0.17 |
| Open Claims | 292,928 | 610,026 | 1184.7 | 2381.8 | 26,865 | 52,315 | 0.99 (0.98 - 1.01) | 0.41 | 0.99 (0.90 - 1.09) | 0.87 |
| OptumEHR | 20,428 | 51,411 | 65.1 | 179.8 | 1,999 | 5,864 | 0.98 (0.93 - 1.04) | 0.57 | 1.03 (0.96 - 1.09) | 0.48 |
| VA | 13,322 | 98,467 | 32.2 | 509.3 | 621 | 9,154 | 1.03 (0.94 - 1.13) | 0.56 | 1.05 (0.95 - 1.16) | 0.35 |
| GermanyDA | 12,474 | 2,955 | 51.2 | 17.3 | 80 | 33 | 0.83 (0.53 - 1.32) | 0.42 | 0.86 (0.54 - 1.35) | 0.51 |
| SIDIAP | 25,045 | 7,879 | 99.9 | 55.5 | 1,174 | 578 | 0.91 (0.81 - 1.03) | 0.12 | 0.89 (0.79 - 1.00) | 0.08 |
| Meta-analysis | 393,694 | 765,299 | 1515.2 | 2900.0 | 34,691 | 69,673 | 0.98 (0.95 - 1.00) | 0.08 | 0.98 (0.90 - 1.06) | 0.56 |
| **PS-matching / intent-to-treat TAR** | | | | | | | | | | |
| CCAE | 1,665 | 2,933 | 0.6 | 1.0 | 19 | 41 | 0.88 (0.45 - 1.65) | 0.70 | 0.87 (0.45 - 1.68) | 0.70 |
| OptumDOD | 21,835 | 55,648 | 60.7 | 164.7 | 2,608 | 7,239 | 0.93 (0.88 - 0.98) | 0.01 | 0.93 (0.87 - 1.00) | 0.08 |
| MDCR | 14,380 | 28,339 | 38.4 | 78.1 | 1,590 | 3,194 | 0.98 (0.91 - 1.06) | 0.65 | 0.97 (0.89 - 1.06) | 0.57 |
| MDCD | 938 | 2,030 | 2.5 | 6.4 | 118 | 277 | 0.98 (0.75 - 1.28) | 0.90 | 0.85 (0.65 - 1.12) | 0.28 |
| Open Claims | 287,496 | 610,023 | 1161.6 | 2381.8 | 26,154 | 52,314 | 0.99 (0.98 - 1.01) | 0.57 | 0.99 (0.91 - 1.09) | 0.88 |
| OptumEHR | 19,414 | 51,374 | 62.2 | 179.6 | 1,909 | 5,858 | 1.01 (0.94 - 1.07) | 0.87 | 1.05 (0.98 - 1.13) | 0.21 |
| VA | 12,415 | 70,431 | 31.1 | 304.0 | 592 | 5,418 | 1.01 (0.90 - 1.12) | 0.91 | 1.03 (0.92 - 1.15) | 0.63 |
| GermanyDA | 12,439 | 2,528 | 51.2 | 13.8 | 80 | 26 | 0.92 (0.54 - 1.65) | 0.78 | 0.93 (0.53 - 1.64) | 0.81 |
| SIDIAP | 24,934 | 5,941 | 99.8 | 38.6 | 1,172 | 409 | 0.87 (0.76 - 1.00) | 0.05 | 0.88 (0.76 - 1.01) | 0.10 |
| Meta-analysis | 396,861 | 832,297 | 1513.1 | 3180.5 | 34,263 | 74,804 | 0.98 (0.95 - 1.01) | 0.16 | 0.98 (0.90 - 1.06) | 0.60 |

##### 6.6.8 Genitourinary infection

**Supplementary Table 205:** Exposures, outcomes and risk estimates for Genitourinary infection in DPP4I (T) vs SU (C) new-user cohorts under four different propensity score (PS) adjustment / time-at-risk (TAR) designs across data sources. We report total patient exposure time in 1,000 patient-year (KYr) units, uncalibrated and calibrated hazard ratios (HRs) and their 95% confidence intervals (CIs). Greyed out entries did not pass study diagnostics and are included only for completeness.

| | Patients | | Exposure (KYr) | | Outcomes | | Uncalibrated | | Calibrated | |
|---|---|---|---|---|---|---|---|---|---|---|
| | T | C | T | C | T | C | HR (95% CI) | $p$ | HR (95% CI) | $p$ |
| **PS-stratification / on-treatment TAR** | | | | | | | | | | |
| CCAE | 1,613 | 2,697 | 0.5 | 0.7 | 19 | 33 | 0.98 (0.53 - 1.75) | 0.95 | 1.09 (0.60 - 1.99) | 0.76 |
| OptumDOD | 19,139 | 48,391 | 15.2 | 47.6 | 1,272 | 3,916 | 0.93 (0.87 - 1.00) | 0.04 | 0.94 (0.87 - 1.01) | 0.19 |
| MDCR | 13,747 | 26,273 | 13.2 | 25.1 | 852 | 1,605 | 1.01 (0.92 - 1.10) | 0.89 | 1.01 (0.92 - 1.11) | 0.82 |
| MDCD | 798 | 1,741 | 0.5 | 1.2 | 51 | 108 | 1.04 (0.73 - 1.47) | 0.81 | 0.91 (0.64 - 1.30) | 0.66 |
| Open Claims | 247,187 | 528,233 | 222.8 | 600.3 | 10,583 | 24,933 | 1.03 (1.01 - 1.05) | 0.02 | 1.02 (0.91 - 1.16) | 0.71 |
| OptumEHR | 18,902 | 48,235 | 9.3 | 27.6 | 633 | 1,855 | 1.02 (0.93 - 1.12) | 0.68 | 1.08 (0.98 - 1.20) | 0.22 |
| VA | 13,255 | 96,687 | 13.7 | 107.5 | 231 | 2,185 | 0.85 (0.72 - 0.99) | 0.04 | 0.87 (0.74 - 1.02) | 0.09 |
| GermanyDA | 9,479 | 2,314 | 10.0 | 2.4 | 407 | 100 | 1.04 (0.81 - 1.34) | 0.76 | 1.21 (0.94 - 1.57) | 0.24 |
| SIDIAP | 20,527 | 7,170 | 53.0 | 18.6 | 2,390 | 689 | 1.02 (0.92 - 1.12) | 0.72 | 1.01 (0.85 - 1.21) | 0.82 |
| Meta-analysis | 333,273 | 667,821 | 324.4 | 723.6 | 16,214 | 33,263 | 1.00 (0.97 - 1.04) | 0.85 | 1.01 (0.91 - 1.11) | 0.87 |
| **PS-matching / on-treatment TAR** | | | | | | | | | | |
| CCAE | 1,466 | 2,615 | 0.4 | 0.7 | 19 | 32 | 0.94 (0.47 - 1.81) | 0.85 | 0.95 (0.48 - 1.88) | 0.79 |
| OptumDOD | 18,599 | 48,346 | 14.7 | 47.5 | 1,224 | 3,914 | 0.98 (0.90 - 1.07) | 0.67 | 1.00 (0.91 - 1.09) | 0.84 |
| MDCR | 12,924 | 25,413 | 12.4 | 24.3 | 796 | 1,566 | 1.01 (0.90 - 1.14) | 0.84 | 0.97 (0.86 - 1.10) | 0.70 |
| MDCD | 766 | 1,729 | 0.5 | 1.1 | 47 | 105 | 0.94 (0.55 - 1.55) | 0.80 | 0.88 (0.52 - 1.48) | 0.65 |
| Open Claims | 243,347 | 528,224 | 218.2 | 600.3 | 10,246 | 24,933 | 1.01 (0.97 - 1.04) | 0.76 | 1.02 (0.93 - 1.11) | 0.74 |
| OptumEHR | 18,003 | 48,199 | 8.8 | 27.6 | 596 | 1,851 | 1.03 (0.91 - 1.16) | 0.65 | 1.06 (0.93 - 1.20) | 0.44 |
| VA | 12,386 | 69,459 | 13.1 | 73.6 | 220 | 1,431 | 0.90 (0.73 - 1.09) | 0.28 | 0.90 (0.74 - 1.11) | 0.36 |
| GermanyDA | 9,452 | 1,968 | 10.0 | 2.0 | 405 | 84 | 0.97 (0.71 - 1.33) | 0.83 | 1.00 (0.72 - 1.37) | 0.70 |
| SIDIAP | 20,451 | 5,348 | 52.9 | 13.1 | 2,388 | 505 | 1.02 (0.89 - 1.16) | 0.82 | 1.08 (0.77 - 1.51) | 0.69 |
| Meta-analysis | 337,394 | 731,301 | 330.8 | 790.3 | 15,941 | 34,421 | 1.00 (0.97 - 1.03) | 0.90 | 1.00 (0.92 - 1.10) | 0.87 |
| **PS-stratification / intent-to-treat TAR** | | | | | | | | | | |
| CCAE | 1,613 | 2,697 | 0.6 | 0.9 | 22 | 42 | 0.92 (0.53 - 1.56) | 0.77 | 0.97 (0.56 - 1.67) | 0.81 |
| OptumDOD | 19,139 | 48,391 | 49.1 | 130.8 | 3,948 | 10,428 | 0.97 (0.93 - 1.00) | 0.07 | 0.97 (0.92 - 1.02) | 0.29 |
| MDCR | 13,747 | 26,273 | 34.8 | 68.8 | 2,353 | 4,697 | 0.98 (0.93 - 1.03) | 0.34 | 0.98 (0.91 - 1.05) | 0.65 |
| MDCD | 798 | 1,741 | 1.9 | 5.1 | 182 | 400 | 1.20 (0.99 - 1.44) | 0.06 | 1.06 (0.88 - 1.27) | 0.62 |
| Open Claims | 247,187 | 528,233 | 949.0 | 1964.3 | 46,279 | 90,943 | 1.00 (0.99 - 1.01) | 0.57 | 1.00 (0.91 - 1.10) | 0.92 |
| OptumEHR | 18,902 | 48,235 | 57.9 | 162.5 | 2,754 | 7,848 | 0.99 (0.95 - 1.04) | 0.83 | 1.04 (0.98 - 1.10) | 0.28 |
| VA | 13,255 | 96,687 | 32.0 | 496.7 | 571 | 10,716 | 0.86 (0.78 - 0.95) | <0.01 | 0.88 (0.80 - 0.97) | 0.02 |
| GermanyDA | 9,479 | 2,314 | 34.9 | 11.6 | 1,310 | 453 | 1.05 (0.94 - 1.19) | 0.38 | 1.09 (0.97 - 1.23) | 0.24 |
| SIDIAP | 20,527 | 7,170 | 78.2 | 44.8 | 3,801 | 2,168 | 0.95 (0.89 - 1.01) | 0.08 | 0.92 (0.86 - 0.99) | 0.06 |
| Meta-analysis | 333,273 | 667,821 | 1213.3 | 2399.8 | 60,782 | 117,285 | 0.99 (0.97 - 1.01) | 0.28 | 0.99 (0.91 - 1.07) | 0.77 |
| **PS-matching / intent-to-treat TAR** | | | | | | | | | | |
| CCAE | 1,466 | 2,615 | 0.5 | 0.9 | 22 | 40 | 0.87 (0.46 - 1.60) | 0.67 | 0.87 (0.46 - 1.62) | 0.67 |
| OptumDOD | 18,599 | 48,346 | 47.8 | 130.7 | 3,825 | 10,412 | 0.97 (0.92 - 1.01) | 0.17 | 0.97 (0.91 - 1.03) | 0.43 |
| MDCR | 12,924 | 25,413 | 32.9 | 66.5 | 2,207 | 4,586 | 0.98 (0.92 - 1.05) | 0.60 | 0.97 (0.90 - 1.05) | 0.54 |
| MDCD | 766 | 1,729 | 1.8 | 5.1 | 173 | 395 | 1.25 (0.99 - 1.57) | 0.06 | 1.08 (0.86 - 1.37) | 0.54 |
| Open Claims | 243,347 | 528,224 | 934.1 | 1964.2 | 45,188 | 90,941 | 0.99 (0.98 - 1.01) | 0.34 | 0.99 (0.91 - 1.08) | 0.86 |
| OptumEHR | 18,003 | 48,199 | 55.4 | 162.3 | 2,641 | 7,838 | 1.05 (0.99 - 1.11) | 0.09 | 1.10 (1.03 - 1.17) | 0.01 |
| VA | 12,386 | 69,459 | 30.9 | 299.1 | 551 | 6,087 | 0.91 (0.82 - 1.02) | 0.10 | 0.94 (0.83 - 1.05) | 0.29 |
| GermanyDA | 9,452 | 1,968 | 34.8 | 9.4 | 1,306 | 346 | 1.05 (0.91 - 1.22) | 0.49 | 1.06 (0.92 - 1.23) | 0.52 |
| SIDIAP | 20,451 | 5,348 | 78.1 | 31.0 | 3,797 | 1,519 | 0.96 (0.89 - 1.03) | 0.23 | 0.97 (0.89 - 1.04) | 0.48 |
| Meta-analysis | 338,604 | 734,045 | 1220.9 | 2680.0 | 59,806 | 122,466 | 0.99 (0.98 - 1.00) | 0.24 | 0.99 (0.92 - 1.07) | 0.86 |

##### 6.6.9 Hyperkalemia

**Supplementary Table 206:** Exposures, outcomes and risk estimates for Hyperkalemia in DPP4I (T) vs SU (C) new-user cohorts under four different propensity score (PS) adjustment / time-at-risk (TAR) designs across data sources. We report total patient exposure time in 1,000 patient-year (KYr) units, uncalibrated and calibrated hazard ratios (HRs) and their 95% confidence intervals (CIs). Greyed out entries did not pass study diagnostics and are included only for completeness.

| | Patients | | Exposure (KYr) | | Outcomes | | Uncalibrated | | Calibrated | |
|---|---|---|---|---|---|---|---|---|---|---|
| | T | C | T | C | T | C | HR (95% CI) | $p$ | HR (95% CI) | $p$ |
| **PS-stratification / on-treatment TAR** | | | | | | | | | | |
| CCAE | 2,034 | 3,301 | 0.6 | 0.9 | 6 | 14 | 0.87 (0.30 - 2.25) | 0.79 | 0.98 (0.36 - 2.67) | 0.89 |
| OptumDOD | 25,334 | 61,820 | 20.6 | 62.8 | 688 | 2,047 | 1.00 (0.91 - 1.09) | 0.93 | 1.00 (0.91 - 1.11) | 0.88 |
| MDCR | 17,425 | 32,657 | 17.3 | 31.8 | 277 | 556 | 0.93 (0.80 - 1.08) | 0.35 | 0.93 (0.80 - 1.09) | 0.42 |
| MDCD | 1,112 | 2,305 | 0.7 | 1.6 | 26 | 48 | 1.19 (0.70 - 1.96) | 0.51 | 1.04 (0.62 - 1.74) | 0.84 |
| Open Claims | 331,544 | 682,393 | 304.6 | 780.3 | 3,844 | 9,285 | 1.05 (1.01 - 1.09) | 0.02 | 1.04 (0.92 - 1.18) | 0.53 |
| OptumEHR | 22,494 | 56,312 | 11.5 | 33.6 | 305 | 809 | 1.27 (1.10 - 1.46) | <0.01 | 1.35 (1.17 - 1.56) | <0.01 |
| VA | 14,128 | 101,963 | 14.4 | 112.5 | 226 | 2,204 | 0.95 (0.80 - 1.11) | 0.51 | 0.96 (0.82 - 1.14) | 0.68 |
| GermanyDA | 11,553 | 2,842 | 12.6 | 3.1 | 243 | 52 | 0.97 (0.69 - 1.38) | 0.85 | 1.13 (0.79 - 1.61) | 0.57 |
| SIDIAP | 24,462 | 7,741 | 64.2 | 21.3 | 889 | 283 | 1.01 (0.87 - 1.19) | 0.86 | 1.01 (0.81 - 1.26) | 0.84 |
| Meta-analysis | 435,958 | 849,371 | 432.1 | 935.3 | 6,278 | 13,094 | 1.04 (0.96 - 1.13) | 0.30 | 1.05 (0.93 - 1.18) | 0.47 |
| **PS-matching / on-treatment TAR** | | | | | | | | | | |
| CCAE | 1,846 | 3,228 | 0.5 | 0.9 | 6 | 14 | 0.85 (0.26 - 2.40) | 0.78 | 0.87 (0.29 - 2.62) | 0.80 |
| OptumDOD | 24,539 | 61,759 | 19.9 | 62.8 | 668 | 2,044 | 0.97 (0.86 - 1.10) | 0.68 | 0.99 (0.88 - 1.12) | 0.86 |
| MDCR | 16,228 | 31,625 | 16.0 | 30.8 | 260 | 545 | 0.93 (0.75 - 1.15) | 0.52 | 0.90 (0.72 - 1.11) | 0.34 |
| MDCD | 1,051 | 2,289 | 0.7 | 1.5 | 26 | 48 | 0.73 (0.32 - 1.54) | 0.43 | 0.69 (0.32 - 1.50) | 0.36 |
| Open Claims | 325,065 | 682,389 | 296.7 | 780.3 | 3,739 | 9,285 | 1.03 (0.97 - 1.08) | 0.34 | 1.04 (0.94 - 1.15) | 0.49 |
| OptumEHR | 21,328 | 56,273 | 10.7 | 33.6 | 285 | 806 | 1.28 (1.06 - 1.55) | <0.01 | 1.32 (1.09 - 1.60) | <0.01 |
| VA | 13,147 | 73,463 | 13.7 | 77.4 | 212 | 1,406 | 1.07 (0.87 - 1.31) | 0.53 | 1.08 (0.88 - 1.32) | 0.52 |
| GermanyDA | 11,527 | 2,429 | 12.6 | 2.6 | 242 | 41 | 1.23 (0.76 - 2.07) | 0.43 | 1.26 (0.76 - 2.10) | 0.42 |
| SIDIAP | 24,353 | 5,873 | 64.2 | 15.3 | 885 | 206 | 1.03 (0.83 - 1.27) | 0.81 | 1.09 (0.75 - 1.59) | 0.68 |
| Meta-analysis | 439,084 | 919,328 | 434.9 | 1005.1 | 6,323 | 14,395 | 1.03 (0.99 - 1.08) | 0.19 | 1.03 (0.94 - 1.13) | 0.53 |
| **PS-stratification / intent-to-treat TAR** | | | | | | | | | | |
| CCAE | 2,034 | 3,301 | 0.7 | 1.1 | 6 | 16 | 0.75 (0.26 - 1.89) | 0.57 | 0.79 (0.30 - 2.12) | 0.65 |
| OptumDOD | 25,334 | 61,820 | 72.7 | 190.7 | 2,308 | 6,120 | 0.99 (0.95 - 1.04) | 0.82 | 1.00 (0.94 - 1.06) | 0.86 |
| MDCR | 17,425 | 32,657 | 48.7 | 94.4 | 903 | 1,910 | 0.98 (0.90 - 1.06) | 0.57 | 0.98 (0.89 - 1.08) | 0.74 |
| MDCD | 1,112 | 2,305 | 3.2 | 7.8 | 92 | 237 | 0.98 (0.76 - 1.26) | 0.87 | 0.86 (0.67 - 1.12) | 0.30 |
| Open Claims | 331,544 | 682,393 | 1383.3 | 2736.3 | 17,459 | 36,579 | 0.96 (0.94 - 0.98) | <0.01 | 0.96 (0.87 - 1.05) | 0.38 |
| OptumEHR | 22,494 | 56,312 | 74.6 | 205.2 | 1,289 | 3,919 | 1.00 (0.94 - 1.07) | 0.90 | 1.05 (0.97 - 1.13) | 0.27 |
| VA | 14,128 | 101,963 | 33.7 | 525.0 | 556 | 10,153 | 0.99 (0.89 - 1.09) | 0.80 | 1.01 (0.91 - 1.12) | 0.84 |
| GermanyDA | 11,553 | 2,842 | 45.6 | 15.9 | 876 | 278 | 0.99 (0.85 - 1.15) | 0.87 | 1.02 (0.87 - 1.19) | 0.74 |
| SIDIAP | 24,462 | 7,741 | 96.6 | 53.3 | 1,371 | 847 | 0.93 (0.84 - 1.03) | 0.15 | 0.91 (0.82 - 1.00) | 0.10 |
| Meta-analysis | 435,958 | 849,371 | 1725.3 | 3304.8 | 24,304 | 49,906 | 0.97 (0.95 - 0.99) | <0.01 | 0.97 (0.90 - 1.05) | 0.43 |
| **PS-matching / intent-to-treat TAR** | | | | | | | | | | |
| CCAE | 1,846 | 3,228 | 0.6 | 1.1 | 6 | 16 | 0.64 (0.20 - 1.71) | 0.41 | 0.63 (0.22 - 1.84) | 0.41 |
| OptumDOD | 24,539 | 61,759 | 70.6 | 190.5 | 2,234 | 6,107 | 1.02 (0.96 - 1.09) | 0.49 | 1.03 (0.95 - 1.10) | 0.54 |
| MDCR | 16,228 | 31,625 | 45.5 | 91.5 | 852 | 1,867 | 0.98 (0.88 - 1.09) | 0.65 | 0.97 (0.86 - 1.08) | 0.58 |
| MDCD | 1,051 | 2,289 | 3.0 | 7.7 | 89 | 233 | 0.84 (0.61 - 1.14) | 0.27 | 0.73 (0.53 - 0.99) | 0.06 |
| Open Claims | 325,065 | 682,389 | 1354.3 | 2736.3 | 17,116 | 36,578 | 0.97 (0.95 - 1.00) | 0.02 | 0.97 (0.89 - 1.06) | 0.57 |
| OptumEHR | 21,328 | 56,273 | 71.1 | 205.0 | 1,232 | 3,913 | 0.99 (0.92 - 1.07) | 0.86 | 1.04 (0.96 - 1.13) | 0.41 |
| VA | 13,147 | 73,463 | 32.5 | 316.2 | 534 | 5,717 | 1.01 (0.91 - 1.13) | 0.82 | 1.04 (0.92 - 1.16) | 0.57 |
| GermanyDA | 11,527 | 2,429 | 45.5 | 12.8 | 874 | 217 | 1.03 (0.86 - 1.24) | 0.77 | 1.04 (0.86 - 1.25) | 0.71 |
| SIDIAP | 24,353 | 5,873 | 96.5 | 37.5 | 1,366 | 573 | 0.90 (0.80 - 1.02) | 0.09 | 0.91 (0.81 - 1.03) | 0.18 |
| Meta-analysis | 439,084 | 919,328 | 1719.7 | 3598.6 | 24,303 | 55,221 | 0.98 (0.96 - 1.00) | 0.03 | 0.98 (0.91 - 1.06) | 0.60 |

##### 6.6.10 Hypoglycemia

**Supplementary Table 207:** Exposures, outcomes and risk estimates for Hypoglycemia in DPP4I (T) vs SU (C) new-user cohorts under four different propensity score (PS) adjustment / time-at-risk (TAR) designs across data sources. We report total patient exposure time in 1,000 patient-year (KYr) units, uncalibrated and calibrated hazard ratios (HRs) and their 95% confidence intervals (CIs). Greyed out entries did not pass study diagnostics and are included only for completeness.

| | Patients | | Exposure (KYr) | | Outcomes | | Uncalibrated | | Calibrated | |
|---|---|---|---|---|---|---|---|---|---|---|
| | T | C | T | C | T | C | HR (95% CI) | $p$ | HR (95% CI) | $p$ |
| **PS-stratification / on-treatment TAR** | | | | | | | | | | |
| CCAE | 2,051 | 3,341 | 0.6 | 0.9 | <5 | 19 | 0.23 (0.05 - 0.68) | 0.02 | 0.25 (0.07 - 0.92) | 0.04 |
| OptumDOD | 26,793 | 65,348 | 22.3 | 67.4 | 176 | 1,838 | 0.27 (0.23 - 0.31) | <0.01 | 0.27 (0.23 - 0.31) | <0.01 |
| MDCR | 17,761 | 33,426 | 17.7 | 32.7 | 79 | 630 | 0.24 (0.18 - 0.30) | <0.01 | 0.24 (0.19 - 0.30) | <0.01 |
| MDCD | 1,174 | 2,405 | 0.8 | 1.6 | 7 | 63 | 0.20 (0.08 - 0.42) | <0.01 | 0.17 (0.08 - 0.40) | <0.01 |
| Open Claims | 338,626 | 696,050 | 313.9 | 800.5 | 1,048 | 9,476 | 0.25 (0.24 - 0.27) | <0.01 | 0.25 (0.22 - 0.29) | <0.01 |
| OptumEHR | 23,136 | 57,847 | 12.0 | 35.0 | 103 | 771 | 0.39 (0.31 - 0.48) | <0.01 | 0.41 (0.33 - 0.51) | <0.01 |
| VA | 15,174 | 109,673 | 15.7 | 123.3 | 38 | 1,028 | 0.29 (0.20 - 0.41) | <0.01 | 0.30 (0.21 - 0.42) | <0.01 |
| GermanyDA | 12,542 | 2,979 | 13.9 | 3.3 | 27 | 29 | 0.32 (0.18 - 0.60) | <0.01 | 0.38 (0.20 - 0.70) | <0.01 |
| SIDIAP | 26,423 | 8,087 | 70.7 | 22.4 | 59 | 110 | 0.16 (0.11 - 0.23) | <0.01 | 0.16 (0.11 - 0.24) | <0.01 |
| Meta-analysis | 448,506 | 869,483 | 451.9 | 963.9 | <1,504 | 12,936 | 0.26 (0.21 - 0.32) | <0.01 | 0.26 (0.21 - 0.32) | <0.01 |
| **PS-matching / on-treatment TAR** | | | | | | | | | | |
| CCAE | 1,868 | 3,269 | 0.5 | 0.9 | <5 | 19 | 0.23 (0.04 - 0.86) | 0.07 | 0.24 (0.05 - 1.16) | 0.08 |
| OptumDOD | 25,923 | 65,278 | 21.5 | 67.4 | 166 | 1,836 | 0.22 (0.18 - 0.27) | <0.01 | 0.22 (0.18 - 0.28) | <0.01 |
| MDCR | 16,573 | 32,541 | 16.5 | 31.8 | 74 | 618 | 0.20 (0.14 - 0.27) | <0.01 | 0.19 (0.14 - 0.26) | <0.01 |
| MDCD | 1,104 | 2,386 | 0.8 | 1.6 | 7 | 63 | 0.13 (0.03 - 0.37) | <0.01 | 0.13 (0.04 - 0.43) | <0.01 |
| Open Claims | 332,096 | 696,046 | 305.8 | 800.5 | 1,010 | 9,475 | 0.21 (0.19 - 0.22) | <0.01 | 0.21 (0.18 - 0.23) | <0.01 |
| OptumEHR | 21,940 | 57,804 | 11.2 | 35.0 | 98 | 770 | 0.32 (0.24 - 0.42) | <0.01 | 0.33 (0.25 - 0.44) | <0.01 |
| VA | 14,102 | 79,115 | 14.9 | 84.5 | 37 | 721 | 0.32 (0.21 - 0.47) | <0.01 | 0.32 (0.22 - 0.48) | <0.01 |
| GermanyDA | 12,506 | 2,544 | 13.9 | 2.8 | 27 | 22 | 0.45 (0.19 - 1.10) | 0.08 | 0.47 (0.20 - 1.13) | 0.10 |
| SIDIAP | 26,297 | 6,142 | 70.6 | 16.2 | 59 | 84 | 0.10 (0.05 - 0.18) | <0.01 | 0.11 (0.05 - 0.21) | <0.01 |
| Meta-analysis | 452,409 | 945,125 | 455.6 | 1040.6 | <1,483 | 13,608 | 0.23 (0.18 - 0.29) | <0.01 | 0.23 (0.18 - 0.29) | <0.01 |
| **PS-stratification / intent-to-treat TAR** | | | | | | | | | | |
| CCAE | 2,051 | 3,341 | 0.7 | 1.2 | <5 | 21 | 0.21 (0.05 - 0.63) | 0.02 | 0.22 (0.06 - 0.80) | 0.02 |
| OptumDOD | 26,793 | 65,348 | 79.6 | 203.9 | 966 | 4,714 | 0.51 (0.48 - 0.55) | <0.01 | 0.51 (0.47 - 0.55) | <0.01 |
| MDCR | 17,761 | 33,426 | 50.3 | 96.3 | 402 | 1,678 | 0.46 (0.41 - 0.52) | <0.01 | 0.47 (0.41 - 0.53) | <0.01 |
| MDCD | 1,174 | 2,405 | 3.4 | 8.0 | 46 | 215 | 0.47 (0.33 - 0.65) | <0.01 | 0.41 (0.30 - 0.58) | <0.01 |
| Open Claims | 338,626 | 696,050 | 1429.9 | 2790.4 | 8,070 | 31,818 | 0.49 (0.48 - 0.51) | <0.01 | 0.49 (0.45 - 0.54) | <0.01 |
| OptumEHR | 23,136 | 57,847 | 78.6 | 214.1 | 530 | 2,851 | 0.53 (0.48 - 0.58) | <0.01 | 0.55 (0.50 - 0.61) | <0.01 |
| VA | 15,174 | 109,673 | 36.9 | 580.1 | 140 | 4,804 | 0.51 (0.42 - 0.60) | <0.01 | 0.52 (0.43 - 0.62) | <0.01 |
| GermanyDA | 12,542 | 2,979 | 51.5 | 17.2 | 113 | 86 | 0.50 (0.36 - 0.69) | <0.01 | 0.52 (0.38 - 0.71) | <0.01 |
| SIDIAP | 26,423 | 8,087 | 106.2 | 57.7 | 195 | 294 | 0.43 (0.34 - 0.52) | <0.01 | 0.42 (0.34 - 0.51) | <0.01 |
| Meta-analysis | 450,565 | 872,561 | 1807.7 | 3401.1 | <10,394 | 41,804 | 0.50 (0.49 - 0.51) | <0.01 | 0.50 (0.46 - 0.54) | <0.01 |
| **PS-matching / intent-to-treat TAR** | | | | | | | | | | |
| CCAE | 1,868 | 3,269 | 0.6 | 1.1 | <5 | 21 | 0.19 (0.03 - 0.69) | 0.04 | 0.19 (0.04 - 0.91) | 0.04 |
| OptumDOD | 25,923 | 65,278 | 77.3 | 203.6 | 930 | 4,705 | 0.46 (0.42 - 0.51) | <0.01 | 0.46 (0.42 - 0.51) | <0.01 |
| MDCR | 16,573 | 32,541 | 47.2 | 93.7 | 377 | 1,658 | 0.41 (0.36 - 0.48) | <0.01 | 0.41 (0.35 - 0.48) | <0.01 |
| MDCD | 1,104 | 2,386 | 3.2 | 7.9 | 45 | 211 | 0.41 (0.27 - 0.61) | <0.01 | 0.36 (0.24 - 0.54) | <0.01 |
| Open Claims | 332,096 | 696,046 | 1400.3 | 2790.4 | 7,895 | 31,816 | 0.47 (0.45 - 0.48) | <0.01 | 0.47 (0.43 - 0.51) | <0.01 |
| OptumEHR | 21,940 | 57,804 | 74.9 | 213.8 | 510 | 2,845 | 0.51 (0.46 - 0.57) | <0.01 | 0.54 (0.48 - 0.60) | <0.01 |
| VA | 14,102 | 79,115 | 35.5 | 347.4 | 136 | 2,745 | 0.51 (0.41 - 0.62) | <0.01 | 0.52 (0.42 - 0.64) | <0.01 |
| GermanyDA | 12,506 | 2,544 | 51.4 | 13.8 | 113 | 63 | 0.47 (0.32 - 0.69) | <0.01 | 0.47 (0.32 - 0.70) | <0.01 |
| SIDIAP | 26,297 | 6,142 | 106.0 | 40.4 | 195 | 200 | 0.40 (0.31 - 0.51) | <0.01 | 0.41 (0.32 - 0.52) | <0.01 |
| Meta-analysis | 453,766 | 948,179 | 1801.6 | 3724.5 | <10,250 | 44,388 | 0.47 (0.46 - 0.48) | <0.01 | 0.47 (0.43 - 0.51) | <0.01 |

##### 6.6.11 Hypotension

**Supplementary Table 208:** Exposures, outcomes and risk estimates for Hypotension in DPP4I (T) vs SU (C) new-user cohorts under four different propensity score (PS) adjustment / time-at-risk (TAR) designs across data sources. We report total patient exposure time in 1,000 patient-year (KYr) units, uncalibrated and calibrated hazard ratios (HRs) and their 95% confidence intervals (CIs). Greyed out entries did not pass study diagnostics and are included only for completeness.

| | Patients | | Exposure (KYr) | | Outcomes | | Uncalibrated | | Calibrated | |
|---|---|---|---|---|---|---|---|---|---|---|
| | T | C | T | C | T | C | HR (95% CI) | $p$ | HR (95% CI) | $p$ |
| **PS-stratification / on-treatment TAR** | | | | | | | | | | |
| CCAE | 2,020 | 3,295 | 0.6 | 0.9 | 10 | 11 | 1.61 (0.64 - 4.01) | 0.31 | 1.80 (0.72 - 4.51) | 0.22 |
| OptumDOD | 25,211 | 61,517 | 20.6 | 63.3 | 715 | 2,001 | 1.06 (0.97 - 1.16) | 0.22 | 1.07 (0.97 - 1.17) | 0.26 |
| MDCR | 16,916 | 31,823 | 16.7 | 31.0 | 429 | 772 | 1.09 (0.96 - 1.23) | 0.20 | 1.09 (0.96 - 1.24) | 0.24 |
| MDCD | 1,074 | 2,211 | 0.7 | 1.5 | 43 | 65 | 1.28 (0.84 - 1.93) | 0.24 | 1.13 (0.74 - 1.71) | 0.61 |
| Open Claims | 325,257 | 670,854 | 299.7 | 771.9 | 4,467 | 10,242 | 1.08 (1.04 - 1.12) | <0.01 | 1.08 (0.95 - 1.22) | 0.27 |
| OptumEHR | 22,478 | 56,159 | 11.5 | 33.6 | 322 | 976 | 1.09 (0.95 - 1.25) | 0.20 | 1.16 (1.01 - 1.33) | 0.07 |
| VA | 14,124 | 102,850 | 14.5 | 114.6 | 219 | 1,851 | 1.02 (0.86 - 1.21) | 0.80 | 1.04 (0.88 - 1.23) | 0.66 |
| GermanyDA | 12,256 | 2,919 | 13.6 | 3.2 | 89 | 18 | 1.30 (0.75 - 2.39) | 0.37 | 1.52 (0.85 - 2.74) | 0.19 |
| SIDIAP | 26,228 | 8,073 | 70.1 | 22.5 | 183 | 25 | 1.52 (0.99 - 2.45) | 0.07 | 1.52 (0.94 - 2.45) | 0.10 |
| Meta-analysis | 431,440 | 836,851 | 433.5 | 928.0 | 6,258 | 14,110 | 1.08 (1.05 - 1.12) | <0.01 | 1.09 (0.99 - 1.20) | 0.09 |
| **PS-matching / on-treatment TAR** | | | | | | | | | | |
| CCAE | 1,835 | 3,224 | 0.5 | 0.9 | 8 | 10 | 1.53 (0.48 - 4.63) | 0.46 | 1.55 (0.50 - 4.81) | 0.46 |
| OptumDOD | 24,356 | 61,434 | 19.8 | 63.3 | 691 | 1,997 | 1.10 (0.97 - 1.24) | 0.12 | 1.12 (0.99 - 1.27) | 0.10 |
| MDCR | 15,772 | 30,879 | 15.5 | 30.1 | 390 | 750 | 1.07 (0.89 - 1.28) | 0.46 | 1.03 (0.86 - 1.23) | 0.78 |
| MDCD | 1,019 | 2,194 | 0.7 | 1.5 | 41 | 65 | 1.35 (0.76 - 2.40) | 0.30 | 1.28 (0.72 - 2.28) | 0.43 |
| Open Claims | 318,926 | 670,851 | 292.0 | 771.9 | 4,325 | 10,242 | 1.08 (1.03 - 1.14) | <0.01 | 1.09 (0.99 - 1.21) | 0.09 |
| OptumEHR | 21,284 | 56,118 | 10.7 | 33.5 | 298 | 971 | 1.10 (0.92 - 1.31) | 0.28 | 1.13 (0.95 - 1.35) | 0.20 |
| VA | 13,173 | 73,224 | 13.8 | 77.5 | 207 | 1,176 | 1.06 (0.86 - 1.30) | 0.59 | 1.07 (0.86 - 1.32) | 0.58 |
| GermanyDA | 12,221 | 2,496 | 13.5 | 2.7 | 89 | 15 | 1.09 (0.53 - 2.43) | 0.83 | 1.12 (0.52 - 2.41) | 0.77 |
| SIDIAP | 26,107 | 6,126 | 70.0 | 16.2 | 182 | 23 | 1.49 (0.87 - 2.70) | 0.17 | 1.58 (0.83 - 3.02) | 0.18 |
| Meta-analysis | 434,693 | 906,546 | 436.6 | 997.6 | 6,231 | 15,249 | 1.09 (1.04 - 1.13) | <0.01 | 1.09 (0.99 - 1.20) | 0.08 |
| **PS-stratification / intent-to-treat TAR** | | | | | | | | | | |
| CCAE | 2,020 | 3,295 | 0.7 | 1.1 | 12 | 18 | 1.23 (0.56 - 2.62) | 0.60 | 1.29 (0.59 - 2.81) | 0.53 |
| OptumDOD | 25,211 | 61,517 | 72.3 | 189.4 | 2,499 | 6,678 | 0.98 (0.94 - 1.03) | 0.50 | 0.98 (0.93 - 1.04) | 0.67 |
| MDCR | 16,916 | 31,823 | 46.4 | 90.3 | 1,415 | 2,947 | 1.00 (0.93 - 1.06) | 0.89 | 1.00 (0.92 - 1.09) | 0.88 |
| MDCD | 1,074 | 2,211 | 2.9 | 7.3 | 147 | 302 | 1.25 (1.01 - 1.54) | 0.04 | 1.10 (0.89 - 1.36) | 0.42 |
| Open Claims | 325,257 | 670,854 | 1351.2 | 2684.7 | 22,168 | 44,867 | 0.99 (0.97 - 1.01) | 0.25 | 0.99 (0.90 - 1.09) | 0.82 |
| OptumEHR | 22,478 | 56,159 | 74.3 | 204.7 | 1,429 | 4,255 | 1.05 (0.98 - 1.12) | 0.14 | 1.09 (1.02 - 1.17) | 0.02 |
| VA | 14,124 | 102,850 | 34.1 | 535.1 | 538 | 9,807 | 1.02 (0.92 - 1.12) | 0.73 | 1.04 (0.94 - 1.16) | 0.48 |
| GermanyDA | 12,256 | 2,919 | 50.1 | 16.9 | 266 | 86 | 1.13 (0.87 - 1.50) | 0.37 | 1.17 (0.89 - 1.54) | 0.28 |
| SIDIAP | 26,228 | 8,073 | 105.5 | 58.4 | 301 | 135 | 1.08 (0.85 - 1.37) | 0.54 | 1.05 (0.83 - 1.33) | 0.70 |
| Meta-analysis | 433,509 | 839,963 | 1711.0 | 3265.5 | 28,260 | 59,341 | 1.00 (0.98 - 1.02) | 0.94 | 1.00 (0.92 - 1.08) | 0.91 |
| **PS-matching / intent-to-treat TAR** | | | | | | | | | | |
| CCAE | 1,835 | 3,224 | 0.6 | 1.1 | 10 | 17 | 1.10 (0.43 - 2.66) | 0.84 | 1.09 (0.44 - 2.73) | 0.83 |
| OptumDOD | 24,356 | 61,434 | 70.0 | 189.1 | 2,431 | 6,654 | 1.00 (0.94 - 1.06) | 0.98 | 1.00 (0.94 - 1.08) | 0.87 |
| MDCR | 15,772 | 30,879 | 43.4 | 87.7 | 1,326 | 2,896 | 0.98 (0.89 - 1.07) | 0.61 | 0.97 (0.88 - 1.06) | 0.54 |
| MDCD | 1,019 | 2,194 | 2.8 | 7.2 | 144 | 295 | 1.52 (1.17 - 1.97) | <0.01 | 1.32 (1.01 - 1.71) | 0.06 |
| Open Claims | 318,926 | 670,851 | 1323.1 | 2684.7 | 21,641 | 44,867 | 1.00 (0.98 - 1.02) | 0.71 | 1.00 (0.92 - 1.10) | 0.91 |
| OptumEHR | 21,284 | 56,118 | 70.8 | 204.5 | 1,359 | 4,245 | 1.04 (0.97 - 1.13) | 0.27 | 1.09 (1.01 - 1.19) | 0.05 |
| VA | 13,173 | 73,224 | 32.9 | 315.8 | 516 | 5,431 | 1.01 (0.90 - 1.13) | 0.90 | 1.03 (0.92 - 1.16) | 0.63 |
| GermanyDA | 12,221 | 2,496 | 50.0 | 13.5 | 266 | 72 | 1.10 (0.81 - 1.51) | 0.53 | 1.12 (0.82 - 1.52) | 0.53 |
| SIDIAP | 26,107 | 6,126 | 105.3 | 40.8 | 300 | 89 | 1.08 (0.82 - 1.45) | 0.57 | 1.10 (0.82 - 1.46) | 0.55 |
| Meta-analysis | 436,058 | 909,634 | 1704.2 | 3556.9 | 28,011 | 64,618 | 1.01 (0.99 - 1.02) | 0.44 | 1.01 (0.93 - 1.09) | 0.84 |

##### 6.6.12 Joint pain

**Supplementary Table 209:** Exposures, outcomes and risk estimates for Joint pain in DPP4I (T) vs SU (C) new-user cohorts under four different propensity score (PS) adjustment / time-at-risk (TAR) designs across data sources. We report total patient exposure time in 1,000 patient-year (KYr) units, uncalibrated and calibrated hazard ratios (HRs) and their 95% confidence intervals (CIs). Greyed out entries did not pass study diagnostics and are included only for completeness.

| | Patients | | Exposure (KYr) | | Outcomes | | Uncalibrated | | Calibrated | |
|---|---|---|---|---|---|---|---|---|---|---|
| | T | C | T | C | T | C | HR (95% CI) | $p$ | HR (95% CI) | $p$ |
| **PS-stratification / on-treatment TAR** | | | | | | | | | | |
| CCAE | 1,946 | 3,238 | 0.5 | 0.9 | <5 | 6 | 0.85 (0.21 - 3.05) | 0.82 | 0.95 (0.25 - 3.60) | 0.91 |
| OptumDOD | 25,533 | 62,681 | 21.0 | 65.0 | 336 | 815 | 1.11 (0.97 - 1.27) | 0.13 | 1.12 (0.97 - 1.28) | 0.14 |
| MDCR | 17,206 | 32,465 | 17.1 | 31.8 | 187 | 289 | 1.16 (0.96 - 1.41) | 0.13 | 1.17 (0.96 - 1.42) | 0.14 |
| MDCD | 1,107 | 2,316 | 0.8 | 1.6 | 11 | 26 | 0.86 (0.39 - 1.75) | 0.68 | 0.75 (0.36 - 1.58) | 0.46 |
| Open Claims | 322,794 | 668,806 | 298.4 | 771.9 | 2,141 | 4,522 | 1.07 (1.02 - 1.13) | <0.01 | 1.07 (0.94 - 1.22) | 0.34 |
| OptumEHR | 22,153 | 55,801 | 11.4 | 33.7 | 123 | 346 | 1.11 (0.88 - 1.38) | 0.37 | 1.17 (0.94 - 1.47) | 0.19 |
| VA | 13,570 | 99,112 | 14.1 | 111.4 | 63 | 643 | 1.04 (0.76 - 1.40) | 0.79 | 1.06 (0.78 - 1.44) | 0.71 |
| GermanyDA | 10,863 | 2,683 | 11.8 | 2.9 | 228 | 64 | 0.93 (0.68 - 1.28) | 0.64 | 1.08 (0.78 - 1.50) | 0.67 |
| SIDIAP | 21,576 | 7,328 | 56.6 | 19.3 | 1,244 | 499 | 0.84 (0.74 - 0.95) | <0.01 | 0.83 (0.69 - 1.01) | 0.13 |
| Meta-analysis | 421,232 | 832,080 | 417.1 | 926.3 | 4,270 | 6,561 | 1.03 (0.92 - 1.14) | 0.63 | 1.03 (0.90 - 1.19) | 0.68 |
| **PS-matching / on-treatment TAR** | | | | | | | | | | |
| CCAE | 1,779 | 3,167 | 0.5 | 0.9 | <5 | 6 | 0.70 (0.14 - 2.91) | 0.65 | 0.72 (0.16 - 3.23) | 0.67 |
| OptumDOD | 24,737 | 62,613 | 20.3 | 64.9 | 317 | 813 | 1.11 (0.93 - 1.33) | 0.24 | 1.13 (0.95 - 1.36) | 0.20 |
| MDCR | 16,065 | 31,596 | 15.9 | 30.9 | 167 | 287 | 1.07 (0.82 - 1.40) | 0.61 | 1.03 (0.79 - 1.34) | 0.84 |
| MDCD | 1,038 | 2,299 | 0.7 | 1.6 | 10 | 26 | 1.26 (0.39 - 3.86) | 0.70 | 1.18 (0.38 - 3.71) | 0.77 |
| Open Claims | 317,021 | 668,790 | 291.2 | 771.9 | 2,046 | 4,522 | 1.04 (0.96 - 1.12) | 0.33 | 1.05 (0.94 - 1.17) | 0.42 |
| OptumEHR | 21,055 | 55,760 | 10.7 | 33.7 | 114 | 346 | 1.07 (0.80 - 1.44) | 0.63 | 1.11 (0.82 - 1.49) | 0.52 |
| VA | 12,640 | 71,549 | 13.5 | 76.6 | 61 | 437 | 0.97 (0.66 - 1.39) | 0.87 | 0.98 (0.67 - 1.42) | 0.90 |
| GermanyDA | 10,836 | 2,280 | 11.8 | 2.4 | 228 | 52 | 1.04 (0.71 - 1.57) | 0.85 | 1.07 (0.72 - 1.61) | 0.70 |
| SIDIAP | 21,398 | 5,461 | 56.4 | 13.6 | 1,243 | 382 | 0.83 (0.70 - 0.97) | 0.02 | 0.88 (0.62 - 1.24) | 0.49 |
| Meta-analysis | 424,790 | 900,348 | 420.5 | 995.6 | 4,186 | 6,865 | 1.01 (0.91 - 1.11) | 0.87 | 1.01 (0.89 - 1.15) | 0.88 |
| **PS-stratification / intent-to-treat TAR** | | | | | | | | | | |
| CCAE | 1,946 | 3,238 | 0.7 | 1.1 | 7 | 6 | 1.39 (0.45 - 4.42) | 0.58 | 1.46 (0.46 - 4.59) | 0.52 |
| OptumDOD | 25,533 | 62,681 | 75.2 | 199.9 | 1,093 | 2,453 | 1.07 (0.99 - 1.15) | 0.08 | 1.07 (0.99 - 1.16) | 0.15 |
| MDCR | 17,206 | 32,465 | 48.3 | 94.9 | 526 | 930 | 1.06 (0.94 - 1.18) | 0.35 | 1.06 (0.94 - 1.20) | 0.38 |
| MDCD | 1,107 | 2,316 | 3.2 | 7.8 | 45 | 114 | 0.93 (0.64 - 1.33) | 0.69 | 0.82 (0.57 - 1.19) | 0.31 |
| Open Claims | 322,794 | 668,806 | 1358.5 | 2715.6 | 9,473 | 17,065 | 1.02 (1.00 - 1.05) | 0.06 | 1.02 (0.93 - 1.13) | 0.64 |
| OptumEHR | 22,153 | 55,801 | 74.5 | 209.0 | 696 | 1,662 | 1.12 (1.02 - 1.23) | 0.02 | 1.17 (1.06 - 1.29) | <0.01 |
| VA | 13,570 | 99,112 | 33.3 | 528.3 | 155 | 2,805 | 1.04 (0.86 - 1.25) | 0.66 | 1.07 (0.88 - 1.29) | 0.51 |
| GermanyDA | 10,863 | 2,683 | 42.7 | 14.6 | 699 | 258 | 0.96 (0.82 - 1.13) | 0.60 | 0.99 (0.84 - 1.17) | 0.77 |
| SIDIAP | 21,576 | 7,328 | 85.4 | 48.7 | 1,883 | 1,210 | 0.91 (0.84 - 0.99) | 0.03 | 0.89 (0.81 - 0.97) | 0.02 |
| Meta-analysis | 423,299 | 835,170 | 1695.6 | 3303.3 | 14,424 | 23,716 | 1.02 (0.96 - 1.08) | 0.48 | 1.02 (0.93 - 1.12) | 0.66 |
| **PS-matching / intent-to-treat TAR** | | | | | | | | | | |
| CCAE | 1,779 | 3,167 | 0.6 | 1.1 | 6 | 6 | 1.23 (0.34 - 4.48) | 0.76 | 1.22 (0.33 - 4.46) | 0.76 |
| OptumDOD | 24,737 | 62,613 | 73.2 | 199.6 | 1,040 | 2,446 | 1.11 (1.01 - 1.22) | 0.03 | 1.11 (1.01 - 1.23) | 0.05 |
| MDCR | 16,065 | 31,596 | 45.3 | 92.4 | 480 | 921 | 1.05 (0.91 - 1.21) | 0.48 | 1.04 (0.90 - 1.20) | 0.61 |
| MDCD | 1,038 | 2,299 | 3.0 | 7.7 | 43 | 110 | 0.90 (0.57 - 1.39) | 0.65 | 0.78 (0.50 - 1.22) | 0.30 |
| Open Claims | 317,021 | 668,790 | 1332.8 | 2715.6 | 9,146 | 17,064 | 1.01 (0.98 - 1.04) | 0.49 | 1.01 (0.92 - 1.11) | 0.84 |
| OptumEHR | 21,055 | 55,760 | 71.3 | 208.8 | 653 | 1,659 | 1.13 (1.01 - 1.26) | 0.04 | 1.18 (1.05 - 1.33) | <0.01 |
| VA | 12,640 | 71,549 | 32.1 | 318.2 | 153 | 1,636 | 1.03 (0.83 - 1.26) | 0.79 | 1.05 (0.86 - 1.30) | 0.63 |
| GermanyDA | 10,836 | 2,280 | 42.7 | 11.7 | 698 | 198 | 0.91 (0.75 - 1.10) | 0.33 | 0.92 (0.75 - 1.12) | 0.46 |
| SIDIAP | 21,398 | 5,461 | 85.2 | 33.5 | 1,881 | 878 | 0.90 (0.82 - 0.99) | 0.03 | 0.91 (0.82 - 1.00) | 0.09 |
| Meta-analysis | 426,153 | 903,414 | 1690.8 | 3599.9 | 14,101 | 24,936 | 1.02 (0.95 - 1.08) | 0.64 | 1.02 (0.92 - 1.12) | 0.74 |

##### 6.6.13 Lower extremity amputation

**Supplementary Table 210:** Exposures, outcomes and risk estimates for Lower extremity amputation in DPP4I (T) vs SU (C) new-user cohorts under four different propensity score (PS) adjustment / time-at-risk (TAR) designs across data sources. We report total patient exposure time in 1,000 patient-year (KYr) units, uncalibrated and calibrated hazard ratios (HRs) and their 95% confidence intervals (CIs). Greyed out entries did not pass study diagnostics and are included only for completeness.

| | Patients | | Exposure (KYr) | | Outcomes | | Uncalibrated | | Calibrated | |
|---|---|---|---|---|---|---|---|---|---|---|
| | T | C | T | C | T | C | HR (95% CI) | $p$ | HR (95% CI) | $p$ |
| **PS-stratification / on-treatment TAR** | | | | | | | | | | |
| CCAE | 2,085 | 3,378 | 0.6 | 0.9 | <5 | <5 | 1.74 (0.07 - 44.0) | 0.74 | 1.94 (0.08 - 49.2) | 0.69 |
| OptumDOD | 27,175 | 66,157 | 22.7 | 69.1 | 17 | 94 | 0.63 (0.36 - 1.05) | 0.09 | 0.64 (0.37 - 1.09) | 0.10 |
| MDCR | 17,911 | 33,651 | 17.9 | 33.1 | 16 | 36 | 0.91 (0.48 - 1.66) | 0.77 | 0.92 (0.49 - 1.71) | 0.78 |
| MDCD | 1,205 | 2,474 | 0.8 | 1.7 | 0 | <5 | | | | |
| Open Claims | 343,958 | 706,044 | 319.2 | 815.2 | 165 | 532 | 0.90 (0.75 - 1.08) | 0.26 | 0.90 (0.72 - 1.11) | 0.33 |
| OptumEHR | 23,319 | 58,314 | 12.1 | 35.8 | 9 | 45 | 0.77 (0.34 - 1.59) | 0.51 | 0.82 (0.38 - 1.78) | 0.62 |
| VA | 15,277 | 110,322 | 15.8 | 124.5 | <10 | 93 | 0.53 (0.21 - 1.18) | 0.16 | 0.54 (0.23 - 1.29) | 0.17 |
| GermanyDA | 12,634 | 2,999 | 14.1 | 3.3 | 0 | 0 | | | | |
| SIDIAP | 26,434 | 8,082 | 70.7 | 22.6 | 23 | 8 | 2.08 (0.88 - 5.36) | 0.11 | 2.08 (0.83 - 5.18) | 0.12 |
| Meta-analysis | 438,797 | 872,248 | 442.6 | 975.7 | 230 | 715 | 0.89 (0.76 - 1.04) | 0.16 | 0.89 (0.75 - 1.07) | 0.23 |
| **PS-matching / on-treatment TAR** | | | | | | | | | | |
| CCAE | 1,893 | 3,305 | 0.5 | 0.9 | <5 | <5 | | | | |
| OptumDOD | 26,286 | 66,087 | 21.8 | 69.0 | 17 | 94 | 0.71 (0.35 - 1.32) | 0.30 | 0.72 (0.37 - 1.39) | 0.33 |
| MDCR | 16,701 | 32,770 | 16.6 | 32.2 | 16 | 36 | 1.42 (0.59 - 3.35) | 0.43 | 1.36 (0.57 - 3.26) | 0.49 |
| MDCD | 1,135 | 2,455 | 0.8 | 1.7 | 0 | <5 | | | | |
| Open Claims | 337,296 | 706,040 | 310.9 | 815.2 | 162 | 532 | 0.91 (0.71 - 1.15) | 0.44 | 0.92 (0.71 - 1.19) | 0.52 |
| OptumEHR | 22,103 | 58,271 | 11.4 | 35.7 | 8 | 45 | 0.87 (0.35 - 1.97) | 0.75 | 0.90 (0.38 - 2.13) | 0.81 |
| VA | 14,202 | 79,712 | 15.0 | 85.4 | <10 | 68 | 0.69 (0.22 - 1.80) | 0.49 | 0.70 (0.24 - 1.99) | 0.50 |
| GermanyDA | 12,597 | 2,566 | 14.0 | 2.8 | 0 | 0 | | | | |
| SIDIAP | 26,308 | 6,141 | 70.6 | 16.2 | 23 | 5 | 1.46 (0.46 - 5.52) | 0.55 | 1.55 (0.43 - 5.56) | 0.51 |
| Meta-analysis | 442,896 | 949,021 | 446.3 | 1053.8 | <236 | 780 | 0.91 (0.74 - 1.12) | 0.36 | 0.91 (0.73 - 1.14) | 0.41 |
| **PS-stratification / intent-to-treat TAR** | | | | | | | | | | |
| CCAE | 2,085 | 3,378 | 0.7 | 1.2 | <5 | <5 | 1.79 (0.07 - 45.2) | 0.72 | 1.88 (0.07 - 47.6) | 0.70 |
| OptumDOD | 27,175 | 66,157 | 82.3 | 216.1 | 113 | 372 | 0.94 (0.75 - 1.17) | 0.59 | 0.94 (0.75 - 1.18) | 0.61 |
| MDCR | 17,911 | 33,651 | 51.2 | 99.8 | 64 | 148 | 0.99 (0.73 - 1.35) | 0.97 | 1.00 (0.73 - 1.37) | 0.96 |
| MDCD | 1,205 | 2,474 | 3.6 | 8.7 | 9 | 23 | 1.06 (0.45 - 2.30) | 0.90 | 0.93 (0.41 - 2.11) | 0.86 |
| Open Claims | 343,958 | 706,044 | 1468.8 | 2902.7 | 923 | 2,585 | 0.80 (0.74 - 0.86) | <0.01 | 0.80 (0.71 - 0.90) | <0.01 |
| OptumEHR | 23,319 | 58,314 | 80.4 | 222.6 | 51 | 236 | 0.84 (0.61 - 1.15) | 0.29 | 0.88 (0.64 - 1.21) | 0.42 |
| VA | 15,277 | 110,322 | 37.3 | 596.1 | 29 | 667 | 0.70 (0.46 - 1.05) | 0.10 | 0.72 (0.47 - 1.10) | 0.13 |
| GermanyDA | 12,634 | 2,999 | 52.2 | 17.7 | 0 | 0 | | | | |
| SIDIAP | 26,434 | 8,082 | 106.5 | 58.7 | 39 | 34 | 1.26 (0.75 - 2.12) | 0.38 | 1.23 (0.73 - 2.07) | 0.44 |
| Meta-analysis | 440,002 | 874,722 | 1792.8 | 3508.5 | 1,199 | 3,398 | 0.88 (0.78 - 1.00) | 0.06 | 0.88 (0.76 - 1.02) | 0.10 |
| **PS-matching / intent-to-treat TAR** | | | | | | | | | | |
| CCAE | 1,893 | 3,305 | 0.6 | 1.1 | <5 | <5 | | | | |
| OptumDOD | 26,286 | 66,087 | 79.9 | 215.8 | 113 | 371 | 0.89 (0.68 - 1.15) | 0.38 | 0.89 (0.68 - 1.17) | 0.41 |
| MDCR | 16,701 | 32,770 | 48.0 | 97.2 | 62 | 144 | 1.05 (0.70 - 1.55) | 0.82 | 1.04 (0.69 - 1.55) | 0.86 |
| MDCD | 1,135 | 2,455 | 3.4 | 8.6 | 9 | 23 | 1.03 (0.34 - 2.88) | 0.96 | 0.89 (0.31 - 2.60) | 0.83 |
| Open Claims | 337,296 | 706,040 | 1438.4 | 2902.7 | 911 | 2,585 | 0.82 (0.75 - 0.90) | <0.01 | 0.82 (0.73 - 0.93) | <0.01 |
| OptumEHR | 22,103 | 58,271 | 76.7 | 222.3 | 48 | 235 | 0.85 (0.58 - 1.22) | 0.39 | 0.89 (0.61 - 1.30) | 0.55 |
| VA | 14,202 | 79,712 | 36.0 | 356.6 | 29 | 384 | 0.64 (0.38 - 1.02) | 0.08 | 0.65 (0.40 - 1.07) | 0.10 |
| GermanyDA | 12,597 | 2,566 | 52.1 | 14.1 | 0 | 0 | | | | |
| SIDIAP | 26,308 | 6,141 | 106.4 | 41.1 | 39 | 16 | 1.56 (0.85 - 2.99) | 0.17 | 1.57 (0.84 - 2.96) | 0.16 |
| Meta-analysis | 444,031 | 951,476 | 1788.7 | 3844.3 | 1,211 | 3,758 | 0.84 (0.78 - 0.91) | <0.01 | 0.84 (0.76 - 0.94) | <0.01 |

##### 6.6.14 Nausea

**Supplementary Table 211:** Exposures, outcomes and risk estimates for Nausea in DPP4I (T) vs SU (C) new-user cohorts under four different propensity score (PS) adjustment / time-at-risk (TAR) designs across data sources. We report total patient exposure time in 1,000 patient-year (KYr) units, uncalibrated and calibrated hazard ratios (HRs) and their 95% confidence intervals (CIs). Greyed out entries did not pass study diagnostics and are included only for completeness.

| | Patients | | Exposure (KYr) | | Outcomes | | Uncalibrated | | Calibrated | |
|---|---|---|---|---|---|---|---|---|---|---|
| | T | C | T | C | T | C | HR (95% CI) | $p$ | HR (95% CI) | $p$ |
| **PS-stratification / on-treatment TAR** | | | | | | | | | | |
| CCAE | 1,852 | 3,051 | 0.5 | 0.8 | 16 | 24 | 1.04 (0.53 - 1.99) | 0.90 | 1.17 (0.60 - 2.26) | 0.66 |
| OptumDOD | 23,106 | 56,570 | 18.8 | 57.8 | 854 | 2,589 | 0.94 (0.86 - 1.02) | 0.12 | 0.95 (0.87 - 1.03) | 0.29 |
| MDCR | 15,971 | 29,913 | 15.8 | 29.0 | 541 | 1,072 | 0.90 (0.81 - 1.00) | 0.05 | 0.90 (0.81 - 1.01) | 0.11 |
| MDCD | 911 | 1,956 | 0.6 | 1.3 | 45 | 76 | 1.21 (0.81 - 1.77) | 0.34 | 1.06 (0.71 - 1.57) | 0.77 |
| Open Claims | 297,457 | 614,955 | 274.0 | 708.6 | 6,039 | 14,967 | 0.97 (0.94 - 1.00) | 0.06 | 0.97 (0.85 - 1.09) | 0.59 |
| OptumEHR | 21,214 | 53,261 | 10.8 | 31.4 | 372 | 1,194 | 0.94 (0.83 - 1.06) | 0.31 | 0.99 (0.87 - 1.13) | 0.84 |
| VA | 14,379 | 104,723 | 14.9 | 117.7 | 109 | 966 | 1.05 (0.83 - 1.32) | 0.70 | 1.07 (0.84 - 1.35) | 0.60 |
| GermanyDA | 11,730 | 2,799 | 13.0 | 3.1 | 140 | 36 | 1.05 (0.69 - 1.62) | 0.83 | 1.22 (0.79 - 1.88) | 0.42 |
| SIDIAP | 25,547 | 7,960 | 68.2 | 22.1 | 531 | 127 | 1.04 (0.83 - 1.31) | 0.74 | 1.04 (0.79 - 1.36) | 0.81 |
| Meta-analysis | 397,788 | 770,465 | 401.5 | 854.1 | 8,538 | 20,085 | 0.96 (0.94 - 0.99) | <0.01 | 0.97 (0.88 - 1.06) | 0.47 |
| **PS-matching / on-treatment TAR** | | | | | | | | | | |
| CCAE | 1,680 | 2,983 | 0.5 | 0.8 | 15 | 24 | 0.93 (0.43 - 1.93) | 0.85 | 0.95 (0.45 - 2.00) | 0.80 |
| OptumDOD | 22,321 | 56,486 | 18.1 | 57.7 | 817 | 2,583 | 0.95 (0.85 - 1.06) | 0.35 | 0.97 (0.87 - 1.08) | 0.61 |
| MDCR | 14,919 | 28,985 | 14.7 | 28.1 | 496 | 1,055 | 0.96 (0.82 - 1.12) | 0.58 | 0.92 (0.79 - 1.07) | 0.34 |
| MDCD | 881 | 1,941 | 0.6 | 1.3 | 41 | 74 | 1.30 (0.74 - 2.25) | 0.36 | 1.22 (0.70 - 2.13) | 0.50 |
| Open Claims | 291,664 | 614,935 | 267.0 | 708.6 | 5,835 | 14,967 | 0.97 (0.93 - 1.01) | 0.13 | 0.98 (0.89 - 1.08) | 0.68 |
| OptumEHR | 20,090 | 53,221 | 10.0 | 31.4 | 350 | 1,191 | 0.98 (0.84 - 1.15) | 0.82 | 1.01 (0.86 - 1.19) | 0.82 |
| VA | 13,385 | 75,977 | 14.2 | 81.4 | 102 | 662 | 1.05 (0.78 - 1.39) | 0.75 | 1.06 (0.79 - 1.41) | 0.73 |
| GermanyDA | 11,697 | 2,384 | 12.9 | 2.6 | 139 | 29 | 1.27 (0.75 - 2.22) | 0.39 | 1.31 (0.76 - 2.25) | 0.39 |
| SIDIAP | 25,436 | 6,027 | 68.1 | 15.8 | 530 | 97 | 1.00 (0.75 - 1.34) | 1.00 | 1.06 (0.70 - 1.62) | 0.78 |
| Meta-analysis | 402,073 | 842,939 | 406.0 | 927.7 | 8,325 | 20,682 | 0.97 (0.94 - 1.01) | 0.10 | 0.97 (0.89 - 1.07) | 0.54 |
| **PS-stratification / intent-to-treat TAR** | | | | | | | | | | |
| CCAE | 1,852 | 3,051 | 0.6 | 1.1 | 22 | 34 | 1.08 (0.61 - 1.88) | 0.78 | 1.14 (0.65 - 2.00) | 0.66 |
| OptumDOD | 23,106 | 56,570 | 64.7 | 167.8 | 2,823 | 7,645 | 0.92 (0.88 - 0.96) | <0.01 | 0.92 (0.87 - 0.97) | 0.02 |
| MDCR | 15,971 | 29,913 | 42.8 | 82.8 | 1,701 | 3,496 | 0.93 (0.87 - 0.99) | 0.02 | 0.93 (0.86 - 1.01) | 0.15 |
| MDCD | 911 | 1,956 | 2.3 | 5.8 | 166 | 362 | 1.18 (0.97 - 1.43) | 0.10 | 1.04 (0.85 - 1.26) | 0.73 |
| Open Claims | 297,457 | 614,955 | 1210.4 | 2410.2 | 28,492 | 57,566 | 0.96 (0.95 - 0.97) | <0.01 | 0.96 (0.87 - 1.05) | 0.38 |
| OptumEHR | 21,214 | 53,261 | 68.9 | 190.8 | 1,780 | 5,087 | 0.98 (0.93 - 1.04) | 0.48 | 1.02 (0.96 - 1.09) | 0.59 |
| VA | 14,379 | 104,723 | 34.9 | 556.1 | 287 | 4,944 | 0.98 (0.85 - 1.12) | 0.76 | 1.00 (0.87 - 1.15) | 0.91 |
| GermanyDA | 11,730 | 2,799 | 47.1 | 15.8 | 495 | 199 | 0.94 (0.78 - 1.13) | 0.50 | 0.97 (0.80 - 1.17) | 0.73 |
| SIDIAP | 25,547 | 7,960 | 102.4 | 56.6 | 888 | 481 | 0.91 (0.80 - 1.04) | 0.16 | 0.89 (0.78 - 1.01) | 0.11 |
| Meta-analysis | 397,788 | 770,465 | 1539.2 | 2930.9 | 36,367 | 74,870 | 0.95 (0.93 - 0.97) | <0.01 | 0.95 (0.88 - 1.03) | 0.22 |
| **PS-matching / intent-to-treat TAR** | | | | | | | | | | |
| CCAE | 1,680 | 2,983 | 0.6 | 1.0 | 21 | 34 | 0.94 (0.50 - 1.73) | 0.84 | 0.93 (0.50 - 1.75) | 0.78 |
| OptumDOD | 22,321 | 56,486 | 62.7 | 167.5 | 2,718 | 7,625 | 0.93 (0.88 - 0.98) | <0.01 | 0.93 (0.87 - 0.99) | 0.07 |
| MDCR | 14,919 | 28,985 | 40.1 | 80.2 | 1,590 | 3,431 | 0.95 (0.87 - 1.02) | 0.17 | 0.94 (0.86 - 1.02) | 0.18 |
| MDCD | 881 | 1,941 | 2.2 | 5.8 | 160 | 354 | 1.18 (0.93 - 1.50) | 0.17 | 1.02 (0.81 - 1.30) | 0.80 |
| Open Claims | 291,664 | 614,935 | 1185.5 | 2410.2 | 27,789 | 57,565 | 0.96 (0.95 - 0.98) | <0.01 | 0.96 (0.88 - 1.05) | 0.41 |
| OptumEHR | 20,090 | 53,221 | 65.6 | 190.6 | 1,689 | 5,079 | 0.99 (0.93 - 1.06) | 0.81 | 1.04 (0.96 - 1.12) | 0.38 |
| VA | 13,385 | 75,977 | 33.7 | 336.9 | 275 | 2,961 | 0.99 (0.85 - 1.15) | 0.88 | 1.01 (0.87 - 1.18) | 0.87 |
| GermanyDA | 11,697 | 2,384 | 47.0 | 12.6 | 493 | 154 | 0.89 (0.71 - 1.11) | 0.29 | 0.90 (0.72 - 1.12) | 0.40 |
| SIDIAP | 25,436 | 6,027 | 102.3 | 39.6 | 887 | 337 | 0.87 (0.75 - 1.02) | 0.08 | 0.88 (0.75 - 1.03) | 0.13 |
| Meta-analysis | 402,073 | 842,939 | 1539.8 | 3244.4 | 35,622 | 77,540 | 0.96 (0.95 - 0.98) | <0.01 | 0.96 (0.89 - 1.04) | 0.31 |

##### 6.6.15 Peripheral edema

**Supplementary Table 212:** Exposures, outcomes and risk estimates for Peripheral edema in DPP4I (T) vs SU (C) new-user cohorts under four different propensity score (PS) adjustment / time-at-risk (TAR) designs across data sources. We report total patient exposure time in 1,000 patient-year (KYr) units, uncalibrated and calibrated hazard ratios (HRs) and their 95% confidence intervals (CIs). Greyed out entries did not pass study diagnostics and are included only for completeness.

| | Patients | | Exposure (KYr) | | Outcomes | | Uncalibrated | | Calibrated | |
|---|---|---|---|---|---|---|---|---|---|---|
| | T | C | T | C | T | C | HR (95% CI) | $p$ | HR (95% CI) | $p$ |
| **PS-stratification / on-treatment TAR** | | | | | | | | | | |
| CCAE | 1,647 | 2,733 | 0.4 | 0.7 | 22 | 55 | 0.64 (0.38 - 1.07) | 0.10 | 0.72 (0.43 - 1.22) | 0.24 |
| OptumDOD | 19,396 | 48,325 | 15.0 | 46.6 | 1,589 | 4,879 | 0.96 (0.90 - 1.02) | 0.16 | 0.97 (0.90 - 1.03) | 0.44 |
| MDCR | 13,304 | 25,259 | 12.6 | 23.2 | 1,091 | 2,210 | 0.93 (0.86 - 1.01) | 0.07 | 0.94 (0.86 - 1.01) | 0.19 |
| MDCD | 717 | 1,662 | 0.5 | 1.0 | 56 | 139 | 0.88 (0.63 - 1.22) | 0.46 | 0.77 (0.55 - 1.08) | 0.17 |
| Open Claims | 250,191 | 526,163 | 224.7 | 589.8 | 10,726 | 28,111 | 0.95 (0.93 - 0.97) | <0.01 | 0.94 (0.84 - 1.07) | 0.37 |
| OptumEHR | 18,904 | 47,411 | 9.2 | 26.3 | 603 | 2,271 | 0.83 (0.75 - 0.91) | <0.01 | 0.88 (0.79 - 0.97) | 0.04 |
| VA | 11,571 | 87,019 | 11.8 | 94.1 | 444 | 4,083 | 0.85 (0.76 - 0.95) | <0.01 | 0.87 (0.77 - 0.98) | 0.02 |
| GermanyDA | 10,333 | 2,524 | 11.3 | 2.6 | 407 | 117 | 0.76 (0.60 - 0.96) | 0.02 | 0.89 (0.69 - 1.14) | 0.48 |
| SIDIAP | 23,708 | 7,689 | 62.3 | 20.8 | 1,353 | 373 | 0.89 (0.78 - 1.02) | 0.10 | 0.89 (0.73 - 1.09) | 0.35 |
| Meta-analysis | 338,200 | 661,766 | 335.9 | 711.1 | 15,847 | 38,155 | 0.91 (0.86 - 0.96) | <0.01 | 0.91 (0.82 - 1.01) | 0.08 |
| **PS-matching / on-treatment TAR** | | | | | | | | | | |
| CCAE | 1,510 | 2,673 | 0.4 | 0.7 | 19 | 54 | 0.59 (0.29 - 1.13) | 0.13 | 0.60 (0.30 - 1.19) | 0.17 |
| OptumDOD | 18,789 | 48,260 | 14.5 | 46.5 | 1,526 | 4,872 | 0.95 (0.88 - 1.03) | 0.22 | 0.97 (0.90 - 1.05) | 0.55 |
| MDCR | 12,380 | 24,417 | 11.6 | 22.4 | 1,000 | 2,150 | 0.91 (0.82 - 1.01) | 0.07 | 0.87 (0.78 - 0.97) | 0.03 |
| MDCD | 692 | 1,646 | 0.4 | 1.0 | 53 | 138 | 0.87 (0.55 - 1.35) | 0.54 | 0.82 (0.52 - 1.28) | 0.42 |
| Open Claims | 245,855 | 526,162 | 219.6 | 589.8 | 10,382 | 28,111 | 0.93 (0.90 - 0.96) | <0.01 | 0.94 (0.86 - 1.03) | 0.20 |
| OptumEHR | 17,871 | 47,359 | 8.5 | 26.2 | 566 | 2,266 | 0.84 (0.75 - 0.95) | <0.01 | 0.87 (0.77 - 0.99) | 0.06 |
| VA | 10,786 | 61,870 | 11.2 | 63.7 | 416 | 2,806 | 0.82 (0.70 - 0.94) | <0.01 | 0.82 (0.71 - 0.96) | 0.02 |
| GermanyDA | 10,309 | 2,163 | 11.2 | 2.2 | 406 | 107 | 0.79 (0.60 - 1.06) | 0.11 | 0.82 (0.61 - 1.10) | 0.28 |
| SIDIAP | 23,568 | 5,788 | 62.2 | 14.8 | 1,346 | 293 | 0.88 (0.74 - 1.04) | 0.13 | 0.93 (0.65 - 1.32) | 0.71 |
| Meta-analysis | 341,760 | 720,338 | 339.8 | 767.4 | 15,714 | 40,797 | 0.91 (0.87 - 0.94) | <0.01 | 0.91 (0.83 - 1.00) | 0.04 |
| **PS-stratification / intent-to-treat TAR** | | | | | | | | | | |
| CCAE | 1,647 | 2,733 | 0.6 | 0.9 | 30 | 75 | 0.66 (0.42 - 1.02) | 0.07 | 0.70 (0.45 - 1.09) | 0.14 |
| OptumDOD | 19,396 | 48,325 | 48.4 | 128.5 | 4,665 | 12,468 | 0.97 (0.94 - 1.01) | 0.13 | 0.97 (0.93 - 1.02) | 0.41 |
| MDCR | 13,304 | 25,259 | 32.4 | 62.8 | 3,001 | 6,171 | 0.94 (0.90 - 0.98) | <0.01 | 0.94 (0.88 - 1.01) | 0.17 |
| MDCD | 717 | 1,662 | 1.6 | 4.7 | 195 | 510 | 1.02 (0.85 - 1.21) | 0.83 | 0.90 (0.75 - 1.07) | 0.30 |
| Open Claims | 250,191 | 526,163 | 966.3 | 1947.9 | 45,308 | 94,615 | 0.96 (0.94 - 0.97) | <0.01 | 0.95 (0.87 - 1.05) | 0.33 |
| OptumEHR | 18,904 | 47,411 | 57.6 | 155.1 | 2,998 | 9,370 | 0.92 (0.88 - 0.96) | <0.01 | 0.96 (0.91 - 1.01) | 0.17 |
| VA | 11,571 | 87,019 | 27.2 | 425.4 | 984 | 17,311 | 0.91 (0.85 - 0.98) | 0.01 | 0.93 (0.86 - 1.01) | 0.12 |
| GermanyDA | 10,333 | 2,524 | 40.0 | 13.2 | 1,337 | 504 | 0.91 (0.81 - 1.02) | 0.12 | 0.94 (0.84 - 1.06) | 0.41 |
| SIDIAP | 23,708 | 7,689 | 93.1 | 52.1 | 2,120 | 1,136 | 0.94 (0.87 - 1.03) | 0.18 | 0.92 (0.84 - 1.01) | 0.12 |
| Meta-analysis | 340,178 | 664,732 | 1247.3 | 2377.3 | 59,708 | 124,957 | 0.95 (0.94 - 0.97) | <0.01 | 0.95 (0.88 - 1.03) | 0.20 |
| **PS-matching / intent-to-treat TAR** | | | | | | | | | | |
| CCAE | 1,510 | 2,673 | 0.5 | 0.9 | 27 | 73 | 0.83 (0.48 - 1.40) | 0.50 | 0.83 (0.49 - 1.41) | 0.52 |
| OptumDOD | 18,789 | 48,260 | 47.0 | 128.3 | 4,510 | 12,436 | 0.98 (0.94 - 1.02) | 0.32 | 0.98 (0.93 - 1.04) | 0.61 |
| MDCR | 12,380 | 24,417 | 30.3 | 60.6 | 2,783 | 6,039 | 0.92 (0.86 - 0.97) | <0.01 | 0.91 (0.85 - 0.97) | 0.02 |
| MDCD | 692 | 1,646 | 1.6 | 4.6 | 187 | 503 | 0.93 (0.75 - 1.15) | 0.49 | 0.80 (0.65 - 1.00) | 0.07 |
| Open Claims | 245,855 | 526,162 | 948.8 | 1947.9 | 44,234 | 94,615 | 0.96 (0.94 - 0.97) | <0.01 | 0.96 (0.88 - 1.04) | 0.32 |
| OptumEHR | 17,871 | 47,359 | 54.8 | 154.9 | 2,840 | 9,348 | 0.91 (0.86 - 0.95) | <0.01 | 0.95 (0.89 - 1.01) | 0.18 |
| VA | 10,786 | 61,870 | 26.3 | 255.2 | 941 | 10,224 | 0.89 (0.81 - 0.96) | <0.01 | 0.91 (0.83 - 0.99) | 0.06 |
| GermanyDA | 10,309 | 2,163 | 39.9 | 10.6 | 1,334 | 405 | 0.88 (0.77 - 1.01) | 0.07 | 0.89 (0.77 - 1.02) | 0.19 |
| SIDIAP | 23,568 | 5,788 | 93.0 | 36.3 | 2,112 | 800 | 0.95 (0.86 - 1.05) | 0.32 | 0.96 (0.87 - 1.06) | 0.50 |
| Meta-analysis | 343,065 | 723,282 | 1247.1 | 2611.1 | 59,011 | 134,550 | 0.94 (0.91 - 0.96) | <0.01 | 0.94 (0.87 - 1.02) | 0.11 |

##### 6.6.16 Photosensitivity

**Supplementary Table 213:** Exposures, outcomes and risk estimates for Photosensitivity in DPP4I (T) vs SU (C) new-user cohorts under four different propensity score (PS) adjustment / time-at-risk (TAR) designs across data sources. We report total patient exposure time in 1,000 patient-year (KYr) units, uncalibrated and calibrated hazard ratios (HRs) and their 95% confidence intervals (CIs). Greyed out entries did not pass study diagnostics and are included only for completeness.

| | Patients | | Exposure (KYr) | | Outcomes | | Uncalibrated | | Calibrated | |
|---|---|---|---|---|---|---|---|---|---|---|
| | T | C | T | C | T | C | HR (95% CI) | $p$ | HR (95% CI) | $p$ |
| **PS-stratification / on-treatment TAR** | | | | | | | | | | |
| CCAE | 2,057 | 3,331 | 0.6 | 0.9 | <5 | <5 | 0.64 (0.09 - 3.52) | 0.64 | 0.72 (0.11 - 4.60) | 0.73 |
| OptumDOD | 26,869 | 65,494 | 22.3 | 68.2 | 39 | 122 | 1.14 (0.77 - 1.66) | 0.49 | 1.15 (0.79 - 1.69) | 0.48 |
| MDCR | 17,593 | 33,139 | 17.5 | 32.5 | 71 | 105 | 1.24 (0.90 - 1.70) | 0.18 | 1.24 (0.90 - 1.71) | 0.19 |
| MDCD | 1,205 | 2,471 | 0.8 | 1.7 | 0 | 0 | | | | |
| Open Claims | 339,369 | 698,040 | 314.5 | 804.6 | 193 | 433 | 1.09 (0.92 - 1.30) | 0.32 | 1.09 (0.88 - 1.34) | 0.44 |
| OptumEHR | 23,122 | 57,750 | 12.0 | 35.3 | 22 | 84 | 0.97 (0.57 - 1.57) | 0.90 | 1.03 (0.62 - 1.70) | 0.92 |
| VA | 15,025 | 108,925 | 15.5 | 122.6 | <10 | 204 | 0.50 (0.15 - 1.30) | 0.22 | 0.51 (0.17 - 1.52) | 0.23 |
| GermanyDA | 12,586 | 2,992 | 14.0 | 3.3 | <5 | <5 | 0.90 (0.10 - 8.16) | 0.92 | 1.05 (0.12 - 9.54) | 0.95 |
| SIDIAP | 26,410 | 8,090 | 70.7 | 22.6 | 8 | <5 | 0.59 (0.16 - 2.68) | 0.47 | 0.59 (0.14 - 2.45) | 0.47 |
| Meta-analysis | 406,953 | 854,423 | 366.4 | 940.6 | 325 | 744 | 1.12 (0.97 - 1.28) | 0.12 | 1.12 (0.95 - 1.32) | 0.18 |
| **PS-matching / on-treatment TAR** | | | | | | | | | | |
| CCAE | 1,867 | 3,259 | 0.5 | 0.9 | <5 | <5 | 0.50 (0.02 - 5.22) | 0.62 | 0.51 (0.03 - 7.62) | 0.63 |
| OptumDOD | 25,998 | 65,423 | 21.5 | 68.1 | 36 | 122 | 1.24 (0.75 - 2.01) | 0.39 | 1.26 (0.77 - 2.07) | 0.36 |
| MDCR | 16,401 | 32,260 | 16.2 | 31.6 | 66 | 102 | 1.38 (0.91 - 2.10) | 0.13 | 1.33 (0.87 - 2.02) | 0.19 |
| MDCD | 1,138 | 2,452 | 0.8 | 1.7 | 0 | 0 | | | | |
| Open Claims | 332,799 | 698,036 | 306.4 | 804.6 | 188 | 433 | 1.11 (0.88 - 1.39) | 0.37 | 1.12 (0.88 - 1.43) | 0.36 |
| OptumEHR | 21,924 | 57,708 | 11.2 | 35.3 | 22 | 84 | 0.89 (0.47 - 1.63) | 0.72 | 0.92 (0.49 - 1.72) | 0.80 |
| VA | 13,973 | 78,397 | 14.8 | 83.7 | <10 | 128 | 0.83 (0.24 - 2.18) | 0.74 | 0.84 (0.28 - 2.53) | 0.75 |
| GermanyDA | 12,549 | 2,560 | 14.0 | 2.8 | <5 | <5 | | | | |
| SIDIAP | 26,284 | 6,139 | 70.6 | 16.2 | 8 | <5 | 0.50 (0.09 - 2.85) | 0.44 | 0.53 (0.09 - 3.11) | 0.48 |
| Meta-analysis | 411,095 | 931,824 | 370.2 | 1023.3 | <322 | 869 | 1.14 (0.96 - 1.36) | 0.14 | 1.14 (0.94 - 1.39) | 0.18 |
| **PS-stratification / intent-to-treat TAR** | | | | | | | | | | |
| CCAE | 2,057 | 3,331 | 0.7 | 1.2 | <5 | <5 | 0.66 (0.09 - 3.64) | 0.66 | 0.69 (0.11 - 4.47) | 0.70 |
| OptumDOD | 26,869 | 65,494 | 80.8 | 212.8 | 99 | 305 | 0.95 (0.75 - 1.20) | 0.68 | 0.95 (0.75 - 1.21) | 0.69 |
| MDCR | 17,593 | 33,139 | 50.0 | 97.6 | 155 | 315 | 0.97 (0.79 - 1.19) | 0.79 | 0.98 (0.79 - 1.20) | 0.84 |
| MDCD | 1,205 | 2,471 | 3.6 | 8.7 | <5 | 7 | 0.54 (0.07 - 2.54) | 0.49 | 0.47 (0.08 - 2.79) | 0.41 |
| Open Claims | 339,369 | 698,040 | 1447.4 | 2869.1 | 505 | 994 | 1.02 (0.92 - 1.14) | 0.67 | 1.02 (0.89 - 1.18) | 0.76 |
| OptumEHR | 23,122 | 57,750 | 79.5 | 219.8 | 117 | 327 | 1.26 (1.00 - 1.58) | 0.04 | 1.32 (1.05 - 1.66) | 0.02 |
| VA | 15,025 | 108,925 | 36.7 | 586.7 | 11 | 685 | 0.84 (0.42 - 1.52) | 0.60 | 0.86 (0.45 - 1.64) | 0.65 |
| GermanyDA | 12,586 | 2,992 | 51.9 | 17.6 | 16 | 6 | 1.35 (0.49 - 4.16) | 0.58 | 1.40 (0.48 - 4.09) | 0.54 |
| SIDIAP | 26,410 | 8,090 | 106.5 | 58.8 | 14 | 7 | 0.95 (0.33 - 2.93) | 0.93 | 0.93 (0.31 - 2.76) | 0.89 |
| Meta-analysis | 406,953 | 854,423 | 1657.8 | 3399.4 | 876 | 1,941 | 1.04 (0.95 - 1.12) | 0.41 | 1.04 (0.93 - 1.16) | 0.54 |
| **PS-matching / intent-to-treat TAR** | | | | | | | | | | |
| CCAE | 1,867 | 3,259 | 0.6 | 1.1 | <5 | <5 | 0.76 (0.10 - 4.65) | 0.78 | 0.75 (0.11 - 5.17) | 0.78 |
| OptumDOD | 25,998 | 65,423 | 78.5 | 212.5 | 95 | 304 | 1.07 (0.81 - 1.40) | 0.63 | 1.07 (0.82 - 1.41) | 0.61 |
| MDCR | 16,401 | 32,260 | 46.8 | 95.1 | 144 | 309 | 1.06 (0.83 - 1.36) | 0.64 | 1.05 (0.82 - 1.35) | 0.71 |
| MDCD | 1,138 | 2,452 | 3.4 | 8.6 | <5 | 7 | | | | |
| Open Claims | 332,799 | 698,036 | 1417.6 | 2869.1 | 494 | 994 | 1.05 (0.93 - 1.18) | 0.43 | 1.05 (0.91 - 1.21) | 0.53 |
| OptumEHR | 21,924 | 57,708 | 75.8 | 219.5 | 114 | 327 | 1.18 (0.90 - 1.53) | 0.21 | 1.24 (0.95 - 1.61) | 0.12 |
| VA | 13,973 | 78,397 | 35.4 | 349.4 | 10 | 334 | 0.65 (0.29 - 1.27) | 0.26 | 0.66 (0.31 - 1.40) | 0.28 |
| GermanyDA | 12,549 | 2,560 | 51.8 | 14.1 | 16 | <5 | 1.32 (0.42 - 5.01) | 0.66 | 1.33 (0.38 - 4.63) | 0.65 |
| SIDIAP | 26,284 | 6,139 | 106.4 | 41.1 | 14 | <5 | 1.11 (0.35 - 4.21) | 0.87 | 1.12 (0.32 - 3.88) | 0.86 |
| Meta-analysis | 411,095 | 931,824 | 1654.0 | 3745.6 | 857 | 2,268 | 1.06 (0.97 - 1.16) | 0.21 | 1.06 (0.94 - 1.19) | 0.32 |

##### 6.6.17 Venous thromboembolism

**Supplementary Table 214:** Exposures, outcomes and risk estimates for Venous thromboembolism in DPP4I (T) vs SU (C) new-user cohorts under four different propensity score (PS) adjustment / time-at-risk (TAR) designs across data sources. We report total patient exposure time in 1,000 patient-year (KYr) units, uncalibrated and calibrated hazard ratios (HRs) and their 95% confidence intervals (CIs). Greyed out entries did not pass study diagnostics and are included only for completeness.

| | Patients | | Exposure (KYr) | | Outcomes | | Uncalibrated | | Calibrated | |
|---|---|---|---|---|---|---|---|---|---|---|
| | T | C | T | C | T | C | HR (95% CI) | $p$ | HR (95% CI) | $p$ |
| **PS-stratification / on-treatment TAR** | | | | | | | | | | |
| CCAE | 2,016 | 3,284 | 0.6 | 0.9 | 7 | 7 | 1.81 (0.61 - 5.42) | 0.29 | 2.03 (0.68 - 6.07) | 0.21 |
| OptumDOD | 26,091 | 63,541 | 21.6 | 65.8 | 313 | 999 | 0.97 (0.84 - 1.10) | 0.61 | 0.97 (0.85 - 1.12) | 0.72 |
| MDCR | 17,192 | 32,221 | 17.1 | 31.5 | 229 | 518 | 0.82 (0.70 - 0.97) | 0.02 | 0.83 (0.70 - 0.98) | 0.03 |
| MDCD | 1,140 | 2,364 | 0.8 | 1.6 | 18 | 27 | 1.66 (0.85 - 3.14) | 0.13 | 1.46 (0.76 - 2.80) | 0.28 |
| Open Claims | 329,000 | 676,714 | 304.5 | 779.1 | 2,221 | 6,397 | 0.87 (0.83 - 0.91) | <0.01 | 0.87 (0.76 - 0.99) | 0.03 |
| OptumEHR | 22,532 | 56,350 | 11.6 | 34.0 | 116 | 497 | 0.76 (0.62 - 0.94) | 0.01 | 0.81 (0.65 - 1.01) | 0.08 |
| VA | 14,702 | 107,070 | 15.1 | 120.3 | 84 | 717 | 0.95 (0.72 - 1.23) | 0.69 | 0.96 (0.73 - 1.26) | 0.79 |
| GermanyDA | 11,576 | 2,782 | 12.8 | 3.1 | 130 | 36 | 0.84 (0.56 - 1.28) | 0.41 | 0.98 (0.64 - 1.49) | 0.81 |
| SIDIAP | 25,562 | 7,942 | 68.3 | 22.1 | 358 | 96 | 1.05 (0.81 - 1.36) | 0.74 | 1.04 (0.77 - 1.41) | 0.81 |
| Meta-analysis | 433,093 | 841,914 | 436.8 | 937.2 | 3,385 | 8,570 | 0.88 (0.83 - 0.93) | <0.01 | 0.89 (0.80 - 0.99) | 0.03 |
| **PS-matching / on-treatment TAR** | | | | | | | | | | |
| CCAE | 1,830 | 3,214 | 0.5 | 0.9 | 7 | 7 | 2.79 (0.65 - 14.1) | 0.19 | 2.83 (0.61 - 13.2) | 0.19 |
| OptumDOD | 25,247 | 63,481 | 20.9 | 65.7 | 303 | 997 | 0.94 (0.79 - 1.12) | 0.52 | 0.96 (0.81 - 1.15) | 0.68 |
| MDCR | 16,035 | 31,196 | 15.9 | 30.5 | 217 | 506 | 0.82 (0.66 - 1.03) | 0.10 | 0.79 (0.63 - 1.00) | 0.06 |
| MDCD | 1,078 | 2,346 | 0.7 | 1.6 | 18 | 27 | 2.80 (1.14 - 7.04) | 0.03 | 2.64 (1.06 - 6.57) | 0.04 |
| Open Claims | 322,630 | 676,711 | 296.6 | 779.1 | 2,155 | 6,397 | 0.89 (0.83 - 0.95) | <0.01 | 0.90 (0.81 - 1.00) | 0.06 |
| OptumEHR | 21,336 | 56,310 | 10.9 | 34.0 | 114 | 495 | 0.74 (0.57 - 0.96) | 0.03 | 0.77 (0.59 - 1.00) | 0.06 |
| VA | 13,685 | 76,974 | 14.4 | 82.0 | 81 | 484 | 1.17 (0.84 - 1.62) | 0.35 | 1.18 (0.85 - 1.64) | 0.35 |
| GermanyDA | 11,547 | 2,382 | 12.8 | 2.6 | 129 | 32 | 0.68 (0.41 - 1.16) | 0.15 | 0.71 (0.42 - 1.19) | 0.23 |
| SIDIAP | 25,424 | 6,006 | 68.2 | 15.9 | 356 | 75 | 1.26 (0.90 - 1.81) | 0.19 | 1.34 (0.84 - 2.14) | 0.24 |
| Meta-analysis | 436,982 | 915,406 | 440.3 | 1011.4 | 3,373 | 9,013 | 0.90 (0.85 - 0.95) | <0.01 | 0.90 (0.81 - 1.00) | 0.04 |
| **PS-stratification / intent-to-treat TAR** | | | | | | | | | | |
| CCAE | 2,016 | 3,284 | 0.7 | 1.1 | 9 | 11 | 1.34 (0.53 - 3.32) | 0.53 | 1.41 (0.56 - 3.56) | 0.47 |
| OptumDOD | 26,091 | 63,541 | 77.4 | 202.1 | 1,102 | 3,186 | 0.92 (0.86 - 0.99) | 0.02 | 0.92 (0.85 - 1.00) | 0.07 |
| MDCR | 17,192 | 32,221 | 48.1 | 93.0 | 758 | 1,762 | 0.86 (0.79 - 0.94) | <0.01 | 0.87 (0.78 - 0.96) | 0.01 |
| MDCD | 1,140 | 2,364 | 3.2 | 8.1 | 62 | 139 | 1.19 (0.86 - 1.63) | 0.28 | 1.05 (0.76 - 1.45) | 0.77 |
| Open Claims | 329,000 | 676,714 | 1387.5 | 2744.5 | 11,435 | 24,885 | 0.92 (0.90 - 0.94) | <0.01 | 0.92 (0.83 - 1.01) | 0.07 |
| OptumEHR | 22,532 | 56,350 | 76.3 | 210.8 | 642 | 2,057 | 0.93 (0.84 - 1.02) | 0.10 | 0.96 (0.87 - 1.06) | 0.49 |
| VA | 14,702 | 107,070 | 35.9 | 572.3 | 226 | 4,025 | 0.93 (0.79 - 1.08) | 0.32 | 0.95 (0.81 - 1.11) | 0.52 |
| GermanyDA | 11,576 | 2,782 | 46.6 | 15.7 | 485 | 165 | 1.09 (0.90 - 1.34) | 0.39 | 1.13 (0.92 - 1.38) | 0.27 |
| SIDIAP | 25,562 | 7,942 | 102.6 | 57.0 | 561 | 324 | 0.93 (0.79 - 1.09) | 0.37 | 0.91 (0.77 - 1.06) | 0.26 |
| Meta-analysis | 435,109 | 845,198 | 1742.4 | 3332.2 | 15,054 | 32,529 | 0.92 (0.90 - 0.94) | <0.01 | 0.92 (0.85 - 0.99) | 0.03 |
| **PS-matching / intent-to-treat TAR** | | | | | | | | | | |
| CCAE | 1,830 | 3,214 | 0.6 | 1.1 | 9 | 11 | 1.80 (0.63 - 5.30) | 0.28 | 1.79 (0.61 - 5.21) | 0.29 |
| OptumDOD | 25,247 | 63,481 | 75.1 | 201.9 | 1,067 | 3,177 | 0.87 (0.80 - 0.95) | <0.01 | 0.88 (0.80 - 0.97) | 0.01 |
| MDCR | 16,035 | 31,196 | 45.1 | 90.1 | 719 | 1,736 | 0.86 (0.76 - 0.96) | <0.01 | 0.85 (0.75 - 0.96) | 0.01 |
| MDCD | 1,078 | 2,346 | 3.1 | 8.0 | 62 | 136 | 1.20 (0.80 - 1.78) | 0.36 | 1.04 (0.70 - 1.55) | 0.84 |
| Open Claims | 322,630 | 676,711 | 1358.7 | 2744.5 | 11,170 | 24,884 | 0.91 (0.89 - 0.94) | <0.01 | 0.91 (0.83 - 1.00) | 0.05 |
| OptumEHR | 21,336 | 56,310 | 72.7 | 210.5 | 615 | 2,053 | 0.94 (0.84 - 1.05) | 0.26 | 0.98 (0.88 - 1.10) | 0.80 |
| VA | 13,685 | 76,974 | 34.6 | 341.0 | 217 | 2,285 | 0.92 (0.77 - 1.09) | 0.31 | 0.94 (0.79 - 1.12) | 0.49 |
| GermanyDA | 11,547 | 2,382 | 46.5 | 12.6 | 484 | 119 | 1.19 (0.94 - 1.51) | 0.15 | 1.20 (0.94 - 1.53) | 0.18 |
| SIDIAP | 25,424 | 6,006 | 102.4 | 39.7 | 559 | 232 | 1.01 (0.84 - 1.22) | 0.93 | 1.02 (0.84 - 1.23) | 0.84 |
| Meta-analysis | 438,812 | 918,620 | 1739.0 | 3649.4 | 14,902 | 34,633 | 0.91 (0.89 - 0.94) | <0.01 | 0.91 (0.85 - 0.99) | 0.02 |

##### 6.6.18 Vomiting

**Supplementary Table 215:** Exposures, outcomes and risk estimates for Vomiting in DPP4I (T) vs SU (C) new-user cohorts under four different propensity score (PS) adjustment / time-at-risk (TAR) designs across data sources. We report total patient exposure time in 1,000 patient-year (KYr) units, uncalibrated and calibrated hazard ratios (HRs) and their 95% confidence intervals (CIs). Greyed out entries did not pass study diagnostics and are included only for completeness.

| | Patients | | Exposure (KYr) | | Outcomes | | Uncalibrated | | Calibrated | |
|---|---|---|---|---|---|---|---|---|---|---|
| | T | C | T | C | T | C | HR (95% CI) | $p$ | HR (95% CI) | $p$ |
| **PS-stratification / on-treatment TAR** | | | | | | | | | | |
| CCAE | 1,913 | 3,146 | 0.5 | 0.8 | 13 | 19 | 1.12 (0.53 - 2.28) | 0.76 | 1.25 (0.60 - 2.60) | 0.56 |
| OptumDOD | 24,209 | 59,039 | 19.8 | 60.8 | 662 | 2,033 | 0.94 (0.85 - 1.03) | 0.17 | 0.95 (0.86 - 1.04) | 0.33 |
| MDCR | 16,422 | 30,773 | 16.2 | 29.9 | 436 | 913 | 0.89 (0.79 - 1.00) | 0.05 | 0.89 (0.79 - 1.01) | 0.10 |
| MDCD | 965 | 2,025 | 0.6 | 1.4 | 38 | 56 | 1.39 (0.89 - 2.14) | 0.15 | 1.22 (0.78 - 1.89) | 0.42 |
| Open Claims | 308,206 | 635,811 | 284.5 | 733.8 | 4,904 | 11,912 | 0.99 (0.95 - 1.02) | 0.50 | 0.98 (0.87 - 1.11) | 0.80 |
| OptumEHR | 22,044 | 55,074 | 11.3 | 33.0 | 266 | 836 | 0.99 (0.85 - 1.14) | 0.85 | 1.05 (0.90 - 1.21) | 0.62 |
| VA | 14,661 | 106,319 | 15.1 | 119.7 | 98 | 727 | 1.34 (1.03 - 1.72) | 0.02 | 1.36 (1.06 - 1.76) | 0.02 |
| GermanyDA | 11,722 | 2,797 | 13.0 | 3.1 | 140 | 37 | 1.03 (0.68 - 1.58) | 0.89 | 1.20 (0.78 - 1.84) | 0.46 |
| SIDIAP | 25,484 | 7,948 | 68.0 | 22.0 | 557 | 130 | 1.06 (0.85 - 1.33) | 0.62 | 1.06 (0.81 - 1.38) | 0.73 |
| Meta-analysis | 410,965 | 796,613 | 414.0 | 884.8 | 7,016 | 15,936 | 0.98 (0.94 - 1.01) | 0.19 | 0.98 (0.89 - 1.08) | 0.67 |
| **PS-matching / on-treatment TAR** | | | | | | | | | | |
| CCAE | 1,726 | 3,076 | 0.5 | 0.8 | 13 | 19 | 0.82 (0.33 - 1.93) | 0.67 | 0.84 (0.34 - 2.04) | 0.70 |
| OptumDOD | 23,400 | 58,951 | 19.1 | 60.7 | 633 | 2,031 | 0.92 (0.81 - 1.04) | 0.18 | 0.94 (0.83 - 1.06) | 0.35 |
| MDCR | 15,323 | 29,831 | 15.0 | 29.0 | 400 | 898 | 0.95 (0.80 - 1.12) | 0.54 | 0.91 (0.76 - 1.08) | 0.32 |
| MDCD | 928 | 2,009 | 0.6 | 1.3 | 35 | 55 | 1.34 (0.75 - 2.38) | 0.33 | 1.26 (0.70 - 2.25) | 0.45 |
| Open Claims | 302,249 | 635,789 | 277.2 | 733.8 | 4,733 | 11,911 | 0.99 (0.94 - 1.04) | 0.68 | 1.00 (0.91 - 1.10) | 0.90 |
| OptumEHR | 20,877 | 55,034 | 10.6 | 32.9 | 254 | 836 | 1.05 (0.87 - 1.26) | 0.61 | 1.08 (0.90 - 1.31) | 0.45 |
| VA | 13,637 | 77,013 | 14.4 | 82.6 | 91 | 499 | 1.50 (1.10 - 2.04) | 0.01 | 1.52 (1.11 - 2.07) | 0.01 |
| GermanyDA | 11,689 | 2,384 | 12.9 | 2.6 | 139 | 30 | 1.27 (0.76 - 2.24) | 0.38 | 1.31 (0.76 - 2.27) | 0.38 |
| SIDIAP | 25,374 | 6,017 | 67.9 | 15.8 | 556 | 98 | 1.06 (0.80 - 1.42) | 0.70 | 1.12 (0.74 - 1.71) | 0.61 |
| Meta-analysis | 415,203 | 870,104 | 418.3 | 959.5 | 6,854 | 16,377 | 0.99 (0.95 - 1.03) | 0.74 | 0.99 (0.91 - 1.09) | 0.87 |
| **PS-stratification / intent-to-treat TAR** | | | | | | | | | | |
| CCAE | 1,913 | 3,146 | 0.7 | 1.1 | 15 | 27 | 0.94 (0.48 - 1.78) | 0.86 | 0.99 (0.52 - 1.91) | 0.85 |
| OptumDOD | 24,209 | 59,039 | 68.8 | 178.8 | 2,324 | 6,395 | 0.93 (0.88 - 0.98) | <0.01 | 0.93 (0.88 - 0.99) | 0.04 |
| MDCR | 16,422 | 30,773 | 44.6 | 85.9 | 1,411 | 3,052 | 0.90 (0.84 - 0.96) | <0.01 | 0.90 (0.83 - 0.98) | 0.03 |
| MDCD | 965 | 2,025 | 2.5 | 6.3 | 136 | 308 | 1.12 (0.91 - 1.39) | 0.28 | 0.99 (0.80 - 1.23) | 0.84 |
| Open Claims | 308,206 | 635,811 | 1268.1 | 2518.2 | 23,825 | 48,340 | 0.96 (0.94 - 0.97) | <0.01 | 0.96 (0.87 - 1.05) | 0.38 |
| OptumEHR | 22,044 | 55,074 | 72.9 | 200.7 | 1,329 | 3,853 | 1.00 (0.94 - 1.07) | 0.95 | 1.04 (0.97 - 1.12) | 0.29 |
| VA | 14,661 | 106,319 | 35.7 | 567.5 | 246 | 3,977 | 1.11 (0.95 - 1.28) | 0.19 | 1.13 (0.97 - 1.32) | 0.13 |
| GermanyDA | 11,722 | 2,797 | 47.1 | 15.8 | 498 | 201 | 0.94 (0.78 - 1.13) | 0.48 | 0.97 (0.80 - 1.17) | 0.72 |
| SIDIAP | 25,484 | 7,948 | 102.2 | 56.5 | 921 | 503 | 0.90 (0.80 - 1.03) | 0.13 | 0.88 (0.77 - 1.01) | 0.08 |
| Meta-analysis | 413,041 | 799,733 | 1614.6 | 3076.2 | 30,467 | 62,694 | 0.95 (0.92 - 0.98) | <0.01 | 0.95 (0.88 - 1.03) | 0.20 |
| **PS-matching / intent-to-treat TAR** | | | | | | | | | | |
| CCAE | 1,726 | 3,076 | 0.6 | 1.1 | 15 | 27 | 0.76 (0.32 - 1.68) | 0.52 | 0.76 (0.33 - 1.74) | 0.53 |
| OptumDOD | 23,400 | 58,951 | 66.8 | 178.5 | 2,241 | 6,378 | 0.95 (0.89 - 1.01) | 0.08 | 0.95 (0.89 - 1.02) | 0.23 |
| MDCR | 15,323 | 29,831 | 41.7 | 83.3 | 1,314 | 2,994 | 0.92 (0.85 - 1.01) | 0.07 | 0.91 (0.83 - 1.00) | 0.08 |
| MDCD | 928 | 2,009 | 2.4 | 6.3 | 131 | 302 | 1.04 (0.80 - 1.34) | 0.79 | 0.90 (0.69 - 1.16) | 0.46 |
| Open Claims | 302,249 | 635,789 | 1242.2 | 2518.1 | 23,226 | 48,338 | 0.97 (0.95 - 0.99) | <0.01 | 0.97 (0.89 - 1.06) | 0.45 |
| OptumEHR | 20,877 | 55,034 | 69.4 | 200.5 | 1,273 | 3,849 | 1.01 (0.94 - 1.09) | 0.78 | 1.06 (0.97 - 1.15) | 0.23 |
| VA | 13,637 | 77,013 | 34.4 | 342.5 | 234 | 2,303 | 1.13 (0.96 - 1.34) | 0.14 | 1.16 (0.98 - 1.38) | 0.10 |
| GermanyDA | 11,689 | 2,384 | 47.0 | 12.6 | 496 | 155 | 0.88 (0.71 - 1.10) | 0.27 | 0.89 (0.72 - 1.12) | 0.38 |
| SIDIAP | 25,374 | 6,017 | 102.0 | 39.5 | 920 | 352 | 0.87 (0.75 - 1.01) | 0.06 | 0.88 (0.75 - 1.02) | 0.11 |
| Meta-analysis | 416,572 | 873,200 | 1611.8 | 3395.0 | 29,856 | 64,713 | 0.97 (0.95 - 0.98) | <0.01 | 0.97 (0.90 - 1.04) | 0.38 |

### 7 Results of the sensitivity analysis on the follow-up strategy

#### 7.1 GLP1RA vs SGLT2I

##### 7.1.1 Abnormal weight gain

**Supplementary Table 216:** Exposures, outcomes and risk estimates for Abnormal weight gain in GLP1RA (T) vs SGLT2I (C) new-user cohorts under two different propensity score (PS) adjustment across data sources. We report total patient exposure time in 1,000 patient-year (KYr) units, uncalibrated and calibrated hazard ratios (HRs) and their 95% confidence intervals (CIs). Greyed out entries did not pass study diagnostics and are included only for completeness.

| | Patients | | Exposure (KYr) | | Outcomes | | Uncalibrated | | Calibrated | |
|---|---|---|---|---|---|---|---|---|---|---|
| | T | C | T | C | T | C | HR (95% CI) | $p$ | HR (95% CI) | $p$ |
| **PS-stratification / on-treatment 2 TAR** | | | | | | | | | | |
| OptumDOD | 5,009 | 11,275 | 2.4 | 6.1 | 15 | 18 | 1.22 (0.57 - 2.57) | 0.60 | 1.25 (0.59 - 2.65) | 0.56 |
| MDCR | 1,427 | 3,444 | 0.8 | 2.2 | 5 | <5 | 3.22 (0.81 - 13.5) | 0.10 | 2.78 (0.68 - 11.3) | 0.16 |
| Open Claims | 81,047 | 177,824 | 45.1 | 109.8 | 90 | 118 | 1.50 (1.11 - 2.02) | <0.01 | 1.44 (1.06 - 1.94) | 0.02 |
| OptumEHR | 3,184 | 7,560 | 0.8 | 1.9 | 8 | 5 | 3.79 (1.16 - 13.7) | 0.03 | 4.06 (1.18 - 14.0) | 0.03 |
| Meta-analysis | 86,056 | 189,099 | 47.4 | 115.9 | 105 | 136 | 1.46 (1.11 - 1.93) | <0.01 | 1.40 (1.06 - 1.86) | 0.02 |
| **PS-matching / on-treatment 2 TAR** | | | | | | | | | | |
| OptumDOD | 4,556 | 11,259 | 2.2 | 6.1 | 11 | 18 | 0.65 (0.14 - 2.43) | 0.56 | 0.74 (0.18 - 3.12) | 0.68 |
| MDCR | 1,348 | 3,436 | 0.8 | 2.2 | 5 | <5 | | 0.21 | | 0.23 |
| Open Claims | 74,946 | 177,769 | 42.3 | 109.8 | 75 | 118 | 1.74 (1.14 - 2.66) | 0.01 | 1.63 (1.06 - 2.50) | 0.03 |
| OptumEHR | 3,008 | 7,548 | 0.7 | 1.9 | 7 | <5 | | | | |
| Meta-analysis | 79,502 | 189,028 | 44.5 | 115.9 | 86 | 136 | 1.37 (0.60 - 3.13) | 0.46 | 1.33 (0.58 - 3.05) | 0.50 |

##### 7.1.2 Abnormal weight loss

**Supplementary Table 217:** Exposures, outcomes and risk estimates for Abnormal weight loss in GLP1RA (T) vs SGLT2I (C) new-user cohorts under two different propensity score (PS) adjustment across data sources. We report total patient exposure time in 1,000 patient-year (KYr) units, uncalibrated and calibrated hazard ratios (HRs) and their 95% confidence intervals (CIs). Greyed out entries did not pass study diagnostics and are included only for completeness.

| | Patients | | Exposure (KYr) | | Outcomes | | Uncalibrated | | Calibrated | |
|---|---|---|---|---|---|---|---|---|---|---|
| | T | C | T | C | T | C | HR (95% CI) | $p$ | HR (95% CI) | $p$ |
| **PS-stratification / on-treatment 2 TAR** | | | | | | | | | | |
| OptumDOD | 5,123 | 11,094 | 2.4 | 6.0 | 55 | 149 | 0.97 (0.69 - 1.34) | 0.86 | 0.99 (0.71 - 1.38) | 0.89 |
| MDCR | 1,458 | 3,392 | 0.8 | 2.2 | 13 | 40 | 1.00 (0.50 - 1.89) | 0.99 | 0.86 (0.44 - 1.68) | 0.67 |
| Open Claims | 83,825 | 176,400 | 46.5 | 108.8 | 330 | 970 | 0.90 (0.78 - 1.02) | 0.10 | 0.86 (0.75 - 0.98) | 0.03 |
| OptumEHR | 3,296 | 7,530 | 0.8 | 1.9 | 7 | 27 | 0.52 (0.20 - 1.18) | 0.14 | 0.55 (0.23 - 1.34) | 0.20 |
| Meta-analysis | 93,702 | 198,416 | 50.5 | 118.8 | 405 | 1,186 | 0.90 (0.80 - 1.01) | 0.08 | 0.87 (0.76 - 0.98) | 0.03 |
| **PS-matching / on-treatment 2 TAR** | | | | | | | | | | |
| OptumDOD | 4,578 | 11,078 | 2.2 | 6.0 | 51 | 149 | 1.02 (0.66 - 1.55) | 0.94 | 1.16 (0.75 - 1.77) | 0.52 |
| MDCR | 1,363 | 3,384 | 0.8 | 2.2 | 13 | 39 | 1.11 (0.48 - 2.41) | 0.80 | 1.05 (0.47 - 2.35) | 0.87 |
| Open Claims | 76,528 | 176,352 | 43.1 | 108.8 | 303 | 967 | 0.81 (0.67 - 0.96) | 0.02 | 0.75 (0.63 - 0.90) | <0.01 |
| OptumEHR | 3,081 | 7,518 | 0.7 | 1.9 | 6 | 27 | 0.60 (0.16 - 1.80) | 0.40 | 0.56 (0.17 - 1.86) | 0.35 |
| Meta-analysis | 85,550 | 198,332 | 46.8 | 118.8 | 373 | 1,182 | 0.84 (0.71 - 0.98) | 0.03 | 0.81 (0.69 - 0.96) | 0.02 |

##### 7.1.3 Acute pancreatitis

**Supplementary Table 218:** Exposures, outcomes and risk estimates for Acute pancreatitis in GLP1RA (T) vs SGLT2I (C) new-user cohorts under two different propensity score (PS) adjustment across data sources. We report total patient exposure time in 1,000 patient-year (KYr) units, uncalibrated and calibrated hazard ratios (HRs) and their 95% confidence intervals (CIs). Greyed out entries did not pass study diagnostics and are included only for completeness.

| | Patients | | Exposure (KYr) | | Outcomes | | Uncalibrated | | Calibrated | |
|---|---|---|---|---|---|---|---|---|---|---|
| | T | C | T | C | T | C | HR (95% CI) | $p$ | HR (95% CI) | $p$ |
| **PS-stratification / on-treatment 2 TAR** | | | | | | | | | | |
| OptumDOD | 5,302 | 11,512 | 2.5 | 6.3 | 7 | 18 | 0.92 (0.34 - 2.20) | 0.86 | 0.94 (0.37 - 2.38) | 0.90 |
| MDCR | 1,487 | 3,480 | 0.8 | 2.3 | <5 | <5 | 2.23 (0.36 - 12.3) | 0.37 | 1.92 (0.33 - 11.3) | 0.47 |
| Open Claims | 86,346 | 182,041 | 47.9 | 112.4 | 77 | 146 | 1.38 (1.02 - 1.84) | 0.03 | 1.32 (0.98 - 1.77) | 0.07 |
| OptumEHR | 3,385 | 7,801 | 0.8 | 1.9 | <5 | <5 | 2.30 (0.39 - 13.8) | 0.36 | 2.46 (0.42 - 14.6) | 0.32 |
| Meta-analysis | 91,648 | 193,553 | 50.4 | 118.7 | 84 | 164 | 1.33 (1.00 - 1.76) | 0.05 | 1.28 (0.96 - 1.69) | 0.09 |
| **PS-matching / on-treatment 2 TAR** | | | | | | | | | | |
| OptumDOD | 4,737 | 11,496 | 2.3 | 6.2 | 6 | 18 | 0.49 (0.14 - 1.43) | 0.24 | 0.56 (0.17 - 1.81) | 0.34 |
| MDCR | 1,385 | 3,472 | 0.8 | 2.3 | <5 | <5 | 1.50 (0.18 - 10.5) | 0.70 | 1.41 (0.18 - 10.8) | 0.75 |
| Open Claims | 78,761 | 181,987 | 44.4 | 112.4 | 72 | 146 | 1.42 (0.95 - 2.12) | 0.09 | 1.33 (0.89 - 2.00) | 0.17 |
| OptumEHR | 3,162 | 7,789 | 0.8 | 1.9 | <5 | <5 | 0.00 (0.00 - 11.7) | 0.23 | 0.00 (0.00 - 93059) | 0.23 |
| Meta-analysis | 83,498 | 193,483 | 46.7 | 118.6 | 78 | 164 | 0.97 (0.36 - 2.63) | 0.96 | 0.95 (0.35 - 2.56) | 0.91 |

##### 7.1.4 All-cause mortality

**Supplementary Table 219:** Exposures, outcomes and risk estimates for All-cause mortality in GLP1RA (T) vs SGLT2I (C) new-user cohorts under two different propensity score (PS) adjustment across data sources. We report total patient exposure time in 1,000 patient-year (KYr) units, uncalibrated and calibrated hazard ratios (HRs) and their 95% confidence intervals (CIs). Greyed out entries did not pass study diagnostics and are included only for completeness.

| | Patients | | Exposure (KYr) | | Outcomes | | Uncalibrated | | Calibrated | |
|---|---|---|---|---|---|---|---|---|---|---|
| | T | C | T | C | T | C | HR (95% CI) | $p$ | HR (95% CI) | $p$ |
| **PS-stratification / on-treatment 2 TAR** | | | | | | | | | | |
| OptumDOD | 5,336 | 11,614 | 2.5 | 6.3 | 14 | 40 | 0.91 (0.46 - 1.70) | 0.78 | 0.93 (0.49 - 1.79) | 0.83 |
| MDCR | 1,493 | 3,510 | 0.8 | 2.3 | 0 | <5 | | | | |
| Open Claims | 87,367 | 184,807 | 48.4 | 114.0 | 0 | 0 | | | | |
| OptumEHR | 3,403 | 7,828 | 0.8 | 2.0 | <5 | 7 | 0.42 (0.06 - 1.90) | 0.32 | 0.45 (0.08 - 2.53) | 0.36 |
| Meta-analysis | 5,336 | 11,614 | 2.5 | 6.3 | 14 | 40 | 0.91 (0.46 - 1.70) | 0.78 | 0.88 (0.46 - 1.69) | 0.69 |
| **PS-matching / on-treatment 2 TAR** | | | | | | | | | | |
| OptumDOD | 4,772 | 11,598 | 2.3 | 6.3 | 13 | 40 | 2.12 (0.82 - 5.39) | 0.12 | 2.41 (0.94 - 6.20) | 0.07 |
| MDCR | 1,393 | 3,502 | 0.8 | 2.3 | 0 | <5 | | | | |
| Open Claims | 79,722 | 184,753 | 44.9 | 114.0 | 0 | 0 | | | | |
| OptumEHR | 3,177 | 7,816 | 0.8 | 2.0 | <5 | 7 | 0.42 (0.02 - 3.10) | 0.50 | 0.40 (0.03 - 4.81) | 0.47 |
| Meta-analysis | 4,772 | 11,598 | 2.3 | 6.3 | 13 | 40 | 2.12 (0.82 - 5.39) | 0.12 | 2.06 (0.80 - 5.30) | 0.13 |

##### 7.1.5 Bone fracture

**Supplementary Table 220:** Exposures, outcomes and risk estimates for Bone fracture in GLP1RA (T) vs SGLT2I (C) new-user cohorts under two different propensity score (PS) adjustment across data sources. We report total patient exposure time in 1,000 patient-year (KYr) units, uncalibrated and calibrated hazard ratios (HRs) and their 95% confidence intervals (CIs). Greyed out entries did not pass study diagnostics and are included only for completeness.

| | Patients | | Exposure (KYr) | | Outcomes | | Uncalibrated | | Calibrated | |
|---|---|---|---|---|---|---|---|---|---|---|
| | T | C | T | C | T | C | HR (95% CI) | $p$ | HR (95% CI) | $p$ |
| **PS-stratification / on-treatment 2 TAR** | | | | | | | | | | |
| OptumDOD | 4,701 | 10,423 | 2.2 | 5.6 | 80 | 172 | 1.03 (0.77 - 1.37) | 0.83 | 1.06 (0.79 - 1.41) | 0.73 |
| MDCR | 1,295 | 3,143 | 0.7 | 2.0 | 18 | 62 | 0.65 (0.36 - 1.12) | 0.14 | 0.56 (0.32 - 0.99) | 0.05 |
| Open Claims | 70,647 | 155,987 | 39.0 | 96.2 | 678 | 1,571 | 0.98 (0.89 - 1.08) | 0.68 | 0.94 (0.85 - 1.04) | 0.24 |
| OptumEHR | 3,067 | 7,175 | 0.7 | 1.8 | 15 | 33 | 0.75 (0.38 - 1.41) | 0.39 | 0.81 (0.42 - 1.54) | 0.52 |
| Meta-analysis | 79,710 | 176,728 | 42.6 | 105.6 | 791 | 1,838 | 0.97 (0.89 - 1.06) | 0.51 | 0.93 (0.85 - 1.03) | 0.18 |
| **PS-matching / on-treatment 2 TAR** | | | | | | | | | | |
| OptumDOD | 4,216 | 10,409 | 2.0 | 5.6 | 67 | 172 | 0.90 (0.60 - 1.33) | 0.60 | 1.02 (0.68 - 1.53) | 0.86 |
| MDCR | 1,212 | 3,136 | 0.7 | 2.0 | 17 | 62 | 0.72 (0.32 - 1.50) | 0.40 | 0.68 (0.31 - 1.47) | 0.35 |
| Open Claims | 65,162 | 155,863 | 36.5 | 96.2 | 614 | 1,568 | 0.96 (0.84 - 1.10) | 0.58 | 0.90 (0.79 - 1.04) | 0.18 |
| OptumEHR | 2,874 | 7,163 | 0.7 | 1.8 | 12 | 33 | 0.86 (0.36 - 1.91) | 0.72 | 0.80 (0.35 - 1.85) | 0.62 |
| Meta-analysis | 73,464 | 176,571 | 39.9 | 105.5 | 710 | 1,835 | 0.95 (0.84 - 1.07) | 0.39 | 0.92 (0.81 - 1.05) | 0.23 |

##### 7.1.6 Diabetic ketoacidosis

**Supplementary Table 221:** Exposures, outcomes and risk estimates for Diabetic ketoacidosis in GLP1RA (T) vs SGLT2I (C) new-user cohorts under two different propensity score (PS) adjustment across data sources. We report total patient exposure time in 1,000 patient-year (KYr) units, uncalibrated and calibrated hazard ratios (HRs) and their 95% confidence intervals (CIs). Greyed out entries did not pass study diagnostics and are included only for completeness.

| | Patients | | Exposure (KYr) | | Outcomes | | Uncalibrated | | Calibrated | |
|---|---|---|---|---|---|---|---|---|---|---|
| | T | C | T | C | T | C | HR (95% CI) | $p$ | HR (95% CI) | $p$ |
| **PS-stratification / on-treatment 2 TAR** | | | | | | | | | | |
| OptumDOD | 5,330 | 11,606 | 2.5 | 6.3 | <5 | 17 | 0.35 (0.08 - 1.09) | 0.12 | 0.36 (0.10 - 1.33) | 0.12 |
| MDCR | 1,492 | 3,506 | 0.8 | 2.3 | 0 | <5 | | | | |
| Open Claims | 87,275 | 184,585 | 48.4 | 113.9 | 28 | 114 | 0.54 (0.35 - 0.83) | <0.01 | 0.52 (0.34 - 0.81) | <0.01 |
| OptumEHR | 3,402 | 7,821 | 0.8 | 2.0 | <5 | <5 | 1.11 (0.05 - 12.6) | 0.94 | 1.19 (0.07 - 19.1) | 0.90 |
| Meta-analysis | 92,605 | 196,191 | 50.9 | 120.2 | <33 | 131 | 0.52 (0.34 - 0.79) | <0.01 | 0.50 (0.33 - 0.76) | <0.01 |
| **PS-matching / on-treatment 2 TAR** | | | | | | | | | | |
| OptumDOD | 4,769 | 11,590 | 2.3 | 6.3 | <5 | 17 | 0.78 (0.16 - 2.80) | 0.72 | 0.88 (0.21 - 3.64) | 0.86 |
| MDCR | 1,391 | 3,498 | 0.8 | 2.3 | 0 | <5 | | | | |
| Open Claims | 79,633 | 184,531 | 44.9 | 113.9 | 25 | 113 | 0.48 (0.27 - 0.80) | <0.01 | 0.45 (0.26 - 0.77) | <0.01 |
| OptumEHR | 3,176 | 7,809 | 0.8 | 2.0 | <5 | <5 | 2.00 (0.07 - 56.4) | 0.68 | 1.87 (0.07 - 52.8) | 0.71 |
| Meta-analysis | 84,402 | 196,121 | 47.2 | 120.2 | <30 | 130 | 0.51 (0.31 - 0.84) | <0.01 | 0.49 (0.30 - 0.82) | <0.01 |

##### 7.1.7 Diarrhea

**Supplementary Table 222:** Exposures, outcomes and risk estimates for Diarrhea in GLP1RA (T) vs SGLT2I (C) new-user cohorts under two different propensity score (PS) adjustment across data sources. We report total patient exposure time in 1,000 patient-year (KYr) units, uncalibrated and calibrated hazard ratios (HRs) and their 95% confidence intervals (CIs). Greyed out entries did not pass study diagnostics and are included only for completeness.

| | Patients | | Exposure (KYr) | | Outcomes | | Uncalibrated | | Calibrated | |
|---|---|---|---|---|---|---|---|---|---|---|
| | T | C | T | C | T | C | HR (95% CI) | $p$ | HR (95% CI) | $p$ |
| **PS-stratification / on-treatment 2 TAR** | | | | | | | | | | |
| OptumDOD | 4,449 | 9,875 | 2.1 | 5.3 | 112 | 207 | 1.20 (0.94 - 1.54) | 0.14 | 1.23 (0.96 - 1.58) | 0.12 |
| MDCR | 1,281 | 3,032 | 0.7 | 2.0 | 18 | 52 | 0.83 (0.45 - 1.45) | 0.52 | 0.71 (0.40 - 1.27) | 0.26 |
| Open Claims | 72,445 | 158,033 | 40.1 | 97.6 | 862 | 1,489 | 1.33 (1.21 - 1.45) | <0.01 | 1.27 (1.15 - 1.39) | <0.01 |
| OptumEHR | 2,980 | 6,910 | 0.7 | 1.7 | 29 | 64 | 0.89 (0.55 - 1.40) | 0.61 | 0.95 (0.60 - 1.52) | 0.76 |
| Meta-analysis | 81,155 | 177,850 | 43.6 | 106.6 | 1,021 | 1,812 | 1.18 (0.98 - 1.42) | 0.09 | 1.13 (0.93 - 1.37) | 0.21 |
| **PS-matching / on-treatment 2 TAR** | | | | | | | | | | |
| OptumDOD | 3,981 | 9,860 | 1.9 | 5.3 | 100 | 205 | 1.19 (0.85 - 1.66) | 0.31 | 1.35 (0.97 - 1.89) | 0.09 |
| MDCR | 1,195 | 3,025 | 0.7 | 2.0 | 17 | 52 | 0.99 (0.44 - 2.12) | 0.97 | 0.93 (0.42 - 2.05) | 0.84 |
| Open Claims | 66,618 | 157,992 | 37.4 | 97.6 | 766 | 1,487 | 1.38 (1.22 - 1.56) | <0.01 | 1.29 (1.14 - 1.47) | <0.01 |
| OptumEHR | 2,775 | 6,899 | 0.7 | 1.7 | 28 | 64 | 0.93 (0.52 - 1.62) | 0.79 | 0.87 (0.49 - 1.54) | 0.63 |
| Meta-analysis | 74,569 | 177,776 | 40.6 | 106.5 | 911 | 1,808 | 1.28 (1.08 - 1.51) | <0.01 | 1.24 (1.05 - 1.47) | 0.02 |

##### 7.1.8 Genitourinary infection

**Supplementary Table 223:** Exposures, outcomes and risk estimates for Genitourinary infection in GLP1RA (T) vs SGLT2I (C) new-user cohorts under two different propensity score (PS) adjustment across data sources. We report total patient exposure time in 1,000 patient-year (KYr) units, uncalibrated and calibrated hazard ratios (HRs) and their 95% confidence intervals (CIs). Greyed out entries did not pass study diagnostics and are included only for completeness.

| | Patients | | Exposure (KYr) | | Outcomes | | Uncalibrated | | Calibrated | |
|---|---|---|---|---|---|---|---|---|---|---|
| | T | C | T | C | T | C | HR (95% CI) | $p$ | HR (95% CI) | $p$ |
| **PS-stratification / on-treatment 2 TAR** | | | | | | | | | | |
| OptumDOD | 3,876 | 9,018 | 1.8 | 4.8 | 128 | 292 | 1.09 (0.87 - 1.36) | 0.43 | 1.12 (0.89 - 1.40) | 0.35 |
| MDCR | 1,118 | 2,894 | 0.6 | 1.9 | 34 | 89 | 1.01 (0.66 - 1.53) | 0.95 | 0.87 (0.57 - 1.33) | 0.55 |
| Open Claims | 60,348 | 137,023 | 33.6 | 85.9 | 1,194 | 2,538 | 1.09 (1.01 - 1.17) | 0.03 | 1.04 (0.96 - 1.12) | 0.41 |
| OptumEHR | 2,754 | 6,565 | 0.6 | 1.6 | 40 | 80 | 1.09 (0.72 - 1.62) | 0.68 | 1.17 (0.78 - 1.75) | 0.49 |
| Meta-analysis | 68,096 | 155,500 | 36.6 | 94.2 | 1,396 | 2,999 | 1.09 (1.01 - 1.16) | 0.02 | 1.04 (0.96 - 1.13) | 0.31 |
| **PS-matching / on-treatment 2 TAR** | | | | | | | | | | |
| OptumDOD | 3,491 | 9,007 | 1.6 | 4.8 | 119 | 292 | 0.83 (0.61 - 1.11) | 0.22 | 0.94 (0.70 - 1.27) | 0.70 |
| MDCR | 1,054 | 2,886 | 0.6 | 1.9 | 32 | 88 | 0.92 (0.49 - 1.67) | 0.79 | 0.87 (0.47 - 1.61) | 0.68 |
| Open Claims | 56,132 | 136,980 | 31.7 | 85.9 | 1,060 | 2,536 | 0.98 (0.89 - 1.08) | 0.69 | 0.92 (0.83 - 1.02) | 0.15 |
| OptumEHR | 2,609 | 6,555 | 0.6 | 1.6 | 37 | 80 | 0.97 (0.52 - 1.75) | 0.92 | 0.91 (0.49 - 1.67) | 0.71 |
| Meta-analysis | 63,286 | 155,428 | 34.5 | 94.1 | 1,248 | 2,996 | 0.96 (0.88 - 1.06) | 0.42 | 0.93 (0.84 - 1.04) | 0.22 |

##### 7.1.9 Hyperkalemia

**Supplementary Table 224:** Exposures, outcomes and risk estimates for Hyperkalemia in GLP1RA (T) vs SGLT2I (C) new-user cohorts under two different propensity score (PS) adjustment across data sources. We report total patient exposure time in 1,000 patient-year (KYr) units, uncalibrated and calibrated hazard ratios (HRs) and their 95% confidence intervals (CIs). Greyed out entries did not pass study diagnostics and are included only for completeness.

| | Patients | | Exposure (KYr) | | Outcomes | | Uncalibrated | | Calibrated | |
|---|---|---|---|---|---|---|---|---|---|---|
| | T | C | T | C | T | C | HR (95% CI) | $p$ | HR (95% CI) | $p$ |
| **PS-stratification / on-treatment 2 TAR** | | | | | | | | | | |
| OptumDOD | 5,051 | 10,938 | 2.4 | 5.9 | 59 | 145 | 1.12 (0.81 - 1.54) | 0.49 | 1.15 (0.83 - 1.58) | 0.43 |
| MDCR | 1,457 | 3,408 | 0.8 | 2.2 | 9 | 24 | 0.92 (0.38 - 2.06) | 0.85 | 0.79 (0.34 - 1.86) | 0.60 |
| Open Claims | 84,511 | 178,385 | 46.8 | 110.1 | 255 | 755 | 0.93 (0.80 - 1.08) | 0.36 | 0.89 (0.76 - 1.04) | 0.16 |
| OptumEHR | 3,303 | 7,603 | 0.8 | 1.9 | 7 | 26 | 0.54 (0.21 - 1.22) | 0.17 | 0.57 (0.24 - 1.40) | 0.23 |
| Meta-analysis | 94,322 | 200,334 | 50.7 | 120.2 | 330 | 950 | 0.95 (0.83 - 1.08) | 0.44 | 0.91 (0.79 - 1.05) | 0.21 |
| **PS-matching / on-treatment 2 TAR** | | | | | | | | | | |
| OptumDOD | 4,513 | 10,924 | 2.2 | 5.9 | 55 | 144 | 0.97 (0.61 - 1.51) | 0.89 | 1.10 (0.70 - 1.74) | 0.69 |
| MDCR | 1,357 | 3,400 | 0.8 | 2.2 | 8 | 23 | 1.00 (0.35 - 2.58) | 0.99 | 0.95 (0.35 - 2.59) | 0.88 |
| Open Claims | 77,107 | 178,338 | 43.4 | 110.1 | 236 | 754 | 0.97 (0.79 - 1.19) | 0.77 | 0.91 (0.74 - 1.12) | 0.38 |
| OptumEHR | 3,089 | 7,592 | 0.7 | 1.9 | 6 | 26 | 0.25 (0.04 - 0.92) | 0.09 | 0.24 (0.05 - 1.14) | 0.08 |
| Meta-analysis | 86,066 | 200,254 | 47.1 | 120.1 | 305 | 947 | 0.95 (0.80 - 1.14) | 0.61 | 0.93 (0.77 - 1.12) | 0.44 |

###### 7.1.10 Hypoglycemia

**Supplementary Table 225:** Exposures, outcomes and risk estimates for Hypoglycemia in GLP1RA (T) vs SGLT2I (C) new-user cohorts under two different propensity score (PS) adjustment across data sources. We report total patient exposure time in 1,000 patient-year (KYr) units, uncalibrated and calibrated hazard ratios (HRs) and their 95% confidence intervals (CIs). Greyed out entries did not pass study diagnostics and are included only for completeness.

| | Patients | | Exposure (KYr) | | Outcomes | | Uncalibrated | | Calibrated | |
|---|---|---|---|---|---|---|---|---|---|---|
| | T | C | T | C | T | C | HR (95% CI) | $p$ | HR (95% CI) | $p$ |
| **PS-stratification / on-treatment 2 TAR** | | | | | | | | | | |
| OptumDOD | 5,239 | 11,439 | 2.5 | 6.2 | 12 | 36 | 0.63 (0.30 - 1.22) | 0.19 | 0.64 (0.32 - 1.29) | 0.22 |
| MDCR | 1,473 | 3,474 | 0.8 | 2.3 | <5 | <5 | 0.82 (0.04 - 6.67) | 0.88 | 0.71 (0.05 - 9.15) | 0.79 |
| Open Claims | 85,575 | 181,772 | 47.4 | 112.2 | 104 | 159 | 1.52 (1.16 - 1.98) | <0.01 | 1.46 (1.11 - 1.90) | <0.01 |
| OptumEHR | 3,355 | 7,748 | 0.8 | 1.9 | <5 | <5 | 0.66 (0.03 - 5.31) | 0.75 | 0.71 (0.06 - 9.08) | 0.79 |
| Meta-analysis | 90,814 | 193,211 | 49.9 | 118.5 | 116 | 195 | 1.04 (0.44 - 2.46) | 0.93 | 1.00 (0.42 - 2.37) | 0.98 |
| **PS-matching / on-treatment 2 TAR** | | | | | | | | | | |
| OptumDOD | 4,701 | 11,424 | 2.3 | 6.2 | 10 | 36 | 0.44 (0.12 - 1.27) | 0.17 | 0.50 (0.15 - 1.61) | 0.24 |
| MDCR | 1,373 | 3,466 | 0.8 | 2.3 | <5 | <5 | 0.88 (0.04 - 9.66) | 0.93 | 0.83 (0.05 - 13.0) | 0.89 |
| Open Claims | 78,221 | 181,718 | 44.1 | 112.2 | 94 | 159 | 1.89 (1.32 - 2.71) | <0.01 | 1.77 (1.23 - 2.54) | <0.01 |
| OptumEHR | 3,140 | 7,736 | 0.8 | 1.9 | <5 | <5 | | | | |
| Meta-analysis | 82,922 | 193,142 | 46.4 | 118.4 | 104 | 195 | 1.01 (0.24 - 4.20) | 0.98 | 0.99 (0.24 - 4.08) | 0.98 |

##### 7.1.11 Hypotension

**Supplementary Table 226:** Exposures, outcomes and risk estimates for Hypotension in GLP1RA (T) vs SGLT2I (C) new-user cohorts under two different propensity score (PS) adjustment across data sources. We report total patient exposure time in 1,000 patient-year (KYr) units, uncalibrated and calibrated hazard ratios (HRs) and their 95% confidence intervals (CIs). Greyed out entries did not pass study diagnostics and are included only for completeness.

| | Patients | | Exposure (KYr) | | Outcomes | | Uncalibrated | | Calibrated | |
|---|---|---|---|---|---|---|---|---|---|---|
| | T | C | T | C | T | C | HR (95% CI) | $p$ | HR (95% CI) | $p$ |
| **PS-stratification / on-treatment 2 TAR** | | | | | | | | | | |
| OptumDOD | 4,991 | 10,826 | 2.4 | 5.9 | 66 | 195 | 0.87 (0.64 - 1.16) | 0.35 | 0.89 (0.65 - 1.20) | 0.44 |
| MDCR | 1,417 | 3,342 | 0.8 | 2.2 | 15 | 53 | 0.81 (0.44 - 1.44) | 0.50 | 0.70 (0.39 - 1.28) | 0.25 |
| Open Claims | 82,501 | 174,134 | 45.7 | 107.6 | 479 | 1,261 | 1.03 (0.92 - 1.15) | 0.65 | 0.98 (0.87 - 1.10) | 0.76 |
| OptumEHR | 3,277 | 7,517 | 0.8 | 1.9 | 14 | 35 | 0.99 (0.50 - 1.85) | 0.98 | 1.06 (0.55 - 2.04) | 0.83 |
| Meta-analysis | 92,186 | 195,819 | 49.6 | 117.5 | 574 | 1,544 | 1.00 (0.90 - 1.11) | 0.95 | 0.96 (0.86 - 1.07) | 0.48 |
| **PS-matching / on-treatment 2 TAR** | | | | | | | | | | |
| OptumDOD | 4,483 | 10,804 | 2.1 | 5.8 | 60 | 195 | 0.75 (0.49 - 1.11) | 0.16 | 0.85 (0.57 - 1.27) | 0.44 |
| MDCR | 1,319 | 3,334 | 0.8 | 2.2 | 15 | 53 | 1.14 (0.51 - 2.44) | 0.74 | 1.08 (0.49 - 2.36) | 0.84 |
| Open Claims | 75,427 | 174,062 | 42.5 | 107.6 | 431 | 1,254 | 0.95 (0.81 - 1.11) | 0.49 | 0.89 (0.76 - 1.04) | 0.16 |
| OptumEHR | 3,053 | 7,506 | 0.7 | 1.9 | 13 | 35 | 1.59 (0.73 - 3.37) | 0.24 | 1.49 (0.69 - 3.20) | 0.33 |
| Meta-analysis | 84,282 | 195,706 | 46.1 | 117.5 | 519 | 1,537 | 0.94 (0.82 - 1.08) | 0.40 | 0.91 (0.79 - 1.06) | 0.25 |

##### 7.1.12 Joint pain

**Supplementary Table 227:** Exposures, outcomes and risk estimates for Joint pain in GLP1RA (T) vs SGLT2I (C) new-user cohorts under two different propensity score (PS) adjustment across data sources. We report total patient exposure time in 1,000 patient-year (KYr) units, uncalibrated and calibrated hazard ratios (HRs) and their 95% confidence intervals (CIs). Greyed out entries did not pass study diagnostics and are included only for completeness.

| | Patients | | Exposure (KYr) | | Outcomes | | Uncalibrated | | Calibrated | |
|---|---|---|---|---|---|---|---|---|---|---|
| | T | C | T | C | T | C | HR (95% CI) | $p$ | HR (95% CI) | $p$ |
| **PS-stratification / on-treatment 2 TAR** | | | | | | | | | | |
| OptumDOD | 4,874 | 10,899 | 2.3 | 5.9 | 41 | 75 | 1.14 (0.75 - 1.70) | 0.54 | 1.16 (0.77 - 1.75) | 0.48 |
| MDCR | 1,390 | 3,368 | 0.8 | 2.2 | 8 | 23 | 0.63 (0.25 - 1.43) | 0.29 | 0.54 (0.23 - 1.29) | 0.17 |
| Open Claims | 79,450 | 172,024 | 44.2 | 106.5 | 282 | 545 | 1.06 (0.91 - 1.23) | 0.48 | 1.01 (0.86 - 1.18) | 0.88 |
| OptumEHR | 3,155 | 7,405 | 0.8 | 1.8 | 7 | 24 | 0.62 (0.24 - 1.43) | 0.29 | 0.66 (0.27 - 1.63) | 0.37 |
| Meta-analysis | 88,869 | 193,696 | 48.1 | 116.4 | 338 | 667 | 1.04 (0.90 - 1.19) | 0.60 | 1.00 (0.86 - 1.15) | 0.90 |
| **PS-matching / on-treatment 2 TAR** | | | | | | | | | | |
| OptumDOD | 4,400 | 10,883 | 2.1 | 5.9 | 38 | 75 | 0.79 (0.44 - 1.39) | 0.43 | 0.90 (0.50 - 1.61) | 0.72 |
| MDCR | 1,312 | 3,360 | 0.8 | 2.2 | 7 | 23 | 0.40 (0.06 - 1.78) | 0.29 | 0.38 (0.07 - 2.09) | 0.27 |
| Open Claims | 73,133 | 171,974 | 41.3 | 106.4 | 247 | 545 | 0.96 (0.77 - 1.19) | 0.72 | 0.90 (0.72 - 1.12) | 0.36 |
| OptumEHR | 2,958 | 7,395 | 0.7 | 1.8 | 7 | 24 | 0.35 (0.05 - 1.43) | 0.21 | 0.32 (0.06 - 1.71) | 0.19 |
| Meta-analysis | 81,803 | 193,612 | 44.9 | 116.4 | 299 | 667 | 0.91 (0.75 - 1.12) | 0.38 | 0.89 (0.72 - 1.09) | 0.26 |

###### 7.1.13 Lower extremity amputation

**Supplementary Table 228:** Exposures, outcomes and risk estimates for Lower extremity amputation in GLP1RA (T) vs SGLT2I (C) new-user cohorts under two different propensity score (PS) adjustment across data sources. We report total patient exposure time in 1,000 patient-year (KYr) units, uncalibrated and calibrated hazard ratios (HRs) and their 95% confidence intervals (CIs). Greyed out entries did not pass study diagnostics and are included only for completeness.

| | Patients | | Exposure (KYr) | | Outcomes | | Uncalibrated | | Calibrated | |
|---|---|---|---|---|---|---|---|---|---|---|
| | T | C | T | C | T | C | HR (95% CI) | $p$ | HR (95% CI) | $p$ |
| **PS-stratification / on-treatment 2 TAR** | | | | | | | | | | |
| OptumDOD | 5,323 | 11,589 | 2.5 | 6.3 | <5 | <5 | 0.54 (0.02 - 4.71) | 0.64 | 0.55 (0.04 - 7.65) | 0.65 |
| MDCR | 1,492 | 3,500 | 0.8 | 2.3 | 0 | <5 | | | | |
| Open Claims | 87,174 | 184,471 | 48.3 | 113.8 | 13 | 57 | 0.58 (0.30 - 1.05) | 0.09 | 0.55 (0.29 - 1.04) | 0.07 |
| OptumEHR | 3,400 | 7,821 | 0.8 | 2.0 | 0 | 0 | | | | |
| Meta-analysis | 87,174 | 184,471 | 48.3 | 113.8 | 13 | 57 | 0.58 (0.30 - 1.05) | 0.09 | 0.55 (0.29 - 1.05) | 0.07 |
| **PS-matching / on-treatment 2 TAR** | | | | | | | | | | |
| OptumDOD | 4,758 | 11,574 | 2.3 | 6.3 | 0 | <5 | | | | |
| MDCR | 1,392 | 3,492 | 0.8 | 2.3 | 0 | <5 | | | | |
| Open Claims | 79,536 | 184,417 | 44.8 | 113.8 | 13 | 57 | 0.50 (0.21 - 1.07) | 0.10 | 0.47 (0.21 - 1.06) | 0.07 |
| OptumEHR | 3,174 | 7,809 | 0.8 | 2.0 | 0 | 0 | | | | |
| Meta-analysis | 79,536 | 184,417 | 44.8 | 113.8 | 13 | 57 | 0.50 (0.21 - 1.07) | 0.10 | 0.49 (0.22 - 1.10) | 0.09 |

###### 7.1.14 Nausea

**Supplementary Table 229:** Exposures, outcomes and risk estimates for Nausea in GLP1RA (T) vs SGLT2I (C) new-user cohorts under two different propensity score (PS) adjustment across data sources. We report total patient exposure time in 1,000 patient-year (KYr) units, uncalibrated and calibrated hazard ratios (HRs) and their 95% confidence intervals (CIs). Greyed out entries did not pass study diagnostics and are included only for completeness.

| | Patients | | Exposure (KYr) | | Outcomes | | Uncalibrated | | Calibrated | |
|---|---|---|---|---|---|---|---|---|---|---|
| | T | C | T | C | T | C | HR (95% CI) | $p$ | HR (95% CI) | $p$ |
| **PS-stratification / on-treatment 2 TAR** | | | | | | | | | | |
| OptumDOD | 4,438 | 9,971 | 2.1 | 5.4 | 163 | 199 | 1.75 (1.40 - 2.20) | <0.01 | 1.79 (1.43 - 2.26) | <0.01 |
| MDCR | 1,332 | 3,120 | 0.7 | 2.0 | 27 | 62 | 0.96 (0.58 - 1.54) | 0.86 | 0.82 (0.50 - 1.35) | 0.45 |
| Open Claims | 72,522 | 158,462 | 40.2 | 98.7 | 1,156 | 1,483 | 1.71 (1.57 - 1.85) | <0.01 | 1.63 (1.49 - 1.78) | <0.01 |
| OptumEHR | 3,028 | 7,090 | 0.7 | 1.8 | 51 | 53 | 1.87 (1.24 - 2.82) | <0.01 | 2.00 (1.32 - 3.03) | <0.01 |
| Meta-analysis | 81,320 | 178,643 | 43.8 | 107.9 | 1,397 | 1,797 | 1.69 (1.57 - 1.83) | <0.01 | 1.63 (1.50 - 1.77) | <0.01 |
| **PS-matching / on-treatment 2 TAR** | | | | | | | | | | |
| OptumDOD | 3,990 | 9,957 | 1.9 | 5.4 | 141 | 199 | 1.94 (1.44 - 2.63) | <0.01 | 2.21 (1.63 - 2.99) | <0.01 |
| MDCR | 1,244 | 3,113 | 0.7 | 2.0 | 26 | 62 | 0.88 (0.44 - 1.68) | 0.71 | 0.83 (0.42 - 1.63) | 0.61 |
| Open Claims | 66,731 | 158,415 | 37.6 | 98.7 | 1,029 | 1,482 | 1.90 (1.70 - 2.13) | <0.01 | 1.78 (1.59 - 2.00) | <0.01 |
| OptumEHR | 2,835 | 7,079 | 0.7 | 1.8 | 48 | 53 | 1.97 (1.17 - 3.31) | 0.01 | 1.84 (1.09 - 3.10) | 0.03 |
| Meta-analysis | 74,800 | 178,564 | 40.8 | 107.9 | 1,244 | 1,796 | 1.87 (1.69 - 2.07) | <0.01 | 1.82 (1.63 - 2.03) | <0.01 |

##### 7.1.15 Peripheral edema

**Supplementary Table 230:** Exposures, outcomes and risk estimates for Peripheral edema in GLP1RA (T) vs SGLT2I (C) new-user cohorts under two different propensity score (PS) adjustment across data sources. We report total patient exposure time in 1,000 patient-year (KYr) units, uncalibrated and calibrated hazard ratios (HRs) and their 95% confidence intervals (CIs). Greyed out entries did not pass study diagnostics and are included only for completeness.

| | Patients | | Exposure (KYr) | | Outcomes | | Uncalibrated | | Calibrated | |
|---|---|---|---|---|---|---|---|---|---|---|
| | T | C | T | C | T | C | HR (95% CI) | $p$ | HR (95% CI) | $p$ |
| **PS-stratification / on-treatment 2 TAR** | | | | | | | | | | |
| OptumDOD | 3,713 | 8,491 | 1.7 | 4.6 | 120 | 298 | 0.97 (0.77 - 1.21) | 0.77 | 0.99 (0.79 - 1.25) | 0.84 |
| MDCR | 1,085 | 2,647 | 0.6 | 1.7 | 34 | 74 | 1.27 (0.81 - 1.97) | 0.29 | 1.10 (0.70 - 1.71) | 0.70 |
| Open Claims | 58,688 | 132,741 | 32.5 | 83.0 | 928 | 2,095 | 1.12 (1.04 - 1.22) | <0.01 | 1.08 (0.98 - 1.17) | 0.14 |
| OptumEHR | 2,573 | 6,272 | 0.6 | 1.5 | 27 | 42 | 1.34 (0.78 - 2.27) | 0.28 | 1.44 (0.84 - 2.45) | 0.20 |
| Meta-analysis | 66,059 | 150,151 | 35.5 | 90.8 | 1,109 | 2,509 | 1.11 (1.03 - 1.20) | <0.01 | 1.07 (0.98 - 1.17) | 0.13 |
| **PS-matching / on-treatment 2 TAR** | | | | | | | | | | |
| OptumDOD | 3,349 | 8,482 | 1.6 | 4.6 | 109 | 298 | 1.03 (0.76 - 1.38) | 0.86 | 1.17 (0.87 - 1.57) | 0.33 |
| MDCR | 1,021 | 2,639 | 0.6 | 1.7 | 28 | 74 | 0.72 (0.33 - 1.43) | 0.37 | 0.67 (0.32 - 1.40) | 0.32 |
| Open Claims | 54,529 | 132,608 | 30.7 | 82.9 | 849 | 2,088 | 1.08 (0.96 - 1.20) | 0.19 | 1.01 (0.90 - 1.13) | 0.83 |
| OptumEHR | 2,426 | 6,262 | 0.6 | 1.5 | 25 | 42 | 1.91 (0.93 - 3.90) | 0.08 | 1.79 (0.87 - 3.67) | 0.13 |
| Meta-analysis | 61,325 | 149,991 | 33.4 | 90.7 | 1,011 | 2,502 | 1.08 (0.97 - 1.19) | 0.17 | 1.04 (0.93 - 1.17) | 0.46 |

##### 7.1.16 Photosensitivity

**Supplementary Table 231:** Exposures, outcomes and risk estimates for Photosensitivity in GLP1RA (T) vs SGLT2I (C) new-user cohorts under two different propensity score (PS) adjustment across data sources. We report total patient exposure time in 1,000 patient-year (KYr) units, uncalibrated and calibrated hazard ratios (HRs) and their 95% confidence intervals (CIs). Greyed out entries did not pass study diagnostics and are included only for completeness.

| | Patients | | Exposure (KYr) | | Outcomes | | Uncalibrated | | Calibrated | |
|---|---|---|---|---|---|---|---|---|---|---|
| | T | C | T | C | T | C | HR (95% CI) | $p$ | HR (95% CI) | $p$ |
| **PS-stratification / on-treatment 2 TAR** | | | | | | | | | | |
| OptumDOD | 5,293 | 11,516 | 2.5 | 6.3 | <5 | <5 | 1.59 (0.07 - 13.8) | 0.73 | 1.63 (0.12 - 22.2) | 0.72 |
| MDCR | 1,461 | 3,464 | 0.8 | 2.3 | <5 | <5 | 2.29 (0.27 - 15.3) | 0.42 | 1.97 (0.26 - 14.7) | 0.51 |
| Open Claims | 85,859 | 181,939 | 47.5 | 112.1 | 17 | 26 | 1.94 (0.97 - 3.75) | 0.05 | 1.85 (0.94 - 3.64) | 0.07 |
| OptumEHR | 3,366 | 7,758 | 0.8 | 1.9 | <5 | 0 | | | | |
| Meta-analysis | 85,859 | 181,939 | 47.5 | 112.1 | 17 | 26 | 1.94 (0.97 - 3.75) | 0.05 | 1.86 (0.95 - 3.66) | 0.07 |
| **PS-matching / on-treatment 2 TAR** | | | | | | | | | | |
| OptumDOD | 4,739 | 11,501 | 2.3 | 6.2 | <5 | <5 | 2.36 (0.10 - 27.4) | 0.55 | 2.68 (0.16 - 44.0) | 0.49 |
| MDCR | 1,365 | 3,456 | 0.8 | 2.2 | <5 | <5 | 2.81 (0.30 - 25.7) | 0.36 | 2.65 (0.29 - 24.4) | 0.40 |
| Open Claims | 78,393 | 181,885 | 44.1 | 112.1 | 14 | 26 | 1.46 (0.63 - 3.25) | 0.37 | 1.36 (0.60 - 3.10) | 0.46 |
| OptumEHR | 3,146 | 7,746 | 0.8 | 1.9 | <5 | 0 | | | | |
| Meta-analysis | 78,393 | 181,885 | 44.1 | 112.1 | 14 | 26 | 1.46 (0.63 - 3.25) | 0.37 | 1.41 (0.62 - 3.21) | 0.41 |

###### 7.1.17 Venous thromboembolism

**Supplementary Table 232:** Exposures, outcomes and risk estimates for Venous thromboembolism in GLP1RA (T) vs SGLT2I (C) new-user cohorts under two different propensity score (PS) adjustment across data sources. We report total patient exposure time in 1,000 patient-year (KYr) units, uncalibrated and calibrated hazard ratios (HRs) and their 95% confidence intervals (CIs). Greyed out entries did not pass study diagnostics and are included only for completeness.

| | Patients | | Exposure (KYr) | | Outcomes | | Uncalibrated | | Calibrated | |
|---|---|---|---|---|---|---|---|---|---|---|
| | T | C | T | C | T | C | HR (95% CI) | $p$ | HR (95% CI) | $p$ |
| **PS-stratification / on-treatment 2 TAR** | | | | | | | | | | |
| OptumDOD | 5,063 | 11,177 | 2.4 | 6.1 | 26 | 57 | 0.88 (0.53 - 1.44) | 0.61 | 0.90 (0.54 - 1.49) | 0.68 |
| MDCR | 1,418 | 3,367 | 0.8 | 2.2 | 7 | 18 | 1.16 (0.44 - 2.75) | 0.74 | 1.00 (0.40 - 2.50) | 0.93 |
| Open Claims | 82,510 | 176,274 | 45.9 | 109.0 | 239 | 567 | 0.96 (0.82 - 1.13) | 0.62 | 0.92 (0.78 - 1.08) | 0.33 |
| OptumEHR | 3,263 | 7,536 | 0.8 | 1.9 | 8 | 14 | 1.00 (0.38 - 2.47) | 0.99 | 1.07 (0.42 - 2.73) | 0.86 |
| Meta-analysis | 92,254 | 198,354 | 49.8 | 119.1 | 280 | 656 | 0.96 (0.83 - 1.11) | 0.58 | 0.92 (0.79 - 1.08) | 0.31 |
| **PS-matching / on-treatment 2 TAR** | | | | | | | | | | |
| OptumDOD | 4,546 | 11,162 | 2.2 | 6.0 | 23 | 57 | 1.11 (0.59 - 2.03) | 0.74 | 1.26 (0.68 - 2.34) | 0.46 |
| MDCR | 1,333 | 3,360 | 0.8 | 2.2 | 7 | 18 | 0.52 (0.14 - 1.51) | 0.27 | 0.49 (0.15 - 1.59) | 0.24 |
| Open Claims | 75,451 | 176,226 | 42.6 | 108.9 | 217 | 567 | 1.02 (0.82 - 1.26) | 0.85 | 0.96 (0.77 - 1.19) | 0.70 |
| OptumEHR | 3,046 | 7,524 | 0.7 | 1.9 | 7 | 14 | 1.31 (0.38 - 4.25) | 0.67 | 1.22 (0.36 - 4.12) | 0.75 |
| Meta-analysis | 84,376 | 198,272 | 46.3 | 119.0 | 254 | 656 | 1.02 (0.83 - 1.24) | 0.87 | 0.99 (0.81 - 1.21) | 0.88 |

###### 7.1.18 Vomiting

**Supplementary Table 233:** Exposures, outcomes and risk estimates for Vomiting in GLP1RA (T) vs SGLT2I (C) new-user cohorts under two different propensity score (PS) adjustment across data sources. We report total patient exposure time in 1,000 patient-year (KYr) units, uncalibrated and calibrated hazard ratios (HRs) and their 95% confidence intervals (CIs). Greyed out entries did not pass study diagnostics and are included only for completeness.

| | Patients | | Exposure (KYr) | | Outcomes | | Uncalibrated | | Calibrated | |
|---|---|---|---|---|---|---|---|---|---|---|
| | T | C | T | C | T | C | HR (95% CI) | $p$ | HR (95% CI) | $p$ |
| **PS-stratification / on-treatment 2 TAR** | | | | | | | | | | |
| OptumDOD | 4,765 | 10,476 | 2.2 | 5.7 | 118 | 144 | 1.75 (1.34 - 2.29) | <0.01 | 1.80 (1.37 - 2.35) | <0.01 |
| MDCR | 1,385 | 3,271 | 0.8 | 2.1 | 22 | 45 | 1.17 (0.67 - 2.00) | 0.57 | 1.01 (0.58 - 1.75) | 0.89 |
| Open Claims | 76,917 | 165,476 | 42.6 | 102.8 | 844 | 1,119 | 1.70 (1.54 - 1.87) | <0.01 | 1.62 (1.47 - 1.79) | <0.01 |
| OptumEHR | 3,174 | 7,398 | 0.8 | 1.8 | 26 | 35 | 1.67 (0.97 - 2.85) | 0.06 | 1.79 (1.04 - 3.08) | 0.04 |
| Meta-analysis | 86,241 | 186,621 | 46.4 | 112.4 | 1,010 | 1,343 | 1.69 (1.54 - 1.84) | <0.01 | 1.62 (1.47 - 1.79) | <0.01 |
| **PS-matching / on-treatment 2 TAR** | | | | | | | | | | |
| OptumDOD | 4,261 | 10,462 | 2.0 | 5.7 | 100 | 144 | 1.70 (1.19 - 2.43) | <0.01 | 1.93 (1.35 - 2.76) | <0.01 |
| MDCR | 1,296 | 3,263 | 0.7 | 2.1 | 22 | 45 | 1.24 (0.58 - 2.59) | 0.58 | 1.17 (0.55 - 2.48) | 0.71 |
| Open Claims | 70,503 | 165,428 | 39.7 | 102.7 | 756 | 1,118 | 1.82 (1.60 - 2.07) | <0.01 | 1.71 (1.49 - 1.95) | <0.01 |
| OptumEHR | 2,968 | 7,387 | 0.7 | 1.8 | 24 | 35 | 1.79 (0.90 - 3.52) | 0.10 | 1.67 (0.84 - 3.32) | 0.16 |
| Meta-analysis | 79,028 | 186,540 | 43.1 | 112.4 | 902 | 1,342 | 1.79 (1.59 - 2.01) | <0.01 | 1.74 (1.53 - 1.97) | <0.01 |

#### 7.2 GLP1RA vs DPP4I

##### 7.2.1 Abnormal weight gain

**Supplementary Table 234:** Exposures, outcomes and risk estimates for Abnormal weight gain in GLP1RA (T) vs DPP4I (C) new-user cohorts under two different propensity score (PS) adjustment across data sources. We report total patient exposure time in 1,000 patient-year (KYr) units, uncalibrated and calibrated hazard ratios (HRs) and their 95% confidence intervals (CIs). Greyed out entries did not pass study diagnostics and are included only for completeness.

| | Patients | | Exposure (KYr) | | Outcomes | | Uncalibrated | | Calibrated | |
|---|---|---|---|---|---|---|---|---|---|---|
| | T | C | T | C | T | C | HR (95% CI) | $p$ | HR (95% CI) | $p$ |
| **PS-stratification / on-treatment 2 TAR** | | | | | | | | | | |
| OptumDOD | 5,281 | 26,500 | 2.5 | 18.8 | 16 | 103 | 1.19 (0.62 - 2.15) | 0.58 | 1.15 (0.62 - 2.14) | 0.66 |
| MDCR | 1,915 | 17,725 | 1.1 | 15.3 | 6 | 36 | 3.17 (1.08 - 7.97) | 0.02 | 2.75 (1.01 - 7.48) | 0.05 |
| Open Claims | 81,161 | 334,884 | 45.1 | 257.6 | 90 | 436 | 1.09 (0.83 - 1.42) | 0.53 | 1.06 (0.78 - 1.45) | 0.70 |
| OptumEHR | 3,350 | 22,797 | 0.8 | 9.8 | 8 | 40 | 2.21 (0.88 - 4.95) | 0.07 | 1.71 (0.72 - 4.08) | 0.24 |
| Meta-analysis | 84,511 | 357,681 | 45.9 | 267.4 | 98 | 476 | 1.37 (0.72 - 2.62) | 0.34 | 1.35 (0.69 - 2.61) | 0.38 |
| **PS-matching / on-treatment 2 TAR** | | | | | | | | | | |
| OptumDOD | 4,536 | 25,289 | 2.3 | 18.0 | 12 | 94 | 0.96 (0.39 - 2.09) | 0.92 | 0.98 (0.42 - 2.26) | 0.94 |
| MDCR | 1,725 | 16,893 | 1.0 | 14.6 | 6 | 35 | 3.15 (0.92 - 9.25) | 0.05 | 2.73 (0.86 - 8.68) | 0.09 |
| Open Claims | 66,636 | 331,260 | 39.8 | 254.9 | 65 | 430 | 1.02 (0.68 - 1.48) | 0.93 | 0.97 (0.65 - 1.45) | 0.88 |
| OptumEHR | 3,089 | 22,640 | 0.8 | 9.8 | 7 | 39 | 2.40 (0.80 - 6.80) | 0.11 | 1.97 (0.67 - 5.74) | 0.22 |
| Meta-analysis | 74,261 | 379,189 | 42.8 | 282.6 | 84 | 563 | 1.09 (0.78 - 1.53) | 0.60 | 1.06 (0.74 - 1.51) | 0.77 |

##### 7.2.2 Abnormal weight loss

**Supplementary Table 235:** Exposures, outcomes and risk estimates for Abnormal weight loss in GLP1RA (T) vs DPP4I (C) new-user cohorts under two different propensity score (PS) adjustment across data sources. We report total patient exposure time in 1,000 patient-year (KYr) units, uncalibrated and calibrated hazard ratios (HRs) and their 95% confidence intervals (CIs). Greyed out entries did not pass study diagnostics and are included only for completeness.

| | Patients | | Exposure (KYr) | | Outcomes | | Uncalibrated | | Calibrated | |
|---|---|---|---|---|---|---|---|---|---|---|
| | T | C | T | C | T | C | HR (95% CI) | $p$ | HR (95% CI) | $p$ |
| **PS-stratification / on-treatment 2 TAR** | | | | | | | | | | |
| OptumDOD | 5,409 | 25,475 | 2.6 | 18.0 | 55 | 381 | 1.39 (1.00 - 1.92) | 0.05 | 1.35 (0.97 - 1.87) | 0.08 |
| MDCR | 1,954 | 17,148 | 1.1 | 14.7 | 15 | 225 | 1.08 (0.59 - 1.87) | 0.78 | 0.94 (0.53 - 1.68) | 0.84 |
| Open Claims | 83,942 | 326,105 | 46.5 | 250.0 | 330 | 2,394 | 0.99 (0.86 - 1.12) | 0.83 | 0.96 (0.79 - 1.18) | 0.72 |
| OptumEHR | 3,464 | 22,394 | 0.9 | 9.6 | 7 | 161 | 0.62 (0.25 - 1.29) | 0.25 | 0.48 (0.21 - 1.09) | 0.09 |
| Meta-analysis | 87,406 | 348,499 | 47.4 | 259.6 | 337 | 2,555 | 0.94 (0.71 - 1.24) | 0.66 | 0.92 (0.68 - 1.26) | 0.62 |
| **PS-matching / on-treatment 2 TAR** | | | | | | | | | | |
| OptumDOD | 4,571 | 24,676 | 2.3 | 17.5 | 54 | 362 | 1.38 (0.92 - 2.05) | 0.11 | 1.41 (0.94 - 2.12) | 0.10 |
| MDCR | 1,735 | 16,470 | 1.0 | 14.2 | 14 | 209 | 1.50 (0.75 - 2.77) | 0.22 | 1.30 (0.68 - 2.50) | 0.43 |
| Open Claims | 67,430 | 323,455 | 40.4 | 247.9 | 291 | 2,344 | 0.96 (0.81 - 1.13) | 0.64 | 0.92 (0.75 - 1.12) | 0.43 |
| OptumEHR | 3,142 | 22,259 | 0.8 | 9.6 | 7 | 159 | 1.08 (0.42 - 2.38) | 0.86 | 0.89 (0.37 - 2.10) | 0.78 |
| Meta-analysis | 76,878 | 386,860 | 44.5 | 289.2 | 366 | 3,074 | 1.13 (0.87 - 1.47) | 0.36 | 1.09 (0.81 - 1.47) | 0.57 |

##### 7.2.3 Acute pancreatitis

**Supplementary Table 236:** Exposures, outcomes and risk estimates for Acute pancreatitis in GLP1RA (T) vs DPP4I (C) new-user cohorts under two different propensity score (PS) adjustment across data sources. We report total patient exposure time in 1,000 patient-year (KYr) units, uncalibrated and calibrated hazard ratios (HRs) and their 95% confidence intervals (CIs). Greyed out entries did not pass study diagnostics and are included only for completeness.

| | Patients | | Exposure (KYr) | | Outcomes | | Uncalibrated | | Calibrated | |
|---|---|---|---|---|---|---|---|---|---|---|
| | T | C | T | C | T | C | HR (95% CI) | $p$ | HR (95% CI) | $p$ |
| **PS-stratification / on-treatment 2 TAR** | | | | | | | | | | |
| OptumDOD | 5,591 | 26,987 | 2.7 | 19.3 | 7 | 64 | 1.21 (0.48 - 2.65) | 0.66 | 1.17 (0.50 - 2.76) | 0.72 |
| MDCR | 1,985 | 17,814 | 1.1 | 15.3 | <5 | 49 | 1.69 (0.46 - 4.79) | 0.38 | 1.47 (0.45 - 4.74) | 0.52 |
| Open Claims | 86,463 | 340,319 | 48.0 | 261.6 | 77 | 596 | 0.95 (0.72 - 1.23) | 0.69 | 0.92 (0.68 - 1.26) | 0.63 |
| OptumEHR | 3,555 | 23,251 | 0.9 | 10.1 | <5 | 20 | 1.75 (0.39 - 5.56) | 0.41 | 1.35 (0.36 - 5.13) | 0.66 |
| Meta-analysis | 86,463 | 340,319 | 48.0 | 261.6 | 77 | 596 | 0.95 (0.72 - 1.23) | 0.69 | 0.93 (0.69 - 1.26) | 0.65 |
| **PS-matching / on-treatment 2 TAR** | | | | | | | | | | |
| OptumDOD | 4,723 | 25,870 | 2.4 | 18.5 | 7 | 60 | 0.90 (0.29 - 2.31) | 0.85 | 0.92 (0.33 - 2.60) | 0.88 |
| MDCR | 1,770 | 16,989 | 1.1 | 14.7 | <5 | 44 | 1.31 (0.29 - 4.27) | 0.70 | 1.14 (0.29 - 4.40) | 0.85 |
| Open Claims | 69,581 | 336,618 | 41.7 | 258.8 | 68 | 581 | 0.95 (0.68 - 1.31) | 0.76 | 0.91 (0.64 - 1.28) | 0.59 |
| OptumEHR | 3,229 | 23,093 | 0.8 | 10.0 | <5 | 20 | 0.48 (0.03 - 2.57) | 0.53 | 0.39 (0.04 - 3.92) | 0.43 |
| Meta-analysis | 76,074 | 379,477 | 45.1 | 292.0 | <80 | 685 | 0.96 (0.71 - 1.31) | 0.80 | 0.93 (0.66 - 1.29) | 0.66 |

##### 7.2.4 All-cause mortality

**Supplementary Table 237:** Exposures, outcomes and risk estimates for All-cause mortality in GLP1RA (T) vs DPP4I (C) new-user cohorts under two different propensity score (PS) adjustment across data sources. We report total patient exposure time in 1,000 patient-year (KYr) units, uncalibrated and calibrated hazard ratios (HRs) and their 95% confidence intervals (CIs). Greyed out entries did not pass study diagnostics and are included only for completeness.

| | Patients | | Exposure (KYr) | | Outcomes | | Uncalibrated | | Calibrated | |
|---|---|---|---|---|---|---|---|---|---|---|
| | T | C | T | C | T | C | HR (95% CI) | $p$ | HR (95% CI) | $p$ |
| **PS-stratification / on-treatment 2 TAR** | | | | | | | | | | |
| OptumDOD | 5,626 | 27,232 | 2.7 | 19.4 | 15 | 227 | 0.77 (0.42 - 1.33) | 0.38 | 0.75 (0.42 - 1.34) | 0.33 |
| MDCR | 1,993 | 17,961 | 1.1 | 15.5 | <5 | 43 | 2.10 (0.33 - 7.46) | 0.35 | 1.82 (0.38 - 8.69) | 0.45 |
| Open Claims | 87,486 | 344,544 | 48.5 | 265.0 | 0 | <5 | | | | |
| OptumEHR | 3,573 | 23,343 | 0.9 | 10.1 | <5 | 113 | 0.31 (0.05 - 1.02) | 0.13 | 0.24 (0.05 - 1.09) | 0.07 |
| Meta-analysis | 3,573 | 23,343 | 0.9 | 10.1 | <5 | 113 | 0.31 (0.05 - 1.02) | 0.13 | 0.31 (0.07 - 1.39) | 0.13 |
| **PS-matching / on-treatment 2 TAR** | | | | | | | | | | |
| OptumDOD | 4,755 | 26,062 | 2.4 | 18.6 | 14 | 196 | 0.82 (0.39 - 1.60) | 0.58 | 0.84 (0.41 - 1.70) | 0.63 |
| MDCR | 1,778 | 17,109 | 1.1 | 14.8 | <5 | 34 | 1.17 (0.18 - 4.48) | 0.85 | 1.01 (0.20 - 5.12) | 0.96 |
| Open Claims | 70,428 | 340,897 | 42.2 | 262.3 | 0 | <5 | | | | |
| OptumEHR | 3,242 | 23,184 | 0.8 | 10.0 | <5 | 107 | 0.46 (0.07 - 1.62) | 0.33 | 0.38 (0.08 - 1.79) | 0.22 |
| Meta-analysis | 7,997 | 49,246 | 3.2 | 28.7 | <19 | 303 | 0.74 (0.39 - 1.41) | 0.36 | 0.72 (0.37 - 1.38) | 0.32 |

##### 7.2.5 Bone fracture

**Supplementary Table 238:** Exposures, outcomes and risk estimates for Bone fracture in GLP1RA (T) vs DPP4I (C) new-user cohorts under two different propensity score (PS) adjustment across data sources. We report total patient exposure time in 1,000 patient-year (KYr) units, uncalibrated and calibrated hazard ratios (HRs) and their 95% confidence intervals (CIs). Greyed out entries did not pass study diagnostics and are included only for completeness.

| | Patients | | Exposure (KYr) | | Outcomes | | Uncalibrated | | Calibrated | |
|---|---|---|---|---|---|---|---|---|---|---|
| | T | C | T | C | T | C | HR (95% CI) | $p$ | HR (95% CI) | $p$ |
| **PS-stratification / on-treatment 2 TAR** | | | | | | | | | | |
| OptumDOD | 4,964 | 24,065 | 2.3 | 16.9 | 86 | 582 | 1.16 (0.89 - 1.50) | 0.26 | 1.12 (0.86 - 1.46) | 0.40 |
| MDCR | 1,744 | 15,701 | 1.0 | 13.3 | 26 | 460 | 0.87 (0.56 - 1.32) | 0.54 | 0.76 (0.49 - 1.17) | 0.22 |
| Open Claims | 70,749 | 291,457 | 39.0 | 222.1 | 680 | 5,161 | 0.89 (0.81 - 0.98) | 0.01 | 0.87 (0.73 - 1.04) | 0.15 |
| OptumEHR | 3,230 | 21,423 | 0.8 | 9.1 | 17 | 217 | 1.10 (0.62 - 1.81) | 0.73 | 0.85 (0.50 - 1.46) | 0.58 |
| Meta-analysis | 73,979 | 312,880 | 39.8 | 231.2 | 697 | 5,378 | 0.90 (0.82 - 0.98) | 0.02 | 0.88 (0.74 - 1.05) | 0.15 |
| **PS-matching / on-treatment 2 TAR** | | | | | | | | | | |
| OptumDOD | 4,206 | 22,818 | 2.1 | 16.0 | 73 | 530 | 1.02 (0.73 - 1.41) | 0.91 | 1.04 (0.75 - 1.45) | 0.80 |
| MDCR | 1,549 | 15,031 | 0.9 | 12.8 | 26 | 430 | 1.05 (0.63 - 1.67) | 0.85 | 0.91 (0.56 - 1.48) | 0.72 |
| Open Claims | 57,737 | 287,634 | 34.3 | 219.1 | 587 | 5,018 | 0.85 (0.75 - 0.96) | <0.01 | 0.81 (0.69 - 0.95) | 0.03 |
| OptumEHR | 2,949 | 21,287 | 0.7 | 9.1 | 16 | 214 | 1.16 (0.60 - 2.09) | 0.63 | 0.95 (0.51 - 1.78) | 0.83 |
| Meta-analysis | 66,441 | 346,770 | 38.0 | 257.0 | 702 | 6,192 | 0.91 (0.78 - 1.05) | 0.21 | 0.88 (0.72 - 1.07) | 0.19 |

##### 7.2.6 Diabetic ketoacidosis

**Supplementary Table 239:** Exposures, outcomes and risk estimates for Diabetic ketoacidosis in GLP1RA (T) vs DPP4I (C) new-user cohorts under two different propensity score (PS) adjustment across data sources. We report total patient exposure time in 1,000 patient-year (KYr) units, uncalibrated and calibrated hazard ratios (HRs) and their 95% confidence intervals (CIs). Greyed out entries did not pass study diagnostics and are included only for completeness.

| | Patients | | Exposure (KYr) | | Outcomes | | Uncalibrated | | Calibrated | |
|---|---|---|---|---|---|---|---|---|---|---|
| | T | C | T | C | T | C | HR (95% CI) | $p$ | HR (95% CI) | $p$ |
| **PS-stratification / on-treatment 2 TAR** | | | | | | | | | | |
| OptumDOD | 5,619 | 27,194 | 2.7 | 19.4 | <5 | 29 | 0.46 (0.10 - 1.56) | 0.27 | 0.45 (0.11 - 1.77) | 0.25 |
| MDCR | 1,992 | 17,928 | 1.1 | 15.4 | 0 | 14 | | | | |
| Open Claims | 87,394 | 344,081 | 48.4 | 264.6 | 28 | 190 | 0.91 (0.57 - 1.41) | 0.69 | 0.89 (0.55 - 1.44) | 0.64 |
| OptumEHR | 3,572 | 23,333 | 0.9 | 10.1 | <5 | 8 | 0.39 (0.02 - 2.92) | 0.46 | 0.30 (0.02 - 3.73) | 0.35 |
| Meta-analysis | 87,394 | 344,081 | 48.4 | 264.6 | 28 | 190 | 0.91 (0.57 - 1.41) | 0.69 | 0.90 (0.56 - 1.44) | 0.65 |
| **PS-matching / on-treatment 2 TAR** | | | | | | | | | | |
| OptumDOD | 4,750 | 26,038 | 2.4 | 18.6 | <5 | 27 | 0.59 (0.13 - 2.05) | 0.46 | 0.60 (0.15 - 2.42) | 0.48 |
| MDCR | 1,777 | 17,083 | 1.1 | 14.8 | 0 | 13 | | | | |
| Open Claims | 70,336 | 340,456 | 42.1 | 261.9 | 24 | 189 | 0.94 (0.52 - 1.60) | 0.82 | 0.89 (0.50 - 1.58) | 0.70 |
| OptumEHR | 3,239 | 23,174 | 0.8 | 10.0 | <5 | 8 | 0.72 (0.03 - 6.65) | 0.81 | 0.59 (0.04 - 8.32) | 0.70 |
| Meta-analysis | 75,086 | 366,494 | 44.5 | 280.5 | <29 | 216 | 0.88 (0.52 - 1.48) | 0.62 | 0.85 (0.50 - 1.45) | 0.54 |

##### 7.2.7 Diarrhea

**Supplementary Table 240:** Exposures, outcomes and risk estimates for Diarrhea in GLP1RA (T) vs DPP4I (C) new-user cohorts under two different propensity score (PS) adjustment across data sources. We report total patient exposure time in 1,000 patient-year (KYr) units, uncalibrated and calibrated hazard ratios (HRs) and their 95% confidence intervals (CIs). Greyed out entries did not pass study diagnostics and are included only for completeness.

| | Patients | | Exposure (KYr) | | Outcomes | | Uncalibrated | | Calibrated | |
|---|---|---|---|---|---|---|---|---|---|---|
| | T | C | T | C | T | C | HR (95% CI) | $p$ | HR (95% CI) | $p$ |
| **PS-stratification / on-treatment 2 TAR** | | | | | | | | | | |
| OptumDOD | 4,704 | 22,558 | 2.2 | 15.7 | 116 | 694 | 1.09 (0.87 - 1.37) | 0.45 | 1.06 (0.84 - 1.33) | 0.65 |
| MDCR | 1,736 | 15,423 | 1.0 | 13.0 | 36 | 466 | 0.98 (0.66 - 1.40) | 0.91 | 0.85 (0.58 - 1.23) | 0.41 |
| Open Claims | 72,549 | 292,867 | 40.1 | 223.0 | 863 | 4,959 | 1.10 (1.01 - 1.20) | 0.03 | 1.07 (0.90 - 1.28) | 0.45 |
| OptumEHR | 3,136 | 20,427 | 0.8 | 8.6 | 32 | 310 | 1.29 (0.86 - 1.89) | 0.20 | 1.00 (0.67 - 1.50) | 0.82 |
| Meta-analysis | 75,685 | 313,294 | 40.9 | 231.6 | 895 | 5,269 | 1.11 (1.02 - 1.20) | 0.02 | 1.09 (0.92 - 1.28) | 0.32 |
| **PS-matching / on-treatment 2 TAR** | | | | | | | | | | |
| OptumDOD | 3,968 | 21,606 | 2.0 | 15.1 | 101 | 650 | 1.06 (0.79 - 1.40) | 0.70 | 1.08 (0.81 - 1.44) | 0.61 |
| MDCR | 1,556 | 14,634 | 0.9 | 12.4 | 35 | 433 | 1.42 (0.92 - 2.13) | 0.10 | 1.23 (0.81 - 1.88) | 0.34 |
| Open Claims | 58,708 | 289,666 | 35.0 | 220.5 | 729 | 4,836 | 1.14 (1.03 - 1.27) | 0.01 | 1.09 (0.93 - 1.27) | 0.35 |
| OptumEHR | 2,846 | 20,254 | 0.7 | 8.5 | 31 | 307 | 1.39 (0.88 - 2.13) | 0.14 | 1.14 (0.73 - 1.78) | 0.57 |
| Meta-analysis | 67,078 | 346,160 | 38.7 | 256.6 | 896 | 6,226 | 1.16 (1.05 - 1.27) | <0.01 | 1.11 (0.95 - 1.31) | 0.19 |

##### 7.2.8 Genitourinary infection

**Supplementary Table 241:** Exposures, outcomes and risk estimates for Genitourinary infection in GLP1RA (T) vs DPP4I (C) new-user cohorts under two different propensity score (PS) adjustment across data sources. We report total patient exposure time in 1,000 patient-year (KYr) units, uncalibrated and calibrated hazard ratios (HRs) and their 95% confidence intervals (CIs). Greyed out entries did not pass study diagnostics and are included only for completeness.

| | Patients | | Exposure (KYr) | | Outcomes | | Uncalibrated | | Calibrated | |
|---|---|---|---|---|---|---|---|---|---|---|
| | T | C | T | C | T | C | HR (95% CI) | $p$ | HR (95% CI) | $p$ |
| **PS-stratification / on-treatment 2 TAR** | | | | | | | | | | |
| OptumDOD | 4,094 | 19,137 | 1.9 | 13.1 | 136 | 1,102 | 1.02 (0.83 - 1.25) | 0.84 | 0.99 (0.81 - 1.21) | 0.82 |
| MDCR | 1,515 | 13,747 | 0.8 | 11.5 | 46 | 728 | 0.91 (0.65 - 1.25) | 0.57 | 0.79 (0.57 - 1.09) | 0.17 |
| Open Claims | 60,438 | 247,138 | 33.7 | 185.8 | 1,195 | 8,843 | 0.94 (0.88 - 1.01) | 0.08 | 0.92 (0.78 - 1.09) | 0.35 |
| OptumEHR | 2,901 | 18,900 | 0.7 | 7.8 | 44 | 497 | 1.19 (0.84 - 1.64) | 0.31 | 0.92 (0.66 - 1.29) | 0.67 |
| Meta-analysis | 63,339 | 266,038 | 34.4 | 193.7 | 1,239 | 9,340 | 1.00 (0.82 - 1.21) | 0.97 | 0.98 (0.77 - 1.25) | 0.86 |
| **PS-matching / on-treatment 2 TAR** | | | | | | | | | | |
| OptumDOD | 3,448 | 18,130 | 1.7 | 12.4 | 125 | 1,011 | 1.05 (0.81 - 1.33) | 0.72 | 1.07 (0.83 - 1.37) | 0.62 |
| MDCR | 1,355 | 13,361 | 0.8 | 11.2 | 45 | 699 | 0.91 (0.61 - 1.32) | 0.62 | 0.79 (0.53 - 1.16) | 0.25 |
| Open Claims | 49,225 | 243,775 | 29.5 | 183.4 | 1,038 | 8,545 | 0.96 (0.87 - 1.04) | 0.31 | 0.91 (0.79 - 1.05) | 0.27 |
| OptumEHR | 2,645 | 18,728 | 0.6 | 7.8 | 42 | 483 | 1.01 (0.66 - 1.50) | 0.97 | 0.83 (0.55 - 1.25) | 0.39 |
| Meta-analysis | 56,673 | 293,994 | 32.6 | 214.7 | 1,250 | 10,738 | 0.96 (0.89 - 1.04) | 0.37 | 0.93 (0.80 - 1.09) | 0.35 |

##### 7.2.9 Hyperkalemia

**Supplementary Table 242:** Exposures, outcomes and risk estimates for Hyperkalemia in GLP1RA (T) vs DPP4I (C) new-user cohorts under two different propensity score (PS) adjustment across data sources. We report total patient exposure time in 1,000 patient-year (KYr) units, uncalibrated and calibrated hazard ratios (HRs) and their 95% confidence intervals (CIs). Greyed out entries did not pass study diagnostics and are included only for completeness.

| | Patients | | Exposure (KYr) | | Outcomes | | Uncalibrated | | Calibrated | |
|---|---|---|---|---|---|---|---|---|---|---|
| | T | C | T | C | T | C | HR (95% CI) | $p$ | HR (95% CI) | $p$ |
| **PS-stratification / on-treatment 2 TAR** | | | | | | | | | | |
| OptumDOD | 5,330 | 25,331 | 2.5 | 17.7 | 60 | 586 | 0.82 (0.61 - 1.10) | 0.20 | 0.80 (0.59 - 1.07) | 0.15 |
| MDCR | 1,951 | 17,425 | 1.1 | 14.9 | 12 | 238 | 0.72 (0.36 - 1.31) | 0.31 | 0.62 (0.33 - 1.18) | 0.16 |
| Open Claims | 84,627 | 331,478 | 46.8 | 252.9 | 256 | 3,156 | 0.63 (0.55 - 0.73) | <0.01 | 0.62 (0.50 - 0.76) | <0.01 |
| OptumEHR | 3,469 | 22,492 | 0.9 | 9.6 | 7 | 231 | 0.36 (0.15 - 0.74) | 0.01 | 0.28 (0.13 - 0.62) | <0.01 |
| Meta-analysis | 88,096 | 353,970 | 47.7 | 262.5 | 263 | 3,387 | 0.55 (0.35 - 0.88) | 0.01 | 0.54 (0.34 - 0.89) | 0.01 |
| **PS-matching / on-treatment 2 TAR** | | | | | | | | | | |
| OptumDOD | 4,496 | 24,476 | 2.3 | 17.1 | 57 | 556 | 0.96 (0.67 - 1.36) | 0.83 | 0.98 (0.69 - 1.40) | 0.87 |
| MDCR | 1,735 | 16,684 | 1.0 | 14.4 | 11 | 212 | 0.89 (0.37 - 1.88) | 0.77 | 0.77 (0.34 - 1.74) | 0.54 |
| Open Claims | 67,983 | 327,967 | 40.7 | 250.2 | 222 | 3,021 | 0.64 (0.53 - 0.76) | <0.01 | 0.61 (0.50 - 0.75) | <0.01 |
| OptumEHR | 3,137 | 22,354 | 0.8 | 9.5 | 5 | 230 | 0.25 (0.07 - 0.62) | <0.01 | 0.20 (0.07 - 0.58) | <0.01 |
| Meta-analysis | 77,351 | 391,481 | 44.7 | 291.2 | 295 | 4,019 | 0.70 (0.49 - 1.01) | 0.06 | 0.67 (0.46 - 1.00) | 0.05 |

##### 7.2.10 Hypoglycemia

**Supplementary Table 243:** Exposures, outcomes and risk estimates for Hypoglycemia in GLP1RA (T) vs DPP4I (C) new-user cohorts under two different propensity score (PS) adjustment across data sources. We report total patient exposure time in 1,000 patient-year (KYr) units, uncalibrated and calibrated hazard ratios (HRs) and their 95% confidence intervals (CIs). Greyed out entries did not pass study diagnostics and are included only for completeness.

| | Patients | | Exposure (KYr) | | Outcomes | | Uncalibrated | | Calibrated | |
|---|---|---|---|---|---|---|---|---|---|---|
| | T | C | T | C | T | C | HR (95% CI) | $p$ | HR (95% CI) | $p$ |
| **PS-stratification / on-treatment 2 TAR** | | | | | | | | | | |
| OptumDOD | 5,527 | 26,790 | 2.6 | 19.1 | 13 | 96 | 0.83 (0.42 - 1.56) | 0.59 | 0.81 (0.42 - 1.56) | 0.52 |
| MDCR | 1,968 | 17,761 | 1.1 | 15.3 | <5 | 48 | 0.44 (0.07 - 1.57) | 0.30 | 0.38 (0.08 - 1.82) | 0.23 |
| Open Claims | 85,691 | 338,560 | 47.5 | 260.5 | 104 | 562 | 1.09 (0.85 - 1.39) | 0.48 | 1.07 (0.80 - 1.42) | 0.67 |
| OptumEHR | 3,524 | 23,134 | 0.9 | 10.0 | <5 | 47 | 0.24 (0.01 - 1.23) | 0.22 | 0.19 (0.02 - 1.81) | 0.15 |
| Meta-analysis | 89,215 | 361,694 | 48.4 | 270.5 | <109 | 609 | 0.80 (0.24 - 2.63) | 0.71 | 0.79 (0.24 - 2.61) | 0.69 |
| **PS-matching / on-treatment 2 TAR** | | | | | | | | | | |
| OptumDOD | 4,676 | 25,682 | 2.4 | 18.4 | 11 | 92 | 0.69 (0.28 - 1.52) | 0.39 | 0.71 (0.31 - 1.64) | 0.43 |
| MDCR | 1,756 | 16,935 | 1.1 | 14.7 | <5 | 44 | 0.28 (0.02 - 1.51) | 0.28 | 0.24 (0.02 - 2.44) | 0.23 |
| Open Claims | 69,093 | 335,043 | 41.4 | 257.9 | 87 | 551 | 1.12 (0.82 - 1.50) | 0.47 | 1.06 (0.77 - 1.46) | 0.72 |
| OptumEHR | 3,200 | 22,978 | 0.8 | 9.9 | <5 | 46 | 0.19 (0.01 - 1.00) | 0.16 | 0.16 (0.02 - 1.54) | 0.11 |
| Meta-analysis | 78,725 | 400,638 | 45.6 | 300.8 | <108 | 733 | 0.80 (0.44 - 1.45) | 0.47 | 0.77 (0.42 - 1.42) | 0.41 |

###### 7.2.11 Hypotension

**Supplementary Table 244:** Exposures, outcomes and risk estimates for Hypotension in GLP1RA (T) vs DPP4I (C) new-user cohorts under two different propensity score (PS) adjustment across data sources. We report total patient exposure time in 1,000 patient-year (KYr) units, uncalibrated and calibrated hazard ratios (HRs) and their 95% confidence intervals (CIs). Greyed out entries did not pass study diagnostics and are included only for completeness.

| | Patients | | Exposure (KYr) | | Outcomes | | Uncalibrated | | Calibrated | |
|---|---|---|---|---|---|---|---|---|---|---|
| | T | C | T | C | T | C | HR (95% CI) | $p$ | HR (95% CI) | $p$ |
| **PS-stratification / on-treatment 2 TAR** | | | | | | | | | | |
| OptumDOD | 5,272 | 25,208 | 2.5 | 17.7 | 68 | 598 | 1.04 (0.77 - 1.37) | 0.81 | 1.00 (0.75 - 1.34) | 0.88 |
| MDCR | 1,902 | 16,916 | 1.1 | 14.4 | 21 | 373 | 0.94 (0.56 - 1.47) | 0.79 | 0.81 (0.50 - 1.31) | 0.41 |
| Open Claims | 82,616 | 325,192 | 45.7 | 248.9 | 480 | 3,655 | 0.95 (0.85 - 1.06) | 0.39 | 0.93 (0.77 - 1.12) | 0.47 |
| OptumEHR | 3,445 | 22,476 | 0.8 | 9.6 | 16 | 234 | 0.93 (0.52 - 1.55) | 0.78 | 0.72 (0.41 - 1.25) | 0.27 |
| Meta-analysis | 86,061 | 347,668 | 46.6 | 258.6 | 496 | 3,889 | 0.95 (0.85 - 1.06) | 0.37 | 0.94 (0.78 - 1.12) | 0.47 |
| **PS-matching / on-treatment 2 TAR** | | | | | | | | | | |
| OptumDOD | 4,460 | 24,064 | 2.2 | 16.9 | 58 | 534 | 0.91 (0.62 - 1.29) | 0.59 | 0.93 (0.64 - 1.33) | 0.69 |
| MDCR | 1,690 | 16,146 | 1.0 | 13.9 | 20 | 331 | 1.00 (0.55 - 1.72) | 0.99 | 0.87 (0.49 - 1.55) | 0.65 |
| Open Claims | 66,534 | 321,705 | 39.8 | 246.3 | 427 | 3,503 | 1.03 (0.89 - 1.17) | 0.71 | 0.98 (0.82 - 1.16) | 0.82 |
| OptumEHR | 3,127 | 22,351 | 0.8 | 9.6 | 16 | 232 | 1.37 (0.74 - 2.39) | 0.29 | 1.13 (0.63 - 2.02) | 0.70 |
| Meta-analysis | 75,811 | 384,266 | 43.8 | 286.6 | 521 | 4,600 | 1.02 (0.91 - 1.16) | 0.71 | 0.99 (0.82 - 1.18) | 0.86 |

##### 7.2.12 Joint pain

**Supplementary Table 245:** Exposures, outcomes and risk estimates for Joint pain in GLP1RA (T) vs DPP4I (C) new-user cohorts under two different propensity score (PS) adjustment across data sources. We report total patient exposure time in 1,000 patient-year (KYr) units, uncalibrated and calibrated hazard ratios (HRs) and their 95% confidence intervals (CIs). Greyed out entries did not pass study diagnostics and are included only for completeness.

| | Patients | | Exposure (KYr) | | Outcomes | | Uncalibrated | | Calibrated | |
|---|---|---|---|---|---|---|---|---|---|---|
| | T | C | T | C | T | C | HR (95% CI) | $p$ | HR (95% CI) | $p$ |
| **PS-stratification / on-treatment 2 TAR** | | | | | | | | | | |
| OptumDOD | 5,149 | 25,530 | 2.4 | 18.1 | 42 | 297 | 1.01 (0.69 - 1.44) | 0.96 | 0.98 (0.68 - 1.41) | 0.88 |
| MDCR | 1,869 | 17,206 | 1.1 | 14.8 | 8 | 160 | 0.50 (0.22 - 0.99) | 0.07 | 0.43 (0.20 - 0.92) | 0.03 |
| Open Claims | 79,565 | 322,730 | 44.3 | 247.6 | 282 | 1,749 | 0.88 (0.76 - 1.01) | 0.07 | 0.86 (0.69 - 1.06) | 0.16 |
| OptumEHR | 3,316 | 22,151 | 0.8 | 9.5 | 8 | 99 | 1.13 (0.49 - 2.28) | 0.75 | 0.88 (0.41 - 1.90) | 0.75 |
| Meta-analysis | 82,881 | 344,881 | 45.1 | 257.0 | 290 | 1,848 | 0.88 (0.77 - 1.02) | 0.09 | 0.87 (0.71 - 1.06) | 0.17 |
| **PS-matching / on-treatment 2 TAR** | | | | | | | | | | |
| OptumDOD | 4,383 | 24,474 | 2.2 | 17.3 | 35 | 279 | 0.86 (0.53 - 1.36) | 0.54 | 0.88 (0.55 - 1.41) | 0.61 |
| MDCR | 1,678 | 16,305 | 1.0 | 14.1 | 6 | 157 | 0.50 (0.17 - 1.23) | 0.18 | 0.44 (0.16 - 1.18) | 0.11 |
| Open Claims | 64,635 | 319,079 | 38.7 | 244.8 | 242 | 1,725 | 0.83 (0.68 - 0.99) | 0.04 | 0.79 (0.63 - 0.98) | 0.05 |
| OptumEHR | 3,036 | 22,005 | 0.8 | 9.4 | 8 | 97 | 1.16 (0.36 - 3.13) | 0.79 | 0.95 (0.32 - 2.82) | 0.90 |
| Meta-analysis | 73,732 | 381,863 | 42.7 | 285.6 | 291 | 2,258 | 0.83 (0.70 - 0.98) | 0.03 | 0.80 (0.64 - 0.99) | 0.04 |

##### 7.2.13 Lower extremity amputation

**Supplementary Table 246:** Exposures, outcomes and risk estimates for Lower extremity amputation in GLP1RA (T) vs DPP4I (C) new-user cohorts under two different propensity score (PS) adjustment across data sources. We report total patient exposure time in 1,000 patient-year (KYr) units, uncalibrated and calibrated hazard ratios (HRs) and their 95% confidence intervals (CIs). Greyed out entries did not pass study diagnostics and are included only for completeness.

| | Patients | | Exposure (KYr) | | Outcomes | | Uncalibrated | | Calibrated | |
|---|---|---|---|---|---|---|---|---|---|---|
| | T | C | T | C | T | C | HR (95% CI) | $p$ | HR (95% CI) | $p$ |
| **PS-stratification / on-treatment 2 TAR** | | | | | | | | | | |
| OptumDOD | 5,613 | 27,172 | 2.7 | 19.4 | <5 | 13 | 0.53 (0.03 - 3.46) | 0.61 | 0.51 (0.04 - 5.95) | 0.59 |
| MDCR | 1,991 | 17,911 | 1.1 | 15.4 | 0 | 16 | | | | |
| Open Claims | 87,292 | 343,891 | 48.4 | 264.5 | 13 | 139 | 0.79 (0.41 - 1.42) | 0.46 | 0.77 (0.41 - 1.47) | 0.44 |
| OptumEHR | 3,570 | 23,317 | 0.9 | 10.1 | 0 | 7 | | | | |
| Meta-analysis | 87,292 | 343,891 | 48.4 | 264.5 | 13 | 139 | 0.79 (0.41 - 1.42) | 0.46 | 0.78 (0.41 - 1.48) | 0.44 |
| **PS-matching / on-treatment 2 TAR** | | | | | | | | | | |
| OptumDOD | 4,744 | 26,015 | 2.4 | 18.6 | <5 | 13 | | | | |
| MDCR | 1,776 | 17,071 | 1.1 | 14.8 | 0 | 12 | | | | |
| Open Claims | 70,275 | 340,250 | 42.1 | 261.8 | 13 | 133 | 0.88 (0.40 - 1.79) | 0.74 | 0.84 (0.39 - 1.79) | 0.66 |
| OptumEHR | 3,238 | 23,158 | 0.8 | 10.0 | 0 | 7 | | | | |
| Meta-analysis | 70,275 | 340,250 | 42.1 | 261.8 | 13 | 133 | 0.88 (0.40 - 1.79) | 0.74 | 0.85 (0.40 - 1.82) | 0.68 |

##### 7.2.14 Nausea

**Supplementary Table 247:** Exposures, outcomes and risk estimates for Nausea in GLP1RA (T) vs DPP4I (C) new-user cohorts under two different propensity score (PS) adjustment across data sources. We report total patient exposure time in 1,000 patient-year (KYr) units, uncalibrated and calibrated hazard ratios (HRs) and their 95% confidence intervals (CIs). Greyed out entries did not pass study diagnostics and are included only for completeness.

| | Patients | | Exposure (KYr) | | Outcomes | | Uncalibrated | | Calibrated | |
|---|---|---|---|---|---|---|---|---|---|---|
| | T | C | T | C | T | C | HR (95% CI) | $p$ | HR (95% CI) | $p$ |
| **PS-stratification / on-treatment 2 TAR** | | | | | | | | | | |
| OptumDOD | 4,695 | 23,103 | 2.2 | 16.2 | 168 | 734 | 1.47 (1.20 - 1.79) | <0.01 | 1.42 (1.16 - 1.73) | <0.01 |
| MDCR | 1,799 | 15,971 | 1.0 | 13.6 | 38 | 464 | 1.03 (0.70 - 1.46) | 0.89 | 0.89 (0.61 - 1.29) | 0.55 |
| Open Claims | 72,632 | 297,396 | 40.3 | 228.1 | 1,158 | 4,948 | 1.46 (1.36 - 1.58) | <0.01 | 1.43 (1.21 - 1.70) | <0.01 |
| OptumEHR | 3,189 | 21,212 | 0.8 | 9.0 | 55 | 293 | 2.28 (1.63 - 3.14) | <0.01 | 1.77 (1.26 - 2.47) | <0.01 |
| Meta-analysis | 75,821 | 318,608 | 41.0 | 237.1 | 1,213 | 5,241 | 1.77 (1.15 - 2.72) | <0.01 | 1.74 (1.11 - 2.74) | 0.02 |
| **PS-matching / on-treatment 2 TAR** | | | | | | | | | | |
| OptumDOD | 3,990 | 21,854 | 2.0 | 15.3 | 146 | 691 | 1.73 (1.35 - 2.22) | <0.01 | 1.77 (1.38 - 2.28) | <0.01 |
| MDCR | 1,612 | 15,197 | 1.0 | 13.0 | 35 | 422 | 1.10 (0.71 - 1.65) | 0.66 | 0.95 (0.63 - 1.46) | 0.81 |
| Open Claims | 58,946 | 293,827 | 35.2 | 225.4 | 981 | 4,844 | 1.56 (1.42 - 1.71) | <0.01 | 1.48 (1.28 - 1.71) | <0.01 |
| OptumEHR | 2,914 | 21,085 | 0.7 | 8.9 | 47 | 287 | 2.26 (1.49 - 3.38) | <0.01 | 1.85 (1.23 - 2.80) | <0.01 |
| Meta-analysis | 67,462 | 351,963 | 38.9 | 262.6 | 1,209 | 6,244 | 1.61 (1.37 - 1.89) | <0.01 | 1.55 (1.26 - 1.91) | <0.01 |

##### 7.2.15 Peripheral edema

**Supplementary Table 248:** Exposures, outcomes and risk estimates for Peripheral edema in GLP1RA (T) vs DPP4I (C) new-user cohorts under two different propensity score (PS) adjustment across data sources. We report total patient exposure time in 1,000 patient-year (KYr) units, uncalibrated and calibrated hazard ratios (HRs) and their 95% confidence intervals (CIs). Greyed out entries did not pass study diagnostics and are included only for completeness.

| | Patients | | Exposure (KYr) | | Outcomes | | Uncalibrated | | Calibrated | |
|---|---|---|---|---|---|---|---|---|---|---|
| | T | C | T | C | T | C | HR (95% CI) | $p$ | HR (95% CI) | $p$ |
| **PS-stratification / on-treatment 2 TAR** | | | | | | | | | | |
| OptumDOD | 3,920 | 19,394 | 1.8 | 13.0 | 128 | 1,352 | 0.72 (0.58 - 0.88) | <0.01 | 0.70 (0.57 - 0.85) | <0.01 |
| MDCR | 1,471 | 13,304 | 0.8 | 11.0 | 53 | 932 | 1.02 (0.75 - 1.37) | 0.89 | 0.89 (0.65 - 1.20) | 0.45 |
| Open Claims | 58,785 | 250,135 | 32.6 | 187.8 | 930 | 8,815 | 0.80 (0.74 - 0.87) | <0.01 | 0.78 (0.66 - 0.93) | <0.01 |
| OptumEHR | 2,721 | 18,902 | 0.7 | 7.8 | 31 | 455 | 0.98 (0.65 - 1.41) | 0.90 | 0.76 (0.51 - 1.12) | 0.21 |
| Meta-analysis | 61,506 | 269,037 | 33.2 | 195.5 | 961 | 9,270 | 0.81 (0.75 - 0.87) | <0.01 | 0.79 (0.68 - 0.93) | <0.01 |
| **PS-matching / on-treatment 2 TAR** | | | | | | | | | | |
| OptumDOD | 3,331 | 18,604 | 1.6 | 12.5 | 110 | 1,260 | 0.62 (0.48 - 0.80) | <0.01 | 0.63 (0.49 - 0.82) | <0.01 |
| MDCR | 1,315 | 12,867 | 0.8 | 10.7 | 49 | 870 | 0.66 (0.43 - 0.96) | 0.04 | 0.57 (0.38 - 0.85) | <0.01 |
| Open Claims | 48,713 | 246,417 | 28.9 | 185.0 | 820 | 8,482 | 0.80 (0.72 - 0.88) | <0.01 | 0.76 (0.66 - 0.88) | <0.01 |
| OptumEHR | 2,522 | 18,727 | 0.6 | 7.7 | 28 | 445 | 0.85 (0.51 - 1.36) | 0.52 | 0.70 (0.43 - 1.14) | 0.17 |
| Meta-analysis | 55,881 | 296,615 | 31.9 | 215.9 | 1,007 | 11,057 | 0.74 (0.63 - 0.86) | <0.01 | 0.71 (0.58 - 0.87) | <0.01 |

##### 7.2.16 Photosensitivity

**Supplementary Table 249:** Exposures, outcomes and risk estimates for Photosensitivity in GLP1RA (T) vs DPP4I (C) new-user cohorts under two different propensity score (PS) adjustment across data sources. We report total patient exposure time in 1,000 patient-year (KYr) units, uncalibrated and calibrated hazard ratios (HRs) and their 95% confidence intervals (CIs). Greyed out entries did not pass study diagnostics and are included only for completeness.

| | Patients | | Exposure (KYr) | | Outcomes | | Uncalibrated | | Calibrated | |
|---|---|---|---|---|---|---|---|---|---|---|
| | T | C | T | C | T | C | HR (95% CI) | $p$ | HR (95% CI) | $p$ |
| **PS-stratification / on-treatment 2 TAR** | | | | | | | | | | |
| OptumDOD | 5,575 | 26,866 | 2.7 | 19.1 | <5 | 34 | 1.54 (0.32 - 5.10) | 0.54 | 1.49 (0.37 - 5.93) | 0.57 |
| MDCR | 1,951 | 17,593 | 1.1 | 15.1 | <5 | 62 | 0.88 (0.14 - 2.92) | 0.86 | 0.76 (0.17 - 3.45) | 0.72 |
| Open Claims | 85,977 | 339,304 | 47.6 | 260.7 | 18 | 172 | 1.69 (0.93 - 2.87) | 0.07 | 1.65 (0.92 - 2.95) | 0.09 |
| OptumEHR | 3,534 | 23,120 | 0.9 | 10.0 | <5 | 20 | 1.42 (0.20 - 5.67) | 0.68 | 1.10 (0.21 - 5.86) | 0.91 |
| Meta-analysis | 85,977 | 339,304 | 47.6 | 260.7 | 18 | 172 | 1.69 (0.93 - 2.87) | 0.07 | 1.66 (0.93 - 2.97) | 0.09 |
| **PS-matching / on-treatment 2 TAR** | | | | | | | | | | |
| OptumDOD | 4,715 | 25,730 | 2.4 | 18.3 | <5 | 33 | 1.42 (0.20 - 6.07) | 0.69 | 1.45 (0.27 - 7.92) | 0.67 |
| MDCR | 1,735 | 16,778 | 1.0 | 14.5 | <5 | 58 | 0.98 (0.15 - 3.43) | 0.98 | 0.85 (0.18 - 4.01) | 0.84 |
| Open Claims | 69,216 | 335,699 | 41.4 | 258.0 | 15 | 169 | 1.97 (0.99 - 3.66) | 0.04 | 1.88 (0.97 - 3.65) | 0.07 |
| OptumEHR | 3,214 | 22,963 | 0.8 | 9.9 | <5 | 19 | 2.63 (0.30 - 16.8) | 0.35 | 2.16 (0.29 - 16.1) | 0.45 |
| Meta-analysis | 75,666 | 378,207 | 44.8 | 290.8 | <25 | 260 | 1.73 (0.98 - 3.06) | 0.06 | 1.67 (0.93 - 2.99) | 0.09 |

##### 7.2.17 Venous thromboembolism

**Supplementary Table 250:** Exposures, outcomes and risk estimates for Venous thromboembolism in GLP1RA (T) vs DPP4I (C) new-user cohorts under two different propensity score (PS) adjustment across data sources. We report total patient exposure time in 1,000 patient-year (KYr) units, uncalibrated and calibrated hazard ratios (HRs) and their 95% confidence intervals (CIs). Greyed out entries did not pass study diagnostics and are included only for completeness.

| | Patients | | Exposure (KYr) | | Outcomes | | Uncalibrated | | Calibrated | |
|---|---|---|---|---|---|---|---|---|---|---|
| | T | C | T | C | T | C | HR (95% CI) | $p$ | HR (95% CI) | $p$ |
| **PS-stratification / on-treatment 2 TAR** | | | | | | | | | | |
| OptumDOD | 5,345 | 26,088 | 2.6 | 18.5 | 27 | 277 | 0.74 (0.46 - 1.15) | 0.20 | 0.72 (0.46 - 1.14) | 0.16 |
| MDCR | 1,904 | 17,192 | 1.1 | 14.8 | 8 | 190 | 0.62 (0.27 - 1.26) | 0.23 | 0.54 (0.25 - 1.17) | 0.12 |
| Open Claims | 82,626 | 328,935 | 45.9 | 252.7 | 240 | 1,848 | 0.95 (0.81 - 1.10) | 0.51 | 0.93 (0.75 - 1.15) | 0.51 |
| OptumEHR | 3,430 | 22,530 | 0.8 | 9.7 | 11 | 89 | 1.93 (0.93 - 3.65) | 0.06 | 1.50 (0.75 - 2.98) | 0.27 |
| Meta-analysis | 86,056 | 351,465 | 46.8 | 262.4 | 251 | 1,937 | 1.25 (0.63 - 2.46) | 0.52 | 1.23 (0.61 - 2.45) | 0.56 |
| **PS-matching / on-treatment 2 TAR** | | | | | | | | | | |
| OptumDOD | 4,544 | 24,889 | 2.3 | 17.7 | 24 | 255 | 0.84 (0.48 - 1.40) | 0.52 | 0.86 (0.50 - 1.47) | 0.59 |
| MDCR | 1,697 | 16,337 | 1.0 | 14.1 | 7 | 175 | 0.50 (0.17 - 1.21) | 0.17 | 0.43 (0.16 - 1.17) | 0.10 |
| Open Claims | 66,724 | 325,663 | 40.0 | 250.3 | 215 | 1,791 | 1.00 (0.82 - 1.20) | 0.98 | 0.95 (0.76 - 1.18) | 0.67 |
| OptumEHR | 3,126 | 22,391 | 0.8 | 9.6 | 10 | 85 | 2.12 (0.94 - 4.40) | 0.05 | 1.74 (0.81 - 3.77) | 0.16 |
| Meta-analysis | 76,091 | 389,280 | 44.1 | 291.6 | 256 | 2,306 | 1.00 (0.84 - 1.18) | 0.97 | 0.96 (0.77 - 1.19) | 0.72 |

##### 7.2.18 Vomiting

**Supplementary Table 251:** Exposures, outcomes and risk estimates for Vomiting in GLP1RA (T) vs DPP4I (C) new-user cohorts under two different propensity score (PS) adjustment across data sources. We report total patient exposure time in 1,000 patient-year (KYr) units, uncalibrated and calibrated hazard ratios (HRs) and their 95% confidence intervals (CIs). Greyed out entries did not pass study diagnostics and are included only for completeness.

| | Patients | | Exposure (KYr) | | Outcomes | | Uncalibrated | | Calibrated | |
|---|---|---|---|---|---|---|---|---|---|---|
| | T | C | T | C | T | C | HR (95% CI) | $p$ | HR (95% CI) | $p$ |
| **PS-stratification / on-treatment 2 TAR** | | | | | | | | | | |
| OptumDOD | 5,030 | 24,206 | 2.4 | 17.1 | 121 | 567 | 1.63 (1.29 - 2.06) | <0.01 | 1.58 (1.25 - 2.00) | <0.01 |
| MDCR | 1,861 | 16,422 | 1.0 | 14.0 | 30 | 366 | 1.08 (0.70 - 1.61) | 0.72 | 0.94 (0.62 - 1.42) | 0.76 |
| Open Claims | 77,029 | 308,143 | 42.7 | 236.6 | 845 | 3,985 | 1.44 (1.32 - 1.58) | <0.01 | 1.41 (1.18 - 1.68) | <0.01 |
| OptumEHR | 3,338 | 22,042 | 0.8 | 9.4 | 28 | 206 | 2.08 (1.32 - 3.16) | <0.01 | 1.61 (1.04 - 2.51) | 0.05 |
| Meta-analysis | 80,367 | 330,185 | 43.5 | 246.0 | 873 | 4,191 | 1.62 (1.16 - 2.27) | <0.01 | 1.60 (1.11 - 2.29) | 0.01 |
| **PS-matching / on-treatment 2 TAR** | | | | | | | | | | |
| OptumDOD | 4,252 | 23,228 | 2.1 | 16.4 | 108 | 539 | 1.55 (1.15 - 2.07) | <0.01 | 1.59 (1.18 - 2.13) | <0.01 |
| MDCR | 1,660 | 15,632 | 1.0 | 13.4 | 28 | 326 | 1.23 (0.76 - 1.94) | 0.39 | 1.07 (0.67 - 1.71) | 0.78 |
| Open Claims | 62,159 | 304,538 | 37.2 | 233.8 | 716 | 3,876 | 1.49 (1.34 - 1.66) | <0.01 | 1.42 (1.22 - 1.66) | <0.01 |
| OptumEHR | 3,040 | 21,828 | 0.8 | 9.4 | 23 | 203 | 2.02 (1.13 - 3.46) | 0.01 | 1.66 (0.95 - 2.91) | 0.08 |
| Meta-analysis | 71,111 | 365,226 | 41.0 | 272.9 | 875 | 4,944 | 1.50 (1.36 - 1.65) | <0.01 | 1.44 (1.22 - 1.70) | <0.01 |

#### 7.3 GLP1RA vs SU

##### 7.3.1 Abnormal weight gain

**Supplementary Table 252:** Exposures, outcomes and risk estimates for Abnormal weight gain in GLP1RA (T) vs SU (C) new-user cohorts under two different propensity score (PS) adjustment across data sources. We report total patient exposure time in 1,000 patient-year (KYr) units, uncalibrated and calibrated hazard ratios (HRs) and their 95% confidence intervals (CIs). Greyed out entries did not pass study diagnostics and are included only for completeness.

| | Patients | | Exposure (KYr) | | Outcomes | | Uncalibrated | | Calibrated | |
|---|---|---|---|---|---|---|---|---|---|---|
| | T | C | T | C | T | C | HR (95% CI) | $p$ | HR (95% CI) | $p$ |
| **PS-stratification / on-treatment 2 TAR** | | | | | | | | | | |
| OptumDOD | 5,281 | 64,799 | 2.5 | 61.2 | 16 | 369 | 0.91 (0.51 - 1.52) | 0.74 | 0.89 (0.52 - 1.53) | 0.68 |
| MDCR | 2,056 | 36,650 | 1.2 | 33.2 | 6 | 112 | 1.07 (0.39 - 2.45) | 0.89 | 0.92 (0.37 - 2.31) | 0.86 |
| Open Claims | 81,161 | 689,985 | 45.1 | 699.6 | 90 | 1,429 | 0.83 (0.65 - 1.04) | 0.12 | 0.79 (0.59 - 1.05) | 0.11 |
| OptumEHR | 3,350 | 57,248 | 0.8 | 30.6 | 8 | 176 | 1.72 (0.74 - 3.51) | 0.17 | 1.42 (0.60 - 3.40) | 0.47 |
| Meta-analysis | 81,161 | 689,985 | 45.1 | 699.6 | 90 | 1,429 | 0.83 (0.65 - 1.04) | 0.12 | 0.79 (0.58 - 1.06) | 0.11 |
| **PS-matching / on-treatment 2 TAR** | | | | | | | | | | |
| OptumDOD | 4,871 | 53,498 | 2.4 | 49.2 | 13 | 291 | 0.58 (0.25 - 1.20) | 0.18 | 0.59 (0.27 - 1.30) | 0.19 |
| MDCR | 1,896 | 28,137 | 1.1 | 25.8 | 6 | 76 | 1.03 (0.33 - 2.65) | 0.96 | 0.91 (0.32 - 2.57) | 0.85 |
| Open Claims | 75,457 | 642,107 | 42.8 | 650.4 | 72 | 1,263 | 0.68 (0.50 - 0.91) | 0.01 | 0.65 (0.48 - 0.89) | <0.01 |
| OptumEHR | 3,118 | 46,569 | 0.8 | 24.4 | 6 | 137 | 1.46 (0.47 - 3.76) | 0.47 | 1.20 (0.35 - 4.07) | 0.79 |
| Meta-analysis | 85,342 | 770,311 | 47.1 | 749.8 | 97 | 1,767 | 0.72 (0.56 - 0.94) | 0.01 | 0.69 (0.51 - 0.93) | 0.01 |

##### 7.3.2 Abnormal weight loss

**Supplementary Table 253:** Exposures, outcomes and risk estimates for Abnormal weight loss in GLP1RA (T) vs SU (C) new-user cohorts under two different propensity score (PS) adjustment across data sources. We report total patient exposure time in 1,000 patient-year (KYr) units, uncalibrated and calibrated hazard ratios (HRs) and their 95% confidence intervals (CIs). Greyed out entries did not pass study diagnostics and are included only for completeness.

| | Patients | | Exposure (KYr) | | Outcomes | | Uncalibrated | | Calibrated | |
|---|---|---|---|---|---|---|---|---|---|---|
| | T | C | T | C | T | C | HR (95% CI) | $p$ | HR (95% CI) | $p$ |
| **PS-stratification / on-treatment 2 TAR** | | | | | | | | | | |
| OptumDOD | 5,409 | 62,245 | 2.6 | 58.8 | 55 | 1,026 | 1.29 (0.95 - 1.73) | 0.10 | 1.26 (0.93 - 1.70) | 0.15 |
| MDCR | 2,094 | 35,329 | 1.2 | 32.1 | 15 | 391 | 1.03 (0.56 - 1.76) | 0.92 | 0.89 (0.50 - 1.58) | 0.70 |
| Open Claims | 83,942 | 672,909 | 46.5 | 682.8 | 330 | 4,903 | 1.32 (1.17 - 1.49) | <0.01 | 1.25 (1.01 - 1.55) | 0.05 |
| OptumEHR | 3,464 | 56,005 | 0.9 | 29.8 | 7 | 427 | 0.71 (0.30 - 1.45) | 0.41 | 0.59 (0.25 - 1.42) | 0.29 |
| Meta-analysis | 83,942 | 672,909 | 46.5 | 682.8 | 330 | 4,903 | 1.32 (1.17 - 1.49) | <0.01 | 1.25 (1.00 - 1.56) | 0.05 |
| **PS-matching / on-treatment 2 TAR** | | | | | | | | | | |
| OptumDOD | 4,912 | 52,504 | 2.4 | 48.5 | 53 | 793 | 1.20 (0.83 - 1.71) | 0.31 | 1.22 (0.85 - 1.75) | 0.29 |
| MDCR | 1,909 | 27,582 | 1.1 | 25.2 | 14 | 281 | 0.99 (0.48 - 1.86) | 0.99 | 0.88 (0.45 - 1.73) | 0.71 |
| Open Claims | 77,147 | 630,400 | 43.8 | 639.0 | 302 | 4,296 | 1.23 (1.06 - 1.43) | <0.01 | 1.18 (0.99 - 1.40) | 0.09 |
| OptumEHR | 3,182 | 45,909 | 0.8 | 24.0 | 7 | 315 | 1.00 (0.41 - 2.08) | 0.99 | 0.82 (0.29 - 2.30) | 0.73 |
| Meta-analysis | 87,150 | 756,395 | 48.2 | 736.7 | 376 | 5,685 | 1.21 (1.06 - 1.39) | <0.01 | 1.16 (0.95 - 1.41) | 0.15 |

##### 7.3.3 Acute pancreatitis

**Supplementary Table 254:** Exposures, outcomes and risk estimates for Acute pancreatitis in GLP1RA (T) vs SU (C) new-user cohorts under two different propensity score (PS) adjustment across data sources. We report total patient exposure time in 1,000 patient-year (KYr) units, uncalibrated and calibrated hazard ratios (HRs) and their 95% confidence intervals (CIs). Greyed out entries did not pass study diagnostics and are included only for completeness.

| | Patients | | Exposure (KYr) | | Outcomes | | Uncalibrated | | Calibrated | |
|---|---|---|---|---|---|---|---|---|---|---|
| | T | C | T | C | T | C | HR (95% CI) | $p$ | HR (95% CI) | $p$ |
| **PS-stratification / on-treatment 2 TAR** | | | | | | | | | | |
| OptumDOD | 5,591 | 65,491 | 2.7 | 62.1 | 7 | 230 | 0.84 (0.34 - 1.77) | 0.68 | 0.82 (0.36 - 1.86) | 0.63 |
| MDCR | 2,126 | 36,544 | 1.2 | 33.1 | <5 | 119 | 1.05 (0.30 - 2.80) | 0.93 | 0.91 (0.30 - 2.78) | 0.87 |
| Open Claims | 86,463 | 696,817 | 48.0 | 706.3 | 77 | 1,512 | 0.99 (0.76 - 1.25) | 0.91 | 0.93 (0.69 - 1.26) | 0.66 |
| OptumEHR | 3,555 | 58,038 | 0.9 | 31.1 | <5 | 78 | 1.55 (0.36 - 4.58) | 0.50 | 1.29 (0.34 - 4.85) | 0.72 |
| Meta-analysis | 86,463 | 696,817 | 48.0 | 706.3 | 77 | 1,512 | 0.99 (0.76 - 1.25) | 0.91 | 0.93 (0.68 - 1.27) | 0.65 |
| **PS-matching / on-treatment 2 TAR** | | | | | | | | | | |
| OptumDOD | 5,083 | 54,435 | 2.5 | 50.2 | 7 | 170 | 0.68 (0.20 - 1.79) | 0.49 | 0.69 (0.23 - 2.09) | 0.51 |
| MDCR | 1,939 | 28,098 | 1.2 | 25.7 | <5 | 87 | 1.08 (0.25 - 3.30) | 0.91 | 0.95 (0.26 - 3.49) | 0.93 |
| Open Claims | 79,335 | 650,891 | 45.1 | 659.1 | 72 | 1,337 | 1.04 (0.78 - 1.38) | 0.77 | 1.00 (0.74 - 1.35) | 0.94 |
| OptumEHR | 3,266 | 47,414 | 0.8 | 25.0 | <5 | 59 | 1.48 (0.21 - 6.09) | 0.65 | 1.21 (0.20 - 7.24) | 0.84 |
| Meta-analysis | 86,357 | 733,424 | 48.8 | 735.0 | <84 | 1,594 | 1.02 (0.78 - 1.34) | 0.89 | 0.97 (0.71 - 1.32) | 0.86 |

##### 7.3.4 All-cause mortality

**Supplementary Table 255:** Exposures, outcomes and risk estimates for All-cause mortality in GLP1RA (T) vs SU (C) new-user cohorts under two different propensity score (PS) adjustment across data sources. We report total patient exposure time in 1,000 patient-year (KYr) units, uncalibrated and calibrated hazard ratios (HRs) and their 95% confidence intervals (CIs). Greyed out entries did not pass study diagnostics and are included only for completeness.

| | Patients | | Exposure (KYr) | | Outcomes | | Uncalibrated | | Calibrated | |
|---|---|---|---|---|---|---|---|---|---|---|
| | T | C | T | C | T | C | HR (95% CI) | $p$ | HR (95% CI) | $p$ |
| **PS-stratification / on-treatment 2 TAR** | | | | | | | | | | |
| OptumDOD | 5,626 | 66,287 | 2.7 | 62.9 | 15 | 960 | 0.64 (0.36 - 1.04) | 0.10 | 0.62 (0.36 - 1.06) | 0.08 |
| MDCR | 2,135 | 36,988 | 1.2 | 33.6 | <5 | 135 | 1.32 (0.21 - 4.53) | 0.73 | 1.14 (0.24 - 5.34) | 0.87 |
| Open Claims | 87,486 | 707,423 | 48.5 | 717.2 | 0 | 11 | | | | |
| OptumEHR | 3,573 | 58,370 | 0.9 | 31.3 | <5 | 429 | 0.31 (0.05 - 0.98) | 0.12 | 0.26 (0.06 - 1.18) | 0.09 |
| **PS-matching / on-treatment 2 TAR** | | | | | | | | | | |
| OptumDOD | 5,114 | 54,931 | 2.5 | 50.7 | 15 | 632 | 0.88 (0.48 - 1.49) | 0.66 | 0.89 (0.51 - 1.57) | 0.70 |
| MDCR | 1,949 | 28,390 | 1.2 | 26.0 | <5 | 73 | 1.64 (0.26 - 5.85) | 0.54 | 1.45 (0.30 - 6.93) | 0.64 |
| Open Claims | 80,327 | 659,963 | 45.6 | 668.4 | 0 | 9 | | | | |
| OptumEHR | 3,282 | 47,614 | 0.8 | 25.1 | <5 | 289 | 0.38 (0.06 - 1.29) | 0.22 | 0.31 (0.06 - 1.63) | 0.19 |
| Meta-analysis | 10,345 | 130,935 | 4.5 | 101.8 | <25 | 994 | 0.86 (0.52 - 1.41) | 0.55 | 0.82 (0.48 - 1.38) | 0.45 |

##### 7.3.5 Bone fracture

**Supplementary Table 256:** Exposures, outcomes and risk estimates for Bone fracture in GLP1RA (T) vs SU (C) new-user cohorts under two different propensity score (PS) adjustment across data sources. We report total patient exposure time in 1,000 patient-year (KYr) units, uncalibrated and calibrated hazard ratios (HRs) and their 95% confidence intervals (CIs). Greyed out entries did not pass study diagnostics and are included only for completeness.

| | Patients | | Exposure (KYr) | | Outcomes | | Uncalibrated | | Calibrated | |
|---|---|---|---|---|---|---|---|---|---|---|
| | T | C | T | C | T | C | HR (95% CI) | $p$ | HR (95% CI) | $p$ |
| **PS-stratification / on-treatment 2 TAR** | | | | | | | | | | |
| OptumDOD | 4,964 | 59,394 | 2.3 | 55.3 | 86 | 1,909 | 1.04 (0.82 - 1.31) | 0.74 | 1.01 (0.80 - 1.29) | 0.85 |
| MDCR | 1,876 | 32,430 | 1.1 | 28.9 | 31 | 1,020 | 0.78 (0.52 - 1.13) | 0.21 | 0.68 (0.46 - 1.00) | 0.05 |
| Open Claims | 70,749 | 605,180 | 39.0 | 608.8 | 680 | 13,742 | 0.84 (0.78 - 0.92) | <0.01 | 0.80 (0.66 - 0.97) | 0.03 |
| OptumEHR | 3,230 | 53,600 | 0.8 | 28.1 | 17 | 790 | 0.89 (0.51 - 1.42) | 0.64 | 0.73 (0.39 - 1.38) | 0.42 |
| Meta-analysis | 70,749 | 605,180 | 39.0 | 608.8 | 680 | 13,742 | 0.84 (0.78 - 0.92) | <0.01 | 0.80 (0.65 - 0.98) | 0.03 |
| **PS-matching / on-treatment 2 TAR** | | | | | | | | | | |
| OptumDOD | 4,534 | 49,318 | 2.2 | 44.7 | 79 | 1,453 | 1.03 (0.76 - 1.35) | 0.87 | 1.04 (0.78 - 1.39) | 0.79 |
| MDCR | 1,721 | 25,443 | 1.0 | 22.8 | 31 | 772 | 0.89 (0.57 - 1.35) | 0.61 | 0.79 (0.51 - 1.22) | 0.30 |
| Open Claims | 65,480 | 562,671 | 36.9 | 565.4 | 628 | 12,371 | 0.82 (0.74 - 0.90) | <0.01 | 0.78 (0.68 - 0.89) | <0.01 |
| OptumEHR | 2,985 | 43,566 | 0.7 | 22.5 | 15 | 575 | 1.12 (0.61 - 1.90) | 0.70 | 0.92 (0.39 - 2.15) | 0.84 |
| Meta-analysis | 74,720 | 680,998 | 40.9 | 655.4 | 753 | 15,171 | 0.89 (0.76 - 1.04) | 0.13 | 0.85 (0.68 - 1.05) | 0.12 |

##### 7.3.6 Diabetic ketoacidosis

**Supplementary Table 257:** Exposures, outcomes and risk estimates for Diabetic ketoacidosis in GLP1RA (T) vs SU (C) new-user cohorts under two different propensity score (PS) adjustment across data sources. We report total patient exposure time in 1,000 patient-year (KYr) units, uncalibrated and calibrated hazard ratios (HRs) and their 95% confidence intervals (CIs). Greyed out entries did not pass study diagnostics and are included only for completeness.

| | Patients | | Exposure (KYr) | | Outcomes | | Uncalibrated | | Calibrated | |
|---|---|---|---|---|---|---|---|---|---|---|
| | T | C | T | C | T | C | HR (95% CI) | $p$ | HR (95% CI) | $p$ |
| **PS-stratification / on-treatment 2 TAR** | | | | | | | | | | |
| OptumDOD | 5,619 | 66,185 | 2.7 | 62.8 | <5 | 90 | 0.71 (0.16 - 2.19) | 0.61 | 0.70 (0.19 - 2.55) | 0.58 |
| MDCR | 2,134 | 36,952 | 1.2 | 33.6 | 0 | 35 | | | | |
| Open Claims | 87,394 | 706,213 | 48.4 | 716.2 | 28 | 482 | 1.08 (0.70 - 1.60) | 0.72 | 1.02 (0.65 - 1.60) | 0.93 |
| OptumEHR | 3,572 | 58,335 | 0.9 | 31.3 | <5 | 25 | 2.02 (0.11 - 11.0) | 0.55 | 1.67 (0.16 - 17.5) | 0.67 |
| Meta-analysis | 87,394 | 706,213 | 48.4 | 716.2 | 28 | 482 | 1.08 (0.70 - 1.60) | 0.72 | 1.02 (0.65 - 1.60) | 0.93 |
| **PS-matching / on-treatment 2 TAR** | | | | | | | | | | |
| OptumDOD | 5,109 | 54,873 | 2.5 | 50.7 | <5 | 68 | 1.77 (0.36 - 6.15) | 0.43 | 1.80 (0.43 - 7.44) | 0.42 |
| MDCR | 1,949 | 28,376 | 1.2 | 26.0 | 0 | 20 | | | | |
| Open Claims | 80,238 | 658,824 | 45.6 | 667.4 | 28 | 416 | 1.07 (0.67 - 1.63) | 0.78 | 1.02 (0.65 - 1.60) | 0.94 |
| OptumEHR | 3,280 | 47,587 | 0.8 | 25.1 | <5 | 17 | 2.61 (0.14 - 15.9) | 0.43 | 2.14 (0.18 - 25.1) | 0.55 |
| Meta-analysis | 85,347 | 713,697 | 48.1 | 718.1 | <33 | 484 | 1.11 (0.73 - 1.70) | 0.61 | 1.06 (0.68 - 1.66) | 0.79 |

##### 7.3.7 Diarrhea

**Supplementary Table 258:** Exposures, outcomes and risk estimates for Diarrhea in GLP1RA (T) vs SU (C) new-user cohorts under two different propensity score (PS) adjustment across data sources. We report total patient exposure time in 1,000 patient-year (KYr) units, uncalibrated and calibrated hazard ratios (HRs) and their 95% confidence intervals (CIs). Greyed out entries did not pass study diagnostics and are included only for completeness.

| | Patients | | Exposure (KYr) | | Outcomes | | Uncalibrated | | Calibrated | |
|---|---|---|---|---|---|---|---|---|---|---|
| | T | C | T | C | T | C | HR (95% CI) | $p$ | HR (95% CI) | $p$ |
| **PS-stratification / on-treatment 2 TAR** | | | | | | | | | | |
| OptumDOD | 4,704 | 55,726 | 2.2 | 51.3 | 116 | 2,327 | 1.00 (0.81 - 1.22) | 1.00 | 0.98 (0.79 - 1.20) | 0.80 |
| MDCR | 1,864 | 32,189 | 1.1 | 28.7 | 40 | 1,000 | 1.03 (0.72 - 1.43) | 0.88 | 0.89 (0.63 - 1.26) | 0.52 |
| Open Claims | 72,549 | 609,869 | 40.1 | 613.0 | 863 | 12,704 | 1.06 (0.98 - 1.14) | 0.13 | 1.00 (0.83 - 1.22) | 0.89 |
| OptumEHR | 3,136 | 51,403 | 0.8 | 26.4 | 32 | 1,090 | 1.11 (0.75 - 1.59) | 0.58 | 0.92 (0.54 - 1.57) | 0.79 |
| Meta-analysis | 72,549 | 609,869 | 40.1 | 613.0 | 863 | 12,704 | 1.06 (0.98 - 1.14) | 0.13 | 1.00 (0.82 - 1.23) | 0.90 |
| **PS-matching / on-treatment 2 TAR** | | | | | | | | | | |
| OptumDOD | 4,274 | 46,675 | 2.1 | 42.0 | 106 | 1,866 | 0.98 (0.75 - 1.25) | 0.85 | 0.99 (0.77 - 1.28) | 0.84 |
| MDCR | 1,722 | 24,974 | 1.0 | 22.5 | 39 | 742 | 1.26 (0.83 - 1.84) | 0.26 | 1.11 (0.74 - 1.65) | 0.62 |
| Open Claims | 67,070 | 567,394 | 38.0 | 570.0 | 791 | 11,419 | 1.05 (0.95 - 1.14) | 0.34 | 1.00 (0.88 - 1.13) | 0.87 |
| OptumEHR | 2,890 | 42,214 | 0.7 | 21.4 | 29 | 822 | 1.32 (0.85 - 1.96) | 0.20 | 1.08 (0.50 - 2.30) | 0.84 |
| Meta-analysis | 75,956 | 681,257 | 41.8 | 655.8 | 965 | 14,849 | 1.06 (0.97 - 1.15) | 0.20 | 1.01 (0.85 - 1.19) | 0.87 |

##### 7.3.8 Genitourinary infection

**Supplementary Table 259:** Exposures, outcomes and risk estimates for Genitourinary infection in GLP1RA (T) vs SU (C) new-user cohorts under two different propensity score (PS) adjustment across data sources. We report total patient exposure time in 1,000 patient-year (KYr) units, uncalibrated and calibrated hazard ratios (HRs) and their 95% confidence intervals (CIs). Greyed out entries did not pass study diagnostics and are included only for completeness.

| | Patients | | Exposure (KYr) | | Outcomes | | Uncalibrated | | Calibrated | |
|---|---|---|---|---|---|---|---|---|---|---|
| | T | C | T | C | T | C | HR (95% CI) | $p$ | HR (95% CI) | $p$ |
| **PS-stratification / on-treatment 2 TAR** | | | | | | | | | | |
| OptumDOD | 4,094 | 48,387 | 1.9 | 43.5 | 136 | 3,589 | 0.92 (0.76 - 1.10) | 0.37 | 0.90 (0.74 - 1.08) | 0.29 |
| MDCR | 1,633 | 28,860 | 0.9 | 25.6 | 51 | 1,589 | 0.91 (0.66 - 1.21) | 0.53 | 0.79 (0.58 - 1.06) | 0.14 |
| Open Claims | 60,438 | 528,093 | 33.7 | 530.2 | 1,195 | 21,938 | 0.98 (0.92 - 1.05) | 0.64 | 0.93 (0.77 - 1.12) | 0.48 |
| OptumEHR | 2,901 | 48,228 | 0.7 | 24.6 | 44 | 1,537 | 1.27 (0.91 - 1.72) | 0.14 | 1.05 (0.64 - 1.73) | 0.83 |
| Meta-analysis | 60,438 | 528,093 | 33.7 | 530.2 | 1,195 | 21,938 | 0.98 (0.92 - 1.05) | 0.64 | 0.93 (0.77 - 1.13) | 0.47 |
| **PS-matching / on-treatment 2 TAR** | | | | | | | | | | |
| OptumDOD | 3,764 | 39,980 | 1.8 | 35.1 | 130 | 2,703 | 0.89 (0.71 - 1.10) | 0.28 | 0.90 (0.72 - 1.12) | 0.37 |
| MDCR | 1,519 | 22,564 | 0.9 | 20.1 | 49 | 1,131 | 0.96 (0.67 - 1.34) | 0.81 | 0.85 (0.60 - 1.20) | 0.37 |
| Open Claims | 56,324 | 490,658 | 32.0 | 492.1 | 1,105 | 19,476 | 0.92 (0.85 - 0.99) | 0.04 | 0.88 (0.79 - 0.99) | 0.08 |
| OptumEHR | 2,680 | 39,588 | 0.6 | 19.9 | 41 | 1,148 | 1.09 (0.74 - 1.56) | 0.65 | 0.89 (0.43 - 1.87) | 0.79 |
| Meta-analysis | 64,287 | 592,790 | 35.3 | 567.2 | 1,325 | 24,458 | 0.93 (0.86 - 0.99) | 0.03 | 0.88 (0.75 - 1.04) | 0.13 |

##### 7.3.9 Hyperkalemia

**Supplementary Table 260:** Exposures, outcomes and risk estimates for Hyperkalemia in GLP1RA (T) vs SU (C) new-user cohorts under two different propensity score (PS) adjustment across data sources. We report total patient exposure time in 1,000 patient-year (KYr) units, uncalibrated and calibrated hazard ratios (HRs) and their 95% confidence intervals (CIs). Greyed out entries did not pass study diagnostics and are included only for completeness.

| | Patients | | Exposure (KYr) | | Outcomes | | Uncalibrated | | Calibrated | |
|---|---|---|---|---|---|---|---|---|---|---|
| | T | C | T | C | T | C | HR (95% CI) | $p$ | HR (95% CI) | $p$ |
| **PS-stratification / on-treatment 2 TAR** | | | | | | | | | | |
| OptumDOD | 5,330 | 61,816 | 2.5 | 57.3 | 60 | 1,820 | 0.84 (0.63 - 1.09) | 0.20 | 0.81 (0.62 - 1.08) | 0.17 |
| MDCR | 2,092 | 35,848 | 1.2 | 32.3 | 12 | 520 | 0.69 (0.36 - 1.23) | 0.25 | 0.60 (0.32 - 1.11) | 0.11 |
| Open Claims | 84,627 | 682,223 | 46.8 | 686.2 | 256 | 7,987 | 0.65 (0.57 - 0.74) | <0.01 | 0.62 (0.49 - 0.77) | <0.01 |
| OptumEHR | 3,469 | 56,303 | 0.9 | 29.7 | 7 | 600 | 0.52 (0.22 - 1.03) | 0.10 | 0.43 (0.18 - 1.02) | 0.08 |
| Meta-analysis | 84,627 | 682,223 | 46.8 | 686.2 | 256 | 7,987 | 0.65 (0.57 - 0.74) | <0.01 | 0.62 (0.49 - 0.77) | <0.01 |
| **PS-matching / on-treatment 2 TAR** | | | | | | | | | | |
| OptumDOD | 4,839 | 51,345 | 2.3 | 46.5 | 59 | 1,376 | 0.89 (0.64 - 1.20) | 0.45 | 0.90 (0.65 - 1.23) | 0.52 |
| MDCR | 1,912 | 27,765 | 1.1 | 25.2 | 11 | 361 | 0.77 (0.36 - 1.49) | 0.47 | 0.68 (0.33 - 1.39) | 0.29 |
| Open Claims | 77,661 | 637,350 | 44.0 | 640.5 | 242 | 6,937 | 0.62 (0.53 - 0.72) | <0.01 | 0.59 (0.49 - 0.70) | <0.01 |
| OptumEHR | 3,179 | 46,045 | 0.8 | 23.8 | 6 | 435 | 0.50 (0.17 - 1.17) | 0.16 | 0.41 (0.13 - 1.30) | 0.17 |
| Meta-analysis | 87,591 | 762,505 | 48.3 | 736.1 | 318 | 9,109 | 0.70 (0.54 - 0.90) | <0.01 | 0.67 (0.50 - 0.90) | <0.01 |

##### 7.3.10 Hypoglycemia

**Supplementary Table 261:** Exposures, outcomes and risk estimates for Hypoglycemia in GLP1RA (T) vs SU (C) new-user cohorts under two different propensity score (PS) adjustment across data sources. We report total patient exposure time in 1,000 patient-year (KYr) units, uncalibrated and calibrated hazard ratios (HRs) and their 95% confidence intervals (CIs). Greyed out entries did not pass study diagnostics and are included only for completeness.

| | Patients | | Exposure (KYr) | | Outcomes | | Uncalibrated | | Calibrated | |
|---|---|---|---|---|---|---|---|---|---|---|
| | T | C | T | C | T | C | HR (95% CI) | $p$ | HR (95% CI) | $p$ |
| **PS-stratification / on-treatment 2 TAR** | | | | | | | | | | |
| OptumDOD | 5,527 | 65,344 | 2.6 | 61.4 | 13 | 1,633 | 0.20 (0.11 - 0.33) | <0.01 | 0.19 (0.11 - 0.34) | <0.01 |
| MDCR | 2,108 | 36,653 | 1.2 | 33.1 | <5 | 629 | 0.09 (0.01 - 0.28) | <0.01 | 0.08 (0.02 - 0.34) | <0.01 |
| Open Claims | 85,691 | 695,883 | 47.5 | 703.4 | 104 | 8,148 | 0.22 (0.18 - 0.27) | <0.01 | 0.21 (0.16 - 0.27) | <0.01 |
| OptumEHR | 3,524 | 57,838 | 0.9 | 30.8 | <5 | 581 | 0.07 (0.00 - 0.30) | 0.01 | 0.05 (0.01 - 0.50) | 0.01 |
| Meta-analysis | 85,691 | 695,883 | 47.5 | 703.4 | 104 | 8,148 | 0.22 (0.18 - 0.27) | <0.01 | 0.21 (0.16 - 0.27) | <0.01 |
| **PS-matching / on-treatment 2 TAR** | | | | | | | | | | |
| OptumDOD | 5,038 | 54,266 | 2.5 | 49.6 | 11 | 1,228 | 0.16 (0.08 - 0.30) | <0.01 | 0.16 (0.08 - 0.32) | <0.01 |
| MDCR | 1,928 | 28,184 | 1.2 | 25.7 | <5 | 424 | 0.07 (0.00 - 0.31) | 0.02 | 0.06 (0.01 - 0.53) | 0.01 |
| Open Claims | 78,843 | 649,860 | 44.8 | 656.3 | 95 | 7,059 | 0.21 (0.16 - 0.26) | <0.01 | 0.20 (0.15 - 0.25) | <0.01 |
| OptumEHR | 3,246 | 47,264 | 0.8 | 24.7 | <5 | 434 | 0.09 (0.00 - 0.39) | 0.03 | 0.07 (0.01 - 0.69) | 0.03 |
| Meta-analysis | 89,055 | 779,574 | 49.2 | 756.3 | <116 | 9,145 | 0.20 (0.16 - 0.24) | <0.01 | 0.19 (0.14 - 0.24) | <0.01 |

##### 7.3.11 Hypotension

**Supplementary Table 262:** Exposures, outcomes and risk estimates for Hypotension in GLP1RA (T) vs SU (C) new-user cohorts under two different propensity score (PS) adjustment across data sources. We report total patient exposure time in 1,000 patient-year (KYr) units, uncalibrated and calibrated hazard ratios (HRs) and their 95% confidence intervals (CIs). Greyed out entries did not pass study diagnostics and are included only for completeness.

| | Patients | | Exposure (KYr) | | Outcomes | | Uncalibrated | | Calibrated | |
|---|---|---|---|---|---|---|---|---|---|---|
| | T | C | T | C | T | C | HR (95% CI) | $p$ | HR (95% CI) | $p$ |
| **PS-stratification / on-treatment 2 TAR** | | | | | | | | | | |
| OptumDOD | 5,272 | 61,513 | 2.5 | 57.7 | 68 | 1,818 | 0.94 (0.71 - 1.21) | 0.64 | 0.91 (0.70 - 1.19) | 0.53 |
| MDCR | 2,042 | 34,961 | 1.2 | 31.5 | 23 | 726 | 1.10 (0.68 - 1.69) | 0.69 | 0.95 (0.60 - 1.50) | 0.82 |
| Open Claims | 82,616 | 670,684 | 45.7 | 678.3 | 480 | 8,813 | 1.06 (0.95 - 1.17) | 0.28 | 1.00 (0.82 - 1.23) | 0.90 |
| OptumEHR | 3,445 | 56,150 | 0.8 | 29.6 | 16 | 701 | 1.04 (0.59 - 1.70) | 0.89 | 0.86 (0.45 - 1.65) | 0.70 |
| Meta-analysis | 82,616 | 670,684 | 45.7 | 678.3 | 480 | 8,813 | 1.06 (0.95 - 1.17) | 0.28 | 1.00 (0.81 - 1.24) | 0.90 |
| **PS-matching / on-treatment 2 TAR** | | | | | | | | | | |
| OptumDOD | 4,794 | 51,335 | 2.3 | 47.1 | 63 | 1,369 | 0.94 (0.67 - 1.29) | 0.71 | 0.95 (0.69 - 1.32) | 0.78 |
| MDCR | 1,865 | 26,924 | 1.1 | 24.4 | 23 | 471 | 0.99 (0.57 - 1.63) | 0.98 | 0.88 (0.52 - 1.48) | 0.63 |
| Open Claims | 75,965 | 625,422 | 43.1 | 631.7 | 437 | 7,526 | 1.00 (0.88 - 1.13) | 0.99 | 0.95 (0.82 - 1.11) | 0.59 |
| OptumEHR | 3,164 | 45,943 | 0.8 | 23.8 | 16 | 479 | 1.25 (0.69 - 2.12) | 0.44 | 1.02 (0.44 - 2.38) | 0.88 |
| Meta-analysis | 85,788 | 749,624 | 47.4 | 727.1 | 539 | 9,845 | 1.00 (0.90 - 1.12) | 1.00 | 0.95 (0.79 - 1.15) | 0.61 |

##### 7.3.12 Joint pain

**Supplementary Table 263:** Exposures, outcomes and risk estimates for Joint pain in GLP1RA (T) vs SU (C) new-user cohorts under two different propensity score (PS) adjustment across data sources. We report total patient exposure time in 1,000 patient-year (KYr) units, uncalibrated and calibrated hazard ratios (HRs) and their 95% confidence intervals (CIs). Greyed out entries did not pass study diagnostics and are included only for completeness.

| | Patients | | Exposure (KYr) | | Outcomes | | Uncalibrated | | Calibrated | |
|---|---|---|---|---|---|---|---|---|---|---|
| | T | C | T | C | T | C | HR (95% CI) | $p$ | HR (95% CI) | $p$ |
| **PS-stratification / on-treatment 2 TAR** | | | | | | | | | | |
| OptumDOD | 5,149 | 62,677 | 2.4 | 59.1 | 42 | 746 | 1.05 (0.74 - 1.46) | 0.79 | 1.02 (0.72 - 1.44) | 0.88 |
| MDCR | 2,007 | 35,645 | 1.2 | 32.3 | 8 | 284 | 0.58 (0.25 - 1.15) | 0.16 | 0.50 (0.24 - 1.07) | 0.08 |
| Open Claims | 79,565 | 668,637 | 44.3 | 678.3 | 282 | 3,958 | 0.95 (0.83 - 1.08) | 0.44 | 0.90 (0.72 - 1.12) | 0.36 |
| OptumEHR | 3,316 | 55,792 | 0.8 | 29.6 | 8 | 277 | 1.12 (0.49 - 2.19) | 0.77 | 0.93 (0.40 - 2.14) | 0.86 |
| Meta-analysis | 79,565 | 668,637 | 44.3 | 678.3 | 282 | 3,958 | 0.95 (0.83 - 1.08) | 0.44 | 0.90 (0.71 - 1.13) | 0.35 |
| **PS-matching / on-treatment 2 TAR** | | | | | | | | | | |
| OptumDOD | 4,732 | 51,869 | 2.3 | 47.7 | 39 | 606 | 1.14 (0.72 - 1.73) | 0.57 | 1.15 (0.74 - 1.79) | 0.53 |
| MDCR | 1,835 | 27,180 | 1.1 | 24.8 | 6 | 219 | 0.28 (0.07 - 0.80) | 0.04 | 0.25 (0.07 - 0.86) | 0.03 |
| Open Claims | 73,542 | 622,035 | 41.8 | 630.5 | 256 | 3,618 | 0.90 (0.76 - 1.07) | 0.23 | 0.86 (0.72 - 1.04) | 0.16 |
| OptumEHR | 3,052 | 45,476 | 0.8 | 23.7 | 7 | 221 | 0.85 (0.33 - 1.87) | 0.71 | 0.70 (0.24 - 2.03) | 0.56 |
| Meta-analysis | 83,161 | 746,560 | 46.0 | 726.8 | 308 | 4,664 | 0.91 (0.78 - 1.06) | 0.23 | 0.87 (0.70 - 1.07) | 0.19 |

##### 7.3.13 Lower extremity amputation

**Supplementary Table 264:** Exposures, outcomes and risk estimates for Lower extremity amputation in GLP1RA (T) vs SU (C) new-user cohorts under two different propensity score (PS) adjustment across data sources. We report total patient exposure time in 1,000 patient-year (KYr) units, uncalibrated and calibrated hazard ratios (HRs) and their 95% confidence intervals (CIs). Greyed out entries did not pass study diagnostics and are included only for completeness.

| | Patients | | Exposure (KYr) | | Outcomes | | Uncalibrated | | Calibrated | |
|---|---|---|---|---|---|---|---|---|---|---|
| | T | C | T | C | T | C | HR (95% CI) | $p$ | HR (95% CI) | $p$ |
| **PS-stratification / on-treatment 2 TAR** | | | | | | | | | | |
| OptumDOD | 5,613 | 66,153 | 2.7 | 62.8 | <5 | 81 | 0.33 (0.02 - 1.67) | 0.34 | 0.33 (0.03 - 3.11) | 0.33 |
| MDCR | 2,133 | 36,894 | 1.2 | 33.5 | 0 | 33 | | | | |
| Open Claims | 87,292 | 705,868 | 48.4 | 715.5 | 13 | 457 | 0.62 (0.33 - 1.07) | 0.11 | 0.59 (0.32 - 1.08) | 0.09 |
| OptumEHR | 3,570 | 58,305 | 0.9 | 31.3 | 0 | 24 | | | | |
| Meta-analysis | 87,292 | 705,868 | 48.4 | 715.5 | 13 | 457 | 0.62 (0.33 - 1.07) | 0.11 | 0.59 (0.32 - 1.08) | 0.09 |
| **PS-matching / on-treatment 2 TAR** | | | | | | | | | | |
| OptumDOD | 5,103 | 54,852 | 2.5 | 50.7 | <5 | 62 | | | | |
| MDCR | 1,947 | 28,351 | 1.2 | 26.0 | 0 | 26 | | | | |
| Open Claims | 80,154 | 658,385 | 45.5 | 666.7 | 13 | 385 | 0.65 (0.32 - 1.21) | 0.22 | 0.63 (0.32 - 1.23) | 0.18 |
| OptumEHR | 3,280 | 47,533 | 0.8 | 25.0 | 0 | 16 | | | | |
| Meta-analysis | 80,154 | 658,385 | 45.5 | 666.7 | 13 | 385 | 0.65 (0.32 - 1.21) | 0.22 | 0.62 (0.31 - 1.25) | 0.18 |

##### 7.3.14 Nausea

**Supplementary Table 265:** Exposures, outcomes and risk estimates for Nausea in GLP1RA (T) vs SU (C) new-user cohorts under two different propensity score (PS) adjustment across data sources. We report total patient exposure time in 1,000 patient-year (KYr) units, uncalibrated and calibrated hazard ratios (HRs) and their 95% confidence intervals (CIs). Greyed out entries did not pass study diagnostics and are included only for completeness.

| | Patients | | Exposure (KYr) | | Outcomes | | Uncalibrated | | Calibrated | |
|---|---|---|---|---|---|---|---|---|---|---|
| | T | C | T | C | T | C | HR (95% CI) | $p$ | HR (95% CI) | $p$ |
| **PS-stratification / on-treatment 2 TAR** | | | | | | | | | | |
| OptumDOD | 4,695 | 56,566 | 2.2 | 52.7 | 168 | 2,350 | 1.57 (1.32 - 1.87) | <0.01 | 1.53 (1.28 - 1.84) | <0.01 |
| MDCR | 1,932 | 32,962 | 1.1 | 29.7 | 41 | 1,026 | 0.96 (0.67 - 1.33) | 0.81 | 0.83 (0.59 - 1.17) | 0.31 |
| Open Claims | 72,632 | 614,797 | 40.3 | 624.5 | 1,158 | 12,942 | 1.42 (1.33 - 1.52) | <0.01 | 1.35 (1.12 - 1.63) | <0.01 |
| OptumEHR | 3,189 | 53,253 | 0.8 | 27.8 | 55 | 931 | 2.39 (1.76 - 3.19) | <0.01 | 1.98 (1.22 - 3.22) | 0.03 |
| Meta-analysis | 72,632 | 614,797 | 40.3 | 624.5 | 1,158 | 12,942 | 1.42 (1.33 - 1.52) | <0.01 | 1.35 (1.10 - 1.64) | <0.01 |
| **PS-matching / on-treatment 2 TAR** | | | | | | | | | | |
| OptumDOD | 4,285 | 46,818 | 2.1 | 42.5 | 153 | 1,814 | 1.67 (1.34 - 2.06) | <0.01 | 1.69 (1.37 - 2.10) | <0.01 |
| MDCR | 1,775 | 25,420 | 1.1 | 23.1 | 39 | 761 | 0.96 (0.63 - 1.43) | 0.86 | 0.85 (0.57 - 1.28) | 0.45 |
| Open Claims | 67,082 | 572,010 | 38.0 | 580.1 | 1,063 | 11,545 | 1.49 (1.37 - 1.61) | <0.01 | 1.42 (1.26 - 1.60) | <0.01 |
| OptumEHR | 2,932 | 43,666 | 0.7 | 22.5 | 50 | 689 | 2.27 (1.57 - 3.23) | <0.01 | 1.86 (0.90 - 3.87) | 0.19 |
| Meta-analysis | 76,074 | 687,914 | 41.9 | 668.1 | 1,305 | 14,809 | 1.54 (1.16 - 2.06) | <0.01 | 1.47 (1.07 - 2.03) | 0.02 |

##### 7.3.15 Peripheral edema

**Supplementary Table 266:** Exposures, outcomes and risk estimates for Peripheral edema in GLP1RA (T) vs SU (C) new-user cohorts under two different propensity score (PS) adjustment across data sources. We report total patient exposure time in 1,000 patient-year (KYr) units, uncalibrated and calibrated hazard ratios (HRs) and their 95% confidence intervals (CIs). Greyed out entries did not pass study diagnostics and are included only for completeness.

| | Patients | | Exposure (KYr) | | Outcomes | | Uncalibrated | | Calibrated | |
|---|---|---|---|---|---|---|---|---|---|---|
| | T | C | T | C | T | C | HR (95% CI) | $p$ | HR (95% CI) | $p$ |
| **PS-stratification / on-treatment 2 TAR** | | | | | | | | | | |
| OptumDOD | 3,920 | 48,322 | 1.8 | 42.8 | 128 | 4,419 | 0.67 (0.56 - 0.81) | <0.01 | 0.66 (0.54 - 0.79) | <0.01 |
| MDCR | 1,587 | 27,977 | 0.9 | 24.0 | 58 | 2,192 | 0.81 (0.60 - 1.06) | 0.13 | 0.70 (0.53 - 0.93) | 0.02 |
| Open Claims | 58,785 | 526,028 | 32.6 | 521.7 | 930 | 24,638 | 0.75 (0.70 - 0.80) | <0.01 | 0.71 (0.58 - 0.85) | <0.01 |
| OptumEHR | 2,721 | 47,402 | 0.7 | 23.5 | 31 | 1,880 | 0.72 (0.49 - 1.02) | 0.08 | 0.60 (0.35 - 1.01) | 0.13 |
| Meta-analysis | 58,785 | 526,028 | 32.6 | 521.7 | 930 | 24,638 | 0.75 (0.70 - 0.80) | <0.01 | 0.71 (0.58 - 0.86) | <0.01 |
| **PS-matching / on-treatment 2 TAR** | | | | | | | | | | |
| OptumDOD | 3,601 | 40,002 | 1.7 | 34.7 | 118 | 3,378 | 0.67 (0.53 - 0.84) | <0.01 | 0.68 (0.54 - 0.85) | <0.01 |
| MDCR | 1,475 | 21,406 | 0.8 | 18.5 | 52 | 1,541 | 0.68 (0.46 - 0.97) | 0.04 | 0.60 (0.41 - 0.87) | <0.01 |
| Open Claims | 54,897 | 488,033 | 31.0 | 483.6 | 875 | 21,457 | 0.72 (0.66 - 0.78) | <0.01 | 0.68 (0.61 - 0.77) | <0.01 |
| OptumEHR | 2,528 | 38,649 | 0.6 | 19.0 | 29 | 1,355 | 0.72 (0.46 - 1.08) | 0.13 | 0.59 (0.28 - 1.27) | 0.27 |
| Meta-analysis | 62,501 | 588,090 | 34.2 | 555.8 | 1,074 | 27,731 | 0.71 (0.66 - 0.76) | <0.01 | 0.68 (0.57 - 0.80) | <0.01 |

##### 7.3.16 Photosensitivity

**Supplementary Table 267:** Exposures, outcomes and risk estimates for Photosensitivity in GLP1RA (T) vs SU (C) new-user cohorts under two different propensity score (PS) adjustment across data sources. We report total patient exposure time in 1,000 patient-year (KYr) units, uncalibrated and calibrated hazard ratios (HRs) and their 95% confidence intervals (CIs). Greyed out entries did not pass study diagnostics and are included only for completeness.

| | Patients | | Exposure (KYr) | | Outcomes | | Uncalibrated | | Calibrated | |
|---|---|---|---|---|---|---|---|---|---|---|
| | T | C | T | C | T | C | HR (95% CI) | $p$ | HR (95% CI) | $p$ |
| **PS-stratification / on-treatment 2 TAR** | | | | | | | | | | |
| OptumDOD | 5,575 | 65,490 | 2.7 | 61.9 | <5 | 115 | 1.61 (0.38 - 4.59) | 0.45 | 1.57 (0.45 - 5.46) | 0.48 |
| MDCR | 2,092 | 36,363 | 1.2 | 32.9 | <5 | 106 | 0.93 (0.22 - 2.73) | 0.91 | 0.80 (0.23 - 2.86) | 0.73 |
| Open Claims | 85,977 | 697,868 | 47.6 | 706.3 | 18 | 398 | 1.72 (0.99 - 2.81) | 0.04 | 1.63 (0.94 - 2.83) | 0.08 |
| OptumEHR | 3,534 | 57,741 | 0.9 | 30.9 | <5 | 75 | 2.92 (0.46 - 9.87) | 0.17 | 2.41 (0.50 - 11.7) | 0.29 |
| Meta-analysis | 85,977 | 697,868 | 47.6 | 706.3 | 18 | 398 | 1.72 (0.99 - 2.81) | 0.04 | 1.63 (0.94 - 2.83) | 0.08 |
| **PS-matching / on-treatment 2 TAR** | | | | | | | | | | |
| OptumDOD | 5,067 | 54,291 | 2.5 | 50.0 | <5 | 78 | 1.93 (0.41 - 6.26) | 0.34 | 1.96 (0.50 - 7.61) | 0.33 |
| MDCR | 1,909 | 27,815 | 1.2 | 25.4 | <5 | 72 | 1.74 (0.39 - 5.46) | 0.41 | 1.53 (0.41 - 5.76) | 0.53 |
| Open Claims | 79,003 | 650,817 | 44.8 | 658.0 | 16 | 332 | 1.98 (1.04 - 3.50) | 0.03 | 1.89 (1.02 - 3.49) | 0.04 |
| OptumEHR | 3,251 | 47,209 | 0.8 | 24.9 | <5 | 47 | 1.64 (0.22 - 6.92) | 0.57 | 1.34 (0.22 - 8.35) | 0.76 |
| Meta-analysis | 85,979 | 732,923 | 48.4 | 733.4 | <26 | 482 | 1.93 (1.16 - 3.23) | 0.01 | 1.84 (1.08 - 3.14) | 0.02 |

##### 7.3.17 Venous thromboembolism

**Supplementary Table 268:** Exposures, outcomes and risk estimates for Venous thromboembolism in GLP1RA (T) vs SU (C) new-user cohorts under two different propensity score (PS) adjustment across data sources. We report total patient exposure time in 1,000 patient-year (KYr) units, uncalibrated and calibrated hazard ratios (HRs) and their 95% confidence intervals (CIs). Greyed out entries did not pass study diagnostics and are included only for completeness.

| | Patients | | Exposure (KYr) | | Outcomes | | Uncalibrated | | Calibrated | |
|---|---|---|---|---|---|---|---|---|---|---|
| | T | C | T | C | T | C | HR (95% CI) | $p$ | HR (95% CI) | $p$ |
| **PS-stratification / on-treatment 2 TAR** | | | | | | | | | | |
| OptumDOD | 5,345 | 63,537 | 2.6 | 59.8 | 27 | 903 | 0.86 (0.56 - 1.28) | 0.47 | 0.84 (0.55 - 1.27) | 0.41 |
| MDCR | 2,043 | 35,380 | 1.2 | 31.9 | 9 | 499 | 0.53 (0.25 - 1.01) | 0.08 | 0.46 (0.23 - 0.93) | 0.03 |
| Open Claims | 82,626 | 676,546 | 45.9 | 684.2 | 240 | 5,628 | 0.77 (0.67 - 0.88) | <0.01 | 0.73 (0.58 - 0.91) | <0.01 |
| OptumEHR | 3,430 | 56,341 | 0.8 | 29.9 | 11 | 375 | 1.44 (0.72 - 2.59) | 0.26 | 1.19 (0.57 - 2.51) | 0.68 |
| Meta-analysis | 82,626 | 676,546 | 45.9 | 684.2 | 240 | 5,628 | 0.77 (0.67 - 0.88) | <0.01 | 0.73 (0.58 - 0.92) | <0.01 |
| **PS-matching / on-treatment 2 TAR** | | | | | | | | | | |
| OptumDOD | 4,865 | 52,372 | 2.4 | 47.9 | 25 | 649 | 0.89 (0.53 - 1.41) | 0.63 | 0.90 (0.55 - 1.47) | 0.68 |
| MDCR | 1,868 | 27,207 | 1.1 | 24.8 | 9 | 341 | 0.61 (0.26 - 1.26) | 0.22 | 0.54 (0.24 - 1.19) | 0.13 |
| Open Claims | 76,024 | 630,961 | 43.3 | 637.3 | 227 | 4,871 | 0.78 (0.66 - 0.92) | <0.01 | 0.75 (0.62 - 0.90) | <0.01 |
| OptumEHR | 3,157 | 46,144 | 0.8 | 24.1 | 9 | 272 | 1.38 (0.62 - 2.70) | 0.39 | 1.13 (0.43 - 2.98) | 0.82 |
| Meta-analysis | 85,914 | 756,684 | 47.6 | 734.0 | 270 | 6,133 | 0.80 (0.69 - 0.93) | <0.01 | 0.77 (0.62 - 0.94) | 0.01 |

##### 7.3.18 Vomiting

**Supplementary Table 269:** Exposures, outcomes and risk estimates for Vomiting in GLP1RA (T) vs SU (C) new-user cohorts under two different propensity score (PS) adjustment across data sources. We report total patient exposure time in 1,000 patient-year (KYr) units, uncalibrated and calibrated hazard ratios (HRs) and their 95% confidence intervals (CIs). Greyed out entries did not pass study diagnostics and are included only for completeness.

| | Patients | | Exposure (KYr) | | Outcomes | | Uncalibrated | | Calibrated | |
|---|---|---|---|---|---|---|---|---|---|---|
| | T | C | T | C | T | C | HR (95% CI) | $p$ | HR (95% CI) | $p$ |
| **PS-stratification / on-treatment 2 TAR** | | | | | | | | | | |
| OptumDOD | 5,030 | 59,035 | 2.4 | 55.4 | 121 | 1,837 | 1.43 (1.15 - 1.75) | <0.01 | 1.39 (1.13 - 1.71) | <0.01 |
| MDCR | 2,000 | 33,833 | 1.1 | 30.5 | 33 | 868 | 0.98 (0.66 - 1.41) | 0.92 | 0.85 (0.58 - 1.24) | 0.42 |
| Open Claims | 77,029 | 635,649 | 42.7 | 645.9 | 845 | 10,262 | 1.40 (1.29 - 1.51) | <0.01 | 1.32 (1.09 - 1.60) | <0.01 |
| OptumEHR | 3,338 | 55,066 | 0.8 | 29.1 | 28 | 642 | 1.94 (1.26 - 2.86) | <0.01 | 1.61 (0.92 - 2.81) | 0.18 |
| Meta-analysis | 77,029 | 635,649 | 42.7 | 645.9 | 845 | 10,262 | 1.40 (1.29 - 1.51) | <0.01 | 1.32 (1.08 - 1.61) | <0.01 |
| **PS-matching / on-treatment 2 TAR** | | | | | | | | | | |
| OptumDOD | 4,567 | 49,167 | 2.2 | 44.9 | 111 | 1,404 | 1.45 (1.12 - 1.86) | <0.01 | 1.47 (1.14 - 1.90) | <0.01 |
| MDCR | 1,838 | 26,183 | 1.1 | 23.7 | 32 | 653 | 1.03 (0.64 - 1.57) | 0.91 | 0.91 (0.58 - 1.42) | 0.67 |
| Open Claims | 70,903 | 593,021 | 40.2 | 601.6 | 786 | 9,093 | 1.41 (1.28 - 1.55) | <0.01 | 1.35 (1.19 - 1.53) | <0.01 |
| OptumEHR | 3,065 | 45,087 | 0.8 | 23.4 | 27 | 467 | 2.06 (1.28 - 3.22) | <0.01 | 1.69 (0.77 - 3.71) | 0.28 |
| Meta-analysis | 80,373 | 713,458 | 44.3 | 693.7 | 956 | 11,617 | 1.42 (1.30 - 1.54) | <0.01 | 1.35 (1.14 - 1.60) | <0.01 |

#### 7.4 SGLT2I vs DPP4I

##### 7.4.1 Abnormal weight gain

**Supplementary Table 270:** Exposures, outcomes and risk estimates for Abnormal weight gain in SGLT2I (T) vs DPP4I (C) new-user cohorts under two different propensity score (PS) adjustment across data sources. We report total patient exposure time in 1,000 patient-year (KYr) units, uncalibrated and calibrated hazard ratios (HRs) and their 95% confidence intervals (CIs). Greyed out entries did not pass study diagnostics and are included only for completeness.

| | Patients | | Exposure (KYr) | | Outcomes | | Uncalibrated | | Calibrated | |
|---|---|---|---|---|---|---|---|---|---|---|
| | T | C | T | C | T | C | HR (95% CI) | $p$ | HR (95% CI) | $p$ |
| **PS-stratification / on-treatment 2 TAR** | | | | | | | | | | |
| OptumDOD | 11,275 | 21,260 | 6.1 | 14.5 | 18 | 75 | 0.68 (0.38 - 1.15) | 0.17 | 0.65 (0.37 - 1.13) | 0.13 |
| MDCR | 3,444 | 9,872 | 2.2 | 7.7 | <5 | 20 | 1.00 (0.26 - 2.98) | 1.00 | 1.01 (0.30 - 3.41) | 0.96 |
| Open Claims | 177,824 | 332,881 | 109.8 | 255.7 | 118 | 431 | 0.79 (0.62 - 0.98) | 0.04 | 0.80 (0.59 - 1.08) | 0.15 |
| OptumEHR | 7,560 | 19,569 | 1.9 | 8.6 | 5 | 32 | 1.10 (0.35 - 2.85) | 0.86 | 0.96 (0.34 - 2.74) | 0.93 |
| VA | 7,350 | 14,321 | 4.0 | 12.1 | <10 | <10 | 1.50 (0.40 - 5.54) | 0.55 | 1.52 (0.41 - 5.62) | 0.53 |
| GermanyDA | 4,137 | 10,716 | 3.0 | 10.7 | 0 | 12 | | | | |
| SIDIAP | 3,707 | 20,269 | 4.0 | 36.5 | <5 | 49 | 1.29 (0.37 - 3.37) | 0.66 | 1.33 (0.44 - 4.02) | 0.61 |
| Meta-analysis | 211,160 | 418,172 | 128.1 | 335.0 | <161 | <617 | 0.81 (0.66 - 0.99) | 0.04 | 0.81 (0.62 - 1.07) | 0.13 |
| **PS-matching / on-treatment 2 TAR** | | | | | | | | | | |
| OptumDOD | 9,027 | 20,878 | 5.3 | 14.2 | 17 | 72 | 0.73 (0.35 - 1.42) | 0.37 | 0.69 (0.34 - 1.38) | 0.30 |
| MDCR | 2,867 | 9,846 | 2.0 | 7.6 | <5 | 20 | 1.18 (0.24 - 4.63) | 0.82 | 1.19 (0.27 - 5.22) | 0.82 |
| Open Claims | 135,354 | 331,887 | 90.5 | 255.0 | 98 | 428 | 0.86 (0.62 - 1.18) | 0.37 | 0.86 (0.58 - 1.26) | 0.45 |
| OptumEHR | 6,731 | 19,496 | 1.7 | 8.6 | 5 | 31 | 2.17 (0.49 - 9.06) | 0.30 | 2.01 (0.46 - 8.66) | 0.35 |
| VA | 4,515 | 13,918 | 2.8 | 11.5 | <10 | <10 | 2.52 (0.36 - 23.2) | 0.38 | 2.68 (0.33 - 21.5) | 0.36 |
| GermanyDA | 3,406 | 10,647 | 2.6 | 10.6 | 0 | 12 | | | | |
| SIDIAP | 3,081 | 19,652 | 3.6 | 34.9 | <5 | 44 | 1.94 (0.52 - 5.59) | 0.27 | 2.07 (0.63 - 6.76) | 0.23 |
| Meta-analysis | 157,060 | 401,759 | 103.2 | 320.3 | <130 | 595 | 0.92 (0.70 - 1.20) | 0.53 | 0.91 (0.64 - 1.29) | 0.58 |

##### 7.4.2 Abnormal weight loss

**Supplementary Table 271:** Exposures, outcomes and risk estimates for Abnormal weight loss in SGLT2I (T) vs DPP4I (C) new-user cohorts under two different propensity score (PS) adjustment across data sources. We report total patient exposure time in 1,000 patient-year (KYr) units, uncalibrated and calibrated hazard ratios (HRs) and their 95% confidence intervals (CIs). Greyed out entries did not pass study diagnostics and are included only for completeness.

| | Patients | | Exposure (KYr) | | Outcomes | | Uncalibrated | | Calibrated | |
|---|---|---|---|---|---|---|---|---|---|---|
| | T | C | T | C | T | C | HR (95% CI) | $p$ | HR (95% CI) | $p$ |
| **PS-stratification / on-treatment 2 TAR** | | | | | | | | | | |
| OptumDOD | 11,094 | 20,351 | 6.0 | 13.8 | 149 | 294 | 1.34 (1.07 - 1.67) | <0.01 | 1.29 (1.03 - 1.61) | 0.03 |
| MDCR | 3,392 | 9,519 | 2.2 | 7.3 | 40 | 126 | 1.35 (0.90 - 1.99) | 0.14 | 1.37 (0.92 - 2.04) | 0.13 |
| Open Claims | 176,400 | 324,139 | 108.8 | 248.1 | 970 | 2,374 | 1.14 (1.04 - 1.24) | <0.01 | 1.16 (0.94 - 1.43) | 0.19 |
| OptumEHR | 7,530 | 19,221 | 1.9 | 8.4 | 27 | 139 | 1.11 (0.69 - 1.71) | 0.66 | 0.96 (0.61 - 1.52) | 0.85 |
| VA | 7,184 | 13,921 | 3.9 | 11.7 | 62 | 134 | 1.75 (1.16 - 2.60) | <0.01 | 1.77 (1.19 - 2.65) | <0.01 |
| GermanyDA | 4,053 | 10,447 | 2.9 | 10.3 | 33 | 84 | 1.16 (0.72 - 1.84) | 0.52 | 1.06 (0.66 - 1.69) | 0.76 |
| SIDIAP | 3,673 | 20,124 | 4.0 | 36.1 | 32 | 202 | 1.16 (0.74 - 1.77) | 0.50 | 1.21 (0.78 - 1.87) | 0.42 |
| Meta-analysis | 213,326 | 417,722 | 129.6 | 335.8 | 1,313 | 3,353 | 1.23 (1.09 - 1.38) | <0.01 | 1.23 (0.98 - 1.53) | 0.07 |
| **PS-matching / on-treatment 2 TAR** | | | | | | | | | | |
| OptumDOD | 8,849 | 20,010 | 5.2 | 13.6 | 135 | 283 | 1.25 (0.93 - 1.68) | 0.13 | 1.18 (0.88 - 1.59) | 0.28 |
| MDCR | 2,818 | 9,480 | 2.0 | 7.3 | 38 | 126 | 1.17 (0.67 - 1.99) | 0.57 | 1.17 (0.68 - 2.03) | 0.58 |
| Open Claims | 133,917 | 323,111 | 89.5 | 247.3 | 802 | 2,354 | 1.13 (1.00 - 1.26) | 0.04 | 1.12 (0.88 - 1.43) | 0.36 |
| OptumEHR | 6,698 | 19,157 | 1.7 | 8.4 | 25 | 138 | 1.17 (0.65 - 2.05) | 0.59 | 1.08 (0.61 - 1.92) | 0.79 |
| VA | 4,387 | 13,541 | 2.7 | 11.2 | 46 | 128 | 1.89 (1.14 - 3.08) | 0.01 | 2.01 (1.22 - 3.31) | <0.01 |
| GermanyDA | 3,336 | 10,392 | 2.5 | 10.3 | 24 | 84 | 1.08 (0.60 - 1.91) | 0.80 | 1.03 (0.57 - 1.84) | 0.78 |
| SIDIAP | 3,060 | 19,520 | 3.5 | 34.5 | 26 | 197 | 1.05 (0.60 - 1.74) | 0.86 | 1.12 (0.66 - 1.91) | 0.69 |
| Meta-analysis | 163,065 | 415,211 | 107.2 | 332.6 | 1,096 | 3,310 | 1.16 (1.05 - 1.28) | <0.01 | 1.15 (0.90 - 1.46) | 0.26 |

##### 7.4.3 Acute pancreatitis

**Supplementary Table 272:** Exposures, outcomes and risk estimates for Acute pancreatitis in SGLT2I (T) vs DPP4I (C) new-user cohorts under two different propensity score (PS) adjustment across data sources. We report total patient exposure time in 1,000 patient-year (KYr) units, uncalibrated and calibrated hazard ratios (HRs) and their 95% confidence intervals (CIs). Greyed out entries did not pass study diagnostics and are included only for completeness.

| | Patients | | Exposure (KYr) | | Outcomes | | Uncalibrated | | Calibrated | |
|---|---|---|---|---|---|---|---|---|---|---|
| | T | C | T | C | T | C | HR (95% CI) | $p$ | HR (95% CI) | $p$ |
| **PS-stratification / on-treatment 2 TAR** | | | | | | | | | | |
| OptumDOD | 11,512 | 21,644 | 6.3 | 14.8 | 18 | 53 | 0.97 (0.52 - 1.72) | 0.91 | 0.93 (0.51 - 1.69) | 0.81 |
| MDCR | 3,480 | 9,962 | 2.3 | 7.7 | <5 | 20 | 1.11 (0.31 - 3.12) | 0.86 | 1.12 (0.35 - 3.57) | 0.85 |
| Open Claims | 182,041 | 338,295 | 112.4 | 259.7 | 146 | 589 | 0.73 (0.59 - 0.89) | <0.01 | 0.74 (0.56 - 0.98) | 0.04 |
| OptumEHR | 7,801 | 19,970 | 1.9 | 8.8 | <5 | 17 | 0.83 (0.18 - 2.68) | 0.78 | 0.72 (0.19 - 2.76) | 0.63 |
| VA | 7,438 | 14,470 | 4.1 | 12.2 | <10 | 15 | 0.19 (0.01 - 1.26) | 0.19 | 0.20 (0.02 - 2.23) | 0.19 |
| GermanyDA | 4,175 | 10,792 | 3.0 | 10.7 | 0 | 0 | | | | |
| SIDIAP | 3,736 | 20,481 | 4.1 | 36.8 | 0 | 18 | | | | |
| Meta-analysis | 212,272 | 404,341 | 126.9 | 303.2 | <184 | 694 | 0.75 (0.62 - 0.90) | <0.01 | 0.75 (0.58 - 0.98) | 0.04 |
| **PS-matching / on-treatment 2 TAR** | | | | | | | | | | |
| OptumDOD | 9,221 | 21,253 | 5.4 | 14.5 | 15 | 49 | 0.91 (0.43 - 1.85) | 0.80 | 0.86 (0.41 - 1.79) | 0.68 |
| MDCR | 2,902 | 9,936 | 2.1 | 7.7 | <5 | 20 | 1.61 (0.33 - 6.21) | 0.53 | 1.61 (0.37 - 7.03) | 0.53 |
| Open Claims | 138,478 | 337,302 | 92.6 | 259.0 | 122 | 586 | 0.76 (0.58 - 0.99) | 0.04 | 0.76 (0.54 - 1.06) | 0.12 |
| OptumEHR | 6,949 | 19,896 | 1.8 | 8.8 | <5 | 17 | 0.78 (0.17 - 2.56) | 0.71 | 0.72 (0.19 - 2.77) | 0.63 |
| VA | 4,551 | 14,060 | 2.9 | 11.7 | <10 | 15 | | | | |
| GermanyDA | 3,434 | 10,733 | 2.6 | 10.7 | 0 | 0 | | | | |
| SIDIAP | 3,110 | 19,850 | 3.6 | 35.2 | 0 | 17 | | | | |
| Meta-analysis | 157,550 | 388,387 | 101.9 | 290.0 | <147 | 672 | 0.79 (0.62 - 1.01) | 0.06 | 0.78 (0.57 - 1.08) | 0.14 |

##### 7.4.4 All-cause mortality

**Supplementary Table 273:** Exposures, outcomes and risk estimates for All-cause mortality in SGLT2I (T) vs DPP4I (C) new-user cohorts under two different propensity score (PS) adjustment across data sources. We report total patient exposure time in 1,000 patient-year (KYr) units, uncalibrated and calibrated hazard ratios (HRs) and their 95% confidence intervals (CIs). Greyed out entries did not pass study diagnostics and are included only for completeness.

| | Patients | | Exposure (KYr) | | Outcomes | | Uncalibrated | | Calibrated | |
|---|---|---|---|---|---|---|---|---|---|---|
| | T | C | T | C | T | C | HR (95% CI) | $p$ | HR (95% CI) | $p$ |
| **PS-stratification / on-treatment 2 TAR** | | | | | | | | | | |
| OptumDOD | 11,614 | 21,849 | 6.3 | 15.0 | 40 | 188 | 0.72 (0.49 - 1.04) | 0.08 | 0.69 (0.47 - 1.01) | 0.06 |
| MDCR | 3,510 | 10,031 | 2.3 | 7.8 | <5 | 21 | 1.38 (0.21 - 4.95) | 0.69 | 1.39 (0.29 - 6.70) | 0.68 |
| Open Claims | 184,807 | 342,501 | 114.0 | 263.0 | 0 | <5 | | | | |
| OptumEHR | 7,828 | 20,055 | 2.0 | 8.8 | 7 | 105 | 0.44 (0.18 - 0.93) | 0.05 | 0.39 (0.17 - 0.88) | 0.03 |
| VA | 7,529 | 14,605 | 4.1 | 12.3 | 51 | 187 | 0.58 (0.39 - 0.84) | <0.01 | 0.58 (0.40 - 0.85) | <0.01 |
| GermanyDA | 4,175 | 10,792 | 3.0 | 10.7 | <5 | 9 | 1.19 (0.23 - 4.92) | 0.82 | 1.09 (0.23 - 5.06) | 0.91 |
| SIDIAP | 3,746 | 20,524 | 4.1 | 36.9 | 86 | 1,138 | 0.85 (0.66 - 1.07) | 0.18 | 0.88 (0.68 - 1.13) | 0.36 |
| Meta-analysis | 34,227 | 87,064 | 18.8 | 80.8 | <189 | 1,639 | 0.71 (0.57 - 0.89) | <0.01 | 0.71 (0.53 - 0.96) | 0.02 |
| **PS-matching / on-treatment 2 TAR** | | | | | | | | | | |
| OptumDOD | 9,301 | 21,451 | 5.5 | 14.7 | 32 | 178 | 0.55 (0.32 - 0.91) | 0.03 | 0.52 (0.31 - 0.88) | 0.02 |
| MDCR | 2,927 | 10,005 | 2.1 | 7.8 | <5 | 20 | 1.12 (0.17 - 4.61) | 0.89 | 1.13 (0.21 - 5.93) | 0.89 |
| Open Claims | 140,534 | 341,473 | 94.0 | 262.3 | 0 | <5 | | | | |
| OptumEHR | 6,967 | 19,980 | 1.8 | 8.8 | 6 | 101 | 0.47 (0.17 - 1.10) | 0.11 | 0.44 (0.17 - 1.10) | 0.08 |
| VA | 4,600 | 14,186 | 2.9 | 11.8 | 32 | 181 | 0.53 (0.31 - 0.87) | 0.02 | 0.56 (0.34 - 0.95) | 0.04 |
| GermanyDA | 3,434 | 10,733 | 2.6 | 10.7 | <5 | 9 | 0.83 (0.11 - 4.58) | 0.85 | 0.79 (0.12 - 5.09) | 0.81 |
| SIDIAP | 3,116 | 19,888 | 3.6 | 35.3 | 67 | 1,054 | 0.70 (0.51 - 0.95) | 0.03 | 0.75 (0.55 - 1.03) | 0.10 |
| Meta-analysis | 26,911 | 85,510 | 15.8 | 78.2 | <142 | 1,534 | 0.63 (0.50 - 0.79) | <0.01 | 0.62 (0.45 - 0.85) | <0.01 |

##### 7.4.5 Bone fracture

**Supplementary Table 274:** Exposures, outcomes and risk estimates for Bone fracture in SGLT2I (T) vs DPP4I (C) new-user cohorts under two different propensity score (PS) adjustment across data sources. We report total patient exposure time in 1,000 patient-year (KYr) units, uncalibrated and calibrated hazard ratios (HRs) and their 95% confidence intervals (CIs). Greyed out entries did not pass study diagnostics and are included only for completeness.

| | Patients | | Exposure (KYr) | | Outcomes | | Uncalibrated | | Calibrated | |
|---|---|---|---|---|---|---|---|---|---|---|
| | T | C | T | C | T | C | HR (95% CI) | $p$ | HR (95% CI) | $p$ |
| **PS-stratification / on-treatment 2 TAR** | | | | | | | | | | |
| OptumDOD | 10,423 | 19,212 | 5.6 | 12.9 | 172 | 452 | 1.04 (0.85 - 1.26) | 0.72 | 1.00 (0.82 - 1.21) | 0.87 |
| MDCR | 3,143 | 8,713 | 2.0 | 6.7 | 62 | 226 | 1.15 (0.83 - 1.57) | 0.38 | 1.17 (0.85 - 1.60) | 0.36 |
| Open Claims | 155,987 | 289,690 | 96.2 | 220.4 | 1,571 | 5,120 | 0.91 (0.85 - 0.97) | <0.01 | 0.93 (0.76 - 1.13) | 0.48 |
| OptumEHR | 7,175 | 18,312 | 1.8 | 8.0 | 33 | 179 | 1.19 (0.78 - 1.76) | 0.41 | 1.03 (0.69 - 1.56) | 0.84 |
| VA | 6,699 | 13,160 | 3.7 | 11.1 | 35 | 103 | 0.98 (0.58 - 1.61) | 0.94 | 0.99 (0.60 - 1.66) | 0.91 |
| GermanyDA | 3,627 | 9,166 | 2.6 | 9.0 | 46 | 234 | 0.88 (0.61 - 1.24) | 0.49 | 0.80 (0.56 - 1.14) | 0.26 |
| SIDIAP | 3,268 | 17,523 | 3.6 | 31.2 | 50 | 609 | 0.91 (0.66 - 1.24) | 0.58 | 0.95 (0.69 - 1.31) | 0.73 |
| Meta-analysis | 190,322 | 375,776 | 115.4 | 299.3 | 1,969 | 6,923 | 0.96 (0.88 - 1.04) | 0.32 | 0.96 (0.78 - 1.18) | 0.68 |
| **PS-matching / on-treatment 2 TAR** | | | | | | | | | | |
| OptumDOD | 8,335 | 18,905 | 4.9 | 12.7 | 155 | 437 | 1.36 (1.04 - 1.76) | 0.02 | 1.28 (0.99 - 1.67) | 0.08 |
| MDCR | 2,618 | 8,623 | 1.8 | 6.6 | 60 | 220 | 1.50 (0.98 - 2.27) | 0.06 | 1.50 (0.98 - 2.29) | 0.08 |
| Open Claims | 118,920 | 288,961 | 79.4 | 219.9 | 1,346 | 5,090 | 0.93 (0.85 - 1.01) | 0.08 | 0.92 (0.73 - 1.16) | 0.51 |
| OptumEHR | 6,382 | 18,251 | 1.6 | 8.0 | 28 | 178 | 1.10 (0.66 - 1.80) | 0.70 | 1.02 (0.62 - 1.68) | 0.84 |
| VA | 4,146 | 12,701 | 2.6 | 10.5 | 21 | 95 | 0.73 (0.37 - 1.37) | 0.35 | 0.78 (0.40 - 1.49) | 0.46 |
| GermanyDA | 2,976 | 9,138 | 2.2 | 8.9 | 42 | 229 | 0.87 (0.55 - 1.34) | 0.55 | 0.83 (0.53 - 1.30) | 0.45 |
| SIDIAP | 2,700 | 17,009 | 3.1 | 30.0 | 40 | 562 | 0.99 (0.66 - 1.45) | 0.96 | 1.06 (0.71 - 1.57) | 0.74 |
| Meta-analysis | 146,077 | 373,588 | 95.6 | 296.7 | 1,692 | 6,811 | 1.06 (0.89 - 1.26) | 0.53 | 1.05 (0.79 - 1.39) | 0.76 |

##### 7.4.6 Diabetic ketoacidosis

**Supplementary Table 275:** Exposures, outcomes and risk estimates for Diabetic ketoacidosis in SGLT2I (T) vs DPP4I (C) new-user cohorts under two different propensity score (PS) adjustment across data sources. We report total patient exposure time in 1,000 patient-year (KYr) units, uncalibrated and calibrated hazard ratios (HRs) and their 95% confidence intervals (CIs). Greyed out entries did not pass study diagnostics and are included only for completeness.

| | Patients | | Exposure (KYr) | | Outcomes | | Uncalibrated | | Calibrated | |
|---|---|---|---|---|---|---|---|---|---|---|
| | T | C | T | C | T | C | HR (95% CI) | $p$ | HR (95% CI) | $p$ |
| **PS-stratification / on-treatment 2 TAR** | | | | | | | | | | |
| OptumDOD | 11,606 | 21,817 | 6.3 | 14.9 | 17 | 26 | 1.82 (0.89 - 3.64) | 0.10 | 1.75 (0.86 - 3.54) | 0.12 |
| MDCR | 3,506 | 10,008 | 2.3 | 7.8 | <5 | 7 | 0.87 (0.04 - 6.24) | 0.92 | 0.88 (0.07 - 10.8) | 0.92 |
| Open Claims | 184,585 | 342,039 | 113.9 | 262.7 | 114 | 189 | 1.65 (1.26 - 2.16) | <0.01 | 1.68 (1.21 - 2.34) | <0.01 |
| OptumEHR | 7,821 | 20,047 | 2.0 | 8.8 | <5 | 8 | 0.85 (0.12 - 4.24) | 0.86 | 0.74 (0.12 - 4.51) | 0.75 |
| VA | 7,524 | 14,600 | 4.1 | 12.3 | <10 | <10 | | 0.06 | | 0.06 |
| GermanyDA | 4,175 | 10,792 | 3.0 | 10.7 | 0 | 0 | | | | |
| SIDIAP | 3,746 | 20,524 | 4.1 | 36.9 | 0 | 0 | | | | |
| Meta-analysis | 203,715 | 378,456 | 124.3 | 289.9 | <141 | <225 | 1.70 (1.32 - 2.19) | <0.01 | 1.70 (1.24 - 2.33) | <0.01 |
| **PS-matching / on-treatment 2 TAR** | | | | | | | | | | |
| OptumDOD | 9,292 | 21,424 | 5.5 | 14.6 | 14 | 25 | 1.63 (0.67 - 3.82) | 0.27 | 1.54 (0.65 - 3.67) | 0.33 |
| MDCR | 2,922 | 9,982 | 2.1 | 7.7 | <5 | 7 | 0.82 (0.04 - 6.32) | 0.88 | 0.82 (0.06 - 10.5) | 0.88 |
| Open Claims | 140,351 | 341,015 | 93.8 | 262.0 | 96 | 189 | 1.39 (0.99 - 1.95) | 0.06 | 1.39 (0.93 - 2.06) | 0.12 |
| OptumEHR | 6,961 | 19,973 | 1.8 | 8.8 | <5 | 8 | 0.60 (0.03 - 5.43) | 0.71 | 0.56 (0.04 - 7.73) | 0.66 |
| VA | 4,598 | 14,181 | 2.9 | 11.8 | <10 | <10 | | 0.19 | | 0.19 |
| GermanyDA | 3,434 | 10,733 | 2.6 | 10.7 | 0 | 0 | | | | |
| SIDIAP | 3,116 | 19,888 | 3.6 | 35.3 | 0 | 0 | | | | |
| Meta-analysis | 149,643 | 362,439 | 99.3 | 276.6 | 110 | 214 | 1.42 (1.04 - 1.95) | 0.03 | 1.40 (0.96 - 2.06) | 0.08 |

##### 7.4.7 Diarrhea

**Supplementary Table 276:** Exposures, outcomes and risk estimates for Diarrhea in SGLT2I (T) vs DPP4I (C) new-user cohorts under two different propensity score (PS) adjustment across data sources. We report total patient exposure time in 1,000 patient-year (KYr) units, uncalibrated and calibrated hazard ratios (HRs) and their 95% confidence intervals (CIs). Greyed out entries did not pass study diagnostics and are included only for completeness.

| | Patients | | Exposure (KYr) | | Outcomes | | Uncalibrated | | Calibrated | |
|---|---|---|---|---|---|---|---|---|---|---|
| | T | C | T | C | T | C | HR (95% CI) | $p$ | HR (95% CI) | $p$ |
| **PS-stratification / on-treatment 2 TAR** | | | | | | | | | | |
| OptumDOD | 9,875 | 17,929 | 5.3 | 12.0 | 207 | 554 | 0.95 (0.79 - 1.13) | 0.56 | 0.91 (0.76 - 1.09) | 0.34 |
| MDCR | 3,032 | 8,448 | 2.0 | 6.4 | 52 | 243 | 0.74 (0.53 - 1.02) | 0.07 | 0.75 (0.54 - 1.04) | 0.10 |
| Open Claims | 158,033 | 291,037 | 97.6 | 221.2 | 1,489 | 4,928 | 0.86 (0.81 - 0.92) | <0.01 | 0.88 (0.72 - 1.08) | 0.24 |
| OptumEHR | 6,910 | 17,477 | 1.7 | 7.5 | 64 | 267 | 1.23 (0.90 - 1.65) | 0.18 | 1.07 (0.79 - 1.45) | 0.70 |
| VA | 6,417 | 12,666 | 3.5 | 10.7 | 59 | 204 | 0.80 (0.56 - 1.14) | 0.23 | 0.82 (0.57 - 1.16) | 0.28 |
| GermanyDA | 4,111 | 10,663 | 3.0 | 10.6 | 6 | 20 | 2.13 (0.62 - 6.26) | 0.20 | 1.94 (0.61 - 6.15) | 0.26 |
| SIDIAP | 3,550 | 19,181 | 3.9 | 34.2 | 32 | 466 | 0.50 (0.33 - 0.73) | <0.01 | 0.52 (0.35 - 0.77) | <0.01 |
| Meta-analysis | 191,928 | 377,401 | 116.9 | 302.5 | 1,909 | 6,682 | 0.86 (0.70 - 1.05) | 0.15 | 0.86 (0.65 - 1.14) | 0.30 |
| **PS-matching / on-treatment 2 TAR** | | | | | | | | | | |
| OptumDOD | 7,859 | 17,646 | 4.6 | 11.8 | 187 | 535 | 0.96 (0.76 - 1.22) | 0.76 | 0.91 (0.72 - 1.15) | 0.46 |
| MDCR | 2,517 | 8,362 | 1.8 | 6.3 | 47 | 237 | 0.70 (0.45 - 1.06) | 0.10 | 0.70 (0.46 - 1.08) | 0.13 |
| Open Claims | 119,369 | 290,175 | 79.9 | 220.6 | 1,259 | 4,901 | 0.90 (0.82 - 0.98) | 0.02 | 0.89 (0.71 - 1.12) | 0.36 |
| OptumEHR | 6,117 | 17,357 | 1.5 | 7.5 | 56 | 266 | 1.27 (0.83 - 1.89) | 0.26 | 1.17 (0.77 - 1.77) | 0.48 |
| VA | 3,980 | 12,179 | 2.5 | 10.1 | 40 | 196 | 0.77 (0.47 - 1.21) | 0.26 | 0.81 (0.51 - 1.30) | 0.41 |
| GermanyDA | 3,389 | 10,605 | 2.6 | 10.5 | 6 | 20 | 3.71 (1.02 - 12.7) | 0.04 | 3.52 (1.00 - 12.4) | 0.05 |
| SIDIAP | 2,946 | 18,612 | 3.4 | 32.7 | 23 | 446 | 0.30 (0.16 - 0.52) | <0.01 | 0.32 (0.18 - 0.58) | <0.01 |
| Meta-analysis | 146,177 | 374,936 | 96.3 | 299.5 | 1,618 | 6,601 | 0.85 (0.59 - 1.24) | 0.41 | 0.84 (0.55 - 1.30) | 0.45 |

##### 7.4.8 Genitourinary infection

**Supplementary Table 277:** Exposures, outcomes and risk estimates for Genitourinary infection in SGLT2I (T) vs DPP4I (C) new-user cohorts under two different propensity score (PS) adjustment across data sources. We report total patient exposure time in 1,000 patient-year (KYr) units, uncalibrated and calibrated hazard ratios (HRs) and their 95% confidence intervals (CIs). Greyed out entries did not pass study diagnostics and are included only for completeness.

| | Patients | | Exposure (KYr) | | Outcomes | | Uncalibrated | | Calibrated | |
|---|---|---|---|---|---|---|---|---|---|---|
| | T | C | T | C | T | C | HR (95% CI) | $p$ | HR (95% CI) | $p$ |
| **PS-stratification / on-treatment 2 TAR** | | | | | | | | | | |
| OptumDOD | 9,018 | 15,153 | 4.8 | 10.0 | 292 | 848 | 0.91 (0.78 - 1.05) | 0.20 | 0.87 (0.75 - 1.01) | 0.10 |
| MDCR | 2,894 | 7,564 | 1.9 | 5.7 | 89 | 346 | 0.99 (0.76 - 1.28) | 0.96 | 1.01 (0.77 - 1.31) | 0.85 |
| Open Claims | 137,023 | 245,572 | 85.9 | 184.4 | 2,538 | 8,773 | 0.86 (0.82 - 0.91) | <0.01 | 0.88 (0.72 - 1.07) | 0.23 |
| OptumEHR | 6,565 | 16,105 | 1.6 | 6.8 | 80 | 421 | 1.00 (0.77 - 1.30) | 0.97 | 0.88 (0.67 - 1.14) | 0.39 |
| VA | 6,609 | 12,626 | 3.6 | 10.6 | 43 | 177 | 0.61 (0.39 - 0.91) | 0.02 | 0.61 (0.40 - 0.94) | 0.03 |
| GermanyDA | 3,202 | 8,059 | 2.3 | 7.7 | 81 | 322 | 1.18 (0.89 - 1.55) | 0.23 | 1.07 (0.81 - 1.42) | 0.59 |
| SIDIAP | 2,815 | 15,113 | 3.1 | 26.5 | 136 | 1,264 | 1.08 (0.88 - 1.31) | 0.44 | 1.12 (0.92 - 1.37) | 0.34 |
| Meta-analysis | 168,126 | 320,192 | 103.2 | 251.8 | 3,259 | 12,151 | 0.94 (0.85 - 1.04) | 0.23 | 0.94 (0.76 - 1.17) | 0.58 |
| **PS-matching / on-treatment 2 TAR** | | | | | | | | | | |
| OptumDOD | 7,055 | 14,990 | 4.1 | 9.9 | 254 | 823 | 0.95 (0.78 - 1.14) | 0.57 | 0.89 (0.74 - 1.08) | 0.28 |
| MDCR | 2,386 | 7,538 | 1.7 | 5.7 | 79 | 344 | 0.83 (0.58 - 1.15) | 0.27 | 0.83 (0.59 - 1.17) | 0.33 |
| Open Claims | 102,627 | 244,869 | 69.8 | 184.0 | 2,172 | 8,707 | 0.92 (0.87 - 0.99) | 0.02 | 0.92 (0.74 - 1.15) | 0.49 |
| OptumEHR | 5,785 | 16,004 | 1.4 | 6.8 | 75 | 409 | 1.07 (0.76 - 1.48) | 0.70 | 0.99 (0.71 - 1.38) | 0.78 |
| VA | 4,043 | 12,255 | 2.6 | 10.2 | 23 | 166 | 0.54 (0.29 - 0.94) | 0.04 | 0.57 (0.32 - 1.03) | 0.07 |
| GermanyDA | 2,640 | 8,032 | 2.0 | 7.6 | 73 | 321 | 1.12 (0.80 - 1.55) | 0.49 | 1.07 (0.77 - 1.48) | 0.61 |
| SIDIAP | 2,325 | 14,631 | 2.8 | 25.4 | 116 | 1,184 | 1.06 (0.83 - 1.34) | 0.62 | 1.13 (0.89 - 1.44) | 0.37 |
| Meta-analysis | 126,861 | 318,319 | 84.4 | 249.6 | 2,792 | 11,954 | 0.93 (0.88 - 0.99) | 0.02 | 0.92 (0.74 - 1.16) | 0.49 |

##### 7.4.9 Hyperkalemia

**Supplementary Table 278:** Exposures, outcomes and risk estimates for Hyperkalemia in SGLT2I (T) vs DPP4I (C) new-user cohorts under two different propensity score (PS) adjustment across data sources. We report total patient exposure time in 1,000 patient-year (KYr) units, uncalibrated and calibrated hazard ratios (HRs) and their 95% confidence intervals (CIs). Greyed out entries did not pass study diagnostics and are included only for completeness.

| | Patients | | Exposure (KYr) | | Outcomes | | Uncalibrated | | Calibrated | |
|---|---|---|---|---|---|---|---|---|---|---|
| | T | C | T | C | T | C | HR (95% CI) | $p$ | HR (95% CI) | $p$ |
| **PS-stratification / on-treatment 2 TAR** | | | | | | | | | | |
| OptumDOD | 10,938 | 20,150 | 5.9 | 13.5 | 145 | 483 | 0.82 (0.67 - 1.01) | 0.06 | 0.79 (0.64 - 0.97) | 0.03 |
| MDCR | 3,408 | 9,650 | 2.2 | 7.4 | 24 | 145 | 0.72 (0.44 - 1.15) | 0.19 | 0.73 (0.45 - 1.19) | 0.22 |
| Open Claims | 178,385 | 329,485 | 110.1 | 251.0 | 755 | 3,131 | 0.67 (0.62 - 0.74) | <0.01 | 0.69 (0.56 - 0.85) | <0.01 |
| OptumEHR | 7,603 | 19,295 | 1.9 | 8.4 | 26 | 208 | 0.61 (0.39 - 0.93) | 0.03 | 0.53 (0.34 - 0.82) | <0.01 |
| VA | 6,726 | 13,484 | 3.7 | 11.2 | 64 | 167 | 0.97 (0.67 - 1.40) | 0.87 | 0.98 (0.68 - 1.43) | 0.87 |
| GermanyDA | 3,819 | 9,818 | 2.7 | 9.6 | 41 | 183 | 0.67 (0.45 - 0.97) | 0.04 | 0.61 (0.41 - 0.89) | 0.02 |
| SIDIAP | 3,509 | 18,795 | 3.8 | 33.4 | 56 | 461 | 0.91 (0.65 - 1.24) | 0.56 | 0.94 (0.68 - 1.30) | 0.71 |
| Meta-analysis | 214,388 | 420,677 | 130.5 | 334.6 | 1,111 | 4,778 | 0.75 (0.66 - 0.85) | <0.01 | 0.75 (0.60 - 0.94) | 0.01 |
| **PS-matching / on-treatment 2 TAR** | | | | | | | | | | |
| OptumDOD | 8,761 | 19,826 | 5.1 | 13.3 | 126 | 466 | 0.77 (0.58 - 1.02) | 0.07 | 0.73 (0.55 - 0.96) | 0.03 |
| MDCR | 2,844 | 9,629 | 2.0 | 7.4 | 23 | 145 | 0.66 (0.35 - 1.17) | 0.17 | 0.66 (0.36 - 1.21) | 0.20 |
| Open Claims | 136,020 | 328,574 | 90.9 | 250.4 | 583 | 3,092 | 0.66 (0.58 - 0.74) | <0.01 | 0.66 (0.51 - 0.83) | <0.01 |
| OptumEHR | 6,746 | 19,232 | 1.7 | 8.3 | 21 | 208 | 0.53 (0.29 - 0.91) | 0.03 | 0.49 (0.28 - 0.87) | 0.02 |
| VA | 4,211 | 13,135 | 2.6 | 10.7 | 37 | 160 | 1.04 (0.64 - 1.65) | 0.87 | 1.11 (0.69 - 1.78) | 0.69 |
| GermanyDA | 3,155 | 9,764 | 2.4 | 9.6 | 29 | 180 | 0.59 (0.35 - 0.96) | 0.04 | 0.56 (0.34 - 0.93) | 0.03 |
| SIDIAP | 2,919 | 18,303 | 3.4 | 32.2 | 38 | 436 | 0.84 (0.53 - 1.28) | 0.43 | 0.90 (0.58 - 1.39) | 0.64 |
| Meta-analysis | 164,656 | 418,463 | 108.2 | 331.9 | 857 | 4,687 | 0.70 (0.62 - 0.78) | <0.01 | 0.69 (0.54 - 0.88) | <0.01 |

##### 7.4.10 Hypoglycemia

**Supplementary Table 279:** Exposures, outcomes and risk estimates for Hypoglycemia in SGLT2I (T) vs DPP4I (C) new-user cohorts under two different propensity score (PS) adjustment across data sources. We report total patient exposure time in 1,000 patient-year (KYr) units, uncalibrated and calibrated hazard ratios (HRs) and their 95% confidence intervals (CIs). Greyed out entries did not pass study diagnostics and are included only for completeness.

| | Patients | | Exposure (KYr) | | Outcomes | | Uncalibrated | | Calibrated | |
|---|---|---|---|---|---|---|---|---|---|---|
| | T | C | T | C | T | C | HR (95% CI) | $p$ | HR (95% CI) | $p$ |
| **PS-stratification / on-treatment 2 TAR** | | | | | | | | | | |
| OptumDOD | 11,439 | 21,482 | 6.2 | 14.7 | 36 | 85 | 1.21 (0.78 - 1.85) | 0.39 | 1.16 (0.75 - 1.80) | 0.51 |
| MDCR | 3,474 | 9,898 | 2.3 | 7.7 | <5 | 28 | 0.37 (0.09 - 1.11) | 0.13 | 0.37 (0.10 - 1.35) | 0.13 |
| Open Claims | 181,772 | 336,560 | 112.2 | 258.6 | 159 | 558 | 0.73 (0.60 - 0.89) | <0.01 | 0.75 (0.57 - 0.98) | 0.04 |
| OptumEHR | 7,748 | 19,865 | 1.9 | 8.7 | <5 | 44 | 0.34 (0.08 - 0.99) | 0.09 | 0.30 (0.08 - 1.05) | 0.06 |
| VA | 7,425 | 14,475 | 4.0 | 12.2 | <10 | 17 | 2.37 (0.69 - 7.41) | 0.15 | 2.41 (0.73 - 7.89) | 0.15 |
| GermanyDA | 4,153 | 10,715 | 3.0 | 10.6 | <5 | 21 | 1.14 (0.26 - 3.49) | 0.84 | 1.04 (0.28 - 3.80) | 0.92 |
| SIDIAP | 3,739 | 20,488 | 4.1 | 36.8 | 0 | 13 | | | | |
| Meta-analysis | 216,011 | 412,995 | 129.7 | 312.6 | <220 | 753 | 0.86 (0.57 - 1.31) | 0.49 | 0.86 (0.55 - 1.36) | 0.53 |
| **PS-matching / on-treatment 2 TAR** | | | | | | | | | | |
| OptumDOD | 9,150 | 21,098 | 5.4 | 14.4 | 31 | 82 | 1.24 (0.72 - 2.11) | 0.42 | 1.17 (0.69 - 2.01) | 0.56 |
| MDCR | 2,896 | 9,872 | 2.1 | 7.6 | <5 | 28 | 0.69 (0.10 - 3.38) | 0.68 | 0.69 (0.12 - 4.13) | 0.69 |
| Open Claims | 138,168 | 335,542 | 92.4 | 257.9 | 134 | 554 | 0.75 (0.57 - 0.97) | 0.03 | 0.75 (0.53 - 1.04) | 0.10 |
| OptumEHR | 6,900 | 19,792 | 1.7 | 8.7 | <5 | 44 | 0.37 (0.06 - 1.40) | 0.22 | 0.34 (0.07 - 1.70) | 0.19 |
| VA | 4,550 | 13,997 | 2.8 | 11.6 | <10 | 16 | 2.35 (0.53 - 9.46) | 0.25 | 2.49 (0.59 - 10.5) | 0.22 |
| GermanyDA | 3,415 | 10,656 | 2.6 | 10.6 | <5 | 21 | 0.31 (0.02 - 1.72) | 0.33 | 0.30 (0.03 - 3.02) | 0.31 |
| SIDIAP | 3,112 | 19,855 | 3.6 | 35.2 | 0 | 13 | | | | |
| Meta-analysis | 165,079 | 410,957 | 107.1 | 310.8 | <190 | 745 | 0.88 (0.60 - 1.29) | 0.50 | 0.87 (0.56 - 1.35) | 0.53 |

##### 7.4.11 Hypotension

**Supplementary Table 280:** Exposures, outcomes and risk estimates for Hypotension in SGLT2I (T) vs DPP4I (C) new-user cohorts under two different propensity score (PS) adjustment across data sources. We report total patient exposure time in 1,000 patient-year (KYr) units, uncalibrated and calibrated hazard ratios (HRs) and their 95% confidence intervals (CIs). Greyed out entries did not pass study diagnostics and are included only for completeness.

| | Patients | | Exposure (KYr) | | Outcomes | | Uncalibrated | | Calibrated | |
|---|---|---|---|---|---|---|---|---|---|---|
| | T | C | T | C | T | C | HR (95% CI) | $p$ | HR (95% CI) | $p$ |
| **PS-stratification / on-treatment 2 TAR** | | | | | | | | | | |
| OptumDOD | 10,826 | 20,117 | 5.9 | 13.6 | 195 | 459 | 1.17 (0.97 - 1.41) | 0.10 | 1.12 (0.93 - 1.36) | 0.26 |
| MDCR | 3,342 | 9,375 | 2.2 | 7.2 | 53 | 209 | 1.18 (0.83 - 1.65) | 0.35 | 1.19 (0.84 - 1.69) | 0.33 |
| Open Claims | 174,134 | 323,216 | 107.6 | 247.1 | 1,261 | 3,635 | 0.96 (0.89 - 1.03) | 0.26 | 0.98 (0.80 - 1.20) | 0.83 |
| OptumEHR | 7,517 | 19,273 | 1.9 | 8.4 | 35 | 211 | 0.83 (0.56 - 1.22) | 0.37 | 0.73 (0.49 - 1.08) | 0.14 |
| VA | 6,697 | 13,463 | 3.7 | 11.3 | 103 | 150 | 1.40 (1.01 - 1.94) | 0.05 | 1.42 (1.02 - 1.97) | 0.04 |
| GermanyDA | 4,027 | 10,466 | 2.9 | 10.4 | 19 | 67 | 0.78 (0.42 - 1.40) | 0.42 | 0.71 (0.39 - 1.29) | 0.28 |
| SIDIAP | 3,705 | 20,297 | 4.0 | 36.4 | 19 | 101 | 1.48 (0.79 - 2.62) | 0.20 | 1.53 (0.84 - 2.79) | 0.17 |
| Meta-analysis | 210,248 | 416,207 | 128.1 | 334.3 | 1,685 | 4,832 | 1.07 (0.93 - 1.24) | 0.33 | 1.07 (0.85 - 1.36) | 0.55 |
| **PS-matching / on-treatment 2 TAR** | | | | | | | | | | |
| OptumDOD | 8,713 | 19,803 | 5.1 | 13.3 | 158 | 438 | 1.13 (0.88 - 1.43) | 0.33 | 1.07 (0.83 - 1.36) | 0.63 |
| MDCR | 2,781 | 9,357 | 2.0 | 7.2 | 46 | 207 | 1.30 (0.83 - 2.01) | 0.24 | 1.31 (0.84 - 2.04) | 0.27 |
| Open Claims | 133,224 | 322,265 | 89.1 | 246.4 | 991 | 3,597 | 0.98 (0.89 - 1.08) | 0.73 | 0.98 (0.78 - 1.24) | 0.86 |
| OptumEHR | 6,705 | 19,208 | 1.7 | 8.4 | 31 | 208 | 0.75 (0.45 - 1.20) | 0.25 | 0.69 (0.43 - 1.13) | 0.16 |
| VA | 4,192 | 13,096 | 2.6 | 10.8 | 55 | 145 | 1.38 (0.88 - 2.15) | 0.16 | 1.47 (0.94 - 2.30) | 0.11 |
| GermanyDA | 3,327 | 10,411 | 2.5 | 10.3 | 15 | 66 | 0.72 (0.34 - 1.45) | 0.38 | 0.69 (0.33 - 1.42) | 0.33 |
| SIDIAP | 3,086 | 19,670 | 3.6 | 34.8 | 17 | 95 | 1.48 (0.70 - 2.94) | 0.28 | 1.58 (0.77 - 3.25) | 0.22 |
| Meta-analysis | 162,028 | 413,810 | 106.6 | 331.2 | 1,313 | 4,756 | 1.04 (0.92 - 1.17) | 0.52 | 1.03 (0.80 - 1.32) | 0.83 |

##### 7.4.12 Joint pain

**Supplementary Table 281:** Exposures, outcomes and risk estimates for Joint pain in SGLT2I (T) vs DPP4I (C) new-user cohorts under two different propensity score (PS) adjustment across data sources. We report total patient exposure time in 1,000 patient-year (KYr) units, uncalibrated and calibrated hazard ratios (HRs) and their 95% confidence intervals (CIs). Greyed out entries did not pass study diagnostics and are included only for completeness.

| | Patients | | Exposure (KYr) | | Outcomes | | Uncalibrated | | Calibrated | |
|---|---|---|---|---|---|---|---|---|---|---|
| | T | C | T | C | T | C | HR (95% CI) | $p$ | HR (95% CI) | $p$ |
| **PS-stratification / on-treatment 2 TAR** | | | | | | | | | | |
| OptumDOD | 10,899 | 20,405 | 5.9 | 13.8 | 75 | 230 | 0.86 (0.64 - 1.13) | 0.28 | 0.82 (0.62 - 1.09) | 0.19 |
| MDCR | 3,368 | 9,564 | 2.2 | 7.4 | 23 | 93 | 0.81 (0.48 - 1.31) | 0.41 | 0.82 (0.50 - 1.36) | 0.45 |
| Open Claims | 172,024 | 320,780 | 106.5 | 245.7 | 545 | 1,736 | 0.84 (0.76 - 0.94) | <0.01 | 0.86 (0.69 - 1.07) | 0.20 |
| OptumEHR | 7,405 | 18,976 | 1.8 | 8.3 | 24 | 80 | 1.78 (1.06 - 2.91) | 0.03 | 1.55 (0.94 - 2.58) | 0.10 |
| VA | 6,617 | 12,920 | 3.6 | 11.0 | 19 | 49 | 0.98 (0.50 - 1.88) | 0.96 | 1.00 (0.51 - 1.93) | 0.93 |
| GermanyDA | 3,534 | 9,208 | 2.5 | 8.9 | 34 | 178 | 0.87 (0.57 - 1.30) | 0.51 | 0.79 (0.52 - 1.20) | 0.29 |
| SIDIAP | 2,847 | 16,060 | 3.1 | 28.3 | 56 | 600 | 0.91 (0.66 - 1.22) | 0.55 | 0.94 (0.69 - 1.29) | 0.71 |
| Meta-analysis | 206,694 | 407,913 | 125.7 | 323.4 | 776 | 2,966 | 0.87 (0.80 - 0.96) | <0.01 | 0.88 (0.71 - 1.08) | 0.21 |
| **PS-matching / on-treatment 2 TAR** | | | | | | | | | | |
| OptumDOD | 8,712 | 20,040 | 5.1 | 13.5 | 67 | 223 | 0.88 (0.60 - 1.26) | 0.49 | 0.83 (0.57 - 1.20) | 0.33 |
| MDCR | 2,822 | 9,539 | 2.0 | 7.4 | 19 | 93 | 0.68 (0.31 - 1.36) | 0.30 | 0.68 (0.32 - 1.43) | 0.32 |
| Open Claims | 130,810 | 319,866 | 87.8 | 245.1 | 457 | 1,727 | 0.79 (0.68 - 0.92) | <0.01 | 0.79 (0.61 - 1.02) | 0.08 |
| OptumEHR | 6,588 | 18,909 | 1.6 | 8.2 | 21 | 79 | 1.36 (0.65 - 2.71) | 0.40 | 1.25 (0.61 - 2.56) | 0.55 |
| VA | 4,078 | 12,531 | 2.6 | 10.5 | 10 | 48 | 1.36 (0.48 - 3.62) | 0.55 | 1.45 (0.53 - 3.99) | 0.48 |
| GermanyDA | 2,887 | 9,166 | 2.2 | 8.9 | 32 | 178 | 0.83 (0.50 - 1.34) | 0.47 | 0.79 (0.48 - 1.30) | 0.39 |
| SIDIAP | 2,361 | 15,541 | 2.8 | 27.1 | 53 | 564 | 0.89 (0.61 - 1.27) | 0.54 | 0.95 (0.66 - 1.37) | 0.73 |
| Meta-analysis | 158,258 | 405,592 | 104.0 | 320.7 | 659 | 2,912 | 0.83 (0.73 - 0.93) | <0.01 | 0.82 (0.64 - 1.05) | 0.12 |

##### 7.4.13 Lower extremity amputation

**Supplementary Table 282:** Exposures, outcomes and risk estimates for Lower extremity amputation in SGLT2I (T) vs DPP4I (C) new-user cohorts under two different propensity score (PS) adjustment across data sources. We report total patient exposure time in 1,000 patient-year (KYr) units, uncalibrated and calibrated hazard ratios (HRs) and their 95% confidence intervals (CIs). Greyed out entries did not pass study diagnostics and are included only for completeness.

| | Patients | | Exposure (KYr) | | Outcomes | | Uncalibrated | | Calibrated | |
|---|---|---|---|---|---|---|---|---|---|---|
| | T | C | T | C | T | C | HR (95% CI) | $p$ | HR (95% CI) | $p$ |
| **PS-stratification / on-treatment 2 TAR** | | | | | | | | | | |
| OptumDOD | 11,589 | 21,794 | 6.3 | 14.9 | <5 | 12 | 1.27 (0.33 - 4.00) | 0.71 | 1.22 (0.35 - 4.24) | 0.75 |
| MDCR | 3,500 | 10,004 | 2.3 | 7.7 | <5 | 6 | 4.12 (0.74 - 18.8) | 0.09 | 4.18 (0.83 - 21.1) | 0.08 |
| Open Claims | 184,471 | 341,853 | 113.8 | 262.6 | 57 | 139 | 1.26 (0.88 - 1.79) | 0.20 | 1.28 (0.86 - 1.93) | 0.23 |
| OptumEHR | 7,821 | 20,033 | 2.0 | 8.8 | 0 | 6 | | | | |
| VA | 7,511 | 14,572 | 4.1 | 12.3 | <10 | <10 | 1.83 (0.49 - 7.03) | 0.37 | 1.86 (0.49 - 7.08) | 0.36 |
| GermanyDA | 4,175 | 10,792 | 3.0 | 10.7 | 0 | 0 | | | | |
| SIDIAP | 3,745 | 20,500 | 4.1 | 36.9 | 0 | 8 | | | | |
| Meta-analysis | 191,982 | 356,425 | 117.9 | 274.8 | <67 | <149 | 1.29 (0.91 - 1.83) | 0.15 | 1.29 (0.87 - 1.92) | 0.20 |
| **PS-matching / on-treatment 2 TAR** | | | | | | | | | | |
| OptumDOD | 9,281 | 21,401 | 5.5 | 14.6 | <5 | 12 | 1.47 (0.36 - 5.26) | 0.57 | 1.39 (0.36 - 5.31) | 0.63 |
| MDCR | 2,921 | 9,978 | 2.1 | 7.7 | <5 | 6 | 2.25 (0.26 - 17.0) | 0.45 | 2.25 (0.28 - 18.2) | 0.45 |
| Open Claims | 140,306 | 340,833 | 93.8 | 261.8 | 45 | 135 | 1.55 (0.91 - 2.59) | 0.10 | 1.54 (0.88 - 2.70) | 0.13 |
| OptumEHR | 6,961 | 19,958 | 1.8 | 8.8 | 0 | 6 | | | | |
| VA | 4,587 | 14,155 | 2.9 | 11.7 | <10 | <10 | | 0.22 | | 0.21 |
| GermanyDA | 3,434 | 10,733 | 2.6 | 10.7 | 0 | 0 | | | | |
| SIDIAP | 3,114 | 19,867 | 3.6 | 35.2 | 0 | 7 | | | | |
| Meta-analysis | 140,306 | 340,833 | 93.8 | 261.8 | 45 | 135 | 1.55 (0.91 - 2.59) | 0.10 | 1.53 (0.87 - 2.69) | 0.14 |

##### 7.4.14 Nausea

**Supplementary Table 283:** Exposures, outcomes and risk estimates for Nausea in SGLT2I (T) vs DPP4I (C) new-user cohorts under two different propensity score (PS) adjustment across data sources. We report total patient exposure time in 1,000 patient-year (KYr) units, uncalibrated and calibrated hazard ratios (HRs) and their 95% confidence intervals (CIs). Greyed out entries did not pass study diagnostics and are included only for completeness.

| | Patients | | Exposure (KYr) | | Outcomes | | Uncalibrated | | Calibrated | |
|---|---|---|---|---|---|---|---|---|---|---|
| | T | C | T | C | T | C | HR (95% CI) | $p$ | HR (95% CI) | $p$ |
| **PS-stratification / on-treatment 2 TAR** | | | | | | | | | | |
| OptumDOD | 9,971 | 18,322 | 5.4 | 12.3 | 199 | 595 | 0.85 (0.71 - 1.01) | 0.07 | 0.82 (0.68 - 0.98) | 0.04 |
| MDCR | 3,120 | 8,773 | 2.0 | 6.7 | 62 | 253 | 1.02 (0.75 - 1.38) | 0.89 | 1.04 (0.76 - 1.41) | 0.82 |
| Open Claims | 158,462 | 295,534 | 98.7 | 226.4 | 1,483 | 4,903 | 0.86 (0.81 - 0.92) | <0.01 | 0.88 (0.72 - 1.08) | 0.23 |
| OptumEHR | 7,090 | 18,119 | 1.8 | 7.8 | 53 | 258 | 1.19 (0.85 - 1.63) | 0.30 | 1.04 (0.75 - 1.44) | 0.80 |
| VA | 6,967 | 13,696 | 3.8 | 11.6 | 23 | 84 | 0.69 (0.37 - 1.26) | 0.24 | 0.70 (0.38 - 1.30) | 0.27 |
| GermanyDA | 3,908 | 9,999 | 2.8 | 9.9 | 18 | 105 | 0.76 (0.43 - 1.27) | 0.31 | 0.69 (0.40 - 1.19) | 0.19 |
| SIDIAP | 3,589 | 19,649 | 4.0 | 35.2 | 16 | 302 | 0.52 (0.29 - 0.86) | 0.02 | 0.54 (0.31 - 0.93) | 0.03 |
| Meta-analysis | 193,107 | 384,092 | 118.5 | 309.9 | 1,854 | 6,500 | 0.87 (0.82 - 0.92) | <0.01 | 0.87 (0.71 - 1.06) | 0.16 |
| **PS-matching / on-treatment 2 TAR** | | | | | | | | | | |
| OptumDOD | 7,961 | 18,031 | 4.7 | 12.1 | 177 | 576 | 0.83 (0.66 - 1.05) | 0.13 | 0.79 (0.62 - 1.00) | 0.06 |
| MDCR | 2,606 | 8,753 | 1.9 | 6.7 | 55 | 252 | 0.94 (0.63 - 1.37) | 0.75 | 0.94 (0.63 - 1.39) | 0.77 |
| Open Claims | 120,491 | 294,522 | 81.3 | 225.7 | 1,249 | 4,869 | 0.82 (0.76 - 0.90) | <0.01 | 0.82 (0.65 - 1.03) | 0.11 |
| OptumEHR | 6,300 | 17,988 | 1.6 | 7.8 | 48 | 256 | 1.16 (0.76 - 1.75) | 0.48 | 1.07 (0.71 - 1.63) | 0.73 |
| VA | 4,301 | 13,319 | 2.7 | 11.1 | 10 | 81 | 0.59 (0.21 - 1.37) | 0.26 | 0.62 (0.25 - 1.58) | 0.32 |
| GermanyDA | 3,205 | 9,959 | 2.4 | 9.8 | 15 | 105 | 0.64 (0.33 - 1.18) | 0.18 | 0.61 (0.32 - 1.16) | 0.15 |
| SIDIAP | 2,988 | 19,075 | 3.5 | 33.8 | 11 | 288 | 0.43 (0.20 - 0.83) | 0.02 | 0.46 (0.23 - 0.94) | 0.04 |
| Meta-analysis | 147,852 | 381,647 | 98.0 | 306.9 | 1,565 | 6,427 | 0.83 (0.77 - 0.89) | <0.01 | 0.82 (0.65 - 1.03) | 0.09 |

##### 7.4.15 Peripheral edema

**Supplementary Table 284:** Exposures, outcomes and risk estimates for Peripheral edema in SGLT2I (T) vs DPP4I (C) new-user cohorts under two different propensity score (PS) adjustment across data sources. We report total patient exposure time in 1,000 patient-year (KYr) units, uncalibrated and calibrated hazard ratios (HRs) and their 95% confidence intervals (CIs). Greyed out entries did not pass study diagnostics and are included only for completeness.

| | Patients | | Exposure (KYr) | | Outcomes | | Uncalibrated | | Calibrated | |
|---|---|---|---|---|---|---|---|---|---|---|
| | T | C | T | C | T | C | HR (95% CI) | $p$ | HR (95% CI) | $p$ |
| **PS-stratification / on-treatment 2 TAR** | | | | | | | | | | |
| OptumDOD | 8,491 | 15,272 | 4.6 | 9.9 | 298 | 1,017 | 0.74 (0.65 - 0.86) | <0.01 | 0.72 (0.62 - 0.83) | <0.01 |
| MDCR | 2,647 | 7,141 | 1.7 | 5.3 | 74 | 477 | 0.63 (0.47 - 0.81) | <0.01 | 0.63 (0.48 - 0.83) | <0.01 |
| Open Claims | 132,741 | 248,533 | 83.0 | 186.3 | 2,095 | 8,752 | 0.71 (0.67 - 0.74) | <0.01 | 0.72 (0.59 - 0.88) | <0.01 |
| OptumEHR | 6,272 | 16,106 | 1.5 | 6.8 | 42 | 391 | 0.66 (0.46 - 0.91) | 0.02 | 0.57 (0.41 - 0.81) | <0.01 |
| VA | 5,373 | 11,017 | 3.0 | 9.2 | 94 | 333 | 0.83 (0.62 - 1.10) | 0.21 | 0.84 (0.63 - 1.13) | 0.27 |
| GermanyDA | 3,389 | 8,757 | 2.4 | 8.5 | 51 | 329 | 0.58 (0.42 - 0.80) | <0.01 | 0.53 (0.38 - 0.74) | <0.01 |
| SIDIAP | 3,190 | 17,958 | 3.6 | 31.8 | 40 | 764 | 0.52 (0.37 - 0.73) | <0.01 | 0.54 (0.38 - 0.77) | <0.01 |
| Meta-analysis | 162,103 | 324,784 | 99.8 | 257.8 | 2,694 | 12,063 | 0.70 (0.67 - 0.74) | <0.01 | 0.70 (0.58 - 0.85) | <0.01 |
| **PS-matching / on-treatment 2 TAR** | | | | | | | | | | |
| OptumDOD | 6,834 | 15,067 | 4.0 | 9.8 | 261 | 986 | 0.69 (0.56 - 0.83) | <0.01 | 0.65 (0.53 - 0.79) | <0.01 |
| MDCR | 2,188 | 7,101 | 1.5 | 5.3 | 65 | 472 | 0.66 (0.46 - 0.91) | 0.02 | 0.66 (0.47 - 0.93) | 0.03 |
| Open Claims | 102,215 | 247,958 | 69.0 | 186.0 | 1,779 | 8,691 | 0.72 (0.67 - 0.77) | <0.01 | 0.71 (0.57 - 0.89) | <0.01 |
| OptumEHR | 5,642 | 15,998 | 1.4 | 6.8 | 41 | 386 | 0.72 (0.47 - 1.07) | 0.11 | 0.66 (0.44 - 1.00) | 0.06 |
| VA | 3,400 | 10,752 | 2.2 | 8.9 | 62 | 316 | 0.69 (0.47 - 1.00) | 0.06 | 0.73 (0.50 - 1.08) | 0.13 |
| GermanyDA | 2,793 | 8,715 | 2.1 | 8.4 | 43 | 327 | 0.60 (0.39 - 0.90) | 0.02 | 0.57 (0.38 - 0.87) | 0.01 |
| SIDIAP | 2,656 | 17,306 | 3.2 | 30.2 | 35 | 713 | 0.57 (0.37 - 0.84) | <0.01 | 0.61 (0.41 - 0.92) | 0.02 |
| Meta-analysis | 125,728 | 322,897 | 83.4 | 255.3 | 2,286 | 11,891 | 0.70 (0.66 - 0.75) | <0.01 | 0.70 (0.55 - 0.87) | <0.01 |

##### 7.4.16 Photosensitivity

**Supplementary Table 285:** Exposures, outcomes and risk estimates for Photosensitivity in SGLT2I (T) vs DPP4I (C) new-user cohorts under two different propensity score (PS) adjustment across data sources. We report total patient exposure time in 1,000 patient-year (KYr) units, uncalibrated and calibrated hazard ratios (HRs) and their 95% confidence intervals (CIs). Greyed out entries did not pass study diagnostics and are included only for completeness.

| | Patients | | Exposure (KYr) | | Outcomes | | Uncalibrated | | Calibrated | |
|---|---|---|---|---|---|---|---|---|---|---|
| | T | C | T | C | T | C | HR (95% CI) | $p$ | HR (95% CI) | $p$ |
| **PS-stratification / on-treatment 2 TAR** | | | | | | | | | | |
| OptumDOD | 11,516 | 21,588 | 6.3 | 14.8 | <5 | 8 | 2.17 (0.38 - 9.22) | 0.34 | 2.08 (0.42 - 10.3) | 0.37 |
| MDCR | 3,464 | 9,818 | 2.3 | 7.6 | <5 | 20 | 0.98 (0.22 - 3.08) | 0.97 | 0.99 (0.26 - 3.72) | 0.97 |
| Open Claims | 181,939 | 337,293 | 112.1 | 258.8 | 26 | 163 | 0.93 (0.58 - 1.42) | 0.74 | 0.94 (0.58 - 1.53) | 0.82 |
| OptumEHR | 7,758 | 19,847 | 1.9 | 8.7 | 0 | 20 | | | | |
| VA | 7,386 | 14,331 | 4.0 | 12.1 | 0 | <10 | | | | |
| GermanyDA | 4,167 | 10,750 | 3.0 | 10.7 | 0 | 0 | | | | |
| SIDIAP | 3,737 | 20,476 | 4.1 | 36.8 | 0 | 5 | | | | |
| Meta-analysis | 185,403 | 347,111 | 114.4 | 266.3 | <31 | 183 | 0.93 (0.61 - 1.42) | 0.74 | 0.93 (0.59 - 1.48) | 0.77 |
| **PS-matching / on-treatment 2 TAR** | | | | | | | | | | |
| OptumDOD | 9,215 | 21,198 | 5.4 | 14.5 | <5 | 8 | 1.49 (0.18 - 8.97) | 0.69 | 1.41 (0.20 - 9.85) | 0.73 |
| MDCR | 2,887 | 9,793 | 2.1 | 7.6 | <5 | 20 | 0.53 (0.03 - 3.18) | 0.60 | 0.53 (0.05 - 5.71) | 0.61 |
| Open Claims | 138,358 | 336,236 | 92.4 | 258.0 | 25 | 162 | 0.99 (0.56 - 1.66) | 0.96 | 0.98 (0.55 - 1.76) | 0.95 |
| OptumEHR | 6,908 | 19,770 | 1.7 | 8.7 | 0 | 20 | | | | |
| VA | 4,518 | 13,831 | 2.8 | 11.4 | 0 | <10 | | | | |
| GermanyDA | 3,424 | 10,691 | 2.6 | 10.6 | 0 | 0 | | | | |
| SIDIAP | 3,110 | 19,842 | 3.6 | 35.2 | 0 | <5 | | | | |
| Meta-analysis | 141,245 | 346,029 | 94.4 | 265.6 | <30 | 182 | 0.96 (0.56 - 1.63) | 0.87 | 0.95 (0.53 - 1.68) | 0.85 |

##### 7.4.17 Venous thromboembolism

**Supplementary Table 286:** Exposures, outcomes and risk estimates for Venous thromboembolism in SGLT2I (T) vs DPP4I (C) new-user cohorts under two different propensity score (PS) adjustment across data sources. We report total patient exposure time in 1,000 patient-year (KYr) units, uncalibrated and calibrated hazard ratios (HRs) and their 95% confidence intervals (CIs). Greyed out entries did not pass study diagnostics and are included only for completeness.

| | Patients | | Exposure (KYr) | | Outcomes | | Uncalibrated | | Calibrated | |
|---|---|---|---|---|---|---|---|---|---|---|
| | T | C | T | C | T | C | HR (95% CI) | $p$ | HR (95% CI) | $p$ |
| **PS-stratification / on-treatment 2 TAR** | | | | | | | | | | |
| OptumDOD | 11,177 | 20,859 | 6.1 | 14.2 | 57 | 212 | 0.71 (0.51 - 0.98) | 0.04 | 0.68 (0.49 - 0.95) | 0.03 |
| MDCR | 3,367 | 9,538 | 2.2 | 7.4 | 18 | 86 | 0.91 (0.50 - 1.56) | 0.74 | 0.92 (0.52 - 1.63) | 0.78 |
| Open Claims | 176,274 | 326,954 | 109.0 | 250.8 | 567 | 1,843 | 0.90 (0.81 - 1.00) | 0.06 | 0.92 (0.74 - 1.14) | 0.47 |
| OptumEHR | 7,536 | 19,333 | 1.9 | 8.5 | 14 | 80 | 1.09 (0.58 - 1.92) | 0.78 | 0.95 (0.52 - 1.74) | 0.86 |
| VA | 7,154 | 14,015 | 3.9 | 11.8 | 16 | 60 | 0.46 (0.23 - 0.88) | 0.02 | 0.46 (0.23 - 0.91) | 0.03 |
| GermanyDA | 3,859 | 9,869 | 2.7 | 9.7 | 25 | 102 | 1.14 (0.68 - 1.84) | 0.61 | 1.04 (0.63 - 1.70) | 0.79 |
| SIDIAP | 3,597 | 19,712 | 3.9 | 35.4 | 18 | 182 | 0.95 (0.54 - 1.60) | 0.86 | 0.99 (0.57 - 1.71) | 0.87 |
| Meta-analysis | 212,964 | 420,280 | 129.6 | 337.8 | 715 | 2,565 | 0.89 (0.81 - 0.98) | 0.01 | 0.89 (0.72 - 1.10) | 0.27 |
| **PS-matching / on-treatment 2 TAR** | | | | | | | | | | |
| OptumDOD | 8,952 | 20,503 | 5.3 | 14.0 | 49 | 204 | 0.77 (0.51 - 1.14) | 0.21 | 0.73 (0.48 - 1.09) | 0.13 |
| MDCR | 2,809 | 9,513 | 2.0 | 7.4 | 17 | 86 | 1.37 (0.65 - 2.73) | 0.39 | 1.37 (0.67 - 2.82) | 0.41 |
| Open Claims | 134,282 | 325,993 | 89.9 | 250.2 | 472 | 1,823 | 0.88 (0.76 - 1.01) | 0.07 | 0.87 (0.68 - 1.13) | 0.32 |
| OptumEHR | 6,729 | 19,270 | 1.7 | 8.4 | 14 | 79 | 1.29 (0.57 - 2.68) | 0.52 | 1.19 (0.55 - 2.57) | 0.67 |
| VA | 4,363 | 13,526 | 2.7 | 11.2 | <10 | 57 | 0.20 (0.05 - 0.58) | 0.01 | 0.21 (0.06 - 0.74) | 0.02 |
| GermanyDA | 3,149 | 9,826 | 2.4 | 9.7 | 21 | 102 | 1.59 (0.83 - 2.93) | 0.15 | 1.51 (0.80 - 2.84) | 0.22 |
| SIDIAP | 2,991 | 19,131 | 3.4 | 33.9 | 17 | 168 | 1.04 (0.56 - 1.82) | 0.89 | 1.11 (0.62 - 2.00) | 0.73 |
| Meta-analysis | 163,275 | 417,762 | 107.4 | 334.6 | <600 | 2,519 | 0.90 (0.79 - 1.02) | 0.10 | 0.89 (0.69 - 1.15) | 0.37 |

##### 7.4.18 Vomiting

**Supplementary Table 287:** Exposures, outcomes and risk estimates for Vomiting in SGLT2I (T) vs DPP4I (C) new-user cohorts under two different propensity score (PS) adjustment across data sources. We report total patient exposure time in 1,000 patient-year (KYr) units, uncalibrated and calibrated hazard ratios (HRs) and their 95% confidence intervals (CIs). Greyed out entries did not pass study diagnostics and are included only for completeness.

| | Patients | | Exposure (KYr) | | Outcomes | | Uncalibrated | | Calibrated | |
|---|---|---|---|---|---|---|---|---|---|---|
| | T | C | T | C | T | C | HR (95% CI) | $p$ | HR (95% CI) | $p$ |
| **PS-stratification / on-treatment 2 TAR** | | | | | | | | | | |
| OptumDOD | 10,476 | 19,270 | 5.7 | 13.0 | 144 | 455 | 0.90 (0.73 - 1.10) | 0.30 | 0.86 (0.70 - 1.06) | 0.18 |
| MDCR | 3,271 | 9,092 | 2.1 | 7.0 | 45 | 199 | 0.93 (0.64 - 1.31) | 0.67 | 0.94 (0.65 - 1.35) | 0.74 |
| Open Claims | 165,476 | 306,254 | 102.8 | 234.8 | 1,119 | 3,950 | 0.84 (0.78 - 0.91) | <0.01 | 0.86 (0.70 - 1.05) | 0.16 |
| OptumEHR | 7,398 | 18,868 | 1.8 | 8.2 | 35 | 173 | 1.31 (0.87 - 1.93) | 0.17 | 1.14 (0.77 - 1.70) | 0.54 |
| VA | 7,106 | 13,967 | 3.9 | 11.8 | 19 | 73 | 0.76 (0.39 - 1.40) | 0.39 | 0.77 (0.40 - 1.45) | 0.42 |
| GermanyDA | 3,907 | 9,993 | 2.8 | 9.9 | 18 | 106 | 0.75 (0.43 - 1.27) | 0.31 | 0.69 (0.40 - 1.19) | 0.19 |
| SIDIAP | 3,583 | 19,596 | 3.9 | 35.1 | 18 | 315 | 0.56 (0.33 - 0.92) | 0.03 | 0.59 (0.35 - 0.98) | 0.05 |
| Meta-analysis | 201,217 | 397,040 | 123.1 | 319.8 | 1,398 | 5,271 | 0.85 (0.80 - 0.91) | <0.01 | 0.85 (0.70 - 1.04) | 0.12 |
| **PS-matching / on-treatment 2 TAR** | | | | | | | | | | |
| OptumDOD | 8,391 | 18,971 | 5.0 | 12.8 | 131 | 442 | 0.92 (0.69 - 1.20) | 0.53 | 0.86 (0.65 - 1.14) | 0.32 |
| MDCR | 2,712 | 9,071 | 1.9 | 6.9 | 40 | 199 | 1.00 (0.64 - 1.53) | 1.00 | 1.00 (0.65 - 1.55) | 0.88 |
| Open Claims | 125,626 | 305,455 | 84.5 | 234.2 | 944 | 3,930 | 0.83 (0.75 - 0.92) | <0.01 | 0.83 (0.66 - 1.05) | 0.13 |
| OptumEHR | 6,581 | 18,727 | 1.7 | 8.2 | 34 | 172 | 1.54 (0.93 - 2.49) | 0.09 | 1.42 (0.87 - 2.32) | 0.18 |
| VA | 4,384 | 13,577 | 2.8 | 11.3 | 11 | 69 | 0.56 (0.20 - 1.31) | 0.22 | 0.59 (0.23 - 1.51) | 0.28 |
| GermanyDA | 3,205 | 9,953 | 2.4 | 9.8 | 15 | 106 | 0.65 (0.33 - 1.19) | 0.18 | 0.61 (0.32 - 1.17) | 0.15 |
| SIDIAP | 2,983 | 19,027 | 3.5 | 33.7 | 13 | 298 | 0.51 (0.25 - 0.92) | 0.04 | 0.54 (0.28 - 1.04) | 0.07 |
| Meta-analysis | 153,882 | 394,781 | 101.8 | 316.9 | 1,188 | 5,216 | 0.86 (0.76 - 0.97) | 0.02 | 0.85 (0.66 - 1.09) | 0.20 |

#### 7.5 SGLT2I vs SU

##### 7.5.1 Abnormal weight gain

**Supplementary Table 288:** Exposures, outcomes and risk estimates for Abnormal weight gain in SGLT2I (T) vs SU (C) new-user cohorts under two different propensity score (PS) adjustment across data sources. We report total patient exposure time in 1,000 patient-year (KYr) units, uncalibrated and calibrated hazard ratios (HRs) and their 95% confidence intervals (CIs). Greyed out entries did not pass study diagnostics and are included only for completeness.

| | Patients | | Exposure (KYr) | | Outcomes | | Uncalibrated | | Calibrated | |
|---|---|---|---|---|---|---|---|---|---|---|
| | T | C | T | C | T | C | HR (95% CI) | $p$ | HR (95% CI) | $p$ |
| **PS-stratification / on-treatment 2 TAR** | | | | | | | | | | |
| OptumDOD | 11,275 | 48,470 | 6.1 | 43.8 | 18 | 278 | 0.44 (0.26 - 0.72) | <0.01 | 0.43 (0.26 - 0.72) | <0.01 |
| MDCR | 3,444 | 15,316 | 2.2 | 12.3 | <5 | 52 | 0.36 (0.10 - 0.99) | 0.08 | 0.34 (0.11 - 1.06) | 0.07 |
| Open Claims | 177,824 | 682,286 | 109.8 | 689.3 | 118 | 1,412 | 0.54 (0.44 - 0.65) | <0.01 | 0.55 (0.41 - 0.72) | <0.01 |
| OptumEHR | 7,560 | 45,158 | 1.9 | 24.2 | 5 | 143 | 0.49 (0.17 - 1.14) | 0.14 | 0.47 (0.18 - 1.21) | 0.12 |
| VA | 7,350 | 52,957 | 4.0 | 47.3 | <10 | 61 | 1.11 (0.36 - 2.89) | 0.85 | 1.21 (0.43 - 3.40) | 0.73 |
| GermanyDA | 4,116 | 1,537 | 3.0 | 1.5 | 0 | <5 | | | | |
| SIDIAP | 3,707 | 2,113 | 4.0 | 4.1 | <5 | 9 | 0.46 (0.08 - 2.25) | 0.37 | 0.52 (0.10 - 2.78) | 0.45 |
| Meta-analysis | 196,659 | 775,914 | 117.8 | 757.4 | 141 | 1,833 | 0.52 (0.43 - 0.63) | <0.01 | 0.52 (0.40 - 0.68) | <0.01 |
| **PS-matching / on-treatment 2 TAR** | | | | | | | | | | |
| OptumDOD | 10,483 | 47,455 | 5.8 | 43.0 | 18 | 264 | 0.40 (0.20 - 0.74) | <0.01 | 0.39 (0.20 - 0.76) | <0.01 |
| MDCR | 2,983 | 14,556 | 2.0 | 11.7 | <5 | 47 | 0.29 (0.04 - 1.10) | 0.13 | 0.28 (0.06 - 1.38) | 0.12 |
| Open Claims | 163,428 | 665,817 | 103.1 | 673.6 | 106 | 1,349 | 0.54 (0.42 - 0.69) | <0.01 | 0.55 (0.41 - 0.75) | <0.01 |
| OptumEHR | 7,048 | 42,481 | 1.7 | 22.7 | <5 | 128 | 0.28 (0.04 - 0.98) | 0.11 | 0.28 (0.06 - 1.31) | 0.11 |
| VA | 5,185 | 33,189 | 3.2 | 26.2 | <10 | 31 | 1.03 (0.23 - 3.39) | 0.96 | 1.16 (0.30 - 4.47) | 0.83 |
| GermanyDA | 4,057 | 883 | 3.0 | 0.8 | 0 | <5 | | | | |
| SIDIAP | 3,280 | 958 | 3.7 | 1.5 | <5 | <5 | 0.53 (0.10 - 3.07) | 0.47 | 0.73 (0.13 - 4.12) | 0.72 |
| Meta-analysis | 183,942 | 770,309 | 112.7 | 751.1 | <134 | 1,788 | 0.51 (0.40 - 0.64) | <0.01 | 0.51 (0.38 - 0.68) | <0.01 |

##### 7.5.2 Abnormal weight loss

**Supplementary Table 289:** Exposures, outcomes and risk estimates for Abnormal weight loss in SGLT2I (T) vs SU (C) new-user cohorts under two different propensity score (PS) adjustment across data sources. We report total patient exposure time in 1,000 patient-year (KYr) units, uncalibrated and calibrated hazard ratios (HRs) and their 95% confidence intervals (CIs). Greyed out entries did not pass study diagnostics and are included only for completeness.

| | Patients | | Exposure (KYr) | | Outcomes | | Uncalibrated | | Calibrated | |
|---|---|---|---|---|---|---|---|---|---|---|
| | T | C | T | C | T | C | HR (95% CI) | $p$ | HR (95% CI) | $p$ |
| **PS-stratification / on-treatment 2 TAR** | | | | | | | | | | |
| OptumDOD | 11,094 | 46,524 | 6.0 | 42.2 | 149 | 713 | 1.59 (1.30 - 1.94) | <0.01 | 1.55 (1.26 - 1.89) | <0.01 |
| MDCR | 3,392 | 14,708 | 2.2 | 11.9 | 40 | 171 | 1.79 (1.18 - 2.67) | <0.01 | 1.69 (1.12 - 2.55) | 0.01 |
| Open Claims | 176,400 | 665,366 | 108.8 | 672.9 | 970 | 4,790 | 1.45 (1.34 - 1.57) | <0.01 | 1.48 (1.20 - 1.82) | <0.01 |
| OptumEHR | 7,530 | 44,122 | 1.9 | 23.6 | 27 | 327 | 1.31 (0.83 - 1.99) | 0.22 | 1.25 (0.81 - 1.93) | 0.33 |
| VA | 7,184 | 51,321 | 3.9 | 45.9 | 62 | 366 | 1.77 (1.22 - 2.55) | <0.01 | 1.93 (1.33 - 2.80) | <0.01 |
| GermanyDA | 4,032 | 1,500 | 2.9 | 1.5 | 33 | 8 | 1.99 (0.64 - 7.02) | 0.26 | 1.93 (0.58 - 6.44) | 0.29 |
| SIDIAP | 3,673 | 2,113 | 4.0 | 4.0 | 32 | 19 | 1.43 (0.61 - 3.32) | 0.41 | 1.61 (0.69 - 3.75) | 0.30 |
| Meta-analysis | 195,024 | 756,012 | 116.6 | 738.6 | 1,146 | 5,830 | 1.47 (1.36 - 1.57) | <0.01 | 1.46 (1.20 - 1.79) | <0.01 |
| **PS-matching / on-treatment 2 TAR** | | | | | | | | | | |
| OptumDOD | 10,284 | 45,740 | 5.7 | 41.5 | 138 | 688 | 1.56 (1.22 - 1.99) | <0.01 | 1.53 (1.20 - 1.95) | <0.01 |
| MDCR | 2,932 | 13,950 | 2.0 | 11.3 | 38 | 163 | 1.72 (1.05 - 2.75) | 0.03 | 1.63 (1.01 - 2.65) | 0.05 |
| Open Claims | 162,116 | 649,510 | 102.1 | 657.8 | 912 | 4,585 | 1.45 (1.31 - 1.59) | <0.01 | 1.48 (1.21 - 1.83) | <0.01 |
| OptumEHR | 7,005 | 41,529 | 1.7 | 22.2 | 26 | 287 | 1.05 (0.59 - 1.77) | 0.87 | 1.03 (0.59 - 1.79) | 0.87 |
| VA | 5,071 | 32,451 | 3.1 | 25.7 | 46 | 193 | 1.47 (0.90 - 2.35) | 0.12 | 1.65 (1.02 - 2.68) | 0.04 |
| GermanyDA | 3,974 | 869 | 2.9 | 0.8 | 33 | <5 | 3.82 (0.68 - 72.3) | 0.26 | 4.59 (0.44 - 47.5) | 0.21 |
| SIDIAP | 3,257 | 957 | 3.7 | 1.5 | 26 | 6 | 1.38 (0.44 - 5.21) | 0.61 | 1.89 (0.55 - 6.52) | 0.33 |
| Meta-analysis | 182,337 | 750,729 | 111.5 | 732.7 | 1,114 | 5,723 | 1.46 (1.34 - 1.59) | <0.01 | 1.46 (1.19 - 1.78) | <0.01 |

##### 7.5.3 Acute pancreatitis

**Supplementary Table 290:** Exposures, outcomes and risk estimates for Acute pancreatitis in SGLT2I (T) vs SU (C) new-user cohorts under two different propensity score (PS) adjustment across data sources. We report total patient exposure time in 1,000 patient-year (KYr) units, uncalibrated and calibrated hazard ratios (HRs) and their 95% confidence intervals (CIs). Greyed out entries did not pass study diagnostics and are included only for completeness.

| | Patients | | Exposure (KYr) | | Outcomes | | Uncalibrated | | Calibrated | |
|---|---|---|---|---|---|---|---|---|---|---|
| | T | C | T | C | T | C | HR (95% CI) | $p$ | HR (95% CI) | $p$ |
| **PS-stratification / on-treatment 2 TAR** | | | | | | | | | | |
| OptumDOD | 11,512 | 49,009 | 6.3 | 44.5 | 18 | 156 | 0.98 (0.56 - 1.61) | 0.93 | 0.95 (0.56 - 1.61) | 0.85 |
| MDCR | 3,480 | 15,305 | 2.3 | 12.4 | <5 | 44 | 0.60 (0.17 - 1.60) | 0.37 | 0.56 (0.18 - 1.73) | 0.32 |
| Open Claims | 182,041 | 689,073 | 112.4 | 695.9 | 146 | 1,486 | 0.72 (0.60 - 0.86) | <0.01 | 0.74 (0.56 - 0.96) | 0.03 |
| OptumEHR | 7,801 | 45,830 | 1.9 | 24.7 | <5 | 56 | 0.91 (0.21 - 2.68) | 0.89 | 0.87 (0.24 - 3.08) | 0.83 |
| VA | 7,438 | 53,201 | 4.1 | 47.5 | <10 | 79 | 0.29 (0.02 - 1.51) | 0.29 | 0.32 (0.03 - 3.09) | 0.32 |
| GermanyDA | 4,154 | 1,547 | 3.0 | 1.5 | 0 | 0 | | | | |
| SIDIAP | 3,736 | 2,128 | 4.1 | 4.1 | 0 | 0 | | | | |
| Meta-analysis | 201,354 | 783,912 | 120.6 | 765.1 | <169 | 1,698 | 0.75 (0.62 - 0.91) | <0.01 | 0.75 (0.58 - 0.98) | 0.04 |
| **PS-matching / on-treatment 2 TAR** | | | | | | | | | | |
| OptumDOD | 10,688 | 47,974 | 6.0 | 43.6 | 17 | 153 | 1.12 (0.61 - 1.96) | 0.71 | 1.09 (0.61 - 1.97) | 0.77 |
| MDCR | 3,011 | 14,560 | 2.1 | 11.8 | <5 | 43 | 0.57 (0.13 - 1.85) | 0.42 | 0.55 (0.14 - 2.08) | 0.38 |
| Open Claims | 167,109 | 672,969 | 105.4 | 680.6 | 142 | 1,429 | 0.76 (0.61 - 0.94) | 0.01 | 0.78 (0.59 - 1.04) | 0.10 |
| OptumEHR | 7,259 | 43,132 | 1.8 | 23.2 | <5 | 53 | 1.41 (0.20 - 6.14) | 0.69 | 1.39 (0.25 - 7.64) | 0.71 |
| VA | 5,219 | 33,500 | 3.2 | 26.5 | <10 | 38 | 0.36 (0.02 - 2.00) | 0.39 | 0.40 (0.04 - 4.12) | 0.44 |
| GermanyDA | 4,095 | 892 | 3.0 | 0.8 | 0 | 0 | | | | |
| SIDIAP | 3,307 | 967 | 3.8 | 1.5 | 0 | 0 | | | | |
| Meta-analysis | 188,067 | 778,635 | 115.3 | 759.1 | <169 | 1,678 | 0.81 (0.63 - 1.04) | 0.10 | 0.81 (0.60 - 1.11) | 0.19 |

##### 7.5.4 All-cause mortality

**Supplementary Table 291:** Exposures, outcomes and risk estimates for All-cause mortality in SGLT2I (T) vs SU (C) new-user cohorts under two different propensity score (PS) adjustment across data sources. We report total patient exposure time in 1,000 patient-year (KYr) units, uncalibrated and calibrated hazard ratios (HRs) and their 95% confidence intervals (CIs). Greyed out entries did not pass study diagnostics and are included only for completeness.

| | Patients | | Exposure (KYr) | | Outcomes | | Uncalibrated | | Calibrated | |
|---|---|---|---|---|---|---|---|---|---|---|
| | T | C | T | C | T | C | HR (95% CI) | $p$ | HR (95% CI) | $p$ |
| **PS-stratification / on-treatment 2 TAR** | | | | | | | | | | |
| OptumDOD | 11,614 | 49,630 | 6.3 | 45.1 | 40 | 696 | 0.54 (0.38 - 0.75) | <0.01 | 0.53 (0.38 - 0.74) | <0.01 |
| MDCR | 3,510 | 15,524 | 2.3 | 12.5 | <5 | 29 | 1.86 (0.29 - 6.76) | 0.44 | 1.76 (0.36 - 8.52) | 0.49 |
| Open Claims | 184,807 | 699,596 | 114.0 | 706.7 | 0 | 11 | | | | |
| OptumEHR | 7,828 | 46,129 | 2.0 | 24.8 | 7 | 372 | 0.47 (0.20 - 0.94) | 0.06 | 0.45 (0.20 - 0.98) | 0.05 |
| VA | 7,529 | 53,862 | 4.1 | 48.1 | 51 | 864 | 0.55 (0.39 - 0.77) | <0.01 | 0.60 (0.43 - 0.84) | <0.01 |
| GermanyDA | 4,154 | 1,547 | 3.0 | 1.5 | <5 | 0 | | | | |
| SIDIAP | 3,746 | 2,136 | 4.1 | 4.1 | 86 | 84 | 0.69 (0.43 - 1.10) | 0.12 | 0.78 (0.49 - 1.24) | 0.33 |
| Meta-analysis | 19,442 | 95,759 | 8.3 | 69.9 | 47 | 1,068 | 0.53 (0.39 - 0.73) | <0.01 | 0.53 (0.37 - 0.76) | <0.01 |
| **PS-matching / on-treatment 2 TAR** | | | | | | | | | | |
| OptumDOD | 10,783 | 48,541 | 6.0 | 44.1 | 40 | 652 | 0.62 (0.41 - 0.91) | 0.02 | 0.60 (0.40 - 0.90) | 0.02 |
| MDCR | 3,040 | 14,747 | 2.1 | 11.9 | <5 | 23 | 0.98 (0.14 - 4.22) | 0.98 | 0.93 (0.17 - 5.12) | 0.94 |
| Open Claims | 169,729 | 683,066 | 107.0 | 691.0 | 0 | 11 | | | | |
| OptumEHR | 7,282 | 43,385 | 1.8 | 23.3 | 6 | 330 | 0.50 (0.19 - 1.10) | 0.12 | 0.49 (0.20 - 1.19) | 0.12 |
| VA | 5,286 | 33,867 | 3.3 | 26.8 | 41 | 485 | 0.47 (0.29 - 0.72) | <0.01 | 0.53 (0.34 - 0.83) | <0.01 |
| GermanyDA | 4,095 | 892 | 3.0 | 0.8 | <5 | 0 | | | | |
| SIDIAP | 3,314 | 971 | 3.8 | 1.5 | 66 | 31 | 0.53 (0.27 - 1.03) | 0.06 | 0.74 (0.38 - 1.43) | 0.39 |
| Meta-analysis | 18,065 | 91,926 | 7.8 | 67.4 | 46 | 982 | 0.60 (0.41 - 0.86) | <0.01 | 0.60 (0.40 - 0.90) | 0.01 |

##### 7.5.5 Bone fracture

**Supplementary Table 292:** Exposures, outcomes and risk estimates for Bone fracture in SGLT2I (T) vs SU (C) new-user cohorts under two different propensity score (PS) adjustment across data sources. We report total patient exposure time in 1,000 patient-year (KYr) units, uncalibrated and calibrated hazard ratios (HRs) and their 95% confidence intervals (CIs). Greyed out entries did not pass study diagnostics and are included only for completeness.

| | Patients | | Exposure (KYr) | | Outcomes | | Uncalibrated | | Calibrated | |
|---|---|---|---|---|---|---|---|---|---|---|
| | T | C | T | C | T | C | HR (95% CI) | $p$ | HR (95% CI) | $p$ |
| **PS-stratification / on-treatment 2 TAR** | | | | | | | | | | |
| OptumDOD | 10,423 | 44,290 | 5.6 | 39.5 | 172 | 1,351 | 0.96 (0.81 - 1.14) | 0.69 | 0.94 (0.79 - 1.12) | 0.50 |
| MDCR | 3,143 | 13,509 | 2.0 | 10.6 | 62 | 391 | 0.93 (0.68 - 1.25) | 0.62 | 0.88 (0.64 - 1.19) | 0.44 |
| Open Claims | 155,987 | 598,249 | 96.2 | 599.8 | 1,571 | 13,495 | 0.86 (0.81 - 0.91) | <0.01 | 0.87 (0.71 - 1.07) | 0.21 |
| OptumEHR | 7,175 | 42,034 | 1.8 | 22.1 | 33 | 600 | 1.00 (0.67 - 1.43) | 0.99 | 0.95 (0.65 - 1.38) | 0.78 |
| VA | 6,699 | 48,394 | 3.7 | 43.2 | 35 | 427 | 1.23 (0.78 - 1.90) | 0.36 | 1.34 (0.86 - 2.09) | 0.21 |
| GermanyDA | 3,608 | 1,340 | 2.6 | 1.3 | 46 | 38 | 1.34 (0.73 - 2.44) | 0.35 | 1.30 (0.71 - 2.39) | 0.41 |
| SIDIAP | 3,268 | 1,874 | 3.6 | 3.6 | 50 | 54 | 0.78 (0.44 - 1.36) | 0.38 | 0.87 (0.49 - 1.54) | 0.65 |
| Meta-analysis | 173,585 | 684,573 | 103.6 | 661.4 | 1,776 | 15,446 | 0.89 (0.81 - 0.98) | 0.02 | 0.89 (0.72 - 1.09) | 0.26 |
| **PS-matching / on-treatment 2 TAR** | | | | | | | | | | |
| OptumDOD | 9,696 | 43,445 | 5.3 | 38.7 | 166 | 1,305 | 0.98 (0.79 - 1.21) | 0.86 | 0.96 (0.77 - 1.19) | 0.72 |
| MDCR | 2,745 | 12,892 | 1.8 | 10.2 | 61 | 367 | 1.10 (0.74 - 1.59) | 0.63 | 1.05 (0.71 - 1.53) | 0.81 |
| Open Claims | 143,566 | 583,748 | 90.5 | 585.9 | 1,477 | 12,987 | 0.85 (0.79 - 0.92) | <0.01 | 0.88 (0.72 - 1.07) | 0.21 |
| OptumEHR | 6,659 | 39,434 | 1.6 | 20.7 | 32 | 529 | 0.98 (0.62 - 1.49) | 0.92 | 0.96 (0.62 - 1.50) | 0.82 |
| VA | 4,736 | 30,474 | 2.9 | 24.1 | 24 | 224 | 1.28 (0.70 - 2.19) | 0.40 | 1.43 (0.81 - 2.54) | 0.22 |
| GermanyDA | 3,561 | 791 | 2.6 | 0.7 | 46 | 20 | 1.00 (0.45 - 2.34) | 1.00 | 1.20 (0.53 - 2.75) | 0.62 |
| SIDIAP | 2,892 | 861 | 3.3 | 1.3 | 44 | 12 | 1.23 (0.52 - 3.26) | 0.66 | 1.69 (0.67 - 4.26) | 0.29 |
| Meta-analysis | 162,666 | 679,519 | 99.3 | 655.6 | 1,736 | 15,188 | 0.91 (0.81 - 1.03) | 0.13 | 0.91 (0.73 - 1.14) | 0.41 |

##### 7.5.6 Diabetic ketoacidosis

**Supplementary Table 293:** Exposures, outcomes and risk estimates for Diabetic ketoacidosis in SGLT2I (T) vs SU (C) new-user cohorts under two different propensity score (PS) adjustment across data sources. We report total patient exposure time in 1,000 patient-year (KYr) units, uncalibrated and calibrated hazard ratios (HRs) and their 95% confidence intervals (CIs). Greyed out entries did not pass study diagnostics and are included only for completeness.

| | Patients | | Exposure (KYr) | | Outcomes | | Uncalibrated | | Calibrated | |
|---|---|---|---|---|---|---|---|---|---|---|
| | T | C | T | C | T | C | HR (95% CI) | $p$ | HR (95% CI) | $p$ |
| **PS-stratification / on-treatment 2 TAR** | | | | | | | | | | |
| OptumDOD | 11,606 | 49,554 | 6.3 | 45.0 | 17 | 69 | 2.02 (1.06 - 3.68) | 0.03 | 1.96 (1.06 - 3.65) | 0.03 |
| MDCR | 3,506 | 15,501 | 2.3 | 12.5 | <5 | 14 | 0.76 (0.04 - 4.79) | 0.82 | 0.71 (0.06 - 8.00) | 0.78 |
| Open Claims | 184,585 | 698,404 | 113.9 | 705.8 | 114 | 477 | 1.67 (1.32 - 2.10) | <0.01 | 1.70 (1.25 - 2.30) | <0.01 |
| OptumEHR | 7,821 | 46,097 | 2.0 | 24.9 | <5 | 24 | 2.06 (0.30 - 8.13) | 0.39 | 1.96 (0.38 - 10.1) | 0.43 |
| VA | 7,524 | 53,832 | 4.1 | 48.1 | <10 | 10 | 3.06 (0.49 - 19.7) | 0.24 | 3.32 (0.52 - 21.2) | 0.20 |
| GermanyDA | 4,154 | 1,547 | 3.0 | 1.5 | 0 | 0 | | | | |
| SIDIAP | 3,746 | 2,136 | 4.1 | 4.1 | 0 | <5 | | | | |
| Meta-analysis | 196,191 | 747,958 | 120.2 | 750.8 | 131 | 546 | 1.71 (1.38 - 2.12) | <0.01 | 1.70 (1.28 - 2.27) | <0.01 |
| **PS-matching / on-treatment 2 TAR** | | | | | | | | | | |
| OptumDOD | 10,774 | 48,470 | 6.0 | 44.1 | 15 | 66 | 2.68 (1.16 - 5.91) | 0.02 | 2.62 (1.16 - 5.90) | 0.02 |
| MDCR | 3,036 | 14,734 | 2.1 | 11.9 | <5 | 13 | 1.09 (0.06 - 6.96) | 0.94 | 1.04 (0.09 - 11.7) | 0.97 |
| Open Claims | 169,532 | 681,857 | 106.9 | 690.0 | 106 | 457 | 1.87 (1.41 - 2.47) | <0.01 | 1.92 (1.37 - 2.68) | <0.01 |
| OptumEHR | 7,275 | 43,358 | 1.8 | 23.3 | <5 | 22 | 2.09 (0.11 - 12.7) | 0.55 | 2.05 (0.19 - 22.3) | 0.56 |
| VA | 5,284 | 33,852 | 3.3 | 26.7 | <10 | <10 | 3.03 (0.46 - 18.4) | 0.24 | 3.40 (0.54 - 21.6) | 0.20 |
| GermanyDA | 4,095 | 892 | 3.0 | 0.8 | 0 | 0 | | | | |
| SIDIAP | 3,314 | 971 | 3.8 | 1.5 | 0 | <5 | | | | |
| Meta-analysis | 180,306 | 730,327 | 112.9 | 734.1 | 121 | 523 | 1.94 (1.49 - 2.53) | <0.01 | 1.94 (1.41 - 2.68) | <0.01 |

##### 7.5.7 Diarrhea

**Supplementary Table 294:** Exposures, outcomes and risk estimates for Diarrhea in SGLT2I (T) vs SU (C) new-user cohorts under two different propensity score (PS) adjustment across data sources. We report total patient exposure time in 1,000 patient-year (KYr) units, uncalibrated and calibrated hazard ratios (HRs) and their 95% confidence intervals (CIs). Greyed out entries did not pass study diagnostics and are included only for completeness.

| | Patients | | Exposure (KYr) | | Outcomes | | Uncalibrated | | Calibrated | |
|---|---|---|---|---|---|---|---|---|---|---|
| | T | C | T | C | T | C | HR (95% CI) | $p$ | HR (95% CI) | $p$ |
| **PS-stratification / on-treatment 2 TAR** | | | | | | | | | | |
| OptumDOD | 9,875 | 41,297 | 5.3 | 36.6 | 207 | 1,610 | 0.91 (0.78 - 1.06) | 0.25 | 0.88 (0.75 - 1.04) | 0.17 |
| MDCR | 3,032 | 13,000 | 2.0 | 10.3 | 52 | 385 | 0.71 (0.51 - 0.98) | 0.04 | 0.67 (0.48 - 0.93) | 0.02 |
| Open Claims | 158,033 | 602,791 | 97.6 | 603.8 | 1,489 | 12,463 | 0.82 (0.77 - 0.87) | <0.01 | 0.83 (0.68 - 1.02) | 0.09 |
| OptumEHR | 6,910 | 40,186 | 1.7 | 20.7 | 64 | 791 | 1.08 (0.81 - 1.43) | 0.58 | 1.03 (0.78 - 1.37) | 0.80 |
| VA | 6,417 | 46,833 | 3.5 | 41.3 | 59 | 757 | 0.90 (0.64 - 1.24) | 0.52 | 0.98 (0.70 - 1.36) | 0.85 |
| GermanyDA | 4,090 | 1,526 | 2.9 | 1.5 | 6 | <5 | 9.46 (2.06 - 43.2) | <0.01 | 9.21 (2.01 - 42.2) | <0.01 |
| SIDIAP | 3,550 | 1,997 | 3.9 | 3.8 | 32 | 39 | 0.39 (0.18 - 0.83) | 0.02 | 0.44 (0.20 - 0.94) | 0.04 |
| Meta-analysis | 174,818 | 684,274 | 104.6 | 661.2 | 1,760 | 14,864 | 0.89 (0.77 - 1.02) | 0.08 | 0.88 (0.70 - 1.11) | 0.30 |
| **PS-matching / on-treatment 2 TAR** | | | | | | | | | | |
| OptumDOD | 9,161 | 40,356 | 5.0 | 35.8 | 198 | 1,562 | 0.90 (0.75 - 1.09) | 0.29 | 0.88 (0.73 - 1.07) | 0.24 |
| MDCR | 2,638 | 12,081 | 1.8 | 9.6 | 45 | 352 | 0.81 (0.52 - 1.21) | 0.32 | 0.77 (0.50 - 1.17) | 0.24 |
| Open Claims | 145,007 | 588,132 | 91.5 | 590.0 | 1,387 | 12,049 | 0.77 (0.72 - 0.83) | <0.01 | 0.79 (0.65 - 0.97) | 0.03 |
| OptumEHR | 6,404 | 37,784 | 1.6 | 19.5 | 57 | 706 | 1.11 (0.78 - 1.55) | 0.56 | 1.09 (0.77 - 1.55) | 0.65 |
| VA | 4,546 | 29,232 | 2.8 | 22.8 | 43 | 417 | 0.87 (0.57 - 1.28) | 0.49 | 0.98 (0.65 - 1.46) | 0.85 |
| GermanyDA | 4,031 | 878 | 2.9 | 0.8 | 6 | 0 | | | | |
| SIDIAP | 3,144 | 906 | 3.6 | 1.4 | 28 | 14 | 0.43 (0.14 - 1.30) | 0.14 | 0.59 (0.19 - 1.82) | 0.37 |
| Meta-analysis | 163,210 | 678,353 | 99.9 | 654.9 | 1,687 | 14,669 | 0.85 (0.74 - 0.99) | 0.04 | 0.85 (0.68 - 1.08) | 0.19 |

##### 7.5.8 Genitourinary infection

**Supplementary Table 295:** Exposures, outcomes and risk estimates for Genitourinary infection in SGLT2I (T) vs SU (C) new-user cohorts under two different propensity score (PS) adjustment across data sources. We report total patient exposure time in 1,000 patient-year (KYr) units, uncalibrated and calibrated hazard ratios (HRs) and their 95% confidence intervals (CIs). Greyed out entries did not pass study diagnostics and are included only for completeness.

| | Patients | | Exposure (KYr) | | Outcomes | | Uncalibrated | | Calibrated | |
|---|---|---|---|---|---|---|---|---|---|---|
| | T | C | T | C | T | C | HR (95% CI) | $p$ | HR (95% CI) | $p$ |
| **PS-stratification / on-treatment 2 TAR** | | | | | | | | | | |
| OptumDOD | 9,018 | 35,987 | 4.8 | 31.4 | 292 | 2,392 | 0.88 (0.77 - 1.00) | 0.06 | 0.85 (0.75 - 0.98) | 0.04 |
| MDCR | 2,894 | 11,879 | 1.9 | 9.3 | 89 | 564 | 0.99 (0.76 - 1.27) | 0.93 | 0.93 (0.72 - 1.21) | 0.65 |
| Open Claims | 137,023 | 521,831 | 85.9 | 522.2 | 2,538 | 21,571 | 0.88 (0.84 - 0.92) | <0.01 | 0.90 (0.73 - 1.09) | 0.30 |
| OptumEHR | 6,565 | 37,673 | 1.6 | 19.3 | 80 | 1,165 | 1.07 (0.83 - 1.37) | 0.57 | 1.02 (0.80 - 1.31) | 0.79 |
| VA | 6,609 | 46,264 | 3.6 | 41.0 | 43 | 807 | 0.61 (0.42 - 0.89) | 0.01 | 0.67 (0.46 - 0.98) | 0.04 |
| GermanyDA | 3,188 | 1,172 | 2.3 | 1.1 | 81 | 39 | 1.22 (0.74 - 2.03) | 0.43 | 1.19 (0.72 - 1.97) | 0.51 |
| SIDIAP | 2,815 | 1,678 | 3.1 | 3.1 | 136 | 136 | 0.98 (0.70 - 1.36) | 0.89 | 1.10 (0.78 - 1.54) | 0.61 |
| Meta-analysis | 152,606 | 595,491 | 92.3 | 572.9 | 2,910 | 25,128 | 0.88 (0.85 - 0.92) | <0.01 | 0.88 (0.73 - 1.07) | 0.20 |
| **PS-matching / on-treatment 2 TAR** | | | | | | | | | | |
| OptumDOD | 8,360 | 35,704 | 4.6 | 31.2 | 278 | 2,349 | 0.95 (0.81 - 1.11) | 0.54 | 0.93 (0.79 - 1.09) | 0.43 |
| MDCR | 2,515 | 11,338 | 1.7 | 9.0 | 83 | 523 | 1.03 (0.74 - 1.41) | 0.86 | 0.98 (0.71 - 1.35) | 0.82 |
| Open Claims | 126,202 | 509,338 | 80.8 | 510.6 | 2,380 | 20,722 | 0.91 (0.87 - 0.97) | <0.01 | 0.94 (0.78 - 1.14) | 0.53 |
| OptumEHR | 6,100 | 35,316 | 1.5 | 18.0 | 76 | 1,042 | 1.12 (0.83 - 1.49) | 0.44 | 1.10 (0.82 - 1.49) | 0.56 |
| VA | 4,658 | 29,538 | 2.9 | 23.3 | 28 | 435 | 0.67 (0.40 - 1.06) | 0.10 | 0.75 (0.46 - 1.22) | 0.26 |
| GermanyDA | 3,143 | 685 | 2.2 | 0.6 | 81 | 18 | 1.63 (0.79 - 3.72) | 0.22 | 1.96 (0.90 - 4.27) | 0.11 |
| SIDIAP | 2,499 | 758 | 2.9 | 1.2 | 125 | 56 | 1.09 (0.71 - 1.70) | 0.68 | 1.50 (0.96 - 2.34) | 0.11 |
| Meta-analysis | 143,177 | 591,696 | 88.5 | 568.7 | 2,817 | 24,636 | 0.93 (0.88 - 0.97) | <0.01 | 0.93 (0.77 - 1.12) | 0.43 |

##### 7.5.9 Hyperkalemia

**Supplementary Table 296:** Exposures, outcomes and risk estimates for Hyperkalemia in SGLT2I (T) vs SU (C) new-user cohorts under two different propensity score (PS) adjustment across data sources. We report total patient exposure time in 1,000 patient-year (KYr) units, uncalibrated and calibrated hazard ratios (HRs) and their 95% confidence intervals (CIs). Greyed out entries did not pass study diagnostics and are included only for completeness.

| | Patients | | Exposure (KYr) | | Outcomes | | Uncalibrated | | Calibrated | |
|---|---|---|---|---|---|---|---|---|---|---|
| | T | C | T | C | T | C | HR (95% CI) | $p$ | HR (95% CI) | $p$ |
| **PS-stratification / on-treatment 2 TAR** | | | | | | | | | | |
| OptumDOD | 10,938 | 45,992 | 5.9 | 40.8 | 145 | 1,349 | 0.83 (0.69 - 1.00) | 0.05 | 0.81 (0.67 - 0.98) | 0.04 |
| MDCR | 3,408 | 14,878 | 2.2 | 11.9 | 24 | 253 | 0.68 (0.41 - 1.07) | 0.11 | 0.64 (0.40 - 1.04) | 0.08 |
| Open Claims | 178,385 | 674,573 | 110.1 | 676.1 | 755 | 7,852 | 0.73 (0.67 - 0.79) | <0.01 | 0.75 (0.60 - 0.92) | <0.01 |
| OptumEHR | 7,603 | 44,391 | 1.9 | 23.5 | 26 | 461 | 0.91 (0.58 - 1.36) | 0.65 | 0.86 (0.56 - 1.32) | 0.51 |
| VA | 6,726 | 49,326 | 3.7 | 43.5 | 64 | 758 | 0.98 (0.70 - 1.35) | 0.91 | 1.07 (0.77 - 1.48) | 0.71 |
| GermanyDA | 3,801 | 1,459 | 2.7 | 1.4 | 41 | 16 | 0.79 (0.38 - 1.72) | 0.54 | 0.77 (0.36 - 1.65) | 0.52 |
| SIDIAP | 3,509 | 2,023 | 3.8 | 3.9 | 56 | 48 | 0.65 (0.36 - 1.15) | 0.14 | 0.73 (0.41 - 1.30) | 0.31 |
| Meta-analysis | 196,926 | 764,956 | 117.9 | 740.4 | 926 | 9,662 | 0.77 (0.69 - 0.86) | <0.01 | 0.77 (0.62 - 0.95) | 0.02 |
| **PS-matching / on-treatment 2 TAR** | | | | | | | | | | |
| OptumDOD | 10,178 | 45,090 | 5.6 | 40.1 | 137 | 1,307 | 0.80 (0.64 - 1.00) | 0.05 | 0.78 (0.62 - 0.98) | 0.04 |
| MDCR | 2,961 | 14,148 | 2.0 | 11.3 | 23 | 232 | 0.70 (0.38 - 1.21) | 0.23 | 0.67 (0.38 - 1.19) | 0.18 |
| Open Claims | 164,246 | 659,135 | 103.6 | 661.5 | 664 | 7,480 | 0.70 (0.64 - 0.78) | <0.01 | 0.72 (0.59 - 0.89) | <0.01 |
| OptumEHR | 7,051 | 41,907 | 1.7 | 22.1 | 22 | 418 | 0.97 (0.57 - 1.57) | 0.89 | 0.95 (0.57 - 1.59) | 0.83 |
| VA | 4,822 | 31,325 | 3.0 | 24.6 | 49 | 378 | 0.96 (0.63 - 1.42) | 0.83 | 1.07 (0.71 - 1.62) | 0.74 |
| GermanyDA | 3,746 | 828 | 2.7 | 0.7 | 40 | 10 | 0.74 (0.28 - 2.22) | 0.57 | 0.89 (0.31 - 2.53) | 0.72 |
| SIDIAP | 3,133 | 916 | 3.6 | 1.4 | 43 | 17 | 0.66 (0.30 - 1.47) | 0.30 | 0.90 (0.41 - 2.01) | 0.75 |
| Meta-analysis | 184,436 | 760,280 | 113.0 | 735.1 | 846 | 9,437 | 0.73 (0.66 - 0.81) | <0.01 | 0.73 (0.59 - 0.91) | <0.01 |

##### 7.5.10 Hypoglycemia

**Supplementary Table 297:** Exposures, outcomes and risk estimates for Hypoglycemia in SGLT2I (T) vs SU (C) new-user cohorts under two different propensity score (PS) adjustment across data sources. We report total patient exposure time in 1,000 patient-year (KYr) units, uncalibrated and calibrated hazard ratios (HRs) and their 95% confidence intervals (CIs). Greyed out entries did not pass study diagnostics and are included only for completeness.

| | Patients | | Exposure (KYr) | | Outcomes | | Uncalibrated | | Calibrated | |
|---|---|---|---|---|---|---|---|---|---|---|
| | T | C | T | C | T | C | HR (95% CI) | $p$ | HR (95% CI) | $p$ |
| **PS-stratification / on-treatment 2 TAR** | | | | | | | | | | |
| OptumDOD | 11,439 | 48,868 | 6.2 | 44.0 | 36 | 1,228 | 0.26 (0.18 - 0.36) | <0.01 | 0.25 (0.18 - 0.36) | <0.01 |
| MDCR | 3,474 | 15,356 | 2.3 | 12.3 | <5 | 286 | 0.07 (0.02 - 0.20) | <0.01 | 0.07 (0.02 - 0.23) | <0.01 |
| Open Claims | 181,772 | 688,223 | 112.2 | 693.2 | 159 | 8,052 | 0.14 (0.12 - 0.17) | <0.01 | 0.14 (0.11 - 0.19) | <0.01 |
| OptumEHR | 7,748 | 45,662 | 1.9 | 24.4 | <5 | 461 | 0.08 (0.02 - 0.22) | <0.01 | 0.08 (0.02 - 0.26) | <0.01 |
| VA | 7,425 | 53,312 | 4.0 | 47.5 | <10 | 422 | 0.20 (0.07 - 0.44) | <0.01 | 0.21 (0.09 - 0.52) | <0.01 |
| GermanyDA | 4,132 | 1,538 | 3.0 | 1.5 | <5 | 11 | 0.36 (0.07 - 1.58) | 0.21 | 0.35 (0.07 - 1.70) | 0.20 |
| SIDIAP | 3,739 | 2,133 | 4.1 | 4.1 | 0 | 18 | | | | |
| Meta-analysis | 200,959 | 782,753 | 120.4 | 761.6 | <200 | 9,741 | 0.17 (0.10 - 0.29) | <0.01 | 0.17 (0.09 - 0.30) | <0.01 |
| **PS-matching / on-treatment 2 TAR** | | | | | | | | | | |
| OptumDOD | 10,623 | 47,820 | 5.9 | 43.1 | 33 | 1,178 | 0.25 (0.17 - 0.37) | <0.01 | 0.25 (0.17 - 0.36) | <0.01 |
| MDCR | 3,002 | 14,595 | 2.1 | 11.7 | <5 | 250 | 0.08 (0.01 - 0.25) | <0.01 | 0.07 (0.02 - 0.33) | <0.01 |
| Open Claims | 167,032 | 671,919 | 105.4 | 677.7 | 149 | 7,673 | 0.13 (0.11 - 0.16) | <0.01 | 0.14 (0.10 - 0.18) | <0.01 |
| OptumEHR | 7,209 | 42,966 | 1.8 | 22.9 | <5 | 416 | 0.10 (0.02 - 0.27) | <0.01 | 0.10 (0.03 - 0.33) | <0.01 |
| VA | 5,220 | 33,278 | 3.2 | 26.1 | <10 | 224 | 0.18 (0.04 - 0.49) | <0.01 | 0.20 (0.06 - 0.68) | <0.01 |
| GermanyDA | 4,074 | 889 | 3.0 | 0.8 | <5 | <5 | 0.00 (0.00 - 2.91) | 0.20 | 0.00 (0.00 - 31002) | 0.20 |
| SIDIAP | 3,311 | 970 | 3.8 | 1.5 | 0 | 9 | | | | |
| Meta-analysis | 187,866 | 777,300 | 115.1 | 755.4 | <192 | 9,517 | 0.15 (0.09 - 0.25) | <0.01 | 0.15 (0.09 - 0.26) | <0.01 |

##### 7.5.11 Hypotension

**Supplementary Table 298:** Exposures, outcomes and risk estimates for Hypotension in SGLT2I (T) vs SU (C) new-user cohorts under two different propensity score (PS) adjustment across data sources. We report total patient exposure time in 1,000 patient-year (KYr) units, uncalibrated and calibrated hazard ratios (HRs) and their 95% confidence intervals (CIs). Greyed out entries did not pass study diagnostics and are included only for completeness.

| | Patients | | Exposure (KYr) | | Outcomes | | Uncalibrated | | Calibrated | |
|---|---|---|---|---|---|---|---|---|---|---|
| | T | C | T | C | T | C | HR (95% CI) | $p$ | HR (95% CI) | $p$ |
| **PS-stratification / on-treatment 2 TAR** | | | | | | | | | | |
| OptumDOD | 10,826 | 45,890 | 5.9 | 41.2 | 195 | 1,289 | 1.16 (0.98 - 1.36) | 0.09 | 1.12 (0.95 - 1.33) | 0.22 |
| MDCR | 3,342 | 14,481 | 2.2 | 11.6 | 53 | 295 | 1.61 (1.12 - 2.25) | <0.01 | 1.52 (1.07 - 2.15) | 0.03 |
| Open Claims | 174,134 | 663,104 | 107.6 | 668.3 | 1,261 | 8,668 | 1.08 (1.01 - 1.16) | 0.02 | 1.10 (0.90 - 1.36) | 0.37 |
| OptumEHR | 7,517 | 44,224 | 1.9 | 23.5 | 35 | 528 | 1.00 (0.68 - 1.44) | 0.98 | 0.95 (0.66 - 1.39) | 0.80 |
| VA | 6,697 | 49,540 | 3.7 | 43.9 | 103 | 616 | 1.54 (1.16 - 2.05) | <0.01 | 1.68 (1.26 - 2.23) | <0.01 |
| GermanyDA | 4,007 | 1,503 | 2.9 | 1.5 | 19 | 8 | 1.27 (0.34 - 5.27) | 0.73 | 1.24 (0.31 - 4.88) | 0.76 |
| SIDIAP | 3,705 | 2,123 | 4.0 | 4.1 | 19 | 7 | 2.33 (0.66 - 8.95) | 0.20 | 2.63 (0.71 - 9.70) | 0.16 |
| Meta-analysis | 192,477 | 753,218 | 115.3 | 733.0 | 1,491 | 10,485 | 1.09 (1.03 - 1.16) | <0.01 | 1.09 (0.89 - 1.32) | 0.40 |
| **PS-matching / on-treatment 2 TAR** | | | | | | | | | | |
| OptumDOD | 10,106 | 45,167 | 5.6 | 40.6 | 184 | 1,258 | 1.30 (1.06 - 1.58) | 0.01 | 1.27 (1.04 - 1.55) | 0.03 |
| MDCR | 2,894 | 13,836 | 2.0 | 11.1 | 46 | 267 | 1.53 (0.98 - 2.34) | 0.06 | 1.46 (0.94 - 2.26) | 0.10 |
| Open Claims | 160,749 | 647,650 | 101.4 | 653.6 | 1,121 | 8,250 | 1.07 (0.99 - 1.16) | 0.08 | 1.10 (0.90 - 1.35) | 0.36 |
| OptumEHR | 6,986 | 41,566 | 1.7 | 22.0 | 32 | 463 | 1.04 (0.65 - 1.60) | 0.88 | 1.02 (0.65 - 1.61) | 0.85 |
| VA | 4,826 | 31,081 | 3.0 | 24.4 | 75 | 333 | 1.45 (1.00 - 2.07) | 0.05 | 1.63 (1.13 - 2.35) | 0.01 |
| GermanyDA | 3,951 | 866 | 2.9 | 0.8 | 18 | <5 | 1.20 (0.25 - 8.82) | 0.84 | 1.44 (0.24 - 8.52) | 0.69 |
| SIDIAP | 3,283 | 963 | 3.7 | 1.5 | 18 | <5 | 2.72 (0.43 - 53.2) | 0.42 | 3.74 (0.33 - 41.9) | 0.29 |
| Meta-analysis | 180,735 | 748,219 | 110.7 | 727.4 | 1,383 | 10,238 | 1.17 (1.01 - 1.36) | 0.04 | 1.17 (0.93 - 1.49) | 0.18 |

##### 7.5.12 Joint pain

**Supplementary Table 299:** Exposures, outcomes and risk estimates for Joint pain in SGLT2I (T) vs SU (C) new-user cohorts under two different propensity score (PS) adjustment across data sources. We report total patient exposure time in 1,000 patient-year (KYr) units, uncalibrated and calibrated hazard ratios (HRs) and their 95% confidence intervals (CIs). Greyed out entries did not pass study diagnostics and are included only for completeness.

| | Patients | | Exposure (KYr) | | Outcomes | | Uncalibrated | | Calibrated | |
|---|---|---|---|---|---|---|---|---|---|---|
| | T | C | T | C | T | C | HR (95% CI) | $p$ | HR (95% CI) | $p$ |
| **PS-stratification / on-treatment 2 TAR** | | | | | | | | | | |
| OptumDOD | 10,899 | 46,674 | 5.9 | 42.2 | 75 | 534 | 0.91 (0.69 - 1.18) | 0.47 | 0.88 (0.68 - 1.15) | 0.37 |
| MDCR | 3,368 | 14,797 | 2.2 | 11.9 | 23 | 113 | 0.97 (0.57 - 1.58) | 0.89 | 0.91 (0.55 - 1.52) | 0.73 |
| Open Claims | 172,024 | 661,104 | 106.5 | 668.3 | 545 | 3,909 | 0.87 (0.79 - 0.96) | <0.01 | 0.88 (0.71 - 1.10) | 0.29 |
| OptumEHR | 7,405 | 43,892 | 1.8 | 23.4 | 24 | 216 | 1.45 (0.89 - 2.27) | 0.12 | 1.38 (0.86 - 2.21) | 0.19 |
| VA | 6,617 | 47,471 | 3.6 | 42.3 | 19 | 237 | 1.12 (0.60 - 2.00) | 0.72 | 1.21 (0.66 - 2.23) | 0.54 |
| GermanyDA | 3,517 | 1,353 | 2.5 | 1.3 | 34 | 31 | 0.90 (0.47 - 1.74) | 0.74 | 0.87 (0.45 - 1.69) | 0.66 |
| SIDIAP | 2,847 | 1,738 | 3.1 | 3.2 | 56 | 84 | 0.62 (0.39 - 0.97) | 0.04 | 0.70 (0.44 - 1.10) | 0.15 |
| Meta-analysis | 190,328 | 751,670 | 114.2 | 733.9 | 644 | 4,659 | 0.95 (0.77 - 1.17) | 0.62 | 0.95 (0.72 - 1.25) | 0.70 |
| **PS-matching / on-treatment 2 TAR** | | | | | | | | | | |
| OptumDOD | 10,129 | 45,692 | 5.6 | 41.3 | 75 | 520 | 0.87 (0.62 - 1.21) | 0.43 | 0.85 (0.61 - 1.19) | 0.37 |
| MDCR | 2,921 | 14,040 | 2.0 | 11.3 | 22 | 106 | 1.04 (0.50 - 2.03) | 0.92 | 0.99 (0.49 - 1.99) | 0.92 |
| Open Claims | 158,135 | 644,855 | 100.0 | 652.8 | 506 | 3,784 | 0.85 (0.75 - 0.96) | <0.01 | 0.87 (0.70 - 1.09) | 0.25 |
| OptumEHR | 6,872 | 41,316 | 1.7 | 22.0 | 22 | 200 | 1.39 (0.72 - 2.54) | 0.31 | 1.36 (0.72 - 2.56) | 0.35 |
| VA | 4,695 | 29,907 | 2.9 | 23.8 | 13 | 113 | 0.95 (0.41 - 1.98) | 0.89 | 1.06 (0.48 - 2.34) | 0.88 |
| GermanyDA | 3,453 | 771 | 2.4 | 0.7 | 34 | 12 | 2.22 (0.78 - 7.95) | 0.18 | 2.66 (0.83 - 8.54) | 0.11 |
| SIDIAP | 2,523 | 773 | 2.9 | 1.1 | 53 | 32 | 0.59 (0.29 - 1.19) | 0.14 | 0.80 (0.40 - 1.64) | 0.56 |
| Meta-analysis | 178,057 | 745,903 | 109.4 | 727.5 | 625 | 4,610 | 0.87 (0.78 - 0.97) | 0.01 | 0.87 (0.70 - 1.08) | 0.21 |

##### 7.5.13 Lower extremity amputation

**Supplementary Table 300:** Exposures, outcomes and risk estimates for Lower extremity amputation in SGLT2I (T) vs SU (C) new-user cohorts under two different propensity score (PS) adjustment across data sources. We report total patient exposure time in 1,000 patient-year (KYr) units, uncalibrated and calibrated hazard ratios (HRs) and their 95% confidence intervals (CIs). Greyed out entries did not pass study diagnostics and are included only for completeness.

| | Patients | | Exposure (KYr) | | Outcomes | | Uncalibrated | | Calibrated | |
|---|---|---|---|---|---|---|---|---|---|---|
| | T | C | T | C | T | C | HR (95% CI) | $p$ | HR (95% CI) | $p$ |
| **PS-stratification / on-treatment 2 TAR** | | | | | | | | | | |
| OptumDOD | 11,589 | 49,525 | 6.3 | 45.0 | <5 | 65 | 0.61 (0.18 - 1.55) | 0.37 | 0.59 (0.20 - 1.74) | 0.34 |
| MDCR | 3,500 | 15,493 | 2.3 | 12.5 | <5 | 13 | 1.32 (0.26 - 4.98) | 0.72 | 1.25 (0.28 - 5.50) | 0.77 |
| Open Claims | 184,471 | 698,060 | 113.8 | 705.1 | 57 | 449 | 1.19 (0.87 - 1.61) | 0.25 | 1.22 (0.85 - 1.75) | 0.30 |
| OptumEHR | 7,821 | 46,072 | 2.0 | 24.8 | 0 | 16 | | | | |
| VA | 7,511 | 53,702 | 4.1 | 47.9 | <10 | 40 | 1.86 (0.47 - 6.46) | 0.35 | 2.02 (0.55 - 7.45) | 0.29 |
| GermanyDA | 4,154 | 1,547 | 3.0 | 1.5 | 0 | 0 | | | | |
| SIDIAP | 3,745 | 2,134 | 4.1 | 4.1 | 0 | 0 | | | | |
| Meta-analysis | 196,060 | 747,585 | 120.1 | 750.0 | <62 | 514 | 1.05 (0.63 - 1.76) | 0.85 | 1.05 (0.60 - 1.82) | 0.87 |
| **PS-matching / on-treatment 2 TAR** | | | | | | | | | | |
| OptumDOD | 10,760 | 48,449 | 6.0 | 44.0 | <5 | 63 | 0.67 (0.15 - 2.03) | 0.55 | 0.66 (0.18 - 2.38) | 0.52 |
| MDCR | 3,034 | 14,725 | 2.1 | 11.9 | <5 | 12 | 1.28 (0.17 - 6.51) | 0.79 | 1.22 (0.20 - 7.52) | 0.83 |
| Open Claims | 169,437 | 681,660 | 106.8 | 689.5 | 55 | 417 | 1.29 (0.90 - 1.82) | 0.16 | 1.33 (0.89 - 1.97) | 0.17 |
| OptumEHR | 7,278 | 43,336 | 1.8 | 23.3 | 0 | 14 | | | | |
| VA | 5,273 | 33,797 | 3.2 | 26.7 | <10 | 20 | 3.15 (0.55 - 13.4) | 0.16 | 3.54 (0.72 - 17.4) | 0.12 |
| GermanyDA | 4,095 | 892 | 3.0 | 0.8 | 0 | 0 | | | | |
| SIDIAP | 3,313 | 970 | 3.8 | 1.5 | 0 | 0 | | | | |
| Meta-analysis | 180,197 | 730,109 | 112.8 | 733.5 | <60 | 480 | 1.23 (0.88 - 1.73) | 0.23 | 1.23 (0.84 - 1.82) | 0.29 |

##### 7.5.14 Nausea

**Supplementary Table 301:** Exposures, outcomes and risk estimates for Nausea in SGLT2I (T) vs SU (C) new-user cohorts under two different propensity score (PS) adjustment across data sources. We report total patient exposure time in 1,000 patient-year (KYr) units, uncalibrated and calibrated hazard ratios (HRs) and their 95% confidence intervals (CIs). Greyed out entries did not pass study diagnostics and are included only for completeness.

| | Patients | | Exposure (KYr) | | Outcomes | | Uncalibrated | | Calibrated | |
|---|---|---|---|---|---|---|---|---|---|---|
| | T | C | T | C | T | C | HR (95% CI) | $p$ | HR (95% CI) | $p$ |
| **PS-stratification / on-treatment 2 TAR** | | | | | | | | | | |
| OptumDOD | 9,971 | 41,942 | 5.4 | 37.6 | 199 | 1,626 | 0.88 (0.75 - 1.03) | 0.11 | 0.85 (0.72 - 1.00) | 0.07 |
| MDCR | 3,120 | 13,382 | 2.0 | 10.7 | 62 | 407 | 0.98 (0.72 - 1.31) | 0.89 | 0.93 (0.68 - 1.25) | 0.65 |
| Open Claims | 158,462 | 607,675 | 98.7 | 615.0 | 1,483 | 12,728 | 0.82 (0.77 - 0.87) | <0.01 | 0.83 (0.68 - 1.02) | 0.09 |
| OptumEHR | 7,090 | 41,646 | 1.8 | 21.9 | 53 | 699 | 1.21 (0.88 - 1.63) | 0.22 | 1.15 (0.85 - 1.57) | 0.39 |
| VA | 6,967 | 50,402 | 3.8 | 45.1 | 23 | 344 | 0.80 (0.46 - 1.35) | 0.42 | 0.87 (0.51 - 1.49) | 0.63 |
| GermanyDA | 3,888 | 1,428 | 2.8 | 1.4 | 18 | 15 | 1.58 (0.59 - 4.24) | 0.37 | 1.54 (0.57 - 4.12) | 0.40 |
| SIDIAP | 3,589 | 2,054 | 4.0 | 3.9 | 16 | 29 | 0.40 (0.16 - 0.96) | 0.05 | 0.45 (0.18 - 1.13) | 0.10 |
| Meta-analysis | 175,523 | 691,263 | 105.9 | 674.5 | 1,735 | 15,053 | 0.91 (0.75 - 1.10) | 0.34 | 0.91 (0.70 - 1.19) | 0.48 |
| **PS-matching / on-treatment 2 TAR** | | | | | | | | | | |
| OptumDOD | 9,287 | 41,144 | 5.2 | 36.9 | 190 | 1,581 | 0.94 (0.77 - 1.13) | 0.50 | 0.91 (0.75 - 1.11) | 0.41 |
| MDCR | 2,721 | 12,944 | 1.8 | 10.4 | 59 | 392 | 1.06 (0.72 - 1.52) | 0.77 | 1.01 (0.69 - 1.46) | 0.86 |
| Open Claims | 145,552 | 593,196 | 92.7 | 601.1 | 1,394 | 12,265 | 0.83 (0.77 - 0.89) | <0.01 | 0.85 (0.70 - 1.04) | 0.12 |
| OptumEHR | 6,581 | 39,191 | 1.6 | 20.6 | 49 | 626 | 1.15 (0.80 - 1.64) | 0.44 | 1.14 (0.79 - 1.64) | 0.53 |
| VA | 4,955 | 31,759 | 3.1 | 25.2 | 13 | 176 | 0.82 (0.38 - 1.59) | 0.58 | 0.92 (0.45 - 1.88) | 0.81 |
| GermanyDA | 3,830 | 831 | 2.8 | 0.7 | 18 | <5 | 3.51 (0.59 - 67.5) | 0.30 | 4.22 (0.39 - 45.4) | 0.24 |
| SIDIAP | 3,188 | 933 | 3.7 | 1.5 | 12 | 10 | 0.85 (0.21 - 3.60) | 0.82 | 1.17 (0.28 - 4.87) | 0.83 |
| Meta-analysis | 164,141 | 686,475 | 101.3 | 668.9 | 1,692 | 14,864 | 0.92 (0.80 - 1.06) | 0.25 | 0.92 (0.73 - 1.16) | 0.48 |

##### 7.5.15 Peripheral edema

**Supplementary Table 302:** Exposures, outcomes and risk estimates for Peripheral edema in SGLT2I (T) vs SU (C) new-user cohorts under two different propensity score (PS) adjustment across data sources. We report total patient exposure time in 1,000 patient-year (KYr) units, uncalibrated and calibrated hazard ratios (HRs) and their 95% confidence intervals (CIs). Greyed out entries did not pass study diagnostics and are included only for completeness.

| | Patients | | Exposure (KYr) | | Outcomes | | Uncalibrated | | Calibrated | |
|---|---|---|---|---|---|---|---|---|---|---|
| | T | C | T | C | T | C | HR (95% CI) | $p$ | HR (95% CI) | $p$ |
| **PS-stratification / on-treatment 2 TAR** | | | | | | | | | | |
| OptumDOD | 8,491 | 35,457 | 4.6 | 30.4 | 298 | 3,015 | 0.69 (0.60 - 0.78) | <0.01 | 0.67 (0.58 - 0.76) | <0.01 |
| MDCR | 2,647 | 11,067 | 1.7 | 8.4 | 74 | 812 | 0.56 (0.43 - 0.73) | <0.01 | 0.53 (0.41 - 0.70) | <0.01 |
| Open Claims | 132,741 | 519,881 | 83.0 | 513.9 | 2,095 | 24,190 | 0.66 (0.63 - 0.69) | <0.01 | 0.67 (0.55 - 0.82) | <0.01 |
| OptumEHR | 6,272 | 36,844 | 1.5 | 18.5 | 42 | 1,388 | 0.48 (0.34 - 0.66) | <0.01 | 0.46 (0.33 - 0.63) | <0.01 |
| VA | 5,373 | 40,662 | 3.0 | 35.3 | 94 | 1,582 | 0.71 (0.54 - 0.91) | <0.01 | 0.77 (0.59 - 0.99) | 0.05 |
| GermanyDA | 3,374 | 1,282 | 2.4 | 1.2 | 51 | 61 | 0.43 (0.25 - 0.73) | <0.01 | 0.42 (0.24 - 0.72) | <0.01 |
| SIDIAP | 3,190 | 1,916 | 3.6 | 3.6 | 40 | 77 | 0.61 (0.35 - 1.03) | 0.07 | 0.69 (0.40 - 1.18) | 0.20 |
| Meta-analysis | 147,504 | 592,182 | 89.1 | 562.7 | 2,435 | 28,593 | 0.66 (0.63 - 0.69) | <0.01 | 0.66 (0.54 - 0.79) | <0.01 |
| **PS-matching / on-treatment 2 TAR** | | | | | | | | | | |
| OptumDOD | 7,985 | 34,773 | 4.4 | 29.8 | 280 | 2,917 | 0.63 (0.54 - 0.73) | <0.01 | 0.61 (0.52 - 0.72) | <0.01 |
| MDCR | 2,338 | 10,699 | 1.6 | 8.2 | 67 | 763 | 0.62 (0.44 - 0.85) | <0.01 | 0.59 (0.42 - 0.82) | <0.01 |
| Open Claims | 123,465 | 508,290 | 78.6 | 503.3 | 1,964 | 23,222 | 0.67 (0.64 - 0.71) | <0.01 | 0.69 (0.57 - 0.84) | <0.01 |
| OptumEHR | 5,847 | 34,817 | 1.4 | 17.5 | 42 | 1,253 | 0.52 (0.36 - 0.74) | <0.01 | 0.51 (0.35 - 0.74) | <0.01 |
| VA | 3,926 | 25,913 | 2.4 | 20.1 | 67 | 848 | 0.69 (0.50 - 0.94) | 0.02 | 0.78 (0.57 - 1.07) | 0.13 |
| GermanyDA | 3,323 | 734 | 2.4 | 0.6 | 50 | 29 | 0.42 (0.23 - 0.80) | <0.01 | 0.51 (0.27 - 0.96) | 0.06 |
| SIDIAP | 2,854 | 856 | 3.3 | 1.3 | 37 | 28 | 0.53 (0.24 - 1.16) | 0.12 | 0.72 (0.33 - 1.62) | 0.45 |
| Meta-analysis | 139,635 | 588,579 | 86.0 | 558.8 | 2,353 | 28,155 | 0.66 (0.62 - 0.70) | <0.01 | 0.66 (0.54 - 0.80) | <0.01 |

##### 7.5.16 Photosensitivity

**Supplementary Table 303:** Exposures, outcomes and risk estimates for Photosensitivity in SGLT2I (T) vs SU (C) new-user cohorts under two different propensity score (PS) adjustment across data sources. We report total patient exposure time in 1,000 patient-year (KYr) units, uncalibrated and calibrated hazard ratios (HRs) and their 95% confidence intervals (CIs). Greyed out entries did not pass study diagnostics and are included only for completeness.

| | Patients | | Exposure (KYr) | | Outcomes | | Uncalibrated | | Calibrated | |
|---|---|---|---|---|---|---|---|---|---|---|
| | T | C | T | C | T | C | HR (95% CI) | $p$ | HR (95% CI) | $p$ |
| **PS-stratification / on-treatment 2 TAR** | | | | | | | | | | |
| OptumDOD | 11,516 | 49,068 | 6.3 | 44.5 | <5 | 36 | 0.97 (0.22 - 3.07) | 0.97 | 0.94 (0.25 - 3.56) | 0.93 |
| MDCR | 3,464 | 15,230 | 2.3 | 12.3 | <5 | 29 | 0.79 (0.17 - 2.59) | 0.73 | 0.74 (0.19 - 2.91) | 0.67 |
| Open Claims | 181,939 | 690,142 | 112.1 | 696.0 | 26 | 381 | 0.96 (0.61 - 1.43) | 0.84 | 0.97 (0.61 - 1.55) | 0.91 |
| OptumEHR | 7,758 | 45,589 | 1.9 | 24.5 | 0 | 61 | | | | |
| VA | 7,386 | 52,854 | 4.0 | 47.2 | 0 | 27 | | | | |
| GermanyDA | 4,146 | 1,544 | 3.0 | 1.5 | 0 | <5 | | | | |
| SIDIAP | 3,737 | 2,132 | 4.1 | 4.1 | 0 | <5 | | | | |
| Meta-analysis | 193,455 | 739,210 | 118.4 | 740.5 | <31 | 417 | 0.96 (0.64 - 1.43) | 0.83 | 0.96 (0.61 - 1.49) | 0.84 |
| **PS-matching / on-treatment 2 TAR** | | | | | | | | | | |
| OptumDOD | 10,694 | 48,019 | 6.0 | 43.6 | <5 | 33 | 2.94 (0.41 - 13.0) | 0.22 | 2.87 (0.51 - 16.3) | 0.23 |
| MDCR | 3,000 | 14,491 | 2.0 | 11.7 | <5 | 29 | 1.04 (0.21 - 3.77) | 0.96 | 0.99 (0.24 - 4.14) | 0.96 |
| Open Claims | 167,162 | 673,821 | 105.2 | 680.5 | 25 | 346 | 1.02 (0.61 - 1.61) | 0.95 | 1.04 (0.62 - 1.75) | 0.87 |
| OptumEHR | 7,222 | 42,897 | 1.8 | 23.0 | 0 | 53 | | | | |
| VA | 5,202 | 33,266 | 3.2 | 26.3 | 0 | <10 | | | | |
| GermanyDA | 4,087 | 891 | 3.0 | 0.8 | 0 | 0 | | | | |
| SIDIAP | 3,308 | 970 | 3.8 | 1.5 | 0 | 0 | | | | |
| Meta-analysis | 180,856 | 736,331 | 113.2 | 735.8 | <35 | 408 | 1.09 (0.70 - 1.70) | 0.70 | 1.09 (0.68 - 1.76) | 0.72 |

##### 7.5.17 Venous thromboembolism

**Supplementary Table 304:** Exposures, outcomes and risk estimates for Venous thromboembolism in SGLT2I (T) vs SU (C) new-user cohorts under two different propensity score (PS) adjustment across data sources. We report total patient exposure time in 1,000 patient-year (KYr) units, uncalibrated and calibrated hazard ratios (HRs) and their 95% confidence intervals (CIs). Greyed out entries did not pass study diagnostics and are included only for completeness.

| | Patients | | Exposure (KYr) | | Outcomes | | Uncalibrated | | Calibrated | |
|---|---|---|---|---|---|---|---|---|---|---|
| | T | C | T | C | T | C | HR (95% CI) | $p$ | HR (95% CI) | $p$ |
| **PS-stratification / on-treatment 2 TAR** | | | | | | | | | | |
| OptumDOD | 11,177 | 47,431 | 6.1 | 42.8 | 57 | 597 | 0.81 (0.60 - 1.08) | 0.17 | 0.79 (0.59 - 1.06) | 0.13 |
| MDCR | 3,367 | 14,720 | 2.2 | 11.8 | 18 | 187 | 0.68 (0.39 - 1.15) | 0.18 | 0.65 (0.37 - 1.12) | 0.13 |
| Open Claims | 176,274 | 668,987 | 109.0 | 674.2 | 567 | 5,547 | 0.77 (0.70 - 0.85) | <0.01 | 0.79 (0.64 - 0.98) | 0.04 |
| OptumEHR | 7,536 | 44,398 | 1.9 | 23.7 | 14 | 282 | 0.97 (0.53 - 1.66) | 0.93 | 0.93 (0.52 - 1.64) | 0.79 |
| VA | 7,154 | 51,672 | 3.9 | 46.1 | 16 | 246 | 0.56 (0.29 - 1.03) | 0.07 | 0.61 (0.33 - 1.14) | 0.13 |
| GermanyDA | 3,838 | 1,426 | 2.7 | 1.4 | 25 | 20 | 0.72 (0.31 - 1.65) | 0.43 | 0.70 (0.30 - 1.61) | 0.42 |
| SIDIAP | 3,597 | 2,051 | 3.9 | 3.9 | 18 | 22 | 0.56 (0.24 - 1.28) | 0.17 | 0.63 (0.28 - 1.45) | 0.30 |
| Meta-analysis | 194,987 | 760,816 | 116.9 | 740.7 | 638 | 6,426 | 0.78 (0.72 - 0.85) | <0.01 | 0.78 (0.63 - 0.96) | 0.02 |
| **PS-matching / on-treatment 2 TAR** | | | | | | | | | | |
| OptumDOD | 10,388 | 46,555 | 5.8 | 42.1 | 52 | 567 | 0.83 (0.57 - 1.17) | 0.30 | 0.81 (0.56 - 1.16) | 0.26 |
| MDCR | 2,917 | 13,883 | 2.0 | 11.2 | 16 | 169 | 1.17 (0.60 - 2.13) | 0.64 | 1.11 (0.59 - 2.10) | 0.75 |
| Open Claims | 162,134 | 652,654 | 102.3 | 658.7 | 534 | 5,257 | 0.74 (0.66 - 0.83) | <0.01 | 0.76 (0.61 - 0.94) | 0.02 |
| OptumEHR | 7,012 | 41,767 | 1.7 | 22.2 | 14 | 248 | 1.21 (0.61 - 2.21) | 0.57 | 1.19 (0.62 - 2.27) | 0.61 |
| VA | 5,042 | 32,270 | 3.1 | 25.4 | 12 | 120 | 0.62 (0.24 - 1.40) | 0.29 | 0.70 (0.29 - 1.68) | 0.43 |
| GermanyDA | 3,782 | 828 | 2.7 | 0.7 | 25 | 9 | 0.54 (0.17 - 1.94) | 0.32 | 0.65 (0.19 - 2.22) | 0.50 |
| SIDIAP | 3,161 | 924 | 3.6 | 1.4 | 16 | 12 | 1.23 (0.42 - 4.11) | 0.72 | 1.70 (0.54 - 5.34) | 0.39 |
| Meta-analysis | 182,451 | 754,859 | 111.8 | 734.2 | 616 | 6,241 | 0.83 (0.67 - 1.02) | 0.07 | 0.83 (0.63 - 1.09) | 0.18 |

##### 7.5.18 Vomiting

**Supplementary Table 305:** Exposures, outcomes and risk estimates for Vomiting in SGLT2I (T) vs SU (C) new-user cohorts under two different propensity score (PS) adjustment across data sources. We report total patient exposure time in 1,000 patient-year (KYr) units, uncalibrated and calibrated hazard ratios (HRs) and their 95% confidence intervals (CIs). Greyed out entries did not pass study diagnostics and are included only for completeness.

| | Patients | | Exposure (KYr) | | Outcomes | | Uncalibrated | | Calibrated | |
|---|---|---|---|---|---|---|---|---|---|---|
| | T | C | T | C | T | C | HR (95% CI) | $p$ | HR (95% CI) | $p$ |
| **PS-stratification / on-treatment 2 TAR** | | | | | | | | | | |
| OptumDOD | 10,476 | 43,978 | 5.7 | 39.6 | 144 | 1,276 | 0.82 (0.68 - 0.99) | 0.04 | 0.80 (0.66 - 0.96) | 0.03 |
| MDCR | 3,271 | 13,927 | 2.1 | 11.2 | 45 | 325 | 0.97 (0.67 - 1.36) | 0.86 | 0.92 (0.64 - 1.31) | 0.65 |
| Open Claims | 165,476 | 628,373 | 102.8 | 636.2 | 1,119 | 10,083 | 0.81 (0.76 - 0.87) | <0.01 | 0.83 (0.67 - 1.02) | 0.08 |
| OptumEHR | 7,398 | 43,245 | 1.8 | 23.0 | 35 | 484 | 1.21 (0.83 - 1.73) | 0.30 | 1.15 (0.80 - 1.68) | 0.47 |
| VA | 7,106 | 51,346 | 3.9 | 46.0 | 19 | 261 | 0.71 (0.38 - 1.25) | 0.25 | 0.77 (0.43 - 1.39) | 0.39 |
| GermanyDA | 3,887 | 1,428 | 2.8 | 1.4 | 18 | 15 | 1.58 (0.59 - 4.25) | 0.36 | 1.54 (0.57 - 4.14) | 0.40 |
| SIDIAP | 3,583 | 2,045 | 3.9 | 3.9 | 18 | 27 | 0.46 (0.19 - 1.07) | 0.08 | 0.52 (0.21 - 1.24) | 0.15 |
| Meta-analysis | 183,350 | 715,596 | 110.3 | 698.8 | 1,298 | 11,843 | 0.82 (0.77 - 0.88) | <0.01 | 0.82 (0.67 - 1.00) | 0.05 |
| **PS-matching / on-treatment 2 TAR** | | | | | | | | | | |
| OptumDOD | 9,734 | 43,246 | 5.4 | 39.0 | 140 | 1,235 | 0.90 (0.72 - 1.11) | 0.34 | 0.88 (0.71 - 1.09) | 0.28 |
| MDCR | 2,837 | 13,465 | 1.9 | 10.8 | 44 | 312 | 0.87 (0.55 - 1.34) | 0.54 | 0.83 (0.53 - 1.30) | 0.43 |
| Open Claims | 151,965 | 613,909 | 96.5 | 622.4 | 1,039 | 9,697 | 0.82 (0.75 - 0.89) | <0.01 | 0.84 (0.69 - 1.02) | 0.10 |
| OptumEHR | 6,867 | 40,661 | 1.7 | 21.6 | 34 | 425 | 1.42 (0.92 - 2.13) | 0.10 | 1.40 (0.92 - 2.13) | 0.14 |
| VA | 5,037 | 32,331 | 3.1 | 25.6 | 12 | 131 | 0.89 (0.37 - 1.95) | 0.79 | 1.00 (0.43 - 2.32) | 0.94 |
| GermanyDA | 3,829 | 831 | 2.8 | 0.7 | 18 | <5 | 3.51 (0.59 - 67.5) | 0.30 | 4.22 (0.39 - 45.4) | 0.24 |
| SIDIAP | 3,184 | 931 | 3.7 | 1.5 | 14 | 10 | 0.94 (0.25 - 3.88) | 0.93 | 1.30 (0.33 - 5.12) | 0.73 |
| Meta-analysis | 171,403 | 711,281 | 105.5 | 693.7 | 1,257 | 11,669 | 0.92 (0.76 - 1.11) | 0.39 | 0.92 (0.71 - 1.20) | 0.54 |

#### 7.6 DPP4I vs SU

##### 7.6.1 Abnormal weight gain

**Supplementary Table 306:** Exposures, outcomes and risk estimates for Abnormal weight gain in DPP4I (T) vs SU (C) new-user cohorts under two different propensity score (PS) adjustment across data sources. We report total patient exposure time in 1,000 patient-year (KYr) units, uncalibrated and calibrated hazard ratios (HRs) and their 95% confidence intervals (CIs). Greyed out entries did not pass study diagnostics and are included only for completeness.

| | Patients | | Exposure (KYr) | | Outcomes | | Uncalibrated | | Calibrated | |
|---|---|---|---|---|---|---|---|---|---|---|
| | T | C | T | C | T | C | HR (95% CI) | $p$ | HR (95% CI) | $p$ |
| **PS-stratification / on-treatment 2 TAR** | | | | | | | | | | |
| CCAE | 2,023 | 3,289 | 0.5 | 0.9 | <5 | <5 | 2.67 (0.44 - 20.6) | 0.32 | 2.85 (0.41 - 19.6) | 0.29 |
| OptumDOD | 26,503 | 64,803 | 18.8 | 61.2 | 103 | 369 | 0.80 (0.63 - 1.00) | 0.05 | 0.81 (0.64 - 1.02) | 0.08 |
| MDCR | 17,725 | 33,412 | 15.3 | 30.0 | 36 | 108 | 0.57 (0.38 - 0.84) | <0.01 | 0.57 (0.38 - 0.85) | <0.01 |
| MDCD | 1,168 | 2,416 | 0.7 | 1.4 | <5 | 7 | 0.64 (0.13 - 2.49) | 0.55 | 0.58 (0.13 - 2.53) | 0.47 |
| Open Claims | 334,949 | 690,159 | 257.7 | 699.8 | 436 | 1,429 | 0.72 (0.64 - 0.80) | <0.01 | 0.72 (0.61 - 0.85) | <0.01 |
| OptumEHR | 22,799 | 57,257 | 9.8 | 30.6 | 40 | 176 | 0.73 (0.50 - 1.04) | 0.10 | 0.77 (0.51 - 1.15) | 0.23 |
| VA | 15,017 | 108,943 | 12.9 | 111.9 | <10 | 154 | 0.37 (0.16 - 0.77) | 0.01 | 0.39 (0.18 - 0.86) | 0.02 |
| GermanyDA | 12,548 | 2,982 | 12.8 | 3.1 | 13 | 5 | 0.68 (0.21 - 2.53) | 0.55 | 0.79 (0.22 - 2.76) | 0.71 |
| SIDIAP | 26,193 | 8,059 | 54.9 | 19.1 | 88 | 37 | 0.95 (0.61 - 1.50) | 0.82 | 0.94 (0.60 - 1.47) | 0.79 |
| Meta-analysis | 428,169 | 853,690 | 356.6 | 840.7 | 703 | 2,119 | 0.73 (0.67 - 0.80) | <0.01 | 0.73 (0.64 - 0.84) | <0.01 |
| **PS-matching / on-treatment 2 TAR** | | | | | | | | | | |
| CCAE | 1,842 | 3,216 | 0.5 | 0.8 | <5 | <5 | 2.56 (0.24 - 56.1) | 0.50 | 2.57 (0.17 - 39.4) | 0.50 |
| OptumDOD | 25,675 | 64,733 | 18.2 | 61.2 | 99 | 369 | 0.92 (0.68 - 1.23) | 0.57 | 0.95 (0.70 - 1.27) | 0.73 |
| MDCR | 16,554 | 32,528 | 14.2 | 29.1 | 31 | 107 | 0.44 (0.25 - 0.76) | <0.01 | 0.43 (0.25 - 0.75) | <0.01 |
| MDCD | 1,105 | 2,399 | 0.6 | 1.4 | <5 | 7 | | | | |
| Open Claims | 328,595 | 690,156 | 251.2 | 699.8 | 413 | 1,429 | 0.58 (0.50 - 0.68) | <0.01 | 0.59 (0.50 - 0.69) | <0.01 |
| OptumEHR | 21,633 | 57,215 | 9.2 | 30.5 | 39 | 176 | 0.63 (0.38 - 1.00) | 0.06 | 0.65 (0.40 - 1.05) | 0.09 |
| VA | 13,986 | 78,621 | 12.3 | 76.2 | <10 | 105 | 0.27 (0.06 - 0.81) | 0.04 | 0.28 (0.08 - 1.01) | 0.05 |
| GermanyDA | 12,512 | 2,552 | 12.8 | 2.6 | 13 | <5 | 0.81 (0.16 - 6.26) | 0.83 | 0.83 (0.13 - 5.18) | 0.84 |
| SIDIAP | 26,068 | 6,113 | 54.8 | 13.9 | 88 | 30 | 0.94 (0.53 - 1.71) | 0.84 | 1.01 (0.52 - 1.96) | 0.90 |
| Meta-analysis | 432,511 | 929,366 | 359.8 | 910.8 | <680 | 2,216 | 0.66 (0.51 - 0.85) | <0.01 | 0.66 (0.51 - 0.87) | <0.01 |

##### 7.6.2 Abnormal weight loss

**Supplementary Table 307:** Exposures, outcomes and risk estimates for Abnormal weight loss in DPP4I (T) vs SU (C) new-user cohorts under two different propensity score (PS) adjustment across data sources. We report total patient exposure time in 1,000 patient-year (KYr) units, uncalibrated and calibrated hazard ratios (HRs) and their 95% confidence intervals (CIs). Greyed out entries did not pass study diagnostics and are included only for completeness.

| | Patients | | Exposure (KYr) | | Outcomes | | Uncalibrated | | Calibrated | |
|---|---|---|---|---|---|---|---|---|---|---|
| | T | C | T | C | T | C | HR (95% CI) | $p$ | HR (95% CI) | $p$ |
| **PS-stratification / on-treatment 2 TAR** | | | | | | | | | | |
| CCAE | 2,031 | 3,286 | 0.5 | 0.9 | 9 | 13 | 0.95 (0.37 - 2.35) | 0.92 | 1.01 (0.40 - 2.56) | 0.88 |
| OptumDOD | 25,478 | 62,249 | 18.0 | 58.8 | 381 | 1,026 | 1.13 (0.99 - 1.28) | 0.06 | 1.15 (1.01 - 1.30) | 0.05 |
| MDCR | 17,148 | 32,177 | 14.7 | 28.9 | 225 | 365 | 1.16 (0.97 - 1.38) | 0.09 | 1.17 (0.97 - 1.39) | 0.14 |
| MDCD | 1,115 | 2,298 | 0.6 | 1.4 | 17 | 36 | 0.99 (0.52 - 1.79) | 0.96 | 0.90 (0.48 - 1.66) | 0.73 |
| Open Claims | 326,168 | 673,081 | 250.0 | 683.0 | 2,397 | 4,904 | 1.25 (1.18 - 1.31) | <0.01 | 1.24 (1.09 - 1.42) | <0.01 |
| OptumEHR | 22,396 | 56,014 | 9.6 | 29.8 | 161 | 427 | 1.20 (0.99 - 1.46) | 0.07 | 1.26 (0.97 - 1.64) | 0.13 |
| VA | 14,599 | 105,915 | 12.5 | 109.0 | 138 | 777 | 1.35 (1.08 - 1.68) | <0.01 | 1.41 (1.13 - 1.76) | <0.01 |
| GermanyDA | 12,258 | 2,910 | 12.4 | 3.0 | 98 | 21 | 1.00 (0.60 - 1.74) | 0.99 | 1.16 (0.68 - 1.97) | 0.59 |
| SIDIAP | 26,053 | 8,065 | 54.5 | 19.2 | 254 | 53 | 0.91 (0.65 - 1.29) | 0.59 | 0.90 (0.64 - 1.27) | 0.57 |
| Meta-analysis | 432,647 | 840,080 | 360.6 | 824.9 | 3,542 | 6,845 | 1.18 (1.10 - 1.27) | <0.01 | 1.19 (1.05 - 1.34) | <0.01 |
| **PS-matching / on-treatment 2 TAR** | | | | | | | | | | |
| CCAE | 1,848 | 3,215 | 0.5 | 0.8 | 7 | 13 | 0.78 (0.26 - 2.11) | 0.64 | 0.78 (0.28 - 2.22) | 0.66 |
| OptumDOD | 24,685 | 62,182 | 17.4 | 58.7 | 364 | 1,026 | 1.09 (0.92 - 1.28) | 0.32 | 1.12 (0.95 - 1.33) | 0.20 |
| MDCR | 15,980 | 31,323 | 13.6 | 28.1 | 216 | 362 | 1.12 (0.88 - 1.42) | 0.36 | 1.08 (0.85 - 1.38) | 0.53 |
| MDCD | 1,066 | 2,280 | 0.6 | 1.4 | 17 | 36 | 0.86 (0.31 - 2.17) | 0.76 | 0.80 (0.30 - 2.11) | 0.66 |
| Open Claims | 319,989 | 673,077 | 243.9 | 683.0 | 2,308 | 4,904 | 1.17 (1.09 - 1.25) | <0.01 | 1.18 (1.08 - 1.29) | <0.01 |
| OptumEHR | 21,223 | 55,973 | 9.0 | 29.8 | 147 | 426 | 1.12 (0.87 - 1.44) | 0.37 | 1.17 (0.90 - 1.50) | 0.26 |
| VA | 13,594 | 76,432 | 11.9 | 74.2 | 132 | 540 | 1.24 (0.93 - 1.65) | 0.14 | 1.29 (0.97 - 1.73) | 0.09 |
| GermanyDA | 12,223 | 2,498 | 12.4 | 2.5 | 98 | 18 | 1.74 (0.84 - 4.08) | 0.17 | 1.76 (0.80 - 3.90) | 0.18 |
| SIDIAP | 25,925 | 6,116 | 54.5 | 13.9 | 254 | 46 | 0.91 (0.58 - 1.46) | 0.68 | 0.97 (0.55 - 1.71) | 0.88 |
| Meta-analysis | 436,533 | 913,096 | 363.7 | 892.4 | 3,543 | 7,371 | 1.15 (1.09 - 1.22) | <0.01 | 1.16 (1.06 - 1.28) | <0.01 |

##### 7.6.3 Acute pancreatitis

**Supplementary Table 308:** Exposures, outcomes and risk estimates for Acute pancreatitis in DPP4I (T) vs SU (C) new-user cohorts under two different propensity score (PS) adjustment across data sources. We report total patient exposure time in 1,000 patient-year (KYr) units, uncalibrated and calibrated hazard ratios (HRs) and their 95% confidence intervals (CIs). Greyed out entries did not pass study diagnostics and are included only for completeness.

| | Patients | | Exposure (KYr) | | Outcomes | | Uncalibrated | | Calibrated | |
|---|---|---|---|---|---|---|---|---|---|---|
| | T | C | T | C | T | C | HR (95% CI) | $p$ | HR (95% CI) | $p$ |
| **PS-stratification / on-treatment 2 TAR** | | | | | | | | | | |
| CCAE | 2,074 | 3,359 | 0.5 | 0.9 | <5 | <5 | 0.79 (0.11 - 4.14) | 0.80 | 0.84 (0.14 - 5.22) | 0.85 |
| OptumDOD | 26,990 | 65,495 | 19.3 | 62.1 | 64 | 230 | 0.86 (0.64 - 1.15) | 0.32 | 0.88 (0.66 - 1.18) | 0.39 |
| MDCR | 17,814 | 33,321 | 15.3 | 30.0 | 49 | 108 | 1.00 (0.69 - 1.42) | 1.00 | 1.00 (0.70 - 1.44) | 0.95 |
| MDCD | 1,190 | 2,425 | 0.7 | 1.5 | 6 | 7 | 1.60 (0.48 - 5.20) | 0.44 | 1.45 (0.44 - 4.77) | 0.54 |
| Open Claims | 340,386 | 696,991 | 261.7 | 706.5 | 598 | 1,512 | 1.02 (0.92 - 1.12) | 0.71 | 1.02 (0.87 - 1.19) | 0.83 |
| OptumEHR | 23,253 | 58,047 | 10.1 | 31.1 | 20 | 78 | 0.92 (0.53 - 1.53) | 0.76 | 0.96 (0.55 - 1.69) | 0.90 |
| VA | 15,177 | 109,510 | 13.0 | 112.6 | 15 | 156 | 0.95 (0.50 - 1.71) | 0.88 | 1.00 (0.54 - 1.85) | 0.96 |
| GermanyDA | 12,634 | 2,999 | 12.9 | 3.1 | 0 | 0 | | | | |
| SIDIAP | 26,418 | 8,085 | 55.4 | 19.3 | 26 | 6 | 1.17 (0.43 - 3.61) | 0.77 | 1.16 (0.40 - 3.35) | 0.79 |
| Meta-analysis | 434,861 | 861,939 | 361.8 | 848.9 | 757 | 1,934 | 1.00 (0.92 - 1.09) | 0.99 | 1.01 (0.88 - 1.15) | 0.90 |
| **PS-matching / on-treatment 2 TAR** | | | | | | | | | | |
| CCAE | 1,880 | 3,286 | 0.5 | 0.9 | <5 | <5 | 0.46 (0.02 - 3.67) | 0.55 | 0.46 (0.04 - 5.88) | 0.55 |
| OptumDOD | 26,101 | 65,429 | 18.5 | 62.0 | 60 | 230 | 0.86 (0.58 - 1.24) | 0.43 | 0.89 (0.61 - 1.30) | 0.54 |
| MDCR | 16,586 | 32,348 | 14.2 | 29.0 | 46 | 103 | 0.73 (0.43 - 1.19) | 0.22 | 0.70 (0.42 - 1.17) | 0.18 |
| MDCD | 1,122 | 2,406 | 0.7 | 1.4 | 5 | 6 | 1.23 (0.24 - 5.35) | 0.79 | 1.15 (0.24 - 5.40) | 0.86 |
| Open Claims | 333,698 | 696,989 | 254.9 | 706.5 | 574 | 1,512 | 1.03 (0.91 - 1.18) | 0.62 | 1.05 (0.91 - 1.21) | 0.55 |
| OptumEHR | 22,038 | 58,004 | 9.4 | 31.1 | 19 | 78 | 1.45 (0.79 - 2.62) | 0.22 | 1.51 (0.83 - 2.75) | 0.19 |
| VA | 14,102 | 79,293 | 12.4 | 76.9 | 14 | 108 | 1.10 (0.52 - 2.17) | 0.79 | 1.15 (0.56 - 2.35) | 0.71 |
| GermanyDA | 12,597 | 2,566 | 12.8 | 2.6 | 0 | 0 | | | | |
| SIDIAP | 26,291 | 6,137 | 55.3 | 14.0 | 26 | 6 | 1.20 (0.36 - 4.67) | 0.78 | 1.29 (0.35 - 4.83) | 0.71 |
| Meta-analysis | 438,816 | 938,200 | 364.7 | 919.6 | 739 | 2,037 | 1.01 (0.90 - 1.14) | 0.82 | 1.02 (0.89 - 1.18) | 0.74 |

##### 7.6.4 All-cause mortality

**Supplementary Table 309:** Exposures, outcomes and risk estimates for All-cause mortality in DPP4I (T) vs SU (C) new-user cohorts under two different propensity score (PS) adjustment across data sources. We report total patient exposure time in 1,000 patient-year (KYr) units, uncalibrated and calibrated hazard ratios (HRs) and their 95% confidence intervals (CIs). Greyed out entries did not pass study diagnostics and are included only for completeness.

| | Patients | | Exposure (KYr) | | Outcomes | | Uncalibrated | | Calibrated | |
|---|---|---|---|---|---|---|---|---|---|---|
| | T | C | T | C | T | C | HR (95% CI) | $p$ | HR (95% CI) | $p$ |
| **PS-stratification / on-treatment 2 TAR** | | | | | | | | | | |
| CCAE | 2,089 | 3,385 | 0.5 | 0.9 | 0 | <5 | | | | |
| OptumDOD | 27,235 | 66,291 | 19.4 | 62.9 | 227 | 960 | 0.82 (0.70 - 0.95) | <0.01 | 0.83 (0.71 - 0.97) | 0.03 |
| MDCR | 17,961 | 33,739 | 15.5 | 30.4 | 43 | 117 | 0.95 (0.65 - 1.36) | 0.80 | 0.96 (0.66 - 1.38) | 0.82 |
| MDCD | 1,210 | 2,481 | 0.7 | 1.5 | 13 | 10 | 2.87 (1.20 - 7.13) | 0.02 | 2.61 (1.07 - 6.38) | 0.04 |
| Open Claims | 344,611 | 707,599 | 265.1 | 717.4 | <5 | 11 | 0.44 (0.07 - 1.72) | 0.32 | 0.44 (0.09 - 2.25) | 0.32 |
| OptumEHR | 23,345 | 58,379 | 10.1 | 31.3 | 113 | 429 | 0.95 (0.76 - 1.18) | 0.66 | 1.00 (0.75 - 1.32) | 0.90 |
| VA | 15,312 | 110,592 | 13.1 | 113.7 | 196 | 2,011 | 0.82 (0.69 - 0.98) | 0.03 | 0.86 (0.72 - 1.02) | 0.11 |
| GermanyDA | 12,634 | 2,999 | 12.9 | 3.1 | 10 | <5 | 2.28 (0.33 - 47.4) | 0.52 | 2.64 (0.22 - 31.8) | 0.45 |
| SIDIAP | 26,465 | 8,098 | 55.5 | 19.3 | 1,586 | 361 | 1.14 (1.00 - 1.31) | 0.05 | 1.13 (0.98 - 1.31) | 0.19 |
| Meta-analysis | 98,292 | 172,101 | 101.3 | 145.5 | 1,987 | 1,900 | 1.00 (0.84 - 1.19) | 0.99 | 1.00 (0.82 - 1.23) | 0.94 |
| **PS-matching / on-treatment 2 TAR** | | | | | | | | | | |
| CCAE | 1,896 | 3,312 | 0.5 | 0.9 | 0 | <5 | | | | |
| OptumDOD | 26,347 | 66,220 | 18.7 | 62.9 | 215 | 957 | 0.84 (0.69 - 1.03) | 0.09 | 0.87 (0.71 - 1.07) | 0.19 |
| MDCR | 16,748 | 32,853 | 14.3 | 29.5 | 42 | 113 | 1.24 (0.76 - 2.00) | 0.39 | 1.20 (0.74 - 1.96) | 0.47 |
| MDCD | 1,140 | 2,462 | 0.7 | 1.5 | 13 | 10 | 1.99 (0.36 - 9.28) | 0.40 | 1.85 (0.37 - 9.36) | 0.46 |
| Open Claims | 337,948 | 707,595 | 258.3 | 717.4 | <5 | 11 | 1.19 (0.14 - 10.1) | 0.88 | 1.20 (0.14 - 10.2) | 0.87 |
| OptumEHR | 22,131 | 58,336 | 9.4 | 31.2 | 111 | 429 | 0.99 (0.75 - 1.29) | 0.92 | 1.02 (0.78 - 1.35) | 0.85 |
| VA | 14,229 | 79,981 | 12.5 | 77.6 | 184 | 1,351 | 0.81 (0.65 - 1.00) | 0.06 | 0.85 (0.68 - 1.05) | 0.16 |
| GermanyDA | 12,597 | 2,566 | 12.8 | 2.6 | 10 | 0 | | | | |
| SIDIAP | 26,339 | 6,148 | 55.4 | 14.1 | 1,579 | 259 | 1.15 (0.95 - 1.38) | 0.15 | 1.23 (0.85 - 1.78) | 0.35 |
| Meta-analysis | 108,303 | 249,089 | 111.1 | 216.9 | 2,149 | 3,142 | 0.97 (0.83 - 1.13) | 0.70 | 0.98 (0.82 - 1.17) | 0.82 |

##### 7.6.5 Bone fracture

**Supplementary Table 310:** Exposures, outcomes and risk estimates for Bone fracture in DPP4I (T) vs SU (C) new-user cohorts under two different propensity score (PS) adjustment across data sources. We report total patient exposure time in 1,000 patient-year (KYr) units, uncalibrated and calibrated hazard ratios (HRs) and their 95% confidence intervals (CIs). Greyed out entries did not pass study diagnostics and are included only for completeness.

| | Patients | | Exposure (KYr) | | Outcomes | | Uncalibrated | | Calibrated | |
|---|---|---|---|---|---|---|---|---|---|---|
| | T | C | T | C | T | C | HR (95% CI) | $p$ | HR (95% CI) | $p$ |
| **PS-stratification / on-treatment 2 TAR** | | | | | | | | | | |
| CCAE | 1,879 | 3,098 | 0.5 | 0.8 | 7 | 20 | 0.59 (0.23 - 1.36) | 0.25 | 0.63 (0.25 - 1.54) | 0.34 |
| OptumDOD | 24,067 | 59,398 | 16.9 | 55.3 | 582 | 1,910 | 0.97 (0.87 - 1.06) | 0.48 | 0.98 (0.89 - 1.09) | 0.76 |
| MDCR | 15,701 | 29,545 | 13.3 | 26.1 | 460 | 928 | 0.95 (0.85 - 1.07) | 0.41 | 0.96 (0.84 - 1.08) | 0.56 |
| MDCD | 988 | 2,135 | 0.6 | 1.3 | 20 | 59 | 0.80 (0.46 - 1.32) | 0.40 | 0.72 (0.43 - 1.23) | 0.24 |
| Open Claims | 291,512 | 605,340 | 222.1 | 609.0 | 5,163 | 13,744 | 0.97 (0.94 - 1.00) | 0.06 | 0.97 (0.85 - 1.10) | 0.63 |
| OptumEHR | 21,425 | 53,609 | 9.1 | 28.1 | 217 | 790 | 0.88 (0.75 - 1.03) | 0.13 | 0.92 (0.73 - 1.18) | 0.58 |
| VA | 13,821 | 101,522 | 11.9 | 104.1 | 110 | 1,068 | 1.06 (0.83 - 1.33) | 0.64 | 1.10 (0.87 - 1.40) | 0.43 |
| GermanyDA | 10,829 | 2,636 | 10.8 | 2.7 | 286 | 71 | 0.95 (0.71 - 1.29) | 0.75 | 1.10 (0.82 - 1.49) | 0.53 |
| SIDIAP | 23,069 | 7,470 | 47.7 | 17.4 | 924 | 322 | 1.10 (0.95 - 1.28) | 0.20 | 1.09 (0.93 - 1.28) | 0.39 |
| Meta-analysis | 389,470 | 763,231 | 321.1 | 740.5 | 7,659 | 17,844 | 0.97 (0.94 - 1.00) | 0.03 | 0.97 (0.88 - 1.08) | 0.61 |
| **PS-matching / on-treatment 2 TAR** | | | | | | | | | | |
| CCAE | 1,706 | 3,011 | 0.5 | 0.8 | 7 | 20 | 0.65 (0.22 - 1.65) | 0.39 | 0.65 (0.24 - 1.76) | 0.41 |
| OptumDOD | 23,325 | 59,331 | 16.3 | 55.2 | 559 | 1,905 | 0.92 (0.80 - 1.05) | 0.22 | 0.95 (0.83 - 1.09) | 0.50 |
| MDCR | 14,674 | 28,642 | 12.4 | 25.2 | 421 | 913 | 0.93 (0.79 - 1.10) | 0.41 | 0.90 (0.76 - 1.07) | 0.28 |
| MDCD | 945 | 2,119 | 0.5 | 1.2 | 19 | 59 | 1.09 (0.53 - 2.18) | 0.80 | 1.02 (0.50 - 2.06) | 0.85 |
| Open Claims | 285,984 | 605,337 | 216.6 | 609.0 | 5,004 | 13,744 | 0.98 (0.94 - 1.03) | 0.50 | 1.00 (0.93 - 1.07) | 0.87 |
| OptumEHR | 20,324 | 53,563 | 8.6 | 28.0 | 206 | 788 | 0.90 (0.73 - 1.11) | 0.32 | 0.94 (0.76 - 1.15) | 0.57 |
| VA | 12,914 | 73,134 | 11.3 | 70.9 | 106 | 697 | 1.07 (0.79 - 1.43) | 0.66 | 1.11 (0.83 - 1.50) | 0.50 |
| GermanyDA | 10,788 | 2,256 | 10.8 | 2.3 | 285 | 60 | 0.99 (0.68 - 1.48) | 0.98 | 1.01 (0.68 - 1.50) | 0.72 |
| SIDIAP | 22,974 | 5,624 | 47.7 | 12.5 | 924 | 230 | 1.14 (0.93 - 1.40) | 0.23 | 1.22 (0.83 - 1.78) | 0.38 |
| Meta-analysis | 393,634 | 833,017 | 324.6 | 805.1 | 7,531 | 18,416 | 0.98 (0.94 - 1.02) | 0.28 | 0.99 (0.91 - 1.08) | 0.79 |

##### 7.6.6 Diabetic ketoacidosis

**Supplementary Table 311:** Exposures, outcomes and risk estimates for Diabetic ketoacidosis in DPP4I (T) vs SU (C) new-user cohorts under two different propensity score (PS) adjustment across data sources. We report total patient exposure time in 1,000 patient-year (KYr) units, uncalibrated and calibrated hazard ratios (HRs) and their 95% confidence intervals (CIs). Greyed out entries did not pass study diagnostics and are included only for completeness.

| | Patients | | Exposure (KYr) | | Outcomes | | Uncalibrated | | Calibrated | |
|---|---|---|---|---|---|---|---|---|---|---|
| | T | C | T | C | T | C | HR (95% CI) | $p$ | HR (95% CI) | $p$ |
| **PS-stratification / on-treatment 2 TAR** | | | | | | | | | | |
| CCAE | 2,088 | 3,383 | 0.5 | 0.9 | 0 | 0 | | | | |
| OptumDOD | 27,197 | 66,189 | 19.4 | 62.8 | 29 | 90 | 1.00 (0.63 - 1.53) | 0.99 | 1.01 (0.65 - 1.58) | 0.95 |
| MDCR | 17,928 | 33,703 | 15.4 | 30.4 | 14 | 35 | 0.91 (0.46 - 1.71) | 0.79 | 0.92 (0.48 - 1.76) | 0.80 |
| MDCD | 1,207 | 2,474 | 0.7 | 1.5 | 0 | 5 | | | | |
| Open Claims | 344,148 | 706,389 | 264.7 | 716.4 | 190 | 482 | 1.02 (0.86 - 1.22) | 0.78 | 1.02 (0.83 - 1.27) | 0.83 |
| OptumEHR | 23,335 | 58,344 | 10.1 | 31.3 | 8 | 25 | 1.03 (0.42 - 2.30) | 0.94 | 1.08 (0.45 - 2.58) | 0.87 |
| VA | 15,307 | 110,551 | 13.1 | 113.7 | <10 | 25 | 0.37 (0.02 - 2.45) | 0.42 | 0.38 (0.03 - 4.43) | 0.44 |
| GermanyDA | 12,634 | 2,999 | 12.9 | 3.1 | 0 | 0 | | | | |
| SIDIAP | 26,465 | 8,098 | 55.5 | 19.3 | 0 | <5 | | | | |
| Meta-analysis | 412,608 | 864,625 | 309.7 | 840.9 | 241 | 632 | 1.01 (0.87 - 1.18) | 0.85 | 1.02 (0.85 - 1.22) | 0.82 |
| **PS-matching / on-treatment 2 TAR** | | | | | | | | | | |
| CCAE | 1,895 | 3,310 | 0.5 | 0.9 | 0 | 0 | | | | |
| OptumDOD | 26,305 | 66,120 | 18.7 | 62.8 | 27 | 90 | 1.06 (0.59 - 1.86) | 0.84 | 1.10 (0.61 - 1.96) | 0.76 |
| MDCR | 16,716 | 32,819 | 14.3 | 29.5 | 13 | 35 | 0.62 (0.24 - 1.41) | 0.29 | 0.60 (0.25 - 1.45) | 0.26 |
| MDCD | 1,137 | 2,455 | 0.7 | 1.5 | 0 | 5 | | | | |
| Open Claims | 337,485 | 706,385 | 257.9 | 716.4 | 187 | 482 | 1.06 (0.84 - 1.34) | 0.61 | 1.07 (0.85 - 1.36) | 0.57 |
| OptumEHR | 22,121 | 58,301 | 9.5 | 31.3 | 7 | 25 | 0.43 (0.10 - 1.34) | 0.21 | 0.44 (0.12 - 1.65) | 0.23 |
| VA | 14,226 | 79,950 | 12.5 | 77.6 | <10 | 18 | | | | |
| GermanyDA | 12,597 | 2,566 | 12.8 | 2.6 | 0 | 0 | | | | |
| SIDIAP | 26,339 | 6,148 | 55.4 | 14.1 | 0 | <5 | | | | |
| Meta-analysis | 402,627 | 863,625 | 300.3 | 839.9 | 234 | 632 | 1.01 (0.82 - 1.24) | 0.94 | 1.02 (0.82 - 1.27) | 0.87 |

##### 7.6.7 Diarrhea

**Supplementary Table 312:** Exposures, outcomes and risk estimates for Diarrhea in DPP4I (T) vs SU (C) new-user cohorts under two different propensity score (PS) adjustment across data sources. We report total patient exposure time in 1,000 patient-year (KYr) units, uncalibrated and calibrated hazard ratios (HRs) and their 95% confidence intervals (CIs). Greyed out entries did not pass study diagnostics and are included only for completeness.

| | Patients | | Exposure (KYr) | | Outcomes | | Uncalibrated | | Calibrated | |
|---|---|---|---|---|---|---|---|---|---|---|
| | T | C | T | C | T | C | HR (95% CI) | $p$ | HR (95% CI) | $p$ |
| **PS-stratification / on-treatment 2 TAR** | | | | | | | | | | |
| CCAE | 1,822 | 2,998 | 0.5 | 0.8 | 17 | 34 | 0.77 (0.41 - 1.40) | 0.40 | 0.82 (0.44 - 1.52) | 0.57 |
| OptumDOD | 22,561 | 55,730 | 15.7 | 51.3 | 694 | 2,327 | 0.89 (0.81 - 0.97) | <0.01 | 0.91 (0.82 - 0.99) | 0.06 |
| MDCR | 15,423 | 29,180 | 13.0 | 25.7 | 466 | 926 | 0.93 (0.83 - 1.05) | 0.24 | 0.93 (0.83 - 1.06) | 0.40 |
| MDCD | 976 | 2,046 | 0.6 | 1.2 | 29 | 60 | 0.87 (0.54 - 1.38) | 0.57 | 0.79 (0.50 - 1.27) | 0.35 |
| Open Claims | 292,928 | 610,026 | 223.0 | 613.2 | 4,961 | 12,708 | 0.97 (0.94 - 1.00) | 0.09 | 0.97 (0.85 - 1.10) | 0.65 |
| OptumEHR | 20,428 | 51,411 | 8.6 | 26.4 | 310 | 1,090 | 0.91 (0.79 - 1.04) | 0.16 | 0.95 (0.76 - 1.19) | 0.71 |
| VA | 13,322 | 98,467 | 11.4 | 99.9 | 214 | 1,725 | 1.10 (0.92 - 1.29) | 0.28 | 1.14 (0.96 - 1.36) | 0.15 |
| GermanyDA | 12,474 | 2,955 | 12.7 | 3.0 | 27 | 7 | 1.07 (0.45 - 2.88) | 0.88 | 1.24 (0.49 - 3.14) | 0.65 |
| SIDIAP | 25,045 | 7,879 | 52.2 | 18.6 | 583 | 116 | 1.05 (0.83 - 1.33) | 0.69 | 1.04 (0.82 - 1.32) | 0.79 |
| Meta-analysis | 391,657 | 762,225 | 326.3 | 740.2 | 7,087 | 17,268 | 0.94 (0.90 - 0.99) | 0.01 | 0.95 (0.85 - 1.06) | 0.34 |
| **PS-matching / on-treatment 2 TAR** | | | | | | | | | | |
| CCAE | 1,665 | 2,933 | 0.4 | 0.8 | 15 | 34 | 0.68 (0.29 - 1.45) | 0.34 | 0.68 (0.30 - 1.52) | 0.37 |
| OptumDOD | 21,835 | 55,648 | 15.2 | 51.2 | 664 | 2,326 | 0.89 (0.79 - 1.00) | 0.05 | 0.92 (0.82 - 1.03) | 0.20 |
| MDCR | 14,380 | 28,339 | 12.1 | 24.9 | 425 | 903 | 0.87 (0.73 - 1.02) | 0.09 | 0.84 (0.71 - 0.99) | 0.06 |
| MDCD | 938 | 2,030 | 0.5 | 1.2 | 29 | 57 | 0.74 (0.38 - 1.37) | 0.35 | 0.69 (0.36 - 1.31) | 0.27 |
| Open Claims | 287,496 | 610,023 | 217.6 | 613.2 | 4,796 | 12,708 | 0.95 (0.90 - 0.99) | 0.02 | 0.96 (0.89 - 1.03) | 0.33 |
| OptumEHR | 19,414 | 51,374 | 8.1 | 26.3 | 290 | 1,089 | 0.89 (0.74 - 1.05) | 0.17 | 0.92 (0.77 - 1.10) | 0.41 |
| VA | 12,415 | 70,431 | 10.9 | 67.4 | 203 | 1,188 | 1.12 (0.90 - 1.39) | 0.29 | 1.17 (0.94 - 1.45) | 0.18 |
| GermanyDA | 12,439 | 2,528 | 12.6 | 2.6 | 27 | 7 | 0.99 (0.33 - 3.74) | 0.99 | 1.01 (0.30 - 3.43) | 0.90 |
| SIDIAP | 24,934 | 5,941 | 52.1 | 13.5 | 582 | 93 | 1.00 (0.74 - 1.35) | 0.98 | 1.07 (0.69 - 1.65) | 0.79 |
| Meta-analysis | 395,516 | 829,247 | 329.5 | 801.0 | 7,031 | 18,405 | 0.94 (0.90 - 0.97) | <0.01 | 0.95 (0.87 - 1.03) | 0.21 |

##### 7.6.8 Genitourinary infection

**Supplementary Table 313:** Exposures, outcomes and risk estimates for Genitourinary infection in DPP4I (T) vs SU (C) new-user cohorts under two different propensity score (PS) adjustment across data sources. We report total patient exposure time in 1,000 patient-year (KYr) units, uncalibrated and calibrated hazard ratios (HRs) and their 95% confidence intervals (CIs). Greyed out entries did not pass study diagnostics and are included only for completeness.

| | Patients | | Exposure (KYr) | | Outcomes | | Uncalibrated | | Calibrated | |
|---|---|---|---|---|---|---|---|---|---|---|
| | T | C | T | C | T | C | HR (95% CI) | $p$ | HR (95% CI) | $p$ |
| **PS-stratification / on-treatment 2 TAR** | | | | | | | | | | |
| CCAE | 1,613 | 2,697 | 0.4 | 0.7 | 18 | 33 | 0.97 (0.52 - 1.74) | 0.91 | 1.03 (0.56 - 1.89) | 0.82 |
| OptumDOD | 19,139 | 48,391 | 13.1 | 43.6 | 1,102 | 3,590 | 0.93 (0.87 - 1.00) | 0.05 | 0.95 (0.88 - 1.02) | 0.24 |
| MDCR | 13,747 | 26,273 | 11.5 | 23.1 | 728 | 1,447 | 1.00 (0.91 - 1.10) | 0.96 | 1.00 (0.90 - 1.11) | 0.86 |
| MDCD | 798 | 1,741 | 0.4 | 1.0 | 38 | 90 | 0.99 (0.66 - 1.47) | 0.98 | 0.90 (0.61 - 1.35) | 0.63 |
| Open Claims | 247,187 | 528,233 | 185.9 | 530.3 | 8,846 | 21,946 | 1.02 (1.00 - 1.05) | 0.08 | 1.02 (0.90 - 1.16) | 0.74 |
| OptumEHR | 18,902 | 48,235 | 7.8 | 24.6 | 497 | 1,537 | 1.01 (0.90 - 1.12) | 0.88 | 1.06 (0.86 - 1.30) | 0.66 |
| VA | 13,255 | 96,687 | 11.4 | 98.2 | 185 | 1,926 | 0.82 (0.69 - 0.98) | 0.03 | 0.86 (0.72 - 1.03) | 0.11 |
| GermanyDA | 9,479 | 2,314 | 9.2 | 2.2 | 375 | 90 | 1.05 (0.81 - 1.37) | 0.72 | 1.22 (0.93 - 1.58) | 0.17 |
| SIDIAP | 20,527 | 7,170 | 41.9 | 16.1 | 1,841 | 570 | 1.02 (0.91 - 1.14) | 0.72 | 1.01 (0.89 - 1.14) | 0.76 |
| Meta-analysis | 333,273 | 667,821 | 270.3 | 641.6 | 13,450 | 29,327 | 1.00 (0.96 - 1.04) | 0.93 | 1.00 (0.91 - 1.11) | 0.89 |
| **PS-matching / on-treatment 2 TAR** | | | | | | | | | | |
| CCAE | 1,466 | 2,615 | 0.4 | 0.7 | 18 | 32 | 0.94 (0.47 - 1.81) | 0.85 | 0.94 (0.47 - 1.86) | 0.78 |
| OptumDOD | 18,599 | 48,346 | 12.7 | 43.5 | 1,059 | 3,588 | 0.96 (0.88 - 1.05) | 0.41 | 0.99 (0.90 - 1.09) | 0.82 |
| MDCR | 12,924 | 25,413 | 10.7 | 22.3 | 682 | 1,410 | 0.98 (0.86 - 1.11) | 0.74 | 0.95 (0.83 - 1.08) | 0.49 |
| MDCD | 766 | 1,729 | 0.4 | 1.0 | 36 | 87 | 0.92 (0.51 - 1.61) | 0.77 | 0.85 (0.48 - 1.53) | 0.61 |
| Open Claims | 243,347 | 528,224 | 182.1 | 530.3 | 8,563 | 21,946 | 1.00 (0.96 - 1.03) | 0.90 | 1.01 (0.95 - 1.08) | 0.81 |
| OptumEHR | 18,003 | 48,199 | 7.4 | 24.5 | 463 | 1,534 | 1.03 (0.90 - 1.18) | 0.66 | 1.07 (0.93 - 1.23) | 0.41 |
| VA | 12,386 | 69,459 | 10.9 | 66.7 | 174 | 1,266 | 0.93 (0.74 - 1.16) | 0.52 | 0.97 (0.77 - 1.21) | 0.78 |
| GermanyDA | 9,452 | 1,968 | 9.2 | 1.9 | 373 | 75 | 1.06 (0.76 - 1.48) | 0.75 | 1.07 (0.77 - 1.50) | 0.63 |
| SIDIAP | 20,451 | 5,348 | 41.9 | 11.5 | 1,839 | 431 | 1.00 (0.86 - 1.16) | 0.96 | 1.07 (0.75 - 1.52) | 0.75 |
| Meta-analysis | 337,394 | 731,301 | 275.6 | 702.4 | 13,207 | 30,369 | 0.99 (0.96 - 1.02) | 0.65 | 1.00 (0.92 - 1.09) | 0.87 |

##### 7.6.9 Hyperkalemia

**Supplementary Table 314:** Exposures, outcomes and risk estimates for Hyperkalemia in DPP4I (T) vs SU (C) new-user cohorts under two different propensity score (PS) adjustment across data sources. We report total patient exposure time in 1,000 patient-year (KYr) units, uncalibrated and calibrated hazard ratios (HRs) and their 95% confidence intervals (CIs). Greyed out entries did not pass study diagnostics and are included only for completeness.

| | Patients | | Exposure (KYr) | | Outcomes | | Uncalibrated | | Calibrated | |
|---|---|---|---|---|---|---|---|---|---|---|
| | T | C | T | C | T | C | HR (95% CI) | $p$ | HR (95% CI) | $p$ |
| **PS-stratification / on-treatment 2 TAR** | | | | | | | | | | |
| CCAE | 2,034 | 3,301 | 0.5 | 0.9 | 6 | 14 | 0.88 (0.30 - 2.25) | 0.80 | 0.93 (0.34 - 2.56) | 0.87 |
| OptumDOD | 25,334 | 61,820 | 17.7 | 57.3 | 586 | 1,820 | 1.00 (0.91 - 1.10) | 0.99 | 1.02 (0.92 - 1.13) | 0.77 |
| MDCR | 17,425 | 32,657 | 14.9 | 29.1 | 238 | 498 | 0.96 (0.81 - 1.12) | 0.59 | 0.96 (0.81 - 1.14) | 0.67 |
| MDCD | 1,112 | 2,305 | 0.6 | 1.4 | 19 | 43 | 0.97 (0.54 - 1.70) | 0.93 | 0.89 (0.50 - 1.57) | 0.68 |
| Open Claims | 331,544 | 682,393 | 253.0 | 686.3 | 3,158 | 7,992 | 1.05 (1.00 - 1.09) | 0.04 | 1.04 (0.92 - 1.19) | 0.52 |
| OptumEHR | 22,494 | 56,312 | 9.6 | 29.7 | 231 | 600 | 1.30 (1.10 - 1.53) | <0.01 | 1.37 (1.07 - 1.74) | 0.03 |
| VA | 14,128 | 101,963 | 11.9 | 102.9 | 173 | 1,933 | 0.90 (0.75 - 1.07) | 0.25 | 0.94 (0.78 - 1.13) | 0.51 |
| GermanyDA | 11,553 | 2,842 | 11.6 | 2.9 | 221 | 48 | 0.98 (0.69 - 1.42) | 0.92 | 1.14 (0.79 - 1.63) | 0.50 |
| SIDIAP | 24,462 | 7,741 | 50.5 | 18.2 | 702 | 240 | 1.00 (0.84 - 1.20) | 0.99 | 0.99 (0.82 - 1.19) | 0.80 |
| Meta-analysis | 435,958 | 849,371 | 358.5 | 825.8 | 5,161 | 11,255 | 1.04 (0.97 - 1.12) | 0.26 | 1.05 (0.93 - 1.18) | 0.44 |
| **PS-matching / on-treatment 2 TAR** | | | | | | | | | | |
| CCAE | 1,846 | 3,228 | 0.5 | 0.8 | 6 | 14 | 0.85 (0.26 - 2.40) | 0.78 | 0.86 (0.28 - 2.59) | 0.79 |
| OptumDOD | 24,539 | 61,759 | 17.1 | 57.3 | 568 | 1,817 | 1.00 (0.88 - 1.14) | 0.99 | 1.03 (0.91 - 1.18) | 0.65 |
| MDCR | 16,228 | 31,625 | 13.8 | 28.2 | 225 | 487 | 0.94 (0.75 - 1.18) | 0.62 | 0.92 (0.73 - 1.15) | 0.47 |
| MDCD | 1,051 | 2,289 | 0.6 | 1.4 | 19 | 43 | 0.51 (0.20 - 1.17) | 0.13 | 0.47 (0.19 - 1.15) | 0.11 |
| Open Claims | 325,065 | 682,389 | 246.5 | 686.3 | 3,074 | 7,992 | 1.01 (0.95 - 1.07) | 0.68 | 1.02 (0.95 - 1.11) | 0.62 |
| OptumEHR | 21,328 | 56,273 | 9.0 | 29.6 | 213 | 598 | 1.28 (1.02 - 1.59) | 0.03 | 1.32 (1.06 - 1.66) | 0.02 |
| VA | 13,147 | 73,463 | 11.3 | 70.2 | 160 | 1,228 | 1.02 (0.81 - 1.28) | 0.87 | 1.06 (0.84 - 1.34) | 0.63 |
| GermanyDA | 11,527 | 2,429 | 11.6 | 2.4 | 220 | 38 | 1.23 (0.75 - 2.12) | 0.42 | 1.25 (0.75 - 2.11) | 0.43 |
| SIDIAP | 24,353 | 5,873 | 50.4 | 13.3 | 698 | 176 | 1.05 (0.83 - 1.34) | 0.67 | 1.13 (0.76 - 1.68) | 0.61 |
| Meta-analysis | 439,084 | 919,328 | 360.8 | 889.5 | 5,183 | 12,393 | 1.02 (0.97 - 1.07) | 0.42 | 1.03 (0.94 - 1.13) | 0.52 |

##### 7.6.10 Hypoglycemia

**Supplementary Table 315:** Exposures, outcomes and risk estimates for Hypoglycemia in DPP4I (T) vs SU (C) new-user cohorts under two different propensity score (PS) adjustment across data sources. We report total patient exposure time in 1,000 patient-year (KYr) units, uncalibrated and calibrated hazard ratios (HRs) and their 95% confidence intervals (CIs). Greyed out entries did not pass study diagnostics and are included only for completeness.

| | Patients | | Exposure (KYr) | | Outcomes | | Uncalibrated | | Calibrated | |
|---|---|---|---|---|---|---|---|---|---|---|
| | T | C | T | C | T | C | HR (95% CI) | $p$ | HR (95% CI) | $p$ |
| **PS-stratification / on-treatment 2 TAR** | | | | | | | | | | |
| CCAE | 2,051 | 3,341 | 0.5 | 0.9 | <5 | 19 | 0.14 (0.02 - 0.52) | 0.02 | 0.15 (0.03 - 0.74) | 0.02 |
| OptumDOD | 26,793 | 65,348 | 19.1 | 61.4 | 96 | 1,633 | 0.17 (0.13 - 0.20) | <0.01 | 0.17 (0.14 - 0.21) | <0.01 |
| MDCR | 17,761 | 33,426 | 15.3 | 29.9 | 48 | 570 | 0.16 (0.12 - 0.21) | <0.01 | 0.16 (0.12 - 0.22) | <0.01 |
| MDCD | 1,174 | 2,405 | 0.7 | 1.4 | 6 | 50 | 0.22 (0.08 - 0.49) | <0.01 | 0.20 (0.08 - 0.49) | <0.01 |
| Open Claims | 338,626 | 696,050 | 260.6 | 703.5 | 563 | 8,151 | 0.16 (0.15 - 0.18) | <0.01 | 0.16 (0.14 - 0.19) | <0.01 |
| OptumEHR | 23,136 | 57,847 | 10.0 | 30.8 | 47 | 581 | 0.23 (0.17 - 0.31) | <0.01 | 0.24 (0.17 - 0.35) | <0.01 |
| VA | 15,174 | 109,673 | 13.1 | 112.4 | 18 | 883 | 0.17 (0.10 - 0.27) | <0.01 | 0.18 (0.11 - 0.29) | <0.01 |
| GermanyDA | 12,542 | 2,979 | 12.8 | 3.1 | 23 | 29 | 0.26 (0.13 - 0.49) | <0.01 | 0.30 (0.16 - 0.57) | <0.01 |
| SIDIAP | 26,423 | 8,087 | 55.4 | 19.2 | 18 | 85 | 0.07 (0.04 - 0.12) | <0.01 | 0.07 (0.04 - 0.12) | <0.01 |
| Meta-analysis | 448,506 | 869,483 | 374.4 | 850.2 | <806 | 11,118 | 0.17 (0.14 - 0.21) | <0.01 | 0.17 (0.13 - 0.21) | <0.01 |
| **PS-matching / on-treatment 2 TAR** | | | | | | | | | | |
| CCAE | 1,868 | 3,269 | 0.5 | 0.9 | <5 | 19 | 0.23 (0.04 - 0.86) | 0.07 | 0.23 (0.05 - 1.14) | 0.08 |
| OptumDOD | 25,923 | 65,278 | 18.4 | 61.3 | 88 | 1,631 | 0.15 (0.11 - 0.19) | <0.01 | 0.15 (0.12 - 0.20) | <0.01 |
| MDCR | 16,573 | 32,541 | 14.2 | 29.1 | 44 | 560 | 0.13 (0.08 - 0.19) | <0.01 | 0.12 (0.08 - 0.19) | <0.01 |
| MDCD | 1,104 | 2,386 | 0.6 | 1.4 | 6 | 50 | 0.16 (0.04 - 0.44) | <0.01 | 0.15 (0.04 - 0.50) | <0.01 |
| Open Claims | 332,096 | 696,046 | 254.0 | 703.5 | 544 | 8,150 | 0.14 (0.13 - 0.16) | <0.01 | 0.15 (0.13 - 0.17) | <0.01 |
| OptumEHR | 21,940 | 57,804 | 9.4 | 30.7 | 45 | 580 | 0.24 (0.16 - 0.34) | <0.01 | 0.25 (0.17 - 0.36) | <0.01 |
| VA | 14,102 | 79,115 | 12.4 | 76.5 | 17 | 609 | 0.17 (0.09 - 0.29) | <0.01 | 0.18 (0.10 - 0.32) | <0.01 |
| GermanyDA | 12,506 | 2,544 | 12.7 | 2.6 | 23 | 22 | 0.44 (0.18 - 1.07) | 0.07 | 0.45 (0.18 - 1.08) | 0.08 |
| SIDIAP | 26,297 | 6,142 | 55.3 | 14.0 | 18 | 65 | 0.05 (0.02 - 0.12) | <0.01 | 0.05 (0.02 - 0.14) | <0.01 |
| Meta-analysis | 452,409 | 945,125 | 377.5 | 920.1 | <790 | 11,686 | 0.16 (0.12 - 0.21) | <0.01 | 0.16 (0.12 - 0.21) | <0.01 |

##### 7.6.11 Hypotension

**Supplementary Table 316:** Exposures, outcomes and risk estimates for Hypotension in DPP4I (T) vs SU (C) new-user cohorts under two different propensity score (PS) adjustment across data sources. We report total patient exposure time in 1,000 patient-year (KYr) units, uncalibrated and calibrated hazard ratios (HRs) and their 95% confidence intervals (CIs). Greyed out entries did not pass study diagnostics and are included only for completeness.

| | Patients | | Exposure (KYr) | | Outcomes | | Uncalibrated | | Calibrated | |
|---|---|---|---|---|---|---|---|---|---|---|
| | T | C | T | C | T | C | HR (95% CI) | $p$ | HR (95% CI) | $p$ |
| **PS-stratification / on-treatment 2 TAR** | | | | | | | | | | |
| CCAE | 2,020 | 3,295 | 0.5 | 0.9 | 10 | 10 | 1.86 (0.73 - 4.77) | 0.20 | 1.98 (0.77 - 5.10) | 0.17 |
| OptumDOD | 25,211 | 61,517 | 17.7 | 57.7 | 598 | 1,818 | 1.02 (0.93 - 1.13) | 0.63 | 1.04 (0.94 - 1.15) | 0.47 |
| MDCR | 16,916 | 31,823 | 14.4 | 28.4 | 373 | 685 | 1.12 (0.98 - 1.28) | 0.10 | 1.12 (0.98 - 1.29) | 0.18 |
| MDCD | 1,074 | 2,211 | 0.6 | 1.3 | 33 | 53 | 1.25 (0.78 - 1.98) | 0.34 | 1.14 (0.72 - 1.82) | 0.60 |
| Open Claims | 325,257 | 670,854 | 249.0 | 678.5 | 3,656 | 8,817 | 1.08 (1.04 - 1.13) | <0.01 | 1.08 (0.95 - 1.23) | 0.25 |
| OptumEHR | 22,478 | 56,159 | 9.6 | 29.6 | 234 | 701 | 1.15 (0.98 - 1.34) | 0.09 | 1.20 (0.95 - 1.52) | 0.19 |
| VA | 14,124 | 102,850 | 12.1 | 104.8 | 161 | 1,575 | 0.99 (0.81 - 1.19) | 0.90 | 1.03 (0.85 - 1.25) | 0.77 |
| GermanyDA | 12,256 | 2,919 | 12.4 | 3.0 | 78 | 18 | 1.15 (0.65 - 2.13) | 0.65 | 1.33 (0.73 - 2.42) | 0.35 |
| SIDIAP | 26,228 | 8,073 | 54.9 | 19.2 | 144 | 18 | 1.57 (0.95 - 2.75) | 0.10 | 1.56 (0.91 - 2.66) | 0.12 |
| Meta-analysis | 431,440 | 836,851 | 359.2 | 818.6 | 5,126 | 12,120 | 1.08 (1.05 - 1.12) | <0.01 | 1.09 (0.98 - 1.21) | 0.10 |
| **PS-matching / on-treatment 2 TAR** | | | | | | | | | | |
| CCAE | 1,835 | 3,224 | 0.5 | 0.8 | 8 | 9 | 1.88 (0.58 - 5.98) | 0.29 | 1.88 (0.58 - 6.05) | 0.30 |
| OptumDOD | 24,356 | 61,434 | 17.0 | 57.6 | 578 | 1,814 | 1.09 (0.96 - 1.24) | 0.17 | 1.13 (0.99 - 1.29) | 0.09 |
| MDCR | 15,772 | 30,879 | 13.4 | 27.5 | 341 | 664 | 1.17 (0.96 - 1.41) | 0.11 | 1.13 (0.93 - 1.37) | 0.24 |
| MDCD | 1,019 | 2,194 | 0.6 | 1.3 | 31 | 53 | 1.18 (0.60 - 2.25) | 0.63 | 1.09 (0.56 - 2.13) | 0.78 |
| Open Claims | 318,926 | 670,851 | 242.6 | 678.5 | 3,541 | 8,817 | 1.08 (1.02 - 1.14) | <0.01 | 1.09 (1.01 - 1.18) | 0.06 |
| OptumEHR | 21,284 | 56,118 | 9.0 | 29.6 | 215 | 698 | 1.19 (0.97 - 1.45) | 0.09 | 1.24 (1.01 - 1.52) | 0.05 |
| VA | 13,173 | 73,224 | 11.5 | 70.3 | 153 | 985 | 1.00 (0.78 - 1.27) | 0.99 | 1.04 (0.81 - 1.33) | 0.78 |
| GermanyDA | 12,221 | 2,496 | 12.4 | 2.5 | 78 | 15 | 1.07 (0.51 - 2.40) | 0.87 | 1.08 (0.50 - 2.34) | 0.81 |
| SIDIAP | 26,107 | 6,126 | 54.8 | 14.0 | 143 | 18 | 1.48 (0.77 - 3.10) | 0.27 | 1.59 (0.74 - 3.42) | 0.26 |
| Meta-analysis | 432,858 | 903,322 | 361.3 | 881.4 | 5,080 | 13,064 | 1.09 (1.04 - 1.14) | <0.01 | 1.10 (1.01 - 1.20) | 0.03 |

##### 7.6.12 Joint pain

**Supplementary Table 317:** Exposures, outcomes and risk estimates for Joint pain in DPP4I (T) vs SU (C) new-user cohorts under two different propensity score (PS) adjustment across data sources. We report total patient exposure time in 1,000 patient-year (KYr) units, uncalibrated and calibrated hazard ratios (HRs) and their 95% confidence intervals (CIs). Greyed out entries did not pass study diagnostics and are included only for completeness.

| | Patients | | Exposure (KYr) | | Outcomes | | Uncalibrated | | Calibrated | |
|---|---|---|---|---|---|---|---|---|---|---|
| | T | C | T | C | T | C | HR (95% CI) | $p$ | HR (95% CI) | $p$ |
| **PS-stratification / on-treatment 2 TAR** | | | | | | | | | | |
| CCAE | 1,946 | 3,238 | 0.5 | 0.9 | <5 | 6 | 0.85 (0.21 - 3.04) | 0.81 | 0.91 (0.24 - 3.43) | 0.88 |
| OptumDOD | 25,533 | 62,681 | 18.1 | 59.1 | 297 | 746 | 1.12 (0.97 - 1.29) | 0.11 | 1.14 (0.99 - 1.32) | 0.09 |
| MDCR | 17,206 | 32,465 | 14.8 | 29.1 | 160 | 264 | 1.15 (0.94 - 1.41) | 0.18 | 1.16 (0.94 - 1.43) | 0.22 |
| MDCD | 1,107 | 2,316 | 0.6 | 1.4 | 8 | 23 | 0.71 (0.29 - 1.59) | 0.43 | 0.64 (0.27 - 1.51) | 0.32 |
| Open Claims | 322,794 | 668,806 | 247.6 | 678.5 | 1,749 | 3,960 | 1.05 (0.99 - 1.11) | 0.10 | 1.05 (0.92 - 1.20) | 0.50 |
| OptumEHR | 22,153 | 55,801 | 9.5 | 29.6 | 99 | 278 | 1.17 (0.91 - 1.50) | 0.21 | 1.23 (0.91 - 1.67) | 0.23 |
| VA | 13,570 | 99,112 | 11.7 | 101.6 | 57 | 587 | 1.12 (0.81 - 1.53) | 0.48 | 1.17 (0.85 - 1.62) | 0.35 |
| GermanyDA | 10,863 | 2,683 | 10.8 | 2.7 | 213 | 61 | 0.91 (0.66 - 1.26) | 0.56 | 1.05 (0.76 - 1.46) | 0.69 |
| SIDIAP | 21,576 | 7,328 | 44.3 | 16.6 | 964 | 424 | 0.86 (0.75 - 0.98) | 0.03 | 0.85 (0.73 - 0.98) | 0.06 |
| Meta-analysis | 421,232 | 832,080 | 345.6 | 817.0 | 3,490 | 5,756 | 1.03 (0.93 - 1.14) | 0.57 | 1.04 (0.90 - 1.19) | 0.62 |
| **PS-matching / on-treatment 2 TAR** | | | | | | | | | | |
| CCAE | 1,779 | 3,167 | 0.5 | 0.8 | <5 | 6 | 0.70 (0.14 - 2.91) | 0.65 | 0.71 (0.16 - 3.19) | 0.66 |
| OptumDOD | 24,737 | 62,613 | 17.4 | 59.1 | 279 | 744 | 1.15 (0.95 - 1.39) | 0.14 | 1.19 (0.98 - 1.44) | 0.08 |
| MDCR | 16,065 | 31,596 | 13.7 | 28.3 | 142 | 262 | 1.01 (0.76 - 1.34) | 0.95 | 0.98 (0.74 - 1.30) | 0.87 |
| MDCD | 1,038 | 2,299 | 0.6 | 1.4 | 8 | 23 | 1.49 (0.40 - 5.39) | 0.55 | 1.39 (0.38 - 5.08) | 0.63 |
| Open Claims | 317,021 | 668,790 | 241.7 | 678.5 | 1,671 | 3,960 | 1.01 (0.93 - 1.09) | 0.85 | 1.02 (0.92 - 1.13) | 0.72 |
| OptumEHR | 21,055 | 55,760 | 8.9 | 29.6 | 92 | 278 | 1.00 (0.72 - 1.36) | 0.98 | 1.03 (0.75 - 1.42) | 0.84 |
| VA | 12,640 | 71,549 | 11.2 | 69.3 | 55 | 392 | 1.14 (0.76 - 1.67) | 0.53 | 1.18 (0.80 - 1.76) | 0.42 |
| GermanyDA | 10,836 | 2,280 | 10.8 | 2.3 | 213 | 50 | 0.96 (0.64 - 1.47) | 0.86 | 0.98 (0.64 - 1.49) | 0.73 |
| SIDIAP | 21,398 | 5,461 | 44.1 | 11.8 | 963 | 326 | 0.80 (0.67 - 0.96) | 0.01 | 0.86 (0.59 - 1.24) | 0.48 |
| Meta-analysis | 424,790 | 900,348 | 348.4 | 880.2 | 3,423 | 6,035 | 1.00 (0.89 - 1.11) | 0.93 | 1.01 (0.88 - 1.15) | 0.91 |

##### 7.6.13 Lower extremity amputation

**Supplementary Table 318:** Exposures, outcomes and risk estimates for Lower extremity amputation in DPP4I (T) vs SU (C) new-user cohorts under two different propensity score (PS) adjustment across data sources. We report total patient exposure time in 1,000 patient-year (KYr) units, uncalibrated and calibrated hazard ratios (HRs) and their 95% confidence intervals (CIs). Greyed out entries did not pass study diagnostics and are included only for completeness.

| | Patients | | Exposure (KYr) | | Outcomes | | Uncalibrated | | Calibrated | |
|---|---|---|---|---|---|---|---|---|---|---|
| | T | C | T | C | T | C | HR (95% CI) | $p$ | HR (95% CI) | $p$ |
| **PS-stratification / on-treatment 2 TAR** | | | | | | | | | | |
| CCAE | 2,085 | 3,378 | 0.5 | 0.9 | <5 | <5 | 1.88 (0.07 - 47.4) | 0.70 | 2.00 (0.08 - 50.5) | 0.68 |
| OptumDOD | 27,175 | 66,157 | 19.4 | 62.8 | 13 | 81 | 0.60 (0.31 - 1.05) | 0.10 | 0.61 (0.33 - 1.12) | 0.11 |
| MDCR | 17,911 | 33,651 | 15.4 | 30.3 | 16 | 30 | 1.16 (0.60 - 2.18) | 0.65 | 1.16 (0.61 - 2.22) | 0.65 |
| MDCD | 1,205 | 2,474 | 0.7 | 1.5 | 0 | <5 | | | | |
| Open Claims | 343,958 | 706,044 | 264.6 | 715.7 | 139 | 457 | 0.93 (0.76 - 1.13) | 0.46 | 0.93 (0.74 - 1.17) | 0.53 |
| OptumEHR | 23,319 | 58,314 | 10.1 | 31.3 | 7 | 24 | 1.01 (0.38 - 2.38) | 0.99 | 1.05 (0.41 - 2.69) | 0.91 |
| VA | 15,277 | 110,322 | 13.1 | 113.4 | <10 | 76 | 0.63 (0.24 - 1.44) | 0.31 | 0.66 (0.27 - 1.60) | 0.36 |
| GermanyDA | 12,634 | 2,999 | 12.9 | 3.1 | 0 | 0 | | | | |
| SIDIAP | 26,434 | 8,082 | 55.4 | 19.3 | 17 | 5 | 2.27 (0.79 - 7.56) | 0.16 | 2.24 (0.73 - 6.94) | 0.16 |
| Meta-analysis | 412,363 | 864,166 | 309.5 | 840.0 | 175 | 592 | 0.91 (0.77 - 1.09) | 0.31 | 0.92 (0.75 - 1.12) | 0.41 |
| **PS-matching / on-treatment 2 TAR** | | | | | | | | | | |
| CCAE | 1,893 | 3,305 | 0.5 | 0.9 | <5 | <5 | | | | |
| OptumDOD | 26,286 | 66,087 | 18.7 | 62.7 | 13 | 81 | 0.58 (0.26 - 1.19) | 0.17 | 0.60 (0.28 - 1.30) | 0.19 |
| MDCR | 16,701 | 32,770 | 14.3 | 29.4 | 16 | 30 | 1.42 (0.59 - 3.35) | 0.43 | 1.38 (0.58 - 3.29) | 0.47 |
| MDCD | 1,135 | 2,455 | 0.7 | 1.5 | 0 | <5 | | | | |
| Open Claims | 337,296 | 706,040 | 257.8 | 715.7 | 137 | 457 | 0.99 (0.76 - 1.28) | 0.94 | 1.00 (0.76 - 1.31) | 0.96 |
| OptumEHR | 22,103 | 58,271 | 9.5 | 31.3 | 7 | 24 | 1.02 (0.34 - 2.77) | 0.97 | 1.06 (0.37 - 3.02) | 0.92 |
| VA | 14,202 | 79,712 | 12.4 | 77.3 | <10 | 56 | 1.02 (0.31 - 2.93) | 0.97 | 1.07 (0.35 - 3.28) | 0.91 |
| GermanyDA | 12,597 | 2,566 | 12.8 | 2.6 | 0 | 0 | | | | |
| SIDIAP | 26,308 | 6,141 | 55.3 | 14.1 | 17 | <5 | 2.47 (0.58 - 16.8) | 0.29 | 2.65 (0.48 - 14.7) | 0.27 |
| Meta-analysis | 416,588 | 942,880 | 312.6 | 916.3 | <183 | 648 | 0.97 (0.77 - 1.22) | 0.80 | 0.98 (0.77 - 1.25) | 0.87 |

##### 7.6.14 Nausea

**Supplementary Table 319:** Exposures, outcomes and risk estimates for Nausea in DPP4I (T) vs SU (C) new-user cohorts under two different propensity score (PS) adjustment across data sources. We report total patient exposure time in 1,000 patient-year (KYr) units, uncalibrated and calibrated hazard ratios (HRs) and their 95% confidence intervals (CIs). Greyed out entries did not pass study diagnostics and are included only for completeness.

| | Patients | | Exposure (KYr) | | Outcomes | | Uncalibrated | | Calibrated | |
|---|---|---|---|---|---|---|---|---|---|---|
| | T | C | T | C | T | C | HR (95% CI) | $p$ | HR (95% CI) | $p$ |
| **PS-stratification / on-treatment 2 TAR** | | | | | | | | | | |
| CCAE | 1,852 | 3,051 | 0.5 | 0.8 | 16 | 24 | 1.05 (0.54 - 1.99) | 0.89 | 1.11 (0.57 - 2.16) | 0.75 |
| OptumDOD | 23,106 | 56,570 | 16.2 | 52.7 | 734 | 2,350 | 0.93 (0.86 - 1.02) | 0.13 | 0.95 (0.87 - 1.04) | 0.36 |
| MDCR | 15,971 | 29,913 | 13.6 | 26.7 | 464 | 961 | 0.91 (0.81 - 1.03) | 0.13 | 0.92 (0.81 - 1.04) | 0.28 |
| MDCD | 911 | 1,956 | 0.5 | 1.1 | 38 | 70 | 1.10 (0.72 - 1.65) | 0.66 | 1.00 (0.66 - 1.51) | 0.83 |
| Open Claims | 297,457 | 614,955 | 228.2 | 624.6 | 4,950 | 12,946 | 0.96 (0.93 - 1.00) | 0.03 | 0.96 (0.85 - 1.09) | 0.57 |
| OptumEHR | 21,214 | 53,261 | 9.0 | 27.8 | 293 | 931 | 1.00 (0.87 - 1.14) | 0.96 | 1.04 (0.83 - 1.31) | 0.75 |
| VA | 14,379 | 104,723 | 12.4 | 107.5 | 95 | 822 | 1.23 (0.95 - 1.58) | 0.11 | 1.29 (0.99 - 1.66) | 0.06 |
| GermanyDA | 11,730 | 2,799 | 11.9 | 2.9 | 131 | 34 | 1.03 (0.67 - 1.61) | 0.90 | 1.19 (0.77 - 1.85) | 0.44 |
| SIDIAP | 25,547 | 7,960 | 53.4 | 18.9 | 409 | 103 | 1.03 (0.80 - 1.34) | 0.83 | 1.02 (0.78 - 1.33) | 0.86 |
| Meta-analysis | 397,788 | 770,465 | 333.3 | 755.5 | 7,035 | 17,419 | 0.96 (0.93 - 0.99) | <0.01 | 0.97 (0.87 - 1.07) | 0.50 |
| **PS-matching / on-treatment 2 TAR** | | | | | | | | | | |
| CCAE | 1,680 | 2,983 | 0.4 | 0.8 | 15 | 24 | 0.98 (0.45 - 2.04) | 0.95 | 0.98 (0.46 - 2.09) | 0.82 |
| OptumDOD | 22,321 | 56,486 | 15.6 | 52.7 | 702 | 2,344 | 0.93 (0.83 - 1.05) | 0.25 | 0.97 (0.86 - 1.09) | 0.60 |
| MDCR | 14,919 | 28,985 | 12.7 | 25.8 | 421 | 944 | 0.93 (0.79 - 1.10) | 0.41 | 0.91 (0.77 - 1.07) | 0.28 |
| MDCD | 881 | 1,941 | 0.5 | 1.1 | 35 | 68 | 1.31 (0.71 - 2.37) | 0.38 | 1.22 (0.66 - 2.23) | 0.55 |
| Open Claims | 291,664 | 614,935 | 222.4 | 624.6 | 4,784 | 12,946 | 0.96 (0.92 - 1.01) | 0.11 | 0.97 (0.91 - 1.05) | 0.56 |
| OptumEHR | 20,090 | 53,221 | 8.4 | 27.8 | 279 | 930 | 1.04 (0.87 - 1.24) | 0.63 | 1.08 (0.91 - 1.29) | 0.42 |
| VA | 13,385 | 75,977 | 11.8 | 73.8 | 90 | 552 | 1.18 (0.85 - 1.61) | 0.30 | 1.23 (0.90 - 1.69) | 0.22 |
| GermanyDA | 11,697 | 2,384 | 11.8 | 2.4 | 130 | 28 | 1.21 (0.71 - 2.17) | 0.50 | 1.23 (0.70 - 2.16) | 0.50 |
| SIDIAP | 25,436 | 6,027 | 53.4 | 13.7 | 408 | 80 | 1.00 (0.73 - 1.40) | 1.00 | 1.07 (0.68 - 1.69) | 0.79 |
| Meta-analysis | 402,073 | 842,939 | 337.0 | 822.6 | 6,864 | 17,916 | 0.97 (0.93 - 1.01) | 0.10 | 0.98 (0.90 - 1.07) | 0.61 |

##### 7.6.15 Peripheral edema

**Supplementary Table 320:** Exposures, outcomes and risk estimates for Peripheral edema in DPP4I (T) vs SU (C) new-user cohorts under two different propensity score (PS) adjustment across data sources. We report total patient exposure time in 1,000 patient-year (KYr) units, uncalibrated and calibrated hazard ratios (HRs) and their 95% confidence intervals (CIs). Greyed out entries did not pass study diagnostics and are included only for completeness.

| | Patients | | Exposure (KYr) | | Outcomes | | Uncalibrated | | Calibrated | |
|---|---|---|---|---|---|---|---|---|---|---|
| | T | C | T | C | T | C | HR (95% CI) | $p$ | HR (95% CI) | $p$ |
| **PS-stratification / on-treatment 2 TAR** | | | | | | | | | | |
| CCAE | 1,647 | 2,733 | 0.4 | 0.7 | 22 | 55 | 0.66 (0.38 - 1.08) | 0.11 | 0.70 (0.41 - 1.18) | 0.24 |
| OptumDOD | 19,396 | 48,325 | 13.0 | 42.8 | 1,352 | 4,419 | 0.94 (0.88 - 1.00) | 0.07 | 0.96 (0.89 - 1.03) | 0.34 |
| MDCR | 13,304 | 25,259 | 11.0 | 21.5 | 932 | 2,031 | 0.91 (0.84 - 0.99) | 0.03 | 0.91 (0.83 - 1.00) | 0.18 |
| MDCD | 717 | 1,662 | 0.4 | 0.9 | 48 | 125 | 0.90 (0.63 - 1.27) | 0.56 | 0.82 (0.58 - 1.16) | 0.29 |
| Open Claims | 250,191 | 526,163 | 187.8 | 521.8 | 8,817 | 24,643 | 0.93 (0.91 - 0.96) | <0.01 | 0.93 (0.82 - 1.06) | 0.28 |
| OptumEHR | 18,904 | 47,411 | 7.8 | 23.5 | 455 | 1,880 | 0.79 (0.71 - 0.88) | <0.01 | 0.83 (0.67 - 1.02) | 0.13 |
| VA | 11,571 | 87,019 | 9.9 | 86.4 | 350 | 3,651 | 0.82 (0.72 - 0.93) | <0.01 | 0.85 (0.75 - 0.97) | 0.03 |
| GermanyDA | 10,333 | 2,524 | 10.3 | 2.5 | 378 | 108 | 0.75 (0.59 - 0.96) | 0.02 | 0.87 (0.68 - 1.12) | 0.33 |
| SIDIAP | 23,708 | 7,689 | 49.0 | 17.9 | 1,041 | 319 | 0.85 (0.74 - 0.99) | 0.04 | 0.85 (0.72 - 0.99) | 0.08 |
| Meta-analysis | 338,200 | 661,766 | 279.7 | 631.7 | 13,045 | 33,580 | 0.89 (0.83 - 0.94) | <0.01 | 0.89 (0.79 - 1.00) | 0.05 |
| **PS-matching / on-treatment 2 TAR** | | | | | | | | | | |
| CCAE | 1,510 | 2,673 | 0.4 | 0.7 | 19 | 54 | 0.55 (0.26 - 1.06) | 0.09 | 0.55 (0.27 - 1.11) | 0.12 |
| OptumDOD | 18,789 | 48,260 | 12.5 | 42.8 | 1,295 | 4,412 | 0.95 (0.87 - 1.03) | 0.19 | 0.98 (0.90 - 1.07) | 0.66 |
| MDCR | 12,380 | 24,417 | 10.2 | 20.7 | 853 | 1,980 | 0.88 (0.78 - 0.98) | 0.02 | 0.85 (0.76 - 0.96) | 0.02 |
| MDCD | 692 | 1,646 | 0.4 | 0.9 | 46 | 124 | 0.91 (0.56 - 1.47) | 0.71 | 0.85 (0.52 - 1.39) | 0.54 |
| Open Claims | 245,855 | 526,162 | 183.6 | 521.8 | 8,534 | 24,643 | 0.92 (0.89 - 0.95) | <0.01 | 0.93 (0.87 - 0.99) | 0.07 |
| OptumEHR | 17,871 | 47,359 | 7.2 | 23.5 | 427 | 1,879 | 0.82 (0.71 - 0.94) | <0.01 | 0.85 (0.74 - 0.98) | 0.05 |
| VA | 10,786 | 61,870 | 9.4 | 58.0 | 328 | 2,502 | 0.80 (0.68 - 0.94) | <0.01 | 0.83 (0.70 - 0.98) | 0.04 |
| GermanyDA | 10,309 | 2,163 | 10.3 | 2.1 | 377 | 99 | 0.76 (0.57 - 1.02) | 0.06 | 0.77 (0.57 - 1.04) | 0.14 |
| SIDIAP | 23,568 | 5,788 | 48.9 | 12.9 | 1,034 | 256 | 0.88 (0.73 - 1.07) | 0.20 | 0.94 (0.65 - 1.37) | 0.77 |
| Meta-analysis | 341,760 | 720,338 | 282.9 | 683.4 | 12,913 | 35,949 | 0.89 (0.85 - 0.93) | <0.01 | 0.90 (0.82 - 0.98) | 0.02 |

##### 7.6.16 Photosensitivity

**Supplementary Table 321:** Exposures, outcomes and risk estimates for Photosensitivity in DPP4I (T) vs SU (C) new-user cohorts under two different propensity score (PS) adjustment across data sources. We report total patient exposure time in 1,000 patient-year (KYr) units, uncalibrated and calibrated hazard ratios (HRs) and their 95% confidence intervals (CIs). Greyed out entries did not pass study diagnostics and are included only for completeness.

| | Patients | | Exposure (KYr) | | Outcomes | | Uncalibrated | | Calibrated | |
|---|---|---|---|---|---|---|---|---|---|---|
| | T | C | T | C | T | C | HR (95% CI) | $p$ | HR (95% CI) | $p$ |
| **PS-stratification / on-treatment 2 TAR** | | | | | | | | | | |
| CCAE | 2,057 | 3,331 | 0.5 | 0.9 | <5 | <5 | 1.02 (0.13 - 6.60) | 0.99 | 1.08 (0.15 - 7.76) | 0.92 |
| OptumDOD | 26,869 | 65,494 | 19.1 | 61.9 | 34 | 115 | 1.13 (0.74 - 1.67) | 0.56 | 1.15 (0.77 - 1.73) | 0.50 |
| MDCR | 17,593 | 33,139 | 15.1 | 29.7 | 62 | 97 | 1.17 (0.83 - 1.63) | 0.37 | 1.17 (0.83 - 1.64) | 0.39 |
| MDCD | 1,205 | 2,471 | 0.7 | 1.5 | 0 | 0 | | | | |
| Open Claims | 339,369 | 698,040 | 260.8 | 706.5 | 172 | 398 | 1.11 (0.92 - 1.33) | 0.28 | 1.11 (0.89 - 1.38) | 0.38 |
| OptumEHR | 23,122 | 57,750 | 10.0 | 30.9 | 20 | 75 | 0.98 (0.56 - 1.63) | 0.93 | 1.02 (0.59 - 1.79) | 0.93 |
| VA | 15,025 | 108,925 | 12.9 | 111.8 | <10 | 192 | 0.70 (0.21 - 1.79) | 0.52 | 0.73 (0.25 - 2.16) | 0.57 |
| GermanyDA | 12,586 | 2,992 | 12.8 | 3.1 | <5 | <5 | 0.91 (0.10 - 8.25) | 0.93 | 1.06 (0.12 - 9.57) | 0.95 |
| SIDIAP | 26,410 | 8,090 | 55.4 | 19.3 | 6 | <5 | 0.60 (0.14 - 3.35) | 0.53 | 0.59 (0.12 - 2.91) | 0.52 |
| Meta-analysis | 406,953 | 854,423 | 304.9 | 829.1 | 288 | 685 | 1.11 (0.96 - 1.28) | 0.16 | 1.12 (0.94 - 1.33) | 0.21 |
| **PS-matching / on-treatment 2 TAR** | | | | | | | | | | |
| CCAE | 1,867 | 3,259 | 0.5 | 0.9 | <5 | <5 | 1.00 (0.04 - 25.3) | 1.00 | 1.00 (0.04 - 25.3) | 0.95 |
| OptumDOD | 25,998 | 65,423 | 18.4 | 61.9 | 31 | 115 | 1.17 (0.68 - 1.97) | 0.55 | 1.21 (0.71 - 2.06) | 0.48 |
| MDCR | 16,401 | 32,260 | 14.0 | 28.9 | 57 | 94 | 1.33 (0.86 - 2.06) | 0.20 | 1.29 (0.83 - 2.00) | 0.26 |
| MDCD | 1,138 | 2,452 | 0.7 | 1.5 | 0 | 0 | | | | |
| Open Claims | 332,799 | 698,036 | 254.1 | 706.5 | 168 | 398 | 1.11 (0.87 - 1.41) | 0.40 | 1.12 (0.88 - 1.44) | 0.37 |
| OptumEHR | 21,924 | 57,708 | 9.4 | 30.9 | 20 | 75 | 0.79 (0.38 - 1.52) | 0.49 | 0.82 (0.41 - 1.63) | 0.57 |
| VA | 13,973 | 78,397 | 12.3 | 75.8 | <10 | 121 | 0.94 (0.27 - 2.52) | 0.92 | 0.98 (0.32 - 3.02) | 0.97 |
| GermanyDA | 12,549 | 2,560 | 12.8 | 2.6 | <5 | <5 | | | | |
| SIDIAP | 26,284 | 6,139 | 55.3 | 14.0 | 6 | <5 | 0.59 (0.10 - 3.37) | 0.55 | 0.64 (0.11 - 3.73) | 0.62 |
| Meta-analysis | 411,095 | 931,824 | 308.1 | 903.9 | <286 | 803 | 1.12 (0.93 - 1.35) | 0.23 | 1.13 (0.93 - 1.38) | 0.23 |

##### 7.6.17 Venous thromboembolism

**Supplementary Table 322:** Exposures, outcomes and risk estimates for Venous thromboembolism in DPP4I (T) vs SU (C) new-user cohorts under two different propensity score (PS) adjustment across data sources. We report total patient exposure time in 1,000 patient-year (KYr) units, uncalibrated and calibrated hazard ratios (HRs) and their 95% confidence intervals (CIs). Greyed out entries did not pass study diagnostics and are included only for completeness.

| | Patients | | Exposure (KYr) | | Outcomes | | Uncalibrated | | Calibrated | |
|---|---|---|---|---|---|---|---|---|---|---|
| | T | C | T | C | T | C | HR (95% CI) | $p$ | HR (95% CI) | $p$ |
| **PS-stratification / on-treatment 2 TAR** | | | | | | | | | | |
| CCAE | 2,016 | 3,284 | 0.5 | 0.9 | 7 | 7 | 1.82 (0.61 - 5.44) | 0.29 | 1.93 (0.64 - 5.81) | 0.25 |
| OptumDOD | 26,091 | 63,541 | 18.5 | 59.8 | 277 | 903 | 1.00 (0.86 - 1.15) | 0.96 | 1.01 (0.88 - 1.17) | 0.85 |
| MDCR | 17,192 | 32,221 | 14.8 | 28.8 | 190 | 470 | 0.80 (0.67 - 0.95) | 0.01 | 0.80 (0.67 - 0.96) | 0.03 |
| MDCD | 1,140 | 2,364 | 0.7 | 1.4 | 17 | 22 | 2.11 (1.05 - 4.13) | 0.03 | 1.91 (0.97 - 3.79) | 0.07 |
| Open Claims | 329,000 | 676,714 | 252.8 | 684.4 | 1,848 | 5,629 | 0.86 (0.82 - 0.91) | <0.01 | 0.86 (0.75 - 0.98) | 0.03 |
| OptumEHR | 22,532 | 56,350 | 9.7 | 29.9 | 89 | 375 | 0.80 (0.62 - 1.01) | 0.07 | 0.84 (0.62 - 1.13) | 0.29 |
| VA | 14,702 | 107,070 | 12.6 | 109.7 | 62 | 598 | 0.92 (0.67 - 1.24) | 0.58 | 0.96 (0.70 - 1.31) | 0.78 |
| GermanyDA | 11,576 | 2,782 | 11.7 | 2.8 | 123 | 36 | 0.80 (0.53 - 1.23) | 0.30 | 0.93 (0.61 - 1.41) | 0.71 |
| SIDIAP | 25,562 | 7,942 | 53.6 | 18.9 | 259 | 78 | 0.94 (0.70 - 1.28) | 0.71 | 0.94 (0.69 - 1.27) | 0.68 |
| Meta-analysis | 433,093 | 841,914 | 361.7 | 826.1 | 2,803 | 7,513 | 0.88 (0.82 - 0.96) | <0.01 | 0.89 (0.79 - 1.01) | 0.07 |
| **PS-matching / on-treatment 2 TAR** | | | | | | | | | | |
| CCAE | 1,830 | 3,214 | 0.5 | 0.8 | 7 | 7 | 2.79 (0.65 - 14.1) | 0.19 | 2.79 (0.60 - 13.0) | 0.20 |
| OptumDOD | 25,247 | 63,481 | 17.8 | 59.8 | 269 | 901 | 0.96 (0.80 - 1.16) | 0.69 | 1.00 (0.83 - 1.20) | 0.90 |
| MDCR | 16,035 | 31,196 | 13.7 | 27.9 | 179 | 459 | 0.83 (0.64 - 1.05) | 0.13 | 0.80 (0.62 - 1.03) | 0.09 |
| MDCD | 1,078 | 2,346 | 0.6 | 1.4 | 17 | 22 | 4.10 (1.51 - 11.9) | <0.01 | 3.82 (1.36 - 10.7) | 0.01 |
| Open Claims | 322,630 | 676,711 | 246.2 | 684.4 | 1,795 | 5,629 | 0.87 (0.81 - 0.94) | <0.01 | 0.88 (0.81 - 0.97) | 0.02 |
| OptumEHR | 21,336 | 56,310 | 9.0 | 29.9 | 88 | 374 | 0.87 (0.65 - 1.16) | 0.35 | 0.91 (0.68 - 1.21) | 0.52 |
| VA | 13,685 | 76,974 | 12.0 | 74.3 | 59 | 405 | 1.30 (0.90 - 1.87) | 0.16 | 1.36 (0.94 - 1.97) | 0.11 |
| GermanyDA | 11,547 | 2,382 | 11.7 | 2.4 | 122 | 32 | 0.68 (0.41 - 1.17) | 0.15 | 0.69 (0.41 - 1.17) | 0.20 |
| SIDIAP | 25,424 | 6,006 | 53.5 | 13.7 | 258 | 60 | 1.07 (0.71 - 1.65) | 0.75 | 1.15 (0.68 - 1.95) | 0.64 |
| Meta-analysis | 436,982 | 915,406 | 364.6 | 893.8 | 2,787 | 7,882 | 0.91 (0.84 - 0.99) | 0.03 | 0.92 (0.82 - 1.03) | 0.16 |

###### 7.6.18 Vomiting

**Supplementary Table 323:** Exposures, outcomes and risk estimates for Vomiting in DPP4I (T) vs SU (C) new-user cohorts under two different propensity score (PS) adjustment across data sources. We report total patient exposure time in 1,000 patient-year (KYr) units, uncalibrated and calibrated hazard ratios (HRs) and their 95% confidence intervals (CIs). Greyed out entries did not pass study diagnostics and are included only for completeness.

| | Patients | | Exposure (KYr) | | Outcomes | | Uncalibrated | | Calibrated | |
|---|---|---|---|---|---|---|---|---|---|---|
| | T | C | T | C | T | C | HR (95% CI) | $p$ | HR (95% CI) | $p$ |
| **PS-stratification / on-treatment 2 TAR** | | | | | | | | | | |
| CCAE | 1,913 | 3,146 | 0.5 | 0.8 | 12 | 18 | 1.07 (0.49 - 2.23) | 0.86 | 1.14 (0.53 - 2.43) | 0.75 |
| OptumDOD | 24,209 | 59,039 | 17.1 | 55.4 | 567 | 1,837 | 0.94 (0.85 - 1.04) | 0.21 | 0.96 (0.86 - 1.06) | 0.44 |
| MDCR | 16,422 | 30,773 | 14.0 | 27.5 | 366 | 801 | 0.91 (0.80 - 1.03) | 0.14 | 0.91 (0.79 - 1.04) | 0.27 |
| MDCD | 965 | 2,025 | 0.6 | 1.2 | 32 | 50 | 1.35 (0.83 - 2.14) | 0.22 | 1.22 (0.76 - 1.96) | 0.42 |
| Open Claims | 308,206 | 635,811 | 236.6 | 646.1 | 3,986 | 10,267 | 0.97 (0.94 - 1.01) | 0.17 | 0.97 (0.86 - 1.11) | 0.68 |
| OptumEHR | 22,044 | 55,074 | 9.4 | 29.1 | 206 | 642 | 1.03 (0.87 - 1.22) | 0.70 | 1.08 (0.85 - 1.38) | 0.58 |
| VA | 14,661 | 106,319 | 12.6 | 109.3 | 82 | 629 | 1.45 (1.09 - 1.90) | <0.01 | 1.51 (1.14 - 2.00) | <0.01 |
| GermanyDA | 11,722 | 2,797 | 11.9 | 2.9 | 131 | 35 | 1.01 (0.66 - 1.57) | 0.96 | 1.17 (0.76 - 1.81) | 0.49 |
| SIDIAP | 25,484 | 7,948 | 53.3 | 18.8 | 424 | 102 | 1.09 (0.84 - 1.42) | 0.52 | 1.08 (0.83 - 1.40) | 0.62 |
| Meta-analysis | 410,965 | 796,613 | 343.4 | 781.7 | 5,724 | 13,752 | 0.97 (0.94 - 1.00) | 0.08 | 0.98 (0.88 - 1.08) | 0.65 |
| **PS-matching / on-treatment 2 TAR** | | | | | | | | | | |
| CCAE | 1,726 | 3,076 | 0.5 | 0.8 | 12 | 18 | 0.78 (0.29 - 1.91) | 0.60 | 0.78 (0.30 - 2.00) | 0.62 |
| OptumDOD | 23,400 | 58,951 | 16.4 | 55.4 | 541 | 1,835 | 0.90 (0.79 - 1.02) | 0.11 | 0.93 (0.81 - 1.06) | 0.32 |
| MDCR | 15,323 | 29,831 | 13.0 | 26.6 | 334 | 786 | 0.91 (0.76 - 1.09) | 0.32 | 0.88 (0.73 - 1.06) | 0.22 |
| MDCD | 928 | 2,009 | 0.5 | 1.2 | 30 | 49 | 1.48 (0.80 - 2.74) | 0.21 | 1.38 (0.74 - 2.57) | 0.33 |
| Open Claims | 302,249 | 635,789 | 230.6 | 646.1 | 3,853 | 10,266 | 0.99 (0.94 - 1.04) | 0.67 | 1.00 (0.93 - 1.08) | 0.89 |
| OptumEHR | 20,877 | 55,034 | 8.8 | 29.0 | 197 | 642 | 1.03 (0.84 - 1.28) | 0.76 | 1.07 (0.87 - 1.33) | 0.55 |
| VA | 13,637 | 77,013 | 12.0 | 74.8 | 78 | 427 | 1.57 (1.10 - 2.22) | 0.01 | 1.63 (1.15 - 2.33) | <0.01 |
| GermanyDA | 11,689 | 2,384 | 11.8 | 2.4 | 130 | 29 | 1.17 (0.68 - 2.11) | 0.58 | 1.19 (0.68 - 2.10) | 0.57 |
| SIDIAP | 25,374 | 6,017 | 53.2 | 13.7 | 423 | 79 | 1.06 (0.77 - 1.48) | 0.72 | 1.14 (0.72 - 1.80) | 0.63 |
| Meta-analysis | 415,203 | 870,104 | 346.9 | 849.9 | 5,598 | 14,131 | 0.99 (0.94 - 1.03) | 0.54 | 1.00 (0.91 - 1.09) | 0.88 |

### 8 Bibliography

[1] Voss EA, Boyce RD, Ryan PB, van der Lei J, Rijnbeek PR, Schuemie MJ. Accuracy of an automated knowledge base for identifying drug adverse reactions. J Biomed Inform. 2017;66:72-81.

[2] Khera R, Aminorroaya A, Dhingra LS, Thangaraj PM, Camargos AP, Bu F, et al. Comparative Effectiveness of Second-Line Antihyperglycemic Agents for Cardiovascular Outcomes. JACC. 2024;84(10):904-17. Available from: https://www.jacc.org/doi/abs/10.1016/j.jacc.2024.05.069.

[3] Winnenburg R, Sorbello A, Ripple A, Harpaz R, Tonning J, Szarfman A, et al. Leveraging MEDLINE indexing for pharmacovigilance–Inherent limitations and mitigation strategies. Journal of Biomedical Informatics. 2015;57:425-35.

[4] Duke J, Friedlin J, Li X. Consistency in the safety labeling of bioequivalent medications. Pharmacoepidemiology and Drug Safety. 2013;22(3):294-301.

[5] Evans S, Waller PC, Davis S. Use of proportional reporting ratios (PRRs) for signal generation from spontaneous adverse drug reaction reports. Pharmacoepidemiology and Drug Safety. 2001;10(6):483-6.

[6] Banda JM, Evans L, Vanguri RS, Tatonetti NP, Ryan PB, Shah NH. A curated and standardized adverse drug event resource to accelerate drug safety research. Scientific data. 2016;3:160026.
